# Proteogenomic signature of risk of Alzheimer’s disease and related dementia risk in individuals with a history of major depression disorder

**DOI:** 10.1101/2024.09.11.24313493

**Authors:** Breno Satler Diniz, Zhiduo Chen, David C. Steffens, Luke Pilling, Richard H. Fortinsky, George A. Kuchel, Chia-Ling Kuo

## Abstract

The mechanisms linking a history of major depressive disorder (MDD) to an increased risk of Alzheimer’s disease and related dementia (ADRD) are not fully understood. Using the UK Biobank available proteomic and genomic data, we evaluated the biological mechanisms linking both conditions. In participants with a history of MDD at baseline (n=3,615), we found that plasma levels of NfL, GFAP, PSG1 were associated with higher risk (HR=1.38; 1.37; 1.34, respectively; all adjusted p-values<0.05), while VGF, GET3, and HPGDS were associated with lower risk of incident ADRD (n=150) (HR=0.73; 0.71; 0.66, respectively; all adjusted p-values<0.05) during a mean follow-up of 13.7 years (SD=2.2). Two-sample Mendelian randomization analysis using cis-pQTLs genetic instruments revealed that a lower protein expression of apolipoprotein E and higher IL-10 receptor subunit B were causally linked to incident ADRD. Finally, we developed a Proteomic Risk Score (PrRS_MDD-ADRD_), which showed strong discriminative power (C-statistic = 0.84) to identify participants with MDD that developed ADRD upon follow-up. In addition to demonstrating an association between plasma proteins associated with inflammation and future ADRD risk in individuals with MDD, our findings include an element of causality using Mendelian Randomization (MR) and PrRS_MDD-ADRD_ can be useful to identify individuals with the highest risk to develop ADRD in a highly vulnerable population.

## Introduction

Major depressive disorder (MDD) is a highly heterogeneous, multifactorial condition with multiple concurrent risk factors, as well as pathophysiological processes playing a significant role in its phenotypic manifestation and long-term outcomes^1, 2^. The elevated burden of disease associated with MDD^3^ is due not only to the severity of psychopathology but also to its association with adverse health outcomes and multiple other diseases of aging^4^. Epidemiological studies have consistently shown that a history of MDD across the lifespan significantly increases the risk of incident Alzheimer’s disease and related dementia (ADRD) in older adults^5, 6^. The association between MDD and the risk of ADRD cannot be underestimated. For example, there are over 7 million people living with ADRD in the US, with 11.1% to 14.7% of these cases attributable to major depression^7^. Therefore, preventing MDD in the general population, or reducing the risk of development of ADRD among those with MDD can significantly lower the incidence of ADRD in older ages^7–9^.

Despite well-known associations, underlying mechanisms linking MDD to a higher risk of developing ADRD are unclear. For example, previous studies did not suggest that a major depressive episode increases brain amyloid burden^10–12^, the primary pathological mechanism of Alzheimer’s disease, despite some conflicting results^13^. On the other hand, the presence of mild cognitive impairment (MCI) during an MDD episode increases the risk of incident ADRD^14^ and is associated with greater cortical and hippocampal atrophy and dysregulation in multiple biological pathways implicated in aging^15, 16^ that are relevant to the ADRD physiopathology, including increased pro-inflammatory burden, loss of proteostasis control, cellular senescence, and metabolic control^17–19^. Also, a recent study demonstrated a significant genetic correlation between MDD and AD and a potential causal link between MDD and AD using a generalized data-summary Mendelian randomization approach^20^.

Despite the relevance of these past studies, they did not provide definitive evidence of which biological mechanisms may underlie the elevated risk of ADRD among individuals with MDD. Moreover, the results were based on relatively small sample sizes. Large-scale population studies, including identifying MDD cases at baseline and incident ADRD cases, along with multi-omics measurements (e.g., genomic and proteomic data), are necessary to identify the biological processes linking MDD to a higher risk of ADRD. For example, the UK Biobank (UKB, https://www.ukbiobank.ac.uk/) is one of the largest biomedical databases containing data from up to half a million UK participants. Through linkage with electronic health records from primary care and hospital settings, the UKB includes data on the history of MDD and a variety of long-term outcomes, including the diagnosis of ADRD. In addition, the UKB offers in-depth biological information, including genomic and proteomic data, which enables the discovery of over 14,000 protein quantitative loci (pQTL). Identifying pQTLs and disease associations can provide more robust causal inference information via approaches such as Mendelian Randomization supporting drug development or repurposing strategies^21, 22^. Our primary goal was to investigate the proteogenomic signatures of MDD that are distinctively associated with the risk of ADRD by integrating proteomic and genomic available from the UKB.

## Methods

### UK Biobank Pharma Proteomics Project (UKB PPP) cohort

A total of 53,018 active participants were included in the UKB Pharma Proteomics Project (UKB PPP) cohort^22^. The plasma proteomic analysis was done using the Olink® Explore 3072 assay, covering 2,923 proteins. Normalized protein expression (NPX) levels were calculated to account for technical variations^22^. After removing three proteins with high missing rates (GLIPR1 99.7%, NPM1 74.0%, and PCOLCE 63.6%), the median missing rate per protein was 14.7% (interquartile range 3.0% to 17.4%), and the median missing rate per individual was 0.5% (interquartile range 0.1% to 7.5%). Missing proteomic data were imputed using the *k*-nearest neighbor approach (*k*=10), with higher weights to neighbors sharing higher similarities across proteins^23^.

### Baseline cohort

The “*baseline cohort*” included 42,807 UKB PPP participants after excluding participants at baseline with (1) pre-existing psychiatric disorders (schizophrenia, unspecified nonorganic psychosis, manic episode, and bipolar affective disorder); (2) pre-existing ADRD or dementia in other diseases, (3) diagnosis of MDD before the age of 18 years; (4) any missing baseline covariate data (age at recruitment, self-reported sex, ethnicity, and education, body mass index [BMI], hypertension diagnosis (yes/no), diabetes diagnosis (yes/no), *APOE* e4 carrier status (yes/no), antidepressant use (yes/no), and selection by the UKB PPP consortium (yes/no) (**Figure S1**). A total of 3,615 individuals had a history of MDD at baseline (n=3,615) (**Figure S1**). Cases of MDD and incident ADRD were identified using the first occurrence data derived by the UKB, which integrated multi-source data based on ICD-10 codes (primary care, hospital admissions, death registry, and baseline self-reported medical condition data) (**Table S1**). Data extraction was conducted using the field IDs presented in **Table S1**. The anatomical therapeutic chemical codes of antidepressants used to confirm antidepressant use at baseline are provided in **Table S2**.

### Association between protein expression and incident ADRD in participants with a history of MDD at baseline

We applied the inverse normal transformation to individual proteins (n=2,920) in the baseline cohort to correct distributional skewness and unify the scales into z-scores^24^. The association of each protein with incident ADRD was modeled using a Cox regression model, adjusting for baseline covariates (age at recruitment, self-reported sex, ethnicity, education, BMI, hypertension diagnosis (yes/no), diabetes diagnosis (yes/no), *APOE* e4 carrier status (yes/no), antidepressant use (yes/no), and selection by the UKB PPP consortium (yes/no). A sensitivity analysis was performed by excluding *APOE* e4 carrier status from the covariates. P-values were corrected for multiple testing using the Benjamini-Hochberg false discovery rate (FDR) approach^25^, and adjusted p-values smaller than 5% were considered statistically significant. Proteins significant at the FDR-adjusted level of 5% were jointly modeled in a Cox regression model adjusting for covariates to evaluate their dependency.

### Two-sample Mendelian randomization analysis

Observational study data provide limited evidence about causal relationships between an exposure (e.g., protein expression) and an outcome (e.g., incidence of ADRD) due to the lack of experimental control, unmeasured confounding, and risk of reverse causality that are intrinsic to observational study designs^26, 27^. Mendelian randomization (MR) analyses can help overcome these limitations by using genetic variants. Genotypes are fixed at conception as instrumental variables to examine exposure-outcome relationships^28^. The focus on protein quantitative trait loci (pQTL) helps to understand how common and rare genetic variation influences protein levels^29^ and identify proteins to target for drug development.

Two-sample Mendelian Randomization (MR) methods were applied to assess the causal effect of each protein on incident ADRD in UKB European-descent participants with a history of MDD at baseline. Autosomal cis-protein quantitative trait loci (cis-pQTL) were used as genetic instruments for protein expression levels. The selection of cis-pQTL was based on the genome-wide association study (GWAS) summary statistics using UKB European-descent participants from the Sun et al. discovery cohort^22^. They were further validated in the subset with a history of MDD at baseline (“*MR: baseline MDD cohort*” [n=3,896], see **Figure S2**). To avoid bias from sample overlap, we estimated the associations between cis-pQTL and incident ADRD using UKB European-descent participants with a history of MDD at baseline who were not in the UKB PPP (referred to as the “*MR: incident ADRD in MDD cohort*” [n=30,903], **Figure S2**).

#### Selection of genetic instruments

Genetic variants that showed significant associations with each protein, with a p-value< 5×10^−8^ and were located within 1 Mb of the coding gene region (i.e., cis-gene region)^30^ were extracted from the Sun et al^22^. discovery GWAS summary statistics. These variants were identified through individual associations with each protein following the inverse normal transformation, using a two-step procedure involving standard linear regression models^22^.

Those with a minor allele frequency smaller than 0.01 or an INFO score <0.7 (low imputation accuracy) were excluded. For each protein, linkage disequilibrium (LD) clumping was performed on the remaining variants to select independent cis-pQTL. The selection started from the most significant variant with the smallest p-value and then the next after excluding those in LD (r^2^>0.01) or within 500 kb of that variant (clumping window 500 kb). The LD between genetic variants was assessed using genome-wide genotype data from 5,000 randomly selected unrelated UKB participants of European descent. This procedure was repeated until no further cis-pQTL was identified.

We excluded cis-pQTL that showed a discrepancy in β greater than 0.1 for associations with the coded protein (i.e., >0.1 SD change in the transformed NPX per effect allele increase) between the Sun et al. discovery cohort^22^ and the MR baseline MDD cohort. Similar covariate adjustments were made in the MR baseline MDD cohort, including age at baseline assessment (age in short), age^2^, sex, age × sex, age^2^ × sex, genotyping array, top 10 genetic principal components in the UKB, and the consortium selection.

We conducted a sensitivity analysis using cis-pQTL as selected by Sun et al.^22^, who used similar criteria but a more stringent GWAS significance level (p<1.7×10^−11^) and the clumping windows of 10,000 kb followed by 500 kb to account for potential long-range LD, while also merging overlapped clumps.

### Associations between genetic instruments for protein expression levels and incident ADRD in MDD

Cox regression models were used adjusting for age at baseline assessment (age in short), age^2^, sex, age × sex, age^2^ × sex, genotyping array, and top 10 genetic principal components in the UKB. Both associations of cis-pQTL with a protein (from Sun et al.^22^) and incident ADRD were expressed as the effects per copy increase in the level-increasing allele. In the MR analysis, we reported the hazard ratio (HR) for incident ADRD per SD increase in the genetically determined NPX.

### Two-sample Mendelian randomization methods

The primary two-sample MR analysis was conducted using the inverse-variance weighted (IVW) method^31^. A fixed-effect model was used when there were three variants or fewer, and a random-effects model otherwise. Additionally, we applied the MR-Egger regression and MR-Robust Adjusted Profile Score (MR-RAPS) methods^32, 33^. Comparing the results across different methods allows us to evaluate the robustness of our findings. Based on the IVW results, proteins significant at the FDR level of 5% were further examined for the 1) strength of cis-pQTL (weak instrument if the IVW F-statistic<10), 2) heterogeneity in the causal effect estimates of cis-pQTL (significant heterogeneity if the IVW Cochran’s Q test p<0.01), and 3) pleiotropy (MR-Egger intercept test p<0.01) for those with little evidence against the no measurement error assumption of MR-Egger (I^2^≥0.9). When the NOME assumption is not met, an inflated type I error is expected for the pleiotropy test^34^. After excluding proteins that failed in any of the examinations, we compared the IVW and MR-RAPS results, with similar results between methods suggesting finding robustness.

### Prediction model for incident ADRD in participants with a history of MDD at baseline

We evaluated the prediction for incident ADRD using all included proteins (n=2,920) in participants with proteomic data and a history of MDD at baseline. Proteins were selected by a least absolute shrinkage and selection operator (LASSO) Cox regression model, where the regularization parameter lambda that determined the shrinkage of regression coefficients associated with the proteins for a parsimonious model was chosen for close-to-optimal deviance within one standard error of the minimal deviance (i.e., one-standard-error rule)^35^ using the 10-fold cross-validation. The selected proteins were carried forward to fit a Gomperz model^36^ to develop a proteomic risk score (PrRS_MDD-ADRD_) to estimate the 10-year risk of ADRD. We compared different prediction models, including sociodemographic factors and *APOE* e4 carrier status, for the prediction of incident ADRD in MDD using Harrell’s C-index^37^.

To further validate PrRS_MDD-ADRD_, we examined its correlations with intermediate phenotypes of ADRD, i.e., cognitive function measures and brain MRI image-derived phenotypes. We selected five cognitive function measures from the baseline or first imaging visit, depending on when they were first implemented in the UK Biobank,

1. reaction time (processing speed);
2. digit spam forward test (working memory);
3. symbol digit substitution (executive function);
4. trail making test B (executive function);
5. matrix pattern completion (non-verbal reasoning);

showing moderate to high concurrent validity with well-validated reference tests and test–retest reliability^38^. Their UKB field IDs were provided in **Table S1**, with measurement details described elsewhere^38^. We also selected brain MRI T1 structural and T2-weighted image-derived phenotypes (IDPs), including regional gray matter volumes, subcortical volumes, and white matter hyperintensities (UKB filed IDs in **Table S1**). We calculated Spearman correlations between PrRS_MDD-ADRD_ and cognitive function measures or IDPs, and p-values were adjusted using the FDR approach. Cognitive function measures from the first imaging visit were adjusted for the time gap between the baseline and first imaging visits. Brain MRI measures were adjusted for the between-visit time gap and head size.

All the statistical tests were two-sided. The statistical analyses were performed in R version 4.2.3. The R packages used included “survival” for fitting Cox regression models^39^, “glmnet” for fitting LASSO Cox regression models^40^, “flexsurv”^41^ for fitting Gompertz models, “gwasRtools”^42^ for LD clumping, “MendelianRandomization”^43^ for two-sample MR analyses, and “stat” for multiple testing adjustments.

## Results

In the baseline cohort (n=42,807, **Figure S1**), 3,615 were diagnosed with MDD before or at baseline. The characterization of participants with and without a history of MDD at baseline is reported in **Table S3**. During a mean follow-up of 13.3 years (SD=2.2), the incidence of ADRD was higher in participants with a history of MDD than in those who were MDD-free at baseline (4.1% versus 3%), including the incidence of ADRD subtypes, such as Alzheimer’s disease (1.8% versus 1.4%) and vascular dementia (0.9% versus 0.5%). A history of MDD was significantly associated with higher risks of ADRD (HR=1.81, 95% CI 1.52 to 2.15, p=1.37×10^−11^), Alzheimer’s disease (HR=1.72, 95% CI 1.32 to 2.23, p=4.79×10^−5^), and vascular dementia (HR=2.38, 95% CI 1.65 to 3.43, p=3.72×10^−6^) after adjusting for baseline covariates, age, sex, ethnicity, education, BMI, smoking status, diabetes diagnosis, hypertension diagnosis, and *APOE* e4 carrier status.

### Identification of proteins associated with incident ADRD in participants with a history of MDD

Of the 2,920 proteins tested, six were significantly associated with the risk of ADRD in participants with a history of MDD at baseline (FDR-adjusted p < 0.05). Higher expression of GFAP (glial fibrillary acidic protein), NFL (neurofilament light chain protein), and PSG1 (pregnancy-specific beta-1-glycoprotein 1) were associated with increased risk of ADRD, while higher expression of VGF (neurosecretory protein VGF), GST3 (guided entry of tail-anchored proteins factor 3, ATPase), and HPGDS (hematopoietic prostaglandin D synthase) were associated with decreased risk of ADRD upon follow-up (**Table S4**, **Figure 1**). These associations remained statistically significant after excluding *APOE* e4 carrier status from the covariates, but GLYR1 (Glyoxylate Reductase 1 Homolog) also became statistically significant (**Table S5**). In the joint model with GFAP, NEFL, PSG1, GET3, HPGDS, and VGF, adjusting for covariates including *APOE* e4 carrier status, the hazard ratios associated with these proteins little changed compared to those from models with one protein at a time and covariates.

**Figure 1.**
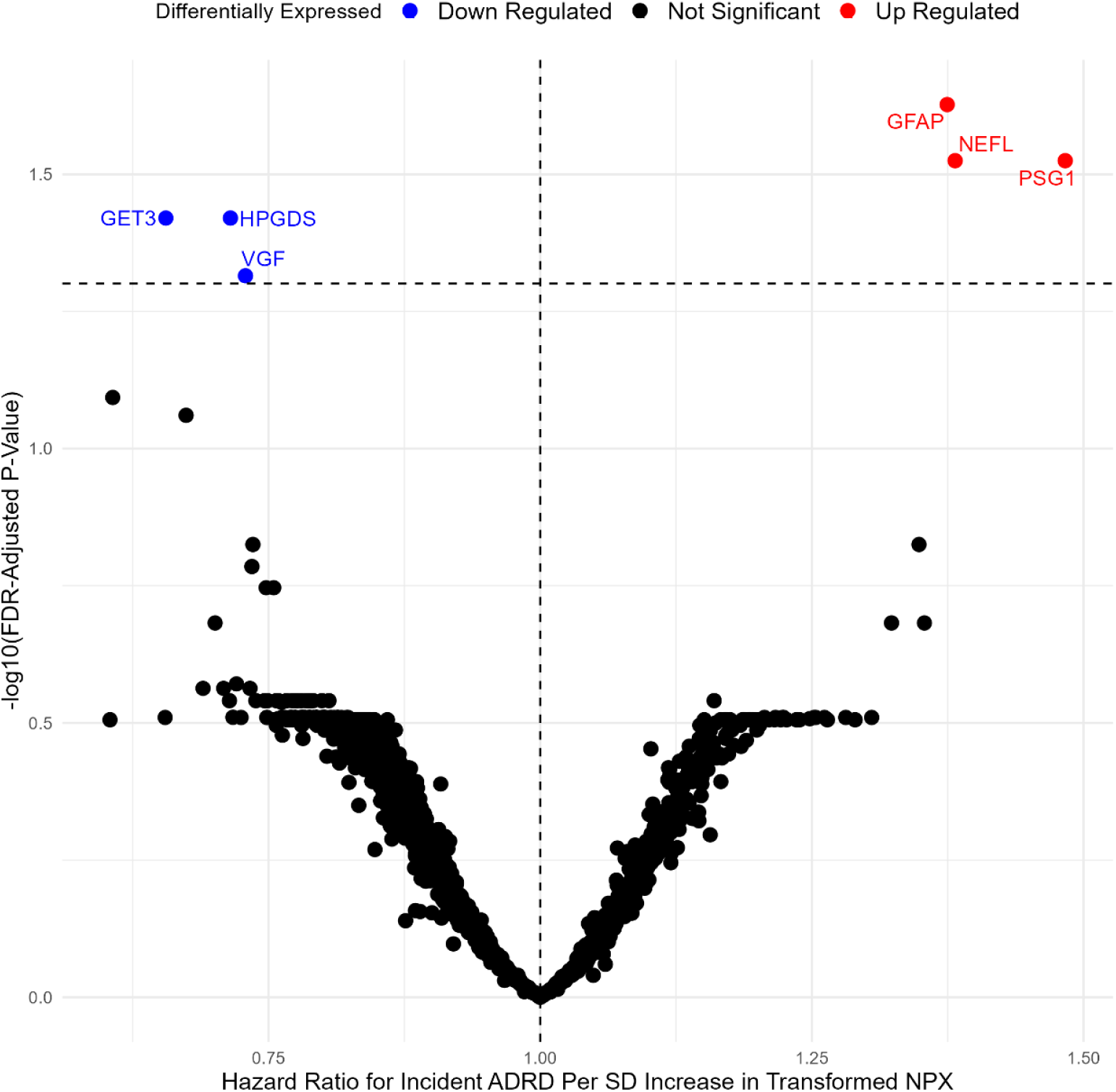
Proteins significantly associated with incident ADRD in participants with a history of MDD at ine

### A two-sample Mendelian randomization analysis

Of 2,920 proteins, 2,003 had one or more autosomal cis-pQTL. After excluding 3,670 associations between cis-pQTL and their coded proteins that showed a significant discrepancy (absolute difference in standardized *β* greater than 0.1) between general whites in the discovery cohort of Sun et al. and its subset with a history of MDD at baseline, 1,982 proteins remained. Half of the proteins had 5 or less cis-pQTL (range 1 to 81). Individual cis-pQTL showed an ignorable discrepancy in effect allele frequencies (≤0.021) between the two cohorts (whites vs. non-whites with a history of MDD).

Genetically determined lower apolipoprotein E and higher IL10RB protein expression levels were significantly associated with incident ADRD in MDD (IVW HR=1.81 and 1.41 per SD, FDR-adjusted p-values 5.79×10^−10^ and 0.035, respectively). **Figure 2** shows the ratio estimate (log(HR)) of per allele association with incident ADD to per allele association with the expression of APOE or IL10RB. Both IVW and MR-RAPS showed similar results (**Table S6**). Additionally, there was no evidence of weak instruments (IVW F-statistic>10), heterogeneity among causal estimates (IVW Cochran’s Q test p>0.01), and pleiotropy (MR-Egger intercept test p>0.01) (**Table S6**).

**Figure 2.**
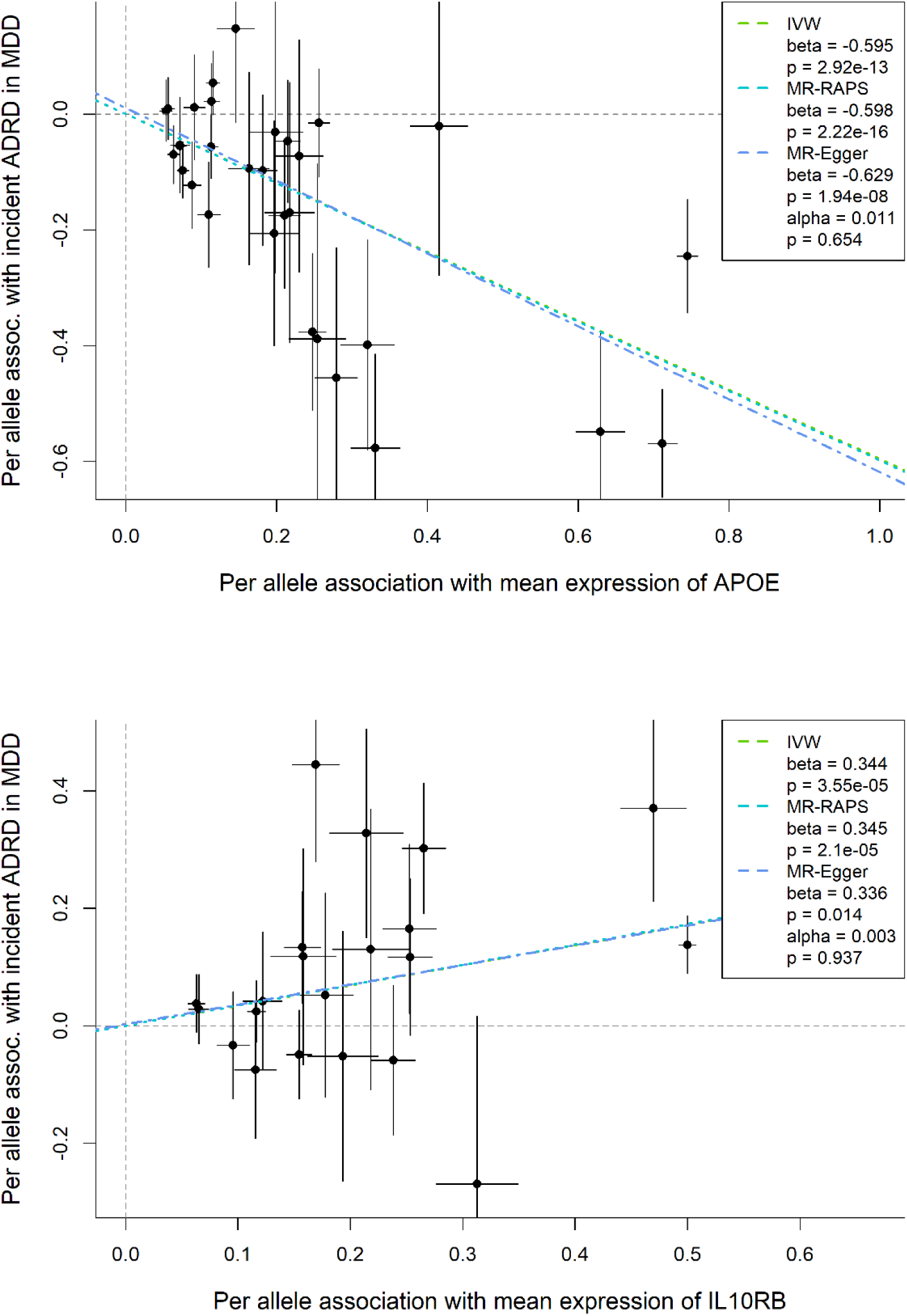
MR-Egger plots to show causal estimates from IVW vs. other MR methods for the effects of APOE and IL10RB (FDR-adjusted p<0.05) on incident ADRD in participants with a history of MDD at ine. The slope estimates represent log(HR) per SD increase in genetically determined APOE or B.

We conducted a sensitivity analysis using the IVW method with 1 or 2 cis-pQTL per protein (n=1,734), as reported in Sun et al^22^., removing cis-pQTL with a minor allele frequency smaller than 0.01. The hazard ratios associated with genetically determined protein expression levels were moderately correlated (Pearson correlation coefficient 0.76) across proteins (n=1,718) between analyses using this set of cis-pQTL and cis-pQTL identified in our setting (primary analysis) (**Table S7**).

### Prediction of ADRD risk in individuals with a history of MDD

We fitted a LASSO Cox regression model to identify the protein set that could best predict the future risk of ADRD among participants with a history of MDD at baseline. The hyperparameter lambda (λ) was chosen as 0.009731. Nineteen proteins were selected by a LASSO Cox regression model (**Table S8**). These proteins were used to develop a Proteomic Risk Score (PrRS_MD-ADRD_) for incident ADRD in a Gomperz model (Gomperz parameter estimates in **Table S9**). The PrRS_MDD-ADRD_ showed a strong discriminative power separating incident ADRD cases and controls within 10-year of follow-up among participants with a history of MDD at baseline (C-statistics = 0.84, SE = 0.016) (**Table 1**). Its discriminative power was higher than common risk factors and predictors of ADRD in the general population (e.g., age, sex, education, and *APOE* e4 carrier status, whether considered individually or in combination). Interestingly, the discriminative power of the model with PrRS_MD-ADRD_ alone was higher than the model with PrRS_MDD-ADRD_, age, sex, education, and *APOE* carrier status.

### Correlations between PrRS_MDD-ADRD_ with intermediate phenotypes of ADRD

We examined the correlations between PrRS_MDD-ADRD_ and intermediate phenotypes of ADRD in participants with a history of MDD at baseline. It is worth noting that the sample size was reduced due to limited overlaps between cohorts and incomplete data, ranging from 556 (numeric memory) to 4527 individuals (reaction time test) for cognitive function measures and from 568 (white matter hyperintensities phenotypes) to 591 (regional cortical volume phenotypes) for brain MRI IDPs. An increased PrRS_MDD-ADRD_ was significantly correlated with worse cognitive performance among individuals with a history of MDD at baseline (**Figure 3**). These cognitive domains are commonly affected in individuals with a history of MDD and are strong predictors of the future development of ADRD^44^. We also observed significant correlations of PrRS_MDD-ADRD_ with atrophy in multiple cortical and subcortical regions and increased cerebrovascular burden in brain regions critical for cognitive and emotional processing and implicated in both MDD and ADRD^45^ (**Figure 4, Table S10)**. These results reinforce the robustness of PrRS_MDD-ADRD_.

**Figure 3-.**
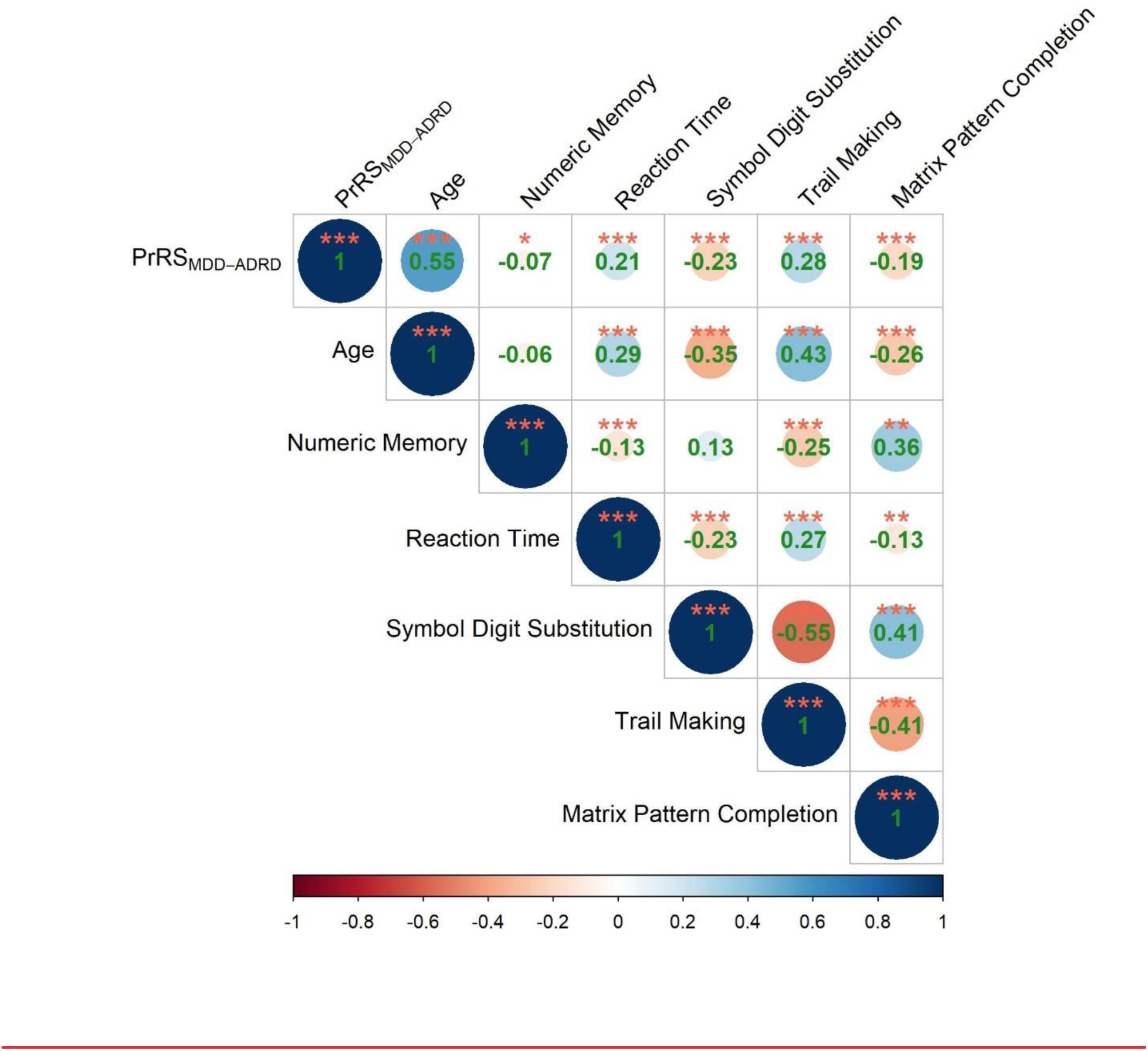
Spearman correlations between PrRS_MDD-ADRD_, age, and cognitive performance measures baseline (reaction time, numeric memory, fluid intelligence score) or first imaging visit (symbol substitution, trail making, matrix pattern completion). Significance: ***p<0.001, **0.01<p<0.001, <p<0.05.

**Figure 4.**
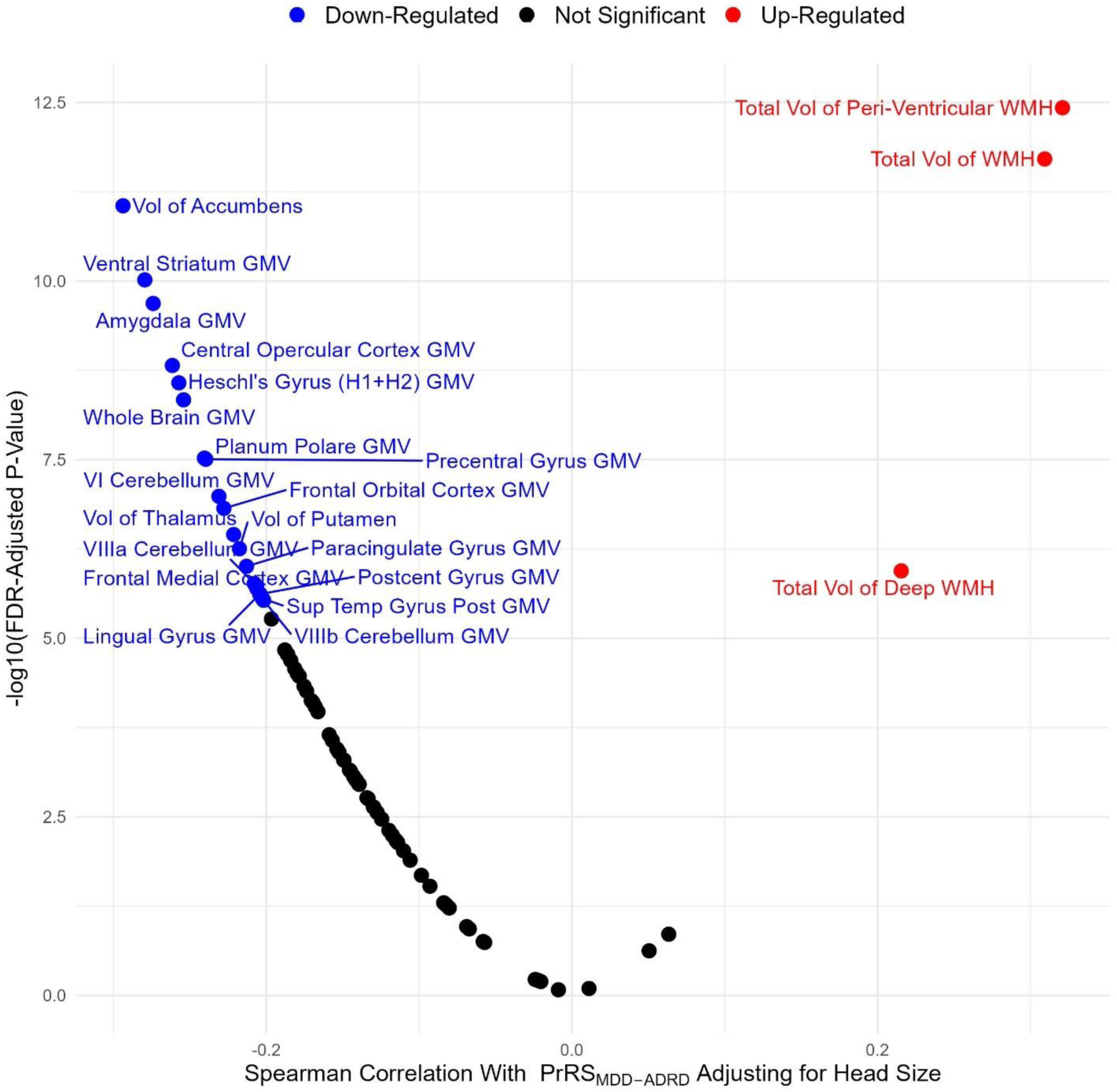
Spearman correlations of PrRS_MDD-ADRD_ with T1 structural and T2-weighted brain MRI image-ed phenotypes (IDPs), adjusting for head size. IDPs labeled if FDR-adjusted p<0.05 and Spearman lation >0.2.

## Discussion

Prior studies showed a significant genetic correlation between MDD and ADRD, a significant overlap of biological processes between MDD and ADRD (including inflammation-related pathways), and that amyloid-related pathways are causally linked to MDD and ADRD using bi-directional Mendelian randomization. However, these studies relied on cross-sectional data, summary-based GWAS data, and analyses of post-mortem brain tissues, and despite the robustness of the findings, these samples and study designs can introduce significant biases to the results. Moreover, these studies did not provide a biologically based predictive model to identify individuals with MDD that have the highest risk to progress to ADRD over time.

Our results are consistent with these prior observations, but significantly extend them in several ways. First, using proteomic data from the UKB, we found that a small set of proteins was significantly associated with the incidence of ADRD among individuals with MDD, including 2 proteins (i.e., NfL and GFAP) that have been extensively associated with the risk of ADRD in the general population. Our Mendelian randomization approach relied on the identification of cis-pQTLs as genetic instruments instead of GWAS summary statistics, which can provide more robust causal inference evidence^21, 22^. Using this approach, we determined that genetically determined protein expression of apolipoprotein E and IL-10 receptor subunit B are causally related to the elevated risk of ADRD in individuals with MDD. Finally, we developed a proteomic risk score (PRS_MDD-ADRD_) with strong predictive power to identify those with MDD that will progress to ADRD over a long-term follow-up. Importantly, PrRS_MDD-ADRD_ was also associated with intermediate phenotypes relevant to both MDD and ADRD, such as worse cognitive performance, cortical brain atrophy in areas relevant to both conditions, and cerebrovascular disease burden. Therefore, PrRS_MDD-ADRD_ can be used in clinical trials as a biomarker to identify those with the highest risk of developing ADRD to test interventions aiming at reducing the risk of ADRD in a highly vulnerable population.

We identified a small set of proteins that were significantly associated with the risk of ADRD in MDD. The GFAP and NfL are well-established markers of astroglial activation and neurodegeneration, respectively, higher levels of these proteins in the blood are associated with progression from mild cognitive impairment to clinical dementia in multiple cohorts^46^. Our findings, thus, support the role of unspecific neurodegenerative and neuroinflammatory abnormalities as markers related to the progression to ADRD in individuals with MDD. VGF is a multifunctional polyprotein, primarily secreted by neurons and involved in neuroplasticity, neurogenesis, and energy metabolism^47^. Previous studies showed that VGF levels are reduced in MDD (both in plasma and CSF), and lower levels are associated with more severe cognitive impairment^48, 49^. Also, lower levels of VGF have been reported in older adults with MCI and AD and related to Alzheimer’s disease neuropathological changes^50, 51^.

We found novel proteins associated with the risk of ADRD among individuals with MDD that have not been previously reported in the literature. HPGDS is responsible for the conversion of prostaglandin H2 (PGH2) to prostaglandin G2 (PGD2) in the immune cells. The primary effect of PGD2 is the regulation of inflammatory processes through the recruitment of CD4+ TH_2_ cells and it has been associated with allergic reactions and asthma development^52^. However, PGD2 is also the most abundant prostaglandin in the brain and previous studies have reported lower levels in individuals with MDD^53^, but higher levels of PGD2 have been reported in AD^54^. Therefore, our results showing that higher levels of HPGDS are associated with reduced risk of ADRD are contradictory and warrant further investigation. GET3 is a protein chaperone and important in the cytoplasmic protein trafficking and protection against oxidative stress damage, playing a major role in maintaining proteostasis^55^. PSG1 is a protein that is mostly secreted during pregnancy in the placenta, but is also secreted by multiple tissues in non-pregnant people^56^. It has a potent immunomodulatory effect by activating the TGF-β signaling pathway^57^ and higher expression of PSG1 is associated with poor prognosis in multiple cancers^58^. Further investigations are necessary to clarify their roles in both conditions and how they can lead to a higher risk of ADRD in individuals with MDD.

The Mendelian randomization analyses revealed that genetically determined protein expression of apolipoprotein E and IL-10 receptor is causally linked to the risk of ADRD among individuals with MDD. The *APOE* gene is a well-established risk factor for ADRD and the presence of its ε4 allele significantly increases the ADRD risk, while the ε2 allele is protective against it in the general population^59, 60^. The presence of the allele ε4 leads to structural modification, reduced lipidation potential, and lower protein expression levels^61–63^ of apolipoprotein E, and the net effect is reduced clearance of toxic amyloid-β proteins in the brain and a greater propensity to amyloid-β aggregation and development of neuritic plaques. A recent large-scale proteogenomic study showed that *APOE* is also a pQTL and the allele ε2 leads to higher genetically determined protein expression of apolipoprotein E compared to the allele ε4^22^. Such finding provides additional mechanistic evidence why different *APOE* gene polymorphisms confer a protective or harmful effect against ADRD development.

On the other hand, a genetically determined higher expression of IL-10 receptor subunit B (IL-10RB) is causally linked to ADRD among MDD individuals. The IL-10 cytokine, through the interaction of its receptor IL-10, has a primarily anti-inflammatory effect and is a master regulator of the resolution of the inflammatory response^64^. IL-10 has a similar immunomodulatory effect in the brain and is associated with the response against acute insults to the brain, e.g., acute brain injuries and stroke^65^. However, the hyperactivation of the IL-10/IL-10R system can be detrimental, preventing the resolution of tissue damage, autoimmune conditions, and immunological escape of tumors ^66, 67^. Interestingly, the primary intracellular signaling pathway activated by IL-10/IL-10R is the JAK/STAT signaling pathway. The overactivation of this pathway can lead to the inhibition of pro-apoptotic factors and induction of cellular senescence^68, 69^. It is worth noting that elevated senescence burden has been associated with major depression and cognitive impairment across the lifespan^18, 70^ and that IL-10 is overexpressed in immunosenescent cells^71^, thus increased activation of IL-10 in this context may be more reflective of this cytokine’s role in cellular senescence than its anti-inflammatory properties.

Overall, our findings from the Mendelian randomization and observational analyses provide a robust mechanistic explanation for the higher risk of ADRD in individuals with MDD. First, a genetically determined reduction in the apolipoprotein E expression can promote the aggregation of the amyloid-β protein in the brain. Coupled with impaired control of the immune response by the genetically determined higher expression of IL-10RB, there is a reduction in the capacity of brain tissues to resolve the local insults secondary to the amyloid-β accumulation. Over time, the imbalance of amyloid-β accumulation and lower damage resolution capacity in individuals with MDD can lead to additional development of astrogliosis, neuronal injury, cellular senescence, reduced neurotrophic support, impaired proteostasis and metabolic control, culminating in the progression of neurodegenerative changes and the development of ADRD. It is important to note that other characteristics of a depressive episode, that were not captured in the current study, like chronic perceived stress, medical comorbidities, and poor lifestyle and behaviors, can contribute to the intensification of these pathophysiological processes and moderate the risk of ADRD in individuals with MDD.

Antidepressant treatment may have a mild effect on improving cognitive performance in individuals with MDD^72^, although they do not seem to have a robust effect preventing ADRD in individuals with MDD^73^. Therefore, more specific interventions are needed, and our results point to more specific treatment targets for interventions aiming to mitigate the risk of ADRD in this group. For example, several drugs that inhibit the JAK/STAT pathway, a major pathway activated by the IL-10 receptor, are clinically available (e.g., baricitinib and tofacitinib) and could be repurposed aiming the prevention of ADRD in individuals with MDD; however, their side effect profile and low brain penetrance may preclude its effectiveness for this purpose^74^. On the other hand, there has been a growing interest in modulating apolipoprotein E effects as a treatment target for ADRD and several compounds have been developed and tested in animal models^75^ and they could be also promising in the prevention of ADRD in individuals with MDD.

To the best of our knowledge, we were the first to develop a proteomic risk score estimating the 10-year risk of ADRD in individuals with a history of MDD. Our model (PrRS_MDD-ADRD_) had a strong discriminative performance, with a C-statistics of 0.84. Interestingly, PrRS_MDD-ADRD_ alone showed a stronger predictive performance than well-established risk factors for ADRD in the general population, including APOE genotype, socio-demographic variables (age, education, and sex), or their combination. Importantly, the PrRS_MDD-ADRD_ was also associated with intermediate phenotypes of ADRD like worse cognitive performance, atrophy in cortical and subcortical brain regions, and cerebrovascular burden. These findings support the robustness of PrRS_MDD-ADRD_ to predict ADRD development in MDD populations and for its potential use in observational studies and clinical trials aiming to evaluate the association between MDD and ADRD.

Our results should be interpreted in light of the study limitations. The UK Biobank sample is relatively healthier, with better socioeconomic status, and predominantly white compared to the general UK population. Thus, our findings may not be generalizable to the general population or other geographical locations. The identification of MDD cases was based on electronic health records. Despite of the strong validity of EHR for the identification of MDD cases in the UK Biobank^76^, the lack of formal psychiatric interview for the majority of UK Biobank participants does not allow for a fine-grained characterization of the major depressive episode, such as currently depressed or in remission, age of onset, chronicity and number of prior episodes, trajectories of depressive symptoms after the diagnosis, treatment response which are all variables that can influence the risk of ADRD in this population^77, 78^. Similarly, the identification of ADRD was based on EHR and there is no information about AD-related biomarkers (amyloid-β and phosphorylated Tau protein). Thus, there is a risk of misclassification of cases that could have influenced the current results. We were not able to validate our models and results using an external validation study. This is due to the absence of large-scale epidemiologic studies with the identification of MDD cases and future cases of ADRD that also have used the OLINK Explore 3072^®^ proteomic assay. Therefore, our findings must be replicated and validated in future studies, including large and diverse sample sizes.

Despite the study limitation, our analyses have unique strengths such as the large sample of individuals with MDD, proteomic data, and a long follow-up. Despite UKB participants being younger, healthier, and better off socio-economically we were still able to detect the association between MDD and ADRD and to develop a robust predictive model based on the proteomic data. Finally, the Mendelian randomization analysis, using cis-pQTLs as genetic instruments, allows one to move beyond typically descriptions of associations with all the multitudes of different and unavoidable biases of observational studies and towards studies involving inference of biological causation.

## Data Availability

All data produced in the present work are contained in the manuscript

## Acknowledgments

This work was supported by NIH grant P30AG067988. Access to UK Biobank data was granted under application no.92647 “Research to Inform the Field of Precision Gerontology” (PI: Richard H. Fortinski).

