## Supplementary material for "Proteogenomic signature of risk of Alzheimer’s disease and related dementia risk in individuals with a history of major depression disorder": Table

**Table 1 Incident ADRD prediction in participants with a history of MDD at baseline using proteins, sociodemographic factors, and *APOE* e4 carrier status**

| Model | Outcome | Predictors | C-Statistic (SE) |
|---|---|---|---|
| 1 | Incident ADRD | Age | 0.754 (0.019) |
| 2 | Incident ADRD | *APOE* e4 carrier status | 0.588 (0.020) |
| 3 | Incident ADRD | Age + Sex + Education + *APOE* e4 carrier status | 0.765 (0.019) |
| **4** | **Incident ADRD** | **$PrRS_{MD\text{-}ADRD}$** | **0.837 (0.016)** |
| 5 | Incident ADRD | $PrRS_{MD\text{-}ADRD}$ + Age + Sex + Education + *APOE* e4 allele(s) | 0.796 (0.019) |
