## Supplementary material for "Proteogenomic signature of risk of Alzheimer’s disease and related dementia risk in individuals with a history of major depression disorder"

**Supplementary Tables**

**Table S1 UK Biobank field IDs to extract data for the present project**
**Table S2 A list of antidepressant ATC codes and the corresponding UK Biobank codes in the baseline self-reported medication data to identify participants using antidepressants at baseline assessment**
**Table S3 Baseline participant characteristics of incident ADRD cases versus those who remained ADRD-free during follow-up in the baseline MDD cohort**
**Table S4 Associations between protein expression levels and incident ADRD in participants with a history of MDD at baseline, adjusting for covariates including *APOE* e4 carrier status**
**Table S5 Associations between protein expression levels and incident ADRD in participants with a history of MDD at baseline, adjusting for covariates without *APOE* e4 carrier status**
**Table S6 Two-sample Mendelian randomization analysis results using cis-pQTL identified in this present study to test the causal effects of proteins on incident ADRD in participants with a history of MDD at baseline**
**Table S7 Two-sample Mendelian randomization analysis results using cis-pQTL reported by Sun et al. to test the causal effects of proteins on incident ADRD in participants with a history of MDD at baseline**
**Table S8 19 Hazard ratios for incident ADRD associated with the 19 proteins selected by a LASSO Cox regression model**
**Table S9 Parameter estimates associated the 19 proteins selected by a LASSO Cox regression model in a Gomperz regression model**
**Table S10 Spearman correlations of $PrRS_{MDD\text{-}ADRD}$ with T1 structural and T2-weighted brain MRI image-derived phenotypes (IDPs)**

**Table S1 UK Biobank field IDs to extract data for the present project**

Instance 0: initial assessment visit (2006-2010) at which participants were recruited and consent given

Instance 2 : Imaging visit (2014+)

| Category | Field | Field ID | ICD-10 code | Instance |
|---|---|---|---|---|
| Major depressive disorder (MDD) | 1. Depressive episode | 130894 | F32 | - |
| | 2. Recurrent depressive disorder | 130896 | F33 | - |
| | 3. Persistent mood [affective] disorders | 130898 | F34 | - |
| | 4. Other mood [affective] disorders | 130900 | F38 | - |
| | 5. Unspecified mood [affective] disorder | 130902 | F39 | - |
| Excluded: Other psychiatric disorders | 1. Schizophrenia | 130874 | F20 | - |
| | 2. Unspecified nonorganic psychosis | 130888 | F29 | - |
| | 3. Manic episode | 130890 | F30 | - |
| | 4. Bipolar affective disorder | 130892 | F31 | - |
| Alzheimer's disease and related dementia (ADRD) | Date of ADRD first reported | | | |
| | 1. AD | 131036 | G30 | - |
| | 2. Dementia in AD | 130836 | F00 | - |
| | 3. Vascular dementia | 130838 | F01 | - |
| | 4. Unspecified dementia | 130842 | F03 | - |
| | 5. Other degenerative diseases of nervous system, not elsewhere classified | 131038 | G31 | - |
| Excluded: dementia in other diseases | Date of F02 first reported (dementia in other diseases classified elsewhere) | 130840 | F02 | - |
| OLINK protein biomarkers | OLINK protein biomarkers (record table 1072) | 30900 | - | - |
| Association or Mendelian randomization covariates | 1. Age at recruitment | 21022 | - | 0 |
| | 2. Sex | 31 | - | - |
| | 3. Ethnic background | 21000 | - | - |
| | 4. Qualifications (education) | 6138 | - | - |
| | 5. Body mass index (BMI) | 21001 | - | 0 |
| | 6. Smoking status | 20116 | - | 0 |
| | 7. APOE e4 status | 22418 | - | - |
| | 8. Date of hypertension first reported | | | |
| | - Essential (primary) hypertension | 131286 | I10 | - |
| | hypertensive heart disease | | | - |
| | - Hypertensive heart disease | 131288 | I11 | - |
| | - Hypertensive re-l disease | 131290 | I12 | - |
| | - Hypertensive heart and re-l disease | 131292 | I13 | - |
| | - Secondary hypertension | 131294 | I15 | - |
| | 9. Date of diabetes first reported | | | |
| | - Insulin-dependent diabetes mellitus | 130706 | E10 | - |
| | - Non-insulin-dependent diabetes mellitus | 130708 | E11 | - |
| | - Malnutrition-related diabetes mellitus | 130710 | E12 | - |
| | - Other specified diabetes mellitus | 130712 | E13 | - |
| | - Unspecified diabetes mellitus | 130714 | E14 | 0 |
| | 10. Treatment/medication code | 20003 | | |
| | | | - | |
| | 11. Genetic principal components | 22009 | | |
| | 12. Genotype measurement batch | 22000 | | |
| Other considerations | | | | |
| Recruitment | Date of attending assessment center | 53 | - | 0 |
| Death register | Date of death | 40000 | - | - |
| Genotypes | Genetic ethnic grouping | 22006 | - | - |
| Recruitment | Date of attending assessment centre | 53 | | 0, 2 |
| Cognitive function | | | | |
| Reaction time | Mean time to correctly identify matches | 20023 | - | 0 |
| Numeric memory | Maximum digits remembered correctly | 4282 | - | 0 |
| Symbol digit substitution | Number of symbol digit matches made correctly | 23324 | - | 2 |
| Trail making | Duration to complete alphanumeric path (trail #2) | 6350 | - | 2 |
| Matrix pattern completion | Number of puzzles correctly solved | 6373 | - | 2 |
| Brain MRI | | | | |
| T2-weighted brain MRI | Total volume of peri-ventricular white matter hyperintensities | 24485 | | |
| T2-weighted brain MRI | Total volume of white matter hyperintensities (from T1 and T2_FLAIR images) | 25781 | | |
| T2-weighted brain MRI | Total volume of deep white matter hyperintensities | 24486 | | |
| T1 structural brain MRI | Volumetric scaling from T1 head image to standard space | 25000 | | |
| Freesurfer ASEG | Volume of TotalGray in the whole brain generated by subcortical volumetric segme[illegible] | 26518 | | |
| Subcortical volumes (FIRST) | Volume of accumbens (from T1 brain image) | 25023/25024 | | |
| Subcortical volumes (FIRST) | Volume of thalamus (from T1 brain image) | 25011/25012 | | |
| Subcortical volumes (FIRST) | Volume of putamen (from T1 brain image) | 25015/25016 | | |
| Subcortical volumes (FIRST) | Volume of caudate (from T1 brain image) | 25013/25014 | | |
| Subcortical volumes (FIRST) | Volume of hippocampus (from T1 brain image) | 25019/25020 | | |
| Subcortical volumes (FIRST) | Volume of pallidum (from T1 brain image) | 25017/25018 | | |
| Subcortical volumes (FIRST) | Volume of amygdala (from T1 brain image) | 25021/25022 | | |
| Regional grey matter volumes (FAST) | Volume of grey matter in Ventral Striatum | 25890/25891 | | |
| Regional grey matter volumes (FAST) | Volume of grey matter in Amygdala | 25888/25889 | | |
| Regional grey matter volumes (FAST) | Volume of grey matter in Central Opercular Cortex | 25864/25865 | | |
| Regional grey matter volumes (FAST) | Volume of grey matter in Heschl's Gyrus (includes H1 and H2) | 25870/25871 | | |
| Regional grey matter volumes (FAST) | Volume of grey matter in Planum Polare | 25868/25869 | | |
| Regional grey matter volumes (FAST) | Volume of grey matter in Precentral Gyrus | 25794/25795 | | |
| Regional grey matter volumes (FAST) | Volume of grey matter in VI Cerebellum | 25897/25899 | | |
| Regional grey matter volumes (FAST) | Volume of grey matter in Frontal Orbital Cortex | 25846/25847 | | |
| Regional grey matter volumes (FAST) | Volume of grey matter in Paracingulate Gyrus | 25836/25837 | | |
| Regional grey matter volumes (FAST) | Volume of grey matter in VIIIa Cerebellum | 25909/25911 | | |
| Regional grey matter volumes (FAST) | Volume of grey matter in Frontal Medial Cortex | 25830/25831 | | |
| Regional grey matter volumes (FAST) | Volume of grey matter in Lingual Gyrus | 25852/25853 | | |
| Regional grey matter volumes (FAST) | Volume of grey matter in Postcentral Gyrus | 25814/25815 | | |
| Regional grey matter volumes (FAST) | Volume of grey matter in Superior Temporal Gyrus, posterior division | 25800/25801 | | |
| Regional grey matter volumes (FAST) | Volume of grey matter in VIIIb Cerebellum | 25912/25914 | | |
| Regional grey matter volumes (FAST) | Volume of grey matter in Precuneous Cortex | 25842/25843 | | |
| Regional grey matter volumes (FAST) | Volume of grey matter in Lateral Occipital Cortex, superior division | 25824/25825 | | |
| Regional grey matter volumes (FAST) | Volume of grey matter in Temporal Pole | 25796/25797 | | |
| Regional grey matter volumes (FAST) | Volume of grey matter in Hippocampus | 25886/25887 | | |
| Regional grey matter volumes (FAST) | Volume of grey matter in Temporal Fusiform Cortex, posterior division | 25856/25857 | | |
| Regional grey matter volumes (FAST) | Volume of grey matter in X Cerebellum | 25918/25920 | | |

| | | |
|---|---|---|
| Regional grey matter volumes (FAST) | Volume of grey matter in Superior Frontal Gyrus | 25786/25787 |
| Regional grey matter volumes (FAST) | Volume of grey matter in Frontal Pole | 25782/25783 |
| Regional grey matter volumes (FAST) | Volume of grey matter in Superior Temporal Gyrus, anterior division | 25798/25799 |
| Regional grey matter volumes (FAST) | Volume of grey matter in Middle Temporal Gyrus, posterior division | 25804/25805 |
| Regional grey matter volumes (FAST) | Volume of grey matter in Lateral Occipital Cortex, inferior division | 25826/25827 |
| Regional grey matter volumes (FAST) | Volume of grey matter in Supramarginal Gyrus, posterior division | 25820/25821 |
| Regional grey matter volumes (FAST) | Volume of grey matter in Crus I Cerebellum | 25900/25902 |
| Regional grey matter volumes (FAST) | Volume of grey matter in Vermis VIIIa Cerebellum | 25910 |
| Regional grey matter volumes (FAST) | Volume of grey matter in Temporal Fusiform Cortex, anterior division | 25854/25855 |
| Regional grey matter volumes (FAST) | Volume of grey matter in VIIb Cerebellum | 25906/25908 |
| Regional grey matter volumes (FAST) | Volume of grey matter in Supracalcarine Cortex | 25874/25875 |
| Regional grey matter volumes (FAST) | Volume of grey matter in V Cerebellum | 25895/25896 |
| Regional grey matter volumes (FAST) | Volume of grey matter in Inferior Temporal Gyrus, posterior division | 25810/25811 |
| Regional grey matter volumes (FAST) | Volume of grey matter in Occipital Fusiform Gyrus | 25860/25861 |
| Regional grey matter volumes (FAST) | Volume of grey matter in Brain-Stem | 25892 |
| Regional grey matter volumes (FAST) | Volume of grey matter in Parahippocampal Gyrus, anterior division | 25848/25849 |
| Regional grey matter volumes (FAST) | Volume of grey matter in Vermis VI Cerebellum | 25898 |
| Regional grey matter volumes (FAST) | Volume of grey matter in Intracalcarine Cortex | 25828/25829 |
| Regional grey matter volumes (FAST) | Volume of grey matter in Middle Frontal Gyrus | 25788/25789 |
| Regional grey matter volumes (FAST) | Volume of grey matter in Occipital Pole | 25876/25877 |
| Regional grey matter volumes (FAST) | Volume of grey matter in Planum Temporale | 25872/25873 |
| Regional grey matter volumes (FAST) | Volume of grey matter in Insular Cortex | 25784/25785 |
| Regional grey matter volumes (FAST) | Volume of grey matter in Vermis VIIb Cerebellum | 25907 |
| Regional grey matter volumes (FAST) | Volume of grey matter in Crus II Cerebellum | 25903/25905 |
| Regional grey matter volumes (FAST) | Volume of grey matter in Frontal Operculum Cortex | 25862/25863 |
| Regional grey matter volumes (FAST) | Volume of grey matter in Putamen | 25882/25883 |
| Regional grey matter volumes (FAST) | Volume of grey matter in Superior Parietal Lobule | 25816/25817 |
| Regional grey matter volumes (FAST) | Volume of grey matter in Vermis VIIIb Cerebellum | 25913 |
| Regional grey matter volumes (FAST) | Volume of grey matter in Inferior Temporal Gyrus, anterior division | 25808/25809 |
| Regional grey matter volumes (FAST) | Volume of grey matter in I-IV Cerebellum | 25893/25894 |
| Regional grey matter volumes (FAST) | Volume of grey matter in Angular Gyrus | 25822/25823 |
| Regional grey matter volumes (FAST) | Volume of grey matter in Inferior Frontal Gyrus, pars opercularis | 25792/25793 |
| Regional grey matter volumes (FAST) | Volume of grey matter in Middle Temporal Gyrus, anterior division | 25802/25803 |
| Regional grey matter volumes (FAST) | Volume of grey matter in Parietal Operculum Cortex | 25866/25867 |
| Regional grey matter volumes (FAST) | Volume of grey matter in Subcallosal Cortex | 25834/25835 |
| Regional grey matter volumes (FAST) | Volume of grey matter in Cuneal Cortex | 25844/25845 |
| Regional grey matter volumes (FAST) | Volume of grey matter in Juxtapositional Lobule Cortex (formerly Supplementary Mo | 25832/25833 |
| Regional grey matter volumes (FAST) | Volume of grey matter in Temporal Occipital Fusiform Cortex | 25858/25859 |
| Regional grey matter volumes (FAST) | Volume of grey matter in IX Cerebellum | 25915/25917 |
| Regional grey matter volumes (FAST) | Volume of grey matter in Supramarginal Gyrus, anterior division | 25818/25819 |
| Regional grey matter volumes (FAST) | Volume of grey matter in Vermis X Cerebellum | 25919 |
| Regional grey matter volumes (FAST) | Volume of grey matter in Parahippocampal Gyrus, posterior division | 25850/25851 |
| Regional grey matter volumes (FAST) | Volume of grey matter in Vermis IX Cerebellum | 25916 |
| Regional grey matter volumes (FAST) | Volume of grey matter in Inferior Frontal Gyrus, pars triangularis | 25790/25791 |
| Regional grey matter volumes (FAST) | Volume of grey matter in Pallidum | 25884/25885 |
| Regional grey matter volumes (FAST) | Volume of grey matter in Inferior Temporal Gyrus, temporooccipital part | 25812/25813 |
| Regional grey matter volumes (FAST) | Volume of grey matter in Middle Temporal Gyrus, temporooccipital part | 25806/25807 |
| Regional grey matter volumes (FAST) | Volume of grey matter in Caudate | 25880/25881 |
| Regional grey matter volumes (FAST) | Volume of grey matter in Cingulate Gyrus, anterior division | 25838/25839 |
| Regional grey matter volumes (FAST) | Volume of grey matter in Thalamus | 25878/25879 |
| Regional grey matter volumes (FAST) | Volume of grey matter in Vermis Crus II Cerebellum | 25904 |
| Regional grey matter volumes (FAST) | Volume of grey matter in Vermis Crus I Cerebellum | 25901 |
| Regional grey matter volumes (FAST) | Volume of grey matter in Cingulate Gyrus, posterior division | 25840/25841 |

**Table S2 A list of antidepressant ATC codes and the corresponding UK Biobank codes in the baseline self-reported medication data to identify participants using antidepressants at baseline assessment**

| Category | UK Biobank Coding | Medication ATC[1] code | Drug Class | Drug Name |
|---|---|---|---|---|
| imipramine | 1140879630 | N06AA02 | Non-selective monoamine reuptake inhibitors | Imipramine |
| clomipramine | 1140879620 | N06AA04 | Non-selective monoamine reuptake inhibitors | Clomipramine |
| anafranil 10mg capsule | 1140867690 | N06AA04 | Non-selective monoamine reuptake inhibitors | Clomipramine |
| trimipramine | 1140867756 | N06AA06 | Non-selective monoamine reuptake inhibitors | Trimipramine |
| surmontil 10mg tablet | 1140867758 | N06AA06 | Non-selective monoamine reuptake inhibitors | Trimipramine |
| lofepramine | 1140867726 | N06AA07 | Non-selective monoamine reuptake inhibitors | Lofepramine |
| amitriptyline | 1140879616 | N06AA09 | Non-selective monoamine reuptake inhibitors | Amitriptyline |
| nortriptyline | 1140867818 | N06AA10 | Non-selective monoamine reuptake inhibitors | Nortriptyline |
| allegron 10mg tablet | 1140867820 | N06AA10 | Non-selective monoamine reuptake inhibitors | Nortriptyline |
| doxepin | 1140867640 | N06AA12 | Non-selective monoamine reuptake inhibitors | Doxepin |
| dosulepin | 1140909806 | N06AA16 | Non-selective monoamine reuptake inhibitors | Dosulepin |
| dothiepin | 1140879628 | N06AA16 | Non-selective monoamine reuptake inhibitors | Dosulepin |
| prothiaden 25mg capsule | 1140867624 | N06AA16 | Non-selective monoamine reuptake inhibitors | Dosulepin |
| fluoxetine | 1140879540 | N06AB03 | Selective serotonin reuptake Inhibitors | Fluoxetine |
| prozac 20mg capsule | 1140867876 | N06AB03 | Selective serotonin reuptake Inhibitors | Fluoxetine |
| oxactin 20mg capsule | 1141174756 | N06AB03 | Selective serotonin reuptake Inhibitors | Fluoxetine |
| citalopram | 1140921600 | N06AB04 | Selective serotonin reuptake Inhibitors | Citalopram |
| cipramil 10mg tablet | 1141151946 | N06AB04 | Selective serotonin reuptake Inhibitors | Citalopram |
| paroxetine | 1140867888 | N06AB05 | Selective serotonin reuptake Inhibitors | Paroxetine |
| seroxat 20mg tablet | 1140882236 | N06AB05 | Selective serotonin reuptake Inhibitors | Paroxetine |
| sertraline | 1140867878 | N06AB06 | Selective serotonin reuptake Inhibitors | Sertraline |
| lustral 50mg tablet | 1140867884 | N06AB06 | Selective serotonin reuptake Inhibitors | Sertraline |
| fluvoxamine | 1140879544 | N06AB08 | Selective serotonin reuptake Inhibitors | Fluvoxamine |
| escitalopram | 1141180212 | N06AB10 | Selective serotonin reuptake Inhibitors | Escitalopram |
| cipralex 5mg tablet | 1141190158 | N06AB10 | Selective serotonin reuptake Inhibitors | Escitalopram |
| phenelzine | 1140867850 | N06AF03 | Monoamine oxidase inhibitors, non-selective | Phenelzine |
| tranylcypromine | 1140867914 | N06AF04 | Monoamine oxidase inhibitors, non-selective | Tranylcypromine |
| moclobemide | 1140867920 | N06AG02 | Monoamine oxidase A inhibitors | Moclobemide |
| tryptophan product | 1140867960 | N06AX02 | Other | Tryptophan |
| trazodone | 1140879634 | N06AX05 | Other | Trazodone |
| mirtazapine | 1141152732 | N06AX11 | Other | Mirtazapine |
| zispin 30mg tablet | 1141152736 | N06AX11 | Other | Mirtazapine |
| zyban 150mg m/r tablet | 1141176858 | N06AX12 | Other | Bupropion |
| bupropion | 1141176854 | N06AX12 | Other | Bupropion |
| venlafaxine | 1140916282 | N06AX16 | Other | Venlafaxine |
| efexor 37.5mg tablet | 1140916288 | N06AX16 | Other | Venlafaxine |
| reboxetine | 1141151978 | N06AX18 | Other | Reboxetine |
| duloxetine | 1141200564 | N06AX21 | Other | Duloxetine |
| cymbalta 30mg gastro-resistant | 1141201834 | N06AX21 | Other | Duloxetine |
| yentreve 20mg gastro-resistant | 1141200570 | N06AX21 | Other | Duloxetine |
| st john's wort/hypericum [ctsu] | 1201 | N06AX25 | Other | St. John'S Wort |

[1]ATC code: Anatomical Therapeutic Chemical code

**Table S3 Baseline participant characteristics of incident ADRD cases versus those who remained ADRD-free during follow-up in the baseline MDD cohort**

| Baseline participant characteristics | Levels | Control (n=3,465)[1] | ADRD (n=150)[1] | P-Value[2] |
|---|---|---|---|---|
| Age at Recruitment | | 55.3 ± 7.8;<br>56 (40, 70) | 62.4 ± 6.5;<br>64 (42, 70) | 3.70E-33 |
| Sex | Female | 2254 (65.1%) | 87 (58%) | 0.048 |
| | Male | 1211 (34.9%) | 63 (42%) | |
| Ethnicity | White | 3311 (95.6%) | 148 (98.7%) | 0.115 |
| | Asian | 50 (1.4%) | 0 (0%) | |
| | Black | 38 (1.1%) | 1 (0.7%) | |
| | Other | 66 (1.9%) | 1 (0.7%) | |
| Education | College or University degree | 1019 (29.4%) | 34 (22.7%) | 8.50E-04 |
| | A levels/AS levels or equivalent | 396 (11.4%) | 13 (8.7%) | |
| | O levels/GCSEs or equivalent | 729 (21%) | 27 (18%) | |
| | CSEs or equivalent | 213 (6.1%) | 7 (4.7%) | |
| | NVQ or HND or HNC or equivalent | 235 (6.8%) | 6 (4%) | |
| | Other professional qualifications e.g., nursing, teaching | 185 (5.3%) | 11 (7.3%) | |
| | None of the above | 688 (19.9%) | 52 (34.7%) | |
| BMI | | 28.2 ± 5.4; 27.4 (15.8, 57) | 28.7 ± 6.3; 27.3 (17.1, 56.5) | 0.268 |
| Smoking Status | Never | 1637 (47.2%) | 70 (46.7%) | |
| | Previous | 1256 (36.2%) | 59 (39.3%) | |
| | Current | 572 (16.5%) | 21 (14%) | 0.649 |
| APOE e4 Carrier Status | No | 2425 (70%) | 74 (49.3%) | 2.90E-07 |
| | Yes | 1040 (30%) | 76 (50.7%) | |
| Diabetes Status | No | 3252 (93.9%) | 131 (87.3%) | 0.001 |
| | Yes | 213 (6.1%) | 19 (12.7%) | |
| Hypertension Status | No | 2401 (69.3%) | 85 (56.7%) | 4.90E-04 |
| | Yes | 1064 (30.7%) | 65 (43.3%) | |
| Antidepressant Use | No | 1741 (50.2%) | 63 (42%) | 0.038 |
| | Yes | 1724 (49.8%) | 87 (58%) | |
| UKB PPP Consortium Selection | No | 3040 (87.7%) | 87 (58%) | 6.50E-21 |
| | Yes | 425 (12.3%) | 63 (42%) | |

[1]mean ± SD; median (minimum, maximum) or frequency (percentage); [2]p-values from unadjusted Cox regression models

**Table S4 Associations between protein expression levels and incident ADRD in participants with a history of MDD at baseline, adjusting for covariates including *APOE* e4 carrier status**

| Olink ID | Assay Target | Protein panel | Gene symbol | UniProt | Gene CHROM | Gene start | Gene end | HR | Lower | Upper | Unadj P | FDR-Adj P |
|---|---|---|---|---|---|---|---|---|---|---|---|---|
| OID21247 | GFAP | Oncology | GFAP | P14136 | 17 | 44903159 | 44916937 | 1.37 | 1.20 | 1.58 | 8.08E-06 | 0.024 |
| OID20871 | NEFL | Neurology | NEFL | P07196 | 8 | 24950955 | 24956721 | 1.38 | 1.19 | 1.60 | 2.15E-05 | 0.030 |
| OID20916 | PSG1 | Neurology | PSG1 | P11464 | 19 | 42866464 | 42879822 | 1.48 | 1.23 | 1.79 | 3.07E-05 | 0.030 |
| OID30303 | GET3 | Cardiometabolic_II | GET3 | O43681 | 19 | 12737139 | 12748323 | 0.66 | 0.53 | 0.81 | 6.51E-05 | 0.038 |
| OID21266 | HPGDS | Oncology | HPGDS | O60760 | 4 | 94298535 | 94342876 | 0.71 | 0.61 | 0.84 | 6.46E-05 | 0.038 |
| OID31067 | VGF | Neurology_II | VGF | O15240 | 7 | 101162509 | 101165569 | 0.73 | 0.62 | 0.85 | 9.95E-05 | 0.048 |
| OID31364 | GLYR1 | Oncology_II | GLYR1 | Q49A26 | 16 | 4803203 | 4847288 | 0.61 | 0.47 | 0.79 | 1.93E-04 | 0.081 |
| OID31287 | CEP290 | Oncology_II | CEP290 | O15078 | 12 | 88049016 | 88142099 | 0.67 | 0.55 | 0.83 | 2.38E-04 | 0.087 |
| OID30571 | AKAP12 | Inflammation_II | AKAP12 | Q02952 | 6 | 151239967 | 151358559 | 0.74 | 0.62 | 0.87 | 5.12E-04 | 0.150 |
| OID30702 | FGL1 | Inflammation_II | FGL1 | Q08830 | 8 | 17864380 | 17910365 | 1.35 | 1.14 | 1.59 | 4.78E-04 | 0.150 |
| OID30574 | GHR | Inflammation_II | GHR | P10912 | 5 | 42423439 | 42721878 | 0.73 | 0.62 | 0.88 | 6.18E-04 | 0.164 |
| OID21479 | CPXM1 | Oncology | CPXM1 | Q96SM3 | 20 | 2794074 | 2800627 | 0.75 | 0.64 | 0.89 | 7.98E-04 | 0.179 |
| OID30604 | SERPINI1 | Inflammation_II | SERPINI1 | Q99574 | 3 | 167735243 | 167825569 | 0.75 | 0.63 | 0.89 | 7.59E-04 | 0.179 |
| OID21389 | EBI3_IL27 | Oncology | EBI3;IL27 | Q14213;Q8NEV9 | 19;16 | 4229523;28499362 | 4237528;28512051 | 1.32 | 1.12 | 1.56 | 1.02E-03 | 0.208 |
| OID20281 | SERPINA11 | Cardiometabolic | SERPINA11 | Q86U17 | 14 | 94442464 | 94452800 | 1.35 | 1.13 | 1.62 | 1.14E-03 | 0.208 |
| OID31476 | SNX18 | Oncology_II | SNX18 | Q96RF0 | 5 | 54517759 | 54546586 | 0.70 | 0.57 | 0.87 | 1.14E-03 | 0.208 |
| OID31004 | GART | Neurology_II | GART | P22102 | 21 | 33503931 | 33543491 | 0.72 | 0.59 | 0.88 | 1.56E-03 | 0.268 |
| OID30985 | CDK5RAP3 | Neurology_II | CDK5RAP3 | Q96JB5 | 17 | 47967810 | 47981781 | 0.73 | 0.60 | 0.89 | 1.73E-03 | 0.273 |
| OID31518 | DDX1 | Oncology_II | DDX1 | Q92499 | 2 | 15591178 | 15634346 | 0.71 | 0.57 | 0.88 | 1.81E-03 | 0.273 |
| OID31041 | GID8 | Neurology_II | GID8 | Q9NWU2 | 20 | 62938147 | 62948475 | 0.69 | 0.55 | 0.87 | 1.87E-03 | 0.273 |
| OID30260 | AHNAK | Cardiometabolic_II | AHNAK | Q09666 | 11 | 62433542 | 62556235 | 0.76 | 0.63 | 0.92 | 4.50E-03 | 0.288 |
| OID31117 | AP3S2 | Neurology_II | AP3S2 | P59780 | 15 | 89830599 | 89894638 | 0.77 | 0.64 | 0.92 | 3.83E-03 | 0.288 |
| OID30815 | CRYM | Neurology_II | CRYM | Q14894 | 16 | 21238874 | 21303083 | 0.75 | 0.61 | 0.91 | 3.91E-03 | 0.288 |
| OID20613 | CXCL6 | Inflammation | CXCL6 | P80162 | 4 | 73836640 | 73849064 | 0.79 | 0.67 | 0.93 | 4.15E-03 | 0.288 |
| OID21066 | DKK1 | Neurology | DKK1 | O94907 | 10 | 52314281 | 52318042 | 0.79 | 0.67 | 0.92 | 2.84E-03 | 0.288 |
| OID31040 | FGD3 | Neurology_II | FGD3 | Q5JSP0 | 9 | 92947523 | 93036236 | 0.78 | 0.66 | 0.93 | 4.36E-03 | 0.288 |
| OID30611 | GP5 | Inflammation_II | GP5 | P40197 | 3 | 194394821 | 194399266 | 0.81 | 0.70 | 0.93 | 3.74E-03 | 0.288 |
| OID30718 | HGFAC | Inflammation_II | HGFAC | Q04756 | 4 | 3441968 | 3449486 | 0.79 | 0.67 | 0.92 | 3.26E-03 | 0.288 |
| OID20232 | KYAT1 | Cardiometabolic | KYAT1 | Q16773 | 9 | 128832942 | 128882494 | 0.78 | 0.66 | 0.92 | 3.33E-03 | 0.288 |
| OID30637 | LZTFL1 | Inflammation_II | LZTFL1 | Q9NQ48 | 3 | 45823316 | 45916042 | 0.78 | 0.66 | 0.93 | 4.54E-03 | 0.288 |
| OID20588 | MGMT | Inflammation | MGMT | P16455 | 10 | 129467190 | 129770983 | 0.80 | 0.68 | 0.93 | 4.53E-03 | 0.288 |
| OID30638 | MOCS2 | Inflammation_II | MOCS2 | O96007 | 5 | 53095679 | 53110063 | 0.78 | 0.65 | 0.92 | 4.48E-03 | 0.288 |
| OID30120 | MYL1 | Cardiometabolic_II | MYL1 | P05976 | 2 | 210290150 | 210315174 | 1.16 | 1.05 | 1.28 | 4.36E-03 | 0.288 |
| OID20191 | NPTXR | Cardiometabolic | NPTXR | O95502 | 22 | 38818452 | 38844028 | 0.76 | 0.63 | 0.91 | 2.38E-03 | 0.288 |
| OID30201 | OPLAH | Cardiometabolic_II | OPLAH | O14841 | 8 | 144051266 | 144063965 | 0.77 | 0.65 | 0.91 | 2.38E-03 | 0.288 |
| OID31003 | PCBP2 | Neurology_II | PCBP2 | Q15366 | 12 | 53452102 | 53481162 | 0.79 | 0.67 | 0.93 | 4.39E-03 | 0.288 |
| OID30511 | PER3 | Inflammation_II | PER3 | P56645 | 1 | 7784291 | 7845177 | 0.77 | 0.65 | 0.92 | 2.93E-03 | 0.288 |
| OID30764 | PF4 | Inflammation_II | PF4 | P02776 | 4 | 73980811 | 73982027 | 0.78 | 0.66 | 0.92 | 3.37E-03 | 0.288 |
| OID20425 | RAB37 | Inflammation | RAB37 | Q96AX2 | 17 | 74670578 | 74747335 | 0.74 | 0.60 | 0.91 | 3.97E-03 | 0.288 |
| OID31376 | RFC4 | Oncology_II | RFC4 | P35249 | 3 | 186789880 | 186807058 | 0.75 | 0.62 | 0.90 | 2.53E-03 | 0.288 |
| OID20342 | SERPINE1 | Cardiometabolic | SERPINE1 | P05121 | 7 | 101127104 | 101139247 | 0.79 | 0.67 | 0.92 | 2.95E-03 | 0.288 |
| OID30215 | SNX5 | Cardiometabolic_II | SNX5 | Q9Y5X3 | 20 | 17941597 | 17968980 | 0.75 | 0.62 | 0.91 | 3.49E-03 | 0.288 |
| OID31216 | SSH3 | Oncology_II | SSH3 | Q8TE77 | 11 | 67303478 | 67312607 | 0.76 | 0.63 | 0.91 | 3.67E-03 | 0.288 |
| OID30682 | TREML1 | Inflammation_II | TREML1 | Q86YW5 | 6 | 41149337 | 41154347 | 0.77 | 0.65 | 0.92 | 2.90E-03 | 0.288 |
| OID21524 | VEGFC | Oncology | VEGFC | P49767 | 4 | 176683538 | 176792922 | 0.79 | 0.68 | 0.92 | 2.85E-03 | 0.288 |
| OID31350 | ZNF830 | Oncology_II | ZNF830 | Q96NB3 | 17 | 34961540 | 34963777 | 0.71 | 0.57 | 0.90 | 3.82E-03 | 0.288 |
| OID20117 | ADH4 | Cardiometabolic | ADH4 | P08319 | 4 | 99123657 | 99157792 | 0.76 | 0.63 | 0.92 | 4.67E-03 | 0.290 |
| OID31179 | VCPKMT | Oncology_II | VCPKMT | Q9H867 | 14 | 50108632 | 50116600 | 0.72 | 0.57 | 0.90 | 5.08E-03 | 0.309 |
| OID30612 | ADH1B | Inflammation_II | ADH1B | P00325 | 4 | 99304971 | 99352760 | 0.78 | 0.65 | 0.94 | 1.01E-02 | 0.309 |
| OID30706 | AHSG | Inflammation_II | AHSG | P02765 | 3 | 186613060 | 186621318 | 0.79 | 0.67 | 0.94 | 7.21E-03 | 0.309 |
| OID20740 | ANGPT1 | Inflammation | ANGPT1 | Q15389 | 8 | 107249482 | 107498055 | 0.82 | 0.71 | 0.96 | 1.09E-02 | 0.309 |
| OID21070 | APP | Neurology | APP | P05067 | 21 | 25880550 | 26171128 | 0.81 | 0.69 | 0.95 | 1.14E-02 | 0.309 |
| OID31259 | BRD1 | Oncology_II | BRD1 | O95696 | 22 | 49773283 | 49827873 | 0.78 | 0.65 | 0.94 | 1.07E-02 | 0.309 |
| OID31095 | C7orf50 | Neurology_II | C7orf50 | Q9BRJ6 | 7 | 996986 | 1138260 | 0.65 | 0.48 | 0.90 | 8.99E-03 | 0.309 |
| OID20709 | CCN2 | Inflammation | CCN2 | P29279 | 6 | 131948176 | 131951372 | 0.79 | 0.67 | 0.94 | 6.84E-03 | 0.309 |
| OID31076 | CDAN1 | Neurology_II | CDAN1 | Q8IWY9 | 15 | 42723544 | 42737128 | 0.76 | 0.63 | 0.93 | 6.66E-03 | 0.309 |
| OID30894 | CGN | Neurology_II | CGN | Q9P2M7 | 1 | 151510510 | 151538692 | 0.77 | 0.63 | 0.94 | 9.08E-03 | 0.309 |
| OID30957 | CLGN | Neurology_II | CLGN | O14967 | 4 | 140388453 | 140427661 | 0.78 | 0.65 | 0.94 | 7.83E-03 | 0.309 |
| OID20347 | CNDP1 | Cardiometabolic | CNDP1 | Q96KN2 | 18 | 74534500 | 74587212 | 0.82 | 0.71 | 0.95 | 8.88E-03 | 0.309 |
| OID30546 | DGKA | Inflammation_II | DGKA | P23743 | 12 | 55927319 | 55954027 | 0.79 | 0.67 | 0.94 | 8.09E-03 | 0.309 |
| OID31434 | DNAJB14 | Oncology_II | DNAJB14 | Q8TBM8 | 4 | 99896248 | 99946618 | 0.79 | 0.66 | 0.94 | 8.47E-03 | 0.309 |
| OID31010 | EDF1 | Neurology_II | EDF1 | O60869 | 9 | 136862119 | 136866308 | 0.76 | 0.62 | 0.93 | 8.98E-03 | 0.309 |
| OID20905 | EREG | Neurology | EREG | O14944 | 4 | 74365145 | 74388749 | 0.79 | 0.67 | 0.94 | 7.87E-03 | 0.309 |
| OID31317 | ERI1 | Oncology_II | ERI1 | Q8IV48 | 8 | 9002147 | 9116746 | 0.77 | 0.63 | 0.94 | 1.05E-02 | 0.309 |
| OID21256 | FUS | Oncology | FUS | P35637 | 16 | 31180138 | 31191605 | 0.76 | 0.63 | 0.93 | 6.09E-03 | 0.309 |
| OID21423 | GALNT2 | Oncology | GALNT2 | Q10471 | 1 | 230057990 | 230282122 | 0.81 | 0.69 | 0.95 | 1.07E-02 | 0.309 |
| OID20251 | GDF15 | Cardiometabolic | GDF15 | Q99988 | 19 | 18374731 | 18389176 | 1.25 | 1.05 | 1.49 | 1.14E-02 | 0.309 |
| OID31438 | GORASP2 | Oncology_II | GORASP2 | Q9H8Y8 | 2 | 170928464 | 170967130 | 0.79 | 0.67 | 0.94 | 8.84E-03 | 0.309 |
| OID20166 | GSTA1 | Cardiometabolic | GSTA1 | P08263 | 6 | 52791371 | 52803860 | 0.77 | 0.64 | 0.94 | 9.62E-03 | 0.309 |
| OID31283 | IFIT3 | Oncology_II | IFIT3 | O14879 | 10 | 89327307 | 89377473 | 0.79 | 0.66 | 0.95 | 1.13E-02 | 0.309 |
| OID20155 | IGFBPL1 | Cardiometabolic | IGFBPL1 | Q8WX77 | 9 | 38406528 | 38424454 | 1.22 | 1.06 | 1.42 | 7.30E-03 | 0.309 |
| OID30806 | KIRREL1 | Neurology_II | KIRREL1 | Q96J84 | 1 | 157993273 | 158100262 | 0.75 | 0.60 | 0.94 | 1.12E-02 | 0.309 |
| OID20438 | LRRN1 | Inflammation | LRRN1 | Q6UXK5 | 3 | 3799431 | 3849834 | 0.80 | 0.69 | 0.94 | 5.19E-03 | 0.309 |
| OID21530 | LYPD3 | Oncology | LYPD3 | O95274 | 19 | 43460787 | 43465608 | 1.22 | 1.05 | 1.42 | 8.23E-03 | 0.309 |
| OID20275 | MSMB | Cardiometabolic | MSMB | P08118 | 10 | 46033307 | 46048180 | 1.25 | 1.05 | 1.49 | 1.05E-02 | 0.309 |
| OID21212 | NFKBIE | Oncology | NFKBIE | O00221 | 6 | 44258166 | 44265788 | 0.79 | 0.66 | 0.94 | 9.47E-03 | 0.309 |
| OID30252 | NIT1 | Cardiometabolic_II | NIT1 | Q86X76 | 1 | 161118086 | 161125445 | 0.82 | 0.70 | 0.95 | 7.47E-03 | 0.309 |
| OID30390 | NPTX2 | Cardiometabolic_II | NPTX2 | P47972 | 7 | 98617285 | 98629869 | 1.28 | 1.07 | 1.54 | 7.83E-03 | 0.309 |
| OID31301 | PARD3 | Oncology_II | PARD3 | Q8TEW0 | 10 | 34109560 | 34815325 | 0.79 | 0.65 | 0.95 | 1.15E-02 | 0.309 |
| OID20313 | PDGFA | Cardiometabolic | PDGFA | P04085 | 7 | 497258 | 520296 | 0.81 | 0.70 | 0.95 | 9.83E-03 | 0.309 |
| OID21183 | PDGFC | Oncology | PDGFC | Q9NRA1 | 4 | 156760454 | 156971799 | 0.81 | 0.70 | 0.95 | 9.55E-03 | 0.309 |
| OID30666 | PENK | Inflammation_II | PENK | P01210 | 8 | 56436674 | 56446671 | 0.79 | 0.67 | 0.94 | 7.14E-03 | 0.309 |
| OID31167 | PHLDB2 | Oncology_II | PHLDB2 | Q86SQ0 | 3 | 111732497 | 111976517 | 0.77 | 0.63 | 0.92 | 5.62E-03 | 0.309 |
| OID30634 | PHYKPL | Inflammation_II | PHYKPL | Q8IUZ5 | 5 | 178208471 | 178232802 | 0.79 | 0.67 | 0.95 | 9.54E-03 | 0.309 |
| OID20144 | PLPBP | Cardiometabolic | PLPBP | O94903 | 8 | 37762595 | 37779768 | 0.81 | 0.68 | 0.95 | 1.14E-02 | 0.309 |
| OID30761 | PPBP | Inflammation_II | PPBP | P02775 | 4 | 73986439 | 73988190 | 0.80 | 0.68 | 0.94 | 6.40E-03 | 0.309 |
| OID21172 | PQBP1 | Oncology | PQBP1 | O60828 | X | 48890197 | 48903402 | 0.72 | 0.58 | 0.91 | 6.27E-03 | 0.309 |
| OID21169 | PSRC1 | Oncology | PSRC1 | Q6PGN9 | 1 | 109279556 | 109283186 | 0.77 | 0.65 | 0.93 | 5.52E-03 | 0.309 |
| OID20570 | PTX3 | Inflammation | PTX3 | P26022 | 3 | 157436850 | 157443633 | 1.31 | 1.08 | 1.58 | 6.55E-03 | 0.309 |
| OID31254 | RAD51 | Oncology_II | RAD51 | Q06609 | 15 | 40694774 | 40732340 | 0.75 | 0.60 | 0.93 | 8.52E-03 | 0.309 |
| OID31337 | RAPGEF2 | Oncology_II | RAPGEF2 | Q9Y4G8 | 4 | 159103013 | 159360174 | 0.79 | 0.66 | 0.94 | 8.80E-03 | 0.309 |
| OID31236 | RBM25 | Oncology_II | RBM25 | P49756 | 14 | 73058532 | 73123899 | 0.80 | 0.67 | 0.95 | 1.10E-02 | 0.309 |
| OID31060 | RGCC | Neurology_II | RGCC | Q9H4X1 | 13 | 41457550 | 41470871 | 0.80 | 0.68 | 0.94 | 7.30E-03 | 0.309 |
| OID30284 | RPL14 | Cardiometabolic_II | RPL14 | P50914 | 3 | 40457292 | 40468587 | 0.77 | 0.64 | 0.93 | 6.63E-03 | 0.309 |
| OID30961 | RRAS | Neurology_II | RRAS | P10301 | 19 | 49635292 | 49640143 | 0.80 | 0.67 | 0.95 | 1.15E-02 | 0.309 |
| OID31012 | SAFB2 | Neurology_II | SAFB2 | Q14151 | 19 | 5586999 | 5624046 | 0.78 | 0.65 | 0.94 | 9.90E-03 | 0.309 |
| OID30763 | SERPINA5 | Inflammation_II | SERPINA5 | P05154 | 14 | 94561442 | 94593118 | 0.78 | 0.65 | 0.94 | 1.09E-02 | 0.309 |
| OID30359 | SERPINE2 | Cardiometabolic_II | SERPINE2 | P07093 | 2 | 223975045 | 224039318 | 0.80 | 0.68 | 0.94 | 8.43E-03 | 0.309 |
| OID20539 | SLC39A5 | Inflammation | SLC39A5 | Q6ZMH5 | 12 | 56230049 | 56237846 | 0.77 | 0.65 | 0.93 | 5.91E-03 | 0.309 |
| OID30853 | SMS | Neurology_II | SMS | P52788 | X | 21940709 | 21994837 | 0.77 | 0.64 | 0.94 | 8.69E-03 | 0.309 |
| OID21511 | SPARC | Oncology | SPARC | P09486 | 5 | 151661096 | 151686975 | 0.81 | 0.69 | 0.95 | 8.29E-03 | 0.309 |

| | | | | | | | | | | | | |
|---|---|---|---|---|---|---|---|---|---|---|---|---|
| OID21447 | TACSTD2 | Oncology | TACSTD2 | P09758 | 1 | 58575433 | 58577252 | 1.26 | 1.07 | 1.49 | 6.00E-03 | 0.309 |
| OID31417 | TGFB2 | Oncology_II | TGFB2 | P61812 | 1 | 218345336 | 218444619 | 0.78 | 0.66 | 0.94 | 7.48E-03 | 0.309 |
| OID31375 | THTPA | Oncology_II | THTPA | Q9BU02 | 14 | 23555988 | 23560271 | 0.80 | 0.68 | 0.94 | 8.00E-03 | 0.309 |
| OID31320 | TIGIT | Oncology_II | TIGIT | Q495A1 | 3 | 114276913 | 114310288 | 0.77 | 0.64 | 0.93 | 5.46E-03 | 0.309 |
| OID21147 | TIMP4 | Neurology | TIMP4 | Q99727 | 3 | 12153068 | 12158912 | 1.22 | 1.05 | 1.42 | 1.13E-02 | 0.309 |
| OID30128 | TRPV3 | Cardiometabolic_II | TRPV3 | Q8NET8 | 17 | 3510502 | 3557812 | 1.21 | 1.05 | 1.38 | 6.68E-03 | 0.309 |
| OID30193 | VIPR1 | Cardiometabolic_II | VIPR1 | P32241 | 3 | 42489299 | 42537573 | 0.78 | 0.64 | 0.95 | 1.15E-02 | 0.309 |
| OID31404 | YJU2 | Oncology_II | YJU2 | Q9BW85 | 19 | 4247080 | 4269088 | 0.78 | 0.65 | 0.94 | 1.08E-02 | 0.309 |
| OID20703 | ANGPTL4 | Inflammation | ANGPTL4 | Q9BY76 | 19 | 8363289 | 8374370 | 1.25 | 1.05 | 1.48 | 1.21E-02 | 0.311 |
| OID31525 | CETN3 | Oncology_II | CETN3 | O15182 | 5 | 90392257 | 90409766 | 0.81 | 0.68 | 0.95 | 1.21E-02 | 0.311 |
| OID21046 | ENO2 | Neurology | ENO2 | P09104 | 12 | 6913745 | 6923698 | 0.81 | 0.68 | 0.95 | 1.22E-02 | 0.311 |
| OID20838 | GGA1 | Neurology | GGA1 | Q9UJY5 | 22 | 37608725 | 37633564 | 0.76 | 0.61 | 0.94 | 1.20E-02 | 0.311 |
| OID30289 | GRHPR | Cardiometabolic_II | GRHPR | Q9UBQ7 | 9 | 37422666 | 37436990 | 0.81 | 0.69 | 0.95 | 1.20E-02 | 0.311 |
| OID30901 | NRXN3 | Neurology_II | NRXN3 | Q9Y4C0 | 14 | 78170373 | 79868291 | 0.77 | 0.63 | 0.94 | 1.22E-02 | 0.311 |
| OID30877 | ADGRV1 | Neurology_II | ADGRV1 | Q8WXG9 | 5 | 90529344 | 91164437 | 0.79 | 0.65 | 0.96 | 1.88E-02 | 0.312 |
| OID31142 | AK2 | Neurology_II | AK2 | P54819 | 1 | 33007986 | 33080996 | 0.82 | 0.70 | 0.97 | 1.84E-02 | 0.312 |
| OID20895 | AKT1S1 | Neurology | AKT1S1 | Q96B36 | 19 | 49869033 | 49878459 | 0.81 | 0.69 | 0.96 | 1.27E-02 | 0.312 |
| OID31052 | AKT2 | Neurology_II | AKT2 | P31751 | 19 | 40230317 | 40285536 | 0.82 | 0.70 | 0.96 | 1.43E-02 | 0.312 |
| OID20211 | ANGPTL1 | Cardiometabolic | ANGPTL1 | O95841 | 1 | 178849535 | 178871077 | 1.20 | 1.02 | 1.42 | 2.90E-02 | 0.312 |
| OID30673 | APOB | Inflammation_II | APOB | P04114 | 2 | 21001429 | 21044073 | 0.84 | 0.72 | 0.98 | 2.49E-02 | 0.312 |
| OID31024 | ARFIP1 | Neurology_II | ARFIP1 | P53367 | 4 | 152779937 | 152918463 | 0.81 | 0.68 | 0.97 | 1.84E-02 | 0.312 |
| OID31339 | ARG2 | Oncology_II | ARG2 | P78540 | 14 | 67619920 | 67651708 | 0.80 | 0.66 | 0.97 | 2.39E-02 | 0.312 |
| OID30653 | ARHGAP45 | Inflammation_II | ARHGAP45 | Q92619 | 19 | 1065923 | 1086628 | 0.83 | 0.71 | 0.99 | 3.41E-02 | 0.312 |
| OID31133 | ARHGEF1 | Neurology_II | ARHGEF1 | Q92888 | 19 | 41883173 | 41930150 | 0.84 | 0.72 | 0.98 | 2.73E-02 | 0.312 |
| OID30861 | ARID3A | Neurology_II | ARID3A | Q99856 | 19 | 925781 | 975939 | 0.80 | 0.67 | 0.96 | 1.69E-02 | 0.312 |
| OID30168 | ARNTL | Cardiometabolic_II | ARNTL | O00327 | 11 | 13276652 | 13387266 | 1.17 | 1.03 | 1.34 | 1.93E-02 | 0.312 |
| OID30283 | ASRGL1 | Cardiometabolic_II | ASRGL1 | Q7L266 | 11 | 62337448 | 62393412 | 0.83 | 0.71 | 0.97 | 1.77E-02 | 0.312 |
| OID31466 | ATG16L1 | Oncology_II | ATG16L1 | Q676U5 | 2 | 233210051 | 233295674 | 0.83 | 0.70 | 0.98 | 2.96E-02 | 0.312 |
| OID20843 | ATP6V1F | Neurology | ATP6V1F | Q16864 | 7 | 128862856 | 128865847 | 0.80 | 0.67 | 0.96 | 1.68E-02 | 0.312 |
| OID30242 | ATP6V1G1 | Cardiometabolic_II | ATP6V1G1 | O75348 | 9 | 114587706 | 114598915 | 0.81 | 0.68 | 0.96 | 1.55E-02 | 0.312 |
| OID31149 | ATXN3 | Neurology_II | ATXN3 | P54252 | 14 | 92044496 | 92106621 | 0.82 | 0.69 | 0.97 | 1.96E-02 | 0.312 |
| OID20998 | BCAN | Neurology | BCAN | Q96GW7 | 1 | 156641390 | 156659532 | 0.82 | 0.70 | 0.97 | 1.96E-02 | 0.312 |
| OID31421 | BCL7B | Oncology_II | BCL7B | Q9BQE9 | 7 | 73536356 | 73557690 | 0.79 | 0.65 | 0.97 | 2.31E-02 | 0.312 |
| OID30373 | BDNF | Cardiometabolic_II | BDNF | P23560 | 11 | 27654893 | 27722058 | 0.83 | 0.71 | 0.97 | 1.97E-02 | 0.312 |
| OID30220 | BECN1 | Cardiometabolic_II | BECN1 | Q14457 | 17 | 42810134 | 42833350 | 0.81 | 0.68 | 0.97 | 2.19E-02 | 0.312 |
| OID21198 | BGN | Oncology | BGN | P21810 | X | 153494980 | 153509546 | 1.22 | 1.04 | 1.43 | 1.69E-02 | 0.312 |
| OID30541 | BLNK | Inflammation_II | BLNK | Q8WV28 | 10 | 96189171 | 96271587 | 0.79 | 0.64 | 0.98 | 2.92E-02 | 0.312 |
| OID31106 | BLOC1S3 | Neurology_II | BLOC1S3 | Q6QNY0 | 19 | 45178784 | 45216933 | 0.81 | 0.68 | 0.98 | 2.61E-02 | 0.312 |
| OID31506 | BRAP | Oncology_II | BRAP | Q7Z569 | 12 | 111642146 | 111685956 | 0.84 | 0.71 | 0.98 | 2.66E-02 | 0.312 |
| OID31049 | C1QTNF6 | Neurology_II | C1QTNF6 | Q9BXI9 | 22 | 37180166 | 37199385 | 0.81 | 0.67 | 0.97 | 2.42E-02 | 0.312 |
| OID30766 | C7 | Inflammation_II | C7 | P10643 | 5 | 40909492 | 40984643 | 1.21 | 1.02 | 1.43 | 2.55E-02 | 0.312 |
| OID21096 | CA6 | Neurology | CA6 | P23280 | 1 | 8945867 | 8975092 | 0.83 | 0.69 | 0.99 | 3.33E-02 | 0.312 |
| OID30649 | CACYBP | Inflammation_II | CACYBP | Q9HB71 | 1 | 174999163 | 175012027 | 0.84 | 0.72 | 0.98 | 3.01E-02 | 0.312 |
| OID20181 | CANT1 | Cardiometabolic | CANT1 | Q8WVQ1 | 17 | 78991716 | 79009867 | 0.81 | 0.69 | 0.96 | 1.34E-02 | 0.312 |
| OID30921 | CASC3 | Neurology_II | CASC3 | O15234 | 17 | 40140318 | 40172171 | 0.83 | 0.69 | 0.99 | 3.40E-02 | 0.312 |
| OID20305 | CASP3 | Cardiometabolic | CASP3 | P42574 | 4 | 184627696 | 184649509 | 0.84 | 0.71 | 0.98 | 2.72E-02 | 0.312 |
| OID30530 | CASP9 | Inflammation_II | CASP9 | P55211 | 1 | 15490832 | 15526534 | 0.82 | 0.69 | 0.98 | 2.88E-02 | 0.312 |
| OID20179 | CBLIF | Cardiometabolic | CBLIF | P27352 | 11 | 59829273 | 59845499 | 0.86 | 0.75 | 0.99 | 3.42E-02 | 0.312 |
| OID30200 | CBS | Cardiometabolic_II | CBS | P35520 | 21 | 43053191 | 43076943 | 0.81 | 0.68 | 0.97 | 2.31E-02 | 0.312 |
| OID31077 | CCAR2 | Neurology_II | CCAR2 | Q8N163 | 8 | 22604757 | 22620964 | 0.77 | 0.62 | 0.97 | 2.93E-02 | 0.312 |
| OID20655 | CCL13 | Inflammation | CCL13 | Q99616 | 17 | 34356480 | 34358610 | 0.83 | 0.71 | 0.98 | 2.67E-02 | 0.312 |
| OID20412 | CCL5 | Cardiometabolic | CCL5 | P13501 | 17 | 35871491 | 35880793 | 0.82 | 0.70 | 0.96 | 1.60E-02 | 0.312 |
| OID21298 | CD207 | Oncology | CD207 | Q9UJ71 | 2 | 70830211 | 70835816 | 1.20 | 1.02 | 1.42 | 2.80E-02 | 0.312 |
| OID20680 | CD276 | Inflammation | CD276 | Q5ZPR3 | 15 | 73683966 | 73714514 | 1.19 | 1.01 | 1.40 | 3.48E-02 | 0.312 |
| OID20616 | CD40LG | Inflammation | CD40LG | P29965 | X | 136648158 | 136660390 | 0.83 | 0.71 | 0.97 | 2.02E-02 | 0.312 |
| OID20649 | CD6 | Inflammation | CD6 | P30203 | 11 | 60971680 | 61020377 | 0.85 | 0.72 | 0.99 | 3.40E-02 | 0.312 |
| OID20988 | CD63 | Neurology | CD63 | P08962 | 12 | 55725323 | 55729707 | 0.83 | 0.71 | 0.98 | 2.72E-02 | 0.312 |
| OID20836 | CETN2 | Neurology | CETN2 | P41208 | X | 152826994 | 152830757 | 0.81 | 0.68 | 0.96 | 1.59E-02 | 0.312 |
| OID30972 | CHM | Neurology_II | CHM | P24386 | X | 85861180 | 86047561 | 0.81 | 0.69 | 0.97 | 1.85E-02 | 0.312 |
| OID21501 | CIAPIN1 | Oncology | CIAPIN1 | Q6FI81 | 16 | 57428187 | 57447420 | 0.83 | 0.70 | 0.98 | 2.43E-02 | 0.312 |
| OID31503 | CIRBP | Oncology_II | CIRBP | Q14011 | 19 | 1259384 | 1274880 | 0.83 | 0.71 | 0.98 | 2.47E-02 | 0.312 |
| OID20831 | CLSPN | Neurology | CLSPN | Q9HAW4 | 1 | 35720218 | 35769978 | 0.80 | 0.65 | 0.98 | 3.47E-02 | 0.312 |
| OID30285 | CNP | Cardiometabolic_II | CNP | P09543 | 17 | 41966763 | 41977740 | 0.84 | 0.71 | 0.98 | 3.05E-02 | 0.312 |
| OID30305 | COCH | Cardiometabolic_II | COCH | O43405 | 14 | 30874514 | 30895065 | 0.83 | 0.70 | 0.98 | 2.51E-02 | 0.312 |
| OID20742 | CRKL | Inflammation | CRKL | P46109 | 22 | 20917407 | 20953747 | 0.84 | 0.71 | 0.98 | 2.60E-02 | 0.312 |
| OID31081 | CRYBB2 | Neurology_II | CRYBB2 | P43320 | 22 | 25212564 | 25231870 | 1.15 | 1.03 | 1.29 | 1.52E-02 | 0.312 |
| OID30183 | CSDE1 | Cardiometabolic_II | CSDE1 | O75534 | 1 | 114716913 | 114758676 | 0.82 | 0.68 | 0.98 | 3.24E-02 | 0.312 |
| OID20228 | CTSL | Cardiometabolic | CTSL | P07711 | 9 | 87724051 | 87731469 | 1.20 | 1.04 | 1.39 | 1.50E-02 | 0.312 |
| OID30664 | DAAM1 | Inflammation_II | DAAM1 | Q9Y4D1 | 14 | 59188646 | 59371405 | 0.84 | 0.71 | 0.99 | 3.32E-02 | 0.312 |
| OID30657 | DCTD | Inflammation_II | DCTD | P32321 | 4 | 182890060 | 182917936 | 0.83 | 0.71 | 0.97 | 2.06E-02 | 0.312 |
| OID30493 | DENR | Inflammation_II | DENR | O43583 | 12 | 122752824 | 122771064 | 0.81 | 0.67 | 0.97 | 2.37E-02 | 0.312 |
| OID20620 | DFFA | Inflammation | DFFA | O00273 | 1 | 10456522 | 10472529 | 0.83 | 0.70 | 0.98 | 3.00E-02 | 0.312 |
| OID31071 | DLG4 | Neurology_II | DLG4 | P78352 | 17 | 7187187 | 7219841 | 0.82 | 0.68 | 0.97 | 2.24E-02 | 0.312 |
| OID30661 | DNAJB2 | Inflammation_II | DNAJB2 | P25686 | 2 | 219279342 | 219286898 | 0.83 | 0.70 | 0.99 | 3.47E-02 | 0.312 |
| OID30613 | DNAJB6 | Inflammation_II | DNAJB6 | O75190 | 7 | 157335381 | 157417439 | 0.82 | 0.69 | 0.96 | 1.63E-02 | 0.312 |
| OID30580 | DTD1 | Inflammation_II | DTD1 | Q8TEA8 | 20 | 18587942 | 18766644 | 0.81 | 0.68 | 0.95 | 1.25E-02 | 0.312 |
| OID20827 | DUSP3 | Neurology | DUSP3 | P51452 | 17 | 43766125 | 43778977 | 0.80 | 0.67 | 0.97 | 2.24E-02 | 0.312 |
| OID31415 | DYNLT3 | Oncology_II | DYNLT3 | P51808 | X | 37836757 | 37847571 | 0.82 | 0.69 | 0.98 | 2.74E-02 | 0.312 |
| OID31442 | EIF2AK2 | Oncology_II | EIF2AK2 | P19525 | 2 | 37099210 | 37157522 | 0.82 | 0.69 | 0.97 | 2.22E-02 | 0.312 |
| OID30270 | EIF2S2 | Cardiometabolic_II | EIF2S2 | P20042 | 20 | 34088309 | 34112243 | 0.80 | 0.66 | 0.97 | 2.46E-02 | 0.312 |
| OID30083 | EXOSC10 | Cardiometabolic_II | EXOSC10 | Q01780 | 1 | 11066618 | 11099869 | 0.76 | 0.60 | 0.95 | 1.51E-02 | 0.312 |
| OID30248 | FAM172A | Cardiometabolic_II | FAM172A | Q8WUF8 | 5 | 93617725 | 94111699 | 0.83 | 0.70 | 0.98 | 2.83E-02 | 0.312 |
| OID30378 | FBLN2 | Cardiometabolic_II | FBLN2 | P98095 | 3 | 13549125 | 13638422 | 1.20 | 1.02 | 1.40 | 2.78E-02 | 0.312 |
| OID21215 | FCGR2B | Oncology | FCGR2B | P31994 | 1 | 161663143 | 161678654 | 0.82 | 0.69 | 0.98 | 2.89E-02 | 0.312 |
| OID30690 | FCN1 | Inflammation_II | FCN1 | O00602 | 9 | 134903232 | 134917912 | 0.85 | 0.73 | 0.99 | 3.24E-02 | 0.312 |
| OID30395 | FGFBP2 | Cardiometabolic_II | FGFBP2 | Q9BYJ0 | 4 | 15960245 | 15969309 | 0.84 | 0.71 | 0.99 | 3.28E-02 | 0.312 |
| OID21271 | FOXO3 | Oncology | FOXO3 | O43524 | 6 | 108559835 | 108684774 | 0.82 | 0.69 | 0.98 | 3.24E-02 | 0.312 |
| OID20861 | GDNF | Neurology | GDNF | P39905 | 5 | 37812677 | 37840041 | 1.19 | 1.02 | 1.39 | 2.48E-02 | 0.312 |
| OID31062 | GIGYF2 | Neurology_II | GIGYF2 | Q6Y7W6 | 2 | 232697299 | 232860605 | 0.82 | 0.69 | 0.98 | 2.95E-02 | 0.312 |
| OID30639 | GIMAP7 | Inflammation_II | GIMAP7 | Q8NHV1 | 7 | 150514872 | 150521073 | 0.80 | 0.66 | 0.97 | 2.41E-02 | 0.312 |
| OID30594 | GLA | Inflammation_II | GLA | P06280 | X | 101393273 | 101408012 | 0.84 | 0.71 | 0.98 | 3.01E-02 | 0.312 |
| OID30340 | GMFG | Cardiometabolic_II | GMFG | O60234 | 19 | 39328353 | 39342372 | 0.82 | 0.70 | 0.97 | 1.73E-02 | 0.312 |
| OID30623 | GMPR2 | Inflammation_II | GMPR2 | Q9P2T1 | 14 | 24232422 | 24239242 | 0.83 | 0.70 | 0.98 | 2.54E-02 | 0.312 |
| OID30597 | GNPDA2 | Inflammation_II | GNPDA2 | Q8TDQ7 | 4 | 44682200 | 44726588 | 0.82 | 0.69 | 0.96 | 1.37E-02 | 0.312 |
| OID20192 | GRAP2 | Cardiometabolic | GRAP2 | O75791 | 22 | 39901084 | 39973721 | 0.82 | 0.69 | 0.96 | 1.31E-02 | 0.312 |
| OID30264 | GRP | Cardiometabolic_II | GRP | P07492 | 18 | 59220158 | 59230774 | 0.84 | 0.72 | 0.98 | 3.12E-02 | 0.312 |
| OID30700 | GSR | Inflammation_II | GSR | P00390 | 8 | 30678066 | 30727846 | 0.81 | 0.69 | 0.97 | 1.81E-02 | 0.312 |
| OID30946 | GTPBP2 | Neurology_II | GTPBP2 | Q9BX10 | 6 | 43605316 | 43629264 | 0.82 | 0.69 | 0.98 | 2.63E-02 | 0.312 |
| OID20663 | GZMA | Inflammation | GZMA | P12544 | 5 | 55102646 | 55110252 | 0.83 | 0.71 | 0.99 | 3.30E-02 | 0.312 |
| OID21422 | HAVCR1 | Oncology | HAVCR1 | Q96D42 | 5 | 157029413 | 157059119 | 1.24 | 1.04 | 1.46 | 1.45E-02 | 0.312 |
| OID31136 | HDGFL2 | Neurology_II | HDGFL2 | Q7Z4V5 | 19 | 4472297 | 4502208 | 0.81 | 0.67 | 0.97 | 2.16E-02 | 0.312 |
| OID31107 | HHEX | Neurology_II | HHEX | Q03014 | 10 | 92689955 | 92695647 | 0.83 | 0.70 | 0.98 | 2.97E-02 | 0.312 |
| OID30214 | HMGCL | Cardiometabolic_II | HMGCL | P35914 | 1 | 23801885 | 23838620 | 0.82 | 0.69 | 0.97 | 2.44E-02 | 0.312 |

| | | | | | | | | | | | | |
|---|---|---|---|---|---|---|---|---|---|---|---|---|
| OID31424 | HSPA2 | Oncology_II | HSPA2 | P54652 | 14 | 64535905 | 64546173 | 0.80 | 0.66 | 0.97 | 2.05E-02 | 0.312 |
| OID30962 | IFT20 | Neurology_II | IFT20 | Q8IY31 | 17 | 28328325 | 28335489 | 0.82 | 0.69 | 0.98 | 3.26E-02 | 0.312 |
| OID20325 | IGFBP2 | Cardiometabolic | IGFBP2 | P18065 | 2 | 216632828 | 216664436 | 1.22 | 1.02 | 1.47 | 3.15E-02 | 0.312 |
| OID30123 | IGHMBP2 | Cardiometabolic_II | IGHMBP2 | P38935 | 11 | 68903863 | 68940602 | 0.80 | 0.67 | 0.97 | 2.10E-02 | 0.312 |
| OID20562 | IL15 | Inflammation | IL15 | P40933 | 4 | 141636583 | 141733987 | 1.19 | 1.04 | 1.36 | 1.43E-02 | 0.312 |
| OID20633 | IL16 | Inflammation | IL16 | Q14005 | 15 | 81159575 | 81314058 | 0.80 | 0.66 | 0.96 | 1.69E-02 | 0.312 |
| OID31407 | IL22 | Oncology_II | IL22 | Q9GZX6 | 12 | 68248242 | 68253604 | 1.19 | 1.03 | 1.37 | 2.20E-02 | 0.312 |
| OID31112 | IMPACT | Neurology_II | IMPACT | Q9P2X3 | 18 | 24426634 | 24453531 | 0.84 | 0.71 | 0.98 | 3.21E-02 | 0.312 |
| OID30259 | INPP5D | Cardiometabolic_II | INPP5D | Q92835 | 2 | 233059967 | 233207903 | 0.81 | 0.68 | 0.96 | 1.49E-02 | 0.312 |
| OID20577 | IRAK4 | Inflammation | IRAK4 | Q9NWZ3 | 12 | 43758944 | 43789543 | 0.84 | 0.72 | 0.98 | 3.06E-02 | 0.312 |
| OID31497 | IST1 | Oncology_II | IST1 | P53990 | 16 | 71845996 | 71931199 | 0.83 | 0.71 | 0.97 | 2.17E-02 | 0.312 |
| OID20504 | ITM2A | Inflammation | ITM2A | O43736 | X | 79360384 | 79367667 | 1.20 | 1.02 | 1.40 | 2.52E-02 | 0.312 |
| OID30586 | JAM3 | Inflammation_II | JAM3 | Q9BX67 | 11 | 134069071 | 134152001 | 0.82 | 0.69 | 0.98 | 2.87E-02 | 0.312 |
| OID31520 | JPT2 | Oncology_II | JPT2 | Q9H910 | 16 | 1678256 | 1702280 | 0.82 | 0.70 | 0.96 | 1.43E-02 | 0.312 |
| OID31148 | KCTD5 | Neurology_II | KCTD5 | Q9NXV2 | 16 | 2682523 | 2709030 | 0.77 | 0.63 | 0.95 | 1.29E-02 | 0.312 |
| OID30241 | KHK | Cardiometabolic_II | KHK | P50053 | 2 | 27086747 | 27100762 | 0.81 | 0.68 | 0.96 | 1.27E-02 | 0.312 |
| OID21391 | KLK4 | Oncology | KLK4 | Q9Y5K2 | 19 | 50906351 | 50911395 | 1.19 | 1.02 | 1.40 | 2.92E-02 | 0.312 |
| OID30464 | KLRF1 | Inflammation_II | KLRF1 | Q9NZS2 | 12 | 9827481 | 9845007 | 1.17 | 1.03 | 1.35 | 2.03E-02 | 0.312 |
| OID20800 | KRT14 | Neurology | KRT14 | P02533 | 17 | 41582279 | 41586895 | 1.17 | 1.03 | 1.32 | 1.43E-02 | 0.312 |
| OID20150 | LACTB2 | Cardiometabolic | LACTB2 | Q53H82 | 8 | 70635318 | 70669185 | 0.83 | 0.70 | 0.98 | 2.67E-02 | 0.312 |
| OID20640 | LAT | Inflammation | LAT | O43561 | 16 | 28984826 | 28990784 | 0.83 | 0.71 | 0.97 | 1.87E-02 | 0.312 |
| OID21292 | LAT2 | Oncology | LAT2 | Q9GZY6 | 7 | 74199652 | 74229834 | 0.83 | 0.71 | 0.99 | 3.33E-02 | 0.312 |
| OID31068 | LDLRAP1 | Neurology_II | LDLRAP1 | Q5SW96 | 1 | 25543606 | 25568886 | 0.84 | 0.71 | 0.98 | 3.06E-02 | 0.312 |
| OID20154 | LGALS1 | Cardiometabolic | LGALS1 | P09382 | 22 | 37675636 | 37679802 | 0.82 | 0.68 | 0.98 | 3.25E-02 | 0.312 |
| OID20606 | LIFR | Inflammation | LIFR | P42702 | 5 | 38474668 | 38608354 | 1.20 | 1.03 | 1.40 | 1.99E-02 | 0.312 |
| OID20254 | LTBP2 | Cardiometabolic | LTBP2 | Q14767 | 14 | 74498183 | 74612378 | 1.24 | 1.03 | 1.48 | 2.21E-02 | 0.312 |
| OID21350 | LYN | Oncology | LYN | P07948 | 8 | 55879835 | 56014169 | 0.84 | 0.71 | 0.99 | 3.31E-02 | 0.312 |
| OID31180 | LYZL2 | Oncology_II | LYZL2 | Q7Z4W2 | 10 | 30611779 | 30629762 | 0.79 | 0.65 | 0.97 | 2.44E-02 | 0.312 |
| OID31293 | MAPKAPK2 | Oncology_II | MAPKAPK2 | P49137 | 1 | 206684905 | 206734281 | 0.82 | 0.69 | 0.97 | 1.88E-02 | 0.312 |
| OID30658 | MARS1 | Inflammation_II | MARS1 | P56192 | 12 | 57475445 | 57517569 | 0.83 | 0.71 | 0.98 | 3.06E-02 | 0.312 |
| OID20815 | METAP1 | Neurology | METAP1 | P53582 | 4 | 98995659 | 99062809 | 0.82 | 0.69 | 0.98 | 2.67E-02 | 0.312 |
| OID20711 | MGLL | Inflammation | MGLL | Q99685 | 3 | 127689062 | 128052190 | 0.84 | 0.72 | 0.99 | 3.25E-02 | 0.312 |
| OID20482 | MILR1 | Inflammation | MILR1 | Q7Z6M3 | 17 | 64449037 | 64468643 | 1.20 | 1.03 | 1.41 | 1.80E-02 | 0.312 |
| OID31156 | MINK1 | Neurology_II | MINK1 | Q8N4C8 | 17 | 4833340 | 4898061 | 0.84 | 0.72 | 0.99 | 3.46E-02 | 0.312 |
| OID20959 | MITD1 | Neurology | MITD1 | Q8WV92 | 2 | 99161427 | 99181058 | 0.83 | 0.71 | 0.97 | 1.90E-02 | 0.312 |
| OID21083 | MMP3 | Neurology | MMP3 | P08254 | 11 | 102835801 | 102843609 | 1.24 | 1.02 | 1.50 | 2.73E-02 | 0.312 |
| OID20092 | MPHOSPH8 | Cardiometabolic | MPHOSPH8 | Q99549 | 13 | 19633659 | 19673441 | 0.80 | 0.67 | 0.95 | 1.32E-02 | 0.312 |
| OID30891 | MRI1 | Neurology_II | MRI1 | Q9BV20 | 19 | 13764522 | 13774282 | 0.83 | 0.70 | 0.98 | 2.60E-02 | 0.312 |
| OID31078 | MTHFD2 | Neurology_II | MTHFD2 | P13995 | 2 | 74186172 | 74217565 | 0.79 | 0.64 | 0.96 | 2.01E-02 | 0.312 |
| OID30188 | NAA10 | Cardiometabolic_II | NAA10 | P41227 | X | 153929225 | 153935080 | 0.83 | 0.72 | 0.97 | 2.19E-02 | 0.312 |
| OID31114 | NAPRT | Neurology_II | NAPRT | Q6XQN6 | 8 | 143574785 | 143578649 | 0.81 | 0.67 | 0.98 | 3.24E-02 | 0.312 |
| OID20442 | NBN | Inflammation | NBN | O60934 | 8 | 89933331 | 90003228 | 0.79 | 0.64 | 0.98 | 2.85E-02 | 0.312 |
| OID20545 | NFATC1 | Inflammation | NFATC1 | O95644 | 18 | 79395856 | 79529325 | 0.82 | 0.69 | 0.97 | 1.82E-02 | 0.312 |
| OID30160 | NFKB1 | Cardiometabolic_II | NFKB1 | P19838 | 4 | 102501330 | 102617302 | 0.82 | 0.70 | 0.97 | 2.28E-02 | 0.312 |
| OID30134 | NFYA | Cardiometabolic_II | NFYA | P23511 | 6 | 41072974 | 41102403 | 0.76 | 0.60 | 0.95 | 1.67E-02 | 0.312 |
| OID30614 | NHLRC3 | Inflammation_II | NHLRC3 | Q5JS37 | 13 | 39038306 | 39050109 | 0.81 | 0.69 | 0.96 | 1.35E-02 | 0.312 |
| OID30336 | NIT2 | Cardiometabolic_II | NIT2 | Q9NQR4 | 3 | 100334739 | 100361635 | 0.84 | 0.71 | 0.99 | 3.45E-02 | 0.312 |
| OID31021 | NMT1 | Neurology_II | NMT1 | P30419 | 17 | 44957992 | 45109016 | 0.79 | 0.65 | 0.96 | 1.60E-02 | 0.312 |
| OID20049 | NPPB | Cardiometabolic | NPPB | P16860 | 1 | 11857464 | 11858945 | 1.20 | 1.02 | 1.41 | 2.55E-02 | 0.312 |
| OID31484 | NUDT16 | Oncology_II | NUDT16 | Q96DE0 | 3 | 131381671 | 131388830 | 0.82 | 0.69 | 0.96 | 1.48E-02 | 0.312 |
| OID31465 | OGA | Oncology_II | OGA | O60502 | 10 | 101784443 | 101818465 | 0.81 | 0.68 | 0.96 | 1.53E-02 | 0.312 |
| OID21352 | OMG | Oncology | OMG | P23515 | 17 | 31272013 | 31297539 | 0.84 | 0.72 | 0.98 | 2.84E-02 | 0.312 |
| OID30835 | OMP | Neurology_II | OMP | P47874 | 11 | 77102840 | 77103331 | 1.17 | 1.02 | 1.34 | 2.04E-02 | 0.312 |
| OID31286 | PAFAH2 | Oncology_II | PAFAH2 | Q99487 | 1 | 25959767 | 25998117 | 0.80 | 0.67 | 0.95 | 1.34E-02 | 0.312 |
| OID20500 | PARP1 | Inflammation | PARP1 | P09874 | 1 | 226360210 | 226408154 | 0.77 | 0.61 | 0.97 | 2.37E-02 | 0.312 |
| OID30591 | PAXX | Inflammation_II | PAXX | Q9BUH6 | 9 | 136992422 | 136993984 | 0.81 | 0.67 | 0.96 | 1.87E-02 | 0.312 |
| OID20741 | PDGFB | Inflammation | PDGFB | P01127 | 22 | 39223359 | 39244982 | 0.84 | 0.72 | 0.97 | 2.19E-02 | 0.312 |
| OID30573 | PDIA3 | Inflammation_II | PDIA3 | P30101 | 15 | 43746394 | 43773279 | 0.82 | 0.69 | 0.98 | 3.24E-02 | 0.312 |
| OID21356 | PFKFB2 | Oncology | PFKFB2 | O60825 | 1 | 207034366 | 207081024 | 0.82 | 0.70 | 0.97 | 2.06E-02 | 0.312 |
| OID20326 | PI3 | Cardiometabolic | PI3 | P19957 | 20 | 45174902 | 45176544 | 1.19 | 1.03 | 1.39 | 2.27E-02 | 0.312 |
| OID21102 | PIGR | Neurology | PIGR | P01833 | 1 | 206928522 | 206946466 | 1.23 | 1.03 | 1.47 | 2.57E-02 | 0.312 |
| OID31088 | PLEKHO1 | Neurology_II | PLEKHO1 | Q53GL0 | 1 | 150149183 | 150164720 | 0.82 | 0.69 | 0.98 | 2.52E-02 | 0.312 |
| OID20445 | PNPT1 | Inflammation | PNPT1 | Q8TCS8 | 2 | 55634061 | 55693863 | 0.78 | 0.63 | 0.95 | 1.55E-02 | 0.312 |
| OID20176 | PPP1R2 | Cardiometabolic | PPP1R2 | P41236 | 3 | 195514428 | 195543386 | 0.83 | 0.70 | 0.99 | 3.34E-02 | 0.312 |
| OID31502 | PPP2R5A | Oncology_II | PPP2R5A | Q15172 | 1 | 212285410 | 212361853 | 0.83 | 0.70 | 0.98 | 2.80E-02 | 0.312 |
| OID21225 | PRKRA | Oncology | PRKRA | O75569 | 2 | 178431292 | 178451512 | 0.82 | 0.69 | 0.97 | 2.06E-02 | 0.312 |
| OID20543 | PROK1 | Inflammation | PROK1 | P58294 | 1 | 110451149 | 110457358 | 1.29 | 1.04 | 1.60 | 1.95E-02 | 0.312 |
| OID20511 | PSPN | Inflammation | PSPN | O60542 | 19 | 6375148 | 6379058 | 0.77 | 0.61 | 0.98 | 3.26E-02 | 0.312 |
| OID30596 | PSTPIP2 | Inflammation_II | PSTPIP2 | Q9H939 | 18 | 45983536 | 46072272 | 0.85 | 0.73 | 0.99 | 3.39E-02 | 0.312 |
| OID20717 | PTPN6 | Inflammation | PTPN6 | P29350 | 12 | 6946468 | 6961316 | 0.84 | 0.72 | 0.98 | 2.71E-02 | 0.312 |
| OID31461 | PTPRK | Oncology_II | PTPRK | Q15262 | 6 | 127968785 | 128520616 | 0.79 | 0.65 | 0.96 | 1.67E-02 | 0.312 |
| OID21166 | RAD23B | Oncology | RAD23B | P54727 | 9 | 107283137 | 107332192 | 0.82 | 0.69 | 0.98 | 2.96E-02 | 0.312 |
| OID30507 | RBPMS | Inflammation_II | RBPMS | Q93062 | 8 | 30384511 | 30572256 | 0.77 | 0.61 | 0.97 | 2.81E-02 | 0.312 |
| OID30999 | RGS10 | Neurology_II | RGS10 | O43665 | 10 | 119499817 | 119542719 | 0.83 | 0.70 | 0.98 | 3.08E-02 | 0.312 |
| OID30342 | RILPL2 | Cardiometabolic_II | RILPL2 | Q969X0 | 12 | 123415039 | 123436684 | 0.84 | 0.72 | 0.99 | 3.44E-02 | 0.312 |
| OID30995 | RLN1 | Neurology_II | RLN1 | P04808 | 9 | 5334930 | 5339876 | 0.60 | 0.39 | 0.93 | 2.15E-02 | 0.312 |
| OID31291 | RNF43 | Oncology_II | RNF43 | Q68DV7 | 17 | 58353676 | 58417595 | 0.79 | 0.63 | 0.98 | 3.36E-02 | 0.312 |
| OID31324 | RPE | Oncology_II | RPE | Q96AT9 | 2 | 210002565 | 210022260 | 0.82 | 0.69 | 0.98 | 3.32E-02 | 0.312 |
| OID30874 | SAG | Neurology_II | SAG | P10523 | 2 | 233307816 | 233347055 | 0.81 | 0.68 | 0.97 | 1.88E-02 | 0.312 |
| OID30278 | SCPEP1 | Cardiometabolic_II | SCPEP1 | Q9HB40 | 17 | 56978129 | 57006768 | 0.81 | 0.68 | 0.97 | 1.90E-02 | 0.312 |
| OID30269 | SEC31A | Cardiometabolic_II | SEC31A | O94979 | 4 | 82818509 | 82901166 | 0.83 | 0.71 | 0.98 | 2.68E-02 | 0.312 |
| OID20379 | SELP | Cardiometabolic | SELP | P16109 | 1 | 169588849 | 169630193 | 0.84 | 0.72 | 0.99 | 3.21E-02 | 0.312 |
| OID21020 | SEMA4D | Neurology | SEMA4D | Q92854 | 9 | 89360787 | 89498130 | 0.83 | 0.70 | 0.97 | 1.95E-02 | 0.312 |
| OID21088 | SERPINB1 | Neurology | SERPINB1 | P30740 | 6 | 2832332 | 2841959 | 0.83 | 0.70 | 0.99 | 3.45E-02 | 0.312 |
| OID20790 | SESTD1 | Neurology | SESTD1 | Q86VW0 | 2 | 179101678 | 179264832 | 0.82 | 0.70 | 0.98 | 2.43E-02 | 0.312 |
| OID21222 | SH2B3 | Oncology | SH2B3 | Q9UQQ2 | 12 | 111405923 | 111451623 | 0.83 | 0.71 | 0.97 | 2.16E-02 | 0.312 |
| OID20514 | SH2D1A | Inflammation | SH2D1A | O60880 | X | 124227868 | 124373197 | 0.80 | 0.66 | 0.97 | 2.62E-02 | 0.312 |
| OID31334 | SH3GL3 | Oncology_II | SH3GL3 | Q99963 | 15 | 83447228 | 83618743 | 0.84 | 0.71 | 0.99 | 3.26E-02 | 0.312 |
| OID31017 | SLA2 | Neurology_II | SLA2 | Q9H6Q3 | 20 | 36612318 | 36646196 | 0.84 | 0.72 | 0.99 | 3.42E-02 | 0.312 |
| OID31418 | SMAD3 | Oncology_II | SMAD3 | P84022 | 15 | 67063763 | 67195173 | 0.83 | 0.70 | 0.99 | 3.44E-02 | 0.312 |
| OID30052 | SNU13 | Cardiometabolic_II | SNU13 | P55769 | 22 | 41673933 | 41690504 | 0.78 | 0.63 | 0.98 | 3.38E-02 | 0.312 |
| OID20135 | SNX9 | Cardiometabolic | SNX9 | Q9Y5X1 | 6 | 157700387 | 157945077 | 0.83 | 0.70 | 0.98 | 3.28E-02 | 0.312 |
| OID20759 | SPON1 | Inflammation | SPON1 | Q9HCB6 | 11 | 13962723 | 14268133 | 1.20 | 1.03 | 1.40 | 1.83E-02 | 0.312 |
| OID30624 | ST13 | Inflammation_II | ST13 | P50502 | 22 | 40824535 | 40856639 | 0.83 | 0.71 | 0.98 | 2.84E-02 | 0.312 |
| OID21277 | STAT5B | Oncology | STAT5B | P51692 | 17 | 42199177 | 42276707 | 0.84 | 0.71 | 0.98 | 3.15E-02 | 0.312 |
| OID20140 | STK4 | Cardiometabolic | STK4 | Q13043 | 20 | 44966479 | 45080021 | 0.82 | 0.69 | 0.96 | 1.63E-02 | 0.312 |
| OID31524 | SUGP1 | Oncology_II | SUGP1 | Q8IWZ8 | 19 | 19276018 | 19320509 | 0.80 | 0.66 | 0.97 | 2.10E-02 | 0.312 |
| OID21285 | TACC3 | Oncology | TACC3 | Q9Y6A5 | 4 | 1712858 | 1745171 | 0.83 | 0.71 | 0.97 | 2.14E-02 | 0.312 |
| OID31343 | TARS1 | Oncology_II | TARS1 | P26639 | 5 | 33440696 | 33468091 | 0.82 | 0.69 | 0.98 | 2.63E-02 | 0.312 |
| OID30616 | TBCA | Inflammation_II | TBCA | O75347 | 5 | 77691166 | 77868780 | 0.82 | 0.69 | 0.96 | 1.57E-02 | 0.312 |
| OID21220 | TBL1X | Oncology | TBL1X | O60907 | X | 9463320 | 9741037 | 0.83 | 0.70 | 0.98 | 2.48E-02 | 0.312 |
| OID20987 | TCL1A | Neurology | TCL1A | P56279 | 14 | 95709947 | 95714196 | 0.82 | 0.70 | 0.97 | 2.08E-02 | 0.312 |
| OID20816 | TDGF1 | Neurology | TDGF1 | P13385 | 3 | 46574534 | 46582457 | 1.19 | 1.02 | 1.38 | 2.66E-02 | 0.312 |

| | | | | | | | | | | | | |
|---|---|---|---|---|---|---|---|---|---|---|---|---|
| OID31403 | TDP1 | Oncology_II | TDP1 | Q9NUW8 | 14 | 89954939 | 90044764 | 0.79 | 0.65 | 0.98 | 3.00E-02 | 0.312 |
| OID20303 | TIE1 | Cardiometabolic | TIE1 | P35590 | 1 | 43300982 | 43323108 | 1.26 | 1.03 | 1.55 | 2.32E-02 | 0.312 |
| OID30940 | TMCO5A | Neurology_II | TMCO5A | Q8N6Q1 | 15 | 37921939 | 37967724 | 0.80 | 0.67 | 0.96 | 1.44E-02 | 0.312 |
| OID31446 | TNFAIP2 | Oncology_II | TNFAIP2 | Q03169 | 14 | 103121476 | 103137439 | 0.81 | 0.67 | 0.99 | 3.48E-02 | 0.312 |
| OID20433 | TNFAIP8 | Inflammation | TNFAIP8 | O95379 | 5 | 119268692 | 119399688 | 0.83 | 0.70 | 0.98 | 3.15E-02 | 0.312 |
| OID30629 | TNFAIP8L2 | Inflammation_II | TNFAIP8L2 | Q6P589 | 1 | 151156649 | 151159749 | 0.81 | 0.68 | 0.96 | 1.84E-02 | 0.312 |
| OID20480 | TNFRSF13C | Inflammation | TNFRSF13C | Q96RJ3 | 22 | 41922032 | 41926806 | 0.83 | 0.70 | 0.98 | 3.14E-02 | 0.312 |
| OID30609 | TPD52L2 | Inflammation_II | TPD52L2 | O43399 | 20 | 63865228 | 63891545 | 0.84 | 0.72 | 0.98 | 2.29E-02 | 0.312 |
| OID30338 | TPK1 | Cardiometabolic_II | TPK1 | Q9H3S4 | 7 | 144451941 | 144836395 | 0.83 | 0.71 | 0.98 | 2.47E-02 | 0.312 |
| OID31368 | UBE2B | Oncology_II | UBE2B | P63146 | 5 | 134371184 | 134392108 | 0.81 | 0.68 | 0.97 | 2.02E-02 | 0.312 |
| OID30654 | UBXN1 | Inflammation_II | UBXN1 | Q04323 | 11 | 62676498 | 62679117 | 0.82 | 0.69 | 0.98 | 2.77E-02 | 0.312 |
| OID31486 | UFD1 | Oncology_II | UFD1 | Q92890 | 22 | 19449911 | 19479202 | 0.83 | 0.71 | 0.98 | 2.80E-02 | 0.312 |
| OID21367 | USO1 | Oncology | USO1 | O60763 | 4 | 75724577 | 75814286 | 0.83 | 0.71 | 0.97 | 2.15E-02 | 0.312 |
| OID20110 | USP8 | Cardiometabolic | USP8 | P40818 | 15 | 50424380 | 50514421 | 0.82 | 0.69 | 0.97 | 2.35E-02 | 0.312 |
| OID31386 | VPS4B | Oncology_II | VPS4B | O75351 | 18 | 63389190 | 63422483 | 0.81 | 0.68 | 0.96 | 1.29E-02 | 0.312 |
| OID20478 | YTHDF3 | Inflammation | YTHDF3 | Q7Z739 | 8 | 63168553 | 63212786 | 0.83 | 0.71 | 0.97 | 2.03E-02 | 0.312 |
| OID30605 | YWHAQ | Inflammation_II | YWHAQ | P27348 | 2 | 9583967 | 9630997 | 0.82 | 0.69 | 0.97 | 1.87E-02 | 0.312 |
| OID21205 | ZBTB16 | Oncology | ZBTB16 | Q05516 | 11 | 114059041 | 114256765 | 0.81 | 0.68 | 0.97 | 2.06E-02 | 0.312 |
| OID31347 | ZNRF4 | Oncology_II | ZNRF4 | Q8WWF5 | 19 | 5455417 | 5456856 | 0.81 | 0.68 | 0.97 | 1.85E-02 | 0.312 |
| OID21377 | GALNT7 | Oncology | GALNT7 | Q86SF2 | 4 | 173168811 | 173323967 | 0.85 | 0.73 | 0.99 | 3.53E-02 | 0.316 |
| OID20583 | ANXA11 | Inflammation | ANXA11 | P50995 | 10 | 80150889 | 80205572 | 0.86 | 0.74 | 0.99 | 3.56E-02 | 0.316 |
| OID20721 | CKMT1A_CKMT1 | Inflammation | (MT1B;CKMT1A | P12532;P12532 | 15;15 | 43593054;43692886 | 43604901;43699222 | 1.20 | 1.01 | 1.43 | 3.58E-02 | 0.316 |
| OID20059 | DUOX2 | Cardiometabolic | DUOX2 | Q9NRD8 | 15 | 45092650 | 45114172 | 0.81 | 0.67 | 0.99 | 3.60E-02 | 0.316 |
| OID30425 | HIF1A | Inflammation_II | HIF1A | Q16665 | 14 | 61695513 | 61748259 | 0.79 | 0.64 | 0.99 | 3.61E-02 | 0.316 |
| OID31410 | PCSK7 | Oncology_II | PCSK7 | Q16549 | 11 | 117204337 | 117232525 | 0.82 | 0.69 | 0.99 | 3.60E-02 | 0.316 |
| OID20870 | TARBP2 | Neurology | TARBP2 | Q15633 | 12 | 53500921 | 53506431 | 0.84 | 0.71 | 0.99 | 3.61E-02 | 0.316 |
| OID20098 | THPO | Cardiometabolic | THPO | P40225 | 3 | 184371935 | 184381968 | 0.84 | 0.71 | 0.99 | 3.61E-02 | 0.316 |
| OID31001 | CCDC50 | Neurology_II | CCDC50 | Q8IVM0 | 3 | 191329085 | 191398659 | 0.79 | 0.64 | 0.99 | 3.67E-02 | 0.316 |
| OID20093 | DNAJB8 | Cardiometabolic | DNAJB8 | Q8NHS0 | 3 | 128462437 | 128466968 | 0.81 | 0.67 | 0.99 | 3.76E-02 | 0.316 |
| OID30519 | EVI5 | Inflammation_II | EVI5 | O60447 | 1 | 92508696 | 92792404 | 0.84 | 0.71 | 0.99 | 3.72E-02 | 0.316 |
| OID30459 | FGF20 | Inflammation_II | FGF20 | Q9NP95 | 8 | 16992181 | 17002345 | 0.80 | 0.65 | 0.99 | 3.82E-02 | 0.316 |
| OID31026 | GOLGA3 | Neurology_II | GOLGA3 | Q08378 | 12 | 132768902 | 132829143 | 0.83 | 0.70 | 0.99 | 3.79E-02 | 0.316 |
| OID20795 | IFNL1 | Neurology | IFNL1 | Q8IU54 | 19 | 39296407 | 39298673 | 1.16 | 1.01 | 1.34 | 3.79E-02 | 0.316 |
| OID21071 | ITGAM | Neurology | ITGAM | P11215 | 16 | 31259967 | 31332892 | 0.84 | 0.71 | 0.99 | 3.66E-02 | 0.316 |
| OID30523 | LATS1 | Inflammation_II | LATS1 | O95835 | 6 | 149658153 | 149718105 | 0.84 | 0.71 | 0.99 | 3.76E-02 | 0.316 |
| OID31327 | MNAT1 | Oncology_II | MNAT1 | P51948 | 14 | 60734742 | 60969965 | 0.81 | 0.66 | 0.99 | 3.69E-02 | 0.316 |
| OID30268 | NFE2 | Cardiometabolic_II | NFE2 | Q16621 | 12 | 54292111 | 54301015 | 0.84 | 0.71 | 0.99 | 3.80E-02 | 0.316 |
| OID30158 | NFX1 | Cardiometabolic_II | NFX1 | Q12986 | 9 | 33290512 | 33371157 | 0.84 | 0.71 | 0.99 | 3.74E-02 | 0.316 |
| OID20125 | NTproBNP | Cardiometabolic | NPPB | P16860 | 1 | 11857464 | 11858945 | 1.17 | 1.01 | 1.36 | 3.82E-02 | 0.316 |
| OID31306 | SAT2 | Oncology_II | SAT2 | Q96F10 | 17 | 7626234 | 7627876 | 0.81 | 0.66 | 0.99 | 3.64E-02 | 0.316 |
| OID30745 | SELENOP | Inflammation_II | SELENOP | P49908 | 5 | 42799880 | 42887392 | 0.84 | 0.71 | 0.99 | 3.82E-02 | 0.316 |
| OID21521 | SRC | Oncology | SRC | P12931 | 20 | 37344685 | 37406050 | 0.84 | 0.72 | 0.99 | 3.71E-02 | 0.316 |
| OID20955 | STAMBP | Neurology | STAMBP | O95630 | 2 | 73828916 | 73873659 | 0.84 | 0.71 | 0.99 | 3.79E-02 | 0.316 |
| OID31422 | TADA3 | Oncology_II | TADA3 | O75528 | 3 | 9779967 | 9793011 | 0.84 | 0.71 | 0.99 | 3.67E-02 | 0.316 |
| OID21232 | TP53 | Oncology | TP53 | P04637 | 17 | 7661779 | 7687538 | 0.84 | 0.72 | 0.99 | 3.72E-02 | 0.316 |
| OID30640 | VTI1A | Inflammation_II | VTI1A | Q96AJ9 | 10 | 112446998 | 112818744 | 0.83 | 0.69 | 0.99 | 3.76E-02 | 0.316 |
| OID20077 | STK11 | Cardiometabolic | STK11 | Q15831 | 19 | 1177558 | 1228431 | 0.84 | 0.72 | 0.99 | 3.84E-02 | 0.317 |
| OID21480 | DCXR | Oncology | DCXR | Q7Z4W1 | 17 | 82035136 | 82037709 | 0.82 | 0.68 | 0.99 | 3.86E-02 | 0.317 |
| OID30670 | TXN | Inflammation_II | TXN | P10599 | 9 | 110243810 | 110256507 | 0.82 | 0.68 | 0.99 | 3.87E-02 | 0.317 |
| OID31478 | AP3B1 | Oncology_II | AP3B1 | O00203 | 5 | 78000522 | 78294762 | 0.84 | 0.71 | 0.99 | 3.90E-02 | 0.318 |
| OID21338 | ERP44 | Oncology | ERP44 | Q9BS26 | 9 | 99979185 | 100099052 | 0.84 | 0.71 | 0.99 | 3.89E-02 | 0.318 |
| OID30899 | KCNH2 | Neurology_II | KCNH2 | Q12809 | 7 | 150944961 | 150978321 | 0.80 | 0.65 | 0.99 | 3.91E-02 | 0.318 |
| OID30986 | KLHL41 | Neurology_II | KLHL41 | O60662 | 2 | 169509702 | 169526258 | 0.81 | 0.67 | 0.99 | 3.91E-02 | 0.318 |
| OID31336 | ARHGAP30 | Oncology_II | ARHGAP30 | Q7Z6I6 | 1 | 161046946 | 161069970 | 0.78 | 0.62 | 0.99 | 4.01E-02 | 0.319 |
| OID20594 | BANK1 | Inflammation | BANK1 | Q8NDB2 | 4 | 101411286 | 102074812 | 0.85 | 0.73 | 0.99 | 4.16E-02 | 0.319 |
| OID30717 | BCHE | Inflammation_II | BCHE | P06276 | 3 | 165772904 | 165837462 | 0.84 | 0.70 | 0.99 | 4.20E-02 | 0.319 |
| OID30556 | CD226 | Inflammation_II | CD226 | Q15762 | 18 | 69831158 | 69961803 | 0.85 | 0.72 | 0.99 | 4.18E-02 | 0.319 |
| OID31251 | CEACAM18 | Oncology_II | CEACAM18 | A8MTB9 | 19 | 51478539 | 51491301 | 1.15 | 1.01 | 1.31 | 4.19E-02 | 0.319 |
| OID30400 | CKB | Cardiometabolic_II | CKB | P12277 | 14 | 103519667 | 103522833 | 1.20 | 1.01 | 1.43 | 4.05E-02 | 0.319 |
| OID20701 | CRIM1 | Inflammation | CRIM1 | Q9NZV1 | 2 | 36355778 | 36551135 | 1.18 | 1.01 | 1.39 | 4.18E-02 | 0.319 |
| OID30272 | CRYZL1 | Cardiometabolic_II | CRYZL1 | O95825 | 21 | 33589341 | 33643926 | 0.85 | 0.72 | 0.99 | 4.04E-02 | 0.319 |
| OID20698 | EGF | Inflammation | EGF | P01133 | 4 | 109912883 | 110013766 | 0.85 | 0.73 | 0.99 | 4.15E-02 | 0.319 |
| OID31381 | FNTA | Oncology_II | FNTA | P49354 | 8 | 43034194 | 43085788 | 0.82 | 0.68 | 0.99 | 3.99E-02 | 0.319 |
| OID21242 | GSTA3 | Oncology | GSTA3 | Q16772 | 6 | 52896639 | 52909698 | 0.82 | 0.68 | 0.99 | 4.19E-02 | 0.319 |
| OID20187 | LEP | Cardiometabolic | LEP | P41159 | 7 | 128241278 | 128257629 | 0.76 | 0.58 | 0.99 | 4.22E-02 | 0.319 |
| OID21406 | LGALS7_LGALS | Oncology | ALS7B;LGALS7 | P47929;P47929 | 19;19 | 38789200;38770968 | 38791754;38773517 | 1.18 | 1.01 | 1.39 | 4.02E-02 | 0.319 |
| OID31269 | LSM8 | Oncology_II | LSM8 | O95777 | 7 | 118184144 | 118204035 | 0.81 | 0.66 | 0.99 | 4.07E-02 | 0.319 |
| OID21513 | MSRA | Oncology | MSRA | Q9UJ68 | 8 | 10054292 | 10428891 | 0.84 | 0.72 | 0.99 | 4.21E-02 | 0.319 |
| OID31475 | NAP1L4 | Oncology_II | NAP1L4 | Q99733 | 11 | 2944431 | 2992377 | 0.85 | 0.72 | 0.99 | 4.18E-02 | 0.319 |
| OID30568 | NUMB | Inflammation_II | NUMB | P49757 | 14 | 73275107 | 73458617 | 0.85 | 0.72 | 0.99 | 4.04E-02 | 0.319 |
| OID31487 | OTUD6B | Oncology_II | OTUD6B | Q8N6M0 | 8 | 91070196 | 91087095 | 0.84 | 0.71 | 0.99 | 4.18E-02 | 0.319 |
| OID31119 | PRKAR2A | Neurology_II | PRKAR2A | P13861 | 3 | 48744597 | 48847874 | 0.84 | 0.71 | 0.99 | 4.17E-02 | 0.319 |
| OID21519 | S100A4 | Oncology | S100A4 | P26447 | 1 | 153543613 | 153550136 | 0.85 | 0.72 | 0.99 | 4.04E-02 | 0.319 |
| OID21506 | SNAP29 | Oncology | SNAP29 | O95721 | 22 | 20859007 | 20891214 | 0.85 | 0.72 | 0.99 | 4.02E-02 | 0.319 |
| OID30644 | SNX15 | Inflammation_II | SNX15 | Q9NRS6 | 11 | 65027439 | 65040572 | 0.83 | 0.70 | 0.99 | 4.08E-02 | 0.319 |
| OID31086 | SPTLC1 | Neurology_II | SPTLC1 | O15269 | 9 | 92000087 | 92115413 | 0.80 | 0.65 | 0.99 | 4.10E-02 | 0.319 |
| OID30536 | STAT2 | Inflammation_II | STAT2 | P52630 | 12 | 56341597 | 56360203 | 0.84 | 0.71 | 0.99 | 4.14E-02 | 0.319 |
| OID20942 | TBCC | Neurology | TBCC | Q15814 | 6 | 42744498 | 42746103 | 0.85 | 0.72 | 0.99 | 4.18E-02 | 0.319 |
| OID31185 | ZP4 | Oncology_II | ZP4 | Q12836 | 1 | 237877864 | 237890922 | 0.79 | 0.64 | 0.99 | 4.22E-02 | 0.319 |
| OID20806 | AMFR | Neurology | AMFR | Q9UKV5 | 16 | 56361452 | 56425545 | 0.82 | 0.68 | 0.99 | 4.37E-02 | 0.320 |
| OID31135 | CASP7 | Neurology_II | CASP7 | P55210 | 10 | 113679162 | 113730907 | 0.84 | 0.71 | 0.99 | 4.30E-02 | 0.320 |
| OID31495 | CEACAM16 | Oncology_II | CEACAM16 | Q2WEN9 | 19 | 44699151 | 44710718 | 0.84 | 0.71 | 1.00 | 4.37E-02 | 0.320 |
| OID31080 | DXO | Neurology_II | DXO | O77932 | 6 | 31969810 | 31972290 | 0.84 | 0.71 | 0.99 | 4.34E-02 | 0.320 |
| OID31022 | EIF4G3 | Neurology_II | EIF4G3 | O43432 | 1 | 20806292 | 21177285 | 0.84 | 0.71 | 0.99 | 4.29E-02 | 0.320 |
| OID31037 | HNRNPUL1 | Neurology_II | HNRNPUL1 | Q9BUJ2 | 19 | 41262496 | 41307787 | 0.81 | 0.66 | 0.99 | 4.26E-02 | 0.320 |
| OID30232 | HSBP1 | Cardiometabolic_II | HSBP1 | O75506 | 16 | 83719311 | 83819737 | 0.84 | 0.71 | 1.00 | 4.41E-02 | 0.320 |
| OID21288 | MPI | Oncology | MPI | P34949 | 15 | 74890005 | 74902219 | 0.84 | 0.70 | 0.99 | 4.24E-02 | 0.320 |
| OID20850 | PMVK | Neurology | PMVK | Q15126 | 1 | 154924740 | 154936719 | 0.86 | 0.74 | 1.00 | 4.42E-02 | 0.320 |
| OID30350 | PPM1F | Cardiometabolic_II | PPM1F | P49593 | 22 | 21919425 | 21952848 | 0.84 | 0.71 | 1.00 | 4.38E-02 | 0.320 |
| OID20644 | PRDX5 | Inflammation | PRDX5 | P30044 | 11 | 64318121 | 64321811 | 0.86 | 0.73 | 1.00 | 4.38E-02 | 0.320 |
| OID30659 | PTRHD1 | Inflammation_II | PTRHD1 | Q6GMV3 | 2 | 24789728 | 24793391 | 0.85 | 0.72 | 1.00 | 4.35E-02 | 0.320 |
| OID31028 | REEP4 | Neurology_II | REEP4 | Q9H6H4 | 8 | 22138020 | 22141951 | 0.83 | 0.70 | 0.99 | 4.35E-02 | 0.320 |
| OID21179 | RNF41 | Oncology | RNF41 | Q9H4P4 | 12 | 56202179 | 56221933 | 0.84 | 0.70 | 0.99 | 4.31E-02 | 0.320 |
| OID30630 | SLC9A3R1 | Inflammation_II | SLC9A3R1 | O14745 | 17 | 74748628 | 74769353 | 0.84 | 0.71 | 1.00 | 4.40E-02 | 0.320 |
| OID20182 | SSC4D | Cardiometabolic | SSC4D | Q8WTU2 | 7 | 76389334 | 76409697 | 0.82 | 0.68 | 0.99 | 4.31E-02 | 0.320 |
| OID20072 | SUSD1 | Cardiometabolic | SUSD1 | Q6UWL2 | 9 | 112040783 | 112175297 | 0.86 | 0.74 | 1.00 | 4.36E-02 | 0.320 |
| OID31303 | ZNF75D | Oncology_II | ZNF75D | P51815 | X | 135248920 | 135344109 | 0.82 | 0.67 | 0.99 | 4.39E-02 | 0.320 |
| OID20857 | CERT | Neurology | CERT1 | Q9Y5P4 | 5 | 75356345 | 75512138 | 0.85 | 0.72 | 1.00 | 4.45E-02 | 0.321 |
| OID30167 | ATP6V1G2 | Cardiometabolic_II | ATP6V1G2 | O95670 | 6 | 31544444 | 31548427 | 1.16 | 1.00 | 1.33 | 4.47E-02 | 0.321 |
| OID21226 | DDX58 | Oncology | DDX58 | O95786 | 9 | 32455302 | 32526208 | 0.82 | 0.67 | 1.00 | 4.46E-02 | 0.321 |
| OID20555 | CASP2 | Inflammation | CASP2 | P42575 | 7 | 143288215 | 143307696 | 0.84 | 0.71 | 1.00 | 4.51E-02 | 0.321 |
| OID21477 | TJAP1 | Oncology | TJAP1 | Q5JTD0 | 6 | 43477523 | 43506556 | 0.84 | 0.71 | 1.00 | 4.49E-02 | 0.321 |
| OID30386 | TWF2 | Cardiometabolic_II | TWF2 | Q6IBS0 | 3 | 52228612 | 52246788 | 0.84 | 0.71 | 1.00 | 4.51E-02 | 0.321 |

| | | | | | | | | | | | | |
|---|---|---|---|---|---|---|---|---|---|---|---|---|
| OID30197 | GNPDA1 | Cardiometabolic_II | GNPDA1 | P46926 | 5 | 141991749 | 142013041 | 0.81 | 0.65 | 1.00 | 4.56E-02 | 0.322 |
| OID21180 | SMAD1 | Oncology | SMAD1 | Q15797 | 4 | 145481194 | 145559176 | 0.84 | 0.71 | 1.00 | 4.54E-02 | 0.322 |
| OID20194 | THOP1 | Cardiometabolic | THOP1 | P52888 | 19 | 2785503 | 2815807 | 0.83 | 0.70 | 1.00 | 4.56E-02 | 0.322 |
| OID21016 | VTA1 | Neurology | VTA1 | Q9NP79 | 6 | 142147162 | 142224685 | 0.85 | 0.72 | 1.00 | 4.57E-02 | 0.322 |
| OID31367 | LACRT | Oncology_II | LACRT | Q9GZZ8 | 12 | 54630811 | 54634895 | 0.83 | 0.68 | 1.00 | 4.60E-02 | 0.323 |
| OID30323 | PDAP1 | Cardiometabolic_II | PDAP1 | Q13442 | 7 | 99392048 | 99408597 | 0.85 | 0.72 | 1.00 | 4.62E-02 | 0.325 |
| OID30130 | ABRAXAS2 | Cardiometabolic_II | ABRAXAS2 | Q15018 | 10 | 124801819 | 124836667 | 0.84 | 0.71 | 1.00 | 4.64E-02 | 0.325 |
| OID30655 | ADD1 | Inflammation_II | ADD1 | P35611 | 4 | 2843844 | 2930076 | 0.85 | 0.72 | 1.00 | 5.02E-02 | 0.326 |
| OID20374 | APOM | Cardiometabolic | APOM | O95445 | 6 | 31652416 | 31658210 | 0.85 | 0.73 | 1.00 | 4.79E-02 | 0.326 |
| OID30221 | CCER2 | Cardiometabolic_II | CCER2 | I3L3R5 | 19 | 38908980 | 38912186 | 1.15 | 1.00 | 1.33 | 4.67E-02 | 0.326 |
| OID21470 | CDC37 | Oncology | CDC37 | Q16543 | 19 | 10391133 | 10420121 | 0.85 | 0.72 | 1.00 | 4.94E-02 | 0.326 |
| OID20054 | CHEK2 | Cardiometabolic | CHEK2 | O96017 | 22 | 28687743 | 28742422 | 0.85 | 0.72 | 1.00 | 4.84E-02 | 0.326 |
| OID30884 | CLEC2L | Neurology_II | CLEC2L | P0C7M8 | 7 | 139523685 | 139544985 | 0.82 | 0.66 | 1.00 | 4.98E-02 | 0.326 |
| OID31406 | CMIP | Oncology_II | CMIP | Q8IY22 | 16 | 81444808 | 81711762 | 0.85 | 0.72 | 1.00 | 4.83E-02 | 0.326 |
| OID20208 | COL4A1 | Cardiometabolic | COL4A1 | P02462 | 13 | 110148963 | 110307157 | 1.18 | 1.00 | 1.38 | 5.02E-02 | 0.326 |
| OID20086 | COMT | Cardiometabolic | COMT | P21964 | 22 | 19941371 | 19969975 | 0.85 | 0.73 | 1.00 | 5.04E-02 | 0.326 |
| OID20995 | CST5 | Neurology | CST5 | P28325 | 20 | 23875934 | 23879748 | 0.84 | 0.71 | 1.00 | 4.93E-02 | 0.326 |
| OID30505 | DAPK2 | Inflammation_II | DAPK2 | Q9UIK4 | 15 | 63907036 | 64072033 | 0.82 | 0.67 | 1.00 | 4.93E-02 | 0.326 |
| OID31125 | EIF1AX | Neurology_II | EIF1AX | P47813 | X | 20124525 | 20141838 | 0.82 | 0.68 | 1.00 | 4.72E-02 | 0.326 |
| OID30557 | EIF4E | Inflammation_II | EIF4E | P06730 | 4 | 98879276 | 98929133 | 0.85 | 0.72 | 1.00 | 4.83E-02 | 0.326 |
| OID21323 | ERBIN | Oncology | ERBIN | Q96RT1 | 5 | 65926556 | 66082546 | 0.85 | 0.72 | 1.00 | 5.00E-02 | 0.326 |
| OID31499 | FKBPL | Oncology_II | FKBPL | Q9UIM3 | 6 | 32128707 | 32130288 | 0.84 | 0.71 | 1.00 | 4.97E-02 | 0.326 |
| OID30600 | GAPDH | Inflammation_II | GAPDH | P04406 | 12 | 6534512 | 6538374 | 0.87 | 0.75 | 1.00 | 4.83E-02 | 0.326 |
| OID30243 | HS1BP3 | Cardiometabolic_II | HS1BP3 | Q53T59 | 2 | 20560448 | 20651130 | 0.85 | 0.71 | 1.00 | 4.98E-02 | 0.326 |
| OID20484 | ICA1 | Inflammation | ICA1 | Q05084 | 7 | 8113184 | 8262687 | 0.84 | 0.71 | 1.00 | 4.79E-02 | 0.326 |
| OID20494 | JCHAIN | Inflammation | JCHAIN | P01591 | 4 | 70655541 | 70681817 | 0.86 | 0.74 | 1.00 | 4.72E-02 | 0.326 |
| OID20669 | KYNU | Inflammation | KYNU | Q16719 | 2 | 142877657 | 143055833 | 0.84 | 0.70 | 1.00 | 4.91E-02 | 0.326 |
| OID31250 | NEK7 | Oncology_II | NEK7 | Q8TDX7 | 1 | 198156994 | 198322420 | 0.82 | 0.67 | 1.00 | 4.72E-02 | 0.326 |
| OID20510 | NUB1 | Inflammation | NUB1 | Q9Y5A7 | 7 | 151341772 | 151378449 | 0.84 | 0.71 | 1.00 | 4.82E-02 | 0.326 |
| OID30584 | PDE5A | Inflammation_II | PDE5A | O76074 | 4 | 119494397 | 119628804 | 0.85 | 0.73 | 1.00 | 4.96E-02 | 0.326 |
| OID20775 | PNLIPRP2 | Inflammation | PNLIPRP2 | P54317 | 10 | 116620953 | 116645143 | 1.20 | 1.00 | 1.44 | 4.79E-02 | 0.326 |
| OID30941 | PRKAG3 | Neurology_II | PRKAG3 | Q9UGI9 | 2 | 218822308 | 218832086 | 0.84 | 0.70 | 1.00 | 4.88E-02 | 0.326 |
| OID20203 | RNASE3 | Cardiometabolic | RNASE3 | P12724 | 14 | 20891385 | 20892348 | 0.83 | 0.69 | 1.00 | 4.97E-02 | 0.326 |
| OID31038 | SATB1 | Neurology_II | SATB1 | Q01826 | 3 | 18345377 | 18445621 | 0.80 | 0.65 | 1.00 | 5.03E-02 | 0.326 |
| OID21161 | SCP2 | Oncology | SCP2 | P22307 | 1 | 52927276 | 53051698 | 0.82 | 0.67 | 1.00 | 4.82E-02 | 0.326 |
| OID20216 | SDC4 | Cardiometabolic | SDC4 | P31431 | 20 | 45325288 | 45348424 | 0.86 | 0.75 | 1.00 | 5.04E-02 | 0.326 |
| OID20218 | SNAP23 | Cardiometabolic | SNAP23 | O00161 | 15 | 42491233 | 42545356 | 0.86 | 0.73 | 1.00 | 4.98E-02 | 0.326 |
| OID31120 | TNIP1 | Neurology_II | TNIP1 | Q15025 | 5 | 151029945 | 151093577 | 0.85 | 0.72 | 1.00 | 4.92E-02 | 0.326 |
| OID31330 | TNPO1 | Oncology_II | TNPO1 | Q92973 | 5 | 72816312 | 72916733 | 0.84 | 0.71 | 1.00 | 5.01E-02 | 0.326 |
| OID31482 | TRIM25 | Oncology_II | TRIM25 | Q14258 | 17 | 56836387 | 56914080 | 0.85 | 0.72 | 1.00 | 4.79E-02 | 0.326 |
| OID30185 | TSNAX | Cardiometabolic_II | TSNAX | Q99598 | 1 | 231528541 | 231566524 | 0.84 | 0.70 | 1.00 | 4.89E-02 | 0.326 |
| OID31494 | YARS1 | Oncology_II | YARS1 | P54577 | 1 | 32775237 | 32818031 | 0.85 | 0.73 | 1.00 | 4.74E-02 | 0.326 |
| OID30356 | ACADSB | Cardiometabolic_II | ACADSB | P45954 | 10 | 123008979 | 123058290 | 0.85 | 0.72 | 1.00 | 5.07E-02 | 0.326 |
| OID20829 | HSP90B1 | Neurology | HSP90B1 | P14625 | 12 | 103930107 | 103953931 | 0.80 | 0.64 | 1.00 | 5.11E-02 | 0.326 |
| OID20506 | IFNLR1 | Inflammation | IFNLR1 | Q8IU57 | 1 | 24154168 | 24187959 | 0.84 | 0.70 | 1.00 | 5.12E-02 | 0.326 |
| OID20544 | IKBKG | Inflammation | IKBKG | Q9Y6K9 | X | 154541199 | 154565046 | 0.85 | 0.72 | 1.00 | 5.09E-02 | 0.326 |
| OID20596 | KRT19 | Inflammation | KRT19 | P08727 | 17 | 41523617 | 41528308 | 1.16 | 1.00 | 1.34 | 5.14E-02 | 0.326 |
| OID30677 | LCAT | Inflammation_II | LCAT | P04180 | 16 | 67939750 | 67944131 | 0.84 | 0.70 | 1.00 | 5.15E-02 | 0.326 |
| OID21437 | METAP2 | Oncology | METAP2 | P50579 | 12 | 95473520 | 95515839 | 0.85 | 0.73 | 1.00 | 5.13E-02 | 0.326 |
| OID20474 | TANK | Inflammation | TANK | Q92844 | 2 | 161136908 | 161236230 | 0.84 | 0.71 | 1.00 | 5.14E-02 | 0.326 |
| OID31016 | TSC22D1 | Neurology_II | TSC22D1 | Q15714 | 13 | 44432143 | 44577147 | 0.84 | 0.70 | 1.00 | 5.07E-02 | 0.326 |
| OID20825 | GLT8D2 | Neurology | GLT8D2 | Q9H1C3 | 12 | 103988987 | 104064183 | 0.81 | 0.66 | 1.00 | 5.17E-02 | 0.327 |
| OID31061 | PIBF1 | Neurology_II | PIBF1 | Q8WXW3 | 13 | 72782133 | 73016461 | 0.85 | 0.72 | 1.00 | 5.19E-02 | 0.327 |
| OID31147 | GCC1 | Neurology_II | GCC1 | Q96CN9 | 7 | 127580628 | 127593611 | 0.85 | 0.73 | 1.00 | 5.24E-02 | 0.329 |
| OID30933 | MFAP3L | Neurology_II | MFAP3L | O75121 | 4 | 169986597 | 170033031 | 0.82 | 0.68 | 1.00 | 5.26E-02 | 0.329 |
| OID21425 | SCLY | Oncology | SCLY | Q96I15 | 2 | 238060924 | 238099413 | 0.84 | 0.70 | 1.00 | 5.28E-02 | 0.329 |
| OID21097 | SPINK1 | Neurology | SPINK1 | P00995 | 5 | 147824572 | 147831671 | 1.16 | 1.00 | 1.34 | 5.28E-02 | 0.329 |
| OID20720 | SPINT2 | Inflammation | SPINT2 | O43291 | 19 | 38244035 | 38292615 | 0.85 | 0.73 | 1.00 | 5.25E-02 | 0.329 |
| OID31054 | STXBP1 | Neurology_II | STXBP1 | P61764 | 9 | 127579370 | 127696027 | 0.85 | 0.73 | 1.00 | 5.29E-02 | 0.329 |
| OID30414 | TALDO1 | Cardiometabolic_II | TALDO1 | P37837 | 11 | 747415 | 765012 | 0.84 | 0.70 | 1.00 | 5.23E-02 | 0.329 |
| OID20932 | SERPINB9 | Neurology | SERPINB9 | P50453 | 6 | 2887270 | 2903309 | 0.86 | 0.73 | 1.00 | 5.31E-02 | 0.329 |
| OID20719 | CSF1 | Inflammation | CSF1 | P09603 | 1 | 109910242 | 109930992 | 1.17 | 1.00 | 1.38 | 5.35E-02 | 0.330 |
| OID21140 | FYB1 | Neurology | FYB1 | O15117 | 5 | 39105252 | 39274528 | 0.86 | 0.73 | 1.00 | 5.35E-02 | 0.330 |
| OID20869 | XRCC4 | Neurology | XRCC4 | Q13426 | 5 | 83077498 | 83353787 | 0.84 | 0.71 | 1.00 | 5.36E-02 | 0.330 |
| OID20648 | HCLS1 | Inflammation | HCLS1 | P14317 | 3 | 121631399 | 121660927 | 0.86 | 0.73 | 1.00 | 5.39E-02 | 0.331 |
| OID30409 | HPSE | Cardiometabolic_II | HPSE | Q9Y251 | 4 | 83292461 | 83335153 | 0.85 | 0.73 | 1.00 | 5.42E-02 | 0.331 |
| OID20158 | IL19 | Cardiometabolic | IL19 | Q9UHD0 | 1 | 206770764 | 206842981 | 1.15 | 1.00 | 1.34 | 5.41E-02 | 0.331 |
| OID30636 | SYAP1 | Inflammation_II | SYAP1 | Q96A49 | X | 16719612 | 16765340 | 0.84 | 0.71 | 1.00 | 5.42E-02 | 0.331 |
| OID20515 | FOXO1 | Inflammation | FOXO1 | Q12778 | 13 | 40555667 | 40666641 | 0.85 | 0.72 | 1.00 | 5.45E-02 | 0.332 |
| OID31111 | DOK1 | Neurology_II | DOK1 | Q99704 | 2 | 74549026 | 74557551 | 0.86 | 0.73 | 1.00 | 5.51E-02 | 0.333 |
| OID20138 | DOK2 | Cardiometabolic | DOK2 | O60496 | 8 | 21908873 | 21913690 | 0.85 | 0.72 | 1.00 | 5.49E-02 | 0.333 |
| OID21402 | FGF21 | Oncology | FGF21 | Q9NSA1 | 19 | 48755524 | 48758333 | 1.17 | 1.00 | 1.38 | 5.54E-02 | 0.333 |
| OID21314 | INPPL1 | Oncology | INPPL1 | O15357 | 11 | 72223701 | 72239147 | 0.85 | 0.72 | 1.00 | 5.50E-02 | 0.333 |
| OID20773 | LGMN | Inflammation | LGMN | Q99538 | 14 | 92703807 | 92748679 | 0.85 | 0.71 | 1.00 | 5.54E-02 | 0.333 |
| OID30662 | SDK2 | Inflammation_II | SDK2 | Q58EX2 | 17 | 73334384 | 73644445 | 0.84 | 0.71 | 1.00 | 5.52E-02 | 0.333 |
| OID20926 | WWP2 | Neurology | WWP2 | O00308 | 16 | 69762328 | 69941741 | 0.85 | 0.72 | 1.00 | 5.50E-02 | 0.333 |
| OID21341 | ATP6V1D | Oncology | ATP6V1D | Q9Y5K8 | 14 | 67294371 | 67360265 | 0.83 | 0.69 | 1.00 | 5.56E-02 | 0.333 |
| OID20899 | MAP4K5 | Neurology | MAP4K5 | Q9Y4K4 | 14 | 50418501 | 50561126 | 0.86 | 0.73 | 1.00 | 5.58E-02 | 0.333 |
| OID31132 | SNRPB2 | Neurology_II | SNRPB2 | P08579 | 20 | 16729961 | 16742564 | 0.76 | 0.58 | 1.01 | 5.57E-02 | 0.333 |
| OID30645 | SNCA | Inflammation_II | SNCA | P37840 | 4 | 89700345 | 89838315 | 0.85 | 0.73 | 1.00 | 5.60E-02 | 0.333 |
| OID30121 | ARL2BP | Cardiometabolic_II | ARL2BP | Q9Y2Y0 | 16 | 57245259 | 57253635 | 0.85 | 0.71 | 1.00 | 5.61E-02 | 0.333 |
| OID20534 | PSIP1 | Inflammation | PSIP1 | O75475 | 9 | 15464066 | 15510995 | 0.83 | 0.68 | 1.00 | 5.62E-02 | 0.333 |
| OID30231 | ENTR1 | Cardiometabolic_II | ENTR1 | Q96C92 | 9 | 136401922 | 136410614 | 0.84 | 0.69 | 1.00 | 5.63E-02 | 0.334 |
| OID30237 | GADD45GIP1 | Cardiometabolic_II | GADD45GIP1 | Q8TAE8 | 19 | 12953119 | 12957223 | 0.84 | 0.71 | 1.00 | 5.68E-02 | 0.336 |
| OID20123 | ANXA4 | Cardiometabolic | ANXA4 | P09525 | 2 | 69644425 | 69827112 | 0.85 | 0.72 | 1.01 | 5.74E-02 | 0.338 |
| OID30994 | ARHGEF10 | Neurology_II | ARHGEF10 | O15013 | 8 | 1823926 | 1958641 | 0.85 | 0.72 | 1.01 | 5.78E-02 | 0.338 |
| OID30170 | ASS1 | Cardiometabolic_II | ASS1 | P00966 | 9 | 130444961 | 130501274 | 0.84 | 0.71 | 1.01 | 5.83E-02 | 0.338 |
| OID21239 | FLI1 | Oncology | FLI1 | Q01543 | 11 | 128686535 | 128813267 | 0.85 | 0.72 | 1.01 | 5.75E-02 | 0.338 |
| OID21086 | HARS1 | Neurology | HARS1 | P12081 | 5 | 140673035 | 140691537 | 0.86 | 0.73 | 1.01 | 5.84E-02 | 0.338 |
| OID20708 | LY6D | Inflammation | LY6D | Q14210 | 8 | 142784882 | 142786539 | 1.15 | 1.00 | 1.34 | 5.81E-02 | 0.338 |
| OID20317 | PLXNB2 | Cardiometabolic | PLXNB2 | O15031 | 22 | 50274979 | 50307646 | 1.17 | 0.99 | 1.38 | 5.78E-02 | 0.338 |
| OID21027 | PPCDC | Neurology | PPCDC | Q96CD2 | 15 | 75023586 | 75117462 | 0.85 | 0.71 | 1.01 | 5.78E-02 | 0.338 |
| OID20536 | SAMD9L | Inflammation | SAMD9L | Q8IVG5 | 7 | 93130056 | 93148385 | 0.85 | 0.72 | 1.01 | 5.81E-02 | 0.338 |
| OID20371 | TFF3 | Cardiometabolic | TFF3 | Q07654 | 21 | 42311667 | 42315409 | 1.15 | 1.00 | 1.32 | 5.82E-02 | 0.338 |
| OID31140 | ZHX2 | Neurology_II | ZHX2 | Q9Y6X8 | 8 | 122781655 | 122974510 | 0.78 | 0.61 | 1.01 | 5.83E-02 | 0.338 |
| OID30590 | APPL2 | Inflammation_II | APPL2 | Q8NEU8 | 12 | 105173297 | 105236203 | 0.86 | 0.73 | 1.01 | 5.99E-02 | 0.339 |
| OID20193 | CDHR5 | Cardiometabolic | CDHR5 | Q9HBB8 | 11 | 616577 | 626078 | 0.86 | 0.74 | 1.01 | 6.01E-02 | 0.339 |
| OID21399 | DCTN1 | Oncology | DCTN1 | Q14203 | 2 | 74361154 | 74392087 | 0.85 | 0.72 | 1.01 | 6.02E-02 | 0.339 |
| OID21037 | ENO1 | Neurology | ENO1 | P06733 | 1 | 8861000 | 8879190 | 0.85 | 0.72 | 1.01 | 5.92E-02 | 0.339 |
| OID21173 | GSAP | Oncology | GSAP | A4D1B5 | 7 | 77310751 | 77416349 | 0.82 | 0.66 | 1.01 | 5.94E-02 | 0.339 |
| OID21181 | MAGED1 | Oncology | MAGED1 | Q9Y5V3 | X | 51803007 | 51902354 | 0.84 | 0.70 | 1.01 | 5.96E-02 | 0.339 |
| OID30669 | MDH1 | Inflammation_II | MDH1 | P40925 | 2 | 63588609 | 63607197 | 0.84 | 0.70 | 1.01 | 5.94E-02 | 0.339 |
| OID21489 | MDK | Oncology | MDK | P21741 | 11 | 46380756 | 46383837 | 0.84 | 0.71 | 1.01 | 6.00E-02 | 0.339 |

| | | | | | | | | | | | | |
|---|---|---|---|---|---|---|---|---|---|---|---|---|
| OID31393 | NACC1 | Oncology_II | NACC1 | Q96RE7 | 19 | 13116862 | 13141147 | 0.85 | 0.72 | 1.01 | 5.97E-02 | 0.339 |
| OID31063 | PDLIM5 | Neurology_II | PDLIM5 | Q96HC4 | 4 | 94451857 | 94668227 | 0.86 | 0.73 | 1.01 | 5.96E-02 | 0.339 |
| OID31155 | PLCB2 | Neurology_II | PLCB2 | Q00722 | 15 | 40278176 | 40307935 | 0.85 | 0.73 | 1.01 | 5.93E-02 | 0.339 |
| OID31360 | PPP1CC | Oncology_II | PPP1CC | P36873 | 12 | 110719680 | 110742939 | 0.85 | 0.72 | 1.01 | 6.00E-02 | 0.339 |
| OID30648 | SFRP4 | Inflammation_II | SFRP4 | Q6FHJ7 | 7 | 37905932 | 38025695 | 0.86 | 0.73 | 1.01 | 5.98E-02 | 0.339 |
| OID31471 | SMNDC1 | Oncology_II | SMNDC1 | O75940 | 10 | 110290730 | 110304938 | 0.81 | 0.65 | 1.01 | 5.95E-02 | 0.339 |
| OID20250 | IL1RL1 | Cardiometabolic | IL1RL1 | Q01638 | 2 | 102311502 | 102352037 | 1.19 | 0.99 | 1.43 | 6.07E-02 | 0.340 |
| OID31419 | UNG | Oncology_II | UNG | P13051 | 12 | 109097597 | 109126725 | 0.85 | 0.72 | 1.01 | 6.06E-02 | 0.340 |
| OID20595 | CD200R1 | Inflammation | CD200R1 | Q8TD46 | 3 | 112921205 | 112975103 | 0.85 | 0.72 | 1.01 | 6.10E-02 | 0.341 |
| OID31463 | FAM13A | Oncology_II | FAM13A | O94988 | 4 | 88725955 | 89111398 | 0.85 | 0.72 | 1.01 | 6.10E-02 | 0.341 |
| OID30893 | NAA80 | Neurology_II | NAA80 | Q93015 | 3 | 50296402 | 50299416 | 0.85 | 0.72 | 1.01 | 6.16E-02 | 0.343 |
| OID30969 | COL28A1 | Neurology_II | COL28A1 | Q2UY09 | 7 | 7356203 | 7535873 | 1.16 | 0.99 | 1.35 | 6.18E-02 | 0.344 |
| OID20867 | PRTFDC1 | Neurology | PRTFDC1 | Q9NRG1 | 10 | 24848614 | 24952606 | 0.86 | 0.74 | 1.01 | 6.21E-02 | 0.344 |
| OID30218 | RNF5 | Cardiometabolic_II | RNF5 | Q99942 | 6 | 32178405 | 32180793 | 0.86 | 0.74 | 1.01 | 6.21E-02 | 0.344 |
| OID20184 | DIABLO | Cardiometabolic | DIABLO | Q9NR28 | 12 | 122207663 | 122227456 | 0.86 | 0.73 | 1.01 | 6.22E-02 | 0.344 |
| OID30294 | NT5C | Cardiometabolic_II | NT5C | Q8TCD5 | 17 | 75130225 | 75131757 | 0.86 | 0.74 | 1.01 | 6.24E-02 | 0.344 |
| OID31055 | ASPSCR1 | Neurology_II | ASPSCR1 | Q9BZE9 | 17 | 81976807 | 82017406 | 0.85 | 0.72 | 1.01 | 6.28E-02 | 0.345 |
| OID20966 | CD274 | Neurology | CD274 | Q9NZQ7 | 9 | 5450503 | 5470566 | 1.16 | 0.99 | 1.35 | 6.27E-02 | 0.345 |
| OID30773 | F11 | Inflammation_II | F11 | P03951 | 4 | 186266189 | 186289681 | 0.84 | 0.70 | 1.01 | 6.29E-02 | 0.345 |
| OID20643 | PPP1R9B | Inflammation | PPP1R9B | Q96SB3 | 17 | 50133737 | 50150677 | 0.86 | 0.73 | 1.01 | 6.29E-02 | 0.345 |
| OID30474 | BAG4 | Inflammation_II | BAG4 | O95429 | 8 | 38176533 | 38213301 | 0.84 | 0.70 | 1.01 | 6.33E-02 | 0.345 |
| OID21405 | CTSV | Oncology | CTSV | O60911 | 9 | 97029677 | 97156556 | 0.85 | 0.71 | 1.01 | 6.31E-02 | 0.345 |
| OID21126 | IL1RAP | Neurology | IL1RAP | Q9NPH3 | 3 | 190514051 | 190659750 | 0.85 | 0.71 | 1.01 | 6.34E-02 | 0.345 |
| OID31087 | OPHN1 | Neurology_II | OPHN1 | O60890 | X | 67949349 | 68433913 | 0.86 | 0.73 | 1.01 | 6.33E-02 | 0.345 |
| OID31207 | TRIM26 | Oncology_II | TRIM26 | Q12899 | 6 | 30184455 | 30213427 | 0.83 | 0.68 | 1.01 | 6.36E-02 | 0.345 |
| OID20509 | SPINK4 | Inflammation | SPINK4 | O60575 | 9 | 33218365 | 33248567 | 1.16 | 0.99 | 1.35 | 6.40E-02 | 0.347 |
| OID30127 | ELOB | Cardiometabolic_II | ELOB | Q15370 | 16 | 2771414 | 2777289 | 0.83 | 0.69 | 1.01 | 6.43E-02 | 0.347 |
| OID21462 | WFDC12 | Oncology | WFDC12 | Q8WWY7 | 20 | 45123425 | 45124465 | 1.18 | 0.99 | 1.40 | 6.44E-02 | 0.347 |
| OID30680 | GPI | Inflammation_II | GPI | P06744 | 19 | 34359480 | 34402413 | 0.85 | 0.71 | 1.01 | 6.45E-02 | 0.348 |
| OID21465 | HBEGF | Oncology | HBEGF | Q99075 | 5 | 140332843 | 140346603 | 0.86 | 0.74 | 1.01 | 6.47E-02 | 0.348 |
| OID20370 | KIT | Cardiometabolic | KIT | P10721 | 4 | 54657267 | 54740783 | 0.86 | 0.73 | 1.01 | 6.49E-02 | 0.348 |
| OID20554 | ARHGEF12 | Inflammation | ARHGEF12 | Q9NZN5 | 11 | 120336413 | 120489937 | 0.86 | 0.73 | 1.01 | 6.55E-02 | 0.349 |
| OID30308 | DTYMK | Cardiometabolic_II | DTYMK | P23919 | 2 | 241675747 | 241686944 | 0.86 | 0.73 | 1.01 | 6.55E-02 | 0.349 |
| OID30570 | ITGA2 | Inflammation_II | ITGA2 | P17301 | 5 | 52989340 | 53094779 | 0.85 | 0.72 | 1.01 | 6.55E-02 | 0.349 |
| OID31215 | PITHD1 | Oncology_II | PITHD1 | Q9GZP4 | 1 | 23778418 | 23788232 | 1.14 | 0.99 | 1.30 | 6.52E-02 | 0.349 |
| OID31522 | ACOT13 | Oncology_II | ACOT13 | Q9NPJ3 | 6 | 24667035 | 24705065 | 0.86 | 0.73 | 1.01 | 6.57E-02 | 0.349 |
| OID20136 | NTRK2 | Cardiometabolic | NTRK2 | Q16620 | 9 | 84668375 | 85095751 | 0.85 | 0.72 | 1.01 | 6.57E-02 | 0.349 |
| OID30292 | BNIP2 | Cardiometabolic_II | BNIP2 | Q12982 | 15 | 59659146 | 59689339 | 0.85 | 0.71 | 1.01 | 6.61E-02 | 0.349 |
| OID20923 | CRADD | Neurology | CRADD | P78560 | 12 | 93677375 | 93894840 | 0.86 | 0.73 | 1.01 | 6.59E-02 | 0.349 |
| OID20487 | IL1RL2 | Inflammation | IL1RL2 | Q9HB29 | 2 | 102187006 | 102240002 | 0.86 | 0.73 | 1.01 | 6.63E-02 | 0.349 |
| OID20855 | ILKAP | Neurology | ILKAP | Q9H0C8 | 2 | 238170402 | 238203708 | 0.85 | 0.71 | 1.01 | 6.67E-02 | 0.349 |
| OID31312 | KIAA1549L | Oncology_II | KIAA1549L | Q6ZVL6 | 11 | 33376108 | 33674102 | 0.85 | 0.71 | 1.01 | 6.66E-02 | 0.349 |
| OID20103 | S100P | Cardiometabolic | S100P | P25815 | 4 | 6693878 | 6697170 | 1.15 | 0.99 | 1.34 | 6.62E-02 | 0.349 |
| OID30331 | SARG | Cardiometabolic_II | C1orf116 | Q9BW04 | 1 | 207018522 | 207032756 | 0.86 | 0.73 | 1.01 | 6.68E-02 | 0.349 |
| OID30310 | SRPX | Cardiometabolic_II | SRPX | P78539 | X | 38149336 | 38220924 | 1.18 | 0.99 | 1.42 | 6.66E-02 | 0.349 |
| OID20993 | TBCB | Neurology | TBCB | Q99426 | 19 | 36114289 | 36125947 | 0.86 | 0.74 | 1.01 | 6.68E-02 | 0.349 |
| OID31272 | VSTM2B | Oncology_II | VSTM2B | A6NLU5 | 19 | 29525994 | 29564551 | 0.82 | 0.66 | 1.01 | 6.72E-02 | 0.350 |
| OID30106 | CPTP | Cardiometabolic_II | CPTP | Q5TA50 | 1 | 1324756 | 1328896 | 0.84 | 0.70 | 1.01 | 6.76E-02 | 0.350 |
| OID20683 | NCK2 | Inflammation | NCK2 | O43639 | 2 | 105744912 | 105894274 | 0.86 | 0.73 | 1.01 | 6.74E-02 | 0.350 |
| OID21115 | NUDT5 | Neurology | NUDT5 | Q9UKK9 | 10 | 12165330 | 12196144 | 0.85 | 0.72 | 1.01 | 6.76E-02 | 0.350 |
| OID30398 | SBSN | Cardiometabolic_II | SBSN | Q6UWP8 | 19 | 35523367 | 35528351 | 1.15 | 0.99 | 1.34 | 6.77E-02 | 0.350 |
| OID21091 | GP6 | Neurology | GP6 | Q9HCN6 | 19 | 55013705 | 55038264 | 0.86 | 0.74 | 1.01 | 6.80E-02 | 0.351 |
| OID20958 | CC2D1A | Neurology | CC2D1A | Q6P1N0 | 19 | 13906201 | 13930879 | 0.85 | 0.72 | 1.01 | 6.82E-02 | 0.352 |
| OID20441 | CEP164 | Inflammation | CEP164 | Q9UPV0 | 11 | 117314557 | 117413266 | 0.83 | 0.69 | 1.01 | 6.84E-02 | 0.352 |
| OID30847 | BRME1 | Neurology_II | BRME1 | Q0VDD7 | 19 | 13882348 | 13906452 | 1.10 | 0.99 | 1.22 | 6.86E-02 | 0.353 |
| OID30837 | BOLA1 | Neurology_II | BOLA1 | Q9Y3E2 | 1 | 149887890 | 149900798 | 0.84 | 0.69 | 1.01 | 6.88E-02 | 0.353 |
| OID20553 | NT5C3A | Inflammation | NT5C3A | Q9H0P0 | 7 | 33014130 | 33062796 | 0.86 | 0.74 | 1.01 | 6.90E-02 | 0.354 |
| OID21036 | ANXA3 | Neurology | ANXA3 | P12429 | 4 | 78551747 | 78610451 | 0.86 | 0.73 | 1.01 | 6.94E-02 | 0.354 |
| OID20378 | CD14 | Cardiometabolic | CD14 | P08571 | 5 | 140631728 | 140633701 | 1.15 | 0.99 | 1.34 | 6.93E-02 | 0.354 |
| OID30297 | PTGR1 | Cardiometabolic_II | PTGR1 | Q14914 | 9 | 111549722 | 111599893 | 0.86 | 0.72 | 1.01 | 6.94E-02 | 0.354 |
| OID30787 | FN1 | Inflammation_II | FN1 | P02751 | 2 | 215360440 | 215436073 | 0.85 | 0.72 | 1.01 | 6.99E-02 | 0.355 |
| OID20950 | RHOC | Neurology | RHOC | P08134 | 1 | 112701127 | 112707434 | 0.86 | 0.73 | 1.01 | 7.01E-02 | 0.356 |
| OID30236 | AAMDC | Cardiometabolic_II | AAMDC | Q9H7C9 | 11 | 77821109 | 77918432 | 0.86 | 0.73 | 1.01 | 7.03E-02 | 0.356 |
| OID31073 | MORC3 | Neurology_II | MORC3 | Q14149 | 21 | 36320189 | 36386148 | 0.83 | 0.69 | 1.02 | 7.05E-02 | 0.357 |
| OID30067 | ADRA2A | Cardiometabolic_II | ADRA2A | P08913 | 10 | 111077029 | 111080907 | 0.83 | 0.68 | 1.02 | 7.07E-02 | 0.357 |
| OID30859 | DGCR6 | Neurology_II | DGCR6 | Q14129 | 22 | 18906028 | 18914238 | 0.83 | 0.67 | 1.02 | 7.08E-02 | 0.357 |
| OID21073 | MIF | Neurology | MIF | P14174 | 22 | 23894383 | 23895227 | 0.86 | 0.74 | 1.01 | 7.13E-02 | 0.359 |
| OID30174 | GP1BB | Cardiometabolic_II | GP1BB | P13224 | 22 | 19723539 | 19724771 | 0.86 | 0.73 | 1.01 | 7.15E-02 | 0.359 |
| OID30860 | ITPRIP | Neurology_II | ITPRIP | Q8IWB1 | 10 | 104309698 | 104338465 | 0.83 | 0.68 | 1.02 | 7.18E-02 | 0.359 |
| OID20821 | LYPD1 | Neurology | LYPD1 | Q8N2G4 | 2 | 132643286 | 132671579 | 0.82 | 0.66 | 1.02 | 7.19E-02 | 0.359 |
| OID31468 | MAP2K1 | Oncology_II | MAP2K1 | Q02750 | 15 | 66386837 | 66491656 | 0.85 | 0.72 | 1.01 | 7.20E-02 | 0.359 |
| OID31462 | TMED8 | Oncology_II | TMED8 | Q6PL24 | 14 | 77335029 | 77377094 | 0.86 | 0.73 | 1.01 | 7.20E-02 | 0.359 |
| OID21078 | AHSP | Neurology | AHSP | Q9NZD4 | 16 | 31527900 | 31528803 | 0.85 | 0.71 | 1.02 | 7.33E-02 | 0.360 |
| OID20558 | BCR | Inflammation | BCR | P11274 | 22 | 23179704 | 23318037 | 0.86 | 0.73 | 1.01 | 7.45E-02 | 0.360 |
| OID20225 | CA13 | Cardiometabolic | CA13 | Q8N1Q1 | 8 | 85220587 | 85284073 | 0.87 | 0.74 | 1.01 | 7.45E-02 | 0.360 |
| OID21401 | CA14 | Oncology | CA14 | Q9ULX7 | 1 | 150257251 | 150265078 | 0.85 | 0.71 | 1.02 | 7.34E-02 | 0.360 |
| OID21265 | CD28 | Oncology | CD28 | P10747 | 2 | 203706475 | 203738912 | 0.86 | 0.73 | 1.01 | 7.48E-02 | 0.360 |
| OID20667 | CDSN | Inflammation | CDSN | Q15517 | 6 | 31115087 | 31120446 | 1.15 | 0.99 | 1.34 | 7.42E-02 | 0.360 |
| OID30327 | CHMP6 | Cardiometabolic_II | CHMP6 | Q96FZ7 | 17 | 80991598 | 81009517 | 0.86 | 0.74 | 1.01 | 7.51E-02 | 0.360 |
| OID30319 | CRYBB1 | Cardiometabolic_II | CRYBB1 | P53674 | 22 | 26599278 | 26618027 | 0.86 | 0.72 | 1.02 | 7.51E-02 | 0.360 |
| OID30431 | DDX4 | Inflammation_II | DDX4 | Q9NQI0 | 5 | 55738017 | 55817157 | 0.84 | 0.69 | 1.02 | 7.47E-02 | 0.360 |
| OID20295 | DLK1 | Cardiometabolic | DLK1 | P80370 | 14 | 100725705 | 100738224 | 0.86 | 0.73 | 1.02 | 7.49E-02 | 0.360 |
| OID31394 | DUSP13 | Oncology_II | DUSP13 | Q6B8I1 | 10 | 75094432 | 75109221 | 0.85 | 0.72 | 1.02 | 7.38E-02 | 0.360 |
| OID20918 | EBAG9 | Neurology | EBAG9 | O00559 | 8 | 109539711 | 109565996 | 0.86 | 0.73 | 1.01 | 7.28E-02 | 0.360 |
| OID20710 | GLOD4 | Inflammation | GLOD4 | Q9HC38 | 17 | 757097 | 782294 | 0.86 | 0.73 | 1.01 | 7.35E-02 | 0.360 |
| OID20615 | HPCAL1 | Inflammation | HPCAL1 | P37235 | 2 | 10302889 | 10427617 | 0.87 | 0.74 | 1.01 | 7.39E-02 | 0.360 |
| OID21063 | MSR1 | Neurology | MSR1 | P21757 | 8 | 16107878 | 16567490 | 1.17 | 0.98 | 1.40 | 7.48E-02 | 0.360 |
| OID31035 | NARS1 | Neurology_II | NARS1 | O43776 | 18 | 57600656 | 57622213 | 0.87 | 0.75 | 1.01 | 7.40E-02 | 0.360 |
| OID30055 | NOP56 | Cardiometabolic_II | NOP56 | O00567 | 20 | 2652593 | 2658393 | 0.83 | 0.67 | 1.02 | 7.38E-02 | 0.360 |
| OID20623 | NUDC | Inflammation | NUDC | Q9Y266 | 1 | 26900238 | 26946871 | 0.86 | 0.74 | 1.01 | 7.44E-02 | 0.360 |
| OID31300 | OGT | Oncology_II | OGT | O15294 | X | 71533087 | 71575892 | 0.84 | 0.69 | 1.02 | 7.25E-02 | 0.360 |
| OID30688 | PEPD | Inflammation_II | PEPD | P12955 | 19 | 33386950 | 33521823 | 0.86 | 0.74 | 1.01 | 7.29E-02 | 0.360 |
| OID30234 | RECK | Cardiometabolic_II | RECK | O95980 | 9 | 36036913 | 36124455 | 0.86 | 0.72 | 1.01 | 7.30E-02 | 0.360 |
| OID21104 | THBS2 | Neurology | THBS2 | P35442 | 6 | 169215780 | 169254050 | 1.15 | 0.99 | 1.35 | 7.26E-02 | 0.360 |
| OID20180 | TINAGL1 | Cardiometabolic | TINAGL1 | Q9GZM7 | 1 | 31576485 | 31587686 | 1.16 | 0.99 | 1.38 | 7.31E-02 | 0.360 |
| OID30554 | VEGFB | Inflammation_II | VEGFB | P49765 | 11 | 64234584 | 64239264 | 1.15 | 0.99 | 1.34 | 7.33E-02 | 0.360 |
| OID31379 | AHSA1 | Oncology_II | AHSA1 | O95433 | 14 | 77457870 | 77469472 | 0.84 | 0.69 | 1.02 | 7.53E-02 | 0.360 |
| OID31224 | GRSF1 | Oncology_II | GRSF1 | Q12849 | 4 | 70815783 | 70839897 | 0.85 | 0.72 | 1.02 | 7.56E-02 | 0.361 |
| OID30166 | ARL13B | Cardiometabolic_II | ARL13B | Q3SXY8 | 3 | 93980139 | 94055678 | 0.85 | 0.72 | 1.02 | 7.57E-02 | 0.361 |
| OID21495 | TXNDC15 | Oncology | TXNDC15 | Q96J42 | 5 | 134874371 | 134901635 | 1.15 | 0.99 | 1.35 | 7.60E-02 | 0.362 |
| OID20517 | BID | Inflammation | BID | P55957 | 22 | 17734138 | 17774770 | 0.85 | 0.72 | 1.02 | 7.63E-02 | 0.363 |
| OID20808 | CDHR1 | Neurology | CDHR1 | Q96JP9 | 10 | 84194537 | 84219621 | 0.85 | 0.71 | 1.02 | 7.64E-02 | 0.363 |
| OID21324 | SRP14 | Oncology | SRP14 | P37108 | 15 | 40035690 | 40039181 | 0.85 | 0.71 | 1.02 | 7.67E-02 | 0.363 |

| | | | | | | | | | | | | |
|---|---|---|---|---|---|---|---|---|---|---|---|---|
| OID30113 | SART1 | Cardiometabolic_II | SART1 | O43290 | 11 | 65961728 | 65980137 | 0.80 | 0.63 | 1.02 | 7.68E-02 | 0.364 |
| OID21294 | ADGRG1 | Oncology | ADGRG1 | Q9Y653 | 16 | 57610652 | 57665580 | 1.13 | 0.99 | 1.30 | 7.79E-02 | 0.365 |
| OID21363 | DNAJB1 | Oncology | DNAJB1 | P25685 | 19 | 14514769 | 14560391 | 0.86 | 0.73 | 1.02 | 7.78E-02 | 0.365 |
| OID31261 | GADD45B | Oncology_II | GADD45B | O75293 | 19 | 2476122 | 2478259 | 0.81 | 0.64 | 1.02 | 7.76E-02 | 0.365 |
| OID20485 | IRAK1 | Inflammation | IRAK1 | P51617 | X | 154010506 | 154019902 | 0.86 | 0.72 | 1.02 | 7.77E-02 | 0.365 |
| OID30452 | RAP1A | Inflammation_II | RAP1A | P62834 | 1 | 111542218 | 111716691 | 0.83 | 0.67 | 1.02 | 7.80E-02 | 0.365 |
| OID20258 | REG3A | Cardiometabolic | REG3A | Q06141 | 2 | 79157003 | 79159753 | 1.15 | 0.98 | 1.35 | 7.76E-02 | 0.365 |
| OID21165 | SFTPA1 | Oncology | SFTPA1 | Q8IWL2 | 10 | 79610939 | 79615455 | 0.84 | 0.70 | 1.02 | 7.74E-02 | 0.365 |
| OID21476 | ATOX1 | Oncology | ATOX1 | O00244 | 5 | 151742316 | 151772532 | 0.86 | 0.73 | 1.02 | 7.82E-02 | 0.365 |
| OID21359 | APBB1IP | Oncology | APBB1IP | Q7Z5R6 | 10 | 26438341 | 26567803 | 0.84 | 0.68 | 1.02 | 7.93E-02 | 0.366 |
| OID21357 | AREG | Oncology | AREG | P15514 | 4 | 74445136 | 74455005 | 1.15 | 0.98 | 1.34 | 7.95E-02 | 0.366 |
| OID31454 | BCL2 | Oncology_II | BCL2 | P10415 | 18 | 63123346 | 63320128 | 0.83 | 0.67 | 1.02 | 8.05E-02 | 0.366 |
| OID30279 | BPIFA2 | Cardiometabolic_II | BPIFA2 | Q96DR5 | 20 | 33161768 | 33181412 | 0.86 | 0.72 | 1.02 | 8.23E-02 | 0.366 |
| OID21061 | CLEC1B | Neurology | CLEC1B | Q9P126 | 12 | 9985642 | 10013424 | 0.87 | 0.74 | 1.02 | 8.09E-02 | 0.366 |
| OID30286 | COL2A1 | Cardiometabolic_II | COL2A1 | P02458 | 12 | 47972967 | 48004554 | 0.86 | 0.73 | 1.02 | 8.30E-02 | 0.366 |
| OID20293 | CST6 | Cardiometabolic | CST6 | Q15828 | 11 | 66012008 | 66013505 | 1.16 | 0.98 | 1.36 | 8.10E-02 | 0.366 |
| OID20207 | CXCL5 | Cardiometabolic | CXCL5 | P42830 | 4 | 73995642 | 73998677 | 0.86 | 0.73 | 1.02 | 8.01E-02 | 0.366 |
| OID31104 | DNAJC21 | Neurology_II | DNAJC21 | Q5F1R6 | 5 | 34929559 | 34958964 | 0.86 | 0.72 | 1.02 | 8.27E-02 | 0.366 |
| OID31472 | DNAJC9 | Oncology_II | DNAJC9 | Q8WXX5 | 10 | 73183362 | 73247255 | 0.86 | 0.73 | 1.02 | 8.20E-02 | 0.366 |
| OID31458 | DNM1 | Oncology_II | DNM1 | Q05193 | 9 | 128191655 | 128255248 | 0.86 | 0.74 | 1.02 | 7.95E-02 | 0.366 |
| OID21460 | DSG3 | Oncology | DSG3 | P32926 | 18 | 31447741 | 31478702 | 1.16 | 0.98 | 1.38 | 8.00E-02 | 0.366 |
| OID30335 | EEF1D | Cardiometabolic_II | EEF1D | P29692 | 8 | 143579697 | 143599541 | 0.86 | 0.73 | 1.02 | 8.02E-02 | 0.366 |
| OID31029 | ELAC1 | Neurology_II | ELAC1 | Q9H777 | 18 | 50967991 | 50988121 | 0.87 | 0.74 | 1.02 | 8.20E-02 | 0.366 |
| OID30209 | ENO3 | Cardiometabolic_II | ENO3 | P13929 | 17 | 4948092 | 4957131 | 0.84 | 0.70 | 1.02 | 7.95E-02 | 0.366 |
| OID30578 | ERP29 | Inflammation_II | ERP29 | P30040 | 12 | 112013348 | 112023449 | 0.86 | 0.74 | 1.02 | 7.94E-02 | 0.366 |
| OID20151 | FADD | Cardiometabolic | FADD | Q13158 | 11 | 70203296 | 70207390 | 0.86 | 0.73 | 1.02 | 8.28E-02 | 0.366 |
| OID20639 | FCRL2 | Inflammation | FCRL2 | Q96LA5 | 1 | 157745733 | 157777132 | 0.87 | 0.74 | 1.02 | 8.27E-02 | 0.366 |
| OID21192 | FEN1 | Oncology | FEN1 | P39748 | 11 | 61792911 | 61797238 | 0.84 | 0.69 | 1.02 | 8.27E-02 | 0.366 |
| OID30253 | GATD3 | Cardiometabolic_II | GATD3 | P0DPI2 | 21 | 44133610 | 44210114 | 0.86 | 0.72 | 1.02 | 8.24E-02 | 0.366 |
| OID30139 | GBP6 | Cardiometabolic_II | GBP6 | Q6ZN66 | 1 | 89364059 | 89388160 | 0.83 | 0.67 | 1.02 | 8.04E-02 | 0.366 |
| OID20107 | GLO1 | Cardiometabolic | GLO1 | Q04760 | 6 | 38675925 | 38703145 | 0.86 | 0.72 | 1.02 | 8.18E-02 | 0.366 |
| OID31013 | GPR158 | Neurology_II | GPR158 | Q5T848 | 10 | 25174802 | 25602229 | 0.85 | 0.71 | 1.02 | 8.08E-02 | 0.366 |
| OID30674 | NRGN | Inflammation_II | NRGN | Q92686 | 11 | 124739942 | 124747210 | 0.86 | 0.73 | 1.02 | 8.28E-02 | 0.366 |
| OID30610 | PCBD1 | Inflammation_II | PCBD1 | P61457 | 10 | 70882280 | 70888565 | 0.84 | 0.70 | 1.02 | 7.93E-02 | 0.366 |
| OID20764 | PLAUR | Inflammation | PLAUR | Q03405 | 19 | 43646095 | 43670547 | 1.17 | 0.98 | 1.39 | 7.96E-02 | 0.366 |
| OID30125 | PMM2 | Cardiometabolic_II | PMM2 | O15305 | 16 | 8788823 | 8862534 | 0.86 | 0.73 | 1.02 | 7.90E-02 | 0.366 |
| OID30647 | RABEP1 | Inflammation_II | RABEP1 | Q15276 | 17 | 5282265 | 5386340 | 0.86 | 0.73 | 1.02 | 8.29E-02 | 0.366 |
| OID21218 | SFTPA2 | Oncology | SFTPA2 | Q8IWL1 | 10 | 79555852 | 79560407 | 1.15 | 0.98 | 1.35 | 8.16E-02 | 0.366 |
| OID20302 | ST6GAL1 | Cardiometabolic | ST6GAL1 | P15907 | 3 | 186930325 | 187078553 | 0.86 | 0.72 | 1.02 | 8.10E-02 | 0.366 |
| OID20388 | TFPI | Cardiometabolic | TFPI | P10646 | 2 | 187464230 | 187565760 | 0.86 | 0.72 | 1.02 | 8.01E-02 | 0.366 |
| OID31143 | TIMM8A | Neurology_II | TIMM8A | O60220 | X | 101345661 | 101348742 | 0.86 | 0.73 | 1.02 | 8.09E-02 | 0.366 |
| OID31335 | TRDMT1 | Oncology_II | TRDMT1 | O14717 | 10 | 17137336 | 17202054 | 0.85 | 0.71 | 1.02 | 8.02E-02 | 0.366 |
| OID21120 | TREML2 | Neurology | TREML2 | Q5T2D2 | 6 | 41189749 | 41201149 | 0.86 | 0.73 | 1.02 | 8.20E-02 | 0.366 |
| OID20526 | TRIM5 | Inflammation | TRIM5 | Q9C035 | 11 | 5663195 | 5938619 | 0.86 | 0.73 | 1.02 | 8.14E-02 | 0.366 |
| OID30692 | VASP | Inflammation_II | VASP | P50552 | 19 | 45506579 | 45526989 | 0.86 | 0.73 | 1.02 | 7.86E-02 | 0.366 |
| OID30795 | VAV3 | Neurology_II | VAV3 | Q9UKW4 | 1 | 107571161 | 107965180 | 0.86 | 0.73 | 1.02 | 8.29E-02 | 0.366 |
| OID31011 | PPP1R14A | Neurology_II | PPP1R14A | Q96A00 | 19 | 38251237 | 38256532 | 0.86 | 0.72 | 1.02 | 8.31E-02 | 0.366 |
| OID30485 | CD36 | Inflammation_II | CD36 | P16671 | 7 | 80369575 | 80679277 | 0.86 | 0.72 | 1.02 | 8.39E-02 | 0.367 |
| OID21143 | DBI | Neurology | DBI | P07108 | 2 | 119366924 | 119372550 | 0.86 | 0.73 | 1.02 | 8.38E-02 | 0.367 |
| OID21210 | LSM1 | Oncology | LSM1 | O15116 | 8 | 38163335 | 38176730 | 0.85 | 0.70 | 1.02 | 8.37E-02 | 0.367 |
| OID20143 | PLIN3 | Cardiometabolic | PLIN3 | O60664 | 19 | 4838341 | 4867694 | 0.86 | 0.72 | 1.02 | 8.35E-02 | 0.367 |
| OID21194 | RASSF2 | Oncology | RASSF2 | P50749 | 20 | 4780023 | 4823608 | 0.84 | 0.69 | 1.02 | 8.38E-02 | 0.367 |
| OID20114 | BAG6 | Cardiometabolic | BAG6 | P46379 | 6 | 31639028 | 31652705 | 0.86 | 0.73 | 1.02 | 8.46E-02 | 0.369 |
| OID21217 | BIRC2 | Oncology | BIRC2 | Q13490 | 11 | 102347211 | 102378670 | 0.86 | 0.73 | 1.02 | 8.62E-02 | 0.371 |
| OID21238 | CDC27 | Oncology | CDC27 | P30260 | 17 | 47117703 | 47189422 | 0.86 | 0.72 | 1.02 | 8.59E-02 | 0.371 |
| OID30619 | CRELD1 | Inflammation_II | CRELD1 | Q96HD1 | 3 | 9933793 | 9945413 | 0.86 | 0.73 | 1.02 | 8.55E-02 | 0.371 |
| OID30667 | CSH1 | Inflammation_II | CSH1 | P0DML2 | 17 | 63894909 | 63896661 | 0.85 | 0.71 | 1.02 | 8.53E-02 | 0.371 |
| OID30925 | DYNLT1 | Neurology_II | DYNLT1 | P63172 | 6 | 158636474 | 158644743 | 0.86 | 0.72 | 1.02 | 8.61E-02 | 0.371 |
| OID20240 | LDLR | Cardiometabolic | LDLR | P01130 | 19 | 11089462 | 11133820 | 0.86 | 0.72 | 1.02 | 8.61E-02 | 0.371 |
| OID20552 | MAP2K6 | Inflammation | MAP2K6 | P52564 | 17 | 69414697 | 69553865 | 0.87 | 0.74 | 1.02 | 8.62E-02 | 0.371 |
| OID30983 | NRN1 | Neurology_II | NRN1 | Q9NPD7 | 6 | 5997999 | 6007605 | 0.84 | 0.69 | 1.02 | 8.58E-02 | 0.371 |
| OID20467 | PADI2 | Inflammation | PADI2 | Q9Y2J8 | 1 | 17066761 | 17119451 | 1.13 | 0.98 | 1.29 | 8.54E-02 | 0.371 |
| OID30698 | C9 | Inflammation_II | C9 | P02748 | 5 | 39284140 | 39371324 | 1.15 | 0.98 | 1.36 | 8.64E-02 | 0.372 |
| OID20868 | TST | Neurology | TST | Q16762 | 22 | 37010859 | 37020183 | 0.86 | 0.73 | 1.02 | 8.66E-02 | 0.372 |
| OID20051 | HNRNPK | Cardiometabolic | HNRNPK | P61978 | 9 | 83968083 | 83980616 | 0.86 | 0.73 | 1.02 | 8.69E-02 | 0.372 |
| OID20672 | MMP1 | Inflammation | MMP1 | P03956 | 11 | 102789401 | 102798160 | 0.87 | 0.74 | 1.02 | 8.70E-02 | 0.372 |
| OID20944 | GPC5 | Neurology | GPC5 | P78333 | 13 | 91398621 | 92873682 | 0.87 | 0.74 | 1.02 | 8.75E-02 | 0.373 |
| OID31027 | RNASEH2A | Neurology_II | RNASEH2A | O75792 | 19 | 12806584 | 12813640 | 0.84 | 0.69 | 1.03 | 8.75E-02 | 0.373 |
| OID21335 | DRG2 | Oncology | DRG2 | P55039 | 17 | 18087892 | 18107970 | 0.86 | 0.72 | 1.02 | 8.78E-02 | 0.374 |
| OID31134 | ESYT2 | Neurology_II | ESYT2 | A0FGR8 | 7 | 158730995 | 158830253 | 0.86 | 0.73 | 1.02 | 8.80E-02 | 0.374 |
| OID31346 | RAB44 | Oncology_II | RAB44 | Q7Z6P3 | 6 | 36697826 | 36733184 | 0.82 | 0.64 | 1.03 | 8.80E-02 | 0.374 |
| OID31039 | CACNB3 | Neurology_II | CACNB3 | P54284 | 12 | 48813794 | 48828941 | 0.87 | 0.74 | 1.02 | 8.97E-02 | 0.374 |
| OID21171 | CEACAM5 | Oncology | CEACAM5 | P06731 | 19 | 41708585 | 41730433 | 1.14 | 0.98 | 1.31 | 8.98E-02 | 0.374 |
| OID20399 | CHI3L1 | Cardiometabolic | CHI3L1 | P36222 | 1 | 203178931 | 203186704 | 1.14 | 0.98 | 1.33 | 8.94E-02 | 0.374 |
| OID30222 | COMMD1 | Cardiometabolic_II | COMMD1 | Q8N668 | 2 | 61888724 | 62147247 | 0.86 | 0.73 | 1.02 | 8.90E-02 | 0.374 |
| OID20872 | FHIT | Neurology | FHIT | P49789 | 3 | 59747277 | 61251459 | 0.87 | 0.74 | 1.02 | 8.94E-02 | 0.374 |
| OID30112 | IGF2BP3 | Cardiometabolic_II | IGF2BP3 | O00425 | 7 | 23310209 | 23470491 | 0.84 | 0.69 | 1.03 | 8.94E-02 | 0.374 |
| OID30907 | LRP2BP | Neurology_II | LRP2BP | Q9P2M1 | 4 | 185363872 | 185395924 | 0.85 | 0.71 | 1.02 | 8.89E-02 | 0.374 |
| OID31212 | PFDN4 | Oncology_II | PFDN4 | Q9NQP4 | 20 | 54208087 | 54228052 | 0.85 | 0.70 | 1.03 | 8.95E-02 | 0.374 |
| OID30104 | SCGB2A2 | Cardiometabolic_II | SCGB2A2 | Q13296 | 11 | 62270158 | 62273160 | 0.85 | 0.70 | 1.03 | 9.00E-02 | 0.374 |
| OID31353 | SMAD2 | Oncology_II | SMAD2 | Q15796 | 18 | 47808957 | 47931146 | 0.86 | 0.73 | 1.02 | 8.99E-02 | 0.374 |
| OID20068 | TIA1 | Cardiometabolic | TIA1 | P31483 | 2 | 70209444 | 70248660 | 0.87 | 0.73 | 1.02 | 8.85E-02 | 0.374 |
| OID31411 | TMEM106A | Oncology_II | TMEM106A | Q96A25 | 17 | 43211835 | 43220041 | 0.86 | 0.73 | 1.02 | 8.90E-02 | 0.374 |
| OID31428 | USP25 | Oncology_II | USP25 | Q9UHP3 | 21 | 15729982 | 15880064 | 0.87 | 0.74 | 1.02 | 8.85E-02 | 0.374 |
| OID31002 | VWC2L | Neurology_II | VWC2L | B2RUY7 | 2 | 214411054 | 214578976 | 0.83 | 0.67 | 1.03 | 8.88E-02 | 0.374 |
| OID31366 | WFDC1 | Oncology_II | WFDC1 | Q9HC57 | 16 | 84294846 | 84329844 | 0.85 | 0.71 | 1.03 | 8.99E-02 | 0.374 |
| OID21015 | RWDD1 | Neurology | RWDD1 | Q9H446 | 6 | 116571409 | 116597675 | 0.87 | 0.74 | 1.02 | 9.07E-02 | 0.377 |
| OID30498 | BCL2L15 | Inflammation_II | BCL2L15 | Q5TBC7 | 1 | 113876816 | 113887581 | 0.84 | 0.69 | 1.03 | 9.11E-02 | 0.377 |
| OID30463 | RELB | Inflammation_II | RELB | Q01201 | 19 | 45001461 | 45038192 | 0.84 | 0.69 | 1.03 | 9.12E-02 | 0.377 |
| OID20322 | SPP1 | Cardiometabolic | SPP1 | P10451 | 4 | 87975667 | 87983532 | 1.14 | 0.98 | 1.34 | 9.11E-02 | 0.377 |
| OID20475 | SPRY2 | Inflammation | SPRY2 | O43597 | 13 | 80335976 | 80341126 | 0.87 | 0.73 | 1.02 | 9.12E-02 | 0.377 |
| OID30778 | APCS | Inflammation_II | APCS | P02743 | 1 | 159587826 | 159588865 | 0.85 | 0.70 | 1.03 | 9.14E-02 | 0.377 |
| OID20685 | PLA2G4A | Inflammation | PLA2G4A | P47712 | 1 | 186828949 | 186988981 | 0.87 | 0.74 | 1.02 | 9.17E-02 | 0.377 |
| OID21077 | STIP1 | Neurology | STIP1 | P31948 | 11 | 64185272 | 64204543 | 0.86 | 0.73 | 1.02 | 9.20E-02 | 0.378 |
| OID30642 | ACYP1 | Inflammation_II | ACYP1 | P07311 | 14 | 75053237 | 75069483 | 0.87 | 0.73 | 1.02 | 9.23E-02 | 0.379 |
| OID20774 | DAG1 | Inflammation | DAG1 | Q14118 | 3 | 49468703 | 49535618 | 0.87 | 0.73 | 1.02 | 9.27E-02 | 0.380 |
| OID30239 | ECHS1 | Cardiometabolic_II | ECHS1 | P30084 | 10 | 133362485 | 133373354 | 0.86 | 0.73 | 1.02 | 9.28E-02 | 0.380 |
| OID31477 | SWAP70 | Oncology_II | SWAP70 | Q9UH65 | 11 | 9664077 | 9752993 | 0.85 | 0.70 | 1.03 | 9.32E-02 | 0.381 |
| OID20768 | LTBR | Inflammation | LTBR | P36941 | 12 | 6375045 | 6391571 | 1.14 | 0.98 | 1.32 | 9.37E-02 | 0.381 |
| OID31362 | RAB2B | Oncology_II | RAB2B | Q8WUD1 | 14 | 21459020 | 21476960 | 0.87 | 0.73 | 1.02 | 9.35E-02 | 0.381 |
| OID20415 | REG1A | Cardiometabolic | REG1A | P05451 | 2 | 79120362 | 79123409 | 1.14 | 0.98 | 1.32 | 9.36E-02 | 0.381 |
| OID21202 | ARHGAP1 | Oncology | ARHGAP1 | Q07960 | 11 | 46677080 | 46700619 | 0.86 | 0.73 | 1.03 | 9.55E-02 | 0.382 |
| OID30753 | C1R | Inflammation_II | C1R | P00736 | 12 | 7080214 | 7092540 | 0.83 | 0.67 | 1.03 | 9.50E-02 | 0.382 |

| | | | | | | | | | | | | |
|---|---|---|---|---|---|---|---|---|---|---|---|---|
| OID21319 | CDKN1A | Oncology | CDKN1A | P38936 | 6 | 36676460 | 36687337 | 0.87 | 0.74 | 1.02 | 9.52E-02 | 0.382 |
| OID20330 | FAP | Cardiometabolic | FAP | Q12884 | 2 | 162170684 | 162245151 | 0.88 | 0.76 | 1.02 | 9.45E-02 | 0.382 |
| OID20977 | IPCEF1 | Neurology | IPCEF1 | Q8WWN9 | 6 | 154154496 | 154356803 | 0.87 | 0.74 | 1.02 | 9.48E-02 | 0.382 |
| OID31191 | KIR3DL2 | Oncology_II | KIR3DL2 | P43630 | 19 | 54850443 | 54867207 | 0.84 | 0.68 | 1.03 | 9.44E-02 | 0.382 |
| OID30865 | KRT8 | Neurology_II | KRT8 | P05787 | 12 | 52897187 | 52949954 | 1.12 | 0.98 | 1.27 | 9.40E-02 | 0.382 |
| OID31160 | MORF4L2 | Oncology_II | MORF4L2 | Q15014 | X | 103675496 | 103688158 | 0.85 | 0.71 | 1.03 | 9.57E-02 | 0.382 |
| OID31409 | MTSS2 | Oncology_II | MTSS2 | Q765P7 | 16 | 70661204 | 70686053 | 0.87 | 0.74 | 1.02 | 9.54E-02 | 0.382 |
| OID30313 | MYDGF | Cardiometabolic_II | MYDGF | Q969H8 | 19 | 4641374 | 4670362 | 0.87 | 0.73 | 1.03 | 9.55E-02 | 0.382 |
| OID31412 | NUBP1 | Oncology_II | NUBP1 | P53384 | 16 | 10743786 | 10769351 | 0.86 | 0.72 | 1.03 | 9.42E-02 | 0.382 |
| OID20266 | REG1B | Cardiometabolic | REG1B | P48304 | 2 | 79085023 | 79088019 | 1.14 | 0.98 | 1.34 | 9.53E-02 | 0.382 |
| OID30846 | STEAP4 | Neurology_II | STEAP4 | Q687X5 | 7 | 88270892 | 88306894 | 0.84 | 0.68 | 1.03 | 9.44E-02 | 0.382 |
| OID20953 | TNFSF14 | Neurology | TNFSF14 | O43557 | 19 | 6661253 | 6670588 | 0.86 | 0.73 | 1.03 | 9.55E-02 | 0.382 |
| OID21135 | TXNRD1 | Neurology | TXNRD1 | Q16881 | 12 | 104215779 | 104350307 | 0.86 | 0.72 | 1.03 | 9.48E-02 | 0.382 |
| OID20779 | MPIG6B | Inflammation | MPIG6B | O95866 | 6 | 31718594 | 31726714 | 0.88 | 0.76 | 1.02 | 9.61E-02 | 0.383 |
| OID30186 | PACS2 | Cardiometabolic_II | PACS2 | Q86VP3 | 14 | 105300563 | 105398147 | 0.86 | 0.73 | 1.03 | 9.64E-02 | 0.383 |
| OID30736 | ECM1 | Inflammation_II | ECM1 | Q16610 | 1 | 150508062 | 150513789 | 0.88 | 0.76 | 1.02 | 9.66E-02 | 0.384 |
| OID30370 | ADAMTSL2 | Cardiometabolic_II | ADAMTSL2 | Q86TH1 | 9 | 133532164 | 133575519 | 1.15 | 0.97 | 1.35 | 9.86E-02 | 0.384 |
| OID21463 | ANGPT2 | Oncology | ANGPT2 | O15123 | 8 | 6499632 | 6563409 | 1.15 | 0.97 | 1.37 | 9.86E-02 | 0.384 |
| OID30097 | CD2 | Cardiometabolic_II | CD2 | P06729 | 1 | 116754430 | 116769229 | 0.85 | 0.70 | 1.03 | 9.71E-02 | 0.384 |
| OID31338 | FAM171B | Oncology_II | FAM171B | Q6P995 | 2 | 186694060 | 186765959 | 0.85 | 0.71 | 1.03 | 9.83E-02 | 0.384 |
| OID31128 | GBP1 | Neurology_II | GBP1 | P32455 | 1 | 89051882 | 89065360 | 0.87 | 0.75 | 1.02 | 9.76E-02 | 0.384 |
| OID30309 | MAMDC2 | Cardiometabolic_II | MAMDC2 | Q7Z304 | 9 | 70043848 | 70226972 | 1.14 | 0.98 | 1.33 | 9.73E-02 | 0.384 |
| OID31085 | OSBPL2 | Neurology_II | OSBPL2 | Q9H1P3 | 20 | 62231922 | 62296213 | 0.84 | 0.68 | 1.03 | 9.83E-02 | 0.384 |
| OID30548 | PPM1B | Inflammation_II | PPM1B | O75688 | 2 | 44167969 | 44244384 | 1.12 | 0.98 | 1.28 | 9.86E-02 | 0.384 |
| OID20822 | PXN | Neurology | PXN | P49023 | 12 | 120210439 | 120265771 | 0.86 | 0.71 | 1.03 | 9.84E-02 | 0.384 |
| OID30357 | RAB10 | Cardiometabolic_II | RAB10 | P61026 | 2 | 26034084 | 26137454 | 0.87 | 0.73 | 1.03 | 9.71E-02 | 0.384 |
| OID30757 | SERPINF1 | Inflammation_II | SERPINF1 | P36955 | 17 | 1762029 | 1777565 | 0.85 | 0.71 | 1.03 | 9.77E-02 | 0.384 |
| OID31453 | SSNA1 | Oncology_II | SSNA1 | O43805 | 9 | 137188660 | 137190370 | 0.87 | 0.73 | 1.03 | 9.84E-02 | 0.384 |
| OID20078 | TSPAN1 | Cardiometabolic | TSPAN1 | O60635 | 1 | 46175073 | 46185962 | 0.86 | 0.71 | 1.03 | 9.82E-02 | 0.384 |
| OID30108 | TTN | Cardiometabolic_II | TTN | Q8WZ42 | 2 | 178525989 | 178830802 | 0.85 | 0.71 | 1.03 | 9.84E-02 | 0.384 |
| OID30225 | SCRIB | Cardiometabolic_II/Infla | SCRIB | Q14160 | 8 | 143790920 | 143815773 | 0.86 | 0.72 | 1.03 | 9.91E-02 | 0.385 |
| OID20061 | CTF1 | Cardiometabolic | CTF1 | Q16619 | 16 | 30896614 | 30903547 | 0.87 | 0.73 | 1.03 | 9.91E-02 | 0.385 |
| OID20714 | SHMT1 | Inflammation | SHMT1 | P34896 | 17 | 18327860 | 18363563 | 0.87 | 0.74 | 1.03 | 9.93E-02 | 0.386 |
| OID30920 | GNGT1 | Neurology_II | GNGT1 | P63211 | 7 | 93591573 | 93911265 | 0.86 | 0.71 | 1.03 | 9.97E-02 | 0.386 |
| OID30722 | ITIH1 | Inflammation_II | ITIH1 | P19827 | 3 | 52777595 | 52792068 | 0.87 | 0.74 | 1.03 | 9.97E-02 | 0.386 |
| OID31053 | CSNK2A1 | Neurology_II | CSNK2A1 | P68400 | 20 | 472498 | 543835 | 0.87 | 0.73 | 1.03 | 9.99E-02 | 0.386 |
| OID30244 | AHNAK2 | Cardiometabolic_II | AHNAK2 | Q8IVF2 | 14 | 104937244 | 104978374 | 0.86 | 0.72 | 1.03 | 1.01E-01 | 0.388 |
| OID20930 | CHMP1A | Neurology | CHMP1A | Q9HD42 | 16 | 89640816 | 89657738 | 0.88 | 0.75 | 1.03 | 1.01E-01 | 0.388 |
| OID20319 | EGFR | Cardiometabolic | EGFR | P00533 | 7 | 55019017 | 55211628 | 0.87 | 0.74 | 1.03 | 1.01E-01 | 0.388 |
| OID31354 | NENF | Oncology_II | NENF | Q9UMX5 | 1 | 212432920 | 212446379 | 0.86 | 0.71 | 1.03 | 1.01E-01 | 0.388 |
| OID30206 | PAGR1 | Cardiometabolic_II | PAGR1 | Q9BTK6 | 16 | 29816152 | 29822489 | 0.86 | 0.71 | 1.03 | 1.01E-01 | 0.388 |
| OID20453 | IL20 | Inflammation | IL20 | Q9NYY1 | 1 | 206865623 | 206869223 | 0.86 | 0.71 | 1.03 | 1.01E-01 | 0.389 |
| OID20112 | ITGB1BP2 | Cardiometabolic | ITGB1BP2 | Q9UKP3 | X | 71301750 | 71305371 | 0.88 | 0.75 | 1.03 | 1.01E-01 | 0.389 |
| OID20849 | NXPH1 | Neurology | NXPH1 | P58417 | 7 | 8433609 | 8752961 | 0.87 | 0.74 | 1.03 | 1.02E-01 | 0.389 |
| OID20582 | AXIN1 | Inflammation | AXIN1 | O15169 | 16 | 287440 | 352723 | 0.87 | 0.74 | 1.03 | 1.02E-01 | 0.390 |
| OID21457 | GFRA1 | Oncology | GFRA1 | P56159 | 10 | 116056925 | 116276803 | 1.14 | 0.97 | 1.33 | 1.02E-01 | 0.390 |
| OID21351 | ICOSLG | Oncology | ICOSLG | O75144 | 21 | 44217014 | 44240966 | 1.14 | 0.97 | 1.33 | 1.02E-01 | 0.390 |
| OID31102 | NT5C1A | Neurology_II | NT5C1A | Q9BXI3 | 1 | 39651229 | 39672107 | 0.85 | 0.70 | 1.03 | 1.02E-01 | 0.390 |
| OID31137 | FTCD | Neurology_II | FTCD | O95954 | 21 | 46136160 | 46155579 | 0.86 | 0.72 | 1.03 | 1.03E-01 | 0.392 |
| OID31512 | CYB5R2 | Oncology_II | CYB5R2 | Q6BCY4 | 11 | 7665100 | 7677222 | 0.86 | 0.72 | 1.03 | 1.03E-01 | 0.393 |
| OID21344 | CA12 | Oncology | CA12 | O43570 | 15 | 63321378 | 63381846 | 1.14 | 0.97 | 1.32 | 1.04E-01 | 0.393 |
| OID20084 | CEP43 | Cardiometabolic | CEP43 | O95684 | 6 | 166999317 | 167094789 | 0.87 | 0.74 | 1.03 | 1.04E-01 | 0.393 |
| OID31313 | EFCAB2 | Oncology_II | EFCAB2 | Q5VUJ9 | 1 | 244969682 | 245127164 | 0.86 | 0.72 | 1.03 | 1.04E-01 | 0.393 |
| OID30739 | F12 | Inflammation_II | F12 | P00748 | 5 | 177402133 | 177416583 | 0.88 | 0.75 | 1.03 | 1.04E-01 | 0.393 |
| OID21514 | FURIN | Oncology | FURIN | P09958 | 15 | 90868588 | 90883564 | 0.86 | 0.72 | 1.03 | 1.04E-01 | 0.393 |
| OID21259 | GFER | Oncology | GFER | P55789 | 16 | 1984193 | 1987749 | 0.87 | 0.73 | 1.03 | 1.05E-01 | 0.393 |
| OID20784 | REG4 | Inflammation | REG4 | Q9BYZ8 | 1 | 119794017 | 119811580 | 1.14 | 0.97 | 1.35 | 1.04E-01 | 0.393 |
| OID30358 | TIMM10 | Cardiometabolic_II | TIMM10 | P62072 | 11 | 57528464 | 57530803 | 0.87 | 0.73 | 1.03 | 1.05E-01 | 0.393 |
| OID31082 | KLRK1 | Neurology_II | KLRK1 | P26718 | 12 | 10372353 | 10391874 | 0.88 | 0.75 | 1.03 | 1.05E-01 | 0.394 |
| OID31498 | ENDOU | Oncology_II | ENDOU | P21128 | 12 | 47709734 | 47725567 | 1.14 | 0.97 | 1.35 | 1.05E-01 | 0.395 |
| OID20690 | SIGLEC1 | Inflammation | SIGLEC1 | Q9BZZ2 | 20 | 3686970 | 3712600 | 1.14 | 0.97 | 1.35 | 1.06E-01 | 0.396 |
| OID21245 | CEACAM3 | Oncology | CEACAM3 | P40198 | 19 | 41796587 | 41811554 | 1.12 | 0.98 | 1.28 | 1.06E-01 | 0.396 |
| OID30360 | CMC1 | Cardiometabolic_II | CMC1 | Q7Z7K0 | 3 | 28241584 | 28325142 | 0.87 | 0.74 | 1.03 | 1.06E-01 | 0.396 |
| OID30397 | CSPG4 | Cardiometabolic_II | CSPG4 | Q6UVK1 | 15 | 75674322 | 75712848 | 1.15 | 0.97 | 1.36 | 1.07E-01 | 0.398 |
| OID31195 | SMC3 | Oncology_II | SMC3 | Q9UQE7 | 10 | 110567684 | 110606048 | 0.86 | 0.71 | 1.03 | 1.07E-01 | 0.399 |
| OID20611 | TNFSF10 | Inflammation | TNFSF10 | P50591 | 3 | 172505508 | 172523475 | 0.87 | 0.74 | 1.03 | 1.07E-01 | 0.399 |
| OID21471 | HTRA2 | Oncology | HTRA2 | O43464 | 2 | 74529725 | 74533332 | 0.87 | 0.73 | 1.03 | 1.08E-01 | 0.400 |
| OID30617 | ANXA1 | Inflammation_II | ANXA1 | P04083 | 9 | 73151865 | 73170393 | 0.85 | 0.70 | 1.04 | 1.09E-01 | 0.401 |
| OID20401 | CCL14 | Cardiometabolic | CCL14 | Q16627 | 17 | 35983288 | 35987004 | 0.88 | 0.74 | 1.03 | 1.09E-01 | 0.401 |
| OID21334 | DCTN2 | Oncology | DCTN2 | Q13561 | 12 | 57529633 | 57547224 | 0.87 | 0.73 | 1.03 | 1.09E-01 | 0.401 |
| OID31507 | GIPC3 | Oncology_II | GIPC3 | Q8TF64 | 19 | 3585478 | 3593541 | 0.88 | 0.75 | 1.03 | 1.09E-01 | 0.401 |
| OID30436 | HDAC8 | Inflammation_II | HDAC8 | Q9BY41 | X | 72329516 | 72573101 | 1.12 | 0.98 | 1.28 | 1.09E-01 | 0.401 |
| OID31396 | LAMTOR5 | Oncology_II | LAMTOR5 | O43504 | 1 | 110401249 | 110407942 | 0.86 | 0.71 | 1.04 | 1.10E-01 | 0.401 |
| OID31372 | LARP1 | Oncology_II | LARP1 | Q6PKG0 | 5 | 154682986 | 154817605 | 0.86 | 0.72 | 1.03 | 1.08E-01 | 0.401 |
| OID21041 | LGALS8 | Neurology | LGALS8 | O00214 | 1 | 236518000 | 236552981 | 0.87 | 0.73 | 1.03 | 1.09E-01 | 0.401 |
| OID30257 | RBPMS2 | Cardiometabolic_II | RBPMS2 | Q6ZRY4 | 15 | 64739891 | 64775589 | 0.87 | 0.74 | 1.03 | 1.10E-01 | 0.401 |
| OID31319 | TAB2 | Oncology_II | TAB2 | Q9NYJ8 | 6 | 149218641 | 149411613 | 0.87 | 0.74 | 1.03 | 1.09E-01 | 0.401 |
| OID20234 | TYMP | Cardiometabolic | TYMP | P19971 | 22 | 50525752 | 50530032 | 0.87 | 0.73 | 1.03 | 1.09E-01 | 0.401 |
| OID31181 | UNC79 | Oncology_II | UNC79 | Q9P2D8 | 14 | 93333219 | 93707876 | 0.86 | 0.72 | 1.03 | 1.10E-01 | 0.401 |
| OID20607 | CD84 | Inflammation | CD84 | Q9UIB8 | 1 | 160541095 | 160579516 | 0.88 | 0.75 | 1.03 | 1.10E-01 | 0.402 |
| OID20678 | IFNGR1 | Inflammation | IFNGR1 | P15260 | 6 | 137197485 | 137219449 | 1.14 | 0.97 | 1.33 | 1.10E-01 | 0.402 |
| OID21339 | KAZALD1 | Oncology | KAZALD1 | Q96I82 | 10 | 101061989 | 101068131 | 1.15 | 0.97 | 1.36 | 1.11E-01 | 0.402 |
| OID20108 | PAG1 | Cardiometabolic | PAG1 | Q9NWQ8 | 8 | 80967810 | 81112068 | 0.87 | 0.73 | 1.03 | 1.11E-01 | 0.402 |
| OID20447 | RABGAP1L | Inflammation | RABGAP1L | Q5R372 | 1 | 174159410 | 174995308 | 0.87 | 0.73 | 1.03 | 1.10E-01 | 0.402 |
| OID20964 | TNFRSF6B | Neurology | TNFRSF6B | O95407 | 20 | 63696652 | 63698684 | 1.12 | 0.97 | 1.29 | 1.10E-01 | 0.402 |
| OID20576 | IL4R | Inflammation | IL4R | P24394 | 16 | 27313668 | 27364778 | 1.13 | 0.97 | 1.31 | 1.11E-01 | 0.402 |
| OID20881 | KEL | Neurology | KEL | P23276 | 7 | 142941114 | 142962363 | 1.15 | 0.97 | 1.35 | 1.11E-01 | 0.402 |
| OID30923 | TBR1 | Neurology_II | TBR1 | Q16650 | 2 | 161416297 | 161425870 | 0.84 | 0.69 | 1.04 | 1.12E-01 | 0.404 |
| OID31380 | ALDH2 | Oncology_II | ALDH2 | P05091 | 12 | 111766887 | 111817532 | 0.85 | 0.70 | 1.04 | 1.12E-01 | 0.404 |
| OID20380 | CD46 | Cardiometabolic | CD46 | P15529 | 1 | 207752054 | 207795513 | 0.87 | 0.73 | 1.03 | 1.13E-01 | 0.404 |
| OID31164 | CDK1 | Oncology_II | CDK1 | P06493 | 10 | 60778331 | 60794852 | 1.14 | 0.97 | 1.34 | 1.15E-01 | 0.404 |
| OID30582 | DIPK2B | Inflammation_II | DIPK2B | Q9H7Y0 | X | 45148373 | 45200901 | 0.87 | 0.73 | 1.03 | 1.13E-01 | 0.404 |
| OID21113 | DSG2 | Neurology | DSG2 | Q14126 | 18 | 31498177 | 31549008 | 1.13 | 0.97 | 1.32 | 1.15E-01 | 0.404 |
| OID20689 | ENPP7 | Inflammation | ENPP7 | Q6UWV6 | 17 | 79730943 | 79742219 | 1.14 | 0.97 | 1.34 | 1.14E-01 | 0.404 |
| OID21520 | ERBB2 | Oncology | ERBB2 | P04626 | 17 | 39687914 | 39730426 | 0.87 | 0.73 | 1.03 | 1.13E-01 | 0.404 |
| OID20618 | FKBP1B | Inflammation | FKBP1B | P68106 | 2 | 24049701 | 24063681 | 0.87 | 0.74 | 1.03 | 1.13E-01 | 0.404 |
| OID30631 | GCHFR | Inflammation_II | GCHFR | P30047 | 15 | 40764068 | 40767708 | 0.85 | 0.69 | 1.04 | 1.14E-01 | 0.404 |
| OID30304 | GGCT | Cardiometabolic_II | GGCT | O75223 | 7 | 30496621 | 30504841 | 0.87 | 0.73 | 1.03 | 1.15E-01 | 0.404 |
| OID30663 | GLRX5 | Inflammation_II | GLRX5 | Q86SX6 | 14 | 95533503 | 95544724 | 0.87 | 0.74 | 1.03 | 1.15E-01 | 0.404 |
| OID20619 | IDS | Inflammation | IDS | P22304 | X | 149476988 | 149521096 | 0.89 | 0.76 | 1.03 | 1.15E-01 | 0.404 |
| OID20601 | IL5RA | Inflammation | IL5RA | Q01344 | 3 | 3066324 | 3126613 | 1.14 | 0.97 | 1.34 | 1.15E-01 | 0.404 |
| OID21185 | KIR3DL1 | Oncology | KIR3DL1 | P43629 | 19 | 54816468 | 54830778 | 0.87 | 0.74 | 1.03 | 1.14E-01 | 0.404 |
| OID31032 | LCN15 | Neurology_II | LCN15 | Q6UWW0 | 9 | 136759634 | 136766255 | 1.12 | 0.97 | 1.29 | 1.15E-01 | 0.404 |

| | | | | | | | | | | | | |
|---|---|---|---|---|---|---|---|---|---|---|---|---|
| OID30191 | LRCH4 | Cardiometabolic_II | LRCH4 | O75427 | 7 | 100574011 | 100586129 | 0.87 | 0.73 | 1.03 | 1.15E-01 | 0.404 |
| OID30895 | LRFN2 | Neurology_II | LRFN2 | Q9ULH4 | 6 | 40391591 | 40587364 | 0.85 | 0.70 | 1.04 | 1.15E-01 | 0.404 |
| OID20285 | MEGF9 | Cardiometabolic | MEGF9 | Q9H1U4 | 9 | 120600811 | 120714470 | 0.87 | 0.74 | 1.03 | 1.14E-01 | 0.404 |
| OID30465 | PIKFYVE | Inflammation_II | PIKFYVE | Q9Y2I7 | 2 | 208266255 | 208358746 | 0.86 | 0.71 | 1.04 | 1.15E-01 | 0.404 |
| OID21011 | PVR | Neurology | PVR | P15151 | 19 | 44643798 | 44666162 | 1.14 | 0.97 | 1.33 | 1.14E-01 | 0.404 |
| OID21082 | SIGLEC5 | Neurology | SIGLEC5 | O15389 | 19 | 51611927 | 51645545 | 1.17 | 0.96 | 1.41 | 1.13E-01 | 0.404 |
| OID21031 | SULT1A1 | Neurology | SULT1A1 | P50225 | 16 | 28603944 | 28623625 | 0.88 | 0.75 | 1.03 | 1.13E-01 | 0.404 |
| OID21118 | TMSB10 | Neurology | TMSB10 | P63313 | 2 | 84905656 | 84906671 | 0.87 | 0.73 | 1.03 | 1.14E-01 | 0.404 |
| OID30555 | VAMP8 | Inflammation_II | VAMP8 | Q9BV40 | 2 | 85561562 | 85582031 | 0.87 | 0.74 | 1.03 | 1.15E-01 | 0.404 |
| OID20091 | WASF1 | Cardiometabolic | WASF1 | Q92558 | 6 | 110099819 | 110180004 | 0.88 | 0.74 | 1.03 | 1.15E-01 | 0.404 |
| OID31452 | ZFYVE19 | Oncology_II | ZFYVE19 | Q96K21 | 15 | 40807086 | 40815084 | 0.87 | 0.74 | 1.03 | 1.13E-01 | 0.404 |
| OID30064 | CACNA1H | Cardiometabolic_II | CACNA1H | O95180 | 16 | 1153106 | 1221771 | 1.12 | 0.97 | 1.29 | 1.16E-01 | 0.406 |
| OID21410 | DPY30 | Oncology | DPY30 | Q9C005 | 2 | 31867809 | 32039805 | 0.86 | 0.72 | 1.04 | 1.16E-01 | 0.406 |
| OID31258 | NFKB2 | Oncology_II | NFKB2 | Q00653 | 10 | 102394110 | 102402524 | 0.82 | 0.65 | 1.05 | 1.16E-01 | 0.406 |
| OID30656 | SPART | Inflammation_II | SPART | Q8N0X7 | 13 | 36301638 | 36370180 | 0.87 | 0.74 | 1.03 | 1.16E-01 | 0.406 |
| OID20859 | CARHSP1 | Neurology | CARHSP1 | Q9Y2V2 | 16 | 8852942 | 8869012 | 0.87 | 0.73 | 1.04 | 1.17E-01 | 0.406 |
| OID20903 | DARS1 | Neurology | DARS1 | P14868 | 2 | 135905881 | 135986100 | 0.88 | 0.75 | 1.03 | 1.16E-01 | 0.406 |
| OID31123 | WASHC3 | Neurology_II | WASHC3 | Q9Y3C0 | 12 | 102012840 | 102062149 | 0.87 | 0.73 | 1.04 | 1.17E-01 | 0.408 |
| OID31355 | ENSA | Oncology_II | ENSA | O43768 | 1 | 150600851 | 150629612 | 0.87 | 0.74 | 1.03 | 1.18E-01 | 0.408 |
| OID20547 | CLEC4C | Inflammation | CLEC4C | Q8WTT0 | 12 | 7729383 | 7751605 | 0.88 | 0.75 | 1.03 | 1.19E-01 | 0.409 |
| OID31333 | COL4A4 | Oncology_II | COL4A4 | P53420 | 2 | 227002714 | 227164453 | 0.91 | 0.81 | 1.02 | 1.19E-01 | 0.409 |
| OID31072 | DHRS4L2 | Neurology_II | DHRS4L2 | Q6PKH6 | 14 | 23969874 | 24006408 | 0.86 | 0.71 | 1.04 | 1.18E-01 | 0.409 |
| OID30229 | EHD3 | Cardiometabolic_II | EHD3 | Q9NZN3 | 2 | 31234152 | 31269451 | 0.88 | 0.75 | 1.03 | 1.18E-01 | 0.409 |
| OID30295 | GALNT5 | Cardiometabolic_II | GALNT5 | Q7Z7M9 | 2 | 157257705 | 157318491 | 1.15 | 0.97 | 1.37 | 1.18E-01 | 0.409 |
| OID20551 | GOPC | Inflammation | GOPC | Q9HD26 | 6 | 117560269 | 117602542 | 0.88 | 0.75 | 1.03 | 1.18E-01 | 0.409 |
| OID20097 | GYS1 | Cardiometabolic | GYS1 | P13807 | 19 | 48968130 | 48993310 | 0.88 | 0.75 | 1.03 | 1.18E-01 | 0.409 |
| OID21085 | NID2 | Neurology | NID2 | Q14112 | 14 | 52004809 | 52069059 | 0.88 | 0.75 | 1.03 | 1.19E-01 | 0.409 |
| OID31348 | AP1G2 | Oncology_II | AP1G2 | O75843 | 14 | 23559565 | 23568070 | 0.87 | 0.74 | 1.04 | 1.19E-01 | 0.410 |
| OID30844 | BCAT2 | Neurology_II | BCAT2 | O15382 | 19 | 48795062 | 48811029 | 0.87 | 0.73 | 1.04 | 1.20E-01 | 0.411 |
| OID21000 | EIF4B | Neurology | EIF4B | P23588 | 12 | 53006282 | 53042215 | 0.88 | 0.76 | 1.03 | 1.20E-01 | 0.411 |
| OID20637 | CD22 | Inflammation | CD22 | P20273 | 19 | 35319261 | 35347361 | 0.88 | 0.75 | 1.03 | 1.21E-01 | 0.412 |
| OID20833 | IVD | Neurology | IVD | P26440 | 15 | 40405485 | 40435947 | 0.88 | 0.75 | 1.03 | 1.20E-01 | 0.412 |
| OID31152 | BCL2L1 | Neurology_II | BCL2L1 | Q07817 | 20 | 31664452 | 31723989 | 0.88 | 0.75 | 1.03 | 1.21E-01 | 0.413 |
| OID30643 | NAGPA | Inflammation_II | NAGPA | Q9UK23 | 16 | 5024844 | 5034141 | 0.88 | 0.75 | 1.03 | 1.21E-01 | 0.413 |
| OID31009 | PHACTR2 | Neurology_II | PHACTR2 | O75167 | 6 | 143536845 | 143831185 | 0.88 | 0.75 | 1.03 | 1.21E-01 | 0.413 |
| OID20865 | ATXN10 | Neurology | ATXN10 | Q9UBB4 | 22 | 45671798 | 45845307 | 0.88 | 0.75 | 1.03 | 1.21E-01 | 0.413 |
| OID30182 | PALM3 | Cardiometabolic_II | PALM3 | A6NDB9 | 19 | 14053363 | 14062076 | 0.85 | 0.70 | 1.04 | 1.22E-01 | 0.414 |
| OID30830 | DCUN1D1 | Neurology_II | DCUN1D1 | Q96GG9 | 3 | 182938074 | 182985953 | 0.87 | 0.73 | 1.04 | 1.22E-01 | 0.414 |
| OID20201 | REN | Cardiometabolic | REN | P00797 | 1 | 204154819 | 204190324 | 1.13 | 0.97 | 1.32 | 1.22E-01 | 0.414 |
| OID20524 | DAPP1 | Inflammation | DAPP1 | Q9UN19 | 4 | 99816827 | 99870190 | 0.89 | 0.76 | 1.03 | 1.23E-01 | 0.416 |
| OID20247 | PLAT | Cardiometabolic | PLAT | P00750 | 8 | 42174718 | 42207709 | 0.86 | 0.72 | 1.04 | 1.23E-01 | 0.416 |
| OID21023 | SERPINB6 | Neurology | SERPINB6 | P35237 | 6 | 2948159 | 2972165 | 0.87 | 0.74 | 1.04 | 1.23E-01 | 0.416 |
| OID20681 | DBNL | Inflammation | DBNL | Q9UJU6 | 7 | 44044640 | 44069456 | 0.88 | 0.75 | 1.04 | 1.23E-01 | 0.416 |
| OID30325 | RAB11FIP3 | Cardiometabolic_II | RAB11FIP3 | O75154 | 16 | 425649 | 523011 | 0.88 | 0.75 | 1.03 | 1.23E-01 | 0.416 |
| OID20853 | ING1 | Neurology | ING1 | Q9UK53 | 13 | 110712736 | 110723339 | 0.86 | 0.70 | 1.04 | 1.24E-01 | 0.416 |
| OID31423 | SDCCAG8 | Oncology_II | SDCCAG8 | Q86SQ7 | 1 | 243256034 | 243500091 | 0.88 | 0.75 | 1.03 | 1.23E-01 | 0.416 |
| OID21412 | ANGPTL7 | Oncology | ANGPTL7 | O43827 | 1 | 11189355 | 11195981 | 1.12 | 0.97 | 1.31 | 1.24E-01 | 0.416 |
| OID31064 | OTUD7B | Neurology_II | OTUD7B | Q6GQQ9 | 1 | 149937812 | 150010726 | 0.88 | 0.74 | 1.04 | 1.24E-01 | 0.416 |
| OID30451 | NME1 | Inflammation_II | NME1 | P15531 | 17 | 51153559 | 51162428 | 0.87 | 0.73 | 1.04 | 1.25E-01 | 0.420 |
| OID31273 | VPS28 | Oncology_II | VPS28 | Q9UK41 | 8 | 144423601 | 144428563 | 0.88 | 0.74 | 1.04 | 1.26E-01 | 0.421 |
| OID20579 | DECR1 | Inflammation | DECR1 | Q16698 | 8 | 90001405 | 90053633 | 0.88 | 0.75 | 1.04 | 1.26E-01 | 0.421 |
| OID20892 | PFDN2 | Neurology | PFDN2 | Q9UHV9 | 1 | 161100556 | 161118055 | 0.87 | 0.73 | 1.04 | 1.26E-01 | 0.421 |
| OID21420 | PVALB | Oncology | PVALB | P20472 | 22 | 36800684 | 36819479 | 0.88 | 0.75 | 1.04 | 1.26E-01 | 0.421 |
| OID30775 | CD5L | Inflammation_II | CD5L | O43866 | 1 | 157830911 | 157898256 | 0.88 | 0.75 | 1.04 | 1.26E-01 | 0.422 |
| OID30796 | CDKL5 | Neurology_II | CDKL5 | O76039 | X | 18425583 | 18653629 | 0.86 | 0.71 | 1.04 | 1.27E-01 | 0.424 |
| OID30178 | MRPL24 | Cardiometabolic_II | MRPL24 | Q96A35 | 1 | 156737303 | 156741590 | 0.86 | 0.70 | 1.04 | 1.27E-01 | 0.424 |
| OID30998 | PAFAH1B3 | Neurology_II | PAFAH1B3 | Q15102 | 19 | 42297033 | 42303546 | 0.86 | 0.72 | 1.04 | 1.28E-01 | 0.425 |
| OID20127 | CNST | Cardiometabolic | CNST | Q6PJW8 | 1 | 246566444 | 246668595 | 0.89 | 0.76 | 1.04 | 1.29E-01 | 0.426 |
| OID20406 | DPP4 | Cardiometabolic | DPP4 | P27487 | 2 | 161992245 | 162074394 | 0.88 | 0.75 | 1.04 | 1.28E-01 | 0.426 |
| OID30438 | EP300 | Inflammation_II | EP300 | Q09472 | 22 | 41092592 | 41180077 | 0.86 | 0.72 | 1.04 | 1.29E-01 | 0.426 |
| OID30851 | CBLN1 | Neurology_II | CBLN1 | P23435 | 16 | 49277917 | 49281838 | 0.86 | 0.70 | 1.05 | 1.29E-01 | 0.428 |
| OID20625 | EIF4G1 | Inflammation | EIF4G1 | Q04637 | 3 | 184314495 | 184335358 | 0.88 | 0.75 | 1.04 | 1.30E-01 | 0.429 |
| OID21436 | RILP | Oncology | RILP | Q96NA2 | 17 | 1646145 | 1649866 | 0.88 | 0.74 | 1.04 | 1.30E-01 | 0.429 |
| OID30466 | BABAM1 | Inflammation_II | BABAM1 | Q9NWV8 | 19 | 17267376 | 17281249 | 0.87 | 0.73 | 1.04 | 1.31E-01 | 0.429 |
| OID20965 | LAIR2 | Neurology | LAIR2 | Q6ISS4 | 19 | 54497879 | 54510687 | 1.15 | 0.96 | 1.37 | 1.30E-01 | 0.429 |
| OID21160 | PARK7 | Neurology | PARK7 | Q99497 | 1 | 7954291 | 7985505 | 0.88 | 0.75 | 1.04 | 1.30E-01 | 0.429 |
| OID30437 | PTPN9 | Inflammation_II | PTPN9 | P43378 | 15 | 75463251 | 75579315 | 0.86 | 0.71 | 1.04 | 1.31E-01 | 0.429 |
| OID20650 | VEGFA | Inflammation | VEGFA | P15692 | 6 | 43770184 | 43786487 | 0.88 | 0.74 | 1.04 | 1.31E-01 | 0.431 |
| OID21236 | MED18 | Oncology | MED18 | Q9BUE0 | 1 | 28329002 | 28335965 | 0.88 | 0.75 | 1.04 | 1.33E-01 | 0.434 |
| OID30150 | PYY | Cardiometabolic_II | PYY | P10082 | 17 | 43952733 | 44004469 | 1.13 | 0.96 | 1.31 | 1.33E-01 | 0.434 |
| OID30779 | SERPINF2 | Inflammation_II | SERPINF2 | P08697 | 17 | 1742836 | 1755265 | 0.87 | 0.73 | 1.04 | 1.33E-01 | 0.434 |
| OID21133 | FCER2 | Neurology | FCER2 | P06734 | 19 | 7688758 | 7702146 | 0.88 | 0.75 | 1.04 | 1.33E-01 | 0.434 |
| OID21317 | CNPY4 | Oncology | CNPY4 | Q8N129 | 7 | 100119634 | 100125508 | 0.88 | 0.75 | 1.04 | 1.33E-01 | 0.434 |
| OID30266 | LIPF | Cardiometabolic_II | LIPF | P07098 | 10 | 88664441 | 88678814 | 0.89 | 0.76 | 1.04 | 1.34E-01 | 0.435 |
| OID30140 | PRKD2 | Cardiometabolic_II | PRKD2 | Q9BZL6 | 19 | 46674275 | 46717127 | 0.86 | 0.71 | 1.05 | 1.34E-01 | 0.435 |
| OID30490 | XIAP | Inflammation_II | XIAP | P98170 | X | 123859712 | 123913972 | 0.88 | 0.74 | 1.04 | 1.34E-01 | 0.435 |
| OID30216 | ITPR1 | Cardiometabolic_II | ITPR1 | Q14643 | 3 | 4493345 | 4847506 | 0.87 | 0.73 | 1.04 | 1.34E-01 | 0.435 |
| OID30408 | ITGBL1 | Cardiometabolic_II | ITGBL1 | O95965 | 13 | 101452593 | 101720856 | 1.13 | 0.96 | 1.34 | 1.34E-01 | 0.436 |
| OID30122 | RAB33A | Cardiometabolic_II | RAB33A | Q14088 | X | 130171962 | 130184870 | 0.88 | 0.74 | 1.04 | 1.35E-01 | 0.436 |
| OID31084 | USP28 | Neurology_II | USP28 | Q96RU2 | 11 | 113797874 | 113875572 | 0.86 | 0.70 | 1.05 | 1.34E-01 | 0.436 |
| OID30198 | NUDT10 | Cardiometabolic_II | NUDT10 | Q8NFP7 | X | 51332231 | 51337525 | 0.86 | 0.70 | 1.05 | 1.35E-01 | 0.437 |
| OID31439 | SH3BP1 | Oncology_II | SH3BP1 | Q9Y3L3 | 22 | 37634654 | 37656117 | 0.86 | 0.71 | 1.05 | 1.35E-01 | 0.437 |
| OID21154 | TFF1 | Neurology | TFF1 | P04155 | 21 | 42362282 | 42366535 | 1.13 | 0.96 | 1.32 | 1.36E-01 | 0.437 |
| OID21311 | AARSD1 | Oncology | AARSD1 | Q9BTE6 | 17 | 42950526 | 42964498 | 0.87 | 0.73 | 1.04 | 1.36E-01 | 0.437 |
| OID20952 | TXLNA | Neurology | TXLNA | P40222 | 1 | 32179675 | 32198285 | 0.88 | 0.75 | 1.04 | 1.36E-01 | 0.438 |
| OID30080 | ACSL1 | Cardiometabolic_II | ACSL1 | P33121 | 4 | 184755595 | 184826818 | 0.86 | 0.71 | 1.05 | 1.36E-01 | 0.438 |
| OID30217 | COQ7 | Cardiometabolic_II | COQ7 | Q99807 | 16 | 19067614 | 19080095 | 0.87 | 0.73 | 1.04 | 1.36E-01 | 0.438 |
| OID20067 | VSTM2L | Cardiometabolic | VSTM2L | Q96N03 | 20 | 37903111 | 37945350 | 0.87 | 0.73 | 1.04 | 1.37E-01 | 0.438 |
| OID30164 | GSTM4 | Cardiometabolic_II | GSTM4 | Q03013 | 1 | 109656099 | 109665496 | 0.85 | 0.69 | 1.05 | 1.37E-01 | 0.439 |
| OID21386 | CBLN4 | Oncology | CBLN4 | Q9NTU7 | 20 | 55997357 | 56005519 | 0.89 | 0.75 | 1.04 | 1.38E-01 | 0.441 |
| OID20199 | AHCY | Cardiometabolic | AHCY | P23526 | 20 | 34280268 | 34311802 | 0.88 | 0.75 | 1.04 | 1.38E-01 | 0.441 |
| OID21424 | RTN4R | Oncology | RTN4R | Q9BZR6 | 22 | 20241415 | 20283246 | 0.87 | 0.73 | 1.05 | 1.38E-01 | 0.441 |
| OID30675 | ABO | Inflammation_II | ABO | P16442 | 9 | 133233278 | 133276024 | 1.13 | 0.96 | 1.33 | 1.39E-01 | 0.442 |
| OID21017 | EZR | Neurology | EZR | P15311 | 6 | 158765741 | 158819368 | 0.87 | 0.73 | 1.04 | 1.39E-01 | 0.442 |
| OID21216 | GFOD2 | Oncology | GFOD2 | Q3B7J2 | 16 | 67674531 | 67719339 | 1.12 | 0.96 | 1.30 | 1.39E-01 | 0.442 |
| OID31358 | GPD1 | Oncology_II | GPD1 | P21695 | 12 | 50103982 | 50111313 | 0.87 | 0.71 | 1.05 | 1.39E-01 | 0.442 |
| OID30608 | LEG1 | Inflammation_II | C6orf58 | Q6P5S2 | 6 | 127519455 | 127591820 | 0.88 | 0.74 | 1.04 | 1.39E-01 | 0.442 |
| OID20394 | ADA2 | Cardiometabolic | ADA2 | Q9NZK5 | 22 | 17178790 | 17258235 | 1.13 | 0.96 | 1.32 | 1.40E-01 | 0.442 |
| OID31511 | GPR15L | Oncology_II | C10orf99 | Q6UWK7 | 10 | 84173801 | 84185294 | 1.14 | 0.96 | 1.35 | 1.39E-01 | 0.442 |
| OID30263 | ANXA2 | Cardiometabolic_II | ANXA2 | P07355 | 15 | 60347134 | 60402883 | 0.87 | 0.73 | 1.05 | 1.40E-01 | 0.444 |
| OID21348 | YES1 | Oncology | YES1 | P07947 | 18 | 721588 | 812546 | 0.88 | 0.75 | 1.04 | 1.40E-01 | 0.444 |
| OID20769 | LAMA4 | Inflammation | LAMA4 | Q16363 | 6 | 112107931 | 112254939 | 0.89 | 0.76 | 1.04 | 1.41E-01 | 0.444 |
| OID30989 | MYO6 | Neurology_II | MYO6 | Q9UM54 | 6 | 75749201 | 75919537 | 0.86 | 0.71 | 1.05 | 1.41E-01 | 0.444 |

| | | | | | | | | | | | | |
|---|---|---|---|---|---|---|---|---|---|---|---|---|
| OID31385 | ACRBP | Oncology_II | ACRBP | Q8NEB7 | 12 | 6638075 | 6647433 | 0.87 | 0.73 | 1.05 | 1.41E-01 | 0.445 |
| OID20446 | ARTN | Inflammation | ARTN | Q5T4W7 | 1 | 43933320 | 43937240 | 1.10 | 0.97 | 1.26 | 1.42E-01 | 0.445 |
| OID30754 | BTD | Inflammation_II | BTD | P43251 | 3 | 15601341 | 15722311 | 0.89 | 0.76 | 1.04 | 1.41E-01 | 0.445 |
| OID30883 | C2orf69 | Neurology_II | C2orf69 | Q8N8R5 | 2 | 199911293 | 199955935 | 0.86 | 0.71 | 1.05 | 1.41E-01 | 0.445 |
| OID30345 | CDA | Cardiometabolic_II | CDA | P32320 | 1 | 20589086 | 20618903 | 0.88 | 0.74 | 1.05 | 1.42E-01 | 0.445 |
| OID31479 | CWC15 | Oncology_II | CWC15 | Q9P013 | 11 | 94962620 | 94973586 | 0.86 | 0.70 | 1.05 | 1.42E-01 | 0.445 |
| OID21303 | HS3ST3B1 | Oncology | HS3ST3B1 | Q9Y662 | 17 | 14301081 | 14349404 | 0.89 | 0.76 | 1.04 | 1.42E-01 | 0.445 |
| OID20462 | SIT1 | Inflammation | SIT1 | Q9Y3P8 | 9 | 35649295 | 35650931 | 0.88 | 0.75 | 1.04 | 1.42E-01 | 0.445 |
| OID20502 | SRPK2 | Inflammation | SRPK2 | P78362 | 7 | 105110704 | 105399308 | 0.88 | 0.74 | 1.04 | 1.42E-01 | 0.445 |
| OID20071 | VAMP5 | Cardiometabolic | VAMP5 | O95183 | 2 | 85584431 | 85593406 | 0.87 | 0.72 | 1.05 | 1.42E-01 | 0.445 |
| OID20153 | CXCL8 | Cardiometabolic/Inflam | CXCL8 | P10145 | 4 | 73740541 | 73743716 | 0.87 | 0.73 | 1.05 | 1.44E-01 | 0.445 |
| OID20921 | ABHD14B | Neurology | ABHD14B | Q96IU4 | 3 | 51968510 | 51983409 | 0.88 | 0.75 | 1.04 | 1.44E-01 | 0.445 |
| OID20089 | ADAMTS16 | Cardiometabolic | ADAMTS16 | Q8TE57 | 5 | 5140330 | 5320304 | 0.87 | 0.72 | 1.05 | 1.44E-01 | 0.445 |
| OID21434 | APEX1 | Oncology | APEX1 | P27695 | 14 | 20455191 | 20457772 | 0.88 | 0.74 | 1.04 | 1.44E-01 | 0.445 |
| OID20525 | EDAR | Inflammation | EDAR | Q9UNE0 | 2 | 108894471 | 108989372 | 0.88 | 0.74 | 1.04 | 1.43E-01 | 0.445 |
| OID21371 | GALNT10 | Oncology | GALNT10 | Q86SR1 | 5 | 154190730 | 154420984 | 0.88 | 0.74 | 1.04 | 1.43E-01 | 0.445 |
| OID21175 | HGS | Oncology | HGS | O14964 | 17 | 81683326 | 81703138 | 0.88 | 0.75 | 1.04 | 1.44E-01 | 0.445 |
| OID20065 | HK2 | Cardiometabolic | HK2 | P52789 | 2 | 74834127 | 74893359 | 0.89 | 0.75 | 1.04 | 1.44E-01 | 0.445 |
| OID20907 | IMPA1 | Neurology | IMPA1 | P29218 | 8 | 81656914 | 81686331 | 0.88 | 0.74 | 1.04 | 1.44E-01 | 0.445 |
| OID20863 | NOS1 | Neurology | NOS1 | P29475 | 12 | 117208142 | 117452170 | 1.13 | 0.96 | 1.33 | 1.44E-01 | 0.445 |
| OID30194 | PNMA1 | Cardiometabolic_II | PNMA1 | Q8ND90 | 14 | 73711783 | 73714384 | 0.88 | 0.75 | 1.04 | 1.44E-01 | 0.445 |
| OID30804 | SCN3B | Neurology_II | SCN3B | Q9NY72 | 11 | 123629187 | 123655244 | 0.87 | 0.72 | 1.05 | 1.43E-01 | 0.445 |
| OID30277 | SCN4B | Cardiometabolic_II | SCN4B | Q8IWT1 | 11 | 118133377 | 118152888 | 0.88 | 0.75 | 1.04 | 1.44E-01 | 0.445 |
| OID31425 | STAM | Oncology_II | STAM | Q92783 | 10 | 17644151 | 17716824 | 0.87 | 0.73 | 1.05 | 1.45E-01 | 0.445 |
| OID20967 | TNFRSF10A | Neurology | TNFRSF10A | O00220 | 8 | 23190452 | 23225102 | 1.13 | 0.96 | 1.34 | 1.45E-01 | 0.446 |
| OID30727 | APOE | Inflammation_II | APOE | P02649 | 19 | 44905791 | 44909393 | 0.83 | 0.65 | 1.07 | 1.46E-01 | 0.447 |
| OID20617 | CKAP4 | Inflammation | CKAP4 | Q07065 | 12 | 106237881 | 106304279 | 1.12 | 0.96 | 1.31 | 1.46E-01 | 0.447 |
| OID30807 | CNTF | Neurology_II | CNTF | P26441 | 11 | 58622665 | 58625733 | 0.89 | 0.75 | 1.04 | 1.46E-01 | 0.448 |
| OID21414 | CASP8 | Oncology | CASP8 | Q14790 | 2 | 201233443 | 201287711 | 0.88 | 0.73 | 1.05 | 1.47E-01 | 0.449 |
| OID30533 | GIT1 | Inflammation_II | GIT1 | Q9Y2X7 | 17 | 29573475 | 29594054 | 0.89 | 0.75 | 1.04 | 1.47E-01 | 0.450 |
| OID30938 | PFDN6 | Neurology_II | PFDN6 | O15212 | 6 | 33289302 | 33298401 | 0.87 | 0.72 | 1.05 | 1.47E-01 | 0.450 |
| OID30987 | S100A14 | Neurology_II | S100A14 | Q9HCY8 | 1 | 153614255 | 153616986 | 0.88 | 0.75 | 1.04 | 1.48E-01 | 0.450 |
| OID31374 | SNX2 | Oncology_II | SNX2 | O60749 | 5 | 122775079 | 122834543 | 0.88 | 0.74 | 1.05 | 1.48E-01 | 0.450 |
| OID30782 | TTR | Inflammation_II | TTR | P02766 | 18 | 31557009 | 31598833 | 0.88 | 0.75 | 1.04 | 1.47E-01 | 0.450 |
| OID21280 | ABL1 | Oncology | ABL1 | P00519 | 9 | 130713016 | 130887675 | 0.88 | 0.74 | 1.05 | 1.49E-01 | 0.451 |
| OID21295 | CDKN2D | Oncology | CDKN2D | P55273 | 19 | 10566460 | 10569059 | 0.89 | 0.76 | 1.04 | 1.49E-01 | 0.451 |
| OID31270 | PRUNE2 | Oncology_II | PRUNE2 | Q8WUY3 | 9 | 76611376 | 76906114 | 0.87 | 0.73 | 1.05 | 1.49E-01 | 0.451 |
| OID30355 | ADAMTSL5 | Cardiometabolic_II | ADAMTSL5 | Q6ZMM2 | 19 | 1505022 | 1513604 | 0.89 | 0.75 | 1.04 | 1.50E-01 | 0.453 |
| OID30892 | DDX25 | Neurology_II | DDX25 | Q9UHL0 | 11 | 125903348 | 125943702 | 0.87 | 0.73 | 1.05 | 1.50E-01 | 0.453 |
| OID20503 | FGF2 | Inflammation | FGF2 | P09038 | 4 | 122826708 | 122898236 | 0.89 | 0.75 | 1.04 | 1.50E-01 | 0.453 |
| OID20900 | IL18RAP | Neurology | IL18RAP | O95256 | 2 | 102418689 | 102452565 | 0.88 | 0.74 | 1.05 | 1.50E-01 | 0.453 |
| OID21345 | INPP1 | Oncology | INPP1 | P49441 | 2 | 190343570 | 190371665 | 0.88 | 0.74 | 1.05 | 1.50E-01 | 0.453 |
| OID30274 | MECR | Cardiometabolic_II | MECR | Q9BV79 | 1 | 29192657 | 29230942 | 0.88 | 0.75 | 1.05 | 1.50E-01 | 0.453 |
| OID31197 | SPRING1 | Oncology_II | SPRING1 | Q9H741 | 12 | 116710171 | 116738070 | 0.87 | 0.71 | 1.05 | 1.50E-01 | 0.453 |
| OID31238 | THAP12 | Oncology_II | THAP12 | O43422 | 11 | 76349898 | 76381132 | 0.87 | 0.71 | 1.05 | 1.50E-01 | 0.453 |
| OID20177 | CD2AP | Cardiometabolic | CD2AP | Q9Y5K6 | 6 | 47477789 | 47627263 | 0.89 | 0.75 | 1.04 | 1.51E-01 | 0.453 |
| OID20418 | TIMP1 | Cardiometabolic | TIMP1 | P01033 | X | 47582408 | 47586789 | 0.88 | 0.74 | 1.05 | 1.51E-01 | 0.454 |
| OID30321 | UBE2L6 | Cardiometabolic_II | UBE2L6 | O14933 | 11 | 57551656 | 57568284 | 0.88 | 0.75 | 1.05 | 1.51E-01 | 0.454 |
| OID20115 | MSTN | Cardiometabolic | MSTN | O14793 | 2 | 190055700 | 190062729 | 0.88 | 0.73 | 1.05 | 1.52E-01 | 0.455 |
| OID21231 | GNE | Oncology | GNE | Q9Y223 | 9 | 36214441 | 36277042 | 0.89 | 0.76 | 1.04 | 1.52E-01 | 0.455 |
| OID31196 | BTNL10 | Oncology_II | BTNL10 | A8MVZ5 | 1 | 228510425 | 228512305 | 0.86 | 0.71 | 1.06 | 1.53E-01 | 0.457 |
| OID20121 | CCL27 | Cardiometabolic | CCL27 | Q9Y4X3 | 9 | 34661880 | 34664048 | 0.88 | 0.74 | 1.05 | 1.53E-01 | 0.458 |
| OID30817 | LYPLA2 | Neurology_II | LYPLA2 | O95372 | 1 | 23791145 | 23795539 | 0.88 | 0.73 | 1.05 | 1.54E-01 | 0.459 |
| OID31444 | MCTS1 | Oncology_II | MCTS1 | Q9ULC4 | X | 120594010 | 120621159 | 0.88 | 0.73 | 1.05 | 1.54E-01 | 0.459 |
| OID30689 | RNASE6 | Inflammation_II | RNASE6 | Q93091 | 14 | 20781268 | 20782467 | 1.11 | 0.96 | 1.28 | 1.54E-01 | 0.459 |
| OID30058 | TPM3 | Cardiometabolic_II | TPM3 | P06753 | 1 | 154155308 | 154194648 | 0.89 | 0.75 | 1.05 | 1.54E-01 | 0.459 |
| OID30748 | ADIPOQ | Inflammation_II | ADIPOQ | Q15848 | 3 | 186842704 | 186858463 | 1.15 | 0.95 | 1.38 | 1.57E-01 | 0.460 |
| OID20864 | C2CD2L | Neurology | C2CD2L | O14523 | 11 | 119102198 | 119118544 | 0.88 | 0.74 | 1.05 | 1.55E-01 | 0.460 |
| OID30875 | CDH4 | Neurology_II | CDH4 | P55283 | 20 | 61252261 | 61940617 | 1.11 | 0.96 | 1.28 | 1.55E-01 | 0.460 |
| OID20172 | FCRL1 | Cardiometabolic | FCRL1 | Q96LA6 | 1 | 157794403 | 157820120 | 0.89 | 0.76 | 1.05 | 1.55E-01 | 0.460 |
| OID30839 | FH | Neurology_II | FH | P07954 | 1 | 241497511 | 241519799 | 0.88 | 0.74 | 1.05 | 1.57E-01 | 0.460 |
| OID31447 | HDDC2 | Oncology_II | HDDC2 | Q7Z4H3 | 6 | 125219962 | 125302078 | 0.88 | 0.75 | 1.05 | 1.56E-01 | 0.460 |
| OID30866 | HTR1B | Neurology_II | HTR1B | P28222 | 6 | 77460924 | 77463491 | 0.88 | 0.73 | 1.05 | 1.56E-01 | 0.460 |
| OID20461 | IL3RA | Inflammation | IL3RA | P26951 | X | 1336616 | 1382689 | 0.88 | 0.75 | 1.05 | 1.55E-01 | 0.460 |
| OID21059 | LRPAP1 | Neurology | LRPAP1 | P30533 | 4 | 3503612 | 3532446 | 0.87 | 0.73 | 1.05 | 1.57E-01 | 0.460 |
| OID31315 | MTUS1 | Oncology_II | MTUS1 | Q9ULD2 | 8 | 17643795 | 17801094 | 0.88 | 0.75 | 1.05 | 1.55E-01 | 0.460 |
| OID20912 | PTPRN2 | Neurology | PTPRN2 | Q92932 | 7 | 157539056 | 158587823 | 0.89 | 0.75 | 1.05 | 1.56E-01 | 0.460 |
| OID30968 | SEZ6 | Neurology_II | SEZ6 | Q53EL9 | 17 | 28954901 | 29006440 | 1.11 | 0.96 | 1.27 | 1.56E-01 | 0.460 |
| OID20058 | TCL1B | Cardiometabolic | TCL1B | O95988 | 14 | 95686426 | 95692628 | 0.87 | 0.72 | 1.05 | 1.56E-01 | 0.460 |
| OID20507 | TRAF2 | Inflammation | TRAF2 | Q12933 | 9 | 136881912 | 136926607 | 0.88 | 0.75 | 1.05 | 1.56E-01 | 0.460 |
| OID30539 | ERMAP | Inflammation_II | ERMAP | Q96PL5 | 1 | 42817122 | 42844991 | 0.89 | 0.75 | 1.05 | 1.57E-01 | 0.461 |
| OID21224 | NDUFS6 | Oncology | NDUFS6 | O75380 | 5 | 1801407 | 1816048 | 0.87 | 0.72 | 1.05 | 1.57E-01 | 0.461 |
| OID31042 | BRSK2 | Neurology_II | BRSK2 | Q8IWQ3 | 11 | 1389899 | 1462689 | 0.88 | 0.74 | 1.05 | 1.58E-01 | 0.461 |
| OID21387 | CALCOCO1 | Oncology | CALCOCO1 | Q9P1Z2 | 12 | 53708517 | 53727745 | 0.89 | 0.76 | 1.05 | 1.59E-01 | 0.461 |
| OID21004 | CCL2 | Neurology | CCL2 | P13500 | 17 | 34255218 | 34257203 | 0.89 | 0.75 | 1.05 | 1.59E-01 | 0.461 |
| OID20722 | FIS1 | Inflammation | FIS1 | Q9Y3D6 | 7 | 101239458 | 101252316 | 0.89 | 0.75 | 1.05 | 1.58E-01 | 0.461 |
| OID31263 | HRAS | Oncology_II | HRAS | P01112 | 11 | 532242 | 537321 | 0.87 | 0.71 | 1.06 | 1.58E-01 | 0.461 |
| OID20585 | IL17RB | Inflammation | IL17RB | Q9NRM6 | 3 | 53846568 | 53865794 | 1.13 | 0.95 | 1.34 | 1.59E-01 | 0.461 |
| OID31172 | JMJD1C | Oncology_II | JMJD1C | Q15652 | 10 | 63167221 | 63521850 | 1.11 | 0.96 | 1.28 | 1.59E-01 | 0.461 |
| OID21475 | PPP1R12A | Oncology | PPP1R12A | O14974 | 12 | 79773563 | 79935460 | 0.89 | 0.75 | 1.05 | 1.59E-01 | 0.461 |
| OID30621 | PRKG1 | Inflammation_II | PRKG1 | Q13976 | 10 | 50990888 | 52298423 | 0.89 | 0.76 | 1.05 | 1.59E-01 | 0.461 |
| OID21163 | RABEPK | Oncology | RABEPK | Q7Z6M1 | 9 | 125200542 | 125234161 | 0.88 | 0.73 | 1.05 | 1.58E-01 | 0.461 |
| OID30204 | AIF1L | Cardiometabolic_II | AIF1L | Q9BQI0 | 9 | 131096476 | 131123152 | 0.86 | 0.70 | 1.06 | 1.60E-01 | 0.462 |
| OID21093 | INHBC | Neurology | INHBC | P55103 | 12 | 57434784 | 57452062 | 0.88 | 0.74 | 1.05 | 1.60E-01 | 0.462 |
| OID30151 | PDZD2 | Cardiometabolic_II | PDZD2 | O15018 | 5 | 31639131 | 32110932 | 0.88 | 0.74 | 1.05 | 1.60E-01 | 0.462 |
| OID21343 | RTBDN | Oncology | RTBDN | Q9BSG5 | 19 | 12825478 | 12835428 | 0.89 | 0.76 | 1.05 | 1.60E-01 | 0.462 |
| OID31513 | MTIF3 | Oncology_II | MTIF3 | Q9H2K0 | 13 | 27435643 | 27450591 | 0.89 | 0.75 | 1.05 | 1.60E-01 | 0.462 |
| OID30651 | PINLYP | Inflammation_II | PINLYP | A6NC86 | 19 | 43575801 | 43583964 | 0.89 | 0.76 | 1.05 | 1.61E-01 | 0.463 |
| OID20331 | PPIB | Cardiometabolic | PPIB | P23284 | 15 | 64155740 | 64163134 | 0.89 | 0.76 | 1.05 | 1.61E-01 | 0.463 |
| OID31297 | RGL2 | Oncology_II | RGL2 | O15211 | 6 | 33291654 | 33298942 | 0.86 | 0.70 | 1.06 | 1.61E-01 | 0.463 |
| OID30190 | LPP | Cardiometabolic_II | LPP | Q93052 | 3 | 188153284 | 188890671 | 0.89 | 0.75 | 1.05 | 1.61E-01 | 0.464 |
| OID31074 | CDH23 | Neurology_II | CDH23 | Q9H251 | 10 | 71396920 | 71815947 | 0.88 | 0.74 | 1.05 | 1.63E-01 | 0.464 |
| OID20665 | FASLG | Inflammation | FASLG | P48023 | 1 | 172659103 | 172666876 | 0.88 | 0.74 | 1.05 | 1.62E-01 | 0.464 |
| OID30714 | GSN | Inflammation_II | GSN | P06396 | 9 | 121207794 | 121332843 | 0.89 | 0.76 | 1.05 | 1.63E-01 | 0.464 |
| OID21320 | KIFBP | Oncology | KIFBP | Q96EK5 | 10 | 68988803 | 69043544 | 0.89 | 0.76 | 1.05 | 1.62E-01 | 0.464 |
| OID30227 | OXCT1 | Cardiometabolic_II | OXCT1 | P55809 | 5 | 41730065 | 41870425 | 0.88 | 0.74 | 1.05 | 1.63E-01 | 0.464 |
| OID30959 | RPGR | Neurology_II | RPGR | Q92834 | X | 38269163 | 38327544 | 1.10 | 0.96 | 1.26 | 1.62E-01 | 0.464 |
| OID30978 | SORBS1 | Neurology_II | SORBS1 | Q9BX66 | 10 | 95311771 | 95561414 | 0.86 | 0.70 | 1.06 | 1.62E-01 | 0.464 |
| OID21331 | ARSB | Oncology | ARSB | P15848 | 5 | 78777209 | 78986087 | 0.88 | 0.74 | 1.05 | 1.65E-01 | 0.471 |
| OID20828 | FGR | Neurology | FGR | P09769 | 1 | 27612064 | 27635185 | 0.87 | 0.71 | 1.06 | 1.65E-01 | 0.471 |
| OID21264 | LYAR | Oncology | LYAR | Q9NX58 | 4 | 4267701 | 4290154 | 0.86 | 0.69 | 1.07 | 1.66E-01 | 0.471 |
| OID31034 | RBM17 | Neurology_II | RBM17 | Q96I25 | 10 | 6089034 | 6117457 | 0.89 | 0.75 | 1.05 | 1.66E-01 | 0.471 |
| OID30311 | BPIFB2 | Cardiometabolic_II | BPIFB2 | Q8N4F0 | 20 | 33007704 | 33023703 | 0.88 | 0.74 | 1.05 | 1.66E-01 | 0.471 |

| | | | | | | | | | | | | |
|---|---|---|---|---|---|---|---|---|---|---|---|---|
| OID31430 | ITIH5 | Oncology_II | ITIH5 | Q86UX2 | 10 | 7559270 | 7666998 | 0.89 | 0.75 | 1.05 | 1.66E-01 | 0.471 |
| OID30219 | LONP1 | Cardiometabolic_II | LONP1 | P36776 | 19 | 5691834 | 5720572 | 0.89 | 0.75 | 1.05 | 1.67E-01 | 0.471 |
| OID30583 | MXRA8 | Inflammation_II | MXRA8 | Q9BRK3 | 1 | 1352689 | 1361777 | 0.88 | 0.74 | 1.05 | 1.66E-01 | 0.471 |
| OID31456 | PCDH7 | Oncology_II | PCDH7 | O60245 | 4 | 30720369 | 31146805 | 0.87 | 0.72 | 1.06 | 1.66E-01 | 0.471 |
| OID30888 | SOWAHA | Neurology_II | SOWAHA | Q2M3V2 | 5 | 132813302 | 132816786 | 0.88 | 0.72 | 1.06 | 1.66E-01 | 0.471 |
| OID21330 | GFRA2 | Oncology | GFRA2 | O00451 | 8 | 21690398 | 21812357 | 1.12 | 0.95 | 1.31 | 1.67E-01 | 0.472 |
| OID21450 | SPINK6 | Oncology | SPINK6 | Q6UWN8 | 5 | 148202794 | 148215137 | 1.14 | 0.95 | 1.37 | 1.67E-01 | 0.472 |
| OID21098 | SUSD2 | Neurology | SUSD2 | Q9UGT4 | 22 | 24181487 | 24189106 | 1.12 | 0.95 | 1.33 | 1.67E-01 | 0.472 |
| OID30848 | TFAP2A | Neurology_II | TFAP2A | P05549 | 6 | 10393186 | 10419659 | 1.11 | 0.96 | 1.28 | 1.67E-01 | 0.472 |
| OID20381 | GP1BA | Cardiometabolic | GP1BA | P07359 | 17 | 4932277 | 4935023 | 0.89 | 0.76 | 1.05 | 1.68E-01 | 0.472 |
| OID21451 | EDA2R | Oncology | EDA2R | Q9HAV5 | X | 66595637 | 66639298 | 1.14 | 0.95 | 1.38 | 1.68E-01 | 0.472 |
| OID31382 | RALY | Oncology_II | RALY | Q9UKM9 | 20 | 33993646 | 34108308 | 0.87 | 0.71 | 1.06 | 1.68E-01 | 0.472 |
| OID31047 | ALDH5A1 | Neurology_II | ALDH5A1 | P51649 | 6 | 24494867 | 24537207 | 0.88 | 0.74 | 1.05 | 1.68E-01 | 0.472 |
| OID20731 | TREM2 | Inflammation | TREM2 | Q9NZC2 | 6 | 41158506 | 41163186 | 1.13 | 0.95 | 1.34 | 1.69E-01 | 0.472 |
| OID21197 | AKT3 | Oncology | AKT3 | Q9Y243 | 1 | 243488233 | 243851079 | 0.89 | 0.75 | 1.05 | 1.69E-01 | 0.473 |
| OID30420 | DAND5 | Inflammation_II | DAND5 | Q8N907 | 19 | 12965159 | 12974760 | 1.11 | 0.96 | 1.30 | 1.69E-01 | 0.473 |
| OID30774 | SERPINA4 | Inflammation_II | SERPINA4 | P29622 | 14 | 94561442 | 94569913 | 0.89 | 0.75 | 1.05 | 1.69E-01 | 0.473 |
| OID20642 | TPSAB1 | Inflammation | TPSAB1 | Q15661 | 16 | 1240379 | 1242554 | 0.90 | 0.76 | 1.05 | 1.69E-01 | 0.473 |
| OID20459 | MVK | Inflammation | MVK | Q03426 | 12 | 109573255 | 109598125 | 0.88 | 0.74 | 1.05 | 1.70E-01 | 0.473 |
| OID20837 | STK24 | Neurology | STK24 | Q9Y6E0 | 13 | 98445185 | 98577940 | 0.89 | 0.76 | 1.05 | 1.70E-01 | 0.473 |
| OID30549 | DBN1 | Inflammation_II | DBN1 | Q16643 | 5 | 177456608 | 177474401 | 0.89 | 0.74 | 1.05 | 1.70E-01 | 0.473 |
| OID21502 | ADAMTS8 | Oncology | ADAMTS8 | Q9UP79 | 11 | 130404923 | 130428609 | 0.89 | 0.75 | 1.05 | 1.71E-01 | 0.476 |
| OID31020 | LRTM2 | Neurology_II | LRTM2 | Q8N967 | 12 | 1820267 | 1836753 | 0.89 | 0.74 | 1.05 | 1.72E-01 | 0.477 |
| OID20777 | PON3 | Inflammation | PON3 | Q15166 | 7 | 95359872 | 95396375 | 1.15 | 0.94 | 1.39 | 1.72E-01 | 0.477 |
| OID21411 | FGF23 | Oncology | FGF23 | Q9GZV9 | 12 | 4368227 | 4379712 | 1.12 | 0.95 | 1.30 | 1.73E-01 | 0.478 |
| OID30751 | LRG1 | Inflammation_II | LRG1 | P02750 | 19 | 4536402 | 4540036 | 1.13 | 0.95 | 1.34 | 1.73E-01 | 0.478 |
| OID30469 | PTP4A3 | Inflammation_II | PTP4A3 | O75365 | 8 | 141391995 | 141432454 | 0.88 | 0.74 | 1.06 | 1.73E-01 | 0.478 |
| OID30388 | TREH | Cardiometabolic_II | TREH | O43280 | 11 | 118657316 | 118679690 | 0.89 | 0.76 | 1.05 | 1.72E-01 | 0.478 |
| OID21087 | PEBP1 | Neurology | PEBP1 | P30086 | 12 | 118136124 | 118145584 | 0.89 | 0.75 | 1.05 | 1.73E-01 | 0.478 |
| OID30427 | UBE2Z | Inflammation_II | UBE2Z | Q9H832 | 17 | 48908407 | 48929056 | 0.88 | 0.72 | 1.06 | 1.73E-01 | 0.478 |
| OID30092 | KRT17 | Cardiometabolic_II | KRT17 | Q04695 | 17 | 41619442 | 41624842 | 1.11 | 0.95 | 1.29 | 1.74E-01 | 0.479 |
| OID30396 | COL15A1 | Cardiometabolic_II | COL15A1 | P39059 | 9 | 98943179 | 99070792 | 0.89 | 0.76 | 1.05 | 1.74E-01 | 0.479 |
| OID20564 | FCAR | Inflammation | FCAR | P24071 | 19 | 54874235 | 54891420 | 1.11 | 0.95 | 1.30 | 1.75E-01 | 0.480 |
| OID30455 | MKI67 | Inflammation_II | MKI67 | P46013 | 10 | 128096659 | 128126423 | 0.86 | 0.69 | 1.07 | 1.75E-01 | 0.480 |
| OID30086 | ACY3 | Cardiometabolic_II | ACY3 | Q96HD9 | 11 | 67642555 | 67650730 | 0.89 | 0.75 | 1.05 | 1.75E-01 | 0.482 |
| OID30872 | TXNL1 | Neurology_II | TXNL1 | O43396 | 18 | 56597209 | 56651600 | 0.88 | 0.73 | 1.06 | 1.76E-01 | 0.482 |
| OID30496 | REPS1 | Inflammation_II | REPS1 | Q96D71 | 6 | 138903493 | 138988261 | 0.89 | 0.75 | 1.06 | 1.77E-01 | 0.484 |
| OID31065 | FKBP14 | Neurology_II | FKBP14 | Q9NWM8 | 7 | 30010587 | 30026702 | 0.89 | 0.76 | 1.05 | 1.78E-01 | 0.486 |
| OID20937 | FKBP5 | Neurology | FKBP5 | Q13451 | 6 | 35573585 | 35728583 | 0.89 | 0.76 | 1.05 | 1.78E-01 | 0.486 |
| OID21279 | SEPTIN9 | Oncology | SEPTIN9 | Q9UHD8 | 17 | 77280569 | 77500596 | 0.88 | 0.73 | 1.06 | 1.78E-01 | 0.486 |
| OID30320 | LECT2 | Cardiometabolic_II | LECT2 | O14960 | 5 | 135922279 | 135954983 | 0.88 | 0.73 | 1.06 | 1.78E-01 | 0.486 |
| OID20726 | ANGPTL2 | Inflammation | ANGPTL2 | Q9UKU9 | 9 | 127087348 | 127122635 | 1.12 | 0.95 | 1.33 | 1.78E-01 | 0.487 |
| OID20581 | ITGA11 | Inflammation | ITGA11 | Q9UKX5 | 15 | 68296532 | 68432163 | 0.89 | 0.75 | 1.05 | 1.79E-01 | 0.487 |
| OID21281 | VPS53 | Oncology | VPS53 | Q5VIR6 | 17 | 508503 | 721717 | 0.89 | 0.75 | 1.05 | 1.79E-01 | 0.487 |
| OID21426 | CNTN2 | Oncology | CNTN2 | Q02246 | 1 | 205042937 | 205078289 | 0.89 | 0.74 | 1.06 | 1.79E-01 | 0.487 |
| OID30740 | IGLC2 | Inflammation_II | IGLC2 | P0DOY2 | 22 | 22900976 | 22901437 | 0.86 | 0.69 | 1.07 | 1.79E-01 | 0.487 |
| OID20941 | IFNGR2 | Neurology | IFNGR2 | P38484 | 21 | 33403413 | 33479348 | 0.89 | 0.74 | 1.06 | 1.80E-01 | 0.488 |
| OID31305 | MTHFSD | Oncology_II | MTHFSD | Q2M296 | 16 | 86530178 | 86555235 | 0.90 | 0.76 | 1.05 | 1.80E-01 | 0.489 |
| OID31508 | CDC26 | Oncology_II | CDC26 | Q8NHZ8 | 9 | 113255835 | 113275572 | 0.89 | 0.74 | 1.06 | 1.80E-01 | 0.489 |
| OID21187 | RRM2 | Oncology | RRM2 | P31350 | 2 | 10120698 | 10211725 | 0.88 | 0.73 | 1.06 | 1.80E-01 | 0.489 |
| OID31323 | SAP18 | Oncology_II | SAP18 | O00422 | 13 | 21140119 | 21149097 | 1.10 | 0.95 | 1.28 | 1.81E-01 | 0.489 |
| OID21080 | CD164 | Neurology | CD164 | Q04900 | 6 | 109366514 | 109382467 | 0.88 | 0.73 | 1.06 | 1.81E-01 | 0.489 |
| OID20877 | MAX | Neurology | MAX | P61244 | 14 | 65006174 | 65102695 | 0.89 | 0.76 | 1.05 | 1.82E-01 | 0.491 |
| OID20436 | LAP3 | Inflammation | LAP3 | P28838 | 4 | 17577198 | 17607972 | 0.89 | 0.75 | 1.06 | 1.82E-01 | 0.491 |
| OID30306 | MAN2B2 | Cardiometabolic_II | MAN2B2 | Q9Y2E5 | 4 | 6575189 | 6623362 | 0.89 | 0.76 | 1.05 | 1.82E-01 | 0.491 |
| OID30908 | MN1 | Neurology_II | MN1 | Q10571 | 22 | 27748277 | 27801756 | 0.89 | 0.74 | 1.06 | 1.82E-01 | 0.491 |
| OID20969 | PSME1 | Neurology | PSME1 | Q06323 | 14 | 24136163 | 24138967 | 0.89 | 0.75 | 1.06 | 1.83E-01 | 0.492 |
| OID21174 | RRM2B | Oncology | RRM2B | Q7LG56 | 8 | 102204502 | 102238961 | 0.89 | 0.76 | 1.05 | 1.83E-01 | 0.492 |
| OID31121 | TXNDC9 | Neurology_II | TXNDC9 | O14530 | 2 | 99318982 | 99340702 | 0.89 | 0.75 | 1.06 | 1.83E-01 | 0.492 |
| OID21026 | VCAN | Neurology | VCAN | P13611 | 5 | 83471618 | 83582303 | 1.12 | 0.95 | 1.31 | 1.83E-01 | 0.492 |
| OID20559 | CLIP2 | Inflammation | CLIP2 | Q9UDT6 | 7 | 74289407 | 74405935 | 0.90 | 0.76 | 1.05 | 1.84E-01 | 0.493 |
| OID30831 | ECSCR | Neurology_II | ECSCR | Q19T08 | 5 | 139448560 | 139462743 | 0.87 | 0.72 | 1.07 | 1.84E-01 | 0.493 |
| OID21268 | ITGB1BP1 | Oncology | ITGB1BP1 | O14713 | 2 | 9403475 | 9423528 | 1.11 | 0.95 | 1.31 | 1.84E-01 | 0.493 |
| OID30228 | PECR | Cardiometabolic_II | PECR | Q9BY49 | 2 | 215996329 | 216082955 | 0.89 | 0.74 | 1.06 | 1.84E-01 | 0.493 |
| OID30212 | LMOD1 | Cardiometabolic_II/Infla | LMOD1 | P29536 | 1 | 201896456 | 201946588 | 1.11 | 0.95 | 1.28 | 1.85E-01 | 0.493 |
| OID30684 | F10 | Inflammation_II | F10 | P00742 | 13 | 113122799 | 113149529 | 0.90 | 0.76 | 1.05 | 1.86E-01 | 0.493 |
| OID20604 | GZMB | Inflammation | GZMB | P10144 | 14 | 24630954 | 24634267 | 0.88 | 0.73 | 1.06 | 1.86E-01 | 0.493 |
| OID30314 | L3HYPDH | Cardiometabolic_II | L3HYPDH | Q96EM0 | 14 | 59460363 | 59484408 | 0.89 | 0.76 | 1.06 | 1.86E-01 | 0.493 |
| OID30450 | LRIG3 | Inflammation_II | LRIG3 | Q6UXM1 | 12 | 58872149 | 58920504 | 1.11 | 0.95 | 1.29 | 1.86E-01 | 0.493 |
| OID21099 | MESD | Neurology | MESD | Q14696 | 15 | 80946289 | 80989828 | 0.90 | 0.76 | 1.05 | 1.86E-01 | 0.493 |
| OID31420 | MPRIP | Oncology_II | MPRIP | Q6WCQ1 | 17 | 17042457 | 17217679 | 0.89 | 0.75 | 1.06 | 1.86E-01 | 0.493 |
| OID30251 | MYL3 | Cardiometabolic_II | MYL3 | P08590 | 3 | 46835111 | 46882172 | 0.89 | 0.74 | 1.06 | 1.86E-01 | 0.493 |
| OID20354 | NCAM1 | Cardiometabolic | NCAM1 | P13591 | 11 | 112961247 | 113278436 | 1.12 | 0.95 | 1.32 | 1.85E-01 | 0.493 |
| OID31369 | NDST1 | Oncology_II | NDST1 | P52848 | 5 | 150485818 | 150558211 | 0.89 | 0.74 | 1.06 | 1.86E-01 | 0.493 |
| OID21509 | P4HB | Oncology | P4HB | P07237 | 17 | 81843159 | 81860856 | 0.89 | 0.75 | 1.06 | 1.85E-01 | 0.493 |
| OID21254 | SLAMF6 | Oncology | SLAMF6 | Q96DU3 | 1 | 160485030 | 160523262 | 0.89 | 0.75 | 1.06 | 1.86E-01 | 0.493 |
| OID21008 | CDH3 | Neurology | CDH3 | P22223 | 16 | 68636189 | 68727468 | 1.11 | 0.95 | 1.28 | 1.87E-01 | 0.494 |
| OID21432 | ACP6 | Oncology | ACP6 | Q9NPH0 | 1 | 147629652 | 147670524 | 0.91 | 0.78 | 1.05 | 1.88E-01 | 0.494 |
| OID30267 | BOLA2_BOLA2B | Cardiometabolic_II | 3OLA2B;BOLA2 | Q9H3K6;Q9H3K6 | 16;16 | 30192932;29453590 | 30194306;29454351 | 0.89 | 0.76 | 1.06 | 1.89E-01 | 0.494 |
| OID20223 | CHRDL2 | Cardiometabolic | CHRDL2 | Q6WN34 | 11 | 74696429 | 74731426 | 1.13 | 0.94 | 1.35 | 1.89E-01 | 0.494 |
| OID20874 | CLSTN1 | Neurology | CLSTN1 | O94985 | 1 | 9728926 | 9823984 | 0.88 | 0.73 | 1.06 | 1.88E-01 | 0.494 |
| OID30723 | F2 | Inflammation_II | F2 | P00734 | 11 | 46719196 | 46739506 | 0.89 | 0.75 | 1.06 | 1.88E-01 | 0.494 |
| OID30950 | LYSMD3 | Neurology_II | LYSMD3 | Q7Z3D4 | 5 | 90515611 | 90529584 | 0.88 | 0.74 | 1.06 | 1.89E-01 | 0.494 |
| OID30633 | MYOM3 | Inflammation_II | MYOM3 | Q5VTT5 | 1 | 24056035 | 24112135 | 0.89 | 0.75 | 1.06 | 1.88E-01 | 0.494 |
| OID20729 | PDLIM7 | Inflammation | PDLIM7 | Q9NR12 | 5 | 177483394 | 177497606 | 0.89 | 0.76 | 1.06 | 1.89E-01 | 0.494 |
| OID30480 | TERF1 | Inflammation_II | TERF1 | P54274 | 8 | 73008856 | 73048123 | 0.89 | 0.75 | 1.06 | 1.89E-01 | 0.494 |
| OID20621 | TGFB1 | Inflammation | TGFB1 | P01137 | 19 | 41301587 | 41353922 | 0.89 | 0.75 | 1.06 | 1.89E-01 | 0.494 |
| OID30964 | UROS | Neurology_II | UROS | P10746 | 10 | 125784980 | 125823258 | 0.88 | 0.73 | 1.06 | 1.88E-01 | 0.494 |
| OID20936 | VWC2 | Neurology | VWC2 | Q2TAL6 | 7 | 49773638 | 49921950 | 1.12 | 0.95 | 1.32 | 1.89E-01 | 0.494 |
| OID30181 | YOD1 | Cardiometabolic_II | YOD1 | Q5VVQ6 | 1 | 207043849 | 207052980 | 0.89 | 0.75 | 1.06 | 1.89E-01 | 0.494 |
| OID30503 | NEDD9 | Inflammation_II | NEDD9 | Q14511 | 6 | 11183298 | 11382348 | 0.88 | 0.73 | 1.07 | 1.90E-01 | 0.495 |
| OID30942 | TTF2 | Neurology_II | TTF2 | Q9UNY4 | 1 | 117060326 | 117107453 | 0.90 | 0.76 | 1.06 | 1.90E-01 | 0.495 |
| OID30869 | CHP1 | Neurology_II | CHP1 | Q99653 | 15 | 41230839 | 41281887 | 0.89 | 0.75 | 1.06 | 1.90E-01 | 0.495 |
| OID31115 | NGRN | Neurology_II | NGRN | Q9NPE2 | 15 | 90265659 | 90275778 | 0.88 | 0.74 | 1.06 | 1.90E-01 | 0.495 |
| OID21244 | AKR1B1 | Oncology | AKR1B1 | P15121 | 7 | 134442356 | 134459284 | 0.90 | 0.76 | 1.06 | 1.91E-01 | 0.496 |
| OID30678 | NEXN | Inflammation_II | NEXN | Q0ZGT2 | 1 | 77888513 | 77943895 | 0.89 | 0.75 | 1.06 | 1.91E-01 | 0.496 |
| OID30711 | VNN1 | Inflammation_II | VNN1 | O95497 | 6 | 132680849 | 132714055 | 0.90 | 0.77 | 1.05 | 1.91E-01 | 0.496 |
| OID20402 | PROC | Cardiometabolic | PROC | P04070 | 2 | 127418427 | 127429242 | 0.90 | 0.77 | 1.05 | 1.91E-01 | 0.497 |
| OID30247 | DNAJC6 | Cardiometabolic_II | DNAJC6 | O75061 | 1 | 65248219 | 65415871 | 0.90 | 0.76 | 1.06 | 1.92E-01 | 0.497 |
| OID30889 | MICALL2 | Neurology_II | MICALL2 | Q8IY33 | 7 | 1428465 | 1459470 | 0.89 | 0.75 | 1.06 | 1.92E-01 | 0.497 |
| OID20688 | SCGB1A1 | Inflammation | SCGB1A1 | P11684 | 11 | 62405103 | 62423195 | 0.90 | 0.77 | 1.05 | 1.92E-01 | 0.498 |
| OID20241 | CA4 | Cardiometabolic | CA4 | P22748 | 17 | 60149942 | 60170899 | 1.11 | 0.95 | 1.31 | 1.95E-01 | 0.499 |
| OID20522 | EPO | Inflammation | EPO | P01588 | 7 | 100720468 | 100723700 | 1.11 | 0.95 | 1.31 | 1.95E-01 | 0.499 |

| | | | | | | | | | | | | |
|---|---|---|---|---|---|---|---|---|---|---|---|---|
| OID20641 | GMPR | Inflammation | GMPR | P36959 | 6 | 16238587 | 16295549 | 0.90 | 0.77 | 1.06 | 1.93E-01 | 0.499 |
| OID20890 | GPKOW | Neurology | GPKOW | Q92917 | X | 49113407 | 49123735 | 0.87 | 0.70 | 1.07 | 1.95E-01 | 0.499 |
| OID20209 | LILRA5 | Cardiometabolic | LILRA5 | A6NI73 | 19 | 54307070 | 54313166 | 1.12 | 0.94 | 1.32 | 1.95E-01 | 0.499 |
| OID31459 | MEP1A | Oncology_II | MEP1A | Q16819 | 6 | 46793389 | 46839778 | 1.12 | 0.95 | 1.31 | 1.94E-01 | 0.499 |
| OID21388 | NPY | Oncology | NPY | P01303 | 7 | 24284188 | 24291862 | 0.90 | 0.76 | 1.06 | 1.95E-01 | 0.499 |
| OID21360 | OGFR | Oncology | OGFR | Q9NZT2 | 20 | 62804835 | 62814000 | 0.89 | 0.75 | 1.06 | 1.95E-01 | 0.499 |
| OID30440 | POLR2A | Inflammation_II | POLR2A | P24928 | 17 | 7484366 | 7514616 | 0.89 | 0.75 | 1.06 | 1.95E-01 | 0.499 |
| OID30258 | RANBP1 | Cardiometabolic_II | RANBP1 | P43487 | 22 | 20115938 | 20127355 | 0.90 | 0.76 | 1.06 | 1.95E-01 | 0.499 |
| OID31210 | STAU1 | Oncology_II | STAU1 | O95793 | 20 | 49113339 | 49188367 | 0.88 | 0.73 | 1.06 | 1.94E-01 | 0.499 |
| OID21286 | TBC1D23 | Oncology | TBC1D23 | Q9NUY8 | 3 | 100260992 | 100325251 | 0.90 | 0.76 | 1.06 | 1.94E-01 | 0.499 |
| OID21482 | TNFRSF12A | Oncology | TNFRSF12A | Q9NP84 | 16 | 3018445 | 3022383 | 1.11 | 0.95 | 1.31 | 1.95E-01 | 0.499 |
| OID20139 | AGXT | Cardiometabolic | AGXT | P21549 | 2 | 240868824 | 240880502 | 0.89 | 0.75 | 1.06 | 1.95E-01 | 0.499 |
| OID31150 | BAP18 | Neurology_II | C17orf49 | Q8IXM2 | 17 | 7014495 | 7017525 | 0.88 | 0.72 | 1.07 | 1.96E-01 | 0.499 |
| OID30781 | F13B | Inflammation_II | F13B | P05160 | 1 | 197038741 | 197067260 | 0.89 | 0.75 | 1.06 | 1.96E-01 | 0.499 |
| OID31043 | DDHD2 | Neurology_II | DDHD2 | O94830 | 8 | 38225218 | 38275558 | 0.90 | 0.76 | 1.06 | 1.97E-01 | 0.501 |
| OID31399 | MINDY1 | Oncology_II | MINDY1 | Q8N5J2 | 1 | 150996549 | 151008376 | 0.90 | 0.77 | 1.06 | 1.97E-01 | 0.501 |
| OID21002 | PLA2G10 | Neurology | PLA2G10 | O15496 | 16 | 14672548 | 14694308 | 1.12 | 0.94 | 1.33 | 1.97E-01 | 0.501 |
| OID20386 | SOD1 | Cardiometabolic | SOD1 | P00441 | 21 | 31659666 | 31668931 | 0.89 | 0.74 | 1.06 | 1.97E-01 | 0.501 |
| OID20548 | CEACAM21 | Inflammation | CEACAM21 | Q3KPI0 | 19 | 41549518 | 41586844 | 1.11 | 0.95 | 1.29 | 1.98E-01 | 0.503 |
| OID21315 | LAG3 | Oncology | LAG3 | P18627 | 12 | 6772512 | 6778455 | 0.89 | 0.75 | 1.06 | 1.98E-01 | 0.503 |
| OID30979 | SPAG1 | Neurology_II | SPAG1 | Q07617 | 8 | 100157906 | 100259278 | 0.88 | 0.73 | 1.07 | 1.98E-01 | 0.503 |
| OID20572 | EGLN1 | Inflammation | EGLN1 | Q9GZT9 | 1 | 231363751 | 231422287 | 0.88 | 0.72 | 1.07 | 1.99E-01 | 0.503 |
| OID20242 | EPHB4 | Cardiometabolic | EPHB4 | P54760 | 7 | 100802565 | 100827523 | 1.10 | 0.95 | 1.28 | 1.99E-01 | 0.504 |
| OID30809 | GLP1R | Neurology_II | GLP1R | P43220 | 6 | 39048781 | 39091303 | 0.90 | 0.76 | 1.06 | 1.99E-01 | 0.504 |
| OID30302 | EHBP1 | Cardiometabolic_II | EHBP1 | Q8NDI1 | 2 | 62673851 | 63046487 | 0.90 | 0.76 | 1.06 | 2.00E-01 | 0.505 |
| OID30376 | LILRA3 | Cardiometabolic_II | LILRA3 | Q8N6C8 | 19 | 54294163 | 54306461 | 1.16 | 0.93 | 1.44 | 2.00E-01 | 0.506 |
| OID30741 | AFM | Inflammation_II | AFM | P43652 | 4 | 73481745 | 73504001 | 0.90 | 0.76 | 1.06 | 2.01E-01 | 0.506 |
| OID31523 | ARF6 | Oncology_II | ARF6 | P62330 | 14 | 49893079 | 49897054 | 0.90 | 0.76 | 1.06 | 2.01E-01 | 0.506 |
| OID30966 | GBA | Neurology_II | GBA | P04062 | 1 | 155234452 | 155244699 | 0.90 | 0.77 | 1.06 | 2.01E-01 | 0.506 |
| OID20826 | RASA1 | Neurology | RASA1 | P20936 | 5 | 87267883 | 87391931 | 0.89 | 0.75 | 1.06 | 2.01E-01 | 0.506 |
| OID20131 | VSIR | Cardiometabolic | VSIR | Q9H7M9 | 10 | 71747556 | 71773520 | 0.90 | 0.76 | 1.06 | 2.01E-01 | 0.506 |
| OID20160 | PLXNB3 | Cardiometabolic | PLXNB3 | Q9ULL4 | X | 153764196 | 153779346 | 0.90 | 0.76 | 1.06 | 2.02E-01 | 0.506 |
| OID20846 | FKBP7 | Neurology | FKBP7 | Q9Y680 | 2 | 178463664 | 178478600 | 0.88 | 0.73 | 1.07 | 2.02E-01 | 0.507 |
| OID31340 | GPRC5C | Oncology_II | GPRC5C | Q9NQ84 | 17 | 74424851 | 74451653 | 0.89 | 0.75 | 1.06 | 2.02E-01 | 0.507 |
| OID30965 | SV2A | Neurology_II | SV2A | Q7L0J3 | 1 | 149903318 | 149917844 | 0.89 | 0.75 | 1.06 | 2.02E-01 | 0.507 |
| OID20255 | TGFBR3 | Cardiometabolic | TGFBR3 | Q03167 | 1 | 91680343 | 91906335 | 1.11 | 0.95 | 1.29 | 2.02E-01 | 0.507 |
| OID20373 | RARRES2 | Cardiometabolic | RARRES2 | Q99969 | 7 | 150338317 | 150341662 | 0.89 | 0.74 | 1.07 | 2.03E-01 | 0.507 |
| OID30526 | ZBP1 | Inflammation_II | ZBP1 | Q9H171 | 20 | 57603846 | 57620576 | 0.87 | 0.71 | 1.08 | 2.03E-01 | 0.507 |
| OID30518 | ANKMY2 | Inflammation_II | ANKMY2 | Q8IV38 | 7 | 16599779 | 16645817 | 0.90 | 0.76 | 1.06 | 2.03E-01 | 0.507 |
| OID30709 | AGT | Inflammation_II | AGT | P01019 | 1 | 230690776 | 230745576 | 1.10 | 0.95 | 1.29 | 2.06E-01 | 0.509 |
| OID30749 | APOC1 | Inflammation_II | APOC1 | P02654 | 19 | 44914247 | 44919349 | 0.89 | 0.74 | 1.07 | 2.06E-01 | 0.509 |
| OID20377 | CD55 | Cardiometabolic | CD55 | P08174 | 1 | 207321532 | 207386804 | 0.90 | 0.77 | 1.06 | 2.07E-01 | 0.509 |
| OID31426 | CHCHD6 | Oncology_II | CHCHD6 | Q9BRQ6 | 3 | 126704240 | 126960420 | 0.88 | 0.73 | 1.07 | 2.07E-01 | 0.509 |
| OID30911 | CNTNAP4 | Neurology_II | CNTNAP4 | Q9C0A0 | 16 | 76277278 | 76560757 | 0.89 | 0.73 | 1.07 | 2.05E-01 | 0.509 |
| OID21349 | DTX3 | Oncology | DTX3 | Q8N9I9 | 12 | 57604622 | 57609804 | 0.89 | 0.74 | 1.07 | 2.05E-01 | 0.509 |
| OID31460 | EFCAB14 | Oncology_II | EFCAB14 | O75071 | 1 | 46674659 | 46719146 | 0.90 | 0.75 | 1.06 | 2.05E-01 | 0.509 |
| OID21043 | FABP5 | Neurology | FABP5 | Q01469 | 8 | 81280536 | 81284777 | 0.89 | 0.75 | 1.06 | 2.05E-01 | 0.509 |
| OID20443 | FCRL3 | Inflammation | FCRL3 | Q96P31 | 1 | 157674321 | 157700769 | 0.91 | 0.78 | 1.06 | 2.06E-01 | 0.509 |
| OID31059 | IGBP1 | Neurology_II | IGBP1 | P78318 | X | 70133447 | 70166324 | 0.90 | 0.76 | 1.06 | 2.06E-01 | 0.509 |
| OID21518 | KLK11 | Oncology | KLK11 | Q9UBX7 | 19 | 51022216 | 51028039 | 0.90 | 0.77 | 1.06 | 2.05E-01 | 0.509 |
| OID20819 | PAK4 | Neurology | PAK4 | O96013 | 19 | 39125770 | 39182816 | 0.90 | 0.76 | 1.06 | 2.06E-01 | 0.509 |
| OID30344 | PGD | Cardiometabolic_II | PGD | P52209 | 1 | 10398592 | 10420511 | 0.89 | 0.75 | 1.06 | 2.04E-01 | 0.509 |
| OID30672 | RNASE1 | Inflammation_II | RNASE1 | P07998 | 14 | 20801228 | 20802855 | 0.89 | 0.75 | 1.06 | 2.07E-01 | 0.509 |
| OID20761 | SKAP2 | Inflammation | SKAP2 | O75563 | 7 | 26667068 | 26995239 | 0.90 | 0.77 | 1.06 | 2.07E-01 | 0.509 |
| OID20213 | SORT1 | Cardiometabolic | SORT1 | Q99523 | 1 | 109309568 | 109397918 | 0.90 | 0.76 | 1.06 | 2.07E-01 | 0.509 |
| OID20735 | TNFRSF11B | Inflammation | TNFRSF11B | O00300 | 8 | 118923557 | 118951885 | 1.12 | 0.94 | 1.33 | 2.07E-01 | 0.509 |
| OID20053 | VIM | Cardiometabolic | VIM | P08670 | 10 | 17228241 | 17237593 | 0.91 | 0.79 | 1.05 | 2.05E-01 | 0.509 |
| OID30339 | SYTL4 | Cardiometabolic_II | SYTL4 | Q96C24 | X | 100671783 | 100732123 | 0.90 | 0.76 | 1.06 | 2.08E-01 | 0.510 |
| OID30271 | NAGK | Cardiometabolic_II | NAGK | Q9UJ70 | 2 | 71064344 | 71079808 | 0.89 | 0.75 | 1.06 | 2.08E-01 | 0.511 |
| OID20657 | MERTK | Inflammation | MERTK | Q12866 | 2 | 111898607 | 112029561 | 1.11 | 0.94 | 1.31 | 2.08E-01 | 0.511 |
| OID20718 | HSPA1A | Inflammation | HSPA1A | P0DMV8 | 6 | 31815543 | 31817946 | 0.90 | 0.75 | 1.06 | 2.09E-01 | 0.512 |
| OID30487 | IL31 | Inflammation_II | IL31 | Q6EBC2 | 12 | 122172029 | 122174221 | 0.89 | 0.73 | 1.07 | 2.09E-01 | 0.513 |
| OID20244 | CES1 | Cardiometabolic | CES1 | P23141 | 16 | 55802851 | 55833337 | 0.90 | 0.76 | 1.06 | 2.10E-01 | 0.513 |
| OID30136 | EDEM2 | Cardiometabolic_II | EDEM2 | Q9BV94 | 20 | 35115364 | 35147336 | 0.88 | 0.73 | 1.07 | 2.10E-01 | 0.513 |
| OID20973 | CCS | Neurology | CCS | O14618 | 11 | 66593153 | 66606019 | 0.90 | 0.76 | 1.06 | 2.10E-01 | 0.513 |
| OID20457 | SCGN | Inflammation | SCGN | O76038 | 6 | 25652201 | 25701783 | 1.11 | 0.94 | 1.30 | 2.10E-01 | 0.513 |
| OID30316 | TCOF1 | Cardiometabolic_II | TCOF1 | Q13428 | 5 | 150357629 | 150400308 | 0.88 | 0.71 | 1.08 | 2.10E-01 | 0.513 |
| OID20147 | IL6ST | Cardiometabolic | IL6ST | P40189 | 5 | 55935095 | 55995022 | 1.11 | 0.94 | 1.30 | 2.12E-01 | 0.515 |
| OID31345 | PRC1 | Oncology_II | PRC1 | O43663 | 15 | 90966040 | 90995629 | 0.86 | 0.69 | 1.09 | 2.12E-01 | 0.515 |
| OID30737 | PROS1 | Inflammation_II | PROS1 | P07225 | 3 | 93873051 | 93980003 | 0.90 | 0.76 | 1.06 | 2.12E-01 | 0.515 |
| OID20516 | VASH1 | Inflammation | VASH1 | Q7L8A9 | 14 | 76761468 | 76783015 | 0.90 | 0.76 | 1.06 | 2.12E-01 | 0.515 |
| OID20256 | VWF | Cardiometabolic | VWF | P04275 | 12 | 5948877 | 6124770 | 1.11 | 0.94 | 1.31 | 2.12E-01 | 0.515 |
| OID21014 | STC1 | Neurology | STC1 | P52823 | 8 | 23841929 | 23854806 | 1.11 | 0.94 | 1.31 | 2.13E-01 | 0.516 |
| OID21234 | AIF1 | Oncology | AIF1 | P55008 | 6 | 31615217 | 31617021 | 0.90 | 0.76 | 1.06 | 2.13E-01 | 0.517 |
| OID30707 | AMY1A_AMY1B | Inflammation_II | ;AMY1B;AMY1C | B6;P0DTE7;P0DTE8 | 1;1;1 | )3687415;103745323 | )3696454;103758726 | 0.90 | 0.76 | 1.06 | 2.14E-01 | 0.517 |
| OID30974 | FBN2 | Neurology_II | FBN2 | P35556 | 5 | 128257909 | 128659185 | 0.90 | 0.77 | 1.06 | 2.14E-01 | 0.517 |
| OID30354 | RAB27B | Cardiometabolic_II | RAB27B | O00194 | 18 | 54717860 | 54895516 | 0.91 | 0.78 | 1.06 | 2.14E-01 | 0.517 |
| OID20691 | F2R | Inflammation | F2R | P25116 | 5 | 76716126 | 76735770 | 0.90 | 0.76 | 1.06 | 2.14E-01 | 0.517 |
| OID20106 | DCN | Cardiometabolic | DCN | P07585 | 12 | 91140484 | 91183217 | 1.11 | 0.94 | 1.31 | 2.14E-01 | 0.518 |
| OID31031 | SUOX | Neurology_II | SUOX | P51687 | 12 | 55997180 | 56006641 | 0.89 | 0.75 | 1.07 | 2.15E-01 | 0.518 |
| OID31165 | PRAME | Oncology_II | PRAME | P78395 | 22 | 22547701 | 22559361 | 0.90 | 0.76 | 1.06 | 2.15E-01 | 0.518 |
| OID31218 | BCL7A | Oncology_II | BCL7A | Q4VC05 | 12 | 122019422 | 122062044 | 0.89 | 0.75 | 1.07 | 2.16E-01 | 0.519 |
| OID31005 | FAM171A2 | Neurology_II | FAM171A2 | A8MVW0 | 17 | 44353215 | 44363853 | 0.89 | 0.74 | 1.07 | 2.16E-01 | 0.519 |
| OID20787 | TFF2 | Inflammation | TFF2 | Q03403 | 21 | 42346357 | 42350997 | 1.10 | 0.94 | 1.29 | 2.16E-01 | 0.519 |
| OID31278 | TMED1 | Oncology_II | TMED1 | Q13445 | 19 | 10832067 | 10836318 | 0.89 | 0.74 | 1.07 | 2.16E-01 | 0.519 |
| OID30881 | AMIGO1 | Neurology_II | AMIGO1 | Q86WK6 | 1 | 109504178 | 109509738 | 0.89 | 0.74 | 1.07 | 2.16E-01 | 0.519 |
| OID20477 | IL17C | Inflammation | IL17C | Q9P0M4 | 16 | 88638572 | 88640468 | 1.10 | 0.94 | 1.29 | 2.17E-01 | 0.519 |
| OID21010 | CD300C | Neurology | CD300C | Q08708 | 17 | 74541073 | 74546115 | 1.11 | 0.94 | 1.30 | 2.18E-01 | 0.519 |
| OID20771 | CHRDL1 | Inflammation | CHRDL1 | Q9BU40 | X | 110673856 | 110795819 | 1.12 | 0.94 | 1.33 | 2.18E-01 | 0.519 |
| OID21150 | CTRB1 | Neurology | CTRB1 | P17538 | 16 | 75218988 | 75226338 | 0.91 | 0.78 | 1.06 | 2.18E-01 | 0.519 |
| OID30332 | FCAMR | Cardiometabolic_II | FCAMR | Q8WWV6 | 1 | 206957965 | 206970625 | 0.89 | 0.75 | 1.07 | 2.17E-01 | 0.519 |
| OID30147 | FDX2 | Cardiometabolic_II | FDX2 | Q6P4F2 | 19 | 10310045 | 10316015 | 0.89 | 0.75 | 1.07 | 2.18E-01 | 0.519 |
| OID31437 | GPHA2 | Oncology_II | GPHA2 | Q96T91 | 11 | 64934471 | 64935893 | 0.88 | 0.72 | 1.08 | 2.19E-01 | 0.519 |
| OID20398 | IGFBP3 | Cardiometabolic | IGFBP3 | P17936 | 7 | 45912245 | 45921874 | 0.92 | 0.80 | 1.05 | 2.17E-01 | 0.519 |
| OID21153 | PAMR1 | Neurology | PAMR1 | Q6UXH9 | 11 | 35431823 | 35530300 | 0.90 | 0.75 | 1.07 | 2.18E-01 | 0.519 |
| OID21229 | SUGT1 | Oncology | SUGT1 | Q9Y2Z0 | 13 | 52652709 | 52700909 | 0.90 | 0.77 | 1.06 | 2.17E-01 | 0.519 |
| OID20341 | TCN2 | Cardiometabolic | TCN2 | P20062 | 22 | 30607003 | 30627271 | 1.10 | 0.95 | 1.27 | 2.18E-01 | 0.519 |
| OID30476 | THSD1 | Inflammation_II | THSD1 | Q9NS62 | 13 | 52377167 | 52416373 | 1.11 | 0.94 | 1.30 | 2.18E-01 | 0.519 |
| OID21428 | DAB2 | Oncology | DAB2 | P98082 | 5 | 39371675 | 39462300 | 0.90 | 0.77 | 1.06 | 2.20E-01 | 0.523 |
| OID30534 | DEFB103A_DEF | Inflammation_II | 03B;DEFB103A | P81534;P81534 | 8;8 | 7428888;7881392 | 7430348;7882663 | 0.90 | 0.75 | 1.07 | 2.21E-01 | 0.523 |
| OID31344 | LRRFIP1 | Oncology_II | LRRFIP1 | Q32MZ4 | 2 | 237627587 | 237813682 | 0.90 | 0.76 | 1.07 | 2.21E-01 | 0.523 |
| OID21365 | RBP2 | Oncology | RBP2 | P50120 | 3 | 139452884 | 139480747 | 1.10 | 0.94 | 1.29 | 2.21E-01 | 0.523 |
| OID31030 | RTN4IP1 | Neurology_II | RTN4IP1 | Q8WWV3 | 6 | 106570771 | 106629498 | 0.90 | 0.76 | 1.06 | 2.20E-01 | 0.523 |

| | | | | | | | | | | | | |
|---|---|---|---|---|---|---|---|---|---|---|---|---|
| OID30135 | ENPEP | Cardiometabolic_II | ENPEP | Q07075 | 4 | 110365733 | 110565285 | 1.10 | 0.94 | 1.30 | 2.21E-01 | 0.523 |
| OID20835 | PLIN1 | Neurology | PLIN1 | O60240 | 15 | 89664367 | 89679427 | 0.89 | 0.74 | 1.07 | 2.21E-01 | 0.523 |
| OID31178 | MYH7B | Oncology_II | MYH7B | A7E2Y1 | 20 | 34955810 | 35002437 | 0.90 | 0.75 | 1.07 | 2.22E-01 | 0.523 |
| OID20927 | APRT | Neurology | APRT | P07741 | 16 | 88809339 | 88811937 | 0.90 | 0.76 | 1.06 | 2.23E-01 | 0.524 |
| OID31413 | CCDC134 | Oncology_II | CCDC134 | Q9H6E4 | 22 | 41800679 | 41832164 | 0.90 | 0.76 | 1.07 | 2.22E-01 | 0.524 |
| OID20353 | CDH1 | Cardiometabolic | CDH1 | P12830 | 16 | 68737292 | 68835537 | 0.89 | 0.75 | 1.07 | 2.23E-01 | 0.524 |
| OID20165 | CLEC5A | Cardiometabolic | CLEC5A | Q9NY25 | 7 | 141927357 | 141947007 | 1.11 | 0.94 | 1.31 | 2.23E-01 | 0.524 |
| OID30762 | ITIH4 | Inflammation_II | ITIH4 | Q14624 | 3 | 52812962 | 52830688 | 0.89 | 0.74 | 1.07 | 2.23E-01 | 0.524 |
| OID30628 | CTSE | Inflammation_II | CTSE | P14091 | 1 | 206008535 | 206023909 | 0.90 | 0.76 | 1.07 | 2.24E-01 | 0.525 |
| OID31280 | TEX33 | Oncology_II | TEX33 | O43247 | 22 | 36991120 | 37007851 | 0.89 | 0.73 | 1.08 | 2.24E-01 | 0.525 |
| OID31357 | DHPS | Oncology_II | DHPS | P49366 | 19 | 12675717 | 12681880 | 0.89 | 0.74 | 1.07 | 2.24E-01 | 0.526 |
| OID31351 | AMOTL2 | Oncology_II | AMOTL2 | Q9Y2J4 | 3 | 134355347 | 134375479 | 0.90 | 0.76 | 1.07 | 2.25E-01 | 0.526 |
| OID21444 | CCN4 | Oncology | CCN4 | O95388 | 8 | 133191039 | 133231690 | 0.90 | 0.75 | 1.07 | 2.26E-01 | 0.526 |
| OID30660 | CHAD | Inflammation_II | CHAD | O15335 | 17 | 50464496 | 50468906 | 0.91 | 0.78 | 1.06 | 2.26E-01 | 0.526 |
| OID31171 | CYB5A | Oncology_II | CYB5A | P00167 | 18 | 74250846 | 74291973 | 0.89 | 0.74 | 1.07 | 2.25E-01 | 0.526 |
| OID21273 | PDCD1LG2 | Oncology | PDCD1LG2 | Q9BQ51 | 9 | 5510531 | 5571282 | 1.11 | 0.94 | 1.30 | 2.25E-01 | 0.526 |
| OID31093 | PRSS53 | Neurology_II | PRSS53 | Q2L4Q9 | 16 | 31083437 | 31089628 | 1.11 | 0.94 | 1.32 | 2.25E-01 | 0.526 |
| OID30111 | SAT1 | Cardiometabolic_II | SAT1 | P21673 | X | 23783173 | 23786210 | 0.89 | 0.74 | 1.07 | 2.26E-01 | 0.526 |
| OID30369 | SLC4A1 | Cardiometabolic_II | SLC4A1 | P02730 | 17 | 44248390 | 44268141 | 1.11 | 0.94 | 1.30 | 2.25E-01 | 0.526 |
| OID21505 | WFDC2 | Oncology | WFDC2 | Q14508 | 20 | 45469753 | 45481532 | 1.11 | 0.94 | 1.31 | 2.26E-01 | 0.526 |
| OID20896 | AGR2 | Neurology | AGR2 | O95994 | 7 | 16791811 | 16833433 | 1.10 | 0.94 | 1.28 | 2.27E-01 | 0.526 |
| OID21448 | NUCB2 | Oncology | NUCB2 | P80303 | 11 | 17208153 | 17349980 | 0.90 | 0.76 | 1.07 | 2.26E-01 | 0.526 |
| OID30207 | SIL1 | Cardiometabolic_II | SIL1 | Q9H173 | 5 | 138946724 | 139293557 | 0.88 | 0.72 | 1.08 | 2.26E-01 | 0.526 |
| OID20597 | PLXNA4 | Inflammation | PLXNA4 | Q9HCM2 | 7 | 132123340 | 132648688 | 0.91 | 0.78 | 1.06 | 2.27E-01 | 0.526 |
| OID20707 | MANF | Inflammation | MANF | P55145 | 3 | 51385291 | 51389397 | 0.91 | 0.78 | 1.06 | 2.27E-01 | 0.527 |
| OID20173 | GPNMB | Cardiometabolic | GPNMB | Q14956 | 7 | 23235967 | 23275108 | 0.90 | 0.77 | 1.06 | 2.27E-01 | 0.527 |
| OID21021 | PDCD5 | Neurology | PDCD5 | O14737 | 19 | 32581190 | 32587453 | 0.90 | 0.76 | 1.07 | 2.27E-01 | 0.527 |
| OID20934 | DKK4 | Neurology | DKK4 | Q9UBT3 | 8 | 42374063 | 42377229 | 0.90 | 0.76 | 1.07 | 2.28E-01 | 0.527 |
| OID30187 | FHIP2A | Cardiometabolic_II | FHIP2A | Q5W0V3 | 10 | 114821744 | 114899832 | 0.90 | 0.75 | 1.07 | 2.28E-01 | 0.527 |
| OID31190 | ARHGAP5 | Oncology_II | ARHGAP5 | Q13017 | 14 | 32076114 | 32159728 | 0.90 | 0.76 | 1.07 | 2.29E-01 | 0.528 |
| OID30975 | INPP5J | Neurology_II | INPP5J | Q15735 | 22 | 31122731 | 31134697 | 1.09 | 0.95 | 1.25 | 2.29E-01 | 0.528 |
| OID21490 | NECTIN4 | Oncology | NECTIN4 | Q96NY8 | 1 | 161070998 | 161089558 | 1.10 | 0.94 | 1.27 | 2.29E-01 | 0.528 |
| OID21200 | CCT5 | Oncology | CCT5 | P48643 | 5 | 10249929 | 10266389 | 0.90 | 0.75 | 1.07 | 2.29E-01 | 0.528 |
| OID30696 | ACE | Inflammation_II | ACE | P12821 | 17 | 63477061 | 63498380 | 0.90 | 0.76 | 1.07 | 2.30E-01 | 0.529 |
| OID20586 | LTA | Inflammation | LTA | P01374 | 6 | 31572054 | 31574324 | 0.90 | 0.77 | 1.07 | 2.30E-01 | 0.529 |
| OID20627 | DNAJA2 | Inflammation | DNAJA2 | O60884 | 16 | 46955362 | 46973674 | 0.91 | 0.77 | 1.07 | 2.30E-01 | 0.529 |
| OID20887 | HMOX2 | Neurology | HMOX2 | P30519 | 16 | 4474690 | 4510347 | 0.89 | 0.74 | 1.07 | 2.30E-01 | 0.530 |
| OID30887 | FARSA | Neurology_II | FARSA | Q9Y285 | 19 | 12922479 | 12934037 | 0.90 | 0.75 | 1.07 | 2.31E-01 | 0.530 |
| OID20238 | PLTP | Cardiometabolic | PLTP | P55058 | 20 | 45898621 | 45912155 | 1.11 | 0.94 | 1.31 | 2.31E-01 | 0.530 |
| OID30862 | IGDCC3 | Neurology_II | IGDCC3 | Q8IVU1 | 15 | 65327127 | 65378002 | 0.88 | 0.72 | 1.08 | 2.31E-01 | 0.530 |
| OID20697 | CXCL10 | Inflammation | CXCL10 | P02778 | 4 | 76021118 | 76023497 | 0.90 | 0.75 | 1.07 | 2.32E-01 | 0.531 |
| OID20940 | CDCP1 | Neurology | CDCP1 | Q9H5V8 | 3 | 45082277 | 45146422 | 1.11 | 0.93 | 1.33 | 2.33E-01 | 0.532 |
| OID30117 | ZCCHC8 | Cardiometabolic_II | ZCCHC8 | Q6NZY4 | 12 | 122471599 | 122501073 | 0.89 | 0.72 | 1.08 | 2.33E-01 | 0.533 |
| OID20188 | LPL | Cardiometabolic | LPL | P06858 | 8 | 19901717 | 19967259 | 1.13 | 0.93 | 1.37 | 2.34E-01 | 0.534 |
| OID30144 | CALY | Cardiometabolic_II | CALY | Q9NYX4 | 10 | 133324072 | 133336935 | 1.10 | 0.94 | 1.29 | 2.34E-01 | 0.535 |
| OID30821 | CEND1 | Neurology_II | CEND1 | Q8N111 | 11 | 787115 | 790113 | 1.07 | 0.96 | 1.20 | 2.34E-01 | 0.535 |
| OID20956 | DNMBP | Neurology | DNMBP | Q6XZF7 | 10 | 99875577 | 100009947 | 0.90 | 0.75 | 1.07 | 2.35E-01 | 0.535 |
| OID21028 | SMPD1 | Neurology | SMPD1 | P17405 | 11 | 6390440 | 6394998 | 1.10 | 0.94 | 1.30 | 2.35E-01 | 0.535 |
| OID30326 | TCTN3 | Cardiometabolic_II | TCTN3 | Q6NUS6 | 10 | 95659823 | 95694143 | 0.90 | 0.76 | 1.07 | 2.35E-01 | 0.535 |
| OID30693 | PNLIP | Inflammation_II | PNLIP | P16233 | 10 | 116545931 | 116567855 | 0.92 | 0.79 | 1.06 | 2.36E-01 | 0.536 |
| OID30233 | FSHB | Cardiometabolic_II | FSHB | P01225 | 11 | 30231014 | 30235261 | 0.85 | 0.65 | 1.11 | 2.37E-01 | 0.538 |
| OID30506 | RNF168 | Inflammation_II | RNF168 | Q8IYW5 | 3 | 196468783 | 196503768 | 0.89 | 0.74 | 1.08 | 2.37E-01 | 0.538 |
| OID31457 | DUT | Oncology_II | DUT | P33316 | 15 | 48331011 | 48343373 | 0.90 | 0.77 | 1.07 | 2.37E-01 | 0.538 |
| OID31342 | MAMDC4 | Oncology_II | MAMDC4 | Q6UXC1 | 9 | 136850943 | 136860799 | 0.91 | 0.77 | 1.07 | 2.38E-01 | 0.540 |
| OID31025 | SEPTIN3 | Neurology_II | SEPTIN3 | Q9UH03 | 22 | 41969443 | 41998221 | 1.10 | 0.94 | 1.30 | 2.39E-01 | 0.540 |
| OID20676 | SMPDL3A | Inflammation | SMPDL3A | Q92484 | 6 | 122789049 | 122809720 | 1.10 | 0.94 | 1.30 | 2.39E-01 | 0.540 |
| OID21497 | KDR | Oncology | KDR | P35968 | 4 | 55078481 | 55125595 | 0.91 | 0.77 | 1.07 | 2.40E-01 | 0.542 |
| OID31145 | SH3GLB2 | Neurology_II | SH3GLB2 | Q9NR46 | 9 | 129007036 | 129028331 | 0.91 | 0.77 | 1.07 | 2.40E-01 | 0.542 |
| OID30265 | ZP3 | Cardiometabolic_II | ZP3 | P21754 | 7 | 76397518 | 76442071 | 0.91 | 0.78 | 1.06 | 2.40E-01 | 0.542 |
| OID30952 | CAMLG | Neurology_II | CAMLG | P49069 | 5 | 134738495 | 134752157 | 0.89 | 0.74 | 1.08 | 2.40E-01 | 0.542 |
| OID30949 | AP2B1 | Neurology_II | AP2B1 | P63010 | 17 | 35578046 | 35726413 | 0.90 | 0.76 | 1.07 | 2.41E-01 | 0.543 |
| OID30909 | OSTN | Neurology_II | OSTN | P61366 | 3 | 191199241 | 191265615 | 1.09 | 0.94 | 1.27 | 2.41E-01 | 0.543 |
| OID30906 | LRTM1 | Neurology_II | LRTM1 | Q9HBL6 | 3 | 54918231 | 54967088 | 1.08 | 0.95 | 1.23 | 2.41E-01 | 0.543 |
| OID20137 | ACY1 | Cardiometabolic | ACY1 | Q03154 | 3 | 51983340 | 51989197 | 0.90 | 0.75 | 1.08 | 2.42E-01 | 0.543 |
| OID21230 | CHAC2 | Oncology | CHAC2 | Q8WUX2 | 2 | 53767804 | 53775196 | 0.90 | 0.76 | 1.07 | 2.42E-01 | 0.543 |
| OID20358 | CTSD | Cardiometabolic | CTSD | P07339 | 11 | 1752752 | 1764573 | 1.09 | 0.94 | 1.27 | 2.42E-01 | 0.543 |
| OID21001 | ADAM22 | Neurology | ADAM22 | Q9P0K1 | 7 | 87934143 | 88202889 | 0.91 | 0.77 | 1.07 | 2.42E-01 | 0.544 |
| OID20267 | IL2RA | Cardiometabolic | IL2RA | P01589 | 10 | 6010689 | 6062370 | 1.09 | 0.94 | 1.27 | 2.43E-01 | 0.545 |
| OID20758 | ESM1 | Inflammation | ESM1 | Q9NQ30 | 5 | 54977867 | 55022671 | 1.11 | 0.93 | 1.32 | 2.44E-01 | 0.546 |
| OID31092 | CLSTN3 | Neurology_II | CLSTN3 | Q9BQT9 | 12 | 7129698 | 7158945 | 0.90 | 0.76 | 1.07 | 2.44E-01 | 0.546 |
| OID30301 | ELN | Cardiometabolic_II | ELN | P15502 | 7 | 74027789 | 74069907 | 1.12 | 0.93 | 1.36 | 2.44E-01 | 0.546 |
| OID20933 | GFRA3 | Neurology | GFRA3 | O60609 | 5 | 138252380 | 138274621 | 0.91 | 0.77 | 1.07 | 2.44E-01 | 0.546 |
| OID30143 | TPR | Cardiometabolic_II | TPR | P12270 | 1 | 186311652 | 186375693 | 0.89 | 0.72 | 1.09 | 2.45E-01 | 0.547 |
| OID30575 | BMP10 | Inflammation_II | BMP10 | O95393 | 2 | 68860909 | 68871397 | 0.91 | 0.77 | 1.07 | 2.45E-01 | 0.547 |
| OID20745 | CCL17 | Inflammation | CCL17 | Q92583 | 16 | 57404767 | 57416063 | 0.91 | 0.78 | 1.07 | 2.45E-01 | 0.547 |
| OID30794 | UPK3A | Neurology_II | UPK3A | O75631 | 22 | 45284949 | 45295874 | 1.08 | 0.95 | 1.23 | 2.46E-01 | 0.548 |
| OID31519 | PCYT2 | Oncology_II | PCYT2 | Q99447 | 17 | 81900958 | 81911432 | 0.91 | 0.77 | 1.07 | 2.46E-01 | 0.548 |
| OID20624 | TNFSF12 | Inflammation | TNFSF12 | O43508 | 17 | 7549058 | 7557890 | 0.91 | 0.77 | 1.07 | 2.46E-01 | 0.548 |
| OID20894 | TIGAR | Neurology | TIGAR | Q9NQ88 | 12 | 4307763 | 4360028 | 0.90 | 0.75 | 1.08 | 2.47E-01 | 0.548 |
| OID21263 | GCG | Oncology | GCG | P01275 | 2 | 162142882 | 162152404 | 0.91 | 0.77 | 1.07 | 2.49E-01 | 0.552 |
| OID21116 | IL1R1 | Neurology | IL1R1 | P14778 | 2 | 102064544 | 102179874 | 1.10 | 0.94 | 1.28 | 2.49E-01 | 0.552 |
| OID31440 | LRRC37A2 | Oncology_II | LRRC37A2 | A6NM11 | 17 | 46511508 | 46556910 | 0.91 | 0.78 | 1.07 | 2.49E-01 | 0.552 |
| OID21282 | CDHR2 | Oncology | CDHR2 | Q9BYE9 | 5 | 176542511 | 176595974 | 0.89 | 0.72 | 1.09 | 2.49E-01 | 0.552 |
| OID20055 | CRHR1 | Cardiometabolic | CRHR1 | P34998 | 17 | 45784280 | 45835828 | 0.90 | 0.75 | 1.08 | 2.50E-01 | 0.552 |
| OID30785 | PLG | Inflammation_II | PLG | P00747 | 6 | 160702238 | 160754097 | 0.90 | 0.75 | 1.08 | 2.50E-01 | 0.552 |
| OID20064 | SERPINB5 | Cardiometabolic | SERPINB5 | P36952 | 18 | 63476958 | 63505085 | 1.10 | 0.93 | 1.30 | 2.49E-01 | 0.552 |
| OID30602 | DDI2 | Inflammation_II | DDI2 | Q5TDH0 | 1 | 15617458 | 15669044 | 0.90 | 0.76 | 1.08 | 2.50E-01 | 0.554 |
| OID31070 | CAMSAP1 | Neurology_II | CAMSAP1 | Q5T5Y3 | 9 | 135808487 | 135907546 | 0.91 | 0.77 | 1.07 | 2.51E-01 | 0.554 |
| OID31116 | CHCHD10 | Neurology_II | CHCHD10 | Q8WYQ3 | 22 | 23765834 | 23767972 | 0.90 | 0.74 | 1.08 | 2.51E-01 | 0.554 |
| OID30857 | SCIN | Neurology_II | SCIN | Q9Y6U3 | 7 | 12570577 | 12660182 | 0.90 | 0.76 | 1.08 | 2.51E-01 | 0.554 |
| OID21034 | LBR | Neurology | LBR | Q14739 | 1 | 225401502 | 225428925 | 0.90 | 0.75 | 1.08 | 2.52E-01 | 0.555 |
| OID20762 | CXCL1 | Inflammation | CXCL1 | P09341 | 4 | 73869393 | 73871308 | 0.90 | 0.75 | 1.08 | 2.53E-01 | 0.556 |
| OID20589 | HEXIM1 | Inflammation | HEXIM1 | O94992 | 17 | 45148475 | 45152099 | 0.91 | 0.77 | 1.07 | 2.53E-01 | 0.557 |
| OID30488 | AKR7L | Inflammation_II | AKR7L | Q8NHP1 | 1 | 19265982 | 19274194 | 0.90 | 0.76 | 1.07 | 2.54E-01 | 0.557 |
| OID30922 | CD164L2 | Neurology_II | CD164L2 | Q6UWJ8 | 1 | 27379176 | 27383333 | 0.91 | 0.76 | 1.07 | 2.54E-01 | 0.557 |
| OID30362 | CEACAM6 | Cardiometabolic_II | CEACAM6 | P40199 | 19 | 41750977 | 41772211 | 1.10 | 0.93 | 1.30 | 2.54E-01 | 0.557 |
| OID30330 | IGSF9 | Cardiometabolic_II | IGSF9 | Q9P2J2 | 1 | 159927039 | 159945613 | 0.90 | 0.75 | 1.08 | 2.54E-01 | 0.557 |
| OID30760 | KLKB1 | Inflammation_II | KLKB1 | P03952 | 4 | 186226438 | 186258471 | 0.91 | 0.77 | 1.07 | 2.54E-01 | 0.557 |
| OID30996 | MYCBP2 | Neurology_II | MYCBP2 | O75592 | 13 | 77042474 | 77327094 | 0.90 | 0.76 | 1.07 | 2.54E-01 | 0.557 |
| OID20314 | ACP5 | Cardiometabolic | ACP5 | P13686 | 19 | 11574660 | 11579008 | 1.10 | 0.93 | 1.31 | 2.55E-01 | 0.557 |
| OID30066 | MED21 | Cardiometabolic_II | MED21 | Q13503 | 12 | 27022546 | 27066343 | 0.89 | 0.72 | 1.09 | 2.55E-01 | 0.558 |
| OID31276 | CASP4 | Oncology_II | CASP4 | P49662 | 11 | 104942866 | 104969366 | 0.90 | 0.76 | 1.08 | 2.55E-01 | 0.558 |
| OID30499 | CEMIP2 | Inflammation_II | CEMIP2 | Q9UHN6 | 9 | 71683366 | 71816690 | 1.08 | 0.95 | 1.23 | 2.57E-01 | 0.558 |

| | | | | | | | | | | | | |
|---|---|---|---|---|---|---|---|---|---|---|---|---|
| OID31192 | CLASP1 | Oncology_II | CLASP1 | Q7Z460 | 2 | 121337776 | 121649476 | 0.90 | 0.75 | 1.08 | 2.58E-01 | 0.558 |
| OID20276 | CTSB | Cardiometabolic | CTSB | P07858 | 8 | 11842524 | 11869533 | 1.10 | 0.93 | 1.29 | 2.58E-01 | 0.558 |
| OID31208 | CYTH3 | Oncology_II | CYTH3 | O43739 | 7 | 6161776 | 6272644 | 0.90 | 0.75 | 1.08 | 2.58E-01 | 0.558 |
| OID21313 | DKKL1 | Oncology | DKKL1 | Q9UK85 | 19 | 49361783 | 49375116 | 1.11 | 0.93 | 1.32 | 2.56E-01 | 0.558 |
| OID21337 | ELOA | Oncology | ELOA | Q14241 | 1 | 23743448 | 23762059 | 0.89 | 0.73 | 1.09 | 2.56E-01 | 0.558 |
| OID20471 | GALNT3 | Inflammation | GALNT3 | Q14435 | 2 | 165747588 | 165794659 | 1.10 | 0.94 | 1.28 | 2.57E-01 | 0.558 |
| OID20813 | KCNIP4 | Neurology | KCNIP4 | Q6PIL6 | 4 | 20728606 | 21948772 | 0.89 | 0.74 | 1.08 | 2.56E-01 | 0.558 |
| OID20725 | OMD | Inflammation | OMD | Q99983 | 9 | 92412380 | 92424471 | 1.11 | 0.93 | 1.32 | 2.58E-01 | 0.558 |
| OID20922 | PHOSPHO1 | Neurology | PHOSPHO1 | Q8TCT1 | 17 | 49223366 | 49230787 | 1.10 | 0.93 | 1.30 | 2.56E-01 | 0.558 |
| OID20444 | PRKAB1 | Inflammation | PRKAB1 | Q9Y478 | 12 | 119667864 | 119681624 | 0.90 | 0.76 | 1.08 | 2.57E-01 | 0.558 |
| OID30635 | RIDA | Inflammation_II | RIDA | P52758 | 8 | 98102344 | 98117171 | 0.90 | 0.75 | 1.08 | 2.58E-01 | 0.558 |
| OID30432 | STX5 | Inflammation_II | STX5 | Q13190 | 11 | 62806860 | 62832051 | 0.89 | 0.73 | 1.09 | 2.57E-01 | 0.558 |
| OID21032 | SUMF2 | Neurology | SUMF2 | Q8NBJ7 | 7 | 56064002 | 56080670 | 0.91 | 0.77 | 1.07 | 2.58E-01 | 0.558 |
| OID30641 | CD72 | Inflammation_II | CD72 | P21854 | 9 | 35609982 | 35646810 | 0.91 | 0.77 | 1.07 | 2.59E-01 | 0.559 |
| OID31244 | MANEAL | Oncology_II | MANEAL | Q5VSG8 | 1 | 37793847 | 37801137 | 0.91 | 0.78 | 1.07 | 2.59E-01 | 0.559 |
| OID21375 | SIRT2 | Oncology | SIRT2 | Q8IXJ6 | 19 | 38878555 | 38899862 | 0.91 | 0.78 | 1.07 | 2.59E-01 | 0.559 |
| OID30103 | TEF | Cardiometabolic_II | TEF | Q10587 | 22 | 41367333 | 41399326 | 0.90 | 0.75 | 1.08 | 2.59E-01 | 0.559 |
| OID31307 | KIAA1549 | Oncology_II | KIAA1549 | Q9HCM3 | 7 | 138831381 | 138981389 | 0.91 | 0.78 | 1.07 | 2.60E-01 | 0.561 |
| OID21035 | LAMP2 | Neurology | LAMP2 | P13473 | X | 120426148 | 120469365 | 0.91 | 0.78 | 1.07 | 2.60E-01 | 0.561 |
| OID20310 | PTPRF | Cardiometabolic | PTPRF | P10586 | 1 | 43525187 | 43623666 | 0.91 | 0.77 | 1.07 | 2.61E-01 | 0.561 |
| OID21415 | RSPO3 | Oncology | RSPO3 | Q9BXY4 | 6 | 127118671 | 127199481 | 0.91 | 0.77 | 1.07 | 2.62E-01 | 0.563 |
| OID21322 | ST3GAL1 | Oncology | ST3GAL1 | Q11201 | 8 | 133454848 | 133571940 | 1.08 | 0.94 | 1.25 | 2.62E-01 | 0.563 |
| OID30053 | THRAP3 | Cardiometabolic_II | THRAP3 | Q9Y2W1 | 1 | 36224432 | 36305357 | 1.08 | 0.94 | 1.25 | 2.62E-01 | 0.563 |
| OID31177 | MAPRE3 | Oncology_II | MAPRE3 | Q9UPY8 | 2 | 26970637 | 27027219 | 0.90 | 0.75 | 1.08 | 2.63E-01 | 0.563 |
| OID30082 | ITPA | Cardiometabolic_II | ITPA | Q9BY32 | 20 | 3208868 | 3223870 | 0.91 | 0.77 | 1.07 | 2.63E-01 | 0.564 |
| OID20289 | CPB1 | Cardiometabolic | CPB1 | P15086 | 3 | 148791102 | 148860187 | 0.91 | 0.77 | 1.07 | 2.64E-01 | 0.566 |
| OID20304 | SIRPA | Cardiometabolic | SIRPA | P78324 | 20 | 1894167 | 1940592 | 1.10 | 0.93 | 1.30 | 2.64E-01 | 0.566 |
| OID31292 | SHPK | Oncology_II | SHPK | Q9UHJ6 | 17 | 3608240 | 3636250 | 0.90 | 0.76 | 1.08 | 2.65E-01 | 0.566 |
| OID30353 | TOMM20 | Cardiometabolic_II | TOMM20 | Q15388 | 1 | 235109341 | 235128837 | 0.91 | 0.77 | 1.07 | 2.65E-01 | 0.566 |
| OID30812 | RANBP2 | Neurology_II | RANBP2 | P49792 | 2 | 108719482 | 108785810 | 0.91 | 0.77 | 1.07 | 2.65E-01 | 0.567 |
| OID21189 | UXS1 | Oncology | UXS1 | Q8NBZ7 | 2 | 106093308 | 106194301 | 0.90 | 0.75 | 1.08 | 2.66E-01 | 0.568 |
| OID21467 | ADM | Oncology | ADM | P35318 | 11 | 10305073 | 10307397 | 1.12 | 0.92 | 1.37 | 2.68E-01 | 0.568 |
| OID30871 | ARHGEF5 | Neurology_II | ARHGEF5 | Q12774 | 7 | 144355288 | 144380632 | 1.09 | 0.94 | 1.27 | 2.68E-01 | 0.568 |
| OID30581 | CST1 | Inflammation_II | CST1 | P01037 | 20 | 23747562 | 23751268 | 0.91 | 0.78 | 1.07 | 2.68E-01 | 0.568 |
| OID31491 | IFNAR1 | Oncology_II | IFNAR1 | P17181 | 21 | 33324387 | 33359864 | 0.91 | 0.77 | 1.08 | 2.68E-01 | 0.568 |
| OID21416 | ITGAV | Oncology | ITGAV | P06756 | 2 | 186590056 | 186680901 | 0.91 | 0.77 | 1.08 | 2.68E-01 | 0.568 |
| OID21439 | MMP12 | Oncology | MMP12 | P39900 | 11 | 102862736 | 102874982 | 1.10 | 0.93 | 1.31 | 2.68E-01 | 0.568 |
| OID30377 | MUC2 | Cardiometabolic_II | MUC2 | Q02817 | 11 | 1074875 | 1110511 | 1.09 | 0.94 | 1.27 | 2.67E-01 | 0.568 |
| OID31281 | UPK3BL1 | Oncology_II | UPK3BL1 | B0FP48 | 7 | 102637025 | 102642791 | 0.89 | 0.72 | 1.10 | 2.67E-01 | 0.568 |
| OID30708 | APOL1 | Inflammation_II | APOL1 | O14791 | 22 | 36253071 | 36267530 | 0.90 | 0.76 | 1.08 | 2.69E-01 | 0.570 |
| OID21114 | SOD2 | Neurology | SOD2 | P04179 | 6 | 159669069 | 159762529 | 1.09 | 0.93 | 1.29 | 2.69E-01 | 0.570 |
| OID20549 | PSMG3 | Inflammation | PSMG3 | Q9BT73 | 7 | 1567332 | 1571005 | 0.91 | 0.76 | 1.08 | 2.69E-01 | 0.571 |
| OID30394 | CD248 | Cardiometabolic_II | CD248 | Q9HCU0 | 11 | 66314494 | 66317044 | 1.10 | 0.93 | 1.30 | 2.70E-01 | 0.571 |
| OID20530 | LSP1 | Inflammation | LSP1 | P33241 | 11 | 1850904 | 1892267 | 0.91 | 0.76 | 1.08 | 2.70E-01 | 0.571 |
| OID20270 | ESAM | Cardiometabolic | ESAM | Q96AP7 | 11 | 124752583 | 124762290 | 0.91 | 0.77 | 1.08 | 2.71E-01 | 0.572 |
| OID21018 | CD8A | Neurology | CD8A | P01732 | 2 | 86784610 | 86808396 | 0.91 | 0.78 | 1.07 | 2.71E-01 | 0.572 |
| OID21060 | CPA2 | Neurology | CPA2 | P48052 | 7 | 130266863 | 130289798 | 1.09 | 0.93 | 1.28 | 2.72E-01 | 0.573 |
| OID31510 | NFU1 | Oncology_II | NFU1 | Q9UMS0 | 2 | 69396113 | 69437628 | 0.91 | 0.77 | 1.08 | 2.72E-01 | 0.573 |
| OID30529 | S100A13 | Inflammation_II | S100A13 | Q99584 | 1 | 153618787 | 153631360 | 0.91 | 0.77 | 1.08 | 2.72E-01 | 0.574 |
| OID20893 | CASP10 | Neurology | CASP10 | Q92851 | 2 | 201182898 | 201229406 | 0.91 | 0.78 | 1.07 | 2.73E-01 | 0.574 |
| OID21255 | ARHGAP25 | Oncology | ARHGAP25 | P42331 | 2 | 68679601 | 68826833 | 0.91 | 0.76 | 1.08 | 2.73E-01 | 0.575 |
| OID30990 | SPACA5_SPAC/ | Neurology_II | ACA5B;SPACA5 | Q96QH8;Q96QH8 | X;X | 48130626;48004336 | 48132620;48009733 | 0.90 | 0.74 | 1.09 | 2.73E-01 | 0.575 |
| OID21209 | CEP20 | Oncology | CEP20 | Q96NB1 | 16 | 15865719 | 15888625 | 0.91 | 0.77 | 1.08 | 2.75E-01 | 0.577 |
| OID21177 | RARRES1 | Oncology | RARRES1 | P49788 | 3 | 158696892 | 158732489 | 0.90 | 0.74 | 1.09 | 2.75E-01 | 0.577 |
| OID20654 | C1QA | Inflammation | C1QA | P02745 | 1 | 22636628 | 22639678 | 1.10 | 0.93 | 1.30 | 2.75E-01 | 0.578 |
| OID30626 | FUOM | Inflammation_II | FUOM | A2VDF0 | 10 | 133355158 | 133358025 | 0.92 | 0.78 | 1.07 | 2.76E-01 | 0.579 |
| OID30569 | AFAP1 | Inflammation_II | AFAP1 | Q8N556 | 4 | 7758714 | 7939926 | 0.91 | 0.77 | 1.08 | 2.77E-01 | 0.579 |
| OID30361 | LRRC59 | Cardiometabolic_II | LRRC59 | Q96AG4 | 17 | 50375059 | 50397523 | 0.91 | 0.77 | 1.08 | 2.77E-01 | 0.579 |
| OID20392 | LCN2 | Cardiometabolic | LCN2 | P80188 | 9 | 128149071 | 128153453 | 0.91 | 0.77 | 1.08 | 2.77E-01 | 0.580 |
| OID30791 | IFNL2 | Neurology_II | IFNL2 | Q8IZJ0 | 19 | 39268396 | 39270188 | 0.91 | 0.76 | 1.08 | 2.78E-01 | 0.581 |
| OID31275 | ALMS1 | Oncology_II | ALMS1 | Q8TCU4 | 2 | 73385758 | 73625166 | 0.90 | 0.75 | 1.09 | 2.78E-01 | 0.581 |
| OID30255 | DLL4 | Cardiometabolic_II | DLL4 | Q9NR61 | 15 | 40929340 | 40939073 | 0.88 | 0.71 | 1.10 | 2.78E-01 | 0.581 |
| OID20748 | LGALS4 | Inflammation | LGALS4 | P56470 | 19 | 38801671 | 38812945 | 1.09 | 0.93 | 1.28 | 2.79E-01 | 0.581 |
| OID20687 | MMP10 | Inflammation | MMP10 | P09238 | 11 | 102770502 | 102780628 | 0.91 | 0.77 | 1.08 | 2.79E-01 | 0.581 |
| OID31378 | NMI | Oncology_II | NMI | Q13287 | 2 | 151270470 | 151289894 | 0.91 | 0.77 | 1.08 | 2.79E-01 | 0.581 |
| OID30444 | PALLD | Inflammation_II | PALLD | Q8WX93 | 4 | 168497052 | 168928457 | 0.90 | 0.75 | 1.09 | 2.79E-01 | 0.581 |
| OID30202 | PYDC1 | Cardiometabolic_II | PYDC1 | Q8WXC3 | 16 | 31215962 | 31217135 | 1.08 | 0.94 | 1.25 | 2.79E-01 | 0.581 |
| OID30544 | FGF3 | Inflammation_II | FGF3 | P11487 | 11 | 69809968 | 69819416 | 1.08 | 0.94 | 1.25 | 2.81E-01 | 0.584 |
| OID31008 | GNAS | Neurology_II | GNAS | O95467 | 20 | 58839718 | 58911192 | 0.91 | 0.76 | 1.08 | 2.81E-01 | 0.584 |
| OID20245 | FABP4 | Cardiometabolic | FABP4 | P15090 | 8 | 81478419 | 81483236 | 0.89 | 0.71 | 1.10 | 2.82E-01 | 0.584 |
| OID30565 | NXPH3 | Inflammation_II | NXPH3 | O95157 | 17 | 49575871 | 49583827 | 0.91 | 0.77 | 1.08 | 2.82E-01 | 0.584 |
| OID20382 | FCN2 | Cardiometabolic | FCN2 | Q15485 | 9 | 134880810 | 134887523 | 0.91 | 0.77 | 1.08 | 2.82E-01 | 0.585 |
| OID20458 | CXCL14 | Inflammation | CXCL14 | O95715 | 5 | 135570679 | 135579279 | 0.89 | 0.72 | 1.10 | 2.83E-01 | 0.586 |
| OID30982 | TPSD1 | Neurology_II | TPSD1 | Q9BZJ3 | 16 | 1256059 | 1259008 | 0.91 | 0.78 | 1.08 | 2.83E-01 | 0.586 |
| OID20674 | CCL25 | Inflammation | CCL25 | O15444 | 19 | 8052318 | 8062660 | 1.10 | 0.93 | 1.30 | 2.84E-01 | 0.587 |
| OID20420 | IL10RA | Inflammation | IL10RA | Q13651 | 11 | 117986370 | 118003037 | 0.90 | 0.74 | 1.09 | 2.84E-01 | 0.587 |
| OID21110 | CD99 | Neurology | CD99 | P14209 | X | 2691187 | 2741309 | 0.90 | 0.75 | 1.09 | 2.85E-01 | 0.588 |
| OID30442 | ST8SIA1 | Inflammation_II | ST8SIA1 | Q92185 | 12 | 22063773 | 22437041 | 0.89 | 0.73 | 1.10 | 2.85E-01 | 0.588 |
| OID20943 | SCARB2 | Neurology | SCARB2 | Q14108 | 4 | 76158737 | 76234536 | 1.09 | 0.93 | 1.28 | 2.85E-01 | 0.588 |
| OID30742 | PGLYRP2 | Inflammation_II | PGLYRP2 | Q96PD5 | 19 | 15468645 | 15498956 | 0.92 | 0.78 | 1.08 | 2.85E-01 | 0.588 |
| OID20906 | SKAP1 | Neurology | SKAP1 | Q86WV1 | 17 | 48133442 | 48430275 | 0.91 | 0.77 | 1.08 | 2.86E-01 | 0.589 |
| OID21214 | SLAMF8 | Oncology | SLAMF8 | Q9P0V8 | 1 | 159826811 | 159837492 | 1.09 | 0.93 | 1.29 | 2.86E-01 | 0.589 |
| OID30161 | ANK2 | Cardiometabolic_II | ANK2 | Q01484 | 4 | 112818032 | 113384221 | 1.08 | 0.93 | 1.26 | 2.87E-01 | 0.590 |
| OID31265 | CA7 | Oncology_II | CA7 | P43166 | 16 | 66844414 | 66854153 | 0.91 | 0.77 | 1.08 | 2.87E-01 | 0.590 |
| OID30705 | RNASE4 | Inflammation_II | RNASE4 | P34096 | 14 | 20684560 | 20701216 | 1.09 | 0.93 | 1.27 | 2.87E-01 | 0.590 |
| OID21196 | KIR2DL3 | Oncology | KIR2DL3 | P43628 | 19 | 54738513 | 54753052 | 1.09 | 0.93 | 1.28 | 2.88E-01 | 0.591 |
| OID20440 | IL24 | Inflammation | IL24 | Q13007 | 1 | 206897443 | 206904139 | 0.92 | 0.78 | 1.08 | 2.88E-01 | 0.591 |
| OID21468 | IDUA | Oncology | IDUA | P35475 | 4 | 986997 | 1004564 | 0.92 | 0.78 | 1.08 | 2.89E-01 | 0.592 |
| OID31271 | TRIM24 | Oncology_II | TRIM24 | O15164 | 7 | 138460259 | 138589996 | 0.91 | 0.77 | 1.08 | 2.89E-01 | 0.592 |
| OID31485 | CDC42BPB | Oncology_II | CDC42BPB | Q9Y5S2 | 14 | 102932380 | 103057549 | 0.91 | 0.78 | 1.08 | 2.90E-01 | 0.593 |
| OID30980 | ATXN2L | Neurology_II | ATXN2L | Q8WWM7 | 16 | 28822999 | 28837237 | 0.91 | 0.75 | 1.09 | 2.90E-01 | 0.593 |
| OID30756 | CLEC3B | Inflammation_II | CLEC3B | P05452 | 3 | 45001548 | 45036071 | 0.91 | 0.77 | 1.08 | 2.90E-01 | 0.593 |
| OID21076 | BCAM | Neurology | BCAM | P50895 | 19 | 44809071 | 44821421 | 1.09 | 0.93 | 1.29 | 2.91E-01 | 0.593 |
| OID20885 | GKN1 | Neurology | GKN1 | Q9NS71 | 2 | 68974573 | 68980980 | 1.09 | 0.93 | 1.28 | 2.91E-01 | 0.593 |
| OID21022 | NSFL1C | Neurology | NSFL1C | Q9UNZ2 | 20 | 1442162 | 1473842 | 0.91 | 0.77 | 1.08 | 2.91E-01 | 0.594 |
| OID20206 | CD69 | Cardiometabolic | CD69 | Q07108 | 12 | 9752486 | 9760901 | 0.92 | 0.78 | 1.08 | 2.92E-01 | 0.595 |
| OID20901 | CRIP2 | Neurology | CRIP2 | P52943 | 14 | 105472962 | 105480162 | 0.91 | 0.76 | 1.09 | 2.93E-01 | 0.595 |
| OID20909 | GGT5 | Neurology | GGT5 | P36269 | 22 | 24219654 | 24245108 | 1.09 | 0.93 | 1.29 | 2.93E-01 | 0.595 |
| OID31392 | KIAA2013 | Oncology_II | KIAA2013 | Q8IYS2 | 1 | 11919591 | 11926428 | 0.90 | 0.73 | 1.10 | 2.93E-01 | 0.595 |
| OID21409 | SLITRK2 | Oncology | SLITRK2 | Q9H156 | X | 145817829 | 145829856 | 1.09 | 0.93 | 1.28 | 2.93E-01 | 0.596 |
| OID31481 | KAZN | Oncology_II | KAZN | Q674X7 | 1 | 13892792 | 15118043 | 0.92 | 0.78 | 1.08 | 2.94E-01 | 0.596 |
| OID20749 | LHPP | Inflammation | LHPP | Q9H008 | 10 | 124461823 | 124617888 | 0.91 | 0.76 | 1.09 | 2.94E-01 | 0.596 |
| OID21269 | ACAA1 | Oncology | ACAA1 | P09110 | 3 | 38103129 | 38137242 | 0.91 | 0.77 | 1.08 | 2.94E-01 | 0.597 |

| | | | | | | | | | | | | |
|---|---|---|---|---|---|---|---|---|---|---|---|---|
| OID20345 | VASN | Cardiometabolic | VASN | Q6EMK4 | 16 | 4371848 | 4383538 | 0.92 | 0.78 | 1.08 | 2.95E-01 | 0.598 |
| OID21346 | RET | Oncology | RET | P07949 | 10 | 43077064 | 43130351 | 0.92 | 0.78 | 1.08 | 2.98E-01 | 0.603 |
| OID20578 | ICAM4 | Inflammation | ICAM4 | Q14773 | 19 | 10286955 | 10288522 | 1.09 | 0.93 | 1.29 | 2.99E-01 | 0.605 |
| OID20066 | GRK5 | Cardiometabolic | GRK5 | P34947 | 10 | 119207571 | 119459745 | 0.91 | 0.75 | 1.09 | 3.00E-01 | 0.607 |
| OID20928 | CLEC10A | Neurology | CLEC10A | Q8IUN9 | 17 | 7074537 | 7080307 | 0.92 | 0.78 | 1.08 | 3.01E-01 | 0.607 |
| OID31310 | IFIT1 | Oncology_II | IFIT1 | P09914 | 10 | 89392546 | 89406487 | 0.92 | 0.78 | 1.08 | 3.01E-01 | 0.607 |
| OID20605 | IL32 | Inflammation | IL32 | P24001 | 16 | 3065297 | 3082192 | 1.09 | 0.93 | 1.29 | 3.01E-01 | 0.607 |
| OID20268 | PDGFRB | Cardiometabolic | PDGFRB | P09619 | 5 | 150113839 | 150155872 | 1.10 | 0.92 | 1.30 | 3.01E-01 | 0.607 |
| OID31352 | S100A3 | Oncology_II | S100A3 | P33764 | 1 | 153547329 | 153549258 | 0.89 | 0.71 | 1.11 | 3.01E-01 | 0.607 |
| OID20603 | CD200 | Inflammation | CD200 | P41217 | 3 | 112332347 | 112362812 | 0.91 | 0.77 | 1.09 | 3.02E-01 | 0.608 |
| OID31521 | NDUFB7 | Oncology_II | NDUFB7 | P17568 | 19 | 14566078 | 14572066 | 0.92 | 0.78 | 1.08 | 3.02E-01 | 0.608 |
| OID21355 | VPS37A | Oncology | VPS37A | Q8NEZ2 | 8 | 17246931 | 17302427 | 0.92 | 0.78 | 1.08 | 3.02E-01 | 0.608 |
| OID30870 | CORO6 | Neurology_II | CORO6 | Q6QEF8 | 17 | 29614756 | 29622912 | 0.91 | 0.77 | 1.09 | 3.04E-01 | 0.610 |
| OID20339 | CD59 | Cardiometabolic | CD59 | P13987 | 11 | 33703010 | 33736479 | 1.09 | 0.93 | 1.27 | 3.04E-01 | 0.610 |
| OID30792 | ARMCX2 | Neurology_II | ARMCX2 | Q7L311 | X | 101655281 | 101659850 | 0.91 | 0.77 | 1.09 | 3.05E-01 | 0.611 |
| OID21006 | CNTN5 | Neurology | CNTN5 | O94779 | 11 | 99020949 | 100358885 | 0.91 | 0.76 | 1.09 | 3.05E-01 | 0.611 |
| OID20501 | FABP9 | Inflammation | FABP9 | Q0Z7S8 | 8 | 81458253 | 81461579 | 1.08 | 0.93 | 1.26 | 3.05E-01 | 0.611 |
| OID20593 | MICB_MICA | Inflammation | MICB;MICA | Q29980;Q29983 | 6;6 | 31494881;31399784 | 31511124;31415315 | 1.10 | 0.92 | 1.32 | 3.05E-01 | 0.611 |
| OID20465 | NRTN | Inflammation | NRTN | Q99748 | 19 | 5805067 | 5828324 | 0.91 | 0.77 | 1.09 | 3.05E-01 | 0.611 |
| OID21182 | CAMKK1 | Oncology | CAMKK1 | Q8N5S9 | 17 | 3860315 | 3894891 | 0.92 | 0.78 | 1.08 | 3.06E-01 | 0.611 |
| OID20262 | CD209 | Cardiometabolic | CD209 | Q9NNX6 | 19 | 7739993 | 7747564 | 1.09 | 0.92 | 1.29 | 3.07E-01 | 0.611 |
| OID20162 | EIF4EBP1 | Cardiometabolic | EIF4EBP1 | Q13541 | 8 | 38030534 | 38060365 | 0.91 | 0.77 | 1.09 | 3.06E-01 | 0.611 |
| OID30132 | HADH | Cardiometabolic_II | HADH | Q16836 | 4 | 107989714 | 108035241 | 1.07 | 0.94 | 1.22 | 3.07E-01 | 0.611 |
| OID30841 | IL13RA2 | Neurology_II | IL13RA2 | Q14627 | X | 115003975 | 115019977 | 0.92 | 0.78 | 1.08 | 3.06E-01 | 0.611 |
| OID31099 | PTPRR | Neurology_II | PTPRR | Q15256 | 12 | 70638073 | 70920738 | 0.92 | 0.78 | 1.08 | 3.07E-01 | 0.611 |
| OID20602 | SLAMF7 | Inflammation | SLAMF7 | Q9NQ25 | 1 | 160739057 | 160754821 | 1.09 | 0.93 | 1.27 | 3.07E-01 | 0.611 |
| OID31220 | SLC28A1 | Oncology_II | SLC28A1 | O00337 | 15 | 84884654 | 84945798 | 0.91 | 0.77 | 1.09 | 3.07E-01 | 0.611 |
| OID20202 | GDF2 | Cardiometabolic | GDF2 | Q9UK05 | 10 | 47322454 | 47327588 | 1.09 | 0.92 | 1.30 | 3.08E-01 | 0.612 |
| OID30074 | MAP1LC3B2 | Cardiometabolic_II | MAP1LC3B2 | A6NCE7 | 12 | 116548105 | 116576606 | 0.91 | 0.76 | 1.09 | 3.09E-01 | 0.613 |
| OID30149 | PROCR | Cardiometabolic_II | PROCR | Q9UNN8 | 20 | 35172072 | 35216240 | 0.90 | 0.73 | 1.10 | 3.09E-01 | 0.613 |
| OID30349 | ADGRF5 | Cardiometabolic_II | ADGRF5 | Q8IZF2 | 6 | 46852522 | 46954943 | 0.92 | 0.78 | 1.08 | 3.11E-01 | 0.615 |
| OID20804 | C19orf12 | Neurology | C19orf12 | Q9NSK7 | 19 | 29698937 | 29715789 | 0.91 | 0.76 | 1.09 | 3.11E-01 | 0.615 |
| OID20715 | FGF19 | Inflammation | FGF19 | O95750 | 11 | 69698238 | 69704022 | 1.09 | 0.92 | 1.28 | 3.10E-01 | 0.615 |
| OID30071 | HIP1 | Cardiometabolic_II | HIP1 | O00291 | 7 | 75533298 | 75738962 | 0.90 | 0.74 | 1.10 | 3.11E-01 | 0.615 |
| OID21290 | LPCAT2 | Oncology | LPCAT2 | Q7L5N7 | 16 | 55509072 | 55586666 | 0.89 | 0.72 | 1.11 | 3.10E-01 | 0.615 |
| OID21458 | NCS1 | Oncology | NCS1 | P62166 | 9 | 130172404 | 130237303 | 1.09 | 0.92 | 1.29 | 3.10E-01 | 0.615 |
| OID30462 | NPHS2 | Inflammation_II | NPHS2 | Q9NP85 | 1 | 179550539 | 179575952 | 1.09 | 0.93 | 1.27 | 3.10E-01 | 0.615 |
| OID20587 | PIK3AP1 | Inflammation | PIK3AP1 | Q6ZUJ8 | 10 | 96593315 | 96720514 | 0.92 | 0.77 | 1.09 | 3.12E-01 | 0.616 |
| OID30592 | HMCN2 | Inflammation_II | HMCN2 | Q8NDA2 | 9 | 130265760 | 130434123 | 0.92 | 0.79 | 1.08 | 3.12E-01 | 0.616 |
| OID20328 | CCL15 | Cardiometabolic | CCL15 | Q16663 | 17 | 35996440 | 36001553 | 1.09 | 0.92 | 1.28 | 3.12E-01 | 0.616 |
| OID30932 | DIPK1C | Neurology_II | DIPK1C | Q0P6D2 | 18 | 74434775 | 74457944 | 0.91 | 0.76 | 1.09 | 3.14E-01 | 0.618 |
| OID21379 | GCNT1 | Oncology | GCNT1 | Q02742 | 9 | 76419850 | 76651203 | 0.92 | 0.77 | 1.09 | 3.15E-01 | 0.621 |
| OID30685 | SHBG | Inflammation_II | SHBG | P04278 | 17 | 7613946 | 7633382 | 1.10 | 0.92 | 1.32 | 3.15E-01 | 0.621 |
| OID31222 | SHC1 | Oncology_II | SHC1 | P29353 | 1 | 154962298 | 154974395 | 0.92 | 0.78 | 1.08 | 3.16E-01 | 0.621 |
| OID20780 | B4GALT1 | Inflammation | B4GALT1 | P15291 | 9 | 33104082 | 33167356 | 1.09 | 0.92 | 1.28 | 3.16E-01 | 0.621 |
| OID31151 | HEPACAM2 | Neurology_II | HEPACAM2 | A8MVW5 | 7 | 93188534 | 93226469 | 1.09 | 0.92 | 1.28 | 3.16E-01 | 0.621 |
| OID30589 | TNFRSF17 | Inflammation_II | TNFRSF17 | Q02223 | 16 | 11965210 | 11968068 | 0.92 | 0.79 | 1.08 | 3.16E-01 | 0.621 |
| OID31314 | CENPJ | Oncology_II | CENPJ | Q9HC77 | 13 | 24882629 | 24922889 | 0.92 | 0.77 | 1.09 | 3.17E-01 | 0.622 |
| OID20535 | IL7 | Inflammation | IL7 | P13232 | 8 | 78675743 | 78805523 | 0.92 | 0.78 | 1.08 | 3.17E-01 | 0.622 |
| OID21012 | TXNDC5 | Neurology | TXNDC5 | Q8NBS9 | 6 | 7881517 | 7910788 | 0.92 | 0.77 | 1.09 | 3.18E-01 | 0.622 |
| OID30934 | STX1B | Neurology_II | STX1B | P61266 | 16 | 30989256 | 31010638 | 0.91 | 0.76 | 1.09 | 3.18E-01 | 0.624 |
| OID30096 | UBQLN3 | Cardiometabolic_II | UBQLN3 | Q9H347 | 11 | 5507300 | 5509958 | 1.07 | 0.94 | 1.22 | 3.19E-01 | 0.625 |
| OID20099 | CLEC1A | Cardiometabolic | CLEC1A | Q8NC01 | 12 | 10069554 | 10111627 | 1.08 | 0.93 | 1.26 | 3.19E-01 | 0.625 |
| OID30632 | TP53I3 | Inflammation_II | TP53I3 | Q53FA7 | 2 | 24077433 | 24085861 | 0.92 | 0.77 | 1.09 | 3.20E-01 | 0.625 |
| OID30885 | TRIM40 | Neurology_II | TRIM40 | Q6P9F5 | 6 | 30136124 | 30148735 | 0.92 | 0.78 | 1.08 | 3.20E-01 | 0.625 |
| OID31492 | MTSS1 | Oncology_II | MTSS1 | O43312 | 8 | 124550784 | 124728473 | 0.92 | 0.78 | 1.08 | 3.20E-01 | 0.625 |
| OID21456 | CRISP2 | Oncology | CRISP2 | P16562 | 6 | 49692358 | 49713590 | 0.92 | 0.77 | 1.09 | 3.23E-01 | 0.626 |
| OID31299 | CTAG1A_CTAG1 | Oncology_II | TAG1A;CTAG1B | P78358;P78358 | X;X | 4585133;154617609 | 54586816;154619282 | 0.91 | 0.76 | 1.09 | 3.22E-01 | 0.626 |
| OID21507 | FGFBP1 | Oncology | FGFBP1 | Q14512 | 4 | 15935577 | 15938740 | 1.09 | 0.92 | 1.29 | 3.23E-01 | 0.626 |
| OID20960 | FKBP4 | Neurology | FKBP4 | Q02790 | 12 | 2794970 | 2805423 | 0.92 | 0.78 | 1.08 | 3.22E-01 | 0.626 |
| OID30967 | MAG | Neurology_II | MAG | P20916 | 19 | 35292125 | 35313807 | 0.92 | 0.79 | 1.08 | 3.23E-01 | 0.626 |
| OID30501 | MCEMP1 | Inflammation_II | MCEMP1 | Q8IX19 | 19 | 7677088 | 7679829 | 0.92 | 0.79 | 1.08 | 3.22E-01 | 0.626 |
| OID20468 | PRKCQ | Inflammation | PRKCQ | Q04759 | 10 | 6427143 | 6580301 | 1.08 | 0.93 | 1.25 | 3.22E-01 | 0.626 |
| OID30913 | RBP1 | Neurology_II | RBP1 | P09455 | 3 | 139517434 | 139539829 | 0.92 | 0.77 | 1.09 | 3.22E-01 | 0.626 |
| OID20272 | TNFSF13B | Cardiometabolic | TNFSF13B | Q9Y275 | 13 | 108251240 | 108308484 | 1.08 | 0.92 | 1.27 | 3.22E-01 | 0.626 |
| OID21270 | TPMT | Oncology | TPMT | P51580 | 6 | 18128311 | 18155077 | 0.92 | 0.78 | 1.09 | 3.23E-01 | 0.626 |
| OID31094 | KIAA0319 | Neurology_II | KIAA0319 | Q5VV43 | 6 | 24544104 | 24646191 | 0.92 | 0.78 | 1.09 | 3.24E-01 | 0.628 |
| OID30391 | PRAP1 | Cardiometabolic_II | PRAP1 | Q96NZ9 | 10 | 133347368 | 133352683 | 0.91 | 0.75 | 1.10 | 3.25E-01 | 0.629 |
| OID30078 | FDX1 | Cardiometabolic_II | FDX1 | P10109 | 11 | 110429948 | 110464884 | 0.91 | 0.77 | 1.09 | 3.25E-01 | 0.629 |
| OID20220 | GH1 | Cardiometabolic | GH1 | P01241 | 17 | 63917200 | 63918839 | 1.09 | 0.91 | 1.31 | 3.25E-01 | 0.629 |
| OID21289 | SEZ6L2 | Oncology | SEZ6L2 | Q6UXD5 | 16 | 29871159 | 29899550 | 0.91 | 0.76 | 1.09 | 3.27E-01 | 0.631 |
| OID31097 | GSTT2B | Neurology_II | GSTT2B | P0CG30 | 22 | 23957414 | 23961195 | 0.92 | 0.78 | 1.08 | 3.27E-01 | 0.632 |
| OID21383 | ALPP | Oncology | ALPP | P05187 | 2 | 232378724 | 232382889 | 1.10 | 0.91 | 1.32 | 3.28E-01 | 0.633 |
| OID30615 | BMPER | Inflammation_II | BMPER | Q8N8U9 | 7 | 33904308 | 34156427 | 0.92 | 0.78 | 1.09 | 3.30E-01 | 0.634 |
| OID20152 | CGREF1 | Cardiometabolic | CGREF1 | Q99674 | 2 | 27098889 | 27119128 | 1.08 | 0.92 | 1.26 | 3.29E-01 | 0.634 |
| OID20307 | CNTN1 | Cardiometabolic | CNTN1 | Q12860 | 12 | 40692439 | 41072415 | 1.09 | 0.92 | 1.30 | 3.29E-01 | 0.634 |
| OID30223 | DMD | Cardiometabolic_II | DMD | P11532 | X | 31097677 | 33339609 | 0.91 | 0.76 | 1.10 | 3.29E-01 | 0.634 |
| OID20744 | DNPH1 | Inflammation | DNPH1 | O43598 | 6 | 43225629 | 43229481 | 0.92 | 0.78 | 1.09 | 3.30E-01 | 0.634 |
| OID30445 | FOXJ3 | Inflammation_II | FOXJ3 | Q9UPW0 | 1 | 42176539 | 42335877 | 0.91 | 0.76 | 1.10 | 3.30E-01 | 0.634 |
| OID30334 | VIT | Cardiometabolic_II | VIT | Q6UXI7 | 2 | 36696690 | 36814794 | 0.92 | 0.77 | 1.09 | 3.30E-01 | 0.634 |
| OID20118 | CLTA | Cardiometabolic | CLTA | P09496 | 9 | 36190856 | 36304781 | 0.92 | 0.77 | 1.09 | 3.32E-01 | 0.637 |
| OID21503 | SIAE | Oncology | SIAE | Q9HAT2 | 11 | 124633113 | 124695707 | 0.92 | 0.78 | 1.09 | 3.32E-01 | 0.637 |
| OID30603 | UROD | Inflammation_II | UROD | P06132 | 1 | 45010950 | 45015575 | 0.92 | 0.78 | 1.09 | 3.32E-01 | 0.637 |
| OID30522 | NFAT5 | Inflammation_II | NFAT5 | O94916 | 16 | 69565094 | 69704666 | 0.92 | 0.78 | 1.09 | 3.34E-01 | 0.639 |
| OID31253 | GABARAPL1 | Oncology_II | GABARAPL1 | Q9H0R8 | 12 | 10212458 | 10223128 | 0.92 | 0.77 | 1.09 | 3.34E-01 | 0.640 |
| OID31246 | TMOD4 | Oncology_II | TMOD4 | Q9NZQ9 | 1 | 151169986 | 151176284 | 1.08 | 0.92 | 1.27 | 3.35E-01 | 0.641 |
| OID20204 | SOST | Cardiometabolic | SOST | Q9BQB4 | 17 | 43753738 | 43758791 | 0.92 | 0.78 | 1.09 | 3.35E-01 | 0.642 |
| OID20978 | AFP | Neurology | AFP | P02771 | 4 | 73431138 | 73456174 | 0.92 | 0.77 | 1.09 | 3.36E-01 | 0.643 |
| OID31405 | PXDNL | Oncology_II | PXDNL | A1KZ92 | 8 | 51319577 | 51809445 | 0.91 | 0.76 | 1.10 | 3.37E-01 | 0.643 |
| OID20614 | PCDH1 | Inflammation | PCDH1 | Q08174 | 5 | 141853090 | 141879246 | 1.07 | 0.93 | 1.24 | 3.39E-01 | 0.646 |
| OID21472 | CD33 | Oncology | CD33 | P20138 | 19 | 51225064 | 51243860 | 1.08 | 0.92 | 1.28 | 3.39E-01 | 0.646 |
| OID21354 | DPP6 | Oncology | DPP6 | P42658 | 7 | 153887097 | 154894285 | 0.92 | 0.78 | 1.09 | 3.40E-01 | 0.648 |
| OID20432 | ARNT | Inflammation | ARNT | P27540 | 1 | 150809713 | 150876708 | 0.91 | 0.76 | 1.10 | 3.41E-01 | 0.649 |
| OID30927 | DOC2B | Neurology_II | DOC2B | Q14184 | 17 | 142789 | 181650 | 0.91 | 0.74 | 1.11 | 3.41E-01 | 0.649 |
| OID20119 | EPHX2 | Cardiometabolic | EPHX2 | P34913 | 8 | 27490781 | 27545564 | 0.91 | 0.76 | 1.10 | 3.41E-01 | 0.649 |
| OID30772 | CFI | Inflammation_II | CFI | P05156 | 4 | 109731008 | 109802150 | 0.91 | 0.76 | 1.10 | 3.42E-01 | 0.649 |
| OID30607 | PRR4 | Inflammation_II | PRR4 | Q16378 | 12 | 10845849 | 10849475 | 0.92 | 0.78 | 1.09 | 3.42E-01 | 0.649 |
| OID20156 | MNDA | Cardiometabolic | MNDA | P41218 | 1 | 158831351 | 158849506 | 0.92 | 0.77 | 1.09 | 3.42E-01 | 0.649 |
| OID30545 | NECTIN1 | Inflammation_II | NECTIN1 | Q15223 | 11 | 119623408 | 119729200 | 1.08 | 0.93 | 1.25 | 3.42E-01 | 0.650 |
| OID20647 | CD160 | Inflammation | CD160 | O95971 | 1 | 145719471 | 145739288 | 0.93 | 0.79 | 1.09 | 3.44E-01 | 0.652 |
| OID30956 | AIDA | Neurology_II | AIDA | Q96BJ3 | 1 | 222668013 | 222713210 | 0.92 | 0.77 | 1.09 | 3.45E-01 | 0.652 |
| OID30832 | HDAC9 | Neurology_II | HDAC9 | Q9UKV0 | 7 | 18086949 | 19002416 | 0.93 | 0.79 | 1.09 | 3.45E-01 | 0.652 |
| OID20253 | NOTCH3 | Cardiometabolic | NOTCH3 | Q9UM47 | 19 | 15159038 | 15200995 | 1.09 | 0.91 | 1.29 | 3.46E-01 | 0.652 |

| | | | | | | | | | | | | |
|---|---|---|---|---|---|---|---|---|---|---|---|---|
| OID20146 | PRKAR1A | Cardiometabolic | PRKAR1A | P10644 | 17 | 68511780 | 68551319 | 0.93 | 0.79 | 1.09 | 3.45E-01 | 0.652 |
| OID20174 | QDPR | Cardiometabolic | QDPR | P09417 | 4 | 17460261 | 17512206 | 0.92 | 0.78 | 1.09 | 3.45E-01 | 0.652 |
| OID20130 | S100A11 | Cardiometabolic | S100A11 | P31949 | 1 | 152032506 | 152047907 | 0.91 | 0.75 | 1.11 | 3.44E-01 | 0.652 |
| OID21048 | SCARF2 | Neurology | SCARF2 | Q96GP6 | 22 | 20424584 | 20437826 | 1.08 | 0.92 | 1.28 | 3.46E-01 | 0.652 |
| OID30798 | TJP3 | Neurology_II | TJP3 | O95049 | 19 | 3708362 | 3750813 | 1.07 | 0.93 | 1.24 | 3.46E-01 | 0.652 |
| OID20702 | TNFRSF13B | Inflammation | TNFRSF13B | O14836 | 17 | 16929816 | 16972118 | 0.92 | 0.78 | 1.09 | 3.46E-01 | 0.652 |
| OID20592 | TNFSF11 | Inflammation | TNFSF11 | O14788 | 13 | 42562736 | 42608013 | 0.93 | 0.79 | 1.09 | 3.44E-01 | 0.652 |
| OID20264 | DKK3 | Cardiometabolic | DKK3 | Q9UBP4 | 11 | 11956207 | 12009769 | 1.08 | 0.92 | 1.27 | 3.47E-01 | 0.652 |
| OID20519 | METAP1D | Inflammation | METAP1D | Q6UB28 | 2 | 171999943 | 172082430 | 0.92 | 0.78 | 1.09 | 3.47E-01 | 0.652 |
| OID20297 | SCARF1 | Cardiometabolic | SCARF1 | Q14162 | 17 | 1633858 | 1645744 | 0.92 | 0.78 | 1.09 | 3.47E-01 | 0.652 |
| OID30189 | VSIG10L | Cardiometabolic_II | VSIG10L | Q86VR7 | 19 | 51331536 | 51342139 | 0.92 | 0.78 | 1.09 | 3.47E-01 | 0.652 |
| OID30467 | WASL | Inflammation_II | WASL | O00401 | 7 | 123681943 | 123749003 | 0.92 | 0.78 | 1.09 | 3.47E-01 | 0.652 |
| OID31209 | PDIA5 | Oncology_II | PDIA5 | Q14554 | 3 | 123067025 | 123225227 | 0.92 | 0.77 | 1.10 | 3.48E-01 | 0.652 |
| OID31433 | PTPRH | Oncology_II | PTPRH | Q9HD43 | 19 | 55181247 | 55209506 | 1.08 | 0.92 | 1.28 | 3.48E-01 | 0.652 |
| OID30916 | CABP2 | Neurology_II | CABP2 | Q9NPB3 | 11 | 67518912 | 67524517 | 0.92 | 0.78 | 1.09 | 3.48E-01 | 0.653 |
| OID20073 | RCOR1 | Cardiometabolic | RCOR1 | Q9UKL0 | 14 | 102592649 | 102730561 | 0.91 | 0.75 | 1.11 | 3.49E-01 | 0.654 |
| OID21252 | EGFL7 | Oncology | EGFL7 | Q9UHF1 | 9 | 136658856 | 136672678 | 0.92 | 0.78 | 1.09 | 3.51E-01 | 0.655 |
| OID21504 | IL13RA1 | Oncology | IL13RA1 | P78552 | X | 118727133 | 118794535 | 1.08 | 0.92 | 1.28 | 3.50E-01 | 0.655 |
| OID21152 | LILRA2 | Neurology | LILRA2 | Q8N149 | 19 | 54572920 | 54590287 | 1.09 | 0.91 | 1.29 | 3.50E-01 | 0.655 |
| OID20830 | MAPT | Neurology | MAPT | P10636 | 17 | 45894527 | 46028334 | 1.08 | 0.92 | 1.28 | 3.51E-01 | 0.655 |
| OID21186 | MZT1 | Oncology | MZT1 | Q08AG7 | 13 | 72708367 | 72727629 | 0.92 | 0.77 | 1.10 | 3.51E-01 | 0.655 |
| OID20812 | SIGLEC15 | Neurology | SIGLEC15 | Q6ZMC9 | 18 | 45825675 | 45844094 | 0.92 | 0.78 | 1.09 | 3.51E-01 | 0.655 |
| OID31429 | TRIM58 | Oncology_II | TRIM58 | Q8NG06 | 1 | 247857187 | 247880138 | 0.93 | 0.79 | 1.09 | 3.51E-01 | 0.655 |
| OID20456 | IL17F | Inflammation | IL17F | Q96PD4 | 6 | 52236681 | 52244500 | 1.07 | 0.93 | 1.24 | 3.52E-01 | 0.656 |
| OID20793 | ODAM | Neurology | ODAM | A1E959 | 4 | 70195725 | 70204576 | 0.92 | 0.78 | 1.09 | 3.53E-01 | 0.657 |
| OID31432 | KIR2DS4 | Oncology_II | KIR2DS4 | P43632 | 19 | 54832676 | 54848569 | 0.92 | 0.78 | 1.09 | 3.53E-01 | 0.657 |
| OID20818 | SSB | Neurology | SSB | P05455 | 2 | 169791933 | 169812064 | 0.91 | 0.76 | 1.11 | 3.55E-01 | 0.660 |
| OID20974 | ADGRB3 | Neurology | ADGRB3 | O60242 | 6 | 68635282 | 69390571 | 1.08 | 0.92 | 1.27 | 3.56E-01 | 0.661 |
| OID21342 | EPHA2 | Oncology | EPHA2 | P29317 | 1 | 16124337 | 16156069 | 1.08 | 0.92 | 1.25 | 3.56E-01 | 0.661 |
| OID20094 | SLITRK6 | Cardiometabolic | SLITRK6 | Q9H5Y7 | 13 | 85792790 | 85806683 | 0.93 | 0.79 | 1.09 | 3.56E-01 | 0.661 |
| OID30138 | RIPK4 | Cardiometabolic_II | RIPK4 | P57078 | 21 | 41739369 | 41767089 | 0.92 | 0.78 | 1.09 | 3.56E-01 | 0.661 |
| OID30365 | ISM2 | Cardiometabolic_II | ISM2 | Q6H9L7 | 14 | 77474394 | 77498816 | 0.92 | 0.78 | 1.09 | 3.58E-01 | 0.663 |
| OID31045 | PCDH12 | Neurology_II | PCDH12 | Q9NPG4 | 5 | 141943581 | 141969741 | 0.93 | 0.79 | 1.09 | 3.58E-01 | 0.663 |
| OID20133 | PDGFRA | Cardiometabolic | PDGFRA | P16234 | 4 | 54229280 | 54298245 | 1.08 | 0.92 | 1.27 | 3.58E-01 | 0.663 |
| OID30876 | SPTBN2 | Neurology_II | SPTBN2 | O15020 | 11 | 66682497 | 66744670 | 0.92 | 0.78 | 1.09 | 3.58E-01 | 0.663 |
| OID21393 | ITGB7 | Oncology | ITGB7 | P26010 | 12 | 53191323 | 53207282 | 0.93 | 0.79 | 1.09 | 3.59E-01 | 0.664 |
| OID31467 | PHLDB1 | Oncology_II | PHLDB1 | Q86UU1 | 11 | 118606440 | 118658031 | 0.92 | 0.77 | 1.10 | 3.59E-01 | 0.664 |
| OID20363 | AXL | Cardiometabolic | AXL | P30530 | 19 | 41219223 | 41261766 | 1.08 | 0.92 | 1.26 | 3.60E-01 | 0.664 |
| OID21195 | CA11 | Oncology | CA11 | O75493 | 19 | 48637946 | 48646187 | 0.91 | 0.74 | 1.11 | 3.60E-01 | 0.664 |
| OID31075 | TAX1BP1 | Neurology_II | TAX1BP1 | Q86VP1 | 7 | 27739331 | 27844564 | 0.92 | 0.78 | 1.09 | 3.60E-01 | 0.664 |
| OID30981 | SOX2 | Neurology_II | SOX2 | P48431 | 3 | 181711925 | 181714436 | 0.93 | 0.79 | 1.09 | 3.61E-01 | 0.666 |
| OID30428 | TLR4 | Inflammation_II | TLR4 | O00206 | 9 | 117704175 | 117724735 | 0.92 | 0.78 | 1.10 | 3.62E-01 | 0.667 |
| OID20449 | IL22RA1 | Inflammation | IL22RA1 | Q8N6P7 | 1 | 24119771 | 24143140 | 0.92 | 0.77 | 1.10 | 3.62E-01 | 0.667 |
| OID20338 | PGLYRP1 | Cardiometabolic | PGLYRP1 | O75594 | 19 | 46019153 | 46023053 | 0.92 | 0.77 | 1.10 | 3.63E-01 | 0.667 |
| OID21369 | RBP5 | Oncology | RBP5 | P82980 | 12 | 7115736 | 7128889 | 0.92 | 0.77 | 1.10 | 3.63E-01 | 0.667 |
| OID21249 | CRACR2A | Oncology | CRACR2A | Q9BSW2 | 12 | 3606633 | 3764819 | 0.93 | 0.78 | 1.09 | 3.64E-01 | 0.668 |
| OID30780 | GC | Inflammation_II | GC | P02774 | 4 | 71741696 | 71804041 | 0.93 | 0.78 | 1.09 | 3.63E-01 | 0.668 |
| OID30385 | PPIF | Cardiometabolic_II | PPIF | P30405 | 10 | 79347469 | 79355334 | 0.92 | 0.77 | 1.10 | 3.64E-01 | 0.668 |
| OID20739 | SIRPB1 | Inflammation | SIRPB1 | O00241 | 20 | 1561385 | 1620061 | 1.08 | 0.92 | 1.26 | 3.64E-01 | 0.668 |
| OID20852 | SLC39A14 | Neurology | SLC39A14 | Q15043 | 8 | 22367278 | 22434129 | 1.08 | 0.91 | 1.28 | 3.65E-01 | 0.668 |
| OID20677 | EPHA1 | Inflammation | EPHA1 | P21709 | 7 | 143390289 | 143408856 | 0.92 | 0.77 | 1.10 | 3.65E-01 | 0.668 |
| OID20532 | HLA-E | Inflammation | HLA-E | P13747 | 6 | 30489509 | 30494194 | 1.08 | 0.92 | 1.26 | 3.65E-01 | 0.669 |
| OID30072 | CSF2 | Cardiometabolic_II | CSF2 | P04141 | 5 | 132073789 | 132076170 | 1.08 | 0.92 | 1.27 | 3.66E-01 | 0.669 |
| OID20463 | FXYD5 | Inflammation | FXYD5 | Q96DB9 | 19 | 35154730 | 35169881 | 0.93 | 0.78 | 1.10 | 3.66E-01 | 0.669 |
| OID30849 | DEFB104A_DEF | Neurology_II | 04B;DEFB104A | Q8WTQ1;Q8WTQ1 | 8;8 | 7470308;7836436 | 7475082;7841242 | 0.91 | 0.75 | 1.11 | 3.66E-01 | 0.670 |
| OID20756 | AGER | Inflammation | AGER | Q15109 | 6 | 32180968 | 32184322 | 0.93 | 0.78 | 1.09 | 3.68E-01 | 0.670 |
| OID20975 | HNMT | Neurology | HNMT | P50135 | 2 | 137964020 | 138016364 | 0.92 | 0.78 | 1.10 | 3.67E-01 | 0.670 |
| OID30598 | MFAP4 | Inflammation_II | MFAP4 | P55083 | 17 | 19383442 | 19387190 | 1.08 | 0.91 | 1.29 | 3.68E-01 | 0.670 |
| OID21121 | MYOC | Neurology | MYOC | Q99972 | 1 | 171635417 | 171652688 | 0.92 | 0.78 | 1.10 | 3.67E-01 | 0.670 |
| OID21382 | VAT1 | Oncology | VAT1 | Q99536 | 17 | 43014607 | 43025123 | 1.08 | 0.92 | 1.27 | 3.67E-01 | 0.670 |
| OID20178 | NADK | Cardiometabolic | NADK | O95544 | 1 | 1751232 | 1780457 | 0.92 | 0.77 | 1.10 | 3.68E-01 | 0.670 |
| OID30595 | MENT | Inflammation_II | C1orf56 | Q9BUN1 | 1 | 151047751 | 151051986 | 0.92 | 0.77 | 1.10 | 3.69E-01 | 0.671 |
| OID21038 | CHGB | Neurology | CHGB | P05060 | 20 | 5911510 | 5925353 | 0.93 | 0.79 | 1.09 | 3.69E-01 | 0.671 |
| OID20599 | CD70 | Inflammation | CD70 | P32970 | 19 | 6583183 | 6604103 | 0.93 | 0.79 | 1.09 | 3.69E-01 | 0.671 |
| OID30620 | C1QTNF9 | Inflammation_II | C1QTNF9 | P0C862 | 13 | 24307166 | 24322535 | 0.93 | 0.79 | 1.09 | 3.70E-01 | 0.672 |
| OID30880 | ERC2 | Neurology_II | ERC2 | O15083 | 3 | 55508311 | 56469095 | 0.92 | 0.77 | 1.10 | 3.70E-01 | 0.672 |
| OID20972 | BAG3 | Neurology | BAG3 | O95817 | 10 | 119651380 | 119677819 | 0.93 | 0.79 | 1.09 | 3.73E-01 | 0.674 |
| OID20334 | CCL16 | Cardiometabolic | CCL16 | O15467 | 17 | 35976493 | 35981497 | 1.09 | 0.90 | 1.31 | 3.72E-01 | 0.674 |
| OID30803 | GABRA4 | Neurology_II | GABRA4 | P48169 | 4 | 46918900 | 46993581 | 0.93 | 0.78 | 1.10 | 3.73E-01 | 0.674 |
| OID20352 | ITIH3 | Cardiometabolic | ITIH3 | Q06033 | 3 | 52794768 | 52809009 | 1.08 | 0.91 | 1.29 | 3.72E-01 | 0.674 |
| OID21103 | MMP9 | Neurology | MMP9 | P14780 | 20 | 46008908 | 46016561 | 0.93 | 0.79 | 1.09 | 3.72E-01 | 0.674 |
| OID31161 | PCNA | Oncology_II | PCNA | P12004 | 20 | 5114953 | 5126626 | 0.92 | 0.77 | 1.10 | 3.72E-01 | 0.674 |
| OID30567 | RALB | Inflammation_II | RALB | P11234 | 2 | 120240064 | 120294710 | 0.93 | 0.78 | 1.10 | 3.73E-01 | 0.674 |
| OID20820 | SIRT5 | Neurology | SIRT5 | Q9NXA8 | 6 | 13574227 | 13615158 | 0.92 | 0.77 | 1.10 | 3.73E-01 | 0.674 |
| OID30816 | KIF20B | Neurology_II | KIF20B | Q96Q89 | 10 | 89701590 | 89774939 | 1.06 | 0.93 | 1.22 | 3.74E-01 | 0.674 |
| OID30540 | NPHS1 | Inflammation_II | NPHS1 | O60500 | 19 | 35825372 | 35869287 | 0.93 | 0.79 | 1.09 | 3.74E-01 | 0.674 |
| OID20915 | CSF2RA | Neurology | CSF2RA | P15509 | X | 1268800 | 1310381 | 0.93 | 0.79 | 1.09 | 3.74E-01 | 0.675 |
| OID20528 | ITGA6 | Inflammation | ITGA6 | P23229 | 2 | 172427354 | 172506459 | 0.93 | 0.78 | 1.10 | 3.75E-01 | 0.676 |
| OID20600 | TGFA | Inflammation | TGFA | P01135 | 2 | 70447284 | 70554193 | 1.07 | 0.92 | 1.26 | 3.75E-01 | 0.676 |
| OID20782 | FSTL3 | Inflammation | FSTL3 | O95633 | 19 | 676392 | 683392 | 1.08 | 0.91 | 1.27 | 3.77E-01 | 0.678 |
| OID21089 | IL17RA | Neurology | IL17RA | Q96F46 | 22 | 17084954 | 17115694 | 1.08 | 0.91 | 1.27 | 3.77E-01 | 0.678 |
| OID30415 | ASAH1 | Cardiometabolic_II | ASAH1 | Q13510 | 8 | 18055992 | 18084998 | 0.93 | 0.79 | 1.10 | 3.78E-01 | 0.679 |
| OID30731 | C1S | Inflammation_II | C1S | P09871 | 12 | 6988259 | 7071032 | 0.92 | 0.77 | 1.10 | 3.78E-01 | 0.679 |
| OID20120 | TP53INP1 | Cardiometabolic | TP53INP1 | Q96A56 | 8 | 94925972 | 94949378 | 1.07 | 0.92 | 1.23 | 3.78E-01 | 0.679 |
| OID21128 | ALDH1A1 | Neurology | ALDH1A1 | P00352 | 9 | 72900671 | 73080442 | 0.92 | 0.77 | 1.10 | 3.80E-01 | 0.680 |
| OID30784 | CFP | Inflammation_II | CFP | P27918 | X | 47623172 | 47630305 | 0.92 | 0.76 | 1.11 | 3.80E-01 | 0.680 |
| OID20069 | CNPY2 | Cardiometabolic | CNPY2 | Q9Y2B0 | 12 | 56309842 | 56316119 | 1.07 | 0.92 | 1.23 | 3.81E-01 | 0.680 |
| OID30993 | DSCAM | Neurology_II | DSCAM | O60469 | 21 | 40010999 | 40847158 | 1.08 | 0.91 | 1.27 | 3.80E-01 | 0.680 |
| OID30535 | FGF16 | Inflammation_II | FGF16 | O43320 | X | 77447389 | 77457278 | 0.92 | 0.77 | 1.10 | 3.80E-01 | 0.680 |
| OID30521 | NEDD4L | Inflammation_II | NEDD4L | Q96PU5 | 18 | 58044226 | 58401540 | 0.93 | 0.79 | 1.09 | 3.81E-01 | 0.680 |
| OID30724 | SERPINA7 | Inflammation_II | SERPINA7 | P05543 | X | 106032435 | 106038727 | 0.93 | 0.78 | 1.10 | 3.80E-01 | 0.680 |
| OID30195 | TOR1AIP1 | Cardiometabolic_II | TOR1AIP1 | Q5JTV8 | 1 | 179882042 | 179920077 | 0.92 | 0.75 | 1.11 | 3.81E-01 | 0.680 |
| OID21191 | LTBP3 | Oncology | LTBP3 | Q9NS15 | 11 | 65538559 | 65558930 | 0.92 | 0.77 | 1.11 | 3.81E-01 | 0.680 |
| OID30065 | ATP1B3 | Cardiometabolic_II | ATP1B3 | P54709 | 3 | 141876124 | 141926549 | 0.92 | 0.76 | 1.11 | 3.82E-01 | 0.681 |
| OID30281 | B3GNT7 | Cardiometabolic_II | B3GNT7 | Q8NFL0 | 2 | 231395710 | 231401164 | 0.93 | 0.78 | 1.10 | 3.82E-01 | 0.681 |
| OID20505 | BACH1 | Inflammation | BACH1 | O14867 | 21 | 29194071 | 29630751 | 0.93 | 0.79 | 1.09 | 3.82E-01 | 0.681 |
| OID21184 | PPM1A | Oncology | PPM1A | P35813 | 14 | 60245752 | 60299087 | 0.93 | 0.78 | 1.10 | 3.83E-01 | 0.681 |
| OID21494 | NELL1 | Oncology | NELL1 | Q92832 | 11 | 20669551 | 21575686 | 0.93 | 0.80 | 1.09 | 3.83E-01 | 0.681 |
| OID21435 | CPVL | Oncology | CPVL | Q9H3G5 | 7 | 28995235 | 29195451 | 1.08 | 0.91 | 1.28 | 3.83E-01 | 0.682 |
| OID30525 | BNIP3L | Inflammation_II | BNIP3L | O60238 | 8 | 26383054 | 26505636 | 0.92 | 0.77 | 1.10 | 3.84E-01 | 0.682 |
| OID20409 | CA1 | Cardiometabolic | CA1 | P00915 | 8 | 85327608 | 85379014 | 0.92 | 0.78 | 1.10 | 3.84E-01 | 0.682 |
| OID20243 | CDH5 | Cardiometabolic | CDH5 | P33151 | 16 | 66366622 | 66404784 | 1.07 | 0.92 | 1.26 | 3.85E-01 | 0.682 |
| OID31464 | COL24A1 | Oncology_II | COL24A1 | Q17RW2 | 1 | 85729233 | 86156943 | 0.93 | 0.80 | 1.09 | 3.85E-01 | 0.682 |

| | | | | | | | | | | | | |
|---|---|---|---|---|---|---|---|---|---|---|---|---|
| OID31108 | FGFBP3 | Neurology_II | FGFBP3 | Q8TAT2 | 10 | 91906584 | 91909486 | 0.93 | 0.80 | 1.09 | 3.84E-01 | 0.682 |
| OID20128 | LEPR | Cardiometabolic | LEPR | P48357 | 1 | 65420652 | 65641559 | 0.92 | 0.77 | 1.10 | 3.86E-01 | 0.682 |
| OID20858 | MRPL46 | Neurology | MRPL46 | Q9H2W6 | 15 | 88459477 | 88467390 | 0.93 | 0.78 | 1.10 | 3.85E-01 | 0.682 |
| OID30997 | PPP1R14D | Neurology_II | PPP1R14D | Q9NXH3 | 15 | 40815451 | 40828709 | 0.93 | 0.80 | 1.09 | 3.84E-01 | 0.682 |
| OID21453 | SORD | Oncology | SORD | Q00796 | 15 | 45023147 | 45077185 | 0.93 | 0.78 | 1.10 | 3.85E-01 | 0.682 |
| OID20426 | IL4 | Inflammation | IL4 | P05112 | 5 | 132673986 | 132682678 | 1.08 | 0.91 | 1.28 | 3.86E-01 | 0.683 |
| OID20215 | DDC | Cardiometabolic | DDC | P20711 | 7 | 50458436 | 50565405 | 0.93 | 0.79 | 1.09 | 3.88E-01 | 0.684 |
| OID20935 | EFNA4 | Neurology | EFNA4 | P52798 | 1 | 155063737 | 155069553 | 1.07 | 0.92 | 1.25 | 3.88E-01 | 0.684 |
| OID20666 | IL12B | Inflammation | IL12B | P29460 | 5 | 159314780 | 159330487 | 1.07 | 0.91 | 1.26 | 3.87E-01 | 0.684 |
| OID20364 | PRSS2 | Cardiometabolic | PRSS2 | P07478 | 7 | 142760398 | 142774564 | 0.93 | 0.79 | 1.09 | 3.88E-01 | 0.684 |
| OID31274 | TPPP2 | Oncology_II | TPPP2 | P59282 | 14 | 21024109 | 21036276 | 0.93 | 0.80 | 1.09 | 3.87E-01 | 0.684 |
| OID30890 | CLIC5 | Neurology_II | CLIC5 | Q9NZA1 | 6 | 45880827 | 46080348 | 0.93 | 0.78 | 1.10 | 3.89E-01 | 0.685 |
| OID20954 | MASP1 | Neurology | MASP1 | P48740 | 3 | 187217282 | 187291980 | 0.93 | 0.79 | 1.09 | 3.89E-01 | 0.685 |
| OID20376 | ITGB1 | Cardiometabolic | ITGB1 | P05556 | 10 | 32887273 | 33005792 | 0.93 | 0.79 | 1.10 | 3.91E-01 | 0.687 |
| OID30515 | C1QL2 | Inflammation_II | C1QL2 | Q7Z5L3 | 2 | 119156243 | 119158751 | 0.93 | 0.78 | 1.10 | 3.91E-01 | 0.687 |
| OID30401 | STAB2 | Cardiometabolic_II | STAB2 | Q8WWQ8 | 12 | 103587273 | 103766719 | 0.93 | 0.78 | 1.10 | 3.91E-01 | 0.687 |
| OID21332 | SCAMP3 | Oncology | SCAMP3 | O14828 | 1 | 155255979 | 155262430 | 0.93 | 0.79 | 1.10 | 3.92E-01 | 0.688 |
| OID30169 | KRT6C | Cardiometabolic_II | KRT6C | P48668 | 12 | 52468516 | 52473805 | 1.07 | 0.92 | 1.25 | 3.93E-01 | 0.690 |
| OID31288 | SEPTIN7 | Oncology_II | SEPTIN7 | Q16181 | 7 | 35800932 | 35907110 | 0.92 | 0.76 | 1.11 | 3.93E-01 | 0.690 |
| OID30786 | SERPINA3 | Inflammation_II | SERPINA3 | P01011 | 14 | 94612384 | 94624055 | 1.08 | 0.91 | 1.28 | 3.95E-01 | 0.692 |
| OID21246 | ATG4A | Oncology | ATG4A | Q8WYN0 | X | 108091668 | 108154671 | 0.93 | 0.78 | 1.10 | 3.95E-01 | 0.692 |
| OID21486 | NBL1 | Oncology | NBL1 | P41271 | 1 | 19596979 | 19658456 | 1.07 | 0.91 | 1.25 | 3.97E-01 | 0.695 |
| OID20684 | TIMP3 | Inflammation | TIMP3 | P35625 | 22 | 32801705 | 32863041 | 0.93 | 0.79 | 1.10 | 3.97E-01 | 0.695 |
| OID31515 | PALM | Oncology_II | PALM | O75781 | 19 | 708935 | 748329 | 0.93 | 0.78 | 1.10 | 3.97E-01 | 0.695 |
| OID21483 | CD1C | Oncology | CD1C | P29017 | 1 | 158289923 | 158294774 | 0.93 | 0.80 | 1.09 | 3.98E-01 | 0.695 |
| OID30576 | CD300A | Inflammation_II | CD300A | Q9UGN4 | 17 | 74466399 | 74484794 | 0.93 | 0.79 | 1.10 | 3.98E-01 | 0.695 |
| OID30449 | GAD2 | Inflammation_II | GAD2 | Q05329 | 10 | 26216665 | 26304558 | 0.93 | 0.78 | 1.10 | 3.98E-01 | 0.695 |
| OID30726 | SAA4 | Inflammation_II | SAA4 | P35542 | 11 | 18231355 | 18236802 | 0.89 | 0.67 | 1.17 | 3.98E-01 | 0.695 |
| OID21159 | GRN | Neurology | GRN | P28799 | 17 | 44345246 | 44353106 | 0.93 | 0.78 | 1.10 | 3.99E-01 | 0.695 |
| OID20408 | PTGDS | Cardiometabolic | PTGDS | P41222 | 9 | 136975092 | 136981742 | 1.07 | 0.92 | 1.25 | 3.99E-01 | 0.695 |
| OID30902 | CRTAP | Neurology_II | CRTAP | O75718 | 3 | 33114014 | 33147773 | 0.93 | 0.78 | 1.10 | 3.99E-01 | 0.695 |
| OID30944 | DDX53 | Neurology_II | DDX53 | Q86TM3 | X | 22999960 | 23003589 | 1.07 | 0.92 | 1.25 | 4.00E-01 | 0.695 |
| OID21251 | LYPD8 | Oncology | LYPD8 | Q6UX82 | 1 | 248739415 | 248755759 | 1.07 | 0.92 | 1.23 | 4.00E-01 | 0.695 |
| OID20087 | MMP7 | Cardiometabolic | MMP7 | P09237 | 11 | 102520508 | 102530750 | 1.08 | 0.91 | 1.27 | 4.00E-01 | 0.695 |
| OID21211 | RUVBL1 | Oncology | RUVBL1 | Q9Y265 | 3 | 128064778 | 128153914 | 1.06 | 0.92 | 1.23 | 4.00E-01 | 0.695 |
| OID31500 | CD86 | Oncology_II | CD86 | P42081 | 3 | 122055362 | 122121139 | 0.93 | 0.78 | 1.10 | 4.01E-01 | 0.695 |
| OID20429 | LILRB4 | Inflammation | LILRB4 | Q8NHJ6 | 19 | 54643889 | 54670359 | 1.07 | 0.91 | 1.27 | 4.01E-01 | 0.695 |
| OID30805 | CNGB3 | Neurology_II | CNGB3 | Q9NQW8 | 8 | 86553977 | 86743675 | 0.93 | 0.78 | 1.11 | 4.02E-01 | 0.696 |
| OID30715 | CRISP3 | Inflammation_II | CRISP3 | P54108 | 6 | 49727376 | 49744437 | 0.93 | 0.78 | 1.10 | 4.02E-01 | 0.696 |
| OID30152 | ENOX2 | Cardiometabolic_II | ENOX2 | Q16206 | X | 130622325 | 130903234 | 0.92 | 0.77 | 1.11 | 4.03E-01 | 0.698 |
| OID30443 | FGF6 | Inflammation_II | FGF6 | P10767 | 12 | 4428155 | 4445815 | 0.93 | 0.77 | 1.11 | 4.04E-01 | 0.698 |
| OID30364 | KLK3 | Cardiometabolic_II | KLK3 | P07288 | 19 | 50854915 | 50860764 | 0.89 | 0.68 | 1.17 | 4.03E-01 | 0.698 |
| OID21084 | WARS | Neurology | WARS1 | P23381 | 14 | 100333790 | 100376805 | 0.93 | 0.79 | 1.10 | 4.04E-01 | 0.698 |
| OID20996 | ASAH2 | Neurology | ASAH2 | Q9NR71 | 10 | 50182778 | 50279720 | 0.93 | 0.79 | 1.10 | 4.04E-01 | 0.698 |
| OID31056 | AZI2 | Neurology_II | AZI2 | Q9H6S1 | 3 | 28315003 | 28349050 | 0.93 | 0.78 | 1.10 | 4.04E-01 | 0.698 |
| OID31496 | EGFLAM | Oncology_II | EGFLAM | Q63HQ2 | 5 | 38258409 | 38465480 | 0.93 | 0.79 | 1.10 | 4.05E-01 | 0.699 |
| OID20839 | PBLD | Neurology | PBLD | P30039 | 10 | 68282660 | 68333049 | 0.93 | 0.78 | 1.10 | 4.06E-01 | 0.699 |
| OID20190 | SIGLEC7 | Cardiometabolic | SIGLEC7 | Q9Y286 | 19 | 51142299 | 51153526 | 0.93 | 0.78 | 1.10 | 4.06E-01 | 0.700 |
| OID31229 | IL3 | Oncology_II | IL3 | P08700 | 5 | 132060655 | 132063204 | 1.07 | 0.91 | 1.26 | 4.07E-01 | 0.700 |
| OID30538 | MTDH | Inflammation_II | MTDH | Q86UE4 | 8 | 97644184 | 97730260 | 0.93 | 0.79 | 1.10 | 4.06E-01 | 0.700 |
| OID30905 | SOX9 | Neurology_II | SOX9 | P48436 | 17 | 72121020 | 72126416 | 0.94 | 0.80 | 1.09 | 4.06E-01 | 0.700 |
| OID20400 | CST3 | Cardiometabolic | CST3 | P01034 | 20 | 23626706 | 23638473 | 0.93 | 0.79 | 1.10 | 4.08E-01 | 0.701 |
| OID21454 | LEFTY2 | Oncology | LEFTY2 | O00292 | 1 | 225936598 | 225941383 | 0.93 | 0.78 | 1.11 | 4.08E-01 | 0.701 |
| OID30110 | MTR | Cardiometabolic_II | MTR | Q99707 | 1 | 236795260 | 236921278 | 0.93 | 0.79 | 1.10 | 4.08E-01 | 0.701 |
| OID21316 | CD38 | Oncology | CD38 | P28907 | 4 | 15778275 | 15853232 | 0.92 | 0.76 | 1.12 | 4.09E-01 | 0.701 |
| OID31019 | SNAPIN | Neurology_II | SNAPIN | O95295 | 1 | 153658703 | 153661852 | 0.93 | 0.79 | 1.10 | 4.09E-01 | 0.701 |
| OID20590 | CLEC4G | Inflammation | CLEC4G | Q6UXB4 | 19 | 7728957 | 7733906 | 0.93 | 0.79 | 1.10 | 4.10E-01 | 0.702 |
| OID30732 | CLU | Inflammation_II | CLU | P10909 | 8 | 27596917 | 27614700 | 0.90 | 0.70 | 1.16 | 4.11E-01 | 0.702 |
| OID31252 | EPN1 | Oncology_II | EPN1 | Q9Y6I3 | 19 | 55675226 | 55709858 | 0.93 | 0.77 | 1.11 | 4.11E-01 | 0.702 |
| OID31127 | FNDC1 | Neurology_II | FNDC1 | Q4ZHG4 | 6 | 159169400 | 159272108 | 0.94 | 0.80 | 1.10 | 4.10E-01 | 0.702 |
| OID21013 | FUT3_FUT5 | Neurology | FUT3;FUT5 | P21217;Q11128 | 19;19 | 5842888;5865826 | 5851471;5870540 | 1.07 | 0.91 | 1.27 | 4.09E-01 | 0.702 |
| OID20085 | IRAG2 | Cardiometabolic | IRAG2 | Q12912 | 12 | 25004342 | 25108335 | 0.93 | 0.79 | 1.10 | 4.11E-01 | 0.702 |
| OID21498 | NT5E | Oncology | NT5E | P21589 | 6 | 85449584 | 85495791 | 0.93 | 0.79 | 1.10 | 4.11E-01 | 0.702 |
| OID20421 | PAPPA | Inflammation | PAPPA | Q13219 | 9 | 116153791 | 116402321 | 0.93 | 0.79 | 1.10 | 4.11E-01 | 0.702 |
| OID30090 | RBM19 | Cardiometabolic_II | RBM19 | Q9Y4C8 | 12 | 113816738 | 113966325 | 1.08 | 0.90 | 1.29 | 4.10E-01 | 0.702 |
| OID30081 | UGDH | Cardiometabolic_II | UGDH | O60701 | 4 | 39498755 | 39528311 | 0.93 | 0.79 | 1.10 | 4.11E-01 | 0.702 |
| OID31493 | PSCA | Oncology_II | PSCA | O43653 | 8 | 142670308 | 142682725 | 1.07 | 0.91 | 1.25 | 4.12E-01 | 0.702 |
| OID20278 | DPT | Cardiometabolic | DPT | Q07507 | 1 | 168695468 | 168729206 | 1.08 | 0.89 | 1.32 | 4.12E-01 | 0.702 |
| OID31138 | MAP2 | Neurology_II | MAP2 | P11137 | 2 | 209424047 | 209734147 | 0.93 | 0.78 | 1.11 | 4.13E-01 | 0.704 |
| OID31226 | PDXDC1 | Oncology_II | PDXDC1 | Q6P996 | 16 | 14974591 | 15139339 | 0.91 | 0.74 | 1.13 | 4.13E-01 | 0.704 |
| OID20976 | CX3CL1 | Neurology | CX3CL1 | P78423 | 16 | 57372477 | 57385044 | 1.07 | 0.91 | 1.25 | 4.14E-01 | 0.705 |
| OID30153 | SPINK8 | Cardiometabolic_II | SPINK8 | P0C7L1 | 3 | 48306842 | 48333661 | 0.93 | 0.78 | 1.11 | 4.15E-01 | 0.705 |
| OID21111 | FCRL5 | Neurology | FCRL5 | Q96RD9 | 1 | 157513377 | 157552515 | 0.93 | 0.78 | 1.11 | 4.16E-01 | 0.705 |
| OID20195 | GLRX | Cardiometabolic | GLRX | P35754 | 5 | 95751319 | 95822726 | 0.93 | 0.78 | 1.11 | 4.16E-01 | 0.705 |
| OID30105 | NEB | Cardiometabolic_II | NEB | P20929 | 2 | 151485336 | 151734487 | 0.93 | 0.79 | 1.10 | 4.15E-01 | 0.705 |
| OID30858 | DEFB118 | Neurology_II | DEFB118 | Q96PH6 | 20 | 31368601 | 31373923 | 1.06 | 0.92 | 1.23 | 4.16E-01 | 0.706 |
| OID30307 | HBZ | Cardiometabolic_II | HBZ | P02008 | 16 | 142728 | 154503 | 1.07 | 0.91 | 1.25 | 4.16E-01 | 0.706 |
| OID21241 | CEP85 | Oncology | CEP85 | Q6P2H3 | 1 | 26234200 | 26278808 | 0.93 | 0.79 | 1.10 | 4.17E-01 | 0.707 |
| OID31279 | PPIE | Oncology_II | PPIE | Q9UNP9 | 1 | 39692182 | 39763914 | 0.92 | 0.76 | 1.12 | 4.18E-01 | 0.707 |
| OID20454 | RGS8 | Inflammation | RGS8 | P57771 | 1 | 182641816 | 182684576 | 0.93 | 0.78 | 1.11 | 4.18E-01 | 0.708 |
| OID20908 | PTPN1 | Neurology | PTPN1 | P18031 | 20 | 50510321 | 50585241 | 0.93 | 0.79 | 1.10 | 4.19E-01 | 0.708 |
| OID31332 | CDH22 | Oncology_II | CDH22 | Q9UJ99 | 20 | 46173739 | 46308498 | 0.93 | 0.78 | 1.11 | 4.20E-01 | 0.709 |
| OID30433 | RICTOR | Inflammation_II | RICTOR | Q6R327 | 5 | 38937920 | 39074399 | 0.92 | 0.76 | 1.12 | 4.19E-01 | 0.709 |
| OID30156 | EIF5 | Cardiometabolic_II | EIF5 | P55010 | 14 | 103333544 | 103345025 | 1.06 | 0.92 | 1.22 | 4.20E-01 | 0.709 |
| OID30290 | GGACT | Cardiometabolic_II | GGACT | Q9BVM4 | 13 | 100530164 | 100589528 | 0.93 | 0.79 | 1.10 | 4.21E-01 | 0.709 |
| OID20767 | MATN2 | Inflammation | MATN2 | O00339 | 8 | 97868840 | 98036724 | 0.93 | 0.79 | 1.10 | 4.21E-01 | 0.709 |
| OID20083 | PTN | Cardiometabolic | PTN | P21246 | 7 | 137227341 | 137343774 | 0.93 | 0.79 | 1.11 | 4.21E-01 | 0.709 |
| OID21007 | ROBO2 | Neurology | ROBO2 | Q9HCK4 | 3 | 75906695 | 77649964 | 0.93 | 0.79 | 1.10 | 4.20E-01 | 0.709 |
| OID31194 | GIMAP8 | Oncology_II | GIMAP8 | Q8ND71 | 7 | 150450630 | 150479393 | 0.93 | 0.77 | 1.12 | 4.22E-01 | 0.709 |
| OID21493 | TGFBR2 | Oncology | TGFBR2 | P37173 | 3 | 30606601 | 30694142 | 1.07 | 0.91 | 1.25 | 4.22E-01 | 0.709 |
| OID21287 | AIFM1 | Oncology | AIFM1 | O95831 | X | 130124666 | 130165879 | 0.93 | 0.79 | 1.10 | 4.23E-01 | 0.710 |
| OID20490 | FGF5 | Inflammation | FGF5 | P12034 | 4 | 80266639 | 80336680 | 1.08 | 0.90 | 1.29 | 4.23E-01 | 0.710 |
| OID30520 | NDUFA5 | Inflammation_II | NDUFA5 | Q16718 | 7 | 123536997 | 123557904 | 0.94 | 0.80 | 1.10 | 4.23E-01 | 0.711 |
| OID30790 | CFH | Inflammation_II | CFH | P08603 | 1 | 196651754 | 196752476 | 0.92 | 0.75 | 1.13 | 4.24E-01 | 0.711 |
| OID30945 | MDM1 | Neurology_II | MDM1 | Q8TC05 | 12 | 68272443 | 68332381 | 0.94 | 0.80 | 1.10 | 4.25E-01 | 0.712 |
| OID30852 | TMPRSS11B | Neurology_II | TMPRSS11B | Q86T26 | 4 | 68226653 | 68245694 | 1.06 | 0.91 | 1.24 | 4.25E-01 | 0.712 |
| OID21139 | ITGA5 | Neurology | ITGA5 | P08648 | 12 | 54395261 | 54419266 | 0.93 | 0.78 | 1.11 | 4.26E-01 | 0.713 |
| OID20638 | LAMP3 | Inflammation | LAMP3 | Q9UQV4 | 3 | 183122215 | 183163839 | 1.08 | 0.90 | 1.30 | 4.25E-01 | 0.713 |
| OID21042 | CXCL11 | Neurology | CXCL11 | O14625 | 4 | 76033682 | 76041415 | 0.93 | 0.78 | 1.11 | 4.27E-01 | 0.713 |
| OID30374 | CYTL1 | Cardiometabolic_II | CYTL1 | Q9NRR1 | 4 | 5014586 | 5019458 | 1.07 | 0.91 | 1.26 | 4.27E-01 | 0.713 |
| OID20132 | PCDH17 | Cardiometabolic | PCDH17 | O14917 | 13 | 57631744 | 57729311 | 1.07 | 0.91 | 1.25 | 4.27E-01 | 0.713 |
| OID20760 | ATP5IF1 | Inflammation | ATP5IF1 | Q9UII2 | 1 | 28236109 | 28246906 | 0.94 | 0.80 | 1.10 | 4.28E-01 | 0.715 |
| OID30424 | TPSG1 | Inflammation_II | TPSG1 | Q9NRR2 | 16 | 1221651 | 1225268 | 0.93 | 0.79 | 1.11 | 4.28E-01 | 0.715 |

| | | | | | | | | | | | | |
|---|---|---|---|---|---|---|---|---|---|---|---|---|
| OID20757 | IL1R2 | Inflammation | IL1R2 | P27930 | 2 | 101991960 | 102028544 | 0.94 | 0.80 | 1.10 | 4.29E-01 | 0.715 |
| OID30826 | BRD2 | Neurology_II | BRD2 | P25440 | 6 | 32968594 | 32981505 | 0.93 | 0.78 | 1.11 | 4.30E-01 | 0.716 |
| OID20854 | CASP1 | Neurology | CASP1 | P29466 | 11 | 105025443 | 105035250 | 1.07 | 0.91 | 1.25 | 4.30E-01 | 0.716 |
| OID30133 | GAMT | Cardiometabolic_II | GAMT | Q14353 | 19 | 1397026 | 1401570 | 1.05 | 0.93 | 1.19 | 4.30E-01 | 0.716 |
| OID30062 | ATP2B4 | Cardiometabolic_II | ATP2B4 | P23634 | 1 | 203626832 | 203744081 | 0.93 | 0.78 | 1.11 | 4.30E-01 | 0.716 |
| OID20866 | MUC13 | Neurology | MUC13 | Q9H3R2 | 3 | 124905442 | 124953819 | 0.94 | 0.80 | 1.10 | 4.31E-01 | 0.717 |
| OID21203 | RANGAP1 | Oncology | RANGAP1 | P46060 | 22 | 41244779 | 41286187 | 0.93 | 0.78 | 1.11 | 4.31E-01 | 0.717 |
| OID31445 | SLC9A3R2 | Oncology_II | SLC9A3R2 | Q15599 | 16 | 2025356 | 2039026 | 0.93 | 0.76 | 1.12 | 4.31E-01 | 0.717 |
| OID30912 | PDRG1 | Neurology_II | PDRG1 | Q9NUG6 | 20 | 31944337 | 31952046 | 1.06 | 0.92 | 1.22 | 4.32E-01 | 0.717 |
| OID30384 | TNFSF8 | Cardiometabolic_II | TNFSF8 | P32971 | 9 | 114893343 | 114930595 | 1.06 | 0.91 | 1.24 | 4.32E-01 | 0.717 |
| OID20159 | ACAN | Cardiometabolic | ACAN | P16112 | 15 | 88803436 | 88875353 | 1.07 | 0.90 | 1.28 | 4.32E-01 | 0.717 |
| OID30971 | TEX101 | Neurology_II | TEX101 | Q9BY14 | 19 | 43401496 | 43418597 | 0.91 | 0.72 | 1.15 | 4.32E-01 | 0.717 |
| OID20753 | MEPE | Inflammation | MEPE | Q9NQ76 | 4 | 87821398 | 87846814 | 0.94 | 0.80 | 1.10 | 4.33E-01 | 0.718 |
| OID31146 | PLSCR3 | Neurology_II | PLSCR3 | Q9NRY6 | 17 | 7389727 | 7394842 | 0.94 | 0.79 | 1.10 | 4.34E-01 | 0.718 |
| OID20508 | SULT2A1 | Inflammation | SULT2A1 | Q06520 | 19 | 47870467 | 47886315 | 1.07 | 0.91 | 1.25 | 4.33E-01 | 0.718 |
| OID31383 | LTB | Oncology_II | LTB | Q06643 | 6 | 31580525 | 31582522 | 0.93 | 0.78 | 1.11 | 4.35E-01 | 0.720 |
| OID20994 | BMP4 | Neurology | BMP4 | P12644 | 14 | 53949736 | 53958761 | 1.07 | 0.90 | 1.27 | 4.37E-01 | 0.722 |
| OID20705 | ERBB3 | Inflammation | ERBB3 | P21860 | 12 | 56076799 | 56103505 | 0.93 | 0.79 | 1.11 | 4.37E-01 | 0.722 |
| OID20056 | TSLP | Cardiometabolic | TSLP | Q969D9 | 5 | 111070062 | 111078026 | 0.93 | 0.79 | 1.11 | 4.37E-01 | 0.722 |
| OID20333 | AMY2A | Cardiometabolic | AMY2A | P04746 | 1 | 103617427 | 103625780 | 0.94 | 0.80 | 1.10 | 4.38E-01 | 0.723 |
| OID30180 | C1GALT1C1 | Cardiometabolic_II | C1GALT1C1 | Q96EU7 | X | 120625674 | 120630054 | 0.93 | 0.79 | 1.11 | 4.39E-01 | 0.723 |
| OID21340 | CES3 | Oncology | CES3 | Q6UWW8 | 16 | 66961245 | 66975149 | 0.93 | 0.79 | 1.11 | 4.39E-01 | 0.723 |
| OID30699 | PGA4 | Inflammation_II | PGA4 | P0DJD7 | 11 | 61222347 | 61231694 | 0.95 | 0.82 | 1.09 | 4.39E-01 | 0.723 |
| OID20088 | PON2 | Cardiometabolic | PON2 | Q15165 | 7 | 95404862 | 95435329 | 0.94 | 0.80 | 1.10 | 4.38E-01 | 0.723 |
| OID20416 | GGH | Cardiometabolic | GGH | Q92820 | 8 | 63014881 | 63039407 | 1.07 | 0.91 | 1.26 | 4.40E-01 | 0.724 |
| OID31284 | ID4 | Oncology_II | ID4 | P47928 | 6 | 19837370 | 19842197 | 0.92 | 0.76 | 1.13 | 4.40E-01 | 0.724 |
| OID30109 | GASK1A | Cardiometabolic_II | GASK1A | Q9UFP1 | 3 | 42979087 | 43067898 | 0.94 | 0.79 | 1.11 | 4.40E-01 | 0.724 |
| OID31483 | ADGRE1 | Oncology_II | ADGRE1 | Q14246 | 19 | 6887566 | 6940459 | 0.94 | 0.80 | 1.10 | 4.41E-01 | 0.724 |
| OID30770 | HRG | Inflammation_II | HRG | P04196 | 3 | 186660216 | 186678234 | 0.93 | 0.79 | 1.11 | 4.41E-01 | 0.724 |
| OID21201 | KLK12 | Oncology | KLK12 | Q9UKR0 | 19 | 51029092 | 51035230 | 1.06 | 0.91 | 1.25 | 4.41E-01 | 0.724 |
| OID20608 | RAB6A | Inflammation | RAB6A | P20340 | 11 | 73675638 | 73761137 | 0.94 | 0.80 | 1.10 | 4.42E-01 | 0.725 |
| OID20917 | DRAXIN | Neurology | DRAXIN | Q8NBI3 | 1 | 11691710 | 11725857 | 1.07 | 0.91 | 1.25 | 4.42E-01 | 0.725 |
| OID20109 | ADAM15 | Cardiometabolic | ADAM15 | Q13444 | 1 | 155050566 | 155062775 | 1.07 | 0.90 | 1.26 | 4.43E-01 | 0.725 |
| OID21162 | ANKRD54 | Oncology | ANKRD54 | Q6NXT1 | 22 | 37830855 | 37849327 | 0.93 | 0.78 | 1.11 | 4.43E-01 | 0.725 |
| OID30827 | CD3D | Neurology_II | CD3D | P04234 | 11 | 118339075 | 118342705 | 1.06 | 0.91 | 1.24 | 4.43E-01 | 0.725 |
| OID31308 | CEACAM20 | Oncology_II | CEACAM20 | Q6UY09 | 19 | 44501677 | 44529788 | 0.94 | 0.80 | 1.10 | 4.43E-01 | 0.725 |
| OID20452 | AOC1 | Inflammation | AOC1 | P19801 | 7 | 150824627 | 150861504 | 0.94 | 0.79 | 1.11 | 4.44E-01 | 0.725 |
| OID20257 | PEAR1 | Cardiometabolic | PEAR1 | Q5VY43 | 1 | 156893698 | 156916429 | 0.94 | 0.80 | 1.11 | 4.44E-01 | 0.725 |
| OID20149 | PM20D1 | Cardiometabolic | PM20D1 | Q6GTS8 | 1 | 205828025 | 205850132 | 0.94 | 0.80 | 1.11 | 4.45E-01 | 0.725 |
| OID21347 | PPME1 | Oncology | PPME1 | Q9Y570 | 11 | 74171267 | 74254703 | 0.94 | 0.79 | 1.11 | 4.45E-01 | 0.725 |
| OID30829 | RAB3GAP1 | Neurology_II | RAB3GAP1 | Q15042 | 2 | 135052289 | 135176667 | 0.92 | 0.76 | 1.13 | 4.44E-01 | 0.725 |
| OID30601 | RLN2 | Inflammation_II | RLN2 | P04090 | 9 | 5299864 | 5304716 | 0.88 | 0.62 | 1.23 | 4.45E-01 | 0.725 |
| OID21240 | STXBP3 | Oncology | STXBP3 | O00186 | 1 | 108746674 | 108809523 | 0.94 | 0.80 | 1.11 | 4.45E-01 | 0.725 |
| OID30559 | TOP2B | Inflammation_II | TOP2B | Q02880 | 3 | 25597978 | 25664909 | 0.94 | 0.79 | 1.11 | 4.45E-01 | 0.725 |
| OID31328 | CINP | Oncology_II | CINP | Q9BW66 | 14 | 102341102 | 102362916 | 0.93 | 0.78 | 1.11 | 4.46E-01 | 0.725 |
| OID30527 | CSF3R | Inflammation_II | CSF3R | Q99062 | 1 | 36466043 | 36483278 | 0.94 | 0.79 | 1.11 | 4.47E-01 | 0.725 |
| OID20897 | DPEP1 | Neurology | DPEP1 | P16444 | 16 | 89613308 | 89638456 | 1.07 | 0.90 | 1.27 | 4.47E-01 | 0.725 |
| OID20391 | FCGR2A | Cardiometabolic | FCGR2A | P12318 | 1 | 161505430 | 161524013 | 0.94 | 0.81 | 1.10 | 4.46E-01 | 0.725 |
| OID21109 | SPINT1 | Neurology | SPINT1 | O43278 | 15 | 40844018 | 40858207 | 1.06 | 0.91 | 1.25 | 4.47E-01 | 0.725 |
| OID21464 | WIF1 | Oncology | WIF1 | Q9Y5W5 | 12 | 65050626 | 65121305 | 1.07 | 0.90 | 1.26 | 4.47E-01 | 0.725 |
| OID21466 | CCL8 | Oncology | CCL8 | P80075 | 17 | 34319435 | 34321402 | 0.94 | 0.79 | 1.11 | 4.48E-01 | 0.727 |
| OID20308 | BPIFB1 | Cardiometabolic | BPIFB1 | Q8TDL5 | 20 | 33273480 | 33309871 | 1.07 | 0.90 | 1.26 | 4.49E-01 | 0.728 |
| OID20781 | LGALS9 | Inflammation | LGALS9 | O00182 | 17 | 27629798 | 27649560 | 0.93 | 0.78 | 1.12 | 4.50E-01 | 0.728 |
| OID21329 | SEMA4C | Oncology | SEMA4C | Q9C0C4 | 2 | 96859718 | 96870757 | 0.94 | 0.79 | 1.11 | 4.49E-01 | 0.728 |
| OID20348 | IL18BP | Cardiometabolic | IL18BP | O95998 | 11 | 71998613 | 72005715 | 1.06 | 0.91 | 1.24 | 4.50E-01 | 0.728 |
| OID31090 | STX3 | Neurology_II | STX3 | Q13277 | 11 | 59713456 | 59805882 | 0.94 | 0.79 | 1.11 | 4.50E-01 | 0.728 |
| OID21188 | ADCYAP1R1 | Oncology | ADCYAP1R1 | P41586 | 7 | 31052308 | 31111479 | 0.93 | 0.76 | 1.13 | 4.51E-01 | 0.729 |
| OID21151 | F11R | Neurology | F11R | Q9Y624 | 1 | 160995211 | 161021343 | 0.94 | 0.80 | 1.11 | 4.51E-01 | 0.729 |
| OID20200 | FABP2 | Cardiometabolic | FABP2 | P12104 | 4 | 119317250 | 119322138 | 1.06 | 0.91 | 1.24 | 4.51E-01 | 0.729 |
| OID21075 | GGT1 | Neurology | GGT1 | P19440 | 22 | 24594811 | 24629005 | 0.93 | 0.78 | 1.12 | 4.51E-01 | 0.729 |
| OID30720 | CFHR2 | Inflammation_II | CFHR2 | P36980 | 1 | 196943738 | 196959622 | 0.94 | 0.80 | 1.10 | 4.52E-01 | 0.729 |
| OID30926 | SLC1A4 | Neurology_II | SLC1A4 | P43007 | 2 | 64988477 | 65023865 | 0.93 | 0.78 | 1.12 | 4.52E-01 | 0.729 |
| OID30850 | ATXN2 | Neurology_II | ATXN2 | Q99700 | 12 | 111443485 | 111599676 | 1.06 | 0.91 | 1.23 | 4.55E-01 | 0.733 |
| OID20405 | COMP | Cardiometabolic | COMP | P49747 | 19 | 18782773 | 18791305 | 0.94 | 0.80 | 1.11 | 4.55E-01 | 0.733 |
| OID20634 | NFASC | Inflammation | NFASC | O94856 | 1 | 204828651 | 205022822 | 0.93 | 0.78 | 1.12 | 4.55E-01 | 0.733 |
| OID21057 | NTRK3 | Neurology | NTRK3 | Q16288 | 15 | 87859751 | 88256768 | 1.07 | 0.90 | 1.28 | 4.55E-01 | 0.733 |
| OID20860 | KRT5 | Neurology | KRT5 | P13647 | 12 | 52514575 | 52520530 | 0.94 | 0.80 | 1.11 | 4.56E-01 | 0.734 |
| OID30079 | ATP5F1D | Cardiometabolic_II | ATP5F1D | P30049 | 19 | 1241746 | 1244825 | 1.04 | 0.93 | 1.17 | 4.57E-01 | 0.734 |
| OID21040 | CD74 | Neurology | CD74 | P04233 | 5 | 150401637 | 150412969 | 1.06 | 0.90 | 1.26 | 4.57E-01 | 0.734 |
| OID30406 | CPQ | Cardiometabolic_II | CPQ | Q9Y646 | 8 | 96645242 | 97149654 | 0.93 | 0.78 | 1.12 | 4.57E-01 | 0.734 |
| OID21155 | TNFRSF1A | Neurology | TNFRSF1A | P19438 | 12 | 6328757 | 6342114 | 1.06 | 0.90 | 1.25 | 4.57E-01 | 0.734 |
| OID20111 | BMP6 | Cardiometabolic | BMP6 | P22004 | 6 | 7726099 | 7881728 | 0.94 | 0.80 | 1.11 | 4.58E-01 | 0.735 |
| OID30210 | IGSF21 | Cardiometabolic_II | IGSF21 | Q96ID5 | 1 | 18107798 | 18378483 | 0.93 | 0.78 | 1.12 | 4.58E-01 | 0.735 |
| OID30686 | APOA2 | Inflammation_II | APOA2 | P02652 | 1 | 161222292 | 161223631 | 1.06 | 0.91 | 1.24 | 4.59E-01 | 0.735 |
| OID30471 | ADAMTS4 | Inflammation_II | ADAMTS4 | O75173 | 1 | 161184302 | 161199054 | 0.94 | 0.79 | 1.11 | 4.60E-01 | 0.735 |
| OID20299 | CCN3 | Cardiometabolic | CCN3 | P48745 | 8 | 119416446 | 119424434 | 0.94 | 0.79 | 1.11 | 4.59E-01 | 0.735 |
| OID30163 | KIF1C | Cardiometabolic_II | KIF1C | O43896 | 17 | 4997950 | 5028401 | 0.93 | 0.77 | 1.12 | 4.60E-01 | 0.735 |
| OID21079 | DSC2 | Neurology | DSC2 | Q02487 | 18 | 31058840 | 31102522 | 1.06 | 0.90 | 1.25 | 4.62E-01 | 0.737 |
| OID31006 | HJV | Neurology_II | HJV | Q6ZVN8 | 1 | 146017468 | 146036746 | 1.06 | 0.91 | 1.24 | 4.62E-01 | 0.737 |
| OID31267 | TCP11 | Oncology_II | TCP11 | Q8WWU5 | 6 | 35118071 | 35148610 | 1.05 | 0.92 | 1.21 | 4.61E-01 | 0.737 |
| OID21364 | TMPRSS15 | Oncology | TMPRSS15 | P98073 | 21 | 18269116 | 18485879 | 1.06 | 0.91 | 1.25 | 4.61E-01 | 0.737 |
| OID20052 | CEBPB | Cardiometabolic | CEBPB | P17676 | 20 | 50190830 | 50192668 | 0.94 | 0.79 | 1.11 | 4.63E-01 | 0.738 |
| OID20970 | LAYN | Neurology | LAYN | Q6UX15 | 11 | 111540280 | 111561745 | 1.06 | 0.91 | 1.24 | 4.63E-01 | 0.738 |
| OID31282 | SLK | Oncology_II | SLK | Q9H2G2 | 10 | 103967140 | 104029233 | 0.93 | 0.76 | 1.13 | 4.63E-01 | 0.738 |
| OID31176 | ESPL1 | Oncology_II | ESPL1 | Q14674 | 12 | 53268299 | 53293638 | 0.94 | 0.79 | 1.11 | 4.64E-01 | 0.739 |
| OID20803 | ARID4B | Neurology | ARID4B | Q4LE39 | 1 | 235131634 | 235328219 | 0.93 | 0.75 | 1.14 | 4.64E-01 | 0.739 |
| OID30382 | MYH9 | Cardiometabolic_II | MYH9 | P35579 | 22 | 36281280 | 36388010 | 0.94 | 0.80 | 1.11 | 4.65E-01 | 0.739 |
| OID20369 | TFRC | Cardiometabolic | TFRC | P02786 | 3 | 196027183 | 196082096 | 0.94 | 0.79 | 1.11 | 4.65E-01 | 0.739 |
| OID30249 | CALCOCO2 | Cardiometabolic_II | CALCOCO2 | Q13137 | 17 | 48831018 | 48866522 | 0.94 | 0.80 | 1.11 | 4.66E-01 | 0.740 |
| OID30337 | HEPH | Cardiometabolic_II | HEPH | Q9BQS7 | X | 66162671 | 66268867 | 0.94 | 0.79 | 1.11 | 4.66E-01 | 0.740 |
| OID20419 | IL2 | Inflammation | IL2 | P60568 | 4 | 122451470 | 122456725 | 0.94 | 0.80 | 1.11 | 4.66E-01 | 0.740 |
| OID30918 | PSMD5 | Neurology_II | PSMD5 | Q16401 | 9 | 120815496 | 120842951 | 0.94 | 0.78 | 1.12 | 4.66E-01 | 0.740 |
| OID31395 | DTNB | Oncology_II | DTNB | O60941 | 2 | 25377198 | 25673647 | 0.93 | 0.77 | 1.12 | 4.67E-01 | 0.740 |
| OID30704 | PON1 | Inflammation_II | PON1 | P27169 | 7 | 95297676 | 95324532 | 0.94 | 0.80 | 1.11 | 4.67E-01 | 0.740 |
| OID30671 | CGB3_CGB5_C( | Inflammation_II | B5;CGB8;CGB3 | 86;P0DN86;P0DN86 | 19;19;19 | ;49047638;49022869 | ;49049111;49024333 | 0.93 | 0.76 | 1.13 | 4.68E-01 | 0.740 |
| OID30703 | DBH | Inflammation_II | DBH | P09172 | 9 | 133636363 | 133659329 | 0.94 | 0.81 | 1.10 | 4.68E-01 | 0.740 |
| OID30502 | TGFBR1 | Inflammation_II | TGFBR1 | P36897 | 9 | 99104038 | 99154192 | 0.94 | 0.79 | 1.11 | 4.68E-01 | 0.740 |
| OID30769 | APOA1 | Inflammation_II | APOA1 | P02647 | 11 | 116835751 | 116837622 | 0.93 | 0.76 | 1.13 | 4.69E-01 | 0.741 |
| OID20696 | IL10RB | Inflammation | IL10RB | Q08334 | 21 | 33266367 | 33310187 | 1.06 | 0.91 | 1.24 | 4.70E-01 | 0.741 |
| OID31268 | OFD1 | Oncology_II | OFD1 | O75665 | X | 13734743 | 13777955 | 1.06 | 0.91 | 1.22 | 4.69E-01 | 0.741 |
| OID21094 | PIK3IP1 | Neurology | PIK3IP1 | Q96FE7 | 22 | 31281594 | 31292534 | 1.06 | 0.90 | 1.25 | 4.70E-01 | 0.741 |
| OID20809 | IL34 | Neurology | IL34 | Q6ZMJ4 | 16 | 70579895 | 70660682 | 1.06 | 0.90 | 1.25 | 4.70E-01 | 0.741 |
| OID20778 | FABP1 | Inflammation | FABP1 | P07148 | 2 | 88122982 | 88128062 | 1.06 | 0.90 | 1.26 | 4.71E-01 | 0.743 |

| | | | | | | | | | | | | |
|---|---|---|---|---|---|---|---|---|---|---|---|---|
| OID20693 | CCL23 | Inflammation | CCL23 | P55773 | 17 | 36013056 | 36017972 | 0.94 | 0.80 | 1.11 | 4.72E-01 | 0.743 |
| OID20100 | ENTPD6 | Cardiometabolic | ENTPD6 | O75354 | 20 | 25195693 | 25228075 | 1.06 | 0.90 | 1.26 | 4.72E-01 | 0.743 |
| OID31295 | FUT1 | Oncology_II | FUT1 | P19526 | 19 | 48748011 | 48755390 | 1.05 | 0.91 | 1.22 | 4.72E-01 | 0.743 |
| OID21193 | FCRLB | Oncology | FCRLB | Q6BAA4 | 1 | 161721563 | 161728143 | 0.94 | 0.79 | 1.12 | 4.73E-01 | 0.743 |
| OID30404 | SLURP1 | Cardiometabolic_II | SLURP1 | P55000 | 8 | 142740949 | 142742406 | 1.05 | 0.92 | 1.21 | 4.72E-01 | 0.743 |
| OID20902 | PPP3R1 | Neurology | PPP3R1 | P63098 | 2 | 68178857 | 68256237 | 0.94 | 0.79 | 1.12 | 4.75E-01 | 0.745 |
| OID30077 | SLC12A2 | Cardiometabolic_II | SLC12A2 | P55011 | 5 | 128083766 | 128189677 | 0.94 | 0.78 | 1.12 | 4.75E-01 | 0.745 |
| OID21310 | VNN2 | Oncology | VNN2 | O95498 | 6 | 132743870 | 132763459 | 0.94 | 0.80 | 1.11 | 4.75E-01 | 0.745 |
| OID21491 | FLT4 | Oncology | FLT4 | P35916 | 5 | 180601506 | 180649624 | 0.94 | 0.80 | 1.11 | 4.75E-01 | 0.746 |
| OID30738 | CFD | Inflammation_II | CFD | P00746 | 19 | 859453 | 867884 | 0.94 | 0.79 | 1.12 | 4.77E-01 | 0.746 |
| OID20550 | COL9A1 | Inflammation | COL9A1 | P20849 | 6 | 70216040 | 70303084 | 0.94 | 0.80 | 1.11 | 4.77E-01 | 0.746 |
| OID21168 | GH2 | Oncology | GH2 | P01242 | 17 | 63880215 | 63881944 | 0.94 | 0.80 | 1.11 | 4.77E-01 | 0.746 |
| OID30375 | PDIA4 | Cardiometabolic_II | PDIA4 | P13667 | 7 | 149003062 | 149028662 | 0.94 | 0.79 | 1.11 | 4.77E-01 | 0.746 |
| OID30668 | PRDX2 | Inflammation_II | PRDX2 | P32119 | 19 | 12796820 | 12801800 | 0.94 | 0.78 | 1.12 | 4.76E-01 | 0.746 |
| OID31058 | PSMD1 | Neurology_II | PSMD1 | Q99460 | 2 | 231056845 | 231173116 | 0.94 | 0.79 | 1.11 | 4.77E-01 | 0.746 |
| OID20847 | PTS | Neurology | PTS | Q03393 | 11 | 112226367 | 112269955 | 0.94 | 0.78 | 1.12 | 4.77E-01 | 0.746 |
| OID20904 | MAD1L1 | Neurology | MAD1L1 | Q9Y6D9 | 7 | 1815793 | 2233243 | 0.93 | 0.77 | 1.13 | 4.78E-01 | 0.747 |
| OID30416 | LAMP1 | Cardiometabolic_II | LAMP1 | P11279 | 13 | 113297239 | 113323672 | 0.94 | 0.79 | 1.11 | 4.79E-01 | 0.747 |
| OID20186 | SEMA3F | Cardiometabolic | SEMA3F | Q13275 | 3 | 50155045 | 50189075 | 1.06 | 0.90 | 1.26 | 4.79E-01 | 0.747 |
| OID30755 | CFB | Inflammation_II | CFB | P00751 | 6 | 31945650 | 31952086 | 1.07 | 0.89 | 1.28 | 4.80E-01 | 0.748 |
| OID20466 | DGKZ | Inflammation | DGKZ | Q13574 | 11 | 46332905 | 46380554 | 1.06 | 0.90 | 1.25 | 4.80E-01 | 0.748 |
| OID21374 | S100A12 | Oncology | S100A12 | P80511 | 1 | 153373711 | 153375621 | 1.06 | 0.90 | 1.25 | 4.80E-01 | 0.748 |
| OID30381 | PRG2 | Cardiometabolic_II | PRG2 | P13727 | 11 | 57386780 | 57390650 | 1.06 | 0.90 | 1.25 | 4.81E-01 | 0.749 |
| OID30389 | BGLAP | Cardiometabolic_II | BGLAP | P02818 | 1 | 156242184 | 156243317 | 0.95 | 0.81 | 1.10 | 4.83E-01 | 0.751 |
| OID30287 | TMEM132A | Cardiometabolic_II | TMEM132A | Q24JP5 | 11 | 60924460 | 60937159 | 0.94 | 0.80 | 1.11 | 4.83E-01 | 0.751 |
| OID31234 | TPRKB | Oncology_II | TPRKB | Q9Y3C4 | 2 | 73729104 | 73737400 | 1.05 | 0.92 | 1.19 | 4.85E-01 | 0.754 |
| OID21449 | CD5 | Oncology | CD5 | P06127 | 11 | 61102489 | 61127852 | 0.94 | 0.80 | 1.11 | 4.86E-01 | 0.755 |
| OID20910 | CLPP | Neurology | CLPP | Q16740 | 19 | 6361531 | 6370242 | 0.94 | 0.80 | 1.11 | 4.87E-01 | 0.755 |
| OID20324 | LILRB5 | Cardiometabolic | LILRB5 | O75023 | 19 | 54249421 | 54257301 | 0.95 | 0.81 | 1.11 | 4.88E-01 | 0.755 |
| OID21047 | OXT | Neurology | OXT | P01178 | 20 | 3071620 | 3072517 | 0.94 | 0.79 | 1.12 | 4.86E-01 | 0.755 |
| OID31214 | PDIA2 | Oncology_II | PDIA2 | Q13087 | 16 | 283164 | 287215 | 0.94 | 0.79 | 1.12 | 4.88E-01 | 0.755 |
| OID20439 | PREB | Inflammation | PREB | Q9HCU5 | 2 | 27130756 | 27134666 | 0.94 | 0.79 | 1.12 | 4.88E-01 | 0.755 |
| OID30948 | SCN2B | Neurology_II | SCN2B | O60939 | 11 | 118162806 | 118176639 | 0.95 | 0.81 | 1.11 | 4.87E-01 | 0.755 |
| OID30056 | SKIV2L | Cardiometabolic_II | SKIV2L | Q15477 | 6 | 31959116 | 31969818 | 1.05 | 0.92 | 1.20 | 4.88E-01 | 0.755 |
| OID30811 | SNAP25 | Neurology_II | SNAP25 | P60880 | 20 | 10172395 | 10308258 | 0.94 | 0.79 | 1.12 | 4.87E-01 | 0.755 |
| OID21525 | DLL1 | Oncology | DLL1 | O00548 | 6 | 170282206 | 170306565 | 1.06 | 0.90 | 1.24 | 4.88E-01 | 0.755 |
| OID20170 | CDH2 | Cardiometabolic | CDH2 | P19022 | 18 | 27932879 | 28177946 | 1.06 | 0.89 | 1.27 | 4.89E-01 | 0.756 |
| OID21496 | TEK | Oncology | TEK | Q02763 | 9 | 27109141 | 27230174 | 1.06 | 0.90 | 1.25 | 4.90E-01 | 0.757 |
| OID20754 | CDON | Inflammation | CDON | Q4KMG0 | 11 | 125955796 | 126063335 | 0.94 | 0.80 | 1.11 | 4.90E-01 | 0.757 |
| OID20279 | CRTAC1 | Cardiometabolic | CRTAC1 | Q9NQ79 | 10 | 97865000 | 98030828 | 1.06 | 0.89 | 1.27 | 4.91E-01 | 0.757 |
| OID21395 | HAO1 | Oncology | HAO1 | Q9UJM8 | 20 | 7882985 | 7940458 | 0.94 | 0.79 | 1.12 | 4.91E-01 | 0.757 |
| OID21204 | MAVS | Oncology | MAVS | Q7Z434 | 20 | 3846799 | 3876123 | 0.94 | 0.80 | 1.12 | 4.91E-01 | 0.757 |
| OID20351 | SSC5D | Cardiometabolic | SSC5D | A1L4H1 | 19 | 55488404 | 55519099 | 0.94 | 0.79 | 1.12 | 4.91E-01 | 0.757 |
| OID30797 | SYNGAP1 | Neurology_II | SYNGAP1 | Q96PV0 | 6 | 33419661 | 33453689 | 1.06 | 0.90 | 1.25 | 4.91E-01 | 0.757 |
| OID31400 | VSIG10 | Oncology_II | VSIG10 | Q8N0Z9 | 12 | 118063593 | 118136026 | 0.94 | 0.78 | 1.13 | 4.92E-01 | 0.757 |
| OID31241 | FGF7 | Oncology_II | FGF7 | P21781 | 15 | 49423237 | 49488775 | 0.94 | 0.79 | 1.12 | 4.92E-01 | 0.757 |
| OID20082 | ZBTB17 | Cardiometabolic | ZBTB17 | Q13105 | 1 | 15941869 | 15976132 | 0.94 | 0.78 | 1.13 | 4.93E-01 | 0.758 |
| OID20464 | CXCL12 | Inflammation | CXCL12 | P48061 | 10 | 44370165 | 44386493 | 0.95 | 0.81 | 1.11 | 4.94E-01 | 0.759 |
| OID20529 | FCRL6 | Inflammation | FCRL6 | Q6DN72 | 1 | 159800511 | 159816257 | 0.95 | 0.81 | 1.11 | 4.94E-01 | 0.759 |
| OID30060 | EIF2AK3 | Cardiometabolic_II | EIF2AK3 | Q9NZJ5 | 2 | 88556741 | 88691518 | 0.94 | 0.80 | 1.11 | 4.95E-01 | 0.760 |
| OID21253 | MAP3K5 | Oncology | MAP3K5 | Q99683 | 6 | 136557046 | 136792477 | 0.94 | 0.80 | 1.11 | 4.95E-01 | 0.760 |
| OID30208 | M6PR | Cardiometabolic_II | M6PR | P20645 | 12 | 8940361 | 8949761 | 1.06 | 0.90 | 1.24 | 4.96E-01 | 0.761 |
| OID20805 | SCARB1 | Neurology | SCARB1 | Q8WTV0 | 12 | 124776856 | 124882668 | 0.94 | 0.77 | 1.13 | 4.96E-01 | 0.761 |
| OID30250 | EPPK1 | Cardiometabolic_II | EPPK1 | P58107 | 8 | 143857324 | 143878467 | 0.94 | 0.79 | 1.12 | 4.97E-01 | 0.762 |
| OID30713 | MST1 | Inflammation_II | MST1 | P26927 | 3 | 49683947 | 49689501 | 0.95 | 0.82 | 1.10 | 4.97E-01 | 0.762 |
| OID30275 | ADAMTSL4 | Cardiometabolic_II | ADAMTSL4 | Q6UY14 | 1 | 150549369 | 150560937 | 0.95 | 0.81 | 1.11 | 4.98E-01 | 0.762 |
| OID30093 | BRD3 | Cardiometabolic_II | BRD3 | Q15059 | 9 | 134030305 | 134068535 | 0.93 | 0.77 | 1.14 | 4.99E-01 | 0.762 |
| OID21124 | PLAU | Neurology | PLAU | P00749 | 10 | 73909177 | 73917496 | 0.94 | 0.79 | 1.12 | 4.98E-01 | 0.762 |
| OID20372 | F7 | Cardiometabolic | F7 | P08709 | 13 | 113105788 | 113120685 | 0.95 | 0.81 | 1.11 | 4.99E-01 | 0.763 |
| OID20472 | IL5 | Inflammation | IL5 | P05113 | 5 | 132541445 | 132556838 | 0.94 | 0.79 | 1.12 | 5.00E-01 | 0.764 |
| OID30494 | DDX39A | Inflammation_II | DDX39A | O00148 | 19 | 14408798 | 14419383 | 1.06 | 0.89 | 1.26 | 5.02E-01 | 0.767 |
| OID20951 | MDGA1 | Neurology | MDGA1 | Q8NFP4 | 6 | 37630679 | 37699306 | 0.94 | 0.80 | 1.12 | 5.03E-01 | 0.768 |
| OID21446 | TNFRSF19 | Oncology | TNFRSF19 | Q9NS68 | 13 | 23570370 | 23676104 | 1.05 | 0.90 | 1.23 | 5.05E-01 | 0.770 |
| OID30560 | STX7 | Inflammation_II | STX7 | O15400 | 6 | 132445867 | 132513198 | 0.94 | 0.79 | 1.12 | 5.06E-01 | 0.771 |
| OID31015 | TMEM25 | Neurology_II | TMEM25 | Q86YD3 | 11 | 118531041 | 118547280 | 1.06 | 0.90 | 1.24 | 5.06E-01 | 0.771 |
| OID31414 | GABARAP | Oncology_II | GABARAP | O95166 | 17 | 7240008 | 7242449 | 1.05 | 0.90 | 1.23 | 5.07E-01 | 0.772 |
| OID20876 | GSTP1 | Neurology | GSTP1 | P09211 | 11 | 67583742 | 67586656 | 0.95 | 0.81 | 1.11 | 5.07E-01 | 0.772 |
| OID20575 | HSD11B1 | Inflammation | HSD11B1 | P28845 | 1 | 209686178 | 209734949 | 1.06 | 0.90 | 1.25 | 5.08E-01 | 0.772 |
| OID30472 | IL36A | Inflammation_II | IL36A | Q9UHA7 | 2 | 113005459 | 113008044 | 0.94 | 0.80 | 1.12 | 5.09E-01 | 0.773 |
| OID20397 | CD93 | Cardiometabolic | CD93 | Q9NPY3 | 20 | 23079360 | 23086324 | 0.95 | 0.81 | 1.11 | 5.09E-01 | 0.773 |
| OID30878 | NLGN2 | Neurology_II | NLGN2 | Q8NFZ4 | 17 | 7404874 | 7419860 | 0.95 | 0.80 | 1.11 | 5.10E-01 | 0.774 |
| OID20736 | ENPP5 | Inflammation | ENPP5 | Q9UJA9 | 6 | 46159185 | 46170980 | 1.06 | 0.88 | 1.28 | 5.11E-01 | 0.774 |
| OID30500 | IGFL4 | Inflammation_II | IGFL4 | Q6B9Z1 | 19 | 46039182 | 46077118 | 0.94 | 0.79 | 1.12 | 5.10E-01 | 0.774 |
| OID20396 | VCAM1 | Cardiometabolic | VCAM1 | P19320 | 1 | 100719742 | 100739045 | 1.06 | 0.90 | 1.24 | 5.11E-01 | 0.774 |
| OID21267 | BAMBI | Oncology | BAMBI | Q13145 | 10 | 28677510 | 28682932 | 0.95 | 0.80 | 1.11 | 5.11E-01 | 0.774 |
| OID21418 | CD300E | Oncology | CD300E | Q496F6 | 17 | 74609885 | 74623738 | 1.06 | 0.90 | 1.25 | 5.11E-01 | 0.774 |
| OID20751 | CRELD2 | Inflammation | CRELD2 | Q6UXH1 | 22 | 49918167 | 49927540 | 0.95 | 0.80 | 1.12 | 5.12E-01 | 0.775 |
| OID30681 | CSF1R | Inflammation_II | CSF1R | P07333 | 5 | 150053291 | 150113372 | 1.06 | 0.89 | 1.25 | 5.13E-01 | 0.775 |
| OID20357 | HYAL1 | Cardiometabolic | HYAL1 | Q12794 | 3 | 50299890 | 50312381 | 0.95 | 0.81 | 1.11 | 5.13E-01 | 0.775 |
| OID31322 | SERPINH1 | Oncology_II | SERPINH1 | P50454 | 11 | 75562056 | 75572783 | 0.95 | 0.80 | 1.12 | 5.13E-01 | 0.775 |
| OID20221 | SPON2 | Cardiometabolic | SPON2 | Q9BUD6 | 4 | 1166932 | 1208962 | 1.06 | 0.90 | 1.24 | 5.13E-01 | 0.775 |
| OID20340 | AMY2B | Cardiometabolic | AMY2B | P19961 | 1 | 103553815 | 103579534 | 0.95 | 0.81 | 1.11 | 5.16E-01 | 0.775 |
| OID20523 | CCL7 | Inflammation | CCL7 | P80098 | 17 | 34270221 | 34272242 | 0.94 | 0.78 | 1.13 | 5.14E-01 | 0.775 |
| OID20134 | CDH17 | Cardiometabolic | CDH17 | Q12864 | 8 | 94127162 | 94217303 | 0.94 | 0.79 | 1.12 | 5.14E-01 | 0.775 |
| OID31083 | GAST | Neurology_II | GAST | P01350 | 17 | 41712331 | 41715969 | 0.95 | 0.82 | 1.11 | 5.14E-01 | 0.775 |
| OID20455 | IL11 | Inflammation | IL11 | P20809 | 19 | 55364382 | 55370463 | 1.06 | 0.89 | 1.26 | 5.15E-01 | 0.775 |
| OID21361 | IQGAP2 | Oncology | IQGAP2 | Q13576 | 5 | 76403285 | 76708132 | 0.95 | 0.80 | 1.12 | 5.15E-01 | 0.775 |
| OID30842 | PNMA2 | Neurology_II | PNMA2 | Q9UL42 | 8 | 26504701 | 26514092 | 1.06 | 0.89 | 1.25 | 5.14E-01 | 0.775 |
| OID30341 | SGSH | Cardiometabolic_II | SGSH | P51688 | 17 | 80206716 | 80220923 | 0.95 | 0.80 | 1.12 | 5.15E-01 | 0.775 |
| OID21353 | SMAD5 | Oncology | SMAD5 | Q99717 | 5 | 136132845 | 136188747 | 0.94 | 0.80 | 1.12 | 5.15E-01 | 0.775 |
| OID20198 | CLUL1 | Cardiometabolic | CLUL1 | Q15846 | 18 | 596988 | 650334 | 1.06 | 0.89 | 1.26 | 5.18E-01 | 0.779 |
| OID21327 | IL12A_IL12B | Oncology | IL12A;IL12B | P29459;P29460 | 3;5 | [illegible]9988835;159314780 | [illegible]59996019;159330487 | 1.05 | 0.90 | 1.23 | 5.18E-01 | 0.779 |
| OID20230 | HSPB1 | Cardiometabolic | HSPB1 | P04792 | 7 | 76302673 | 76304295 | 0.95 | 0.81 | 1.12 | 5.19E-01 | 0.779 |
| OID20122 | AK1 | Cardiometabolic | AK1 | P00568 | 9 | 127866486 | 127877675 | 0.95 | 0.80 | 1.12 | 5.21E-01 | 0.782 |
| OID31326 | PTTG1 | Oncology_II | PTTG1 | O95997 | 5 | 160421855 | 160428739 | 0.94 | 0.79 | 1.13 | 5.21E-01 | 0.782 |
| OID20700 | IL1RN | Inflammation | IL1RN | P18510 | 2 | 113107214 | 113134016 | 1.06 | 0.88 | 1.28 | 5.22E-01 | 0.783 |
| OID20513 | BCL2L11 | Inflammation | BCL2L11 | O43521 | 2 | 111119378 | 111168445 | 0.95 | 0.80 | 1.12 | 5.24E-01 | 0.785 |
| OID20521 | TBC1D5 | Inflammation | TBC1D5 | Q92609 | 3 | 17157162 | 18444817 | 0.95 | 0.80 | 1.12 | 5.24E-01 | 0.785 |
| OID20422 | IL20RA | Inflammation | IL20RA | Q9UHF4 | 6 | 136999971 | 137045180 | 1.05 | 0.90 | 1.24 | 5.25E-01 | 0.786 |
| OID20329 | PRTN3 | Cardiometabolic | PRTN3 | P24158 | 19 | 840999 | 848175 | 1.06 | 0.89 | 1.25 | 5.26E-01 | 0.787 |
| OID31173 | ANKRA2 | Oncology_II | ANKRA2 | Q9H9E1 | 5 | 73552190 | 73565667 | 0.95 | 0.81 | 1.11 | 5.27E-01 | 0.787 |
| OID21056 | CTSS | Neurology | CTSS | P25774 | 1 | 150730079 | 150765957 | 0.95 | 0.80 | 1.12 | 5.27E-01 | 0.787 |
| OID30914 | CUZD1 | Neurology_II | CUZD1 | Q86UP6 | 10 | 122832158 | 122846175 | 0.94 | 0.78 | 1.14 | 5.28E-01 | 0.787 |

| | | | | | | | | | | | | |
|---|---|---|---|---|---|---|---|---|---|---|---|---|
| OID20287 | ENG | Cardiometabolic | ENG | P17813 | 9 | 127815013 | 127854658 | 1.05 | 0.90 | 1.24 | 5.28E-01 | 0.787 |
| OID21301 | FLT1 | Oncology | FLT1 | P17948 | 13 | 28300346 | 28495145 | 1.05 | 0.90 | 1.24 | 5.29E-01 | 0.787 |
| OID20315 | ITGB2 | Cardiometabolic | ITGB2 | P05107 | 21 | 44885953 | 44931989 | 0.95 | 0.80 | 1.12 | 5.27E-01 | 0.787 |
| OID31473 | PSAPL1 | Oncology_II | PSAPL1 | Q6NUJ1 | 4 | 7430285 | 7434930 | 1.05 | 0.90 | 1.22 | 5.29E-01 | 0.787 |
| OID30897 | SLC44A4 | Neurology_II | SLC44A4 | Q53GD3 | 6 | 31863192 | 31879046 | 1.05 | 0.90 | 1.22 | 5.28E-01 | 0.787 |
| OID21170 | STX16 | Oncology | STX16 | O14662 | 20 | 58651253 | 58679526 | 1.06 | 0.89 | 1.25 | 5.29E-01 | 0.787 |
| OID20476 | TPT1 | Inflammation | TPT1 | P13693 | 13 | 45333471 | 45341284 | 1.05 | 0.90 | 1.22 | 5.29E-01 | 0.787 |
| OID21257 | CRH | Oncology | CRH | P06850 | 8 | 66176376 | 66178464 | 0.95 | 0.80 | 1.12 | 5.30E-01 | 0.788 |
| OID31169 | IGLON5 | Oncology_II | IGLON5 | A6NGN9 | 19 | 51311644 | 51330891 | 0.94 | 0.79 | 1.13 | 5.30E-01 | 0.788 |
| OID21440 | F3 | Oncology | F3 | P13726 | 1 | 94529173 | 94541759 | 1.06 | 0.89 | 1.25 | 5.31E-01 | 0.789 |
| OID21394 | SIGLEC6 | Oncology | SIGLEC6 | O43699 | 19 | 51517819 | 51531856 | 1.05 | 0.90 | 1.22 | 5.33E-01 | 0.791 |
| OID20862 | SLIT2 | Neurology | SLIT2 | O94813 | 4 | 20251905 | 20620561 | 1.05 | 0.89 | 1.25 | 5.32E-01 | 0.791 |
| OID20387 | FCGR3B | Cardiometabolic | FCGR3B | O75015 | 1 | 161623196 | 161631963 | 1.05 | 0.89 | 1.24 | 5.33E-01 | 0.791 |
| OID20947 | TMPRSS5 | Neurology | TMPRSS5 | Q9H3S3 | 11 | 113687547 | 113706373 | 0.95 | 0.80 | 1.12 | 5.33E-01 | 0.791 |
| OID21373 | DEFB4A_DEFB4 | Oncology | EFB4A;DEFB4B | O15263;O15263 | 8;8 | 7894677;7414855 | 7896716;7416863 | 1.05 | 0.89 | 1.24 | 5.34E-01 | 0.791 |
| OID31264 | GTF2IRD1 | Oncology_II | GTF2IRD1 | Q9UHL9 | 7 | 74453790 | 74602605 | 0.95 | 0.79 | 1.13 | 5.34E-01 | 0.791 |
| OID30293 | ECHDC3 | Cardiometabolic_II | ECHDC3 | Q96DC8 | 10 | 11742366 | 11764070 | 0.95 | 0.80 | 1.12 | 5.35E-01 | 0.791 |
| OID20492 | AMN | Inflammation | AMN | Q9BXJ7 | 14 | 102922663 | 102933596 | 1.05 | 0.90 | 1.23 | 5.39E-01 | 0.792 |
| OID30856 | BLOC1S2 | Neurology_II | BLOC1S2 | Q6QNY1 | 10 | 100273280 | 100286680 | 0.95 | 0.82 | 1.11 | 5.38E-01 | 0.792 |
| OID31329 | CENPF | Oncology_II | CENPF | P49454 | 1 | 214603195 | 214664571 | 0.95 | 0.79 | 1.13 | 5.38E-01 | 0.792 |
| OID30351 | FABP3 | Cardiometabolic_II | FABP3 | P05413 | 1 | 31365253 | 31376850 | 0.94 | 0.79 | 1.13 | 5.38E-01 | 0.792 |
| OID31141 | HIP1R | Neurology_II | HIP1R | O75146 | 12 | 122834453 | 122862961 | 0.94 | 0.78 | 1.14 | 5.39E-01 | 0.792 |
| OID20236 | IGFBP7 | Cardiometabolic | IGFBP7 | Q16270 | 4 | 57030773 | 57110385 | 1.05 | 0.90 | 1.23 | 5.38E-01 | 0.792 |
| OID20469 | IL17A | Inflammation | IL17A | Q16552 | 6 | 52186375 | 52190638 | 1.05 | 0.91 | 1.21 | 5.38E-01 | 0.792 |
| OID30475 | IL31RA | Inflammation_II | IL31RA | Q8NI17 | 5 | 55851357 | 55922854 | 0.95 | 0.80 | 1.12 | 5.39E-01 | 0.792 |
| OID31096 | PALM2 | Neurology_II | PALM2AKAP2 | Q8IXS6 | 9 | 109498325 | 110172512 | 0.95 | 0.79 | 1.13 | 5.38E-01 | 0.792 |
| OID31213 | RASGRF1 | Oncology_II | RASGRF1 | Q13972 | 15 | 78959906 | 79090780 | 0.95 | 0.81 | 1.12 | 5.39E-01 | 0.792 |
| OID21050 | THY1 | Neurology | THY1 | P04216 | 11 | 119415476 | 119424985 | 0.95 | 0.81 | 1.12 | 5.37E-01 | 0.792 |
| OID20359 | TNC | Cardiometabolic | TNC | P24821 | 9 | 115019575 | 115118257 | 1.05 | 0.90 | 1.23 | 5.38E-01 | 0.792 |
| OID20259 | XG | Cardiometabolic | XG | P55808 | X | 2752040 | 2816500 | 1.06 | 0.88 | 1.29 | 5.37E-01 | 0.792 |
| OID31113 | YAP1 | Neurology_II | YAP1 | P46937 | 11 | 102110447 | 102233424 | 0.95 | 0.79 | 1.13 | 5.38E-01 | 0.792 |
| OID31098 | HSD17B14 | Neurology_II | HSD17B14 | Q9BPX1 | 19 | 48813018 | 48836510 | 1.05 | 0.90 | 1.23 | 5.39E-01 | 0.792 |
| OID21009 | DDR1 | Neurology | DDR1 | Q08345 | 6 | 30876421 | 30900156 | 1.05 | 0.90 | 1.23 | 5.40E-01 | 0.793 |
| OID20990 | ASGR1 | Neurology | ASGR1 | P07306 | 17 | 7173431 | 7179564 | 0.95 | 0.80 | 1.13 | 5.40E-01 | 0.793 |
| OID30483 | PRSS22 | Inflammation_II | PRSS22 | Q9GZN4 | 16 | 2852730 | 2858170 | 1.05 | 0.90 | 1.21 | 5.41E-01 | 0.794 |
| OID20417 | TGFBI | Cardiometabolic | TGFBI | Q15582 | 5 | 136028988 | 136063818 | 0.95 | 0.80 | 1.12 | 5.43E-01 | 0.797 |
| OID30379 | COL3A1 | Cardiometabolic_II | COL3A1 | P02461 | 2 | 188974373 | 189012746 | 0.95 | 0.80 | 1.12 | 5.44E-01 | 0.797 |
| OID21368 | CCN1 | Oncology | CCN1 | O00622 | 1 | 85580761 | 85584589 | 0.95 | 0.81 | 1.12 | 5.44E-01 | 0.797 |
| OID20167 | IGSF8 | Cardiometabolic | IGSF8 | Q969P0 | 1 | 160091340 | 160098943 | 0.95 | 0.81 | 1.12 | 5.45E-01 | 0.797 |
| OID21248 | LHB | Oncology | LHB | P01229 | 19 | 49015980 | 49017091 | 0.95 | 0.80 | 1.13 | 5.45E-01 | 0.798 |
| OID30513 | ACRV1 | Inflammation_II | ACRV1 | P26436 | 11 | 125671522 | 125680874 | 0.92 | 0.70 | 1.21 | 5.47E-01 | 0.799 |
| OID20653 | TNFRSF4 | Inflammation | TNFRSF4 | P43489 | 1 | 1211340 | 1214153 | 1.05 | 0.90 | 1.21 | 5.47E-01 | 0.799 |
| OID20079 | ACTA2 | Cardiometabolic | ACTA2 | P62736 | 10 | 88935074 | 88991339 | 1.06 | 0.89 | 1.26 | 5.47E-01 | 0.800 |
| OID31000 | DNM3 | Neurology_II | DNM3 | Q9UQ16 | 1 | 171817887 | 172418466 | 0.95 | 0.80 | 1.12 | 5.48E-01 | 0.800 |
| OID20704 | CST7 | Inflammation | CST7 | O76096 | 20 | 24949269 | 24959928 | 0.95 | 0.79 | 1.14 | 5.49E-01 | 0.801 |
| OID30977 | GRIK2 | Neurology_II | GRIK2 | Q13002 | 6 | 100962701 | 102081622 | 0.95 | 0.81 | 1.12 | 5.49E-01 | 0.801 |
| OID21309 | GRPEL1 | Oncology | GRPEL1 | Q9HAV7 | 4 | 7058895 | 7068064 | 1.05 | 0.89 | 1.24 | 5.49E-01 | 0.801 |
| OID21145 | TNFRSF1B | Neurology | TNFRSF1B | P20333 | 1 | 12166991 | 12209228 | 1.05 | 0.90 | 1.22 | 5.49E-01 | 0.801 |
| OID20919 | ULBP2 | Neurology | ULBP2 | Q9BZM5 | 6 | 149942014 | 149949235 | 1.05 | 0.89 | 1.24 | 5.50E-01 | 0.801 |
| OID31488 | ADAM9 | Oncology_II | ADAM9 | Q13443 | 8 | 38996754 | 39105445 | 1.05 | 0.90 | 1.23 | 5.51E-01 | 0.801 |
| OID30203 | ANP32C | Cardiometabolic_II | ANP32C | O43423 | 4 | 164197007 | 16419771 | 0.94 | 0.78 | 1.14 | 5.51E-01 | 0.801 |
| OID20968 | FRZB | Neurology | FRZB | Q92765 | 2 | 182833275 | 182866637 | 0.95 | 0.79 | 1.13 | 5.51E-01 | 0.801 |
| OID20296 | GUSB | Cardiometabolic | GUSB | P08236 | 7 | 65960684 | 65982215 | 1.05 | 0.89 | 1.25 | 5.51E-01 | 0.801 |
| OID30238 | RNASE10 | Cardiometabolic_II | RNASE10 | Q5GAN6 | 14 | 20505537 | 20513884 | 1.05 | 0.89 | 1.24 | 5.51E-01 | 0.801 |
| OID20561 | CXADR | Inflammation | CXADR | P78310 | 21 | 17513043 | 17593579 | 1.05 | 0.90 | 1.21 | 5.53E-01 | 0.803 |
| OID31505 | IFI30 | Oncology_II | IFI30 | P13284 | 19 | 18173162 | 18178117 | 0.95 | 0.79 | 1.14 | 5.53E-01 | 0.803 |
| OID30828 | ART5 | Neurology_II | ART5 | Q96L15 | 11 | 3638512 | 3642316 | 1.05 | 0.90 | 1.22 | 5.54E-01 | 0.803 |
| OID21297 | COX5B | Oncology | COX5B | P10606 | 2 | 97646062 | 97648383 | 0.95 | 0.80 | 1.13 | 5.55E-01 | 0.803 |
| OID20261 | FUCA1 | Cardiometabolic | FUCA1 | P04066 | 1 | 23845077 | 23868290 | 0.95 | 0.81 | 1.12 | 5.55E-01 | 0.803 |
| OID20888 | KLB | Neurology | KLB | Q86Z14 | 4 | 39406930 | 39451533 | 0.95 | 0.81 | 1.12 | 5.56E-01 | 0.803 |
| OID30960 | LRP2 | Neurology_II | LRP2 | P98164 | 2 | 169127109 | 169362534 | 1.04 | 0.91 | 1.19 | 5.56E-01 | 0.803 |
| OID30098 | SHD | Cardiometabolic_II | SHD | Q96IW2 | 19 | 4279266 | 4290722 | 0.95 | 0.80 | 1.13 | 5.55E-01 | 0.803 |
| OID21148 | SPINK5 | Neurology | SPINK5 | Q9NQ38 | 5 | 148025683 | 148137382 | 1.05 | 0.89 | 1.23 | 5.55E-01 | 0.803 |
| OID31449 | TOP1 | Oncology_II | TOP1 | P11387 | 20 | 41028822 | 41124487 | 0.95 | 0.80 | 1.12 | 5.54E-01 | 0.803 |
| OID21336 | UBAC1 | Oncology | UBAC1 | Q9BSL1 | 9 | 135932969 | 135961373 | 0.95 | 0.80 | 1.13 | 5.56E-01 | 0.803 |
| OID30783 | TF | Inflammation_II | TF | P02787 | 3 | 133746040 | 133796641 | 0.95 | 0.81 | 1.12 | 5.56E-01 | 0.803 |
| OID21051 | BST1 | Neurology | BST1 | Q10588 | 4 | 15703065 | 15738313 | 0.95 | 0.82 | 1.12 | 5.57E-01 | 0.803 |
| OID31448 | INSL5 | Oncology_II | INSL5 | Q9Y5Q6 | 1 | 66797740 | 66801276 | 0.95 | 0.80 | 1.13 | 5.58E-01 | 0.803 |
| OID30855 | KLRC1 | Neurology_II | KLRC1 | P26715 | 12 | 10442264 | 10454685 | 0.95 | 0.79 | 1.13 | 5.57E-01 | 0.803 |
| OID21484 | L1CAM | Oncology | L1CAM | P32004 | X | 153861514 | 153886173 | 0.95 | 0.80 | 1.12 | 5.57E-01 | 0.803 |
| OID30489 | MRPS16 | Inflammation_II | MRPS16 | Q9Y3D3 | 10 | 73248843 | 73252693 | 0.94 | 0.78 | 1.14 | 5.58E-01 | 0.803 |
| OID20542 | SCRN1 | Inflammation | SCRN1 | Q12765 | 7 | 29920104 | 29990289 | 0.95 | 0.80 | 1.13 | 5.56E-01 | 0.803 |
| OID31079 | SEPTIN8 | Neurology_II | SEPTIN8 | Q92599 | 5 | 132750819 | 132807241 | 0.95 | 0.79 | 1.13 | 5.58E-01 | 0.803 |
| OID21398 | XCL1 | Oncology | XCL1 | P47992 | 1 | 168576605 | 168582069 | 0.95 | 0.80 | 1.13 | 5.58E-01 | 0.803 |
| OID21366 | CES2 | Oncology | CES2 | O00748 | 16 | 66934444 | 66945096 | 1.05 | 0.89 | 1.23 | 5.61E-01 | 0.806 |
| OID31232 | H2AP | Oncology_II | H2AP | O75409 | X | 37990779 | 37991314 | 1.05 | 0.90 | 1.22 | 5.62E-01 | 0.806 |
| OID31397 | IMMT | Oncology_II | IMMT | Q16891 | 2 | 86143932 | 86195472 | 0.95 | 0.80 | 1.13 | 5.62E-01 | 0.806 |
| OID31018 | LETM1 | Neurology_II | LETM1 | O95202 | 4 | 1811479 | 1856156 | 0.95 | 0.80 | 1.13 | 5.62E-01 | 0.806 |
| OID30917 | MYLPF | Neurology_II | MYL11 | Q96A32 | 16 | 30370934 | 30377991 | 1.05 | 0.89 | 1.25 | 5.61E-01 | 0.806 |
| OID20141 | TGM2 | Cardiometabolic | TGM2 | P21980 | 20 | 38127385 | 38166578 | 0.95 | 0.80 | 1.13 | 5.62E-01 | 0.806 |
| OID20875 | TDRKH | Neurology | TDRKH | Q9Y2W6 | 1 | 151770107 | 151791416 | 0.95 | 0.80 | 1.13 | 5.63E-01 | 0.807 |
| OID31262 | TXK | Oncology_II | TXK | P42681 | 4 | 48066393 | 48134250 | 0.95 | 0.78 | 1.14 | 5.63E-01 | 0.807 |
| OID20189 | ADGRG2 | Cardiometabolic | ADGRG2 | Q8IZP9 | X | 18989307 | 19122637 | 0.95 | 0.80 | 1.13 | 5.64E-01 | 0.807 |
| OID31450 | ZNRD2 | Oncology_II | ZNRD2 | O60232 | 11 | 65570460 | 65573942 | 0.95 | 0.80 | 1.13 | 5.64E-01 | 0.808 |
| OID20537 | SIGLEC10 | Inflammation | SIGLEC10 | Q96LC7 | 19 | 51410020 | 51417803 | 1.05 | 0.89 | 1.24 | 5.65E-01 | 0.809 |
| OID21376 | GPA33 | Oncology | GPA33 | Q99795 | 1 | 167052836 | 167166479 | 0.95 | 0.81 | 1.12 | 5.66E-01 | 0.810 |
| OID30421 | EDNRB | Inflammation_II | EDNRB | P24530 | 13 | 77895481 | 77975529 | 0.94 | 0.77 | 1.15 | 5.67E-01 | 0.811 |
| OID31239 | MRPL52 | Oncology_II | MRPL52 | Q86TS9 | 14 | 22829879 | 22835037 | 0.95 | 0.80 | 1.13 | 5.68E-01 | 0.811 |
| OID20222 | NECTIN2 | Cardiometabolic | NECTIN2 | Q92692 | 19 | 44846175 | 44889223 | 1.05 | 0.89 | 1.23 | 5.68E-01 | 0.811 |
| OID30312 | BCAT1 | Cardiometabolic_II | BCAT1 | P54687 | 12 | 24810024 | 24949101 | 1.05 | 0.89 | 1.24 | 5.69E-01 | 0.812 |
| OID30317 | COX6B1 | Cardiometabolic_II | COX6B1 | P14854 | 19 | 35648323 | 35658782 | 0.95 | 0.81 | 1.12 | 5.69E-01 | 0.813 |
| OID21058 | SCARA5 | Neurology | SCARA5 | Q6ZMJ2 | 8 | 27869883 | 27992673 | 0.95 | 0.81 | 1.13 | 5.71E-01 | 0.814 |
| OID20883 | BRK1 | Neurology | BRK1 | Q8WUW1 | 3 | 10115675 | 10127190 | 1.05 | 0.89 | 1.23 | 5.71E-01 | 0.814 |
| OID31514 | FAM3D | Oncology_II | FAM3D | Q96BQ1 | 3 | 58633946 | 58666834 | 1.05 | 0.89 | 1.23 | 5.71E-01 | 0.814 |
| OID21221 | PSMA1 | Oncology | PSMA1 | P25786 | 11 | 14504874 | 14643635 | 0.95 | 0.81 | 1.13 | 5.73E-01 | 0.816 |
| OID30085 | RYR1 | Cardiometabolic_II | RYR1 | P21817 | 19 | 38433691 | 38595273 | 1.04 | 0.91 | 1.18 | 5.73E-01 | 0.816 |
| OID21138 | ARSA | Neurology | ARSA | P15289 | 22 | 50622754 | 50628173 | 0.95 | 0.81 | 1.13 | 5.74E-01 | 0.817 |
| OID20356 | OSMR | Cardiometabolic | OSMR | Q99650 | 5 | 38845858 | 38945596 | 1.05 | 0.89 | 1.24 | 5.75E-01 | 0.817 |
| OID30676 | PI16 | Inflammation_II | PI16 | Q6UXB8 | 6 | 36948263 | 36964837 | 0.95 | 0.81 | 1.13 | 5.75E-01 | 0.817 |
| OID30146 | ABCA2 | Cardiometabolic_II | ABCA2 | Q9BZC7 | 9 | 137007234 | 137028915 | 1.05 | 0.89 | 1.22 | 5.77E-01 | 0.818 |
| OID30840 | CACNA1C | Neurology_II | CACNA1C | Q13936 | 12 | 1970772 | 2697950 | 0.95 | 0.80 | 1.13 | 5.76E-01 | 0.818 |
| OID30606 | CPA4 | Inflammation_II | CPA4 | Q9UI42 | 7 | 130293134 | 130324180 | 0.96 | 0.82 | 1.12 | 5.76E-01 | 0.818 |
| OID21064 | JAM2 | Neurology | JAM2 | P57087 | 21 | 25639258 | 25717562 | 0.96 | 0.82 | 1.12 | 5.76E-01 | 0.818 |

| | | | | | | | | | | | | |
|---|---|---|---|---|---|---|---|---|---|---|---|---|
| OID20196 | KITLG | Cardiometabolic | KITLG | P21583 | 12 | 88492793 | 88580851 | 0.96 | 0.82 | 1.12 | 5.77E-01 | 0.818 |
| OID30057 | LAMA1 | Cardiometabolic_II | LAMA1 | P25391 | 18 | 6941742 | 7117797 | 0.95 | 0.79 | 1.14 | 5.78E-01 | 0.818 |
| OID30102 | NUDT15 | Cardiometabolic_II | NUDT15 | Q9NV35 | 13 | 48037726 | 48047221 | 0.95 | 0.81 | 1.13 | 5.78E-01 | 0.818 |
| OID30423 | SPRED2 | Inflammation_II | SPRED2 | Q7Z698 | 2 | 65310851 | 65432637 | 0.95 | 0.81 | 1.13 | 5.76E-01 | 0.818 |
| OID30162 | LILRA4 | Cardiometabolic_II | LILRA4 | P59901 | 19 | 54333185 | 54339162 | 0.95 | 0.80 | 1.13 | 5.78E-01 | 0.819 |
| OID30429 | GLI2 | Inflammation_II | GLI2 | P10070 | 2 | 120735623 | 120992653 | 0.95 | 0.80 | 1.13 | 5.80E-01 | 0.819 |
| OID30461 | IL20RB | Inflammation_II | IL20RB | Q6UXL0 | 3 | 136946230 | 137011085 | 1.05 | 0.89 | 1.23 | 5.80E-01 | 0.819 |
| OID30101 | PGLYRP4 | Cardiometabolic_II | PGLYRP4 | Q96LB8 | 1 | 153330120 | 153348841 | 0.95 | 0.80 | 1.13 | 5.79E-01 | 0.819 |
| OID30854 | ELAVL4 | Neurology_II | ELAVL4 | P26378 | 1 | 50024029 | 50203772 | 0.95 | 0.81 | 1.13 | 5.80E-01 | 0.820 |
| OID20801 | CALB2 | Neurology | CALB2 | P22676 | 16 | 71358713 | 71390436 | 0.95 | 0.80 | 1.13 | 5.82E-01 | 0.820 |
| OID30454 | FGF12 | Inflammation_II | FGF12 | P61328 | 3 | 192139390 | 192767764 | 0.95 | 0.79 | 1.14 | 5.82E-01 | 0.820 |
| OID20727 | GAL | Inflammation | GAL | P22466 | 11 | 68683779 | 68691175 | 1.05 | 0.89 | 1.23 | 5.82E-01 | 0.820 |
| OID20488 | IL1A | Inflammation | IL1A | P01583 | 2 | 112773925 | 112784493 | 0.95 | 0.81 | 1.13 | 5.81E-01 | 0.820 |
| OID21123 | OGN | Neurology | OGN | P20774 | 9 | 92383268 | 92404696 | 1.05 | 0.89 | 1.24 | 5.82E-01 | 0.820 |
| OID30075 | REXO2 | Cardiometabolic_II | REXO2 | Q9Y3B8 | 11 | 114439435 | 114450279 | 0.95 | 0.81 | 1.13 | 5.82E-01 | 0.820 |
| OID30205 | SNED1 | Cardiometabolic_II | SNED1 | Q8TER0 | 2 | 240998618 | 241095568 | 1.04 | 0.89 | 1.22 | 5.84E-01 | 0.822 |
| OID21131 | PECAM1 | Neurology | PECAM1 | P16284 | 17 | 64319415 | 64413776 | 0.96 | 0.82 | 1.12 | 5.84E-01 | 0.822 |
| OID30801 | ZPR1 | Neurology_II | ZPR1 | O75312 | 11 | 116773799 | 116788039 | 0.96 | 0.81 | 1.12 | 5.85E-01 | 0.824 |
| OID30585 | ACHE | Inflammation_II | ACHE | P22303 | 7 | 100889994 | 100896974 | 1.05 | 0.89 | 1.24 | 5.87E-01 | 0.825 |
| OID20224 | ROR1 | Cardiometabolic | ROR1 | Q01973 | 1 | 63774017 | 64181498 | 1.05 | 0.89 | 1.23 | 5.87E-01 | 0.825 |
| OID21227 | NAMPT | Oncology | NAMPT | P43490 | 7 | 106248298 | 106285966 | 1.05 | 0.89 | 1.24 | 5.88E-01 | 0.826 |
| OID20145 | ICAM5 | Cardiometabolic | ICAM5 | Q9UMF0 | 19 | 10289952 | 10296778 | 1.05 | 0.88 | 1.25 | 5.89E-01 | 0.827 |
| OID30928 | TPBGL | Neurology_II | TPBGL | P0DKB5 | 11 | 75240774 | 75243704 | 0.96 | 0.81 | 1.13 | 5.89E-01 | 0.827 |
| OID21005 | CDH15 | Neurology | CDH15 | P55291 | 16 | 89171748 | 89195492 | 1.05 | 0.88 | 1.26 | 5.90E-01 | 0.827 |
| OID30886 | EPB41L5 | Neurology_II | EPB41L5 | Q9HCM4 | 2 | 120013077 | 120179119 | 1.04 | 0.89 | 1.23 | 5.91E-01 | 0.827 |
| OID20661 | FLT3LG | Inflammation | FLT3LG | P49771 | 19 | 49474207 | 49486231 | 0.95 | 0.80 | 1.13 | 5.91E-01 | 0.827 |
| OID21442 | FXN | Oncology | FXN | Q16595 | 9 | 69035751 | 69079076 | 0.96 | 0.81 | 1.13 | 5.91E-01 | 0.827 |
| OID30864 | LMOD2 | Neurology_II | LMOD2 | Q6P5Q4 | 7 | 123655866 | 123664290 | 0.95 | 0.80 | 1.14 | 5.91E-01 | 0.827 |
| OID31321 | RNF4 | Oncology_II | RNF4 | P78317 | 4 | 2462220 | 2515857 | 0.95 | 0.81 | 1.13 | 5.91E-01 | 0.827 |
| OID31051 | S100G | Neurology_II | S100G | P29377 | X | 16650158 | 16654670 | 0.96 | 0.82 | 1.12 | 5.91E-01 | 0.827 |
| OID30456 | CCNE1 | Inflammation_II | CCNE1 | P24864 | 19 | 29811991 | 29824312 | 1.04 | 0.89 | 1.22 | 5.92E-01 | 0.827 |
| OID21092 | CNTN3 | Neurology | CNTN3 | Q9P232 | 3 | 74262568 | 74614659 | 0.95 | 0.79 | 1.15 | 5.93E-01 | 0.827 |
| OID31048 | HLA-A | Neurology_II | HLA-A | P04439 | 6 | 29941260 | 29945884 | 0.96 | 0.81 | 1.12 | 5.94E-01 | 0.827 |
| OID21308 | NUDT2 | Oncology | NUDT2 | P50583 | 9 | 34329506 | 34343711 | 0.95 | 0.80 | 1.13 | 5.93E-01 | 0.827 |
| OID20682 | PKLR | Inflammation | PKLR | P30613 | 1 | 155289293 | 155301438 | 1.05 | 0.89 | 1.23 | 5.94E-01 | 0.827 |
| OID30473 | RNF31 | Inflammation_II | RNF31 | Q96EP0 | 14 | 24146683 | 24160660 | 0.95 | 0.79 | 1.14 | 5.93E-01 | 0.827 |
| OID30512 | TSPYL1 | Inflammation_II | TSPYL1 | Q9H0U9 | 6 | 116267760 | 116279930 | 0.95 | 0.81 | 1.13 | 5.93E-01 | 0.827 |
| OID20622 | CXCL17 | Inflammation | CXCL17 | Q6UXB2 | 19 | 42428278 | 42442946 | 0.95 | 0.77 | 1.16 | 5.94E-01 | 0.827 |
| OID21137 | HAVCR2 | Neurology | HAVCR2 | Q8TDQ0 | 5 | 157085832 | 157142869 | 1.05 | 0.89 | 1.23 | 5.95E-01 | 0.828 |
| OID20706 | NELL2 | Inflammation | NELL2 | Q99435 | 12 | 44508275 | 44921848 | 0.95 | 0.80 | 1.13 | 5.95E-01 | 0.828 |
| OID30776 | C3 | Inflammation_II | C3 | P01024 | 19 | 6677704 | 6730562 | 0.95 | 0.78 | 1.15 | 5.95E-01 | 0.828 |
| OID30116 | NPR1 | Cardiometabolic_II | NPR1 | P16066 | 1 | 153678688 | 153693992 | 1.05 | 0.88 | 1.25 | 5.96E-01 | 0.828 |
| OID30380 | PTPRC | Cardiometabolic_II | PTPRC | P08575 | 1 | 198638457 | 198757476 | 1.05 | 0.89 | 1.24 | 5.96E-01 | 0.828 |
| OID30129 | IFNW1 | Cardiometabolic_II | IFNW1 | P05000 | 9 | 21140632 | 21141832 | 0.95 | 0.79 | 1.14 | 5.97E-01 | 0.828 |
| OID20496 | SLAMF1 | Inflammation | SLAMF1 | Q13291 | 1 | 160608106 | 160647044 | 1.05 | 0.88 | 1.24 | 5.97E-01 | 0.828 |
| OID30799 | TUBB3 | Neurology_II | TUBB3 | Q13509 | 16 | 89921392 | 89938761 | 0.96 | 0.81 | 1.13 | 5.97E-01 | 0.829 |
| OID20630 | SERPINB8 | Inflammation | SERPINB8 | P50452 | 18 | 63970029 | 64019779 | 0.95 | 0.79 | 1.14 | 5.98E-01 | 0.829 |
| OID20142 | DPP7 | Cardiometabolic | DPP7 | Q9UHL4 | 9 | 137110546 | 137115177 | 0.96 | 0.81 | 1.13 | 5.99E-01 | 0.830 |
| OID21325 | DSG4 | Oncology | DSG4 | Q86SJ6 | 18 | 31376777 | 31414912 | 1.05 | 0.88 | 1.24 | 5.99E-01 | 0.830 |
| OID30947 | PPT1 | Neurology_II | PPT1 | P50897 | 1 | 40072710 | 40097260 | 0.95 | 0.80 | 1.14 | 5.99E-01 | 0.830 |
| OID21400 | SCG2 | Oncology | SCG2 | P13521 | 2 | 223596940 | 223602361 | 0.96 | 0.81 | 1.13 | 5.99E-01 | 0.830 |
| OID21455 | HDGF | Oncology | HDGF | P51858 | 1 | 156742109 | 156766925 | 0.96 | 0.81 | 1.13 | 6.00E-01 | 0.830 |
| OID20938 | RSPO1 | Neurology | RSPO1 | Q2MKA7 | 1 | 37611350 | 37634892 | 0.96 | 0.81 | 1.13 | 6.00E-01 | 0.830 |
| OID30588 | ADGRD1 | Inflammation_II | ADGRD1 | Q6QNK2 | 12 | 130953907 | 131141469 | 0.95 | 0.79 | 1.14 | 6.01E-01 | 0.831 |
| OID20772 | CCL24 | Inflammation | CCL24 | O00175 | 7 | 75810825 | 75823356 | 1.04 | 0.89 | 1.23 | 6.01E-01 | 0.831 |
| OID21485 | FOLR3 | Oncology | FOLR3 | P41439 | 11 | 72114869 | 72139892 | 1.05 | 0.89 | 1.23 | 6.03E-01 | 0.833 |
| OID31388 | SLC13A1 | Oncology_II | SLC13A1 | Q9BZW2 | 7 | 123113531 | 123199972 | 1.04 | 0.89 | 1.22 | 6.03E-01 | 0.833 |
| OID20263 | MB | Cardiometabolic | MB | P02144 | 22 | 35606764 | 35637951 | 0.95 | 0.79 | 1.15 | 6.05E-01 | 0.834 |
| OID30407 | NOTCH2 | Cardiometabolic_II | NOTCH2 | Q04721 | 1 | 119911553 | 120100779 | 0.96 | 0.80 | 1.14 | 6.05E-01 | 0.834 |
| OID20971 | SNCG | Neurology | SNCG | O76070 | 10 | 86958599 | 86963258 | 1.05 | 0.88 | 1.25 | 6.04E-01 | 0.834 |
| OID30701 | APOF | Inflammation_II | APOF | Q13790 | 12 | 56360568 | 56362857 | 1.06 | 0.85 | 1.31 | 6.06E-01 | 0.834 |
| OID20227 | FBP1 | Cardiometabolic | FBP1 | P09467 | 9 | 94603133 | 94640249 | 0.96 | 0.80 | 1.14 | 6.06E-01 | 0.834 |
| OID21207 | FES | Oncology | FES | P07332 | 15 | 90883695 | 90895776 | 0.96 | 0.81 | 1.13 | 6.05E-01 | 0.834 |
| OID20920 | NGF | Neurology | NGF | P01138 | 1 | 115285904 | 115338770 | 1.04 | 0.90 | 1.19 | 6.07E-01 | 0.836 |
| OID30457 | PRR5 | Inflammation_II | PRR5 | P85299 | 22 | 44668547 | 44737681 | 0.95 | 0.79 | 1.15 | 6.08E-01 | 0.836 |
| OID20752 | CTRC | Inflammation | CTRC | Q99895 | 1 | 15438442 | 15449242 | 0.96 | 0.82 | 1.13 | 6.08E-01 | 0.836 |
| OID20229 | NRCAM | Cardiometabolic | NRCAM | Q92823 | 7 | 108147623 | 108456717 | 0.96 | 0.83 | 1.12 | 6.08E-01 | 0.836 |
| OID21055 | NCAN | Neurology | NCAN | O14594 | 19 | 19211958 | 19252233 | 1.05 | 0.88 | 1.25 | 6.09E-01 | 0.836 |
| OID20074 | TNF | Cardiometabolic/Inflam | TNF | P01375 | 6 | 31575565 | 31578336 | 0.96 | 0.80 | 1.14 | 6.11E-01 | 0.838 |
| OID31277 | ACADM | Oncology_II | ACADM | P11310 | 1 | 75724431 | 75787575 | 1.04 | 0.90 | 1.19 | 6.11E-01 | 0.838 |
| OID20350 | ANPEP | Cardiometabolic | ANPEP | P15144 | 15 | 89784895 | 89815401 | 1.04 | 0.89 | 1.22 | 6.11E-01 | 0.838 |
| OID30063 | CASQ2 | Cardiometabolic_II | CASQ2 | O14958 | 1 | 115700021 | 115768714 | 1.04 | 0.90 | 1.20 | 6.12E-01 | 0.838 |
| OID20609 | CLEC4D | Inflammation | CLEC4D | Q8WXI8 | 12 | 8509475 | 8522366 | 1.04 | 0.88 | 1.24 | 6.12E-01 | 0.838 |
| OID31285 | GAS2 | Oncology_II | GAS2 | O43903 | 11 | 22625509 | 22813001 | 0.95 | 0.79 | 1.15 | 6.12E-01 | 0.838 |
| OID20984 | SFRP1 | Neurology | SFRP1 | Q8N474 | 8 | 41261962 | 41309473 | 1.04 | 0.88 | 1.24 | 6.12E-01 | 0.838 |
| OID20284 | LGALS3 | Cardiometabolic | LGALS3 | P17931 | 14 | 55124110 | 55145423 | 1.04 | 0.88 | 1.24 | 6.12E-01 | 0.838 |
| OID20686 | CCL21 | Inflammation | CCL21 | O00585 | 9 | 34709005 | 34710136 | 1.04 | 0.89 | 1.22 | 6.13E-01 | 0.838 |
| OID31189 | CDC25A | Oncology_II | CDC25A | P30304 | 3 | 48157146 | 48188417 | 1.04 | 0.89 | 1.22 | 6.15E-01 | 0.840 |
| OID20050 | TNNI3 | Cardiometabolic | TNNI3 | P19429 | 19 | 55151767 | 55157773 | 1.04 | 0.88 | 1.23 | 6.15E-01 | 0.840 |
| OID21443 | PRTG | Oncology | PRTG | Q2VWP7 | 15 | 55611544 | 55743152 | 1.04 | 0.88 | 1.23 | 6.16E-01 | 0.841 |
| OID30399 | CILP | Cardiometabolic_II | CILP | O75339 | 15 | 65194760 | 65211473 | 0.96 | 0.80 | 1.14 | 6.16E-01 | 0.841 |
| OID20437 | AMBN | Inflammation | AMBN | Q9NP70 | 4 | 70592256 | 70607288 | 1.04 | 0.89 | 1.21 | 6.18E-01 | 0.842 |
| OID20679 | CTSC | Inflammation | CTSC | P53634 | 11 | 88265069 | 88359684 | 1.04 | 0.88 | 1.23 | 6.17E-01 | 0.842 |
| OID30159 | MMUT | Cardiometabolic_II | MMUT | P22033 | 6 | 49430360 | 49463253 | 0.96 | 0.81 | 1.13 | 6.19E-01 | 0.843 |
| OID20197 | PILRB | Cardiometabolic | PILRB | Q9UKJ0 | 7 | 100352176 | 100367831 | 0.96 | 0.82 | 1.12 | 6.19E-01 | 0.844 |
| OID21403 | TRIAP1 | Oncology | TRIAP1 | O43715 | 12 | 120443964 | 120446384 | 0.96 | 0.81 | 1.14 | 6.20E-01 | 0.844 |
| OID30587 | COL5A1 | Inflammation_II | COL5A1 | P20908 | 9 | 134641803 | 134844843 | 0.96 | 0.81 | 1.13 | 6.21E-01 | 0.846 |
| OID20306 | PTPRS | Cardiometabolic | PTPRS | Q13332 | 19 | 5158495 | 5340812 | 0.96 | 0.82 | 1.12 | 6.22E-01 | 0.847 |
| OID21030 | CCL19 | Neurology | CCL19 | Q99731 | 9 | 34689570 | 34691276 | 1.03 | 0.90 | 1.18 | 6.24E-01 | 0.847 |
| OID20280 | CPA1 | Cardiometabolic | CPA1 | P15085 | 7 | 130380339 | 130388114 | 0.96 | 0.82 | 1.12 | 6.23E-01 | 0.847 |
| OID20411 | ICAM1 | Cardiometabolic | ICAM1 | P05362 | 19 | 10271093 | 10286615 | 1.04 | 0.89 | 1.22 | 6.24E-01 | 0.847 |
| OID20430 | IL13 | Inflammation | IL13 | P35225 | 5 | 132656263 | 132661110 | 0.96 | 0.84 | 1.11 | 6.23E-01 | 0.847 |
| OID21045 | RGMB | Neurology | RGMB | Q6NW40 | 5 | 98768650 | 98798643 | 0.96 | 0.82 | 1.12 | 6.24E-01 | 0.847 |
| OID20645 | ADA | Inflammation | ADA | P00813 | 20 | 44619522 | 44652233 | 0.96 | 0.81 | 1.14 | 6.26E-01 | 0.848 |
| OID31349 | ENOPH1 | Oncology_II | ENOPH1 | Q9UHY7 | 4 | 82430590 | 82461177 | 0.96 | 0.81 | 1.13 | 6.27E-01 | 0.848 |
| OID30622 | EPHA4 | Inflammation_II | EPHA4 | P54764 | 2 | 221418027 | 221574202 | 0.96 | 0.82 | 1.13 | 6.27E-01 | 0.848 |
| OID30245 | MYL4 | Cardiometabolic_II | MYL4 | P12829 | 17 | 47200446 | 47223679 | 0.96 | 0.80 | 1.15 | 6.26E-01 | 0.848 |
| OID30943 | PMS1 | Neurology_II | PMS1 | P54277 | 2 | 189784085 | 189877629 | 0.96 | 0.80 | 1.14 | 6.27E-01 | 0.848 |
| OID30564 | PPL | Inflammation_II | PPL | O60437 | 16 | 4882507 | 4960741 | 1.04 | 0.88 | 1.23 | 6.25E-01 | 0.848 |
| OID31131 | SPRR3 | Neurology_II | SPRR3 | Q9UBC9 | 1 | 153001747 | 153003856 | 1.04 | 0.89 | 1.22 | 6.26E-01 | 0.848 |
| OID30095 | TSPAN15 | Cardiometabolic_II | TSPAN15 | O95858 | 10 | 69451465 | 69507666 | 1.04 | 0.89 | 1.21 | 6.27E-01 | 0.848 |
| OID30868 | MAP1LC3A | Neurology_II | MAP1LC3A | Q9H492 | 20 | 34546854 | 34560345 | 0.96 | 0.82 | 1.13 | 6.28E-01 | 0.848 |
| OID21049 | TNFRSF8 | Neurology | TNFRSF8 | P28908 | 1 | 12063303 | 12144207 | 1.04 | 0.89 | 1.21 | 6.29E-01 | 0.850 |

| | | | | | | | | | | | | |
|---|---|---|---|---|---|---|---|---|---|---|---|---|
| OID21417 | CA9 | Oncology | CA9 | Q16790 | 9 | 35673928 | 35681159 | 1.04 | 0.89 | 1.22 | 6.30E-01 | 0.851 |
| OID30142 | ECI2 | Cardiometabolic_II | ECI2 | O75521 | 6 | 4115693 | 4135597 | 0.96 | 0.81 | 1.13 | 6.31E-01 | 0.851 |
| OID20274 | PLA2G2A | Cardiometabolic | PLA2G2A | P14555 | 1 | 19975431 | 19980416 | 1.04 | 0.89 | 1.22 | 6.30E-01 | 0.851 |
| OID30955 | AKR1B10 | Neurology_II | AKR1B10 | O60218 | 7 | 134527567 | 134541412 | 0.96 | 0.82 | 1.13 | 6.33E-01 | 0.851 |
| OID30721 | C1RL | Inflammation_II | C1RL | Q9NZP8 | 12 | 7089587 | 7109238 | 1.04 | 0.87 | 1.25 | 6.34E-01 | 0.851 |
| OID21108 | CD109 | Neurology | CD109 | Q6YHK3 | 6 | 73695785 | 73828316 | 0.96 | 0.81 | 1.14 | 6.34E-01 | 0.851 |
| OID30291 | CD80 | Cardiometabolic_II | CD80 | P33681 | 3 | 119524293 | 119559614 | 1.04 | 0.89 | 1.22 | 6.33E-01 | 0.851 |
| OID20104 | ENTPD5 | Cardiometabolic | ENTPD5 | O75356 | 14 | 73958010 | 74019399 | 1.04 | 0.88 | 1.23 | 6.33E-01 | 0.851 |
| OID30367 | LCP1 | Cardiometabolic_II | LCP1 | P13796 | 13 | 46125920 | 46211871 | 0.96 | 0.81 | 1.14 | 6.32E-01 | 0.851 |
| OID21293 | LRP1 | Oncology | LRP1 | Q07954 | 12 | 57128483 | 57213361 | 1.04 | 0.88 | 1.23 | 6.34E-01 | 0.851 |
| OID30299 | MMP15 | Cardiometabolic_II | MMP15 | P51511 | 16 | 58025754 | 58046901 | 0.96 | 0.81 | 1.14 | 6.35E-01 | 0.851 |
| OID31066 | NEO1 | Neurology_II | NEO1 | Q92859 | 15 | 73051710 | 73305205 | 1.03 | 0.90 | 1.19 | 6.35E-01 | 0.851 |
| OID31101 | PLXDC2 | Neurology_II | PLXDC2 | Q6UX71 | 10 | 19816239 | 20289856 | 1.03 | 0.90 | 1.19 | 6.34E-01 | 0.851 |
| OID21516 | PPY | Oncology | PPY | P01298 | 17 | 43940804 | 43942476 | 1.04 | 0.89 | 1.22 | 6.34E-01 | 0.851 |
| OID30984 | RBFOX3 | Neurology_II | RBFOX3 | A6NFN3 | 17 | 79089345 | 79611051 | 0.96 | 0.81 | 1.14 | 6.34E-01 | 0.851 |
| OID30752 | SELL | Inflammation_II | SELL | P14151 | 1 | 169690665 | 169711702 | 0.96 | 0.81 | 1.14 | 6.35E-01 | 0.851 |
| OID30261 | SIGLEC8 | Cardiometabolic_II | SIGLEC8 | Q9NYZ4 | 19 | 51450847 | 51458456 | 1.04 | 0.88 | 1.23 | 6.33E-01 | 0.851 |
| OID20981 | TNFRSF10B | Neurology | TNFRSF10B | O14763 | 8 | 23020133 | 23069031 | 1.03 | 0.90 | 1.18 | 6.32E-01 | 0.851 |
| OID20489 | GBP2 | Inflammation | GBP2 | P32456 | 1 | 89106132 | 89150456 | 0.96 | 0.81 | 1.14 | 6.36E-01 | 0.851 |
| OID30863 | SH3BGRL2 | Neurology_II | SH3BGRL2 | Q9UJC5 | 6 | 79631329 | 79703655 | 0.96 | 0.82 | 1.13 | 6.36E-01 | 0.851 |
| OID20237 | UMOD | Cardiometabolic | UMOD | P07911 | 16 | 20333051 | 20356301 | 1.04 | 0.88 | 1.24 | 6.37E-01 | 0.852 |
| OID30410 | POSTN | Cardiometabolic_II | POSTN | Q15063 | 13 | 37562583 | 37598844 | 1.04 | 0.88 | 1.23 | 6.38E-01 | 0.854 |
| OID30453 | RPA2 | Inflammation_II | RPA2 | P15927 | 1 | 27891524 | 27914746 | 0.96 | 0.81 | 1.14 | 6.39E-01 | 0.854 |
| OID30171 | UHRF2 | Cardiometabolic_II | UHRF2 | Q96PU4 | 9 | 6413151 | 6507054 | 1.04 | 0.87 | 1.25 | 6.39E-01 | 0.854 |
| OID21522 | ITGB5 | Oncology | ITGB5 | P18084 | 3 | 124761948 | 124901418 | 0.96 | 0.81 | 1.14 | 6.40E-01 | 0.854 |
| OID31202 | BEX3 | Oncology_II | BEX3 | Q00994 | X | 103376395 | 103378164 | 0.96 | 0.80 | 1.15 | 6.41E-01 | 0.856 |
| OID30383 | PRG3 | Cardiometabolic_II | PRG3 | Q9Y2Y8 | 11 | 57376769 | 57381150 | 0.96 | 0.81 | 1.14 | 6.41E-01 | 0.856 |
| OID20670 | LY9 | Inflammation | LY9 | Q9HBG7 | 1 | 160796074 | 160828261 | 1.04 | 0.89 | 1.21 | 6.42E-01 | 0.856 |
| OID30747 | LPA | Inflammation_II | LPA | P08519 | 6 | 160531482 | 160664275 | 0.96 | 0.83 | 1.13 | 6.43E-01 | 0.857 |
| OID20483 | PRDX3 | Inflammation | PRDX3 | P30048 | 10 | 119167720 | 119178812 | 0.96 | 0.82 | 1.13 | 6.43E-01 | 0.857 |
| OID30495 | CPOX | Inflammation_II | CPOX | P36551 | 3 | 98579446 | 98593648 | 0.96 | 0.83 | 1.12 | 6.44E-01 | 0.857 |
| OID20460 | WNT9A | Inflammation | WNT9A | O14904 | 1 | 227918656 | 227947932 | 1.04 | 0.87 | 1.24 | 6.44E-01 | 0.858 |
| OID21095 | NCAM2 | Neurology | NCAM2 | O15394 | 21 | 20998409 | 21543329 | 1.04 | 0.88 | 1.23 | 6.45E-01 | 0.858 |
| OID20546 | CCL26 | Inflammation | CCL26 | Q9Y258 | 7 | 75769533 | 75789896 | 1.03 | 0.89 | 1.20 | 6.45E-01 | 0.858 |
| OID20318 | GAS6 | Cardiometabolic | GAS6 | Q14393 | 13 | 113820549 | 113864076 | 1.04 | 0.88 | 1.22 | 6.46E-01 | 0.858 |
| OID20999 | ACVRL1 | Neurology | ACVRL1 | P37023 | 12 | 51906908 | 51923361 | 0.96 | 0.81 | 1.14 | 6.47E-01 | 0.859 |
| OID20390 | NRP1 | Cardiometabolic | NRP1 | O14786 | 10 | 33177492 | 33336262 | 1.04 | 0.89 | 1.21 | 6.47E-01 | 0.859 |
| OID21362 | OPTC | Oncology | OPTC | Q9UBM4 | 1 | 203494153 | 203508949 | 1.04 | 0.89 | 1.21 | 6.47E-01 | 0.859 |
| OID20664 | CLSTN2 | Inflammation | CLSTN2 | Q9H4D0 | 3 | 139935185 | 140577397 | 1.04 | 0.88 | 1.23 | 6.48E-01 | 0.860 |
| OID20171 | MCFD2 | Cardiometabolic | MCFD2 | Q8NI22 | 2 | 46901870 | 46941855 | 0.96 | 0.81 | 1.14 | 6.48E-01 | 0.860 |
| OID20327 | RETN | Cardiometabolic | RETN | Q9HD89 | 19 | 7669049 | 7670455 | 1.04 | 0.88 | 1.23 | 6.48E-01 | 0.860 |
| OID30486 | SUMF1 | Inflammation_II | SUMF1 | Q8NBK3 | 3 | 3700814 | 4467273 | 1.04 | 0.88 | 1.24 | 6.49E-01 | 0.861 |
| OID21003 | LXN | Neurology | LXN | Q9BS40 | 3 | 158645822 | 158672648 | 1.04 | 0.88 | 1.23 | 6.51E-01 | 0.863 |
| OID20567 | NCF2 | Inflammation | NCF2 | P19878 | 1 | 183555562 | 183590876 | 0.96 | 0.81 | 1.14 | 6.52E-01 | 0.863 |
| OID30683 | SOD3 | Inflammation_II | SOD3 | P08294 | 4 | 24789912 | 24800842 | 0.95 | 0.78 | 1.17 | 6.53E-01 | 0.864 |
| OID30823 | DEFB116 | Neurology_II | DEFB116 | Q30KQ4 | 20 | 31303212 | 31308585 | 0.96 | 0.82 | 1.13 | 6.54E-01 | 0.865 |
| OID21397 | EPS8L2 | Oncology | EPS8L2 | Q9H6S3 | 11 | 694438 | 727727 | 0.96 | 0.81 | 1.14 | 6.54E-01 | 0.865 |
| OID30532 | TLR1 | Inflammation_II | TLR1 | Q15399 | 4 | 38790677 | 38856817 | 1.04 | 0.88 | 1.22 | 6.54E-01 | 0.865 |
| OID20979 | MATN3 | Neurology | MATN3 | O15232 | 2 | 19992052 | 20012668 | 1.04 | 0.88 | 1.22 | 6.55E-01 | 0.865 |
| OID30679 | ASGR2 | Inflammation_II | ASGR2 | P07307 | 17 | 7101322 | 7115700 | 0.96 | 0.82 | 1.14 | 6.55E-01 | 0.866 |
| OID21039 | ADAM8 | Neurology | ADAM8 | P78325 | 10 | 133262420 | 133276868 | 0.96 | 0.82 | 1.13 | 6.56E-01 | 0.866 |
| OID30744 | C5 | Inflammation_II | C5 | P01031 | 9 | 120932987 | 121075195 | 1.04 | 0.87 | 1.25 | 6.57E-01 | 0.866 |
| OID20360 | CD163 | Cardiometabolic | CD163 | Q86VB7 | 12 | 7470811 | 7503893 | 1.04 | 0.88 | 1.22 | 6.57E-01 | 0.866 |
| OID31309 | EVPL | Oncology_II | EVPL | Q92817 | 17 | 76004502 | 76027306 | 1.04 | 0.88 | 1.23 | 6.57E-01 | 0.866 |
| OID20807 | MMP13 | Neurology | MMP13 | P45452 | 11 | 102942995 | 102955732 | 0.96 | 0.82 | 1.13 | 6.57E-01 | 0.866 |
| OID21156 | CLPS | Neurology | CLPS | P04118 | 6 | 35794982 | 35797344 | 0.97 | 0.82 | 1.13 | 6.58E-01 | 0.867 |
| OID31007 | C1QBP | Neurology_II | C1QBP | Q07021 | 17 | 5432777 | 5448830 | 0.96 | 0.81 | 1.14 | 6.60E-01 | 0.867 |
| OID20175 | GHRL | Cardiometabolic | GHRL | Q9UBU3 | 3 | 10285666 | 10292947 | 0.96 | 0.82 | 1.14 | 6.60E-01 | 0.867 |
| OID30460 | PGR | Inflammation_II | PGR | P06401 | 11 | 101029624 | 101129813 | 1.04 | 0.89 | 1.21 | 6.60E-01 | 0.867 |
| OID30963 | PTH | Neurology_II | PTH | P01270 | 11 | 13492054 | 13496181 | 1.04 | 0.88 | 1.21 | 6.60E-01 | 0.867 |
| OID20810 | ANXA5 | Neurology | ANXA5 | P08758 | 4 | 121667946 | 121696995 | 1.04 | 0.88 | 1.21 | 6.62E-01 | 0.868 |
| OID31247 | CEP350 | Oncology_II | CEP350 | Q5VT06 | 1 | 179954674 | 180114880 | 1.04 | 0.88 | 1.22 | 6.62E-01 | 0.868 |
| OID30819 | GPR101 | Neurology_II | GPR101 | Q96P66 | X | 137023929 | 137033995 | 0.96 | 0.81 | 1.14 | 6.61E-01 | 0.868 |
| OID21429 | LRIG1 | Oncology | LRIG1 | Q96JA1 | 3 | 66378797 | 66501263 | 1.04 | 0.88 | 1.21 | 6.62E-01 | 0.868 |
| OID21164 | VTCN1 | Oncology | VTCN1 | Q7Z7D3 | 1 | 117143587 | 117210960 | 0.96 | 0.82 | 1.13 | 6.61E-01 | 0.868 |
| OID30426 | KDM3A | Inflammation_II | KDM3A | Q9Y4C1 | 2 | 86440647 | 86492716 | 0.96 | 0.81 | 1.15 | 6.64E-01 | 0.869 |
| OID20945 | LY96 | Neurology | LY96 | Q9Y6Y9 | 8 | 73991392 | 74029079 | 1.04 | 0.88 | 1.21 | 6.63E-01 | 0.869 |
| OID30282 | POMC | Cardiometabolic_II | POMC | P01189 | 2 | 25160853 | 25168903 | 1.04 | 0.87 | 1.24 | 6.63E-01 | 0.869 |
| OID31441 | SLMAP | Oncology_II | SLMAP | Q14BN4 | 3 | 57755450 | 57930003 | 0.96 | 0.82 | 1.14 | 6.64E-01 | 0.869 |
| OID30352 | UPB1 | Cardiometabolic_II | UPB1 | Q9UBR1 | 22 | 24494107 | 24528390 | 0.96 | 0.81 | 1.15 | 6.63E-01 | 0.869 |
| OID20949 | GLB1 | Neurology | GLB1 | P16278 | 3 | 32996609 | 33097202 | 0.96 | 0.81 | 1.15 | 6.64E-01 | 0.869 |
| OID30759 | MBL2 | Inflammation_II | MBL2 | P11226 | 10 | 52765380 | 52772784 | 1.04 | 0.88 | 1.22 | 6.65E-01 | 0.869 |
| OID30593 | CELSR2 | Inflammation_II | CELSR2 | Q9HCU4 | 1 | 109249539 | 109275751 | 1.04 | 0.88 | 1.21 | 6.66E-01 | 0.870 |
| OID20286 | CORO1A | Cardiometabolic | CORO1A | P31146 | 16 | 30182827 | 30189076 | 0.96 | 0.82 | 1.14 | 6.67E-01 | 0.870 |
| OID20344 | DEFA1_DEFA1B | Cardiometabolic | DEFA1;DEFA1B | P59665;P59665 | 8;8 | 6977649;6996766 | 6980092;6999198 | 1.04 | 0.88 | 1.23 | 6.67E-01 | 0.870 |
| OID30694 | MRC1 | Inflammation_II | MRC1 | P22897 | 10 | 17809348 | 17911164 | 1.04 | 0.88 | 1.23 | 6.66E-01 | 0.870 |
| OID31311 | PKN3 | Oncology_II | PKN3 | Q6P5Z2 | 9 | 128702503 | 128720916 | 0.96 | 0.80 | 1.15 | 6.66E-01 | 0.870 |
| OID31230 | ATP1B4 | Oncology_II | ATP1B4 | Q9UN42 | X | 120362085 | 120383249 | 1.03 | 0.89 | 1.20 | 6.69E-01 | 0.870 |
| OID21306 | CALB1 | Oncology | CALB1 | P05937 | 8 | 90058608 | 90095475 | 1.04 | 0.88 | 1.23 | 6.69E-01 | 0.870 |
| OID20675 | CXCL9 | Inflammation | CXCL9 | Q07325 | 4 | 76001275 | 76007509 | 1.04 | 0.87 | 1.24 | 6.70E-01 | 0.870 |
| OID20879 | DCTN6 | Neurology | DCTN6 | O00399 | 8 | 30156319 | 30183639 | 0.96 | 0.82 | 1.14 | 6.68E-01 | 0.870 |
| OID20424 | JUN | Inflammation | JUN | P05412 | 1 | 58776845 | 58784048 | 1.03 | 0.88 | 1.21 | 6.69E-01 | 0.870 |
| OID20355 | LBP | Cardiometabolic | LBP | P18428 | 20 | 38346482 | 38377013 | 0.96 | 0.81 | 1.14 | 6.69E-01 | 0.870 |
| OID31356 | RCC1 | Oncology_II | RCC1 | P18754 | 1 | 28505943 | 28539300 | 1.03 | 0.89 | 1.21 | 6.69E-01 | 0.870 |
| OID21326 | STX4 | Oncology | STX4 | Q12846 | 16 | 31032889 | 31042975 | 0.96 | 0.80 | 1.15 | 6.69E-01 | 0.870 |
| OID31289 | TAP1 | Oncology_II | TAP1 | Q03518 | 6 | 32845209 | 32853816 | 0.96 | 0.81 | 1.15 | 6.69E-01 | 0.870 |
| OID21069 | CGA | Neurology | CGA | P01215 | 6 | 87085498 | 87095106 | 1.06 | 0.81 | 1.39 | 6.70E-01 | 0.871 |
| OID31516 | PCDH9 | Oncology_II | PCDH9 | Q9HC56 | 13 | 66302834 | 67230445 | 0.96 | 0.81 | 1.14 | 6.71E-01 | 0.871 |
| OID30650 | SUSD5 | Inflammation_II | SUSD5 | O60279 | 3 | 33150043 | 33218810 | 1.04 | 0.88 | 1.23 | 6.71E-01 | 0.871 |
| OID30412 | TNN | Cardiometabolic_II | TNN | Q9UQP3 | 1 | 175067833 | 175148075 | 1.04 | 0.88 | 1.22 | 6.71E-01 | 0.871 |
| OID31069 | OCLN | Neurology_II | OCLN | Q16625 | 5 | 69492292 | 69558104 | 0.96 | 0.82 | 1.14 | 6.72E-01 | 0.871 |
| OID30867 | RRP15 | Neurology_II | RRP15 | Q9Y3B9 | 1 | 218285293 | 218337983 | 1.04 | 0.88 | 1.22 | 6.72E-01 | 0.871 |
| OID30765 | LGALS3BP | Inflammation_II | LGALS3BP | Q08380 | 17 | 78971238 | 78979947 | 0.96 | 0.81 | 1.15 | 6.73E-01 | 0.872 |
| OID31036 | MRPL58 | Neurology_II | MRPL58 | Q14197 | 17 | 75012670 | 75021261 | 1.03 | 0.89 | 1.20 | 6.73E-01 | 0.872 |
| OID20161 | TYRO3 | Cardiometabolic | TYRO3 | Q06418 | 15 | 41557675 | 41583589 | 0.97 | 0.82 | 1.13 | 6.73E-01 | 0.872 |
| OID20931 | NAAA | Neurology | NAAA | Q02083 | 4 | 75913660 | 75941013 | 0.97 | 0.82 | 1.14 | 6.74E-01 | 0.873 |
| OID20799 | SLC16A1 | Neurology | SLC16A1 | P53985 | 1 | 112911847 | 112957593 | 0.97 | 0.82 | 1.14 | 6.75E-01 | 0.873 |
| OID30177 | CBX2 | Cardiometabolic_II | CBX2 | Q14781 | 17 | 79778148 | 79787983 | 1.03 | 0.89 | 1.20 | 6.76E-01 | 0.873 |
| OID30758 | CFHR4 | Inflammation_II | CFHR4 | Q92496 | 1 | 196888014 | 196918795 | 0.97 | 0.82 | 1.14 | 6.77E-01 | 0.873 |
| OID31373 | EPGN | Oncology_II | EPGN | Q6UW88 | 4 | 74308470 | 74316789 | 1.03 | 0.89 | 1.21 | 6.76E-01 | 0.873 |
| OID20349 | F9 | Cardiometabolic | F9 | P00740 | X | 139530739 | 139563459 | 1.04 | 0.87 | 1.23 | 6.76E-01 | 0.873 |
| OID21404 | PRDX6 | Oncology | PRDX6 | P30041 | 1 | 173477330 | 173488815 | 1.03 | 0.88 | 1.21 | 6.77E-01 | 0.873 |
| OID20763 | PRSS8 | Inflammation | PRSS8 | Q16651 | 16 | 31131433 | 31135727 | 1.04 | 0.87 | 1.25 | 6.77E-01 | 0.873 |

| | | | | | | | | | | | | |
|---|---|---|---|---|---|---|---|---|---|---|---|---|
| OID30517 | TYRP1 | Inflammation_II | TYRP1 | P17643 | 9 | 12685439 | 12710285 | 0.97 | 0.83 | 1.13 | 6.77E-01 | 0.873 |
| OID30929 | CLNS1A | Neurology_II | CLNS1A | P54105 | 11 | 77514936 | 77637794 | 0.96 | 0.81 | 1.15 | 6.78E-01 | 0.873 |
| OID20732 | MZB1 | Inflammation | MZB1 | Q8WU39 | 5 | 139387467 | 139390081 | 1.04 | 0.88 | 1.22 | 6.78E-01 | 0.873 |
| OID20556 | CNTNAP2 | Inflammation | CNTNAP2 | Q9UHC6 | 7 | 146116002 | 148420998 | 0.97 | 0.82 | 1.14 | 6.80E-01 | 0.875 |
| OID30625 | INHBB | Inflammation_II | INHBB | P09529 | 2 | 120346136 | 120351803 | 1.03 | 0.88 | 1.22 | 6.81E-01 | 0.876 |
| OID20957 | TNR | Neurology | TNR | Q92752 | 1 | 175315194 | 175743616 | 1.04 | 0.88 | 1.22 | 6.80E-01 | 0.876 |
| OID20292 | COL6A3 | Cardiometabolic | COL6A3 | P12111 | 2 | 237324003 | 237414328 | 1.03 | 0.88 | 1.22 | 6.82E-01 | 0.876 |
| OID30434 | ESR1 | Inflammation_II | ESR1 | P03372 | 6 | 151656691 | 152129619 | 1.04 | 0.88 | 1.22 | 6.82E-01 | 0.876 |
| OID20269 | MET | Cardiometabolic | MET | P08581 | 7 | 116672196 | 116798377 | 1.04 | 0.87 | 1.23 | 6.82E-01 | 0.876 |
| OID20271 | CA3 | Cardiometabolic | CA3 | P07451 | 8 | 85373436 | 85449040 | 0.96 | 0.81 | 1.15 | 6.83E-01 | 0.877 |
| OID21527 | CD27 | Oncology | CD27 | P26842 | 12 | 6444955 | 6451718 | 0.97 | 0.82 | 1.14 | 6.83E-01 | 0.877 |
| OID30497 | ALPI | Inflammation_II | ALPI | P09923 | 2 | 232456125 | 232460753 | 0.97 | 0.83 | 1.13 | 6.84E-01 | 0.877 |
| OID21438 | GPC1 | Oncology | GPC1 | P35052 | 2 | 240435663 | 240468076 | 1.03 | 0.88 | 1.22 | 6.84E-01 | 0.877 |
| OID31157 | CTRL | Neurology_II | CTRL | P40313 | 16 | 67927640 | 67932365 | 1.03 | 0.88 | 1.21 | 6.85E-01 | 0.878 |
| OID30470 | TP53BP1 | Inflammation_II | TP53BP1 | Q12888 | 15 | 43403061 | 43510728 | 1.04 | 0.88 | 1.22 | 6.85E-01 | 0.878 |
| OID20737 | LAIR1 | Inflammation | LAIR1 | Q6GTX8 | 19 | 54351384 | 54370558 | 1.04 | 0.87 | 1.24 | 6.85E-01 | 0.878 |
| OID30099 | MEGF11 | Cardiometabolic_II | MEGF11 | A6BM72 | 15 | 65895079 | 66253756 | 0.97 | 0.82 | 1.14 | 6.86E-01 | 0.878 |
| OID30070 | RPS10 | Cardiometabolic_II | RPS10 | P46783 | 6 | 34417454 | 34426069 | 0.97 | 0.81 | 1.14 | 6.86E-01 | 0.878 |
| OID21107 | TNXB | Neurology | TNXB | P22105 | 6 | 32041153 | 32115334 | 0.97 | 0.82 | 1.14 | 6.87E-01 | 0.878 |
| OID21199 | BTC | Oncology | BTC | P35070 | 4 | 74744759 | 74794523 | 0.97 | 0.82 | 1.14 | 6.87E-01 | 0.878 |
| OID30155 | FRMD4B | Cardiometabolic_II | FRMD4B | Q9Y2L6 | 3 | 69168782 | 69542583 | 0.97 | 0.82 | 1.14 | 6.87E-01 | 0.878 |
| OID31469 | GPIHBP1 | Oncology_II | GPIHBP1 | Q8IV16 | 8 | 143213218 | 143217170 | 1.04 | 0.87 | 1.23 | 6.88E-01 | 0.879 |
| OID20335 | COL1A1 | Cardiometabolic | COL1A1 | P02452 | 17 | 50184101 | 50201632 | 0.97 | 0.83 | 1.13 | 6.90E-01 | 0.879 |
| OID21233 | FMR1 | Oncology | FMR1 | Q06787 | X | 147911919 | 147951125 | 1.03 | 0.88 | 1.20 | 6.90E-01 | 0.879 |
| OID20301 | LILRB2 | Cardiometabolic | LILRB2 | Q8N423 | 19 | 54273812 | 54281184 | 1.04 | 0.87 | 1.23 | 6.90E-01 | 0.879 |
| OID30825 | OTOA | Neurology_II | OTOA | Q7RTW8 | 16 | 21663968 | 21761935 | 1.03 | 0.88 | 1.22 | 6.90E-01 | 0.879 |
| OID30514 | PSMG4 | Inflammation_II | PSMG4 | Q5JS54 | 6 | 3231403 | 3303373 | 0.96 | 0.81 | 1.15 | 6.90E-01 | 0.879 |
| OID30468 | SSBP1 | Inflammation_II | SSBP1 | Q04837 | 7 | 141738334 | 141787922 | 0.97 | 0.81 | 1.15 | 6.90E-01 | 0.879 |
| OID21500 | CTSF | Oncology | CTSF | Q9UBX1 | 11 | 66563464 | 66568879 | 0.97 | 0.82 | 1.14 | 6.92E-01 | 0.879 |
| OID21125 | EFNA1 | Neurology | EFNA1 | P20827 | 1 | 155127876 | 155134899 | 1.03 | 0.88 | 1.21 | 6.91E-01 | 0.879 |
| OID31223 | ERCC1 | Oncology_II | ERCC1 | P07992 | 19 | 45407334 | 45478828 | 1.04 | 0.87 | 1.24 | 6.91E-01 | 0.879 |
| OID20946 | FLRT2 | Neurology | FLRT2 | O43155 | 14 | 85530144 | 85654428 | 1.03 | 0.88 | 1.22 | 6.92E-01 | 0.879 |
| OID20168 | MEP1B | Cardiometabolic | MEP1B | Q16820 | 18 | 32185069 | 32220404 | 1.03 | 0.88 | 1.22 | 6.91E-01 | 0.879 |
| OID31255 | COL9A2 | Oncology_II | COL9A2 | Q14055 | 1 | 40300489 | 40317813 | 0.97 | 0.82 | 1.14 | 6.92E-01 | 0.879 |
| OID30845 | LELP1 | Neurology_II | LELP1 | Q5T871 | 1 | 153203430 | 153205120 | 0.97 | 0.83 | 1.14 | 6.92E-01 | 0.879 |
| OID30441 | IL21R | Inflammation_II | IL21R | Q9HBE5 | 16 | 27402174 | 27452042 | 0.97 | 0.82 | 1.14 | 6.94E-01 | 0.880 |
| OID20746 | MEGF10 | Inflammation | MEGF10 | Q96KG7 | 5 | 127290796 | 127465737 | 0.97 | 0.82 | 1.14 | 6.95E-01 | 0.880 |
| OID31023 | MRPL28 | Neurology_II | MRPL28 | Q13084 | 16 | 366969 | 371289 | 1.03 | 0.88 | 1.20 | 6.95E-01 | 0.880 |
| OID20219 | SERPINA12 | Cardiometabolic | SERPINA12 | Q8IW75 | 14 | 94487274 | 94517844 | 1.03 | 0.87 | 1.23 | 6.94E-01 | 0.880 |
| OID31290 | TLR2 | Oncology_II | TLR2 | O60603 | 4 | 153684070 | 153706260 | 0.97 | 0.82 | 1.14 | 6.94E-01 | 0.880 |
| OID20671 | CCL20 | Inflammation | CCL20 | P78556 | 2 | 227805739 | 227817564 | 1.03 | 0.88 | 1.22 | 6.96E-01 | 0.881 |
| OID30050 | DENND2B | Cardiometabolic_II | DENND2B | P78524 | 11 | 8693351 | 8910951 | 1.03 | 0.89 | 1.20 | 6.96E-01 | 0.881 |
| OID20891 | ECE1 | Neurology | ECE1 | P42892 | 1 | 21217247 | 21345572 | 0.97 | 0.83 | 1.13 | 6.96E-01 | 0.881 |
| OID30273 | ENPP6 | Cardiometabolic_II | ENPP6 | Q6UWR7 | 4 | 184088706 | 184221230 | 0.97 | 0.81 | 1.15 | 6.96E-01 | 0.881 |
| OID20282 | CXCL16 | Cardiometabolic | CXCL16 | Q9H2A7 | 17 | 4733533 | 4739928 | 0.97 | 0.82 | 1.14 | 6.97E-01 | 0.881 |
| OID31235 | DCDC2C | Oncology_II | DCDC2C | A8MYV0 | 2 | 3703575 | 3848008 | 0.97 | 0.82 | 1.14 | 6.97E-01 | 0.881 |
| OID20428 | IL33 | Inflammation | IL33 | O95760 | 9 | 6215786 | 6257983 | 0.97 | 0.82 | 1.14 | 6.97E-01 | 0.881 |
| OID20361 | SPARCL1 | Cardiometabolic | SPARCL1 | Q14515 | 4 | 87473335 | 87531061 | 0.97 | 0.82 | 1.14 | 6.98E-01 | 0.882 |
| OID20413 | IGFBP6 | Cardiometabolic | IGFBP6 | P24592 | 12 | 53097436 | 53102345 | 0.97 | 0.81 | 1.15 | 6.99E-01 | 0.882 |
| OID20102 | CEACAM8 | Cardiometabolic | CEACAM8 | P31997 | 19 | 42580243 | 42595055 | 0.97 | 0.81 | 1.15 | 7.00E-01 | 0.884 |
| OID30954 | EFNB2 | Neurology_II | EFNB2 | P52799 | 13 | 106489745 | 106535662 | 0.97 | 0.81 | 1.16 | 7.02E-01 | 0.885 |
| OID31377 | PCDHB15 | Oncology_II | PCDHB15 | Q9Y5E8 | 5 | 141245395 | 141249365 | 0.97 | 0.82 | 1.15 | 7.02E-01 | 0.885 |
| OID31526 | RBP7 | Oncology_II | RBP7 | Q96R05 | 1 | 9997206 | 10016021 | 0.97 | 0.80 | 1.16 | 7.02E-01 | 0.885 |
| OID30991 | LEO1 | Neurology_II | LEO1 | Q8WVC0 | 15 | 51938025 | 51971778 | 0.96 | 0.80 | 1.16 | 7.04E-01 | 0.886 |
| OID30224 | EDN1 | Cardiometabolic_II | EDN1 | P05305 | 6 | 12290361 | 12297194 | 1.03 | 0.88 | 1.22 | 7.04E-01 | 0.886 |
| OID20980 | OBP2B | Neurology | OBP2B | Q9NPH6 | 9 | 133205277 | 133209250 | 0.96 | 0.79 | 1.18 | 7.05E-01 | 0.887 |
| OID20101 | IL6 | Cardiometabolic/Inflam | IL6 | P05231 | 7 | 22725884 | 22732002 | 1.03 | 0.88 | 1.21 | 7.05E-01 | 0.887 |
| OID21053 | CPPED1 | Neurology | CPPED1 | Q9BRF8 | 16 | 12659799 | 12803887 | 1.03 | 0.87 | 1.23 | 7.06E-01 | 0.887 |
| OID20491 | CSF3 | Inflammation | CSF3 | P09919 | 17 | 40015361 | 40017813 | 0.97 | 0.82 | 1.15 | 7.06E-01 | 0.887 |
| OID20538 | ISM1 | Inflammation | ISM1 | B1AKI9 | 20 | 13221274 | 13300651 | 0.97 | 0.81 | 1.15 | 7.06E-01 | 0.887 |
| OID30516 | SHH | Inflammation_II | SHH | Q15465 | 7 | 155799980 | 155812463 | 1.03 | 0.87 | 1.22 | 7.06E-01 | 0.887 |
| OID30712 | CAT | Inflammation_II | CAT | P04040 | 11 | 34438934 | 34472060 | 0.97 | 0.81 | 1.16 | 7.08E-01 | 0.887 |
| OID20495 | IFNG | Inflammation | IFNG | P01579 | 12 | 68154768 | 68159740 | 1.03 | 0.88 | 1.22 | 7.08E-01 | 0.887 |
| OID30599 | SEMA3G | Inflammation_II | SEMA3G | Q9NS98 | 3 | 52433035 | 52445103 | 0.97 | 0.82 | 1.14 | 7.07E-01 | 0.887 |
| OID20265 | TNFRSF10C | Cardiometabolic | TNFRSF10C | O14798 | 8 | 23102921 | 23117445 | 1.03 | 0.88 | 1.22 | 7.07E-01 | 0.887 |
| OID31325 | ITGAX | Oncology_II | ITGAX | P20702 | 16 | 31355134 | 31382999 | 0.97 | 0.81 | 1.15 | 7.08E-01 | 0.887 |
| OID20789 | NME3 | Inflammation | NME3 | Q13232 | 16 | 1770320 | 1771561 | 0.97 | 0.81 | 1.16 | 7.10E-01 | 0.890 |
| OID21390 | SIGLEC9 | Oncology | SIGLEC9 | Q9Y336 | 19 | 51124906 | 51136651 | 0.97 | 0.82 | 1.15 | 7.10E-01 | 0.890 |
| OID20298 | CELA3A | Cardiometabolic | CELA3A | P09093 | 1 | 22001657 | 22012542 | 1.03 | 0.88 | 1.20 | 7.11E-01 | 0.890 |
| OID31489 | C9orf40 | Oncology_II | C9orf40 | Q8IXQ3 | 9 | 74946583 | 74952912 | 0.97 | 0.82 | 1.15 | 7.13E-01 | 0.892 |
| OID21318 | CDNF | Oncology | CDNF | Q49AH0 | 10 | 14819245 | 14838575 | 0.97 | 0.83 | 1.14 | 7.14E-01 | 0.892 |
| OID31387 | SMPDL3B | Oncology_II | SMPDL3B | Q92485 | 1 | 27935000 | 27959157 | 0.97 | 0.83 | 1.14 | 7.14E-01 | 0.892 |
| OID21206 | MAEA | Oncology | MAEA | Q7L5Y9 | 4 | 1289887 | 1340147 | 1.03 | 0.88 | 1.21 | 7.15E-01 | 0.893 |
| OID31490 | NCR3LG1 | Oncology_II | NCR3LG1 | Q68D85 | 11 | 17351800 | 17377341 | 1.03 | 0.88 | 1.21 | 7.15E-01 | 0.893 |
| OID30068 | TMED10 | Cardiometabolic_II | TMED10 | P49755 | 14 | 75131469 | 75176612 | 1.03 | 0.89 | 1.18 | 7.15E-01 | 0.893 |
| OID31390 | COMMD9 | Oncology_II | COMMD9 | Q9P000 | 11 | 36269284 | 36289449 | 0.97 | 0.80 | 1.16 | 7.16E-01 | 0.894 |
| OID30393 | CTHRC1 | Cardiometabolic_II | CTHRC1 | Q96CG8 | 8 | 103371538 | 103382989 | 1.03 | 0.86 | 1.25 | 7.17E-01 | 0.895 |
| OID20105 | ACE2 | Cardiometabolic | ACE2 | Q9BYF1 | X | 15494566 | 15607236 | 1.03 | 0.88 | 1.21 | 7.18E-01 | 0.895 |
| OID20366 | CHL1 | Cardiometabolic | CHL1 | O00533 | 3 | 196763 | 409417 | 1.03 | 0.88 | 1.20 | 7.19E-01 | 0.895 |
| OID20913 | NMNAT1 | Neurology | NMNAT1 | Q9HAN9 | 1 | 9943428 | 9985501 | 0.97 | 0.81 | 1.16 | 7.18E-01 | 0.895 |
| OID31398 | SUSD4 | Oncology_II | SUSD4 | Q5VX71 | 1 | 223220819 | 223364233 | 1.03 | 0.87 | 1.22 | 7.19E-01 | 0.895 |
| OID20124 | ACOX1 | Cardiometabolic | ACOX1 | Q15067 | 17 | 75941507 | 75979177 | 0.97 | 0.83 | 1.14 | 7.20E-01 | 0.896 |
| OID21408 | HSPB6 | Oncology | HSPB6 | O14558 | 19 | 35754566 | 35758079 | 1.03 | 0.86 | 1.25 | 7.20E-01 | 0.896 |
| OID21129 | PILRA | Neurology | PILRA | Q9UKJ1 | 7 | 100367530 | 100400096 | 0.97 | 0.83 | 1.14 | 7.20E-01 | 0.896 |
| OID30324 | PTPRB | Cardiometabolic_II | PTPRB | P23467 | 12 | 70515870 | 70637440 | 1.03 | 0.87 | 1.23 | 7.21E-01 | 0.896 |
| OID30246 | APOBR | Cardiometabolic_II | APOBR | Q0VD83 | 16 | 28494643 | 28498964 | 1.03 | 0.87 | 1.22 | 7.22E-01 | 0.896 |
| OID30976 | DLGAP5 | Neurology_II | DLGAP5 | Q15398 | 14 | 55148112 | 55191608 | 0.97 | 0.81 | 1.16 | 7.22E-01 | 0.896 |
| OID30141 | MANSC4 | Cardiometabolic_II | MANSC4 | A6NHS7 | 12 | 27762427 | 27780236 | 1.03 | 0.88 | 1.19 | 7.22E-01 | 0.896 |
| OID30087 | NOS2 | Cardiometabolic_II | NOS2 | P35228 | 17 | 27756766 | 27800529 | 0.97 | 0.82 | 1.15 | 7.22E-01 | 0.896 |
| OID31186 | CPLX2 | Oncology_II | CPLX2 | Q6PUV4 | 5 | 175796310 | 175884021 | 1.03 | 0.87 | 1.23 | 7.23E-01 | 0.897 |
| OID30422 | IL12RB2 | Inflammation_II | IL12RB2 | Q99665 | 1 | 67307364 | 67398724 | 0.97 | 0.81 | 1.16 | 7.23E-01 | 0.897 |
| OID20788 | CXCL3 | Inflammation | CXCL3 | P19876 | 4 | 74036589 | 74038807 | 0.97 | 0.83 | 1.14 | 7.25E-01 | 0.897 |
| OID30157 | HMGCS1 | Cardiometabolic_II | HMGCS1 | Q01581 | 5 | 43287470 | 43313512 | 1.03 | 0.88 | 1.21 | 7.24E-01 | 0.897 |
| OID20652 | IL18R1 | Inflammation | IL18R1 | Q13478 | 2 | 102311529 | 102398777 | 1.03 | 0.87 | 1.22 | 7.24E-01 | 0.897 |
| OID20734 | ROBO1 | Inflammation | ROBO1 | Q9Y6N7 | 3 | 78597239 | 79767998 | 1.03 | 0.88 | 1.21 | 7.25E-01 | 0.897 |
| OID21106 | NOMO1 | Neurology | NOMO1 | Q15155 | 16 | 14833721 | 14896157 | 0.97 | 0.82 | 1.15 | 7.25E-01 | 0.897 |
| OID30572 | AMOT | Inflammation_II | AMOT | Q4VCS5 | X | 112774503 | 112840831 | 0.97 | 0.80 | 1.16 | 7.27E-01 | 0.898 |
| OID21291 | KLK13 | Oncology | KLK13 | Q9UKR3 | 19 | 51055626 | 51065114 | 1.03 | 0.88 | 1.20 | 7.27E-01 | 0.898 |
| OID20580 | LY75 | Inflammation | LY75 | O60449 | 2 | 159803355 | 159904756 | 0.97 | 0.82 | 1.15 | 7.27E-01 | 0.898 |
| OID30531 | SEMA6C | Inflammation_II | SEMA6C | Q9H3T2 | 1 | 151131685 | 151146631 | 1.03 | 0.88 | 1.21 | 7.27E-01 | 0.898 |
| OID31139 | AMPD3 | Neurology_II | AMPD3 | Q01432 | 11 | 10308313 | 10507579 | 0.97 | 0.82 | 1.15 | 7.28E-01 | 0.899 |
| OID21515 | CREG1 | Oncology | CREG1 | O75629 | 1 | 167529117 | 167553805 | 1.03 | 0.88 | 1.21 | 7.28E-01 | 0.899 |
| OID30059 | GCLM | Cardiometabolic_II | GCLM | P48507 | 1 | 93885199 | 93909456 | 0.97 | 0.82 | 1.15 | 7.29E-01 | 0.899 |

| | | | | | | | | | | | | |
|---|---|---|---|---|---|---|---|---|---|---|---|---|
| OID21146 | IGF2R | Neurology | IGF2R | P11717 | 6 | 159969082 | 160113507 | 1.03 | 0.87 | 1.21 | 7.29E-01 | 0.899 |
| OID30814 | IMPG1 | Neurology_II | IMPG1 | Q17R60 | 6 | 75921114 | 76072678 | 1.03 | 0.87 | 1.21 | 7.29E-01 | 0.899 |
| OID31470 | TSPAN8 | Oncology_II | TSPAN8 | P19075 | 12 | 71125085 | 71441898 | 0.97 | 0.82 | 1.15 | 7.29E-01 | 0.899 |
| OID30084 | PDE4D | Cardiometabolic_II | PDE4D | Q08499 | 5 | 58969038 | 60522120 | 0.97 | 0.80 | 1.16 | 7.30E-01 | 0.899 |
| OID30049 | RTKN2 | Cardiometabolic_II | RTKN2 | Q8IZC4 | 10 | 62183035 | 62268844 | 1.03 | 0.88 | 1.19 | 7.30E-01 | 0.899 |
| OID20081 | EDIL3 | Cardiometabolic | EDIL3 | O43854 | 5 | 83940554 | 84384880 | 1.03 | 0.87 | 1.23 | 7.30E-01 | 0.899 |
| OID31296 | SPRR1B | Oncology_II | SPRR1B | P22528 | 1 | 153031203 | 153032900 | 0.97 | 0.81 | 1.15 | 7.31E-01 | 0.899 |
| OID31363 | BTN1A1 | Oncology_II | BTN1A1 | Q13410 | 6 | 26500303 | 26510425 | 0.97 | 0.82 | 1.15 | 7.32E-01 | 0.900 |
| OID20823 | IDI2 | Neurology | IDI2 | Q9BXS1 | 10 | 1018910 | 1025859 | 0.97 | 0.82 | 1.15 | 7.33E-01 | 0.901 |
| OID30528 | ADAMTS1 | Inflammation_II | ADAMTS1 | Q9UHI8 | 21 | 26835755 | 26845409 | 0.97 | 0.83 | 1.14 | 7.34E-01 | 0.901 |
| OID20096 | GZMH | Cardiometabolic | GZMH | P20718 | 14 | 24606480 | 24609699 | 0.97 | 0.82 | 1.15 | 7.34E-01 | 0.901 |
| OID30743 | SERPINA6 | Inflammation_II | SERPINA6 | P08185 | 14 | 94304248 | 94323389 | 0.97 | 0.80 | 1.17 | 7.34E-01 | 0.901 |
| OID20842 | TPPP3 | Neurology | TPPP3 | Q9BW30 | 16 | 67389809 | 67393518 | 0.97 | 0.82 | 1.15 | 7.35E-01 | 0.901 |
| OID21459 | HAGH | Oncology | HAGH | Q16775 | 16 | 1795620 | 1827157 | 1.03 | 0.87 | 1.22 | 7.35E-01 | 0.901 |
| OID31091 | GFRAL | Neurology_II | GFRAL | Q6UXV0 | 6 | 55327469 | 55402493 | 0.97 | 0.83 | 1.14 | 7.35E-01 | 0.901 |
| OID20311 | NOTCH1 | Cardiometabolic | NOTCH1 | P46531 | 9 | 136494433 | 136546048 | 0.97 | 0.83 | 1.14 | 7.36E-01 | 0.901 |
| OID20658 | AGRP | Inflammation | AGRP | O00253 | 16 | 67482571 | 67483547 | 1.03 | 0.87 | 1.21 | 7.37E-01 | 0.902 |
| OID20832 | PADI4 | Neurology | PADI4 | Q9UM07 | 1 | 17308195 | 17364004 | 0.97 | 0.82 | 1.15 | 7.37E-01 | 0.902 |
| OID30543 | CD7 | Inflammation_II | CD7 | P09564 | 17 | 82314868 | 82317608 | 0.97 | 0.83 | 1.14 | 7.40E-01 | 0.905 |
| OID20878 | S100A16 | Neurology | S100A16 | Q96FQ6 | 1 | 153606886 | 153613145 | 1.03 | 0.88 | 1.21 | 7.40E-01 | 0.905 |
| OID20728 | SCG3 | Inflammation | SCG3 | Q8WXD2 | 15 | 51681492 | 51721026 | 0.97 | 0.83 | 1.14 | 7.40E-01 | 0.905 |
| OID20566 | NCR1 | Inflammation | NCR1 | O76036 | 19 | 54906148 | 54916140 | 1.03 | 0.88 | 1.20 | 7.41E-01 | 0.906 |
| OID20792 | NDRG1 | Neurology | NDRG1 | Q92597 | 8 | 133237175 | 133302022 | 0.97 | 0.82 | 1.15 | 7.42E-01 | 0.907 |
| OID20798 | ATF2 | Neurology | ATF2 | P15336 | 2 | 175072250 | 175168382 | 1.03 | 0.86 | 1.23 | 7.43E-01 | 0.907 |
| OID21074 | NPTX1 | Neurology | NPTX1 | Q15818 | 17 | 80466834 | 80477843 | 1.03 | 0.87 | 1.22 | 7.44E-01 | 0.908 |
| OID31130 | DNAJA4 | Neurology_II | DNAJA4 | Q8WW22 | 15 | 78264086 | 78282196 | 0.97 | 0.82 | 1.15 | 7.45E-01 | 0.909 |
| OID30347 | MELTF | Cardiometabolic_II | MELTF | P08582 | 3 | 196980590 | 197029835 | 0.97 | 0.83 | 1.14 | 7.45E-01 | 0.909 |
| OID30833 | SCN2A | Neurology_II | SCN2A | Q99250 | 2 | 165194993 | 165392310 | 0.97 | 0.83 | 1.14 | 7.46E-01 | 0.909 |
| OID30789 | SERPINC1 | Inflammation_II | SERPINC1 | P01008 | 1 | 173903800 | 173917327 | 0.97 | 0.82 | 1.15 | 7.45E-01 | 0.909 |
| OID30750 | SERPINA1 | Inflammation_II | SERPINA1 | P01009 | 14 | 94376747 | 94390693 | 0.97 | 0.82 | 1.15 | 7.46E-01 | 0.909 |
| OID30069 | RAB39B | Cardiometabolic_II | RAB39B | Q96DA2 | X | 155258235 | 155264491 | 0.97 | 0.81 | 1.16 | 7.47E-01 | 0.909 |
| OID21176 | PDP1 | Oncology | PDP1 | Q9P0J1 | 8 | 93857807 | 93926068 | 0.97 | 0.82 | 1.15 | 7.48E-01 | 0.910 |
| OID21033 | PRL | Neurology | PRL | P01236 | 6 | 22287244 | 22302826 | 0.98 | 0.84 | 1.14 | 7.48E-01 | 0.910 |
| OID30579 | HEG1 | Inflammation_II | HEG1 | Q9ULI3 | 3 | 124965710 | 125055997 | 0.97 | 0.82 | 1.15 | 7.48E-01 | 0.910 |
| OID20699 | CRLF1 | Inflammation | CRLF1 | O75462 | 19 | 18572220 | 18607741 | 1.03 | 0.87 | 1.22 | 7.49E-01 | 0.910 |
| OID20527 | DPP10 | Inflammation | DPP10 | Q8N608 | 2 | 114442299 | 115845780 | 1.03 | 0.87 | 1.22 | 7.49E-01 | 0.910 |
| OID20856 | BAX | Neurology | BAX | Q07812 | 19 | 48954815 | 48961798 | 0.97 | 0.82 | 1.15 | 7.51E-01 | 0.910 |
| OID31242 | CDC123 | Oncology_II | CDC123 | O75794 | 10 | 12195965 | 12250589 | 0.97 | 0.81 | 1.16 | 7.52E-01 | 0.910 |
| OID21260 | ENTPD2 | Oncology | ENTPD2 | Q9Y5L3 | 9 | 137048107 | 137054061 | 0.97 | 0.83 | 1.14 | 7.50E-01 | 0.910 |
| OID21372 | FAM3B | Oncology | FAM3B | P58499 | 21 | 41304212 | 41357727 | 1.03 | 0.86 | 1.22 | 7.52E-01 | 0.910 |
| OID21296 | KRT18 | Oncology | KRT18 | P05783 | 12 | 52948871 | 52952906 | 0.97 | 0.81 | 1.17 | 7.52E-01 | 0.910 |
| OID21378 | MUC16 | Oncology | MUC16 | Q8WXI7 | 19 | 8848844 | 8981342 | 1.03 | 0.87 | 1.20 | 7.50E-01 | 0.910 |
| OID20451 | MYO9B | Inflammation | MYO9B | Q13459 | 19 | 17075777 | 17214537 | 0.97 | 0.82 | 1.15 | 7.51E-01 | 0.910 |
| OID21304 | PSMD9 | Oncology | PSMD9 | O00233 | 12 | 121888732 | 121918297 | 0.97 | 0.82 | 1.16 | 7.50E-01 | 0.910 |
| OID20811 | ATP5PO | Neurology | ATP5PO | P48047 | 21 | 33903453 | 33915814 | 1.03 | 0.88 | 1.20 | 7.53E-01 | 0.911 |
| OID20435 | ACTN4 | Inflammation | ACTN4 | O43707 | 19 | 38647649 | 38731589 | 0.97 | 0.81 | 1.17 | 7.54E-01 | 0.911 |
| OID30165 | B3GAT3 | Cardiometabolic_II | B3GAT3 | O94766 | 11 | 62615296 | 62622154 | 0.98 | 0.84 | 1.14 | 7.62E-01 | 0.911 |
| OID30482 | CA8 | Inflammation_II | CA8 | P35219 | 8 | 60185412 | 60281400 | 1.03 | 0.87 | 1.22 | 7.58E-01 | 0.911 |
| OID30820 | CACNB1 | Neurology_II | CACNB1 | Q02641 | 17 | 39173453 | 39197702 | 1.02 | 0.89 | 1.18 | 7.60E-01 | 0.911 |
| OID21431 | CAPG | Oncology | CAPG | P40121 | 2 | 85394753 | 85418432 | 1.03 | 0.86 | 1.22 | 7.56E-01 | 0.911 |
| OID21469 | CD300LF | Oncology | CD300LF | Q8TDQ1 | 17 | 74694311 | 74712978 | 1.03 | 0.87 | 1.22 | 7.58E-01 | 0.911 |
| OID20692 | CD48 | Inflammation | CD48 | P09326 | 1 | 160678746 | 160711831 | 0.97 | 0.83 | 1.15 | 7.58E-01 | 0.911 |
| OID30240 | CEP170 | Cardiometabolic_II | CEP170 | Q5SW79 | 1 | 243124428 | 243255348 | 0.97 | 0.82 | 1.15 | 7.56E-01 | 0.911 |
| OID30403 | CHGA | Cardiometabolic_II | CHGA | P10645 | 14 | 92923150 | 92935285 | 0.98 | 0.86 | 1.12 | 7.57E-01 | 0.911 |
| OID30552 | CLEC12A | Inflammation_II | CLEC12A | Q5QGZ9 | 12 | 9951316 | 9995694 | 1.03 | 0.87 | 1.20 | 7.61E-01 | 0.911 |
| OID21305 | DPEP2 | Oncology | DPEP2 | Q9H4A9 | 16 | 67987394 | 68000586 | 0.97 | 0.83 | 1.15 | 7.62E-01 | 0.911 |
| OID30254 | FSTL1 | Cardiometabolic_II | FSTL1 | Q12841 | 3 | 120392293 | 120450993 | 1.03 | 0.87 | 1.21 | 7.59E-01 | 0.911 |
| OID20520 | HLA-DRA | Inflammation | HLA-DRA | P01903 | 6 | 32439878 | 32445046 | 1.02 | 0.88 | 1.20 | 7.55E-01 | 0.911 |
| OID20824 | LIF | Neurology | LIF | P15018 | 22 | 30240453 | 30246759 | 0.97 | 0.83 | 1.15 | 7.62E-01 | 0.911 |
| OID21452 | LTA4H | Oncology | LTA4H | P09960 | 12 | 96000753 | 96043520 | 1.03 | 0.87 | 1.21 | 7.62E-01 | 0.911 |
| OID31033 | MCEE | Neurology_II | MCEE | Q96PE7 | 2 | 71109684 | 71130239 | 0.98 | 0.83 | 1.15 | 7.61E-01 | 0.911 |
| OID21100 | MPO | Neurology | MPO | P05164 | 17 | 58269855 | 58280935 | 1.03 | 0.86 | 1.22 | 7.60E-01 | 0.911 |
| OID31170 | MUCL3 | Oncology_II | MUCL3 | Q3MIW9 | 6 | 30934523 | 30954221 | 0.97 | 0.82 | 1.16 | 7.56E-01 | 0.911 |
| OID30904 | PGM2 | Neurology_II | PGM2 | Q96G03 | 4 | 37826660 | 37862937 | 1.02 | 0.88 | 1.19 | 7.59E-01 | 0.911 |
| OID30184 | PKD2 | Cardiometabolic_II | PKD2 | Q13563 | 4 | 88007635 | 88077777 | 0.97 | 0.82 | 1.16 | 7.59E-01 | 0.911 |
| OID31122 | PNLIPRP1 | Neurology_II | PNLIPRP1 | P54315 | 10 | 116590385 | 116609175 | 0.98 | 0.83 | 1.14 | 7.55E-01 | 0.911 |
| OID30276 | PRRT3 | Cardiometabolic_II | PRRT3 | Q5FWE3 | 3 | 9945542 | 9952408 | 1.03 | 0.87 | 1.21 | 7.54E-01 | 0.911 |
| OID30561 | PTGES2 | Inflammation_II | PTGES2 | Q9H7Z7 | 9 | 128120693 | 128128462 | 0.97 | 0.82 | 1.15 | 7.61E-01 | 0.911 |
| OID30577 | SCGB3A1 | Inflammation_II | SCGB3A1 | Q96QR1 | 5 | 180590105 | 180591499 | 0.97 | 0.82 | 1.15 | 7.56E-01 | 0.911 |
| OID20560 | SELPLG | Inflammation | SELPLG | Q14242 | 12 | 108621895 | 108633894 | 1.03 | 0.86 | 1.23 | 7.56E-01 | 0.911 |
| OID30387 | SHISA5 | Cardiometabolic_II | SHISA5 | Q8N114 | 3 | 48467798 | 48504826 | 1.03 | 0.87 | 1.21 | 7.62E-01 | 0.911 |
| OID31509 | SPINT3 | Oncology_II | SPINT3 | P49223 | 20 | 45512461 | 45515622 | 1.05 | 0.77 | 1.43 | 7.60E-01 | 0.911 |
| OID20531 | STX8 | Inflammation | STX8 | Q9UNK0 | 17 | 9250471 | 9576591 | 0.97 | 0.82 | 1.16 | 7.60E-01 | 0.911 |
| OID20733 | TNFSF13 | Inflammation | TNFSF13 | O75888 | 17 | 7558292 | 7561608 | 0.97 | 0.82 | 1.16 | 7.59E-01 | 0.911 |
| OID30447 | UNC5D | Inflammation_II | UNC5D | Q6UXZ4 | 8 | 35235475 | 35796550 | 0.97 | 0.82 | 1.15 | 7.56E-01 | 0.911 |
| OID30348 | FAM20A | Cardiometabolic_II | FAM20A | Q96MK3 | 17 | 68535113 | 68601367 | 1.03 | 0.86 | 1.23 | 7.64E-01 | 0.913 |
| OID20533 | TRIM21 | Inflammation | TRIM21 | P19474 | 11 | 4384897 | 4393702 | 0.98 | 0.84 | 1.14 | 7.64E-01 | 0.913 |
| OID31249 | CEP152 | Oncology_II | CEP152 | O94986 | 15 | 48712928 | 48811146 | 0.97 | 0.82 | 1.16 | 7.66E-01 | 0.914 |
| OID30419 | PSAP | Cardiometabolic_II | PSAP | P07602 | 10 | 71816298 | 71851251 | 0.98 | 0.83 | 1.15 | 7.66E-01 | 0.915 |
| OID30930 | CHRM1 | Neurology_II | CHRM1 | P11229 | 11 | 62908679 | 62921807 | 0.98 | 0.83 | 1.15 | 7.67E-01 | 0.915 |
| OID31200 | PSMC3 | Oncology_II | PSMC3 | P17980 | 11 | 47418769 | 47426473 | 0.98 | 0.83 | 1.15 | 7.67E-01 | 0.915 |
| OID30417 | GOT1 | Cardiometabolic_II | GOT1 | P17174 | 10 | 99396870 | 99430624 | 0.97 | 0.82 | 1.16 | 7.68E-01 | 0.916 |
| OID30510 | SLITRK1 | Inflammation_II | SLITRK1 | Q96PX8 | 13 | 83877205 | 83882474 | 0.97 | 0.82 | 1.16 | 7.68E-01 | 0.916 |
| OID30824 | HTR1A | Neurology_II | HTR1A | P08908 | 5 | 63957874 | 63962507 | 1.02 | 0.88 | 1.19 | 7.70E-01 | 0.916 |
| OID30402 | IGDCC4 | Cardiometabolic_II | IGDCC4 | Q8TDY8 | 15 | 65381484 | 65422947 | 1.03 | 0.86 | 1.22 | 7.70E-01 | 0.916 |
| OID21526 | KLK6 | Oncology | KLK6 | Q92876 | 19 | 50958631 | 50969673 | 0.98 | 0.84 | 1.14 | 7.71E-01 | 0.916 |
| OID20057 | MFAP3 | Cardiometabolic | MFAP3 | P55082 | 5 | 154038959 | 154220478 | 1.02 | 0.87 | 1.21 | 7.71E-01 | 0.916 |
| OID30296 | MYBPC1 | Cardiometabolic_II | MYBPC1 | Q00872 | 12 | 101568353 | 101686028 | 0.97 | 0.82 | 1.16 | 7.69E-01 | 0.916 |
| OID20766 | PRELP | Inflammation | PRELP | P51888 | 1 | 203475806 | 203491352 | 0.98 | 0.85 | 1.12 | 7.70E-01 | 0.916 |
| OID20948 | SETMAR | Neurology | SETMAR | Q53H47 | 3 | 4303332 | 4317265 | 0.97 | 0.80 | 1.18 | 7.70E-01 | 0.916 |
| OID31233 | TOP1MT | Oncology_II | TOP1MT | Q969P6 | 8 | 143304384 | 143359979 | 0.98 | 0.83 | 1.15 | 7.71E-01 | 0.916 |
| OID30366 | AMDHD2 | Cardiometabolic_II | AMDHD2 | Q9Y303 | 16 | 2520357 | 2531422 | 1.03 | 0.87 | 1.21 | 7.73E-01 | 0.917 |
| OID30618 | C1QTNF5 | Inflammation_II | C1QTNF5 | Q9BXJ0 | 11 | 119338942 | 119340940 | 0.98 | 0.83 | 1.15 | 7.72E-01 | 0.917 |
| OID31384 | CAPN3 | Oncology_II | CAPN3 | P20807 | 15 | 42359498 | 42412949 | 1.02 | 0.89 | 1.17 | 7.73E-01 | 0.917 |
| OID21274 | IGF1R | Oncology | IGF1R | P08069 | 15 | 98648539 | 98964530 | 1.02 | 0.87 | 1.20 | 7.75E-01 | 0.919 |
| OID20273 | ALCAM | Cardiometabolic | ALCAM | Q13740 | 3 | 105366909 | 105576900 | 1.03 | 0.86 | 1.22 | 7.76E-01 | 0.919 |
| OID31316 | CCND2 | Oncology_II | CCND2 | P30279 | 12 | 4269771 | 4305353 | 1.03 | 0.86 | 1.22 | 7.75E-01 | 0.919 |
| OID31231 | DCUN1D2 | Oncology_II | DCUN1D2 | Q6PH85 | 13 | 113455819 | 113490951 | 1.02 | 0.88 | 1.19 | 7.78E-01 | 0.919 |
| OID20712 | DNER | Inflammation | DNER | Q8NFT8 | 2 | 229357629 | 229714555 | 0.97 | 0.82 | 1.16 | 7.78E-01 | 0.919 |
| OID31118 | DNPEP | Neurology_II | DNPEP | Q9ULA0 | 2 | 219372043 | 219400022 | 0.98 | 0.84 | 1.14 | 7.79E-01 | 0.919 |
| OID20248 | ICAM2 | Cardiometabolic | ICAM2 | P13598 | 17 | 64002594 | 64020634 | 1.02 | 0.87 | 1.20 | 7.78E-01 | 0.919 |
| OID20185 | LRP11 | Cardiometabolic | LRP11 | Q86VZ4 | 6 | 149818757 | 149864359 | 0.98 | 0.83 | 1.15 | 7.77E-01 | 0.919 |
| OID20362 | NID1 | Cardiometabolic | NID1 | P14543 | 1 | 235975830 | 236065109 | 0.98 | 0.83 | 1.15 | 7.78E-01 | 0.919 |

| | | | | | | | | | | | | |
|---|---|---|---|---|---|---|---|---|---|---|---|---|
| OID20235 | PCSK9 | Cardiometabolic | PCSK9 | Q8NBP7 | 1 | 55039447 | 55064852 | 0.98 | 0.83 | 1.15 | 7.78E-01 | 0.919 |
| OID31248 | REST | Oncology_II | REST | Q13127 | 4 | 56907876 | 56966808 | 1.03 | 0.86 | 1.22 | 7.76E-01 | 0.919 |
| OID31436 | SDHB | Oncology_II | SDHB | P21912 | 1 | 17018722 | 17054032 | 0.98 | 0.83 | 1.15 | 7.76E-01 | 0.919 |
| OID20796 | SMARCA2 | Neurology | SMARCA2 | P51531 | 9 | 1980290 | 2193624 | 0.98 | 0.82 | 1.16 | 7.77E-01 | 0.919 |
| OID21523 | KLK10 | Oncology | KLK10 | O43240 | 19 | 51012739 | 51020175 | 0.98 | 0.83 | 1.15 | 7.79E-01 | 0.919 |
| OID20375 | TIMD4 | Cardiometabolic | TIMD4 | Q96H15 | 5 | 156919292 | 156963226 | 1.02 | 0.87 | 1.20 | 7.80E-01 | 0.920 |
| OID20486 | IL12RB1 | Inflammation | IL12RB1 | P42701 | 19 | 18058995 | 18098944 | 1.02 | 0.87 | 1.19 | 7.82E-01 | 0.921 |
| OID20385 | IL6R | Cardiometabolic | IL6R | P08887 | 1 | 154405193 | 154469450 | 0.98 | 0.83 | 1.15 | 7.82E-01 | 0.921 |
| OID20164 | APLP1 | Cardiometabolic | APLP1 | P51693 | 19 | 35867899 | 35879792 | 1.02 | 0.87 | 1.20 | 7.83E-01 | 0.922 |
| OID20992 | FUT8 | Neurology | FUT8 | Q9BYC5 | 14 | 65410592 | 65744121 | 1.02 | 0.87 | 1.20 | 7.85E-01 | 0.922 |
| OID31243 | IL25 | Oncology_II | IL25 | Q9H293 | 14 | 23372809 | 23376403 | 1.02 | 0.88 | 1.19 | 7.83E-01 | 0.922 |
| OID30537 | ITGAL | Inflammation_II | ITGAL | P20701 | 16 | 30472658 | 30523567 | 0.98 | 0.82 | 1.16 | 7.84E-01 | 0.922 |
| OID21384 | MOG | Oncology | MOG | Q16653 | 6 | 29657002 | 29672372 | 0.98 | 0.83 | 1.15 | 7.84E-01 | 0.922 |
| OID31110 | PLB1 | Neurology_II | PLB1 | Q6P1J6 | 2 | 28457145 | 28644142 | 1.02 | 0.86 | 1.22 | 7.84E-01 | 0.922 |
| OID31203 | SLC34A3 | Oncology_II | SLC34A3 | Q8N130 | 9 | 137230757 | 137236555 | 1.02 | 0.87 | 1.21 | 7.83E-01 | 0.922 |
| OID30154 | TMED4 | Cardiometabolic_II | TMED4 | Q7Z7H5 | 7 | 44577894 | 44582287 | 1.02 | 0.88 | 1.19 | 7.84E-01 | 0.922 |
| OID20277 | ICAM3 | Cardiometabolic | ICAM3 | P32942 | 19 | 10333776 | 10339661 | 1.02 | 0.87 | 1.20 | 7.85E-01 | 0.922 |
| OID30936 | MYL6B | Neurology_II | MYL6B | P14649 | 12 | 56152256 | 56159647 | 0.98 | 0.84 | 1.14 | 7.86E-01 | 0.922 |
| OID30484 | PLCB1 | Inflammation_II | PLCB1 | Q9NQ66 | 20 | 8077251 | 8968360 | 0.98 | 0.82 | 1.17 | 7.87E-01 | 0.923 |
| OID30931 | FZD10 | Neurology_II | FZD10 | Q9ULW2 | 12 | 130162459 | 130165740 | 0.98 | 0.81 | 1.17 | 7.87E-01 | 0.923 |
| OID20512 | ALDH3A1 | Inflammation | ALDH3A1 | P30838 | 17 | 19737984 | 19748943 | 0.98 | 0.83 | 1.16 | 7.88E-01 | 0.924 |
| OID30788 | FGA | Inflammation_II | FGA | P02671 | 4 | 154583128 | 154590745 | 0.98 | 0.81 | 1.17 | 7.88E-01 | 0.924 |
| OID20389 | AOC3 | Cardiometabolic | AOC3 | Q16853 | 17 | 42851184 | 42858130 | 1.02 | 0.86 | 1.21 | 7.93E-01 | 0.924 |
| OID21067 | BIN2 | Neurology | BIN2 | Q9UBW5 | 12 | 51281038 | 51324668 | 0.98 | 0.83 | 1.15 | 7.93E-01 | 0.924 |
| OID21392 | CPE | Oncology | CPE | P16870 | 4 | 165361194 | 165498547 | 0.98 | 0.82 | 1.16 | 7.92E-01 | 0.924 |
| OID20070 | CRX | Cardiometabolic | CRX | O43186 | 19 | 47819779 | 47843330 | 0.98 | 0.84 | 1.15 | 7.91E-01 | 0.924 |
| OID31391 | ERN1 | Oncology_II | ERN1 | O75460 | 17 | 64039080 | 64130819 | 0.98 | 0.82 | 1.16 | 7.92E-01 | 0.924 |
| OID21132 | GNLY | Neurology | GNLY | P22749 | 2 | 85685175 | 85698852 | 0.98 | 0.83 | 1.16 | 7.94E-01 | 0.924 |
| OID20217 | HMOX1 | Cardiometabolic | HMOX1 | P09601 | 22 | 35380361 | 35394214 | 1.02 | 0.87 | 1.20 | 7.94E-01 | 0.924 |
| OID30481 | HS6ST2 | Inflammation_II | HS6ST2 | Q96MM7 | X | 132626015 | 132961395 | 0.98 | 0.83 | 1.15 | 7.91E-01 | 0.924 |
| OID30145 | HSD17B3 | Cardiometabolic_II | HSD17B3 | P37058 | 9 | 96235306 | 96302176 | 1.02 | 0.88 | 1.18 | 7.90E-01 | 0.924 |
| OID31370 | INSL4 | Oncology_II | INSL4 | Q14641 | 9 | 5231419 | 5235304 | 1.02 | 0.87 | 1.21 | 7.94E-01 | 0.924 |
| OID21517 | KLK8 | Oncology | KLK8 | O60259 | 19 | 50996007 | 51002711 | 1.02 | 0.87 | 1.20 | 7.90E-01 | 0.924 |
| OID30199 | MYBPC2 | Cardiometabolic_II | MYBPC2 | Q14324 | 19 | 50432892 | 50466321 | 0.98 | 0.81 | 1.18 | 7.93E-01 | 0.924 |
| OID20962 | PRDX1 | Neurology | PRDX1 | Q06830 | 1 | 45510914 | 45542732 | 0.98 | 0.83 | 1.16 | 7.92E-01 | 0.924 |
| OID20986 | RBKS | Neurology | RBKS | Q9H477 | 2 | 27781379 | 27890681 | 0.98 | 0.82 | 1.17 | 7.94E-01 | 0.924 |
| OID20844 | TBC1D17 | Neurology | TBC1D17 | Q9HA65 | 19 | 49877666 | 49888750 | 0.98 | 0.82 | 1.17 | 7.90E-01 | 0.924 |
| OID30478 | TSC1 | Inflammation_II | TSC1 | Q92574 | 9 | 132891348 | 132946874 | 0.98 | 0.82 | 1.17 | 7.90E-01 | 0.924 |
| OID21381 | VMO1 | Oncology | VMO1 | Q7Z5L0 | 17 | 4785285 | 4786433 | 1.02 | 0.88 | 1.19 | 7.89E-01 | 0.924 |
| OID21144 | VSIG4 | Neurology | VSIG4 | Q9Y279 | X | 66021738 | 66040125 | 1.02 | 0.87 | 1.20 | 7.93E-01 | 0.924 |
| OID21421 | VWA1 | Oncology | VWA1 | Q6PCB0 | 1 | 1434861 | 1442882 | 1.02 | 0.86 | 1.21 | 7.92E-01 | 0.924 |
| OID31256 | CD82 | Oncology_II | CD82 | P27701 | 11 | 44564427 | 44620358 | 0.98 | 0.83 | 1.16 | 7.95E-01 | 0.924 |
| OID20231 | THBD | Cardiometabolic | THBD | P07204 | 20 | 23045633 | 23049672 | 1.02 | 0.87 | 1.19 | 7.96E-01 | 0.924 |
| OID20695 | CCL4 | Inflammation | CCL4 | P13236 | 17 | 36103827 | 36105621 | 0.98 | 0.83 | 1.15 | 7.96E-01 | 0.925 |
| OID20963 | LPO | Neurology | LPO | P22079 | 17 | 58218548 | 58268518 | 0.98 | 0.83 | 1.16 | 7.98E-01 | 0.926 |
| OID30061 | MYH4 | Cardiometabolic_II | MYH4 | Q9Y623 | 17 | 10443290 | 10469559 | 0.98 | 0.83 | 1.16 | 7.99E-01 | 0.927 |
| OID21149 | CA2 | Neurology | CA2 | P00918 | 8 | 85463968 | 85481493 | 0.98 | 0.82 | 1.16 | 7.99E-01 | 0.927 |
| OID30230 | IDO1 | Cardiometabolic_II/Infla | IDO1 | P14902 | 8 | 39902275 | 39928790 | 0.98 | 0.82 | 1.16 | 8.00E-01 | 0.928 |
| OID21481 | C4BPB | Oncology | C4BPB | P20851 | 1 | 207088860 | 207099993 | 1.02 | 0.86 | 1.21 | 8.00E-01 | 0.928 |
| OID20628 | CD244 | Inflammation | CD244 | Q9BZW8 | 1 | 160830160 | 160862887 | 0.98 | 0.83 | 1.15 | 8.02E-01 | 0.929 |
| OID21302 | RP2 | Oncology | RP2 | O75695 | X | 46837043 | 46882358 | 0.98 | 0.83 | 1.16 | 8.02E-01 | 0.929 |
| OID31228 | PAIP2B | Oncology_II | PAIP2B | Q9ULR5 | 2 | 71182738 | 71227103 | 1.02 | 0.87 | 1.20 | 8.03E-01 | 0.929 |
| OID31182 | CLINT1 | Oncology_II | CLINT1 | Q14677 | 5 | 157785743 | 157859145 | 0.98 | 0.81 | 1.18 | 8.04E-01 | 0.931 |
| OID21499 | KLK1 | Oncology | KLK1 | P06870 | 19 | 50819146 | 50823787 | 1.02 | 0.87 | 1.20 | 8.04E-01 | 0.931 |
| OID21044 | GOLM2 | Neurology | GOLM2 | Q6P4E1 | 15 | 44288719 | 44415758 | 1.02 | 0.86 | 1.21 | 8.05E-01 | 0.931 |
| OID30793 | AGBL2 | Neurology_II | AGBL2 | Q5U5Z8 | 11 | 47659591 | 47715389 | 0.98 | 0.84 | 1.14 | 8.10E-01 | 0.932 |
| OID20407 | ANGPTL3 | Cardiometabolic | ANGPTL3 | Q9Y5C1 | 1 | 62597520 | 62606313 | 0.98 | 0.83 | 1.16 | 8.11E-01 | 0.932 |
| OID21141 | BLVRB | Neurology | BLVRB | P30043 | 19 | 40447765 | 40465764 | 0.98 | 0.82 | 1.16 | 8.10E-01 | 0.932 |
| OID20626 | BSG | Inflammation | BSG | P35613 | 19 | 571277 | 583494 | 1.02 | 0.87 | 1.20 | 8.09E-01 | 0.932 |
| OID20713 | BTN2A1 | Inflammation | BTN2A1 | Q7KYR7 | 6 | 26457904 | 26476621 | 1.02 | 0.87 | 1.19 | 8.08E-01 | 0.932 |
| OID20540 | BTN3A2 | Inflammation | BTN3A2 | P78410 | 6 | 26365159 | 26378320 | 0.98 | 0.84 | 1.15 | 8.11E-01 | 0.932 |
| OID20367 | C1QTNF1 | Cardiometabolic | C1QTNF1 | Q9BXJ1 | 17 | 79022814 | 79049798 | 0.98 | 0.83 | 1.16 | 8.06E-01 | 0.932 |
| OID20765 | CCL22 | Inflammation | CCL22 | O00626 | 16 | 57358783 | 57366189 | 0.98 | 0.83 | 1.16 | 8.10E-01 | 0.932 |
| OID21112 | CLEC11A | Neurology | CLEC11A | Q9Y240 | 19 | 50723364 | 50725708 | 0.98 | 0.84 | 1.15 | 8.11E-01 | 0.932 |
| OID31365 | EDDM3B | Oncology_II | EDDM3B | P56851 | 14 | 20768404 | 20770948 | 0.97 | 0.74 | 1.27 | 8.10E-01 | 0.932 |
| OID20095 | GPR37 | Cardiometabolic | GPR37 | O15354 | 7 | 124743885 | 124765792 | 0.98 | 0.84 | 1.15 | 8.08E-01 | 0.932 |
| OID30813 | KCNC4 | Neurology_II | KCNC4 | Q03721 | 1 | 110210314 | 110283100 | 1.02 | 0.88 | 1.18 | 8.09E-01 | 0.932 |
| OID30665 | OLFM4 | Inflammation_II | OLFM4 | Q6UX06 | 13 | 53028813 | 53052057 | 1.02 | 0.86 | 1.22 | 8.11E-01 | 0.932 |
| OID20776 | OSCAR | Inflammation | OSCAR | Q8IYS5 | 19 | 54094668 | 54102692 | 0.98 | 0.84 | 1.15 | 8.06E-01 | 0.932 |
| OID30439 | SIRT1 | Inflammation_II | SIRT1 | Q96EB6 | 10 | 67884656 | 67918390 | 0.98 | 0.82 | 1.17 | 8.08E-01 | 0.932 |
| OID31183 | STOML2 | Oncology_II | STOML2 | Q9UJZ1 | 9 | 35099776 | 35103195 | 0.98 | 0.83 | 1.16 | 8.10E-01 | 0.932 |
| OID21219 | STX6 | Oncology | STX6 | O43752 | 1 | 180972712 | 181023121 | 1.02 | 0.87 | 1.20 | 8.08E-01 | 0.932 |
| OID31103 | TMPRSS11D | Neurology_II | TMPRSS11D | O60235 | 4 | 67820876 | 67884002 | 1.02 | 0.87 | 1.19 | 8.09E-01 | 0.932 |
| OID31044 | EFHD1 | Neurology_II | EFHD1 | Q9BUP0 | 2 | 232606057 | 232682776 | 0.98 | 0.83 | 1.16 | 8.12E-01 | 0.932 |
| OID31504 | SERPINI2 | Oncology_II | SERPINI2 | O75830 | 3 | 167441914 | 167478972 | 0.98 | 0.82 | 1.17 | 8.12E-01 | 0.932 |
| OID30054 | TRAF3IP2 | Cardiometabolic_II | TRAF3IP2 | O43734 | 6 | 111555381 | 111606906 | 1.02 | 0.85 | 1.24 | 8.12E-01 | 0.932 |
| OID21178 | ATP6AP2 | Oncology | ATP6AP2 | O75787 | X | 40579372 | 40606848 | 0.98 | 0.82 | 1.17 | 8.15E-01 | 0.933 |
| OID20414 | EFEMP1 | Cardiometabolic | EFEMP1 | Q12805 | 2 | 55865967 | 55924139 | 1.02 | 0.86 | 1.21 | 8.14E-01 | 0.933 |
| OID21510 | ERBB4 | Oncology | ERBB4 | Q15303 | 2 | 211375717 | 212538841 | 1.02 | 0.86 | 1.21 | 8.15E-01 | 0.933 |
| OID30148 | GUK1 | Cardiometabolic_II | GUK1 | Q16774 | 1 | 228139962 | 228148984 | 1.02 | 0.87 | 1.19 | 8.15E-01 | 0.933 |
| OID20662 | VEGFD | Inflammation | VEGFD | O43915 | X | 15345596 | 15384413 | 1.02 | 0.86 | 1.21 | 8.14E-01 | 0.933 |
| OID20573 | CLEC4A | Inflammation | CLEC4A | Q9UMR7 | 12 | 8123617 | 8138607 | 0.98 | 0.83 | 1.16 | 8.16E-01 | 0.933 |
| OID30235 | RNF149 | Cardiometabolic_II | RNF149 | Q8NC42 | 2 | 101271219 | 101308701 | 0.98 | 0.84 | 1.15 | 8.18E-01 | 0.936 |
| OID30089 | NECAP2 | Cardiometabolic_II | NECAP2 | Q9NVZ3 | 1 | 16440721 | 16460078 | 0.98 | 0.83 | 1.16 | 8.19E-01 | 0.936 |
| OID21117 | CNTN4 | Neurology | CNTN4 | Q8IWV2 | 3 | 2098813 | 3057959 | 1.02 | 0.87 | 1.19 | 8.20E-01 | 0.936 |
| OID31266 | TET2 | Oncology_II | TET2 | Q6N021 | 4 | 105145875 | 105279816 | 1.02 | 0.88 | 1.18 | 8.20E-01 | 0.936 |
| OID20794 | PTEN | Neurology | PTEN | P60484 | 10 | 87862638 | 87971930 | 0.98 | 0.84 | 1.15 | 8.21E-01 | 0.938 |
| OID20169 | SDC1 | Cardiometabolic | SDC1 | P18827 | 2 | 20200797 | 20225433 | 0.98 | 0.83 | 1.16 | 8.22E-01 | 0.938 |
| OID30937 | TARM1 | Neurology_II | TARM1 | B6A8C7 | 19 | 54069895 | 54081365 | 0.98 | 0.83 | 1.16 | 8.22E-01 | 0.938 |
| OID30343 | PTPRZ1 | Cardiometabolic_II | PTPRZ1 | P23471 | 7 | 121873089 | 122062036 | 1.02 | 0.85 | 1.22 | 8.23E-01 | 0.939 |
| OID20797 | COPE | Neurology | COPE | O14579 | 19 | 18899514 | 18919387 | 1.02 | 0.87 | 1.20 | 8.24E-01 | 0.939 |
| OID30372 | DDT | Cardiometabolic_II | DDT | P30046 | 22 | 23970542 | 23980525 | 0.98 | 0.82 | 1.17 | 8.24E-01 | 0.939 |
| OID31402 | LUZP2 | Oncology_II | LUZP2 | Q86TE4 | 11 | 24496970 | 25082638 | 1.02 | 0.88 | 1.17 | 8.24E-01 | 0.939 |
| OID20075 | CA5A | Cardiometabolic | CA5A | P35218 | 16 | 87881546 | 87936580 | 0.98 | 0.82 | 1.17 | 8.25E-01 | 0.939 |
| OID30822 | DNLZ | Neurology_II | DNLZ | Q5SXM8 | 9 | 136359480 | 136363744 | 1.02 | 0.86 | 1.20 | 8.26E-01 | 0.940 |
| OID21158 | IGFBP4 | Neurology | IGFBP4 | P22692 | 17 | 40443450 | 40457725 | 1.02 | 0.87 | 1.20 | 8.26E-01 | 0.940 |
| OID31154 | SMTN | Neurology_II | SMTN | P53814 | 22 | 31064105 | 31104757 | 0.98 | 0.83 | 1.16 | 8.25E-01 | 0.940 |
| OID21052 | TNFRSF21 | Neurology | TNFRSF21 | O75509 | 6 | 47231532 | 47309905 | 1.02 | 0.87 | 1.19 | 8.26E-01 | 0.940 |
| OID30114 | EXTL1 | Cardiometabolic_II | EXTL1 | Q92935 | 1 | 26019884 | 26036464 | 0.98 | 0.84 | 1.15 | 8.27E-01 | 0.940 |
| OID21101 | GUCA2A | Neurology | GUCA2A | Q02747 | 1 | 42162690 | 42164745 | 1.02 | 0.87 | 1.18 | 8.28E-01 | 0.941 |
| OID30405 | CLEC4M | Cardiometabolic_II | CLEC4M | Q9H2X3 | 19 | 7763210 | 7769605 | 1.02 | 0.86 | 1.20 | 8.29E-01 | 0.941 |
| OID20541 | MLN | Inflammation | MLN | P12872 | 6 | 33794673 | 33804003 | 0.98 | 0.82 | 1.17 | 8.28E-01 | 0.941 |
| OID21380 | MSLN | Oncology | MSLN | Q13421 | 16 | 760734 | 768865 | 1.02 | 0.86 | 1.20 | 8.29E-01 | 0.941 |

| | | | | | | | | | | | | |
|---|---|---|---|---|---|---|---|---|---|---|---|---|
| OID30131 | MYOM2 | Cardiometabolic_II | MYOM2 | P54296 | 8 | 2045046 | 2165552 | 0.98 | 0.84 | 1.15 | 8.29E-01 | 0.941 |
| OID21081 | NRP2 | Neurology | NRP2 | O60462 | 2 | 205681990 | 205798133 | 0.98 | 0.83 | 1.16 | 8.30E-01 | 0.941 |
| OID21312 | IGSF3 | Oncology | IGSF3 | O75054 | 1 | 116574399 | 116667755 | 0.98 | 0.82 | 1.17 | 8.30E-01 | 0.941 |
| OID21445 | ARG1 | Oncology | ARG1 | P05089 | 6 | 131470832 | 131584332 | 0.98 | 0.83 | 1.16 | 8.32E-01 | 0.942 |
| OID30213 | DMP1 | Cardiometabolic_II | DMP1 | Q13316 | 4 | 87650280 | 87664361 | 0.98 | 0.83 | 1.16 | 8.34E-01 | 0.943 |
| OID20884 | FMNL1 | Neurology | FMNL1 | O95466 | 17 | 45221444 | 45247319 | 0.98 | 0.82 | 1.17 | 8.34E-01 | 0.943 |
| OID30919 | HNF1A | Neurology_II | HNF1A | P20823 | 12 | 120978543 | 121002512 | 0.98 | 0.82 | 1.17 | 8.34E-01 | 0.943 |
| OID30900 | NLGN1 | Neurology_II | NLGN1 | Q8N2Q7 | 3 | 173396284 | 174294372 | 0.98 | 0.82 | 1.18 | 8.34E-01 | 0.943 |
| OID20320 | QPCT | Cardiometabolic | QPCT | Q16769 | 2 | 37342827 | 37373322 | 1.02 | 0.86 | 1.20 | 8.34E-01 | 0.943 |
| OID20321 | RNASET2 | Cardiometabolic | RNASET2 | O00584 | 6 | 166922113 | 166957191 | 0.98 | 0.83 | 1.17 | 8.33E-01 | 0.943 |
| OID20403 | ANG | Cardiometabolic | ANG | P03950 | 14 | 20684177 | 20698971 | 0.98 | 0.83 | 1.16 | 8.36E-01 | 0.943 |
| OID21261 | CFC1 | Oncology | CFC1 | P0CG37 | 2 | 130592165 | 130599575 | 1.02 | 0.86 | 1.20 | 8.37E-01 | 0.943 |
| OID30492 | CSNK1D | Inflammation_II | CSNK1D | P48730 | 17 | 82239023 | 82273700 | 1.02 | 0.86 | 1.20 | 8.37E-01 | 0.943 |
| OID30363 | CTLA4 | Cardiometabolic_II | CTLA4 | P16410 | 2 | 203853888 | 203873965 | 0.98 | 0.83 | 1.16 | 8.35E-01 | 0.943 |
| OID20557 | MAPK9 | Inflammation | MAPK9 | P45984 | 5 | 180233143 | 180292099 | 0.98 | 0.83 | 1.16 | 8.36E-01 | 0.943 |
| OID20574 | OSM | Inflammation | OSM | P13725 | 22 | 30262829 | 30266851 | 0.98 | 0.83 | 1.16 | 8.36E-01 | 0.943 |
| OID20337 | PRCP | Cardiometabolic | PRCP | P42785 | 11 | 82822936 | 82970584 | 1.02 | 0.85 | 1.22 | 8.36E-01 | 0.943 |
| OID20129 | TSHB | Cardiometabolic | TSHB | P01222 | 1 | 115029826 | 115034302 | 1.02 | 0.87 | 1.18 | 8.36E-01 | 0.943 |
| OID20882 | WASF3 | Neurology | WASF3 | Q9UPY6 | 13 | 26557683 | 26688948 | 1.02 | 0.87 | 1.18 | 8.37E-01 | 0.943 |
| OID20448 | NCLN | Inflammation | NCLN | Q969V3 | 19 | 3185563 | 3209575 | 1.02 | 0.86 | 1.20 | 8.37E-01 | 0.943 |
| OID21478 | FGFR2 | Oncology | FGFR2 | P21802 | 10 | 121478332 | 121598458 | 1.02 | 0.86 | 1.21 | 8.38E-01 | 0.943 |
| OID30992 | FCER1A | Neurology_II | FCER1A | P12319 | 1 | 159289714 | 159308224 | 1.01 | 0.88 | 1.17 | 8.39E-01 | 0.944 |
| OID20312 | CHIT1 | Cardiometabolic | CHIT1 | Q13231 | 1 | 203212827 | 203273641 | 1.02 | 0.86 | 1.20 | 8.40E-01 | 0.944 |
| OID20336 | BLMH | Cardiometabolic | BLMH | Q13867 | 17 | 30248203 | 30292056 | 1.02 | 0.87 | 1.19 | 8.41E-01 | 0.946 |
| OID20113 | CTSH | Cardiometabolic | CTSH | P09668 | 15 | 78921058 | 78949574 | 0.98 | 0.84 | 1.16 | 8.42E-01 | 0.946 |
| OID21134 | MFGE8 | Neurology | MFGE8 | Q08431 | 15 | 88898683 | 88913381 | 0.98 | 0.83 | 1.16 | 8.43E-01 | 0.946 |
| OID31211 | MLLT1 | Oncology_II | MLLT1 | Q03111 | 19 | 6210381 | 6279975 | 1.02 | 0.86 | 1.20 | 8.42E-01 | 0.946 |
| OID20723 | SCGB3A2 | Inflammation | SCGB3A2 | Q96PL1 | 5 | 147870682 | 147882191 | 1.02 | 0.87 | 1.19 | 8.44E-01 | 0.947 |
| OID20880 | PTK7 | Neurology | PTK7 | Q13308 | 6 | 43076307 | 43161719 | 0.98 | 0.83 | 1.16 | 8.45E-01 | 0.948 |
| OID21529 | FOLR1 | Oncology | FOLR1 | P15328 | 11 | 72189558 | 72196323 | 0.98 | 0.83 | 1.16 | 8.46E-01 | 0.948 |
| OID30413 | GM2A | Cardiometabolic_II | GM2A | P17900 | 5 | 151212150 | 151270440 | 0.98 | 0.84 | 1.15 | 8.46E-01 | 0.948 |
| OID20591 | NTF3 | Inflammation | NTF3 | P20783 | 12 | 5432108 | 5521536 | 0.98 | 0.83 | 1.16 | 8.46E-01 | 0.948 |
| OID31105 | VSNL1 | Neurology_II | VSNL1 | P62760 | 2 | 17539126 | 17657018 | 1.02 | 0.86 | 1.20 | 8.46E-01 | 0.948 |
| OID30458 | CEBPA | Inflammation_II | CEBPA | P49715 | 19 | 33299934 | 33302534 | 1.01 | 0.87 | 1.18 | 8.49E-01 | 0.949 |
| OID20294 | CSTB | Cardiometabolic | CSTB | P04080 | 21 | 43772511 | 43776330 | 1.02 | 0.85 | 1.22 | 8.48E-01 | 0.949 |
| OID30430 | FOLH1 | Inflammation_II | FOLH1 | Q04609 | 11 | 49145092 | 49208638 | 1.02 | 0.86 | 1.20 | 8.50E-01 | 0.949 |
| OID31187 | GJA8 | Oncology_II | GJA8 | P48165 | 1 | 147902795 | 147909269 | 0.98 | 0.83 | 1.16 | 8.49E-01 | 0.949 |
| OID30802 | GRIN2B | Neurology_II | GRIN2B | Q13224 | 12 | 13437942 | 13982002 | 1.01 | 0.88 | 1.17 | 8.49E-01 | 0.949 |
| OID21136 | IL7R | Neurology | IL7R | P16871 | 5 | 35852695 | 35879603 | 0.98 | 0.83 | 1.17 | 8.48E-01 | 0.949 |
| OID21208 | NTF4 | Oncology | NTF4 | P34130 | 19 | 49061066 | 49065054 | 1.02 | 0.85 | 1.21 | 8.49E-01 | 0.949 |
| OID21487 | PODXL2 | Oncology | PODXL2 | Q9NZ53 | 3 | 127629185 | 127672802 | 1.02 | 0.87 | 1.19 | 8.49E-01 | 0.949 |
| OID21321 | TAFA5 | Oncology | TAFA5 | Q7Z5A7 | 22 | 48489553 | 48850912 | 1.01 | 0.87 | 1.18 | 8.50E-01 | 0.949 |
| OID20939 | WFIKKN1 | Neurology | WFIKKN1 | Q96NZ8 | 16 | 629239 | 634117 | 0.98 | 0.84 | 1.16 | 8.50E-01 | 0.949 |
| OID30550 | PDZK1 | Inflammation_II | PDZK1 | Q5T2W1 | 1 | 145670852 | 145708148 | 1.02 | 0.86 | 1.20 | 8.52E-01 | 0.950 |
| OID30126 | DDA1 | Cardiometabolic_II | DDA1 | Q9BW61 | 19 | 17309518 | 17323298 | 0.99 | 0.84 | 1.15 | 8.53E-01 | 0.951 |
| OID20450 | IL2RB | Inflammation | IL2RB | P14784 | 22 | 37125843 | 37175054 | 0.99 | 0.85 | 1.15 | 8.53E-01 | 0.951 |
| OID20668 | CCL11 | Inflammation | CCL11 | P51671 | 17 | 34285742 | 34288334 | 1.02 | 0.85 | 1.21 | 8.54E-01 | 0.951 |
| OID21142 | RELT | Neurology | RELT | Q969Z4 | 11 | 73376399 | 73397474 | 0.98 | 0.83 | 1.17 | 8.54E-01 | 0.951 |
| OID20834 | NOS3 | Neurology | NOS3 | P29474 | 7 | 150991017 | 151014588 | 1.02 | 0.86 | 1.21 | 8.54E-01 | 0.951 |
| OID20157 | CDH6 | Cardiometabolic | CDH6 | P55285 | 5 | 31193686 | 31329146 | 1.02 | 0.86 | 1.19 | 8.55E-01 | 0.952 |
| OID20300 | HYOU1 | Cardiometabolic | HYOU1 | Q9Y4L1 | 11 | 119044188 | 119057227 | 1.01 | 0.88 | 1.17 | 8.55E-01 | 0.952 |
| OID20080 | MTPN | Cardiometabolic | MTPN | P58546 | 7 | 135926760 | 135977359 | 0.99 | 0.84 | 1.16 | 8.55E-01 | 0.952 |
| OID20404 | FETUB | Cardiometabolic | FETUB | Q9UGM5 | 3 | 186635969 | 186653141 | 0.98 | 0.84 | 1.16 | 8.56E-01 | 0.952 |
| OID21433 | HBQ1 | Oncology | HBQ1 | P09105 | 16 | 180459 | 181179 | 1.02 | 0.85 | 1.21 | 8.56E-01 | 0.952 |
| OID31435 | PBXIP1 | Oncology_II | PBXIP1 | Q96AQ6 | 1 | 154944076 | 154956123 | 0.99 | 0.84 | 1.16 | 8.57E-01 | 0.952 |
| OID20673 | PGF | Inflammation | PGF | P49763 | 14 | 74941834 | 74955626 | 0.98 | 0.82 | 1.18 | 8.57E-01 | 0.952 |
| OID30691 | TGOLN2 | Inflammation_II | TGOLN2 | O43493 | 2 | 85318027 | 85328251 | 1.01 | 0.87 | 1.19 | 8.58E-01 | 0.953 |
| OID20814 | SLC27A4 | Neurology | SLC27A4 | Q6P1M0 | 9 | 128340527 | 128361470 | 0.98 | 0.83 | 1.17 | 8.60E-01 | 0.955 |
| OID30810 | TSPAN7 | Neurology_II | TSPAN7 | P41732 | X | 38561542 | 38688920 | 1.02 | 0.85 | 1.22 | 8.61E-01 | 0.955 |
| OID30176 | CSRP3 | Cardiometabolic_II | CSRP3 | P50461 | 11 | 19182030 | 19210571 | 0.99 | 0.83 | 1.16 | 8.61E-01 | 0.955 |
| OID31451 | ATRAID | Oncology_II | ATRAID | Q6UW56 | 2 | 27212041 | 27217178 | 0.99 | 0.84 | 1.16 | 8.64E-01 | 0.956 |
| OID21299 | DDAH1 | Oncology | DDAH1 | O94760 | 1 | 85318481 | 85578363 | 1.02 | 0.86 | 1.20 | 8.62E-01 | 0.956 |
| OID31318 | GUCY2C | Oncology_II | GUCY2C | P25092 | 12 | 14612632 | 14696599 | 0.98 | 0.83 | 1.17 | 8.63E-01 | 0.956 |
| OID20214 | SFTPD | Cardiometabolic | SFTPD | P35247 | 10 | 79937467 | 79982614 | 0.99 | 0.83 | 1.17 | 8.64E-01 | 0.956 |
| OID20985 | TNFRSF9 | Neurology | TNFRSF9 | Q07011 | 1 | 7915871 | 7943165 | 1.01 | 0.87 | 1.19 | 8.64E-01 | 0.956 |
| OID30446 | YY1 | Inflammation_II | YY1 | P25490 | 14 | 100238298 | 100282788 | 0.99 | 0.83 | 1.16 | 8.62E-01 | 0.956 |
| OID20569 | CCL28 | Inflammation | CCL28 | Q9NRJ3 | 5 | 43376645 | 43412391 | 0.99 | 0.84 | 1.16 | 8.66E-01 | 0.956 |
| OID30094 | ERVV-1 | Cardiometabolic_II | ERVV-1 | B6SEH8 | 19 | 53013921 | 53016123 | 0.98 | 0.83 | 1.18 | 8.65E-01 | 0.956 |
| OID20770 | FST | Inflammation | FST | P19883 | 5 | 53480626 | 53487134 | 1.01 | 0.86 | 1.20 | 8.66E-01 | 0.956 |
| OID21258 | PLXDC1 | Oncology | PLXDC1 | Q8IUK5 | 17 | 39063313 | 39154394 | 1.01 | 0.87 | 1.18 | 8.66E-01 | 0.956 |
| OID31124 | PRND | Neurology_II | PRND | Q9UKY0 | 20 | 4721909 | 4728460 | 0.99 | 0.85 | 1.14 | 8.67E-01 | 0.956 |
| OID20817 | RAB6B | Neurology | RAB6B | Q9NRW1 | 3 | 133824235 | 133895882 | 1.01 | 0.85 | 1.21 | 8.67E-01 | 0.956 |
| OID30958 | RAC3 | Neurology_II | RAC3 | P60763 | 17 | 82031678 | 82034204 | 0.99 | 0.84 | 1.16 | 8.67E-01 | 0.956 |
| OID30333 | SCRG1 | Cardiometabolic_II | SCRG1 | O75711 | 4 | 173384701 | 173406380 | 0.99 | 0.83 | 1.17 | 8.66E-01 | 0.956 |
| OID20783 | TNFRSF14 | Inflammation | TNFRSF14 | Q92956 | 1 | 2555639 | 2565382 | 0.99 | 0.83 | 1.17 | 8.67E-01 | 0.956 |
| OID30818 | FZD8 | Neurology_II | FZD8 | Q9H461 | 10 | 35638247 | 35642296 | 0.99 | 0.84 | 1.16 | 8.67E-01 | 0.956 |
| OID30734 | CPB2 | Inflammation_II | CPB2 | Q96IY4 | 13 | 46053186 | 46105033 | 1.01 | 0.86 | 1.20 | 8.68E-01 | 0.956 |
| OID30735 | ATRN | Inflammation_II | ATRN | O75882 | 20 | 3471018 | 3651118 | 1.01 | 0.85 | 1.20 | 8.69E-01 | 0.957 |
| OID20565 | CD83 | Inflammation | CD83 | Q01151 | 6 | 14117256 | 14136918 | 1.01 | 0.86 | 1.19 | 8.69E-01 | 0.957 |
| OID30137 | DHODH | Cardiometabolic_II | DHODH | Q02127 | 16 | 72008588 | 72027664 | 0.99 | 0.83 | 1.17 | 8.70E-01 | 0.957 |
| OID20246 | IGFBP1 | Cardiometabolic | IGFBP1 | P08833 | 7 | 45888360 | 45893660 | 0.99 | 0.82 | 1.18 | 8.69E-01 | 0.957 |
| OID20651 | ADAM23 | Inflammation | ADAM23 | O75077 | 2 | 206443532 | 206621127 | 0.99 | 0.84 | 1.16 | 8.71E-01 | 0.957 |
| OID30873 | BHLHE40 | Neurology_II | BHLHE40 | O14503 | 3 | 4979437 | 4985323 | 0.99 | 0.84 | 1.16 | 8.71E-01 | 0.957 |
| OID21157 | STC2 | Neurology | STC2 | O76061 | 5 | 173314723 | 173328447 | 0.99 | 0.83 | 1.17 | 8.70E-01 | 0.957 |
| OID20183 | OLR1 | Cardiometabolic | OLR1 | P78380 | 12 | 10158301 | 10172138 | 0.99 | 0.83 | 1.17 | 8.71E-01 | 0.957 |
| OID20427 | IL1B | Inflammation | IL1B | P01584 | 2 | 112829751 | 112836816 | 0.99 | 0.84 | 1.16 | 8.72E-01 | 0.957 |
| OID21228 | CLEC6A | Oncology | CLEC6A | Q6EIG7 | 12 | 8455962 | 8478330 | 1.01 | 0.86 | 1.19 | 8.72E-01 | 0.957 |
| OID30836 | DOCK9 | Neurology_II | DOCK9 | Q9BZ29 | 13 | 98793429 | 99086625 | 1.01 | 0.87 | 1.18 | 8.73E-01 | 0.958 |
| OID30553 | CSF2RB | Inflammation_II | CSF2RB | P32927 | 22 | 36913628 | 36940439 | 0.99 | 0.83 | 1.17 | 8.73E-01 | 0.958 |
| OID31427 | KLK15 | Oncology_II | KLK15 | Q9H2R5 | 19 | 50825289 | 50837213 | 1.01 | 0.86 | 1.19 | 8.74E-01 | 0.958 |
| OID20233 | FAM3C | Cardiometabolic | FAM3C | Q92520 | 7 | 121348878 | 121396364 | 1.01 | 0.86 | 1.20 | 8.75E-01 | 0.959 |
| OID30051 | BHMT2 | Cardiometabolic_II | BHMT2 | Q9H2M3 | 5 | 79069767 | 79090069 | 0.99 | 0.84 | 1.17 | 8.75E-01 | 0.959 |
| OID30192 | ACTN2 | Cardiometabolic_II | ACTN2 | P35609 | 1 | 236664141 | 236764631 | 1.01 | 0.86 | 1.20 | 8.76E-01 | 0.959 |
| OID30843 | CSPG5 | Neurology_II | CSPG5 | O95196 | 3 | 47562238 | 47580792 | 0.99 | 0.86 | 1.13 | 8.77E-01 | 0.959 |
| OID31199 | GAGE2A | Oncology_II | GAGE2A | Q6NT46 | X | 49589496 | 49596824 | 0.99 | 0.84 | 1.16 | 8.76E-01 | 0.959 |
| OID20785 | WFIKKN2 | Inflammation | WFIKKN2 | Q8TEU8 | 17 | 50834650 | 50842353 | 0.99 | 0.83 | 1.17 | 8.77E-01 | 0.959 |
| OID20747 | CRHBP | Inflammation | CRHBP | P24387 | 5 | 76953045 | 76981158 | 1.01 | 0.85 | 1.21 | 8.79E-01 | 0.960 |
| OID20210 | MARCO | Cardiometabolic | MARCO | Q9UEW3 | 2 | 118942194 | 118994660 | 1.01 | 0.86 | 1.20 | 8.78E-01 | 0.960 |
| OID20571 | PTH1R | Inflammation | PTH1R | Q03431 | 3 | 46877721 | 46903799 | 1.01 | 0.85 | 1.21 | 8.79E-01 | 0.960 |
| OID20479 | WAS | Inflammation | WAS | P42768 | X | 48676596 | 48691427 | 1.01 | 0.86 | 1.20 | 8.79E-01 | 0.960 |
| OID21307 | MME | Oncology | MME | P08473 | 3 | 155024124 | 155183704 | 0.99 | 0.84 | 1.16 | 8.80E-01 | 0.960 |
| OID31158 | PBK | Oncology_II | PBK | Q96KB5 | 8 | 27809624 | 27838082 | 1.01 | 0.86 | 1.19 | 8.80E-01 | 0.960 |
| OID21284 | SERPINA9 | Oncology | SERPINA9 | Q86WD7 | 14 | 94462717 | 94479689 | 1.01 | 0.87 | 1.18 | 8.81E-01 | 0.960 |

| | | | | | | | | | | | | |
|---|---|---|---|---|---|---|---|---|---|---|---|---|
| OID21488 | SEZ6L | Oncology | SEZ6L | Q9BYH1 | 22 | 26169462 | 26383597 | 0.99 | 0.83 | 1.17 | 8.81E-01 | 0.960 |
| OID30477 | SMPD3 | Inflammation_II | SMPD3 | Q9NY59 | 16 | 68358327 | 68448508 | 1.01 | 0.86 | 1.20 | 8.81E-01 | 0.960 |
| OID20116 | BOC | Cardiometabolic | BOC | Q9BWV1 | 3 | 113211003 | 113287459 | 0.99 | 0.83 | 1.17 | 8.84E-01 | 0.962 |
| OID20636 | CLEC7A | Inflammation | CLEC7A | Q9BXN2 | 12 | 10116777 | 10130258 | 0.99 | 0.83 | 1.17 | 8.83E-01 | 0.962 |
| OID21474 | DCBLD2 | Oncology | DCBLD2 | Q96PD2 | 3 | 98795941 | 98901695 | 1.01 | 0.86 | 1.19 | 8.83E-01 | 0.962 |
| OID21262 | SORCS2 | Oncology | SORCS2 | Q96PQ0 | 4 | 7192538 | 7742836 | 0.99 | 0.84 | 1.16 | 8.83E-01 | 0.962 |
| OID20612 | TLR3 | Inflammation | TLR3 | O15455 | 4 | 186069155 | 186088073 | 1.01 | 0.86 | 1.19 | 8.83E-01 | 0.962 |
| OID31361 | IL2RG | Oncology_II | IL2RG | P31785 | X | 71107404 | 71112108 | 0.99 | 0.86 | 1.14 | 8.84E-01 | 0.962 |
| OID20841 | GHRHR | Neurology | GHRHR | Q02643 | 7 | 30938669 | 30993254 | 1.01 | 0.87 | 1.17 | 8.86E-01 | 0.963 |
| OID30479 | TRAF3 | Inflammation_II | TRAF3 | Q13114 | 14 | 102777449 | 102911500 | 0.99 | 0.83 | 1.18 | 8.86E-01 | 0.963 |
| OID30115 | CEP112 | Cardiometabolic_II | CEP112 | Q8N8E3 | 17 | 65635537 | 66192133 | 1.01 | 0.87 | 1.18 | 8.86E-01 | 0.963 |
| OID20126 | CLC | Cardiometabolic | CLC | Q05315 | 19 | 39731255 | 39738029 | 0.99 | 0.83 | 1.18 | 8.88E-01 | 0.963 |
| OID30882 | DTX2 | Neurology_II | DTX2 | Q86UW9 | 7 | 76461676 | 76505995 | 0.99 | 0.83 | 1.18 | 8.88E-01 | 0.963 |
| OID20743 | EPCAM | Inflammation | EPCAM | P16422 | 2 | 47345158 | 47387601 | 1.01 | 0.86 | 1.19 | 8.88E-01 | 0.963 |
| OID20316 | FAS | Cardiometabolic | FAS | P25445 | 10 | 88953813 | 89029605 | 1.01 | 0.85 | 1.20 | 8.88E-01 | 0.963 |
| OID20431 | IL10 | Inflammation | IL10 | P22301 | 1 | 206767602 | 206774541 | 0.99 | 0.86 | 1.14 | 8.89E-01 | 0.963 |
| OID30346 | LILRA6 | Cardiometabolic_II | LILRA6 | Q6PI73 | 19 | 54236592 | 54242791 | 0.99 | 0.84 | 1.16 | 8.89E-01 | 0.963 |
| OID21473 | PLA2G15 | Oncology | PLA2G15 | Q8NCC3 | 16 | 68245304 | 68261058 | 0.99 | 0.84 | 1.17 | 8.89E-01 | 0.963 |
| OID30627 | SPINK2 | Inflammation_II | SPINK2 | P20155 | 4 | 56809860 | 56821742 | 1.01 | 0.85 | 1.21 | 8.88E-01 | 0.963 |
| OID21272 | FLT3 | Oncology | FLT3 | P36888 | 13 | 28003274 | 28100592 | 1.01 | 0.87 | 1.18 | 8.90E-01 | 0.964 |
| OID30729 | QSOX1 | Inflammation_II | QSOX1 | O00391 | 1 | 180154869 | 180204030 | 0.99 | 0.81 | 1.20 | 8.90E-01 | 0.964 |
| OID20423 | EIF5A | Inflammation | EIF5A | P63241 | 17 | 7306999 | 7312463 | 1.01 | 0.85 | 1.20 | 8.91E-01 | 0.964 |
| OID31501 | LMNB1 | Oncology_II | LMNB1 | P20700 | 5 | 126776623 | 126837020 | 0.99 | 0.82 | 1.18 | 8.91E-01 | 0.964 |
| OID31260 | ARAF | Oncology_II | ARAF | P10398 | X | 47561205 | 47571908 | 0.99 | 0.85 | 1.16 | 8.93E-01 | 0.965 |
| OID30322 | CPXM2 | Cardiometabolic_II | CPXM2 | Q8N436 | 10 | 123706207 | 123940267 | 1.01 | 0.85 | 1.20 | 8.92E-01 | 0.965 |
| OID30328 | PKD1 | Cardiometabolic_II | PKD1 | P98161 | 16 | 2088708 | 2135898 | 1.01 | 0.85 | 1.20 | 8.93E-01 | 0.965 |
| OID21275 | ADAMTS15 | Oncology | ADAMTS15 | Q8TE58 | 11 | 130448645 | 130476645 | 0.99 | 0.83 | 1.18 | 8.94E-01 | 0.965 |
| OID30100 | NUP50 | Cardiometabolic_II | NUP50 | Q9UKX7 | 22 | 45163925 | 45188017 | 0.99 | 0.83 | 1.18 | 8.94E-01 | 0.965 |
| OID30768 | ORM1 | Inflammation_II | ORM1 | P02763 | 9 | 114323098 | 114326479 | 0.99 | 0.82 | 1.18 | 8.94E-01 | 0.965 |
| OID21427 | CRNN | Oncology | CRNN | Q9UBG3 | 1 | 152409243 | 152414263 | 0.99 | 0.84 | 1.16 | 8.95E-01 | 0.966 |
| OID21019 | MMP8 | Neurology | MMP8 | P22894 | 11 | 102711796 | 102727050 | 0.99 | 0.84 | 1.17 | 8.96E-01 | 0.966 |
| OID20716 | CD58 | Inflammation | CD58 | P19256 | 1 | 116514534 | 116571039 | 0.99 | 0.84 | 1.16 | 8.97E-01 | 0.967 |
| OID31443 | CEACAM19 | Oncology_II | CEACAM19 | Q7Z692 | 19 | 44662278 | 44684359 | 1.01 | 0.86 | 1.18 | 8.97E-01 | 0.967 |
| OID30716 | CFHR5 | Inflammation_II | CFHR5 | Q9BXR6 | 1 | 196975010 | 197009678 | 0.99 | 0.84 | 1.17 | 8.98E-01 | 0.967 |
| OID20694 | IL18 | Inflammation | IL18 | Q14116 | 11 | 112143253 | 112164096 | 1.01 | 0.85 | 1.20 | 8.97E-01 | 0.967 |
| OID20925 | PAEP | Neurology | PAEP | P09466 | 9 | 135561756 | 135566955 | 1.02 | 0.80 | 1.29 | 8.98E-01 | 0.967 |
| OID30551 | IL36G | Inflammation_II | IL36G | Q9NZH8 | 2 | 112973203 | 112985658 | 0.99 | 0.85 | 1.16 | 8.99E-01 | 0.967 |
| OID20343 | MCAM | Cardiometabolic | MCAM | P43121 | 11 | 119308529 | 119321521 | 0.99 | 0.84 | 1.17 | 8.99E-01 | 0.967 |
| OID30118 | ATP1B2 | Cardiometabolic_II | ATP1B2 | P14415 | 17 | 7646627 | 7657770 | 0.99 | 0.85 | 1.16 | 9.01E-01 | 0.968 |
| OID20410 | C2 | Cardiometabolic | C2 | P06681 | 6 | 31897785 | 31945672 | 0.99 | 0.85 | 1.15 | 9.00E-01 | 0.968 |
| OID21283 | POLR2F | Oncology | POLR2F | P61218 | 22 | 37952607 | 38041915 | 0.99 | 0.82 | 1.19 | 9.01E-01 | 0.968 |
| OID20062 | AKR1C4 | Cardiometabolic | AKR1C4 | P17516 | 10 | 5195462 | 5218949 | 1.01 | 0.87 | 1.17 | 9.03E-01 | 0.968 |
| OID31193 | FGF9 | Oncology_II | FGF9 | P31371 | 13 | 21671073 | 21704498 | 1.01 | 0.87 | 1.17 | 9.03E-01 | 0.968 |
| OID30542 | GIPR | Inflammation_II | GIPR | P48546 | 19 | 45668221 | 45683722 | 0.99 | 0.84 | 1.16 | 9.03E-01 | 0.968 |
| OID21065 | RGMA | Neurology | RGMA | Q96B86 | 15 | 93035271 | 93089211 | 1.01 | 0.86 | 1.18 | 9.03E-01 | 0.968 |
| OID21278 | SF3B4 | Oncology | SF3B4 | Q15427 | 1 | 149923317 | 149927803 | 0.99 | 0.84 | 1.17 | 9.03E-01 | 0.968 |
| OID30119 | USP47 | Cardiometabolic_II | USP47 | Q96K76 | 11 | 11841423 | 11961887 | 0.99 | 0.84 | 1.17 | 9.02E-01 | 0.968 |
| OID20750 | TPP1 | Inflammation | TPP1 | O14773 | 11 | 6612768 | 6619448 | 1.01 | 0.85 | 1.20 | 9.04E-01 | 0.969 |
| OID30076 | NXPE4 | Cardiometabolic_II | NXPE4 | Q6UWF7 | 11 | 114570591 | 114595786 | 0.99 | 0.84 | 1.17 | 9.04E-01 | 0.969 |
| OID31227 | EVI2B | Oncology_II | EVI2B | P34910 | 17 | 31303770 | 31314105 | 1.01 | 0.87 | 1.17 | 9.04E-01 | 0.969 |
| OID20239 | MFAP5 | Cardiometabolic | MFAP5 | Q13361 | 12 | 8637346 | 8662888 | 0.99 | 0.83 | 1.18 | 9.05E-01 | 0.969 |
| OID30448 | CD3G | Inflammation_II | CD3G | P09693 | 11 | 118344344 | 118355161 | 0.99 | 0.84 | 1.16 | 9.06E-01 | 0.969 |
| OID31144 | CRYGD | Neurology_II | CRYGD | P07320 | 2 | 208121607 | 208124524 | 0.99 | 0.84 | 1.17 | 9.06E-01 | 0.969 |
| OID31201 | PDE1C | Oncology_II | PDE1C | Q14123 | 7 | 31751179 | 32428131 | 1.01 | 0.86 | 1.19 | 9.06E-01 | 0.969 |
| OID20249 | ADAMTS13 | Cardiometabolic | ADAMTS13 | Q76LX8 | 9 | 133414358 | 133459402 | 1.01 | 0.86 | 1.19 | 9.07E-01 | 0.970 |
| OID30939 | MSLNL | Neurology_II | MSLNL | Q96KJ4 | 16 | 769428 | 783370 | 0.99 | 0.84 | 1.16 | 9.07E-01 | 0.970 |
| OID31188 | CREB3 | Oncology_II | CREB3 | O43889 | 9 | 35732598 | 35736999 | 0.99 | 0.86 | 1.15 | 9.09E-01 | 0.971 |
| OID20755 | ADGRE2 | Inflammation | ADGRE2 | Q9UHX3 | 19 | 14732392 | 14778560 | 0.99 | 0.84 | 1.17 | 9.10E-01 | 0.972 |
| OID21068 | CCN5 | Neurology | CCN5 | O76076 | 20 | 44714844 | 44728509 | 1.01 | 0.84 | 1.22 | 9.10E-01 | 0.972 |
| OID30211 | HRC | Cardiometabolic_II | HRC | P23327 | 19 | 49151198 | 49155396 | 1.01 | 0.87 | 1.18 | 9.11E-01 | 0.972 |
| OID20924 | SPOCK1 | Neurology | SPOCK1 | Q08629 | 5 | 136975298 | 137598379 | 0.99 | 0.84 | 1.17 | 9.11E-01 | 0.972 |
| OID20368 | THBS4 | Cardiometabolic | THBS4 | P35443 | 5 | 79991311 | 80083287 | 0.99 | 0.84 | 1.17 | 9.11E-01 | 0.972 |
| OID30508 | ZNF174 | Inflammation_II | ZNF174 | Q15697 | 16 | 3401215 | 3409364 | 0.99 | 0.83 | 1.19 | 9.12E-01 | 0.972 |
| OID31166 | GAD1 | Oncology_II | GAD1 | Q99259 | 2 | 170813213 | 170861151 | 0.99 | 0.84 | 1.17 | 9.13E-01 | 0.973 |
| OID21250 | BAIAP2 | Oncology | BAIAP2 | Q9UQB8 | 17 | 81035122 | 81117434 | 1.01 | 0.85 | 1.20 | 9.14E-01 | 0.973 |
| OID21119 | CD177 | Neurology | CD177 | Q8N6Q3 | 19 | 43353686 | 43363172 | 1.01 | 0.86 | 1.18 | 9.15E-01 | 0.973 |
| OID30175 | KIF22 | Cardiometabolic_II | KIF22 | Q14807 | 16 | 29790727 | 29805385 | 0.99 | 0.84 | 1.17 | 9.15E-01 | 0.973 |
| OID30262 | MAN1A2 | Cardiometabolic_II | MAN1A2 | O60476 | 1 | 117367449 | 117528872 | 1.01 | 0.86 | 1.19 | 9.15E-01 | 0.973 |
| OID30687 | TCN1 | Inflammation_II | TCN1 | P20061 | 11 | 59852800 | 59866489 | 1.01 | 0.85 | 1.21 | 9.15E-01 | 0.973 |
| OID31198 | VWA5A | Oncology_II | VWA5A | O00534 | 11 | 124115404 | 124147721 | 0.99 | 0.82 | 1.19 | 9.14E-01 | 0.973 |
| OID20260 | ADGRE5 | Cardiometabolic | ADGRE5 | P48960 | 19 | 14380501 | 14408725 | 0.99 | 0.84 | 1.17 | 9.16E-01 | 0.973 |
| OID31206 | CREBZF | Oncology_II | CREBZF | Q9NS37 | 11 | 85657742 | 85682908 | 0.99 | 0.84 | 1.17 | 9.16E-01 | 0.973 |
| OID20873 | KIRREL2 | Neurology | KIRREL2 | Q6UWL6 | 19 | 35855861 | 35867136 | 1.01 | 0.86 | 1.19 | 9.16E-01 | 0.973 |
| OID30970 | CIT | Neurology_II | CIT | O14578 | 12 | 119685791 | 119877320 | 1.01 | 0.85 | 1.20 | 9.17E-01 | 0.974 |
| OID21419 | CLMP | Oncology | CLMP | Q9H6B4 | 11 | 123069872 | 123195248 | 1.01 | 0.84 | 1.21 | 9.18E-01 | 0.974 |
| OID21508 | XPNPEP2 | Oncology | XPNPEP2 | O43895 | X | 129738949 | 129769536 | 0.99 | 0.84 | 1.18 | 9.20E-01 | 0.976 |
| OID30915 | CADPS | Neurology_II | CADPS | Q9ULU8 | 3 | 62398346 | 62875416 | 0.99 | 0.85 | 1.16 | 9.22E-01 | 0.977 |
| OID20383 | HSPG2 | Cardiometabolic | HSPG2 | P98160 | 1 | 21822244 | 21937310 | 1.01 | 0.86 | 1.18 | 9.23E-01 | 0.977 |
| OID31129 | INSL3 | Neurology_II | INSL3 | P51460 | 19 | 17816512 | 17821574 | 0.99 | 0.73 | 1.33 | 9.23E-01 | 0.977 |
| OID30728 | LYVE1 | Inflammation_II | LYVE1 | Q9Y5Y7 | 11 | 10556966 | 10611689 | 0.99 | 0.84 | 1.17 | 9.22E-01 | 0.977 |
| OID31331 | MAGEA3 | Oncology_II | MAGEA3 | P43357 | X | 152698767 | 152702347 | 0.99 | 0.84 | 1.17 | 9.22E-01 | 0.977 |
| OID30988 | NFIC | Neurology_II | NFIC | P08651 | 19 | 3314403 | 3469217 | 1.01 | 0.83 | 1.22 | 9.23E-01 | 0.977 |
| OID30951 | SCT | Neurology_II | SCT | P09683 | 11 | 626309 | 627181 | 0.99 | 0.85 | 1.16 | 9.22E-01 | 0.977 |
| OID20497 | ENAH | Inflammation | ENAH | Q8N8S7 | 1 | 225486829 | 225653142 | 0.99 | 0.83 | 1.18 | 9.25E-01 | 0.977 |
| OID30566 | INSR | Inflammation_II | INSR | P06213 | 19 | 7112255 | 7294414 | 0.99 | 0.85 | 1.16 | 9.25E-01 | 0.977 |
| OID31109 | LMNB2 | Neurology_II | LMNB2 | Q03252 | 19 | 2427638 | 2456959 | 0.99 | 0.84 | 1.18 | 9.24E-01 | 0.977 |
| OID21333 | LRRC25 | Oncology | LRRC25 | Q8N386 | 19 | 18391137 | 18397622 | 0.99 | 0.84 | 1.17 | 9.25E-01 | 0.977 |
| OID30329 | NPL | Cardiometabolic_II | NPL | Q9BXD5 | 1 | 182789293 | 182830384 | 0.99 | 0.83 | 1.18 | 9.24E-01 | 0.977 |
| OID31159 | IZUMO1 | Oncology_II | IZUMO1 | Q8IYV9 | 19 | 48740852 | 48746909 | 1.01 | 0.86 | 1.18 | 9.25E-01 | 0.977 |
| OID30652 | KLK7 | Inflammation_II | KLK7 | P49862 | 19 | 50976468 | 50984099 | 0.99 | 0.84 | 1.17 | 9.26E-01 | 0.977 |
| OID30491 | ADAM12 | Inflammation_II | ADAM12 | O43184 | 10 | 126012381 | 126388477 | 0.99 | 0.83 | 1.19 | 9.28E-01 | 0.978 |
| OID21072 | APOH | Neurology | APOH | P02749 | 17 | 66212033 | 66256525 | 1.01 | 0.85 | 1.19 | 9.28E-01 | 0.978 |
| OID30315 | ASPN | Cardiometabolic_II | ASPN | Q9BXN1 | 9 | 92456205 | 92482506 | 0.99 | 0.86 | 1.15 | 9.28E-01 | 0.978 |
| OID30124 | ATP1B1 | Cardiometabolic_II | ATP1B1 | P05026 | 1 | 169105697 | 169310992 | 0.99 | 0.83 | 1.18 | 9.32E-01 | 0.978 |
| OID21029 | BST2 | Neurology | BST2 | Q10589 | 19 | 17402939 | 17405630 | 1.01 | 0.86 | 1.18 | 9.30E-01 | 0.978 |
| OID20983 | CALCA | Neurology | CALCA | P01258 | 11 | 14966668 | 14972351 | 1.01 | 0.82 | 1.24 | 9.29E-01 | 0.978 |
| OID20635 | CD79B | Inflammation | CD79B | P40259 | 17 | 63928740 | 63932336 | 1.01 | 0.86 | 1.18 | 9.32E-01 | 0.978 |
| OID20365 | CTSZ | Cardiometabolic | CTSZ | Q9UBR2 | 20 | 58985686 | 59008238 | 1.01 | 0.86 | 1.19 | 9.31E-01 | 0.978 |
| OID30935 | DUSP29 | Neurology_II | DUSP29 | Q68J44 | 10 | 75037472 | 75073642 | 0.99 | 0.85 | 1.16 | 9.30E-01 | 0.978 |
| OID31298 | IL9 | Oncology_II | IL9 | P15248 | 5 | 135892246 | 135895841 | 0.99 | 0.85 | 1.16 | 9.32E-01 | 0.978 |
| OID20205 | NPDC1 | Cardiometabolic | NPDC1 | Q9NQX5 | 9 | 137039463 | 137046179 | 1.01 | 0.86 | 1.18 | 9.27E-01 | 0.978 |
| OID20291 | PAM | Cardiometabolic | PAM | P19021 | 5 | 102753981 | 103029730 | 1.01 | 0.86 | 1.18 | 9.29E-01 | 0.978 |
| OID30226 | ROBO4 | Cardiometabolic_II | ROBO4 | Q8WZ75 | 11 | 124883691 | 124898500 | 1.01 | 0.86 | 1.19 | 9.29E-01 | 0.978 |

| | | | | | | | | | | | | |
|---|---|---|---|---|---|---|---|---|---|---|---|---|
| OID30107 | SLC51B | Cardiometabolic_II | SLC51B | Q86UW2 | 15 | 65045387 | 65053397 | 0.99 | 0.85 | 1.16 | 9.32E-01 | 0.978 |
| OID31089 | SYT1 | Neurology_II | SYT1 | P21579 | 12 | 78863993 | 79452008 | 1.01 | 0.87 | 1.16 | 9.31E-01 | 0.978 |
| OID31225 | TAGLN3 | Oncology_II | TAGLN3 | Q9UI15 | 3 | 111998739 | 112013887 | 0.99 | 0.84 | 1.17 | 9.30E-01 | 0.978 |
| OID21512 | TFPI2 | Oncology | TFPI2 | P48307 | 7 | 93885396 | 93890753 | 0.99 | 0.83 | 1.18 | 9.32E-01 | 0.978 |
| OID20148 | DCTPP1 | Cardiometabolic | DCTPP1 | Q9H773 | 16 | 30423615 | 30430030 | 1.01 | 0.85 | 1.20 | 9.33E-01 | 0.978 |
| OID21062 | FOLR2 | Neurology | FOLR2 | P14207 | 11 | 72216601 | 72221950 | 0.99 | 0.84 | 1.17 | 9.33E-01 | 0.978 |
| OID20163 | PDCD6 | Cardiometabolic | PDCD6 | O75340 | 5 | 271621 | 314974 | 1.01 | 0.86 | 1.17 | 9.33E-01 | 0.978 |
| OID31205 | MAPK13 | Oncology_II | MAPK13 | O15264 | 6 | 36127809 | 36144524 | 1.01 | 0.84 | 1.20 | 9.34E-01 | 0.978 |
| OID21243 | AGR3 | Oncology | AGR3 | Q8TD06 | 7 | 16859412 | 16881987 | 1.01 | 0.86 | 1.18 | 9.34E-01 | 0.978 |
| OID30879 | TP73 | Neurology_II | TP73 | O15350 | 1 | 3652516 | 3736201 | 1.01 | 0.86 | 1.18 | 9.35E-01 | 0.979 |
| OID20393 | CR2 | Cardiometabolic | CR2 | P20023 | 1 | 207454230 | 207489895 | 1.01 | 0.86 | 1.18 | 9.36E-01 | 0.979 |
| OID20568 | NPPC | Inflammation | NPPC | P23582 | 2 | 231921809 | 231926396 | 0.99 | 0.83 | 1.18 | 9.36E-01 | 0.980 |
| OID21127 | B4GAT1 | Neurology | B4GAT1 | O43505 | 11 | 66345374 | 66347629 | 0.99 | 0.84 | 1.17 | 9.37E-01 | 0.980 |
| OID30411 | CELA2A | Cardiometabolic_II | CELA2A | P08217 | 1 | 15456728 | 15472091 | 1.01 | 0.87 | 1.16 | 9.37E-01 | 0.980 |
| OID20470 | NFATC3 | Inflammation | NFATC3 | Q12968 | 16 | 68084751 | 68229259 | 1.01 | 0.86 | 1.18 | 9.38E-01 | 0.980 |
| OID20493 | ITGB6 | Inflammation | ITGB6 | P18564 | 2 | 160099667 | 160200313 | 1.01 | 0.85 | 1.20 | 9.38E-01 | 0.980 |
| OID20434 | LTO1 | Inflammation | LTO1 | Q8WV07 | 11 | 69653076 | 69675416 | 1.01 | 0.84 | 1.20 | 9.38E-01 | 0.980 |
| OID31162 | ATF4 | Oncology_II | ATF4 | P18848 | 22 | 39519695 | 39522683 | 0.99 | 0.84 | 1.17 | 9.39E-01 | 0.980 |
| OID21358 | PODXL | Oncology | PODXL | O00592 | 7 | 131500262 | 131558217 | 1.01 | 0.86 | 1.17 | 9.39E-01 | 0.980 |
| OID30771 | A1BG | Inflammation_II | A1BG | P04217 | 19 | 58345178 | 58353492 | 1.01 | 0.84 | 1.21 | 9.42E-01 | 0.981 |
| OID21054 | CPM | Neurology | CPM | P14384 | 12 | 68842197 | 68971570 | 0.99 | 0.82 | 1.20 | 9.41E-01 | 0.981 |
| OID20845 | GBP4 | Neurology | GBP4 | Q96PP9 | 1 | 89181144 | 89198942 | 0.99 | 0.84 | 1.17 | 9.44E-01 | 0.981 |
| OID20656 | HGF | Inflammation | HGF | P14210 | 7 | 81699010 | 81770438 | 1.01 | 0.84 | 1.21 | 9.40E-01 | 0.981 |
| OID31431 | KIR2DL2 | Oncology_II | KIR2DL2 | P43627 | 19 | 54754693 | 54769187 | 1.01 | 0.85 | 1.19 | 9.42E-01 | 0.981 |
| OID20629 | KLRB1 | Inflammation | KLRB1 | Q12918 | 12 | 9594551 | 9607916 | 0.99 | 0.84 | 1.17 | 9.41E-01 | 0.981 |
| OID21492 | MANSC1 | Oncology | MANSC1 | Q9H8J5 | 12 | 12326056 | 12350242 | 1.01 | 0.86 | 1.18 | 9.43E-01 | 0.981 |
| OID21167 | NINJ1 | Oncology | NINJ1 | Q92982 | 9 | 93121496 | 93134251 | 1.01 | 0.84 | 1.20 | 9.43E-01 | 0.981 |
| OID30767 | SERPIND1 | Inflammation_II | SERPIND1 | P05546 | 22 | 20774113 | 20787720 | 0.99 | 0.82 | 1.20 | 9.41E-01 | 0.981 |
| OID30777 | SERPING1 | Inflammation_II | SERPING1 | P05155 | 11 | 57597387 | 57619171 | 0.99 | 0.84 | 1.18 | 9.42E-01 | 0.981 |
| OID30910 | TDO2 | Neurology_II | TDO2 | P48775 | 4 | 155854738 | 155920406 | 0.99 | 0.84 | 1.18 | 9.42E-01 | 0.981 |
| OID20646 | TNFRSF11A | Inflammation | TNFRSF11A | Q9Y6Q6 | 18 | 62325287 | 62391288 | 0.99 | 0.84 | 1.17 | 9.43E-01 | 0.981 |
| OID21370 | AMIGO2 | Oncology | AMIGO2 | Q86SJ2 | 12 | 47075707 | 47079959 | 1.00 | 0.85 | 1.17 | 9.55E-01 | 0.981 |
| OID30733 | APOA4 | Inflammation_II | APOA4 | P06727 | 11 | 116820700 | 116823304 | 0.99 | 0.83 | 1.19 | 9.54E-01 | 0.981 |
| OID30719 | C8B | Inflammation_II | C8B | P07358 | 1 | 56929207 | 56974383 | 0.99 | 0.84 | 1.18 | 9.54E-01 | 0.981 |
| OID20226 | CCDC80 | Cardiometabolic | CCDC80 | Q76M96 | 3 | 112596797 | 112649530 | 0.99 | 0.84 | 1.17 | 9.50E-01 | 0.981 |
| OID20395 | CCL18 | Cardiometabolic | CCL18 | P55774 | 17 | 36064272 | 36072032 | 1.00 | 0.84 | 1.18 | 9.55E-01 | 0.981 |
| OID21130 | CD300LG | Neurology | CD300LG | Q6UXG3 | 17 | 43847148 | 43863639 | 0.99 | 0.85 | 1.17 | 9.51E-01 | 0.981 |
| OID20584 | CD4 | Inflammation | CD4 | P01730 | 12 | 6786858 | 6820799 | 1.00 | 0.84 | 1.18 | 9.55E-01 | 0.981 |
| OID20724 | CD40 | Inflammation | CD40 | P25942 | 20 | 46118278 | 46129863 | 1.00 | 0.85 | 1.17 | 9.55E-01 | 0.981 |
| OID21122 | CD99L2 | Neurology | CD99L2 | Q8TCZ2 | X | 150766336 | 150898816 | 1.01 | 0.85 | 1.20 | 9.48E-01 | 0.981 |
| OID21528 | CEACAM1 | Oncology | CEACAM1 | P13688 | 19 | 42507304 | 42561234 | 1.01 | 0.84 | 1.21 | 9.47E-01 | 0.981 |
| OID31240 | DCLRE1C | Oncology_II | DCLRE1C | Q96SD1 | 10 | 14897359 | 14954432 | 1.00 | 0.85 | 1.18 | 9.54E-01 | 0.981 |
| OID20840 | EPHA10 | Neurology | EPHA10 | Q5JZY3 | 1 | 37713880 | 37765133 | 1.01 | 0.86 | 1.18 | 9.51E-01 | 0.981 |
| OID30646 | FGFR4 | Inflammation_II | FGFR4 | P22455 | 5 | 177086905 | 177098144 | 1.00 | 0.85 | 1.18 | 9.55E-01 | 0.981 |
| OID31221 | FOS | Oncology_II | FOS | P01100 | 14 | 75278826 | 75282230 | 1.01 | 0.86 | 1.18 | 9.51E-01 | 0.981 |
| OID30898 | FRMD7 | Neurology_II | FRMD7 | Q6ZUT3 | X | 132076990 | 132128020 | 1.01 | 0.84 | 1.20 | 9.48E-01 | 0.981 |
| OID20063 | HEBP1 | Cardiometabolic | HEBP1 | Q9NRV9 | 12 | 12974870 | 13000265 | 1.01 | 0.86 | 1.18 | 9.46E-01 | 0.981 |
| OID21385 | HMBS | Oncology | HMBS | P08397 | 11 | 119084866 | 119093834 | 0.99 | 0.84 | 1.18 | 9.48E-01 | 0.981 |
| OID21213 | IKZF2 | Oncology | IKZF2 | Q9UKS7 | 2 | 212999691 | 213152427 | 1.00 | 0.86 | 1.17 | 9.51E-01 | 0.981 |
| OID20481 | IL17D | Inflammation | IL17D | Q8TAD2 | 13 | 20702127 | 20723098 | 0.99 | 0.83 | 1.19 | 9.47E-01 | 0.981 |
| OID30808 | KLF4 | Neurology_II | KLF4 | O43474 | 9 | 107484852 | 107490482 | 1.01 | 0.84 | 1.20 | 9.49E-01 | 0.981 |
| OID20632 | KLRD1 | Inflammation | KLRD1 | Q13241 | 12 | 10226058 | 10329608 | 1.01 | 0.85 | 1.19 | 9.53E-01 | 0.981 |
| OID31257 | MORF4L1 | Oncology_II | MORF4L1 | Q9UBU8 | 15 | 78810487 | 78898139 | 1.01 | 0.85 | 1.18 | 9.52E-01 | 0.981 |
| OID30834 | MYOM1 | Neurology_II | MYOM1 | P52179 | 18 | 3066807 | 3219968 | 1.00 | 0.86 | 1.16 | 9.54E-01 | 0.981 |
| OID30300 | NGFR | Cardiometabolic_II | NGFR | P08138 | 17 | 49495293 | 49515008 | 0.99 | 0.84 | 1.17 | 9.51E-01 | 0.981 |
| OID30418 | NPC2 | Cardiometabolic_II | NPC2 | P61916 | 14 | 74476192 | 74494177 | 1.01 | 0.85 | 1.19 | 9.51E-01 | 0.981 |
| OID31168 | PDCL2 | Oncology_II | PDCL2 | Q8N4E4 | 4 | 55556519 | 55592245 | 1.00 | 0.87 | 1.14 | 9.46E-01 | 0.981 |
| OID20252 | PLA2G1B | Cardiometabolic | PLA2G1B | P04054 | 12 | 120322115 | 120327779 | 0.99 | 0.85 | 1.16 | 9.47E-01 | 0.981 |
| OID31304 | PPP1R12B | Oncology_II | PPP1R12B | O60237 | 1 | 202348699 | 202592706 | 0.99 | 0.84 | 1.18 | 9.49E-01 | 0.981 |
| OID21461 | SMOC1 | Oncology | SMOC1 | Q9H4F8 | 14 | 69854131 | 70032366 | 0.99 | 0.83 | 1.18 | 9.50E-01 | 0.981 |
| OID31359 | TG | Oncology_II | TG | P01266 | 8 | 132866958 | 133134903 | 1.00 | 0.84 | 1.17 | 9.55E-01 | 0.981 |
| OID20851 | VSTM1 | Neurology | VSTM1 | Q6UX27 | 19 | 54040825 | 54063953 | 1.01 | 0.85 | 1.19 | 9.51E-01 | 0.981 |
| OID21407 | KLK14 | Oncology | KLK14 | Q9P0G3 | 19 | 51077495 | 51084245 | 1.00 | 0.85 | 1.19 | 9.56E-01 | 0.982 |
| OID30725 | B2M | Inflammation_II | B2M | P61769 | 15 | 44711358 | 44718851 | 1.00 | 0.85 | 1.16 | 9.57E-01 | 0.983 |
| OID30973 | CCDC28A | Neurology_II | CCDC28A | Q8IWP9 | 6 | 138773769 | 138793319 | 1.00 | 0.84 | 1.17 | 9.58E-01 | 0.983 |
| OID21413 | AMBP | Oncology | AMBP | P02760 | 9 | 114060127 | 114078328 | 1.00 | 0.85 | 1.19 | 9.60E-01 | 0.984 |
| OID30088 | HCG22 | Cardiometabolic_II | HCG22 | E2RYF7 | 6 | 31053450 | 31059890 | 1.00 | 0.85 | 1.17 | 9.60E-01 | 0.984 |
| OID20598 | PTPRM | Inflammation | PTPRM | P28827 | 18 | 7566782 | 8406861 | 1.00 | 0.85 | 1.19 | 9.59E-01 | 0.984 |
| OID20283 | SEMA7A | Cardiometabolic | SEMA7A | O75326 | 15 | 74409289 | 74433958 | 1.00 | 0.84 | 1.18 | 9.59E-01 | 0.984 |
| OID31174 | SLIRP | Oncology_II | SLIRP | Q9GZT3 | 14 | 77708071 | 77761104 | 1.00 | 0.83 | 1.19 | 9.59E-01 | 0.984 |
| OID20786 | AGRN | Inflammation | AGRN | O00468 | 1 | 1020120 | 1056118 | 1.00 | 0.85 | 1.17 | 9.62E-01 | 0.984 |
| OID31184 | BSND | Oncology_II | BSND | Q8WZ55 | 1 | 54998933 | 55017172 | 1.00 | 0.84 | 1.18 | 9.62E-01 | 0.984 |
| OID20610 | CCL3 | Inflammation | CCL3 | P10147 | 17 | 36088256 | 36090169 | 1.00 | 0.82 | 1.23 | 9.61E-01 | 0.984 |
| OID20982 | EPHB6 | Neurology | EPHB6 | O15197 | 7 | 142855061 | 142871094 | 1.00 | 0.86 | 1.17 | 9.62E-01 | 0.984 |
| OID31219 | GATA3 | Oncology_II | GATA3 | P23771 | 10 | 8045378 | 8075198 | 1.00 | 0.85 | 1.19 | 9.61E-01 | 0.984 |
| OID30288 | GIP | Cardiometabolic_II | GIP | P09681 | 17 | 48958554 | 48968596 | 1.00 | 0.86 | 1.16 | 9.61E-01 | 0.984 |
| OID20498 | IL15RA | Inflammation | IL15RA | Q13261 | 10 | 5943639 | 5978187 | 1.00 | 0.86 | 1.18 | 9.62E-01 | 0.984 |
| OID20886 | ANXA10 | Neurology | ANXA10 | Q9UJ72 | 4 | 168092537 | 168187736 | 1.00 | 0.86 | 1.17 | 9.63E-01 | 0.984 |
| OID31341 | DYNC1H1 | Oncology_II | DYNC1H1 | Q14204 | 14 | 101964573 | 102056443 | 1.00 | 0.83 | 1.21 | 9.64E-01 | 0.985 |
| OID30172 | PMCH | Cardiometabolic_II | PMCH | P20382 | 12 | 102196459 | 102197833 | 1.00 | 0.84 | 1.18 | 9.64E-01 | 0.985 |
| OID31204 | DNAJA1 | Oncology_II | DNAJA1 | P31689 | 9 | 33025273 | 33039907 | 1.00 | 0.85 | 1.19 | 9.66E-01 | 0.985 |
| OID20323 | LILRB1 | Cardiometabolic | LILRB1 | Q8NHL6 | 19 | 54617158 | 54638022 | 1.00 | 0.84 | 1.18 | 9.65E-01 | 0.985 |
| OID31163 | NMRK2 | Oncology_II | NMRK2 | Q9NPI5 | 19 | 3933069 | 3942416 | 1.00 | 0.85 | 1.17 | 9.66E-01 | 0.985 |
| OID31237 | BTLA | Oncology_II | BTLA | Q7Z6A9 | 3 | 112463966 | 112499472 | 1.00 | 0.85 | 1.17 | 9.68E-01 | 0.986 |
| OID20090 | GP2 | Cardiometabolic | GP2 | P55259 | 16 | 20309574 | 20327808 | 1.00 | 0.84 | 1.18 | 9.68E-01 | 0.986 |
| OID31175 | HMMR | Oncology_II | HMMR | O75330 | 5 | 163460203 | 163491941 | 1.00 | 0.84 | 1.19 | 9.67E-01 | 0.986 |
| OID31245 | KHDC3L | Oncology_II | KHDC3L | Q587J8 | 6 | 73362658 | 73364171 | 1.00 | 0.84 | 1.20 | 9.67E-01 | 0.986 |
| OID31389 | WDR46 | Oncology_II | WDR46 | O15213 | 6 | 33279108 | 33289247 | 1.00 | 0.84 | 1.19 | 9.68E-01 | 0.986 |
| OID30697 | CR1 | Inflammation_II | CR1 | P17927 | 1 | 207496147 | 207641765 | 1.00 | 0.85 | 1.19 | 9.69E-01 | 0.986 |
| OID20288 | ENPP2 | Cardiometabolic | ENPP2 | Q13822 | 8 | 119557086 | 119673453 | 1.00 | 0.83 | 1.22 | 9.69E-01 | 0.986 |
| OID30710 | CTBS | Inflammation_II | CTBS | Q01459 | 1 | 84549611 | 84574480 | 1.00 | 0.83 | 1.19 | 9.70E-01 | 0.987 |
| OID31480 | CD101 | Oncology_II | CD101 | Q93033 | 1 | 117001750 | 117036552 | 1.00 | 0.84 | 1.19 | 9.73E-01 | 0.989 |
| OID30173 | COPB2 | Cardiometabolic_II | COPB2 | P35606 | 3 | 139353946 | 139389736 | 1.00 | 0.84 | 1.19 | 9.73E-01 | 0.989 |
| OID21024 | CXCL13 | Neurology | CXCL13 | O43927 | 4 | 77511753 | 77611834 | 1.00 | 0.84 | 1.18 | 9.74E-01 | 0.989 |
| OID20346 | ART3 | Cardiometabolic | ART3 | Q13508 | 4 | 76011184 | 76112802 | 1.00 | 0.85 | 1.19 | 9.76E-01 | 0.990 |
| OID30730 | PZP | Inflammation_II | PZP | P20742 | 12 | 9148840 | 9208395 | 1.00 | 0.80 | 1.25 | 9.76E-01 | 0.990 |
| OID30298 | SPESP1 | Cardiometabolic_II | SPESP1 | Q6UW49 | 15 | 68818221 | 68946811 | 1.00 | 0.85 | 1.18 | 9.75E-01 | 0.990 |
| OID31371 | VSIG2 | Oncology_II | VSIG2 | Q96IQ7 | 11 | 124747474 | 124752255 | 1.00 | 0.87 | 1.15 | 9.75E-01 | 0.990 |
| OID30924 | LRRC38 | Neurology_II | LRRC38 | Q5VT99 | 1 | 13474973 | 13514003 | 1.00 | 0.86 | 1.16 | 9.76E-01 | 0.990 |
| OID30179 | BRDT | Cardiometabolic_II | BRDT | Q58F21 | 1 | 91949343 | 92014426 | 1.00 | 0.85 | 1.17 | 9.77E-01 | 0.991 |
| OID30509 | NAGA | Inflammation_II | NAGA | P17050 | 22 | 42058334 | 42070842 | 1.00 | 0.85 | 1.18 | 9.78E-01 | 0.992 |
| OID30896 | BATF | Neurology_II | BATF | Q16520 | 14 | 75522455 | 75546993 | 1.00 | 0.85 | 1.17 | 9.79E-01 | 0.992 |
| OID20309 | AZU1 | Cardiometabolic | AZU1 | P20160 | 19 | 825097 | 832018 | 1.00 | 0.84 | 1.19 | 9.81E-01 | 0.993 |

| | | | | | | | | | | | | |
|---|---|---|---|---|---|---|---|---|---|---|---|---|
| OID20660 | CTSO | Inflammation | CTSO | P43234 | 4 | 155921580 | 155953912 | 1.00 | 0.85 | 1.18 | 9.81E-01 | 0.993 |
| OID21531 | MIA | Oncology | MIA | Q16674 | 19 | 40771648 | 40777490 | 1.00 | 0.86 | 1.17 | 9.80E-01 | 0.993 |
| OID21235 | NPTN | Oncology | NPTN | Q9Y639 | 15 | 73560014 | 73634134 | 1.00 | 0.85 | 1.17 | 9.80E-01 | 0.993 |
| OID31294 | TK1 | Oncology_II | TK1 | P04183 | 17 | 78174091 | 78187233 | 1.00 | 0.84 | 1.18 | 9.81E-01 | 0.993 |
| OID21328 | CD302 | Oncology | CD302 | Q8IX05 | 2 | 159768628 | 159798255 | 1.00 | 0.85 | 1.18 | 9.82E-01 | 0.993 |
| OID30800 | CTNNA1 | Neurology_II | CTNNA1 | P35221 | 5 | 138610967 | 138935034 | 1.00 | 0.85 | 1.17 | 9.82E-01 | 0.993 |
| OID20076 | FABP6 | Cardiometabolic | FABP6 | P51161 | 5 | 160187367 | 160238735 | 1.00 | 0.85 | 1.17 | 9.83E-01 | 0.993 |
| OID20730 | SMOC2 | Inflammation | SMOC2 | Q9H3U7 | 6 | 168441151 | 168673445 | 1.00 | 0.84 | 1.18 | 9.83E-01 | 0.993 |
| OID30091 | CYP24A1 | Cardiometabolic_II | CYP24A1 | Q07973 | 20 | 54153446 | 54173986 | 1.00 | 0.85 | 1.17 | 9.84E-01 | 0.994 |
| OID31153 | GIPC2 | Neurology_II | GIPC2 | Q8TF65 | 1 | 77979542 | 78138444 | 1.00 | 0.84 | 1.18 | 9.85E-01 | 0.994 |
| OID20290 | SELE | Cardiometabolic | SELE | P16581 | 1 | 169722640 | 169764705 | 1.00 | 0.85 | 1.18 | 9.85E-01 | 0.994 |
| OID30524 | ACP1 | Inflammation_II | ACP1 | P24666 | 2 | 264140 | 278283 | 1.00 | 0.84 | 1.20 | 9.90E-01 | 0.996 |
| OID30746 | APOD | Inflammation_II | APOD | P05090 | 3 | 195568705 | 195584033 | 1.00 | 0.83 | 1.21 | 9.89E-01 | 0.996 |
| OID31126 | CAPS | Neurology_II | CAPS | Q13938 | 19 | 5912339 | 5916211 | 1.00 | 0.84 | 1.18 | 9.89E-01 | 0.996 |
| OID31302 | CD3E | Oncology_II | CD3E | P07766 | 11 | 118304730 | 118316175 | 1.00 | 0.84 | 1.19 | 9.88E-01 | 0.996 |
| OID20914 | CRTAM | Neurology | CRTAM | O95727 | 11 | 122838500 | 122872643 | 1.00 | 0.86 | 1.17 | 9.90E-01 | 0.996 |
| OID30953 | DCC | Neurology_II | DCC | P43146 | 18 | 52340197 | 53535903 | 1.00 | 0.84 | 1.20 | 9.88E-01 | 0.996 |
| OID21441 | HS6ST1 | Oncology | HS6ST1 | O60243 | 2 | 128236716 | 128318868 | 1.00 | 0.85 | 1.17 | 9.88E-01 | 0.996 |
| OID31217 | MORN4 | Oncology_II | MORN4 | Q8NDC4 | 10 | 97614553 | 97633500 | 1.00 | 0.86 | 1.16 | 9.87E-01 | 0.996 |
| OID21105 | PLA2G7 | Neurology | PLA2G7 | Q13093 | 6 | 46704201 | 46735693 | 1.00 | 0.85 | 1.17 | 9.89E-01 | 0.996 |
| OID30504 | POF1B | Inflammation_II | POF1B | Q8WVV4 | X | 85277396 | 85379717 | 1.00 | 0.85 | 1.17 | 9.90E-01 | 0.996 |
| OID30196 | SEL1L | Cardiometabolic_II | SEL1L | Q9UBV2 | 14 | 81471547 | 81533853 | 1.00 | 0.84 | 1.18 | 9.90E-01 | 0.996 |
| OID20738 | COLEC12 | Inflammation | COLEC12 | Q5KU26 | 18 | 316737 | 500722 | 1.00 | 0.84 | 1.19 | 9.91E-01 | 0.996 |
| OID21025 | CD34 | Neurology | CD34 | P28906 | 1 | 207880972 | 207911402 | 1.00 | 0.85 | 1.18 | 9.92E-01 | 0.996 |
| OID30256 | HSDL2 | Cardiometabolic_II | HSDL2 | Q6YN16 | 9 | 112380080 | 112472405 | 1.00 | 0.85 | 1.17 | 9.91E-01 | 0.996 |
| OID30838 | PCARE | Neurology_II | PCARE | A6NGG8 | 2 | 29060976 | 29074523 | 1.00 | 0.85 | 1.17 | 9.92E-01 | 0.996 |
| OID20889 | ISLR2 | Neurology | ISLR2 | Q6UXK2 | 15 | 74100311 | 74138540 | 1.00 | 0.85 | 1.18 | 9.94E-01 | 0.997 |
| OID31100 | CALCB | Neurology_II | CALCB | P10092 | 11 | 14904997 | 15082342 | 1.00 | 0.84 | 1.19 | 9.96E-01 | 0.998 |
| OID20791 | FOSB | Neurology | FOSB | P53539 | 19 | 45467995 | 45475179 | 1.00 | 0.85 | 1.18 | 9.96E-01 | 0.998 |
| OID30280 | LAMB1 | Cardiometabolic_II | LAMB1 | P07942 | 7 | 107923799 | 108003213 | 1.00 | 0.85 | 1.18 | 9.95E-01 | 0.998 |
| OID21396 | PDCD1 | Oncology | PDCD1 | Q15116 | 2 | 241849884 | 241858894 | 1.00 | 0.86 | 1.16 | 9.96E-01 | 0.998 |
| OID30318 | TIMP2 | Cardiometabolic_II | TIMP2 | P16035 | 17 | 78852977 | 78925387 | 1.00 | 0.86 | 1.16 | 9.96E-01 | 0.998 |
| OID21090 | CLEC14A | Neurology | CLEC14A | Q86T13 | 14 | 38254000 | 38256093 | 1.00 | 0.85 | 1.17 | 9.98E-01 | 0.999 |
| OID20332 | COL18A1 | Cardiometabolic | COL18A1 | P39060 | 21 | 45405165 | 45513720 | 1.00 | 0.84 | 1.18 | 9.97E-01 | 0.999 |
| OID20212 | PRSS27 | Cardiometabolic | PRSS27 | Q9BQR3 | 16 | 2712419 | 2720551 | 1.00 | 0.85 | 1.18 | 9.98E-01 | 0.999 |
| OID20989 | PSME2 | Neurology | PSME2 | Q9UL46 | 14 | 24143362 | 24147570 | 1.00 | 0.84 | 1.19 | 9.98E-01 | 0.999 |
| OID31408 | REG3G | Oncology_II | REG3G | Q6UW15 | 2 | 79025686 | 79028505 | 1.00 | 0.85 | 1.17 | 9.99E-01 | 0.999 |
| OID30903 | SCN3A | Neurology_II | SCN3A | Q9NY46 | 2 | 165087526 | 165204050 | 1.00 | 0.84 | 1.20 | 1.00E+00 | 1.000 |

**Table S5 Associations between protein expression levels and incident ADRD in participants with a history of MDD at baseline, adjusting for covariates without *APOE* e4 carrier status**

| Olink ID | Assay Target | Protein panel | Gene symbol | UniProt | Gene CHROM | Gene start | Gene end | HR | Lower | Upper | Unadj P | FDR-Adj P |
|---|---|---|---|---|---|---|---|---|---|---|---|---|
| OID21247 | GFAP | Oncology | GFAP | P14136 | 17 | 44903159 | 44916937 | 1.43 | 1.25 | 1.64 | 1.81E-07 | 0.001 |
| OID20871 | NEFL | Neurology | NEFL | P07196 | 8 | 24950955 | 24956721 | 1.37 | 1.18 | 1.59 | 2.50E-05 | 0.036 |
| OID30303 | GET3 | Cardiometabolic_II | GET3 | O43681 | 19 | 12737139 | 12748323 | 0.65 | 0.53 | 0.81 | 7.65E-05 | 0.039 |
| OID31364 | GLYR1 | Oncology_II | GLYR1 | Q49A26 | 16 | 4803203 | 4847288 | 0.59 | 0.45 | 0.77 | 8.54E-05 | 0.039 |
| OID21266 | HPGDS | Oncology | HPGDS | O60760 | 4 | 94298535 | 94342876 | 0.72 | 0.61 | 0.85 | 8.45E-05 | 0.039 |
| OID20916 | PSG1 | Neurology | PSG1 | P11464 | 19 | 42866464 | 42879822 | 1.48 | 1.22 | 1.78 | 4.67E-05 | 0.039 |
| OID31067 | VGF | Neurology_II | VGF | O15240 | 7 | 101162509 | 101165569 | 0.73 | 0.62 | 0.86 | 9.25E-05 | 0.039 |
| OID30727 | APOE | Inflammation_II | APOE | P02649 | 19 | 44905791 | 44909393 | 0.64 | 0.51 | 0.81 | 2.02E-04 | 0.074 |
| OID30574 | GHR | Inflammation_II | GHR | P10912 | 5 | 42423439 | 42721878 | 0.72 | 0.60 | 0.86 | 2.56E-04 | 0.083 |
| OID31287 | CEP290 | Oncology_II | CEP290 | O15078 | 12 | 88049016 | 88142099 | 0.69 | 0.56 | 0.85 | 4.46E-04 | 0.130 |
| OID30571 | AKAP12 | Inflammation_II | AKAP12 | Q02952 | 6 | 151239967 | 151358559 | 0.74 | 0.62 | 0.88 | 6.62E-04 | 0.152 |
| OID30702 | FGL1 | Inflammation_II | FGL1 | Q08830 | 8 | 17864380 | 17910365 | 1.33 | 1.13 | 1.56 | 7.59E-04 | 0.152 |
| OID31004 | GART | Neurology_II | GART | P22102 | 21 | 33503931 | 33543491 | 0.70 | 0.57 | 0.86 | 7.44E-04 | 0.152 |
| OID20281 | SERPINA11 | Cardiometabolic | SERPINA11 | Q86U17 | 14 | 94442464 | 94452800 | 1.37 | 1.14 | 1.65 | 7.82E-04 | 0.152 |
| OID30604 | SERPINI1 | Inflammation_II | SERPINI1 | Q99574 | 3 | 167735243 | 167825569 | 0.74 | 0.63 | 0.88 | 7.02E-04 | 0.152 |
| OID21479 | CPXM1 | Oncology | CPXM1 | Q96SM3 | 20 | 2794074 | 2800627 | 0.76 | 0.64 | 0.89 | 9.01E-04 | 0.155 |
| OID21389 | EBI3_IL27 | Oncology | EBI3;IL27 | Q14213;Q8NEV9 | 19;16 | 4229523;28499362 | 4237528;28512051 | 1.33 | 1.12 | 1.57 | 8.72E-04 | 0.155 |
| OID30985 | CDK5RAP3 | Neurology_II | CDK5RAP3 | Q96JB5 | 17 | 47967810 | 47981781 | 0.73 | 0.60 | 0.88 | 1.26E-03 | 0.196 |
| OID31518 | DDX1 | Oncology_II | DDX1 | Q92499 | 2 | 15591178 | 15634346 | 0.70 | 0.57 | 0.87 | 1.27E-03 | 0.196 |
| OID31476 | SNX18 | Oncology_II | SNX18 | Q96RF0 | 5 | 54517759 | 54546586 | 0.71 | 0.57 | 0.87 | 1.35E-03 | 0.197 |
| OID31376 | RFC4 | Oncology_II | RFC4 | P35249 | 3 | 186789880 | 186807058 | 0.74 | 0.61 | 0.89 | 1.83E-03 | 0.254 |
| OID30612 | ADH1B | Inflammation_II | ADH1B | P00325 | 4 | 99304971 | 99352760 | 0.76 | 0.63 | 0.92 | 4.95E-03 | 0.269 |
| OID20117 | ADH4 | Cardiometabolic | ADH4 | P08319 | 4 | 99123657 | 99157792 | 0.76 | 0.63 | 0.91 | 3.13E-03 | 0.269 |
| OID30260 | AHNAK | Cardiometabolic_II | AHNAK | Q09666 | 11 | 62433542 | 62556235 | 0.74 | 0.61 | 0.90 | 2.74E-03 | 0.269 |
| OID31117 | AP3S2 | Neurology_II | AP3S2 | P59780 | 15 | 89830599 | 89894638 | 0.77 | 0.64 | 0.92 | 4.21E-03 | 0.269 |
| OID30815 | CRYM | Neurology_II | CRYM | Q14894 | 16 | 21238874 | 21303083 | 0.74 | 0.61 | 0.91 | 3.41E-03 | 0.269 |
| OID20613 | CXCL6 | Inflammation | CXCL6 | P80162 | 4 | 73836640 | 73849064 | 0.79 | 0.67 | 0.93 | 5.16E-03 | 0.269 |
| OID21066 | DKK1 | Neurology | DKK1 | O94907 | 10 | 52314281 | 52318042 | 0.80 | 0.68 | 0.93 | 4.47E-03 | 0.269 |
| OID31317 | ERI1 | Oncology_II | ERI1 | Q8IV48 | 8 | 9002147 | 9116746 | 0.75 | 0.61 | 0.92 | 5.16E-03 | 0.269 |
| OID31040 | FGD3 | Neurology_II | FGD3 | Q5JSP0 | 9 | 92947523 | 93036236 | 0.78 | 0.66 | 0.92 | 4.08E-03 | 0.269 |
| OID31041 | GID8 | Neurology_II | GID8 | Q9NWU2 | 20 | 62938147 | 62948475 | 0.70 | 0.55 | 0.88 | 2.47E-03 | 0.269 |
| OID30611 | GP5 | Inflammation_II | GP5 | P40197 | 3 | 194394821 | 194399266 | 0.81 | 0.70 | 0.94 | 4.86E-03 | 0.269 |
| OID31283 | IFIT3 | Oncology_II | IFIT3 | O14879 | 10 | 89327307 | 89377473 | 0.77 | 0.64 | 0.92 | 5.24E-03 | 0.269 |
| OID31148 | KCTD5 | Neurology_II | KCTD5 | Q9NXV2 | 16 | 2682523 | 2709030 | 0.75 | 0.61 | 0.92 | 5.53E-03 | 0.269 |
| OID20232 | KYAT1 | Cardiometabolic | KYAT1 | Q16773 | 9 | 128832942 | 128882494 | 0.77 | 0.65 | 0.91 | 2.42E-03 | 0.269 |
| OID20438 | LRRN1 | Inflammation | LRRN1 | Q6UXK5 | 3 | 3799431 | 3849834 | 0.79 | 0.68 | 0.92 | 3.03E-03 | 0.269 |
| OID21530 | LYPD3 | Oncology | LYPD3 | O95274 | 19 | 43460787 | 43465608 | 1.26 | 1.08 | 1.46 | 2.54E-03 | 0.269 |
| OID30637 | LZTFL1 | Inflammation_II | LZTFL1 | Q9NQ48 | 3 | 45823316 | 45916042 | 0.78 | 0.66 | 0.93 | 4.54E-03 | 0.269 |
| OID20588 | MGMT | Inflammation | MGMT | P16455 | 10 | 129467190 | 129770983 | 0.80 | 0.69 | 0.93 | 4.67E-03 | 0.269 |
| OID30638 | MOCS2 | Inflammation_II | MOCS2 | O96007 | 5 | 53095679 | 53110063 | 0.78 | 0.65 | 0.93 | 4.88E-03 | 0.269 |
| OID30120 | MYL1 | Cardiometabolic_II | MYL1 | P05976 | 2 | 210290150 | 210315174 | 1.17 | 1.05 | 1.30 | 4.19E-03 | 0.269 |
| OID30252 | NIT1 | Cardiometabolic_II | NIT1 | Q86X76 | 1 | 161118086 | 161125445 | 0.81 | 0.69 | 0.93 | 4.00E-03 | 0.269 |
| OID30390 | NPTX2 | Cardiometabolic_II | NPTX2 | P47972 | 7 | 98617285 | 98629869 | 1.30 | 1.09 | 1.55 | 3.18E-03 | 0.269 |
| OID20191 | NPTXR | Cardiometabolic | NPTXR | O95502 | 22 | 38818452 | 38844028 | 0.77 | 0.64 | 0.91 | 3.14E-03 | 0.269 |
| OID30201 | OPLAH | Cardiometabolic_II | OPLAH | O14841 | 8 | 144051266 | 144063965 | 0.76 | 0.64 | 0.91 | 2.22E-03 | 0.269 |
| OID31003 | PCBP2 | Neurology_II | PCBP2 | Q15366 | 12 | 53452102 | 53481162 | 0.79 | 0.67 | 0.93 | 3.81E-03 | 0.269 |
| OID30511 | PER3 | Inflammation_II | PER3 | P56645 | 1 | 7784291 | 7845177 | 0.79 | 0.67 | 0.93 | 5.27E-03 | 0.269 |
| OID20570 | PTX3 | Inflammation | PTX3 | P26022 | 3 | 157436850 | 157443633 | 1.33 | 1.10 | 1.61 | 3.58E-03 | 0.269 |
| OID20425 | RAB37 | Inflammation | RAB37 | Q96AX2 | 17 | 74670578 | 74747335 | 0.74 | 0.61 | 0.91 | 4.05E-03 | 0.269 |
| OID30284 | RPL14 | Cardiometabolic_II | RPL14 | P50914 | 3 | 40457292 | 40468587 | 0.75 | 0.61 | 0.90 | 2.92E-03 | 0.269 |
| OID20342 | SERPINE1 | Cardiometabolic | SERPINE1 | P05121 | 7 | 101127104 | 101139247 | 0.80 | 0.68 | 0.93 | 4.08E-03 | 0.269 |
| OID20539 | SLC39A5 | Inflammation | SLC39A5 | Q6ZMH5 | 12 | 56230049 | 56237846 | 0.77 | 0.64 | 0.92 | 3.50E-03 | 0.269 |
| OID30215 | SNX5 | Cardiometabolic_II | SNX5 | Q9Y5X3 | 20 | 17941597 | 17968980 | 0.75 | 0.62 | 0.91 | 3.90E-03 | 0.269 |
| OID31216 | SSH3 | Oncology_II | SSH3 | Q8TE77 | 11 | 67303478 | 67312607 | 0.76 | 0.63 | 0.91 | 3.11E-03 | 0.269 |
| OID31320 | TIGIT | Oncology_II | TIGIT | Q495A1 | 3 | 114276913 | 114310288 | 0.76 | 0.63 | 0.92 | 4.93E-03 | 0.269 |
| OID30682 | TREML1 | Inflammation_II | TREML1 | Q86YW5 | 6 | 41149337 | 41154347 | 0.78 | 0.66 | 0.92 | 4.06E-03 | 0.269 |
| OID30128 | TRPV3 | Cardiometabolic_II | TRPV3 | Q8NET8 | 17 | 3510502 | 3557812 | 1.21 | 1.06 | 1.39 | 5.46E-03 | 0.269 |
| OID31179 | VCPKMT | Oncology_II | VCPKMT | Q9H867 | 14 | 50108632 | 50116600 | 0.72 | 0.57 | 0.91 | 5.52E-03 | 0.269 |
| OID21524 | VEGFC | Oncology | VEGFC | P49767 | 4 | 176683538 | 176792922 | 0.80 | 0.69 | 0.94 | 5.04E-03 | 0.269 |
| OID31350 | ZNF830 | Oncology_II | ZNF830 | Q96NB3 | 17 | 34961540 | 34963777 | 0.70 | 0.56 | 0.88 | 2.29E-03 | 0.269 |
| OID30706 | AHSG | Inflammation_II | AHSG | P02765 | 3 | 186613060 | 186621318 | 0.80 | 0.68 | 0.95 | 9.49E-03 | 0.287 |
| OID31142 | AK2 | Neurology_II | AK2 | P54819 | 1 | 33007986 | 33080996 | 0.82 | 0.70 | 0.97 | 2.05E-02 | 0.287 |
| OID20895 | AKT1S1 | Neurology | AKT1S1 | Q96B36 | 19 | 49869033 | 49878459 | 0.81 | 0.69 | 0.95 | 1.03E-02 | 0.287 |
| OID31052 | AKT2 | Neurology_II | AKT2 | P31751 | 19 | 40230317 | 40285536 | 0.82 | 0.70 | 0.96 | 1.22E-02 | 0.287 |
| OID20740 | ANGPT1 | Inflammation | ANGPT1 | Q15389 | 8 | 107249482 | 107498055 | 0.83 | 0.72 | 0.97 | 1.72E-02 | 0.287 |
| OID20703 | ANGPTL4 | Inflammation | ANGPTL4 | Q9BY76 | 19 | 8363289 | 8374370 | 1.23 | 1.03 | 1.46 | 1.93E-02 | 0.287 |
| OID21070 | APP | Neurology | APP | P05067 | 21 | 25880550 | 26171128 | 0.82 | 0.70 | 0.97 | 1.81E-02 | 0.287 |
| OID31024 | ARFIP1 | Neurology_II | ARFIP1 | P53367 | 4 | 152779937 | 152918463 | 0.81 | 0.68 | 0.96 | 1.70E-02 | 0.287 |
| OID31339 | ARG2 | Oncology_II | ARG2 | P78540 | 14 | 67619920 | 67651708 | 0.80 | 0.66 | 0.97 | 2.14E-02 | 0.287 |
| OID30653 | ARHGAP45 | Inflammation_II | ARHGAP45 | Q92619 | 19 | 1065923 | 1086628 | 0.82 | 0.69 | 0.97 | 2.14E-02 | 0.287 |
| OID31133 | ARHGEF1 | Neurology_II | ARHGEF1 | Q92888 | 19 | 41883173 | 41930150 | 0.83 | 0.71 | 0.97 | 2.10E-02 | 0.287 |
| OID30168 | ARNTL | Cardiometabolic_II | ARNTL | O00327 | 11 | 13276652 | 13387266 | 1.17 | 1.03 | 1.33 | 1.82E-02 | 0.287 |
| OID30283 | ASRGL1 | Cardiometabolic_II | ASRGL1 | Q7L266 | 11 | 62337448 | 62393412 | 0.81 | 0.69 | 0.95 | 1.12E-02 | 0.287 |
| OID20843 | ATP6V1F | Neurology | ATP6V1F | Q16864 | 7 | 128862856 | 128865847 | 0.80 | 0.67 | 0.95 | 1.09E-02 | 0.287 |
| OID30242 | ATP6V1G1 | Cardiometabolic_II | ATP6V1G1 | O75348 | 9 | 114587706 | 114598915 | 0.80 | 0.68 | 0.96 | 1.31E-02 | 0.287 |
| OID31149 | ATXN3 | Neurology_II | ATXN3 | P54252 | 14 | 92044496 | 92106621 | 0.81 | 0.68 | 0.96 | 1.41E-02 | 0.287 |
| OID20998 | BCAN | Neurology | BCAN | Q96GW7 | 1 | 156641390 | 156659532 | 0.82 | 0.70 | 0.97 | 1.97E-02 | 0.287 |
| OID30220 | BECN1 | Cardiometabolic_II | BECN1 | Q14457 | 17 | 42810134 | 42833350 | 0.79 | 0.66 | 0.95 | 1.10E-02 | 0.287 |
| OID31506 | BRAP | Oncology_II | BRAP | Q7Z569 | 12 | 111642146 | 111685956 | 0.82 | 0.70 | 0.97 | 1.84E-02 | 0.287 |
| OID31259 | BRD1 | Oncology_II | BRD1 | O95696 | 22 | 49773283 | 49827873 | 0.80 | 0.66 | 0.97 | 2.27E-02 | 0.287 |
| OID31049 | C1QTNF6 | Neurology_II | C1QTNF6 | Q9BXI9 | 22 | 37180166 | 37199385 | 0.80 | 0.67 | 0.97 | 2.01E-02 | 0.287 |
| OID31095 | C7orf50 | Neurology_II | C7orf50 | Q9BRJ6 | 7 | 996986 | 1138260 | 0.67 | 0.49 | 0.92 | 1.27E-02 | 0.287 |
| OID20181 | CANT1 | Cardiometabolic | CANT1 | Q8WVQ1 | 17 | 78991716 | 79009867 | 0.80 | 0.68 | 0.95 | 9.70E-03 | 0.287 |
| OID30200 | CBS | Cardiometabolic_II | CBS | P35520 | 21 | 43053191 | 43076943 | 0.80 | 0.67 | 0.96 | 1.57E-02 | 0.287 |
| OID20709 | CCN2 | Inflammation | CCN2 | P29279 | 6 | 131948176 | 131951372 | 0.81 | 0.69 | 0.96 | 1.51E-02 | 0.287 |
| OID20680 | CD276 | Inflammation | CD276 | Q5ZPR3 | 15 | 73683966 | 73714514 | 1.21 | 1.03 | 1.43 | 2.11E-02 | 0.287 |
| OID31076 | CDAN1 | Neurology_II | CDAN1 | Q8IWY9 | 15 | 42723544 | 42737128 | 0.78 | 0.64 | 0.95 | 1.41E-02 | 0.287 |
| OID20836 | CETN2 | Neurology | CETN2 | P41208 | X | 152826994 | 152830757 | 0.81 | 0.68 | 0.96 | 1.70E-02 | 0.287 |
| OID31525 | CETN3 | Oncology_II | CETN3 | O15182 | 5 | 90392257 | 90409766 | 0.80 | 0.68 | 0.95 | 1.15E-02 | 0.287 |
| OID30894 | CGN | Neurology_II | CGN | Q9P2M7 | 1 | 151510510 | 151538692 | 0.79 | 0.65 | 0.96 | 1.83E-02 | 0.287 |
| OID30972 | CHM | Neurology_II | CHM | P24386 | X | 85861180 | 86047561 | 0.82 | 0.69 | 0.97 | 2.02E-02 | 0.287 |
| OID21501 | CIAPIN1 | Oncology | CIAPIN1 | Q6FI81 | 16 | 57428187 | 57447420 | 0.82 | 0.69 | 0.97 | 1.98E-02 | 0.287 |
| OID31503 | CIRBP | Oncology_II | CIRBP | Q14011 | 19 | 1259384 | 1274880 | 0.83 | 0.71 | 0.97 | 2.27E-02 | 0.287 |
| OID30400 | CKB | Cardiometabolic_II | CKB | P12277 | 14 | 103519667 | 103522833 | 1.24 | 1.04 | 1.47 | 1.76E-02 | 0.287 |
| OID30957 | CLGN | Neurology_II | CLGN | O14967 | 4 | 140388453 | 140427661 | 0.80 | 0.67 | 0.95 | 1.20E-02 | 0.287 |
| OID20347 | CNDP1 | Cardiometabolic | CNDP1 | Q96KN2 | 18 | 74534500 | 74587212 | 0.83 | 0.72 | 0.96 | 9.63E-03 | 0.287 |
| OID30285 | CNP | Cardiometabolic_II | CNP | P09543 | 17 | 41966763 | 41977740 | 0.83 | 0.70 | 0.97 | 2.16E-02 | 0.287 |
| OID31081 | CRYBB2 | Neurology_II | CRYBB2 | P43320 | 22 | 25212564 | 25231870 | 1.16 | 1.04 | 1.30 | 7.31E-03 | 0.287 |
| OID20228 | CTSL | Cardiometabolic | CTSL | P07711 | 9 | 87724051 | 87731469 | 1.19 | 1.03 | 1.38 | 1.65E-02 | 0.287 |
| OID30657 | DCTD | Inflammation_II | DCTD | P32321 | 4 | 182890060 | 182917936 | 0.83 | 0.71 | 0.97 | 2.06E-02 | 0.287 |
| OID21480 | DCXR | Oncology | DCXR | Q7Z4W1 | 17 | 82035136 | 82037709 | 0.80 | 0.66 | 0.96 | 1.57E-02 | 0.287 |
| OID30493 | DENR | Inflammation_II | DENR | O43583 | 12 | 122752824 | 122771064 | 0.80 | 0.67 | 0.96 | 1.71E-02 | 0.287 |
| OID20620 | DFFA | Inflammation | DFFA | O00273 | 1 | 10456522 | 10472529 | 0.82 | 0.69 | 0.97 | 2.22E-02 | 0.287 |
| OID30546 | DGKA | Inflammation_II | DGKA | P23743 | 12 | 55927319 | 55954027 | 0.78 | 0.66 | 0.93 | 6.56E-03 | 0.287 |
| OID31434 | DNAJB14 | Oncology_II | DNAJB14 | Q8TBM8 | 4 | 99896248 | 99946618 | 0.78 | 0.65 | 0.93 | 6.56E-03 | 0.287 |

| | | | | | | | | | | | | |
|---|---|---|---|---|---|---|---|---|---|---|---|---|
| OID30661 | DNAJB2 | Inflammation_II | DNAJB2 | P25686 | 2 | 219279342 | 219286898 | 0.81 | 0.68 | 0.97 | 1.84E-02 | 0.287 |
| OID30613 | DNAJB6 | Inflammation_II | DNAJB6 | O75190 | 7 | 157335381 | 157417439 | 0.81 | 0.69 | 0.96 | 1.41E-02 | 0.287 |
| OID30580 | DTD1 | Inflammation_II | DTD1 | Q8TEA8 | 20 | 18587942 | 18766644 | 0.81 | 0.68 | 0.96 | 1.26E-02 | 0.287 |
| OID31010 | EDF1 | Neurology_II | EDF1 | O60869 | 9 | 136862119 | 136866308 | 0.77 | 0.63 | 0.95 | 1.25E-02 | 0.287 |
| OID31442 | EIF2AK2 | Oncology_II | EIF2AK2 | P19525 | 2 | 37099210 | 37157522 | 0.81 | 0.68 | 0.96 | 1.67E-02 | 0.287 |
| OID21046 | ENO2 | Neurology | ENO2 | P09104 | 12 | 6913745 | 6923698 | 0.80 | 0.68 | 0.95 | 1.14E-02 | 0.287 |
| OID20905 | EREG | Neurology | EREG | O14944 | 4 | 74365145 | 74388749 | 0.80 | 0.67 | 0.95 | 1.06E-02 | 0.287 |
| OID21256 | FUS | Oncology | FUS | P35637 | 16 | 31180138 | 31191605 | 0.77 | 0.63 | 0.93 | 8.02E-03 | 0.287 |
| OID21423 | GALNT2 | Oncology | GALNT2 | Q10471 | 1 | 230057990 | 230282122 | 0.82 | 0.70 | 0.96 | 1.18E-02 | 0.287 |
| OID20251 | GDF15 | Cardiometabolic | GDF15 | Q99988 | 19 | 18374731 | 18389176 | 1.26 | 1.05 | 1.50 | 1.11E-02 | 0.287 |
| OID20838 | GGA1 | Neurology | GGA1 | Q9UJY5 | 22 | 37608725 | 37633564 | 0.75 | 0.61 | 0.93 | 8.83E-03 | 0.287 |
| OID30639 | GIMAP7 | Inflammation_II | GIMAP7 | Q8NHV1 | 7 | 150514872 | 150521073 | 0.80 | 0.66 | 0.96 | 1.83E-02 | 0.287 |
| OID30594 | GLA | Inflammation_II | GLA | P06280 | X | 101393273 | 101408012 | 0.82 | 0.70 | 0.96 | 1.31E-02 | 0.287 |
| OID30340 | GMFG | Cardiometabolic_II | GMFG | O60234 | 19 | 39328353 | 39342372 | 0.82 | 0.70 | 0.96 | 1.66E-02 | 0.287 |
| OID30623 | GMPR2 | Inflammation_II | GMPR2 | Q9P2T1 | 14 | 24232422 | 24239242 | 0.82 | 0.70 | 0.97 | 2.02E-02 | 0.287 |
| OID30597 | GNPDA2 | Inflammation_II | GNPDA2 | Q8TDQ7 | 4 | 44682200 | 44726588 | 0.83 | 0.70 | 0.97 | 2.07E-02 | 0.287 |
| OID31438 | GORASP2 | Oncology_II | GORASP2 | Q9H8Y8 | 2 | 170928464 | 170967130 | 0.80 | 0.68 | 0.95 | 1.22E-02 | 0.287 |
| OID20192 | GRAP2 | Cardiometabolic | GRAP2 | O75791 | 22 | 39901084 | 39973721 | 0.82 | 0.69 | 0.96 | 1.25E-02 | 0.287 |
| OID30289 | GRHPR | Cardiometabolic_II | GRHPR | Q9UBQ7 | 9 | 37422666 | 37436990 | 0.80 | 0.68 | 0.94 | 8.19E-03 | 0.287 |
| OID20166 | GSTA1 | Cardiometabolic | GSTA1 | P08263 | 6 | 52791371 | 52803860 | 0.77 | 0.64 | 0.94 | 8.41E-03 | 0.287 |
| OID21422 | HAVCR1 | Oncology | HAVCR1 | Q96D42 | 5 | 157029413 | 157059119 | 1.23 | 1.03 | 1.46 | 1.90E-02 | 0.287 |
| OID31136 | HDGFL2 | Neurology_II | HDGFL2 | Q7Z4V5 | 19 | 4472297 | 4502208 | 0.80 | 0.66 | 0.96 | 1.62E-02 | 0.287 |
| OID30718 | HGFAC | Inflammation_II | HGFAC | Q04756 | 4 | 3441968 | 3449486 | 0.81 | 0.69 | 0.94 | 7.53E-03 | 0.287 |
| OID31107 | HHEX | Neurology_II | HHEX | Q03014 | 10 | 92689955 | 92695647 | 0.82 | 0.69 | 0.97 | 2.17E-02 | 0.287 |
| OID30214 | HMGCL | Cardiometabolic_II | HMGCL | P35914 | 1 | 23801885 | 23838620 | 0.81 | 0.68 | 0.97 | 2.26E-02 | 0.287 |
| OID31424 | HSPA2 | Oncology_II | HSPA2 | P54652 | 14 | 64535905 | 64546173 | 0.80 | 0.66 | 0.97 | 2.04E-02 | 0.287 |
| OID20325 | IGFBP2 | Cardiometabolic | IGFBP2 | P18065 | 2 | 216632828 | 216664436 | 1.25 | 1.04 | 1.50 | 1.74E-02 | 0.287 |
| OID20155 | IGFBPL1 | Cardiometabolic | IGFBPL1 | Q8WX77 | 9 | 38406528 | 38424454 | 1.21 | 1.04 | 1.40 | 1.17E-02 | 0.287 |
| OID20562 | IL15 | Inflammation | IL15 | P40933 | 4 | 141636583 | 141733987 | 1.18 | 1.03 | 1.35 | 1.75E-02 | 0.287 |
| OID20633 | IL16 | Inflammation | IL16 | Q14005 | 15 | 81159575 | 81314058 | 0.78 | 0.64 | 0.94 | 7.83E-03 | 0.287 |
| OID31407 | IL22 | Oncology_II | IL22 | Q9GZX6 | 12 | 68248242 | 68253604 | 1.19 | 1.02 | 1.38 | 2.25E-02 | 0.287 |
| OID31112 | IMPACT | Neurology_II | IMPACT | Q9P2X3 | 18 | 24426634 | 24453531 | 0.82 | 0.70 | 0.97 | 2.07E-02 | 0.287 |
| OID30259 | INPP5D | Cardiometabolic_II | INPP5D | Q92835 | 2 | 233059967 | 233207903 | 0.79 | 0.66 | 0.94 | 7.87E-03 | 0.287 |
| OID31497 | IST1 | Oncology_II | IST1 | P53990 | 16 | 71845996 | 71931199 | 0.83 | 0.71 | 0.97 | 1.86E-02 | 0.287 |
| OID21071 | ITGAM | Neurology | ITGAM | P11215 | 16 | 31259967 | 31332892 | 0.83 | 0.70 | 0.97 | 2.18E-02 | 0.287 |
| OID30586 | JAM3 | Inflammation_II | JAM3 | Q9BX67 | 11 | 134069071 | 134152001 | 0.81 | 0.68 | 0.97 | 2.14E-02 | 0.287 |
| OID31520 | JPT2 | Oncology_II | JPT2 | Q9H910 | 16 | 1678256 | 1702280 | 0.81 | 0.69 | 0.96 | 1.18E-02 | 0.287 |
| OID30241 | KHK | Cardiometabolic_II | KHK | P50053 | 2 | 27086747 | 27100762 | 0.80 | 0.67 | 0.94 | 8.39E-03 | 0.287 |
| OID30806 | KIRREL1 | Neurology_II | KIRREL1 | Q96J84 | 1 | 157993273 | 158100262 | 0.75 | 0.60 | 0.94 | 1.26E-02 | 0.287 |
| OID21391 | KLK4 | Oncology | KLK4 | Q9Y5K2 | 19 | 50906351 | 50911395 | 1.21 | 1.03 | 1.41 | 1.83E-02 | 0.287 |
| OID20640 | LAT | Inflammation | LAT | O43561 | 16 | 28984826 | 28990784 | 0.83 | 0.71 | 0.97 | 1.89E-02 | 0.287 |
| OID21406 | LGALS7_LGALS7B | Oncology | GALS7B;LGALS7 | P47929;P47929 | 19;19 | 38789200;38770968 | 38791754;38773517 | 1.22 | 1.04 | 1.43 | 1.64E-02 | 0.287 |
| OID20606 | LIFR | Inflammation | LIFR | P42702 | 5 | 38474668 | 38608354 | 1.22 | 1.04 | 1.42 | 1.32E-02 | 0.287 |
| OID20254 | LTBP2 | Cardiometabolic | LTBP2 | Q14767 | 14 | 74498183 | 74612378 | 1.25 | 1.04 | 1.50 | 1.63E-02 | 0.287 |
| OID31293 | MAPKAPK2 | Oncology_II | MAPKAPK2 | P49137 | 1 | 206684905 | 206734281 | 0.81 | 0.69 | 0.96 | 1.63E-02 | 0.287 |
| OID20815 | METAP1 | Neurology | METAP1 | P53582 | 4 | 98995659 | 99062809 | 0.80 | 0.67 | 0.96 | 1.52E-02 | 0.287 |
| OID20959 | MITD1 | Neurology | MITD1 | Q8WV92 | 2 | 99161427 | 99181058 | 0.83 | 0.71 | 0.97 | 1.96E-02 | 0.287 |
| OID31327 | MNAT1 | Oncology_II | MNAT1 | P51948 | 14 | 60734742 | 60969965 | 0.79 | 0.64 | 0.96 | 1.82E-02 | 0.287 |
| OID20092 | MPHOSPH8 | Cardiometabolic | MPHOSPH8 | Q99549 | 13 | 19633659 | 19673441 | 0.81 | 0.68 | 0.97 | 1.92E-02 | 0.287 |
| OID30891 | MRI1 | Neurology_II | MRI1 | Q9BV20 | 19 | 13764522 | 13774282 | 0.82 | 0.69 | 0.96 | 1.60E-02 | 0.287 |
| OID20275 | MSMB | Cardiometabolic | MSMB | P08118 | 10 | 46033307 | 46048180 | 1.26 | 1.06 | 1.49 | 8.17E-03 | 0.287 |
| OID30188 | NAA10 | Cardiometabolic_II | NAA10 | P41227 | X | 153929225 | 153935080 | 0.83 | 0.71 | 0.97 | 1.85E-02 | 0.287 |
| OID20545 | NFATC1 | Inflammation | NFATC1 | O95644 | 18 | 79395856 | 79529325 | 0.81 | 0.69 | 0.96 | 1.49E-02 | 0.287 |
| OID30160 | NFKB1 | Cardiometabolic_II | NFKB1 | P19838 | 4 | 102501330 | 102617302 | 0.81 | 0.69 | 0.96 | 1.68E-02 | 0.287 |
| OID21212 | NFKBIE | Oncology | NFKBIE | O00221 | 6 | 44258166 | 44265788 | 0.78 | 0.65 | 0.94 | 7.52E-03 | 0.287 |
| OID30158 | NFX1 | Cardiometabolic_II | NFX1 | Q12986 | 9 | 33290512 | 33371157 | 0.82 | 0.69 | 0.97 | 2.14E-02 | 0.287 |
| OID30134 | NFYA | Cardiometabolic_II | NFYA | P23511 | 6 | 41072974 | 41102403 | 0.74 | 0.59 | 0.94 | 1.17E-02 | 0.287 |
| OID30614 | NHLRC3 | Inflammation_II | NHLRC3 | Q5JS37 | 13 | 39038306 | 39050109 | 0.81 | 0.69 | 0.96 | 1.52E-02 | 0.287 |
| OID31021 | NMT1 | Neurology_II | NMT1 | P30419 | 17 | 44957992 | 45109016 | 0.79 | 0.65 | 0.96 | 1.68E-02 | 0.287 |
| OID20049 | NPPB | Cardiometabolic | NPPB | P16860 | 1 | 11857464 | 11858945 | 1.24 | 1.05 | 1.45 | 9.35E-03 | 0.287 |
| OID30901 | NRXN3 | Neurology_II | NRXN3 | Q9Y4C0 | 14 | 78170373 | 79868291 | 0.78 | 0.63 | 0.96 | 1.69E-02 | 0.287 |
| OID20125 | NTproBNP | Cardiometabolic | NPPB | P16860 | 1 | 11857464 | 11858945 | 1.21 | 1.04 | 1.40 | 1.54E-02 | 0.287 |
| OID31484 | NUDT16 | Oncology_II | NUDT16 | Q96DE0 | 3 | 131381671 | 131388830 | 0.81 | 0.68 | 0.95 | 9.98E-03 | 0.287 |
| OID31465 | OGA | Oncology_II | OGA | O60502 | 10 | 101784443 | 101818465 | 0.80 | 0.67 | 0.95 | 1.07E-02 | 0.287 |
| OID31286 | PAFAH2 | Oncology_II | PAFAH2 | Q99487 | 1 | 25959767 | 25998117 | 0.81 | 0.68 | 0.97 | 2.05E-02 | 0.287 |
| OID31301 | PARD3 | Oncology_II | PARD3 | Q8TEW0 | 10 | 34109560 | 34815325 | 0.80 | 0.66 | 0.96 | 1.74E-02 | 0.287 |
| OID30591 | PAXX | Inflammation_II | PAXX | Q9BUH6 | 9 | 136992422 | 136993984 | 0.80 | 0.67 | 0.96 | 1.44E-02 | 0.287 |
| OID20313 | PDGFA | Cardiometabolic | PDGFA | P04085 | 7 | 497258 | 520296 | 0.82 | 0.70 | 0.96 | 1.48E-02 | 0.287 |
| OID21183 | PDGFC | Oncology | PDGFC | Q9NRA1 | 4 | 156760454 | 156971799 | 0.81 | 0.69 | 0.94 | 7.08E-03 | 0.287 |
| OID30666 | PENK | Inflammation_II | PENK | P01210 | 8 | 56436674 | 56446671 | 0.81 | 0.68 | 0.95 | 1.19E-02 | 0.287 |
| OID30764 | PF4 | Inflammation_II | PF4 | P02776 | 4 | 73980811 | 73982027 | 0.80 | 0.68 | 0.94 | 8.02E-03 | 0.287 |
| OID21356 | PFKFB2 | Oncology | PFKFB2 | O60825 | 1 | 207034366 | 207081024 | 0.81 | 0.69 | 0.95 | 1.17E-02 | 0.287 |
| OID31167 | PHLDB2 | Oncology_II | PHLDB2 | Q86SQ0 | 3 | 111732497 | 111976517 | 0.77 | 0.63 | 0.93 | 7.77E-03 | 0.287 |
| OID30634 | PHYKPL | Inflammation_II | PHYKPL | Q8IUZ5 | 5 | 178208471 | 178232802 | 0.79 | 0.66 | 0.94 | 6.74E-03 | 0.287 |
| OID20326 | PI3 | Cardiometabolic | PI3 | P19957 | 20 | 45174902 | 45176544 | 1.21 | 1.04 | 1.41 | 1.57E-02 | 0.287 |
| OID20144 | PLPBP | Cardiometabolic | PLPBP | O94903 | 8 | 37762595 | 37779768 | 0.80 | 0.68 | 0.95 | 8.80E-03 | 0.287 |
| OID20445 | PNPT1 | Inflammation | PNPT1 | Q8TCS8 | 2 | 55634061 | 55693863 | 0.77 | 0.63 | 0.95 | 1.31E-02 | 0.287 |
| OID30761 | PPBP | Inflammation_II | PPBP | P02775 | 4 | 73986439 | 73988190 | 0.82 | 0.70 | 0.96 | 1.35E-02 | 0.287 |
| OID21172 | PQBP1 | Oncology | PQBP1 | O60828 | X | 48890197 | 48903402 | 0.74 | 0.59 | 0.93 | 8.85E-03 | 0.287 |
| OID21225 | PRKRA | Oncology | PRKRA | O75569 | 2 | 178431292 | 178451512 | 0.82 | 0.69 | 0.97 | 2.24E-02 | 0.287 |
| OID20543 | PROK1 | Inflammation | PROK1 | P58294 | 1 | 110451149 | 110457358 | 1.29 | 1.05 | 1.59 | 1.57E-02 | 0.287 |
| OID20511 | PSPN | Inflammation | PSPN | O60542 | 19 | 6375148 | 6379058 | 0.76 | 0.60 | 0.96 | 2.17E-02 | 0.287 |
| OID21169 | PSRC1 | Oncology | PSRC1 | Q6PGN9 | 1 | 109279556 | 109283186 | 0.78 | 0.65 | 0.94 | 7.11E-03 | 0.287 |
| OID31461 | PTPRK | Oncology_II | PTPRK | Q15262 | 6 | 127968785 | 128520616 | 0.79 | 0.65 | 0.96 | 2.00E-02 | 0.287 |
| OID31254 | RAD51 | Oncology_II | RAD51 | Q06609 | 15 | 40694774 | 40732340 | 0.76 | 0.62 | 0.95 | 1.41E-02 | 0.287 |
| OID31337 | RAPGEF2 | Oncology_II | RAPGEF2 | Q9Y4G8 | 4 | 159103013 | 159360174 | 0.79 | 0.66 | 0.95 | 1.11E-02 | 0.287 |
| OID31236 | RBM25 | Oncology_II | RBM25 | P49756 | 14 | 73058532 | 73123899 | 0.81 | 0.68 | 0.96 | 1.65E-02 | 0.287 |
| OID31060 | RGCC | Neurology_II | RGCC | Q9H4X1 | 13 | 41457550 | 41470871 | 0.80 | 0.68 | 0.94 | 7.07E-03 | 0.287 |
| OID30995 | RLN1 | Neurology_II | RLN1 | P04808 | 9 | 5334930 | 5339876 | 0.60 | 0.39 | 0.93 | 2.10E-02 | 0.287 |
| OID30961 | RRAS | Neurology_II | RRAS | P10301 | 19 | 49635292 | 49640143 | 0.81 | 0.68 | 0.96 | 1.75E-02 | 0.287 |
| OID31012 | SAFB2 | Neurology_II | SAFB2 | Q14151 | 19 | 5586999 | 5624046 | 0.78 | 0.65 | 0.94 | 1.03E-02 | 0.287 |
| OID30874 | SAG | Neurology_II | SAG | P10523 | 2 | 233307816 | 233347055 | 0.80 | 0.67 | 0.96 | 1.38E-02 | 0.287 |
| OID30278 | SCPEP1 | Cardiometabolic_II | SCPEP1 | Q9HB40 | 17 | 56978129 | 57006768 | 0.81 | 0.68 | 0.96 | 1.71E-02 | 0.287 |
| OID30269 | SEC31A | Cardiometabolic_II | SEC31A | O94979 | 4 | 82818509 | 82901166 | 0.83 | 0.70 | 0.97 | 2.18E-02 | 0.287 |
| OID30763 | SERPINA5 | Inflammation_II | SERPINA5 | P05154 | 14 | 94561442 | 94593118 | 0.78 | 0.64 | 0.94 | 1.03E-02 | 0.287 |
| OID30359 | SERPINE2 | Cardiometabolic_II | SERPINE2 | P07093 | 2 | 223975045 | 224039318 | 0.80 | 0.68 | 0.95 | 9.56E-03 | 0.287 |
| OID21222 | SH2B3 | Oncology | SH2B3 | Q9UQQ2 | 12 | 111405923 | 111451623 | 0.83 | 0.70 | 0.97 | 2.13E-02 | 0.287 |
| OID20514 | SH2D1A | Inflammation | SH2D1A | O60880 | X | 124227868 | 124373197 | 0.80 | 0.66 | 0.97 | 2.19E-02 | 0.287 |
| OID30853 | SMS | Neurology_II | SMS | P52788 | X | 21940709 | 21994837 | 0.77 | 0.63 | 0.93 | 7.65E-03 | 0.287 |
| OID21511 | SPARC | Oncology | SPARC | P09486 | 5 | 151661096 | 151686975 | 0.83 | 0.71 | 0.97 | 1.75E-02 | 0.287 |
| OID20759 | SPON1 | Inflammation | SPON1 | Q9HCB6 | 11 | 13962723 | 14268133 | 1.21 | 1.05 | 1.41 | 9.42E-03 | 0.287 |
| OID30624 | ST13 | Inflammation_II | ST13 | P50502 | 22 | 40824535 | 40856639 | 0.82 | 0.70 | 0.97 | 1.83E-02 | 0.287 |
| OID20140 | STK4 | Cardiometabolic | STK4 | Q13043 | 20 | 44966479 | 45080021 | 0.82 | 0.70 | 0.97 | 2.12E-02 | 0.287 |
| OID31524 | SUGP1 | Oncology_II | SUGP1 | Q8IWZ8 | 19 | 19276018 | 19320509 | 0.80 | 0.66 | 0.97 | 2.26E-02 | 0.287 |
| OID21447 | TACSTD2 | Oncology | TACSTD2 | P09758 | 1 | 58575433 | 58577252 | 1.24 | 1.05 | 1.46 | 1.01E-02 | 0.287 |
| OID30616 | TBCA | Inflammation_II | TBCA | O75347 | 5 | 77691166 | 77868780 | 0.81 | 0.69 | 0.96 | 1.44E-02 | 0.287 |
| OID20987 | TCL1A | Neurology | TCL1A | P56279 | 14 | 95709947 | 95714196 | 0.82 | 0.70 | 0.97 | 2.03E-02 | 0.287 |

| | | | | | | | | | | | | |
|---|---|---|---|---|---|---|---|---|---|---|---|---|
| OID31417 | TGFB2 | Oncology_II | TGFB2 | P61812 | 1 | 218345336 | 218444619 | 0.80 | 0.67 | 0.95 | 1.25E-02 | 0.287 |
| OID20194 | THOP1 | Cardiometabolic | THOP1 | P52888 | 19 | 2785503 | 2815807 | 0.81 | 0.67 | 0.97 | 1.93E-02 | 0.287 |
| OID31375 | THTPA | Oncology_II | THTPA | Q9BU02 | 14 | 23555988 | 23560271 | 0.80 | 0.68 | 0.94 | 6.44E-03 | 0.287 |
| OID20303 | TIE1 | Cardiometabolic | TIE1 | P35590 | 1 | 43300982 | 43323108 | 1.26 | 1.03 | 1.54 | 2.20E-02 | 0.287 |
| OID21147 | TIMP4 | Neurology | TIMP4 | Q99727 | 3 | 12153068 | 12158912 | 1.21 | 1.04 | 1.41 | 1.16E-02 | 0.287 |
| OID30940 | TMCO5A | Neurology_II | TMCO5A | Q8N6Q1 | 15 | 37921939 | 37967724 | 0.81 | 0.68 | 0.97 | 1.95E-02 | 0.287 |
| OID31446 | TNFAIP2 | Oncology_II | TNFAIP2 | Q03169 | 14 | 103121476 | 103137439 | 0.79 | 0.65 | 0.96 | 2.01E-02 | 0.287 |
| OID30629 | TNFAIP8L2 | Inflammation_II | TNFAIP8L2 | Q6P589 | 1 | 151156649 | 151159749 | 0.80 | 0.67 | 0.96 | 1.58E-02 | 0.287 |
| OID30609 | TPD52L2 | Inflammation_II | TPD52L2 | O43399 | 20 | 63865228 | 63891545 | 0.84 | 0.72 | 0.98 | 2.27E-02 | 0.287 |
| OID30338 | TPK1 | Cardiometabolic_II | TPK1 | Q9H3S4 | 7 | 144451941 | 144836395 | 0.82 | 0.70 | 0.97 | 1.78E-02 | 0.287 |
| OID31368 | UBE2B | Oncology_II | UBE2B | P63146 | 5 | 134371184 | 134392108 | 0.82 | 0.68 | 0.97 | 2.24E-02 | 0.287 |
| OID30654 | UBXN1 | Inflammation_II | UBXN1 | Q04323 | 11 | 62676498 | 62679117 | 0.81 | 0.68 | 0.97 | 1.99E-02 | 0.287 |
| OID21367 | USO1 | Oncology | USO1 | O60763 | 4 | 75724577 | 75814286 | 0.83 | 0.71 | 0.97 | 2.16E-02 | 0.287 |
| OID30193 | VIPR1 | Cardiometabolic_II | VIPR1 | P32241 | 3 | 42489299 | 42537573 | 0.79 | 0.65 | 0.95 | 1.42E-02 | 0.287 |
| OID31386 | VPS4B | Oncology_II | VPS4B | O75351 | 18 | 63389190 | 63422483 | 0.82 | 0.69 | 0.97 | 1.81E-02 | 0.287 |
| OID31404 | YJU2 | Oncology_II | YJU2 | Q9BW85 | 19 | 4247080 | 4269088 | 0.79 | 0.65 | 0.95 | 1.30E-02 | 0.287 |
| OID30605 | YWHAQ | Inflammation_II | YWHAQ | P27348 | 2 | 9583967 | 9630997 | 0.81 | 0.69 | 0.96 | 1.50E-02 | 0.287 |
| OID21205 | ZBTB16 | Oncology | ZBTB16 | Q05516 | 11 | 114059041 | 114256765 | 0.81 | 0.68 | 0.97 | 2.01E-02 | 0.287 |
| OID31347 | ZNRF4 | Oncology_II | ZNRF4 | Q8WWF5 | 19 | 5455417 | 5456856 | 0.79 | 0.66 | 0.94 | 8.45E-03 | 0.287 |
| OID31466 | ATG16L1 | Oncology_II | ATG16L1 | Q676U5 | 2 | 233210051 | 233295674 | 0.83 | 0.70 | 0.98 | 2.40E-02 | 0.287 |
| OID30541 | BLNK | Inflammation_II | BLNK | Q8WV28 | 10 | 96189171 | 96271587 | 0.79 | 0.64 | 0.97 | 2.40E-02 | 0.287 |
| OID30766 | C7 | Inflammation_II | C7 | P10643 | 5 | 40909492 | 40984643 | 1.21 | 1.03 | 1.44 | 2.37E-02 | 0.287 |
| OID30270 | EIF2S2 | Cardiometabolic_II | EIF2S2 | P20042 | 20 | 34088309 | 34112243 | 0.80 | 0.66 | 0.97 | 2.34E-02 | 0.287 |
| OID20861 | GDNF | Neurology | GDNF | P39905 | 5 | 37812677 | 37840041 | 1.19 | 1.02 | 1.38 | 2.39E-02 | 0.287 |
| OID30700 | GSR | Inflammation_II | GSR | P00390 | 8 | 30678066 | 30727846 | 0.82 | 0.69 | 0.97 | 2.38E-02 | 0.287 |
| OID30658 | MARS1 | Inflammation_II | MARS1 | P56192 | 12 | 57475445 | 57517569 | 0.82 | 0.70 | 0.97 | 2.34E-02 | 0.287 |
| OID21352 | OMG | Oncology | OMG | P23515 | 17 | 31272013 | 31297539 | 0.83 | 0.71 | 0.98 | 2.39E-02 | 0.287 |
| OID20500 | PARP1 | Inflammation | PARP1 | P09874 | 1 | 226360210 | 226408154 | 0.77 | 0.61 | 0.96 | 2.35E-02 | 0.287 |
| OID31410 | PCSK7 | Oncology_II | PCSK7 | Q16549 | 11 | 117204337 | 117232525 | 0.80 | 0.67 | 0.97 | 2.32E-02 | 0.287 |
| OID20717 | PTPN6 | Inflammation | PTPN6 | P29350 | 12 | 6946468 | 6961316 | 0.84 | 0.72 | 0.98 | 2.38E-02 | 0.287 |
| OID21232 | TP53 | Oncology | TP53 | P04637 | 17 | 7661779 | 7687538 | 0.83 | 0.71 | 0.97 | 2.32E-02 | 0.287 |
| OID20478 | YTHDF3 | Inflammation | YTHDF3 | Q7Z739 | 8 | 63168553 | 63212786 | 0.83 | 0.71 | 0.98 | 2.40E-02 | 0.287 |
| OID20442 | NBN | Inflammation | NBN | O60934 | 8 | 89933331 | 90003228 | 0.78 | 0.63 | 0.97 | 2.41E-02 | 0.288 |
| OID30083 | EXOSC10 | Cardiometabolic_II | EXOSC10 | Q01780 | 1 | 11066618 | 11099869 | 0.77 | 0.62 | 0.97 | 2.45E-02 | 0.289 |
| OID30946 | GTPBP2 | Neurology_II | GTPBP2 | Q9BX10 | 6 | 43605316 | 43629264 | 0.82 | 0.69 | 0.97 | 2.45E-02 | 0.289 |
| OID20150 | LACTB2 | Cardiometabolic | LACTB2 | Q53H82 | 8 | 70635318 | 70669185 | 0.83 | 0.70 | 0.98 | 2.46E-02 | 0.290 |
| OID20305 | CASP3 | Cardiometabolic | CASP3 | P42574 | 4 | 184627696 | 184649509 | 0.83 | 0.71 | 0.98 | 2.52E-02 | 0.290 |
| OID20742 | CRKL | Inflammation | CRKL | P46109 | 22 | 20917407 | 20953747 | 0.83 | 0.71 | 0.98 | 2.54E-02 | 0.290 |
| OID21226 | DDX58 | Oncology | DDX58 | O95786 | 9 | 32455302 | 32526208 | 0.80 | 0.65 | 0.97 | 2.55E-02 | 0.290 |
| OID31062 | GIGYF2 | Neurology_II | GIGYF2 | Q6Y7W6 | 2 | 232697299 | 232860605 | 0.81 | 0.68 | 0.97 | 2.53E-02 | 0.290 |
| OID20577 | IRAK4 | Inflammation | IRAK4 | Q9NWZ3 | 12 | 43758944 | 43789543 | 0.84 | 0.72 | 0.98 | 2.54E-02 | 0.290 |
| OID30986 | KLHL41 | Neurology_II | KLHL41 | O60662 | 2 | 169509702 | 169526258 | 0.80 | 0.66 | 0.97 | 2.59E-02 | 0.290 |
| OID20800 | KRT14 | Neurology | KRT14 | P02533 | 17 | 41582279 | 41586895 | 1.15 | 1.02 | 1.30 | 2.59E-02 | 0.290 |
| OID21519 | S100A4 | Oncology | S100A4 | P26447 | 1 | 153543613 | 153550136 | 0.83 | 0.71 | 0.98 | 2.57E-02 | 0.290 |
| OID31306 | SAT2 | Oncology_II | SAT2 | Q96F10 | 17 | 7626234 | 7627876 | 0.79 | 0.65 | 0.97 | 2.54E-02 | 0.290 |
| OID21020 | SEMA4D | Neurology | SEMA4D | Q92854 | 9 | 89360787 | 89498130 | 0.83 | 0.71 | 0.98 | 2.50E-02 | 0.290 |
| OID21088 | SERPINB1 | Neurology | SERPINB1 | P30740 | 6 | 2832332 | 2841959 | 0.82 | 0.69 | 0.98 | 2.59E-02 | 0.290 |
| OID31334 | SH3GL3 | Oncology_II | SH3GL3 | Q99963 | 15 | 83447228 | 83618743 | 0.83 | 0.70 | 0.98 | 2.59E-02 | 0.290 |
| OID20480 | TNFRSF13C | Inflammation | TNFRSF13C | Q96RJ3 | 22 | 41922032 | 41926806 | 0.83 | 0.70 | 0.98 | 2.57E-02 | 0.290 |
| OID30356 | ACADSB | Cardiometabolic_II | ACADSB | P45954 | 10 | 123008979 | 123058290 | 0.84 | 0.71 | 0.99 | 3.42E-02 | 0.290 |
| OID30655 | ADD1 | Inflammation_II | ADD1 | P35611 | 4 | 2843844 | 2930076 | 0.83 | 0.71 | 0.98 | 2.97E-02 | 0.290 |
| OID30877 | ADGRV1 | Neurology_II | ADGRV1 | Q8WXG9 | 5 | 90529344 | 91164437 | 0.81 | 0.66 | 0.98 | 2.88E-02 | 0.290 |
| OID20806 | AMFR | Neurology | AMFR | Q9UKV5 | 16 | 56361452 | 56425545 | 0.81 | 0.67 | 0.98 | 3.27E-02 | 0.290 |
| OID20211 | ANGPTL1 | Cardiometabolic | ANGPTL1 | O95841 | 1 | 178849535 | 178871077 | 1.20 | 1.02 | 1.41 | 3.00E-02 | 0.290 |
| OID31478 | AP3B1 | Oncology_II | AP3B1 | O00203 | 5 | 78000522 | 78294762 | 0.83 | 0.70 | 0.99 | 3.56E-02 | 0.290 |
| OID30861 | ARID3A | Neurology_II | ARID3A | Q99856 | 19 | 925781 | 975939 | 0.82 | 0.69 | 0.98 | 3.22E-02 | 0.290 |
| OID30170 | ASS1 | Cardiometabolic_II | ASS1 | P00966 | 9 | 130444961 | 130501274 | 0.82 | 0.69 | 0.98 | 3.08E-02 | 0.290 |
| OID30167 | ATP6V1G2 | Cardiometabolic_II | ATP6V1G2 | O95670 | 6 | 31544444 | 31548427 | 1.17 | 1.01 | 1.34 | 3.03E-02 | 0.290 |
| OID20594 | BANK1 | Inflammation | BANK1 | Q8NDB2 | 4 | 101411286 | 102074812 | 0.84 | 0.72 | 0.99 | 3.43E-02 | 0.290 |
| OID21198 | BGN | Oncology | BGN | P21810 | X | 153494980 | 153509546 | 1.19 | 1.02 | 1.39 | 3.19E-02 | 0.290 |
| OID31106 | BLOC1S3 | Neurology_II | BLOC1S3 | Q6QNY0 | 19 | 45178784 | 45216933 | 0.82 | 0.68 | 0.98 | 2.87E-02 | 0.290 |
| OID21096 | CA6 | Neurology | CA6 | P23280 | 1 | 8945867 | 8975092 | 0.83 | 0.70 | 0.98 | 3.30E-02 | 0.290 |
| OID30649 | CACYBP | Inflammation_II | CACYBP | Q9HB71 | 1 | 174999163 | 175012027 | 0.84 | 0.71 | 0.98 | 2.75E-02 | 0.290 |
| OID30921 | CASC3 | Neurology_II | CASC3 | O15234 | 17 | 40140318 | 40172171 | 0.82 | 0.69 | 0.98 | 3.04E-02 | 0.290 |
| OID30530 | CASP9 | Inflammation_II | CASP9 | P55211 | 1 | 15490832 | 15526534 | 0.82 | 0.69 | 0.98 | 3.10E-02 | 0.290 |
| OID20179 | CBLIF | Cardiometabolic | CBLIF | P27352 | 11 | 59829273 | 59845499 | 0.86 | 0.75 | 0.99 | 3.12E-02 | 0.290 |
| OID31077 | CCAR2 | Neurology_II | CCAR2 | Q8N163 | 8 | 22604757 | 22620964 | 0.78 | 0.62 | 0.98 | 3.52E-02 | 0.290 |
| OID20655 | CCL13 | Inflammation | CCL13 | Q99616 | 17 | 34356480 | 34358610 | 0.84 | 0.71 | 0.99 | 3.29E-02 | 0.290 |
| OID20412 | CCL5 | Cardiometabolic | CCL5 | P13501 | 17 | 35871491 | 35880793 | 0.84 | 0.72 | 0.98 | 2.77E-02 | 0.290 |
| OID20616 | CD40LG | Inflammation | CD40LG | P29965 | X | 136648158 | 136660390 | 0.84 | 0.72 | 0.98 | 3.04E-02 | 0.290 |
| OID20988 | CD63 | Neurology | CD63 | P08962 | 12 | 55725323 | 55729707 | 0.84 | 0.72 | 0.99 | 3.25E-02 | 0.290 |
| OID20721 | CKMT1A_CKMT1B | Inflammation | CKMT1B;CKMT1A | P12532;P12532 | 15;15 | 43593054;43692886 | 43604901;43699222 | 1.21 | 1.02 | 1.43 | 2.83E-02 | 0.290 |
| OID20831 | CLSPN | Neurology | CLSPN | Q9HAW4 | 1 | 35720218 | 35769978 | 0.80 | 0.65 | 0.98 | 3.18E-02 | 0.290 |
| OID30305 | COCH | Cardiometabolic_II | COCH | O43405 | 14 | 30874514 | 30895065 | 0.83 | 0.70 | 0.98 | 2.63E-02 | 0.290 |
| OID30272 | CRYZL1 | Cardiometabolic_II | CRYZL1 | O95825 | 21 | 33589341 | 33643926 | 0.84 | 0.71 | 0.98 | 3.01E-02 | 0.290 |
| OID20995 | CST5 | Neurology | CST5 | P28325 | 20 | 23875934 | 23879748 | 0.82 | 0.69 | 0.98 | 3.00E-02 | 0.290 |
| OID30664 | DAAM1 | Inflammation_II | DAAM1 | Q9Y4D1 | 14 | 59188646 | 59371405 | 0.83 | 0.70 | 0.98 | 2.78E-02 | 0.290 |
| OID30505 | DAPK2 | Inflammation_II | DAPK2 | Q9UIK4 | 15 | 63907036 | 64072033 | 0.81 | 0.66 | 0.98 | 3.30E-02 | 0.290 |
| OID31071 | DLG4 | Neurology_II | DLG4 | P78352 | 17 | 7187187 | 7219841 | 0.82 | 0.69 | 0.98 | 3.00E-02 | 0.290 |
| OID20827 | DUSP3 | Neurology | DUSP3 | P51452 | 17 | 43766125 | 43778977 | 0.82 | 0.68 | 0.99 | 3.47E-02 | 0.290 |
| OID31080 | DXO | Neurology_II | DXO | O77932 | 6 | 31969810 | 31972290 | 0.83 | 0.70 | 0.99 | 3.55E-02 | 0.290 |
| OID31415 | DYNLT3 | Oncology_II | DYNLT3 | P51808 | X | 37836757 | 37847571 | 0.83 | 0.70 | 0.99 | 3.32E-02 | 0.290 |
| OID30519 | EVI5 | Inflammation_II | EVI5 | O60447 | 1 | 92508696 | 92792404 | 0.83 | 0.70 | 0.98 | 2.85E-02 | 0.290 |
| OID30248 | FAM172A | Cardiometabolic_II | FAM172A | Q8WUF8 | 5 | 93617725 | 94111699 | 0.83 | 0.70 | 0.98 | 3.07E-02 | 0.290 |
| OID30378 | FBLN2 | Cardiometabolic_II | FBLN2 | P98095 | 3 | 13549125 | 13638422 | 1.19 | 1.01 | 1.39 | 3.54E-02 | 0.290 |
| OID21215 | FCGR2B | Oncology | FCGR2B | P31994 | 1 | 161663143 | 161678654 | 0.83 | 0.70 | 0.98 | 3.03E-02 | 0.290 |
| OID21271 | FOXO3 | Oncology | FOXO3 | O43524 | 6 | 108559835 | 108684774 | 0.82 | 0.69 | 0.99 | 3.46E-02 | 0.290 |
| OID30197 | GNPDA1 | Cardiometabolic_II | GNPDA1 | P46926 | 5 | 141991749 | 142013041 | 0.79 | 0.64 | 0.98 | 2.82E-02 | 0.290 |
| OID31026 | GOLGA3 | Neurology_II | GOLGA3 | Q08378 | 12 | 132768902 | 132829143 | 0.82 | 0.69 | 0.98 | 2.65E-02 | 0.290 |
| OID21242 | GSTA3 | Oncology | GSTA3 | Q16772 | 6 | 52896639 | 52909698 | 0.81 | 0.67 | 0.98 | 2.85E-02 | 0.290 |
| OID20663 | GZMA | Inflammation | GZMA | P12544 | 5 | 55102646 | 55110252 | 0.83 | 0.70 | 0.98 | 2.65E-02 | 0.290 |
| OID31037 | HNRNPUL1 | Neurology_II | HNRNPUL1 | Q9BUJ2 | 19 | 41262496 | 41307787 | 0.80 | 0.65 | 0.98 | 3.23E-02 | 0.290 |
| OID30962 | IFT20 | Neurology_II | IFT20 | Q8IY31 | 17 | 28328325 | 28335489 | 0.82 | 0.68 | 0.98 | 2.90E-02 | 0.290 |
| OID30123 | IGHMBP2 | Cardiometabolic_II | IGHMBP2 | P38935 | 11 | 68903863 | 68940602 | 0.81 | 0.68 | 0.98 | 2.73E-02 | 0.290 |
| OID20544 | IKBKG | Inflammation | IKBKG | Q9Y6K9 | X | 154541199 | 154565046 | 0.84 | 0.71 | 0.99 | 3.41E-02 | 0.290 |
| OID20504 | ITM2A | Inflammation | ITM2A | O43736 | X | 79360384 | 79367667 | 1.18 | 1.01 | 1.38 | 3.32E-02 | 0.290 |
| OID20669 | KYNU | Inflammation | KYNU | Q16719 | 2 | 142877657 | 143055833 | 0.83 | 0.70 | 0.99 | 3.52E-02 | 0.290 |
| OID31068 | LDLRAP1 | Neurology_II | LDLRAP1 | Q5SW96 | 1 | 25543606 | 25568886 | 0.83 | 0.71 | 0.98 | 3.10E-02 | 0.290 |
| OID20154 | LGALS1 | Cardiometabolic | LGALS1 | P09382 | 22 | 37675636 | 37679802 | 0.82 | 0.69 | 0.99 | 3.53E-02 | 0.290 |
| OID31180 | LYZL2 | Oncology_II | LYZL2 | Q7Z4W2 | 10 | 30611779 | 30629762 | 0.80 | 0.66 | 0.98 | 3.16E-02 | 0.290 |
| OID20711 | MGLL | Inflammation | MGLL | Q99685 | 3 | 127689062 | 128052190 | 0.84 | 0.72 | 0.99 | 3.49E-02 | 0.290 |
| OID31156 | MINK1 | Neurology_II | MINK1 | Q8N4C8 | 17 | 4833340 | 4898061 | 0.84 | 0.72 | 0.99 | 3.41E-02 | 0.290 |
| OID21513 | MSRA | Oncology | MSRA | Q9UJ68 | 8 | 10054292 | 10428891 | 0.83 | 0.71 | 0.98 | 3.12E-02 | 0.290 |
| OID31078 | MTHFD2 | Neurology_II | MTHFD2 | P13995 | 2 | 74186172 | 74217565 | 0.81 | 0.66 | 0.98 | 3.30E-02 | 0.290 |
| OID31475 | NAP1L4 | Oncology_II | NAP1L4 | Q99733 | 11 | 2944431 | 2992377 | 0.84 | 0.72 | 0.98 | 3.11E-02 | 0.290 |
| OID30268 | NFE2 | Cardiometabolic_II | NFE2 | Q16621 | 12 | 54292111 | 54301015 | 0.84 | 0.71 | 0.99 | 3.54E-02 | 0.290 |
| OID30336 | NIT2 | Cardiometabolic_II | NIT2 | Q9NQR4 | 3 | 100334739 | 100361635 | 0.84 | 0.71 | 0.99 | 3.23E-02 | 0.290 |

| | | | | | | | | | | | | |
|---|---|---|---|---|---|---|---|---|---|---|---|---|
| OID30568 | NUMB | Inflammation_II | NUMB | P49757 | 14 | 73275107 | 73458617 | 0.84 | 0.71 | 0.99 | 3.51E-02 | 0.290 |
| OID30835 | OMP | Neurology_II | OMP | P47874 | 11 | 77102840 | 77103331 | 1.15 | 1.01 | 1.31 | 3.40E-02 | 0.290 |
| OID20741 | PDGFB | Inflammation | PDGFB | P01127 | 22 | 39223359 | 39244982 | 0.84 | 0.72 | 0.98 | 3.09E-02 | 0.290 |
| OID31088 | PLEKHO1 | Neurology_II | PLEKHO1 | Q53GL0 | 1 | 150149183 | 150164720 | 0.83 | 0.70 | 0.98 | 3.02E-02 | 0.290 |
| OID20176 | PPP1R2 | Cardiometabolic | PPP1R2 | P41236 | 3 | 195514428 | 195543386 | 0.83 | 0.70 | 0.99 | 3.46E-02 | 0.290 |
| OID31502 | PPP2R5A | Oncology_II | PPP2R5A | Q15172 | 1 | 212285410 | 212361853 | 0.83 | 0.70 | 0.99 | 3.56E-02 | 0.290 |
| OID31119 | PRKAR2A | Neurology_II | PRKAR2A | P13861 | 3 | 48744597 | 48847874 | 0.83 | 0.71 | 0.98 | 3.23E-02 | 0.290 |
| OID30596 | PSTPIP2 | Inflammation_II | PSTPIP2 | Q9H939 | 18 | 45983536 | 46072272 | 0.84 | 0.72 | 0.98 | 2.96E-02 | 0.290 |
| OID30659 | PTRHD1 | Inflammation_II | PTRHD1 | Q6GMV3 | 2 | 24789728 | 24793391 | 0.84 | 0.71 | 0.98 | 3.15E-02 | 0.290 |
| OID21166 | RAD23B | Oncology | RAD23B | P54727 | 9 | 107283137 | 107332192 | 0.83 | 0.70 | 0.99 | 3.58E-02 | 0.290 |
| OID30999 | RGS10 | Neurology_II | RGS10 | O43665 | 10 | 119499817 | 119542719 | 0.83 | 0.70 | 0.98 | 2.93E-02 | 0.290 |
| OID30342 | RILPL2 | Cardiometabolic_II | RILPL2 | Q969X0 | 12 | 123415039 | 123436684 | 0.84 | 0.72 | 0.99 | 3.36E-02 | 0.290 |
| OID31324 | RPE | Oncology_II | RPE | Q96AT9 | 2 | 210002565 | 210022260 | 0.82 | 0.69 | 0.98 | 3.35E-02 | 0.290 |
| OID21425 | SCLY | Oncology | SCLY | Q96I15 | 2 | 238060924 | 238099413 | 0.82 | 0.69 | 0.98 | 3.22E-02 | 0.290 |
| OID20932 | SERPINB9 | Neurology | SERPINB9 | P50453 | 6 | 2887270 | 2903309 | 0.84 | 0.72 | 0.99 | 3.33E-02 | 0.290 |
| OID20790 | SESTD1 | Neurology | SESTD1 | Q86VW0 | 2 | 179101678 | 179264832 | 0.83 | 0.70 | 0.98 | 2.91E-02 | 0.290 |
| OID31418 | SMAD3 | Oncology_II | SMAD3 | P84022 | 15 | 67063763 | 67195173 | 0.83 | 0.69 | 0.98 | 3.10E-02 | 0.290 |
| OID21506 | SNAP29 | Oncology | SNAP29 | O95721 | 22 | 20859007 | 20891214 | 0.84 | 0.72 | 0.99 | 3.26E-02 | 0.290 |
| OID30052 | SNU13 | Cardiometabolic_II | SNU13 | P55769 | 22 | 41673933 | 41690504 | 0.78 | 0.62 | 0.97 | 2.80E-02 | 0.290 |
| OID30644 | SNX15 | Inflammation_II | SNX15 | Q9NRS6 | 11 | 65027439 | 65040572 | 0.82 | 0.69 | 0.98 | 2.68E-02 | 0.290 |
| OID20135 | SNX9 | Cardiometabolic | SNX9 | Q9Y5X1 | 6 | 157700387 | 157945077 | 0.82 | 0.69 | 0.98 | 2.79E-02 | 0.290 |
| OID21097 | SPINK1 | Neurology | SPINK1 | P00995 | 5 | 147824572 | 147831671 | 1.17 | 1.01 | 1.35 | 3.58E-02 | 0.290 |
| OID21521 | SRC | Oncology | SRC | P12931 | 20 | 37344685 | 37406050 | 0.84 | 0.71 | 0.99 | 3.48E-02 | 0.290 |
| OID20955 | STAMBP | Neurology | STAMBP | O95630 | 2 | 73828916 | 73873659 | 0.83 | 0.71 | 0.99 | 3.42E-02 | 0.290 |
| OID30536 | STAT2 | Inflammation_II | STAT2 | P52630 | 12 | 56341597 | 56360203 | 0.83 | 0.70 | 0.98 | 3.13E-02 | 0.290 |
| OID21277 | STAT5B | Oncology | STAT5B | P51692 | 17 | 42199177 | 42276707 | 0.84 | 0.72 | 0.99 | 3.32E-02 | 0.290 |
| OID20072 | SUSD1 | Cardiometabolic | SUSD1 | Q6UWL2 | 9 | 112040783 | 112175297 | 0.85 | 0.73 | 0.99 | 3.43E-02 | 0.290 |
| OID21285 | TACC3 | Oncology | TACC3 | Q9Y6A5 | 4 | 1712858 | 1745171 | 0.83 | 0.71 | 0.98 | 2.75E-02 | 0.290 |
| OID20870 | TARBP2 | Neurology | TARBP2 | Q15633 | 12 | 53500921 | 53506431 | 0.83 | 0.71 | 0.99 | 3.37E-02 | 0.290 |
| OID21220 | TBL1X | Oncology | TBL1X | O60907 | X | 9463320 | 9741037 | 0.83 | 0.71 | 0.98 | 2.61E-02 | 0.290 |
| OID20816 | TDGF1 | Neurology | TDGF1 | P13385 | 3 | 46574534 | 46582457 | 1.18 | 1.01 | 1.37 | 3.27E-02 | 0.290 |
| OID31403 | TDP1 | Oncology_II | TDP1 | Q9NUW8 | 14 | 89954939 | 90044764 | 0.80 | 0.65 | 0.98 | 3.45E-02 | 0.290 |
| OID20433 | TNFAIP8 | Inflammation | TNFAIP8 | O95379 | 5 | 119268692 | 119399688 | 0.83 | 0.70 | 0.98 | 3.25E-02 | 0.290 |
| OID31120 | TNIP1 | Neurology_II | TNIP1 | Q15025 | 5 | 151029945 | 151093577 | 0.83 | 0.70 | 0.98 | 3.09E-02 | 0.290 |
| OID31482 | TRIM25 | Oncology_II | TRIM25 | Q14258 | 17 | 56836387 | 56914080 | 0.84 | 0.71 | 0.99 | 3.46E-02 | 0.290 |
| OID30185 | TSNAX | Cardiometabolic_II | TSNAX | Q99598 | 1 | 231528541 | 231566524 | 0.83 | 0.69 | 0.98 | 3.29E-02 | 0.290 |
| OID30670 | TXN | Inflammation_II | TXN | P10599 | 9 | 110243810 | 110256507 | 0.81 | 0.67 | 0.98 | 3.15E-02 | 0.290 |
| OID31486 | UFD1 | Oncology_II | UFD1 | Q92890 | 22 | 19449911 | 19479202 | 0.83 | 0.71 | 0.98 | 2.63E-02 | 0.290 |
| OID20110 | USP8 | Cardiometabolic | USP8 | P40818 | 15 | 50424380 | 50514421 | 0.83 | 0.70 | 0.98 | 3.02E-02 | 0.290 |
| OID30640 | VTI1A | Inflammation_II | VTI1A | Q96AJ9 | 10 | 112446998 | 112818744 | 0.82 | 0.68 | 0.98 | 2.66E-02 | 0.290 |
| OID20926 | WWP2 | Neurology | WWP2 | O00308 | 16 | 69762328 | 69941741 | 0.83 | 0.70 | 0.99 | 3.41E-02 | 0.290 |
| OID21477 | TJAP1 | Oncology | TJAP1 | Q5JTD0 | 6 | 43477523 | 43506556 | 0.83 | 0.71 | 0.99 | 3.62E-02 | 0.292 |
| OID20182 | SSC4D | Cardiometabolic | SSC4D | Q8WTU2 | 7 | 76389334 | 76409697 | 0.82 | 0.68 | 0.99 | 3.63E-02 | 0.293 |
| OID30673 | APOB | Inflammation_II | APOB | P04114 | 2 | 21001429 | 21044073 | 0.85 | 0.72 | 0.99 | 3.69E-02 | 0.297 |
| OID20203 | RNASE3 | Cardiometabolic | RNASE3 | P12724 | 14 | 20891385 | 20892348 | 0.82 | 0.68 | 0.99 | 3.71E-02 | 0.297 |
| OID31494 | YARS1 | Oncology_II | YARS1 | P54577 | 1 | 32775237 | 32818031 | 0.85 | 0.72 | 0.99 | 3.70E-02 | 0.297 |
| OID20583 | ANXA11 | Inflammation | ANXA11 | P50995 | 10 | 80150889 | 80205572 | 0.86 | 0.74 | 0.99 | 3.75E-02 | 0.297 |
| OID31421 | BCL7B | Oncology_II | BCL7B | Q9BQE9 | 7 | 73536356 | 73557690 | 0.81 | 0.66 | 0.99 | 3.76E-02 | 0.297 |
| OID30373 | BDNF | Cardiometabolic_II | BDNF | P23560 | 11 | 27654893 | 27722058 | 0.85 | 0.73 | 0.99 | 3.78E-02 | 0.297 |
| OID21470 | CDC37 | Oncology | CDC37 | Q16543 | 19 | 10391133 | 10420121 | 0.84 | 0.71 | 0.99 | 3.79E-02 | 0.297 |
| OID20482 | MILR1 | Inflammation | MILR1 | Q7Z6M3 | 17 | 64449037 | 64468643 | 1.18 | 1.01 | 1.38 | 3.75E-02 | 0.297 |
| OID21083 | MMP3 | Neurology | MMP3 | P08254 | 11 | 102835801 | 102843609 | 1.22 | 1.01 | 1.47 | 3.80E-02 | 0.297 |
| OID31114 | NAPRT | Neurology_II | NAPRT | Q6XQN6 | 8 | 143574785 | 143578649 | 0.82 | 0.67 | 0.99 | 3.80E-02 | 0.297 |
| OID20077 | STK11 | Cardiometabolic | STK11 | Q15831 | 19 | 1177558 | 1228431 | 0.84 | 0.72 | 0.99 | 3.79E-02 | 0.297 |
| OID20644 | PRDX5 | Inflammation | PRDX5 | P30044 | 11 | 64318121 | 64321811 | 0.85 | 0.73 | 0.99 | 3.82E-02 | 0.298 |
| OID30414 | TALDO1 | Cardiometabolic_II | TALDO1 | P37837 | 11 | 747415 | 765012 | 0.83 | 0.69 | 0.99 | 3.83E-02 | 0.298 |
| OID30474 | BAG4 | Inflammation_II | BAG4 | O95429 | 8 | 38176533 | 38213301 | 0.82 | 0.68 | 0.99 | 4.10E-02 | 0.298 |
| OID31135 | CASP7 | Neurology_II | CASP7 | P55210 | 10 | 113679162 | 113730907 | 0.84 | 0.71 | 0.99 | 3.87E-02 | 0.298 |
| OID20857 | CERT | Neurology | CERT1 | Q9Y5P4 | 5 | 75356345 | 75512138 | 0.84 | 0.72 | 0.99 | 4.06E-02 | 0.298 |
| OID30183 | CSDE1 | Cardiometabolic_II | CSDE1 | O75534 | 1 | 114716913 | 114758676 | 0.82 | 0.68 | 0.99 | 3.95E-02 | 0.298 |
| OID30231 | ENTR1 | Cardiometabolic_II | ENTR1 | Q96C92 | 9 | 136401922 | 136410614 | 0.82 | 0.68 | 0.99 | 3.99E-02 | 0.298 |
| OID30395 | FGFBP2 | Cardiometabolic_II | FGFBP2 | Q9BYJ0 | 4 | 15960245 | 15969309 | 0.84 | 0.71 | 0.99 | 4.07E-02 | 0.298 |
| OID31499 | FKBPL | Oncology_II | FKBPL | Q9UIM3 | 6 | 32128707 | 32130288 | 0.84 | 0.70 | 0.99 | 4.04E-02 | 0.298 |
| OID30600 | GAPDH | Inflammation_II | GAPDH | P04406 | 12 | 6534512 | 6538374 | 0.86 | 0.75 | 0.99 | 4.05E-02 | 0.298 |
| OID30425 | HIF1A | Inflammation_II | HIF1A | Q16665 | 14 | 61695513 | 61748259 | 0.79 | 0.64 | 0.99 | 4.03E-02 | 0.298 |
| OID20484 | ICA1 | Inflammation | ICA1 | Q05084 | 7 | 8113184 | 8262687 | 0.84 | 0.71 | 0.99 | 4.06E-02 | 0.298 |
| OID20506 | IFNLR1 | Inflammation | IFNLR1 | Q8IU57 | 1 | 24154168 | 24187959 | 0.83 | 0.69 | 0.99 | 4.00E-02 | 0.298 |
| OID21292 | LAT2 | Oncology | LAT2 | Q9GZY6 | 7 | 74199652 | 74229834 | 0.84 | 0.71 | 0.99 | 3.95E-02 | 0.298 |
| OID30523 | LATS1 | Inflammation_II | LATS1 | O95835 | 6 | 149658153 | 149718105 | 0.84 | 0.71 | 0.99 | 4.04E-02 | 0.298 |
| OID21350 | LYN | Oncology | LYN | P07948 | 8 | 55879835 | 56014169 | 0.84 | 0.72 | 0.99 | 4.09E-02 | 0.298 |
| OID20510 | NUB1 | Inflammation | NUB1 | Q9Y5A7 | 7 | 151341772 | 151378449 | 0.83 | 0.70 | 0.99 | 3.97E-02 | 0.298 |
| OID31487 | OTUD6B | Oncology_II | OTUD6B | Q8N6M0 | 8 | 91070196 | 91087095 | 0.84 | 0.71 | 0.99 | 3.99E-02 | 0.298 |
| OID30584 | PDE5A | Inflammation_II | PDE5A | O76074 | 4 | 119494397 | 119628804 | 0.85 | 0.72 | 0.99 | 4.02E-02 | 0.298 |
| OID30507 | RBPMS | Inflammation_II | RBPMS | Q93062 | 8 | 30384511 | 30572256 | 0.78 | 0.62 | 0.99 | 4.04E-02 | 0.298 |
| OID21179 | RNF41 | Oncology | RNF41 | Q9H4P4 | 12 | 56202179 | 56221933 | 0.83 | 0.70 | 0.99 | 3.93E-02 | 0.298 |
| OID30648 | SFRP4 | Inflammation_II | SFRP4 | Q6FHJ7 | 7 | 37905932 | 38025695 | 0.84 | 0.72 | 0.99 | 4.05E-02 | 0.298 |
| OID31471 | SMNDC1 | Oncology_II | SMNDC1 | O75940 | 10 | 110290730 | 110304938 | 0.79 | 0.63 | 0.99 | 3.99E-02 | 0.298 |
| OID31086 | SPTLC1 | Neurology_II | SPTLC1 | O15269 | 9 | 92000087 | 92115413 | 0.80 | 0.65 | 0.99 | 4.05E-02 | 0.298 |
| OID31343 | TARS1 | Oncology_II | TARS1 | P26639 | 5 | 33440696 | 33468091 | 0.83 | 0.70 | 0.99 | 3.94E-02 | 0.298 |
| OID20371 | TFF3 | Cardiometabolic | TFF3 | Q07654 | 21 | 42311667 | 42315409 | 1.16 | 1.01 | 1.34 | 3.89E-02 | 0.298 |
| OID20098 | THPO | Cardiometabolic | THPO | P40225 | 3 | 184371935 | 184381968 | 0.84 | 0.72 | 0.99 | 3.85E-02 | 0.298 |
| OID31207 | TRIM26 | Oncology_II | TRIM26 | Q12899 | 6 | 30184455 | 30213427 | 0.81 | 0.67 | 0.99 | 4.10E-02 | 0.298 |
| OID21338 | ERP44 | Oncology | ERP44 | Q9BS26 | 9 | 99979185 | 100099052 | 0.84 | 0.71 | 0.99 | 4.13E-02 | 0.299 |
| OID31422 | TADA3 | Oncology_II | TADA3 | O75528 | 3 | 9779967 | 9793011 | 0.84 | 0.71 | 0.99 | 4.13E-02 | 0.299 |
| OID31495 | CEACAM16 | Oncology_II | CEACAM16 | Q2WEN9 | 19 | 44699151 | 44710718 | 0.84 | 0.71 | 0.99 | 4.16E-02 | 0.299 |
| OID31463 | FAM13A | Oncology_II | FAM13A | O94988 | 4 | 88725955 | 89111398 | 0.84 | 0.70 | 0.99 | 4.16E-02 | 0.299 |
| OID20250 | IL1RL1 | Cardiometabolic | IL1RL1 | Q01638 | 2 | 102311502 | 102352037 | 1.21 | 1.01 | 1.45 | 4.17E-02 | 0.299 |
| OID21437 | METAP2 | Oncology | METAP2 | P50579 | 12 | 95473520 | 95515839 | 0.85 | 0.72 | 0.99 | 4.19E-02 | 0.299 |
| OID31291 | RNF43 | Oncology_II | RNF43 | Q68DV7 | 17 | 58353676 | 58417595 | 0.79 | 0.64 | 0.99 | 4.18E-02 | 0.299 |
| OID31017 | SLA2 | Neurology_II | SLA2 | Q9H6Q3 | 20 | 36612318 | 36646196 | 0.85 | 0.72 | 0.99 | 4.19E-02 | 0.299 |
| OID21027 | PPCDC | Neurology | PPCDC | Q96CD2 | 15 | 75023586 | 75117462 | 0.84 | 0.71 | 0.99 | 4.22E-02 | 0.301 |
| OID21288 | MPI | Oncology | MPI | P34949 | 15 | 74890005 | 74902219 | 0.84 | 0.71 | 0.99 | 4.24E-02 | 0.302 |
| OID31055 | ASPSCR1 | Neurology_II | ASPSCR1 | Q9BZE9 | 17 | 81976807 | 82017406 | 0.84 | 0.71 | 0.99 | 4.29E-02 | 0.302 |
| OID30221 | CCER2 | Cardiometabolic_II | CCER2 | I3L3R5 | 19 | 38908980 | 38912186 | 1.16 | 1.00 | 1.33 | 4.35E-02 | 0.302 |
| OID20649 | CD6 | Inflammation | CD6 | P30203 | 11 | 60971680 | 61020377 | 0.85 | 0.73 | 1.00 | 4.35E-02 | 0.302 |
| OID31111 | DOK1 | Neurology_II | DOK1 | Q99704 | 2 | 74549026 | 74557551 | 0.85 | 0.73 | 1.00 | 4.40E-02 | 0.302 |
| OID20059 | DUOX2 | Cardiometabolic | DUOX2 | Q9NRD8 | 15 | 45092650 | 45114172 | 0.82 | 0.67 | 0.99 | 4.39E-02 | 0.302 |
| OID31022 | EIF4G3 | Neurology_II | EIF4G3 | O43432 | 1 | 20806292 | 21177285 | 0.84 | 0.71 | 0.99 | 4.34E-02 | 0.302 |
| OID31381 | FNTA | Oncology_II | FNTA | P49354 | 8 | 43034194 | 43085788 | 0.82 | 0.68 | 0.99 | 4.29E-02 | 0.302 |
| OID21377 | GALNT7 | Oncology | GALNT7 | Q86SF2 | 4 | 173168811 | 173323967 | 0.85 | 0.73 | 1.00 | 4.39E-02 | 0.302 |
| OID30264 | GRP | Cardiometabolic_II | GRP | P07492 | 18 | 59220158 | 59230774 | 0.85 | 0.73 | 1.00 | 4.34E-02 | 0.302 |
| OID21314 | INPPL1 | Oncology | INPPL1 | O15357 | 11 | 72223701 | 72239147 | 0.84 | 0.72 | 0.99 | 4.31E-02 | 0.302 |
| OID30350 | PPM1F | Cardiometabolic_II | PPM1F | P49593 | 22 | 21919425 | 21952848 | 0.84 | 0.71 | 1.00 | 4.39E-02 | 0.302 |
| OID30630 | SLC9A3R1 | Inflammation_II | SLC9A3R1 | O14745 | 17 | 74748628 | 74769353 | 0.84 | 0.71 | 0.99 | 4.29E-02 | 0.302 |
| OID20474 | TANK | Inflammation | TANK | Q92844 | 2 | 161136908 | 161236230 | 0.84 | 0.71 | 1.00 | 4.36E-02 | 0.302 |
| OID20942 | TBCC | Neurology | TBCC | Q15814 | 6 | 42744498 | 42746103 | 0.85 | 0.72 | 1.00 | 4.36E-02 | 0.302 |
| OID20509 | SPINK4 | Inflammation | SPINK4 | O60575 | 9 | 33218365 | 33248567 | 1.17 | 1.00 | 1.37 | 4.41E-02 | 0.302 |

| | | | | | | | | | | | | |
|---|---|---|---|---|---|---|---|---|---|---|---|---|
| OID30556 | CD226 | Inflammation_II | CD226 | Q15762 | 18 | 69831158 | 69961803 | 0.85 | 0.72 | 1.00 | 4.44E-02 | 0.303 |
| OID30459 | FGF20 | Inflammation_II | FGF20 | Q9NP95 | 8 | 16992181 | 17002345 | 0.81 | 0.65 | 1.00 | 4.49E-02 | 0.305 |
| OID20218 | SNAP23 | Cardiometabolic | SNAP23 | O00161 | 15 | 42491233 | 42545356 | 0.85 | 0.73 | 1.00 | 4.49E-02 | 0.305 |
| OID21016 | VTA1 | Neurology | VTA1 | Q9NP79 | 6 | 142147162 | 142224685 | 0.85 | 0.72 | 1.00 | 4.50E-02 | 0.305 |
| OID31140 | ZHX2 | Neurology_II | ZHX2 | Q9Y6X8 | 8 | 122781655 | 122974510 | 0.77 | 0.59 | 0.99 | 4.50E-02 | 0.305 |
| OID21171 | CEACAM5 | Oncology | CEACAM5 | P06731 | 19 | 41708585 | 41730433 | 1.16 | 1.00 | 1.33 | 4.57E-02 | 0.305 |
| OID30557 | EIF4E | Inflammation_II | EIF4E | P06730 | 4 | 98879276 | 98929133 | 0.85 | 0.72 | 1.00 | 4.57E-02 | 0.305 |
| OID21140 | FYB1 | Neurology | FYB1 | O15117 | 5 | 39105252 | 39274528 | 0.85 | 0.73 | 1.00 | 4.52E-02 | 0.305 |
| OID30610 | PCBD1 | Inflammation_II | PCBD1 | P61457 | 10 | 70882280 | 70888565 | 0.82 | 0.68 | 1.00 | 4.58E-02 | 0.305 |
| OID30323 | PDAP1 | Cardiometabolic_II | PDAP1 | Q13442 | 7 | 99392048 | 99408597 | 0.85 | 0.72 | 1.00 | 4.58E-02 | 0.305 |
| OID31155 | PLCB2 | Neurology_II | PLCB2 | Q00722 | 15 | 40278176 | 40307935 | 0.85 | 0.72 | 1.00 | 4.54E-02 | 0.305 |
| OID30386 | TWF2 | Cardiometabolic_II | TWF2 | Q6IBS0 | 3 | 52228612 | 52246788 | 0.84 | 0.71 | 1.00 | 4.53E-02 | 0.305 |
| OID30297 | PTGR1 | Cardiometabolic_II | PTGR1 | Q14914 | 9 | 111549722 | 111599893 | 0.84 | 0.71 | 1.00 | 4.59E-02 | 0.305 |
| OID21161 | SCP2 | Oncology | SCP2 | P22307 | 1 | 52927276 | 53051698 | 0.82 | 0.67 | 1.00 | 4.61E-02 | 0.306 |
| OID30645 | SNCA | Inflammation_II | SNCA | P37840 | 4 | 89700345 | 89838315 | 0.85 | 0.72 | 1.00 | 4.61E-02 | 0.306 |
| OID21102 | PIGR | Neurology | PIGR | P01833 | 1 | 206928522 | 206946466 | 1.20 | 1.00 | 1.43 | 4.63E-02 | 0.306 |
| OID30130 | ABRAXAS2 | Cardiometabolic_II | ABRAXAS2 | Q15018 | 10 | 124801819 | 124836667 | 0.84 | 0.71 | 1.00 | 4.69E-02 | 0.307 |
| OID20554 | ARHGEF12 | Inflammation | ARHGEF12 | Q9NZN5 | 11 | 120336413 | 120489937 | 0.85 | 0.72 | 1.00 | 4.71E-02 | 0.307 |
| OID20555 | CASP2 | Inflammation | CASP2 | P42575 | 7 | 143288215 | 143307696 | 0.84 | 0.71 | 1.00 | 4.71E-02 | 0.307 |
| OID20086 | COMT | Cardiometabolic | COMT | P21964 | 22 | 19941371 | 19969975 | 0.85 | 0.72 | 1.00 | 4.71E-02 | 0.307 |
| OID30464 | KLRF1 | Inflammation_II | KLRF1 | Q9NZS2 | 12 | 9827481 | 9845007 | 1.15 | 1.00 | 1.32 | 4.66E-02 | 0.307 |
| OID30310 | SRPX | Cardiometabolic_II | SRPX | P78539 | X | 38149336 | 38220924 | 1.20 | 1.00 | 1.44 | 4.71E-02 | 0.307 |
| OID30573 | PDIA3 | Inflammation_II | PDIA3 | P30101 | 15 | 43746394 | 43773279 | 0.84 | 0.70 | 1.00 | 4.73E-02 | 0.308 |
| OID31001 | CCDC50 | Neurology_II | CCDC50 | Q8IVM0 | 3 | 191329085 | 191398659 | 0.80 | 0.65 | 1.00 | 4.75E-02 | 0.308 |
| OID30243 | HS1BP3 | Cardiometabolic_II | HS1BP3 | Q53T59 | 2 | 20560448 | 20651130 | 0.84 | 0.71 | 1.00 | 4.81E-02 | 0.308 |
| OID20708 | LY6D | Inflammation | LY6D | Q14210 | 8 | 142784882 | 142786539 | 1.16 | 1.00 | 1.34 | 4.82E-02 | 0.308 |
| OID31061 | PIBF1 | Neurology_II | PIBF1 | Q8WXW3 | 13 | 72782133 | 73016461 | 0.84 | 0.71 | 1.00 | 4.77E-02 | 0.308 |
| OID20775 | PNLIPRP2 | Inflammation | PNLIPRP2 | P54317 | 10 | 116620953 | 116645143 | 1.20 | 1.00 | 1.44 | 4.82E-02 | 0.308 |
| OID31028 | REEP4 | Neurology_II | REEP4 | Q9H6H4 | 8 | 22138020 | 22141951 | 0.83 | 0.70 | 1.00 | 4.79E-02 | 0.308 |
| OID20103 | S100P | Cardiometabolic | S100P | P25815 | 4 | 6693878 | 6697170 | 1.16 | 1.00 | 1.35 | 4.83E-02 | 0.308 |
| OID21180 | SMAD1 | Oncology | SMAD1 | Q15797 | 4 | 145481194 | 145559176 | 0.85 | 0.72 | 1.00 | 4.77E-02 | 0.308 |
| OID30636 | SYAP1 | Inflammation_II | SYAP1 | Q96A49 | X | 16719612 | 16765340 | 0.84 | 0.71 | 1.00 | 4.80E-02 | 0.308 |
| OID21323 | ERBIN | Oncology | ERBIN | Q96RT1 | 5 | 65926556 | 66082546 | 0.85 | 0.72 | 1.00 | 4.99E-02 | 0.312 |
| OID21086 | HARS1 | Neurology | HARS1 | P12081 | 5 | 140673035 | 140691537 | 0.85 | 0.72 | 1.00 | 4.98E-02 | 0.312 |
| OID20648 | HCLS1 | Inflammation | HCLS1 | P14317 | 3 | 121631399 | 121660927 | 0.85 | 0.73 | 1.00 | 4.94E-02 | 0.312 |
| OID20855 | ILKAP | Neurology | ILKAP | Q9H0C8 | 2 | 238170402 | 238203708 | 0.84 | 0.70 | 1.00 | 4.95E-02 | 0.312 |
| OID30677 | LCAT | Inflammation_II | LCAT | P04180 | 16 | 67939750 | 67944131 | 0.84 | 0.70 | 1.00 | 4.95E-02 | 0.312 |
| OID31035 | NARS1 | Neurology_II | NARS1 | O43776 | 18 | 57600656 | 57622213 | 0.86 | 0.73 | 1.00 | 4.96E-02 | 0.312 |
| OID20534 | PSIP1 | Inflammation | PSIP1 | O75475 | 9 | 15464066 | 15510995 | 0.82 | 0.68 | 1.00 | 4.95E-02 | 0.312 |
| OID30218 | RNF5 | Cardiometabolic_II | RNF5 | Q99942 | 6 | 32178405 | 32180793 | 0.85 | 0.73 | 1.00 | 4.98E-02 | 0.312 |
| OID31303 | ZNF75D | Oncology_II | ZNF75D | P51815 | X | 135248920 | 135344109 | 0.82 | 0.67 | 1.00 | 4.96E-02 | 0.312 |
| OID30121 | ARL2BP | Cardiometabolic_II | ARL2BP | Q9Y2Y0 | 16 | 57245259 | 57253635 | 0.84 | 0.71 | 1.00 | 5.03E-02 | 0.313 |
| OID20701 | CRIM1 | Inflammation | CRIM1 | Q9NZV1 | 2 | 36355778 | 36551135 | 1.17 | 1.00 | 1.38 | 5.02E-02 | 0.313 |
| OID31128 | GBP1 | Neurology_II | GBP1 | P32455 | 1 | 89051882 | 89065360 | 0.85 | 0.72 | 1.00 | 5.06E-02 | 0.314 |
| OID31147 | GCC1 | Neurology_II | GCC1 | Q96CN9 | 7 | 127580628 | 127593611 | 0.85 | 0.73 | 1.00 | 5.08E-02 | 0.315 |
| OID21036 | ANXA3 | Neurology | ANXA3 | P12429 | 4 | 78551747 | 78610451 | 0.85 | 0.72 | 1.00 | 5.11E-02 | 0.316 |
| OID20208 | COL4A1 | Cardiometabolic | COL4A1 | P02462 | 13 | 110148963 | 110307157 | 1.18 | 1.00 | 1.38 | 5.13E-02 | 0.316 |
| OID30745 | SELENOP | Inflammation_II | SELENOP | P49908 | 5 | 42799880 | 42887392 | 0.85 | 0.72 | 1.00 | 5.13E-02 | 0.316 |
| OID21298 | CD207 | Oncology | CD207 | Q9UJ71 | 2 | 70830211 | 70835816 | 1.18 | 1.00 | 1.39 | 5.20E-02 | 0.316 |
| OID31406 | CMIP | Oncology_II | CMIP | Q8IY22 | 16 | 81444808 | 81711762 | 0.85 | 0.72 | 1.00 | 5.18E-02 | 0.316 |
| OID30308 | DTYMK | Cardiometabolic_II | DTYMK | P23919 | 2 | 241675747 | 241686944 | 0.85 | 0.72 | 1.00 | 5.16E-02 | 0.316 |
| OID30232 | HSBP1 | Cardiometabolic_II | HSBP1 | O75506 | 16 | 83719311 | 83819737 | 0.85 | 0.72 | 1.00 | 5.18E-02 | 0.316 |
| OID20821 | LYPD1 | Neurology | LYPD1 | Q8N2G4 | 2 | 132643286 | 132671579 | 0.81 | 0.65 | 1.00 | 5.15E-02 | 0.316 |
| OID21324 | SRP14 | Oncology | SRP14 | P37108 | 15 | 40035690 | 40039181 | 0.83 | 0.70 | 1.00 | 5.19E-02 | 0.316 |
| OID30994 | ARHGEF10 | Neurology_II | ARHGEF10 | O15013 | 8 | 1823926 | 1958641 | 0.85 | 0.72 | 1.00 | 5.27E-02 | 0.320 |
| OID20958 | CC2D1A | Neurology | CC2D1A | Q6P1N0 | 19 | 13906201 | 13930879 | 0.84 | 0.71 | 1.00 | 5.28E-02 | 0.320 |
| OID20596 | KRT19 | Inflammation | KRT19 | P08727 | 17 | 41523617 | 41528308 | 1.15 | 1.00 | 1.33 | 5.32E-02 | 0.322 |
| OID20850 | PMVK | Neurology | PMVK | Q15126 | 1 | 154924740 | 154936719 | 0.86 | 0.74 | 1.00 | 5.33E-02 | 0.322 |
| OID31336 | ARHGAP30 | Oncology_II | ARHGAP30 | Q7Z6I6 | 1 | 161046946 | 161069970 | 0.79 | 0.62 | 1.00 | 5.36E-02 | 0.322 |
| OID30590 | APPL2 | Inflammation_II | APPL2 | Q8NEU8 | 12 | 105173297 | 105236203 | 0.85 | 0.73 | 1.00 | 5.38E-02 | 0.323 |
| OID21037 | ENO1 | Neurology | ENO1 | P06733 | 1 | 8861000 | 8879190 | 0.85 | 0.72 | 1.00 | 5.39E-02 | 0.323 |
| OID20899 | MAP4K5 | Neurology | MAP4K5 | Q9Y4K4 | 14 | 50418501 | 50561126 | 0.85 | 0.73 | 1.00 | 5.39E-02 | 0.323 |
| OID20698 | EGF | Inflammation | EGF | P01133 | 4 | 109912883 | 110013766 | 0.86 | 0.73 | 1.00 | 5.42E-02 | 0.323 |
| OID30680 | GPI | Inflammation_II | GPI | P06744 | 19 | 34359480 | 34402413 | 0.84 | 0.71 | 1.00 | 5.41E-02 | 0.323 |
| OID31132 | SNRPB2 | Neurology_II | SNRPB2 | P08579 | 20 | 16729961 | 16742564 | 0.76 | 0.58 | 1.00 | 5.43E-02 | 0.323 |
| OID20138 | DOK2 | Cardiometabolic | DOK2 | O60496 | 8 | 21908873 | 21913690 | 0.85 | 0.72 | 1.00 | 5.48E-02 | 0.325 |
| OID30899 | KCNH2 | Neurology_II | KCNH2 | Q12809 | 7 | 150944961 | 150978321 | 0.81 | 0.66 | 1.00 | 5.52E-02 | 0.327 |
| OID31522 | ACOT13 | Oncology_II | ACOT13 | Q9NPJ3 | 6 | 24667035 | 24705065 | 0.85 | 0.72 | 1.00 | 5.58E-02 | 0.327 |
| OID21078 | AHSP | Neurology | AHSP | Q9NZD4 | 16 | 31527900 | 31528803 | 0.85 | 0.71 | 1.01 | 5.96E-02 | 0.327 |
| OID20123 | ANXA4 | Cardiometabolic | ANXA4 | P09525 | 2 | 69644425 | 69827112 | 0.85 | 0.72 | 1.00 | 5.66E-02 | 0.327 |
| OID21359 | APBB1IP | Oncology | APBB1IP | Q7Z5R6 | 10 | 26438341 | 26567803 | 0.82 | 0.67 | 1.01 | 5.71E-02 | 0.327 |
| OID21357 | AREG | Oncology | AREG | P15514 | 4 | 74445136 | 74455005 | 1.16 | 0.99 | 1.34 | 5.88E-02 | 0.327 |
| OID30837 | BOLA1 | Neurology_II | BOLA1 | Q9Y3E2 | 1 | 149887890 | 149900798 | 0.83 | 0.69 | 1.01 | 6.00E-02 | 0.327 |
| OID30311 | BPIFB2 | Cardiometabolic_II | BPIFB2 | Q8N4F0 | 20 | 33007704 | 33023703 | 0.84 | 0.70 | 1.01 | 5.80E-02 | 0.327 |
| OID20378 | CD14 | Cardiometabolic | CD14 | P08571 | 5 | 140631728 | 140633701 | 1.15 | 1.00 | 1.33 | 5.76E-02 | 0.327 |
| OID30969 | COL28A1 | Neurology_II | COL28A1 | Q2UY09 | 7 | 7356203 | 7535873 | 1.16 | 1.00 | 1.35 | 5.78E-02 | 0.327 |
| OID20293 | CST6 | Cardiometabolic | CST6 | Q15828 | 11 | 66012008 | 66013505 | 1.17 | 0.99 | 1.37 | 5.93E-02 | 0.327 |
| OID21399 | DCTN1 | Oncology | DCTN1 | Q14203 | 2 | 74361154 | 74392087 | 0.85 | 0.72 | 1.01 | 5.96E-02 | 0.327 |
| OID20184 | DIABLO | Cardiometabolic | DIABLO | Q9NR28 | 12 | 122207663 | 122227456 | 0.86 | 0.73 | 1.01 | 6.00E-02 | 0.327 |
| OID20295 | DLK1 | Cardiometabolic | DLK1 | P80370 | 14 | 100725705 | 100738224 | 0.85 | 0.72 | 1.01 | 5.98E-02 | 0.327 |
| OID20093 | DNAJB8 | Cardiometabolic | DNAJB8 | Q8NHS0 | 3 | 128462437 | 128466968 | 0.83 | 0.68 | 1.01 | 5.85E-02 | 0.327 |
| OID30925 | DYNLT1 | Neurology_II | DYNLT1 | P63172 | 6 | 158636474 | 158644743 | 0.84 | 0.70 | 1.00 | 5.61E-02 | 0.327 |
| OID30787 | FN1 | Inflammation_II | FN1 | P02751 | 2 | 215360440 | 215436073 | 0.85 | 0.71 | 1.00 | 5.58E-02 | 0.327 |
| OID20515 | FOXO1 | Inflammation | FOXO1 | Q12778 | 13 | 40555667 | 40666641 | 0.85 | 0.72 | 1.00 | 5.61E-02 | 0.327 |
| OID30237 | GADD45GIP1 | Cardiometabolic_II | GADD45GIP1 | Q8TAE8 | 19 | 12953119 | 12957223 | 0.84 | 0.71 | 1.00 | 5.59E-02 | 0.327 |
| OID20710 | GLOD4 | Inflammation | GLOD4 | Q9HC38 | 17 | 757097 | 782294 | 0.85 | 0.72 | 1.01 | 6.00E-02 | 0.327 |
| OID30409 | HPSE | Cardiometabolic_II | HPSE | Q9Y251 | 4 | 83292461 | 83335153 | 0.86 | 0.73 | 1.01 | 5.84E-02 | 0.327 |
| OID20829 | HSP90B1 | Neurology | HSP90B1 | P14625 | 12 | 103930107 | 103953931 | 0.81 | 0.65 | 1.01 | 5.86E-02 | 0.327 |
| OID20795 | IFNL1 | Neurology | IFNL1 | Q8IU54 | 19 | 39296407 | 39298673 | 1.14 | 1.00 | 1.31 | 5.55E-02 | 0.327 |
| OID20487 | IL1RL2 | Inflammation | IL1RL2 | Q9HB29 | 2 | 102187006 | 102240002 | 0.85 | 0.72 | 1.01 | 5.91E-02 | 0.327 |
| OID30860 | ITPRIP | Neurology_II | ITPRIP | Q8IWB1 | 10 | 104309698 | 104338465 | 0.83 | 0.68 | 1.00 | 5.59E-02 | 0.327 |
| OID20187 | LEP | Cardiometabolic | LEP | P41159 | 7 | 128241278 | 128257629 | 0.77 | 0.59 | 1.01 | 5.73E-02 | 0.327 |
| OID31269 | LSM8 | Oncology_II | LSM8 | O95777 | 7 | 118184144 | 118204035 | 0.82 | 0.67 | 1.01 | 6.00E-02 | 0.327 |
| OID30669 | MDH1 | Inflammation_II | MDH1 | P40925 | 2 | 63588609 | 63607197 | 0.84 | 0.70 | 1.01 | 5.72E-02 | 0.327 |
| OID21073 | MIF | Neurology | MIF | P14174 | 22 | 23894383 | 23895227 | 0.86 | 0.73 | 1.00 | 5.71E-02 | 0.327 |
| OID30893 | NAA80 | Neurology_II | NAA80 | Q93015 | 3 | 50296402 | 50299416 | 0.85 | 0.72 | 1.01 | 5.90E-02 | 0.327 |
| OID31250 | NEK7 | Oncology_II | NEK7 | Q8TDX7 | 1 | 198156994 | 198322420 | 0.83 | 0.68 | 1.01 | 5.96E-02 | 0.327 |
| OID20553 | NT5C3A | Inflammation | NT5C3A | Q9H0P0 | 7 | 33014130 | 33062796 | 0.86 | 0.73 | 1.01 | 5.97E-02 | 0.327 |
| OID21115 | NUDT5 | Neurology | NUDT5 | Q9UKK9 | 10 | 12165330 | 12196144 | 0.85 | 0.71 | 1.01 | 5.92E-02 | 0.327 |
| OID31063 | PDLIM5 | Neurology_II | PDLIM5 | Q96HC4 | 4 | 94451857 | 94668227 | 0.86 | 0.73 | 1.00 | 5.66E-02 | 0.327 |
| OID30647 | RABEP1 | Inflammation_II | RABEP1 | Q15276 | 17 | 5282265 | 5386340 | 0.85 | 0.72 | 1.01 | 5.85E-02 | 0.327 |
| OID20258 | REG3A | Cardiometabolic | REG3A | Q06141 | 2 | 79157003 | 79159753 | 1.17 | 1.00 | 1.37 | 5.63E-02 | 0.327 |
| OID20536 | SAMD9L | Inflammation | SAMD9L | Q8IVG5 | 7 | 93130056 | 93148385 | 0.85 | 0.71 | 1.00 | 5.69E-02 | 0.327 |
| OID30662 | SDK2 | Inflammation_II | SDK2 | Q58EX2 | 17 | 73334384 | 73644445 | 0.84 | 0.70 | 1.01 | 5.68E-02 | 0.327 |
| OID20379 | SELP | Cardiometabolic | SELP | P16109 | 1 | 169588849 | 169630193 | 0.86 | 0.73 | 1.01 | 5.94E-02 | 0.327 |
| OID31054 | STXBP1 | Neurology_II | STXBP1 | P61764 | 9 | 127579370 | 127696027 | 0.86 | 0.73 | 1.01 | 5.83E-02 | 0.327 |
| OID20993 | TBCB | Neurology | TBCB | Q99426 | 19 | 36114289 | 36125947 | 0.86 | 0.73 | 1.01 | 5.88E-02 | 0.327 |

| | | | | | | | | | | | | |
|---|---|---|---|---|---|---|---|---|---|---|---|---|
| OID31016 | TSC22D1 | Neurology_II | TSC22D1 | Q15714 | 13 | 44432143 | 44577147 | 0.84 | 0.70 | 1.01 | 5.86E-02 | 0.327 |
| OID20869 | XRCC4 | Neurology | XRCC4 | Q13426 | 5 | 83077498 | 83353787 | 0.85 | 0.71 | 1.01 | 5.74E-02 | 0.327 |
| OID31185 | ZP4 | Oncology_II | ZP4 | Q12836 | 1 | 237877864 | 237890922 | 0.81 | 0.65 | 1.01 | 5.75E-02 | 0.327 |
| OID30335 | EEF1D | Cardiometabolic_II | EEF1D | P29692 | 8 | 143579697 | 143599541 | 0.85 | 0.72 | 1.01 | 6.02E-02 | 0.327 |
| OID20558 | BCR | Inflammation | BCR | P11274 | 22 | 23179704 | 23318037 | 0.86 | 0.73 | 1.01 | 6.06E-02 | 0.329 |
| OID20317 | PLXNB2 | Cardiometabolic | PLXNB2 | O15031 | 22 | 50274979 | 50307646 | 1.17 | 0.99 | 1.38 | 6.07E-02 | 0.329 |
| OID21091 | GP6 | Neurology | GP6 | Q9HCN6 | 19 | 55013705 | 55038264 | 0.86 | 0.73 | 1.01 | 6.09E-02 | 0.329 |
| OID20158 | IL19 | Cardiometabolic | IL19 | Q9UHD0 | 1 | 206770764 | 206842981 | 1.15 | 0.99 | 1.33 | 6.17E-02 | 0.332 |
| OID30757 | SERPINF1 | Inflammation_II | SERPINF1 | P36955 | 17 | 1762029 | 1777565 | 0.83 | 0.69 | 1.01 | 6.18E-02 | 0.332 |
| OID20714 | SHMT1 | Inflammation | SHMT1 | P34896 | 17 | 18327860 | 18363563 | 0.85 | 0.72 | 1.01 | 6.16E-02 | 0.332 |
| OID30554 | VEGFB | Inflammation_II | VEGFB | P49765 | 11 | 64234584 | 64239264 | 1.16 | 0.99 | 1.36 | 6.16E-02 | 0.332 |
| OID20485 | IRAK1 | Inflammation | IRAK1 | P51617 | X | 154010506 | 154019902 | 0.85 | 0.71 | 1.01 | 6.20E-02 | 0.332 |
| OID30127 | ELOB | Cardiometabolic_II | ELOB | Q15370 | 16 | 2771414 | 2777289 | 0.83 | 0.69 | 1.01 | 6.22E-02 | 0.332 |
| OID30773 | F11 | Inflammation_II | F11 | P03951 | 4 | 186266189 | 186289681 | 0.84 | 0.70 | 1.01 | 6.21E-02 | 0.332 |
| OID31428 | USP25 | Oncology_II | USP25 | Q9UHP3 | 21 | 15729982 | 15880064 | 0.85 | 0.72 | 1.01 | 6.27E-02 | 0.334 |
| OID31038 | SATB1 | Neurology_II | SATB1 | Q01826 | 3 | 18345377 | 18445621 | 0.81 | 0.65 | 1.01 | 6.29E-02 | 0.334 |
| OID31087 | OPHN1 | Neurology_II | OPHN1 | O60890 | X | 67949349 | 68433913 | 0.86 | 0.73 | 1.01 | 6.34E-02 | 0.337 |
| OID31393 | NACC1 | Oncology_II | NACC1 | Q96RE7 | 19 | 13116862 | 13141147 | 0.85 | 0.72 | 1.01 | 6.36E-02 | 0.337 |
| OID30236 | AAMDC | Cardiometabolic_II | AAMDC | Q9H7C9 | 11 | 77821109 | 77918432 | 0.85 | 0.72 | 1.01 | 6.42E-02 | 0.339 |
| OID31360 | PPP1CC | Oncology_II | PPP1CC | P36873 | 12 | 110719680 | 110742939 | 0.85 | 0.72 | 1.01 | 6.42E-02 | 0.339 |
| OID31251 | CEACAM18 | Oncology_II | CEACAM18 | A8MTB9 | 19 | 51478539 | 51491301 | 1.13 | 0.99 | 1.28 | 6.49E-02 | 0.340 |
| OID30106 | CPTP | Cardiometabolic_II | CPTP | Q5TA50 | 1 | 1324756 | 1328896 | 0.84 | 0.70 | 1.01 | 6.45E-02 | 0.340 |
| OID20923 | CRADD | Neurology | CRADD | P78560 | 12 | 93677375 | 93894840 | 0.86 | 0.73 | 1.01 | 6.47E-02 | 0.340 |
| OID21514 | FURIN | Oncology | FURIN | P09958 | 15 | 90868588 | 90883564 | 0.84 | 0.70 | 1.01 | 6.49E-02 | 0.340 |
| OID30113 | SART1 | Cardiometabolic_II | SART1 | O43290 | 11 | 65961728 | 65980137 | 0.79 | 0.62 | 1.01 | 6.49E-02 | 0.340 |
| OID20054 | CHEK2 | Cardiometabolic | CHEK2 | O96017 | 22 | 28687743 | 28742422 | 0.86 | 0.72 | 1.01 | 6.51E-02 | 0.340 |
| OID30331 | SARG | Cardiometabolic_II | C1orf116 | Q9BW04 | 1 | 207018522 | 207032756 | 0.86 | 0.73 | 1.01 | 6.52E-02 | 0.340 |
| OID21061 | CLEC1B | Neurology | CLEC1B | Q9P126 | 12 | 9985642 | 10013424 | 0.86 | 0.73 | 1.01 | 6.53E-02 | 0.340 |
| OID30485 | CD36 | Inflammation_II | CD36 | P16671 | 7 | 80369575 | 80679277 | 0.85 | 0.71 | 1.01 | 6.56E-02 | 0.341 |
| OID21192 | FEN1 | Oncology | FEN1 | P39748 | 11 | 61792911 | 61797238 | 0.83 | 0.68 | 1.01 | 6.60E-02 | 0.341 |
| OID31261 | GADD45B | Oncology_II | GADD45B | O75293 | 19 | 2476122 | 2478259 | 0.80 | 0.63 | 1.01 | 6.58E-02 | 0.341 |
| OID20180 | TINAGL1 | Cardiometabolic | TINAGL1 | Q9GZM7 | 1 | 31576485 | 31587686 | 1.17 | 0.99 | 1.38 | 6.57E-02 | 0.341 |
| OID31462 | TMED8 | Oncology_II | TMED8 | Q6PL24 | 14 | 77335029 | 77377094 | 0.86 | 0.73 | 1.01 | 6.59E-02 | 0.341 |
| OID30717 | BCHE | Inflammation_II | BCHE | P06276 | 3 | 165772904 | 165837462 | 0.85 | 0.71 | 1.01 | 6.63E-02 | 0.342 |
| OID30617 | ANXA1 | Inflammation_II | ANXA1 | P04083 | 9 | 73151865 | 73170393 | 0.83 | 0.68 | 1.01 | 6.67E-02 | 0.342 |
| OID31125 | EIF1AX | Neurology_II | EIF1AX | P47813 | X | 20124525 | 20141838 | 0.84 | 0.69 | 1.01 | 6.66E-02 | 0.342 |
| OID20784 | REG4 | Inflammation | REG4 | Q9BYZ8 | 1 | 119794017 | 119811580 | 1.16 | 0.99 | 1.37 | 6.68E-02 | 0.342 |
| OID20867 | PRTFDC1 | Neurology | PRTFDC1 | Q9NRG1 | 10 | 24848614 | 24952606 | 0.86 | 0.74 | 1.01 | 6.69E-02 | 0.342 |
| OID20114 | BAG6 | Cardiometabolic | BAG6 | P46379 | 6 | 31639028 | 31652705 | 0.86 | 0.72 | 1.01 | 6.74E-02 | 0.343 |
| OID21077 | STIP1 | Neurology | STIP1 | P31948 | 11 | 64185272 | 64204543 | 0.85 | 0.72 | 1.01 | 6.73E-02 | 0.343 |
| OID31029 | ELAC1 | Neurology_II | ELAC1 | Q9H777 | 18 | 50967991 | 50988121 | 0.86 | 0.73 | 1.01 | 6.77E-02 | 0.344 |
| OID30327 | CHMP6 | Cardiometabolic_II | CHMP6 | Q96FZ7 | 17 | 80991598 | 81009517 | 0.86 | 0.73 | 1.01 | 6.81E-02 | 0.345 |
| OID30859 | DGCR6 | Neurology_II | DGCR6 | Q14129 | 22 | 18906028 | 18914238 | 0.83 | 0.68 | 1.01 | 6.81E-02 | 0.345 |
| OID30150 | PYY | Cardiometabolic_II | PYY | P10082 | 17 | 43952733 | 44004469 | 1.16 | 0.99 | 1.35 | 6.84E-02 | 0.346 |
| OID31472 | DNAJC9 | Oncology_II | DNAJC9 | Q8WXX5 | 10 | 73183362 | 73247255 | 0.86 | 0.72 | 1.01 | 6.86E-02 | 0.346 |
| OID31468 | MAP2K1 | Oncology_II | MAP2K1 | Q02750 | 15 | 66386837 | 66491656 | 0.85 | 0.72 | 1.01 | 6.85E-02 | 0.346 |
| OID20667 | CDSN | Inflammation | CDSN | Q15517 | 6 | 31115087 | 31120446 | 1.15 | 0.99 | 1.34 | 6.87E-02 | 0.346 |
| OID20825 | GLT8D2 | Neurology | GLT8D2 | Q9H1C3 | 12 | 103988987 | 104064183 | 0.82 | 0.67 | 1.02 | 6.90E-02 | 0.346 |
| OID20773 | LGMN | Inflammation | LGMN | Q99538 | 14 | 92703807 | 92748679 | 0.85 | 0.72 | 1.01 | 6.89E-02 | 0.346 |
| OID20719 | CSF1 | Inflammation | CSF1 | P09603 | 1 | 109910242 | 109930992 | 1.16 | 0.99 | 1.36 | 6.92E-02 | 0.346 |
| OID20107 | GLO1 | Cardiometabolic | GLO1 | Q04760 | 6 | 38675925 | 38703145 | 0.85 | 0.72 | 1.01 | 6.93E-02 | 0.346 |
| OID30209 | ENO3 | Cardiometabolic_II | ENO3 | P13929 | 17 | 4948092 | 4957131 | 0.84 | 0.70 | 1.01 | 6.95E-02 | 0.347 |
| OID21239 | FLI1 | Oncology | FLI1 | Q01543 | 11 | 128686535 | 128813267 | 0.86 | 0.73 | 1.01 | 6.97E-02 | 0.347 |
| OID30933 | MFAP3L | Neurology_II | MFAP3L | O75121 | 4 | 169986597 | 170033031 | 0.83 | 0.69 | 1.01 | 6.97E-02 | 0.347 |
| OID21363 | DNAJB1 | Oncology | DNAJB1 | P25685 | 19 | 14514769 | 14560391 | 0.86 | 0.73 | 1.01 | 7.00E-02 | 0.347 |
| OID30294 | NT5C | Cardiometabolic_II | NT5C | Q8TCD5 | 17 | 75130225 | 75131757 | 0.87 | 0.74 | 1.01 | 7.00E-02 | 0.347 |
| OID31379 | AHSA1 | Oncology_II | AHSA1 | O95433 | 14 | 77457870 | 77469472 | 0.84 | 0.69 | 1.01 | 7.03E-02 | 0.348 |
| OID30884 | CLEC2L | Neurology_II | CLEC2L | P0C7M8 | 7 | 139523685 | 139544985 | 0.83 | 0.67 | 1.02 | 7.04E-02 | 0.348 |
| OID20643 | PPP1R9B | Inflammation | PPP1R9B | Q96SB3 | 17 | 50133737 | 50150677 | 0.86 | 0.73 | 1.01 | 7.07E-02 | 0.349 |
| OID30642 | ACYP1 | Inflammation_II | ACYP1 | P07311 | 14 | 75053237 | 75069483 | 0.86 | 0.73 | 1.01 | 7.11E-02 | 0.349 |
| OID30631 | GCHFR | Inflammation_II | GCHFR | P30047 | 15 | 40764068 | 40767708 | 0.83 | 0.67 | 1.02 | 7.16E-02 | 0.349 |
| OID31367 | LACRT | Oncology_II | LACRT | Q9GZZ8 | 12 | 54630811 | 54634895 | 0.84 | 0.69 | 1.02 | 7.15E-02 | 0.349 |
| OID20247 | PLAT | Cardiometabolic | PLAT | P00750 | 8 | 42174718 | 42207709 | 0.84 | 0.70 | 1.02 | 7.15E-02 | 0.349 |
| OID20822 | PXN | Neurology | PXN | P49023 | 12 | 120210439 | 120265771 | 0.85 | 0.70 | 1.01 | 7.13E-02 | 0.349 |
| OID21495 | TXNDC15 | Oncology | TXNDC15 | Q96J42 | 5 | 134874371 | 134901635 | 1.16 | 0.99 | 1.36 | 7.15E-02 | 0.349 |
| OID21135 | TXNRD1 | Neurology | TXNRD1 | Q16881 | 12 | 104215779 | 104350307 | 0.85 | 0.71 | 1.01 | 7.12E-02 | 0.349 |
| OID30431 | DDX4 | Inflammation_II | DDX4 | Q9NQI0 | 5 | 55738017 | 55817157 | 0.84 | 0.69 | 1.02 | 7.26E-02 | 0.350 |
| OID31137 | FTCD | Neurology_II | FTCD | O95954 | 21 | 46136160 | 46155579 | 0.85 | 0.71 | 1.01 | 7.20E-02 | 0.350 |
| OID31224 | GRSF1 | Oncology_II | GRSF1 | Q12849 | 4 | 70815783 | 70839897 | 0.85 | 0.71 | 1.01 | 7.25E-02 | 0.350 |
| OID30452 | RAP1A | Inflammation_II | RAP1A | P62834 | 1 | 111542218 | 111716691 | 0.82 | 0.67 | 1.02 | 7.21E-02 | 0.350 |
| OID20950 | RHOC | Neurology | RHOC | P08134 | 1 | 112701127 | 112707434 | 0.86 | 0.73 | 1.01 | 7.24E-02 | 0.350 |
| OID20611 | TNFSF10 | Inflammation | TNFSF10 | P50591 | 3 | 172505508 | 172523475 | 0.86 | 0.73 | 1.01 | 7.26E-02 | 0.350 |
| OID20526 | TRIM5 | Inflammation | TRIM5 | Q9C035 | 11 | 5663195 | 5938619 | 0.86 | 0.72 | 1.01 | 7.24E-02 | 0.350 |
| OID30222 | COMMD1 | Cardiometabolic_II | COMMD1 | Q8N668 | 2 | 61888724 | 62147247 | 0.85 | 0.72 | 1.01 | 7.29E-02 | 0.351 |
| OID20136 | NTRK2 | Cardiometabolic | NTRK2 | Q16620 | 9 | 84668375 | 85095751 | 0.86 | 0.73 | 1.01 | 7.38E-02 | 0.355 |
| OID31215 | PITHD1 | Oncology_II | PITHD1 | Q9GZP4 | 1 | 23778418 | 23788232 | 1.13 | 0.99 | 1.28 | 7.40E-02 | 0.355 |
| OID31458 | DNM1 | Oncology_II | DNM1 | Q05193 | 9 | 128191655 | 128255248 | 0.86 | 0.73 | 1.01 | 7.42E-02 | 0.355 |
| OID31477 | SWAP70 | Oncology_II | SWAP70 | Q9UH65 | 11 | 9664077 | 9752993 | 0.84 | 0.69 | 1.02 | 7.43E-02 | 0.355 |
| OID21476 | ATOX1 | Oncology | ATOX1 | O00244 | 5 | 151742316 | 151772532 | 0.86 | 0.72 | 1.02 | 7.55E-02 | 0.357 |
| OID31454 | BCL2 | Oncology_II | BCL2 | P10415 | 18 | 63123346 | 63320128 | 0.82 | 0.67 | 1.02 | 7.58E-02 | 0.357 |
| OID20517 | BID | Inflammation | BID | P55957 | 22 | 17734138 | 17774770 | 0.85 | 0.71 | 1.02 | 7.49E-02 | 0.357 |
| OID21344 | CA12 | Oncology | CA12 | O43570 | 15 | 63321378 | 63381846 | 1.15 | 0.99 | 1.35 | 7.52E-02 | 0.357 |
| OID30239 | ECHS1 | Cardiometabolic_II | ECHS1 | P30084 | 10 | 133362485 | 133373354 | 0.86 | 0.72 | 1.02 | 7.58E-02 | 0.357 |
| OID20615 | HPCAL1 | Inflammation | HPCAL1 | P37235 | 2 | 10302889 | 10427617 | 0.87 | 0.74 | 1.01 | 7.56E-02 | 0.357 |
| OID20683 | NCK2 | Inflammation | NCK2 | O43639 | 2 | 105744912 | 105894274 | 0.86 | 0.73 | 1.02 | 7.55E-02 | 0.357 |
| OID20143 | PLIN3 | Cardiometabolic | PLIN3 | O60664 | 19 | 4838341 | 4867694 | 0.85 | 0.71 | 1.02 | 7.58E-02 | 0.357 |
| OID21015 | RWDD1 | Neurology | RWDD1 | Q9H446 | 6 | 116571409 | 116597675 | 0.86 | 0.73 | 1.02 | 7.53E-02 | 0.357 |
| OID31143 | TIMM8A | Neurology_II | TIMM8A | O60220 | X | 101345661 | 101348742 | 0.86 | 0.73 | 1.02 | 7.58E-02 | 0.357 |
| OID21402 | FGF21 | Oncology | FGF21 | Q9NSA1 | 19 | 48755524 | 48758333 | 1.15 | 0.98 | 1.35 | 7.64E-02 | 0.359 |
| OID31330 | TNPO1 | Oncology_II | TNPO1 | Q92973 | 5 | 72816312 | 72916733 | 0.86 | 0.73 | 1.02 | 7.66E-02 | 0.359 |
| OID20918 | EBAG9 | Neurology | EBAG9 | O00559 | 8 | 109539711 | 109565996 | 0.86 | 0.73 | 1.02 | 7.67E-02 | 0.359 |
| OID20764 | PLAUR | Inflammation | PLAUR | Q03405 | 19 | 43646095 | 43670547 | 1.17 | 0.98 | 1.38 | 7.70E-02 | 0.360 |
| OID20808 | CDHR1 | Neurology | CDHR1 | Q96JP9 | 10 | 84194537 | 84219621 | 0.85 | 0.70 | 1.02 | 7.73E-02 | 0.360 |
| OID30319 | CRYBB1 | Cardiometabolic_II | CRYBB1 | P53674 | 22 | 26599278 | 26618027 | 0.86 | 0.72 | 1.02 | 7.75E-02 | 0.361 |
| OID31011 | PPP1R14A | Neurology_II | PPP1R14A | Q96A00 | 19 | 38251237 | 38256532 | 0.86 | 0.72 | 1.02 | 7.76E-02 | 0.361 |
| OID21462 | WFDC12 | Oncology | WFDC12 | Q8WWY7 | 20 | 45123425 | 45124465 | 1.17 | 0.98 | 1.39 | 7.79E-02 | 0.362 |
| OID30292 | BNIP2 | Cardiometabolic_II | BNIP2 | Q12982 | 15 | 59659146 | 59689339 | 0.85 | 0.72 | 1.02 | 7.81E-02 | 0.362 |
| OID20552 | MAP2K6 | Inflammation | MAP2K6 | P52564 | 17 | 69414697 | 69553865 | 0.86 | 0.74 | 1.02 | 7.82E-02 | 0.362 |
| OID21341 | ATP6V1D | Oncology | ATP6V1D | Q9Y5K8 | 14 | 67294371 | 67360265 | 0.84 | 0.70 | 1.02 | 7.89E-02 | 0.363 |
| OID20225 | CA13 | Cardiometabolic | CA13 | Q8N1Q1 | 8 | 85220587 | 85284073 | 0.87 | 0.74 | 1.02 | 7.87E-02 | 0.363 |
| OID30253 | GATD3 | Cardiometabolic_II | GATD3 | P0DPI2 | 21 | 44133610 | 44210114 | 0.86 | 0.72 | 1.02 | 7.88E-02 | 0.363 |
| OID30688 | PEPD | Inflammation_II | PEPD | P12955 | 19 | 33386950 | 33521823 | 0.87 | 0.74 | 1.02 | 7.90E-02 | 0.363 |
| OID21120 | TREML2 | Neurology | TREML2 | Q5T2D2 | 6 | 41189749 | 41201149 | 0.86 | 0.73 | 1.02 | 7.87E-02 | 0.363 |
| OID20944 | GPC5 | Neurology | GPC5 | P78333 | 13 | 91398621 | 92873682 | 0.87 | 0.74 | 1.02 | 7.99E-02 | 0.366 |
| OID20494 | JCHAIN | Inflammation | JCHAIN | P01591 | 4 | 70655541 | 70681817 | 0.87 | 0.74 | 1.02 | 8.00E-02 | 0.366 |
| OID20051 | HNRNPK | Cardiometabolic | HNRNPK | P61978 | 9 | 83968083 | 83980616 | 0.86 | 0.73 | 1.02 | 8.03E-02 | 0.367 |
| OID30166 | ARL13B | Cardiometabolic_II | ARL13B | Q3SXY8 | 3 | 93980139 | 94055678 | 0.86 | 0.72 | 1.02 | 8.04E-02 | 0.367 |

| | | | | | | | | | | | | |
|---|---|---|---|---|---|---|---|---|---|---|---|---|
| OID20623 | NUDC | Inflammation | NUDC | Q9Y266 | 1 | 26900238 | 26946871 | 0.87 | 0.74 | 1.02 | 8.11E-02 | 0.368 |
| OID20467 | PADI2 | Inflammation | PADI2 | Q9Y2J8 | 1 | 17066761 | 17119451 | 1.13 | 0.98 | 1.30 | 8.07E-02 | 0.368 |
| OID31346 | RAB44 | Oncology_II | RAB44 | Q7Z6P3 | 6 | 36697826 | 36733184 | 0.81 | 0.64 | 1.03 | 8.11E-02 | 0.368 |
| OID21194 | RASSF2 | Oncology | RASSF2 | P50749 | 20 | 4780023 | 4823608 | 0.84 | 0.68 | 1.02 | 8.10E-02 | 0.368 |
| OID20582 | AXIN1 | Inflammation | AXIN1 | O15169 | 16 | 287440 | 352723 | 0.87 | 0.74 | 1.02 | 8.21E-02 | 0.368 |
| OID20774 | DAG1 | Inflammation | DAG1 | Q14118 | 3 | 49468703 | 49535618 | 0.86 | 0.73 | 1.02 | 8.28E-02 | 0.368 |
| OID21143 | DBI | Neurology | DBI | P07108 | 2 | 119366924 | 119372550 | 0.86 | 0.72 | 1.02 | 8.23E-02 | 0.368 |
| OID30578 | ERP29 | Inflammation_II | ERP29 | P30040 | 12 | 112013348 | 112023449 | 0.87 | 0.74 | 1.02 | 8.20E-02 | 0.368 |
| OID31134 | ESYT2 | Neurology_II | ESYT2 | A0FGR8 | 7 | 158730995 | 158830253 | 0.86 | 0.72 | 1.02 | 8.19E-02 | 0.368 |
| OID30112 | IGF2BP3 | Cardiometabolic_II | IGF2BP3 | O00425 | 7 | 23310209 | 23470491 | 0.84 | 0.69 | 1.02 | 8.20E-02 | 0.368 |
| OID31312 | KIAA1549L | Oncology_II | KIAA1549L | Q6ZVL6 | 11 | 33376108 | 33674102 | 0.85 | 0.72 | 1.02 | 8.28E-02 | 0.368 |
| OID31191 | KIR3DL2 | Oncology_II | KIR3DL2 | P43630 | 19 | 54850443 | 54867207 | 0.83 | 0.68 | 1.02 | 8.24E-02 | 0.368 |
| OID31082 | KLRK1 | Neurology_II | KLRK1 | P26718 | 12 | 10372353 | 10391874 | 0.87 | 0.74 | 1.02 | 8.18E-02 | 0.368 |
| OID31258 | NFKB2 | Oncology_II | NFKB2 | Q00653 | 10 | 102394110 | 102402524 | 0.81 | 0.63 | 1.03 | 8.26E-02 | 0.368 |
| OID30234 | RECK | Cardiometabolic_II | RECK | O95980 | 9 | 36036913 | 36124455 | 0.86 | 0.73 | 1.02 | 8.23E-02 | 0.368 |
| OID30398 | SBSN | Cardiometabolic_II | SBSN | Q6UWP8 | 19 | 35523367 | 35528351 | 1.14 | 0.98 | 1.32 | 8.18E-02 | 0.368 |
| OID31419 | UNG | Oncology_II | UNG | P13051 | 12 | 109097597 | 109126725 | 0.86 | 0.73 | 1.02 | 8.27E-02 | 0.368 |
| OID30674 | NRGN | Inflammation_II | NRGN | Q92686 | 11 | 124739942 | 124747210 | 0.86 | 0.73 | 1.02 | 8.30E-02 | 0.368 |
| OID30692 | VASP | Inflammation_II | VASP | P50552 | 19 | 45506579 | 45526989 | 0.86 | 0.73 | 1.02 | 8.32E-02 | 0.369 |
| OID30357 | RAB10 | Cardiometabolic_II | RAB10 | P61026 | 2 | 26034084 | 26137454 | 0.86 | 0.73 | 1.02 | 8.36E-02 | 0.369 |
| OID21165 | SFTPA1 | Oncology | SFTPA1 | Q8IWL2 | 10 | 79610939 | 79615455 | 0.85 | 0.70 | 1.02 | 8.36E-02 | 0.369 |
| OID31272 | VSTM2B | Oncology_II | VSTM2B | A6NLU5 | 19 | 29525994 | 29564551 | 0.83 | 0.67 | 1.03 | 8.35E-02 | 0.369 |
| OID21463 | ANGPT2 | Oncology | ANGPT2 | O15123 | 8 | 6499632 | 6563409 | 1.16 | 0.98 | 1.38 | 8.39E-02 | 0.370 |
| OID31073 | MORC3 | Neurology_II | MORC3 | Q14149 | 21 | 36320189 | 36386148 | 0.84 | 0.69 | 1.02 | 8.42E-02 | 0.370 |
| OID30795 | VAV3 | Neurology_II | VAV3 | Q9UKW4 | 1 | 107571161 | 107965180 | 0.86 | 0.73 | 1.02 | 8.41E-02 | 0.370 |
| OID21265 | CD28 | Oncology | CD28 | P10747 | 2 | 203706475 | 203738912 | 0.87 | 0.74 | 1.02 | 8.46E-02 | 0.371 |
| OID31409 | MTSS2 | Oncology_II | MTSS2 | Q765P7 | 16 | 70661204 | 70686053 | 0.87 | 0.74 | 1.02 | 8.47E-02 | 0.371 |
| OID30279 | BPIFA2 | Cardiometabolic_II | BPIFA2 | Q96DR5 | 20 | 33161768 | 33181412 | 0.86 | 0.72 | 1.02 | 8.50E-02 | 0.371 |
| OID20892 | PFDN2 | Neurology | PFDN2 | Q9UHV9 | 1 | 161100556 | 161118055 | 0.86 | 0.72 | 1.02 | 8.53E-02 | 0.371 |
| OID30104 | SCGB2A2 | Cardiometabolic_II | SCGB2A2 | Q13296 | 11 | 62270158 | 62273160 | 0.85 | 0.70 | 1.02 | 8.52E-02 | 0.371 |
| OID31452 | ZFYVE19 | Oncology_II | ZFYVE19 | Q96K21 | 15 | 40807086 | 40815084 | 0.86 | 0.73 | 1.02 | 8.50E-02 | 0.371 |
| OID30186 | PACS2 | Cardiometabolic_II | PACS2 | Q86VP3 | 14 | 105300563 | 105398147 | 0.86 | 0.73 | 1.02 | 8.57E-02 | 0.372 |
| OID30498 | BCL2L15 | Inflammation_II | BCL2L15 | Q5TBC7 | 1 | 113876816 | 113887581 | 0.84 | 0.69 | 1.03 | 8.61E-02 | 0.373 |
| OID30643 | NAGPA | Inflammation_II | NAGPA | Q9UK23 | 16 | 5024844 | 5034141 | 0.87 | 0.74 | 1.02 | 8.61E-02 | 0.373 |
| OID31394 | DUSP13 | Oncology_II | DUSP13 | Q6B8I1 | 10 | 75094432 | 75109221 | 0.86 | 0.72 | 1.02 | 8.64E-02 | 0.373 |
| OID20779 | MPIG6B | Inflammation | MPIG6B | O95866 | 6 | 31718594 | 31726714 | 0.88 | 0.75 | 1.02 | 8.64E-02 | 0.373 |
| OID31362 | RAB2B | Oncology_II | RAB2B | Q8WUD1 | 14 | 21459020 | 21476960 | 0.86 | 0.73 | 1.02 | 8.65E-02 | 0.373 |
| OID21401 | CA14 | Oncology | CA14 | Q9ULX7 | 1 | 150257251 | 150265078 | 0.86 | 0.72 | 1.02 | 8.69E-02 | 0.374 |
| OID20859 | CARHSP1 | Neurology | CARHSP1 | Q9Y2V2 | 16 | 8852942 | 8869012 | 0.86 | 0.72 | 1.02 | 8.82E-02 | 0.374 |
| OID30619 | CRELD1 | Inflammation_II | CRELD1 | Q96HD1 | 3 | 9933793 | 9945413 | 0.87 | 0.73 | 1.02 | 8.83E-02 | 0.374 |
| OID21520 | ERBB2 | Oncology | ERBB2 | P04626 | 17 | 39687914 | 39730426 | 0.86 | 0.73 | 1.02 | 8.80E-02 | 0.374 |
| OID30304 | GGCT | Cardiometabolic_II | GGCT | O75223 | 7 | 30496621 | 30504841 | 0.86 | 0.72 | 1.02 | 8.81E-02 | 0.374 |
| OID30174 | GP1BB | Cardiometabolic_II | GP1BB | P13224 | 22 | 19723539 | 19724771 | 0.87 | 0.74 | 1.02 | 8.76E-02 | 0.374 |
| OID21471 | HTRA2 | Oncology | HTRA2 | O43464 | 2 | 74529725 | 74533332 | 0.86 | 0.73 | 1.02 | 8.75E-02 | 0.374 |
| OID21210 | LSM1 | Oncology | LSM1 | O15116 | 8 | 38163335 | 38176730 | 0.85 | 0.70 | 1.02 | 8.73E-02 | 0.374 |
| OID30055 | NOP56 | Cardiometabolic_II | NOP56 | O00567 | 20 | 2652593 | 2658393 | 0.83 | 0.67 | 1.03 | 8.71E-02 | 0.374 |
| OID20685 | PLA2G4A | Inflammation | PLA2G4A | P47712 | 1 | 186828949 | 186988981 | 0.87 | 0.74 | 1.02 | 8.79E-02 | 0.374 |
| OID21436 | RILP | Oncology | RILP | Q96NA2 | 17 | 1646145 | 1649866 | 0.87 | 0.73 | 1.02 | 8.81E-02 | 0.374 |
| OID31353 | SMAD2 | Oncology_II | SMAD2 | Q15796 | 18 | 47808957 | 47931146 | 0.86 | 0.73 | 1.02 | 8.77E-02 | 0.374 |
| OID20720 | SPINT2 | Inflammation | SPINT2 | O43291 | 19 | 38244035 | 38292615 | 0.87 | 0.74 | 1.02 | 8.68E-02 | 0.374 |
| OID20868 | TST | Neurology | TST | Q16762 | 22 | 37010859 | 37020183 | 0.86 | 0.73 | 1.02 | 8.87E-02 | 0.375 |
| OID20853 | ING1 | Neurology | ING1 | Q9UK53 | 13 | 110712736 | 110723339 | 0.84 | 0.69 | 1.03 | 8.90E-02 | 0.375 |
| OID30983 | NRN1 | Neurology_II | NRN1 | Q9NPD7 | 6 | 5997999 | 6007605 | 0.84 | 0.69 | 1.03 | 8.89E-02 | 0.375 |
| OID20374 | APOM | Cardiometabolic | APOM | O95445 | 6 | 31652416 | 31658210 | 0.87 | 0.75 | 1.02 | 8.99E-02 | 0.375 |
| OID30345 | CDA | Cardiometabolic_II | CDA | P32320 | 1 | 20589086 | 20618903 | 0.86 | 0.72 | 1.02 | 9.01E-02 | 0.375 |
| OID21238 | CDC27 | Oncology | CDC27 | P30260 | 17 | 47117703 | 47189422 | 0.86 | 0.72 | 1.02 | 9.01E-02 | 0.375 |
| OID20930 | CHMP1A | Neurology | CHMP1A | Q9HD42 | 16 | 89640816 | 89657738 | 0.87 | 0.74 | 1.02 | 9.01E-02 | 0.375 |
| OID30570 | ITGA2 | Inflammation_II | ITGA2 | P17301 | 5 | 52989340 | 53094779 | 0.86 | 0.73 | 1.02 | 8.96E-02 | 0.375 |
| OID30125 | PMM2 | Cardiometabolic_II | PMM2 | O15305 | 16 | 8788823 | 8862534 | 0.87 | 0.74 | 1.02 | 8.97E-02 | 0.375 |
| OID30656 | SPART | Inflammation_II | SPART | Q8N0X7 | 13 | 36301638 | 36370180 | 0.86 | 0.73 | 1.02 | 8.93E-02 | 0.375 |
| OID31453 | SSNA1 | Oncology_II | SSNA1 | O43805 | 9 | 137188660 | 137190370 | 0.86 | 0.73 | 1.02 | 8.97E-02 | 0.375 |
| OID20642 | TPSAB1 | Inflammation | TPSAB1 | Q15661 | 16 | 1240379 | 1242554 | 0.87 | 0.74 | 1.02 | 8.94E-02 | 0.375 |
| OID20151 | FADD | Cardiometabolic | FADD | Q13158 | 11 | 70203296 | 70207390 | 0.87 | 0.73 | 1.02 | 9.03E-02 | 0.375 |
| OID30846 | STEAP4 | Neurology_II | STEAP4 | Q687X5 | 7 | 88270892 | 88306894 | 0.84 | 0.68 | 1.03 | 9.07E-02 | 0.376 |
| OID20964 | TNFRSF6B | Neurology | TNFRSF6B | O95407 | 20 | 63696652 | 63698684 | 1.13 | 0.98 | 1.30 | 9.08E-02 | 0.376 |
| OID20965 | LAIR2 | Neurology | LAIR2 | Q6ISS4 | 19 | 54497879 | 54510687 | 1.17 | 0.98 | 1.40 | 9.12E-02 | 0.377 |
| OID20507 | TRAF2 | Inflammation | TRAF2 | Q12933 | 9 | 136881912 | 136926607 | 0.86 | 0.73 | 1.02 | 9.11E-02 | 0.377 |
| OID20881 | KEL | Neurology | KEL | P23276 | 7 | 142941114 | 142962363 | 1.15 | 0.98 | 1.36 | 9.14E-02 | 0.377 |
| OID31423 | SDCCAG8 | Oncology_II | SDCCAG8 | Q86SQ7 | 1 | 243256034 | 243500091 | 0.87 | 0.74 | 1.02 | 9.15E-02 | 0.377 |
| OID30778 | APCS | Inflammation_II | APCS | P02743 | 1 | 159587826 | 159588865 | 0.85 | 0.70 | 1.03 | 9.18E-02 | 0.378 |
| OID30883 | C2orf69 | Neurology_II | C2orf69 | Q8N8R5 | 2 | 199911293 | 199955935 | 0.84 | 0.69 | 1.03 | 9.57E-02 | 0.381 |
| OID31039 | CACNB3 | Neurology_II | CACNB3 | P54284 | 12 | 48813794 | 48828941 | 0.87 | 0.74 | 1.02 | 9.38E-02 | 0.381 |
| OID20441 | CEP164 | Inflammation | CEP164 | Q9UPV0 | 11 | 117314557 | 117413266 | 0.84 | 0.69 | 1.03 | 9.58E-02 | 0.381 |
| OID20127 | CNST | Cardiometabolic | CNST | Q6PJW8 | 1 | 246566444 | 246668595 | 0.87 | 0.74 | 1.02 | 9.47E-02 | 0.381 |
| OID30397 | CSPG4 | Cardiometabolic_II | CSPG4 | Q6UVK1 | 15 | 75674322 | 75712848 | 1.15 | 0.98 | 1.37 | 9.47E-02 | 0.381 |
| OID31104 | DNAJC21 | Neurology_II | DNAJC21 | Q5F1R6 | 5 | 34929559 | 34958964 | 0.86 | 0.73 | 1.02 | 9.32E-02 | 0.381 |
| OID21460 | DSG3 | Oncology | DSG3 | P32926 | 18 | 31447741 | 31478702 | 1.15 | 0.98 | 1.37 | 9.56E-02 | 0.381 |
| OID21017 | EZR | Neurology | EZR | P15311 | 6 | 158765741 | 158819368 | 0.86 | 0.72 | 1.03 | 9.52E-02 | 0.381 |
| OID20330 | FAP | Cardiometabolic | FAP | Q12884 | 2 | 162170684 | 162245151 | 0.88 | 0.75 | 1.02 | 9.31E-02 | 0.381 |
| OID21259 | GFER | Oncology | GFER | P55789 | 16 | 1984193 | 1987749 | 0.86 | 0.72 | 1.03 | 9.58E-02 | 0.381 |
| OID30663 | GLRX5 | Inflammation_II | GLRX5 | Q86SX6 | 14 | 95533503 | 95544724 | 0.87 | 0.73 | 1.03 | 9.57E-02 | 0.381 |
| OID21173 | GSAP | Oncology | GSAP | A4D1B5 | 7 | 77310751 | 77416349 | 0.84 | 0.68 | 1.03 | 9.32E-02 | 0.381 |
| OID21351 | ICOSLG | Oncology | ICOSLG | O75144 | 21 | 44217014 | 44240966 | 1.14 | 0.98 | 1.34 | 9.60E-02 | 0.381 |
| OID20619 | IDS | Inflammation | IDS | P22304 | X | 149476988 | 149521096 | 0.88 | 0.77 | 1.02 | 9.60E-02 | 0.381 |
| OID20370 | KIT | Cardiometabolic | KIT | P10721 | 4 | 54657267 | 54740783 | 0.87 | 0.74 | 1.03 | 9.56E-02 | 0.381 |
| OID31372 | LARP1 | Oncology_II | LARP1 | Q6PKG0 | 5 | 154682986 | 154817605 | 0.86 | 0.71 | 1.03 | 9.44E-02 | 0.381 |
| OID31032 | LCN15 | Neurology_II | LCN15 | Q6UWW0 | 9 | 136759634 | 136766255 | 1.13 | 0.98 | 1.29 | 9.58E-02 | 0.381 |
| OID21181 | MAGED1 | Oncology | MAGED1 | Q9Y5V3 | X | 51803007 | 51902354 | 0.86 | 0.72 | 1.03 | 9.56E-02 | 0.381 |
| OID30465 | PIKFYVE | Inflammation_II | PIKFYVE | Q9Y2I7 | 2 | 208266255 | 208358746 | 0.85 | 0.71 | 1.03 | 9.59E-02 | 0.381 |
| OID21011 | PVR | Neurology | PVR | P15151 | 19 | 44643798 | 44666162 | 1.15 | 0.98 | 1.35 | 9.47E-02 | 0.381 |
| OID21163 | RABEPK | Oncology | RABEPK | Q7Z6M1 | 9 | 125200542 | 125234161 | 0.86 | 0.71 | 1.03 | 9.40E-02 | 0.381 |
| OID20216 | SDC4 | Cardiometabolic | SDC4 | P31431 | 20 | 45325288 | 45348424 | 0.88 | 0.76 | 1.02 | 9.43E-02 | 0.381 |
| OID30968 | SEZ6 | Neurology_II | SEZ6 | Q53EL9 | 17 | 28954901 | 29006440 | 1.12 | 0.98 | 1.28 | 9.42E-02 | 0.381 |
| OID30358 | TIMM10 | Cardiometabolic_II | TIMM10 | P62072 | 11 | 57528464 | 57530803 | 0.86 | 0.72 | 1.03 | 9.48E-02 | 0.381 |
| OID20967 | TNFRSF10A | Neurology | TNFRSF10A | O00220 | 8 | 23190452 | 23225102 | 1.15 | 0.98 | 1.36 | 9.55E-02 | 0.381 |
| OID31366 | WFDC1 | Oncology_II | WFDC1 | Q9HC57 | 16 | 84294846 | 84329844 | 0.85 | 0.71 | 1.03 | 9.57E-02 | 0.381 |
| OID21319 | CDKN1A | Oncology | CDKN1A | P38936 | 6 | 36676460 | 36687337 | 0.87 | 0.74 | 1.03 | 9.67E-02 | 0.381 |
| OID30217 | COQ7 | Cardiometabolic_II | COQ7 | Q99807 | 16 | 19067614 | 19080095 | 0.85 | 0.71 | 1.03 | 9.62E-02 | 0.381 |
| OID31512 | CYB5R2 | Oncology_II | CYB5R2 | Q6BCY4 | 11 | 7665100 | 7677222 | 0.86 | 0.72 | 1.03 | 9.63E-02 | 0.381 |
| OID30582 | DIPK2B | Inflammation_II | DIPK2B | Q9H7Y0 | X | 45148373 | 45200901 | 0.87 | 0.73 | 1.03 | 9.65E-02 | 0.381 |
| OID20234 | TYMP | Cardiometabolic | TYMP | P19971 | 22 | 50525752 | 50530032 | 0.86 | 0.73 | 1.03 | 9.67E-02 | 0.381 |
| OID30064 | CACNA1H | Cardiometabolic_II | CACNA1H | O95180 | 16 | 1153106 | 1221771 | 1.13 | 0.98 | 1.30 | 9.71E-02 | 0.382 |
| OID31123 | WASHC3 | Neurology_II | WASHC3 | Q9Y3C0 | 12 | 102012840 | 102062149 | 0.86 | 0.72 | 1.03 | 9.70E-02 | 0.382 |
| OID21405 | CTSV | Oncology | CTSV | O60911 | 9 | 97029677 | 97156556 | 0.87 | 0.73 | 1.03 | 9.73E-02 | 0.382 |
| OID20977 | IPCEF1 | Neurology | IPCEF1 | Q8WWN9 | 6 | 154154496 | 154356803 | 0.87 | 0.74 | 1.03 | 9.74E-02 | 0.382 |
| OID20768 | LTBR | Inflammation | LTBR | P36941 | 12 | 6375045 | 6391571 | 1.14 | 0.98 | 1.32 | 9.78E-02 | 0.383 |
| OID20903 | DARS1 | Neurology | DARS1 | P14868 | 2 | 135905881 | 135986100 | 0.87 | 0.75 | 1.03 | 9.80E-02 | 0.383 |

| | | | | | | | | | | | | |
|---|---|---|---|---|---|---|---|---|---|---|---|---|
| OID30229 | EHD3 | Cardiometabolic_II | EHD3 | Q9NZN3 | 2 | 31234152 | 31269451 | 0.87 | 0.74 | 1.03 | 9.82E-02 | 0.383 |
| OID21154 | TFF1 | Neurology | TFF1 | P04155 | 21 | 42362282 | 42366535 | 1.14 | 0.98 | 1.33 | 9.83E-02 | 0.383 |
| OID20354 | NCAM1 | Cardiometabolic | NCAM1 | P13591 | 11 | 112961247 | 113278436 | 1.15 | 0.97 | 1.35 | 9.87E-02 | 0.384 |
| OID31085 | OSBPL2 | Neurology_II | OSBPL2 | Q9H1P3 | 20 | 62231922 | 62296213 | 0.84 | 0.68 | 1.03 | 9.87E-02 | 0.384 |
| OID31013 | GPR158 | Neurology_II | GPR158 | Q5T848 | 10 | 25174802 | 25602229 | 0.86 | 0.71 | 1.03 | 9.91E-02 | 0.385 |
| OID21041 | LGALS8 | Neurology | LGALS8 | O00214 | 1 | 236518000 | 236552981 | 0.86 | 0.73 | 1.03 | 9.93E-02 | 0.385 |
| OID20475 | SPRY2 | Inflammation | SPRY2 | O43597 | 13 | 80335976 | 80341126 | 0.87 | 0.73 | 1.03 | 9.93E-02 | 0.385 |
| OID31102 | NT5C1A | Neurology_II | NT5C1A | Q9BXI3 | 1 | 39651229 | 39672107 | 0.85 | 0.70 | 1.03 | 9.95E-02 | 0.385 |
| OID20193 | CDHR5 | Cardiometabolic | CDHR5 | Q9HBB8 | 11 | 616577 | 626078 | 0.88 | 0.75 | 1.03 | 1.00E-01 | 0.386 |
| OID31498 | ENDOU | Oncology_II | ENDOU | P21128 | 12 | 47709734 | 47725567 | 1.15 | 0.97 | 1.35 | 1.00E-01 | 0.386 |
| OID20872 | FHIT | Neurology | FHIT | P49789 | 3 | 59747277 | 61251459 | 0.87 | 0.74 | 1.03 | 1.00E-01 | 0.386 |
| OID20447 | RABGAP1L | Inflammation | RABGAP1L | Q5R372 | 1 | 174159410 | 174995308 | 0.86 | 0.73 | 1.03 | 1.00E-01 | 0.386 |
| OID20053 | VIM | Cardiometabolic | VIM | P08670 | 10 | 17228241 | 17237593 | 0.89 | 0.77 | 1.02 | 1.00E-01 | 0.386 |
| OID21489 | MDK | Oncology | MDK | P21741 | 11 | 46380756 | 46383837 | 0.86 | 0.73 | 1.03 | 1.01E-01 | 0.386 |
| OID31319 | TAB2 | Oncology_II | TAB2 | Q9NYJ8 | 6 | 149218641 | 149411613 | 0.87 | 0.74 | 1.03 | 1.01E-01 | 0.387 |
| OID21217 | BIRC2 | Oncology | BIRC2 | Q13490 | 11 | 102347211 | 102378670 | 0.87 | 0.73 | 1.03 | 1.01E-01 | 0.388 |
| OID20415 | REG1A | Cardiometabolic | REG1A | P05451 | 2 | 79120362 | 79123409 | 1.13 | 0.98 | 1.31 | 1.02E-01 | 0.388 |
| OID21023 | SERPINB6 | Neurology | SERPINB6 | P35237 | 6 | 2948159 | 2972165 | 0.87 | 0.73 | 1.03 | 1.02E-01 | 0.388 |
| OID20595 | CD200R1 | Inflammation | CD200R1 | Q8TD46 | 3 | 112921205 | 112975103 | 0.87 | 0.74 | 1.03 | 1.02E-01 | 0.390 |
| OID21218 | SFTPA2 | Oncology | SFTPA2 | Q8IWL1 | 10 | 79555852 | 79560407 | 1.14 | 0.97 | 1.34 | 1.02E-01 | 0.390 |
| OID20579 | DECR1 | Inflammation | DECR1 | Q16698 | 8 | 90001405 | 90053633 | 0.87 | 0.74 | 1.03 | 1.03E-01 | 0.390 |
| OID30244 | AHNAK2 | Cardiometabolic_II | AHNAK2 | Q8IVF2 | 14 | 104937244 | 104978374 | 0.86 | 0.72 | 1.03 | 1.04E-01 | 0.392 |
| OID21434 | APEX1 | Oncology | APEX1 | P27695 | 14 | 20455191 | 20457772 | 0.87 | 0.73 | 1.03 | 1.04E-01 | 0.392 |
| OID21202 | ARHGAP1 | Oncology | ARHGAP1 | Q07960 | 11 | 46677080 | 46700619 | 0.87 | 0.73 | 1.03 | 1.04E-01 | 0.392 |
| OID20244 | CES1 | Cardiometabolic | CES1 | P23141 | 16 | 55802851 | 55833337 | 0.87 | 0.73 | 1.03 | 1.03E-01 | 0.392 |
| OID20639 | FCRL2 | Inflammation | FCRL2 | Q96LA5 | 1 | 157745733 | 157777132 | 0.87 | 0.74 | 1.03 | 1.04E-01 | 0.392 |
| OID30690 | FCN1 | Inflammation_II | FCN1 | O00602 | 9 | 134903232 | 134917912 | 0.88 | 0.76 | 1.03 | 1.04E-01 | 0.393 |
| OID21104 | THBS2 | Neurology | THBS2 | P35442 | 6 | 169215780 | 169254050 | 1.14 | 0.97 | 1.33 | 1.05E-01 | 0.394 |
| OID30225 | SCRIB | Cardiometabolic_II/Inflan | SCRIB | Q14160 | 8 | 143790920 | 143815773 | 0.86 | 0.72 | 1.03 | 1.05E-01 | 0.394 |
| OID30204 | AIF1L | Cardiometabolic_II | AIF1L | Q9BQI0 | 9 | 131096476 | 131123152 | 0.84 | 0.68 | 1.04 | 1.05E-01 | 0.394 |
| OID31459 | MEP1A | Oncology_II | MEP1A | Q16819 | 6 | 46793389 | 46839778 | 1.15 | 0.97 | 1.35 | 1.05E-01 | 0.394 |
| OID30206 | PAGR1 | Cardiometabolic_II | PAGR1 | Q9BTK6 | 16 | 29816152 | 29822489 | 0.86 | 0.71 | 1.03 | 1.05E-01 | 0.394 |
| OID21311 | AARSD1 | Oncology | AARSD1 | Q9BTE6 | 17 | 42950526 | 42964498 | 0.87 | 0.73 | 1.03 | 1.07E-01 | 0.395 |
| OID30370 | ADAMTSL2 | Cardiometabolic_II | ADAMTSL2 | Q86TH1 | 9 | 133532164 | 133575519 | 1.14 | 0.97 | 1.35 | 1.07E-01 | 0.395 |
| OID31348 | AP1G2 | Oncology_II | AP1G2 | O75843 | 14 | 23559565 | 23568070 | 0.87 | 0.73 | 1.03 | 1.07E-01 | 0.395 |
| OID30753 | C1R | Inflammation_II | C1R | P00736 | 12 | 7080214 | 7092540 | 0.84 | 0.68 | 1.04 | 1.07E-01 | 0.395 |
| OID31333 | COL4A4 | Oncology_II | COL4A4 | P53420 | 2 | 227002714 | 227164453 | 0.90 | 0.79 | 1.02 | 1.07E-01 | 0.395 |
| OID21457 | GFRA1 | Oncology | GFRA1 | P56159 | 10 | 116056925 | 116276803 | 1.14 | 0.97 | 1.34 | 1.07E-01 | 0.395 |
| OID21160 | PARK7 | Neurology | PARK7 | Q99497 | 1 | 7954291 | 7985505 | 0.87 | 0.74 | 1.03 | 1.07E-01 | 0.395 |
| OID31027 | RNASEH2A | Neurology_II | RNASEH2A | O75792 | 19 | 12806584 | 12813640 | 0.85 | 0.70 | 1.04 | 1.06E-01 | 0.395 |
| OID21031 | SULT1A1 | Neurology | SULT1A1 | P50225 | 16 | 28603944 | 28623625 | 0.88 | 0.75 | 1.03 | 1.07E-01 | 0.395 |
| OID20068 | TIA1 | Cardiometabolic | TIA1 | P31483 | 2 | 70209444 | 70248660 | 0.87 | 0.74 | 1.03 | 1.07E-01 | 0.395 |
| OID31335 | TRDMT1 | Oncology_II | TRDMT1 | O14717 | 10 | 17137336 | 17202054 | 0.86 | 0.72 | 1.03 | 1.07E-01 | 0.395 |
| OID31181 | UNC79 | Oncology_II | UNC79 | Q9P2D8 | 14 | 93333219 | 93707876 | 0.86 | 0.72 | 1.03 | 1.06E-01 | 0.395 |
| OID30555 | VAMP8 | Inflammation_II | VAMP8 | Q9BV40 | 2 | 85561562 | 85582031 | 0.87 | 0.73 | 1.03 | 1.07E-01 | 0.395 |
| OID30191 | LRCH4 | Cardiometabolic_II | LRCH4 | O75427 | 7 | 100574011 | 100586129 | 0.86 | 0.72 | 1.03 | 1.07E-01 | 0.396 |
| OID30360 | CMC1 | Cardiometabolic_II | CMC1 | Q7Z7K0 | 3 | 28241584 | 28325142 | 0.87 | 0.74 | 1.03 | 1.08E-01 | 0.396 |
| OID20681 | DBNL | Inflammation | DBNL | Q9UJU6 | 7 | 44044640 | 44069456 | 0.88 | 0.74 | 1.03 | 1.08E-01 | 0.396 |
| OID31009 | PHACTR2 | Neurology_II | PHACTR2 | O75167 | 6 | 143536845 | 143831185 | 0.87 | 0.74 | 1.03 | 1.08E-01 | 0.396 |
| OID20604 | GZMB | Inflammation | GZMB | P10144 | 14 | 24630954 | 24634267 | 0.86 | 0.71 | 1.03 | 1.08E-01 | 0.397 |
| OID21063 | MSR1 | Neurology | MSR1 | P21757 | 8 | 16107878 | 16567490 | 1.15 | 0.97 | 1.37 | 1.09E-01 | 0.398 |
| OID20266 | REG1B | Cardiometabolic | REG1B | P48304 | 2 | 79085023 | 79088019 | 1.14 | 0.97 | 1.33 | 1.10E-01 | 0.400 |
| OID20091 | WASF1 | Cardiometabolic | WASF1 | Q92558 | 6 | 110099819 | 110180004 | 0.87 | 0.74 | 1.03 | 1.09E-01 | 0.400 |
| OID20207 | CXCL5 | Cardiometabolic | CXCL5 | P42830 | 4 | 73995642 | 73998677 | 0.87 | 0.74 | 1.03 | 1.10E-01 | 0.400 |
| OID31507 | GIPC3 | Oncology_II | GIPC3 | Q8TF64 | 19 | 3585478 | 3593541 | 0.88 | 0.75 | 1.03 | 1.10E-01 | 0.400 |
| OID30313 | MYDGF | Cardiometabolic_II | MYDGF | Q969H8 | 19 | 4641374 | 4670362 | 0.87 | 0.74 | 1.03 | 1.10E-01 | 0.400 |
| OID20302 | ST6GAL1 | Cardiometabolic | ST6GAL1 | P15907 | 3 | 186930325 | 187078553 | 0.87 | 0.74 | 1.03 | 1.10E-01 | 0.400 |
| OID30754 | BTD | Inflammation_II | BTD | P43251 | 3 | 15601341 | 15722311 | 0.88 | 0.75 | 1.03 | 1.11E-01 | 0.400 |
| OID20319 | EGFR | Cardiometabolic | EGFR | P00533 | 7 | 55019017 | 55211628 | 0.87 | 0.74 | 1.03 | 1.11E-01 | 0.400 |
| OID21126 | IL1RAP | Neurology | IL1RAP | Q9NPH3 | 3 | 190514051 | 190659750 | 0.87 | 0.73 | 1.03 | 1.11E-01 | 0.401 |
| OID30907 | LRP2BP | Neurology_II | LRP2BP | Q9P2M1 | 4 | 185363872 | 185395924 | 0.86 | 0.72 | 1.04 | 1.11E-01 | 0.401 |
| OID31300 | OGT | Oncology_II | OGT | O15294 | X | 71533087 | 71575892 | 0.85 | 0.70 | 1.04 | 1.11E-01 | 0.401 |
| OID20953 | TNFSF14 | Neurology | TNFSF14 | O43557 | 19 | 6661253 | 6670588 | 0.87 | 0.73 | 1.03 | 1.11E-01 | 0.401 |
| OID21000 | EIF4B | Neurology | EIF4B | P23588 | 12 | 53006282 | 53042215 | 0.88 | 0.75 | 1.03 | 1.12E-01 | 0.402 |
| OID21093 | INHBC | Neurology | INHBC | P55103 | 12 | 57434784 | 57452062 | 0.87 | 0.72 | 1.03 | 1.12E-01 | 0.402 |
| OID31411 | TMEM106A | Oncology_II | TMEM106A | Q96A25 | 17 | 43211835 | 43220041 | 0.87 | 0.74 | 1.03 | 1.12E-01 | 0.402 |
| OID20547 | CLEC4C | Inflammation | CLEC4C | Q8WTT0 | 12 | 7729383 | 7751605 | 0.88 | 0.74 | 1.03 | 1.13E-01 | 0.403 |
| OID31338 | FAM171B | Oncology_II | FAM171B | Q6P995 | 2 | 186694060 | 186765959 | 0.86 | 0.71 | 1.04 | 1.13E-01 | 0.403 |
| OID20285 | MEGF9 | Cardiometabolic | MEGF9 | Q9H1U4 | 9 | 120600811 | 120714470 | 0.87 | 0.74 | 1.03 | 1.13E-01 | 0.403 |
| OID31212 | PFDN4 | Oncology_II | PFDN4 | Q9NQP4 | 20 | 54208087 | 54228052 | 0.85 | 0.70 | 1.04 | 1.13E-01 | 0.403 |
| OID20071 | VAMP5 | Cardiometabolic | VAMP5 | O95183 | 2 | 85584431 | 85593406 | 0.86 | 0.72 | 1.04 | 1.13E-01 | 0.403 |
| OID30490 | XIAP | Inflammation_II | XIAP | P98170 | X | 123859712 | 123913972 | 0.87 | 0.73 | 1.03 | 1.13E-01 | 0.403 |
| OID21335 | DRG2 | Oncology | DRG2 | P55039 | 17 | 18087892 | 18107970 | 0.87 | 0.73 | 1.03 | 1.14E-01 | 0.403 |
| OID30295 | GALNT5 | Cardiometabolic_II | GALNT5 | Q7Z7M9 | 2 | 157257705 | 157318491 | 1.15 | 0.97 | 1.36 | 1.14E-01 | 0.403 |
| OID30548 | PPM1B | Inflammation_II | PPM1B | O75688 | 2 | 44167969 | 44244384 | 1.11 | 0.97 | 1.27 | 1.14E-01 | 0.403 |
| OID30325 | RAB11FIP3 | Cardiometabolic_II | RAB11FIP3 | O75154 | 16 | 425649 | 523011 | 0.88 | 0.75 | 1.03 | 1.14E-01 | 0.403 |
| OID20952 | TXLNA | Neurology | TXLNA | P40222 | 1 | 32179675 | 32198285 | 0.87 | 0.74 | 1.03 | 1.14E-01 | 0.403 |
| OID21118 | TMSB10 | Neurology | TMSB10 | P63313 | 2 | 84905656 | 84906671 | 0.87 | 0.73 | 1.03 | 1.14E-01 | 0.404 |
| OID20399 | CHI3L1 | Cardiometabolic | CHI3L1 | P36222 | 1 | 203178931 | 203186704 | 1.13 | 0.97 | 1.31 | 1.15E-01 | 0.405 |
| OID20153 | CXCL8 | Cardiometabolic/Inflamm | CXCL8 | P10145 | 4 | 73740541 | 73743716 | 0.86 | 0.72 | 1.04 | 1.15E-01 | 0.405 |
| OID20966 | CD274 | Neurology | CD274 | Q9NZQ7 | 9 | 5450503 | 5470566 | 1.13 | 0.97 | 1.32 | 1.15E-01 | 0.405 |
| OID30277 | SCN4B | Cardiometabolic_II | SCN4B | Q8IWT1 | 11 | 118133377 | 118152888 | 0.88 | 0.74 | 1.03 | 1.15E-01 | 0.405 |
| OID30941 | PRKAG3 | Neurology_II | PRKAG3 | Q9UGI9 | 2 | 218822308 | 218832086 | 0.87 | 0.73 | 1.04 | 1.15E-01 | 0.405 |
| OID20625 | EIF4G1 | Inflammation | EIF4G1 | Q04637 | 3 | 184314495 | 184335358 | 0.88 | 0.75 | 1.03 | 1.16E-01 | 0.406 |
| OID20576 | IL4R | Inflammation | IL4R | P24394 | 16 | 27313668 | 27364778 | 1.13 | 0.97 | 1.31 | 1.16E-01 | 0.406 |
| OID30463 | RELB | Inflammation_II | RELB | Q01201 | 19 | 45001461 | 45038192 | 0.86 | 0.71 | 1.04 | 1.16E-01 | 0.406 |
| OID30330 | IGSF9 | Cardiometabolic_II | IGSF9 | Q9P2J2 | 1 | 159927039 | 159945613 | 0.87 | 0.72 | 1.04 | 1.16E-01 | 0.406 |
| OID30257 | RBPMS2 | Cardiometabolic_II | RBPMS2 | Q6ZRY4 | 15 | 64739891 | 64775589 | 0.88 | 0.74 | 1.03 | 1.16E-01 | 0.406 |
| OID20828 | FGR | Neurology | FGR | P09769 | 1 | 27612064 | 27635185 | 0.85 | 0.70 | 1.04 | 1.17E-01 | 0.407 |
| OID20240 | LDLR | Cardiometabolic | LDLR | P01130 | 19 | 11089462 | 11133820 | 0.87 | 0.73 | 1.04 | 1.17E-01 | 0.407 |
| OID31152 | BCL2L1 | Neurology_II | BCL2L1 | Q07817 | 20 | 31664452 | 31723989 | 0.88 | 0.75 | 1.03 | 1.17E-01 | 0.407 |
| OID20678 | IFNGR1 | Inflammation | IFNGR1 | P15260 | 6 | 137197485 | 137219449 | 1.13 | 0.97 | 1.32 | 1.17E-01 | 0.407 |
| OID21410 | DPY30 | Oncology | DPY30 | Q9C005 | 2 | 31867809 | 32039805 | 0.87 | 0.72 | 1.04 | 1.18E-01 | 0.409 |
| OID20551 | GOPC | Inflammation | GOPC | Q9HD26 | 6 | 117560269 | 117602542 | 0.88 | 0.75 | 1.03 | 1.18E-01 | 0.410 |
| OID21082 | SIGLEC5 | Neurology | SIGLEC5 | O15389 | 19 | 51611927 | 51645545 | 1.16 | 0.96 | 1.40 | 1.18E-01 | 0.411 |
| OID30198 | NUDT10 | Cardiometabolic_II | NUDT10 | Q8NFP7 | X | 51332231 | 51337525 | 0.85 | 0.69 | 1.04 | 1.19E-01 | 0.411 |
| OID20097 | GYS1 | Cardiometabolic | GYS1 | P13807 | 19 | 48968130 | 48993310 | 0.88 | 0.75 | 1.03 | 1.19E-01 | 0.411 |
| OID30067 | ADRA2A | Cardiometabolic_II | ADRA2A | P08913 | 10 | 111077029 | 111080907 | 0.85 | 0.70 | 1.04 | 1.20E-01 | 0.413 |
| OID20406 | DPP4 | Cardiometabolic | DPP4 | P27487 | 2 | 161992245 | 162074394 | 0.88 | 0.75 | 1.03 | 1.20E-01 | 0.413 |
| OID21465 | HBEGF | Oncology | HBEGF | Q99075 | 5 | 140332843 | 140346603 | 0.88 | 0.75 | 1.03 | 1.20E-01 | 0.413 |
| OID31315 | MTUS1 | Oncology_II | MTUS1 | Q9ULD2 | 8 | 17643795 | 17801094 | 0.87 | 0.73 | 1.04 | 1.20E-01 | 0.413 |
| OID31064 | OTUD7B | Neurology_II | OTUD7B | Q6GQQ9 | 1 | 149937812 | 150010726 | 0.87 | 0.74 | 1.04 | 1.20E-01 | 0.413 |
| OID31313 | EFCAB2 | Oncology_II | EFCAB2 | Q5VUJ9 | 1 | 244969682 | 245127164 | 0.87 | 0.73 | 1.04 | 1.20E-01 | 0.413 |
| OID31172 | JMJD1C | Oncology_II | JMJD1C | Q15652 | 10 | 63167221 | 63521850 | 1.12 | 0.97 | 1.30 | 1.21E-01 | 0.414 |
| OID20607 | CD84 | Inflammation | CD84 | Q9UIB8 | 1 | 160541095 | 160579516 | 0.88 | 0.75 | 1.03 | 1.21E-01 | 0.414 |
| OID21133 | FCER2 | Neurology | FCER2 | P06734 | 19 | 7688758 | 7702146 | 0.88 | 0.75 | 1.03 | 1.22E-01 | 0.417 |
| OID31273 | VPS28 | Oncology_II | VPS28 | Q9UK41 | 8 | 144423601 | 144428563 | 0.87 | 0.74 | 1.04 | 1.22E-01 | 0.417 |

| | | | | | | | | | | | | |
|---|---|---|---|---|---|---|---|---|---|---|---|---|
| OID20865 | ATXN10 | Neurology | ATXN10 | Q9UBB4 | 22 | 45671798 | 45845307 | 0.88 | 0.75 | 1.03 | 1.23E-01 | 0.418 |
| OID21386 | CBLN4 | Oncology | CBLN4 | Q9NTU7 | 20 | 55997357 | 56005519 | 0.88 | 0.75 | 1.03 | 1.23E-01 | 0.418 |
| OID31447 | HDDC2 | Oncology_II | HDDC2 | Q7Z4H3 | 6 | 125219962 | 125302078 | 0.87 | 0.74 | 1.04 | 1.23E-01 | 0.418 |
| OID20863 | NOS1 | Neurology | NOS1 | P29475 | 12 | 117208142 | 117452170 | 1.13 | 0.97 | 1.33 | 1.23E-01 | 0.418 |
| OID30427 | UBE2Z | Inflammation_II | UBE2Z | Q9H832 | 17 | 48908407 | 48929056 | 0.86 | 0.71 | 1.04 | 1.23E-01 | 0.418 |
| OID30964 | UROS | Neurology_II | UROS | P10746 | 10 | 125784980 | 125823258 | 0.86 | 0.72 | 1.04 | 1.23E-01 | 0.418 |
| OID31084 | USP28 | Neurology_II | USP28 | Q96RU2 | 11 | 113797874 | 113875572 | 0.85 | 0.70 | 1.04 | 1.23E-01 | 0.418 |
| OID30698 | C9 | Inflammation_II | C9 | P02748 | 5 | 39284140 | 39371324 | 1.14 | 0.97 | 1.34 | 1.24E-01 | 0.419 |
| OID31020 | LRTM2 | Neurology_II | LRTM2 | Q8N967 | 12 | 1820267 | 1836753 | 0.87 | 0.73 | 1.04 | 1.24E-01 | 0.420 |
| OID30181 | YOD1 | Cardiometabolic_II | YOD1 | Q5VVQ6 | 1 | 207043849 | 207052980 | 0.88 | 0.74 | 1.04 | 1.24E-01 | 0.420 |
| OID21387 | CALCOCO1 | Oncology | CALCOCO1 | Q9P1Z2 | 12 | 53708517 | 53727745 | 0.88 | 0.75 | 1.04 | 1.25E-01 | 0.422 |
| OID21175 | HGS | Oncology | HGS | O14964 | 17 | 81683326 | 81703138 | 0.88 | 0.74 | 1.04 | 1.25E-01 | 0.422 |
| OID21339 | KAZALD1 | Oncology | KAZALD1 | Q96I82 | 10 | 101061989 | 101068131 | 1.14 | 0.96 | 1.34 | 1.25E-01 | 0.422 |
| OID20112 | ITGB1BP2 | Cardiometabolic | ITGB1BP2 | Q9UKP3 | X | 71301750 | 71305371 | 0.88 | 0.76 | 1.04 | 1.26E-01 | 0.424 |
| OID30923 | TBR1 | Neurology_II | TBR1 | Q16650 | 2 | 161416297 | 161425870 | 0.85 | 0.69 | 1.05 | 1.26E-01 | 0.424 |
| OID21331 | ARSB | Oncology | ARSB | P15848 | 5 | 78777209 | 78986087 | 0.87 | 0.73 | 1.04 | 1.26E-01 | 0.424 |
| OID20177 | CD2AP | Cardiometabolic | CD2AP | Q9Y5K6 | 6 | 47477789 | 47627263 | 0.88 | 0.75 | 1.04 | 1.27E-01 | 0.424 |
| OID20524 | DAPP1 | Inflammation | DAPP1 | Q9UN19 | 4 | 99816827 | 99870190 | 0.89 | 0.76 | 1.03 | 1.27E-01 | 0.424 |
| OID21371 | GALNT10 | Oncology | GALNT10 | Q86SR1 | 5 | 154190730 | 154420984 | 0.88 | 0.74 | 1.04 | 1.27E-01 | 0.424 |
| OID31396 | LAMTOR5 | Oncology_II | LAMTOR5 | O43504 | 1 | 110401249 | 110407942 | 0.86 | 0.72 | 1.04 | 1.27E-01 | 0.424 |
| OID30321 | UBE2L6 | Cardiometabolic_II | UBE2L6 | O14933 | 11 | 57551656 | 57568284 | 0.88 | 0.74 | 1.04 | 1.27E-01 | 0.424 |
| OID31439 | SH3BP1 | Oncology_II | SH3BP1 | Q9Y3L3 | 22 | 37634654 | 37656117 | 0.86 | 0.70 | 1.04 | 1.27E-01 | 0.424 |
| OID20380 | CD46 | Cardiometabolic | CD46 | P15529 | 1 | 207752054 | 207795513 | 0.88 | 0.74 | 1.04 | 1.28E-01 | 0.425 |
| OID20084 | CEP43 | Cardiometabolic | CEP43 | O95684 | 6 | 166999317 | 167094789 | 0.88 | 0.75 | 1.04 | 1.28E-01 | 0.425 |
| OID21334 | DCTN2 | Oncology | DCTN2 | Q13561 | 12 | 57529633 | 57547224 | 0.88 | 0.74 | 1.04 | 1.28E-01 | 0.425 |
| OID30182 | PALM3 | Cardiometabolic_II | PALM3 | A6NDB9 | 19 | 14053363 | 14062076 | 0.85 | 0.69 | 1.05 | 1.28E-01 | 0.425 |
| OID31195 | SMC3 | Oncology_II | SMC3 | Q9UQE7 | 10 | 110567684 | 110606048 | 0.86 | 0.71 | 1.04 | 1.28E-01 | 0.425 |
| OID30466 | BABAM1 | Inflammation_II | BABAM1 | Q9NWV8 | 19 | 17267376 | 17281249 | 0.87 | 0.72 | 1.04 | 1.29E-01 | 0.427 |
| OID30438 | EP300 | Inflammation_II | EP300 | Q09472 | 22 | 41092592 | 41180077 | 0.87 | 0.72 | 1.04 | 1.30E-01 | 0.427 |
| OID30722 | ITIH1 | Inflammation_II | ITIH1 | P19827 | 3 | 52777595 | 52792068 | 0.88 | 0.74 | 1.04 | 1.30E-01 | 0.427 |
| OID21185 | KIR3DL1 | Oncology | KIR3DL1 | P43629 | 19 | 54816468 | 54830778 | 0.88 | 0.75 | 1.04 | 1.30E-01 | 0.427 |
| OID21236 | MED18 | Oncology | MED18 | Q9BUE0 | 1 | 28329002 | 28335965 | 0.88 | 0.75 | 1.04 | 1.29E-01 | 0.427 |
| OID30451 | NME1 | Inflammation_II | NME1 | P15531 | 17 | 51153559 | 51162428 | 0.87 | 0.73 | 1.04 | 1.30E-01 | 0.427 |
| OID21360 | OGFR | Oncology | OGFR | Q9NZT2 | 20 | 62804835 | 62814000 | 0.88 | 0.74 | 1.04 | 1.29E-01 | 0.427 |
| OID31425 | STAM | Oncology_II | STAM | Q92783 | 10 | 17644151 | 17716824 | 0.87 | 0.72 | 1.04 | 1.30E-01 | 0.427 |
| OID31002 | VWC2L | Neurology_II | VWC2L | B2RUY7 | 2 | 214411054 | 214578976 | 0.85 | 0.69 | 1.05 | 1.29E-01 | 0.427 |
| OID30748 | ADIPOQ | Inflammation_II | ADIPOQ | Q15848 | 3 | 186842704 | 186858463 | 1.15 | 0.96 | 1.39 | 1.31E-01 | 0.430 |
| OID30140 | PRKD2 | Cardiometabolic_II | PRKD2 | Q9BZL6 | 19 | 46674275 | 46717127 | 0.85 | 0.70 | 1.05 | 1.32E-01 | 0.431 |
| OID30178 | MRPL24 | Cardiometabolic_II | MRPL24 | Q96A35 | 1 | 156737303 | 156741590 | 0.86 | 0.71 | 1.05 | 1.32E-01 | 0.431 |
| OID21280 | ABL1 | Oncology | ABL1 | P00519 | 9 | 130713016 | 130887675 | 0.88 | 0.74 | 1.04 | 1.32E-01 | 0.432 |
| OID30271 | NAGK | Cardiometabolic_II | NAGK | Q9UJ70 | 2 | 71064344 | 71079808 | 0.87 | 0.73 | 1.04 | 1.33E-01 | 0.432 |
| OID30533 | GIT1 | Inflammation_II | GIT1 | Q9Y2X7 | 17 | 29573475 | 29594054 | 0.88 | 0.75 | 1.04 | 1.33E-01 | 0.433 |
| OID20061 | CTF1 | Cardiometabolic | CTF1 | Q16619 | 16 | 30896614 | 30903547 | 0.88 | 0.74 | 1.04 | 1.33E-01 | 0.433 |
| OID20322 | SPP1 | Cardiometabolic | SPP1 | P10451 | 4 | 87975667 | 87983532 | 1.13 | 0.96 | 1.32 | 1.33E-01 | 0.433 |
| OID30436 | HDAC8 | Inflammation_II | HDAC8 | Q9BY41 | X | 72329516 | 72573101 | 1.11 | 0.97 | 1.28 | 1.34E-01 | 0.434 |
| OID21294 | ADGRG1 | Oncology | ADGRG1 | Q9Y653 | 16 | 57610652 | 57665580 | 1.11 | 0.97 | 1.28 | 1.34E-01 | 0.435 |
| OID20139 | AGXT | Cardiometabolic | AGXT | P21549 | 2 | 240868824 | 240880502 | 0.87 | 0.73 | 1.04 | 1.34E-01 | 0.435 |
| OID30895 | LRFN2 | Neurology_II | LRFN2 | Q9ULH4 | 6 | 40391591 | 40587364 | 0.86 | 0.70 | 1.05 | 1.34E-01 | 0.435 |
| OID20502 | SRPK2 | Inflammation | SRPK2 | P78362 | 7 | 105110704 | 105399308 | 0.87 | 0.73 | 1.04 | 1.35E-01 | 0.435 |
| OID21026 | VCAN | Neurology | VCAN | P13611 | 5 | 83471618 | 83582303 | 1.13 | 0.96 | 1.32 | 1.35E-01 | 0.435 |
| OID30306 | MAN2B2 | Cardiometabolic_II | MAN2B2 | Q9Y2E5 | 4 | 6575189 | 6623362 | 0.88 | 0.75 | 1.04 | 1.35E-01 | 0.435 |
| OID30437 | PTPN9 | Inflammation_II | PTPN9 | P43378 | 15 | 75463251 | 75579315 | 0.86 | 0.71 | 1.05 | 1.35E-01 | 0.436 |
| OID20690 | SIGLEC1 | Inflammation | SIGLEC1 | Q9BZZ2 | 20 | 3686970 | 3712600 | 1.13 | 0.96 | 1.33 | 1.35E-01 | 0.436 |
| OID30848 | TFAP2A | Neurology_II | TFAP2A | P05549 | 6 | 10393186 | 10419659 | 1.12 | 0.97 | 1.29 | 1.36E-01 | 0.436 |
| OID20787 | TFF2 | Inflammation | TFF2 | Q03403 | 21 | 42346357 | 42350997 | 1.13 | 0.96 | 1.32 | 1.36E-01 | 0.436 |
| OID30847 | BRME1 | Neurology_II | BRME1 | Q0VDD7 | 19 | 13882348 | 13906452 | 1.08 | 0.98 | 1.20 | 1.36E-01 | 0.436 |
| OID30274 | MECR | Cardiometabolic_II | MECR | Q9BV79 | 1 | 29192657 | 29230942 | 0.88 | 0.75 | 1.04 | 1.36E-01 | 0.436 |
| OID21475 | PPP1R12A | Oncology | PPP1R12A | O14974 | 12 | 79773563 | 79935460 | 0.88 | 0.75 | 1.04 | 1.36E-01 | 0.437 |
| OID20078 | TSPAN1 | Cardiometabolic | TSPAN1 | O60635 | 1 | 46175073 | 46185962 | 0.87 | 0.73 | 1.04 | 1.36E-01 | 0.437 |
| OID20199 | AHCY | Cardiometabolic | AHCY | P23526 | 20 | 34280268 | 34311802 | 0.88 | 0.75 | 1.04 | 1.38E-01 | 0.438 |
| OID31150 | BAP18 | Neurology_II | C17orf49 | Q8IXM2 | 17 | 7014495 | 7017525 | 0.86 | 0.71 | 1.05 | 1.39E-01 | 0.438 |
| OID30097 | CD2 | Cardiometabolic_II | CD2 | P06729 | 1 | 116754430 | 116769229 | 0.87 | 0.72 | 1.05 | 1.39E-01 | 0.438 |
| OID30821 | CEND1 | Neurology_II | CEND1 | Q8N111 | 11 | 787115 | 790113 | 1.09 | 0.97 | 1.22 | 1.39E-01 | 0.438 |
| OID20617 | CKAP4 | Inflammation | CKAP4 | Q07065 | 12 | 106237881 | 106304279 | 1.12 | 0.96 | 1.31 | 1.38E-01 | 0.438 |
| OID30667 | CSH1 | Inflammation_II | CSH1 | P0DML2 | 17 | 63894909 | 63896661 | 0.87 | 0.72 | 1.05 | 1.39E-01 | 0.438 |
| OID31479 | CWC15 | Oncology_II | CWC15 | Q9P013 | 11 | 94962620 | 94973586 | 0.86 | 0.69 | 1.05 | 1.39E-01 | 0.438 |
| OID30736 | ECM1 | Inflammation_II | ECM1 | Q16610 | 1 | 150508062 | 150513789 | 0.89 | 0.77 | 1.04 | 1.38E-01 | 0.438 |
| OID30539 | ERMAP | Inflammation_II | ERMAP | Q96PL5 | 1 | 42817122 | 42844991 | 0.88 | 0.74 | 1.04 | 1.39E-01 | 0.438 |
| OID31305 | MTHFSD | Oncology_II | MTHFSD | Q2M296 | 16 | 86530178 | 86555235 | 0.88 | 0.75 | 1.04 | 1.39E-01 | 0.438 |
| OID31412 | NUBP1 | Oncology_II | NUBP1 | P53384 | 16 | 10743786 | 10769351 | 0.87 | 0.73 | 1.05 | 1.39E-01 | 0.438 |
| OID30998 | PAFAH1B3 | Neurology_II | PAFAH1B3 | Q15102 | 19 | 42297033 | 42303546 | 0.87 | 0.72 | 1.05 | 1.38E-01 | 0.438 |
| OID30469 | PTP4A3 | Inflammation_II | PTP4A3 | O75365 | 8 | 141391995 | 141432454 | 0.87 | 0.73 | 1.04 | 1.38E-01 | 0.438 |
| OID30122 | RAB33A | Cardiometabolic_II | RAB33A | Q14088 | X | 130171962 | 130184870 | 0.88 | 0.74 | 1.04 | 1.39E-01 | 0.438 |
| OID20826 | RASA1 | Neurology | RASA1 | P20936 | 5 | 87267883 | 87391931 | 0.88 | 0.74 | 1.04 | 1.39E-01 | 0.438 |
| OID21348 | YES1 | Oncology | YES1 | P07947 | 18 | 721588 | 812546 | 0.88 | 0.75 | 1.04 | 1.39E-01 | 0.438 |
| OID30332 | FCAMR | Cardiometabolic_II | FCAMR | Q8WWV6 | 1 | 206957965 | 206970625 | 0.87 | 0.73 | 1.04 | 1.40E-01 | 0.439 |
| OID30219 | LONP1 | Cardiometabolic_II | LONP1 | P36776 | 19 | 5691834 | 5720572 | 0.88 | 0.75 | 1.04 | 1.40E-01 | 0.439 |
| OID21004 | CCL2 | Neurology | CCL2 | P13500 | 17 | 34255218 | 34257203 | 0.88 | 0.74 | 1.04 | 1.41E-01 | 0.439 |
| OID21008 | CDH3 | Neurology | CDH3 | P22223 | 16 | 68636189 | 68727468 | 1.12 | 0.96 | 1.30 | 1.41E-01 | 0.439 |
| OID21113 | DSG2 | Neurology | DSG2 | Q14126 | 18 | 31498177 | 31549008 | 1.12 | 0.96 | 1.31 | 1.41E-01 | 0.439 |
| OID30839 | FH | Neurology_II | FH | P07954 | 1 | 241497511 | 241519799 | 0.87 | 0.73 | 1.05 | 1.41E-01 | 0.439 |
| OID21059 | LRPAP1 | Neurology | LRPAP1 | P30533 | 4 | 3503612 | 3532446 | 0.87 | 0.73 | 1.05 | 1.41E-01 | 0.439 |
| OID30301 | ELN | Cardiometabolic_II | ELN | P15502 | 7 | 74027789 | 74069907 | 1.16 | 0.95 | 1.40 | 1.41E-01 | 0.440 |
| OID30503 | NEDD9 | Inflammation_II | NEDD9 | Q14511 | 6 | 11183298 | 11382348 | 0.87 | 0.72 | 1.05 | 1.41E-01 | 0.440 |
| OID30807 | CNTF | Neurology_II | CNTF | P26441 | 11 | 58622665 | 58625733 | 0.88 | 0.75 | 1.04 | 1.42E-01 | 0.441 |
| OID21087 | PEBP1 | Neurology | PEBP1 | P30086 | 12 | 118136124 | 118145584 | 0.88 | 0.74 | 1.04 | 1.42E-01 | 0.441 |
| OID20726 | ANGPTL2 | Inflammation | ANGPTL2 | Q9UKU9 | 9 | 127087348 | 127122635 | 1.13 | 0.96 | 1.34 | 1.42E-01 | 0.441 |
| OID20618 | FKBP1B | Inflammation | FKBP1B | P68106 | 2 | 24049701 | 24063681 | 0.88 | 0.75 | 1.04 | 1.42E-01 | 0.441 |
| OID20525 | EDAR | Inflammation | EDAR | Q9UNE0 | 2 | 108894471 | 108989372 | 0.88 | 0.75 | 1.04 | 1.43E-01 | 0.442 |
| OID30938 | PFDN6 | Neurology_II | PFDN6 | O15212 | 6 | 33289302 | 33298401 | 0.87 | 0.72 | 1.05 | 1.43E-01 | 0.442 |
| OID21414 | CASP8 | Oncology | CASP8 | Q14790 | 2 | 201233443 | 201287711 | 0.88 | 0.73 | 1.05 | 1.43E-01 | 0.443 |
| OID30608 | LEG1 | Inflammation_II | C6orf58 | Q6P5S2 | 6 | 127519455 | 127591820 | 0.88 | 0.74 | 1.04 | 1.43E-01 | 0.443 |
| OID21268 | ITGB1BP1 | Oncology | ITGB1BP1 | O14713 | 2 | 9403475 | 9423528 | 1.13 | 0.96 | 1.33 | 1.44E-01 | 0.443 |
| OID31164 | CDK1 | Oncology_II | CDK1 | P06493 | 10 | 60778331 | 60794852 | 1.13 | 0.96 | 1.33 | 1.44E-01 | 0.444 |
| OID31420 | MPRIP | Oncology_II | MPRIP | Q6WCQ1 | 17 | 17042457 | 17217679 | 0.88 | 0.74 | 1.04 | 1.44E-01 | 0.444 |
| OID31270 | PRUNE2 | Oncology_II | PRUNE2 | Q8WUY3 | 9 | 76611376 | 76906114 | 0.87 | 0.72 | 1.05 | 1.44E-01 | 0.444 |
| OID30711 | VNN1 | Inflammation_II | VNN1 | O95497 | 6 | 132680849 | 132714055 | 0.89 | 0.76 | 1.04 | 1.44E-01 | 0.444 |
| OID30875 | CDH4 | Neurology_II | CDH4 | P55283 | 20 | 61252261 | 61940617 | 1.11 | 0.96 | 1.28 | 1.45E-01 | 0.445 |
| OID31072 | DHRS4L2 | Neurology_II | DHRS4L2 | Q6PKH6 | 14 | 23969874 | 24006408 | 0.87 | 0.72 | 1.05 | 1.45E-01 | 0.445 |
| OID20459 | MVK | Inflammation | MVK | Q03426 | 12 | 109573255 | 109598125 | 0.88 | 0.74 | 1.05 | 1.45E-01 | 0.445 |
| OID30080 | ACSL1 | Cardiometabolic_II | ACSL1 | P33121 | 4 | 184755595 | 184826818 | 0.86 | 0.70 | 1.05 | 1.47E-01 | 0.447 |
| OID30396 | COL15A1 | Cardiometabolic_II | COL15A1 | P39059 | 9 | 98943179 | 99070792 | 0.89 | 0.76 | 1.04 | 1.46E-01 | 0.447 |
| OID30583 | MXRA8 | Inflammation_II | MXRA8 | Q9BRK3 | 1 | 1352689 | 1361777 | 0.88 | 0.73 | 1.05 | 1.47E-01 | 0.447 |
| OID30989 | MYO6 | Neurology_II | MYO6 | Q9UM54 | 6 | 75749201 | 75919537 | 0.86 | 0.71 | 1.05 | 1.47E-01 | 0.447 |
| OID20637 | CD22 | Inflammation | CD22 | P20273 | 19 | 35319261 | 35347361 | 0.89 | 0.76 | 1.04 | 1.48E-01 | 0.448 |
| OID30286 | COL2A1 | Cardiometabolic_II | COL2A1 | P02458 | 12 | 47972967 | 48004554 | 0.89 | 0.75 | 1.04 | 1.47E-01 | 0.448 |
| OID31358 | GPD1 | Oncology_II | GPD1 | P21695 | 12 | 50103982 | 50111313 | 0.87 | 0.72 | 1.05 | 1.47E-01 | 0.448 |

| | | | | | | | | | | | | |
|---|---|---|---|---|---|---|---|---|---|---|---|---|
| OID31160 | MORF4L2 | Oncology_II | MORF4L2 | Q15014 | X | 103675496 | 103688158 | 0.87 | 0.72 | 1.05 | 1.48E-01 | 0.448 |
| OID30139 | GBP6 | Cardiometabolic_II | GBP6 | Q6ZN66 | 1 | 89364059 | 89388160 | 0.85 | 0.69 | 1.06 | 1.48E-01 | 0.448 |
| OID20921 | ABHD14B | Neurology | ABHD14B | Q96IU4 | 3 | 51968510 | 51983409 | 0.89 | 0.75 | 1.05 | 1.50E-01 | 0.451 |
| OID20137 | ACY1 | Cardiometabolic | ACY1 | Q03154 | 3 | 51983340 | 51989197 | 0.87 | 0.73 | 1.05 | 1.50E-01 | 0.451 |
| OID30355 | ADAMTSL5 | Cardiometabolic_II | ADAMTSL5 | Q6ZMM2 | 19 | 1505022 | 1513604 | 0.89 | 0.75 | 1.04 | 1.49E-01 | 0.451 |
| OID21282 | CDHR2 | Oncology | CDHR2 | Q9BYE9 | 5 | 176542511 | 176595974 | 0.86 | 0.70 | 1.05 | 1.50E-01 | 0.451 |
| OID30602 | DDI2 | Inflammation_II | DDI2 | Q5TDH0 | 1 | 15617458 | 15669044 | 0.88 | 0.74 | 1.05 | 1.50E-01 | 0.451 |
| OID21411 | FGF23 | Oncology | FGF23 | Q9GZV9 | 12 | 4368227 | 4379712 | 1.12 | 0.96 | 1.32 | 1.49E-01 | 0.451 |
| OID21216 | GFOD2 | Oncology | GFOD2 | Q3B7J2 | 16 | 67674531 | 67719339 | 1.11 | 0.96 | 1.29 | 1.50E-01 | 0.451 |
| OID21264 | LYAR | Oncology | LYAR | Q9NX58 | 4 | 4267701 | 4290154 | 0.85 | 0.69 | 1.06 | 1.50E-01 | 0.451 |
| OID31354 | NENF | Oncology_II | NENF | Q9UMX5 | 1 | 212432920 | 212446379 | 0.87 | 0.73 | 1.05 | 1.50E-01 | 0.451 |
| OID20058 | TCL1B | Cardiometabolic | TCL1B | O95988 | 14 | 95686426 | 95692628 | 0.87 | 0.72 | 1.05 | 1.50E-01 | 0.451 |
| OID31042 | BRSK2 | Neurology_II | BRSK2 | Q8IWQ3 | 11 | 1389899 | 1462689 | 0.88 | 0.74 | 1.05 | 1.51E-01 | 0.451 |
| OID21451 | EDA2R | Oncology | EDA2R | Q9HAV5 | X | 66595637 | 66639298 | 1.15 | 0.95 | 1.38 | 1.51E-01 | 0.451 |
| OID30263 | ANXA2 | Cardiometabolic_II | ANXA2 | P07355 | 15 | 60347134 | 60402883 | 0.88 | 0.74 | 1.05 | 1.51E-01 | 0.452 |
| OID31511 | GPR15L | Oncology_II | C10orf99 | Q6UWK7 | 10 | 84173801 | 84185294 | 1.13 | 0.96 | 1.34 | 1.51E-01 | 0.452 |
| OID20108 | PAG1 | Cardiometabolic | PAG1 | Q9NWQ8 | 8 | 80967810 | 81112068 | 0.88 | 0.74 | 1.05 | 1.51E-01 | 0.452 |
| OID30194 | PNMA1 | Cardiometabolic_II | PNMA1 | Q8ND90 | 14 | 73711783 | 73714384 | 0.88 | 0.75 | 1.05 | 1.51E-01 | 0.452 |
| OID31355 | ENSA | Oncology_II | ENSA | O43768 | 1 | 150600851 | 150629612 | 0.88 | 0.74 | 1.05 | 1.52E-01 | 0.452 |
| OID20453 | IL20 | Inflammation | IL20 | Q9NYY1 | 1 | 206865623 | 206869223 | 0.87 | 0.72 | 1.05 | 1.52E-01 | 0.452 |
| OID31034 | RBM17 | Neurology_II | RBM17 | Q96I25 | 10 | 6089034 | 6117457 | 0.88 | 0.75 | 1.05 | 1.52E-01 | 0.452 |
| OID20689 | ENPP7 | Inflammation | ENPP7 | Q6UWV6 | 17 | 79730943 | 79742219 | 1.13 | 0.96 | 1.32 | 1.53E-01 | 0.453 |
| OID30309 | MAMDC2 | Cardiometabolic_II | MAMDC2 | Q7Z304 | 9 | 70043848 | 70226972 | 1.12 | 0.96 | 1.30 | 1.53E-01 | 0.453 |
| OID30086 | ACY3 | Cardiometabolic_II | ACY3 | Q96HD9 | 11 | 67642555 | 67650730 | 0.88 | 0.74 | 1.05 | 1.58E-01 | 0.456 |
| OID20394 | ADA2 | Cardiometabolic | ADA2 | Q9NZK5 | 22 | 17178790 | 17258235 | 1.12 | 0.96 | 1.31 | 1.55E-01 | 0.456 |
| OID31047 | ALDH5A1 | Neurology_II | ALDH5A1 | P51649 | 6 | 24494867 | 24537207 | 0.88 | 0.74 | 1.05 | 1.58E-01 | 0.456 |
| OID21412 | ANGPTL7 | Oncology | ANGPTL7 | O43827 | 1 | 11189355 | 11195981 | 1.12 | 0.96 | 1.30 | 1.54E-01 | 0.456 |
| OID30267 | BOLA2_BOLA2B | Cardiometabolic_II | BOLA2B;BOLA2 | Q9H3K6;Q9H3K6 | 16;16 | 30192932;29453590 | 30194306;29454351 | 0.89 | 0.75 | 1.05 | 1.57E-01 | 0.456 |
| OID31196 | BTNL10 | Oncology_II | BTNL10 | A8MVZ5 | 1 | 228510425 | 228512305 | 0.87 | 0.71 | 1.06 | 1.57E-01 | 0.456 |
| OID30851 | CBLN1 | Neurology_II | CBLN1 | P23435 | 16 | 49277917 | 49281838 | 0.87 | 0.71 | 1.06 | 1.58E-01 | 0.456 |
| OID21295 | CDKN2D | Oncology | CDKN2D | P55273 | 19 | 10566460 | 10569059 | 0.89 | 0.76 | 1.04 | 1.55E-01 | 0.456 |
| OID21245 | CEACAM3 | Oncology | CEACAM3 | P40198 | 19 | 41796587 | 41811554 | 1.10 | 0.96 | 1.26 | 1.57E-01 | 0.456 |
| OID30549 | DBN1 | Inflammation_II | DBN1 | Q16643 | 5 | 177456608 | 177474401 | 0.88 | 0.74 | 1.05 | 1.58E-01 | 0.456 |
| OID30247 | DNAJC6 | Cardiometabolic_II | DNAJC6 | O75061 | 1 | 65248219 | 65415871 | 0.89 | 0.75 | 1.05 | 1.59E-01 | 0.456 |
| OID20722 | FIS1 | Inflammation | FIS1 | Q9Y3D6 | 7 | 101239458 | 101252316 | 0.88 | 0.75 | 1.05 | 1.55E-01 | 0.456 |
| OID20941 | IFNGR2 | Neurology | IFNGR2 | P38484 | 21 | 33403413 | 33479348 | 0.88 | 0.74 | 1.05 | 1.55E-01 | 0.456 |
| OID31059 | IGBP1 | Neurology_II | IGBP1 | P78318 | X | 70133447 | 70166324 | 0.89 | 0.75 | 1.05 | 1.58E-01 | 0.456 |
| OID20585 | IL17RB | Inflammation | IL17RB | Q9NRM6 | 3 | 53846568 | 53865794 | 1.13 | 0.95 | 1.33 | 1.58E-01 | 0.456 |
| OID20147 | IL6ST | Cardiometabolic | IL6ST | P40189 | 5 | 55935095 | 55995022 | 1.12 | 0.96 | 1.31 | 1.56E-01 | 0.456 |
| OID21345 | INPP1 | Oncology | INPP1 | P49441 | 2 | 190343570 | 190371665 | 0.88 | 0.74 | 1.05 | 1.57E-01 | 0.456 |
| OID30216 | ITPR1 | Cardiometabolic_II | ITPR1 | Q14643 | 3 | 4493345 | 4847506 | 0.88 | 0.73 | 1.05 | 1.56E-01 | 0.456 |
| OID20833 | IVD | Neurology | IVD | P26440 | 15 | 40405485 | 40435947 | 0.89 | 0.76 | 1.05 | 1.59E-01 | 0.456 |
| OID21518 | KLK11 | Oncology | KLK11 | Q9UBX7 | 19 | 51022216 | 51028039 | 0.89 | 0.76 | 1.05 | 1.57E-01 | 0.456 |
| OID30865 | KRT8 | Neurology_II | KRT8 | P05787 | 12 | 52897187 | 52949954 | 1.10 | 0.96 | 1.26 | 1.55E-01 | 0.456 |
| OID30314 | L3HYPDH | Cardiometabolic_II | L3HYPDH | Q96EM0 | 14 | 59460363 | 59484408 | 0.89 | 0.75 | 1.05 | 1.55E-01 | 0.456 |
| OID21034 | LBR | Neurology | LBR | Q14739 | 1 | 225401502 | 225428925 | 0.87 | 0.72 | 1.05 | 1.56E-01 | 0.456 |
| OID30266 | LIPF | Cardiometabolic_II | LIPF | P07098 | 10 | 88664441 | 88678814 | 0.89 | 0.77 | 1.04 | 1.57E-01 | 0.456 |
| OID30227 | OXCT1 | Cardiometabolic_II | OXCT1 | P55809 | 5 | 41730065 | 41870425 | 0.88 | 0.74 | 1.05 | 1.57E-01 | 0.456 |
| OID31456 | PCDH7 | Oncology_II | PCDH7 | O60245 | 4 | 30720369 | 31146805 | 0.87 | 0.72 | 1.05 | 1.59E-01 | 0.456 |
| OID30228 | PECR | Cardiometabolic_II | PECR | Q9BY49 | 2 | 215996329 | 216082955 | 0.88 | 0.73 | 1.05 | 1.57E-01 | 0.456 |
| OID30258 | RANBP1 | Cardiometabolic_II | RANBP1 | P43487 | 22 | 20115938 | 20127355 | 0.89 | 0.75 | 1.05 | 1.57E-01 | 0.456 |
| OID20201 | REN | Cardiometabolic | REN | P00797 | 1 | 204154819 | 204190324 | 1.12 | 0.96 | 1.31 | 1.56E-01 | 0.456 |
| OID31297 | RGL2 | Oncology_II | RGL2 | O15211 | 6 | 33291654 | 33298942 | 0.86 | 0.71 | 1.06 | 1.55E-01 | 0.456 |
| OID21174 | RRM2B | Oncology | RRM2B | Q7LG56 | 8 | 102204502 | 102238961 | 0.89 | 0.75 | 1.05 | 1.57E-01 | 0.456 |
| OID30774 | SERPINA4 | Inflammation_II | SERPINA4 | P29622 | 14 | 94561442 | 94569913 | 0.89 | 0.75 | 1.05 | 1.59E-01 | 0.456 |
| OID20462 | SIT1 | Inflammation | SIT1 | Q9Y3P8 | 9 | 35649295 | 35650931 | 0.89 | 0.75 | 1.05 | 1.59E-01 | 0.456 |
| OID31238 | THAP12 | Oncology_II | THAP12 | O43422 | 11 | 76349898 | 76381132 | 0.87 | 0.71 | 1.06 | 1.59E-01 | 0.456 |
| OID30476 | THSD1 | Inflammation_II | THSD1 | Q9NS62 | 13 | 52377167 | 52416373 | 1.12 | 0.96 | 1.32 | 1.57E-01 | 0.456 |
| OID20657 | MERTK | Inflammation | MERTK | Q12866 | 2 | 111898607 | 112029561 | 1.12 | 0.96 | 1.32 | 1.59E-01 | 0.456 |
| OID20874 | CLSTN1 | Neurology | CLSTN1 | O94985 | 1 | 9728926 | 9823984 | 0.88 | 0.73 | 1.05 | 1.60E-01 | 0.456 |
| OID30190 | LPP | Cardiometabolic_II | LPP | Q93052 | 3 | 188153284 | 188890671 | 0.89 | 0.75 | 1.05 | 1.60E-01 | 0.457 |
| OID21502 | ADAMTS8 | Oncology | ADAMTS8 | Q9UP79 | 11 | 130404923 | 130428609 | 0.89 | 0.75 | 1.05 | 1.60E-01 | 0.458 |
| OID20907 | IMPA1 | Neurology | IMPA1 | P29218 | 8 | 81656914 | 81686331 | 0.89 | 0.75 | 1.05 | 1.60E-01 | 0.458 |
| OID31513 | MTIF3 | Oncology_II | MTIF3 | Q9H2K0 | 13 | 27435643 | 27450591 | 0.89 | 0.75 | 1.05 | 1.61E-01 | 0.458 |
| OID30092 | KRT17 | Cardiometabolic_II | KRT17 | Q04695 | 17 | 41619442 | 41624842 | 1.12 | 0.96 | 1.31 | 1.61E-01 | 0.459 |
| OID31043 | DDHD2 | Neurology_II | DDHD2 | O94830 | 8 | 38225218 | 38275558 | 0.89 | 0.75 | 1.05 | 1.63E-01 | 0.461 |
| OID30147 | FDX2 | Cardiometabolic_II | FDX2 | Q6P4F2 | 19 | 10310045 | 10316015 | 0.88 | 0.74 | 1.05 | 1.63E-01 | 0.461 |
| OID20769 | LAMA4 | Inflammation | LAMA4 | Q16363 | 6 | 112107931 | 112254939 | 0.89 | 0.76 | 1.05 | 1.62E-01 | 0.461 |
| OID30950 | LYSMD3 | Neurology_II | LYSMD3 | Q7Z3D4 | 5 | 90515611 | 90529584 | 0.88 | 0.73 | 1.05 | 1.63E-01 | 0.461 |
| OID30689 | RNASE6 | Inflammation_II | RNASE6 | Q93091 | 14 | 20781268 | 20782467 | 1.10 | 0.96 | 1.27 | 1.63E-01 | 0.461 |
| OID30987 | S100A14 | Neurology_II | S100A14 | Q9HCY8 | 1 | 153614255 | 153616986 | 0.89 | 0.75 | 1.05 | 1.62E-01 | 0.461 |
| OID21281 | VPS53 | Oncology | VPS53 | Q5VIR6 | 17 | 508503 | 721717 | 0.89 | 0.75 | 1.05 | 1.62E-01 | 0.461 |
| OID20115 | MSTN | Cardiometabolic | MSTN | O14793 | 2 | 190055700 | 190062729 | 0.88 | 0.74 | 1.05 | 1.64E-01 | 0.462 |
| OID30796 | CDKL5 | Neurology_II | CDKL5 | O76039 | X | 18425583 | 18653629 | 0.87 | 0.72 | 1.06 | 1.64E-01 | 0.462 |
| OID21330 | GFRA2 | Oncology | GFRA2 | O00451 | 8 | 21690398 | 21812357 | 1.12 | 0.95 | 1.31 | 1.64E-01 | 0.462 |
| OID30164 | GSTM4 | Cardiometabolic_II | GSTM4 | Q03013 | 1 | 109656099 | 109665496 | 0.87 | 0.71 | 1.06 | 1.64E-01 | 0.462 |
| OID21021 | PDCD5 | Neurology | PDCD5 | O14737 | 19 | 32581190 | 32587453 | 0.89 | 0.75 | 1.05 | 1.65E-01 | 0.462 |
| OID30108 | TTN | Cardiometabolic_II | TTN | Q8WZ42 | 2 | 178525989 | 178830802 | 0.88 | 0.73 | 1.06 | 1.64E-01 | 0.462 |
| OID20401 | CCL14 | Cardiometabolic | CCL14 | Q16627 | 17 | 35983288 | 35987004 | 0.89 | 0.76 | 1.05 | 1.65E-01 | 0.464 |
| OID30779 | SERPINF2 | Inflammation_II | SERPINF2 | P08697 | 17 | 1742836 | 1755265 | 0.88 | 0.73 | 1.05 | 1.65E-01 | 0.464 |
| OID31380 | ALDH2 | Oncology_II | ALDH2 | P05091 | 12 | 111766887 | 111817532 | 0.88 | 0.73 | 1.06 | 1.66E-01 | 0.465 |
| OID20641 | GMPR | Inflammation | GMPR | P36959 | 6 | 16238587 | 16295549 | 0.89 | 0.76 | 1.05 | 1.66E-01 | 0.465 |
| OID21420 | PVALB | Oncology | PVALB | P20472 | 22 | 36800684 | 36819479 | 0.89 | 0.76 | 1.05 | 1.66E-01 | 0.465 |
| OID30388 | TREH | Cardiometabolic_II | TREH | O43280 | 11 | 118657316 | 118679690 | 0.89 | 0.76 | 1.05 | 1.66E-01 | 0.465 |
| OID30161 | ANK2 | Cardiometabolic_II | ANK2 | Q01484 | 4 | 112818032 | 113384221 | 1.11 | 0.96 | 1.28 | 1.67E-01 | 0.465 |
| OID30978 | SORBS1 | Neurology_II | SORBS1 | Q9BX66 | 10 | 95311771 | 95561414 | 0.87 | 0.71 | 1.06 | 1.67E-01 | 0.465 |
| OID21317 | CNPY4 | Oncology | CNPY4 | Q8N129 | 7 | 100119634 | 100125508 | 0.89 | 0.75 | 1.05 | 1.68E-01 | 0.467 |
| OID20572 | EGLN1 | Inflammation | EGLN1 | Q9GZT9 | 1 | 231363751 | 231422287 | 0.87 | 0.71 | 1.06 | 1.68E-01 | 0.467 |
| OID30751 | LRG1 | Inflammation_II | LRG1 | P02750 | 19 | 4536402 | 4540036 | 1.13 | 0.95 | 1.34 | 1.68E-01 | 0.467 |
| OID20386 | SOD1 | Cardiometabolic | SOD1 | P00441 | 21 | 31659666 | 31668931 | 0.88 | 0.74 | 1.05 | 1.68E-01 | 0.467 |
| OID20837 | STK24 | Neurology | STK24 | Q9Y6E0 | 13 | 98445185 | 98577940 | 0.89 | 0.76 | 1.05 | 1.69E-01 | 0.469 |
| OID20927 | APRT | Neurology | APRT | P07741 | 16 | 88809339 | 88811937 | 0.89 | 0.75 | 1.05 | 1.69E-01 | 0.470 |
| OID30117 | ZCCHC8 | Cardiometabolic_II | ZCCHC8 | Q6NZY4 | 12 | 122471599 | 122501073 | 0.87 | 0.71 | 1.06 | 1.70E-01 | 0.470 |
| OID20106 | DCN | Cardiometabolic | DCN | P07585 | 12 | 91140484 | 91183217 | 1.12 | 0.95 | 1.32 | 1.70E-01 | 0.471 |
| OID30739 | F12 | Inflammation_II | F12 | P00748 | 5 | 177402133 | 177416583 | 0.89 | 0.76 | 1.05 | 1.71E-01 | 0.471 |
| OID30187 | FHIP2A | Cardiometabolic_II | FHIP2A | Q5W0V3 | 10 | 114821744 | 114899832 | 0.88 | 0.74 | 1.06 | 1.71E-01 | 0.471 |
| OID20581 | ITGA11 | Inflammation | ITGA11 | Q9UKX5 | 15 | 68296532 | 68432163 | 0.89 | 0.75 | 1.05 | 1.71E-01 | 0.471 |
| OID30344 | PGD | Cardiometabolic_II | PGD | P52209 | 1 | 10398592 | 10420511 | 0.88 | 0.74 | 1.05 | 1.71E-01 | 0.471 |
| OID20969 | PSME1 | Neurology | PSME1 | Q06323 | 14 | 24136163 | 24138967 | 0.89 | 0.75 | 1.05 | 1.71E-01 | 0.471 |
| OID31145 | SH3GLB2 | Neurology_II | SH3GLB2 | Q9NR46 | 9 | 129007036 | 129028331 | 0.89 | 0.76 | 1.05 | 1.71E-01 | 0.471 |
| OID30316 | TCOF1 | Cardiometabolic_II | TCOF1 | Q13428 | 5 | 150357629 | 150400308 | 0.87 | 0.71 | 1.06 | 1.70E-01 | 0.471 |
| OID30526 | ZBP1 | Inflammation_II | ZBP1 | Q9H171 | 20 | 57603846 | 57620576 | 0.86 | 0.70 | 1.07 | 1.71E-01 | 0.471 |
| OID30920 | GNGT1 | Neurology_II | GNGT1 | P63211 | 7 | 93591573 | 93911265 | 0.88 | 0.73 | 1.06 | 1.72E-01 | 0.471 |
| OID30450 | LRIG3 | Inflammation_II | LRIG3 | Q6UXM1 | 12 | 58872149 | 58920504 | 1.11 | 0.96 | 1.28 | 1.72E-01 | 0.471 |
| OID30136 | EDEM2 | Cardiometabolic_II | EDEM2 | Q9BV94 | 20 | 35115364 | 35147336 | 0.87 | 0.72 | 1.06 | 1.72E-01 | 0.471 |
| OID21303 | HS3ST3B1 | Oncology | HS3ST3B1 | Q9Y662 | 17 | 14301081 | 14349404 | 0.90 | 0.77 | 1.05 | 1.73E-01 | 0.473 |
| OID31399 | MINDY1 | Oncology_II | MINDY1 | Q8N5J2 | 1 | 150996549 | 151008376 | 0.89 | 0.76 | 1.05 | 1.73E-01 | 0.473 |

| | | | | | | | | | | | | |
|---|---|---|---|---|---|---|---|---|---|---|---|---|
| OID21153 | PAMR1 | Neurology | PAMR1 | Q6UXH9 | 11 | 35431823 | 35530300 | 0.88 | 0.74 | 1.06 | 1.73E-01 | 0.473 |
| OID30135 | ENPEP | Cardiometabolic_II | ENPEP | Q07075 | 4 | 110365733 | 110565285 | 1.11 | 0.95 | 1.30 | 1.74E-01 | 0.473 |
| OID30626 | FUOM | Inflammation_II | FUOM | A2VDF0 | 10 | 133355158 | 133358025 | 0.90 | 0.76 | 1.05 | 1.74E-01 | 0.473 |
| OID21320 | KIFBP | Oncology | KIFBP | Q96EK5 | 10 | 68988803 | 69043544 | 0.89 | 0.76 | 1.05 | 1.74E-01 | 0.473 |
| OID31444 | MCTS1 | Oncology_II | MCTS1 | Q9ULC4 | X | 120594010 | 120621159 | 0.88 | 0.74 | 1.06 | 1.73E-01 | 0.473 |
| OID30496 | REPS1 | Inflammation_II | REPS1 | Q96D71 | 6 | 138903493 | 138988261 | 0.89 | 0.74 | 1.06 | 1.74E-01 | 0.473 |
| OID20849 | NXPH1 | Neurology | NXPH1 | P58417 | 7 | 8433609 | 8752961 | 0.89 | 0.76 | 1.05 | 1.74E-01 | 0.473 |
| OID21085 | NID2 | Neurology | NID2 | Q14112 | 14 | 52004809 | 52069059 | 0.89 | 0.76 | 1.05 | 1.75E-01 | 0.476 |
| OID20503 | FGF2 | Inflammation | FGF2 | P09038 | 4 | 122826708 | 122898236 | 0.89 | 0.76 | 1.05 | 1.76E-01 | 0.476 |
| OID20388 | TFPI | Cardiometabolic | TFPI | P10646 | 2 | 187464230 | 187565760 | 0.89 | 0.75 | 1.05 | 1.76E-01 | 0.476 |
| OID20067 | VSTM2L | Cardiometabolic | VSTM2L | Q96N03 | 20 | 37903111 | 37945350 | 0.89 | 0.74 | 1.06 | 1.76E-01 | 0.476 |
| OID31385 | ACRBP | Oncology_II | ACRBP | Q8NEB7 | 12 | 6638075 | 6647433 | 0.88 | 0.74 | 1.06 | 1.76E-01 | 0.477 |
| OID20937 | FKBP5 | Neurology | FKBP5 | Q13451 | 6 | 35573585 | 35728583 | 0.89 | 0.76 | 1.05 | 1.77E-01 | 0.477 |
| OID21043 | FABP5 | Neurology | FABP5 | Q01469 | 8 | 81280536 | 81284777 | 0.89 | 0.74 | 1.06 | 1.77E-01 | 0.477 |
| OID20973 | CCS | Neurology | CCS | O14618 | 11 | 66593153 | 66606019 | 0.89 | 0.75 | 1.05 | 1.77E-01 | 0.477 |
| OID31197 | SPRING1 | Oncology_II | SPRING1 | Q9H741 | 12 | 116710171 | 116738070 | 0.87 | 0.71 | 1.06 | 1.77E-01 | 0.477 |
| OID30455 | MKI67 | Inflammation_II | MKI67 | P46013 | 10 | 128096659 | 128126423 | 0.86 | 0.69 | 1.07 | 1.77E-01 | 0.477 |
| OID21244 | AKR1B1 | Oncology | AKR1B1 | P15121 | 7 | 134442356 | 134459284 | 0.90 | 0.76 | 1.05 | 1.79E-01 | 0.478 |
| OID30660 | CHAD | Inflammation_II | CHAD | O15335 | 17 | 50464496 | 50468906 | 0.90 | 0.77 | 1.05 | 1.78E-01 | 0.478 |
| OID30869 | CHP1 | Neurology_II | CHP1 | Q99653 | 15 | 41230839 | 41281887 | 0.88 | 0.74 | 1.06 | 1.78E-01 | 0.478 |
| OID20672 | MMP1 | Inflammation | MMP1 | P03956 | 11 | 102789401 | 102798160 | 0.90 | 0.77 | 1.05 | 1.78E-01 | 0.478 |
| OID30151 | PDZD2 | Cardiometabolic_II | PDZD2 | O15018 | 5 | 31639131 | 32110932 | 0.89 | 0.74 | 1.06 | 1.79E-01 | 0.478 |
| OID20331 | PPIB | Cardiometabolic | PPIB | P23284 | 15 | 64155740 | 64163134 | 0.89 | 0.76 | 1.05 | 1.79E-01 | 0.478 |
| OID30621 | PRKG1 | Inflammation_II | PRKG1 | Q13976 | 10 | 50990888 | 52298423 | 0.89 | 0.76 | 1.05 | 1.78E-01 | 0.478 |
| OID20402 | PROC | Cardiometabolic | PROC | P04070 | 2 | 127418427 | 127429242 | 0.90 | 0.77 | 1.05 | 1.79E-01 | 0.478 |
| OID20912 | PTPRN2 | Neurology | PTPRN2 | Q92932 | 7 | 157539056 | 158587823 | 0.90 | 0.76 | 1.05 | 1.79E-01 | 0.478 |
| OID30959 | RPGR | Neurology_II | RPGR | Q92834 | X | 38269163 | 38327544 | 1.10 | 0.96 | 1.26 | 1.79E-01 | 0.478 |
| OID30942 | TTF2 | Neurology_II | TTF2 | Q9UNY4 | 1 | 117060326 | 117107453 | 0.89 | 0.76 | 1.05 | 1.78E-01 | 0.478 |
| OID20307 | CNTN1 | Cardiometabolic | CNTN1 | Q12860 | 12 | 40692439 | 41072415 | 1.13 | 0.95 | 1.34 | 1.80E-01 | 0.479 |
| OID20748 | LGALS4 | Inflammation | LGALS4 | P56470 | 19 | 38801671 | 38812945 | 1.12 | 0.95 | 1.32 | 1.80E-01 | 0.479 |
| OID30817 | LYPLA2 | Neurology_II | LYPLA2 | O95372 | 1 | 23791145 | 23795539 | 0.88 | 0.74 | 1.06 | 1.80E-01 | 0.479 |
| OID30354 | RAB27B | Cardiometabolic_II | RAB27B | O00194 | 18 | 54717860 | 54895516 | 0.90 | 0.77 | 1.05 | 1.80E-01 | 0.479 |
| OID30518 | ANKMY2 | Inflammation_II | ANKMY2 | Q8IV38 | 7 | 16599779 | 16645817 | 0.89 | 0.76 | 1.05 | 1.82E-01 | 0.480 |
| OID20461 | IL3RA | Inflammation | IL3RA | P26951 | X | 1336616 | 1382689 | 0.89 | 0.75 | 1.06 | 1.81E-01 | 0.480 |
| OID21187 | RRM2 | Oncology | RRM2 | P31350 | 2 | 10120698 | 10211725 | 0.88 | 0.73 | 1.06 | 1.81E-01 | 0.480 |
| OID30053 | THRAP3 | Cardiometabolic_II | THRAP3 | Q9Y2W1 | 1 | 36224432 | 36305357 | 1.10 | 0.96 | 1.26 | 1.81E-01 | 0.480 |
| OID21482 | TNFRSF12A | Oncology | TNFRSF12A | Q9NP84 | 16 | 3018445 | 3022383 | 1.12 | 0.95 | 1.32 | 1.81E-01 | 0.480 |
| OID30794 | UPK3A | Neurology_II | UPK3A | O75631 | 22 | 45284949 | 45295874 | 1.10 | 0.96 | 1.26 | 1.82E-01 | 0.480 |
| OID31053 | CSNK2A1 | Neurology_II | CSNK2A1 | P68400 | 20 | 472498 | 543835 | 0.89 | 0.75 | 1.06 | 1.82E-01 | 0.480 |
| OID20501 | FABP9 | Inflammation | FABP9 | Q0Z7S8 | 8 | 81458253 | 81461579 | 1.11 | 0.95 | 1.29 | 1.83E-01 | 0.482 |
| OID31065 | FKBP14 | Neurology_II | FKBP14 | Q9NWM8 | 7 | 30010587 | 30026702 | 0.90 | 0.76 | 1.05 | 1.83E-01 | 0.482 |
| OID20890 | GPKOW | Neurology | GPKOW | Q92917 | X | 49113407 | 49123735 | 0.87 | 0.71 | 1.07 | 1.83E-01 | 0.482 |
| OID21509 | P4HB | Oncology | P4HB | P07237 | 17 | 81843159 | 81860856 | 0.89 | 0.75 | 1.06 | 1.83E-01 | 0.482 |
| OID30635 | RIDA | Inflammation_II | RIDA | P52758 | 8 | 98102344 | 98117171 | 0.88 | 0.74 | 1.06 | 1.83E-01 | 0.482 |
| OID30740 | IGLC2 | Inflammation_II | IGLC2 | P0DOY2 | 22 | 22900976 | 22901437 | 0.86 | 0.70 | 1.07 | 1.84E-01 | 0.482 |
| OID20188 | LPL | Cardiometabolic | LPL | P06858 | 8 | 19901717 | 19967259 | 1.14 | 0.94 | 1.38 | 1.84E-01 | 0.483 |
| OID20267 | IL2RA | Cardiometabolic | IL2RA | P01589 | 10 | 6010689 | 6062370 | 1.11 | 0.95 | 1.29 | 1.85E-01 | 0.484 |
| OID30684 | F10 | Inflammation_II | F10 | P00742 | 13 | 113122799 | 113149529 | 0.90 | 0.76 | 1.05 | 1.85E-01 | 0.485 |
| OID20597 | PLXNA4 | Inflammation | PLXNA4 | Q9HCM2 | 7 | 132123340 | 132648688 | 0.90 | 0.77 | 1.05 | 1.85E-01 | 0.485 |
| OID20418 | TIMP1 | Cardiometabolic | TIMP1 | P01033 | X | 47582408 | 47586789 | 0.89 | 0.75 | 1.06 | 1.86E-01 | 0.485 |
| OID31457 | DUT | Oncology_II | DUT | P33316 | 15 | 48331011 | 48343373 | 0.89 | 0.75 | 1.06 | 1.86E-01 | 0.485 |
| OID21224 | NDUFS6 | Oncology | NDUFS6 | O75380 | 5 | 1801407 | 1816048 | 0.88 | 0.73 | 1.06 | 1.87E-01 | 0.487 |
| OID21505 | WFDC2 | Oncology | WFDC2 | Q14508 | 20 | 45469753 | 45481532 | 1.12 | 0.95 | 1.32 | 1.87E-01 | 0.487 |
| OID20650 | VEGFA | Inflammation | VEGFA | P15692 | 6 | 43770184 | 43786487 | 0.89 | 0.75 | 1.06 | 1.87E-01 | 0.488 |
| OID21468 | IDUA | Oncology | IDUA | P35475 | 4 | 986997 | 1004564 | 0.90 | 0.76 | 1.05 | 1.88E-01 | 0.489 |
| OID30760 | KLKB1 | Inflammation_II | KLKB1 | P03952 | 4 | 186226438 | 186258471 | 0.89 | 0.76 | 1.06 | 1.88E-01 | 0.489 |
| OID20896 | AGR2 | Neurology | AGR2 | O95994 | 7 | 16791811 | 16833433 | 1.11 | 0.95 | 1.29 | 1.91E-01 | 0.490 |
| OID31508 | CDC26 | Oncology_II | CDC26 | Q8NHZ8 | 9 | 113255835 | 113275572 | 0.89 | 0.75 | 1.06 | 1.90E-01 | 0.490 |
| OID20055 | CRHR1 | Cardiometabolic | CRHR1 | P34998 | 17 | 45784280 | 45835828 | 0.89 | 0.74 | 1.06 | 1.90E-01 | 0.490 |
| OID31171 | CYB5A | Oncology_II | CYB5A | P00167 | 18 | 74250846 | 74291973 | 0.88 | 0.73 | 1.06 | 1.89E-01 | 0.490 |
| OID31357 | DHPS | Oncology_II | DHPS | P49366 | 19 | 12675717 | 12681880 | 0.88 | 0.73 | 1.06 | 1.90E-01 | 0.490 |
| OID20242 | EPHB4 | Cardiometabolic | EPHB4 | P54760 | 7 | 100802565 | 100827523 | 1.11 | 0.95 | 1.29 | 1.90E-01 | 0.490 |
| OID21231 | GNE | Oncology | GNE | Q9Y223 | 9 | 36214441 | 36277042 | 0.90 | 0.76 | 1.06 | 1.91E-01 | 0.490 |
| OID31340 | GPRC5C | Oncology_II | GPRC5C | Q9NQ84 | 17 | 74424851 | 74451653 | 0.89 | 0.74 | 1.06 | 1.90E-01 | 0.490 |
| OID20718 | HSPA1A | Inflammation | HSPA1A | P0DMV8 | 6 | 31815543 | 31817946 | 0.89 | 0.75 | 1.06 | 1.89E-01 | 0.490 |
| OID20436 | LAP3 | Inflammation | LAP3 | P28838 | 4 | 17577198 | 17607972 | 0.89 | 0.75 | 1.06 | 1.91E-01 | 0.490 |
| OID20749 | LHPP | Inflammation | LHPP | Q9H008 | 10 | 124461823 | 124617888 | 0.89 | 0.75 | 1.06 | 1.89E-01 | 0.490 |
| OID21439 | MMP12 | Oncology | MMP12 | P39900 | 11 | 102862736 | 102874982 | 1.12 | 0.94 | 1.34 | 1.90E-01 | 0.490 |
| OID30251 | MYL3 | Cardiometabolic_II | MYL3 | P08590 | 3 | 46835111 | 46882172 | 0.89 | 0.74 | 1.06 | 1.90E-01 | 0.490 |
| OID20160 | PLXNB3 | Cardiometabolic | PLXNB3 | Q9ULL4 | X | 153764196 | 153779346 | 0.90 | 0.76 | 1.06 | 1.91E-01 | 0.490 |
| OID30207 | SIL1 | Cardiometabolic_II | SIL1 | Q9H173 | 5 | 138946724 | 139293557 | 0.87 | 0.71 | 1.07 | 1.91E-01 | 0.490 |
| OID31374 | SNX2 | Oncology_II | SNX2 | O60749 | 5 | 122775079 | 122834543 | 0.89 | 0.75 | 1.06 | 1.90E-01 | 0.490 |
| OID21230 | CHAC2 | Oncology | CHAC2 | Q8WUX2 | 2 | 53767804 | 53775196 | 0.89 | 0.75 | 1.06 | 1.92E-01 | 0.491 |
| OID20223 | CHRDL2 | Cardiometabolic | CHRDL2 | Q6WN34 | 11 | 74696429 | 74731426 | 1.13 | 0.94 | 1.35 | 1.92E-01 | 0.491 |
| OID20761 | SKAP2 | Inflammation | SKAP2 | O75563 | 7 | 26667068 | 26995239 | 0.90 | 0.76 | 1.06 | 1.92E-01 | 0.491 |
| OID21286 | TBC1D23 | Oncology | TBC1D23 | Q9NUY8 | 3 | 100260992 | 100325251 | 0.90 | 0.76 | 1.06 | 1.92E-01 | 0.491 |
| OID30408 | ITGBL1 | Cardiometabolic_II | ITGBL1 | O95965 | 13 | 101452593 | 101720856 | 1.12 | 0.95 | 1.32 | 1.93E-01 | 0.491 |
| OID31344 | LRRFIP1 | Oncology_II | LRRFIP1 | Q32MZ4 | 2 | 237627587 | 237813682 | 0.89 | 0.75 | 1.06 | 1.93E-01 | 0.491 |
| OID20477 | IL17C | Inflammation | IL17C | Q9P0M4 | 16 | 88638572 | 88640468 | 1.11 | 0.95 | 1.29 | 1.93E-01 | 0.491 |
| OID31523 | ARF6 | Oncology_II | ARF6 | P62330 | 14 | 49893079 | 49897054 | 0.90 | 0.76 | 1.06 | 1.94E-01 | 0.491 |
| OID31426 | CHCHD6 | Oncology_II | CHCHD6 | Q9BRQ6 | 3 | 126704240 | 126960420 | 0.88 | 0.73 | 1.07 | 1.94E-01 | 0.491 |
| OID20762 | CXCL1 | Inflammation | CXCL1 | P09341 | 4 | 73869393 | 73871308 | 0.89 | 0.74 | 1.06 | 1.93E-01 | 0.491 |
| OID21424 | RTN4R | Oncology | RTN4R | Q9BZR6 | 22 | 20241415 | 20283246 | 0.89 | 0.74 | 1.06 | 1.93E-01 | 0.491 |
| OID21450 | SPINK6 | Oncology | SPINK6 | Q6UWN8 | 5 | 148202794 | 148215137 | 1.13 | 0.94 | 1.36 | 1.94E-01 | 0.491 |
| OID30741 | AFM | Inflammation_II | AFM | P43652 | 4 | 73481745 | 73504001 | 0.89 | 0.76 | 1.06 | 1.95E-01 | 0.493 |
| OID30111 | SAT1 | Cardiometabolic_II | SAT1 | P21673 | X | 23783173 | 23786210 | 0.88 | 0.73 | 1.07 | 1.95E-01 | 0.493 |
| OID20089 | ADAMTS16 | Cardiometabolic | ADAMTS16 | Q8TE57 | 5 | 5140330 | 5320304 | 0.88 | 0.73 | 1.07 | 1.96E-01 | 0.493 |
| OID21080 | CD164 | Neurology | CD164 | Q04900 | 6 | 109366514 | 109382467 | 0.88 | 0.73 | 1.07 | 1.96E-01 | 0.493 |
| OID20377 | CD55 | Cardiometabolic | CD55 | P08174 | 1 | 207321532 | 207386804 | 0.90 | 0.77 | 1.06 | 1.96E-01 | 0.493 |
| OID20165 | CLEC5A | Cardiometabolic | CLEC5A | Q9NY25 | 7 | 141927357 | 141947007 | 1.12 | 0.94 | 1.32 | 1.96E-01 | 0.493 |
| OID20559 | CLIP2 | Inflammation | CLIP2 | Q9UDT6 | 7 | 74289407 | 74405935 | 0.90 | 0.76 | 1.06 | 1.96E-01 | 0.493 |
| OID20729 | PDLIM7 | Inflammation | PDLIM7 | Q9NR12 | 5 | 177483394 | 177497606 | 0.90 | 0.76 | 1.06 | 1.96E-01 | 0.493 |
| OID20255 | TGFBR3 | Cardiometabolic | TGFBR3 | Q03167 | 1 | 91680343 | 91906335 | 1.11 | 0.95 | 1.30 | 1.96E-01 | 0.493 |
| OID31278 | TMED1 | Oncology_II | TMED1 | Q13445 | 19 | 10832067 | 10836318 | 0.88 | 0.73 | 1.07 | 1.96E-01 | 0.493 |
| OID21254 | SLAMF6 | Oncology | SLAMF6 | Q96DU3 | 1 | 160485030 | 160523262 | 0.90 | 0.76 | 1.06 | 1.97E-01 | 0.494 |
| OID20956 | DNMBP | Neurology | DNMBP | Q6XZF7 | 10 | 99875577 | 100009947 | 0.89 | 0.74 | 1.06 | 1.97E-01 | 0.495 |
| OID20241 | CA4 | Cardiometabolic | CA4 | P22748 | 17 | 60149942 | 60170899 | 1.11 | 0.95 | 1.30 | 1.98E-01 | 0.496 |
| OID30714 | GSN | Inflammation_II | GSN | P06396 | 9 | 121207794 | 121332843 | 0.90 | 0.76 | 1.06 | 1.99E-01 | 0.498 |
| OID21432 | ACP6 | Oncology | ACP6 | Q9NPH0 | 1 | 147629652 | 147670524 | 0.91 | 0.78 | 1.05 | 1.99E-01 | 0.498 |
| OID20601 | IL5RA | Inflammation | IL5RA | Q01344 | 3 | 3066324 | 3126613 | 1.11 | 0.95 | 1.31 | 2.00E-01 | 0.499 |
| OID30320 | LECT2 | Cardiometabolic_II | LECT2 | O14960 | 5 | 135922279 | 135954983 | 0.89 | 0.73 | 1.07 | 2.00E-01 | 0.499 |
| OID20341 | TCN2 | Cardiometabolic | TCN2 | P20062 | 22 | 30607003 | 30627271 | 1.10 | 0.95 | 1.28 | 2.00E-01 | 0.499 |
| OID21035 | LAMP2 | Neurology | LAMP2 | P13473 | X | 120426148 | 120469365 | 0.90 | 0.76 | 1.06 | 2.00E-01 | 0.500 |
| OID20353 | CDH1 | Cardiometabolic | CDH1 | P12830 | 16 | 68737292 | 68835537 | 0.89 | 0.74 | 1.06 | 2.01E-01 | 0.501 |
| OID21002 | PLA2G10 | Neurology | PLA2G10 | O15496 | 16 | 14672548 | 14694308 | 1.12 | 0.94 | 1.32 | 2.01E-01 | 0.501 |
| OID20457 | SCGN | Inflammation | SCGN | O76038 | 6 | 25652201 | 25701783 | 1.11 | 0.95 | 1.30 | 2.01E-01 | 0.501 |
| OID31276 | CASP4 | Oncology_II | CASP4 | P49662 | 11 | 104942866 | 104969366 | 0.89 | 0.75 | 1.06 | 2.04E-01 | 0.501 |

| | | | | | | | | | | | | |
|---|---|---|---|---|---|---|---|---|---|---|---|---|
| OID20674 | CCL25 | Inflammation | CCL25 | O15444 | 19 | 8052318 | 8062660 | 1.12 | 0.94 | 1.33 | 2.03E-01 | 0.501 |
| OID21200 | CCT5 | Oncology | CCT5 | P48643 | 5 | 10249929 | 10266389 | 0.89 | 0.75 | 1.06 | 2.04E-01 | 0.501 |
| OID30830 | DCUN1D1 | Neurology_II | DCUN1D1 | Q96GG9 | 3 | 182938074 | 182985953 | 0.89 | 0.74 | 1.07 | 2.04E-01 | 0.501 |
| OID30302 | EHBP1 | Cardiometabolic_II | EHBP1 | Q8NDI1 | 2 | 62673851 | 63046487 | 0.90 | 0.76 | 1.06 | 2.04E-01 | 0.501 |
| OID20522 | EPO | Inflammation | EPO | P01588 | 7 | 100720468 | 100723700 | 1.11 | 0.94 | 1.31 | 2.05E-01 | 0.501 |
| OID20758 | ESM1 | Inflammation | ESM1 | Q9NQ30 | 5 | 54977867 | 55022671 | 1.12 | 0.94 | 1.33 | 2.05E-01 | 0.501 |
| OID30781 | F13B | Inflammation_II | F13B | P05160 | 1 | 197038741 | 197067260 | 0.89 | 0.75 | 1.06 | 2.02E-01 | 0.501 |
| OID20665 | FASLG | Inflammation | FASLG | P48023 | 1 | 172659103 | 172666876 | 0.89 | 0.75 | 1.06 | 2.02E-01 | 0.501 |
| OID20065 | HK2 | Cardiometabolic | HK2 | P52789 | 2 | 74834127 | 74893359 | 0.90 | 0.77 | 1.06 | 2.05E-01 | 0.501 |
| OID30866 | HTR1B | Neurology_II | HTR1B | P28222 | 6 | 77460924 | 77463491 | 0.89 | 0.74 | 1.07 | 2.04E-01 | 0.501 |
| OID31491 | IFNAR1 | Oncology_II | IFNAR1 | P17181 | 21 | 33324387 | 33359864 | 0.90 | 0.76 | 1.06 | 2.02E-01 | 0.501 |
| OID21315 | LAG3 | Oncology | LAG3 | P18627 | 12 | 6772512 | 6778455 | 0.89 | 0.75 | 1.06 | 2.02E-01 | 0.501 |
| OID30376 | LILRA3 | Cardiometabolic_II | LILRA3 | Q8N6C8 | 19 | 54294163 | 54306461 | 1.15 | 0.93 | 1.44 | 2.03E-01 | 0.501 |
| OID21099 | MESD | Neurology | MESD | Q14696 | 15 | 80946289 | 80989828 | 0.90 | 0.76 | 1.06 | 2.03E-01 | 0.501 |
| OID30633 | MYOM3 | Inflammation_II | MYOM3 | Q5VTT5 | 1 | 24056035 | 24112135 | 0.89 | 0.75 | 1.06 | 2.03E-01 | 0.501 |
| OID20819 | PAK4 | Neurology | PAK4 | O96013 | 19 | 39125770 | 39182816 | 0.90 | 0.75 | 1.06 | 2.03E-01 | 0.501 |
| OID20549 | PSMG3 | Inflammation | PSMG3 | Q9BT73 | 7 | 1567332 | 1571005 | 0.90 | 0.76 | 1.06 | 2.05E-01 | 0.501 |
| OID20373 | RARRES2 | Cardiometabolic | RARRES2 | Q99969 | 7 | 150338317 | 150341662 | 0.89 | 0.74 | 1.07 | 2.05E-01 | 0.501 |
| OID30979 | SPAG1 | Neurology_II | SPAG1 | Q07617 | 8 | 100157906 | 100259278 | 0.89 | 0.73 | 1.07 | 2.04E-01 | 0.501 |
| OID20818 | SSB | Neurology | SSB | P05455 | 2 | 169791933 | 169812064 | 0.88 | 0.73 | 1.07 | 2.03E-01 | 0.501 |
| OID21098 | SUSD2 | Neurology | SUSD2 | Q9UGT4 | 22 | 24181487 | 24189106 | 1.11 | 0.94 | 1.31 | 2.02E-01 | 0.501 |
| OID20121 | CCL27 | Cardiometabolic | CCL27 | Q9Y4X3 | 9 | 34661880 | 34664048 | 0.89 | 0.75 | 1.06 | 2.05E-01 | 0.501 |
| OID21337 | ELOA | Oncology | ELOA | Q14241 | 1 | 23743448 | 23762059 | 0.88 | 0.72 | 1.07 | 2.05E-01 | 0.501 |
| OID30723 | F2 | Inflammation_II | F2 | P00734 | 11 | 46719196 | 46739506 | 0.90 | 0.76 | 1.06 | 2.06E-01 | 0.501 |
| OID30709 | AGT | Inflammation_II | AGT | P01019 | 1 | 230690776 | 230745576 | 1.10 | 0.95 | 1.27 | 2.07E-01 | 0.501 |
| OID30792 | ARMCX2 | Neurology_II | ARMCX2 | Q7L311 | X | 101655281 | 101659850 | 0.89 | 0.74 | 1.07 | 2.07E-01 | 0.501 |
| OID20835 | PLIN1 | Neurology | PLIN1 | O60240 | 15 | 89664367 | 89679427 | 0.89 | 0.74 | 1.07 | 2.07E-01 | 0.501 |
| OID30529 | S100A13 | Inflammation_II | S100A13 | Q99584 | 1 | 153618787 | 153631360 | 0.89 | 0.75 | 1.06 | 2.07E-01 | 0.501 |
| OID20131 | VSIR | Cardiometabolic | VSIR | Q9H7M9 | 10 | 71747556 | 71773520 | 0.90 | 0.76 | 1.06 | 2.06E-01 | 0.501 |
| OID21229 | SUGT1 | Oncology | SUGT1 | Q9Y2Z0 | 13 | 52652709 | 52700909 | 0.90 | 0.77 | 1.06 | 2.07E-01 | 0.502 |
| OID30804 | SCN3B | Neurology_II | SCN3B | Q9NY72 | 11 | 123629187 | 123655244 | 0.89 | 0.74 | 1.07 | 2.08E-01 | 0.502 |
| OID31121 | TXNDC9 | Neurology_II | TXNDC9 | O14530 | 2 | 99318982 | 99340702 | 0.90 | 0.75 | 1.06 | 2.08E-01 | 0.502 |
| OID30775 | CD5L | Inflammation_II | CD5L | O43866 | 1 | 157830911 | 157898256 | 0.90 | 0.77 | 1.06 | 2.08E-01 | 0.502 |
| OID21458 | NCS1 | Oncology | NCS1 | P62166 | 9 | 130172404 | 130237303 | 1.11 | 0.94 | 1.32 | 2.08E-01 | 0.502 |
| OID30685 | SHBG | Inflammation_II | SHBG | P04278 | 17 | 7613946 | 7633382 | 1.13 | 0.94 | 1.35 | 2.08E-01 | 0.502 |
| OID20771 | CHRDL1 | Inflammation | CHRDL1 | Q9BU40 | X | 110673856 | 110795819 | 1.12 | 0.94 | 1.33 | 2.09E-01 | 0.503 |
| OID31307 | KIAA1549 | Oncology_II | KIAA1549 | Q9HCM3 | 7 | 138831381 | 138981389 | 0.90 | 0.77 | 1.06 | 2.09E-01 | 0.503 |
| OID30686 | APOA2 | Inflammation_II | APOA2 | P02652 | 1 | 161222292 | 161223631 | 1.11 | 0.95 | 1.29 | 2.09E-01 | 0.504 |
| OID20621 | TGFB1 | Inflammation | TGFB1 | P01137 | 19 | 41301587 | 41353922 | 0.90 | 0.76 | 1.06 | 2.10E-01 | 0.504 |
| OID31413 | CCDC134 | Oncology_II | CCDC134 | Q9H6E4 | 22 | 41800679 | 41832164 | 0.90 | 0.76 | 1.06 | 2.10E-01 | 0.504 |
| OID30233 | FSHB | Cardiometabolic_II | FSHB | P01225 | 11 | 30231014 | 30235261 | 0.84 | 0.64 | 1.10 | 2.10E-01 | 0.505 |
| OID31460 | EFCAB14 | Oncology_II | EFCAB14 | O75071 | 1 | 46674659 | 46719146 | 0.90 | 0.76 | 1.06 | 2.11E-01 | 0.507 |
| OID20877 | MAX | Neurology | MAX | P61244 | 14 | 65006174 | 65102695 | 0.90 | 0.76 | 1.06 | 2.11E-01 | 0.507 |
| OID21197 | AKT3 | Oncology | AKT3 | Q9Y243 | 1 | 243488233 | 243851079 | 0.90 | 0.76 | 1.06 | 2.12E-01 | 0.507 |
| OID31070 | CAMSAP1 | Neurology_II | CAMSAP1 | Q5T5Y3 | 9 | 135808487 | 135907546 | 0.90 | 0.76 | 1.06 | 2.12E-01 | 0.507 |
| OID30981 | SOX2 | Neurology_II | SOX2 | P48431 | 3 | 181711925 | 181714436 | 0.90 | 0.76 | 1.06 | 2.12E-01 | 0.507 |
| OID20697 | CXCL10 | Inflammation | CXCL10 | P02778 | 4 | 76021118 | 76023497 | 0.89 | 0.74 | 1.07 | 2.13E-01 | 0.508 |
| OID30831 | ECSCR | Neurology_II | ECSCR | Q19T08 | 5 | 139448560 | 139462743 | 0.88 | 0.72 | 1.08 | 2.13E-01 | 0.508 |
| OID20605 | IL32 | Inflammation | IL32 | P24001 | 16 | 3065297 | 3082192 | 1.11 | 0.94 | 1.30 | 2.13E-01 | 0.508 |
| OID21416 | ITGAV | Oncology | ITGAV | P06756 | 2 | 186590056 | 186680901 | 0.90 | 0.76 | 1.06 | 2.13E-01 | 0.508 |
| OID30782 | TTR | Inflammation_II | TTR | P02766 | 18 | 31557009 | 31598833 | 0.90 | 0.76 | 1.06 | 2.13E-01 | 0.508 |
| OID31030 | RTN4IP1 | Neurology_II | RTN4IP1 | Q8WWV3 | 6 | 106570771 | 106629498 | 0.90 | 0.76 | 1.06 | 2.14E-01 | 0.509 |
| OID30641 | CD72 | Inflammation_II | CD72 | P21854 | 9 | 35609982 | 35646810 | 0.90 | 0.76 | 1.06 | 2.14E-01 | 0.509 |
| OID21022 | NSFL1C | Neurology | NSFL1C | Q9UNZ2 | 20 | 1442162 | 1473842 | 0.90 | 0.76 | 1.06 | 2.16E-01 | 0.512 |
| OID30749 | APOC1 | Inflammation_II | APOC1 | P02654 | 19 | 44914247 | 44919349 | 0.89 | 0.74 | 1.07 | 2.17E-01 | 0.513 |
| OID20172 | FCRL1 | Cardiometabolic | FCRL1 | Q96LA6 | 1 | 157794403 | 157820120 | 0.90 | 0.77 | 1.06 | 2.16E-01 | 0.513 |
| OID20589 | HEXIM1 | Inflammation | HEXIM1 | O94992 | 17 | 45148475 | 45152099 | 0.90 | 0.77 | 1.06 | 2.16E-01 | 0.513 |
| OID20900 | IL18RAP | Neurology | IL18RAP | O95256 | 2 | 102418689 | 102452565 | 0.90 | 0.76 | 1.06 | 2.16E-01 | 0.513 |
| OID20777 | PON3 | Inflammation | PON3 | Q15166 | 7 | 95359872 | 95396375 | 1.13 | 0.93 | 1.38 | 2.17E-01 | 0.513 |
| OID31031 | SUOX | Neurology_II | SUOX | P51687 | 12 | 55997180 | 56006641 | 0.89 | 0.75 | 1.07 | 2.17E-01 | 0.513 |
| OID30212 | LMOD1 | Cardiometabolic_II/Inflan | LMOD1 | P29536 | 1 | 201896456 | 201946588 | 1.10 | 0.95 | 1.27 | 2.18E-01 | 0.515 |
| OID30889 | MICALL2 | Neurology_II | MICALL2 | Q8IY33 | 7 | 1428465 | 1459470 | 0.90 | 0.75 | 1.07 | 2.18E-01 | 0.515 |
| OID20587 | PIK3AP1 | Inflammation | PIK3AP1 | Q6ZUJ8 | 10 | 96593315 | 96720514 | 0.90 | 0.76 | 1.07 | 2.18E-01 | 0.515 |
| OID30440 | POLR2A | Inflammation_II | POLR2A | P24928 | 17 | 7484366 | 7514616 | 0.90 | 0.75 | 1.07 | 2.18E-01 | 0.515 |
| OID30391 | PRAP1 | Cardiometabolic_II | PRAP1 | Q96NZ9 | 10 | 133347368 | 133352683 | 0.89 | 0.74 | 1.07 | 2.19E-01 | 0.515 |
| OID31433 | PTPRH | Oncology_II | PTPRH | Q9HD43 | 19 | 55181247 | 55209506 | 1.11 | 0.94 | 1.31 | 2.19E-01 | 0.515 |
| OID30506 | RNF168 | Inflammation_II | RNF168 | Q8IYW5 | 3 | 196468783 | 196503768 | 0.89 | 0.74 | 1.07 | 2.19E-01 | 0.515 |
| OID20804 | C19orf12 | Neurology | C19orf12 | Q9NSK7 | 19 | 29698937 | 29715789 | 0.89 | 0.75 | 1.07 | 2.20E-01 | 0.515 |
| OID30892 | DDX25 | Neurology_II | DDX25 | Q9UHL0 | 11 | 125903348 | 125943702 | 0.89 | 0.74 | 1.07 | 2.20E-01 | 0.515 |
| OID30480 | TERF1 | Inflammation_II | TERF1 | P54274 | 8 | 73008856 | 73048123 | 0.90 | 0.75 | 1.07 | 2.19E-01 | 0.515 |
| OID30326 | TCTN3 | Cardiometabolic_II | TCTN3 | Q6NUS6 | 10 | 95659823 | 95694143 | 0.90 | 0.76 | 1.07 | 2.20E-01 | 0.516 |
| OID31263 | HRAS | Oncology_II | HRAS | P01112 | 11 | 532242 | 537321 | 0.88 | 0.73 | 1.08 | 2.21E-01 | 0.517 |
| OID21269 | ACAA1 | Oncology | ACAA1 | P09110 | 3 | 38103129 | 38137242 | 0.90 | 0.76 | 1.06 | 2.22E-01 | 0.518 |
| OID20444 | PRKAB1 | Inflammation | PRKAB1 | Q9Y478 | 12 | 119667864 | 119681624 | 0.90 | 0.75 | 1.07 | 2.22E-01 | 0.518 |
| OID31323 | SAP18 | Oncology_II | SAP18 | O00422 | 13 | 21140119 | 21149097 | 1.09 | 0.95 | 1.26 | 2.22E-01 | 0.518 |
| OID30888 | SOWAHA | Neurology_II | SOWAHA | Q2M3V2 | 5 | 132813302 | 132816786 | 0.89 | 0.73 | 1.07 | 2.22E-01 | 0.518 |
| OID20548 | CEACAM21 | Inflammation | CEACAM21 | Q3KPI0 | 19 | 41549518 | 41586844 | 1.10 | 0.94 | 1.29 | 2.23E-01 | 0.519 |
| OID30339 | SYTL4 | Cardiometabolic_II | SYTL4 | Q96C24 | X | 100671783 | 100732123 | 0.90 | 0.76 | 1.07 | 2.23E-01 | 0.519 |
| OID21273 | PDCD1LG2 | Oncology | PDCD1LG2 | Q9BQ51 | 9 | 5510531 | 5571282 | 1.10 | 0.94 | 1.30 | 2.23E-01 | 0.520 |
| OID21365 | RBP2 | Oncology | RBP2 | P50120 | 3 | 139452884 | 139480747 | 1.10 | 0.94 | 1.29 | 2.24E-01 | 0.522 |
| OID31115 | NGRN | Neurology_II | NGRN | Q9NPE2 | 15 | 90265659 | 90275778 | 0.89 | 0.74 | 1.07 | 2.24E-01 | 0.522 |
| OID30144 | CALY | Cardiometabolic_II | CALY | Q9NYX4 | 10 | 133324072 | 133336935 | 1.10 | 0.94 | 1.29 | 2.25E-01 | 0.522 |
| OID31378 | NMI | Oncology_II | NMI | Q13287 | 2 | 151270470 | 151289894 | 0.90 | 0.76 | 1.07 | 2.26E-01 | 0.524 |
| OID20162 | EIF4EBP1 | Cardiometabolic | EIF4EBP1 | Q13541 | 8 | 38030534 | 38060365 | 0.90 | 0.76 | 1.07 | 2.27E-01 | 0.525 |
| OID30974 | FBN2 | Neurology_II | FBN2 | P35556 | 5 | 128257909 | 128659185 | 0.91 | 0.77 | 1.06 | 2.27E-01 | 0.525 |
| OID30762 | ITIH4 | Inflammation_II | ITIH4 | Q14624 | 3 | 52812962 | 52830688 | 0.89 | 0.73 | 1.08 | 2.26E-01 | 0.525 |
| OID20688 | SCGB1A1 | Inflammation | SCGB1A1 | P11684 | 11 | 62405103 | 62423195 | 0.91 | 0.78 | 1.06 | 2.26E-01 | 0.525 |
| OID21444 | CCN4 | Oncology | CCN4 | O95388 | 8 | 133191039 | 133231690 | 0.90 | 0.75 | 1.07 | 2.27E-01 | 0.525 |
| OID30651 | PINLYP | Inflammation_II | PINLYP | A6NC86 | 19 | 43575801 | 43583964 | 0.91 | 0.77 | 1.06 | 2.27E-01 | 0.525 |
| OID20564 | FCAR | Inflammation | FCAR | P24071 | 19 | 54874235 | 54891420 | 1.10 | 0.94 | 1.28 | 2.28E-01 | 0.526 |
| OID20707 | MANF | Inflammation | MANF | P55145 | 3 | 51385291 | 51389397 | 0.91 | 0.78 | 1.06 | 2.28E-01 | 0.526 |
| OID21279 | SEPTIN9 | Oncology | SEPTIN9 | Q9UHD8 | 17 | 77280569 | 77500596 | 0.89 | 0.75 | 1.07 | 2.28E-01 | 0.526 |
| OID20893 | CASP10 | Neurology | CASP10 | Q92851 | 2 | 201182898 | 201229406 | 0.91 | 0.77 | 1.06 | 2.29E-01 | 0.527 |
| OID30965 | SV2A | Neurology_II | SV2A | Q7L0J3 | 1 | 149903318 | 149917844 | 0.90 | 0.75 | 1.07 | 2.29E-01 | 0.527 |
| OID30082 | ITPA | Cardiometabolic_II | ITPA | Q9BY32 | 20 | 3208868 | 3223870 | 0.90 | 0.77 | 1.07 | 2.29E-01 | 0.527 |
| OID31299 | CTAG1A_CTAG1B | Oncology_II | CTAG1A;CTAG1B | P78358;P78358 | X;X | 4585133;154617609 | 4586816;154619282 | 0.90 | 0.75 | 1.07 | 2.31E-01 | 0.528 |
| OID20691 | F2R | Inflammation | F2R | P25116 | 5 | 76716126 | 76735770 | 0.90 | 0.77 | 1.07 | 2.30E-01 | 0.528 |
| OID30966 | GBA | Neurology_II | GBA | P04062 | 1 | 155234452 | 155244699 | 0.91 | 0.78 | 1.06 | 2.31E-01 | 0.528 |
| OID30487 | IL31 | Inflammation_II | IL31 | Q6EBC2 | 12 | 122172029 | 122174221 | 0.89 | 0.74 | 1.08 | 2.30E-01 | 0.528 |
| OID21343 | RTBDN | Oncology | RTBDN | Q9BSG5 | 19 | 12825478 | 12835428 | 0.90 | 0.77 | 1.07 | 2.30E-01 | 0.528 |
| OID20846 | FKBP7 | Neurology | FKBP7 | Q9Y680 | 2 | 178463664 | 178478600 | 0.89 | 0.74 | 1.08 | 2.32E-01 | 0.531 |
| OID20310 | PTPRF | Cardiometabolic | PTPRF | P10586 | 1 | 43525187 | 43623666 | 0.90 | 0.77 | 1.07 | 2.32E-01 | 0.531 |
| OID30143 | TPR | Cardiometabolic_II | TPR | P12270 | 1 | 186311652 | 186375693 | 0.88 | 0.72 | 1.08 | 2.32E-01 | 0.531 |
| OID21448 | NUCB2 | Oncology | NUCB2 | P80303 | 11 | 17208153 | 17349980 | 0.90 | 0.76 | 1.07 | 2.34E-01 | 0.533 |
| OID30592 | HMCN2 | Inflammation_II | HMCN2 | Q8NDA2 | 9 | 130265760 | 130434123 | 0.91 | 0.78 | 1.06 | 2.35E-01 | 0.534 |
| OID30169 | KRT6C | Cardiometabolic_II | KRT6C | P48668 | 12 | 52468516 | 52473805 | 1.10 | 0.94 | 1.28 | 2.34E-01 | 0.534 |
| OID20209 | LILRA5 | Cardiometabolic | LILRA5 | A6NI73 | 19 | 54307070 | 54313166 | 1.11 | 0.94 | 1.31 | 2.35E-01 | 0.534 |

| | | | | | | | | | | | | |
|---|---|---|---|---|---|---|---|---|---|---|---|---|
| OID21490 | NECTIN4 | Oncology | NECTIN4 | Q96NY8 | 1 | 161070998 | 161089558 | 1.09 | 0.94 | 1.27 | 2.35E-01 | 0.534 |
| OID20202 | GDF2 | Cardiometabolic | GDF2 | Q9UK05 | 10 | 47322454 | 47327588 | 1.11 | 0.93 | 1.32 | 2.35E-01 | 0.535 |
| OID30872 | TXNL1 | Neurology_II | TXNL1 | O43396 | 18 | 56597209 | 56651600 | 0.89 | 0.74 | 1.08 | 2.35E-01 | 0.535 |
| OID30737 | PROS1 | Inflammation_II | PROS1 | P07225 | 3 | 93873051 | 93980003 | 0.90 | 0.76 | 1.07 | 2.36E-01 | 0.536 |
| OID30707 | AMY1A_AMY1B_AMY | Inflammation_II | A;AMY1B;AMY1C | 6;P0DTE7;P0DTE8 | 1;1;1 | 3687415;103745323 | 3696454;103758726 | 0.90 | 0.76 | 1.07 | 2.38E-01 | 0.538 |
| OID20864 | C2CD2L | Neurology | C2CD2L | O14523 | 11 | 119102198 | 119118544 | 0.90 | 0.76 | 1.07 | 2.37E-01 | 0.538 |
| OID21349 | DTX3 | Oncology | DTX3 | Q8N9I9 | 12 | 57604622 | 57609804 | 0.89 | 0.74 | 1.08 | 2.38E-01 | 0.538 |
| OID21388 | NPY | Oncology | NPY | P01303 | 7 | 24284188 | 24291862 | 0.91 | 0.77 | 1.07 | 2.38E-01 | 0.538 |
| OID20238 | PLTP | Cardiometabolic | PLTP | P55058 | 20 | 45898621 | 45912155 | 1.11 | 0.93 | 1.31 | 2.38E-01 | 0.538 |
| OID30432 | STX5 | Inflammation_II | STX5 | Q13190 | 11 | 62806860 | 62832051 | 0.89 | 0.73 | 1.08 | 2.37E-01 | 0.538 |
| OID20446 | ARTN | Inflammation | ARTN | Q5T4W7 | 1 | 43933320 | 43937240 | 1.08 | 0.95 | 1.23 | 2.40E-01 | 0.542 |
| OID20530 | LSP1 | Inflammation | LSP1 | P33241 | 11 | 1850904 | 1892267 | 0.90 | 0.76 | 1.07 | 2.40E-01 | 0.542 |
| OID20936 | VWC2 | Neurology | VWC2 | Q2TAL6 | 7 | 49773638 | 49921950 | 1.11 | 0.93 | 1.31 | 2.40E-01 | 0.542 |
| OID30887 | FARSA | Neurology_II | FARSA | Q9Y285 | 19 | 12922479 | 12934037 | 0.90 | 0.75 | 1.08 | 2.41E-01 | 0.542 |
| OID30678 | NEXN | Inflammation_II | NEXN | Q0ZGT2 | 1 | 77888513 | 77943895 | 0.90 | 0.76 | 1.07 | 2.41E-01 | 0.543 |
| OID30444 | PALLD | Inflammation_II | PALLD | Q8WX93 | 4 | 168497052 | 168928457 | 0.89 | 0.73 | 1.08 | 2.41E-01 | 0.543 |
| OID31025 | SEPTIN3 | Neurology_II | SEPTIN3 | Q9UH03 | 22 | 41969443 | 41998221 | 1.10 | 0.94 | 1.30 | 2.41E-01 | 0.543 |
| OID30862 | IGDCC3 | Neurology_II | IGDCC3 | Q8IVU1 | 15 | 65327127 | 65378002 | 0.89 | 0.73 | 1.08 | 2.42E-01 | 0.544 |
| OID30844 | BCAT2 | Neurology_II | BCAT2 | O15382 | 19 | 48795062 | 48811029 | 0.90 | 0.75 | 1.08 | 2.44E-01 | 0.546 |
| OID30975 | INPP5J | Neurology_II | INPP5J | Q15735 | 22 | 31122731 | 31134697 | 1.09 | 0.94 | 1.25 | 2.44E-01 | 0.546 |
| OID31440 | LRRC37A2 | Oncology_II | LRRC37A2 | A6NM11 | 17 | 46511508 | 46556910 | 0.91 | 0.78 | 1.07 | 2.44E-01 | 0.546 |
| OID30908 | MN1 | Neurology_II | MN1 | Q10571 | 22 | 27748277 | 27801756 | 0.90 | 0.75 | 1.07 | 2.44E-01 | 0.546 |
| OID20744 | DNPH1 | Inflammation | DNPH1 | O43598 | 6 | 43225629 | 43229481 | 0.91 | 0.77 | 1.07 | 2.45E-01 | 0.547 |
| OID30521 | NEDD4L | Inflammation_II | NEDD4L | Q96PU5 | 18 | 58044226 | 58401540 | 0.91 | 0.77 | 1.07 | 2.45E-01 | 0.547 |
| OID20220 | GH1 | Cardiometabolic | GH1 | P01241 | 17 | 63917200 | 63918839 | 1.11 | 0.93 | 1.33 | 2.46E-01 | 0.549 |
| OID31178 | MYH7B | Oncology_II | MYH7B | A7E2Y1 | 20 | 34955810 | 35002437 | 0.90 | 0.75 | 1.08 | 2.46E-01 | 0.549 |
| OID30675 | ABO | Inflammation_II | ABO | P16442 | 9 | 133233278 | 133276024 | 1.10 | 0.94 | 1.30 | 2.48E-01 | 0.551 |
| OID21018 | CD8A | Neurology | CD8A | P01732 | 2 | 86784610 | 86808396 | 0.91 | 0.78 | 1.07 | 2.47E-01 | 0.551 |
| OID21428 | DAB2 | Oncology | DAB2 | P98082 | 5 | 39371675 | 39462300 | 0.91 | 0.77 | 1.07 | 2.48E-01 | 0.551 |
| OID31151 | HEPACAM2 | Neurology_II | HEPACAM2 | A8MVW5 | 7 | 93188534 | 93226469 | 1.10 | 0.94 | 1.29 | 2.48E-01 | 0.551 |
| OID31345 | PRC1 | Oncology_II | PRC1 | O43663 | 15 | 90966040 | 90995629 | 0.88 | 0.71 | 1.09 | 2.47E-01 | 0.551 |
| OID31382 | RALY | Oncology_II | RALY | Q9UKM9 | 20 | 33993646 | 34108308 | 0.89 | 0.73 | 1.08 | 2.48E-01 | 0.551 |
| OID31210 | STAU1 | Oncology_II | STAU1 | O95793 | 20 | 49113339 | 49188367 | 0.90 | 0.74 | 1.08 | 2.48E-01 | 0.551 |
| OID31280 | TEX33 | Oncology_II | TEX33 | O43247 | 22 | 36991120 | 37007851 | 0.89 | 0.74 | 1.08 | 2.49E-01 | 0.552 |
| OID20627 | DNAJA2 | Inflammation | DNAJA2 | O60884 | 16 | 46955362 | 46973674 | 0.91 | 0.77 | 1.07 | 2.49E-01 | 0.553 |
| OID30361 | LRRC59 | Cardiometabolic_II | LRRC59 | Q96AG4 | 17 | 50375059 | 50397523 | 0.90 | 0.76 | 1.07 | 2.49E-01 | 0.553 |
| OID30202 | PYDC1 | Cardiometabolic_II | PYDC1 | Q8WXC3 | 16 | 31215962 | 31217135 | 1.09 | 0.94 | 1.27 | 2.50E-01 | 0.553 |
| OID31510 | NFU1 | Oncology_II | NFU1 | Q9UMS0 | 2 | 69396113 | 69437628 | 0.91 | 0.77 | 1.07 | 2.50E-01 | 0.554 |
| OID21001 | ADAM22 | Neurology | ADAM22 | Q9P0K1 | 7 | 87934143 | 88202889 | 0.91 | 0.77 | 1.07 | 2.51E-01 | 0.554 |
| OID30628 | CTSE | Inflammation_II | CTSE | P14091 | 1 | 206008535 | 206023909 | 0.91 | 0.77 | 1.07 | 2.52E-01 | 0.554 |
| OID30132 | HADH | Cardiometabolic_II | HADH | Q16836 | 4 | 107989714 | 108035241 | 1.08 | 0.95 | 1.23 | 2.52E-01 | 0.554 |
| OID20352 | ITIH3 | Cardiometabolic | ITIH3 | Q06033 | 3 | 52794768 | 52809009 | 1.11 | 0.93 | 1.32 | 2.52E-01 | 0.554 |
| OID31342 | MAMDC4 | Oncology_II | MAMDC4 | Q6UXC1 | 9 | 136850943 | 136860799 | 0.91 | 0.77 | 1.07 | 2.51E-01 | 0.554 |
| OID20687 | MMP10 | Inflammation | MMP10 | P09238 | 11 | 102770502 | 102780628 | 0.91 | 0.77 | 1.07 | 2.52E-01 | 0.554 |
| OID20178 | NADK | Cardiometabolic | NADK | O95544 | 1 | 1751232 | 1780457 | 0.90 | 0.75 | 1.08 | 2.52E-01 | 0.554 |
| OID20064 | SERPINB5 | Cardiometabolic | SERPINB5 | P36952 | 18 | 63476958 | 63505085 | 1.10 | 0.93 | 1.30 | 2.52E-01 | 0.554 |
| OID20593 | MICB_MICA | Inflammation | MICB;MICA | Q29980;Q29983 | 6;6 | 31494881;31399784 | 31511124;31415315 | 1.11 | 0.93 | 1.33 | 2.52E-01 | 0.554 |
| OID21234 | AIF1 | Oncology | AIF1 | P55008 | 6 | 31615217 | 31617021 | 0.91 | 0.77 | 1.07 | 2.53E-01 | 0.556 |
| OID20793 | ODAM | Neurology | ODAM | A1E959 | 4 | 70195725 | 70204576 | 0.91 | 0.77 | 1.07 | 2.53E-01 | 0.556 |
| OID30922 | CD164L2 | Neurology_II | CD164L2 | Q6UWJ8 | 1 | 27379176 | 27383333 | 0.90 | 0.76 | 1.07 | 2.54E-01 | 0.556 |
| OID31209 | PDIA5 | Oncology_II | PDIA5 | Q14554 | 3 | 123067025 | 123225227 | 0.90 | 0.76 | 1.08 | 2.54E-01 | 0.556 |
| OID31481 | KAZN | Oncology_II | KAZN | Q674X7 | 1 | 13892792 | 15118043 | 0.91 | 0.77 | 1.07 | 2.55E-01 | 0.557 |
| OID20304 | SIRPA | Cardiometabolic | SIRPA | P78324 | 20 | 1894167 | 1940592 | 1.10 | 0.93 | 1.30 | 2.55E-01 | 0.557 |
| OID20757 | IL1R2 | Inflammation | IL1R2 | P27930 | 2 | 101991960 | 102028544 | 0.91 | 0.78 | 1.07 | 2.55E-01 | 0.558 |
| OID20975 | HNMT | Neurology | HNMT | P50135 | 2 | 137964020 | 138016364 | 0.90 | 0.76 | 1.08 | 2.56E-01 | 0.558 |
| OID20398 | IGFBP3 | Cardiometabolic | IGFBP3 | P17936 | 7 | 45912245 | 45921874 | 0.92 | 0.81 | 1.06 | 2.56E-01 | 0.559 |
| OID20974 | ADGRB3 | Neurology | ADGRB3 | O60242 | 6 | 68635282 | 69390571 | 1.10 | 0.93 | 1.29 | 2.60E-01 | 0.562 |
| OID21128 | ALDH1A1 | Neurology | ALDH1A1 | P00352 | 9 | 72900671 | 73080442 | 0.90 | 0.76 | 1.08 | 2.59E-01 | 0.562 |
| OID30620 | C1QTNF9 | Inflammation_II | C1QTNF9 | P0C862 | 13 | 24307166 | 24322535 | 0.91 | 0.78 | 1.07 | 2.60E-01 | 0.562 |
| OID20206 | CD69 | Cardiometabolic | CD69 | Q07108 | 12 | 9752486 | 9760901 | 0.91 | 0.78 | 1.07 | 2.62E-01 | 0.562 |
| OID21060 | CPA2 | Neurology | CPA2 | P48052 | 7 | 130266863 | 130289798 | 1.10 | 0.94 | 1.28 | 2.58E-01 | 0.562 |
| OID20897 | DPEP1 | Neurology | DPEP1 | P16444 | 16 | 89613308 | 89638456 | 1.10 | 0.93 | 1.31 | 2.61E-01 | 0.562 |
| OID20443 | FCRL3 | Inflammation | FCRL3 | Q96P31 | 1 | 157674321 | 157700769 | 0.91 | 0.78 | 1.07 | 2.61E-01 | 0.562 |
| OID30445 | FOXJ3 | Inflammation_II | FOXJ3 | Q9UPW0 | 1 | 42176539 | 42335877 | 0.90 | 0.75 | 1.08 | 2.61E-01 | 0.562 |
| OID20933 | GFRA3 | Neurology | GFRA3 | O60609 | 5 | 138252380 | 138274621 | 0.91 | 0.77 | 1.07 | 2.58E-01 | 0.562 |
| OID20887 | HMOX2 | Neurology | HMOX2 | P30519 | 16 | 4474690 | 4510347 | 0.90 | 0.75 | 1.08 | 2.59E-01 | 0.562 |
| OID30841 | IL13RA2 | Neurology_II | IL13RA2 | Q14627 | X | 115003975 | 115019977 | 0.91 | 0.78 | 1.07 | 2.59E-01 | 0.562 |
| OID31430 | ITIH5 | Oncology_II | ITIH5 | Q86UX2 | 10 | 7559270 | 7666998 | 0.91 | 0.77 | 1.07 | 2.62E-01 | 0.562 |
| OID31521 | NDUFB7 | Oncology_II | NDUFB7 | P17568 | 19 | 14566078 | 14572066 | 0.91 | 0.77 | 1.07 | 2.62E-01 | 0.562 |
| OID20465 | NRTN | Inflammation | NRTN | Q99748 | 19 | 5805067 | 5828324 | 0.90 | 0.76 | 1.08 | 2.58E-01 | 0.562 |
| OID20922 | PHOSPHO1 | Neurology | PHOSPHO1 | Q8TCT1 | 17 | 49223366 | 49230787 | 1.10 | 0.93 | 1.30 | 2.61E-01 | 0.562 |
| OID20146 | PRKAR1A | Cardiometabolic | PRKAR1A | P10644 | 17 | 68511780 | 68551319 | 0.91 | 0.78 | 1.07 | 2.60E-01 | 0.562 |
| OID21346 | RET | Oncology | RET | P07949 | 10 | 43077064 | 43130351 | 0.91 | 0.78 | 1.07 | 2.60E-01 | 0.562 |
| OID21409 | SLITRK2 | Oncology | SLITRK2 | Q9H156 | X | 145817829 | 145829856 | 1.10 | 0.93 | 1.29 | 2.58E-01 | 0.562 |
| OID20213 | SORT1 | Cardiometabolic | SORT1 | Q99523 | 1 | 109309568 | 109397918 | 0.91 | 0.77 | 1.07 | 2.61E-01 | 0.562 |
| OID30990 | SPACA5_SPACA5B | Neurology_II | PACA5B;SPACA5 | Q96QH8;Q96QH8 | X;X | 48130626;48004336 | 48132620;48009733 | 0.90 | 0.75 | 1.08 | 2.58E-01 | 0.562 |
| OID20894 | TIGAR | Neurology | TIGAR | Q9NQ88 | 12 | 4307763 | 4360028 | 0.91 | 0.76 | 1.08 | 2.59E-01 | 0.562 |
| OID31246 | TMOD4 | Oncology_II | TMOD4 | Q9NZQ9 | 1 | 151169986 | 151176284 | 1.09 | 0.94 | 1.27 | 2.62E-01 | 0.562 |
| OID30353 | TOMM20 | Cardiometabolic_II | TOMM20 | Q15388 | 1 | 235109341 | 235128837 | 0.91 | 0.77 | 1.07 | 2.59E-01 | 0.562 |
| OID31234 | TPRKB | Oncology_II | TPRKB | Q9Y3C4 | 2 | 73729104 | 73737400 | 1.08 | 0.94 | 1.23 | 2.62E-01 | 0.562 |
| OID31008 | GNAS | Neurology_II | GNAS | O95467 | 20 | 58839718 | 58911192 | 0.90 | 0.75 | 1.08 | 2.62E-01 | 0.562 |
| OID21014 | STC1 | Neurology | STC1 | P52823 | 8 | 23841929 | 23854806 | 1.10 | 0.93 | 1.29 | 2.63E-01 | 0.562 |
| OID20731 | TREM2 | Inflammation | TREM2 | Q9NZC2 | 6 | 41158506 | 41163186 | 1.10 | 0.93 | 1.30 | 2.63E-01 | 0.562 |
| OID30911 | CNTNAP4 | Neurology_II | CNTNAP4 | Q9C0A0 | 16 | 76277278 | 76560757 | 0.90 | 0.74 | 1.08 | 2.63E-01 | 0.562 |
| OID20381 | GP1BA | Cardiometabolic | GP1BA | P07359 | 17 | 4932277 | 4935023 | 0.91 | 0.78 | 1.07 | 2.65E-01 | 0.566 |
| OID21375 | SIRT2 | Oncology | SIRT2 | Q8IXJ6 | 19 | 38878555 | 38899862 | 0.91 | 0.78 | 1.07 | 2.65E-01 | 0.566 |
| OID30544 | FGF3 | Inflammation_II | FGF3 | P11487 | 11 | 69809968 | 69819416 | 1.09 | 0.94 | 1.26 | 2.66E-01 | 0.566 |
| OID20156 | MNDA | Cardiometabolic | MNDA | P41218 | 1 | 158831351 | 158849506 | 0.91 | 0.76 | 1.08 | 2.66E-01 | 0.566 |
| OID20725 | OMD | Inflammation | OMD | Q99983 | 9 | 92412380 | 92424471 | 1.10 | 0.93 | 1.31 | 2.66E-01 | 0.566 |
| OID31220 | SLC28A1 | Oncology_II | SLC28A1 | O00337 | 15 | 84884654 | 84945798 | 0.91 | 0.76 | 1.08 | 2.66E-01 | 0.566 |
| OID30195 | TOR1AIP1 | Cardiometabolic_II | TOR1AIP1 | Q5JTV8 | 1 | 179882042 | 179920077 | 0.90 | 0.74 | 1.09 | 2.66E-01 | 0.566 |
| OID31097 | GSTT2B | Neurology_II | GSTT2B | P0CG30 | 22 | 23957414 | 23961195 | 0.91 | 0.78 | 1.07 | 2.66E-01 | 0.566 |
| OID30569 | AFAP1 | Inflammation_II | AFAP1 | Q8N556 | 4 | 7758714 | 7939926 | 0.91 | 0.77 | 1.07 | 2.68E-01 | 0.567 |
| OID21255 | ARHGAP25 | Oncology | ARHGAP25 | P42331 | 2 | 68679601 | 68826833 | 0.91 | 0.76 | 1.08 | 2.68E-01 | 0.567 |
| OID21426 | CNTN2 | Oncology | CNTN2 | Q02246 | 1 | 205042937 | 205078289 | 0.91 | 0.76 | 1.08 | 2.67E-01 | 0.567 |
| OID21379 | GCNT1 | Oncology | GCNT1 | Q02742 | 9 | 76419850 | 76651203 | 0.91 | 0.77 | 1.08 | 2.68E-01 | 0.567 |
| OID20420 | IL10RA | Inflammation | IL10RA | Q13651 | 11 | 117986370 | 118003037 | 0.90 | 0.74 | 1.09 | 2.67E-01 | 0.567 |
| OID21290 | LPCAT2 | Oncology | LPCAT2 | Q7L5N7 | 16 | 55509072 | 55586666 | 0.88 | 0.71 | 1.10 | 2.68E-01 | 0.567 |
| OID31519 | PCYT2 | Oncology_II | PCYT2 | Q99447 | 17 | 81900958 | 81911432 | 0.91 | 0.77 | 1.07 | 2.68E-01 | 0.567 |
| OID30693 | PNLIP | Inflammation_II | PNLIP | P16233 | 10 | 116545931 | 116567855 | 0.92 | 0.80 | 1.06 | 2.69E-01 | 0.568 |
| OID20586 | LTA | Inflammation | LTA | P01374 | 6 | 31572054 | 31574324 | 0.91 | 0.77 | 1.08 | 2.70E-01 | 0.570 |
| OID21006 | CNTN5 | Neurology | CNTN5 | O94779 | 11 | 99020949 | 100358885 | 0.90 | 0.76 | 1.08 | 2.70E-01 | 0.571 |
| OID20847 | PTS | Neurology | PTS | Q03393 | 11 | 112226367 | 112269955 | 0.91 | 0.76 | 1.08 | 2.71E-01 | 0.571 |
| OID31275 | ALMS1 | Oncology_II | ALMS1 | Q8TCU4 | 2 | 73385758 | 73625166 | 0.90 | 0.75 | 1.09 | 2.71E-01 | 0.571 |
| OID30575 | BMP10 | Inflammation_II | BMP10 | O95393 | 2 | 68860909 | 68871397 | 0.91 | 0.78 | 1.07 | 2.72E-01 | 0.571 |
| OID30809 | GLP1R | Neurology_II | GLP1R | P43220 | 6 | 39048781 | 39091303 | 0.91 | 0.77 | 1.08 | 2.72E-01 | 0.571 |
| OID31351 | AMOTL2 | Oncology_II | AMOTL2 | Q9Y2J4 | 3 | 134355347 | 134375479 | 0.91 | 0.76 | 1.08 | 2.73E-01 | 0.572 |

| | | | | | | | | | | | | |
|---|---|---|---|---|---|---|---|---|---|---|---|---|
| OID30772 | CFI | Inflammation_II | CFI | P05156 | 4 | 109731008 | 109802150 | 0.90 | 0.74 | 1.09 | 2.72E-01 | 0.572 |
| OID20928 | CLEC10A | Neurology | CLEC10A | Q8IUN9 | 17 | 7074537 | 7080307 | 0.91 | 0.78 | 1.07 | 2.73E-01 | 0.572 |
| OID30581 | CST1 | Inflammation_II | CST1 | P01037 | 20 | 23747562 | 23751268 | 0.91 | 0.78 | 1.07 | 2.73E-01 | 0.572 |
| OID21507 | FGFBP1 | Oncology | FGFBP1 | Q14512 | 4 | 15935577 | 15938740 | 1.10 | 0.93 | 1.30 | 2.73E-01 | 0.572 |
| OID20813 | KCNIP4 | Neurology | KCNIP4 | Q6PIL6 | 4 | 20728606 | 21948772 | 0.90 | 0.74 | 1.09 | 2.73E-01 | 0.572 |
| OID21503 | SIAE | Oncology | SIAE | Q9HAT2 | 11 | 124633113 | 124695707 | 0.91 | 0.77 | 1.08 | 2.73E-01 | 0.572 |
| OID20654 | C1QA | Inflammation | C1QA | P02745 | 1 | 22636628 | 22639678 | 1.10 | 0.93 | 1.30 | 2.73E-01 | 0.572 |
| OID30952 | CAMLG | Neurology_II | CAMLG | P49069 | 5 | 134738495 | 134752157 | 0.90 | 0.75 | 1.08 | 2.75E-01 | 0.573 |
| OID31485 | CDC42BPB | Oncology_II | CDC42BPB | Q9Y5S2 | 14 | 102932380 | 103057549 | 0.91 | 0.77 | 1.08 | 2.76E-01 | 0.573 |
| OID20901 | CRIP2 | Neurology | CRIP2 | P52943 | 14 | 105472962 | 105480162 | 0.90 | 0.75 | 1.08 | 2.76E-01 | 0.573 |
| OID31437 | GPHA2 | Oncology_II | GPHA2 | Q96T91 | 11 | 64934471 | 64935893 | 0.90 | 0.73 | 1.09 | 2.76E-01 | 0.573 |
| OID30785 | PLG | Inflammation_II | PLG | P00747 | 6 | 160702238 | 160754097 | 0.90 | 0.75 | 1.08 | 2.74E-01 | 0.573 |
| OID31165 | PRAME | Oncology_II | PRAME | P78395 | 22 | 22547701 | 22559361 | 0.91 | 0.77 | 1.08 | 2.76E-01 | 0.573 |
| OID30672 | RNASE1 | Inflammation_II | RNASE1 | P07998 | 14 | 20801228 | 20802855 | 0.91 | 0.76 | 1.08 | 2.75E-01 | 0.573 |
| OID30857 | SCIN | Neurology_II | SCIN | Q9Y6U3 | 7 | 12570577 | 12660182 | 0.91 | 0.76 | 1.08 | 2.76E-01 | 0.573 |
| OID30442 | ST8SIA1 | Inflammation_II | ST8SIA1 | Q92185 | 12 | 22063773 | 22437041 | 0.89 | 0.73 | 1.10 | 2.76E-01 | 0.573 |
| OID21393 | ITGB7 | Oncology | ITGB7 | P26010 | 12 | 53191323 | 53207282 | 0.91 | 0.77 | 1.08 | 2.77E-01 | 0.574 |
| OID21322 | ST3GAL1 | Oncology | ST3GAL1 | Q11201 | 8 | 133454848 | 133571940 | 1.08 | 0.94 | 1.25 | 2.77E-01 | 0.574 |
| OID31127 | FNDC1 | Neurology_II | FNDC1 | Q4ZHG4 | 6 | 159169400 | 159272108 | 0.92 | 0.78 | 1.07 | 2.77E-01 | 0.575 |
| OID21189 | UXS1 | Oncology | UXS1 | Q8NBZ7 | 2 | 106093308 | 106194301 | 0.91 | 0.76 | 1.08 | 2.77E-01 | 0.575 |
| OID31075 | TAX1BP1 | Neurology_II | TAX1BP1 | Q86VP1 | 7 | 27739331 | 27844564 | 0.91 | 0.77 | 1.08 | 2.78E-01 | 0.575 |
| OID30265 | ZP3 | Cardiometabolic_II | ZP3 | P21754 | 7 | 76397518 | 76442071 | 0.92 | 0.79 | 1.07 | 2.78E-01 | 0.575 |
| OID21453 | SORD | Oncology | SORD | Q00796 | 15 | 45023147 | 45077185 | 0.91 | 0.77 | 1.08 | 2.79E-01 | 0.577 |
| OID30705 | RNASE4 | Inflammation_II | RNASE4 | P34096 | 14 | 20684560 | 20701216 | 1.09 | 0.93 | 1.27 | 2.79E-01 | 0.577 |
| OID30980 | ATXN2L | Neurology_II | ATXN2L | Q8WWM7 | 16 | 28822999 | 28837237 | 0.90 | 0.75 | 1.09 | 2.81E-01 | 0.581 |
| OID31074 | CDH23 | Neurology_II | CDH23 | Q9H251 | 10 | 71396920 | 71815947 | 0.91 | 0.76 | 1.08 | 2.82E-01 | 0.581 |
| OID30488 | AKR7L | Inflammation_II | AKR7L | Q8NHP1 | 1 | 19265982 | 19274194 | 0.91 | 0.76 | 1.08 | 2.83E-01 | 0.581 |
| OID20069 | CNPY2 | Cardiometabolic | CNPY2 | Q9Y2B0 | 12 | 56309842 | 56316119 | 1.08 | 0.94 | 1.24 | 2.82E-01 | 0.581 |
| OID21116 | IL1R1 | Neurology | IL1R1 | P14778 | 2 | 102064544 | 102179874 | 1.09 | 0.93 | 1.27 | 2.83E-01 | 0.581 |
| OID21186 | MZT1 | Oncology | MZT1 | Q08AG7 | 13 | 72708367 | 72727629 | 0.91 | 0.76 | 1.08 | 2.83E-01 | 0.581 |
| OID31045 | PCDH12 | Neurology_II | PCDH12 | Q9NPG4 | 5 | 141943581 | 141969741 | 0.91 | 0.77 | 1.08 | 2.82E-01 | 0.581 |
| OID20647 | CD160 | Inflammation | CD160 | O95971 | 1 | 145719471 | 145739288 | 0.92 | 0.78 | 1.07 | 2.84E-01 | 0.583 |
| OID21209 | CEP20 | Oncology | CEP20 | Q96NB1 | 16 | 15865719 | 15888625 | 0.91 | 0.77 | 1.08 | 2.84E-01 | 0.583 |
| OID20715 | FGF19 | Inflammation | FGF19 | O95750 | 11 | 69698238 | 69704022 | 1.09 | 0.93 | 1.29 | 2.84E-01 | 0.583 |
| OID30420 | DAND5 | Inflammation_II | DAND5 | Q8N907 | 19 | 12965159 | 12974760 | 1.09 | 0.93 | 1.27 | 2.85E-01 | 0.583 |
| OID31005 | FAM171A2 | Neurology_II | FAM171A2 | A8MVW0 | 17 | 44353215 | 44363853 | 0.91 | 0.76 | 1.08 | 2.85E-01 | 0.583 |
| OID20253 | NOTCH3 | Cardiometabolic | NOTCH3 | Q9UM47 | 19 | 15159038 | 15200995 | 1.10 | 0.92 | 1.31 | 2.85E-01 | 0.583 |
| OID30362 | CEACAM6 | Cardiometabolic_II | CEACAM6 | P40199 | 19 | 41750977 | 41772211 | 1.09 | 0.93 | 1.29 | 2.85E-01 | 0.583 |
| OID31218 | BCL7A | Oncology_II | BCL7A | Q4VC05 | 12 | 122019422 | 122062044 | 0.91 | 0.76 | 1.08 | 2.86E-01 | 0.585 |
| OID31116 | CHCHD10 | Neurology_II | CHCHD10 | Q8WYQ3 | 22 | 23765834 | 23767972 | 0.91 | 0.75 | 1.09 | 2.86E-01 | 0.585 |
| OID20535 | IL7 | Inflammation | IL7 | P13232 | 8 | 78675743 | 78805523 | 0.92 | 0.78 | 1.08 | 2.87E-01 | 0.586 |
| OID20471 | GALNT3 | Inflammation | GALNT3 | Q14435 | 2 | 165747588 | 165794659 | 1.09 | 0.93 | 1.28 | 2.88E-01 | 0.587 |
| OID20440 | IL24 | Inflammation | IL24 | Q13007 | 1 | 206897443 | 206904139 | 0.91 | 0.77 | 1.08 | 2.88E-01 | 0.587 |
| OID21498 | NT5E | Oncology | NT5E | P21589 | 6 | 85449584 | 85495791 | 0.92 | 0.78 | 1.08 | 2.88E-01 | 0.587 |
| OID20268 | PDGFRB | Cardiometabolic | PDGFRB | P09619 | 5 | 150113839 | 150155872 | 1.10 | 0.92 | 1.31 | 2.89E-01 | 0.588 |
| OID20270 | ESAM | Cardiometabolic | ESAM | Q96AP7 | 11 | 124752583 | 124762290 | 0.91 | 0.77 | 1.08 | 2.89E-01 | 0.588 |
| OID30156 | EIF5 | Cardiometabolic_II | EIF5 | P55010 | 14 | 103333544 | 103345025 | 1.08 | 0.93 | 1.25 | 2.91E-01 | 0.591 |
| OID30058 | TPM3 | Cardiometabolic_II | TPM3 | P06753 | 1 | 154155308 | 154194648 | 0.91 | 0.77 | 1.08 | 2.91E-01 | 0.591 |
| OID30603 | UROD | Inflammation_II | UROD | P06132 | 1 | 45010950 | 45015575 | 0.91 | 0.77 | 1.08 | 2.91E-01 | 0.591 |
| OID20109 | ADAM15 | Cardiometabolic | ADAM15 | Q13444 | 1 | 155050566 | 155062775 | 1.09 | 0.93 | 1.29 | 2.92E-01 | 0.593 |
| OID20603 | CD200 | Inflammation | CD200 | P41217 | 3 | 112332347 | 112362812 | 0.91 | 0.77 | 1.08 | 2.95E-01 | 0.595 |
| OID20940 | CDCP1 | Neurology | CDCP1 | Q9H5V8 | 3 | 45082277 | 45146422 | 1.10 | 0.92 | 1.30 | 2.95E-01 | 0.595 |
| OID21249 | CRACR2A | Oncology | CRACR2A | Q9BSW2 | 12 | 3606633 | 3764819 | 0.91 | 0.77 | 1.08 | 2.94E-01 | 0.595 |
| OID30534 | DEFB103A_DEFB103E | Inflammation_II | 3103B;DEFB103A | P81534;P81534 | 8;8 | 7428888;7881392 | 7430348;7882663 | 0.91 | 0.77 | 1.08 | 2.95E-01 | 0.595 |
| OID20245 | FABP4 | Cardiometabolic | FABP4 | P15090 | 8 | 81478419 | 81483236 | 0.89 | 0.72 | 1.11 | 2.97E-01 | 0.595 |
| OID20195 | GLRX | Cardiometabolic | GLRX | P35754 | 5 | 95751319 | 95822726 | 0.91 | 0.77 | 1.08 | 2.94E-01 | 0.595 |
| OID20578 | ICAM4 | Inflammation | ICAM4 | Q14773 | 19 | 10286955 | 10288522 | 1.09 | 0.93 | 1.29 | 2.94E-01 | 0.595 |
| OID20145 | ICAM5 | Cardiometabolic | ICAM5 | Q9UMF0 | 19 | 10289952 | 10296778 | 1.10 | 0.92 | 1.30 | 2.95E-01 | 0.595 |
| OID21196 | KIR2DL3 | Oncology | KIR2DL3 | P43628 | 19 | 54738513 | 54753052 | 1.09 | 0.93 | 1.27 | 2.97E-01 | 0.595 |
| OID21251 | LYPD8 | Oncology | LYPD8 | Q6UX82 | 1 | 248739415 | 248755759 | 1.08 | 0.93 | 1.25 | 2.96E-01 | 0.595 |
| OID30996 | MYCBP2 | Neurology_II | MYCBP2 | O75592 | 13 | 77042474 | 77327094 | 0.91 | 0.76 | 1.09 | 2.97E-01 | 0.595 |
| OID20130 | S100A11 | Cardiometabolic | S100A11 | P31949 | 1 | 152032506 | 152047907 | 0.90 | 0.74 | 1.10 | 2.95E-01 | 0.595 |
| OID30934 | STX1B | Neurology_II | STX1B | P61266 | 16 | 30989256 | 31010638 | 0.91 | 0.77 | 1.08 | 2.95E-01 | 0.595 |
| OID20600 | TGFA | Inflammation | TGFA | P01135 | 2 | 70447284 | 70554193 | 1.09 | 0.93 | 1.27 | 2.97E-01 | 0.595 |
| OID30982 | TPSD1 | Neurology_II | TPSD1 | Q9BZJ3 | 16 | 1256059 | 1259008 | 0.91 | 0.77 | 1.08 | 2.94E-01 | 0.595 |
| OID31271 | TRIM24 | Oncology_II | TRIM24 | O15164 | 7 | 138460259 | 138589996 | 0.91 | 0.77 | 1.08 | 2.96E-01 | 0.595 |
| OID20516 | VASH1 | Inflammation | VASH1 | Q7L8A9 | 14 | 76761468 | 76783015 | 0.91 | 0.77 | 1.08 | 2.96E-01 | 0.595 |
| OID30708 | APOL1 | Inflammation_II | APOL1 | O14791 | 22 | 36253071 | 36267530 | 0.91 | 0.76 | 1.09 | 2.98E-01 | 0.596 |
| OID30394 | CD248 | Cardiometabolic_II | CD248 | Q9HCU0 | 11 | 66314494 | 66317044 | 1.09 | 0.92 | 1.29 | 2.98E-01 | 0.596 |
| OID30927 | DOC2B | Neurology_II | DOC2B | Q14184 | 17 | 142789 | 181650 | 0.90 | 0.73 | 1.10 | 2.98E-01 | 0.596 |
| OID31177 | MAPRE3 | Oncology_II | MAPRE3 | Q9UPY8 | 2 | 26970637 | 27027219 | 0.91 | 0.75 | 1.09 | 2.97E-01 | 0.596 |
| OID30074 | MAP1LC3B2 | Cardiometabolic_II | MAP1LC3B2 | A6NCE7 | 12 | 116548105 | 116576606 | 0.91 | 0.76 | 1.09 | 2.98E-01 | 0.596 |
| OID21369 | RBP5 | Oncology | RBP5 | P82980 | 12 | 7115736 | 7128889 | 0.91 | 0.76 | 1.09 | 2.98E-01 | 0.596 |
| OID20996 | ASAH2 | Neurology | ASAH2 | Q9NR71 | 10 | 50182778 | 50279720 | 0.92 | 0.78 | 1.08 | 2.99E-01 | 0.597 |
| OID20934 | DKK4 | Neurology | DKK4 | Q9UBT3 | 8 | 42374063 | 42377229 | 0.91 | 0.77 | 1.08 | 2.99E-01 | 0.597 |
| OID30949 | AP2B1 | Neurology_II | AP2B1 | P63010 | 17 | 35578046 | 35726413 | 0.91 | 0.77 | 1.08 | 3.01E-01 | 0.597 |
| OID20780 | B4GALT1 | Inflammation | B4GALT1 | P15291 | 9 | 33104082 | 33167356 | 1.09 | 0.93 | 1.28 | 3.01E-01 | 0.597 |
| OID21010 | CD300C | Neurology | CD300C | Q08708 | 17 | 74541073 | 74546115 | 1.09 | 0.93 | 1.28 | 3.02E-01 | 0.597 |
| OID20099 | CLEC1A | Cardiometabolic | CLEC1A | Q8NC01 | 12 | 10069554 | 10111627 | 1.09 | 0.93 | 1.27 | 3.01E-01 | 0.597 |
| OID20449 | IL22RA1 | Inflammation | IL22RA1 | Q8N6P7 | 1 | 24119771 | 24143140 | 0.91 | 0.76 | 1.09 | 3.02E-01 | 0.597 |
| OID30906 | LRTM1 | Neurology_II | LRTM1 | Q9HBL6 | 3 | 54918231 | 54967088 | 1.07 | 0.94 | 1.21 | 3.00E-01 | 0.597 |
| OID30909 | OSTN | Neurology_II | OSTN | P61366 | 3 | 191199241 | 191265615 | 1.08 | 0.93 | 1.25 | 3.02E-01 | 0.597 |
| OID21184 | PPM1A | Oncology | PPM1A | P35813 | 14 | 60245752 | 60299087 | 0.91 | 0.77 | 1.08 | 3.01E-01 | 0.597 |
| OID30589 | TNFRSF17 | Inflammation_II | TNFRSF17 | Q02223 | 16 | 11965210 | 11968068 | 0.92 | 0.79 | 1.08 | 3.01E-01 | 0.597 |
| OID20120 | TP53INP1 | Cardiometabolic | TP53INP1 | Q96A56 | 8 | 94925972 | 94949378 | 1.08 | 0.94 | 1.23 | 3.02E-01 | 0.597 |
| OID30885 | TRIM40 | Neurology_II | TRIM40 | Q6P9F5 | 6 | 30136124 | 30148735 | 0.92 | 0.78 | 1.08 | 3.01E-01 | 0.597 |
| OID31281 | UPK3BL1 | Oncology_II | UPK3BL1 | B0FP48 | 7 | 102637025 | 102642791 | 0.90 | 0.73 | 1.10 | 3.01E-01 | 0.597 |
| OID21114 | SOD2 | Neurology | SOD2 | P04179 | 6 | 159669069 | 159762529 | 1.09 | 0.93 | 1.28 | 3.03E-01 | 0.598 |
| OID21162 | ANKRD54 | Oncology | ANKRD54 | Q6NXT1 | 22 | 37830855 | 37849327 | 0.91 | 0.77 | 1.09 | 3.04E-01 | 0.599 |
| OID30109 | GASK1A | Cardiometabolic_II | GASK1A | Q9UFP1 | 3 | 42979087 | 43067898 | 0.92 | 0.77 | 1.08 | 3.03E-01 | 0.599 |
| OID30632 | TP53I3 | Inflammation_II | TP53I3 | Q53FA7 | 2 | 24077433 | 24085861 | 0.92 | 0.77 | 1.08 | 3.03E-01 | 0.599 |
| OID30565 | NXPH3 | Inflammation_II | NXPH3 | O95157 | 17 | 49575871 | 49583827 | 0.92 | 0.78 | 1.08 | 3.04E-01 | 0.600 |
| OID21383 | ALPP | Oncology | ALPP | P05187 | 2 | 232378724 | 232382889 | 1.10 | 0.91 | 1.33 | 3.05E-01 | 0.600 |
| OID30932 | DIPK1C | Neurology_II | DIPK1C | Q0P6D2 | 18 | 74434775 | 74457944 | 0.91 | 0.76 | 1.09 | 3.05E-01 | 0.600 |
| OID20602 | SLAMF7 | Inflammation | SLAMF7 | Q9NQ25 | 1 | 160739057 | 160754821 | 1.09 | 0.93 | 1.27 | 3.05E-01 | 0.600 |
| OID31429 | TRIM58 | Oncology_II | TRIM58 | Q8NG06 | 1 | 247857187 | 247880138 | 0.92 | 0.78 | 1.08 | 3.06E-01 | 0.602 |
| OID20858 | MRPL46 | Neurology | MRPL46 | Q9H2W6 | 15 | 88459477 | 88467390 | 0.91 | 0.77 | 1.09 | 3.07E-01 | 0.603 |
| OID30696 | ACE | Inflammation_II | ACE | P12821 | 17 | 63477061 | 63498380 | 0.92 | 0.78 | 1.08 | 3.08E-01 | 0.603 |
| OID21467 | ADM | Oncology | ADM | P35318 | 11 | 10305073 | 10307397 | 1.11 | 0.91 | 1.35 | 3.08E-01 | 0.603 |
| OID30415 | ASAH1 | Cardiometabolic_II | ASAH1 | Q13510 | 8 | 18055992 | 18084998 | 0.92 | 0.78 | 1.08 | 3.09E-01 | 0.603 |
| OID20505 | BACH1 | Inflammation | BACH1 | O14867 | 21 | 29194071 | 29630751 | 0.92 | 0.78 | 1.08 | 3.09E-01 | 0.603 |
| OID30255 | DLL4 | Cardiometabolic_II | DLL4 | Q9NR61 | 15 | 40929340 | 40939073 | 0.89 | 0.72 | 1.11 | 3.08E-01 | 0.603 |
| OID30223 | DMD | Cardiometabolic_II | DMD | P11532 | X | 31097677 | 33339609 | 0.91 | 0.76 | 1.09 | 3.09E-01 | 0.603 |
| OID21354 | DPP6 | Oncology | DPP6 | P42658 | 7 | 153887097 | 154894285 | 0.92 | 0.78 | 1.08 | 3.09E-01 | 0.603 |
| OID30791 | IFNL2 | Neurology_II | IFNL2 | Q8IZJ0 | 19 | 39268396 | 39270188 | 0.91 | 0.76 | 1.09 | 3.09E-01 | 0.603 |
| OID30522 | NFAT5 | Inflammation_II | NFAT5 | O94916 | 16 | 69565094 | 69704666 | 0.91 | 0.77 | 1.09 | 3.08E-01 | 0.603 |
| OID21012 | TXNDC5 | Neurology | TXNDC5 | Q8NBS9 | 6 | 7881517 | 7910788 | 0.91 | 0.77 | 1.09 | 3.08E-01 | 0.603 |

| | | | | | | | | | | | | |
|---|---|---|---|---|---|---|---|---|---|---|---|---|
| OID21464 | WIF1 | Oncology | WIF1 | Q9Y5W5 | 12 | 65050626 | 65121305 | 1.09 | 0.92 | 1.28 | 3.09E-01 | 0.603 |
| OID20960 | FKBP4 | Neurology | FKBP4 | Q02790 | 12 | 2794970 | 2805423 | 0.92 | 0.78 | 1.08 | 3.10E-01 | 0.604 |
| OID30103 | TEF | Cardiometabolic_II | TEF | Q10587 | 22 | 41367333 | 41399326 | 0.91 | 0.76 | 1.09 | 3.10E-01 | 0.604 |
| OID20458 | CXCL14 | Inflammation | CXCL14 | O95715 | 5 | 135570679 | 135579279 | 0.90 | 0.73 | 1.10 | 3.11E-01 | 0.605 |
| OID20677 | EPHA1 | Inflammation | EPHA1 | P21709 | 7 | 143390289 | 143408856 | 0.91 | 0.76 | 1.09 | 3.11E-01 | 0.605 |
| OID20174 | QDPR | Cardiometabolic | QDPR | P09417 | 4 | 17460261 | 17512206 | 0.92 | 0.78 | 1.08 | 3.11E-01 | 0.605 |
| OID20087 | MMP7 | Cardiometabolic | MMP7 | P09237 | 11 | 102520508 | 102530750 | 1.09 | 0.92 | 1.30 | 3.12E-01 | 0.606 |
| OID21364 | TMPRSS15 | Oncology | TMPRSS15 | P98073 | 21 | 18269116 | 18485879 | 1.09 | 0.93 | 1.27 | 3.12E-01 | 0.606 |
| OID20745 | CCL17 | Inflammation | CCL17 | Q92583 | 16 | 57404767 | 57416063 | 0.92 | 0.79 | 1.08 | 3.14E-01 | 0.609 |
| OID20909 | GGT5 | Neurology | GGT5 | P36269 | 22 | 24219654 | 24245108 | 1.09 | 0.92 | 1.28 | 3.14E-01 | 0.609 |
| OID21191 | LTBP3 | Oncology | LTBP3 | Q9NS15 | 11 | 65538559 | 65558930 | 0.91 | 0.76 | 1.09 | 3.15E-01 | 0.609 |
| OID20204 | SOST | Cardiometabolic | SOST | Q9BQB4 | 17 | 43753738 | 43758791 | 0.92 | 0.77 | 1.09 | 3.14E-01 | 0.609 |
| OID31244 | MANEAL | Oncology_II | MANEAL | Q5VSG8 | 1 | 37793847 | 37801137 | 0.92 | 0.78 | 1.08 | 3.16E-01 | 0.610 |
| OID30501 | MCEMP1 | Inflammation_II | MCEMP1 | Q8IX19 | 19 | 7677088 | 7679829 | 0.92 | 0.78 | 1.08 | 3.16E-01 | 0.610 |
| OID20358 | CTSD | Cardiometabolic | CTSD | P07339 | 11 | 1752752 | 1764573 | 1.08 | 0.93 | 1.26 | 3.16E-01 | 0.611 |
| OID30576 | CD300A | Inflammation_II | CD300A | Q9UGN4 | 17 | 74466399 | 74484794 | 0.92 | 0.78 | 1.08 | 3.17E-01 | 0.612 |
| OID31292 | SHPK | Oncology_II | SHPK | Q9UHJ6 | 17 | 3608240 | 3636250 | 0.91 | 0.76 | 1.09 | 3.17E-01 | 0.612 |
| OID21456 | CRISP2 | Oncology | CRISP2 | P16562 | 6 | 49692358 | 49713590 | 0.91 | 0.77 | 1.09 | 3.17E-01 | 0.612 |
| OID30786 | SERPINA3 | Inflammation_II | SERPINA3 | P01011 | 14 | 94612384 | 94624055 | 1.09 | 0.92 | 1.29 | 3.17E-01 | 0.612 |
| OID21246 | ATG4A | Oncology | ATG4A | Q8WYN0 | X | 108091668 | 108154671 | 0.91 | 0.77 | 1.09 | 3.19E-01 | 0.613 |
| OID20883 | BRK1 | Neurology | BRK1 | Q8WUW1 | 3 | 10115675 | 10127190 | 1.08 | 0.93 | 1.26 | 3.19E-01 | 0.613 |
| OID20173 | GPNMB | Cardiometabolic | GPNMB | Q14956 | 7 | 23235967 | 23275108 | 0.92 | 0.79 | 1.08 | 3.19E-01 | 0.613 |
| OID20408 | PTGDS | Cardiometabolic | PTGDS | P41222 | 9 | 136975092 | 136981742 | 1.08 | 0.93 | 1.26 | 3.19E-01 | 0.613 |
| OID20906 | SKAP1 | Neurology | SKAP1 | Q86WV1 | 17 | 48133442 | 48430275 | 0.92 | 0.78 | 1.09 | 3.19E-01 | 0.613 |
| OID21313 | DKKL1 | Oncology | DKKL1 | Q9UK85 | 19 | 49361783 | 49375116 | 1.09 | 0.92 | 1.30 | 3.20E-01 | 0.614 |
| OID20532 | HLA-E | Inflammation | HLA-E | P13747 | 6 | 30489509 | 30494194 | 1.09 | 0.92 | 1.27 | 3.20E-01 | 0.614 |
| OID20476 | TPT1 | Inflammation | TPT1 | P13693 | 13 | 45333471 | 45341284 | 1.08 | 0.93 | 1.25 | 3.20E-01 | 0.614 |
| OID20152 | CGREF1 | Cardiometabolic | CGREF1 | Q99674 | 2 | 27098889 | 27119128 | 1.08 | 0.93 | 1.26 | 3.21E-01 | 0.615 |
| OID21263 | GCG | Oncology | GCG | P01275 | 2 | 162142882 | 162152404 | 0.92 | 0.78 | 1.09 | 3.21E-01 | 0.615 |
| OID20756 | AGER | Inflammation | AGER | Q15109 | 6 | 32180968 | 32184322 | 0.92 | 0.78 | 1.09 | 3.23E-01 | 0.615 |
| OID21110 | CD99 | Neurology | CD99 | P14209 | X | 2691187 | 2741309 | 0.91 | 0.75 | 1.10 | 3.22E-01 | 0.615 |
| OID31208 | CYTH3 | Oncology_II | CYTH3 | O43739 | 7 | 6161776 | 6272644 | 0.91 | 0.76 | 1.09 | 3.23E-01 | 0.615 |
| OID20264 | DKK3 | Cardiometabolic | DKK3 | Q9UBP4 | 11 | 11956207 | 12009769 | 1.08 | 0.92 | 1.27 | 3.22E-01 | 0.615 |
| OID20608 | RAB6A | Inflammation | RAB6A | P20340 | 11 | 73675638 | 73761137 | 0.92 | 0.78 | 1.09 | 3.22E-01 | 0.615 |
| OID30056 | SKIV2L | Cardiometabolic_II | SKIV2L | Q15477 | 6 | 31959116 | 31969818 | 1.07 | 0.93 | 1.23 | 3.22E-01 | 0.615 |
| OID30525 | BNIP3L | Inflammation_II | BNIP3L | O60238 | 8 | 26383054 | 26505636 | 0.91 | 0.76 | 1.09 | 3.24E-01 | 0.617 |
| OID20382 | FCN2 | Cardiometabolic | FCN2 | Q15485 | 9 | 134880810 | 134887523 | 0.92 | 0.78 | 1.09 | 3.24E-01 | 0.617 |
| OID31093 | PRSS53 | Neurology_II | PRSS53 | Q2L4Q9 | 16 | 31083437 | 31089628 | 1.09 | 0.92 | 1.30 | 3.24E-01 | 0.617 |
| OID30881 | AMIGO1 | Neurology_II | AMIGO1 | Q86WK6 | 1 | 109504178 | 109509738 | 0.91 | 0.76 | 1.10 | 3.25E-01 | 0.618 |
| OID20289 | CPB1 | Cardiometabolic | CPB1 | P15086 | 3 | 148791102 | 148860187 | 0.92 | 0.78 | 1.09 | 3.25E-01 | 0.618 |
| OID30812 | RANBP2 | Neurology_II | RANBP2 | P49792 | 2 | 108719482 | 108785810 | 0.92 | 0.78 | 1.09 | 3.25E-01 | 0.618 |
| OID20409 | CA1 | Cardiometabolic | CA1 | P00915 | 8 | 85327608 | 85379014 | 0.92 | 0.77 | 1.09 | 3.26E-01 | 0.619 |
| OID30066 | MED21 | Cardiometabolic_II | MED21 | Q13503 | 12 | 27022546 | 27066343 | 0.90 | 0.74 | 1.11 | 3.26E-01 | 0.619 |
| OID20592 | TNFSF11 | Inflammation | TNFSF11 | O14788 | 13 | 42562736 | 42608013 | 0.92 | 0.79 | 1.08 | 3.26E-01 | 0.619 |
| OID20215 | DDC | Cardiometabolic | DDC | P20711 | 7 | 50458436 | 50565405 | 0.92 | 0.78 | 1.08 | 3.27E-01 | 0.620 |
| OID20705 | ERBB3 | Inflammation | ERBB3 | P21860 | 12 | 56076799 | 56103505 | 0.92 | 0.78 | 1.09 | 3.28E-01 | 0.620 |
| OID30704 | PON1 | Inflammation_II | PON1 | P27169 | 7 | 95297676 | 95324532 | 0.92 | 0.78 | 1.08 | 3.27E-01 | 0.620 |
| OID20943 | SCARB2 | Neurology | SCARB2 | Q14108 | 4 | 76158737 | 76234536 | 1.08 | 0.92 | 1.27 | 3.28E-01 | 0.620 |
| OID20314 | ACP5 | Cardiometabolic | ACP5 | P13686 | 19 | 11574660 | 11579008 | 1.09 | 0.92 | 1.29 | 3.30E-01 | 0.624 |
| OID21415 | RSPO3 | Oncology | RSPO3 | Q9BXY4 | 6 | 127118671 | 127199481 | 0.92 | 0.78 | 1.09 | 3.32E-01 | 0.626 |
| OID20085 | IRAG2 | Cardiometabolic | IRAG2 | Q12912 | 12 | 25004342 | 25108335 | 0.92 | 0.78 | 1.09 | 3.33E-01 | 0.628 |
| OID20149 | PM20D1 | Cardiometabolic | PM20D1 | Q6GTS8 | 1 | 205828025 | 205850132 | 0.92 | 0.78 | 1.09 | 3.33E-01 | 0.629 |
| OID31369 | NDST1 | Oncology_II | NDST1 | P52848 | 5 | 150485818 | 150558211 | 0.92 | 0.77 | 1.09 | 3.34E-01 | 0.629 |
| OID21214 | SLAMF8 | Oncology | SLAMF8 | Q9P0V8 | 1 | 159826811 | 159837492 | 1.08 | 0.92 | 1.28 | 3.34E-01 | 0.629 |
| OID20931 | NAAA | Neurology | NAAA | Q02083 | 4 | 75913660 | 75941013 | 0.92 | 0.79 | 1.09 | 3.34E-01 | 0.629 |
| OID30341 | SGSH | Cardiometabolic_II | SGSH | P51688 | 17 | 80206716 | 80220923 | 0.92 | 0.78 | 1.09 | 3.35E-01 | 0.630 |
| OID30769 | APOA1 | Inflammation_II | APOA1 | P02647 | 11 | 116835751 | 116837622 | 0.91 | 0.74 | 1.11 | 3.35E-01 | 0.630 |
| OID31391 | ERN1 | Oncology_II | ERN1 | O75460 | 17 | 64039080 | 64130819 | 0.92 | 0.78 | 1.09 | 3.36E-01 | 0.631 |
| OID30071 | HIP1 | Cardiometabolic_II | HIP1 | O00291 | 7 | 75533298 | 75738962 | 0.91 | 0.74 | 1.11 | 3.36E-01 | 0.631 |
| OID20830 | MAPT | Neurology | MAPT | P10636 | 17 | 45894527 | 46028334 | 1.08 | 0.92 | 1.28 | 3.36E-01 | 0.631 |
| OID31099 | PTPRR | Neurology_II | PTPRR | Q15256 | 12 | 70638073 | 70920738 | 0.92 | 0.79 | 1.08 | 3.36E-01 | 0.631 |
| OID20256 | VWF | Cardiometabolic | VWF | P04275 | 12 | 5948877 | 6124770 | 1.08 | 0.92 | 1.28 | 3.36E-01 | 0.631 |
| OID21075 | GGT1 | Neurology | GGT1 | P19440 | 22 | 24594811 | 24629005 | 0.92 | 0.77 | 1.09 | 3.37E-01 | 0.632 |
| OID30545 | NECTIN1 | Inflammation_II | NECTIN1 | Q15223 | 11 | 119623408 | 119729200 | 1.08 | 0.93 | 1.25 | 3.38E-01 | 0.633 |
| OID21032 | SUMF2 | Neurology | SUMF2 | Q8NBJ7 | 7 | 56064002 | 56080670 | 0.92 | 0.78 | 1.09 | 3.38E-01 | 0.633 |
| OID31500 | CD86 | Oncology_II | CD86 | P42081 | 3 | 122055362 | 122121139 | 0.92 | 0.77 | 1.09 | 3.39E-01 | 0.633 |
| OID30870 | CORO6 | Neurology_II | CORO6 | Q6QEF8 | 17 | 29614756 | 29622912 | 0.92 | 0.77 | 1.09 | 3.40E-01 | 0.634 |
| OID20519 | METAP1D | Inflammation | METAP1D | Q6UB28 | 2 | 171999943 | 172082430 | 0.92 | 0.78 | 1.09 | 3.40E-01 | 0.635 |
| OID30816 | KIF20B | Neurology_II | KIF20B | Q96Q89 | 10 | 89701590 | 89774939 | 1.07 | 0.93 | 1.22 | 3.41E-01 | 0.636 |
| OID30967 | MAG | Neurology_II | MAG | P20916 | 19 | 35292125 | 35313807 | 0.92 | 0.79 | 1.09 | 3.41E-01 | 0.636 |
| OID21048 | SCARF2 | Neurology | SCARF2 | Q96GP6 | 22 | 20424584 | 20437826 | 1.09 | 0.92 | 1.29 | 3.41E-01 | 0.636 |
| OID20276 | CTSB | Cardiometabolic | CTSB | P07858 | 8 | 11842524 | 11869533 | 1.08 | 0.92 | 1.28 | 3.42E-01 | 0.636 |
| OID21111 | FCRL5 | Neurology | FCRL5 | Q96RD9 | 1 | 157513377 | 157552515 | 0.92 | 0.77 | 1.09 | 3.42E-01 | 0.636 |
| OID30499 | CEMIP2 | Inflammation_II | CEMIP2 | Q9UHN6 | 9 | 71683366 | 71816690 | 1.06 | 0.94 | 1.21 | 3.43E-01 | 0.637 |
| OID31279 | PPIE | Oncology_II | PPIE | Q9UNP9 | 1 | 39692182 | 39763914 | 0.91 | 0.75 | 1.11 | 3.44E-01 | 0.639 |
| OID21150 | CTRB1 | Neurology | CTRB1 | P17538 | 16 | 75218988 | 75226338 | 0.93 | 0.80 | 1.08 | 3.44E-01 | 0.639 |
| OID20839 | PBLD | Neurology | PBLD | P30039 | 10 | 68282660 | 68333049 | 0.92 | 0.78 | 1.09 | 3.44E-01 | 0.639 |
| OID20328 | CCL15 | Cardiometabolic | CCL15 | Q16663 | 17 | 35996440 | 36001553 | 1.08 | 0.92 | 1.27 | 3.46E-01 | 0.641 |
| OID21395 | HAO1 | Oncology | HAO1 | Q9UJM8 | 20 | 7882985 | 7940458 | 0.92 | 0.78 | 1.09 | 3.46E-01 | 0.641 |
| OID30595 | MENT | Inflammation_II | C1orf56 | Q9BUN1 | 1 | 151047751 | 151051986 | 1.09 | 0.91 | 1.29 | 3.46E-01 | 0.641 |
| OID20345 | VASN | Cardiometabolic | VASN | Q6EMK4 | 16 | 4371848 | 4383538 | 0.93 | 0.79 | 1.09 | 3.46E-01 | 0.641 |
| OID21355 | VPS37A | Oncology | VPS37A | Q8NEZ2 | 8 | 17246931 | 17302427 | 0.92 | 0.78 | 1.09 | 3.47E-01 | 0.642 |
| OID21497 | KDR | Oncology | KDR | P35968 | 4 | 55078481 | 55125595 | 0.92 | 0.78 | 1.09 | 3.47E-01 | 0.642 |
| OID30428 | TLR4 | Inflammation_II | TLR4 | O00206 | 9 | 117704175 | 117724735 | 0.92 | 0.78 | 1.09 | 3.48E-01 | 0.644 |
| OID30349 | ADGRF5 | Cardiometabolic_II | ADGRF5 | Q8IZF2 | 6 | 46852522 | 46954943 | 0.93 | 0.79 | 1.09 | 3.49E-01 | 0.644 |
| OID20885 | GKN1 | Neurology | GKN1 | Q9NS71 | 2 | 68974573 | 68980980 | 1.08 | 0.92 | 1.27 | 3.49E-01 | 0.644 |
| OID20133 | PDGFRA | Cardiometabolic | PDGFRA | P16234 | 4 | 54229280 | 54298245 | 1.08 | 0.92 | 1.27 | 3.49E-01 | 0.644 |
| OID30416 | LAMP1 | Cardiometabolic_II | LAMP1 | P11279 | 13 | 113297239 | 113323672 | 0.92 | 0.77 | 1.10 | 3.51E-01 | 0.646 |
| OID30462 | NPHS2 | Inflammation_II | NPHS2 | Q9NP85 | 1 | 179550539 | 179575952 | 1.08 | 0.92 | 1.26 | 3.51E-01 | 0.647 |
| OID30535 | FGF16 | Inflammation_II | FGF16 | O43320 | X | 77447389 | 77457278 | 0.92 | 0.77 | 1.10 | 3.52E-01 | 0.647 |
| OID20781 | LGALS9 | Inflammation | LGALS9 | O00182 | 17 | 27629798 | 27649560 | 0.92 | 0.77 | 1.10 | 3.52E-01 | 0.647 |
| OID30607 | PRR4 | Inflammation_II | PRR4 | Q16378 | 12 | 10845849 | 10849475 | 0.92 | 0.78 | 1.09 | 3.52E-01 | 0.647 |
| OID31194 | GIMAP8 | Oncology_II | GIMAP8 | Q8ND71 | 7 | 150450630 | 150479393 | 0.92 | 0.76 | 1.10 | 3.53E-01 | 0.649 |
| OID20334 | CCL16 | Cardiometabolic | CCL16 | O15467 | 17 | 35976493 | 35981497 | 1.09 | 0.91 | 1.31 | 3.54E-01 | 0.649 |
| OID20908 | PTPN1 | Neurology | PTPN1 | P18031 | 20 | 50510321 | 50585241 | 0.92 | 0.78 | 1.09 | 3.54E-01 | 0.649 |
| OID30334 | VIT | Cardiometabolic_II | VIT | Q6UXI7 | 2 | 36696690 | 36814794 | 0.92 | 0.78 | 1.09 | 3.54E-01 | 0.649 |
| OID20735 | TNFRSF11B | Inflammation | TNFRSF11B | O00300 | 8 | 118923557 | 118951885 | 1.08 | 0.91 | 1.29 | 3.54E-01 | 0.650 |
| OID30871 | ARHGEF5 | Neurology_II | ARHGEF5 | Q12774 | 7 | 144355288 | 144380632 | 1.07 | 0.92 | 1.25 | 3.56E-01 | 0.651 |
| OID20805 | SCARB1 | Neurology | SCARB1 | Q8WTV0 | 12 | 124776856 | 124882668 | 0.91 | 0.75 | 1.11 | 3.56E-01 | 0.652 |
| OID20468 | PRKCQ | Inflammation | PRKCQ | Q04759 | 10 | 6427143 | 6580301 | 1.07 | 0.93 | 1.23 | 3.58E-01 | 0.654 |
| OID30079 | ATP5F1D | Cardiometabolic_II | ATP5F1D | P30049 | 19 | 1241746 | 1244825 | 1.06 | 0.94 | 1.19 | 3.58E-01 | 0.655 |
| OID20634 | NFASC | Inflammation | NFASC | O94856 | 1 | 204828651 | 205022822 | 0.92 | 0.78 | 1.10 | 3.58E-01 | 0.655 |
| OID30742 | PGLYRP2 | Inflammation_II | PGLYRP2 | Q96PD5 | 19 | 15468645 | 15498956 | 0.93 | 0.79 | 1.09 | 3.59E-01 | 0.655 |
| OID21287 | AIFM1 | Oncology | AIFM1 | O95831 | X | 130124666 | 130165879 | 0.92 | 0.78 | 1.09 | 3.62E-01 | 0.655 |
| OID31056 | AZI2 | Neurology_II | AZI2 | Q9H6S1 | 3 | 28315003 | 28349050 | 0.92 | 0.78 | 1.10 | 3.63E-01 | 0.655 |
| OID21316 | CD38 | Oncology | CD38 | P28907 | 4 | 15778275 | 15853232 | 0.91 | 0.75 | 1.11 | 3.61E-01 | 0.655 |
| OID30784 | CFP | Inflammation_II | CFP | P27918 | X | 47623172 | 47630305 | 0.92 | 0.76 | 1.11 | 3.62E-01 | 0.655 |

| | | | | | | | | | | | | |
|---|---|---|---|---|---|---|---|---|---|---|---|---|
| OID20463 | FXYD5 | Inflammation | FXYD5 | Q96DB9 | 19 | 35154730 | 35169881 | 0.92 | 0.78 | 1.09 | 3.62E-01 | 0.655 |
| OID31432 | KIR2DS4 | Oncology_II | KIR2DS4 | P43632 | 19 | 54832676 | 54848569 | 0.93 | 0.78 | 1.09 | 3.59E-01 | 0.655 |
| OID30208 | M6PR | Cardiometabolic_II | M6PR | P20645 | 12 | 8940361 | 8949761 | 1.08 | 0.92 | 1.26 | 3.61E-01 | 0.655 |
| OID31492 | MTSS1 | Oncology_II | MTSS1 | O43312 | 8 | 124550784 | 124728473 | 0.93 | 0.79 | 1.09 | 3.61E-01 | 0.655 |
| OID30377 | MUC2 | Cardiometabolic_II | MUC2 | Q02817 | 11 | 1074875 | 1110511 | 1.08 | 0.92 | 1.26 | 3.59E-01 | 0.655 |
| OID21121 | MYOC | Neurology | MYOC | Q99972 | 1 | 171635417 | 171652688 | 0.92 | 0.78 | 1.10 | 3.61E-01 | 0.655 |
| OID30520 | NDUFA5 | Inflammation_II | NDUFA5 | Q16718 | 7 | 123536997 | 123557904 | 0.93 | 0.78 | 1.09 | 3.61E-01 | 0.655 |
| OID21047 | OXT | Neurology | OXT | P01178 | 20 | 3071620 | 3072517 | 0.92 | 0.78 | 1.10 | 3.61E-01 | 0.655 |
| OID31467 | PHLDB1 | Oncology_II | PHLDB1 | Q86UU1 | 11 | 118606440 | 118658031 | 0.92 | 0.77 | 1.10 | 3.61E-01 | 0.655 |
| OID30913 | RBP1 | Neurology_II | RBP1 | P09455 | 3 | 139517434 | 139539829 | 0.92 | 0.77 | 1.10 | 3.61E-01 | 0.655 |
| OID30726 | SAA4 | Inflammation_II | SAA4 | P35542 | 11 | 18231355 | 18236802 | 0.88 | 0.67 | 1.16 | 3.60E-01 | 0.655 |
| OID30926 | SLC1A4 | Neurology_II | SLC1A4 | P43007 | 2 | 64988477 | 65023865 | 0.92 | 0.76 | 1.10 | 3.59E-01 | 0.655 |
| OID30559 | TOP2B | Inflammation_II | TOP2B | Q02880 | 3 | 25597978 | 25664909 | 0.92 | 0.78 | 1.09 | 3.62E-01 | 0.655 |
| OID21382 | VAT1 | Oncology | VAT1 | Q99536 | 17 | 43014607 | 43025123 | 1.08 | 0.92 | 1.27 | 3.62E-01 | 0.655 |
| OID31414 | GABARAP | Oncology_II | GABARAP | O95166 | 17 | 7240008 | 7242449 | 1.07 | 0.92 | 1.25 | 3.64E-01 | 0.657 |
| OID30527 | CSF3R | Inflammation_II | CSF3R | Q99062 | 1 | 36466043 | 36483278 | 0.92 | 0.78 | 1.10 | 3.65E-01 | 0.658 |
| OID20262 | CD209 | Cardiometabolic | CD209 | Q9NNX6 | 19 | 7739993 | 7747564 | 1.08 | 0.91 | 1.28 | 3.65E-01 | 0.659 |
| OID20976 | CX3CL1 | Neurology | CX3CL1 | P78423 | 16 | 57372477 | 57385044 | 1.08 | 0.92 | 1.26 | 3.66E-01 | 0.659 |
| OID30668 | PRDX2 | Inflammation_II | PRDX2 | P32119 | 19 | 12796820 | 12801800 | 0.92 | 0.77 | 1.10 | 3.66E-01 | 0.659 |
| OID31405 | PXDNL | Oncology_II | PXDNL | A1KZ92 | 8 | 51319577 | 51809445 | 0.92 | 0.76 | 1.11 | 3.66E-01 | 0.659 |
| OID20363 | AXL | Cardiometabolic | AXL | P30530 | 19 | 41219223 | 41261766 | 1.07 | 0.92 | 1.25 | 3.68E-01 | 0.660 |
| OID30093 | BRD3 | Cardiometabolic_II | BRD3 | Q15059 | 9 | 134030305 | 134068535 | 0.91 | 0.75 | 1.11 | 3.68E-01 | 0.660 |
| OID21182 | CAMKK1 | Oncology | CAMKK1 | Q8N5S9 | 17 | 3860315 | 3894891 | 0.93 | 0.78 | 1.10 | 3.68E-01 | 0.660 |
| OID21472 | CD33 | Oncology | CD33 | P20138 | 19 | 51225064 | 51243860 | 1.08 | 0.92 | 1.27 | 3.67E-01 | 0.660 |
| OID31253 | GABARAPL1 | Oncology_II | GABARAPL1 | Q9H0R8 | 12 | 10212458 | 10223128 | 0.92 | 0.77 | 1.10 | 3.68E-01 | 0.660 |
| OID31222 | SHC1 | Oncology_II | SHC1 | P29353 | 1 | 154962298 | 154974395 | 0.93 | 0.78 | 1.09 | 3.68E-01 | 0.660 |
| OID20739 | SIRPB1 | Inflammation | SIRPB1 | O00241 | 20 | 1561385 | 1620061 | 1.08 | 0.92 | 1.26 | 3.68E-01 | 0.660 |
| OID20272 | TNFSF13B | Cardiometabolic | TNFSF13B | Q9Y275 | 13 | 108251240 | 108308484 | 1.08 | 0.92 | 1.26 | 3.68E-01 | 0.660 |
| OID21332 | SCAMP3 | Oncology | SCAMP3 | O14828 | 1 | 155255979 | 155262430 | 0.93 | 0.78 | 1.10 | 3.69E-01 | 0.661 |
| OID21342 | EPHA2 | Oncology | EPHA2 | P29317 | 1 | 16124337 | 16156069 | 1.07 | 0.92 | 1.25 | 3.70E-01 | 0.661 |
| OID31268 | OFD1 | Oncology_II | OFD1 | O75665 | X | 13734743 | 13777955 | 1.07 | 0.92 | 1.24 | 3.70E-01 | 0.661 |
| OID30944 | DDX53 | Neurology_II | DDX53 | Q86TM3 | X | 22999960 | 23003589 | 1.07 | 0.92 | 1.26 | 3.72E-01 | 0.664 |
| OID20529 | FCRL6 | Inflammation | FCRL6 | Q6DN72 | 1 | 159800511 | 159816257 | 0.93 | 0.80 | 1.09 | 3.73E-01 | 0.664 |
| OID20782 | FSTL3 | Inflammation | FSTL3 | O95633 | 19 | 676392 | 683392 | 1.08 | 0.91 | 1.27 | 3.73E-01 | 0.664 |
| OID30829 | RAB3GAP1 | Neurology_II | RAB3GAP1 | Q15042 | 2 | 135052289 | 135176667 | 0.91 | 0.75 | 1.11 | 3.73E-01 | 0.664 |
| OID20190 | SIGLEC7 | Cardiometabolic | SIGLEC7 | Q9Y286 | 19 | 51142299 | 51153526 | 0.92 | 0.78 | 1.10 | 3.72E-01 | 0.664 |
| OID30798 | TJP3 | Neurology_II | TJP3 | O95049 | 19 | 3708362 | 3750813 | 1.07 | 0.92 | 1.24 | 3.72E-01 | 0.664 |
| OID21195 | CA11 | Oncology | CA11 | O75493 | 19 | 48637946 | 48646187 | 0.92 | 0.75 | 1.11 | 3.74E-01 | 0.666 |
| OID30281 | B3GNT7 | Cardiometabolic_II | B3GNT7 | Q8NFL0 | 2 | 231395710 | 231401164 | 0.92 | 0.78 | 1.10 | 3.75E-01 | 0.666 |
| OID20339 | CD59 | Cardiometabolic | CD59 | P13987 | 11 | 33703010 | 33736479 | 1.07 | 0.92 | 1.25 | 3.75E-01 | 0.666 |
| OID31392 | KIAA2013 | Oncology_II | KIAA2013 | Q8IYS2 | 1 | 11919591 | 11926428 | 0.91 | 0.75 | 1.12 | 3.75E-01 | 0.667 |
| OID31094 | KIAA0319 | Neurology_II | KIAA0319 | Q5VV43 | 6 | 24544104 | 24646191 | 0.93 | 0.79 | 1.10 | 3.77E-01 | 0.669 |
| OID30732 | CLU | Inflammation_II | CLU | P10909 | 8 | 27596917 | 27614700 | 0.90 | 0.70 | 1.14 | 3.78E-01 | 0.670 |
| OID20261 | FUCA1 | Cardiometabolic | FUCA1 | P04066 | 1 | 23845077 | 23868290 | 0.93 | 0.79 | 1.09 | 3.78E-01 | 0.670 |
| OID21486 | NBL1 | Oncology | NBL1 | P41271 | 1 | 19596979 | 19658456 | 1.07 | 0.92 | 1.26 | 3.78E-01 | 0.670 |
| OID20429 | LILRB4 | Inflammation | LILRB4 | Q8NHJ6 | 19 | 54643889 | 54670359 | 1.08 | 0.91 | 1.28 | 3.78E-01 | 0.670 |
| OID20432 | ARNT | Inflammation | ARNT | P27540 | 1 | 150809713 | 150876708 | 0.92 | 0.77 | 1.11 | 3.79E-01 | 0.670 |
| OID21347 | PPME1 | Oncology | PPME1 | Q9Y570 | 11 | 74171267 | 74254703 | 0.93 | 0.79 | 1.10 | 3.79E-01 | 0.670 |
| OID20073 | RCOR1 | Cardiometabolic | RCOR1 | Q9UKL0 | 14 | 102592649 | 102730561 | 0.92 | 0.76 | 1.11 | 3.80E-01 | 0.670 |
| OID20820 | SIRT5 | Neurology | SIRT5 | Q9NXA8 | 6 | 13574227 | 13615158 | 0.92 | 0.77 | 1.10 | 3.80E-01 | 0.670 |
| OID30369 | SLC4A1 | Cardiometabolic_II | SLC4A1 | P02730 | 17 | 44248390 | 44268141 | 1.08 | 0.91 | 1.27 | 3.80E-01 | 0.670 |
| OID21042 | CXCL11 | Neurology | CXCL11 | O14625 | 4 | 76033682 | 76041415 | 0.92 | 0.78 | 1.10 | 3.80E-01 | 0.671 |
| OID30615 | BMPER | Inflammation_II | BMPER | Q8N8U9 | 7 | 33904308 | 34156427 | 0.93 | 0.79 | 1.10 | 3.81E-01 | 0.672 |
| OID20803 | ARID4B | Neurology | ARID4B | Q4LE39 | 1 | 235131634 | 235328219 | 0.91 | 0.74 | 1.12 | 3.82E-01 | 0.672 |
| OID31496 | EGFLAM | Oncology_II | EGFLAM | Q63HQ2 | 5 | 38258409 | 38465480 | 0.93 | 0.78 | 1.10 | 3.82E-01 | 0.672 |
| OID21089 | IL17RA | Neurology | IL17RA | Q96F46 | 22 | 17084954 | 17115694 | 1.08 | 0.91 | 1.27 | 3.82E-01 | 0.672 |
| OID20297 | SCARF1 | Cardiometabolic | SCARF1 | Q14162 | 17 | 1633858 | 1645744 | 0.93 | 0.78 | 1.10 | 3.82E-01 | 0.672 |
| OID30502 | TGFBR1 | Inflammation_II | TGFBR1 | P36897 | 9 | 99104038 | 99154192 | 0.93 | 0.78 | 1.10 | 3.82E-01 | 0.672 |
| OID30065 | ATP1B3 | Cardiometabolic_II | ATP1B3 | P54709 | 3 | 141876124 | 141926549 | 0.92 | 0.77 | 1.11 | 3.83E-01 | 0.672 |
| OID30385 | PPIF | Cardiometabolic_II | PPIF | P30405 | 10 | 79347469 | 79355334 | 0.92 | 0.78 | 1.10 | 3.83E-01 | 0.672 |
| OID30293 | ECHDC3 | Cardiometabolic_II | ECHDC3 | Q96DC8 | 10 | 11742366 | 11764070 | 0.93 | 0.78 | 1.10 | 3.84E-01 | 0.673 |
| OID20876 | GSTP1 | Neurology | GSTP1 | P09211 | 11 | 67583742 | 67586656 | 0.93 | 0.80 | 1.09 | 3.84E-01 | 0.673 |
| OID30433 | RICTOR | Inflammation_II | RICTOR | Q6R327 | 5 | 38937920 | 39074399 | 0.92 | 0.76 | 1.11 | 3.84E-01 | 0.673 |
| OID20392 | LCN2 | Cardiometabolic | LCN2 | P80188 | 9 | 128149071 | 128153453 | 0.93 | 0.79 | 1.10 | 3.84E-01 | 0.673 |
| OID20422 | IL20RA | Inflammation | IL20RA | Q9UHF4 | 6 | 136999971 | 137045180 | 1.07 | 0.91 | 1.26 | 3.86E-01 | 0.677 |
| OID21270 | TPMT | Oncology | TPMT | P51580 | 6 | 18128311 | 18155077 | 0.93 | 0.78 | 1.10 | 3.87E-01 | 0.677 |
| OID31310 | IFIT1 | Oncology_II | IFIT1 | P09914 | 10 | 89392546 | 89406487 | 0.93 | 0.79 | 1.10 | 3.87E-01 | 0.677 |
| OID20052 | CEBPB | Cardiometabolic | CEBPB | P17676 | 20 | 50190830 | 50192668 | 0.93 | 0.78 | 1.10 | 3.88E-01 | 0.678 |
| OID30078 | FDX1 | Cardiometabolic_II | FDX1 | P10109 | 11 | 110429948 | 110464884 | 0.92 | 0.77 | 1.11 | 3.88E-01 | 0.678 |
| OID20809 | IL34 | Neurology | IL34 | Q6ZMJ4 | 16 | 70579895 | 70660682 | 1.08 | 0.91 | 1.27 | 3.88E-01 | 0.678 |
| OID31329 | CENPF | Oncology_II | CENPF | P49454 | 1 | 214603195 | 214664571 | 0.93 | 0.77 | 1.10 | 3.89E-01 | 0.678 |
| OID30724 | SERPINA7 | Inflammation_II | SERPINA7 | P05543 | X | 106032435 | 106038727 | 0.93 | 0.79 | 1.10 | 3.89E-01 | 0.679 |
| OID31505 | IFI30 | Oncology_II | IFI30 | P13284 | 19 | 18173162 | 18178117 | 0.92 | 0.77 | 1.11 | 3.91E-01 | 0.681 |
| OID31192 | CLASP1 | Oncology_II | CLASP1 | Q7Z460 | 2 | 121337776 | 121649476 | 0.92 | 0.77 | 1.11 | 3.92E-01 | 0.681 |
| OID20119 | EPHX2 | Cardiometabolic | EPHX2 | P34913 | 8 | 27490781 | 27545564 | 0.92 | 0.77 | 1.11 | 3.92E-01 | 0.681 |
| OID31288 | SEPTIN7 | Oncology_II | SEPTIN7 | Q16181 | 7 | 35800932 | 35907110 | 0.92 | 0.76 | 1.12 | 3.92E-01 | 0.681 |
| OID30701 | APOF | Inflammation_II | APOF | Q13790 | 12 | 56360568 | 56362857 | 1.10 | 0.89 | 1.36 | 3.93E-01 | 0.682 |
| OID21152 | LILRA2 | Neurology | LILRA2 | Q8N149 | 19 | 54572920 | 54590287 | 1.08 | 0.91 | 1.28 | 3.93E-01 | 0.682 |
| OID30149 | PROCR | Cardiometabolic_II | PROCR | Q9UNN8 | 20 | 35172072 | 35216240 | 0.91 | 0.74 | 1.12 | 3.93E-01 | 0.682 |
| OID30850 | ATXN2 | Neurology_II | ATXN2 | Q99700 | 12 | 111443485 | 111599676 | 1.07 | 0.92 | 1.24 | 3.94E-01 | 0.682 |
| OID30389 | BGLAP | Cardiometabolic_II | BGLAP | P02818 | 1 | 156242184 | 156243317 | 0.94 | 0.80 | 1.09 | 3.94E-01 | 0.682 |
| OID20915 | CSF2RA | Neurology | CSF2RA | P15509 | X | 1268800 | 1310381 | 0.93 | 0.79 | 1.10 | 3.94E-01 | 0.682 |
| OID30133 | GAMT | Cardiometabolic_II | GAMT | Q14353 | 19 | 1397026 | 1401570 | 1.05 | 0.94 | 1.18 | 3.94E-01 | 0.682 |
| OID31058 | PSMD1 | Neurology_II | PSMD1 | Q99460 | 2 | 231056845 | 231173116 | 0.93 | 0.79 | 1.10 | 3.94E-01 | 0.682 |
| OID20624 | TNFSF12 | Inflammation | TNFSF12 | O43508 | 17 | 7549058 | 7557890 | 0.93 | 0.79 | 1.10 | 3.94E-01 | 0.682 |
| OID20372 | F7 | Cardiometabolic | F7 | P08709 | 13 | 113105788 | 113120685 | 0.93 | 0.79 | 1.10 | 3.96E-01 | 0.684 |
| OID30780 | GC | Inflammation_II | GC | P02774 | 4 | 71741696 | 71804041 | 0.93 | 0.79 | 1.10 | 3.96E-01 | 0.684 |
| OID20456 | IL17F | Inflammation | IL17F | Q96PD4 | 6 | 52236681 | 52244500 | 1.06 | 0.92 | 1.23 | 3.97E-01 | 0.684 |
| OID20521 | TBC1D5 | Inflammation | TBC1D5 | Q92609 | 3 | 17157162 | 18444817 | 0.93 | 0.78 | 1.10 | 3.96E-01 | 0.684 |
| OID30956 | AIDA | Neurology_II | AIDA | Q96BJ3 | 1 | 222668013 | 222713210 | 0.93 | 0.78 | 1.11 | 3.98E-01 | 0.685 |
| OID30803 | GABRA4 | Neurology_II | GABRA4 | P48169 | 4 | 46918900 | 46993581 | 0.93 | 0.79 | 1.10 | 3.98E-01 | 0.685 |
| OID21139 | ITGA5 | Neurology | ITGA5 | P08648 | 12 | 54395261 | 54419266 | 0.93 | 0.78 | 1.10 | 3.98E-01 | 0.685 |
| OID30540 | NPHS1 | Inflammation_II | NPHS1 | O60500 | 19 | 35825372 | 35869287 | 0.93 | 0.79 | 1.10 | 3.98E-01 | 0.685 |
| OID20287 | ENG | Cardiometabolic | ENG | P17813 | 9 | 127815013 | 127854658 | 1.07 | 0.91 | 1.26 | 3.99E-01 | 0.686 |
| OID30090 | RBM19 | Cardiometabolic_II | RBM19 | Q9Y4C8 | 12 | 113816738 | 113966325 | 1.08 | 0.90 | 1.29 | 3.99E-01 | 0.686 |
| OID21253 | MAP3K5 | Oncology | MAP3K5 | Q99683 | 6 | 136557046 | 136792477 | 0.93 | 0.79 | 1.10 | 3.99E-01 | 0.686 |
| OID21483 | CD1C | Oncology | CD1C | P29017 | 1 | 158289923 | 158294774 | 0.93 | 0.80 | 1.10 | 4.01E-01 | 0.688 |
| OID31373 | EPGN | Oncology_II | EPGN | Q6UW88 | 4 | 74308470 | 74316789 | 1.07 | 0.92 | 1.25 | 4.01E-01 | 0.688 |
| OID30249 | CALCOCO2 | Cardiometabolic_II | CALCOCO2 | Q13137 | 17 | 48831018 | 48866522 | 0.93 | 0.79 | 1.10 | 4.03E-01 | 0.690 |
| OID30997 | PPP1R14D | Neurology_II | PPP1R14D | Q9NXH3 | 15 | 40815451 | 40828709 | 0.94 | 0.80 | 1.09 | 4.03E-01 | 0.690 |
| OID20614 | PCDH1 | Inflammation | PCDH1 | Q08174 | 5 | 141853090 | 141879246 | 1.07 | 0.92 | 1.24 | 4.05E-01 | 0.692 |
| OID21177 | RARRES1 | Oncology | RARRES1 | P49788 | 3 | 158696892 | 158732489 | 0.92 | 0.77 | 1.11 | 4.04E-01 | 0.692 |
| OID21028 | SMPD1 | Neurology | SMPD1 | P17405 | 11 | 6390440 | 6394998 | 1.07 | 0.91 | 1.26 | 4.04E-01 | 0.692 |
| OID30287 | TMEM132A | Cardiometabolic_II | TMEM132A | Q24JP5 | 11 | 60924460 | 60937159 | 0.93 | 0.78 | 1.10 | 4.05E-01 | 0.692 |
| OID20702 | TNFRSF13B | Inflammation | TNFRSF13B | O14836 | 17 | 16929816 | 16972118 | 0.93 | 0.79 | 1.10 | 4.04E-01 | 0.692 |
| OID20066 | GRK5 | Cardiometabolic | GRK5 | P34947 | 10 | 119207571 | 119459745 | 0.93 | 0.77 | 1.11 | 4.06E-01 | 0.693 |
| OID30096 | UBQLN3 | Cardiometabolic_II | UBQLN3 | Q9H347 | 11 | 5507300 | 5509958 | 1.06 | 0.93 | 1.21 | 4.06E-01 | 0.694 |

| | | | | | | | | | | | | |
|---|---|---|---|---|---|---|---|---|---|---|---|---|
| OID31282 | SLK | Oncology_II | SLK | Q9H2G2 | 10 | 103967140 | 104029233 | 0.92 | 0.75 | 1.12 | 4.07E-01 | 0.694 |
| OID30905 | SOX9 | Neurology_II | SOX9 | P48436 | 17 | 72121020 | 72126416 | 0.94 | 0.80 | 1.09 | 4.07E-01 | 0.694 |
| OID20910 | CLPP | Neurology | CLPP | Q16740 | 19 | 6361531 | 6370242 | 0.93 | 0.79 | 1.10 | 4.07E-01 | 0.694 |
| OID31092 | CLSTN3 | Neurology_II | CLSTN3 | Q9BQT9 | 12 | 7129698 | 7158945 | 0.93 | 0.78 | 1.10 | 4.08E-01 | 0.694 |
| OID20416 | GGH | Cardiometabolic | GGH | Q92820 | 8 | 63014881 | 63039407 | 1.07 | 0.91 | 1.26 | 4.07E-01 | 0.694 |
| OID30912 | PDRG1 | Neurology_II | PDRG1 | Q9NUG6 | 20 | 31944337 | 31952046 | 1.06 | 0.92 | 1.23 | 4.08E-01 | 0.694 |
| OID20917 | DRAXIN | Neurology | DRAXIN | Q8NBI3 | 1 | 11691710 | 11725857 | 1.07 | 0.91 | 1.25 | 4.09E-01 | 0.695 |
| OID21203 | RANGAP1 | Oncology | RANGAP1 | P46060 | 22 | 41244779 | 41286187 | 0.93 | 0.78 | 1.11 | 4.09E-01 | 0.695 |
| OID31314 | CENPJ | Oncology_II | CENPJ | Q9HC77 | 13 | 24882279 | 24922889 | 0.93 | 0.78 | 1.11 | 4.10E-01 | 0.696 |
| OID20935 | EFNA4 | Neurology | EFNA4 | P52798 | 1 | 155063737 | 155069553 | 1.07 | 0.91 | 1.25 | 4.10E-01 | 0.696 |
| OID20575 | HSD11B1 | Inflammation | HSD11B1 | P28845 | 1 | 209686178 | 209734949 | 1.07 | 0.91 | 1.26 | 4.10E-01 | 0.696 |
| OID20315 | ITGB2 | Cardiometabolic | ITGB2 | P05107 | 21 | 44885953 | 44931989 | 0.93 | 0.79 | 1.10 | 4.10E-01 | 0.696 |
| OID21109 | SPINT1 | Neurology | SPINT1 | O43278 | 15 | 40844018 | 40858207 | 1.07 | 0.91 | 1.25 | 4.12E-01 | 0.698 |
| OID31265 | CA7 | Oncology_II | CA7 | P43166 | 16 | 66844414 | 66854153 | 0.93 | 0.79 | 1.10 | 4.13E-01 | 0.698 |
| OID30538 | MTDH | Inflammation_II | MTDH | Q86UE4 | 8 | 97644184 | 97730260 | 0.93 | 0.79 | 1.10 | 4.13E-01 | 0.698 |
| OID21007 | ROBO2 | Neurology | ROBO2 | Q9HCK4 | 3 | 75906695 | 77649964 | 0.93 | 0.79 | 1.10 | 4.13E-01 | 0.698 |
| OID21504 | IL13RA1 | Oncology | IL13RA1 | P78552 | X | 118727133 | 118794535 | 1.07 | 0.91 | 1.27 | 4.14E-01 | 0.699 |
| OID30876 | SPTBN2 | Neurology_II | SPTBN2 | O15020 | 11 | 66682497 | 66744670 | 0.93 | 0.78 | 1.11 | 4.13E-01 | 0.699 |
| OID20972 | BAG3 | Neurology | BAG3 | O95817 | 10 | 119651380 | 119677819 | 0.93 | 0.80 | 1.10 | 4.14E-01 | 0.699 |
| OID20994 | BMP4 | Neurology | BMP4 | P12644 | 14 | 53949736 | 53958761 | 1.07 | 0.91 | 1.27 | 4.14E-01 | 0.699 |
| OID30406 | CPQ | Cardiometabolic_II | CPQ | Q9Y646 | 8 | 96645242 | 97149654 | 0.93 | 0.78 | 1.11 | 4.14E-01 | 0.699 |
| OID20200 | FABP2 | Cardiometabolic | FABP2 | P12104 | 4 | 119317250 | 119322138 | 1.07 | 0.91 | 1.25 | 4.15E-01 | 0.699 |
| OID20419 | IL2 | Inflammation | IL2 | P60568 | 4 | 122451470 | 122456725 | 0.93 | 0.79 | 1.10 | 4.15E-01 | 0.699 |
| OID31019 | SNAPIN | Neurology_II | SNAPIN | O95295 | 1 | 153658703 | 153661852 | 0.93 | 0.79 | 1.10 | 4.15E-01 | 0.699 |
| OID20359 | TNC | Cardiometabolic | TNC | P24821 | 9 | 115019575 | 115118257 | 1.07 | 0.91 | 1.25 | 4.17E-01 | 0.701 |
| OID30918 | PSMD5 | Neurology_II | PSMD5 | Q16401 | 9 | 120815496 | 120842951 | 0.93 | 0.78 | 1.11 | 4.17E-01 | 0.701 |
| OID21211 | RUVBL1 | Oncology | RUVBL1 | Q9Y265 | 3 | 128064778 | 128153914 | 1.06 | 0.92 | 1.22 | 4.17E-01 | 0.701 |
| OID21079 | DSC2 | Neurology | DSC2 | Q02487 | 18 | 31058840 | 31102522 | 1.07 | 0.91 | 1.26 | 4.18E-01 | 0.701 |
| OID20760 | ATP5IF1 | Inflammation | ATP5IF1 | Q9UII2 | 1 | 28236109 | 28246906 | 0.94 | 0.80 | 1.10 | 4.19E-01 | 0.702 |
| OID21435 | CPVL | Oncology | CPVL | Q9H3G5 | 7 | 28995235 | 29195451 | 1.07 | 0.91 | 1.27 | 4.18E-01 | 0.702 |
| OID31138 | MAP2 | Neurology_II | MAP2 | P11137 | 2 | 209424047 | 209734147 | 0.93 | 0.78 | 1.11 | 4.18E-01 | 0.702 |
| OID21103 | MMP9 | Neurology | MMP9 | P14780 | 20 | 46008908 | 46016561 | 0.94 | 0.80 | 1.10 | 4.18E-01 | 0.702 |
| OID21494 | NELL1 | Oncology | NELL1 | Q92832 | 11 | 20669551 | 21575686 | 0.94 | 0.81 | 1.09 | 4.19E-01 | 0.702 |
| OID20159 | ACAN | Cardiometabolic | ACAN | P16112 | 15 | 88803436 | 88875353 | 1.07 | 0.90 | 1.28 | 4.19E-01 | 0.702 |
| OID31483 | ADGRE1 | Oncology_II | ADGRE1 | Q14246 | 19 | 6887566 | 6940459 | 0.93 | 0.79 | 1.10 | 4.19E-01 | 0.702 |
| OID30146 | ABCA2 | Cardiometabolic_II | ABCA2 | Q9BZC7 | 9 | 137007234 | 137028915 | 1.07 | 0.91 | 1.25 | 4.21E-01 | 0.702 |
| OID21151 | F11R | Neurology | F11R | Q9Y624 | 1 | 160995211 | 161021343 | 0.93 | 0.79 | 1.10 | 4.21E-01 | 0.702 |
| OID20490 | FGF5 | Inflammation | FGF5 | P12034 | 4 | 80266639 | 80336680 | 1.07 | 0.90 | 1.28 | 4.20E-01 | 0.702 |
| OID20904 | MAD1L1 | Neurology | MAD1L1 | Q9Y6D9 | 7 | 1815793 | 2233243 | 0.92 | 0.76 | 1.12 | 4.21E-01 | 0.702 |
| OID31383 | LTB | Oncology_II | LTB | Q06643 | 6 | 31580525 | 31582522 | 0.93 | 0.78 | 1.11 | 4.21E-01 | 0.703 |
| OID30756 | CLEC3B | Inflammation_II | CLEC3B | P05452 | 3 | 45001548 | 45036071 | 0.93 | 0.79 | 1.11 | 4.22E-01 | 0.703 |
| OID31169 | IGLON5 | Oncology_II | IGLON5 | A6NGN9 | 19 | 51311644 | 51330891 | 0.93 | 0.78 | 1.11 | 4.23E-01 | 0.705 |
| OID31226 | PDXDC1 | Oncology_II | PDXDC1 | Q6P996 | 16 | 14974591 | 15139339 | 0.92 | 0.74 | 1.13 | 4.23E-01 | 0.705 |
| OID31352 | S100A3 | Oncology_II | S100A3 | P33764 | 1 | 153547329 | 153549258 | 0.92 | 0.74 | 1.14 | 4.24E-01 | 0.706 |
| OID20954 | MASP1 | Neurology | MASP1 | P48740 | 3 | 187217282 | 187291980 | 0.94 | 0.80 | 1.10 | 4.24E-01 | 0.706 |
| OID20132 | PCDH17 | Cardiometabolic | PCDH17 | O14917 | 13 | 57631744 | 57729311 | 1.07 | 0.91 | 1.25 | 4.25E-01 | 0.707 |
| OID31141 | HIP1R | Neurology_II | HIP1R | O75146 | 12 | 122834453 | 122862961 | 0.93 | 0.77 | 1.12 | 4.26E-01 | 0.708 |
| OID21057 | NTRK3 | Neurology | NTRK3 | Q16288 | 15 | 87859751 | 88256768 | 1.08 | 0.90 | 1.29 | 4.26E-01 | 0.708 |
| OID30567 | RALB | Inflammation_II | RALB | P11234 | 2 | 120240064 | 120294710 | 0.93 | 0.79 | 1.11 | 4.27E-01 | 0.709 |
| OID30365 | ISM2 | Cardiometabolic_II | ISM2 | Q6H9L7 | 14 | 77474394 | 77498816 | 0.93 | 0.79 | 1.11 | 4.27E-01 | 0.709 |
| OID21159 | GRN | Neurology | GRN | P28799 | 17 | 44345246 | 44353106 | 0.93 | 0.79 | 1.11 | 4.28E-01 | 0.709 |
| OID20122 | AK1 | Cardiometabolic | AK1 | P00568 | 9 | 127866486 | 127877675 | 0.93 | 0.79 | 1.11 | 4.28E-01 | 0.710 |
| OID31146 | PLSCR3 | Neurology_II | PLSCR3 | Q9NRY6 | 17 | 7389727 | 7394842 | 0.94 | 0.79 | 1.10 | 4.29E-01 | 0.711 |
| OID30081 | UGDH | Cardiometabolic_II | UGDH | O60701 | 4 | 39498755 | 39528311 | 0.93 | 0.79 | 1.11 | 4.29E-01 | 0.711 |
| OID21076 | BCAM | Neurology | BCAM | P50895 | 19 | 44809071 | 44821421 | 1.07 | 0.91 | 1.26 | 4.31E-01 | 0.712 |
| OID30849 | DEFB104A_DEFB104E | Neurology_II | 3104B;DEFB104A | Q8WTQ1;Q8WTQ1 | 8;8 | 7470308;7836436 | 7475082;7841242 | 0.92 | 0.75 | 1.13 | 4.31E-01 | 0.712 |
| OID31108 | FGFBP3 | Neurology_II | FGFBP3 | Q8TAT2 | 10 | 91906584 | 91909486 | 0.94 | 0.80 | 1.10 | 4.30E-01 | 0.712 |
| OID31229 | IL3 | Oncology_II | IL3 | P08700 | 5 | 132060655 | 132063204 | 1.07 | 0.91 | 1.26 | 4.32E-01 | 0.712 |
| OID21201 | KLK12 | Oncology | KLK12 | Q9UKR0 | 19 | 51029092 | 51035230 | 1.07 | 0.91 | 1.25 | 4.32E-01 | 0.712 |
| OID30897 | SLC44A4 | Neurology_II | SLC44A4 | Q53GD3 | 6 | 31863192 | 31879046 | 1.06 | 0.92 | 1.22 | 4.31E-01 | 0.712 |
| OID31267 | TCP11 | Oncology_II | TCP11 | Q8WWU5 | 6 | 35118071 | 35148610 | 1.06 | 0.92 | 1.21 | 4.31E-01 | 0.712 |
| OID30805 | CNGB3 | Neurology_II | CNGB3 | Q9NQW8 | 8 | 86553977 | 86743675 | 0.93 | 0.78 | 1.11 | 4.32E-01 | 0.712 |
| OID20356 | OSMR | Cardiometabolic | OSMR | Q99650 | 5 | 38845858 | 38945596 | 1.07 | 0.91 | 1.26 | 4.33E-01 | 0.713 |
| OID20454 | RGS8 | Inflammation | RGS8 | P57771 | 1 | 182641816 | 182684576 | 0.93 | 0.78 | 1.11 | 4.34E-01 | 0.714 |
| OID21241 | CEP85 | Oncology | CEP85 | Q6P2H3 | 1 | 26234200 | 26278808 | 0.93 | 0.79 | 1.11 | 4.34E-01 | 0.714 |
| OID20452 | AOC1 | Inflammation | AOC1 | P19801 | 7 | 150824627 | 150861504 | 0.93 | 0.79 | 1.11 | 4.35E-01 | 0.715 |
| OID20630 | SERPINB8 | Inflammation | SERPINB8 | P50452 | 18 | 63970029 | 64019779 | 0.93 | 0.77 | 1.12 | 4.35E-01 | 0.715 |
| OID30840 | CACNA1C | Neurology_II | CACNA1C | Q13936 | 12 | 1970772 | 2697950 | 0.93 | 0.78 | 1.11 | 4.36E-01 | 0.715 |
| OID30731 | C1S | Inflammation_II | C1S | P09871 | 12 | 6988259 | 7071032 | 0.93 | 0.78 | 1.11 | 4.36E-01 | 0.716 |
| OID20243 | CDH5 | Cardiometabolic | CDH5 | P33151 | 16 | 66366622 | 66404784 | 1.06 | 0.91 | 1.25 | 4.38E-01 | 0.718 |
| OID30290 | GGACT | Cardiometabolic_II | GGACT | Q9BVM4 | 13 | 100530164 | 100589528 | 0.94 | 0.79 | 1.11 | 4.38E-01 | 0.718 |
| OID21252 | EGFL7 | Oncology | EGFL7 | Q9UHF1 | 9 | 136658856 | 136672678 | 0.94 | 0.79 | 1.11 | 4.39E-01 | 0.719 |
| OID20528 | ITGA6 | Inflammation | ITGA6 | P23229 | 2 | 172427354 | 172506459 | 0.94 | 0.79 | 1.11 | 4.39E-01 | 0.719 |
| OID30210 | IGSF21 | Cardiometabolic_II | IGSF21 | Q96ID5 | 1 | 18107798 | 18378483 | 0.93 | 0.78 | 1.11 | 4.40E-01 | 0.719 |
| OID30598 | MFAP4 | Inflammation_II | MFAP4 | P55083 | 17 | 19383442 | 19387190 | 1.07 | 0.90 | 1.27 | 4.40E-01 | 0.719 |
| OID30245 | MYL4 | Cardiometabolic_II | MYL4 | P12829 | 17 | 47200446 | 47223679 | 0.93 | 0.78 | 1.12 | 4.40E-01 | 0.719 |
| OID20278 | DPT | Cardiometabolic | DPT | Q07507 | 1 | 168695468 | 168729206 | 1.08 | 0.89 | 1.31 | 4.41E-01 | 0.720 |
| OID30238 | RNASE10 | Cardiometabolic_II | RNASE10 | Q5GAN6 | 14 | 20505537 | 20513884 | 1.07 | 0.90 | 1.26 | 4.41E-01 | 0.720 |
| OID21496 | TEK | Oncology | TEK | Q02763 | 9 | 27109141 | 27230174 | 1.07 | 0.91 | 1.25 | 4.41E-01 | 0.720 |
| OID30153 | SPINK8 | Cardiometabolic_II | SPINK8 | P0C7L1 | 3 | 48306842 | 48333661 | 0.93 | 0.78 | 1.12 | 4.42E-01 | 0.720 |
| OID30382 | MYH9 | Cardiometabolic_II | MYH9 | P35579 | 22 | 36281280 | 36388010 | 0.94 | 0.80 | 1.11 | 4.44E-01 | 0.723 |
| OID30471 | ADAMTS4 | Inflammation_II | ADAMTS4 | O75173 | 1 | 161184302 | 161199054 | 0.93 | 0.78 | 1.11 | 4.45E-01 | 0.724 |
| OID31190 | ARHGAP5 | Oncology_II | ARHGAP5 | Q13017 | 14 | 32076114 | 32159728 | 0.94 | 0.79 | 1.11 | 4.45E-01 | 0.724 |
| OID20366 | CHL1 | Cardiometabolic | CHL1 | O00533 | 3 | 196763 | 409417 | 1.06 | 0.91 | 1.23 | 4.44E-01 | 0.724 |
| OID31284 | ID4 | Oncology_II | ID4 | P47928 | 6 | 19837370 | 19842197 | 0.93 | 0.76 | 1.13 | 4.45E-01 | 0.724 |
| OID30116 | NPR1 | Cardiometabolic_II | NPR1 | P16066 | 1 | 153678688 | 153693992 | 1.07 | 0.90 | 1.26 | 4.45E-01 | 0.724 |
| OID20082 | ZBTB17 | Cardiometabolic | ZBTB17 | Q13105 | 1 | 15941869 | 15976132 | 0.93 | 0.78 | 1.12 | 4.45E-01 | 0.724 |
| OID20638 | LAMP3 | Inflammation | LAMP3 | Q9UQV4 | 3 | 183122215 | 183163839 | 1.07 | 0.89 | 1.29 | 4.46E-01 | 0.724 |
| OID30916 | CABP2 | Neurology_II | CABP2 | Q9NPB3 | 11 | 67518912 | 67524517 | 0.93 | 0.79 | 1.11 | 4.48E-01 | 0.727 |
| OID20088 | PON2 | Cardiometabolic | PON2 | Q15165 | 7 | 95404862 | 95435329 | 0.94 | 0.80 | 1.10 | 4.49E-01 | 0.728 |
| OID20990 | ASGR1 | Neurology | ASGR1 | P07306 | 17 | 7173431 | 7179564 | 0.94 | 0.79 | 1.11 | 4.49E-01 | 0.728 |
| OID20426 | IL4 | Inflammation | IL4 | P05112 | 5 | 132673986 | 132682678 | 1.07 | 0.90 | 1.26 | 4.50E-01 | 0.728 |
| OID20338 | PGLYRP1 | Cardiometabolic | PGLYRP1 | O75594 | 19 | 46019153 | 46023053 | 0.93 | 0.78 | 1.11 | 4.50E-01 | 0.728 |
| OID21168 | GH2 | Oncology | GH2 | P01242 | 17 | 63880215 | 63881944 | 0.94 | 0.80 | 1.11 | 4.50E-01 | 0.728 |
| OID21310 | VNN2 | Oncology | VNN2 | O95498 | 6 | 132743870 | 132763459 | 0.94 | 0.80 | 1.10 | 4.51E-01 | 0.729 |
| OID20590 | CLEC4G | Inflammation | CLEC4G | Q6UXB4 | 19 | 7728957 | 7733906 | 0.94 | 0.79 | 1.11 | 4.53E-01 | 0.731 |
| OID21045 | RGMB | Neurology | RGMB | Q6NW40 | 5 | 98768650 | 98798643 | 0.94 | 0.81 | 1.10 | 4.52E-01 | 0.731 |
| OID30832 | HDAC9 | Neurology_II | HDAC9 | Q9UKV0 | 7 | 18086949 | 19002416 | 0.94 | 0.80 | 1.10 | 4.53E-01 | 0.732 |
| OID21038 | CHGB | Neurology | CHGB | P05060 | 20 | 5911510 | 5925353 | 0.94 | 0.80 | 1.10 | 4.54E-01 | 0.732 |
| OID31464 | COL24A1 | Oncology_II | COL24A1 | Q17RW2 | 1 | 85729233 | 86156943 | 0.94 | 0.81 | 1.10 | 4.54E-01 | 0.732 |
| OID30977 | GRIK2 | Neurology_II | GRIK2 | Q13002 | 6 | 100962701 | 102081622 | 0.94 | 0.80 | 1.10 | 4.54E-01 | 0.732 |
| OID20753 | MEPE | Inflammation | MEPE | Q9NQ76 | 4 | 87821398 | 87846814 | 0.94 | 0.80 | 1.10 | 4.55E-01 | 0.732 |
| OID21493 | TGFBR2 | Oncology | TGFBR2 | P37173 | 3 | 30606601 | 30694142 | 1.06 | 0.91 | 1.25 | 4.55E-01 | 0.732 |
| OID30085 | RYR1 | Cardiometabolic_II | RYR1 | P21817 | 19 | 38433691 | 38595273 | 1.05 | 0.93 | 1.19 | 4.55E-01 | 0.732 |
| OID30971 | TEX101 | Neurology_II | TEX101 | Q9BY14 | 19 | 43401496 | 43418597 | 0.91 | 0.72 | 1.16 | 4.55E-01 | 0.733 |
| OID30601 | RLN2 | Inflammation_II | RLN2 | P04090 | 9 | 5299864 | 5304716 | 0.88 | 0.63 | 1.23 | 4.56E-01 | 0.734 |
| OID30902 | CRTAP | Neurology_II | CRTAP | O75718 | 3 | 33114014 | 33147773 | 0.94 | 0.79 | 1.11 | 4.57E-01 | 0.734 |

| | | | | | | | | | | | | |
|---|---|---|---|---|---|---|---|---|---|---|---|---|
| OID20100 | ENTPD6 | Cardiometabolic | ENTPD6 | O75354 | 20 | 25195693 | 25228075 | 1.07 | 0.90 | 1.26 | 4.57E-01 | 0.734 |
| OID30443 | FGF6 | Inflammation_II | FGF6 | P10767 | 12 | 4428155 | 4445815 | 0.93 | 0.78 | 1.12 | 4.57E-01 | 0.734 |
| OID30878 | NLGN2 | Neurology_II | NLGN2 | Q8NFZ4 | 17 | 7404874 | 7419860 | 0.94 | 0.80 | 1.11 | 4.57E-01 | 0.734 |
| OID31277 | ACADM | Oncology_II | ACADM | P11310 | 1 | 75724431 | 75787575 | 1.05 | 0.92 | 1.21 | 4.59E-01 | 0.736 |
| OID31015 | TMEM25 | Neurology_II | TMEM25 | Q86YD3 | 11 | 118531041 | 118547280 | 1.06 | 0.90 | 1.25 | 4.59E-01 | 0.736 |
| OID30384 | TNFSF8 | Cardiometabolic_II | TNFSF8 | P32971 | 9 | 114893343 | 114930595 | 1.06 | 0.91 | 1.24 | 4.59E-01 | 0.736 |
| OID21525 | DLL1 | Oncology | DLL1 | O00548 | 6 | 170282206 | 170306565 | 1.06 | 0.90 | 1.25 | 4.61E-01 | 0.737 |
| OID30152 | ENOX2 | Cardiometabolic_II | ENOX2 | Q16206 | X | 130622325 | 130903234 | 0.93 | 0.78 | 1.12 | 4.61E-01 | 0.737 |
| OID20778 | FABP1 | Inflammation | FABP1 | P07148 | 2 | 88122982 | 88128062 | 1.07 | 0.90 | 1.26 | 4.61E-01 | 0.737 |
| OID31232 | H2AP | Oncology_II | H2AP | O75409 | X | 37990779 | 37991314 | 1.06 | 0.91 | 1.23 | 4.61E-01 | 0.737 |
| OID31098 | HSD17B14 | Neurology_II | HSD17B14 | Q9BPX1 | 19 | 48813018 | 48836510 | 1.06 | 0.90 | 1.25 | 4.62E-01 | 0.737 |
| OID20666 | IL12B | Inflammation | IL12B | P29460 | 5 | 159314780 | 159330487 | 1.06 | 0.90 | 1.25 | 4.60E-01 | 0.737 |
| OID20866 | MUC13 | Neurology | MUC13 | Q9H3R2 | 3 | 124905442 | 124953819 | 0.94 | 0.80 | 1.10 | 4.61E-01 | 0.737 |
| OID20141 | TGM2 | Cardiometabolic | TGM2 | P21980 | 20 | 38127385 | 38166578 | 0.94 | 0.79 | 1.11 | 4.62E-01 | 0.737 |
| OID21064 | JAM2 | Neurology | JAM2 | P57087 | 21 | 25639258 | 25717562 | 0.94 | 0.80 | 1.10 | 4.63E-01 | 0.738 |
| OID30110 | MTR | Cardiometabolic_II | MTR | Q99707 | 1 | 236795260 | 236921278 | 0.94 | 0.80 | 1.11 | 4.63E-01 | 0.738 |
| OID30890 | CLIC5 | Neurology_II | CLIC5 | Q9NZA1 | 6 | 45880827 | 46080348 | 0.94 | 0.79 | 1.11 | 4.64E-01 | 0.738 |
| OID20400 | CST3 | Cardiometabolic | CST3 | P01034 | 20 | 23626706 | 23638473 | 0.94 | 0.80 | 1.11 | 4.64E-01 | 0.738 |
| OID20754 | CDON | Inflammation | CDON | Q4KMG0 | 11 | 125955796 | 126063335 | 0.94 | 0.80 | 1.11 | 4.65E-01 | 0.739 |
| OID31241 | FGF7 | Oncology_II | FGF7 | P21781 | 15 | 49423237 | 49488775 | 0.94 | 0.79 | 1.12 | 4.65E-01 | 0.739 |
| OID20348 | IL18BP | Cardiometabolic | IL18BP | O95998 | 11 | 71998613 | 72005715 | 1.06 | 0.91 | 1.24 | 4.65E-01 | 0.739 |
| OID20128 | LEPR | Cardiometabolic | LEPR | P48357 | 1 | 65420652 | 65641559 | 0.94 | 0.79 | 1.12 | 4.65E-01 | 0.739 |
| OID30475 | IL31RA | Inflammation_II | IL31RA | Q8NI17 | 5 | 55851357 | 55922854 | 0.94 | 0.79 | 1.11 | 4.66E-01 | 0.740 |
| OID21188 | ADCYAP1R1 | Oncology | ADCYAP1R1 | P41586 | 7 | 31052308 | 31111479 | 0.93 | 0.76 | 1.13 | 4.67E-01 | 0.740 |
| OID20970 | LAYN | Neurology | LAYN | Q6UX15 | 11 | 111540280 | 111561745 | 1.06 | 0.91 | 1.24 | 4.68E-01 | 0.740 |
| OID20324 | LILRB5 | Cardiometabolic | LILRB5 | O75023 | 19 | 54249421 | 54257301 | 0.94 | 0.81 | 1.10 | 4.68E-01 | 0.740 |
| OID30811 | SNAP25 | Neurology_II | SNAP25 | P60880 | 20 | 10172395 | 10308258 | 1.06 | 0.90 | 1.25 | 4.67E-01 | 0.740 |
| OID21050 | THY1 | Neurology | THY1 | P04216 | 11 | 119415476 | 119424985 | 0.94 | 0.80 | 1.11 | 4.67E-01 | 0.740 |
| OID20951 | MDGA1 | Neurology | MDGA1 | Q8NFP4 | 6 | 37630679 | 37699306 | 0.94 | 0.80 | 1.11 | 4.69E-01 | 0.742 |
| OID20396 | VCAM1 | Cardiometabolic | VCAM1 | P19320 | 1 | 100719742 | 100739045 | 1.06 | 0.90 | 1.24 | 4.70E-01 | 0.744 |
| OID30457 | PRR5 | Inflammation_II | PRR5 | P85299 | 22 | 44668547 | 44737681 | 0.93 | 0.78 | 1.12 | 4.71E-01 | 0.744 |
| OID20949 | GLB1 | Neurology | GLB1 | P16278 | 3 | 32996609 | 33097202 | 0.94 | 0.79 | 1.12 | 4.71E-01 | 0.744 |
| OID30180 | C1GALT1C1 | Cardiometabolic_II | C1GALT1C1 | Q96EU7 | X | 120625674 | 120630054 | 0.94 | 0.79 | 1.12 | 4.72E-01 | 0.745 |
| OID30250 | EPPK1 | Cardiometabolic_II | EPPK1 | P58107 | 8 | 143857324 | 143878467 | 0.94 | 0.79 | 1.12 | 4.72E-01 | 0.745 |
| OID20661 | FLT3LG | Inflammation | FLT3LG | P49771 | 19 | 49474207 | 49486231 | 0.94 | 0.79 | 1.12 | 4.72E-01 | 0.745 |
| OID20284 | LGALS3 | Cardiometabolic | LGALS3 | P17931 | 14 | 55124110 | 55145423 | 1.06 | 0.90 | 1.26 | 4.73E-01 | 0.745 |
| OID20421 | PAPPA | Inflammation | PAPPA | Q13219 | 9 | 116153791 | 116402321 | 0.94 | 0.80 | 1.11 | 4.73E-01 | 0.745 |
| OID21148 | SPINK5 | Neurology | SPINK5 | Q9NQ38 | 5 | 148025683 | 148137382 | 1.06 | 0.90 | 1.24 | 4.73E-01 | 0.745 |
| OID30189 | VSIG10L | Cardiometabolic_II | VSIG10L | Q86VR7 | 19 | 51331536 | 51342139 | 0.94 | 0.79 | 1.11 | 4.73E-01 | 0.745 |
| OID30852 | TMPRSS11B | Neurology_II | TMPRSS11B | Q86T26 | 4 | 68226653 | 68245694 | 1.06 | 0.91 | 1.23 | 4.74E-01 | 0.746 |
| OID30072 | CSF2 | Cardiometabolic_II | CSF2 | P04141 | 5 | 132073789 | 132076170 | 1.06 | 0.90 | 1.25 | 4.75E-01 | 0.746 |
| OID21336 | UBAC1 | Oncology | UBAC1 | Q9BSL1 | 9 | 135932969 | 135961373 | 0.94 | 0.79 | 1.12 | 4.75E-01 | 0.747 |
| OID30275 | ADAMTSL4 | Cardiometabolic_II | ADAMTSL4 | Q6UY14 | 1 | 150549369 | 150560937 | 0.94 | 0.81 | 1.10 | 4.77E-01 | 0.747 |
| OID31048 | HLA-A | Neurology_II | HLA-A | P04439 | 6 | 29941260 | 29945884 | 0.94 | 0.80 | 1.11 | 4.77E-01 | 0.747 |
| OID21296 | KRT18 | Oncology | KRT18 | P05783 | 12 | 52948871 | 52952906 | 0.93 | 0.78 | 1.13 | 4.77E-01 | 0.747 |
| OID30375 | PDIA4 | Cardiometabolic_II | PDIA4 | P13667 | 7 | 149003062 | 149028662 | 0.94 | 0.79 | 1.11 | 4.76E-01 | 0.747 |
| OID20094 | SLITRK6 | Cardiometabolic | SLITRK6 | Q9H5Y7 | 13 | 85792790 | 85806683 | 0.94 | 0.80 | 1.11 | 4.77E-01 | 0.747 |
| OID21204 | MAVS | Oncology | MAVS | Q7Z434 | 20 | 3846799 | 3876123 | 0.94 | 0.79 | 1.11 | 4.77E-01 | 0.747 |
| OID30138 | RIPK4 | Cardiometabolic_II | RIPK4 | P57078 | 21 | 41739369 | 41767089 | 0.94 | 0.79 | 1.11 | 4.78E-01 | 0.748 |
| OID31274 | TPPP2 | Oncology_II | TPPP2 | P59282 | 14 | 21024109 | 21036276 | 0.94 | 0.81 | 1.11 | 4.78E-01 | 0.748 |
| OID20472 | IL5 | Inflammation | IL5 | P05113 | 5 | 132541445 | 132556838 | 0.94 | 0.79 | 1.12 | 4.79E-01 | 0.749 |
| OID20658 | AGRP | Inflammation | AGRP | O00253 | 16 | 67482571 | 67483547 | 1.06 | 0.90 | 1.25 | 4.80E-01 | 0.749 |
| OID30880 | ERC2 | Neurology_II | ERC2 | O15083 | 3 | 55508311 | 56469095 | 0.94 | 0.78 | 1.12 | 4.80E-01 | 0.749 |
| OID30424 | TPSG1 | Inflammation_II | TPSG1 | Q9NRR2 | 16 | 1221651 | 1225268 | 0.94 | 0.79 | 1.12 | 4.80E-01 | 0.749 |
| OID20852 | SLC39A14 | Neurology | SLC39A14 | Q15043 | 8 | 22367278 | 22434129 | 1.06 | 0.90 | 1.25 | 4.81E-01 | 0.751 |
| OID21440 | F3 | Oncology | F3 | P13726 | 1 | 94529173 | 94541759 | 1.06 | 0.90 | 1.26 | 4.82E-01 | 0.751 |
| OID21138 | ARSA | Neurology | ARSA | P15289 | 22 | 50622754 | 50628173 | 0.94 | 0.79 | 1.12 | 4.84E-01 | 0.752 |
| OID20599 | CD70 | Inflammation | CD70 | P32970 | 19 | 6583183 | 6604103 | 0.94 | 0.80 | 1.11 | 4.84E-01 | 0.752 |
| OID31395 | DTNB | Oncology_II | DTNB | O60941 | 2 | 25377198 | 25673647 | 0.94 | 0.78 | 1.13 | 4.83E-01 | 0.752 |
| OID20364 | PRSS2 | Cardiometabolic | PRSS2 | P07478 | 7 | 142760398 | 142774564 | 0.94 | 0.81 | 1.11 | 4.84E-01 | 0.752 |
| OID20676 | SMPDL3A | Inflammation | SMPDL3A | Q92484 | 6 | 122789049 | 122809720 | 1.06 | 0.90 | 1.25 | 4.83E-01 | 0.752 |
| OID20259 | XG | Cardiometabolic | XG | P55808 | X | 2752040 | 2816500 | 1.07 | 0.89 | 1.29 | 4.83E-01 | 0.752 |
| OID21522 | ITGB5 | Oncology | ITGB5 | P18084 | 3 | 124761948 | 124901418 | 0.94 | 0.79 | 1.12 | 4.84E-01 | 0.752 |
| OID20812 | SIGLEC15 | Neurology | SIGLEC15 | Q6ZMC9 | 18 | 45825675 | 45844094 | 0.94 | 0.80 | 1.11 | 4.85E-01 | 0.752 |
| OID20333 | AMY2A | Cardiometabolic | AMY2A | P04746 | 1 | 103617427 | 103625780 | 0.94 | 0.80 | 1.11 | 4.86E-01 | 0.753 |
| OID30381 | PRG2 | Cardiometabolic_II | PRG2 | P13727 | 11 | 57386780 | 57390650 | 1.06 | 0.90 | 1.25 | 4.86E-01 | 0.753 |
| OID20891 | ECE1 | Neurology | ECE1 | P42892 | 1 | 21217247 | 21345572 | 0.95 | 0.81 | 1.10 | 4.87E-01 | 0.755 |
| OID21289 | SEZ6L2 | Oncology | SEZ6L2 | Q6UXD5 | 16 | 29871159 | 29899550 | 0.94 | 0.79 | 1.12 | 4.87E-01 | 0.755 |
| OID21373 | DEFB4A_DEFB4B | Oncology | DEFB4A;DEFB4B | O15263;O15263 | 8;8 | 7894677;7414855 | 7896716;7416863 | 1.06 | 0.90 | 1.25 | 4.89E-01 | 0.756 |
| OID30489 | MRPS16 | Inflammation_II | MRPS16 | Q9Y3D3 | 10 | 73248843 | 73252693 | 0.93 | 0.77 | 1.13 | 4.88E-01 | 0.756 |
| OID21084 | WARS | Neurology | WARS1 | P23381 | 14 | 100333790 | 100376805 | 0.94 | 0.80 | 1.11 | 4.88E-01 | 0.756 |
| OID31285 | GAS2 | Oncology_II | GAS2 | O43903 | 11 | 22625509 | 22813001 | 0.93 | 0.77 | 1.13 | 4.90E-01 | 0.757 |
| OID20299 | CCN3 | Cardiometabolic | CCN3 | P48745 | 8 | 119416446 | 119424434 | 0.94 | 0.79 | 1.12 | 4.91E-01 | 0.757 |
| OID30790 | CFH | Inflammation_II | CFH | P08603 | 1 | 196651754 | 196752476 | 0.93 | 0.76 | 1.14 | 4.91E-01 | 0.757 |
| OID30404 | SLURP1 | Cardiometabolic_II | SLURP1 | P55000 | 8 | 142740949 | 142742406 | 1.05 | 0.92 | 1.20 | 4.91E-01 | 0.757 |
| OID30374 | CYTL1 | Cardiometabolic_II | CYTL1 | Q9NRR1 | 4 | 5014586 | 5019458 | 1.06 | 0.90 | 1.25 | 4.91E-01 | 0.758 |
| OID20875 | TDRKH | Neurology | TDRKH | Q9Y2W6 | 1 | 151770107 | 151791416 | 0.94 | 0.80 | 1.12 | 4.92E-01 | 0.759 |
| OID20947 | TMPRSS5 | Neurology | TMPRSS5 | Q9H3S3 | 11 | 113687547 | 113706373 | 0.94 | 0.79 | 1.12 | 4.92E-01 | 0.759 |
| OID21056 | CTSS | Neurology | CTSS | P25774 | 1 | 150730079 | 150765957 | 0.94 | 0.80 | 1.11 | 4.93E-01 | 0.759 |
| OID20118 | CLTA | Cardiometabolic | CLTA | P09496 | 9 | 36190856 | 36304781 | 0.94 | 0.79 | 1.12 | 4.93E-01 | 0.760 |
| OID21094 | PIK3IP1 | Neurology | PIK3IP1 | Q96FE7 | 22 | 31281594 | 31292534 | 1.06 | 0.90 | 1.24 | 4.94E-01 | 0.761 |
| OID20439 | PREB | Inflammation | PREB | Q9HCU5 | 2 | 27130756 | 27134666 | 0.94 | 0.80 | 1.12 | 4.95E-01 | 0.761 |
| OID30587 | COL5A1 | Inflammation_II | COL5A1 | P20908 | 9 | 134641803 | 134844843 | 0.94 | 0.80 | 1.11 | 4.95E-01 | 0.761 |
| OID21123 | OGN | Neurology | OGN | P20774 | 9 | 92383268 | 92404696 | 1.06 | 0.90 | 1.25 | 4.96E-01 | 0.761 |
| OID31213 | RASGRF1 | Oncology_II | RASGRF1 | Q13972 | 15 | 78959906 | 79090780 | 0.94 | 0.80 | 1.11 | 4.95E-01 | 0.761 |
| OID20186 | SEMA3F | Cardiometabolic | SEMA3F | Q13275 | 3 | 50155045 | 50189075 | 1.06 | 0.89 | 1.26 | 4.96E-01 | 0.761 |
| OID30203 | ANP32C | Cardiometabolic_II | ANP32C | O43423 | 4 | 164197007 | 16419771 | 0.94 | 0.77 | 1.13 | 4.98E-01 | 0.764 |
| OID21155 | TNFRSF1A | Neurology | TNFRSF1A | P19438 | 12 | 6328757 | 6342114 | 1.06 | 0.90 | 1.25 | 5.01E-01 | 0.768 |
| OID20767 | MATN2 | Inflammation | MATN2 | O00339 | 8 | 97868840 | 98036724 | 0.95 | 0.80 | 1.11 | 5.02E-01 | 0.768 |
| OID30205 | SNED1 | Cardiometabolic_II | SNED1 | Q8TER0 | 2 | 240998618 | 241095568 | 1.05 | 0.90 | 1.23 | 5.02E-01 | 0.768 |
| OID21051 | BST1 | Neurology | BST1 | Q10588 | 4 | 15703065 | 15738313 | 0.95 | 0.81 | 1.11 | 5.04E-01 | 0.769 |
| OID20279 | CRTAC1 | Cardiometabolic | CRTAC1 | Q9NQ79 | 10 | 97865000 | 98030828 | 1.06 | 0.89 | 1.26 | 5.04E-01 | 0.769 |
| OID30494 | DDX39A | Inflammation_II | DDX39A | O00148 | 19 | 14408798 | 14419383 | 1.06 | 0.89 | 1.26 | 5.03E-01 | 0.769 |
| OID30858 | DEFB118 | Neurology_II | DEFB118 | Q96PH6 | 20 | 31368601 | 31373923 | 1.05 | 0.91 | 1.22 | 5.03E-01 | 0.769 |
| OID20736 | ENPP5 | Inflammation | ENPP5 | Q9UJA9 | 6 | 46159185 | 46170980 | 1.07 | 0.89 | 1.28 | 5.03E-01 | 0.769 |
| OID31252 | EPN1 | Oncology_II | EPN1 | Q9Y6I3 | 19 | 55675226 | 55709858 | 0.94 | 0.78 | 1.13 | 5.03E-01 | 0.769 |
| OID31493 | PSCA | Oncology_II | PSCA | O43653 | 8 | 142670308 | 142682725 | 1.06 | 0.90 | 1.24 | 5.04E-01 | 0.769 |
| OID20978 | AFP | Neurology | AFP | P02771 | 4 | 73431138 | 73456174 | 0.94 | 0.79 | 1.12 | 5.05E-01 | 0.770 |
| OID30703 | DBH | Inflammation_II | DBH | P09172 | 9 | 133636363 | 133659329 | 0.95 | 0.81 | 1.11 | 5.06E-01 | 0.770 |
| OID30868 | MAP1LC3A | Neurology_II | MAP1LC3A | Q9H492 | 20 | 34546854 | 34560345 | 0.95 | 0.80 | 1.11 | 5.05E-01 | 0.770 |
| OID31079 | SEPTIN8 | Neurology_II | SEPTIN8 | Q92599 | 5 | 132750819 | 132807241 | 0.94 | 0.79 | 1.13 | 5.06E-01 | 0.770 |
| OID21449 | CD5 | Oncology | CD5 | P06127 | 11 | 61102489 | 61127852 | 0.95 | 0.80 | 1.11 | 5.07E-01 | 0.770 |
| OID31295 | FUT1 | Oncology_II | FUT1 | P19526 | 19 | 48748011 | 48755390 | 1.05 | 0.91 | 1.22 | 5.06E-01 | 0.770 |
| OID21446 | TNFRSF19 | Oncology | TNFRSF19 | Q9NS68 | 13 | 23570370 | 23676104 | 1.05 | 0.90 | 1.23 | 5.07E-01 | 0.770 |
| OID20772 | CCL24 | Inflammation | CCL24 | O00175 | 7 | 75810825 | 75823356 | 1.06 | 0.90 | 1.24 | 5.07E-01 | 0.770 |
| OID31230 | ATP1B4 | Oncology_II | ATP1B4 | Q9UN42 | X | 120362085 | 120383249 | 1.05 | 0.90 | 1.23 | 5.08E-01 | 0.771 |
| OID21124 | PLAU | Neurology | PLAU | P00749 | 10 | 73909177 | 73917496 | 0.94 | 0.80 | 1.12 | 5.08E-01 | 0.771 |

| | | | | | | | | | | | | |
|---|---|---|---|---|---|---|---|---|---|---|---|---|
| OID21058 | SCARA5 | Neurology | SCARA5 | Q6ZMJ2 | 8 | 27869883 | 27992673 | 0.95 | 0.80 | 1.12 | 5.08E-01 | 0.771 |
| OID31397 | IMMT | Oncology_II | IMMT | Q16891 | 2 | 86143932 | 86195472 | 0.94 | 0.79 | 1.12 | 5.09E-01 | 0.772 |
| OID30715 | CRISP3 | Inflammation_II | CRISP3 | P54108 | 6 | 49727376 | 49744437 | 0.94 | 0.80 | 1.12 | 5.10E-01 | 0.772 |
| OID20862 | SLIT2 | Neurology | SLIT2 | O94813 | 4 | 20251905 | 20620561 | 1.06 | 0.90 | 1.24 | 5.10E-01 | 0.772 |
| OID20079 | ACTA2 | Cardiometabolic | ACTA2 | P62736 | 10 | 88935074 | 88991339 | 1.06 | 0.89 | 1.26 | 5.10E-01 | 0.772 |
| OID30063 | CASQ2 | Cardiometabolic_II | CASQ2 | O14958 | 1 | 115700021 | 115768714 | 1.05 | 0.91 | 1.21 | 5.11E-01 | 0.772 |
| OID20236 | IGFBP7 | Cardiometabolic | IGFBP7 | Q16270 | 4 | 57030773 | 57110385 | 1.06 | 0.90 | 1.24 | 5.11E-01 | 0.772 |
| OID30062 | ATP2B4 | Cardiometabolic_II | ATP2B4 | P23634 | 1 | 203626832 | 203744081 | 0.94 | 0.79 | 1.12 | 5.11E-01 | 0.772 |
| OID30449 | GAD2 | Inflammation_II | GAD2 | Q05329 | 10 | 26216665 | 26304558 | 0.94 | 0.79 | 1.12 | 5.12E-01 | 0.772 |
| OID20567 | NCF2 | Inflammation | NCF2 | P19878 | 1 | 183555562 | 183590876 | 0.94 | 0.79 | 1.12 | 5.12E-01 | 0.772 |
| OID30820 | CACNB1 | Neurology_II | CACNB1 | Q02641 | 17 | 39173453 | 39197702 | 1.05 | 0.91 | 1.21 | 5.13E-01 | 0.773 |
| OID20699 | CRLF1 | Inflammation | CRLF1 | O75462 | 19 | 18572220 | 18607741 | 1.06 | 0.89 | 1.25 | 5.13E-01 | 0.773 |
| OID20376 | ITGB1 | Cardiometabolic | ITGB1 | P05556 | 10 | 32887273 | 33005792 | 0.95 | 0.80 | 1.12 | 5.12E-01 | 0.773 |
| OID31328 | CINP | Oncology_II | CINP | Q9BW66 | 14 | 102341102 | 102362916 | 0.94 | 0.80 | 1.12 | 5.14E-01 | 0.775 |
| OID21055 | NCAN | Neurology | NCAN | O14594 | 19 | 19211958 | 19252233 | 1.06 | 0.89 | 1.26 | 5.15E-01 | 0.776 |
| OID30588 | ADGRD1 | Inflammation_II | ADGRD1 | Q6QNK2 | 12 | 130953907 | 131141469 | 0.94 | 0.79 | 1.13 | 5.15E-01 | 0.776 |
| OID31161 | PCNA | Oncology_II | PCNA | P12004 | 20 | 5114953 | 5126626 | 0.94 | 0.79 | 1.13 | 5.16E-01 | 0.776 |
| OID21131 | PECAM1 | Neurology | PECAM1 | P16284 | 17 | 64319415 | 64413776 | 0.95 | 0.81 | 1.11 | 5.16E-01 | 0.776 |
| OID30467 | WASL | Inflammation_II | WASL | O00401 | 7 | 123681943 | 123749003 | 0.94 | 0.80 | 1.12 | 5.17E-01 | 0.776 |
| OID30699 | PGA4 | Inflammation_II | PGA4 | P0DJD7 | 11 | 61222347 | 61231694 | 0.95 | 0.83 | 1.10 | 5.17E-01 | 0.777 |
| OID30914 | CUZD1 | Neurology_II | CUZD1 | Q86UP6 | 10 | 122832158 | 122846175 | 0.94 | 0.77 | 1.14 | 5.18E-01 | 0.777 |
| OID31326 | PTTG1 | Oncology_II | PTTG1 | O95997 | 5 | 160421855 | 160428739 | 0.95 | 0.80 | 1.12 | 5.19E-01 | 0.778 |
| OID21267 | BAMBI | Oncology | BAMBI | Q13145 | 10 | 28677510 | 28682932 | 0.95 | 0.80 | 1.12 | 5.19E-01 | 0.778 |
| OID20405 | COMP | Cardiometabolic | COMP | P49747 | 19 | 18782773 | 18791305 | 0.95 | 0.80 | 1.12 | 5.19E-01 | 0.778 |
| OID31006 | HJV | Neurology_II | HJV | Q6ZVN8 | 1 | 146017468 | 146036746 | 1.05 | 0.90 | 1.23 | 5.19E-01 | 0.778 |
| OID30917 | MYLPF | Neurology_II | MYL11 | Q96A32 | 16 | 30370934 | 30377991 | 1.06 | 0.89 | 1.25 | 5.20E-01 | 0.778 |
| OID21326 | STX4 | Oncology | STX4 | Q12846 | 16 | 31032889 | 31042975 | 0.94 | 0.78 | 1.13 | 5.20E-01 | 0.778 |
| OID30826 | BRD2 | Neurology_II | BRD2 | P25440 | 6 | 32968594 | 32981505 | 0.94 | 0.79 | 1.13 | 5.21E-01 | 0.779 |
| OID31400 | VSIG10 | Oncology_II | VSIG10 | Q8N0Z9 | 12 | 118063593 | 118136026 | 0.94 | 0.79 | 1.13 | 5.21E-01 | 0.779 |
| OID20308 | BPIFB1 | Cardiometabolic | BPIFB1 | Q8TDL5 | 20 | 33273480 | 33309871 | 1.06 | 0.89 | 1.25 | 5.21E-01 | 0.779 |
| OID20854 | CASP1 | Neurology | CASP1 | P29466 | 11 | 105025443 | 105035250 | 1.05 | 0.90 | 1.23 | 5.23E-01 | 0.782 |
| OID30564 | PPL | Inflammation_II | PPL | O60437 | 16 | 4882507 | 4960741 | 1.06 | 0.89 | 1.25 | 5.24E-01 | 0.782 |
| OID30422 | IL12RB2 | Inflammation_II | IL12RB2 | Q99665 | 1 | 67307364 | 67398724 | 0.94 | 0.79 | 1.13 | 5.24E-01 | 0.782 |
| OID30758 | CFHR4 | Inflammation_II | CFHR4 | Q92496 | 1 | 196888014 | 196918795 | 0.95 | 0.81 | 1.11 | 5.25E-01 | 0.782 |
| OID31096 | PALM2 | Neurology_II | PALM2AKAP2 | Q8IXS6 | 9 | 109498325 | 110172512 | 0.94 | 0.79 | 1.13 | 5.25E-01 | 0.782 |
| OID20653 | TNFRSF4 | Inflammation | TNFRSF4 | P43489 | 1 | 1211340 | 1214153 | 1.05 | 0.90 | 1.22 | 5.25E-01 | 0.782 |
| OID20269 | MET | Cardiometabolic | MET | P08581 | 7 | 116672196 | 116798377 | 1.06 | 0.89 | 1.25 | 5.25E-01 | 0.782 |
| OID30515 | C1QL2 | Inflammation_II | C1QL2 | Q7Z5L3 | 2 | 119156243 | 119158751 | 0.95 | 0.80 | 1.12 | 5.27E-01 | 0.783 |
| OID30993 | DSCAM | Neurology_II | DSCAM | O60469 | 21 | 40010999 | 40847158 | 1.05 | 0.90 | 1.23 | 5.28E-01 | 0.783 |
| OID30886 | EPB41L5 | Neurology_II | EPB41L5 | Q9HCM4 | 2 | 120013077 | 120179119 | 1.05 | 0.90 | 1.24 | 5.27E-01 | 0.783 |
| OID21193 | FCRLB | Oncology | FCRLB | Q6BAA4 | 1 | 161721563 | 161728143 | 0.95 | 0.80 | 1.12 | 5.28E-01 | 0.783 |
| OID20727 | GAL | Inflammation | GAL | P22466 | 11 | 68683779 | 68691175 | 1.05 | 0.89 | 1.24 | 5.28E-01 | 0.783 |
| OID21105 | PLA2G7 | Neurology | PLA2G7 | Q13093 | 6 | 46704201 | 46735693 | 1.05 | 0.90 | 1.24 | 5.26E-01 | 0.783 |
| OID20986 | RBKS | Neurology | RBKS | Q9H477 | 2 | 27781379 | 27890681 | 0.94 | 0.79 | 1.13 | 5.28E-01 | 0.783 |
| OID30500 | IGFL4 | Inflammation_II | IGFL4 | Q6B9Z1 | 19 | 46039182 | 46077118 | 0.94 | 0.79 | 1.13 | 5.29E-01 | 0.784 |
| OID30098 | SHD | Cardiometabolic_II | SHD | Q96IW2 | 19 | 4279266 | 4290722 | 0.95 | 0.80 | 1.12 | 5.29E-01 | 0.784 |
| OID20550 | COL9A1 | Inflammation | COL9A1 | P20849 | 6 | 70216040 | 70303084 | 0.95 | 0.81 | 1.11 | 5.30E-01 | 0.784 |
| OID30671 | CGB3_CGB5_CGB8 | Inflammation_II | GB5;CGB8;CGB3 | 86;P0DN86;P0DN86 | 19;19;19 | 49047638;49022869 | 49049111;49024333 | 0.94 | 0.77 | 1.14 | 5.30E-01 | 0.785 |
| OID31131 | SPRR3 | Neurology_II | SPRR3 | Q9UBC9 | 1 | 153001747 | 153003856 | 1.05 | 0.90 | 1.23 | 5.30E-01 | 0.785 |
| OID30960 | LRP2 | Neurology_II | LRP2 | P98164 | 2 | 169127109 | 169362534 | 1.05 | 0.91 | 1.21 | 5.32E-01 | 0.787 |
| OID30401 | STAB2 | Cardiometabolic_II | STAB2 | Q8WWQ8 | 12 | 103587273 | 103766719 | 0.95 | 0.80 | 1.12 | 5.32E-01 | 0.787 |
| OID30163 | KIF1C | Cardiometabolic_II | KIF1C | O43896 | 17 | 4997950 | 5028401 | 0.94 | 0.78 | 1.14 | 5.33E-01 | 0.787 |
| OID21516 | PPY | Oncology | PPY | P01298 | 17 | 43940804 | 43942476 | 1.05 | 0.90 | 1.23 | 5.35E-01 | 0.790 |
| OID31332 | CDH22 | Oncology_II | CDH22 | Q9UJ99 | 20 | 46173739 | 46308498 | 0.95 | 0.80 | 1.12 | 5.37E-01 | 0.790 |
| OID21297 | COX5B | Oncology | COX5B | P10606 | 2 | 97646062 | 97648383 | 0.95 | 0.80 | 1.12 | 5.38E-01 | 0.790 |
| OID31231 | DCUN1D2 | Oncology_II | DCUN1D2 | Q6PH85 | 13 | 113455819 | 113490951 | 1.05 | 0.90 | 1.22 | 5.37E-01 | 0.790 |
| OID31176 | ESPL1 | Oncology_II | ESPL1 | Q14674 | 12 | 53268299 | 53293638 | 0.95 | 0.80 | 1.12 | 5.38E-01 | 0.790 |
| OID21301 | FLT1 | Oncology | FLT1 | P17948 | 13 | 28300346 | 28495145 | 1.05 | 0.89 | 1.24 | 5.37E-01 | 0.790 |
| OID30694 | MRC1 | Inflammation_II | MRC1 | P22897 | 10 | 17809348 | 17911164 | 1.05 | 0.89 | 1.24 | 5.36E-01 | 0.790 |
| OID20913 | NMNAT1 | Neurology | NMNAT1 | Q9HAN9 | 1 | 9943428 | 9985501 | 0.94 | 0.79 | 1.13 | 5.35E-01 | 0.790 |
| OID21362 | OPTC | Oncology | OPTC | Q9UBM4 | 1 | 203494153 | 203508949 | 1.05 | 0.90 | 1.22 | 5.37E-01 | 0.790 |
| OID20902 | PPP3R1 | Neurology | PPP3R1 | P63098 | 2 | 68178857 | 68256237 | 0.95 | 0.79 | 1.13 | 5.38E-01 | 0.790 |
| OID21353 | SMAD5 | Oncology | SMAD5 | Q99717 | 5 | 136132845 | 136188747 | 0.95 | 0.80 | 1.12 | 5.36E-01 | 0.790 |
| OID21361 | IQGAP2 | Oncology | IQGAP2 | Q13576 | 5 | 76403285 | 76708132 | 0.95 | 0.80 | 1.12 | 5.38E-01 | 0.790 |
| OID20397 | CD93 | Cardiometabolic | CD93 | Q9NPY3 | 20 | 23079360 | 23086324 | 0.95 | 0.82 | 1.11 | 5.39E-01 | 0.790 |
| OID21454 | LEFTY2 | Oncology | LEFTY2 | O00292 | 1 | 225936598 | 225941383 | 0.95 | 0.79 | 1.13 | 5.39E-01 | 0.790 |
| OID31018 | LETM1 | Neurology_II | LETM1 | O95202 | 4 | 1811479 | 1856156 | 0.95 | 0.79 | 1.13 | 5.38E-01 | 0.790 |
| OID30317 | COX6B1 | Cardiometabolic_II | COX6B1 | P14854 | 19 | 35648323 | 35658782 | 0.95 | 0.81 | 1.12 | 5.39E-01 | 0.790 |
| OID30827 | CD3D | Neurology_II | CD3D | P04234 | 11 | 118339075 | 118342705 | 1.05 | 0.90 | 1.23 | 5.40E-01 | 0.790 |
| OID20508 | SULT2A1 | Inflammation | SULT2A1 | Q06520 | 19 | 47870467 | 47886315 | 1.05 | 0.90 | 1.23 | 5.41E-01 | 0.792 |
| OID20483 | PRDX3 | Inflammation | PRDX3 | P30048 | 10 | 119167720 | 119178812 | 0.95 | 0.81 | 1.12 | 5.42E-01 | 0.793 |
| OID31023 | MRPL28 | Neurology_II | MRPL28 | Q13084 | 16 | 366969 | 371289 | 1.05 | 0.90 | 1.23 | 5.43E-01 | 0.794 |
| OID21442 | FXN | Oncology | FXN | Q16595 | 9 | 69035751 | 69079076 | 0.95 | 0.81 | 1.12 | 5.44E-01 | 0.795 |
| OID31473 | PSAPL1 | Oncology_II | PSAPL1 | Q6NUJ1 | 4 | 7430285 | 7434930 | 1.05 | 0.90 | 1.22 | 5.44E-01 | 0.795 |
| OID30472 | IL36A | Inflammation_II | IL36A | Q9UHA7 | 2 | 113005459 | 113008044 | 0.95 | 0.80 | 1.13 | 5.45E-01 | 0.795 |
| OID30483 | PRSS22 | Inflammation_II | PRSS22 | Q9GZN4 | 16 | 2852730 | 2858170 | 1.05 | 0.90 | 1.22 | 5.45E-01 | 0.795 |
| OID30514 | PSMG4 | Inflammation_II | PSMG4 | Q5JS54 | 6 | 3231403 | 3303373 | 0.95 | 0.79 | 1.13 | 5.45E-01 | 0.795 |
| OID31449 | TOP1 | Oncology_II | TOP1 | P11387 | 20 | 41028822 | 41124487 | 0.95 | 0.81 | 1.12 | 5.46E-01 | 0.796 |
| OID20257 | PEAR1 | Cardiometabolic | PEAR1 | Q5VY43 | 1 | 156893698 | 156916429 | 0.95 | 0.81 | 1.12 | 5.46E-01 | 0.796 |
| OID31101 | PLXDC2 | Neurology_II | PLXDC2 | Q6UX71 | 10 | 19816239 | 20289856 | 1.04 | 0.91 | 1.19 | 5.46E-01 | 0.796 |
| OID30421 | EDNRB | Inflammation_II | EDNRB | P24530 | 13 | 77895481 | 77975529 | 0.94 | 0.77 | 1.15 | 5.47E-01 | 0.796 |
| OID20488 | IL1A | Inflammation | IL1A | P01583 | 2 | 112773925 | 112784493 | 0.95 | 0.81 | 1.12 | 5.47E-01 | 0.796 |
| OID20411 | ICAM1 | Cardiometabolic | ICAM1 | P05362 | 19 | 10271093 | 10286615 | 1.05 | 0.90 | 1.23 | 5.49E-01 | 0.799 |
| OID20696 | IL10RB | Inflammation | IL10RB | Q08334 | 21 | 33266367 | 33310187 | 1.05 | 0.89 | 1.24 | 5.49E-01 | 0.799 |
| OID21257 | CRH | Oncology | CRH | P06850 | 8 | 66176376 | 66178464 | 0.95 | 0.80 | 1.12 | 5.50E-01 | 0.799 |
| OID30049 | RTKN2 | Cardiometabolic_II | RTKN2 | Q8IZC4 | 10 | 62183035 | 62268844 | 1.05 | 0.90 | 1.22 | 5.51E-01 | 0.800 |
| OID21095 | NCAM2 | Neurology | NCAM2 | O15394 | 21 | 20998409 | 21543329 | 1.05 | 0.89 | 1.24 | 5.52E-01 | 0.802 |
| OID31157 | CTRL | Neurology_II | CTRL | P40313 | 16 | 67927640 | 67932365 | 1.05 | 0.90 | 1.22 | 5.52E-01 | 0.802 |
| OID31489 | C9orf40 | Oncology_II | C9orf40 | Q8IXQ3 | 9 | 74946583 | 74952912 | 0.95 | 0.80 | 1.12 | 5.53E-01 | 0.802 |
| OID21306 | CALB1 | Oncology | CALB1 | P05937 | 8 | 90058608 | 90095475 | 1.05 | 0.89 | 1.24 | 5.53E-01 | 0.802 |
| OID30712 | CAT | Inflammation_II | CAT | P04040 | 11 | 34438934 | 34472060 | 0.95 | 0.79 | 1.13 | 5.54E-01 | 0.802 |
| OID20230 | HSPB1 | Cardiometabolic | HSPB1 | P04792 | 7 | 76302673 | 76304295 | 0.95 | 0.81 | 1.12 | 5.54E-01 | 0.802 |
| OID21013 | FUT3_FUT5 | Neurology | FUT3;FUT5 | P21217;Q11128 | 19;19 | 5842888;5865826 | 5851471;5870540 | 1.05 | 0.89 | 1.24 | 5.54E-01 | 0.802 |
| OID20946 | FLRT2 | Neurology | FLRT2 | O43155 | 14 | 85530144 | 85654428 | 1.05 | 0.89 | 1.23 | 5.55E-01 | 0.803 |
| OID20561 | CXADR | Inflammation | CXADR | P78310 | 21 | 17513043 | 17593579 | 1.05 | 0.90 | 1.21 | 5.56E-01 | 0.804 |
| OID20350 | ANPEP | Cardiometabolic | ANPEP | P15144 | 15 | 89784895 | 89815401 | 1.05 | 0.90 | 1.22 | 5.59E-01 | 0.805 |
| OID30744 | C5 | Inflammation_II | C5 | P01031 | 9 | 120932987 | 121075195 | 1.06 | 0.88 | 1.27 | 5.58E-01 | 0.805 |
| OID21040 | CD74 | Neurology | CD74 | P04233 | 5 | 150401637 | 150412969 | 1.05 | 0.89 | 1.24 | 5.59E-01 | 0.805 |
| OID20751 | CRELD2 | Inflammation | CRELD2 | Q6UXH1 | 22 | 49918167 | 49927540 | 0.95 | 0.81 | 1.12 | 5.60E-01 | 0.805 |
| OID20142 | DPP7 | Cardiometabolic | DPP7 | Q9UHL4 | 9 | 137110546 | 137115177 | 0.95 | 0.81 | 1.12 | 5.60E-01 | 0.805 |
| OID20391 | FCGR2A | Cardiometabolic | FCGR2A | P12318 | 1 | 161505430 | 161524013 | 0.96 | 0.82 | 1.11 | 5.59E-01 | 0.805 |
| OID21491 | FLT4 | Oncology | FLT4 | P35916 | 5 | 180601506 | 180649624 | 0.95 | 0.81 | 1.12 | 5.59E-01 | 0.805 |
| OID20979 | MATN3 | Neurology | MATN3 | O15232 | 2 | 19992052 | 20012668 | 1.05 | 0.89 | 1.23 | 5.60E-01 | 0.805 |
| OID20263 | MB | Cardiometabolic | MB | P02144 | 22 | 35606764 | 35637951 | 0.95 | 0.79 | 1.14 | 5.59E-01 | 0.805 |
| OID31356 | RCC1 | Oncology_II | RCC1 | P18754 | 1 | 28505943 | 28539300 | 1.05 | 0.90 | 1.22 | 5.58E-01 | 0.805 |
| OID21374 | S100A12 | Oncology | S100A12 | P80511 | 1 | 153373711 | 153375621 | 1.05 | 0.89 | 1.24 | 5.58E-01 | 0.805 |
| OID31445 | SLC9A3R2 | Oncology_II | SLC9A3R2 | Q15599 | 16 | 2025356 | 2039026 | 0.95 | 0.78 | 1.14 | 5.60E-01 | 0.805 |

| | | | | | | | | | | | | |
|---|---|---|---|---|---|---|---|---|---|---|---|---|
| OID20686 | CCL21 | Inflammation | CCL21 | O00585 | 9 | 34709005 | 34710136 | 1.05 | 0.90 | 1.23 | 5.61E-01 | 0.806 |
| OID20684 | TIMP3 | Inflammation | TIMP3 | P35625 | 22 | 32801705 | 32863041 | 0.95 | 0.81 | 1.12 | 5.61E-01 | 0.806 |
| OID31384 | CAPN3 | Oncology_II | CAPN3 | P20807 | 15 | 42359498 | 42412949 | 1.04 | 0.91 | 1.19 | 5.62E-01 | 0.807 |
| OID20351 | SSC5D | Cardiometabolic | SSC5D | A1L4H1 | 19 | 55488404 | 55519099 | 0.95 | 0.80 | 1.13 | 5.62E-01 | 0.807 |
| OID30679 | ASGR2 | Inflammation_II | ASGR2 | P07307 | 17 | 7101322 | 7115700 | 0.95 | 0.81 | 1.12 | 5.64E-01 | 0.809 |
| OID21340 | CES3 | Oncology | CES3 | Q6UWW8 | 16 | 66961245 | 66975149 | 0.95 | 0.80 | 1.13 | 5.65E-01 | 0.809 |
| OID30720 | CFHR2 | Inflammation_II | CFHR2 | P36980 | 1 | 196943738 | 196959622 | 0.95 | 0.81 | 1.12 | 5.64E-01 | 0.809 |
| OID30929 | CLNS1A | Neurology_II | CLNS1A | P54105 | 11 | 77514936 | 77637794 | 0.95 | 0.80 | 1.13 | 5.65E-01 | 0.809 |
| OID30077 | SLC12A2 | Cardiometabolic_II | SLC12A2 | P55011 | 5 | 128083766 | 128189677 | 0.95 | 0.79 | 1.14 | 5.65E-01 | 0.809 |
| OID31262 | TXK | Oncology_II | TXK | P42681 | 4 | 48066393 | 48134250 | 0.95 | 0.78 | 1.14 | 5.65E-01 | 0.809 |
| OID20609 | CLEC4D | Inflammation | CLEC4D | Q8WXI8 | 12 | 8509475 | 8522366 | 1.05 | 0.89 | 1.24 | 5.66E-01 | 0.810 |
| OID20248 | ICAM2 | Cardiometabolic | ICAM2 | P13598 | 17 | 64002594 | 64020634 | 1.05 | 0.89 | 1.23 | 5.67E-01 | 0.810 |
| OID20920 | NGF | Neurology | NGF | P01138 | 1 | 115285904 | 115338770 | 1.04 | 0.90 | 1.20 | 5.67E-01 | 0.810 |
| OID30947 | PPT1 | Neurology_II | PPT1 | P50897 | 1 | 40072710 | 40097260 | 0.95 | 0.80 | 1.13 | 5.67E-01 | 0.810 |
| OID21074 | NPTX1 | Neurology | NPTX1 | Q15818 | 17 | 80466834 | 80477843 | 1.05 | 0.89 | 1.24 | 5.68E-01 | 0.811 |
| OID30856 | BLOC1S2 | Neurology_II | BLOC1S2 | Q6QNY1 | 10 | 100273280 | 100286680 | 0.96 | 0.82 | 1.12 | 5.69E-01 | 0.812 |
| OID20387 | FCGR3B | Cardiometabolic | FCGR3B | O75015 | 1 | 161623196 | 161631963 | 1.05 | 0.89 | 1.23 | 5.70E-01 | 0.813 |
| OID31450 | ZNRD2 | Oncology_II | ZNRD2 | O60232 | 11 | 65570460 | 65573942 | 0.95 | 0.80 | 1.13 | 5.70E-01 | 0.813 |
| OID30828 | ART5 | Neurology_II | ART5 | Q96L15 | 11 | 3638512 | 3642316 | 1.04 | 0.90 | 1.21 | 5.71E-01 | 0.813 |
| OID21376 | GPA33 | Oncology | GPA33 | Q99795 | 1 | 167052836 | 167166479 | 0.95 | 0.81 | 1.13 | 5.71E-01 | 0.813 |
| OID30060 | EIF2AK3 | Cardiometabolic_II | EIF2AK3 | Q9NZJ5 | 2 | 88556741 | 88691518 | 0.95 | 0.81 | 1.12 | 5.72E-01 | 0.813 |
| OID20469 | IL17A | Inflammation | IL17A | Q16552 | 6 | 52186375 | 52190638 | 1.04 | 0.90 | 1.20 | 5.73E-01 | 0.813 |
| OID30102 | NUDT15 | Cardiometabolic_II | NUDT15 | Q9NV35 | 13 | 48037726 | 48047221 | 0.95 | 0.81 | 1.13 | 5.72E-01 | 0.813 |
| OID31090 | STX3 | Neurology_II | STX3 | Q13277 | 11 | 59713456 | 59805882 | 0.95 | 0.80 | 1.13 | 5.73E-01 | 0.813 |
| OID30068 | TMED10 | Cardiometabolic_II | TMED10 | P49755 | 14 | 75131469 | 75176612 | 1.04 | 0.90 | 1.20 | 5.72E-01 | 0.813 |
| OID30799 | TUBB3 | Neurology_II | TUBB3 | Q13509 | 16 | 89921392 | 89938761 | 0.95 | 0.81 | 1.13 | 5.72E-01 | 0.813 |
| OID31173 | ANKRA2 | Oncology_II | ANKRA2 | Q9H9E1 | 5 | 73552190 | 73565667 | 0.95 | 0.81 | 1.13 | 5.75E-01 | 0.814 |
| OID20491 | CSF3 | Inflammation | CSF3 | P09919 | 17 | 40015361 | 40017813 | 0.95 | 0.80 | 1.13 | 5.74E-01 | 0.814 |
| OID30393 | CTHRC1 | Cardiometabolic_II | CTHRC1 | Q96CG8 | 8 | 103371538 | 103382989 | 1.05 | 0.88 | 1.27 | 5.75E-01 | 0.814 |
| OID20879 | DCTN6 | Neurology | DCTN6 | O00399 | 8 | 30156319 | 30183639 | 0.95 | 0.81 | 1.12 | 5.74E-01 | 0.814 |
| OID31214 | PDIA2 | Oncology_II | PDIA2 | Q13087 | 16 | 283164 | 287215 | 0.95 | 0.80 | 1.13 | 5.74E-01 | 0.814 |
| OID30383 | PRG3 | Cardiometabolic_II | PRG3 | Q9Y2Y8 | 11 | 57376769 | 57381150 | 0.95 | 0.80 | 1.13 | 5.74E-01 | 0.814 |
| OID31488 | ADAM9 | Oncology_II | ADAM9 | Q13443 | 8 | 38996754 | 39105445 | 1.05 | 0.89 | 1.22 | 5.76E-01 | 0.814 |
| OID30955 | AKR1B10 | Neurology_II | AKR1B10 | O60218 | 7 | 134527567 | 134541412 | 0.96 | 0.81 | 1.12 | 5.76E-01 | 0.814 |
| OID20189 | ADGRG2 | Cardiometabolic | ADGRG2 | Q8IZP9 | X | 18989307 | 19122637 | 0.95 | 0.80 | 1.14 | 5.76E-01 | 0.814 |
| OID30943 | PMS1 | Neurology_II | PMS1 | P54277 | 2 | 189784085 | 189877629 | 0.95 | 0.80 | 1.13 | 5.76E-01 | 0.814 |
| OID31308 | CEACAM20 | Oncology_II | CEACAM20 | Q6UY09 | 19 | 44501677 | 44529788 | 0.96 | 0.81 | 1.12 | 5.78E-01 | 0.815 |
| OID30945 | MDM1 | Neurology_II | MDM1 | Q8TC05 | 12 | 68272443 | 68332381 | 0.95 | 0.81 | 1.12 | 5.78E-01 | 0.815 |
| OID20222 | NECTIN2 | Cardiometabolic | NECTIN2 | Q92692 | 19 | 44846175 | 44889223 | 1.05 | 0.89 | 1.23 | 5.77E-01 | 0.815 |
| OID21304 | PSMD9 | Oncology | PSMD9 | O00233 | 12 | 121888732 | 121918297 | 0.95 | 0.80 | 1.13 | 5.77E-01 | 0.815 |
| OID30075 | REXO2 | Cardiometabolic_II | REXO2 | Q9Y3B8 | 11 | 114439435 | 114450279 | 0.95 | 0.81 | 1.13 | 5.78E-01 | 0.815 |
| OID20390 | NRP1 | Cardiometabolic | NRP1 | O14786 | 10 | 33177492 | 33336262 | 1.04 | 0.90 | 1.22 | 5.79E-01 | 0.816 |
| OID20799 | SLC16A1 | Neurology | SLC16A1 | P53985 | 1 | 112911847 | 112957593 | 0.95 | 0.81 | 1.13 | 5.80E-01 | 0.817 |
| OID20357 | HYAL1 | Cardiometabolic | HYAL1 | Q12794 | 3 | 50299890 | 50312381 | 0.96 | 0.82 | 1.12 | 5.81E-01 | 0.818 |
| OID21485 | FOLR3 | Oncology | FOLR3 | P41439 | 11 | 72114869 | 72139892 | 1.05 | 0.89 | 1.24 | 5.82E-01 | 0.819 |
| OID20492 | AMN | Inflammation | AMN | Q9BXJ7 | 14 | 102922663 | 102933596 | 1.05 | 0.89 | 1.23 | 5.85E-01 | 0.819 |
| OID20340 | AMY2B | Cardiometabolic | AMY2B | P19961 | 1 | 103553815 | 103579534 | 0.96 | 0.81 | 1.12 | 5.84E-01 | 0.819 |
| OID21466 | CCL8 | Oncology | CCL8 | P80075 | 17 | 34319435 | 34321402 | 0.95 | 0.81 | 1.13 | 5.84E-01 | 0.819 |
| OID31189 | CDC25A | Oncology_II | CDC25A | P30304 | 3 | 48157146 | 48188417 | 1.04 | 0.89 | 1.22 | 5.85E-01 | 0.819 |
| OID31349 | ENOPH1 | Oncology_II | ENOPH1 | Q9UHY7 | 4 | 82430590 | 82461177 | 0.95 | 0.81 | 1.13 | 5.86E-01 | 0.819 |
| OID30351 | FABP3 | Cardiometabolic_II | FABP3 | P05413 | 1 | 31365253 | 31376850 | 0.95 | 0.79 | 1.14 | 5.85E-01 | 0.819 |
| OID20227 | FBP1 | Cardiometabolic | FBP1 | P09467 | 9 | 94603133 | 94640249 | 0.95 | 0.80 | 1.13 | 5.85E-01 | 0.819 |
| OID21312 | IGSF3 | Oncology | IGSF3 | O75054 | 1 | 116574399 | 116667755 | 0.95 | 0.80 | 1.14 | 5.85E-01 | 0.819 |
| OID20167 | IGSF8 | Cardiometabolic | IGSF8 | Q969P0 | 1 | 160091340 | 160098943 | 0.96 | 0.81 | 1.12 | 5.85E-01 | 0.819 |
| OID20860 | KRT5 | Neurology | KRT5 | P13647 | 12 | 52514575 | 52520530 | 0.96 | 0.81 | 1.13 | 5.83E-01 | 0.819 |
| OID21307 | MME | Oncology | MME | P08473 | 3 | 155024124 | 155183704 | 0.96 | 0.81 | 1.12 | 5.83E-01 | 0.819 |
| OID20542 | SCRN1 | Inflammation | SCRN1 | Q12765 | 7 | 29920104 | 29990289 | 0.95 | 0.80 | 1.13 | 5.86E-01 | 0.819 |
| OID31289 | TAP1 | Oncology_II | TAP1 | Q03518 | 6 | 32845209 | 32853816 | 0.95 | 0.80 | 1.14 | 5.84E-01 | 0.819 |
| OID30512 | TSPYL1 | Inflammation_II | TSPYL1 | Q9H0U9 | 6 | 116267760 | 116279930 | 0.95 | 0.80 | 1.13 | 5.86E-01 | 0.819 |
| OID31105 | VSNL1 | Neurology_II | VSNL1 | P62760 | 2 | 17539126 | 17657018 | 1.05 | 0.89 | 1.23 | 5.86E-01 | 0.819 |
| OID30312 | BCAT1 | Cardiometabolic_II | BCAT1 | P54687 | 12 | 24810024 | 24949101 | 1.05 | 0.89 | 1.23 | 5.88E-01 | 0.821 |
| OID21325 | DSG4 | Oncology | DSG4 | Q86SJ6 | 18 | 31376777 | 31414912 | 1.05 | 0.89 | 1.24 | 5.89E-01 | 0.821 |
| OID30057 | LAMA1 | Cardiometabolic_II | LAMA1 | P25391 | 18 | 6941742 | 7117797 | 0.95 | 0.80 | 1.14 | 5.89E-01 | 0.821 |
| OID30380 | PTPRC | Cardiometabolic_II | PTPRC | P08575 | 1 | 198638457 | 198757476 | 1.05 | 0.89 | 1.24 | 5.89E-01 | 0.821 |
| OID30154 | TMED4 | Cardiometabolic_II | TMED4 | Q7Z7H5 | 7 | 44577894 | 44582287 | 1.04 | 0.90 | 1.21 | 5.88E-01 | 0.821 |
| OID31113 | YAP1 | Neurology_II | YAP1 | P46937 | 11 | 102110447 | 102233424 | 0.95 | 0.80 | 1.13 | 5.89E-01 | 0.821 |
| OID31247 | CEP350 | Oncology_II | CEP350 | Q5VT06 | 1 | 179954674 | 180114880 | 1.05 | 0.89 | 1.23 | 5.90E-01 | 0.821 |
| OID30513 | ACRV1 | Inflammation_II | ACRV1 | P26436 | 11 | 125671522 | 125680874 | 0.93 | 0.71 | 1.21 | 5.90E-01 | 0.821 |
| OID30429 | GLI2 | Inflammation_II | GLI2 | P10070 | 2 | 120735623 | 120992653 | 0.95 | 0.80 | 1.14 | 5.90E-01 | 0.821 |
| OID20197 | PILRB | Cardiometabolic | PILRB | Q9UKJ0 | 7 | 100352176 | 100367831 | 0.96 | 0.82 | 1.12 | 5.91E-01 | 0.821 |
| OID21455 | HDGF | Oncology | HDGF | P51858 | 1 | 156742109 | 156766925 | 0.96 | 0.81 | 1.13 | 5.91E-01 | 0.821 |
| OID20645 | ADA | Inflammation | ADA | P00813 | 20 | 44619522 | 44652233 | 0.95 | 0.80 | 1.13 | 5.92E-01 | 0.821 |
| OID20075 | CA5A | Cardiometabolic | CA5A | P35218 | 16 | 87881546 | 87936580 | 0.95 | 0.80 | 1.13 | 5.93E-01 | 0.821 |
| OID30606 | CPA4 | Inflammation_II | CPA4 | Q9UI42 | 7 | 130293134 | 130324180 | 0.96 | 0.82 | 1.12 | 5.92E-01 | 0.821 |
| OID20096 | GZMH | Cardiometabolic | GZMH | P20718 | 14 | 24606480 | 24609699 | 0.95 | 0.81 | 1.13 | 5.92E-01 | 0.821 |
| OID20417 | TGFBI | Cardiometabolic | TGFBI | Q15582 | 5 | 136028988 | 136063818 | 0.96 | 0.81 | 1.13 | 5.93E-01 | 0.821 |
| OID20111 | BMP6 | Cardiometabolic | BMP6 | P22004 | 6 | 7726099 | 7881728 | 0.96 | 0.81 | 1.13 | 5.94E-01 | 0.822 |
| OID30776 | C3 | Inflammation_II | C3 | P01024 | 19 | 6677704 | 6730562 | 0.95 | 0.78 | 1.15 | 5.94E-01 | 0.822 |
| OID20538 | ISM1 | Inflammation | ISM1 | B1AKI9 | 20 | 13221274 | 13300651 | 0.95 | 0.80 | 1.13 | 5.95E-01 | 0.823 |
| OID21221 | PSMA1 | Oncology | PSMA1 | P25786 | 11 | 14504874 | 14643635 | 0.96 | 0.81 | 1.13 | 5.95E-01 | 0.823 |
| OID21452 | LTA4H | Oncology | LTA4H | P09960 | 12 | 96000753 | 96043520 | 1.05 | 0.89 | 1.23 | 5.95E-01 | 0.823 |
| OID30473 | RNF31 | Inflammation_II | RNF31 | Q96EP0 | 14 | 24146283 | 24160660 | 0.95 | 0.79 | 1.14 | 5.96E-01 | 0.823 |
| OID30291 | CD80 | Cardiometabolic_II | CD80 | P33681 | 3 | 119524293 | 119559614 | 1.05 | 0.89 | 1.23 | 5.96E-01 | 0.823 |
| OID31509 | SPINT3 | Oncology_II | SPINT3 | P49223 | 20 | 45512461 | 45515622 | 1.09 | 0.80 | 1.47 | 5.96E-01 | 0.823 |
| OID20298 | CELA3A | Cardiometabolic | CELA3A | P09093 | 1 | 22001657 | 22012542 | 1.04 | 0.89 | 1.21 | 5.97E-01 | 0.824 |
| OID20464 | CXCL12 | Inflammation | CXCL12 | P48061 | 10 | 44370165 | 44386493 | 0.96 | 0.82 | 1.12 | 5.98E-01 | 0.824 |
| OID30842 | PNMA2 | Neurology_II | PNMA2 | Q9UL42 | 8 | 26504701 | 26514092 | 1.05 | 0.88 | 1.24 | 5.98E-01 | 0.824 |
| OID21403 | TRIAP1 | Oncology | TRIAP1 | O43715 | 12 | 120443964 | 120446384 | 0.95 | 0.80 | 1.13 | 5.98E-01 | 0.824 |
| OID21329 | SEMA4C | Oncology | SEMA4C | Q9C0C4 | 2 | 96859718 | 96870757 | 0.95 | 0.80 | 1.13 | 5.99E-01 | 0.824 |
| OID21030 | CCL19 | Neurology | CCL19 | Q99731 | 9 | 34689570 | 34691276 | 1.04 | 0.90 | 1.19 | 5.99E-01 | 0.825 |
| OID21417 | CA9 | Oncology | CA9 | Q16790 | 9 | 35673928 | 35681159 | 1.04 | 0.89 | 1.23 | 6.01E-01 | 0.825 |
| OID20523 | CCL7 | Inflammation | CCL7 | P80098 | 17 | 34270221 | 34272242 | 0.95 | 0.79 | 1.15 | 6.01E-01 | 0.825 |
| OID20104 | ENTPD5 | Cardiometabolic | ENTPD5 | O75356 | 14 | 73958010 | 74019399 | 1.05 | 0.89 | 1.23 | 6.01E-01 | 0.825 |
| OID31083 | GAST | Neurology_II | GAST | P01350 | 17 | 41712331 | 41715969 | 0.96 | 0.83 | 1.12 | 6.01E-01 | 0.825 |
| OID21327 | IL12A_IL12B | Oncology | IL12A;IL12B | P29459;P29460 | 3;5 | 9988835;159314780 | 9996019;159330487 | 1.04 | 0.89 | 1.22 | 6.01E-01 | 0.825 |
| OID20832 | PADI4 | Neurology | PADI4 | Q9UM07 | 1 | 17308195 | 17364004 | 0.95 | 0.80 | 1.14 | 6.01E-01 | 0.825 |
| OID21170 | STX16 | Oncology | STX16 | O14662 | 20 | 58651253 | 58679526 | 1.05 | 0.88 | 1.24 | 6.01E-01 | 0.825 |
| OID21500 | CTSF | Oncology | CTSF | Q9UBX1 | 11 | 66563464 | 66568879 | 0.96 | 0.81 | 1.13 | 6.03E-01 | 0.826 |
| OID30991 | LEO1 | Neurology_II | LEO1 | Q8WVC0 | 15 | 51938025 | 51971778 | 0.95 | 0.79 | 1.15 | 6.02E-01 | 0.826 |
| OID30410 | POSTN | Cardiometabolic_II | POSTN | Q15063 | 13 | 37562583 | 37598844 | 1.05 | 0.88 | 1.24 | 6.03E-01 | 0.826 |
| OID30364 | KLK3 | Cardiometabolic_II | KLK3 | P07288 | 19 | 50854915 | 50860764 | 0.93 | 0.71 | 1.22 | 6.05E-01 | 0.826 |
| OID30105 | NEB | Cardiometabolic_II | NEB | P20929 | 2 | 151485336 | 151734487 | 0.96 | 0.81 | 1.13 | 6.04E-01 | 0.826 |
| OID20274 | PLA2G2A | Cardiometabolic | PLA2G2A | P14555 | 1 | 19975431 | 19980416 | 1.04 | 0.89 | 1.23 | 6.04E-01 | 0.826 |
| OID30560 | STX7 | Inflammation_II | STX7 | O15400 | 6 | 132445867 | 132513198 | 0.96 | 0.80 | 1.14 | 6.04E-01 | 0.826 |
| OID20369 | TFRC | Cardiometabolic | TFRC | P02786 | 3 | 196027183 | 196082096 | 0.96 | 0.81 | 1.13 | 6.04E-01 | 0.826 |
| OID21145 | TNFRSF1B | Neurology | TNFRSF1B | P20333 | 1 | 12166991 | 12209228 | 1.04 | 0.89 | 1.21 | 6.05E-01 | 0.826 |
| OID30855 | KLRC1 | Neurology_II | KLRC1 | P26715 | 12 | 10442264 | 10454685 | 0.96 | 0.80 | 1.14 | 6.07E-01 | 0.829 |

| | | | | | | | | | | | | |
|---|---|---|---|---|---|---|---|---|---|---|---|---|
| OID20198 | CLUL1 | Cardiometabolic | CLUL1 | Q15846 | 18 | 596988 | 650334 | 1.05 | 0.88 | 1.24 | 6.07E-01 | 0.829 |
| OID21243 | AGR3 | Oncology | AGR3 | Q8TD06 | 7 | 16859412 | 16881987 | 1.04 | 0.89 | 1.21 | 6.13E-01 | 0.830 |
| OID31139 | AMPD3 | Neurology_II | AMPD3 | Q01432 | 11 | 10308313 | 10507579 | 0.96 | 0.81 | 1.13 | 6.09E-01 | 0.830 |
| OID21366 | CES2 | Oncology | CES2 | O00748 | 16 | 66934444 | 66945096 | 1.04 | 0.89 | 1.23 | 6.11E-01 | 0.830 |
| OID30681 | CSF1R | Inflammation_II | CSF1R | P07333 | 5 | 150053291 | 150113372 | 1.05 | 0.88 | 1.24 | 6.09E-01 | 0.830 |
| OID30050 | DENND2B | Cardiometabolic_II | DENND2B | P78524 | 11 | 8693351 | 8910951 | 1.04 | 0.89 | 1.21 | 6.14E-01 | 0.830 |
| OID31130 | DNAJA4 | Neurology_II | DNAJA4 | Q8WW22 | 15 | 78264086 | 78282196 | 0.96 | 0.81 | 1.13 | 6.14E-01 | 0.830 |
| OID31000 | DNM3 | Neurology_II | DNM3 | Q9UQ16 | 1 | 171817887 | 172418466 | 0.96 | 0.81 | 1.13 | 6.12E-01 | 0.830 |
| OID30224 | EDN1 | Cardiometabolic_II | EDN1 | P05305 | 6 | 12290361 | 12297194 | 1.04 | 0.89 | 1.23 | 6.11E-01 | 0.830 |
| OID31514 | FAM3D | Oncology_II | FAM3D | Q96BQ1 | 3 | 58633946 | 58666834 | 1.04 | 0.89 | 1.23 | 6.11E-01 | 0.830 |
| OID21137 | HAVCR2 | Neurology | HAVCR2 | Q8TDQ0 | 5 | 157085832 | 157142869 | 1.04 | 0.88 | 1.23 | 6.12E-01 | 0.830 |
| OID30307 | HBZ | Cardiometabolic_II | HBZ | P02008 | 16 | 142728 | 154503 | 1.04 | 0.89 | 1.22 | 6.10E-01 | 0.830 |
| OID31402 | LUZP2 | Oncology_II | LUZP2 | Q86TE4 | 11 | 24496970 | 25082638 | 1.04 | 0.90 | 1.19 | 6.12E-01 | 0.830 |
| OID30713 | MST1 | Inflammation_II | MST1 | P26927 | 3 | 49683947 | 49689501 | 0.96 | 0.83 | 1.12 | 6.12E-01 | 0.830 |
| OID31490 | NCR3LG1 | Oncology_II | NCR3LG1 | Q68D85 | 11 | 17351800 | 17377341 | 1.04 | 0.89 | 1.22 | 6.12E-01 | 0.830 |
| OID30676 | PI16 | Inflammation_II | PI16 | Q6UXB8 | 6 | 36948263 | 36964837 | 0.96 | 0.81 | 1.13 | 6.11E-01 | 0.830 |
| OID20962 | PRDX1 | Neurology | PRDX1 | Q06830 | 1 | 45510914 | 45542732 | 0.96 | 0.81 | 1.13 | 6.13E-01 | 0.830 |
| OID20984 | SFRP1 | Neurology | SFRP1 | Q8N474 | 8 | 41261962 | 41309473 | 1.04 | 0.88 | 1.24 | 6.14E-01 | 0.830 |
| OID21394 | SIGLEC6 | Oncology | SIGLEC6 | O43699 | 19 | 51517819 | 51531856 | 1.04 | 0.89 | 1.22 | 6.14E-01 | 0.830 |
| OID20971 | SNCG | Neurology | SNCG | O76070 | 10 | 86958599 | 86963258 | 1.05 | 0.88 | 1.25 | 6.11E-01 | 0.830 |
| OID30797 | SYNGAP1 | Neurology_II | SYNGAP1 | Q96PV0 | 6 | 33419661 | 33453689 | 1.04 | 0.89 | 1.23 | 6.12E-01 | 0.830 |
| OID30352 | UPB1 | Cardiometabolic_II | UPB1 | Q9UBR1 | 22 | 24494107 | 24528390 | 0.96 | 0.80 | 1.14 | 6.08E-01 | 0.830 |
| OID21372 | FAM3B | Oncology | FAM3B | P58499 | 21 | 41304212 | 41357727 | 1.05 | 0.88 | 1.24 | 6.15E-01 | 0.831 |
| OID21484 | L1CAM | Oncology | L1CAM | P32004 | X | 153861514 | 153886173 | 0.96 | 0.81 | 1.13 | 6.15E-01 | 0.831 |
| OID20496 | SLAMF1 | Inflammation | SLAMF1 | Q13291 | 1 | 160608106 | 160647044 | 1.04 | 0.88 | 1.23 | 6.15E-01 | 0.831 |
| OID20170 | CDH2 | Cardiometabolic | CDH2 | P19022 | 18 | 27932879 | 28177946 | 1.05 | 0.88 | 1.25 | 6.16E-01 | 0.832 |
| OID30738 | CFD | Inflammation_II | CFD | P00746 | 19 | 859453 | 867884 | 0.96 | 0.81 | 1.14 | 6.17E-01 | 0.832 |
| OID20335 | COL1A1 | Cardiometabolic | COL1A1 | P02452 | 17 | 50184101 | 50201632 | 0.96 | 0.82 | 1.13 | 6.18E-01 | 0.832 |
| OID20282 | CXCL16 | Cardiometabolic | CXCL16 | Q9H2A7 | 17 | 4733533 | 4739928 | 0.96 | 0.81 | 1.13 | 6.17E-01 | 0.832 |
| OID20992 | FUT8 | Neurology | FUT8 | Q9BYC5 | 14 | 65410592 | 65744121 | 1.04 | 0.89 | 1.22 | 6.18E-01 | 0.832 |
| OID31526 | RBP7 | Oncology_II | RBP7 | Q96R05 | 1 | 9997206 | 10016021 | 0.95 | 0.80 | 1.15 | 6.18E-01 | 0.832 |
| OID30867 | RRP15 | Neurology_II | RRP15 | Q9Y3B9 | 1 | 218285293 | 218337983 | 1.04 | 0.89 | 1.22 | 6.18E-01 | 0.832 |
| OID30948 | SCN2B | Neurology_II | SCN2B | O60939 | 11 | 118162806 | 118176639 | 0.96 | 0.82 | 1.13 | 6.18E-01 | 0.832 |
| OID20466 | DGKZ | Inflammation | DGKZ | Q13574 | 11 | 46332905 | 46380554 | 1.04 | 0.88 | 1.23 | 6.19E-01 | 0.832 |
| OID30261 | SIGLEC8 | Cardiometabolic_II | SIGLEC8 | Q9NYZ4 | 19 | 51450847 | 51458456 | 1.04 | 0.88 | 1.23 | 6.19E-01 | 0.832 |
| OID20437 | AMBN | Inflammation | AMBN | Q9NP70 | 4 | 70592256 | 70607288 | 1.04 | 0.89 | 1.21 | 6.21E-01 | 0.833 |
| OID30572 | AMOT | Inflammation_II | AMOT | Q4VCS5 | X | 112774503 | 112840831 | 0.95 | 0.79 | 1.15 | 6.20E-01 | 0.833 |
| OID20344 | DEFA1_DEFA1B | Cardiometabolic | DEFA1;DEFA1B | P59665;P59665 | 8;8 | 6977649;6996766 | 6980092;6999198 | 1.04 | 0.88 | 1.24 | 6.21E-01 | 0.833 |
| OID30059 | GCLM | Cardiometabolic_II | GCLM | P48507 | 1 | 93885199 | 93909456 | 0.96 | 0.81 | 1.13 | 6.21E-01 | 0.833 |
| OID30367 | LCP1 | Cardiometabolic_II | LCP1 | P13796 | 13 | 46125920 | 46211871 | 0.96 | 0.81 | 1.14 | 6.21E-01 | 0.833 |
| OID20723 | SCGB3A2 | Inflammation | SCGB3A2 | Q96PL1 | 5 | 147870682 | 147882191 | 1.04 | 0.89 | 1.22 | 6.22E-01 | 0.833 |
| OID20056 | TSLP | Cardiometabolic | TSLP | Q969D9 | 5 | 111070062 | 111078026 | 0.96 | 0.81 | 1.14 | 6.21E-01 | 0.833 |
| OID21069 | CGA | Neurology | CGA | P01215 | 6 | 87085498 | 87095106 | 1.07 | 0.82 | 1.39 | 6.23E-01 | 0.834 |
| OID20273 | ALCAM | Cardiometabolic | ALCAM | Q13740 | 3 | 105366909 | 105576900 | 1.04 | 0.88 | 1.24 | 6.25E-01 | 0.835 |
| OID20693 | CCL23 | Inflammation | CCL23 | P55773 | 17 | 36013056 | 36017972 | 0.96 | 0.82 | 1.13 | 6.24E-01 | 0.835 |
| OID30984 | RBFOX3 | Neurology_II | RBFOX3 | A6NFN3 | 17 | 79089345 | 79611051 | 0.96 | 0.81 | 1.13 | 6.25E-01 | 0.835 |
| OID20321 | RNASET2 | Cardiometabolic | RNASET2 | O00584 | 6 | 166922113 | 166957191 | 0.96 | 0.81 | 1.14 | 6.25E-01 | 0.835 |
| OID30770 | HRG | Inflammation_II | HRG | P04196 | 3 | 186660216 | 186678234 | 0.96 | 0.81 | 1.14 | 6.27E-01 | 0.837 |
| OID20537 | SIGLEC10 | Inflammation | SIGLEC10 | Q96LC7 | 19 | 51410020 | 51417803 | 1.04 | 0.88 | 1.23 | 6.27E-01 | 0.837 |
| OID30423 | SPRED2 | Inflammation_II | SPRED2 | Q7Z698 | 2 | 65310851 | 65432637 | 0.96 | 0.81 | 1.14 | 6.27E-01 | 0.837 |
| OID30337 | HEPH | Cardiometabolic_II | HEPH | Q9BQS7 | X | 66162671 | 66268867 | 0.96 | 0.80 | 1.14 | 6.29E-01 | 0.837 |
| OID30157 | HMGCS1 | Cardiometabolic_II | HMGCS1 | Q01581 | 5 | 43287470 | 43313512 | 1.04 | 0.89 | 1.22 | 6.28E-01 | 0.837 |
| OID20428 | IL33 | Inflammation | IL33 | O95760 | 9 | 6215786 | 6257983 | 0.96 | 0.82 | 1.13 | 6.29E-01 | 0.837 |
| OID30141 | MANSC4 | Cardiometabolic_II | MANSC4 | A6NHS7 | 12 | 27762427 | 27780236 | 1.04 | 0.89 | 1.21 | 6.29E-01 | 0.837 |
| OID20224 | ROR1 | Cardiometabolic | ROR1 | Q01973 | 1 | 63774017 | 64181498 | 1.04 | 0.89 | 1.22 | 6.29E-01 | 0.837 |
| OID31322 | SERPINH1 | Oncology_II | SERPINH1 | P50454 | 11 | 75562056 | 75572783 | 0.96 | 0.81 | 1.13 | 6.29E-01 | 0.837 |
| OID20675 | CXCL9 | Inflammation | CXCL9 | Q07325 | 4 | 76001275 | 76007509 | 1.04 | 0.88 | 1.24 | 6.29E-01 | 0.837 |
| OID20700 | IL1RN | Inflammation | IL1RN | P18510 | 2 | 113107214 | 113134016 | 1.05 | 0.87 | 1.26 | 6.30E-01 | 0.837 |
| OID21106 | NOMO1 | Neurology | NOMO1 | Q15155 | 16 | 14833721 | 14896157 | 0.96 | 0.81 | 1.13 | 6.30E-01 | 0.837 |
| OID20237 | UMOD | Cardiometabolic | UMOD | P07911 | 16 | 20333051 | 20356301 | 1.04 | 0.88 | 1.24 | 6.30E-01 | 0.838 |
| OID20546 | CCL26 | Inflammation | CCL26 | Q9Y258 | 7 | 75769533 | 75789896 | 1.04 | 0.89 | 1.20 | 6.32E-01 | 0.838 |
| OID21274 | IGF1R | Oncology | IGF1R | P08069 | 15 | 98648539 | 98964530 | 1.04 | 0.88 | 1.22 | 6.31E-01 | 0.838 |
| OID31051 | S100G | Neurology_II | S100G | P29377 | X | 16650158 | 16654670 | 0.96 | 0.82 | 1.13 | 6.32E-01 | 0.838 |
| OID21260 | ENTPD2 | Oncology | ENTPD2 | Q9Y5L3 | 9 | 137048107 | 137054061 | 0.96 | 0.82 | 1.13 | 6.33E-01 | 0.839 |
| OID30171 | UHRF2 | Cardiometabolic_II | UHRF2 | Q96PU4 | 9 | 6413151 | 6507054 | 1.04 | 0.87 | 1.25 | 6.34E-01 | 0.840 |
| OID20801 | CALB2 | Neurology | CALB2 | P22676 | 16 | 71358713 | 71390436 | 0.96 | 0.81 | 1.14 | 6.34E-01 | 0.840 |
| OID21149 | CA2 | Neurology | CA2 | P00918 | 8 | 85463968 | 85481493 | 0.96 | 0.81 | 1.14 | 6.37E-01 | 0.843 |
| OID30819 | GPR101 | Neurology_II | GPR101 | Q96P66 | X | 137023929 | 137033995 | 0.96 | 0.81 | 1.14 | 6.37E-01 | 0.843 |
| OID31390 | COMMD9 | Oncology_II | COMMD9 | Q9P000 | 11 | 36269284 | 36289449 | 0.96 | 0.80 | 1.15 | 6.38E-01 | 0.843 |
| OID20329 | PRTN3 | Cardiometabolic | PRTN3 | P24158 | 19 | 840999 | 848175 | 1.04 | 0.88 | 1.23 | 6.38E-01 | 0.843 |
| OID30379 | COL3A1 | Cardiometabolic_II | COL3A1 | P02461 | 2 | 188974373 | 189012746 | 0.96 | 0.81 | 1.13 | 6.38E-01 | 0.843 |
| OID30402 | IGDCC4 | Cardiometabolic_II | IGDCC4 | Q8TDY8 | 15 | 65381484 | 65422947 | 1.04 | 0.88 | 1.24 | 6.38E-01 | 0.843 |
| OID31311 | PKN3 | Oncology_II | PKN3 | Q6P5Z2 | 9 | 128702503 | 128720916 | 0.96 | 0.80 | 1.14 | 6.39E-01 | 0.844 |
| OID21368 | CCN1 | Oncology | CCN1 | O00622 | 1 | 85580761 | 85584589 | 0.96 | 0.82 | 1.13 | 6.40E-01 | 0.844 |
| OID20286 | CORO1A | Cardiometabolic | CORO1A | P31146 | 16 | 30182827 | 30189076 | 0.96 | 0.81 | 1.14 | 6.40E-01 | 0.844 |
| OID30453 | RPA2 | Inflammation_II | RPA2 | P15927 | 1 | 27891524 | 27914746 | 0.96 | 0.81 | 1.14 | 6.40E-01 | 0.844 |
| OID21443 | PRTG | Oncology | PRTG | Q2VWP7 | 15 | 55611544 | 55743152 | 1.04 | 0.88 | 1.23 | 6.40E-01 | 0.844 |
| OID30963 | PTH | Neurology_II | PTH | P01270 | 11 | 13492054 | 13496181 | 1.04 | 0.89 | 1.21 | 6.40E-01 | 0.844 |
| OID20460 | WNT9A | Inflammation | WNT9A | O14904 | 1 | 227918656 | 227947932 | 1.04 | 0.87 | 1.24 | 6.42E-01 | 0.845 |
| OID20407 | ANGPTL3 | Cardiometabolic | ANGPTL3 | Q9Y5C1 | 1 | 62597520 | 62606313 | 0.96 | 0.81 | 1.14 | 6.43E-01 | 0.845 |
| OID21397 | EPS8L2 | Oncology | EPS8L2 | Q9H6S3 | 11 | 694438 | 727727 | 0.96 | 0.81 | 1.14 | 6.43E-01 | 0.845 |
| OID30084 | PDE4D | Cardiometabolic_II | PDE4D | Q08499 | 5 | 58969038 | 60522120 | 0.96 | 0.80 | 1.15 | 6.42E-01 | 0.845 |
| OID20221 | SPON2 | Cardiometabolic | SPON2 | Q9BUD6 | 4 | 1166932 | 1208962 | 1.04 | 0.88 | 1.22 | 6.44E-01 | 0.847 |
| OID20050 | TNNI3 | Cardiometabolic | TNNI3 | P19429 | 19 | 55151767 | 55157773 | 1.04 | 0.88 | 1.23 | 6.47E-01 | 0.850 |
| OID20271 | CA3 | Cardiometabolic | CA3 | P07451 | 8 | 85373436 | 85449040 | 0.96 | 0.81 | 1.14 | 6.48E-01 | 0.850 |
| OID20704 | CST7 | Inflammation | CST7 | O76096 | 20 | 24949269 | 24959928 | 0.96 | 0.80 | 1.15 | 6.48E-01 | 0.850 |
| OID21398 | XCL1 | Oncology | XCL1 | P47992 | 1 | 168576605 | 168582069 | 0.96 | 0.81 | 1.14 | 6.47E-01 | 0.850 |
| OID31309 | EVPL | Oncology_II | EVPL | Q92817 | 17 | 76004502 | 76027306 | 1.04 | 0.88 | 1.23 | 6.48E-01 | 0.851 |
| OID20280 | CPA1 | Cardiometabolic | CPA1 | P15085 | 7 | 130380339 | 130388114 | 0.97 | 0.83 | 1.12 | 6.49E-01 | 0.851 |
| OID31110 | PLB1 | Neurology_II | PLB1 | Q6P1J6 | 2 | 28457145 | 28644142 | 1.04 | 0.87 | 1.24 | 6.51E-01 | 0.853 |
| OID30177 | CBX2 | Cardiometabolic_II | CBX2 | Q14781 | 17 | 79778148 | 79787983 | 1.03 | 0.89 | 1.20 | 6.53E-01 | 0.854 |
| OID20292 | COL6A3 | Cardiometabolic | COL6A3 | P12111 | 2 | 237324003 | 237414328 | 1.04 | 0.88 | 1.22 | 6.52E-01 | 0.854 |
| OID30372 | DDT | Cardiometabolic_II | DDT | P30046 | 22 | 23970542 | 23980525 | 0.96 | 0.80 | 1.15 | 6.53E-01 | 0.854 |
| OID20349 | F9 | Cardiometabolic | F9 | P00740 | X | 139530739 | 139563459 | 1.04 | 0.88 | 1.23 | 6.52E-01 | 0.854 |
| OID20196 | KITLG | Cardiometabolic | KITLG | P21583 | 12 | 88492793 | 88580851 | 0.96 | 0.82 | 1.13 | 6.53E-01 | 0.854 |
| OID31069 | OCLN | Neurology_II | OCLN | Q16625 | 5 | 69492292 | 69558104 | 0.96 | 0.81 | 1.14 | 6.53E-01 | 0.854 |
| OID20856 | BAX | Neurology | BAX | Q07812 | 19 | 48954815 | 48961798 | 0.96 | 0.81 | 1.14 | 6.55E-01 | 0.855 |
| OID31469 | GPIHBP1 | Oncology_II | GPIHBP1 | Q8IV16 | 8 | 143213218 | 143217170 | 1.04 | 0.88 | 1.23 | 6.55E-01 | 0.855 |
| OID30824 | HTR1A | Neurology_II | HTR1A | P08908 | 5 | 63957874 | 63962507 | 1.04 | 0.89 | 1.21 | 6.54E-01 | 0.855 |
| OID30162 | LILRA4 | Cardiometabolic_II | LILRA4 | P59901 | 19 | 54333185 | 54339162 | 0.96 | 0.81 | 1.14 | 6.55E-01 | 0.855 |
| OID30095 | TSPAN15 | Cardiometabolic_II | TSPAN15 | O95858 | 10 | 69451465 | 69507666 | 1.04 | 0.89 | 1.21 | 6.55E-01 | 0.855 |
| OID21308 | NUDT2 | Oncology | NUDT2 | P50583 | 9 | 34329506 | 34343711 | 0.96 | 0.81 | 1.14 | 6.56E-01 | 0.855 |
| OID31363 | BTN1A1 | Oncology_II | BTN1A1 | Q13410 | 6 | 26500303 | 26510425 | 0.96 | 0.82 | 1.13 | 6.57E-01 | 0.856 |
| OID30836 | DOCK9 | Neurology_II | DOCK9 | Q9BZ29 | 13 | 98793429 | 99086625 | 1.03 | 0.89 | 1.20 | 6.57E-01 | 0.856 |
| OID20963 | LPO | Neurology | LPO | P22079 | 17 | 58218548 | 58268518 | 0.96 | 0.81 | 1.14 | 6.58E-01 | 0.857 |
| OID20999 | ACVRL1 | Neurology | ACVRL1 | P37023 | 12 | 51906908 | 51923361 | 0.96 | 0.82 | 1.13 | 6.59E-01 | 0.857 |
| OID20102 | CEACAM8 | Cardiometabolic | CEACAM8 | P31997 | 19 | 42580243 | 42595055 | 0.96 | 0.81 | 1.15 | 6.59E-01 | 0.857 |

| | | | | | | | | | | | | |
|---|---|---|---|---|---|---|---|---|---|---|---|---|
| OID20968 | FRZB | Neurology | FRZB | Q92765 | 2 | 182833275 | 182866637 | 0.96 | 0.80 | 1.15 | 6.60E-01 | 0.857 |
| OID31516 | PCDH9 | Oncology_II | PCDH9 | Q9HC56 | 13 | 66302834 | 67230445 | 0.96 | 0.81 | 1.14 | 6.59E-01 | 0.857 |
| OID31377 | PCDHB15 | Oncology_II | PCDHB15 | Q9Y5E8 | 5 | 141245395 | 141249365 | 0.96 | 0.81 | 1.14 | 6.59E-01 | 0.857 |
| OID21009 | DDR1 | Neurology | DDR1 | Q08345 | 6 | 30876421 | 30900156 | 1.04 | 0.88 | 1.21 | 6.61E-01 | 0.857 |
| OID20844 | TBC1D17 | Neurology | TBC1D17 | Q9HA65 | 19 | 49877666 | 49888750 | 0.96 | 0.80 | 1.15 | 6.61E-01 | 0.857 |
| OID31290 | TLR2 | Oncology_II | TLR2 | O60603 | 4 | 153684070 | 153706260 | 0.96 | 0.82 | 1.14 | 6.61E-01 | 0.857 |
| OID30593 | CELSR2 | Inflammation_II | CELSR2 | Q9HCU4 | 1 | 109249539 | 109275751 | 1.04 | 0.88 | 1.21 | 6.62E-01 | 0.858 |
| OID20219 | SERPINA12 | Cardiometabolic | SERPINA12 | Q8IW75 | 14 | 94487274 | 94517844 | 1.04 | 0.88 | 1.23 | 6.62E-01 | 0.859 |
| OID21039 | ADAM8 | Neurology | ADAM8 | P78325 | 10 | 133262420 | 133276868 | 0.97 | 0.82 | 1.13 | 6.63E-01 | 0.859 |
| OID30801 | ZPR1 | Neurology_II | ZPR1 | O75312 | 11 | 116773799 | 116788039 | 0.97 | 0.82 | 1.13 | 6.63E-01 | 0.859 |
| OID30142 | ECI2 | Cardiometabolic_II | ECI2 | O75521 | 6 | 4115693 | 4135597 | 0.96 | 0.82 | 1.14 | 6.64E-01 | 0.859 |
| OID30788 | FGA | Inflammation_II | FGA | P02671 | 4 | 154583128 | 154590745 | 0.96 | 0.80 | 1.15 | 6.64E-01 | 0.859 |
| OID31441 | SLMAP | Oncology_II | SLMAP | Q14BN4 | 3 | 57755450 | 57930003 | 0.96 | 0.81 | 1.14 | 6.64E-01 | 0.859 |
| OID30254 | FSTL1 | Cardiometabolic_II | FSTL1 | Q12841 | 3 | 120392293 | 120450993 | 1.04 | 0.88 | 1.23 | 6.66E-01 | 0.861 |
| OID31249 | CEP152 | Oncology_II | CEP152 | O94986 | 15 | 48712928 | 48811146 | 0.96 | 0.81 | 1.15 | 6.67E-01 | 0.862 |
| OID21129 | PILRA | Neurology | PILRA | Q9UKJ1 | 7 | 100367530 | 100400096 | 0.97 | 0.82 | 1.13 | 6.67E-01 | 0.862 |
| OID20668 | CCL11 | Inflammation | CCL11 | P51671 | 17 | 34285742 | 34288334 | 1.04 | 0.87 | 1.23 | 6.70E-01 | 0.865 |
| OID30765 | LGALS3BP | Inflammation_II | LGALS3BP | Q08380 | 17 | 78971238 | 78979947 | 0.96 | 0.81 | 1.15 | 6.71E-01 | 0.865 |
| OID21206 | MAEA | Oncology | MAEA | Q7L5Y9 | 4 | 1289887 | 1340147 | 1.04 | 0.88 | 1.22 | 6.71E-01 | 0.865 |
| OID20327 | RETN | Cardiometabolic | RETN | Q9HD89 | 19 | 7669049 | 7670455 | 1.04 | 0.88 | 1.23 | 6.71E-01 | 0.865 |
| OID21400 | SCG2 | Oncology | SCG2 | P13521 | 2 | 223596940 | 223602361 | 0.97 | 0.82 | 1.14 | 6.71E-01 | 0.865 |
| OID20945 | LY96 | Neurology | LY96 | Q9Y6Y9 | 8 | 73991392 | 74029079 | 1.04 | 0.88 | 1.22 | 6.72E-01 | 0.865 |
| OID20081 | EDIL3 | Cardiometabolic | EDIL3 | O43854 | 5 | 83940554 | 84384880 | 1.04 | 0.87 | 1.23 | 6.73E-01 | 0.866 |
| OID31239 | MRPL52 | Oncology_II | MRPL52 | Q86TS9 | 14 | 22829879 | 22835037 | 0.96 | 0.81 | 1.15 | 6.73E-01 | 0.866 |
| OID20455 | IL11 | Inflammation | IL11 | P20809 | 19 | 55364382 | 55370463 | 1.04 | 0.87 | 1.24 | 6.75E-01 | 0.868 |
| OID21141 | BLVRB | Neurology | BLVRB | P30043 | 19 | 40447765 | 40465764 | 0.96 | 0.81 | 1.14 | 6.76E-01 | 0.869 |
| OID21418 | CD300E | Oncology | CD300E | Q496F6 | 17 | 74609885 | 74623738 | 1.04 | 0.88 | 1.22 | 6.78E-01 | 0.871 |
| OID20134 | CDH17 | Cardiometabolic | CDH17 | Q12864 | 8 | 94127162 | 94217303 | 0.96 | 0.81 | 1.15 | 6.77E-01 | 0.871 |
| OID30755 | CFB | Inflammation_II | CFB | P00751 | 6 | 31945650 | 31952086 | 1.04 | 0.86 | 1.25 | 6.78E-01 | 0.871 |
| OID30954 | EFNB2 | Neurology_II | EFNB2 | P52799 | 13 | 106489745 | 106535662 | 0.96 | 0.81 | 1.15 | 6.78E-01 | 0.871 |
| OID30155 | FRMD4B | Cardiometabolic_II | FRMD4B | Q9Y2L6 | 3 | 69168782 | 69542583 | 0.97 | 0.82 | 1.14 | 6.79E-01 | 0.871 |
| OID30087 | NOS2 | Cardiometabolic_II | NOS2 | P35228 | 17 | 27756766 | 27800529 | 0.96 | 0.81 | 1.15 | 6.79E-01 | 0.871 |
| OID30412 | TNN | Cardiometabolic_II | TNN | Q9UQP3 | 1 | 175067833 | 175148075 | 1.04 | 0.88 | 1.22 | 6.79E-01 | 0.871 |
| OID21473 | PLA2G15 | Oncology | PLA2G15 | Q8NCC3 | 16 | 68245304 | 68261058 | 0.97 | 0.82 | 1.14 | 6.80E-01 | 0.871 |
| OID31436 | SDHB | Oncology_II | SDHB | P21912 | 1 | 17018722 | 17054032 | 0.97 | 0.82 | 1.14 | 6.82E-01 | 0.873 |
| OID30441 | IL21R | Inflammation_II | IL21R | Q9HBE5 | 16 | 27402174 | 27452042 | 0.97 | 0.82 | 1.14 | 6.83E-01 | 0.874 |
| OID30426 | KDM3A | Inflammation_II | KDM3A | Q9Y4C1 | 2 | 86440647 | 86492716 | 0.96 | 0.81 | 1.15 | 6.83E-01 | 0.874 |
| OID30864 | LMOD2 | Neurology_II | LMOD2 | Q6P5Q4 | 7 | 123655866 | 123664290 | 0.96 | 0.81 | 1.15 | 6.83E-01 | 0.874 |
| OID30089 | NECAP2 | Cardiometabolic_II | NECAP2 | Q9NVZ3 | 1 | 16440721 | 16460078 | 0.97 | 0.82 | 1.14 | 6.84E-01 | 0.874 |
| OID21108 | CD109 | Neurology | CD109 | Q6YHK3 | 6 | 73695785 | 73828316 | 0.97 | 0.82 | 1.14 | 6.85E-01 | 0.875 |
| OID30399 | CILP | Cardiometabolic_II | CILP | O75339 | 15 | 65194760 | 65211473 | 0.97 | 0.81 | 1.14 | 6.86E-01 | 0.876 |
| OID20355 | LBP | Cardiometabolic | LBP | P18428 | 20 | 38346482 | 38377013 | 0.97 | 0.82 | 1.14 | 6.85E-01 | 0.876 |
| OID30683 | SOD3 | Inflammation_II | SOD3 | P08294 | 4 | 24789912 | 24800842 | 0.96 | 0.79 | 1.17 | 6.86E-01 | 0.876 |
| OID30747 | LPA | Inflammation_II | LPA | P08519 | 6 | 160531482 | 160664275 | 0.97 | 0.83 | 1.13 | 6.87E-01 | 0.876 |
| OID31388 | SLC13A1 | Oncology_II | SLC13A1 | Q9BZW2 | 7 | 123113531 | 123199972 | 1.03 | 0.88 | 1.21 | 6.87E-01 | 0.876 |
| OID20679 | CTSC | Inflammation | CTSC | P53634 | 11 | 88265069 | 88359684 | 1.03 | 0.88 | 1.22 | 6.88E-01 | 0.877 |
| OID21240 | STXBP3 | Oncology | STXBP3 | O00186 | 1 | 108746674 | 108809523 | 0.97 | 0.82 | 1.14 | 6.89E-01 | 0.877 |
| OID20692 | CD48 | Inflammation | CD48 | P09326 | 1 | 160678746 | 160711831 | 0.97 | 0.82 | 1.14 | 6.89E-01 | 0.878 |
| OID30230 | IDO1 | Cardiometabolic_II/Inflan | IDO1 | P14902 | 8 | 39902275 | 39928790 | 0.97 | 0.81 | 1.15 | 6.90E-01 | 0.878 |
| OID21067 | BIN2 | Neurology | BIN2 | Q9UBW5 | 12 | 51281038 | 51324668 | 0.97 | 0.82 | 1.14 | 6.90E-01 | 0.878 |
| OID20807 | MMP13 | Neurology | MMP13 | P45452 | 11 | 102942995 | 102955732 | 0.97 | 0.83 | 1.13 | 6.90E-01 | 0.878 |
| OID20513 | BCL2L11 | Inflammation | BCL2L11 | O43521 | 2 | 111119378 | 111168445 | 0.97 | 0.82 | 1.14 | 6.91E-01 | 0.878 |
| OID30854 | ELAVL4 | Neurology_II | ELAVL4 | P26378 | 1 | 50024029 | 50203772 | 0.97 | 0.82 | 1.14 | 6.92E-01 | 0.879 |
| OID20556 | CNTNAP2 | Inflammation | CNTNAP2 | Q9UHC6 | 7 | 146116002 | 148420998 | 0.97 | 0.82 | 1.14 | 6.92E-01 | 0.879 |
| OID20424 | JUN | Inflammation | JUN | P05412 | 1 | 58776845 | 58784048 | 1.03 | 0.88 | 1.21 | 6.93E-01 | 0.879 |
| OID30276 | PRRT3 | Cardiometabolic_II | PRRT3 | Q5FWE3 | 3 | 9945542 | 9952408 | 1.03 | 0.88 | 1.22 | 6.94E-01 | 0.881 |
| OID30517 | TYRP1 | Inflammation_II | TYRP1 | P17643 | 9 | 12685439 | 12710285 | 0.97 | 0.83 | 1.14 | 6.94E-01 | 0.881 |
| OID21499 | KLK1 | Oncology | KLK1 | P06870 | 19 | 50819146 | 50823787 | 1.03 | 0.88 | 1.21 | 6.95E-01 | 0.881 |
| OID20763 | PRSS8 | Inflammation | PRSS8 | Q16651 | 16 | 31131433 | 31135727 | 1.04 | 0.87 | 1.24 | 6.95E-01 | 0.881 |
| OID31228 | PAIP2B | Oncology_II | PAIP2B | Q9ULR5 | 2 | 71182738 | 71227103 | 1.03 | 0.88 | 1.21 | 6.96E-01 | 0.882 |
| OID21033 | PRL | Neurology | PRL | P01236 | 6 | 22287244 | 22302826 | 0.97 | 0.83 | 1.13 | 6.96E-01 | 0.882 |
| OID30456 | CCNE1 | Inflammation_II | CCNE1 | P24864 | 19 | 29811991 | 29824312 | 1.03 | 0.88 | 1.21 | 6.97E-01 | 0.882 |
| OID20430 | IL13 | Inflammation | IL13 | P35225 | 5 | 132656263 | 132661110 | 0.97 | 0.84 | 1.13 | 6.97E-01 | 0.882 |
| OID20318 | GAS6 | Cardiometabolic | GAS6 | Q14393 | 13 | 113820549 | 113864076 | 1.03 | 0.88 | 1.22 | 6.99E-01 | 0.883 |
| OID30235 | RNF149 | Cardiometabolic_II | RNF149 | Q8NC42 | 2 | 101271219 | 101308701 | 0.97 | 0.83 | 1.13 | 6.98E-01 | 0.883 |
| OID30825 | OTOA | Neurology_II | OTOA | Q7RTW8 | 16 | 21663968 | 21761935 | 1.03 | 0.88 | 1.22 | 7.00E-01 | 0.884 |
| OID20375 | TIMD4 | Cardiometabolic | TIMD4 | Q96H15 | 5 | 156919292 | 156963226 | 1.03 | 0.88 | 1.21 | 7.00E-01 | 0.884 |
| OID20393 | CR2 | Cardiometabolic | CR2 | P20023 | 1 | 207454230 | 207489895 | 1.03 | 0.88 | 1.21 | 7.00E-01 | 0.884 |
| OID30721 | C1RL | Inflammation_II | C1RL | Q9NZP8 | 12 | 7089587 | 7109238 | 1.04 | 0.87 | 1.24 | 7.01E-01 | 0.884 |
| OID20495 | IFNG | Inflammation | IFNG | P01579 | 12 | 68154768 | 68159740 | 1.03 | 0.88 | 1.21 | 7.01E-01 | 0.884 |
| OID21049 | TNFRSF8 | Neurology | TNFRSF8 | P28908 | 1 | 12063303 | 12144207 | 1.03 | 0.88 | 1.20 | 7.01E-01 | 0.884 |
| OID20083 | PTN | Cardiometabolic | PTN | P21246 | 7 | 137227341 | 137343774 | 0.97 | 0.81 | 1.15 | 7.02E-01 | 0.884 |
| OID31202 | BEX3 | Oncology_II | BEX3 | Q00994 | X | 103376395 | 103378164 | 0.97 | 0.81 | 1.16 | 7.03E-01 | 0.885 |
| OID30417 | GOT1 | Cardiometabolic_II | GOT1 | P17174 | 10 | 99396870 | 99430624 | 0.97 | 0.81 | 1.15 | 7.03E-01 | 0.885 |
| OID20566 | NCR1 | Inflammation | NCR1 | O76036 | 19 | 54906148 | 54916140 | 1.03 | 0.88 | 1.21 | 7.02E-01 | 0.885 |
| OID20533 | TRIM21 | Inflammation | TRIM21 | P19474 | 11 | 4384897 | 4393702 | 0.97 | 0.83 | 1.13 | 7.03E-01 | 0.885 |
| OID20734 | ROBO1 | Inflammation | ROBO1 | Q9Y6N7 | 3 | 78597239 | 79767998 | 1.03 | 0.88 | 1.21 | 7.04E-01 | 0.885 |
| OID30810 | TSPAN7 | Neurology_II | TSPAN7 | P41732 | X | 38561542 | 38688920 | 1.04 | 0.87 | 1.24 | 7.04E-01 | 0.885 |
| OID30447 | UNC5D | Inflammation_II | UNC5D | Q6UXZ4 | 8 | 35235475 | 35796550 | 0.97 | 0.82 | 1.14 | 7.04E-01 | 0.885 |
| OID30159 | MMUT | Cardiometabolic_II | MMUT | P22033 | 6 | 49430360 | 49463253 | 0.97 | 0.82 | 1.15 | 7.05E-01 | 0.885 |
| OID30439 | SIRT1 | Inflammation_II | SIRT1 | Q96EB6 | 10 | 67884656 | 67918390 | 0.97 | 0.81 | 1.16 | 7.05E-01 | 0.885 |
| OID20531 | STX8 | Inflammation | STX8 | Q9UNK0 | 17 | 9250471 | 9576591 | 0.97 | 0.81 | 1.15 | 7.05E-01 | 0.885 |
| OID30322 | CPXM2 | Cardiometabolic_II | CPXM2 | Q8N436 | 10 | 123706207 | 123940267 | 1.03 | 0.87 | 1.22 | 7.06E-01 | 0.885 |
| OID30454 | FGF12 | Inflammation_II | FGF12 | P61328 | 3 | 192139390 | 192767764 | 0.96 | 0.80 | 1.17 | 7.06E-01 | 0.885 |
| OID31089 | SYT1 | Neurology_II | SYT1 | P21579 | 12 | 78863993 | 79452008 | 1.03 | 0.89 | 1.18 | 7.06E-01 | 0.885 |
| OID20062 | AKR1C4 | Cardiometabolic | AKR1C4 | P17516 | 10 | 5195462 | 5218949 | 1.03 | 0.89 | 1.19 | 7.08E-01 | 0.886 |
| OID21092 | CNTN3 | Neurology | CNTN3 | Q9P232 | 3 | 74262568 | 74614659 | 0.97 | 0.80 | 1.16 | 7.08E-01 | 0.886 |
| OID20737 | LAIR1 | Inflammation | LAIR1 | Q6GTX8 | 19 | 54351384 | 54370558 | 1.03 | 0.86 | 1.24 | 7.08E-01 | 0.886 |
| OID31256 | CD82 | Oncology_II | CD82 | P27701 | 11 | 44564427 | 44620358 | 0.97 | 0.82 | 1.14 | 7.10E-01 | 0.887 |
| OID30240 | CEP170 | Cardiometabolic_II | CEP170 | Q5SW79 | 1 | 243124428 | 243255348 | 0.97 | 0.82 | 1.15 | 7.09E-01 | 0.887 |
| OID20311 | NOTCH1 | Cardiometabolic | NOTCH1 | P46531 | 9 | 136494433 | 136546048 | 0.97 | 0.82 | 1.14 | 7.10E-01 | 0.887 |
| OID30783 | TF | Inflammation_II | TF | P02787 | 3 | 133746040 | 133796641 | 0.97 | 0.82 | 1.14 | 7.10E-01 | 0.887 |
| OID20265 | TNFRSF10C | Cardiometabolic | TNFRSF10C | O14798 | 8 | 23102921 | 23117445 | 1.03 | 0.87 | 1.22 | 7.09E-01 | 0.887 |
| OID20479 | WAS | Inflammation | WAS | P42768 | X | 48676596 | 48691427 | 1.03 | 0.87 | 1.22 | 7.10E-01 | 0.887 |
| OID20792 | NDRG1 | Neurology | NDRG1 | Q92597 | 8 | 133237175 | 133302022 | 0.97 | 0.82 | 1.15 | 7.11E-01 | 0.887 |
| OID30343 | PTPRZ1 | Cardiometabolic_II | PTPRZ1 | P23471 | 7 | 121873089 | 122062036 | 1.03 | 0.87 | 1.23 | 7.12E-01 | 0.888 |
| OID20919 | ULBP2 | Neurology | ULBP2 | Q9BZM5 | 6 | 149942014 | 149949235 | 1.03 | 0.87 | 1.22 | 7.12E-01 | 0.888 |
| OID31007 | C1QBP | Neurology_II | C1QBP | Q07021 | 17 | 5432777 | 5448830 | 0.97 | 0.81 | 1.15 | 7.14E-01 | 0.889 |
| OID21408 | HSPB6 | Oncology | HSPB6 | O14558 | 19 | 35754566 | 35758079 | 1.04 | 0.86 | 1.25 | 7.14E-01 | 0.889 |
| OID31448 | INSL5 | Oncology_II | INSL5 | Q9Y5Q6 | 1 | 66797740 | 66801276 | 0.97 | 0.81 | 1.15 | 7.13E-01 | 0.889 |
| OID20796 | SMARCA2 | Neurology | SMARCA2 | P51531 | 9 | 1980290 | 2193624 | 0.97 | 0.82 | 1.15 | 7.15E-01 | 0.889 |
| OID30650 | SUSD5 | Inflammation_II | SUSD5 | O60279 | 3 | 33150043 | 33218810 | 1.03 | 0.87 | 1.22 | 7.15E-01 | 0.889 |
| OID21527 | CD27 | Oncology | CD27 | P26842 | 12 | 6444955 | 6451718 | 0.97 | 0.82 | 1.14 | 7.17E-01 | 0.890 |
| OID30845 | LELP1 | Neurology_II | LELP1 | Q5T871 | 1 | 153203430 | 153205120 | 0.97 | 0.83 | 1.14 | 7.16E-01 | 0.890 |
| OID20776 | OSCAR | Inflammation | OSCAR | Q8IYS5 | 19 | 54094668 | 54102692 | 0.97 | 0.83 | 1.14 | 7.17E-01 | 0.890 |
| OID30833 | SCN2A | Neurology_II | SCN2A | Q99250 | 2 | 165194993 | 165392310 | 0.97 | 0.83 | 1.14 | 7.17E-01 | 0.890 |
| OID30101 | PGLYRP4 | Cardiometabolic_II | PGLYRP4 | Q96LB8 | 1 | 153330120 | 153348841 | 0.97 | 0.81 | 1.15 | 7.18E-01 | 0.892 |

| | | | | | | | | | | | | |
|---|---|---|---|---|---|---|---|---|---|---|---|---|
| OID20788 | CXCL3 | Inflammation | CXCL3 | P19876 | 4 | 74036589 | 74038807 | 0.97 | 0.82 | 1.14 | 7.19E-01 | 0.892 |
| OID30976 | DLGAP5 | Neurology_II | DLGAP5 | Q15398 | 14 | 55148112 | 55191608 | 0.97 | 0.80 | 1.16 | 7.20E-01 | 0.893 |
| OID20706 | NELL2 | Inflammation | NELL2 | Q99435 | 12 | 44508275 | 44921848 | 0.97 | 0.82 | 1.15 | 7.20E-01 | 0.893 |
| OID30486 | SUMF1 | Inflammation_II | SUMF1 | Q8NBK3 | 3 | 3700814 | 4467273 | 1.03 | 0.87 | 1.23 | 7.20E-01 | 0.893 |
| OID31316 | CCND2 | Oncology_II | CCND2 | P30279 | 12 | 4269771 | 4305353 | 1.03 | 0.87 | 1.22 | 7.21E-01 | 0.893 |
| OID30625 | INHBB | Inflammation_II | INHBB | P09529 | 2 | 120346136 | 120351803 | 1.03 | 0.88 | 1.21 | 7.22E-01 | 0.893 |
| OID20670 | LY9 | Inflammation | LY9 | Q9HBG7 | 1 | 160796074 | 160828261 | 1.03 | 0.88 | 1.21 | 7.22E-01 | 0.893 |
| OID30282 | POMC | Cardiometabolic_II | POMC | P01189 | 2 | 25160853 | 25168903 | 1.03 | 0.87 | 1.22 | 7.22E-01 | 0.893 |
| OID20306 | PTPRS | Cardiometabolic | PTPRS | Q13332 | 19 | 5158495 | 5340812 | 0.97 | 0.83 | 1.14 | 7.21E-01 | 0.893 |
| OID20413 | IGFBP6 | Cardiometabolic | IGFBP6 | P24592 | 12 | 53097436 | 53102345 | 0.97 | 0.82 | 1.15 | 7.23E-01 | 0.893 |
| OID20074 | TNF | Cardiometabolic/Inflamm | TNF | P01375 | 6 | 31575565 | 31578336 | 0.97 | 0.81 | 1.15 | 7.23E-01 | 0.894 |
| OID30532 | TLR1 | Inflammation_II | TLR1 | Q15399 | 4 | 38790677 | 38856817 | 1.03 | 0.87 | 1.21 | 7.24E-01 | 0.895 |
| OID30931 | FZD10 | Neurology_II | FZD10 | Q9ULW2 | 12 | 130162459 | 130165740 | 0.97 | 0.81 | 1.16 | 7.25E-01 | 0.896 |
| OID21517 | KLK8 | Oncology | KLK8 | O60259 | 19 | 50996007 | 51002711 | 1.03 | 0.88 | 1.20 | 7.27E-01 | 0.896 |
| OID20171 | MCFD2 | Cardiometabolic | MCFD2 | Q8NI22 | 2 | 46901870 | 46941855 | 0.97 | 0.82 | 1.15 | 7.27E-01 | 0.896 |
| OID30478 | TSC1 | Inflammation_II | TSC1 | Q92574 | 9 | 132891348 | 132946874 | 0.97 | 0.81 | 1.16 | 7.27E-01 | 0.897 |
| OID30484 | PLCB1 | Inflammation_II | PLCB1 | Q9NQ66 | 20 | 8077251 | 8968360 | 0.97 | 0.81 | 1.16 | 7.28E-01 | 0.897 |
| OID21164 | VTCN1 | Oncology | VTCN1 | Q7Z7D3 | 1 | 117143587 | 117210960 | 0.97 | 0.83 | 1.14 | 7.28E-01 | 0.897 |
| OID21318 | CDNF | Oncology | CDNF | Q49AH0 | 10 | 14819245 | 14838575 | 0.97 | 0.83 | 1.14 | 7.28E-01 | 0.897 |
| OID30165 | B3GAT3 | Cardiometabolic_II | B3GAT3 | O94766 | 11 | 62615296 | 62622154 | 0.97 | 0.84 | 1.13 | 7.30E-01 | 0.897 |
| OID20367 | C1QTNF1 | Cardiometabolic | C1QTNF1 | Q9BXJ1 | 17 | 79022814 | 79049788 | 0.97 | 0.82 | 1.15 | 7.31E-01 | 0.897 |
| OID21054 | CPM | Neurology | CPM | P14384 | 12 | 68842197 | 68971570 | 0.97 | 0.80 | 1.17 | 7.31E-01 | 0.897 |
| OID30822 | DNLZ | Neurology_II | DNLZ | Q5SXM8 | 9 | 136359480 | 136363744 | 1.03 | 0.87 | 1.22 | 7.29E-01 | 0.897 |
| OID21207 | FES | Oncology | FES | P07332 | 15 | 90883695 | 90895776 | 0.97 | 0.82 | 1.15 | 7.30E-01 | 0.897 |
| OID21309 | GRPEL1 | Oncology | GRPEL1 | Q9HAV7 | 4 | 7058895 | 7068064 | 1.03 | 0.87 | 1.21 | 7.32E-01 | 0.897 |
| OID30481 | HS6ST2 | Inflammation_II | HS6ST2 | Q96MM7 | X | 132626015 | 132961395 | 0.97 | 0.82 | 1.15 | 7.31E-01 | 0.897 |
| OID20824 | LIF | Neurology | LIF | P15018 | 22 | 30240453 | 30246759 | 0.97 | 0.82 | 1.15 | 7.32E-01 | 0.897 |
| OID21227 | NAMPT | Oncology | NAMPT | P43490 | 7 | 106248298 | 106285966 | 1.03 | 0.87 | 1.22 | 7.31E-01 | 0.897 |
| OID30419 | PSAP | Cardiometabolic_II | PSAP | P07602 | 10 | 71816298 | 71851251 | 0.97 | 0.83 | 1.14 | 7.29E-01 | 0.897 |
| OID30928 | TPBGL | Neurology_II | TPBGL | P0DKB5 | 11 | 75240774 | 75243704 | 0.97 | 0.82 | 1.15 | 7.31E-01 | 0.897 |
| OID20671 | CCL20 | Inflammation | CCL20 | P78556 | 2 | 227805739 | 227817564 | 1.03 | 0.87 | 1.21 | 7.32E-01 | 0.897 |
| OID20360 | CD163 | Cardiometabolic | CD163 | Q86VB7 | 12 | 7470811 | 7503893 | 1.03 | 0.87 | 1.21 | 7.33E-01 | 0.898 |
| OID31186 | CPLX2 | Oncology_II | CPLX2 | Q6PUV4 | 5 | 175796310 | 175884021 | 1.03 | 0.86 | 1.23 | 7.38E-01 | 0.899 |
| OID31091 | GFRAL | Neurology_II | GFRAL | Q6UXV0 | 6 | 55327469 | 55402493 | 0.97 | 0.83 | 1.14 | 7.35E-01 | 0.899 |
| OID31370 | INSL4 | Oncology_II | INSL4 | Q14641 | 9 | 5231419 | 5235304 | 1.03 | 0.87 | 1.22 | 7.37E-01 | 0.899 |
| OID31501 | LMNB1 | Oncology_II | LMNB1 | P20700 | 5 | 126776623 | 126837020 | 0.97 | 0.81 | 1.16 | 7.36E-01 | 0.899 |
| OID21293 | LRP1 | Oncology | LRP1 | Q07954 | 12 | 57128483 | 57213361 | 1.03 | 0.87 | 1.22 | 7.34E-01 | 0.899 |
| OID20185 | LRP11 | Cardiometabolic | LRP11 | Q86VZ4 | 6 | 149818757 | 149864359 | 0.97 | 0.83 | 1.14 | 7.38E-01 | 0.899 |
| OID20057 | MFAP3 | Cardiometabolic | MFAP3 | P55082 | 5 | 154038959 | 154220478 | 1.03 | 0.87 | 1.22 | 7.36E-01 | 0.899 |
| OID20451 | MYO9B | Inflammation | MYO9B | Q13459 | 19 | 17075777 | 17214537 | 0.97 | 0.82 | 1.15 | 7.38E-01 | 0.899 |
| OID20252 | PLA2G1B | Cardiometabolic | PLA2G1B | P04054 | 12 | 120322115 | 120327779 | 1.03 | 0.88 | 1.20 | 7.36E-01 | 0.899 |
| OID31248 | REST | Oncology_II | REST | Q13127 | 4 | 56907876 | 56966808 | 1.03 | 0.87 | 1.23 | 7.37E-01 | 0.899 |
| OID20948 | SETMAR | Neurology | SETMAR | Q53H47 | 3 | 4303332 | 4317265 | 0.97 | 0.80 | 1.17 | 7.35E-01 | 0.899 |
| OID30516 | SHH | Inflammation_II | SHH | Q15465 | 7 | 155799980 | 155812463 | 1.03 | 0.87 | 1.21 | 7.38E-01 | 0.899 |
| OID30468 | SSBP1 | Inflammation_II | SSBP1 | Q04837 | 7 | 141738334 | 141787922 | 0.97 | 0.82 | 1.15 | 7.38E-01 | 0.899 |
| OID21107 | TNXB | Neurology | TNXB | P22105 | 6 | 32041153 | 32115334 | 0.97 | 0.82 | 1.15 | 7.37E-01 | 0.899 |
| OID30585 | ACHE | Inflammation_II | ACHE | P22303 | 7 | 100889994 | 100896974 | 1.03 | 0.87 | 1.21 | 7.39E-01 | 0.899 |
| OID30347 | MELTF | Cardiometabolic_II | MELTF | P08582 | 3 | 196980590 | 197029835 | 0.97 | 0.83 | 1.14 | 7.40E-01 | 0.899 |
| OID30665 | OLFM4 | Inflammation_II | OLFM4 | Q6UX06 | 13 | 53028813 | 53052057 | 1.03 | 0.86 | 1.23 | 7.40E-01 | 0.899 |
| OID20622 | CXCL17 | Inflammation | CXCL17 | Q6UXB2 | 19 | 42428278 | 42442946 | 0.97 | 0.79 | 1.18 | 7.40E-01 | 0.900 |
| OID20277 | ICAM3 | Cardiometabolic | ICAM3 | P32942 | 19 | 10333776 | 10339661 | 1.03 | 0.88 | 1.20 | 7.41E-01 | 0.900 |
| OID20486 | IL12RB1 | Inflammation | IL12RB1 | P42701 | 19 | 18058995 | 18098944 | 1.03 | 0.88 | 1.20 | 7.40E-01 | 0.900 |
| OID21112 | CLEC11A | Neurology | CLEC11A | Q9Y240 | 19 | 50723364 | 50725708 | 0.97 | 0.83 | 1.14 | 7.41E-01 | 0.900 |
| OID20124 | ACOX1 | Cardiometabolic | ACOX1 | Q15067 | 17 | 75941507 | 75979177 | 0.97 | 0.83 | 1.14 | 7.44E-01 | 0.902 |
| OID20389 | AOC3 | Cardiometabolic | AOC3 | Q16853 | 17 | 42851184 | 42858130 | 1.03 | 0.87 | 1.21 | 7.44E-01 | 0.902 |
| OID20752 | CTRC | Inflammation | CTRC | Q99895 | 1 | 15438442 | 15449242 | 0.97 | 0.83 | 1.14 | 7.44E-01 | 0.902 |
| OID21305 | DPEP2 | Oncology | DPEP2 | Q9H4A9 | 16 | 67987394 | 68000586 | 0.97 | 0.83 | 1.15 | 7.45E-01 | 0.902 |
| OID30992 | FCER1A | Neurology_II | FCER1A | P12319 | 1 | 159289714 | 159308224 | 1.02 | 0.89 | 1.18 | 7.46E-01 | 0.902 |
| OID20296 | GUSB | Cardiometabolic | GUSB | P08236 | 7 | 65960684 | 65982215 | 1.03 | 0.87 | 1.22 | 7.45E-01 | 0.902 |
| OID30145 | HSD17B3 | Cardiometabolic_II | HSD17B3 | P37058 | 9 | 96235306 | 96302176 | 1.02 | 0.89 | 1.18 | 7.44E-01 | 0.902 |
| OID21378 | MUC16 | Oncology | MUC16 | Q8WXI7 | 19 | 8848844 | 8981342 | 1.03 | 0.87 | 1.21 | 7.43E-01 | 0.902 |
| OID31515 | PALM | Oncology_II | PALM | O75781 | 19 | 708935 | 748329 | 0.97 | 0.83 | 1.15 | 7.45E-01 | 0.902 |
| OID20233 | FAM3C | Cardiometabolic | FAM3C | Q92520 | 7 | 121348878 | 121396364 | 1.03 | 0.87 | 1.21 | 7.46E-01 | 0.902 |
| OID21199 | BTC | Oncology | BTC | P35070 | 4 | 74744759 | 74794523 | 0.97 | 0.82 | 1.15 | 7.47E-01 | 0.902 |
| OID30148 | GUK1 | Cardiometabolic_II | GUK1 | Q16774 | 1 | 228139962 | 228148984 | 1.03 | 0.87 | 1.21 | 7.47E-01 | 0.903 |
| OID30752 | SELL | Inflammation_II | SELL | P14151 | 1 | 169690665 | 169711702 | 0.97 | 0.82 | 1.15 | 7.48E-01 | 0.904 |
| OID20260 | ADGRE5 | Cardiometabolic | ADGRE5 | P48960 | 19 | 14380501 | 14408725 | 0.97 | 0.83 | 1.15 | 7.49E-01 | 0.904 |
| OID30759 | MBL2 | Inflammation_II | MBL2 | P11226 | 10 | 52765380 | 52772784 | 1.03 | 0.87 | 1.21 | 7.51E-01 | 0.906 |
| OID30460 | PGR | Inflammation_II | PGR | P06401 | 11 | 101029624 | 101129813 | 1.03 | 0.88 | 1.20 | 7.52E-01 | 0.907 |
| OID30537 | ITGAL | Inflammation_II | ITGAL | P20701 | 16 | 30472658 | 30523567 | 0.97 | 0.82 | 1.16 | 7.52E-01 | 0.907 |
| OID30622 | EPHA4 | Inflammation_II | EPHA4 | P54764 | 2 | 221418027 | 221574202 | 0.98 | 0.83 | 1.14 | 7.53E-01 | 0.907 |
| OID20291 | PAM | Cardiometabolic | PAM | P19021 | 5 | 102753981 | 103029730 | 1.03 | 0.88 | 1.20 | 7.53E-01 | 0.907 |
| OID30823 | DEFB116 | Neurology_II | DEFB116 | Q30KQ4 | 20 | 31303212 | 31308585 | 0.97 | 0.83 | 1.15 | 7.55E-01 | 0.908 |
| OID21492 | MANSC1 | Oncology | MANSC1 | Q9H8J5 | 12 | 12326056 | 12350242 | 1.03 | 0.87 | 1.20 | 7.54E-01 | 0.908 |
| OID20732 | MZB1 | Inflammation | MZB1 | Q8WU39 | 5 | 139387467 | 139390081 | 1.03 | 0.87 | 1.21 | 7.55E-01 | 0.909 |
| OID21404 | PRDX6 | Oncology | PRDX6 | P30041 | 1 | 173477330 | 173488815 | 1.03 | 0.87 | 1.20 | 7.56E-01 | 0.909 |
| OID30192 | ACTN2 | Cardiometabolic_II | ACTN2 | P35609 | 1 | 236664141 | 236764631 | 1.03 | 0.87 | 1.21 | 7.56E-01 | 0.909 |
| OID21481 | C4BPB | Oncology | C4BPB | P20851 | 1 | 207088860 | 207099993 | 1.03 | 0.87 | 1.22 | 7.57E-01 | 0.910 |
| OID30061 | MYH4 | Cardiometabolic_II | MYH4 | Q9Y623 | 17 | 10443290 | 10469559 | 0.97 | 0.82 | 1.15 | 7.59E-01 | 0.912 |
| OID30734 | CPB2 | Inflammation_II | CPB2 | Q96IY4 | 13 | 46053186 | 46105033 | 1.03 | 0.86 | 1.22 | 7.60E-01 | 0.912 |
| OID30458 | CEBPA | Inflammation_II | CEBPA | P49715 | 19 | 33299934 | 33302534 | 1.02 | 0.87 | 1.20 | 7.62E-01 | 0.914 |
| OID20798 | ATF2 | Neurology | ATF2 | P15336 | 2 | 175072250 | 175168382 | 1.03 | 0.86 | 1.22 | 7.63E-01 | 0.915 |
| OID31321 | RNF4 | Oncology_II | RNF4 | P78317 | 4 | 2462220 | 2515857 | 0.97 | 0.82 | 1.15 | 7.63E-01 | 0.915 |
| OID21178 | ATP6AP2 | Oncology | ATP6AP2 | O75787 | X | 40579372 | 40606848 | 0.97 | 0.82 | 1.16 | 7.64E-01 | 0.915 |
| OID31242 | CDC123 | Oncology_II | CDC123 | O75794 | 10 | 12195965 | 12250589 | 0.97 | 0.81 | 1.17 | 7.64E-01 | 0.915 |
| OID30919 | HNF1A | Neurology_II | HNF1A | P20823 | 12 | 120978543 | 121002512 | 0.97 | 0.82 | 1.16 | 7.65E-01 | 0.915 |
| OID21142 | RELT | Neurology | RELT | Q969Z4 | 11 | 73376399 | 73397474 | 0.97 | 0.82 | 1.16 | 7.65E-01 | 0.915 |
| OID20662 | VEGFD | Inflammation | VEGFD | O43915 | X | 15345596 | 15384413 | 1.03 | 0.87 | 1.22 | 7.65E-01 | 0.915 |
| OID31264 | GTF2IRD1 | Oncology_II | GTF2IRD1 | Q9UHL9 | 7 | 74453790 | 74602605 | 0.97 | 0.82 | 1.16 | 7.65E-01 | 0.915 |
| OID30099 | MEGF11 | Cardiometabolic_II | MEGF11 | A6BM72 | 15 | 65895079 | 66253756 | 0.98 | 0.83 | 1.15 | 7.65E-01 | 0.915 |
| OID20527 | DPP10 | Inflammation | DPP10 | Q8N608 | 2 | 114442299 | 115845780 | 1.03 | 0.87 | 1.21 | 7.67E-01 | 0.916 |
| OID20673 | PGF | Inflammation | PGF | P49763 | 14 | 74941834 | 74955626 | 0.97 | 0.81 | 1.17 | 7.67E-01 | 0.916 |
| OID30510 | SLITRK1 | Inflammation_II | SLITRK1 | Q96PX8 | 13 | 83877205 | 83882474 | 0.97 | 0.82 | 1.16 | 7.67E-01 | 0.916 |
| OID31183 | STOML2 | Oncology_II | STOML2 | Q9UJZ1 | 9 | 35099776 | 35103195 | 0.98 | 0.83 | 1.15 | 7.67E-01 | 0.916 |
| OID20755 | ADGRE2 | Inflammation | ADGRE2 | Q9UHX3 | 19 | 14732392 | 14778560 | 0.98 | 0.83 | 1.15 | 7.70E-01 | 0.918 |
| OID20660 | CTSO | Inflammation | CTSO | P43234 | 4 | 155921580 | 155953912 | 0.98 | 0.83 | 1.15 | 7.69E-01 | 0.918 |
| OID31122 | PNLIPRP1 | Neurology_II | PNLIPRP1 | P54315 | 10 | 116590385 | 116609175 | 0.98 | 0.84 | 1.14 | 7.70E-01 | 0.918 |
| OID20810 | ANXA5 | Neurology | ANXA5 | P08758 | 4 | 121667946 | 121696995 | 1.02 | 0.88 | 1.20 | 7.70E-01 | 0.918 |
| OID30495 | CPOX | Inflammation_II | CPOX | P36551 | 3 | 98579446 | 98593648 | 0.98 | 0.83 | 1.14 | 7.71E-01 | 0.918 |
| OID30904 | PGM2 | Neurology_II | PGM2 | Q96G03 | 4 | 37826660 | 37862937 | 1.02 | 0.88 | 1.19 | 7.72E-01 | 0.919 |
| OID21321 | TAFA5 | Oncology | TAFA5 | Q7Z5A7 | 22 | 48489553 | 48850912 | 1.02 | 0.88 | 1.19 | 7.73E-01 | 0.920 |
| OID20512 | ALDH3A1 | Inflammation | ALDH3A1 | P30838 | 17 | 19737984 | 19748943 | 0.98 | 0.82 | 1.15 | 7.75E-01 | 0.921 |
| OID21438 | GPC1 | Oncology | GPC1 | P35052 | 2 | 240435663 | 240468076 | 1.02 | 0.87 | 1.20 | 7.74E-01 | 0.921 |
| OID31033 | MCEE | Neurology_II | MCEE | Q96PE7 | 2 | 71109684 | 71130239 | 0.98 | 0.83 | 1.15 | 7.75E-01 | 0.921 |
| OID30324 | PTPRB | Cardiometabolic_II | PTPRB | P23467 | 12 | 70515870 | 70637440 | 1.03 | 0.86 | 1.22 | 7.75E-01 | 0.921 |
| OID21390 | SIGLEC9 | Oncology | SIGLEC9 | Q9Y336 | 19 | 51124906 | 51136651 | 0.98 | 0.83 | 1.15 | 7.74E-01 | 0.921 |
| OID31162 | ATF4 | Oncology_II | ATF4 | P18848 | 22 | 39519695 | 39522683 | 0.98 | 0.83 | 1.15 | 7.76E-01 | 0.922 |

| | | | | | | | | | | | | |
|---|---|---|---|---|---|---|---|---|---|---|---|---|
| OID21005 | CDH15 | Neurology | CDH15 | P55291 | 16 | 89171748 | 89195492 | 1.03 | 0.85 | 1.24 | 7.76E-01 | 0.922 |
| OID20925 | PAEP | Neurology | PAEP | P09466 | 9 | 135561756 | 135566955 | 1.04 | 0.81 | 1.32 | 7.78E-01 | 0.922 |
| OID31470 | TSPAN8 | Oncology_II | TSPAN8 | P19075 | 12 | 71125085 | 71441898 | 0.98 | 0.83 | 1.15 | 7.78E-01 | 0.922 |
| OID30115 | CEP112 | Cardiometabolic_II | CEP112 | Q8N8E3 | 17 | 65635537 | 66192133 | 1.02 | 0.87 | 1.20 | 7.80E-01 | 0.924 |
| OID30710 | CTBS | Inflammation_II | CTBS | Q01459 | 1 | 84549611 | 84574480 | 0.98 | 0.82 | 1.16 | 7.79E-01 | 0.924 |
| OID30935 | DUSP29 | Neurology_II | DUSP29 | Q68J44 | 10 | 75037472 | 75073642 | 1.02 | 0.88 | 1.19 | 7.80E-01 | 0.924 |
| OID30814 | IMPG1 | Neurology_II | IMPG1 | Q17R60 | 6 | 75921114 | 76072678 | 1.02 | 0.87 | 1.20 | 7.81E-01 | 0.924 |
| OID21407 | KLK14 | Oncology | KLK14 | Q9P0G3 | 19 | 51077495 | 51084245 | 1.02 | 0.87 | 1.21 | 7.81E-01 | 0.924 |
| OID30184 | PKD2 | Cardiometabolic_II | PKD2 | Q13563 | 4 | 88007635 | 88077777 | 0.98 | 0.82 | 1.16 | 7.81E-01 | 0.924 |
| OID31451 | ATRAID | Oncology_II | ATRAID | Q6UW56 | 2 | 27212041 | 27217178 | 0.98 | 0.83 | 1.15 | 7.82E-01 | 0.924 |
| OID30407 | NOTCH2 | Cardiometabolic_II | NOTCH2 | Q04721 | 1 | 119911553 | 120100779 | 0.98 | 0.83 | 1.16 | 7.82E-01 | 0.925 |
| OID30076 | NXPE4 | Cardiometabolic_II | NXPE4 | Q6UWF7 | 11 | 114570591 | 114595786 | 0.98 | 0.83 | 1.15 | 7.83E-01 | 0.925 |
| OID20161 | TYRO3 | Cardiometabolic | TYRO3 | Q06418 | 15 | 41557675 | 41583589 | 0.98 | 0.83 | 1.15 | 7.83E-01 | 0.925 |
| OID21233 | FMR1 | Oncology | FMR1 | Q06787 | X | 147911919 | 147951125 | 1.02 | 0.87 | 1.20 | 7.84E-01 | 0.926 |
| OID30838 | PCARE | Neurology_II | PCARE | A6NGG8 | 2 | 29060976 | 29074523 | 1.02 | 0.87 | 1.20 | 7.85E-01 | 0.926 |
| OID21510 | ERBB4 | Oncology | ERBB4 | Q15303 | 2 | 211375717 | 212538841 | 1.02 | 0.86 | 1.22 | 7.85E-01 | 0.926 |
| OID20164 | APLP1 | Cardiometabolic | APLP1 | P51693 | 19 | 35867899 | 35879792 | 1.02 | 0.87 | 1.20 | 7.86E-01 | 0.926 |
| OID20938 | RSPO1 | Neurology | RSPO1 | Q2MKA7 | 1 | 37611350 | 37634892 | 0.98 | 0.83 | 1.15 | 7.86E-01 | 0.927 |
| OID30863 | SH3BGRL2 | Neurology_II | SH3BGRL2 | Q9UJC5 | 6 | 79631329 | 79703655 | 0.98 | 0.83 | 1.15 | 7.87E-01 | 0.927 |
| OID30802 | GRIN2B | Neurology_II | GRIN2B | Q13224 | 12 | 13437942 | 13982002 | 1.02 | 0.88 | 1.18 | 7.87E-01 | 0.927 |
| OID30129 | IFNW1 | Cardiometabolic_II | IFNW1 | P05000 | 9 | 21140632 | 21141832 | 0.98 | 0.81 | 1.17 | 7.87E-01 | 0.927 |
| OID30771 | A1BG | Inflammation_II | A1BG | P04217 | 19 | 58345178 | 58353492 | 1.03 | 0.85 | 1.23 | 7.89E-01 | 0.927 |
| OID20337 | PRCP | Cardiometabolic | PRCP | P42785 | 11 | 82822936 | 82970584 | 0.98 | 0.82 | 1.17 | 7.89E-01 | 0.927 |
| OID30387 | SHISA5 | Cardiometabolic_II | SHISA5 | Q8N114 | 3 | 48467798 | 48504826 | 1.02 | 0.87 | 1.21 | 7.89E-01 | 0.927 |
| OID31387 | SMPDL3B | Oncology_II | SMPDL3B | Q92485 | 1 | 27935000 | 27959157 | 0.98 | 0.84 | 1.15 | 7.89E-01 | 0.927 |
| OID31398 | SUSD4 | Oncology_II | SUSD4 | Q5VX71 | 1 | 223220819 | 223364233 | 1.02 | 0.87 | 1.21 | 7.89E-01 | 0.927 |
| OID20489 | GBP2 | Inflammation | GBP2 | P32456 | 1 | 89106132 | 89150456 | 0.98 | 0.82 | 1.16 | 7.90E-01 | 0.928 |
| OID30482 | CA8 | Inflammation_II | CA8 | P35219 | 8 | 60185412 | 60281400 | 1.02 | 0.86 | 1.21 | 7.91E-01 | 0.929 |
| OID20878 | S100A16 | Neurology | S100A16 | Q96FQ6 | 1 | 153606886 | 153613145 | 1.02 | 0.87 | 1.20 | 7.91E-01 | 0.929 |
| OID20981 | TNFRSF10B | Neurology | TNFRSF10B | O14763 | 8 | 23020133 | 23069031 | 1.02 | 0.89 | 1.16 | 7.92E-01 | 0.929 |
| OID31144 | CRYGD | Neurology_II | CRYGD | P07320 | 2 | 208121607 | 208124524 | 0.98 | 0.83 | 1.15 | 7.92E-01 | 0.929 |
| OID21526 | KLK6 | Oncology | KLK6 | Q92876 | 19 | 50958631 | 50969673 | 0.98 | 0.84 | 1.14 | 7.92E-01 | 0.929 |
| OID20886 | ANXA10 | Neurology | ANXA10 | Q9UJ72 | 4 | 168092537 | 168187736 | 1.02 | 0.88 | 1.18 | 7.95E-01 | 0.929 |
| OID20626 | BSG | Inflammation | BSG | P35613 | 19 | 571277 | 583494 | 1.02 | 0.87 | 1.19 | 7.94E-01 | 0.929 |
| OID30553 | CSF2RB | Inflammation_II | CSF2RB | P32927 | 22 | 36913628 | 36940439 | 0.98 | 0.83 | 1.16 | 7.94E-01 | 0.929 |
| OID20431 | IL10 | Inflammation | IL10 | P22301 | 1 | 206767602 | 206774541 | 1.02 | 0.89 | 1.17 | 7.94E-01 | 0.929 |
| OID20301 | LILRB2 | Cardiometabolic | LILRB2 | Q8N423 | 19 | 54273812 | 54281184 | 1.02 | 0.87 | 1.21 | 7.95E-01 | 0.929 |
| OID21429 | LRIG1 | Oncology | LRIG1 | Q96JA1 | 3 | 66378797 | 66501263 | 1.02 | 0.87 | 1.20 | 7.93E-01 | 0.929 |
| OID20168 | MEP1B | Cardiometabolic | MEP1B | Q16820 | 18 | 32185069 | 32220404 | 1.02 | 0.87 | 1.21 | 7.95E-01 | 0.929 |
| OID20834 | NOS3 | Neurology | NOS3 | P29474 | 7 | 150991017 | 151014588 | 1.02 | 0.86 | 1.21 | 7.96E-01 | 0.930 |
| OID20095 | GPR37 | Cardiometabolic | GPR37 | O15354 | 7 | 124743885 | 124765792 | 0.98 | 0.84 | 1.15 | 7.97E-01 | 0.931 |
| OID21291 | KLK13 | Oncology | KLK13 | Q9UKR3 | 19 | 51055626 | 51065114 | 1.02 | 0.87 | 1.20 | 7.97E-01 | 0.931 |
| OID30273 | ENPP6 | Cardiometabolic_II | ENPP6 | Q6UWR7 | 4 | 184088706 | 184221230 | 0.98 | 0.82 | 1.16 | 7.99E-01 | 0.932 |
| OID21358 | PODXL | Oncology | PODXL | O00592 | 7 | 131500262 | 131558217 | 1.02 | 0.87 | 1.19 | 7.99E-01 | 0.932 |
| OID30552 | CLEC12A | Inflammation_II | CLEC12A | Q5QGZ9 | 12 | 9951316 | 9995694 | 1.02 | 0.87 | 1.20 | 8.00E-01 | 0.932 |
| OID30213 | DMP1 | Cardiometabolic_II | DMP1 | Q13316 | 4 | 87650280 | 87664361 | 0.98 | 0.83 | 1.16 | 8.00E-01 | 0.932 |
| OID30461 | IL20RB | Inflammation_II | IL20RB | Q6UXL0 | 3 | 136946230 | 137011085 | 1.02 | 0.87 | 1.21 | 8.00E-01 | 0.932 |
| OID20682 | PKLR | Inflammation | PKLR | P30613 | 1 | 155289293 | 155301438 | 1.02 | 0.87 | 1.20 | 8.01E-01 | 0.933 |
| OID30528 | ADAMTS1 | Inflammation_II | ADAMTS1 | Q9UHI8 | 21 | 26835755 | 26845409 | 0.98 | 0.83 | 1.15 | 8.02E-01 | 0.933 |
| OID21469 | CD300LF | Oncology | CD300LF | Q8TDQ1 | 17 | 74694311 | 74712978 | 1.02 | 0.86 | 1.21 | 8.02E-01 | 0.933 |
| OID30366 | AMDHD2 | Cardiometabolic_II | AMDHD2 | Q9Y303 | 16 | 2520357 | 2531422 | 1.02 | 0.86 | 1.21 | 8.03E-01 | 0.934 |
| OID20794 | PTEN | Neurology | PTEN | P60484 | 10 | 87862638 | 87971930 | 0.98 | 0.83 | 1.15 | 8.03E-01 | 0.934 |
| OID20300 | HYOU1 | Cardiometabolic | HYOU1 | Q9Y4L1 | 11 | 119044188 | 119057227 | 1.02 | 0.88 | 1.18 | 8.04E-01 | 0.934 |
| OID20435 | ACTN4 | Inflammation | ACTN4 | O43707 | 19 | 38647649 | 38731589 | 0.98 | 0.81 | 1.17 | 8.05E-01 | 0.934 |
| OID30246 | APOBR | Cardiometabolic_II | APOBR | Q0VD83 | 16 | 28494643 | 28498964 | 1.02 | 0.86 | 1.21 | 8.06E-01 | 0.934 |
| OID21431 | CAPG | Oncology | CAPG | P40121 | 2 | 85394753 | 85418432 | 1.02 | 0.86 | 1.21 | 8.04E-01 | 0.934 |
| OID30403 | CHGA | Cardiometabolic_II | CHGA | P10645 | 14 | 92923150 | 92935285 | 0.98 | 0.86 | 1.12 | 8.06E-01 | 0.934 |
| OID30697 | CR1 | Inflammation_II | CR1 | P17927 | 1 | 207496147 | 207641765 | 1.02 | 0.86 | 1.21 | 8.06E-01 | 0.934 |
| OID31223 | ERCC1 | Oncology_II | ERCC1 | P07992 | 19 | 45407334 | 45478828 | 1.02 | 0.86 | 1.22 | 8.06E-01 | 0.934 |
| OID30114 | EXTL1 | Cardiometabolic_II | EXTL1 | Q92935 | 1 | 26019884 | 26036464 | 0.98 | 0.84 | 1.15 | 8.05E-01 | 0.934 |
| OID31325 | ITGAX | Oncology_II | ITGAX | P20702 | 16 | 31355134 | 31382999 | 0.98 | 0.83 | 1.16 | 8.08E-01 | 0.936 |
| OID31504 | SERPINI2 | Oncology_II | SERPINI2 | O75830 | 3 | 167441914 | 167478972 | 0.98 | 0.82 | 1.16 | 8.09E-01 | 0.937 |
| OID30735 | ATRN | Inflammation_II | ATRN | O75882 | 20 | 3471018 | 3651118 | 1.02 | 0.86 | 1.21 | 8.10E-01 | 0.937 |
| OID21132 | GNLY | Neurology | GNLY | P22749 | 2 | 85685175 | 85698852 | 0.98 | 0.83 | 1.16 | 8.10E-01 | 0.938 |
| OID21459 | HAGH | Oncology | HAGH | Q16775 | 16 | 1795620 | 1827157 | 1.02 | 0.86 | 1.21 | 8.10E-01 | 0.938 |
| OID21445 | ARG1 | Oncology | ARG1 | P05089 | 6 | 131470832 | 131584332 | 0.98 | 0.83 | 1.16 | 8.13E-01 | 0.939 |
| OID20789 | NME3 | Inflammation | NME3 | Q13232 | 16 | 1770320 | 1771561 | 0.98 | 0.82 | 1.17 | 8.12E-01 | 0.939 |
| OID30730 | PZP | Inflammation_II | PZP | P20742 | 12 | 9148840 | 9208395 | 0.97 | 0.78 | 1.22 | 8.13E-01 | 0.939 |
| OID20309 | AZU1 | Cardiometabolic | AZU1 | P20160 | 19 | 825097 | 832018 | 0.98 | 0.82 | 1.17 | 8.16E-01 | 0.939 |
| OID30434 | ESR1 | Inflammation_II | ESR1 | P03372 | 6 | 151656691 | 152129619 | 1.02 | 0.86 | 1.21 | 8.16E-01 | 0.939 |
| OID30579 | HEG1 | Inflammation_II | HEG1 | Q9ULI3 | 3 | 124965710 | 125055997 | 0.98 | 0.83 | 1.16 | 8.14E-01 | 0.939 |
| OID20217 | HMOX1 | Cardiometabolic | HMOX1 | P09601 | 22 | 35380361 | 35394214 | 1.02 | 0.87 | 1.20 | 8.15E-01 | 0.939 |
| OID20889 | ISLR2 | Neurology | ISLR2 | Q6UXK2 | 15 | 74100311 | 74138540 | 1.02 | 0.87 | 1.19 | 8.15E-01 | 0.939 |
| OID20746 | MEGF10 | Inflammation | MEGF10 | Q96KG7 | 5 | 127290796 | 127465737 | 0.98 | 0.83 | 1.16 | 8.14E-01 | 0.939 |
| OID30299 | MMP15 | Cardiometabolic_II | MMP15 | P51511 | 16 | 58025754 | 58046901 | 0.98 | 0.83 | 1.16 | 8.16E-01 | 0.939 |
| OID21176 | PDP1 | Oncology | PDP1 | Q9P0J1 | 8 | 93857807 | 93926068 | 0.98 | 0.83 | 1.16 | 8.15E-01 | 0.939 |
| OID20212 | PRSS27 | Cardiometabolic | PRSS27 | Q9BQR3 | 16 | 2712419 | 2720551 | 1.02 | 0.87 | 1.20 | 8.14E-01 | 0.939 |
| OID21072 | APOH | Neurology | APOH | P02749 | 17 | 66212033 | 66256525 | 1.02 | 0.87 | 1.20 | 8.18E-01 | 0.940 |
| OID30543 | CD7 | Inflammation_II | CD7 | P09564 | 17 | 82314868 | 82317608 | 0.98 | 0.83 | 1.15 | 8.18E-01 | 0.940 |
| OID30411 | CELA2A | Cardiometabolic_II | CELA2A | P08217 | 1 | 15456728 | 15472091 | 1.02 | 0.88 | 1.17 | 8.17E-01 | 0.940 |
| OID21380 | MSLN | Oncology | MSLN | Q13421 | 16 | 760734 | 768865 | 1.02 | 0.86 | 1.20 | 8.18E-01 | 0.940 |
| OID30531 | SEMA6C | Inflammation_II | SEMA6C | Q9H3T2 | 1 | 151131685 | 151146631 | 1.02 | 0.87 | 1.19 | 8.17E-01 | 0.940 |
| OID20957 | TNR | Neurology | TNR | Q92752 | 1 | 175315194 | 175743616 | 1.02 | 0.87 | 1.20 | 8.18E-01 | 0.940 |
| OID30733 | APOA4 | Inflammation_II | APOA4 | P06727 | 11 | 116820700 | 116823304 | 0.98 | 0.82 | 1.17 | 8.20E-01 | 0.941 |
| OID30716 | CFHR5 | Inflammation_II | CFHR5 | Q9BXR6 | 1 | 196975010 | 197009678 | 0.98 | 0.83 | 1.16 | 8.20E-01 | 0.941 |
| OID20880 | PTK7 | Neurology | PTK7 | Q13308 | 6 | 43076307 | 43161719 | 0.98 | 0.83 | 1.16 | 8.20E-01 | 0.941 |
| OID30898 | FRMD7 | Neurology_II | FRMD7 | Q6ZUT3 | X | 132076990 | 132128020 | 1.02 | 0.86 | 1.22 | 8.23E-01 | 0.943 |
| OID20580 | LY75 | Inflammation | LY75 | O60449 | 2 | 159803355 | 159904756 | 0.98 | 0.83 | 1.16 | 8.22E-01 | 0.943 |
| OID30497 | ALPI | Inflammation_II | ALPI | P09923 | 2 | 232456125 | 232460753 | 0.98 | 0.84 | 1.15 | 8.23E-01 | 0.943 |
| OID20205 | NPDC1 | Cardiometabolic | NPDC1 | Q9NQX5 | 9 | 137039463 | 137046179 | 1.02 | 0.87 | 1.19 | 8.24E-01 | 0.943 |
| OID31227 | EVI2B | Oncology_II | EVI2B | P34910 | 17 | 31303770 | 31314105 | 1.02 | 0.88 | 1.18 | 8.24E-01 | 0.943 |
| OID20249 | ADAMTS13 | Cardiometabolic | ADAMTS13 | Q76LX8 | 9 | 133414358 | 133459402 | 1.02 | 0.87 | 1.20 | 8.25E-01 | 0.943 |
| OID20817 | RAB6B | Neurology | RAB6B | Q9NRW1 | 3 | 133824235 | 133895882 | 1.02 | 0.86 | 1.20 | 8.24E-01 | 0.943 |
| OID20814 | SLC27A4 | Neurology | SLC27A4 | Q6P1M0 | 9 | 128340527 | 128361470 | 0.98 | 0.83 | 1.16 | 8.25E-01 | 0.944 |
| OID31118 | DNPEP | Neurology_II | DNPEP | Q9ULA0 | 2 | 219372043 | 219400022 | 0.98 | 0.85 | 1.14 | 8.25E-01 | 0.944 |
| OID20414 | EFEMP1 | Cardiometabolic | EFEMP1 | Q12805 | 2 | 55865967 | 55924139 | 1.02 | 0.86 | 1.21 | 8.26E-01 | 0.944 |
| OID20403 | ANG | Cardiometabolic | ANG | P03950 | 14 | 20684177 | 20698971 | 0.98 | 0.83 | 1.16 | 8.27E-01 | 0.944 |
| OID31365 | EDDM3B | Oncology_II | EDDM3B | P56851 | 14 | 20768404 | 20770948 | 0.97 | 0.74 | 1.27 | 8.27E-01 | 0.944 |
| OID21146 | IGF2R | Neurology | IGF2R | P11717 | 6 | 159969082 | 160113507 | 1.02 | 0.86 | 1.21 | 8.27E-01 | 0.944 |
| OID21136 | IL7R | Neurology | IL7R | P16871 | 5 | 35852695 | 35879603 | 0.98 | 0.83 | 1.16 | 8.27E-01 | 0.944 |
| OID30051 | BHMT2 | Cardiometabolic_II | BHMT2 | Q9H2M3 | 5 | 79069767 | 79090069 | 0.98 | 0.83 | 1.16 | 8.28E-01 | 0.945 |
| OID20791 | FOSB | Neurology | FOSB | P53539 | 19 | 45467995 | 45475179 | 1.02 | 0.86 | 1.20 | 8.29E-01 | 0.945 |
| OID31036 | MRPL58 | Neurology_II | MRPL58 | Q14197 | 17 | 75012670 | 75021261 | 1.02 | 0.88 | 1.18 | 8.29E-01 | 0.945 |
| OID30300 | NGFR | Cardiometabolic_II | NGFR | P08138 | 17 | 49495293 | 49515008 | 0.98 | 0.83 | 1.16 | 8.30E-01 | 0.945 |
| OID20235 | PCSK9 | Cardiometabolic | PCSK9 | Q8NBP7 | 1 | 55039447 | 55064852 | 0.98 | 0.83 | 1.16 | 8.30E-01 | 0.945 |
| OID20239 | MFAP5 | Cardiometabolic | MFAP5 | Q13361 | 12 | 8637346 | 8662888 | 0.98 | 0.82 | 1.17 | 8.30E-01 | 0.945 |
| OID31154 | SMTN | Neurology_II | SMTN | P53814 | 22 | 31064105 | 31104757 | 0.98 | 0.83 | 1.16 | 8.31E-01 | 0.946 |

| | | | | | | | | | | | | |
|---|---|---|---|---|---|---|---|---|---|---|---|---|
| OID21053 | CPPED1 | Neurology | CPPED1 | Q9BRF8 | 16 | 12659799 | 12803887 | 1.02 | 0.86 | 1.21 | 8.32E-01 | 0.946 |
| OID30315 | ASPN | Cardiometabolic_II | ASPN | Q9BXN1 | 9 | 92456205 | 92482506 | 0.98 | 0.85 | 1.14 | 8.33E-01 | 0.946 |
| OID20312 | CHIT1 | Cardiometabolic | CHIT1 | Q13231 | 1 | 203212827 | 203273641 | 1.02 | 0.86 | 1.20 | 8.33E-01 | 0.946 |
| OID20076 | FABP6 | Cardiometabolic | FABP6 | P51161 | 5 | 160187367 | 160238735 | 1.02 | 0.87 | 1.19 | 8.33E-01 | 0.946 |
| OID20888 | KLB | Neurology | KLB | Q86Z14 | 4 | 39406930 | 39451533 | 0.98 | 0.83 | 1.16 | 8.33E-01 | 0.946 |
| OID30054 | TRAF3IP2 | Cardiometabolic_II | TRAF3IP2 | O43734 | 6 | 111555381 | 111606906 | 1.02 | 0.84 | 1.24 | 8.34E-01 | 0.946 |
| OID20628 | CD244 | Inflammation | CD244 | Q9BZW8 | 1 | 160830160 | 160862887 | 0.98 | 0.84 | 1.15 | 8.35E-01 | 0.947 |
| OID21219 | STX6 | Oncology | STX6 | O43752 | 1 | 180972712 | 181023121 | 1.02 | 0.86 | 1.20 | 8.35E-01 | 0.947 |
| OID20882 | WASF3 | Neurology | WASF3 | Q9UPY6 | 13 | 26557683 | 26688948 | 1.02 | 0.87 | 1.19 | 8.35E-01 | 0.947 |
| OID30767 | SERPIND1 | Inflammation_II | SERPIND1 | P05546 | 22 | 20774113 | 20787720 | 0.98 | 0.81 | 1.18 | 8.36E-01 | 0.947 |
| OID30069 | RAB39B | Cardiometabolic_II | RAB39B | Q96DA2 | X | 155258235 | 155264491 | 0.98 | 0.82 | 1.17 | 8.36E-01 | 0.948 |
| OID21125 | EFNA1 | Neurology | EFNA1 | P20827 | 1 | 155127876 | 155134899 | 1.02 | 0.87 | 1.19 | 8.37E-01 | 0.948 |
| OID21529 | FOLR1 | Oncology | FOLR1 | P15328 | 11 | 72189558 | 72196323 | 0.98 | 0.83 | 1.16 | 8.38E-01 | 0.949 |
| OID20210 | MARCO | Cardiometabolic | MARCO | Q9UEW3 | 2 | 118942194 | 118994660 | 1.02 | 0.86 | 1.20 | 8.40E-01 | 0.951 |
| OID21003 | LXN | Neurology | LXN | Q9BS40 | 3 | 158645822 | 158672648 | 1.02 | 0.86 | 1.20 | 8.40E-01 | 0.951 |
| OID20105 | ACE2 | Cardiometabolic | ACE2 | Q9BYF1 | X | 15494566 | 15607236 | 1.02 | 0.86 | 1.20 | 8.41E-01 | 0.951 |
| OID20498 | IL15RA | Inflammation | IL15RA | Q13261 | 10 | 5943639 | 5978187 | 1.02 | 0.87 | 1.19 | 8.41E-01 | 0.951 |
| OID30577 | SCGB3A1 | Inflammation_II | SCGB3A1 | Q96QR1 | 5 | 180590105 | 180591499 | 0.98 | 0.83 | 1.16 | 8.41E-01 | 0.951 |
| OID31198 | VWA5A | Oncology_II | VWA5A | O00534 | 11 | 124115404 | 124147721 | 0.98 | 0.82 | 1.18 | 8.42E-01 | 0.951 |
| OID20126 | CLC | Cardiometabolic | CLC | Q05315 | 19 | 39731255 | 39738029 | 0.98 | 0.82 | 1.17 | 8.42E-01 | 0.951 |
| OID20713 | BTN2A1 | Inflammation | BTN2A1 | Q7KYR7 | 6 | 26457904 | 26476621 | 1.02 | 0.87 | 1.19 | 8.43E-01 | 0.951 |
| OID20783 | TNFRSF14 | Inflammation | TNFRSF14 | Q92956 | 1 | 2555639 | 2565382 | 0.98 | 0.83 | 1.16 | 8.43E-01 | 0.951 |
| OID31235 | DCDC2C | Oncology_II | DCDC2C | A8MYV0 | 2 | 3703575 | 3848008 | 0.98 | 0.83 | 1.16 | 8.44E-01 | 0.951 |
| OID21127 | B4GAT1 | Neurology | B4GAT1 | O43505 | 11 | 66345374 | 66347629 | 0.98 | 0.83 | 1.16 | 8.46E-01 | 0.952 |
| OID30137 | DHODH | Cardiometabolic_II | DHODH | Q02127 | 16 | 72008588 | 72027664 | 0.98 | 0.83 | 1.17 | 8.45E-01 | 0.952 |
| OID20766 | PRELP | Inflammation | PRELP | P51888 | 1 | 203475806 | 203491352 | 0.99 | 0.87 | 1.13 | 8.45E-01 | 0.952 |
| OID20985 | TNFRSF9 | Neurology | TNFRSF9 | Q07011 | 1 | 7915871 | 7943165 | 1.02 | 0.87 | 1.19 | 8.46E-01 | 0.952 |
| OID30618 | C1QTNF5 | Inflammation_II | C1QTNF5 | Q9BXJ0 | 11 | 119338942 | 119340940 | 0.98 | 0.83 | 1.16 | 8.47E-01 | 0.953 |
| OID21248 | LHB | Oncology | LHB | P01229 | 19 | 49015980 | 49017091 | 0.98 | 0.83 | 1.17 | 8.47E-01 | 0.953 |
| OID21272 | FLT3 | Oncology | FLT3 | P36888 | 13 | 28003274 | 28100592 | 1.02 | 0.87 | 1.18 | 8.48E-01 | 0.953 |
| OID20591 | NTF3 | Inflammation | NTF3 | P20783 | 12 | 5432108 | 5521536 | 0.98 | 0.83 | 1.16 | 8.47E-01 | 0.953 |
| OID20823 | IDI2 | Neurology | IDI2 | Q9BXS1 | 10 | 1018910 | 1025859 | 0.98 | 0.83 | 1.16 | 8.49E-01 | 0.954 |
| OID20574 | OSM | Inflammation | OSM | P13725 | 22 | 30262829 | 30266851 | 0.98 | 0.83 | 1.16 | 8.49E-01 | 0.954 |
| OID30743 | SERPINA6 | Inflammation_II | SERPINA6 | P08185 | 14 | 94304248 | 94323389 | 0.98 | 0.82 | 1.18 | 8.49E-01 | 0.954 |
| OID20231 | THBD | Cardiometabolic | THBD | P07204 | 20 | 23045633 | 23049672 | 1.02 | 0.87 | 1.19 | 8.49E-01 | 0.954 |
| OID21029 | BST2 | Neurology | BST2 | Q10589 | 19 | 17402939 | 17405630 | 0.98 | 0.84 | 1.16 | 8.50E-01 | 0.954 |
| OID31200 | PSMC3 | Oncology_II | PSMC3 | P17980 | 11 | 47418769 | 47426473 | 0.98 | 0.84 | 1.16 | 8.50E-01 | 0.954 |
| OID20229 | NRCAM | Cardiometabolic | NRCAM | Q92823 | 7 | 108147623 | 108456717 | 0.99 | 0.85 | 1.15 | 8.51E-01 | 0.955 |
| OID20365 | CTSZ | Cardiometabolic | CTSZ | Q9UBR2 | 20 | 58985686 | 59008238 | 0.98 | 0.84 | 1.16 | 8.53E-01 | 0.956 |
| OID31044 | EFHD1 | Neurology_II | EFHD1 | Q9BUP0 | 2 | 232606057 | 232682776 | 0.98 | 0.83 | 1.17 | 8.53E-01 | 0.956 |
| OID31129 | INSL3 | Neurology_II | INSL3 | P51460 | 19 | 17816512 | 17821574 | 1.03 | 0.77 | 1.38 | 8.54E-01 | 0.956 |
| OID30561 | PTGES2 | Inflammation_II | PTGES2 | Q9H7Z7 | 9 | 128120693 | 128128462 | 0.98 | 0.83 | 1.17 | 8.54E-01 | 0.956 |
| OID20571 | PTH1R | Inflammation | PTH1R | Q03431 | 3 | 46877721 | 46903799 | 1.02 | 0.85 | 1.22 | 8.54E-01 | 0.956 |
| OID20841 | GHRHR | Neurology | GHRHR | Q02643 | 7 | 30938669 | 30993254 | 1.01 | 0.88 | 1.17 | 8.55E-01 | 0.957 |
| OID30973 | CCDC28A | Neurology_II | CCDC28A | Q8IWP9 | 6 | 138773769 | 138793319 | 0.98 | 0.83 | 1.16 | 8.56E-01 | 0.957 |
| OID21117 | CNTN4 | Neurology | CNTN4 | Q8IWV2 | 3 | 2098813 | 3057959 | 1.02 | 0.86 | 1.19 | 8.56E-01 | 0.957 |
| OID31187 | GJA8 | Oncology_II | GJA8 | P48165 | 1 | 147902795 | 147909269 | 0.99 | 0.84 | 1.16 | 8.57E-01 | 0.958 |
| OID20336 | BLMH | Cardiometabolic | BLMH | Q13867 | 17 | 30248203 | 30292056 | 1.01 | 0.86 | 1.19 | 8.58E-01 | 0.958 |
| OID20573 | CLEC4A | Inflammation | CLEC4A | Q9UMR7 | 12 | 8123617 | 8138607 | 0.98 | 0.83 | 1.17 | 8.58E-01 | 0.958 |
| OID30491 | ADAM12 | Inflammation_II | ADAM12 | O43184 | 10 | 126012381 | 126388477 | 1.02 | 0.85 | 1.22 | 8.63E-01 | 0.958 |
| OID30746 | APOD | Inflammation_II | APOD | P05090 | 3 | 195568705 | 195584033 | 1.02 | 0.85 | 1.22 | 8.60E-01 | 0.958 |
| OID20540 | BTN3A2 | Inflammation | BTN3A2 | P78410 | 6 | 26365159 | 26378320 | 0.99 | 0.84 | 1.15 | 8.62E-01 | 0.958 |
| OID21025 | CD34 | Neurology | CD34 | P28906 | 1 | 207880972 | 207911402 | 1.01 | 0.86 | 1.20 | 8.61E-01 | 0.958 |
| OID20664 | CLSTN2 | Inflammation | CLSTN2 | Q9H4D0 | 3 | 139935185 | 140577397 | 1.01 | 0.86 | 1.19 | 8.64E-01 | 0.958 |
| OID21024 | CXCL13 | Neurology | CXCL13 | O43927 | 4 | 77511753 | 77611834 | 1.01 | 0.86 | 1.20 | 8.62E-01 | 0.958 |
| OID21474 | DCBLD2 | Oncology | DCBLD2 | Q96PD2 | 3 | 98795941 | 98901695 | 1.01 | 0.86 | 1.20 | 8.61E-01 | 0.958 |
| OID20743 | EPCAM | Inflammation | EPCAM | P16422 | 2 | 47345158 | 47387601 | 1.01 | 0.86 | 1.20 | 8.61E-01 | 0.958 |
| OID20845 | GBP4 | Neurology | GBP4 | Q96PP9 | 1 | 89181144 | 89198942 | 0.99 | 0.84 | 1.16 | 8.60E-01 | 0.958 |
| OID21101 | GUCA2A | Neurology | GUCA2A | Q02747 | 1 | 42162690 | 42164745 | 1.01 | 0.87 | 1.18 | 8.63E-01 | 0.958 |
| OID21158 | IGFBP4 | Neurology | IGFBP4 | P22692 | 17 | 40443450 | 40457725 | 1.01 | 0.86 | 1.19 | 8.63E-01 | 0.958 |
| OID31243 | IL25 | Oncology_II | IL25 | Q9H293 | 14 | 23372809 | 23376403 | 1.01 | 0.87 | 1.18 | 8.64E-01 | 0.958 |
| OID20385 | IL6R | Cardiometabolic | IL6R | P08887 | 1 | 154405193 | 154469450 | 0.99 | 0.84 | 1.16 | 8.59E-01 | 0.958 |
| OID21384 | MOG | Oncology | MOG | Q16653 | 6 | 29657002 | 29672372 | 0.99 | 0.84 | 1.16 | 8.64E-01 | 0.958 |
| OID30509 | NAGA | Inflammation_II | NAGA | P17050 | 22 | 42058334 | 42070842 | 0.98 | 0.83 | 1.17 | 8.60E-01 | 0.958 |
| OID21283 | POLR2F | Oncology | POLR2F | P61218 | 22 | 37952607 | 38041915 | 0.98 | 0.82 | 1.18 | 8.62E-01 | 0.958 |
| OID30599 | SEMA3G | Inflammation_II | SEMA3G | Q9NS98 | 3 | 52433035 | 52445103 | 0.99 | 0.84 | 1.16 | 8.64E-01 | 0.958 |
| OID30937 | TARM1 | Neurology_II | TARM1 | B6A8C7 | 19 | 54069895 | 54081365 | 0.99 | 0.84 | 1.16 | 8.59E-01 | 0.958 |
| OID20129 | TSHB | Cardiometabolic | TSHB | P01222 | 1 | 115029826 | 115034302 | 1.01 | 0.87 | 1.18 | 8.63E-01 | 0.958 |
| OID20541 | MLN | Inflammation | MLN | P12872 | 6 | 33794673 | 33804003 | 0.98 | 0.83 | 1.17 | 8.65E-01 | 0.958 |
| OID20175 | GHRL | Cardiometabolic | GHRL | Q9UBU3 | 3 | 10285666 | 10292947 | 0.99 | 0.83 | 1.16 | 8.66E-01 | 0.959 |
| OID31266 | TET2 | Oncology_II | TET2 | Q6N021 | 4 | 105145875 | 105279816 | 1.01 | 0.87 | 1.18 | 8.66E-01 | 0.959 |
| OID20884 | FMNL1 | Neurology | FMNL1 | O95466 | 17 | 45221444 | 45247319 | 0.99 | 0.83 | 1.18 | 8.67E-01 | 0.960 |
| OID31066 | NEO1 | Neurology_II | NEO1 | Q92859 | 15 | 73051710 | 73305205 | 1.01 | 0.88 | 1.16 | 8.69E-01 | 0.961 |
| OID21302 | RP2 | Oncology | RP2 | O75695 | X | 46837043 | 46882358 | 1.01 | 0.86 | 1.20 | 8.71E-01 | 0.963 |
| OID30627 | SPINK2 | Inflammation_II | SPINK2 | P20155 | 4 | 56809860 | 56821742 | 1.01 | 0.85 | 1.21 | 8.72E-01 | 0.964 |
| OID21156 | CLPS | Neurology | CLPS | P04118 | 6 | 35794982 | 35797344 | 0.99 | 0.85 | 1.15 | 8.72E-01 | 0.964 |
| OID30879 | TP73 | Neurology_II | TP73 | O15350 | 1 | 3652516 | 3736201 | 1.01 | 0.86 | 1.19 | 8.73E-01 | 0.964 |
| OID20320 | QPCT | Cardiometabolic | QPCT | Q16769 | 2 | 37342827 | 37373322 | 1.01 | 0.86 | 1.20 | 8.74E-01 | 0.965 |
| OID21515 | CREG1 | Oncology | CREG1 | O75629 | 1 | 167529117 | 167553805 | 0.99 | 0.84 | 1.16 | 8.75E-01 | 0.965 |
| OID20423 | EIF5A | Inflammation | EIF5A | P63241 | 17 | 7306999 | 7312463 | 1.01 | 0.86 | 1.20 | 8.75E-01 | 0.965 |
| OID21385 | HMBS | Oncology | HMBS | P08397 | 11 | 119084866 | 119093834 | 0.99 | 0.84 | 1.16 | 8.75E-01 | 0.965 |
| OID20404 | FETUB | Cardiometabolic | FETUB | Q9UGM5 | 3 | 186635969 | 186653141 | 0.99 | 0.84 | 1.16 | 8.76E-01 | 0.966 |
| OID30646 | FGFR4 | Inflammation_II | FGFR4 | P22455 | 5 | 177086905 | 177098144 | 0.99 | 0.84 | 1.16 | 8.78E-01 | 0.967 |
| OID31205 | MAPK13 | Oncology_II | MAPK13 | O15264 | 6 | 36127809 | 36144524 | 0.99 | 0.83 | 1.18 | 8.78E-01 | 0.967 |
| OID30346 | LILRA6 | Cardiometabolic_II | LILRA6 | Q6PI73 | 19 | 54236592 | 54242791 | 0.99 | 0.84 | 1.16 | 8.78E-01 | 0.967 |
| OID21275 | ADAMTS15 | Oncology | ADAMTS15 | Q8TE58 | 11 | 130448645 | 130476645 | 0.99 | 0.83 | 1.17 | 8.80E-01 | 0.967 |
| OID20410 | C2 | Cardiometabolic | C2 | P06681 | 6 | 31897785 | 31945672 | 0.99 | 0.85 | 1.15 | 8.81E-01 | 0.967 |
| OID21328 | CD302 | Oncology | CD302 | Q8IX05 | 2 | 159768628 | 159798255 | 1.01 | 0.86 | 1.19 | 8.82E-01 | 0.967 |
| OID30930 | CHRM1 | Neurology_II | CHRM1 | P11229 | 11 | 62908679 | 62921807 | 0.99 | 0.84 | 1.16 | 8.83E-01 | 0.967 |
| OID21392 | CPE | Oncology | CPE | P16870 | 4 | 165361194 | 165498547 | 0.99 | 0.83 | 1.17 | 8.82E-01 | 0.967 |
| OID30800 | CTNNA1 | Neurology_II | CTNNA1 | P35221 | 5 | 138610967 | 138935034 | 0.99 | 0.84 | 1.16 | 8.81E-01 | 0.967 |
| OID30126 | DDA1 | Cardiometabolic_II | DDA1 | Q9BW61 | 19 | 17309518 | 17323298 | 0.99 | 0.85 | 1.15 | 8.83E-01 | 0.967 |
| OID20090 | GP2 | Cardiometabolic | GP2 | P55259 | 16 | 20309574 | 20327808 | 0.99 | 0.84 | 1.17 | 8.83E-01 | 0.967 |
| OID30808 | KLF4 | Neurology_II | KLF4 | O43474 | 9 | 107484852 | 107490482 | 0.99 | 0.83 | 1.18 | 8.80E-01 | 0.967 |
| OID20629 | KLRB1 | Inflammation | KLRB1 | Q12918 | 12 | 9594551 | 9607916 | 0.99 | 0.84 | 1.17 | 8.81E-01 | 0.967 |
| OID31170 | MUCL3 | Oncology_II | MUCL3 | Q3MIW9 | 6 | 30934523 | 30954221 | 0.99 | 0.83 | 1.17 | 8.82E-01 | 0.967 |
| OID30199 | MYBPC2 | Cardiometabolic_II | MYBPC2 | Q14324 | 19 | 50432892 | 50466321 | 0.99 | 0.82 | 1.19 | 8.80E-01 | 0.967 |
| OID30777 | SERPING1 | Inflammation_II | SERPING1 | P05155 | 11 | 57597387 | 57619171 | 0.99 | 0.83 | 1.17 | 8.82E-01 | 0.967 |
| OID21052 | TNFRSF21 | Neurology | TNFRSF21 | O75509 | 6 | 47231532 | 47309905 | 1.01 | 0.87 | 1.18 | 8.80E-01 | 0.967 |
| OID21381 | VMO1 | Oncology | VMO1 | Q7Z5L0 | 17 | 4785285 | 4786433 | 1.01 | 0.86 | 1.19 | 8.79E-01 | 0.967 |
| OID31217 | MORN4 | Oncology_II | MORN4 | Q8NDC4 | 10 | 97614553 | 97633500 | 1.01 | 0.87 | 1.18 | 8.84E-01 | 0.968 |
| OID30070 | RPS10 | Cardiometabolic_II | RPS10 | P46783 | 6 | 34417454 | 34426069 | 0.99 | 0.83 | 1.18 | 8.84E-01 | 0.968 |
| OID20101 | IL6 | Cardiometabolic/Inflamm | IL6 | P05231 | 7 | 22725884 | 22732002 | 1.01 | 0.86 | 1.18 | 8.89E-01 | 0.969 |
| OID21250 | BAIAP2 | Oncology | BAIAP2 | Q9UQB8 | 17 | 81035122 | 81117434 | 0.99 | 0.83 | 1.17 | 8.86E-01 | 0.969 |
| OID30179 | BRDT | Cardiometabolic_II | BRDT | Q58F21 | 1 | 91949343 | 92014426 | 0.99 | 0.84 | 1.16 | 8.86E-01 | 0.969 |
| OID31184 | BSND | Oncology_II | BSND | Q8WZ55 | 1 | 54998933 | 55017172 | 0.99 | 0.83 | 1.18 | 8.88E-01 | 0.969 |
| OID20565 | CD83 | Inflammation | CD83 | Q01151 | 6 | 14117256 | 14136918 | 1.01 | 0.86 | 1.19 | 8.88E-01 | 0.969 |

| | | | | | | | | | | | | |
|---|---|---|---|---|---|---|---|---|---|---|---|---|
| OID30882 | DTX2 | Neurology_II | DTX2 | Q86UW9 | 7 | 76461676 | 76505995 | 0.99 | 0.83 | 1.17 | 8.90E-01 | 0.969 |
| OID20520 | HLA-DRA | Inflammation | HLA-DRA | P01903 | 6 | 32439878 | 32445046 | 1.01 | 0.87 | 1.18 | 8.87E-01 | 0.969 |
| OID20694 | IL18 | Inflammation | IL18 | Q14116 | 11 | 112143253 | 112164096 | 1.01 | 0.85 | 1.20 | 8.88E-01 | 0.969 |
| OID31245 | KHDC3L | Oncology_II | KHDC3L | Q587J8 | 6 | 73362658 | 73364171 | 1.01 | 0.85 | 1.20 | 8.90E-01 | 0.969 |
| OID20434 | LTO1 | Inflammation | LTO1 | Q8WV07 | 11 | 69653076 | 69675416 | 1.01 | 0.85 | 1.20 | 8.91E-01 | 0.969 |
| OID31201 | PDE1C | Oncology_II | PDE1C | Q14123 | 7 | 31751179 | 32428131 | 1.01 | 0.85 | 1.20 | 8.91E-01 | 0.969 |
| OID30903 | SCN3A | Neurology_II | SCN3A | Q9NY46 | 2 | 165087526 | 165204050 | 0.99 | 0.83 | 1.18 | 8.91E-01 | 0.969 |
| OID20169 | SDC1 | Cardiometabolic | SDC1 | P18827 | 2 | 20200797 | 20225433 | 0.99 | 0.84 | 1.17 | 8.89E-01 | 0.969 |
| OID31174 | SLIRP | Oncology_II | SLIRP | Q9GZT3 | 14 | 77708071 | 77761104 | 1.01 | 0.85 | 1.21 | 8.88E-01 | 0.969 |
| OID31296 | SPRR1B | Oncology_II | SPRR1B | P22528 | 1 | 153031203 | 153032900 | 0.99 | 0.83 | 1.18 | 8.89E-01 | 0.969 |
| OID30508 | ZNF174 | Inflammation_II | ZNF174 | Q15697 | 16 | 3401215 | 3409364 | 0.99 | 0.83 | 1.18 | 8.90E-01 | 0.969 |
| OID30094 | ERVV-1 | Cardiometabolic_II | ERVV-1 | B6SEH8 | 19 | 53013921 | 53016123 | 0.99 | 0.83 | 1.18 | 8.92E-01 | 0.969 |
| OID21081 | NRP2 | Neurology | NRP2 | O60462 | 2 | 205681990 | 205798133 | 0.99 | 0.84 | 1.17 | 8.92E-01 | 0.969 |
| OID20569 | CCL28 | Inflammation | CCL28 | Q9NRJ3 | 5 | 43376645 | 43412391 | 0.99 | 0.84 | 1.16 | 8.93E-01 | 0.970 |
| OID30448 | CD3G | Inflammation_II | CD3G | P09693 | 11 | 118344344 | 118355161 | 0.99 | 0.85 | 1.16 | 8.92E-01 | 0.970 |
| OID20362 | NID1 | Cardiometabolic | NID1 | P14543 | 1 | 235975830 | 236065109 | 0.99 | 0.84 | 1.16 | 8.93E-01 | 0.970 |
| OID20765 | CCL22 | Inflammation | CCL22 | O00626 | 16 | 57358783 | 57366189 | 0.99 | 0.84 | 1.17 | 8.93E-01 | 0.970 |
| OID31427 | KLK15 | Oncology_II | KLK15 | Q9H2R5 | 19 | 50825289 | 50837213 | 1.01 | 0.86 | 1.19 | 8.94E-01 | 0.970 |
| OID21478 | FGFR2 | Oncology | FGFR2 | P21802 | 10 | 121478332 | 121598458 | 1.01 | 0.86 | 1.20 | 8.95E-01 | 0.971 |
| OID21523 | KLK10 | Oncology | KLK10 | O43240 | 19 | 51012739 | 51020175 | 0.99 | 0.84 | 1.16 | 8.95E-01 | 0.971 |
| OID30262 | MAN1A2 | Cardiometabolic_II | MAN1A2 | O60476 | 1 | 117367449 | 117528872 | 0.99 | 0.84 | 1.16 | 8.98E-01 | 0.972 |
| OID30550 | PDZK1 | Inflammation_II | PDZK1 | Q5T2W1 | 1 | 145670852 | 145708148 | 1.01 | 0.86 | 1.20 | 8.98E-01 | 0.972 |
| OID20283 | SEMA7A | Cardiometabolic | SEMA7A | O75326 | 15 | 74409289 | 74433958 | 0.99 | 0.84 | 1.17 | 8.97E-01 | 0.972 |
| OID30470 | TP53BP1 | Inflammation_II | TP53BP1 | Q12888 | 15 | 43403061 | 43510728 | 1.01 | 0.86 | 1.19 | 8.97E-01 | 0.972 |
| OID31260 | ARAF | Oncology_II | ARAF | P10398 | X | 47561205 | 47571908 | 0.99 | 0.84 | 1.16 | 8.98E-01 | 0.972 |
| OID30118 | ATP1B2 | Cardiometabolic_II | ATP1B2 | P14415 | 17 | 7646627 | 7657770 | 0.99 | 0.85 | 1.16 | 9.03E-01 | 0.972 |
| OID30873 | BHLHE40 | Neurology_II | BHLHE40 | O14503 | 3 | 4979437 | 4985323 | 0.99 | 0.84 | 1.17 | 9.05E-01 | 0.972 |
| OID21130 | CD300LG | Neurology | CD300LG | Q6UXG3 | 17 | 43847148 | 43863639 | 0.99 | 0.84 | 1.16 | 9.05E-01 | 0.972 |
| OID20738 | COLEC12 | Inflammation | COLEC12 | Q5KU26 | 18 | 316737 | 500722 | 0.99 | 0.83 | 1.17 | 9.01E-01 | 0.972 |
| OID20070 | CRX | Cardiometabolic | CRX | O43186 | 19 | 47819779 | 47843330 | 0.99 | 0.85 | 1.16 | 9.04E-01 | 0.972 |
| OID31193 | FGF9 | Oncology_II | FGF9 | P31371 | 13 | 21671073 | 21704498 | 1.01 | 0.87 | 1.17 | 9.02E-01 | 0.972 |
| OID31166 | GAD1 | Oncology_II | GAD1 | Q99259 | 2 | 170813213 | 170861151 | 0.99 | 0.84 | 1.16 | 9.00E-01 | 0.972 |
| OID31175 | HMMR | Oncology_II | HMMR | O75330 | 5 | 163460203 | 163491941 | 1.01 | 0.85 | 1.20 | 9.02E-01 | 0.972 |
| OID20383 | HSPG2 | Cardiometabolic | HSPG2 | P98160 | 1 | 21822244 | 21937310 | 1.01 | 0.86 | 1.18 | 9.01E-01 | 0.972 |
| OID21213 | IKZF2 | Oncology | IKZF2 | Q9UKS7 | 2 | 212999691 | 213152427 | 1.01 | 0.87 | 1.18 | 9.04E-01 | 0.972 |
| OID30566 | INSR | Inflammation_II | INSR | P06213 | 19 | 7112255 | 7294414 | 0.99 | 0.85 | 1.16 | 9.04E-01 | 0.972 |
| OID30296 | MYBPC1 | Cardiometabolic_II | MYBPC1 | Q00872 | 12 | 101568353 | 101686028 | 0.99 | 0.83 | 1.18 | 9.00E-01 | 0.972 |
| OID30834 | MYOM1 | Neurology_II | MYOM1 | P52179 | 18 | 3066807 | 3219968 | 1.01 | 0.86 | 1.18 | 9.04E-01 | 0.972 |
| OID20448 | NCLN | Inflammation | NCLN | Q969V3 | 19 | 3185563 | 3209575 | 1.01 | 0.86 | 1.19 | 9.02E-01 | 0.972 |
| OID30328 | PKD1 | Cardiometabolic_II | PKD1 | P98161 | 16 | 2088708 | 2135898 | 1.01 | 0.85 | 1.20 | 9.05E-01 | 0.972 |
| OID20560 | SELPLG | Inflammation | SELPLG | Q14242 | 12 | 108621895 | 108633894 | 1.01 | 0.85 | 1.21 | 9.00E-01 | 0.972 |
| OID21157 | STC2 | Neurology | STC2 | O76061 | 5 | 173314723 | 173328447 | 1.01 | 0.85 | 1.20 | 9.01E-01 | 0.972 |
| OID30687 | TCN1 | Inflammation_II | TCN1 | P20061 | 11 | 59852800 | 59866489 | 1.01 | 0.85 | 1.20 | 9.02E-01 | 0.972 |
| OID30318 | TIMP2 | Cardiometabolic_II | TIMP2 | P16035 | 17 | 78852977 | 78925387 | 1.01 | 0.88 | 1.15 | 9.04E-01 | 0.972 |
| OID31103 | TMPRSS11D | Neurology_II | TMPRSS11D | O60235 | 4 | 67820876 | 67884002 | 1.01 | 0.87 | 1.18 | 9.02E-01 | 0.972 |
| OID21019 | MMP8 | Neurology | MMP8 | P22894 | 11 | 102711796 | 102727050 | 0.99 | 0.84 | 1.16 | 9.06E-01 | 0.973 |
| OID30196 | SEL1L | Cardiometabolic_II | SEL1L | Q9UBV2 | 14 | 81471547 | 81533853 | 0.99 | 0.84 | 1.17 | 9.06E-01 | 0.973 |
| OID20797 | COPE | Neurology | COPE | O14579 | 19 | 18899514 | 18919387 | 1.01 | 0.86 | 1.19 | 9.10E-01 | 0.974 |
| OID21299 | DDAH1 | Oncology | DDAH1 | O94760 | 1 | 85318481 | 85578363 | 1.01 | 0.85 | 1.20 | 9.08E-01 | 0.974 |
| OID20288 | ENPP2 | Cardiometabolic | ENPP2 | Q13822 | 8 | 119557086 | 119673453 | 1.01 | 0.83 | 1.22 | 9.09E-01 | 0.974 |
| OID30413 | GM2A | Cardiometabolic_II | GM2A | P17900 | 5 | 151212150 | 151270440 | 0.99 | 0.85 | 1.15 | 9.10E-01 | 0.974 |
| OID21134 | MFGE8 | Neurology | MFGE8 | Q08431 | 15 | 88898683 | 88913381 | 0.99 | 0.84 | 1.17 | 9.10E-01 | 0.974 |
| OID21531 | MIA | Oncology | MIA | Q16674 | 19 | 40771648 | 40777490 | 1.01 | 0.86 | 1.18 | 9.10E-01 | 0.974 |
| OID30900 | NLGN1 | Neurology_II | NLGN1 | Q8N2Q7 | 3 | 173396284 | 174294372 | 0.99 | 0.83 | 1.18 | 9.10E-01 | 0.974 |
| OID30789 | SERPINC1 | Inflammation_II | SERPINC1 | P01008 | 1 | 173903800 | 173917327 | 0.99 | 0.84 | 1.17 | 9.10E-01 | 0.974 |
| OID21278 | SF3B4 | Oncology | SF3B4 | Q15427 | 1 | 149923317 | 149927803 | 0.99 | 0.84 | 1.17 | 9.12E-01 | 0.976 |
| OID21228 | CLEC6A | Oncology | CLEC6A | Q6EIG7 | 12 | 8455962 | 8478330 | 1.01 | 0.86 | 1.19 | 9.12E-01 | 0.976 |
| OID30172 | PMCH | Cardiometabolic_II | PMCH | P20382 | 12 | 102196459 | 102197833 | 0.99 | 0.84 | 1.17 | 9.13E-01 | 0.977 |
| OID30793 | AGBL2 | Neurology_II | AGBL2 | Q5U5Z8 | 11 | 47659591 | 47715389 | 0.99 | 0.85 | 1.15 | 9.15E-01 | 0.977 |
| OID20323 | LILRB1 | Cardiometabolic | LILRB1 | Q8NHL6 | 19 | 54617158 | 54638022 | 0.99 | 0.84 | 1.17 | 9.15E-01 | 0.977 |
| OID21208 | NTF4 | Oncology | NTF4 | P34130 | 19 | 49061066 | 49065054 | 1.01 | 0.85 | 1.20 | 9.15E-01 | 0.977 |
| OID21258 | PLXDC1 | Oncology | PLXDC1 | Q8IUK5 | 17 | 39063313 | 39154394 | 0.99 | 0.85 | 1.16 | 9.15E-01 | 0.977 |
| OID21427 | CRNN | Oncology | CRNN | Q9UBG3 | 1 | 152409243 | 152414263 | 1.01 | 0.86 | 1.19 | 9.17E-01 | 0.978 |
| OID30729 | QSOX1 | Inflammation_II | QSOX1 | O00391 | 1 | 180154869 | 180204030 | 0.99 | 0.82 | 1.20 | 9.17E-01 | 0.978 |
| OID20728 | SCG3 | Inflammation | SCG3 | Q8WXD2 | 15 | 51681492 | 51721026 | 0.99 | 0.84 | 1.16 | 9.16E-01 | 0.978 |
| OID21487 | PODXL2 | Oncology | PODXL2 | Q9NZ53 | 3 | 127629185 | 127672802 | 1.01 | 0.86 | 1.18 | 9.18E-01 | 0.978 |
| OID20873 | KIRREL2 | Neurology | KIRREL2 | Q6UWL6 | 19 | 35855861 | 35867136 | 1.01 | 0.86 | 1.18 | 9.19E-01 | 0.979 |
| OID31199 | GAGE2A | Oncology_II | GAGE2A | Q6NT46 | X | 49589496 | 49596824 | 0.99 | 0.84 | 1.17 | 9.19E-01 | 0.979 |
| OID31255 | COL9A2 | Oncology_II | COL9A2 | Q14055 | 1 | 40300489 | 40317813 | 0.99 | 0.84 | 1.17 | 9.20E-01 | 0.979 |
| OID30910 | TDO2 | Neurology_II | TDO2 | P48775 | 4 | 155854738 | 155920406 | 1.01 | 0.85 | 1.19 | 9.21E-01 | 0.980 |
| OID31221 | FOS | Oncology_II | FOS | P01100 | 14 | 75278826 | 75282230 | 1.01 | 0.86 | 1.18 | 9.21E-01 | 0.981 |
| OID21122 | CD99L2 | Neurology | CD99L2 | Q8TCZ2 | X | 150766336 | 150898816 | 0.99 | 0.84 | 1.18 | 9.22E-01 | 0.981 |
| OID31257 | MORF4L1 | Oncology_II | MORF4L1 | Q9UBU8 | 15 | 78810487 | 78898139 | 1.01 | 0.85 | 1.20 | 9.23E-01 | 0.981 |
| OID20983 | CALCA | Neurology | CALCA | P01258 | 11 | 14966668 | 14972351 | 0.99 | 0.81 | 1.21 | 9.23E-01 | 0.981 |
| OID30175 | KIF22 | Cardiometabolic_II | KIF22 | Q14807 | 16 | 29790727 | 29805385 | 0.99 | 0.84 | 1.17 | 9.24E-01 | 0.981 |
| OID21488 | SEZ6L | Oncology | SEZ6L | Q9BYH1 | 22 | 26169462 | 26383597 | 0.99 | 0.84 | 1.18 | 9.23E-01 | 0.981 |
| OID31408 | REG3G | Oncology_II | REG3G | Q6UW15 | 2 | 79025686 | 79028505 | 0.99 | 0.85 | 1.16 | 9.24E-01 | 0.982 |
| OID30728 | LYVE1 | Inflammation_II | LYVE1 | Q9Y5Y7 | 11 | 10556966 | 10611689 | 1.01 | 0.85 | 1.19 | 9.25E-01 | 0.982 |
| OID21396 | PDCD1 | Oncology | PDCD1 | Q15116 | 2 | 241849884 | 241858894 | 1.01 | 0.87 | 1.17 | 9.26E-01 | 0.982 |
| OID30750 | SERPINA1 | Inflammation_II | SERPINA1 | P01009 | 14 | 94376747 | 94390693 | 0.99 | 0.84 | 1.17 | 9.25E-01 | 0.982 |
| OID21235 | NPTN | Oncology | NPTN | Q9Y639 | 15 | 73560014 | 73634134 | 0.99 | 0.84 | 1.17 | 9.26E-01 | 0.982 |
| OID21508 | XPNPEP2 | Oncology | XPNPEP2 | O43895 | X | 129738949 | 129769536 | 0.99 | 0.84 | 1.17 | 9.26E-01 | 0.982 |
| OID20811 | ATP5PO | Neurology | ATP5PO | P48047 | 21 | 33903453 | 33915814 | 1.01 | 0.86 | 1.17 | 9.30E-01 | 0.982 |
| OID20982 | EPHB6 | Neurology | EPHB6 | O15197 | 7 | 142855061 | 142871094 | 1.01 | 0.87 | 1.17 | 9.28E-01 | 0.982 |
| OID30818 | FZD8 | Neurology_II | FZD8 | Q9H461 | 10 | 35638247 | 35642296 | 0.99 | 0.85 | 1.17 | 9.29E-01 | 0.982 |
| OID20493 | ITGB6 | Inflammation | ITGB6 | P18564 | 2 | 160099667 | 160200313 | 1.01 | 0.85 | 1.20 | 9.29E-01 | 0.982 |
| OID21100 | MPO | Neurology | MPO | P05164 | 17 | 58269855 | 58280935 | 1.01 | 0.85 | 1.20 | 9.29E-01 | 0.982 |
| OID30939 | MSLNL | Neurology_II | MSLNL | Q96KJ4 | 16 | 769428 | 783370 | 1.01 | 0.86 | 1.18 | 9.28E-01 | 0.982 |
| OID30226 | ROBO4 | Cardiometabolic_II | ROBO4 | Q8WZ75 | 11 | 124883691 | 124898500 | 1.01 | 0.86 | 1.19 | 9.27E-01 | 0.982 |
| OID30691 | TGOLN2 | Inflammation_II | TGOLN2 | O43493 | 2 | 85318027 | 85328251 | 1.01 | 0.86 | 1.18 | 9.29E-01 | 0.982 |
| OID20842 | TPPP3 | Neurology | TPPP3 | Q9BW30 | 16 | 67389809 | 67393518 | 0.99 | 0.84 | 1.17 | 9.27E-01 | 0.982 |
| OID31371 | VSIG2 | Oncology_II | VSIG2 | Q96IQ7 | 11 | 124747474 | 124752255 | 1.01 | 0.87 | 1.16 | 9.29E-01 | 0.982 |
| OID20730 | SMOC2 | Inflammation | SMOC2 | Q9H3U7 | 6 | 168441151 | 168673445 | 0.99 | 0.84 | 1.17 | 9.31E-01 | 0.983 |
| OID20724 | CD40 | Inflammation | CD40 | P25942 | 20 | 46118278 | 46129863 | 0.99 | 0.84 | 1.17 | 9.34E-01 | 0.984 |
| OID30405 | CLEC4M | Cardiometabolic_II | CLEC4M | Q9H2X3 | 19 | 7763210 | 7769605 | 1.01 | 0.85 | 1.19 | 9.35E-01 | 0.984 |
| OID30542 | GIPR | Inflammation_II | GIPR | P48546 | 19 | 45668221 | 45683722 | 0.99 | 0.84 | 1.17 | 9.34E-01 | 0.984 |
| OID30256 | HSDL2 | Cardiometabolic_II | HSDL2 | Q6YN16 | 9 | 112380080 | 112472405 | 0.99 | 0.84 | 1.17 | 9.35E-01 | 0.984 |
| OID20246 | IGFBP1 | Cardiometabolic | IGFBP1 | P08833 | 7 | 45888360 | 45893660 | 0.99 | 0.83 | 1.19 | 9.34E-01 | 0.984 |
| OID20989 | PSME2 | Neurology | PSME2 | Q9UL46 | 14 | 24143362 | 24147570 | 0.99 | 0.84 | 1.18 | 9.35E-01 | 0.984 |
| OID30107 | SLC51B | Cardiometabolic_II | SLC51B | Q86UW2 | 15 | 65045387 | 65053397 | 1.01 | 0.86 | 1.17 | 9.35E-01 | 0.984 |
| OID31233 | TOP1MT | Oncology_II | TOP1MT | Q969P6 | 8 | 143304384 | 143359979 | 0.99 | 0.85 | 1.17 | 9.35E-01 | 0.984 |
| OID20750 | TPP1 | Inflammation | TPP1 | O14773 | 11 | 6612768 | 6619448 | 1.01 | 0.85 | 1.20 | 9.33E-01 | 0.984 |
| OID20939 | WFIKKN1 | Neurology | WFIKKN1 | Q96NZ8 | 16 | 629239 | 634117 | 0.99 | 0.84 | 1.17 | 9.35E-01 | 0.984 |
| OID31237 | BTLA | Oncology_II | BTLA | Q7Z6A9 | 3 | 112463966 | 112499472 | 1.01 | 0.86 | 1.18 | 9.37E-01 | 0.984 |
| OID30719 | C8B | Inflammation_II | C8B | P07358 | 1 | 56929207 | 56974383 | 1.01 | 0.85 | 1.20 | 9.37E-01 | 0.984 |
| OID30363 | CTLA4 | Cardiometabolic_II | CTLA4 | P16410 | 2 | 203853888 | 203873965 | 0.99 | 0.84 | 1.18 | 9.37E-01 | 0.984 |
| OID20113 | CTSH | Cardiometabolic | CTSH | P09668 | 15 | 78921058 | 78949574 | 0.99 | 0.85 | 1.17 | 9.37E-01 | 0.984 |

| | | | | | | | | | | | | |
|---|---|---|---|---|---|---|---|---|---|---|---|---|
| OID30173 | COPB2 | Cardiometabolic_II | COPB2 | P35606 | 3 | 139353946 | 139389736 | 0.99 | 0.84 | 1.18 | 9.38E-01 | 0.984 |
| OID30298 | SPESP1 | Cardiometabolic_II | SPESP1 | Q6UW49 | 15 | 68818221 | 68946811 | 1.01 | 0.86 | 1.18 | 9.37E-01 | 0.984 |
| OID31294 | TK1 | Oncology_II | TK1 | P04183 | 17 | 78174091 | 78187233 | 1.01 | 0.85 | 1.20 | 9.38E-01 | 0.984 |
| OID30915 | CADPS | Neurology_II | CADPS | Q9ULU8 | 3 | 62398346 | 62875416 | 0.99 | 0.85 | 1.16 | 9.41E-01 | 0.985 |
| OID30176 | CSRP3 | Cardiometabolic_II | CSRP3 | P50461 | 11 | 19182030 | 19210571 | 1.01 | 0.85 | 1.19 | 9.40E-01 | 0.985 |
| OID20712 | DNER | Inflammation | DNER | Q8NFT8 | 2 | 229357629 | 229714555 | 0.99 | 0.83 | 1.19 | 9.40E-01 | 0.985 |
| OID30813 | KCNC4 | Neurology_II | KCNC4 | Q03721 | 1 | 110210314 | 110283100 | 1.01 | 0.87 | 1.17 | 9.39E-01 | 0.985 |
| OID20343 | MCAM | Cardiometabolic | MCAM | P43121 | 11 | 119308529 | 119321521 | 1.01 | 0.85 | 1.19 | 9.41E-01 | 0.985 |
| OID21167 | NINJ1 | Oncology | NINJ1 | Q92982 | 9 | 93121496 | 93134251 | 0.99 | 0.83 | 1.18 | 9.41E-01 | 0.985 |
| OID30333 | SCRG1 | Cardiometabolic_II | SCRG1 | O75711 | 4 | 173384701 | 173406380 | 0.99 | 0.83 | 1.18 | 9.41E-01 | 0.985 |
| OID30951 | SCT | Neurology_II | SCT | P09683 | 11 | 626309 | 627181 | 0.99 | 0.85 | 1.16 | 9.40E-01 | 0.985 |
| OID20786 | AGRN | Inflammation | AGRN | O00468 | 1 | 1020120 | 1056118 | 0.99 | 0.85 | 1.17 | 9.46E-01 | 0.985 |
| OID21413 | AMBP | Oncology | AMBP | P02760 | 9 | 114060127 | 114078328 | 1.01 | 0.85 | 1.20 | 9.44E-01 | 0.985 |
| OID30896 | BATF | Neurology_II | BATF | Q16520 | 14 | 75522455 | 75546993 | 1.01 | 0.86 | 1.17 | 9.46E-01 | 0.985 |
| OID20395 | CCL18 | Cardiometabolic | CCL18 | P55774 | 17 | 36064272 | 36072032 | 0.99 | 0.84 | 1.17 | 9.45E-01 | 0.985 |
| OID20695 | CCL4 | Inflammation | CCL4 | P13236 | 17 | 36103827 | 36105621 | 0.99 | 0.85 | 1.17 | 9.43E-01 | 0.985 |
| OID20636 | CLEC7A | Inflammation | CLEC7A | Q9BXN2 | 12 | 10116777 | 10130258 | 0.99 | 0.84 | 1.17 | 9.44E-01 | 0.985 |
| OID20148 | DCTPP1 | Cardiometabolic | DCTPP1 | Q9H773 | 16 | 30423615 | 30430030 | 0.99 | 0.84 | 1.18 | 9.44E-01 | 0.985 |
| OID20840 | EPHA10 | Neurology | EPHA10 | Q5JZY3 | 1 | 37713880 | 37765133 | 0.99 | 0.85 | 1.16 | 9.46E-01 | 0.985 |
| OID20652 | IL18R1 | Inflammation | IL18R1 | Q13478 | 2 | 102311529 | 102398777 | 1.01 | 0.85 | 1.19 | 9.47E-01 | 0.985 |
| OID30924 | LRRC38 | Neurology_II | LRRC38 | Q5VT99 | 1 | 13474973 | 13514003 | 1.01 | 0.86 | 1.17 | 9.45E-01 | 0.985 |
| OID20557 | MAPK9 | Inflammation | MAPK9 | P45984 | 5 | 180233143 | 180292099 | 0.99 | 0.84 | 1.18 | 9.47E-01 | 0.985 |
| OID31304 | PPP1R12B | Oncology_II | PPP1R12B | O60237 | 1 | 202348699 | 202592706 | 1.01 | 0.85 | 1.20 | 9.43E-01 | 0.985 |
| OID21065 | RGMA | Neurology | RGMA | Q96B86 | 15 | 93035271 | 93089211 | 1.01 | 0.86 | 1.18 | 9.43E-01 | 0.985 |
| OID20733 | TNFSF13 | Inflammation | TNFSF13 | O75888 | 17 | 7558292 | 7561608 | 0.99 | 0.84 | 1.18 | 9.43E-01 | 0.985 |
| OID30492 | CSNK1D | Inflammation_II | CSNK1D | P48730 | 17 | 82239023 | 82273700 | 1.01 | 0.85 | 1.19 | 9.47E-01 | 0.986 |
| OID20785 | WFIKKN2 | Inflammation | WFIKKN2 | Q8TEU8 | 17 | 50834650 | 50842353 | 1.01 | 0.85 | 1.19 | 9.48E-01 | 0.986 |
| OID20716 | CD58 | Inflammation | CD58 | P19256 | 1 | 116514534 | 116571039 | 1.01 | 0.86 | 1.18 | 9.49E-01 | 0.986 |
| OID20635 | CD79B | Inflammation | CD79B | P40259 | 17 | 63928740 | 63932336 | 1.01 | 0.86 | 1.18 | 9.50E-01 | 0.986 |
| OID31361 | IL2RG | Oncology_II | IL2RG | P31785 | X | 71107404 | 71112108 | 1.00 | 0.87 | 1.16 | 9.48E-01 | 0.986 |
| OID20980 | OBP2B | Neurology | OBP2B | Q9NPH6 | 9 | 133205277 | 133209250 | 0.99 | 0.81 | 1.22 | 9.50E-01 | 0.986 |
| OID31168 | PDCL2 | Oncology_II | PDCL2 | Q8N4E4 | 4 | 55556519 | 55592245 | 1.00 | 0.87 | 1.16 | 9.49E-01 | 0.986 |
| OID31359 | TG | Oncology_II | TG | P01266 | 8 | 132866958 | 133134903 | 0.99 | 0.85 | 1.17 | 9.50E-01 | 0.986 |
| OID20651 | ADAM23 | Inflammation | ADAM23 | O75077 | 2 | 206443532 | 206621127 | 0.99 | 0.85 | 1.17 | 9.50E-01 | 0.986 |
| OID30936 | MYL6B | Neurology_II | MYL6B | P14649 | 12 | 56152256 | 56159647 | 1.00 | 0.86 | 1.17 | 9.51E-01 | 0.986 |
| OID31124 | PRND | Neurology_II | PRND | Q9UKY0 | 20 | 4721909 | 4728460 | 1.00 | 0.86 | 1.15 | 9.51E-01 | 0.986 |
| OID31225 | TAGLN3 | Oncology_II | TAGLN3 | Q9UI15 | 3 | 111998739 | 112013887 | 1.00 | 0.86 | 1.17 | 9.51E-01 | 0.986 |
| OID31240 | DCLRE1C | Oncology_II | DCLRE1C | Q96SD1 | 10 | 14897359 | 14954432 | 1.00 | 0.86 | 1.18 | 9.54E-01 | 0.987 |
| OID20481 | IL17D | Inflammation | IL17D | Q8TAD2 | 13 | 20702127 | 20723098 | 1.01 | 0.84 | 1.20 | 9.53E-01 | 0.987 |
| OID20450 | IL2RB | Inflammation | IL2RB | P14784 | 22 | 37125843 | 37175054 | 1.00 | 0.86 | 1.16 | 9.53E-01 | 0.987 |
| OID30652 | KLK7 | Inflammation_II | KLK7 | P49862 | 19 | 50976468 | 50984099 | 1.01 | 0.85 | 1.19 | 9.53E-01 | 0.987 |
| OID30100 | NUP50 | Cardiometabolic_II | NUP50 | Q9UKX7 | 22 | 45163925 | 45188017 | 0.99 | 0.83 | 1.19 | 9.54E-01 | 0.987 |
| OID30477 | SMPD3 | Inflammation_II | SMPD3 | Q9NY59 | 16 | 68358327 | 68448508 | 1.00 | 0.85 | 1.19 | 9.54E-01 | 0.987 |
| OID30446 | YY1 | Inflammation_II | YY1 | P25490 | 14 | 100238298 | 100282788 | 1.00 | 0.84 | 1.17 | 9.54E-01 | 0.987 |
| OID31100 | CALCB | Neurology_II | CALCB | P10092 | 11 | 14904997 | 15082342 | 1.00 | 0.85 | 1.18 | 9.55E-01 | 0.987 |
| OID21461 | SMOC1 | Oncology | SMOC1 | Q9H4F8 | 14 | 69854131 | 70032366 | 1.01 | 0.84 | 1.20 | 9.55E-01 | 0.987 |
| OID21421 | VWA1 | Oncology | VWA1 | Q6PCB0 | 1 | 1434861 | 1442882 | 1.00 | 0.85 | 1.19 | 9.56E-01 | 0.987 |
| OID20157 | CDH6 | Cardiometabolic | CDH6 | P55285 | 5 | 31193686 | 31329146 | 1.00 | 0.86 | 1.18 | 9.57E-01 | 0.988 |
| OID21062 | FOLR2 | Neurology | FOLR2 | P14207 | 11 | 72216601 | 72221950 | 1.00 | 0.84 | 1.17 | 9.57E-01 | 0.988 |
| OID20770 | FST | Inflammation | FST | P19883 | 5 | 53480626 | 53487134 | 1.00 | 0.84 | 1.18 | 9.58E-01 | 0.988 |
| OID30280 | LAMB1 | Cardiometabolic_II | LAMB1 | P07942 | 7 | 107923799 | 108003213 | 1.00 | 0.85 | 1.18 | 9.58E-01 | 0.988 |
| OID31109 | LMNB2 | Neurology_II | LMNB2 | Q03252 | 19 | 2427638 | 2456959 | 1.00 | 0.84 | 1.18 | 9.58E-01 | 0.988 |
| OID20368 | THBS4 | Cardiometabolic | THBS4 | P35443 | 5 | 79991311 | 80083287 | 1.00 | 0.84 | 1.18 | 9.58E-01 | 0.988 |
| OID20747 | CRHBP | Inflammation | CRHBP | P24387 | 5 | 76953045 | 76981158 | 1.00 | 0.84 | 1.20 | 9.59E-01 | 0.988 |
| OID20290 | SELE | Cardiometabolic | SELE | P16581 | 1 | 169722640 | 169764705 | 1.00 | 0.85 | 1.19 | 9.62E-01 | 0.991 |
| OID20063 | HEBP1 | Cardiometabolic | HEBP1 | Q9NRV9 | 12 | 12974870 | 13000265 | 1.00 | 0.85 | 1.17 | 9.63E-01 | 0.992 |
| OID31163 | NMRK2 | Oncology_II | NMRK2 | Q9NPI5 | 19 | 3933069 | 3942416 | 1.00 | 0.85 | 1.17 | 9.64E-01 | 0.992 |
| OID20346 | ART3 | Cardiometabolic | ART3 | Q13508 | 4 | 76011184 | 76112802 | 1.00 | 0.85 | 1.18 | 9.64E-01 | 0.992 |
| OID20183 | OLR1 | Cardiometabolic | OLR1 | P78380 | 12 | 10158301 | 10172138 | 1.00 | 0.84 | 1.19 | 9.65E-01 | 0.992 |
| OID21284 | SERPINA9 | Oncology | SERPINA9 | Q86WD7 | 14 | 94462717 | 94479689 | 1.00 | 0.86 | 1.17 | 9.65E-01 | 0.992 |
| OID20568 | NPPC | Inflammation | NPPC | P23582 | 2 | 231921809 | 231926396 | 1.00 | 0.85 | 1.19 | 9.66E-01 | 0.993 |
| OID30119 | USP47 | Cardiometabolic_II | USP47 | Q96K76 | 11 | 11841423 | 11961887 | 1.00 | 0.85 | 1.19 | 9.66E-01 | 0.993 |
| OID31188 | CREB3 | Oncology_II | CREB3 | O43889 | 9 | 35732598 | 35736999 | 1.00 | 0.87 | 1.16 | 9.69E-01 | 0.994 |
| OID21441 | HS6ST1 | Oncology | HS6ST1 | O60243 | 2 | 128236716 | 128318868 | 1.00 | 0.85 | 1.17 | 9.69E-01 | 0.994 |
| OID30329 | NPL | Cardiometabolic_II | NPL | Q9BXD5 | 1 | 182789293 | 182830384 | 1.00 | 0.83 | 1.19 | 9.68E-01 | 0.994 |
| OID20163 | PDCD6 | Cardiometabolic | PDCD6 | O75340 | 5 | 271621 | 314974 | 1.00 | 0.86 | 1.16 | 9.69E-01 | 0.994 |
| OID30843 | CSPG5 | Neurology_II | CSPG5 | O95196 | 3 | 47562238 | 47580792 | 1.00 | 0.87 | 1.15 | 9.70E-01 | 0.994 |
| OID31219 | GATA3 | Oncology_II | GATA3 | P23771 | 10 | 8045378 | 8075198 | 1.00 | 0.85 | 1.19 | 9.70E-01 | 0.995 |
| OID30725 | B2M | Inflammation_II | B2M | P61769 | 15 | 44711358 | 44718851 | 1.00 | 0.85 | 1.18 | 9.72E-01 | 0.995 |
| OID31126 | CAPS | Neurology_II | CAPS | Q13938 | 19 | 5912339 | 5916211 | 1.00 | 0.84 | 1.18 | 9.72E-01 | 0.995 |
| OID31443 | CEACAM19 | Oncology_II | CEACAM19 | Q7Z692 | 19 | 44662278 | 44684359 | 1.00 | 0.86 | 1.17 | 9.71E-01 | 0.995 |
| OID20080 | MTPN | Cardiometabolic | MTPN | P58546 | 7 | 135926760 | 135977359 | 1.00 | 0.85 | 1.17 | 9.72E-01 | 0.995 |
| OID20226 | CCDC80 | Cardiometabolic | CCDC80 | Q76M96 | 3 | 112596797 | 112649530 | 1.00 | 0.84 | 1.18 | 9.73E-01 | 0.995 |
| OID21528 | CEACAM1 | Oncology | CEACAM1 | P13688 | 19 | 42507304 | 42561234 | 1.00 | 0.83 | 1.20 | 9.74E-01 | 0.995 |
| OID21261 | CFC1 | Oncology | CFC1 | P0CG37 | 2 | 130592165 | 130599575 | 1.00 | 0.85 | 1.18 | 9.74E-01 | 0.995 |
| OID30091 | CYP24A1 | Cardiometabolic_II | CYP24A1 | Q07973 | 20 | 54153446 | 54173986 | 1.00 | 0.85 | 1.17 | 9.74E-01 | 0.995 |
| OID30958 | RAC3 | Neurology_II | RAC3 | P60763 | 17 | 82031678 | 82034204 | 1.00 | 0.86 | 1.18 | 9.73E-01 | 0.995 |
| OID20214 | SFTPD | Cardiometabolic | SFTPD | P35247 | 10 | 79937467 | 79982614 | 1.00 | 0.84 | 1.18 | 9.74E-01 | 0.995 |
| OID31211 | MLLT1 | Oncology_II | MLLT1 | Q03111 | 19 | 6210381 | 6279975 | 1.00 | 0.86 | 1.18 | 9.75E-01 | 0.996 |
| OID20470 | NFATC3 | Inflammation | NFATC3 | Q12968 | 16 | 68084751 | 68229259 | 1.00 | 0.85 | 1.18 | 9.75E-01 | 0.996 |
| OID21419 | CLMP | Oncology | CLMP | Q9H6B4 | 11 | 123069872 | 123195248 | 1.00 | 0.83 | 1.20 | 9.76E-01 | 0.996 |
| OID30768 | ORM1 | Inflammation_II | ORM1 | P02763 | 9 | 114323098 | 114326479 | 1.00 | 0.83 | 1.19 | 9.76E-01 | 0.996 |
| OID21262 | SORCS2 | Oncology | SORCS2 | Q96PQ0 | 4 | 7192538 | 7742836 | 1.00 | 0.86 | 1.17 | 9.76E-01 | 0.996 |
| OID20584 | CD4 | Inflammation | CD4 | P01730 | 12 | 6786858 | 6820799 | 1.00 | 0.85 | 1.19 | 9.77E-01 | 0.996 |
| OID30124 | ATP1B1 | Cardiometabolic_II | ATP1B1 | P05026 | 1 | 169105697 | 169310992 | 1.00 | 0.85 | 1.19 | 9.78E-01 | 0.996 |
| OID20116 | BOC | Cardiometabolic | BOC | Q9BWV1 | 3 | 113211003 | 113287459 | 1.00 | 0.85 | 1.19 | 9.82E-01 | 0.996 |
| OID20610 | CCL3 | Inflammation | CCL3 | P10147 | 17 | 36088256 | 36090169 | 1.00 | 0.82 | 1.22 | 9.80E-01 | 0.996 |
| OID31302 | CD3E | Oncology_II | CD3E | P07766 | 11 | 118304730 | 118316175 | 1.00 | 0.84 | 1.19 | 9.82E-01 | 0.996 |
| OID20294 | CSTB | Cardiometabolic | CSTB | P04080 | 21 | 43772511 | 43776330 | 1.00 | 0.84 | 1.19 | 9.81E-01 | 0.996 |
| OID30953 | DCC | Neurology_II | DCC | P43146 | 18 | 52340197 | 53535903 | 1.00 | 0.84 | 1.18 | 9.79E-01 | 0.996 |
| OID31341 | DYNC1H1 | Oncology_II | DYNC1H1 | Q14204 | 14 | 101964573 | 102056443 | 1.00 | 0.83 | 1.21 | 9.82E-01 | 0.996 |
| OID30348 | FAM20A | Cardiometabolic_II | FAM20A | Q96MK3 | 17 | 68535113 | 68601367 | 1.00 | 0.84 | 1.19 | 9.81E-01 | 0.996 |
| OID20316 | FAS | Cardiometabolic | FAS | P25445 | 10 | 88953813 | 89029605 | 1.00 | 0.84 | 1.19 | 9.80E-01 | 0.996 |
| OID21044 | GOLM2 | Neurology | GOLM2 | Q6P4E1 | 15 | 44288719 | 44415758 | 1.00 | 0.85 | 1.18 | 9.80E-01 | 0.996 |
| OID31159 | IZUMO1 | Oncology_II | IZUMO1 | Q8IYV9 | 19 | 48740852 | 48746909 | 1.00 | 0.86 | 1.16 | 9.81E-01 | 0.996 |
| OID20632 | KLRD1 | Inflammation | KLRD1 | Q13241 | 12 | 10226058 | 10329608 | 1.00 | 0.84 | 1.19 | 9.83E-01 | 0.996 |
| OID31331 | MAGEA3 | Oncology_II | MAGEA3 | P43357 | X | 152698767 | 152702347 | 1.00 | 0.85 | 1.18 | 9.80E-01 | 0.996 |
| OID31158 | PBK | Oncology_II | PBK | Q96KB5 | 8 | 27809624 | 27838082 | 1.00 | 0.85 | 1.18 | 9.83E-01 | 0.996 |
| OID31435 | PBXIP1 | Oncology_II | PBXIP1 | Q96AQ6 | 1 | 154944076 | 154956123 | 1.00 | 0.85 | 1.17 | 9.81E-01 | 0.996 |
| OID20598 | PTPRM | Inflammation | PTPRM | P28827 | 18 | 7566782 | 8406861 | 1.00 | 0.85 | 1.19 | 9.78E-01 | 0.996 |
| OID30551 | IL36G | Inflammation_II | IL36G | Q9NZH8 | 2 | 112973203 | 112985658 | 1.00 | 0.86 | 1.17 | 9.85E-01 | 0.998 |
| OID20612 | TLR3 | Inflammation | TLR3 | O15455 | 4 | 186069155 | 186088073 | 1.00 | 0.85 | 1.18 | 9.86E-01 | 0.999 |
| OID30479 | TRAF3 | Inflammation_II | TRAF3 | Q13114 | 14 | 102777449 | 102911500 | 1.00 | 0.84 | 1.19 | 9.86E-01 | 0.999 |
| OID21144 | VSIG4 | Neurology | VSIG4 | Q9Y279 | X | 66021738 | 66040125 | 1.00 | 0.85 | 1.18 | 9.86E-01 | 0.999 |
| OID20851 | VSTM1 | Neurology | VSTM1 | Q6UX27 | 19 | 54040825 | 54063953 | 1.00 | 0.85 | 1.18 | 9.87E-01 | 0.999 |
| OID21068 | CCN5 | Neurology | CCN5 | O76076 | 20 | 44714844 | 44728509 | 1.00 | 0.83 | 1.20 | 9.90E-01 | 0.999 |
| OID31480 | CD101 | Oncology_II | CD101 | Q93033 | 1 | 117001750 | 117036552 | 1.00 | 0.84 | 1.19 | 9.89E-01 | 0.999 |

| | | | | | | | | | | | | |
|---|---|---|---|---|---|---|---|---|---|---|---|---|
| OID30430 | FOLH1 | Inflammation_II | FOLH1 | Q04609 | 11 | 49145092 | 49208638 | 1.00 | 0.85 | 1.18 | 9.91E-01 | 0.999 |
| OID31153 | GIPC2 | Neurology_II | GIPC2 | Q8TF65 | 1 | 77979542 | 78138444 | 1.00 | 0.85 | 1.18 | 9.89E-01 | 0.999 |
| OID20656 | HGF | Inflammation | HGF | P14210 | 7 | 81699010 | 81770438 | 1.00 | 0.84 | 1.19 | 9.89E-01 | 0.999 |
| OID20427 | IL1B | Inflammation | IL1B | P01584 | 2 | 112829751 | 112836816 | 1.00 | 0.85 | 1.18 | 9.89E-01 | 0.999 |
| OID21333 | LRRC25 | Oncology | LRRC25 | Q8N386 | 19 | 18391137 | 18397622 | 1.00 | 0.85 | 1.18 | 9.90E-01 | 0.999 |
| OID30418 | NPC2 | Cardiometabolic_II | NPC2 | P61916 | 14 | 74476192 | 74494177 | 1.00 | 0.85 | 1.18 | 9.88E-01 | 0.999 |
| OID30504 | POF1B | Inflammation_II | POF1B | Q8WVV4 | X | 85277396 | 85379717 | 1.00 | 0.85 | 1.17 | 9.90E-01 | 0.999 |
| OID21512 | TFPI2 | Oncology | TFPI2 | P48307 | 7 | 93885396 | 93890753 | 1.00 | 0.84 | 1.19 | 9.90E-01 | 0.999 |
| OID30524 | ACP1 | Inflammation_II | ACP1 | P24666 | 2 | 264140 | 278283 | 1.00 | 0.84 | 1.20 | 9.99E-01 | 1.000 |
| OID21370 | AMIGO2 | Oncology | AMIGO2 | Q86SJ2 | 12 | 47075707 | 47079959 | 1.00 | 0.85 | 1.18 | 9.98E-01 | 1.000 |
| OID30970 | CIT | Neurology_II | CIT | O14578 | 12 | 119685791 | 119877320 | 1.00 | 0.84 | 1.19 | 9.99E-01 | 1.000 |
| OID21090 | CLEC14A | Neurology | CLEC14A | Q86T13 | 14 | 38254000 | 38256093 | 1.00 | 0.85 | 1.17 | 9.98E-01 | 1.000 |
| OID31182 | CLINT1 | Oncology_II | CLINT1 | Q14677 | 5 | 157785743 | 157859145 | 1.00 | 0.83 | 1.20 | 9.93E-01 | 1.000 |
| OID20332 | COL18A1 | Cardiometabolic | COL18A1 | P39060 | 21 | 45405165 | 45513720 | 1.00 | 0.84 | 1.19 | 9.93E-01 | 1.000 |
| OID31206 | CREBZF | Oncology_II | CREBZF | Q9NS37 | 11 | 85657742 | 85682908 | 1.00 | 0.84 | 1.18 | 9.97E-01 | 1.000 |
| OID20914 | CRTAM | Neurology | CRTAM | O95727 | 11 | 122838500 | 122872643 | 1.00 | 0.86 | 1.17 | 9.99E-01 | 1.000 |
| OID31204 | DNAJA1 | Oncology_II | DNAJA1 | P31689 | 9 | 33025273 | 33039907 | 1.00 | 0.85 | 1.18 | 9.97E-01 | 1.000 |
| OID20497 | ENAH | Inflammation | ENAH | Q8N8S7 | 1 | 225486829 | 225653142 | 1.00 | 0.84 | 1.19 | 9.98E-01 | 1.000 |
| OID30288 | GIP | Cardiometabolic_II | GIP | P09681 | 17 | 48958554 | 48968596 | 1.00 | 0.86 | 1.16 | 9.99E-01 | 1.000 |
| OID31318 | GUCY2C | Oncology_II | GUCY2C | P25092 | 12 | 14612632 | 14696599 | 1.00 | 0.84 | 1.19 | 9.93E-01 | 1.000 |
| OID21433 | HBQ1 | Oncology | HBQ1 | P09105 | 16 | 180459 | 181179 | 1.00 | 0.84 | 1.19 | 9.96E-01 | 1.000 |
| OID30088 | HCG22 | Cardiometabolic_II | HCG22 | E2RYF7 | 6 | 31053450 | 31059890 | 1.00 | 0.85 | 1.18 | 9.98E-01 | 1.000 |
| OID30211 | HRC | Cardiometabolic_II | HRC | P23327 | 19 | 49151198 | 49155396 | 1.00 | 0.85 | 1.17 | 9.94E-01 | 1.000 |
| OID31298 | IL9 | Oncology_II | IL9 | P15248 | 5 | 135892246 | 135895841 | 1.00 | 0.85 | 1.18 | 9.98E-01 | 1.000 |
| OID31431 | KIR2DL2 | Oncology_II | KIR2DL2 | P43627 | 19 | 54754693 | 54769187 | 1.00 | 0.85 | 1.18 | 9.98E-01 | 1.000 |
| OID30131 | MYOM2 | Cardiometabolic_II | MYOM2 | P54296 | 8 | 2045046 | 2165552 | 1.00 | 0.85 | 1.17 | 9.93E-01 | 1.000 |
| OID30988 | NFIC | Neurology_II | NFIC | P08651 | 19 | 3314403 | 3469217 | 1.00 | 0.82 | 1.22 | 9.94E-01 | 1.000 |
| OID31203 | SLC34A3 | Oncology_II | SLC34A3 | Q8N130 | 9 | 137230757 | 137236555 | 1.00 | 0.85 | 1.18 | 9.97E-01 | 1.000 |
| OID20361 | SPARCL1 | Cardiometabolic | SPARCL1 | Q14515 | 4 | 87473335 | 87531061 | 1.00 | 0.86 | 1.17 | 9.91E-01 | 1.000 |
| OID20646 | TNFRSF11A | Inflammation | TNFRSF11A | Q9Y6Q6 | 18 | 62325287 | 62391288 | 1.00 | 0.85 | 1.18 | 9.99E-01 | 1.000 |
| OID31389 | WDR46 | Oncology_II | WDR46 | O15213 | 6 | 33279108 | 33289247 | 1.00 | 0.84 | 1.19 | 9.92E-01 | 1.000 |
| OID21119 | CD177 | Neurology | CD177 | Q8N6Q3 | 19 | 43353686 | 43363172 | 1.00 | 0.85 | 1.17 | 1.00E+00 | 1.000 |
| OID20924 | SPOCK1 | Neurology | SPOCK1 | Q08629 | 5 | 136975298 | 137598379 | 1.00 | 0.85 | 1.18 | 1.00E+00 | 1.000 |

**Table S6 Two-sample Mendelian randomization analysis results using cis-pQTL identified in this present study to test the causal effects of proteins on incident ADRD in participants with a history of MDD at baseline**

| Protein | Olink ID | Protein n | Gene syr | UniProt | Gene CH | Gene start | Gene end | cis-pQTL | F<10 | Q p<0.01 | I2<0.9 | Egger intercept p<0.01 | IVW HR | VW HR Lower | VW HR Upper | IVW P | IVW P ADJ | IVW Q | IVW Q P | Fstat | RAPS HR | RAPS HR Lower | RAPS HR Upper | RAPS P | RAPS P ADJ | Med HR | Med HR Lower | Med HR Upper | Med P | Egger HR | ger HR Lower | ger HR Upper | Egger P value | Intercept Est | tercept Lower | tercept Upper | Intercept P |
|---|---|---|---|---|---|---|---|---|---|---|---|---|---|---|---|---|---|---|---|---|---|---|---|---|---|---|---|---|---|---|---|---|---|---|---|---|---|
| APOE | OID3072 | Inflamma | APOE | P02649 | 19 | 44905791 | 44909393 | 31 | 0 | 0 | 0 | 0 | 0.55 | 0.47 | 0.65 | 2.92E-13 | 5.79E-10 | 38.29 | 0.14 | 236.02 | 0.55 | 0.48 | 0.63 | 2.22E-16 | 4.40E-13 | 0.46 | 0.36 | 0.59 | 1.16E-09 | 0.53 | 0.43 | 0.66 | 1.94E-08 | 0.01 | -0.04 | 0.06 | 0.654 |
| IL10RB | OID2069 | Inflamma | IL10RB | Q08334 | 21 | 33266367 | 33310187 | 21 | 0 | 0 | 0 | 0 | 1.41 | 1.20 | 1.66 | 3.55E-05 | 0.035 | 21.23 | 0.38 | 299.09 | 1.41 | 1.20 | 1.66 | 2.10E-05 | 0.021 | 1.32 | 1.10 | 1.59 | 0.004 | 1.40 | 1.07 | 1.83 | 0.014 | 0.00 | -0.06 | 0.07 | 0.937 |
| SPINK6 | OID2145 | Oncology | SPINK6 | Q6UWN8 | 5 | 148202794 | 148215137 | 13 | 0 | 0 | 0 | 0 | 1.67 | 1.26 | 2.22 | 3.81E-04 | 0.151 | 13.36 | 0.34 | 180.15 | 1.68 | 1.28 | 2.20 | 1.82E-04 | 0.120 | 1.55 | 1.14 | 2.09 | 0.005 | 1.77 | 1.05 | 3.00 | 0.032 | -0.01 | -0.10 | 0.08 | 0.789 |
| PILRA | OID2112 | Neurolog | PILRA | Q9UKJ1 | 7 | 100367530 | 100400096 | 70 | 0 | 0 | 0 | 0 | 1.16 | 1.07 | 1.26 | 3.62E-04 | 0.151 | 54.84 | 0.89 | 349.88 | 1.16 | 1.07 | 1.26 | 3.68E-04 | 0.149 | 1.18 | 1.04 | 1.34 | 0.011 | 1.17 | 0.96 | 1.42 | 0.123 | 0.00 | -0.07 | 0.07 | 0.962 |
| PILRB | OID2019 | Cardiom | PILRB | Q9UKJ0 | 7 | 100352176 | 100367831 | 63 | 0 | 0 | 0 | 0 | 1.14 | 1.06 | 1.23 | 3.71E-04 | 0.151 | 51.85 | 0.82 | 468.45 | 1.14 | 1.06 | 1.23 | 3.76E-04 | 0.149 | 1.14 | 1.01 | 1.28 | 0.031 | 1.23 | 1.05 | 1.45 | 0.011 | -0.03 | -0.10 | 0.03 | 0.305 |
| GDNF | OID2086 | Neurolog | GDNF | P39905 | 5 | 37812677 | 37840041 | 11 | 0 | 0 | 0 | 0 | 0.64 | 0.50 | 0.83 | 0.001 | 0.211 | 4.79 | 0.90 | 229.95 | 0.64 | 0.49 | 0.83 | 0.001 | 0.195 | 0.69 | 0.50 | 0.95 | 0.025 | 0.81 | 0.52 | 1.28 | 0.375 | -0.06 | -0.15 | 0.03 | 0.204 |
| ADAMTS | OID2008 | Cardiom | ADAMTS | Q8TE57 | 5 | 5140330 | 5320304 | 12 | 0 | 0 | 0 | 0 | 1.92 | 1.29 | 2.87 | 0.001 | 0.317 | 5.34 | 0.91 | 92.52 | 1.93 | 1.28 | 2.91 | 0.002 | 0.327 | 2.02 | 1.19 | 3.42 | 0.009 | 2.31 | 0.98 | 5.42 | 0.055 | -0.02 | -0.08 | 0.05 | 0.635 |
| DNPH1 | OID2074 | Inflamma | DNPH1 | O43598 | 6 | 43225629 | 43229481 | 7 | 0 | 0 | 0 | 0 | 0.62 | 0.47 | 0.83 | 0.001 | 0.317 | 4.01 | 0.68 | 251.82 | 0.62 | 0.46 | 0.83 | 0.001 | 0.276 | 0.66 | 0.49 | 0.90 | 0.009 | 0.70 | 0.45 | 1.09 | 0.116 | -0.04 | -0.14 | 0.06 | 0.471 |
| TNFRSF1 | OID2048 | Inflamma | TNFRSF1 | Q96RJ3 | 22 | 41922032 | 41926806 | 18 | 0 | 0 | 0 | 0 | 1.40 | 1.14 | 1.73 | 0.001 | 0.317 | 20.42 | 0.25 | 255.14 | 1.41 | 1.16 | 1.70 | 0.000 | 0.158 | 1.42 | 1.09 | 1.85 | 0.010 | 1.14 | 0.84 | 1.54 | 0.410 | 0.06 | -0.01 | 0.12 | 0.073 |
| CD109 | OID2110 | Neurolog | CD109 | Q6YHK3 | 6 | 73695785 | 73828316 | 62 | 0 | 0 | 0 | 0 | 0.84 | 0.75 | 0.94 | 0.002 | 0.319 | 68.70 | 0.23 | 249.38 | 0.84 | 0.76 | 0.93 | 0.001 | 0.200 | 0.84 | 0.71 | 1.00 | 0.054 | 0.78 | 0.62 | 0.97 | 0.025 | 0.03 | -0.04 | 0.09 | 0.430 |
| MDH1 | OID3066 | Inflamma | MDH1 | P40925 | 2 | 63588609 | 63607197 | 3 | 0 | 0 | 1 | 0 | 3.38 | 1.56 | 7.30 | 0.002 | 0.350 | 3.42 | 0.18 | 88.01 | 3.43 | 1.55 | 7.58 | 0.002 | 0.411 | 2.84 | 1.16 | 6.94 | 0.022 | 0.32 | 0.02 | 5.00 | 0.413 | 0.19 | -0.02 | 0.40 | 0.080 |
| FGR | OID2082 | Neurolog | FGR | P09769 | 1 | 27612064 | 27635185 | 2 | 0 | 0 | 0 | 0 | 3.29 | 1.50 | 7.25 | 0.003 | 0.386 | 1.02 | 0.31 | 128.54 | 3.30 | 1.47 | 7.41 | 0.004 | 0.417 | - | - | - | - | - | - | - | - | - | - | - | - |
| PRDX1 | OID2096 | Neurolog | PRDX1 | Q06830 | 1 | 45510914 | 45542732 | 2 | 0 | 0 | 0 | 0 | 0.19 | 0.06 | 0.57 | 0.003 | 0.386 | 0.22 | 0.64 | 55.58 | 0.18 | 0.06 | 0.61 | 0.005 | 0.560 | - | - | - | - | - | - | - | - | - | - | - | - |
| TNFRSF2 | OID2105 | Neurolog | TNFRSF2 | O75509 | 6 | 47231532 | 47309905 | 7 | 0 | 0 | 0 | 0 | 2.36 | 1.34 | 4.14 | 0.003 | 0.386 | 1.79 | 0.94 | 73.84 | 2.36 | 1.32 | 4.22 | 0.004 | 0.417 | 2.28 | 1.14 | 4.56 | 0.020 | 2.16 | 0.38 | 12.37 | 0.388 | 0.01 | -0.16 | 0.18 | 0.916 |
| VEGFA | OID2065 | Inflamma | VEGFA | P15692 | 6 | 43770184 | 43786487 | 22 | 0 | 0 | 0 | 0 | 1.25 | 1.08 | 1.45 | 0.003 | 0.386 | 19.23 | 0.57 | 355.18 | 1.25 | 1.08 | 1.45 | 0.003 | 0.411 | 1.23 | 1.04 | 1.45 | 0.014 | 1.22 | 0.99 | 1.49 | 0.057 | 0.01 | -0.03 | 0.05 | 0.718 |
| TXNRD1 | OID2113 | Neurolog | TXNRD1 | Q16881 | 12 | 104215779 | 104350307 | 1 | 0 | 0 | 0 | 0 | 0.04 | 0.01 | 0.35 | 0.003 | 0.386 | - | - | 38.24 | 0.04 | 0.00 | 0.48 | 0.010 | 0.560 | - | - | - | - | - | - | - | - | - | - | - | - |
| CXCL16 | OID2028 | Cardiom | CXCL16 | Q9H2A7 | 17 | 4733533 | 4739928 | 9 | 0 | 0 | 0 | 0 | 1.72 | 1.20 | 2.46 | 0.003 | 0.386 | 7.55 | 0.48 | 148.70 | 1.72 | 1.20 | 2.48 | 0.003 | 0.417 | 1.99 | 1.29 | 3.09 | 0.002 | 1.86 | 0.64 | 5.38 | 0.254 | -0.01 | -0.14 | 0.12 | 0.878 |
| [illegible] | [illegible] | [illegible] | [illegible] | [illegible] | [illegible] | [illegible] | [illegible] | [illegible] | [illegible] | [illegible] | [illegible] | [illegible] | [illegible] | [illegible] | [illegible] | [illegible] | [illegible] | [illegible] | [illegible] | [illegible] | [illegible] | [illegible] | [illegible] | [illegible] | [illegible] | [illegible] | [illegible] | [illegible] | [illegible] | [illegible] | [illegible] | [illegible] | [illegible] | [illegible] | [illegible] | [illegible] | [illegible] |

| | | | | | | | | | | | | | | | | | | | | | | | | | | | | | | | | | | | | | |
|---|---|---|---|---|---|---|---|---|---|---|---|---|---|---|---|---|---|---|---|---|---|---|---|---|---|---|---|---|---|---|---|---|---|---|---|---|---|
| IL5RA | OID2060 | Inflamma | IL5RA | Q01344 | 3 | 3066324 | 3126613 | 19 | 0 | 0 | 0 | 0 | 0.88 | 0.75 | 1.03 | 0.120 | 0.863 | 13.08 | 0.79 | 340.83 | 0.88 | 0.75 | 1.03 | 0.121 | 0.810 | 0.89 | 0.73 | 1.10 | 0.278 | 0.91 | 0.69 | 1.18 | 0.474 | -0.01 | -0.07 | 0.05 | 0.784 |
| NIT2 | OID3033 | Cardiom | NIT2 | Q9NQR4 | 3 | 100334739 | 100361635 | 1 | 0 | 0 | 0 | 0 | 0.37 | 0.11 | 1.29 | 0.119 | 0.863 | - | - | 97.74 | 0.37 | 0.10 | 1.34 | 0.129 | 0.812 | - | - | - | - | - | - | - | - | - | - | - | - |
| CCL24 | OID2077 | Inflamma | CCL24 | O00175 | 7 | 75810825 | 75823356 | 40 | 0 | 0 | 0 | 0 | 0.93 | 0.85 | 1.02 | 0.118 | 0.863 | 35.18 | 0.64 | 445.81 | 0.93 | 0.84 | 1.02 | 0.118 | 0.810 | 0.87 | 0.76 | 1.00 | 0.054 | 0.95 | 0.80 | 1.13 | 0.587 | -0.01 | -0.07 | 0.05 | 0.725 |
| CPM | OID2105 | Neurolog | CPM | P14384 | 12 | 68842197 | 68971570 | 5 | 0 | 0 | 0 | 0 | 1.48 | 0.90 | 2.43 | 0.120 | 0.863 | 0.91 | 0.92 | 145.19 | 1.48 | 0.90 | 2.45 | 0.124 | 0.810 | 1.69 | 0.94 | 3.03 | 0.077 | 2.13 | 0.59 | 7.63 | 0.246 | -0.07 | -0.28 | 0.15 | 0.545 |
| FOXO1 | OID2051 | Inflamma | FOXO1 | Q12778 | 13 | 40555667 | 40666641 | 3 | 0 | 0 | 0 | 0 | 0.58 | 0.29 | 1.15 | 0.118 | 0.863 | 0.50 | 0.78 | 127.41 | 0.58 | 0.29 | 1.16 | 0.123 | 0.810 | 0.57 | 0.27 | 1.20 | 0.140 | 0.80 | 0.22 | 2.86 | 0.732 | -0.06 | -0.24 | 0.13 | 0.551 |
| ANKRD5 | OID2116 | Oncology | ANKRD5 | Q6NXT1 | 22 | 37830855 | 37849327 | 1 | 0 | 0 | 0 | 0 | 0.30 | 0.06 | 1.36 | 0.118 | 0.863 | - | - | 79.28 | 0.30 | 0.06 | 1.44 | 0.131 | 0.812 | - | - | - | - | - | - | - | - | - | - | - | - |
| PIK3IP1 | OID2109 | Neurolog | PIK3IP1 | Q96FE7 | 22 | 31281594 | 31292534 | 13 | 0 | 0 | 0 | 0 | 0.82 | 0.64 | 1.05 | 0.120 | 0.863 | 11.39 | 0.50 | 219.65 | 0.82 | 0.64 | 1.05 | 0.120 | 0.810 | 0.87 | 0.64 | 1.17 | 0.358 | 0.93 | 0.62 | 1.38 | 0.702 | -0.03 | -0.11 | 0.05 | 0.462 |
| CDH6 | OID2015 | Cardiom | CDH6 | P55285 | 5 | 31193686 | 31329146 | 36 | 0 | 0 | 0 | 0 | 1.12 | 0.97 | 1.28 | 0.121 | 0.866 | 33.34 | 0.55 | 234.72 | 1.12 | 0.97 | 1.28 | 0.121 | 0.810 | 1.06 | 0.85 | 1.31 | 0.609 | 1.01 | 0.77 | 1.31 | 0.962 | 0.03 | -0.03 | 0.09 | 0.381 |
| TRDMT1 | OID3133 | Oncology | TRDMT1 | O14717 | 10 | 17137336 | 17202054 | 1 | 0 | 0 | 0 | 0 | 0.60 | 0.32 | 1.15 | 0.123 | 0.870 | - | - | 369.08 | 0.60 | 0.31 | 1.15 | 0.126 | 0.812 | - | - | - | - | - | - | - | - | - | - | - | - |
| SCN4B | OID3027 | Cardiom | SCN4B | Q8IWT1 | 11 | 118133377 | 118152888 | 26 | 0 | 0 | 0 | 0 | 1.13 | 0.97 | 1.33 | 0.123 | 0.870 | 19.37 | 0.78 | 242.47 | 1.13 | 0.97 | 1.33 | 0.124 | 0.810 | 1.13 | 0.91 | 1.40 | 0.257 | 1.22 | 0.94 | 1.57 | 0.133 | -0.02 | -0.07 | 0.03 | 0.490 |
| PGF | OID2067 | Inflamma | PGF | P49763 | 14 | 74941834 | 74955626 | 2 | 0 | 0 | 0 | 0 | 1.58 | 0.88 | 2.84 | 0.122 | 0.870 | 1.10 | 0.29 | 285.18 | 1.59 | 0.88 | 2.85 | 0.124 | 0.810 | - | - | - | - | - | - | - | - | - | - | - | - |
| CEP152 | OID3124 | Oncology | CEP152 | O94986 | 15 | 48712928 | 48811146 | 1 | 0 | 0 | 0 | 0 | 0.21 | 0.03 | 1.52 | 0.123 | 0.870 | - | - | 52.49 | 0.21 | 0.03 | 1.68 | 0.142 | 0.823 | - | - | - | - | - | - | - | - | - | - | - | - |
| GRN | OID2115 | Neurolog | GRN | P28799 | 17 | 44345246 | 44353106 | 5 | 0 | 0 | 0 | 0 | 0.67 | 0.40 | 1.12 | 0.125 | 0.872 | 7.13 | 0.13 | 247.87 | 0.67 | 0.46 | 0.98 | 0.040 | 0.691 | 0.64 | 0.43 | 0.95 | 0.027 | 0.32 | 0.12 | 0.86 | 0.024 | 0.16 | -0.03 | 0.34 | 0.106 |
| ANGPTL | OID2070 | Inflamma | ANGPTL | Q9BY76 | 19 | 8363289 | 8374370 | 5 | 0 | 0 | 0 | 0 | 0.78 | 0.57 | 1.07 | 0.124 | 0.872 | 3.65 | 0.46 | 318.15 | 0.78 | 0.57 | 1.07 | 0.125 | 0.811 | 0.78 | 0.56 | 1.08 | 0.135 | 0.66 | 0.37 | 1.18 | 0.160 | 0.04 | -0.07 | 0.14 | 0.493 |
| CD207 | OID2129 | Oncology | CD207 | Q9UJ71 | 2 | 70830211 | 70835816 | 16 | 0 | 0 | 0 | 0 | 0.86 | 0.71 | 1.04 | 0.126 | 0.876 | 13.60 | 0.56 | 286.40 | 0.86 | 0.71 | 1.04 | 0.126 | 0.812 | 0.82 | 0.65 | 1.03 | 0.093 | 0.84 | 0.62 | 1.12 | 0.231 | 0.01 | -0.06 | 0.08 | 0.784 |
| AMBN | OID2043 | Inflamma | AMBN | Q9NP70 | 4 | 70592256 | 70607288 | 2 | 0 | 0 | 0 | 0 | 0.44 | 0.15 | 1.26 | 0.126 | 0.876 | 0.05 | 0.82 | 64.32 | 0.44 | 0.15 | 1.31 | 0.138 | 0.821 | - | - | - | - | - | - | - | - | - | - | - | - |
| ANGPTL1 | OID2021 | Cardiom | ANGPTL | O95841 | 1 | 178849535 | 178871077 | 17 | 0 | 0 | 0 | 0 | 0.86 | 0.69 | 1.07 | 0.178 | 0.879 | 11.00 | 0.81 | 199.21 | 0.86 | 0.69 | 1.07 | 0.179 | 0.823 | 0.78 | 0.59 | 1.04 | 0.092 | 0.83 | 0.55 | 1.25 | 0.378 | 0.01 | -0.07 | 0.08 | 0.853 |
| BCAN | OID2099 | Neurolog | BCAN | Q96GW7 | 1 | 156641390 | 156659532 | 3 | 0 | 0 | 0 | 0 | 0.76 | 0.50 | 1.15 | 0.192 | 0.879 | 1.30 | 0.52 | 317.35 | 0.76 | 0.50 | 1.15 | 0.194 | 0.836 | 0.78 | 0.51 | 1.19 | 0.247 | 0.41 | 0.01 | 19.12 | 0.650 | 0.14 | -0.72 | 1.00 | 0.752 |
| CD48 | OID2069 | Inflamma | CD48 | P09326 | 1 | 160678746 | 160711831 | 16 | 0 | 0 | 0 | 0 | 1.13 | 0.96 | 1.34 | 0.144 | 0.879 | 17.81 | 0.27 | 431.17 | 1.13 | 0.97 | 1.32 | 0.111 | 0.809 | 1.09 | 0.91 | 1.29 | 0.343 | 1.09 | 0.86 | 1.40 | 0.473 | 0.01 | -0.05 | 0.08 | 0.685 |
| CTBS | OID3071 | Inflamma | CTBS | Q01459 | 1 | 84549611 | 84574480 | 13 | 0 | 0 | 0 | 0 | 1.13 | 0.95 | 1.36 | 0.168 | 0.879 | 8.56 | 0.74 | 378.27 | 1.13 | 0.95 | 1.36 | 0.169 | 0.823 | 1.16 | 0.93 | 1.44 | 0.190 | 1.23 | 0.98 | 1.54 | 0.071 | -0.04 | -0.10 | 0.02 | 0.240 |
| FCAMR | OID3033 | Cardiom | FCAMR | Q8WWV | 1 | 206957965 | 206970625 | 11 | 0 | 0 | 0 | 0 | 1.18 | 0.93 | 1.51 | 0.180 | 0.879 | 2.81 | 0.99 | 240.21 | 1.18 | 0.92 | 1.52 | 0.182 | 0.824 | 1.20 | 0.91 | 1.57 | 0.192 | 1.23 | 0.88 | 1.73 | 0.225 | -0.01 | -0.07 | 0.05 | 0.730 |
| FCRL6 | OID2052 | Inflamma | FCRL6 | Q6DN72 | 1 | 159800511 | 159816257 | 23 | 0 | 0 | 0 | 0 | 0.92 | 0.81 | 1.03 | 0.156 | 0.879 | 16.41 | 0.80 | 448.63 | 0.92 | 0.81 | 1.03 | 0.157 | 0.823 | 0.92 | 0.80 | 1.05 | 0.220 | 0.96 | 0.82 | 1.12 | 0.561 | -0.02 | -0.07 | 0.03 | 0.395 |
| GIPC2 | OID3115 | Neurolog | GIPC2 | Q8TF65 | 1 | 77979542 | 78138444 | 11 | 0 | 0 | 0 | 0 | 0.83 | 0.64 | 1.07 | 0.145 | 0.879 | 4.29 | 0.93 | 224.80 | 0.83 | 0.64 | 1.07 | 0.146 | 0.823 | 0.77 | 0.56 | 1.06 | 0.105 | 0.70 | 0.43 | 1.14 | 0.150 | 0.04 | -0.06 | 0.13 | 0.432 |
| NTproBN | OID2012 | Cardiom | NPPB | P16860 | 1 | 11857464 | 11858945 | 5 | 0 | 0 | 0 | 0 | 1.28 | 0.90 | 1.80 | 0.166 | 0.879 | 3.21 | 0.52 | 308.40 | 1.28 | 0.90 | 1.80 | 0.167 | 0.823 | 1.23 | 0.85 | 1.77 | 0.266 | 1.08 | 0.61 | 1.91 | 0.785 | 0.03 | -0.06 | 0.12 | 0.475 |
| OPTC | OID2136 | Oncology | OPTC | Q9UBM4 | 1 | 203494153 | 203508949 | 8 | 0 | 0 | 0 | 0 | 0.77 | 0.53 | 1.12 | 0.168 | 0.879 | 5.20 | 0.64 | 144.66 | 0.77 | 0.52 | 1.12 | 0.170 | 0.823 | 0.88 | 0.55 | 1.40 | 0.584 | 0.66 | 0.29 | 1.51 | 0.325 | 0.03 | -0.12 | 0.18 | 0.685 |
| PARP1 | OID2050 | Inflamma | PARP1 | P09874 | 1 | 226360210 | 226408154 | 1 | 0 | 0 | 0 | 0 | 1.78 | 0.78 | 4.08 | 0.173 | 0.879 | - | - | 247.04 | 1.78 | 0.77 | 4.11 | 0.178 | 0.823 | - | - | - | - | - | - | - | - | - | - | - | - |
| PLA2G2 | OID2027 | Cardiom | PLA2G2 | P14555 | 1 | 19975431 | 19980416 | 18 | 0 | 0 | 0 | 0 | 0.84 | 0.66 | 1.06 | 0.137 | 0.879 | 29.13 | 0.03 | 302.90 | 0.84 | 0.70 | 1.00 | 0.051 | 0.702 | 0.81 | 0.66 | 1.00 | 0.054 | 0.99 | 0.66 | 1.51 | 0.980 | -0.05 | -0.16 | 0.05 | 0.327 |
| PMVK | OID2085 | Neurolog | PMVK | Q15126 | 1 | 154924740 | 154936719 | 2 | 0 | 0 | 0 | 0 | 2.97 | 0.67 | 13.21 | 0.153 | 0.879 | 0.33 | 0.57 | 36.16 | 2.98 | 0.62 | 14.40 | 0.174 | 0.823 | - | - | - | - | - | - | - | - | - | - | - | - |
| RSPO1 | OID2093 | Neurolog | RSPO1 | Q2MKA7 | 1 | 37611350 | 37634892 | 4 | 0 | 0 | 0 | 0 | 1.28 | 0.93 | 1.76 | 0.127 | 0.879 | 0.96 | 0.81 | 399.99 | 1.28 | 0.93 | 1.76 | 0.129 | 0.812 | 1.26 | 0.90 | 1.77 | 0.181 | 1.20 | 0.80 | 1.80 | 0.387 | 0.02 | -0.06 | 0.10 | 0.605 |
| S100A14 | OID3098 | Neurolog | S100A14 | Q9HCY8 | 1 | 153614255 | 153616986 | 3 | 0 | 0 | 0 | 0 | 1.48 | 0.89 | 2.45 | 0.133 | 0.879 | 2.09 | 0.35 | 207.75 | 1.48 | 0.89 | 2.46 | 0.135 | 0.812 | 1.44 | 0.85 | 2.43 | 0.173 | 0.54 | 0.12 | 2.45 | 0.429 | 0.15 | -0.06 | 0.36 | 0.168 |
| SCAMP3 | OID2133 | Oncology | SCAMP3 | O14828 | 1 | 155255979 | 155262430 | 3 | 0 | 0 | 1 | 0 | 1.91 | 0.76 | 4.78 | 0.166 | 0.879 | 0.48 | 0.79 | 64.94 | 1.91 | 0.75 | 4.90 | 0.176 | 0.823 | 2.06 | 0.74 | 5.71 | 0.166 | 4.91 | 0.20 | 121.12 | 0.330 | -0.08 | -0.34 | 0.18 | 0.547 |
| TRIM58 | OID3142 | Oncology | TRIM58 | Q8NG06 | 1 | 247857187 | 247880138 | 5 | 0 | 0 | 0 | 0 | 1.34 | 0.86 | 2.08 | 0.201 | 0.879 | 0.31 | 0.99 | 168.61 | 1.34 | 0.85 | 2.09 | 0.205 | 0.842 | 1.36 | 0.85 | 2.18 | 0.206 | 1.34 | 0.35 | 5.09 | 0.667 | 0.00 | -0.25 | 0.25 | 0.996 |
| VAV3 | OID3079 | Neurolog | VAV3 | Q9UKW4 | 1 | 107571161 | 107965180 | 2 | 0 | 0 | 0 | 0 | 2.68 | 0.73 | 9.79 | 0.136 | 0.879 | 0.25 | 0.61 | 45.83 | 2.69 | 0.69 | 10.41 | 0.152 | 0.823 | - | - | - | - | - | - | - | - | - | - | - | - |
| ACP1 | OID3052 | Inflamma | ACP1 | P24666 | 2 | 264140 | 278283 | 10 | 0 | 0 | 0 | 0 | 0.84 | 0.65 | 1.09 | 0.194 | 0.879 | 5.91 | 0.75 | 231.71 | 0.84 | 0.65 | 1.09 | 0.195 | 0.836 | 0.88 | 0.65 | 1.20 | 0.417 | 1.03 | 0.61 | 1.73 | 0.909 | -0.04 | -0.13 | 0.05 | 0.372 |
| ANXA4 | OID2012 | Cardiom | ANXA4 | P09525 | 2 | 69644425 | 69827112 | 1 | 0 | 0 | 0 | 0 | 3.77 | 0.51 | 27.89 | 0.193 | 0.879 | - | - | 37.46 | 3.77 | 0.45 | 31.67 | 0.221 | 0.857 | - | - | - | - | - | - | - | - | - | - | - | - |
| CASP10 | OID2089 | Neurolog | CASP10 | Q92851 | 2 | 201182898 | 201229406 | 1 | 0 | 0 | 0 | 0 | 1.44 | 0.86 | 2.42 | 0.166 | 0.879 | - | - | 571.33 | 1.44 | 0.86 | 2.43 | 0.167 | 0.823 | - | - | - | - | - | - | - | - | - | - | - | - |
| CD302 | OID2132 | Oncology | CD302 | Q8IX05 | 2 | 159768628 | 159798255 | 12 | 0 | 0 | 0 | 0 | 0.82 | 0.62 | 1.08 | 0.163 | 0.879 | 7.82 | 0.73 | 187.14 | 0.82 | 0.62 | 1.09 | 0.165 | 0.823 | 0.87 | 0.63 | 1.20 | 0.405 | 0.91 | 0.58 | 1.41 | 0.663 | -0.02 | -0.11 | 0.06 | 0.558 |
| CGREF1 | OID2015 | Cardiom | CGREF1 | Q99674 | 2 | 27098889 | 27119128 | 43 | 0 | 0 | 0 | 0 | 1.09 | 0.97 | 1.22 | 0.162 | 0.879 | 44.24 | 0.38 | 298.78 | 1.09 | 0.97 | 1.22 | 0.151 | 0.823 | 1.10 | 0.91 | 1.32 | 0.328 | 1.19 | 0.96 | 1.48 | 0.118 | -0.03 | -0.10 | 0.03 | 0.337 |
| DPP4 | OID2040 | Cardiom | DPP4 | P27487 | 2 | 161992245 | 162074394 | 7 | 0 | 0 | 0 | 0 | 1.41 | 0.87 | 2.27 | 0.160 | 0.879 | 9.70 | 0.14 | 172.07 | 1.41 | 0.97 | 2.06 | 0.074 | 0.776 | 1.42 | 0.94 | 2.13 | 0.093 | 1.66 | 0.48 | 5.77 | 0.428 | -0.03 | -0.25 | 0.19 | 0.778 |
| EIF2AK2 | OID3144 | Oncology | EIF2AK2 | P19525 | 2 | 37099210 | 37157522 | 2 | 0 | 0 | 0 | 0 | 1.50 | 0.82 | 2.76 | 0.192 | 0.879 | 2.28 | 0.13 | 214.44 | 1.50 | 0.81 | 2.78 | 0.193 | 0.836 | - | - | - | - | - | - | - | - | - | - | - | - |
| FN1 | OID3078 | Inflamma | FN1 | P02751 | 2 | 215360440 | 215436073 | 12 | 0 | 0 | 0 | 0 | 1.17 | 0.92 | 1.47 | 0.195 | 0.879 | 8.13 | 0.70 | 251.80 | 1.17 | 0.92 | 1.47 | 0.196 | 0.836 | 1.23 | 0.96 | 1.58 | 0.106 | 1.29 | 0.95 | 1.77 | 0.104 | -0.03 | -0.08 | 0.03 | 0.324 |
| IL1R1 | OID2111 | Neurolog | IL1R1 | P14778 | 2 | 102064544 | 102179874 | 7 | 0 | 0 | 0 | 0 | 0.75 | 0.51 | 1.10 | 0.139 | 0.879 | 5.34 | 0.50 | 157.30 | 0.75 | 0.51 | 1.10 | 0.140 | 0.823 | 0.59 | 0.38 | 0.94 | 0.026 | 0.44 | 0.14 | 1.36 | 0.153 | 0.08 | -0.08 | 0.23 | 0.326 |
| MSTN | OID2011 | Cardiom | MSTN | O14793 | 2 | 190055700 | 190062729 | 1 | 0 | 0 | 0 | 0 | 0.27 | 0.04 | 1.66 | 0.157 | 0.879 | - | - | 49.82 | 0.27 | 0.04 | 1.83 | 0.178 | 0.823 | - | - | - | - | - | - | - | - | - | - | - | - |
| RALB | OID3056 | Inflamma | RALB | P11234 | 2 | 120240064 | 120294710 | 2 | 0 | 0 | 0 | 0 | 0.61 | 0.29 | 1.29 | 0.195 | 0.879 | 0.17 | 0.68 | 137.41 | 0.61 | 0.29 | 1.30 | 0.201 | 0.836 | - | - | - | - | - | - | - | - | - | - | - | - |
| RPE | OID3132 | Oncology | RPE | Q96AT9 | 2 | 210002565 | 210022260 | 1 | 0 | 0 | 0 | 0 | 3.62 | 0.69 | 19.09 | 0.129 | 0.879 | - | - | 56.01 | 3.62 | 0.64 | 20.66 | 0.147 | 0.823 | - | - | - | - | - | - | - | - | - | - | - | - |
| SESTD1 | OID2079 | Neurolog | SESTD1 | Q86VW0 | 2 | 179101678 | 179264832 | 2 | 0 | 0 | 0 | 0 | 1.80 | 0.74 | 4.38 | 0.198 | 0.879 | 1.01 | 0.32 | 98.79 | 1.80 | 0.73 | 4.46 | 0.204 | 0.842 | - | - | - | - | - | - | - | - | - | - | - | - |
| SNED1 | OID3020 | Cardiom | SNED1 | Q8TER0 | 2 | 240998618 | 241095568 | 5 | 0 | 0 | 0 | 0 | 1.45 | 0.84 | 2.49 | 0.184 | 0.879 | 4.57 | 0.33 | 119.89 | 1.45 | 0.87 | 2.42 | 0.157 | 0.823 | 1.25 | 0.69 | 2.27 | 0.455 | 0.52 | 0.13 | 2.06 | 0.352 | 0.11 | -0.03 | 0.25 | 0.117 |
| VSNL1 | OID3110 | Neurolog | VSNL1 | P62760 | 2 | 17539126 | 17657018 | 15 | 0 | 0 | 0 | 0 | 0.82 | 0.61 | 1.10 | 0.193 | 0.879 | 8.82 | 0.84 | 148.61 | 0.82 | 0.61 | 1.11 | 0.195 | 0.836 | 0.83 | 0.59 | 1.18 | 0.299 | 0.91 | 0.48 | 1.69 | 0.755 | -0.02 | -0.14 | 0.10 | 0.731 |
| AHSG | OID3070 | Inflamma | AHSG | P02765 | 3 | 186613060 | 186621318 | 20 | 0 | 0 | 0 | 0 | 1.13 | 0.96 | 1.33 | 0.139 | 0.879 | 25.38 | 0.15 | 414.78 | 1.13 | 0.98 | 1.31 | 0.087 | 0.805 | 1.18 | 0.97 | 1.44 | 0.088 | 1.03 | 0.82 | 1.29 | 0.783 | 0.03 | -0.02 | 0.09 | 0.246 |
| CHL1 | OID2036 | Cardiom | CHL1 | O00533 | 3 | 196763 | 409417 | 14 | 0 | 0 | 0 | 0 | 1.14 | 0.94 | 1.37 | 0.194 | 0.879 | 11.95 | 0.53 | 305.72 | 1.14 | 0.94 | 1.38 | 0.194 | 0.836 | 1.18 | 0.92 | 1.50 | 0.192 | 1.33 | 0.83 | 2.13 | 0.239 | -0.05 | -0.17 | 0.08 | 0.476 |
| CNTN4 | OID2111 | Neurolog | CNTN4 | Q8IWV2 | 3 | 2098813 | 3057959 | 24 | 0 | 0 | 0 | 0 | 0.86 | 0.68 | 1.08 | 0.190 | 0.879 | 31.41 | 0.11 | 182.87 | 0.86 | 0.70 | 1.04 | 0.124 | 0.810 | 0.85 | 0.64 | 1.14 | 0.292 | 0.85 | 0.58 | 1.26 | 0.428 | 0.00 | -0.05 | 0.05 | 0.983 |
| CPB1 | OID2028 | Cardiom | CPB1 | P15086 | 3 | 148791102 | 148860187 | 1 | 0 | 0 | 0 | 0 | 1.89 | 0.81 | 4.42 | 0.144 | 0.879 | - | - | 221.44 | 1.89 | 0.80 | 4.46 | 0.149 | 0.823 | - | - | - | - | - | - | - | - | - | - | - | - |
| FETUB | OID2040 | Cardiom | FETUB | Q9UGM5 | 3 | 186635969 | 186653141 | 8 | 0 | 0 | 0 | 0 | 1.28 | 0.88 | 1.86 | 0.199 | 0.879 | 10.17 | 0.18 | 197.40 | 1.28 | 0.94 | 1.75 | 0.121 | 0.810 | 1.37 | 0.93 | 2.00 | 0.107 | 1.52 | 0.82 | 2.83 | 0.186 | -0.04 | -0.13 | 0.06 | 0.482 |
| GASK1A | OID3010 | Cardiom | GASK1A | Q9UFP1 | 3 | 42979087 | 43067898 | 40 | 0 | 0 | 0 | 0 | 1.09 | 0.96 | 1.24 | 0.182 | 0.879 | 36.17 | 0.60 | 221.39 | 1.09 | 0.96 | 1.25 | 0.182 | 0.824 | 1.09 | 0.90 | 1.31 | 0.372 | 1.03 | 0.82 | 1.30 | 0.784 | 0.02 | -0.04 | 0.07 | 0.557 |
| GLB1 | OID2094 | Neurolog | GLB1 | P16278 | 3 | 32996609 | 33097202 | 8 | 0 | 0 | 0 | 0 | 0.77 | 0.54 | 1.10 | 0.153 | 0.879 | 2.44 | 0.93 | 162.00 | 0.77 | 0.53 | 1.11 | 0.156 | 0.823 | 0.78 | 0.52 | 1.15 | 0.202 | 0.71 | 0.37 | 1.39 | 0.318 | 0.01 | -0.08 | 0.10 | 0.793 |
| HRG | OID3077 | Inflamma | HRG | P04196 | 3 | 186660216 | 186678234 | 14 | 0 | 0 | 0 | 0 | 1.16 | 0.95 | 1.42 | 0.149 | 0.879 | 13.72 | 0.39 | 277.63 | 1.16 | 0.95 | 1.42 | 0.138 | 0.821 | 1.07 | 0.84 | 1.36 | 0.571 | 1.11 | 0.77 | 1.58 | 0.578 | 0.01 | -0.07 | 0.09 | 0.741 |
| IL20RB | OID3046 | Inflamma | IL20RB | Q6UXL0 | 3 | 136946230 | 137011085 | 2 | 0 | 0 | 0 | 0 | 0.64 | 0.33 | 1.26 | 0.196 | 0.879 | 0.82 | 0.36 | 163.62 | 0.64 | 0.33 | 1.26 | 0.200 | 0.836 | - | - | - | - | - | - | - | - | - | - | - | - |
| ITIH4 | OID3076 | Inflamma | ITIH4 | Q14624 | 3 | 52812962 | 52830688 | 14 | 0 | 0 | 0 | 0 | 1.25 | 0.92 | 1.69 | 0.155 | 0.879 | 18.12 | 0.15 | 177.58 | 1.25 | 0.96 | 1.62 | 0.094 | 0.806 | 1.24 | 0.88 | 1.74 | 0.223 | 1.00 | 0.44 | 2.25 | 0.994 | 0.05 | -0.11 | 0.21 | 0.559 |
| PRKAR2 | OID3111 | Neurolog | PRKAR2 | P13861 | 3 | 48744597 | 48847874 | 3 | 0 | 0 | 1 | 0 | 2.09 | 0.69 | 6.34 | 0.192 | 0.879 | 0.08 | 0.96 | 42.48 | 2.09 | 0.66 | 6.65 | 0.211 | 0.849 | 2.10 | 0.59 | 7.44 | 0.252 | 1.60 | 0.00 | 1035.16 | 0.888 | 0.02 | -0.38 | 0.42 | 0.934 |
| ROBO2 | OID2100 | Neurolog | ROBO2 | Q9HCK4 | 3 | 75906695 | 77649964 | 9 | 0 | 0 | 1 | 0 | 1.50 | 0.87 | 2.59 | 0.148 | 0.879 | 2.12 | 0.98 | 66.37 | 1.50 | 0.86 | 2.63 | 0.156 | 0.823 | 1.57 | 0.78 | 3.16 | 0.205 | 0.90 | 0.20 | 4.08 | 0.893 | 0.04 | -0.08 | 0.17 | 0.479 |
| SUMF1 | OID3048 | Inflamma | SUMF1 | Q8NBK3 | 3 | 3700814 | 4467273 | 1 | 0 | 0 | 0 | 0 | 1.80 | 0.76 | 4.23 | 0.180 | 0.879 | - | - | 202.41 | 1.80 | 0.76 | 4.27 | 0.185 | 0.828 | - | - | - | - | - | - | - | - | - | - | - | - |
| THPO | OID2009 | Cardiom | THPO | P40225 | 3 | 184371935 | 184381968 | 1 | 0 | 0 | 0 | 0 | 2.95 | 0.64 | 13.57 | 0.164 | 0.879 | - | - | 67.34 | 2.95 | 0.61 | 14.35 | 0.179 | 0.823 | - | - | - | - | - | - | - | - | - | - | - | - |
| TOP2B | OID3055 | Inflamma | TOP2B | Q02880 | 3 | 25597978 | 25664909 | 1 | 0 | 0 | 0 | 0 | 2.33 | 0.71 | 7.65 | 0.165 | 0.879 | - | - | 106.61 | 2.33 | 0.69 | 7.86 | 0.175 | 0.823 | - | - | - | - | - | - | - | - | - | - | - | - |
| CLGN | OID3095 | Neurolog | CLGN | O14967 | 4 | 140388453 | 140427661 | 19 | 0 | 0 | 0 | 0 | 1.17 | 0.92 | 1.49 | 0.192 | 0.879 | 20.76 | 0.29 | 157.23 | 1.17 | 0.94 | 1.47 | 0.161 | 0.823 | 1.17 | 0.85 | 1.61 | 0.349 | 1.16 | 0.80 | 1.69 | 0.438 | 0.00 | -0.08 | 0.08 | 0.947 |
| F11 | OID3077 | Inflamma | F11 | P03951 | 4 | 186266189 | 186289681 | 16 | 0 | 0 | 0 | 0 | 1.21 | 0.94 | 1.56 | 0.136 | 0.879 | 18.16 | 0.25 | 222.51 | 1.21 | 0.96 | 1.52 | 0.101 | 0.806 | 1.29 | 0.99 | 1.68 | 0.058 | 1.42 | 0.95 | 2.10 | 0.084 | -0.03 | -0.09 | 0.03 | 0.313 |
| GRPEL1 | OID2130 | Oncology | GRPEL1 | Q9HAV7 | 4 | 7058895 | 7068064 | 1 | 0 | 0 | 0 | 0 | 5.74 | 0.60 | 55.04 | 0.130 | 0.879 | - | - | 34.07 | 5.74 | 0.50 | 65.67 | 0.160 | 0.823 | - | - | - | - | - | - | - | - | - | - | - | - |
| NAAA | OID2093 | Neurolog | NAAA | Q02083 | 4 | 75913660 | 75941013 | 35 | 0 | 0 | 0 | 0 | 0.89 | 0.76 | 1.04 | 0.150 | 0.879 | 40.49 | 0.21 | 226.27 | 0.89 | 0.77 | 1.03 | 0.116 | 0.810 | 0.85 | 0.68 | 1.07 | 0.159 | 0.90 | 0.69 | 1.17 | 0.419 | 0.00 | -0.07 | 0.06 | 0.929 |
| ATOX1 | OID2147 | Oncology | ATOX1 | O00244 | 5 | 151742316 | 151772532 | 1 | 0 | 0 | 0 | 0 | 0.40 | 0.11 | 1.51 | 0.178 | 0.879 | - | - | 88.58 | 0.40 | 0.10 | 1.57 | 0.190 | 0.833 | - | - | - | - | - | - | - | - | - | - | - | - |
| LECT2 | OID3032 | Cardiom | LECT2 | O14960 | 5 | 135922279 | 135954983 | 39 | 0 | 0 | 0 | 0 | 0.90 | 0.78 | 1.03 | 0.137 | 0.879 | 32.46 | 0.72 | 256.85 | 0.90 | 0.78 | 1.04 | 0.137 | 0.820 | 0.93 | 0.76 | 1.14 | 0.486 | 1.00 | 0.75 | 1.32 | 0.990 | -0.03 | -0.09 | 0.04 | 0.387 |
| SELENO | OID3074 | Inflamma | SELENO | P49908 | 5 | 42799880 | 42887392 | 2 | 0 | 0 | 0 | 0 | 1.97 | 0.82 | 4.72 | 0.130 | 0.879 | 3.52 | 0.06 | 108.36 | 1.99 | 0.82 | 4.82 | 0.128 | 0.812 | - | - | - | - | - | - | - | - | - | - | - | - |
| STC2 | OID2115 | Neurolog | STC2 | O76061 | 5 | 173314723 | 173328447 | 3 | 0 | 0 | 1 | 0 | 0.56 | 0.23 | 1.33 | 0.189 | 0.879 | 0.74 | 0.69 | 76.53 | 0.56 | 0.23 | 1.36 | 0.198 | 0.836 | 0.59 | 0.22 | 1.56 | 0.287 | 2.78 | 0.03 | 241.40 | 0.654 | -0.15 | -0.55 | 0.25 | 0.473 |
| AGER | OID2075 | Inflamma | AGER | Q15109 | 6 | 32180968 | 32184322 | 8 | 0 | 0 | 0 | 0 | 1.29 | 0.91 | 1.84 | 0.158 | 0.879 | 3.78 | 0.81 | 171.10 | 1.29 | 0.90 | 1.84 | 0.160 | 0.823 | 1.46 | 0.95 | 2.26 | 0.086 | 2.04 | 0.66 | 6.27 | 0.213 | -0.07 | -0.23 | 0.09 | 0.399 |
| BTN3A2 | OID2054 | Inflamma | BTN3A2 | P78410 | 6 | 26365159 | 26378320 | 27 | 0 | 0 | 0 | 0 | 1.08 | 0.97 | 1.20 | 0.178 | 0.879 | 20.88 | 0.75 | 523.37 | 1.08 | 0.97 | 1.20 | 0.179 | 0.823 | 1.03 | 0.89 | 1.19 | 0.696 | 1.04 | 0.85 | 1.28 | 0.696 | 0.01 | -0.06 | 0.08 | 0.719 |
| CLPS | OID2115 | Neurolog | CLPS | P04118 | 6 | 35794982 | 35797344 | 46 | 0 | 0 | 0 | 0 | 1.10 | 0.96 | 1.26 | 0.174 | 0.879 | 54.90 | 0.15 | 216.42 | 1.10 | 0.97 | 1.25 | 0.133 | 0.812 | 1.08 | 0.90 | 1.30 | 0.392 | 1.17 | 0.87 | 1.59 | 0.300 | -0.02 | -0.10 | 0.06 | 0.635 |
| DXO | OID3108 | Neurolog | DXO | O77932 | 6 | 31969810 | 31972290 | 9 | 0 | 0 | 0 | 0 | 0.83 | 0.65 | 1.06 | 0.141 | 0.879 | 3.00 | 0.93 | 265.30 | 0.83 | 0.65 | 1.06 | 0.143 | 0.823 | 0.82 | 0.61 | 1.09 | 0.165 | 0.81 | 0.59 | 1.10 | 0.178 | 0.01 | -0.04 | 0.06 | 0.773 |
| GFRAL | OID3109 | Neurolog | GFRAL | Q6UXV0 | 6 | 55327469 | 55402493 | 27 | 0 | 0 | 0 | 0 | 0.88 | 0.75 | 1.04 | 0.137 | 0.879 | 20.25 | 0.78 | 223.69 | 0.88 | 0.75 | 1.04 | 0.138 | 0.821 | 0.84 | 0.67 | 1.06 | 0.138 | 0.98 | 0.64 | 1.50 | 0.939 | -0.03 | -0.13 | 0.07 | 0.583 |
| GRIK2 | OID3097 | Neurolog | GRIK2 | Q13002 | 6 | 100962701 | 102081622 | 5 | 0 | 0 | 0 | 0 | 0.69 | 0.41 | 1.16 | 0.159 | 0.879 | 8.26 | 0.08 | 293.55 | 0.69 | 0.48 | 0.99 | 0.043 | 0.691 | 0.72 | 0.49 | 1.06 | 0.098 | 0.68 | 0.30 | 1.56 | 0.364 | 0.00 | -0.13 | 0.14 | 0.968 |
| LATS1 | OID3052 | Inflamma | LATS1 | O95835 | 6 | 149658153 | 149718105 | 1 | 0 | 0 | 0 | 0 | 0.33 | 0.07 | 1.48 | 0.148 | 0.879 | - | - | 66.66 | 0.33 | 0.07 | 1.57 | 0.164 | 0.823 | - | - | - | - | - | - | - | - | - | - | - | - |
| MEP1A | OID3145 | Oncology | MEP1A | Q16819 | 6 | 46793389 | 46839778 | 4 | 0 | 0 | 0 | 0 | 0.62 | 0.30 | 1.29 | 0.197 | 0.879 | 9.10 | 0.03 | 244.66 | 0.61 | 0.40 | 0.94 | 0.023 | 0.650 | 0.54 | 0.34 | 0.87 | 0.011 | 1.29 | 0.28 | 5.91 | 0.742 | -0.11 | -0.32 | 0.09 | 0.280 |
| MOG | OID2138 | Oncology | MOG | Q16653 | 6 | 29657002 | 29672372 | 11 | 0 | 0 | 0 | 0 | 0.80 | 0.59 | 1.08 | 0.139 | 0.879 | 4.33 | 0.93 | 164.58 | 0.80 | 0.59 | 1.08 | 0.142 | 0.823 | 0.97 | 0.65 | 1.43 | 0.875 | 0.85 | 0.51 | 1.43 | 0.546 | -0.01 | -0.10 | 0.07 | 0.748 |
| PLG | OID3078 | Inflamma | PLG | P00747 | 6 | 160702238 | 160754097 | 27 | 0 | 0 | 0 | 0 | 0.88 | 0.73 | 1.06 | 0.166 | 0.879 | 33.80 | 0.14 | 198.89 | 0.88 | 0.74 | 1.03 | 0.114 | 0.809 | 0.94 | 0.74 | 1.20 | 0.631 | 0.82 | 0.62 | 1.08 | 0.160 | 0.02 | -0.04 | 0.08 | 0.513 |
| PSMG4 | OID3051 | Inflamma | PSMG4 | Q5JS54 | 6 | 3231403 | 3303373 | 2 | 0 | 0 | 0 | 0 | 0.68 | 0.39 | 1.18 | 0.171 | 0.879 | 2.42 | 0.12 | 270.98 | 0.68 | 0.39 | 1.18 | 0.171 | 0.823 | - | - | - | - | - | - | - | - | - | - | - | - |
| PTK7 | OID2088 | Neurolog | PTK7 | Q13308 | 6 | 43076307 | 43161719 | 1 | 0 | 0 | 0 | 0 | 2.82 | 0.65 | 12.26 | 0.167 | 0.879 | - | - | 56.09 | 2.82 | 0.61 | 13.09 | 0.186 | 0.828 | - | - | - | - | - | - | - | - | - | - | - | - |
| RAB44 | OID3134 | Oncology | RAB44 | Q7Z6P3 | 6 | 36697826 | 36733184 | 4 | 0 | 0 | 0 | 0 | 0.67 | 0.39 | 1.14 | 0.137 | 0.879 | 1.45 | 0.69 | 130.41 | 0.67 | 0.39 | 1.14 | 0.141 | 0.823 | 0.68 | 0.37 | 1.23 | 0.200 | 0.56 | 0.15 | 2.05 | 0.381 | 0.03 | -0.19 | 0.26 | 0.771 |
| RNASET | OID2032 | Cardiom | RNASET | O00584 | 6 | 166922113 | 166957191 | 19 | 0 | 0 | 0 | 0 | 0.86 | 0.70 | 1.06 | 0.153 | 0.879 | 18.30 | 0.44 | 208.29 | 0.86 | 0.70 | 1.06 | 0.150 | 0.823 | 0.89 | 0.66 | 1.19 | 0.436 | 0.94 | 0.61 | 1.46 | 0.795 | -0.02 | -0.11 | 0.07 | 0.644 |
| TREML2 | OID2112 | Neurolog | TREML2 | Q5T2D2 | 6 | 41189749 | 41201149 | 13 | 0 | 0 | 0 | 0 | 1.22 | 0.94 | 1.59 | 0.131 | 0.879 | 14.29 | 0.28 | 208.29 | 1.22 | 0.96 | 1.56 | 0.100 | 0.806 | 1.38 | 1.01 | 1.88 | 0.041 | 1.56 | 0.94 | 2.57 | 0.086 | -0.05 | -0.14 | 0.04 | 0.275 |
| COL28A1 | OID3096 | Neurolog | COL28A1 | Q2UY09 | 7 | 7356203 | 7535873 | 25 | 0 | 0 | 0 | 0 | 0.90 | 0.77 | 1.06 | 0.196 | 0.879 | 27.65 | 0.27 | 277.45 | 0.90 | 0.78 | 1.04 | 0.165 | 0.823 | 0.92 | 0.72 | 1.18 | 0.495 | 0.97 | 0.76 | 1.23 | 0.798 | -0.02 | -0.07 | 0.03 | 0.422 |
| GGCT | OID3030 | Cardiom | GGCT | O75223 | 7 | 30496621 | 30504841 | 1 | 0 | 0 | 0 | 0 | 0.53 | 0.21 | 1.30 | 0.164 | 0.879 | - | - | 199.36 | 0.53 | 0.21 | 1.31 | 0.170 | 0.823 | - | - | - | - | - | - | - | - | - | - | - | - |
| GPR37 | OID2009 | Cardiom | GPR37 | O15354 | 7 | 124743885 | 124765792 | 34 | 0 | 0 | 0 | 0 | 1.11 | 0.97 | 1.28 | 0.143 | 0.879 | 20.65 | 0.95 | 249.87 | 1.11 | 0.96 | 1.28 | 0.144 | 0.823 | 1.13 | 0.93 | 1.38 | 0.214 | 1.19 | 0.90 | 1.59 | 0.223 | -0.02 | -0.09 | 0.05 | 0.563 |
| NPTX2 | OID3039 | Cardiom | NPTX2 | P47972 | 7 | 98617285 | 98629869 | 17 | 0 | 0 | 0 | 0 | 0.82 | 0.62 | 1.09 | 0.171 | 0.879 | 18.61 | 0.29 | 192.60 | 0.82 | 0.63 | 1.07 | 0.140 | 0.823 | 0.81 | 0.58 | 1.12 | 0.201 | 0.81 | 0.46 | 1.43 | 0.471 | 0.00 | -0.11 | 0.12 | 0.966 |
| NPY | OID2138 | Oncology | NPY | P01303 | 7 | 24284188 | 24291862 | 5 | 0 | 0 | 0 | 0 | 0.74 | 0.46 | 1.17 | 0.200 | 0.879 | 0.23 | 0.99 | 150.52 | 0.74 | 0.46 | 1.18 | 0.205 | 0.842 | 0.74 | 0.44 | 1.23 | 0.245 | 0.67 | 0.25 | 1.82 | 0.433 | 0.01 | -0.12 | 0.15 | 0.834 |
| PDGFA | OID2031 | Cardiom | PDGFA | P04085 | 7 | 497258 | 520296 | 1 | 0 | 0 | 0 | 0 | 4.15 | 0.62 | 27.80 | 0.143 | 0.879 | - | - | 35.09 | 4.15 | 0.54 | 32.00 | 0.172 | 0.823 | - | - | - | - | - | - | - | - | - | - | - | - |
| LPL | OID2018 | Cardiom | LPL | P06858 | 8 | 19901717 | 19967259 | 7 | 0 | 0 | 0 | 0 | 1.29 | 0.90 | 1.84 | 0.166 | 0.879 | 2.03 | 0.92 | 214.25 | 1.29 | 0.90 | 1.84 | 0.169 | 0.823 | 1.37 | 0.91 | 2.06 | 0.133 | 1.24 | 0.63 | 2.42 | 0.539 | 0.01 | -0.08 | 0.09 | 0.889 |
| LY6D | OID2070 | Inflamma | LY6D | Q14210 | 8 | 142784882 | 142786539 | 20 | 0 | 0 | 0 | 0 | 1.19 | 0.94 | 1.51 | 0.138 | 0.879 | 24.66 | 0.17 | 193.90 | 1.19 | 0.97 | 1.47 | 0.091 | 0.806 | 1.01 | 0.77 | 1.33 | 0.939 | 0.99 | 0.61 | 1.60 | 0.953 | 0.05 | -0.06 | 0.15 | 0.374 |
| ABO | OID3067 | Inflamma | ABO | P16442 | 9 | 133233278 | 133276024 | 38 | 0 | 0 | 0 | 0 | 1.07 | 0.97 | 1.19 | 0.176 | 0.879 | 32.41 | 0.68 | 370.13 | 1.07 | 0.97 | 1.19 | 0.176 | 0.823 | 1.17 | 1.01 | 1.34 | 0.035 | 1.09 | 0.93 | 1.29 | 0.297 | -0.01 | -0.05 | 0.04 | 0.800 |
| COL5A1 | OID3058 | Inflamma | COL5A1 | P20908 | 9 | 134641803 | 134844843 | 8 | 0 | 0 | 0 | 0 | 0.79 | 0.56 | 1.11 | 0.168 | 0.879 | 7.99 | 0.33 | 193.46 | 0.79 | 0.57 | 1.08 | 0.141 | 0.823 | 0.78 | 0.54 | 1.14 | 0.206 | 0.76 | 0.35 | 1.65 | 0.483 | 0.01 | -0.10 | 0.11 | 0.910 |
| GRHPR | OID3028 | Cardiom | GRHPR | Q9UBQ7 | 9 | 37422666 | 37436990 | 8 | 0 | 0 | 0 | 0 | 0.78 | 0.54 | 1.12 | 0.176 | 0.879 | 8.30 | 0.31 | 187.00 | 0.78 | 0.56 | 1.09 | 0.141 | 0.823 | 0.79 | 0.55 | 1.13 | 0.198 | 0.94 | 0.49 | 1.80 | 0.841 | -0.03 | -0.13 | 0.07 | 0.504 |
| LCN2 | OID2039 | Cardiom | LCN2 | P80188 | 9 | 128149071 | 128153453 | 1 | 0 | 0 | 0 | 0 | 0.44 | 0.13 | 1.54 | 0.199 | 0.879 | - | - | 86.58 | 0.44 | 0.12 | 1.59 | 0.211 | 0.849 | - | - | - | - | - | - | - | - | - | - | - | - |
| MEGF9 | OID2028 | Cardiom | MEGF9 | Q9H1U4 | 9 | 120600811 | 120714470 | 10 | 0 | 0 | 0 | 0 | 1.24 | 0.94 | 1.63 | 0.128 | 0.879 | 7.39 | 0.60 | 221.76 | 1.24 | 0.94 | 1.63 | 0.129 | 0.812 | 1.22 | 0.90 | 1.65 | 0.198 | 1.02 | 0.65 | 1.60 | 0.929 | 0.04 | -0.03 | 0.11 | 0.289 |
| PAPPA | OID2042 | Inflamma | PAPPA | Q13219 | 9 | 116153791 | 116402321 | 2 | 0 | 0 | 0 | 0 | 2.59 | 0.72 | 9.34 | 0.146 | 0.879 | 0.03 | 0.87 | 46.50 | 2.59 | 0.68 | 9.89 | 0.163 | 0.823 | - | - | - | - | - | - | - | - | - | - | - | - |
| UBAC1 | OID2133 | Oncology | UBAC1 | Q9BSL1 | 9 | 135932969 | 135961373 | 2 | 0 | 0 | 0 | 0 | 0.44 | 0.15 | 1.30 | 0.139 | 0.879 | 0.00 | 0.96 | 63.56 | 0.44 | 0.15 | 1.35 | 0.152 | 0.823 | - | - | - | - | - | - | - | - | - | - | - | - |
| ECHDC3 | OID3029 | Cardiom | ECHDC3 | Q96DC8 | 10 | 11742366 | 11764070 | 20 | 0 | 0 | 0 | 0 | 0.90 | 0.77 | 1.06 | 0.199 | 0.879 | 16.67 | 0.61 | 291.96 | 0.90 | 0.77 | 1.06 | 0.200 | 0.836 | 0.93 | 0.77 | 1.14 | 0.499 | 0.93 | 0.75 | 1.16 | 0.516 | -0.01 | -0.06 | 0.04 | 0.659 |
| LHPP | OID2074 | Inflamma | LHPP | Q9H008 | 10 | 124461823 | 124617888 | 17 | 0 | 0 | 0 | 0 | 1.15 | 0.93 | 1.43 | 0.194 | 0.879 | 18.90 | 0.27 | 245.17 | 1.15 | 0.95 | 1.41 | 0.158 | 0.823 | 1.05 | 0.81 | 1.35 | 0.722 | 1.18 | 0.82 | 1.71 | 0.375 | -0.01 | -0.08 | 0.06 | 0.873 |
| LYZL2 | OID3118 | Oncology | LYZL2 | Q7Z4W2 | 10 | 30611779 | 30629762 | 4 | 0 | 0 | 0 | 0 | 1.33 | 0.90 | 1.98 | 0.150 | 0.879 | 1.12 | 0.77 | 275.60 | 1.34 | 0.90 | 1.98 | 0.152 | 0.823 | 1.36 | 0.89 | 2.08 | 0.154 | 1.36 | 0.79 | 2.35 | 0.266 | -0.01 | -0.10 | 0.09 | 0.917 |
| PNLIPRP | OID2077 | Inflamma | PNLIPRP | P54317 | 10 | 116620953 | 116645143 | 35 | 0 | 0 | 0 | 0 | 0.92 | 0.83 | 1.03 | 0.144 | 0.879 | 24.76 | 0.88 | 388.07 | 0.92 | 0.83 | 1.03 | 0.145 | 0.823 | 0.95 | 0.82 | 1.10 | 0.467 | 0.98 | 0.82 | 1.17 | 0.825 | -0.02 | -0.07 | 0.03 | 0.410 |
| VTI1A | OID3064 | Inflamma | VTI1A | Q96AJ9 | 10 | 112446998 | 112818744 | 1 | 0 | 0 | 0 | 0 | 0.33 | 0.07 | 1.69 | 0.186 | 0.879 | - | - | 55.03 | 0.33 | 0.06 | 1.82 | 0.205 | 0.842 | - | - | - | - | - | - | - | - | - | - | - | - |
| AHNAK | OID3026 | Cardiom | AHNAK | Q09666 | 11 | 62433542 | 62556235 | 2 | 0 | 0 | 0 | 0 | 1.58 | 0.84 | 2.98 | 0.153 | 0.879 | 0.31 | 0.58 | 239.64 | 1.59 | 0.84 | 3.00 | 0.156 | 0.823 | - | - | - | - | - | - | - | - | - | - | - | - |
| APOA1 | OID3076 | Inflamma | APOA1 | P02647 | 11 | 116835751 | 116837622 | 1 | 0 | 0 | 0 | 0 | 0.31 | 0.06 | 1.48 | 0.141 | 0.879 | - | - | 72.35 | 0.31 | 0.06 | 1.57 | 0.156 | 0.823 | - | - | - | - | - | - | - | - | - | - | - | - |
| B4GAT1 | OID2112 | Neurolog | B4GAT1 | O43505 | 11 | 66345374 | 66347629 | 10 | 0 | 0 | 0 | 0 | 0.81 | 0.58 | 1.12 | 0.199 | 0.879 | 13.53 | 0.14 | 232.92 | 0.81 | 0.62 | 1.05 | 0.114 | 0.809 | 0.85 | 0.62 | 1.16 | 0.311 | 1.10 | 0.58 | 2.09 | 0.765 | -0.07 | -0.19 | 0.05 | 0.270 |
| CALCB | OID3110 | Neurolog | CALCB | P10092 | 11 | 14904997 | 15082342 | 5 | 0 | 0 | 0 | 0 | 1.59 | 0.85 | 2.96 | 0.147 | 0.879 | 4.28 | 0.37 | 83.25 | 1.59 | 0.86 | 2.94 | 0.135 | 0.812 | 1.57 | 0.76 | 3.25 | 0.219 | 0.62 | 0.06 | 6.29 | 0.683 | 0.10 | -0.13 | 0.32 | 0.406 |
| CHRDL2 | OID2022 | Cardiom | CHRDL2 | Q6WN34 | 11 | 74696429 | 74731426 | 11 | 0 | 0 | 0 | 0 | 1.21 | 0.93 | 1.58 | 0.148 | 0.879 | 6.18 | 0.80 | 222.67 | 1.22 | 0.93 | 1.58 | 0.149 | 0.823 | 1.08 | 0.78 | 1.48 | 0.646 | 0.99 | 0.67 | 1.45 | 0.952 | 0.05 | -0.02 | 0.12 | 0.154 |
| CST6 | OID2029 | Cardiom | CST6 | Q15828 | 11 | 66012008 | 66013505 | 5 | 0 | 0 | 1 | 0 | 0.55 | 0.22 | 1.37 | 0.201 | 0.879 | 4.06 | 0.40 | 45.11 | 0.55 | 0.22 | 1.38 | 0.202 | 0.839 | 0.64 | 0.20 | 1.98 | 0.436 | 0.28 | 0.02 | 3.37 | 0.317 | 0.05 | -0.13 | 0.24 | 0.561 |
| IL18BP | OID2034 | Cardiom | IL18BP | O95998 | 11 | 71998613 | 72005715 | 3 | 0 | 0 | 1 | 0 | 2.26 | 0.71 | 7.16 | 0.165 | 0.879 | 3.28 | 0.19 | 40.30 | 2.31 | 0.70 | 7.57 | 0.169 | 0.823 | 2.73 | 0.62 | 12.06 | 0.185 | 1.49 | 0.02 | 100.94 | 0.852 | 0.05 | -0.39 | 0.49 | 0.825 |
| LSP1 | OID2053 | Inflamma | LSP1 | P33241 | 11 | 1850904 | 1892267 | 4 | 0 | 0 | 0 | 0 | 1.43 | 0.84 | 2.42 | 0.187 | 0.879 | 0.84 | 0.84 | 143.42 | 1.43 | 0.84 | 2.44 | 0.192 | 0.833 | 1.56 | 0.87 | 2.80 | 0.132 | 1.95 | 0.59 | 6.41 | 0.274 | -0.04 | -0.16 | 0.09 | 0.571 |
| LUZP2 | OID3140 | Oncology | LUZP2 | Q86TE4 | 11 | 24496970 | 25082638 | 9 | 0 | 0 | 0 | 0 | 1.32 | 0.88 | 2.00 | 0.184 | 0.879 | 0.94 | 1.00 | 102.93 | 1.32 | 0.87 | 2.02 | 0.190 | 0.833 | 1.33 | 0.79 | 2.25 | 0.279 | 1.69 | 0.52 | 5.48 | 0.384 | -0.03 | -0.18 | 0.11 | 0.666 |
| LYVE1 | OID3072 | Inflamma | LYVE1 | Q9Y5Y7 | 11 | 10556966 | 10611689 | 3 | 0 | 0 | 0 | 0 | 1.53 | 0.87 | 2.70 | 0.138 | 0.879 | 1.59 | 0.45 | 167.63 | 1.54 | 0.87 | 2.71 | 0.140 | 0.823 | 1.48 | 0.81 | 2.70 | 0.203 | 1.08 | 0.41 | 2.82 | 0.875 | 0.06 | -0.08 | 0.20 | 0.377 |
| MCAM | OID2034 | Cardiom | MCAM | P43121 | 11 | 119308529 | 119321521 | 3 | 0 | 0 | 0 | 0 | 1.53 | 0.84 | 2.78 | 0.164 | 0.879 | 1.41 | 0.49 | 175.06 | 1.53 | 0.84 | 2.80 | 0.166 | 0.823 | 1.64 | 0.88 | 3.08 | 0.119 | 1.77 | 0.62 | 5.05 | 0.283 | -0.03 | -0.16 | 0.11 | 0.717 |
| NELL1 | OID2149 | Oncology | NELL1 | Q92832 | 11 | 20669551 | 21575686 | 10 | 0 | 0 | 0 | 0 | 1.21 | 0.93 | 1.58 | 0.155 | 0.879 | 7.89 | 0.55 | 241.64 | 1.21 | 0.93 | 1.58 | 0.155 | 0.823 | 1.11 | 0.80 | 1.53 | 0.530 | 1.07 | 0.73 | 1.58 | 0.732 | 0.03 | -0.04 | 0.10 | 0.393 |
| OMP | OID3083 | Neurolog | OMP | P47874 | 11 | 77102840 | 77103331 | 3 | 0 | 0 | 0 | 0 | 1.40 | 0.84 | 2.34 | 0.198 | 0.879 | 0.72 | 0.70 | 201.92 | 1.40 | 0.84 | 2.35 | 0.202 | 0.838 | 1.47 | 0.86 | 2.51 | 0.156 | 1.66 | 0.70 | 3.94 | 0.249 | -0.04 | -0.21 | 0.13 | 0.629 |
| VSIG2 | OID3137 | Oncology | VSIG2 | Q96IQ7 | 11 | 124747474 | 124752255 | 4 | 0 | 0 | 0 | 0 | 0.73 | 0.46 | 1.16 | 0.185 | 0.879 | 3.66 | 0.30 | 247.13 | 0.73 | 0.48 | 1.11 | 0.143 | 0.823 | 0.73 | 0.45 | 1.17 | 0.190 | 3.29 | 0.41 | 26.24 | 0.260 | -0.33 | -0.78 | 0.12 | 0.146 |
| AMIGO2 | OID2137 | Oncology | AMIGO2 | Q86SJ2 | 12 | 47075707 | 47079959 | 10 | 0 | 0 | 0 | 0 | 0.75 | 0.51 | 1.09 | 0.131 | 0.879 | 7.36 | 0.60 | 110.21 | 0.75 | 0.51 | 1.09 | 0.133 | 0.812 | 0.89 | 0.55 | 1.45 | 0.637 | 0.82 | 0.32 | 2.12 | 0.688 | -0.01 | -0.12 | 0.10 | 0.822 |
| CALCOC | OID2138 | Oncology | CALCOC | Q9P1Z2 | 12 | 53708517 | 53727745 | 1 | 0 | 0 | 0 | 0 | 0.49 | 0.18 | 1.29 | 0.148 | 0.879 | - | - | 180.63 | 0.49 | 0.18 | 1.31 | 0.154 | 0.823 | - | - | - | - | - | - | - | - | - | - | - | - |
| CKAP4 | OID2061 | Inflamma | CKAP4 | Q07065 | 12 | 106237881 | 106304279 | 9 | 0 | 0 | 0 | 0 | 0.77 | 0.53 | 1.10 | 0.151 | 0.879 | 1.48 | 0.99 | 146.69 | 0.77 | 0.53 | 1.11 | 0.155 | 0.823 | 0.73 | 0.47 | 1.15 | 0.172 | 0.86 | 0.44 | 1.71 | 0.676 | -0.02 | -0.14 | 0.09 | 0.690 |
| CLEC7A | OID2063 | Inflamma | CLEC7A | Q9BXN2 | 12 | 10116777 | 10130258 | 28 | 0 | 0 | 0 | 0 | 1.11 | 0.97 | 1.27 | 0.133 | 0.879 | 25.01 | 0.57 | 308.46 | 1.11 | 0.97 | 1.28 | 0.134 | 0.812 | 1.03 | 0.86 | 1.25 | 0.721 | 1.12 | 0.91 | 1.38 | 0.285 | 0.00 | -0.05 | 0.04 | 0.914 |
| COL2A1 | OID3028 | Cardiom | COL2A1 | P02458 | 12 | 47972967 | 48004554 | 32 | 0 | 0 | 0 | 0 | 0.92 | 0.80 | 1.04 | 0.192 | 0.879 | 34.21 | 0.32 | 308.04 | 0.92 | 0.81 | 1.04 | 0.170 | 0.823 | 0.97 | 0.81 | 1.15 | 0.701 | 0.82 | 0.65 | 1.03 | 0.092 | 0.03 | -0.02 | 0.09 | 0.254 |
| DTX3 | OID2134 | Oncology | DTX3 | Q8N9I9 | 12 | 57604622 | 57609804 | 1 | 0 | 0 | 0 | 0 | 0.48 | 0.18 | 1.28 | 0.145 | 0.879 | - | - | 184.60 | 0.48 | 0.18 | 1.30 | 0.151 | 0.823 | - | - | - | - | - | - | - | - | - | - | - | - |
| LTBR | OID2076 | Inflamma | LTBR | P36941 | 12 | 6375045 | 6391571 | 8 | 0 | 0 | 0 | 0 | 0.83 | 0.63 | 1.09 | 0.175 | 0.879 | 0.89 | 1.00 | 281.27 | 0.83 | 0.63 | 1.09 | 0.178 | 0.823 | 0.83 | 0.62 | 1.12 | 0.231 | 0.84 | 0.42 | 1.67 | 0.614 | 0.00 | -0.17 | 0.16 | 0.974 |
| OLR1 | OID2018 | Cardiom | OLR1 | P78380 | 12 | 10158301 | 10172138 | 2 | 0 | 0 | 0 | 0 | 0.54 | 0.22 | 1.32 | 0.176 | 0.879 | 0.45 | 0.50 | 100.73 | 0.54 | 0.22 | 1.34 | 0.183 | 0.824 | - | - | - | - | - | - | - | - | - | - | - | - |
| PSMD9 | OID2130 | Oncology | PSMD9 | O00233 | 12 | 121888732 | 121918297 | 2 | 0 | 0 | 0 | 0 | 0.70 | 0.42 | 1.16 | 0.166 | 0.879 | 0.47 | 0.49 | 319.84 | 0.70 | 0.42 | 1.16 | 0.168 | 0.823 | - | - | - | - | - | - | - | - | - | - | - | - |
| GAS6 | OID2031 | Cardiom | GAS6 | Q14393 | 13 | 113820549 | 113864076 | 8 | 0 | 0 | 0 | 0 | 0.79 | 0.55 | 1.12 | 0.187 | 0.879 | 10.91 | 0.14 | 257.37 | 0.79 | 0.59 | 1.05 | 0.099 | 0.806 | 0.83 | 0.60 | 1.13 | 0.226 | 1.06 | 0.53 | 2.12 | 0.868 | -0.07 | -0.20 | 0.07 | 0.327 |
| GPC5 | OID2094 | Neurolog | GPC5 | P78333 | 13 | 91398621 | 92873682 | 39 | 0 | 0 | 0 | 0 | 0.92 | 0.82 | 1.04 | 0.191 | 0.879 | 41.19 | 0.33 | 292.26 | 0.92 | 0.82 | 1.04 | 0.173 | 0.823 | 0.89 | 0.76 | 1.06 | 0.184 | 0.90 | 0.74 | 1.10 | 0.301 | 0.01 | -0.04 | 0.06 | 0.767 |
| ANG | OID2040 | Cardiom | ANG | P03950 | 14 | 20684177 | 20698971 | 16 | 0 | 0 | 0 | 0 | 1.12 | 0.95 | 1.31 | 0.178 | 0.879 | 9.23 | 0.87 | 396.00 | 1.12 | 0.95 | 1.31 | 0.178 | 0.823 | 1.07 | 0.88 | 1.31 | 0.499 | 1.07 | 0.86 | 1.34 | 0.527 | 0.01 | -0.04 | 0.07 | 0.639 |
| ATXN3 | OID3114 | Neurolog | ATXN3 | P54252 | 14 | 92044496 | 92106621 | 11 | 0 | 0 | 0 | 0 | 1.22 | 0.92 | 1.63 | 0.171 | 0.879 | 3.67 | 0.96 | 183.37 | 1.22 | 0.92 | 1.64 | 0.173 | 0.823 | 1.24 | 0.88 | 1.74 | 0.222 | 1.34 | 0.81 | 2.22 | 0.248 | -0.02 | -0.09 | 0.06 | 0.652 |
| COCH | OID3030 | Neurolog | COCH | O43405 | 14 | 30874514 | 30895065 | 14 | 0 | 0 | 0 | 0 | 0.87 | 0.71 | 1.06 | 0.166 | 0.879 | 7.04 | 0.90 | 293.67 | 0.87 | 0.71 | 1.06 | 0.168 | 0.823 | 0.90 | 0.69 | 1.17 | 0.432 | 0.94 | 0.51 | 1.76 | 0.858 | -0.02 | -0.16 | 0.12 | 0.773 |
| FLRT2 | OID2094 | Neurolog | FLRT2 | O43155 | 14 | 85530144 | 85654428 | 21 | 0 | 0 | 0 | 0 | 0.87 | 0.73 | 1.04 | 0.134 | 0.879 | 15.87 | 0.72 | 235.19 | 0.87 | 0.73 | 1.04 | 0.135 | 0.812 | 0.85 | 0.68 | 1.07 | 0.173 | 0.78 | 0.55 | 1.11 | 0.170 | 0.03 | -0.04 | 0.10 | 0.473 |
| RNASE4 | OID3070 | Inflamma | RNASE4 | P34096 | 14 | 20684560 | 20701216 | 8 | 0 | 0 | 0 | 0 | 1.23 | 0.90 | 1.68 | 0.197 | 0.879 | 8.92 | 0.26 | 282.68 | 1.23 | 0.93 | 1.62 | 0.145 | 0.823 | 1.43 | 1.05 | 1.95 | 0.025 | 1.54 | 0.94 | 2.53 | 0.086 | -0.05 | -0.14 | 0.04 | 0.250 |
| SERPINA | OID2128 | Oncology | SERPINA | Q86WD7 | 14 | 94462717 | 94479689 | 21 | 0 | 0 | 0 | 0 | 1.12 | 0.95 | 1.32 | 0.193 | 0.879 | 24.04 | 0.24 | 307.90 | 1.12 | 0.96 | 1.30 | 0.153 | 0.823 | 1.07 | 0.86 | 1.34 | 0.540 | 1.24 | 0.93 | 1.66 | 0.140 | -0.03 | -0.10 | 0.04 | 0.374 |
| CA12 | OID2134 | Oncology | CA12 | O43570 | 15 | 63321378 | 63381846 | 16 | 0 | 0 | 0 | 0 | 1.23 | 0.94 | 1.60 | 0.129 | 0.879 | 17.97 | 0.26 | 171.71 | 1.23 | 0.96 | 1.56 | 0.096 | 0.806 | 1.23 | 0.90 | 1.69 | 0.193 | 1.58 | 0.88 | 2.85 | 0.125 | -0.06 | -0.18 | 0.06 | 0.339 |
| FURIN | OID2151 | Oncology | FURIN | P09958 | 15 | 90868588 | 90883564 | 2 | 0 | 0 | 0 | 0 | 1.41 | 0.86 | 2.32 | 0.176 | 0.879 | 0.59 | 0.44 | 329.13 | 1.41 | 0.85 | 2.33 | 0.178 | 0.823 | - | - | - | - | - | - | - | - | - | - | - | - |
| IGDCC4 | OID3040 | Oncology | IGDCC4 | Q8TDY8 | 15 | 65381484 | 65422947 | 17 | 0 | 0 | 0 | 0 | 1.18 | 0.92 | 1.51 | 0.200 | 0.879 | 18.86 | 0.28 | 189.87 | 1.18 | 0.94 | 1.48 | 0.163 | 0.823 | 1.17 | 0.86 | 1.58 | 0.313 | 1.14 | 0.76 | 1.71 | 0.529 | 0.01 | -0.06 | 0.08 | 0.848 |
| ADGRG1 | OID2129 | Oncology | ADGRG1 | Q9Y653 | 16 | 57610652 | 57665580 | 5 | 0 | 0 | 1 | 0 | 0.58 | 0.25 | 1.32 | 0.192 | 0.879 | 2.88 | 0.58 | 48.99 | 0.57 | 0.25 | 1.34 | 0.200 | 0.836 | 0.40 | 0.15 | 1.11 | 0.079 | 0.16 | 0.02 | 1.15 | 0.069 | 0.10 | -0.04 | 0.24 | 0.160 |
| APRT | OID2092 | Neurolog | APRT | P07741 | 16 | 88809339 | 88811937 | 2 | 0 | 0 | 0 | 0 | 2.12 | 0.73 | 6.18 | 0.167 | 0.879 | 5.50 | 0.02 | 69.04 | 2.19 | 0.74 | 6.46 | 0.155 | 0.823 | - | - | - | - | - | - | - | - | - | - | - | - |
| AXIN1 | OID2058 | Inflamma | AXIN1 | O15169 | 16 | 287440 | 352723 | 1 | 0 | 0 | 0 | 0 | 4.61 | 0.64 | 33.40 | 0.130 | 0.879 | - | - | 41.60 | 4.61 | 0.56 | 37.90 | 0.155 | 0.823 | - | - | - | - | - | - | - | - | - | - | - | - |
| CTRL | OID3115 | Neurolog | CTRL | P40313 | 16 | 67927640 | 67932365 | 2 | 0 | 0 | 0 | 0 | 0.41 | 0.12 | 1.34 | 0.140 | 0.879 | 0.52 | 0.47 | 57.45 | 0.41 | 0.12 | 1.39 | 0.152 | 0.823 | - | - | - | - | - | - | - | - | - | - | - | - |
| CX3CL1 | OID2097 | Neurolog | CX3CL1 | P78423 | 16 | 57372477 | 57385044 | 6 | 0 | 0 | 0 | 0 | 0.73 | 0.47 | 1.14 | 0.167 | 0.879 | 6.36 | 0.27 | 178.83 | 0.73 | 0.49 | 1.08 | 0.119 | 0.810 | 0.66 | 0.44 | 1.00 | 0.052 | 0.47 | 0.21 | 1.06 | 0.069 | 0.08 | -0.05 | 0.22 | 0.213 |
| HBZ | OID3030 | Cardiom | HBZ | P02008 | 16 | 142728 | 154503 | 25 | 0 | 0 | 0 | 0 | 1.07 | 0.96 | 1.20 | 0.198 | 0.879 | 25.82 | 0.36 | 563.24 | 1.07 | 0.97 | 1.19 | 0.182 | 0.824 | 0.98 | 0.84 | 1.14 | 0.768 | 0.99 | 0.80 | 1.23 | 0.945 | 0.03 | -0.05 | 0.11 | 0.410 |
| IL17C | OID2047 | Inflamma | IL17C | Q9P0M4 | 16 | 88638572 | 88640468 | 2 | 0 | 0 | 0 | 0 | 0.61 | 0.29 | 1.26 | 0.181 | 0.879 | 0.16 | 0.69 | 144.83 | 0.61 | 0.29 | 1.28 | 0.187 | 0.830 | - | - | - | - | - | - | - | - | - | - | - | - |
| PRSS53 | OID3109 | Neurolog | PRSS53 | Q2L4Q9 | 16 | 31083437 | 31089628 | 49 | 0 | 0 | 0 | 0 | 0.94 | 0.85 | 1.03 | 0.182 | 0.879 | 53.02 | 0.29 | 394.46 | 0.94 | 0.86 | 1.03 | 0.161 | 0.823 | 0.89 | 0.78 | 1.02 | 0.082 | 0.95 | 0.77 | 1.17 | 0.630 | -0.01 | -0.08 | 0.07 | 0.891 |
| SEZ6L2 | OID2128 | Oncology | SEZ6L2 | Q6UXD5 | 16 | 29871159 | 29899550 | 6 | 0 | 0 | 0 | 0 | 1.37 | 0.88 | 2.11 | 0.159 | 0.879 | 1.34 | 0.93 | 170.98 | 1.37 | 0.88 | 2.12 | 0.162 | 0.823 | 1.55 | 0.93 | 2.58 | 0.091 | 1.57 | 0.71 | 3.45 | 0.262 | -0.02 | -0.14 | 0.09 | 0.681 |
| TPPP3 | OID2084 | Neurolog | TPPP3 | Q9BW30 | 16 | 67389809 | 67393518 | 1 | 0 | 0 | 0 | 0 | 2.96 | 0.60 | 14.65 | 0.184 | 0.879 | - | - | 63.94 | 2.96 | 0.56 | 15.56 | 0.201 | 0.836 | - | - | - | - | - | - | - | - | - | - | - | - |
| ASGR2 | OID3067 | Inflamma | ASGR2 | P07307 | 17 | 7101322 | 7115700 | 9 | 0 | 0 | 0 | 0 | 1.27 | 0.91 | 1.76 | 0.165 | 0.879 | 14.00 | 0.08 | 265.96 | 1.27 | 0.99 | 1.63 | 0.065 | 0.744 | 1.26 | 0.95 | 1.68 | 0.115 | 1.17 | 0.60 | 2.26 | 0.644 | 0.02 | -0.11 | 0.15 | 0.780 |
| CCL2 | OID2100 | Neurolog | CCL2 | P13500 | 17 | 34255218 | 34257203 | 1 | 0 | 0 | 0 | 0 | 0.34 | 0.07 | 1.73 | 0.192 | 0.879 | - | - | 64.73 | 0.34 | 0.06 | 1.83 | 0.208 | 0.849 | - | - | - | - | - | - | - | - | - | - | - | - |
| CD300E | OID2141 | Oncology | CD300E | Q496F6 | 17 | 74609885 | 74623738 | 16 | 0 | 0 | 0 | 0 | 1.15 | 0.93 | 1.43 | 0.190 | 0.879 | 11.08 | 0.75 | 229.55 | 1.16 | 0.93 | 1.43 | 0.192 | 0.833 | 1.20 | 0.92 | 1.56 | 0.183 | 1.29 | 0.83 | 2.03 | 0.261 | -0.03 | -0.12 | 0.07 | 0.572 |
| ICAM2 | OID2024 | Cardiom | ICAM2 | P13598 | 17 | 64002594 | 64020634 | 1 | 0 | 0 | 0 | 0 | 2.91 | 0.59 | 14.36 | 0.189 | 0.879 | - | - | 106.68 | 2.91 | 0.57 | 14.88 | 0.199 | 0.836 | - | - | - | - | - | - | - | - | - | - | - | - |
| LRRC37 | OID3144 | Oncology | LRRC37 | A6NM11 | 17 | 46511508 | 46556910 | 35 | 0 | 0 | 0 | 0 | 0.95 | 0.88 | 1.02 | 0.143 | 0.879 | 37.40 | 0.32 | 864.10 | 0.95 | 0.88 | 1.02 | 0.124 | 0.810 | 0.99 | 0.91 | 1.09 | 0.889 | 1.01 | 0.88 | 1.15 | 0.921 | -0.04 | -0.11 | 0.03 | 0.263 |
| PECAM1 | OID2113 | Neurolog | PECAM1 | P16284 | 17 | 64319415 | 64413776 | 1 | 0 | 0 | 0 | 0 | 3.90 | 0.50 | 30.56 | 0.195 | 0.879 | - | - | 40.48 | 3.90 | 0.44 | 34.47 | 0.221 | 0.857 | - | - | - | - | - | - | - | - | - | - | - | - |
| PYY | OID3015 | Cardiom | PYY | P10082 | 17 | 43952733 | 44004469 | 4 | 0 | 0 | 0 | 0 | 0.77 | 0.52 | 1.13 | 0.177 | 0.879 | 1.40 | 0.71 | 255.87 | 0.77 | 0.52 | 1.13 | 0.179 | 0.823 | 0.78 | 0.52 | 1.17 | 0.236 | 0.64 | 0.20 | 2.05 | 0.453 | 0.04 | -0.20 | 0.28 | 0.749 |
| SAT2 | OID3130 | Oncology | SAT2 | Q96F10 | 17 | 7626234 | 7627876 | 18 | 0 | 0 | 0 | 0 | 1.10 | 0.95 | 1.28 | 0.199 | 0.879 | 16.21 | 0.51 | 375.09 | 1.10 | 0.95 | 1.28 | 0.199 | 0.836 | 1.16 | 0.93 | 1.44 | 0.198 | 1.11 | 0.90 | 1.37 | 0.319 | 0.00 | -0.06 | 0.05 | 0.891 |
| SHPK | OID3129 | Oncology | SHPK | Q9UHJ6 | 17 | 3608240 | 3636250 | 1 | 0 | 0 | 0 | 0 | 0.45 | 0.15 | 1.42 | 0.174 | 0.879 | - | - | 106.46 | 0.45 | 0.14 | 1.45 | 0.183 | 0.824 | - | - | - | - | - | - | - | - | - | - | - | - |
| TRIM25 | OID3148 | Oncology | TRIM25 | Q14258 | 17 | 56836387 | 56914080 | 1 | 0 | 0 | 0 | 0 | 2.19 | 0.76 | 6.32 | 0.146 | 0.879 | - | - | 142.96 | 2.19 | 0.75 | 6.44 | 0.153 | 0.823 | - | - | - | - | - | - | - | - | - | - | - | - |
| WFIKKN | OID2078 | Inflamma | WFIKKN | Q8TEU8 | 17 | 50834650 | 50842353 | 28 | 0 | 0 | 0 | 0 | 0.88 | 0.73 | 1.05 | 0.154 | 0.879 | 23.77 | 0.64 | 175.50 | 0.88 | 0.73 | 1.05 | 0.154 | 0.823 | 0.83 | 0.65 | 1.07 | 0.156 | 0.69 | 0.49 | 0.96 | 0.026 | 0.05 | -0.01 | 0.11 | 0.084 |
| CD226 | OID3055 | Inflamma | CD226 | Q15762 | 18 | 69831158 | 69961803 | 8 | 0 | 0 | 0 | 0 | 1.26 | 0.92 | 1.73 | 0.146 | 0.879 | 5.32 | 0.62 | 190.24 | 1.26 | 0.92 | 1.73 | 0.148 | 0.823 | 1.21 | 0.86 | 1.71 | 0.267 | 1.29 | 0.77 | 2.16 | 0.336 | 0.00 | -0.09 | 0.08 | 0.920 |
| DSC2 | OID2107 | Neurolog | DSC2 | Q02487 | 18 | 31058840 | 31102522 | 6 | 0 | 0 | 0 | 0 | 1.26 | 0.90 | 1.76 | 0.186 | 0.879 | 1.98 | 0.85 | 235.45 | 1.26 | 0.89 | 1.77 | 0.188 | 0.830 | 1.20 | 0.83 | 1.74 | 0.342 | 0.80 | 0.37 | 1.73 | 0.579 | 0.11 | -0.06 | 0.29 | 0.204 |
| SERPINF | OID2063 | Inflamma | SERPINF | P50452 | 18 | 63970029 | 64019779 | 20 | 0 | 0 | 0 | 0 | 1.08 | 0.96 | 1.20 | 0.188 | 0.879 | 19.09 | 0.45 | 653.64 | 1.08 | 0.96 | 1.20 | 0.188 | 0.830 | 1.11 | 0.98 | 1.26 | 0.111 | 1.16 | 0.99 | 1.37 | 0.071 | -0.03 | -0.08 | 0.02 | 0.214 |
| BST2 | OID2102 | Neurolog | BST2 | Q10589 | 19 | 17402939 | 17405630 | 4 | 0 | 0 | 0 | 0 | 0.62 | 0.32 | 1.23 | 0.173 | 0.879 | 4.47 | 0.21 | 134.26 | 0.62 | 0.36 | 1.09 | 0.096 | 0.806 | 0.63 | 0.33 | 1.19 | 0.152 | 1.14 | 0.49 | 2.66 | 0.758 | -0.10 | -0.20 | 0.01 | 0.064 |
| CD33 | OID2147 | Neurolog | CD33 | P20138 | 19 | 51225064 | 51243860 | 31 | 0 | 0 | 0 | 0 | 1.09 | 0.96 | 1.24 | 0.170 | 0.879 | 40.09 | 0.10 | 426.53 | 1.09 | 0.98 | 1.22 | 0.112 | 0.809 | 1.07 | 0.91 | 1.26 | 0.406 | 1.24 | 1.03 | 1.50 | 0.025 | -0.05 | -0.10 | 0.01 | 0.083 |
| CEACAM | OID3130 | Oncology | CEACAM | Q6UY09 | 19 | 44501677 | 44529788 | 3 | 0 | 0 | 0 | 0 | 1.54 | 0.88 | 2.72 | 0.133 | 0.879 | 2.24 | 0.33 | 171.56 | 1.55 | 0.87 | 2.74 | 0.134 | 0.812 | 1.55 | 0.83 | 2.87 | 0.167 | 1.11 | 0.38 | 3.20 | 0.849 | 0.06 | -0.09 | 0.22 | 0.432 |
| KLK3 | OID3036 | Cardiom | KLK3 | P07288 | 19 | 50854915 | 50860764 | 3 | 0 | 0 | 1 | 0 | 2.83 | 0.72 | 11.20 | 0.138 | 0.879 | 1.23 | 0.54 | 76.82 | 2.85 | 0.70 | 11.63 | 0.144 | 0.823 | 2.77 | 0.59 | 12.96 | 0.195 | 0.81 | 0.02 | 39.57 | 0.916 | 0.10 | -0.20 | 0.41 | 0.500 |
| KLK6 | OID2152 | Oncology | KLK6 | Q92876 | 19 | 50958631 | 50969673 | 2 | 0 | 0 | 0 | 0 | 1.40 | 0.85 | 2.31 | 0.184 | 0.879 | 2.82 | 0.09 | 317.95 | 1.40 | 0.85 | 2.31 | 0.184 | 0.824 | - | - | - | - | - | - | - | - | - | - | - | - |
| LGALS4 | OID2074 | Inflamma | LGALS4 | P56470 | 19 | 38801671 | 38812945 | 3 | 0 | 0 | 1 | 0 | 1.66 | 0.86 | 3.19 | 0.132 | 0.879 | 1.57 | 0.46 | 137.08 | 1.66 | 0.85 | 3.22 | 0.135 | 0.812 | 1.54 | 0.75 | 3.19 | 0.242 | 3.21 | 0.94 | 10.96 | 0.062 | -0.08 | -0.20 | 0.04 | 0.211 |
| LILRB5 | OID2032 | Cardiom | LILRB5 | O75023 | 19 | 54249421 | 54257301 | 37 | 0 | 0 | 0 | 0 | 0.94 | 0.86 | 1.03 | 0.196 | 0.879 | 33.90 | 0.57 | 508.22 | 0.94 | 0.86 | 1.03 | 0.197 | 0.836 | 0.97 | 0.85 | 1.10 | 0.599 | 0.96 | 0.84 | 1.10 | 0.563 | -0.01 | -0.05 | 0.04 | 0.709 |
| LRG1 | OID3075 | Inflamma | LRG1 | P02750 | 19 | 4536402 | 4540036 | 22 | 0 | 0 | 0 | 0 | 1.22 | 0.93 | 1.61 | 0.152 | 0.879 | 29.77 | 0.10 | 143.73 | 1.22 | 0.97 | 1.54 | 0.088 | 0.806 | 1.35 | 0.99 | 1.83 | 0.058 | 1.73 | 1.06 | 2.82 | 0.028 | -0.06 | -0.14 | 0.01 | 0.099 |
| LRRC25 | OID2133 | Oncology | LRRC25 | Q8N386 | 19 | 18391137 | 18397622 | 13 | 0 | 0 | 0 | 0 | 1.22 | 0.92 | 1.60 | 0.163 | 0.879 | 14.82 | 0.25 | 211.72 | 1.22 | 0.95 | 1.56 | 0.120 | 0.810 | 1.08 | 0.82 | 1.44 | 0.576 | 0.94 | 0.55 | 1.60 | 0.812 | 0.07 | -0.05 | 0.18 | 0.263 |
| PSG1 | OID2091 | Neurolog | PSG1 | P11464 | 19 | 42866464 | 42879822 | 51 | 0 | 0 | 0 | 0 | 1.07 | 0.97 | 1.19 | 0.180 | 0.879 | 51.11 | 0.43 | 313.28 | 1.07 | 0.97 | 1.19 | 0.175 | 0.823 | 1.09 | 0.94 | 1.27 | 0.241 | 1.04 | 0.87 | 1.25 | 0.672 | 0.01 | -0.05 | 0.07 | 0.697 |
| TBC1D1 | OID2084 | Neurolog | TBC1D1 | Q9HA65 | 19 | 49877666 | 49888750 | 11 | 0 | 0 | 0 | 0 | 1.22 | 0.92 | 1.61 | 0.176 | 0.879 | 10.30 | 0.41 | 188.41 | 1.22 | 0.92 | 1.61 | 0.171 | 0.823 | 1.25 | 0.92 | 1.70 | 0.159 | 1.35 | 0.82 | 2.22 | 0.242 | -0.02 | -0.11 | 0.06 | 0.616 |
| VSTM1 | OID2085 | Neurolog | VSTM1 | Q6UX27 | 19 | 54040825 | 54063953 | 26 | 0 | 0 | 0 | 0 | 0.91 | 0.80 | 1.05 | 0.191 | 0.879 | 27.67 | 0.32 | 335.99 | 0.91 | 0.80 | 1.04 | 0.169 | 0.823 | 0.92 | 0.77 | 1.09 | 0.326 | 0.85 | 0.67 | 1.08 | 0.186 | 0.02 | -0.04 | 0.09 | 0.482 |
| ISM1 | OID2053 | Inflamma | ISM1 | B1AKI9 | 20 | 13221274 | 13300651 | 10 | 0 | 0 | 0 | 0 | 1.27 | 0.89 | 1.80 | 0.187 | 0.879 | 11.77 | 0.23 | 179.75 | 1.27 | 0.93 | 1.73 | 0.131 | 0.812 | 1.04 | 0.67 | 1.60 | 0.863 | 1.22 | 0.53 | 2.82 | 0.634 | 0.01 | -0.11 | 0.12 | 0.930 |
| PLTP | OID2023 | Cardiom | PLTP | P55058 | 20 | 45898621 | 45912155 | 14 | 0 | 0 | 0 | 0 | 0.87 | 0.70 | 1.08 | 0.200 | 0.879 | 11.95 | 0.53 | 258.52 | 0.87 | 0.70 | 1.08 | 0.201 | 0.836 | 0.86 | 0.68 | 1.09 | 0.217 | 0.86 | 0.61 | 1.20 | 0.371 | 0.00 | -0.07 | 0.08 | 0.914 |
| WFDC2 | OID2150 | Oncology | WFDC2 | Q14508 | 20 | 45469753 | 45481532 | 2 | 0 | 0 | 0 | 0 | 0.51 | 0.20 | 1.29 | 0.154 | 0.879 | 0.51 | 0.48 | 102.44 | 0.51 | 0.20 | 1.31 | 0.160 | 0.823 | - | - | - | - | - | - | - | - | - | - | - | - |
| IFNAR1 | OID3149 | Oncology | IFNAR1 | P17181 | 21 | 33324387 | 33359864 | 19 | 0 | 0 | 0 | 0 | 0.87 | 0.72 | 1.06 | 0.165 | 0.879 | 9.03 | 0.96 | 231.22 | 0.87 | 0.72 | 1.06 | 0.166 | 0.823 | 0.91 | 0.72 | 1.14 | 0.404 | 0.94 | 0.69 | 1.28 | 0.690 | -0.02 | -0.08 | 0.05 | 0.581 |
| ADA2 | OID2039 | Cardiom | ADA2 | Q9NZK5 | 22 | 17178790 | 17258235 | 6 | 0 | 0 | 0 | 0 | 1.27 | 0.92 | 1.76 | 0.144 | 0.879 | 5.25 | 0.39 | 278.73 | 1.27 | 0.93 | 1.75 | 0.134 | 0.812 | 1.36 | 0.94 | 1.99 | 0.106 | 2.36 | 0.90 | 6.19 | 0.082 | -0.10 | -0.25 | 0.05 | 0.186 |
| C1QTNF | OID3104 | Neurolog | C1QTNF | Q9BXI9 | 22 | 37180166 | 37199385 | 1 | 0 | 0 | 0 | 0 | 3.12 | 0.66 | 14.70 | 0.149 | 0.879 | - | - | 76.44 | 3.12 | 0.63 | 15.48 | 0.163 | 0.823 | - | - | - | - | - | - | - | - | - | - | - | - |
| MB | OID2026 | Cardiom | MB | P02144 | 22 | 35606764 | 35637951 | 1 | 0 | 0 | 0 | 0 | 0.14 | 0.01 | 1.93 | 0.143 | 0.879 | - | - | 31.90 | 0.14 | 0.01 | 2.39 | 0.175 | 0.823 | - | - | - | - | - | - | - | - | - | - | - | - |
| TST | OID2086 | Neurolog | TST | Q16762 | 22 | 37010859 | 37020183 | 3 | 0 | 0 | 0 | 0 | 0.59 | 0.29 | 1.20 | 0.145 | 0.879 | 0.82 | 0.66 | 100.38 | 0.59 | 0.28 | 1.21 | 0.151 | 0.823 | 0.60 | 0.28 | 1.30 | 0.197 | 1.28 | 0.10 | 16.24 | 0.848 | -0.08 | -0.32 | 0.16 | 0.530 |
| SLA2 | OID3101 | Neurolog | SLA2 | Q9H6Q3 | 20 | 36612318 | 36646196 | 1 | 0 | 0 | 0 | 0 | 0.32 | 0.06 | 1.84 | 0.201 | 0.879 | - | - | 51.75 | 0.32 | 0.05 | 1.99 | 0.222 | 0.857 | - | - | - | - | - | - | - | - | - | - | - | - |
| CD58 | OID2071 | Inflamma | CD58 | P19256 | 1 | 116514534 | 116571039 | 4 | 0 | 0 | 0 | 0 | 1.46 | 0.81 | 2.62 | 0.203 | 0.884 | 3.57 | 0.31 | 161.47 | 1.46 | 0.85 | 2.51 | 0.166 | 0.823 | 1.64 | 0.92 | 2.92 | 0.090 | 1.31 | 0.45 | 3.83 | 0.624 | 0.02 | -0.14 | 0.18 | 0.790 |
| FCRL5 | OID2111 | Neurolog | FCRL5 | Q96RD9 | 1 | 157513377 | 157552515 | 12 | 0 | 0 | 0 | 0 | 1.15 | 0.93 | 1.41 | 0.206 | 0.885 | 12.61 | 0.32 | 361.98 | 1.15 | 0.94 | 1.40 | 0.175 | 0.823 | 1.19 | 0.93 | 1.54 | 0.170 | 1.33 | 0.97 | 1.84 | 0.080 | -0.05 | -0.14 | 0.03 | 0.229 |
| MAP2 | OID3113 | Neurolog | MAP2 | P11137 | 2 | 209424047 | 209734147 | 4 | 0 | 0 | 0 | 0 | 0.59 | 0.26 | 1.34 | 0.207 | 0.885 | 4.22 | 0.24 | 89.86 | 0.58 | 0.29 | 1.18 | 0.135 | 0.812 | 0.54 | 0.24 | 1.23 | 0.145 | 1.07 | 0.39 | 2.91 | 0.895 | -0.11 | -0.24 | 0.02 | 0.101 |
| SERPINI | OID3150 | Oncology | SERPINI | O75830 | 3 | 167441914 | 167478972 | 17 | 0 | 0 | 0 | 0 | 1.17 | 0.92 | 1.48 | 0.207 | 0.885 | 11.67 | 0.77 | 202.43 | 1.17 | 0.92 | 1.48 | 0.208 | 0.849 | 1.09 | 0.82 | 1.45 | 0.572 | 0.93 | 0.63 | 1.36 | 0.698 | 0.07 | -0.02 | 0.16 | 0.135 |
| LYAR | OID2126 | Oncology | LYAR | Q9NX58 | 4 | 4267701 | 4290154 | 1 | 0 | 0 | 0 | 0 | 3.12 | 0.53 | 18.35 | 0.207 | 0.885 | - | - | 47.86 | 3.12 | 0.49 | 20.00 | 0.229 | 0.859 | - | - | - | - | - | - | - | - | - | - | - | - |
| SLC39A1 | OID2085 | Neurolog | SLC39A1 | Q15043 | 8 | 22367278 | 22434129 | 1 | 0 | 0 | 0 | 0 | 0.53 | 0.19 | 1.42 | 0.205 | 0.885 | - | - | 148.23 | 0.53 | 0.19 | 1.44 | 0.212 | 0.851 | - | - | - | - | - | - | - | - | - | - | - | - |
| NUDT2 | OID2130 | Oncology | NUDT2 | P50583 | 9 | 34329506 | 34343711 | 12 | 0 | 0 | 0 | 0 | 0.85 | 0.65 | 1.10 | 0.208 | 0.885 | 8.70 | 0.65 | 205.92 | 0.85 | 0.65 | 1.10 | 0.209 | 0.849 | 0.73 | 0.50 | 1.06 | 0.097 | 0.77 | 0.45 | 1.35 | 0.366 | 0.02 | -0.08 | 0.12 | 0.722 |
| PRG2 | OID3038 | Cardiom | PRG2 | P13727 | 11 | 57386780 | 57390650 | 1 | 0 | 0 | 0 | 0 | 1.85 | 0.71 | 4.81 | 0.205 | 0.885 | - | - | 173.04 | 1.85 | 0.71 | 4.87 | 0.211 | 0.849 | - | - | - | - | - | - | - | - | - | - | - | - |
| NEO1 | OID3106 | Neurolog | NEO1 | Q92859 | 15 | 73051710 | 73305205 | 1 | 0 | 0 | 0 | 0 | 0.39 | 0.09 | 1.67 | 0.206 | 0.885 | - | - | 80.71 | 0.39 | 0.09 | 1.75 | 0.219 | 0.857 | - | - | - | - | - | - | - | - | - | - | - | - |
| TNFRSF1 | OID2064 | Inflamma | TNFRSF1 | Q9Y6Q6 | 18 | 62325287 | 62391288 | 27 | 0 | 0 | 0 | 0 | 1.12 | 0.94 | 1.34 | 0.205 | 0.885 | 30.84 | 0.23 | 230.65 | 1.12 | 0.95 | 1.32 | 0.167 | 0.823 | 0.99 | 0.78 | 1.26 | 0.919 | 1.02 | 0.71 | 1.47 | 0.904 | 0.02 | -0.06 | 0.10 | 0.569 |
| OXT | OID2104 | Neurolog | OXT | P01178 | 20 | 3071620 | 3072517 | 23 | 0 | 0 | 0 | 0 | 1.12 | 0.94 | 1.33 | 0.207 | 0.885 | 18.27 | 0.69 | 229.26 | 1.12 | 0.94 | 1.33 | 0.207 | 0.849 | 1.23 | 0.98 | 1.55 | 0.079 | 1.40 | 1.04 | 1.88 | 0.026 | -0.06 | -0.13 | 0.00 | 0.066 |
| SV2A | OID3096 | Neurolog | SV2A | Q7L0J3 | 1 | 149903318 | 149917844 | 2 | 0 | 0 | 0 | 0 | 1.41 | 0.82 | 2.41 | 0.210 | 0.885 | 0.16 | 0.69 | 271.87 | 1.41 | 0.82 | 2.42 | 0.213 | 0.852 | - | - | - | - | - | - | - | - | - | - | - | - |
| CDCP1 | OID2094 | Neurolog | CDCP1 | Q9H5V8 | 3 | 45082277 | 45146422 | 14 | 0 | 0 | 0 | 0 | 0.83 | 0.61 | 1.11 | 0.210 | 0.885 | 14.37 | 0.35 | 174.62 | 0.83 | 0.62 | 1.10 | 0.188 | 0.830 | 0.84 | 0.59 | 1.19 | 0.320 | 1.09 | 0.51 | 2.31 | 0.823 | -0.06 | -0.20 | 0.09 | 0.431 |
| FGF5 | OID2049 | Inflamma | FGF5 | P12034 | 4 | 80266639 | 80336680 | 36 | 0 | 0 | 0 | 0 | 1.13 | 0.94 | 1.35 | 0.210 | 0.885 | 51.09 | 0.04 | 197.89 | 1.13 | 0.97 | 1.31 | 0.129 | 0.812 | 1.07 | 0.84 | 1.36 | 0.583 | 1.11 | 0.77 | 1.59 | 0.572 | 0.00 | -0.10 | 0.10 | 0.924 |
| SPINK2 | OID3062 | Inflamma | SPINK2 | P20155 | 4 | 56809860 | 56821742 | 19 | 0 | 0 | 0 | 0 | 0.89 | 0.74 | 1.07 | 0.212 | 0.885 | 18.99 | 0.39 | 317.95 | 0.89 | 0.74 | 1.06 | 0.200 | 0.836 | 0.94 | 0.74 | 1.18 | 0.595 | 1.06 | 0.81 | 1.38 | 0.688 | -0.04 | -0.09 | 0.01 | 0.086 |
| SERPINF | OID2093 | Neurolog | SERPINF | P50453 | 6 | 2887270 | 2903309 | 4 | 0 | 0 | 0 | 0 | 0.67 | 0.36 | 1.25 | 0.212 | 0.885 | 2.78 | 0.43 | 99.14 | 0.67 | 0.36 | 1.26 | 0.215 | 0.854 | 0.80 | 0.39 | 1.64 | 0.541 | 1.20 | 0.31 | 4.63 | 0.795 | -0.07 | -0.22 | 0.08 | 0.347 |
| TBCC | OID2094 | Neurolog | TBCC | Q15814 | 6 | 42744498 | 42746103 | 2 | 0 | 0 | 0 | 0 | 1.61 | 0.76 | 3.41 | 0.210 | 0.885 | 0.33 | 0.57 | 179.92 | 1.61 | 0.76 | 3.44 | 0.214 | 0.853 | - | - | - | - | - | - | - | - | - | - | - | - |
| VNN2 | OID2131 | Oncology | VNN2 | O95498 | 6 | 132743870 | 132763459 | 30 | 0 | 0 | 0 | 0 | 0.92 | 0.81 | 1.05 | 0.209 | 0.885 | 20.63 | 0.87 | 311.66 | 0.92 | 0.81 | 1.05 | 0.210 | 0.849 | 0.94 | 0.80 | 1.11 | 0.464 | 0.80 | 0.66 | 0.98 | 0.032 | 0.05 | -0.01 | 0.10 | 0.081 |
| MMP12 | OID2143 | Oncology | MMP12 | P39900 | 11 | 102862736 | 102878982 | 26 | 0 | 0 | 0 | 0 | 0.90 | 0.77 | 1.06 | 0.212 | 0.885 | 22.79 | 0.59 | 258.76 | 0.90 | 0.77 | 1.06 | 0.212 | 0.851 | 0.89 | 0.73 | 1.09 | 0.265 | 0.84 | 0.67 | 1.04 | 0.111 | 0.02 | -0.02 | 0.07 | 0.320 |
| NCAM1 | OID2035 | Cardiom | NCAM1 | P13591 | 11 | 112961247 | 113278436 | 15 | 0 | 0 | 0 | 0 | 1.16 | 0.92 | 1.47 | 0.210 | 0.885 | 11.54 | 0.64 | 207.34 | 1.16 | 0.92 | 1.47 | 0.211 | 0.849 | 1.17 | 0.86 | 1.60 | 0.312 | 1.32 | 0.81 | 2.14 | 0.261 | -0.03 | -0.12 | 0.06 | 0.557 |
| LPCAT2 | OID2129 | Oncology | LPCAT2 | Q7L5N7 | 16 | 55509072 | 55586666 | 1 | 0 | 0 | 0 | 0 | 1.81 | 0.71 | 4.61 | 0.213 | 0.885 | - | - | 197.82 | 1.81 | 0.70 | 4.66 | 0.218 | 0.857 | - | - | - | - | - | - | - | - | - | - | - | - |
| MIF | OID2107 | Neurolog | MIF | P14174 | 22 | 23894383 | 23895227 | 2 | 0 | 0 | 0 | 0 | 1.66 | 0.75 | 3.69 | 0.213 | 0.885 | 0.96 | 0.33 | 122.58 | 1.66 | 0.74 | 3.74 | 0.217 | 0.857 | - | - | - | - | - | - | - | - | - | - | - | - |
| CD5L | OID3077 | Inflamma | CD5L | O43866 | 1 | 157830911 | 157898256 | 7 | 0 | 0 | 0 | 0 | 0.79 | 0.55 | 1.14 | 0.215 | 0.889 | 4.23 | 0.64 | 164.99 | 0.79 | 0.55 | 1.15 | 0.217 | 0.857 | 0.81 | 0.53 | 1.25 | 0.337 | 0.80 | 0.25 | 2.54 | 0.705 | 0.00 | -0.21 | 0.21 | 0.985 |
| LEPR | OID2012 | Cardiom | LEPR | P48357 | 1 | 65420652 | 65641559 | 8 | 0 | 0 | 0 | 0 | 1.24 | 0.88 | 1.75 | 0.214 | 0.889 | 5.23 | 0.63 | 181.39 | 1.24 | 0.88 | 1.76 | 0.216 | 0.855 | 1.32 | 0.86 | 2.01 | 0.202 | 2.38 | 0.79 | 7.22 | 0.125 | -0.14 | -0.37 | 0.09 | 0.226 |
| CASP7 | OID3113 | Neurolog | CASP7 | P55210 | 10 | 113679162 | 113730907 | 2 | 0 | 0 | 0 | 0 | 1.67 | 0.74 | 3.74 | 0.215 | 0.889 | 0.07 | 0.80 | 126.62 | 1.67 | 0.74 | 3.78 | 0.221 | 0.857 | - | - | - | - | - | - | - | - | - | - | - | - |
| LILRB1 | OID2032 | Cardiom | LILRB1 | Q8NHL6 | 19 | 54617158 | 54638022 | 30 | 0 | 0 | 0 | 0 | 0.89 | 0.74 | 1.07 | 0.217 | 0.894 | 43.45 | 0.04 | 260.23 | 0.89 | 0.77 | 1.03 | 0.130 | 0.812 | 0.86 | 0.70 | 1.06 | 0.159 | 1.02 | 0.73 | 1.43 | 0.910 | -0.04 | -0.12 | 0.04 | 0.354 |
| HS1BP3 | OID3024 | Cardiom | HS1BP3 | Q53T59 | 2 | 20560448 | 20651130 | 3 | 0 | 0 | 1 | 0 | 1.79 | 0.71 | 4.48 | 0.217 | 0.895 | 1.51 | 0.47 | 61.86 | 1.79 | 0.70 | 4.61 | 0.225 | 0.859 | 2.16 | 0.76 | 6.11 | 0.149 | 0.69 | 0.07 | 6.98 | 0.753 | 0.08 | -0.09 | 0.25 | 0.380 |
| SIGLEC1 | OID2053 | Inflamma | SIGLEC1 | Q96LC7 | 19 | 51410020 | 51417803 | 10 | 0 | 0 | 0 | 0 | 1.14 | 0.93 | 1.39 | 0.218 | 0.896 | 10.65 | 0.30 | 520.44 | 1.14 | 0.94 | 1.37 | 0.180 | 0.824 | 1.11 | 0.90 | 1.37 | 0.345 | 1.21 | 0.96 | 1.54 | 0.105 | -0.03 | -0.08 | 0.02 | 0.282 |
| AMIGO1 | OID3088 | Neurolog | AMIGO1 | Q86WK6 | 1 | 109504178 | 109509738 | 1 | 0 | 0 | 0 | 0 | 0.29 | 0.04 | 2.29 | 0.241 | 0.896 | - | - | 34.58 | 0.29 | 0.03 | 2.61 | 0.271 | 0.871 | - | - | - | - | - | - | - | - | - | - | - | - |
| BOLA1 | OID3083 | Neurolog | BOLA1 | Q9Y3E2 | 1 | 149887890 | 149900798 | 2 | 0 | 0 | 0 | 0 | 1.62 | 0.73 | 3.58 | 0.236 | 0.896 | 0.02 | 0.90 | 132.24 | 1.62 | 0.72 | 3.62 | 0.242 | 0.859 | - | - | - | - | - | - | - | - | - | - | - | - |
| C4BPB | OID2148 | Oncology | C4BPB | P20851 | 1 | 207088860 | 207099993 | 8 | 0 | 0 | 0 | 0 | 0.80 | 0.54 | 1.18 | 0.261 | 0.896 | 7.22 | 0.41 | 140.73 | 0.80 | 0.54 | 1.18 | 0.254 | 0.859 | 0.82 | 0.52 | 1.31 | 0.414 | 0.85 | 0.40 | 1.82 | 0.675 | -0.01 | -0.11 | 0.09 | 0.851 |
| F13B | OID3078 | Inflamma | F13B | P05160 | 1 | 197038741 | 197067260 | 26 | 0 | 0 | 0 | 0 | 1.11 | 0.94 | 1.31 | 0.231 | 0.896 | 19.85 | 0.75 | 224.56 | 1.11 | 0.94 | 1.31 | 0.232 | 0.859 | 1.22 | 0.96 | 1.55 | 0.109 | 0.83 | 0.52 | 1.31 | 0.416 | 0.08 | -0.04 | 0.19 | 0.179 |
| F3 | OID2144 | Oncology | F3 | P13726 | 1 | 94529173 | 94541759 | 12 | 0 | 0 | 0 | 0 | 1.24 | 0.87 | 1.75 | 0.237 | 0.896 | 6.24 | 0.86 | 124.31 | 1.24 | 0.87 | 1.76 | 0.240 | 0.859 | 1.23 | 0.79 | 1.92 | 0.358 | 1.20 | 0.54 | 2.70 | 0.655 | 0.00 | -0.09 | 0.09 | 0.941 |
| FCER1A | OID3099 | Neurolog | FCER1A | P12319 | 1 | 159289714 | 159308224 | 7 | 0 | 0 | 0 | 0 | 0.74 | 0.44 | 1.22 | 0.238 | 0.896 | 6.45 | 0.37 | 91.06 | 0.73 | 0.45 | 1.21 | 0.222 | 0.857 | 0.68 | 0.36 | 1.27 | 0.223 | 2.31 | 0.43 | 12.35 | 0.329 | -0.17 | -0.42 | 0.07 | 0.162 |
| FCGR2B | OID2121 | Oncology | FCGR2B | P31994 | 1 | 161663143 | 161678654 | 21 | 0 | 0 | 0 | 0 | 0.89 | 0.73 | 1.08 | 0.230 | 0.896 | 8.85 | 0.98 | 200.54 | 0.89 | 0.73 | 1.08 | 0.232 | 0.859 | 0.84 | 0.65 | 1.09 | 0.194 | 0.81 | 0.56 | 1.18 | 0.268 | 0.02 | -0.06 | 0.10 | 0.568 |
| FCRL1 | OID2017 | Cardiom | FCRL1 | Q96LA6 | 1 | 157794403 | 157820120 | 14 | 0 | 0 | 0 | 0 | 1.20 | 0.88 | 1.63 | 0.242 | 0.896 | 16.33 | 0.23 | 157.42 | 1.20 | 0.91 | 1.59 | 0.189 | 0.833 | 1.16 | 0.85 | 1.60 | 0.353 | 1.76 | 0.89 | 3.50 | 0.107 | -0.08 | -0.22 | 0.05 | 0.225 |
| FCRL3 | OID2044 | Inflamma | FCRL3 | Q96P31 | 1 | 157674321 | 157700769 | 43 | 0 | 0 | 0 | 0 | 1.07 | 0.95 | 1.21 | 0.239 | 0.896 | 49.62 | 0.20 | 296.15 | 1.07 | 0.96 | 1.20 | 0.200 | 0.836 | 1.02 | 0.86 | 1.20 | 0.858 | 1.02 | 0.82 | 1.26 | 0.868 | 0.02 | -0.04 | 0.08 | 0.553 |
| GBP4 | OID2084 | Neurolog | GBP4 | Q96PP9 | 1 | 89181144 | 89198942 | 47 | 0 | 0 | 0 | 0 | 0.93 | 0.83 | 1.05 | 0.259 | 0.896 | 34.68 | 0.89 | 231.64 | 0.93 | 0.83 | 1.05 | 0.260 | 0.860 | 0.90 | 0.76 | 1.08 | 0.261 | 0.85 | 0.69 | 1.05 | 0.135 | 0.03 | -0.03 | 0.09 | 0.295 |
| IGSF3 | OID2131 | Oncology | IGSF3 | O75054 | 1 | 116574399 | 116667755 | 5 | 0 | 0 | 1 | 0 | 0.60 | 0.25 | 1.43 | 0.251 | 0.896 | 6.91 | 0.14 | 76.08 | 0.60 | 0.31 | 1.16 | 0.130 | 0.812 | 0.82 | 0.35 | 1.92 | 0.651 | 1.35 | 0.26 | 7.06 | 0.719 | -0.08 | -0.22 | 0.06 | 0.264 |
| IGSF8 | OID2016 | Cardiom | IGSF8 | Q969P0 | 1 | 160091340 | 160098943 | 1 | 0 | 0 | 0 | 0 | 0.64 | 0.30 | 1.35 | 0.244 | 0.896 | - | - | 292.65 | 0.64 | 0.30 | 1.36 | 0.247 | 0.859 | - | - | - | - | - | - | - | - | - | - | - | - |
| IL6R | OID2038 | Cardiom | IL6R | P08887 | 1 | 154405193 | 154469450 | 49 | 0 | 0 | 0 | 0 | 0.94 | 0.86 | 1.04 | 0.233 | 0.896 | 44.98 | 0.60 | 354.37 | 0.94 | 0.86 | 1.04 | 0.233 | 0.859 | 0.95 | 0.83 | 1.09 | 0.466 | 0.95 | 0.80 | 1.12 | 0.541 | 0.00 | -0.05 | 0.05 | 0.945 |
| PGLYRP | OID3010 | Cardiom | PGLYRP | Q96LB8 | 1 | 153330120 | 153348841 | 13 | 0 | 0 | 0 | 0 | 0.89 | 0.74 | 1.08 | 0.244 | 0.896 | 9.66 | 0.65 | 313.47 | 0.89 | 0.74 | 1.08 | 0.244 | 0.859 | 0.95 | 0.77 | 1.19 | 0.664 | 0.96 | 0.75 | 1.23 | 0.730 | -0.03 | -0.10 | 0.04 | 0.371 |
| QSOX1 | OID3072 | Inflamma | QSOX1 | O00391 | 1 | 180154869 | 180204030 | 16 | 0 | 0 | 0 | 0 | 0.90 | 0.76 | 1.07 | 0.231 | 0.896 | 15.17 | 0.44 | 321.64 | 0.90 | 0.76 | 1.07 | 0.228 | 0.859 | 0.93 | 0.76 | 1.15 | 0.507 | 0.90 | 0.68 | 1.18 | 0.439 | 0.00 | -0.07 | 0.07 | 0.971 |
| RABGAP | OID2044 | Inflamma | RABGAP | Q5R372 | 1 | 174159410 | 174995308 | 3 | 0 | 0 | 1 | 0 | 0.62 | 0.27 | 1.40 | 0.250 | 0.896 | 0.20 | 0.91 | 78.45 | 0.62 | 0.27 | 1.43 | 0.260 | 0.860 | 0.65 | 0.26 | 1.64 | 0.362 | 1.51 | 0.02 | 140.76 | 0.859 | -0.07 | -0.43 | 0.29 | 0.695 |
| S100A3 | OID3135 | Oncology | S100A3 | P33764 | 1 | 153547329 | 153549258 | 2 | 0 | 0 | 0 | 0 | 0.40 | 0.09 | 1.84 | 0.239 | 0.896 | 0.18 | 0.67 | 35.05 | 0.40 | 0.08 | 2.00 | 0.262 | 0.860 | - | - | - | - | - | - | - | - | - | - | - | - |
| SCP2 | OID2116 | Oncology | SCP2 | P22307 | 1 | 52927276 | 53051698 | 1 | 0 | 0 | 0 | 0 | 0.29 | 0.03 | 2.46 | 0.253 | 0.896 | - | - | 31.68 | 0.29 | 0.03 | 2.85 | 0.286 | 0.879 | - | - | - | - | - | - | - | - | - | - | - | - |
| UROD | OID3060 | Inflamma | UROD | P06132 | 1 | 45010950 | 45015575 | 21 | 0 | 0 | 0 | 0 | 0.89 | 0.74 | 1.07 | 0.220 | 0.896 | 20.52 | 0.43 | 223.83 | 0.89 | 0.74 | 1.07 | 0.214 | 0.852 | 0.93 | 0.73 | 1.18 | 0.525 | 0.88 | 0.62 | 1.25 | 0.467 | 0.00 | -0.07 | 0.08 | 0.916 |
| ADAM23 | OID2065 | Inflamma | ADAM23 | O75077 | 2 | 206443532 | 206621127 | 30 | 0 | 0 | 0 | 0 | 1.08 | 0.95 | 1.22 | 0.219 | 0.896 | 26.38 | 0.61 | 335.81 | 1.08 | 0.95 | 1.22 | 0.219 | 0.857 | 1.16 | 0.96 | 1.39 | 0.123 | 1.01 | 0.83 | 1.23 | 0.935 | 0.03 | -0.03 | 0.09 | 0.364 |
| GPC1 | OID2143 | Oncology | GPC1 | P35052 | 2 | 240435663 | 240468076 | 14 | 0 | 0 | 0 | 0 | 1.14 | 0.90 | 1.45 | 0.261 | 0.896 | 8.44 | 0.81 | 222.31 | 1.14 | 0.90 | 1.45 | 0.262 | 0.860 | 1.09 | 0.82 | 1.44 | 0.540 | 1.07 | 0.76 | 1.52 | 0.698 | 0.02 | -0.05 | 0.08 | 0.617 |
| HNMT | OID2097 | Neurolog | HNMT | P50135 | 2 | 137964020 | 138016364 | 21 | 0 | 0 | 0 | 0 | 0.92 | 0.79 | 1.07 | 0.259 | 0.896 | 20.17 | 0.45 | 311.77 | 0.92 | 0.79 | 1.07 | 0.257 | 0.859 | 0.96 | 0.78 | 1.17 | 0.664 | 0.86 | 0.70 | 1.05 | 0.147 | 0.02 | -0.03 | 0.08 | 0.360 |
| HS6ST1 | OID2144 | Oncology | HS6ST1 | O60243 | 2 | 128236716 | 128318868 | 11 | 0 | 0 | 0 | 0 | 1.24 | 0.88 | 1.74 | 0.222 | 0.896 | 8.20 | 0.61 | 124.91 | 1.24 | 0.88 | 1.75 | 0.224 | 0.859 | 1.33 | 0.88 | 2.01 | 0.176 | 2.53 | 1.21 | 5.26 | 0.013 | -0.11 | -0.21 | -0.01 | 0.031 |
| INHBB | OID3062 | Inflamma | INHBB | P09529 | 2 | 120346136 | 120351803 | 11 | 0 | 0 | 0 | 0 | 0.85 | 0.64 | 1.13 | 0.254 | 0.896 | 5.79 | 0.83 | 181.54 | 0.85 | 0.63 | 1.13 | 0.256 | 0.859 | 0.93 | 0.68 | 1.28 | 0.666 | 1.14 | 0.64 | 2.02 | 0.662 | -0.06 | -0.17 | 0.04 | 0.246 |
| ALCAM | OID2027 | Cardiom | ALCAM | Q13740 | 3 | 105366909 | 105576900 | 6 | 0 | 0 | 1 | 0 | 1.54 | 0.77 | 3.11 | 0.226 | 0.896 | 5.78 | 0.33 | 71.81 | 1.55 | 0.80 | 3.01 | 0.194 | 0.836 | 1.67 | 0.73 | 3.82 | 0.224 | 0.84 | 0.21 | 3.33 | 0.803 | 0.06 | -0.06 | 0.19 | 0.315 |
| CRELD1 | OID3061 | Inflamma | CRELD1 | Q96HD1 | 3 | 9933793 | 9945413 | 29 | 0 | 0 | 0 | 0 | 0.90 | 0.75 | 1.07 | 0.244 | 0.896 | 40.69 | 0.06 | 258.71 | 0.90 | 0.78 | 1.04 | 0.159 | 0.823 | 0.91 | 0.74 | 1.12 | 0.376 | 0.77 | 0.55 | 1.07 | 0.113 | 0.04 | -0.03 | 0.12 | 0.256 |
| FSTL1 | OID3025 | Cardiom | FSTL1 | Q12841 | 3 | 120392293 | 120450993 | 4 | 0 | 0 | 0 | 0 | 0.70 | 0.39 | 1.28 | 0.247 | 0.896 | 4.41 | 0.22 | 157.34 | 0.70 | 0.43 | 1.15 | 0.161 | 0.823 | 0.76 | 0.43 | 1.33 | 0.334 | 0.20 | 0.00 | 13.63 | 0.455 | 0.17 | -0.39 | 0.72 | 0.554 |
| TIMP4 | OID2114 | Neurolog | TIMP4 | Q99727 | 3 | 12153068 | 12158912 | 12 | 0 | 0 | 0 | 0 | 0.87 | 0.68 | 1.10 | 0.240 | 0.896 | 11.24 | 0.42 | 247.36 | 0.87 | 0.68 | 1.10 | 0.236 | 0.859 | 0.85 | 0.65 | 1.12 | 0.245 | 0.68 | 0.44 | 1.05 | 0.083 | 0.06 | -0.03 | 0.16 | 0.193 |
| CXCL9 | OID2067 | Inflamma | CXCL9 | Q07325 | 4 | 76001275 | 76007509 | 2 | 0 | 0 | 0 | 0 | 0.67 | 0.34 | 1.31 | 0.241 | 0.896 | 0.00 | 0.95 | 201.79 | 0.67 | 0.34 | 1.32 | 0.246 | 0.859 | - | - | - | - | - | - | - | - | - | - | - | - |
| GNPDA2 | OID3059 | Inflamma | GNPDA2 | Q8TDQ7 | 4 | 44682200 | 44726588 | 15 | 0 | 0 | 0 | 0 | 0.84 | 0.62 | 1.12 | 0.237 | 0.896 | 12.96 | 0.53 | 119.25 | 0.84 | 0.62 | 1.13 | 0.238 | 0.859 | 0.87 | 0.60 | 1.27 | 0.468 | 1.81 | 0.50 | 6.48 | 0.364 | -0.15 | -0.40 | 0.09 | 0.225 |
| HADH | OID3013 | Cardiom | HADH | Q16836 | 4 | 107989714 | 108035241 | 1 | 0 | 0 | 0 | 0 | 2.57 | 0.54 | 12.23 | 0.234 | 0.896 | - | - | 55.95 | 2.57 | 0.51 | 13.02 | 0.253 | 0.859 | - | - | - | - | - | - | - | - | - | - | - | - |
| SCARB2 | OID2094 | Neurolog | SCARB2 | Q14108 | 4 | 76158737 | 76234536 | 8 | 0 | 0 | 0 | 0 | 0.83 | 0.61 | 1.13 | 0.233 | 0.896 | 4.36 | 0.74 | 239.48 | 0.83 | 0.61 | 1.13 | 0.235 | 0.859 | 0.83 | 0.58 | 1.18 | 0.301 | 0.75 | 0.44 | 1.27 | 0.280 | 0.02 | -0.07 | 0.12 | 0.632 |
| CXCL14 | OID2045 | Inflamma | CXCL14 | O95715 | 5 | 135570679 | 135579279 | 2 | 0 | 0 | 0 | 0 | 1.93 | 0.65 | 5.76 | 0.238 | 0.896 | 0.06 | 0.81 | 73.02 | 1.93 | 0.63 | 5.92 | 0.250 | 0.859 | - | - | - | - | - | - | - | - | - | - | - | - |
| EGFLAM | OID3149 | Oncology | EGFLAM | Q63HQ2 | 5 | 38258409 | 38465480 | 16 | 0 | 0 | 0 | 0 | 1.14 | 0.92 | 1.42 | 0.226 | 0.896 | 12.49 | 0.64 | 219.80 | 1.14 | 0.92 | 1.42 | 0.228 | 0.859 | 1.06 | 0.81 | 1.40 | 0.666 | 1.01 | 0.65 | 1.55 | 0.974 | 0.03 | -0.07 | 0.14 | 0.507 |
| HAVCR1 | OID2142 | Oncology | HAVCR1 | Q96D42 | 5 | 157029413 | 157059119 | 36 | 0 | 0 | 0 | 0 | 1.10 | 0.93 | 1.29 | 0.257 | 0.896 | 27.83 | 0.80 | 209.84 | 1.10 | 0.93 | 1.29 | 0.257 | 0.859 | 1.11 | 0.89 | 1.39 | 0.339 | 1.13 | 0.82 | 1.57 | 0.450 | -0.01 | -0.08 | 0.06 | 0.824 |
| HCG22 | OID3008 | Cardiom | HCG22 | E2RYF7 | 6 | 31053450 | 31059890 | 23 | 0 | 0 | 0 | 0 | 1.12 | 0.92 | 1.35 | 0.254 | 0.896 | 20.46 | 0.55 | 174.78 | 1.12 | 0.92 | 1.36 | 0.255 | 0.859 | 1.03 | 0.80 | 1.34 | 0.794 | 1.16 | 0.79 | 1.69 | 0.449 | -0.01 | -0.08 | 0.07 | 0.831 |
| NFKBIE | OID2121 | Oncology | NFKBIE | O00221 | 6 | 44258166 | 44265788 | 1 | 0 | 0 | 0 | 0 | 1.33 | 0.83 | 2.14 | 0.236 | 0.896 | - | - | 791.19 | 1.33 | 0.83 | 2.15 | 0.237 | 0.859 | - | - | - | - | - | - | - | - | - | - | - | - |
| PLA2G7 | OID2110 | Neurolog | PLA2G7 | Q13093 | 6 | 46704201 | 46735693 | 3 | 0 | 0 | 0 | 0 | 0.59 | 0.25 | 1.43 | 0.246 | 0.896 | 1.53 | 0.46 | 72.91 | 0.59 | 0.24 | 1.45 | 0.252 | 0.859 | 0.59 | 0.23 | 1.52 | 0.274 | 0.14 | 0.01 | 3.44 | 0.228 | 0.12 | -0.13 | 0.37 | 0.356 |
| STX7 | OID3056 | Inflamma | STX7 | O15400 | 6 | 132445867 | 132513198 | 27 | 0 | 0 | 0 | 0 | 1.11 | 0.93 | 1.34 | 0.250 | 0.896 | 17.23 | 0.90 | 184.62 | 1.11 | 0.93 | 1.34 | 0.252 | 0.859 | 0.99 | 0.78 | 1.27 | 0.966 | 1.00 | 0.74 | 1.34 | 0.983 | 0.03 | -0.03 | 0.08 | 0.352 |
| HIP1 | OID3007 | Cardiom | HIP1 | O00291 | 7 | 75533298 | 75738962 | 2 | 0 | 0 | 0 | 0 | 0.41 | 0.09 | 1.89 | 0.253 | 0.896 | 0.21 | 0.65 | 32.63 | 0.41 | 0.08 | 2.05 | 0.278 | 0.879 | - | - | - | - | - | - | - | - | - | - | - | - |
| LRCH4 | OID3019 | Cardiom | LRCH4 | O75427 | 7 | 100574011 | 100586129 | 1 | 0 | 0 | 0 | 0 | 2.34 | 0.55 | 9.89 | 0.248 | 0.896 | - | - | 73.07 | 2.34 | 0.53 | 10.33 | 0.262 | 0.860 | - | - | - | - | - | - | - | - | - | - | - | - |
| NAMPT | OID2122 | Oncology | NAMPT | P43490 | 7 | 106248298 | 106285966 | 1 | 0 | 0 | 0 | 0 | 3.70 | 0.44 | 30.94 | 0.227 | 0.896 | - | - | 34.31 | 3.70 | 0.38 | 35.61 | 0.257 | 0.859 | - | - | - | - | - | - | - | - | - | - | - | - |
| PON3 | OID2077 | Inflamma | PON3 | Q15166 | 7 | 95359872 | 95396375 | 13 | 0 | 0 | 0 | 0 | 0.86 | 0.67 | 1.11 | 0.252 | 0.896 | 15.57 | 0.21 | 238.78 | 0.86 | 0.69 | 1.08 | 0.191 | 0.833 | 0.90 | 0.68 | 1.19 | 0.474 | 0.75 | 0.51 | 1.11 | 0.146 | 0.04 | -0.04 | 0.11 | 0.350 |
| DECR1 | OID2057 | Inflamma | DECR1 | Q16698 | 8 | 90001405 | 90053633 | 1 | 0 | 0 | 0 | 0 | 3.83 | 0.43 | 33.87 | 0.228 | 0.896 | - | - | 37.74 | 3.83 | 0.38 | 38.61 | 0.255 | 0.859 | - | - | - | - | - | - | - | - | - | - | - | - |
| GGH | OID2041 | Cardiom | GGH | Q92820 | 8 | 63014881 | 63039407 | 27 | 0 | 0 | 0 | 0 | 1.12 | 0.92 | 1.38 | 0.260 | 0.896 | 27.61 | 0.38 | 157.51 | 1.12 | 0.92 | 1.37 | 0.246 | 0.859 | 1.03 | 0.78 | 1.35 | 0.860 | 1.21 | 0.86 | 1.70 | 0.273 | -0.02 | -0.08 | 0.05 | 0.595 |
| STC1 | OID2101 | Neurolog | STC1 | P52823 | 8 | 23841929 | 23854806 | 1 | 0 | 0 | 0 | 0 | 2.35 | 0.59 | 9.46 | 0.228 | 0.896 | - | - | 77.59 | 2.35 | 0.56 | 9.85 | 0.241 | 0.859 | - | - | - | - | - | - | - | - | - | - | - | - |
| AMBP | OID2141 | Oncology | AMBP | P02760 | 9 | 114060127 | 114078328 | 3 | 0 | 0 | 1 | 0 | 1.53 | 0.74 | 3.17 | 0.253 | 0.896 | 0.84 | 0.66 | 115.34 | 1.53 | 0.73 | 3.21 | 0.258 | 0.859 | 1.73 | 0.76 | 3.95 | 0.193 | 0.97 | 0.12 | 7.99 | 0.980 | 0.06 | -0.21 | 0.34 | 0.654 |
| CCL21 | OID2068 | Inflamma | CCL21 | O00585 | 9 | 34709005 | 34710136 | 4 | 0 | 0 | 0 | 0 | 0.66 | 0.32 | 1.36 | 0.258 | 0.896 | 0.41 | 0.94 | 78.15 | 0.66 | 0.31 | 1.38 | 0.267 | 0.868 | 0.71 | 0.31 | 1.62 | 0.418 | 0.28 | 0.01 | 7.16 | 0.442 | 0.09 | -0.23 | 0.40 | 0.597 |
| GCNT1 | OID2137 | Oncology | GCNT1 | Q02742 | 9 | 76419850 | 76651203 | 17 | 0 | 0 | 0 | 0 | 0.89 | 0.73 | 1.08 | 0.221 | 0.896 | 11.19 | 0.80 | 243.44 | 0.89 | 0.73 | 1.08 | 0.222 | 0.857 | 0.83 | 0.65 | 1.07 | 0.149 | 1.02 | 0.75 | 1.38 | 0.919 | -0.04 | -0.11 | 0.03 | 0.262 |
| NCS1 | OID2145 | Oncology | NCS1 | P62166 | 9 | 130172404 | 130237303 | 3 | 0 | 0 | 0 | 0 | 0.65 | 0.32 | 1.33 | 0.240 | 0.896 | 3.07 | 0.21 | 136.24 | 0.65 | 0.32 | 1.33 | 0.242 | 0.859 | 0.61 | 0.28 | 1.32 | 0.212 | 2.25 | 0.03 | 198.77 | 0.722 | -0.16 | -0.74 | 0.41 | 0.577 |
| NINJ1 | OID2116 | Oncology | NINJ1 | Q92982 | 9 | 93121496 | 93134251 | 3 | 0 | 0 | 0 | 0 | 1.26 | 0.86 | 1.84 | 0.231 | 0.896 | 1.19 | 0.55 | 353.92 | 1.26 | 0.86 | 1.84 | 0.232 | 0.859 | 1.18 | 0.80 | 1.74 | 0.400 | 1.04 | 0.53 | 2.06 | 0.904 | 0.04 | -0.07 | 0.14 | 0.514 |
| TRAF2 | OID2050 | Inflamma | TRAF2 | Q12933 | 9 | 136881912 | 136926607 | 1 | 0 | 0 | 0 | 0 | 2.82 | 0.52 | 15.26 | 0.228 | 0.896 | - | - | 58.25 | 2.82 | 0.49 | 16.29 | 0.246 | 0.859 | - | - | - | - | - | - | - | - | - | - | - | - |
| ADAM8 | OID2103 | Neurolog | ADAM8 | P78325 | 10 | 133262420 | 133276868 | 14 | 0 | 0 | 0 | 0 | 1.19 | 0.89 | 1.57 | 0.236 | 0.896 | 17.05 | 0.20 | 185.86 | 1.19 | 0.93 | 1.52 | 0.174 | 0.823 | 1.08 | 0.80 | 1.45 | 0.620 | 1.31 | 0.85 | 2.03 | 0.226 | -0.03 | -0.12 | 0.06 | 0.554 |
| GOT1 | OID3041 | Cardiom | GOT1 | P17174 | 10 | 99396870 | 99430624 | 1 | 0 | 0 | 0 | 0 | 3.73 | 0.38 | 36.85 | 0.260 | 0.896 | - | - | 31.71 | 3.73 | 0.32 | 43.16 | 0.292 | 0.879 | - | - | - | - | - | - | - | - | - | - | - | - |
| PNLIPRP | OID3112 | Neurolog | PNLIPRP | P54315 | 10 | 116590385 | 116609175 | 8 | 0 | 0 | 0 | 0 | 1.23 | 0.88 | 1.71 | 0.231 | 0.896 | 1.11 | 0.99 | 183.93 | 1.23 | 0.88 | 1.72 | 0.235 | 0.859 | 1.20 | 0.82 | 1.75 | 0.349 | 1.38 | 0.66 | 2.92 | 0.395 | -0.02 | -0.14 | 0.10 | 0.724 |
| RET | OID2134 | Oncology | RET | P07949 | 10 | 43077064 | 43130351 | 26 | 0 | 0 | 0 | 0 | 1.10 | 0.93 | 1.30 | 0.249 | 0.896 | 20.86 | 0.70 | 214.75 | 1.10 | 0.93 | 1.31 | 0.250 | 0.859 | 1.12 | 0.89 | 1.39 | 0.333 | 1.13 | 0.78 | 1.65 | 0.515 | -0.01 | -0.11 | 0.09 | 0.876 |
| CRTAM | OID2091 | Neurolog | CRTAM | O95727 | 11 | 122838500 | 122872643 | 12 | 0 | 0 | 0 | 0 | 0.85 | 0.64 | 1.12 | 0.248 | 0.896 | 14.64 | 0.20 | 216.13 | 0.85 | 0.66 | 1.08 | 0.182 | 0.824 | 0.96 | 0.71 | 1.30 | 0.793 | 0.94 | 0.61 | 1.45 | 0.783 | -0.03 | -0.11 | 0.05 | 0.531 |
| IL10RA | OID2042 | Inflamma | IL10RA | Q13651 | 11 | 117986370 | 118003037 | 1 | 0 | 0 | 0 | 0 | 1.46 | 0.76 | 2.82 | 0.253 | 0.896 | - | - | 334.16 | 1.46 | 0.76 | 2.83 | 0.256 | 0.859 | - | - | - | - | - | - | - | - | - | - | - | - |

| | | | | | | | | | | | | | | | | | | | | | | | | | | | | | | | | | | | | | |
|---|---|---|---|---|---|---|---|---|---|---|---|---|---|---|---|---|---|---|---|---|---|---|---|---|---|---|---|---|---|---|---|---|---|---|---|---|---|
| MMP13 | OID2080 | Neurolog | MMP13 | P45452 | 11 | 102942995 | 102955732 | 2 | 0 | 0 | 0 | 0 | 0.67 | 0.35 | 1.30 | 0.239 | 0.896 | 1.49 | 0.22 | 178.03 | 0.67 | 0.35 | 1.30 | 0.241 | 0.859 | - | - | - | - | - | - | - | - | - | - | - | - |
| YAP1 | OID3111 | Neurolog | YAP1 | P46937 | 11 | 102110447 | 102233424 | 1 | 0 | 0 | 0 | 0 | 2.50 | 0.55 | 11.42 | 0.238 | 0.896 | - | - | 81.50 | 2.50 | 0.52 | 11.90 | 0.251 | 0.859 | - | - | - | - | - | - | - | - | - | - | - | - |
| C1RL | OID3072 | Inflamma | C1RL | Q9NZP8 | 12 | 7089587 | 7109238 | 14 | 0 | 0 | 0 | 0 | 1.10 | 0.94 | 1.30 | 0.238 | 0.896 | 4.64 | 0.98 | 477.58 | 1.10 | 0.94 | 1.30 | 0.240 | 0.859 | 1.11 | 0.92 | 1.35 | 0.286 | 1.12 | 0.92 | 1.37 | 0.271 | -0.01 | -0.05 | 0.04 | 0.809 |
| SLC39A5 | OID2053 | Inflamma | SLC39A5 | Q6ZMH5 | 12 | 56230049 | 56237846 | 2 | 0 | 0 | 0 | 0 | 1.90 | 0.64 | 5.61 | 0.245 | 0.896 | 0.28 | 0.60 | 80.17 | 1.90 | 0.63 | 5.76 | 0.255 | 0.859 | - | - | - | - | - | - | - | - | - | - | - | - |
| STAB2 | OID3040 | Cardiom | STAB2 | Q8WWQ | 12 | 103587273 | 103766719 | 20 | 0 | 0 | 0 | 0 | 1.12 | 0.93 | 1.35 | 0.242 | 0.896 | 9.13 | 0.97 | 244.11 | 1.12 | 0.93 | 1.35 | 0.243 | 0.859 | 1.19 | 0.93 | 1.51 | 0.162 | 1.23 | 0.92 | 1.65 | 0.158 | -0.03 | -0.09 | 0.04 | 0.389 |
| VWF | OID2025 | Cardiom | VWF | P04275 | 12 | 5948877 | 6124770 | 2 | 0 | 0 | 0 | 0 | 1.61 | 0.70 | 3.67 | 0.261 | 0.896 | 0.00 | 0.98 | 121.60 | 1.61 | 0.69 | 3.72 | 0.268 | 0.870 | - | - | - | - | - | - | - | - | - | - | - | - |
| ACYP1 | OID3064 | Inflamma | ACYP1 | P07311 | 14 | 75053237 | 75069483 | 3 | 0 | 0 | 0 | 0 | 1.40 | 0.81 | 2.40 | 0.228 | 0.896 | 2.25 | 0.33 | 169.22 | 1.40 | 0.81 | 2.41 | 0.230 | 0.859 | 1.47 | 0.82 | 2.64 | 0.201 | 2.77 | 0.96 | 7.94 | 0.058 | -0.10 | -0.23 | 0.03 | 0.138 |
| FDDM3B | OID3136 | Oncology | FDDM3B | P56851 | 14 | 20768404 | 20770948 | 2 | 0 | 0 | 0 | 0 | 0.69 | 0.37 | 1.29 | 0.246 | 0.896 | 0.54 | 0.46 | 478.60 | 0.69 | 0.37 | 1.29 | 0.247 | 0.859 | - | - | - | - | - | - | - | - | - | - | - | - |
| RNASE3 | OID2020 | Cardiom | RNASE3 | P12724 | 14 | 20891385 | 20892348 | 6 | 0 | 0 | 0 | 0 | 1.19 | 0.88 | 1.61 | 0.260 | 0.896 | 3.77 | 0.58 | 285.32 | 1.19 | 0.88 | 1.62 | 0.261 | 0.860 | 1.19 | 0.85 | 1.65 | 0.313 | 1.15 | 0.74 | 1.80 | 0.535 | 0.01 | -0.09 | 0.11 | 0.846 |
| RNASE6 | OID3068 | Inflamma | RNASE6 | Q93091 | 14 | 20781268 | 20782467 | 14 | 0 | 0 | 0 | 0 | 0.89 | 0.74 | 1.07 | 0.224 | 0.896 | 9.97 | 0.70 | 356.88 | 0.89 | 0.74 | 1.07 | 0.224 | 0.859 | 0.88 | 0.71 | 1.09 | 0.231 | 0.92 | 0.66 | 1.28 | 0.601 | -0.01 | -0.09 | 0.07 | 0.857 |
| PRTG | OID2144 | Oncology | PRTG | Q2VWP7 | 15 | 55611544 | 55743152 | 23 | 0 | 0 | 0 | 0 | 0.90 | 0.77 | 1.07 | 0.228 | 0.896 | 19.92 | 0.59 | 249.77 | 0.90 | 0.77 | 1.07 | 0.229 | 0.859 | 0.99 | 0.79 | 1.23 | 0.907 | 1.14 | 0.84 | 1.55 | 0.397 | -0.07 | -0.15 | 0.01 | 0.076 |
| ARL2BP | OID3012 | Cardiom | ARL2BP | Q9Y2Y0 | 16 | 57245259 | 57253635 | 1 | 0 | 0 | 0 | 0 | 1.24 | 0.87 | 1.79 | 0.239 | 0.896 | - | - | 1282.37 | 1.24 | 0.86 | 1.79 | 0.239 | 0.859 | - | - | - | - | - | - | - | - | - | - | - | - |
| CIAPIN1 | OID2150 | Oncology | CIAPIN1 | Q6FI81 | 16 | 57428187 | 57447420 | 1 | 0 | 0 | 0 | 0 | 2.76 | 0.52 | 14.78 | 0.236 | 0.896 | - | - | 56.02 | 2.76 | 0.48 | 15.80 | 0.254 | 0.859 | - | - | - | - | - | - | - | - | - | - | - | - |
| DPEP1 | OID2089 | Neurolog | DPEP1 | P16444 | 16 | 89613308 | 89638456 | 34 | 0 | 0 | 0 | 0 | 0.94 | 0.85 | 1.04 | 0.241 | 0.896 | 25.59 | 0.82 | 444.19 | 0.94 | 0.85 | 1.04 | 0.241 | 0.859 | 0.89 | 0.77 | 1.03 | 0.132 | 0.86 | 0.71 | 1.04 | 0.117 | 0.03 | -0.03 | 0.09 | 0.263 |
| LAT | OID2064 | Inflamma | LAT | O43561 | 16 | 28984826 | 28990784 | 1 | 0 | 0 | 0 | 0 | 3.46 | 0.41 | 29.55 | 0.256 | 0.896 | - | - | 32.29 | 3.46 | 0.35 | 34.20 | 0.288 | 0.879 | - | - | - | - | - | - | - | - | - | - | - | - |
| PKD1 | OID3032 | Cardiom | PKD1 | P98161 | 16 | 2088708 | 2135898 | 9 | 0 | 0 | 0 | 0 | 0.85 | 0.64 | 1.13 | 0.252 | 0.896 | 7.62 | 0.47 | 237.12 | 0.85 | 0.63 | 1.13 | 0.253 | 0.859 | 0.85 | 0.61 | 1.20 | 0.357 | 0.77 | 0.54 | 1.11 | 0.158 | 0.03 | -0.04 | 0.09 | 0.409 |
| SMPD3 | OID3047 | Inflamma | SMPD3 | Q9NY59 | 16 | 68358327 | 68448508 | 1 | 0 | 0 | 0 | 0 | 3.45 | 0.41 | 28.72 | 0.252 | 0.896 | - | - | 34.31 | 3.45 | 0.36 | 32.91 | 0.282 | 0.879 | - | - | - | - | - | - | - | - | - | - | - | - |
| UMOD | OID2023 | Cardiom | UMOD | P07911 | 16 | 20333051 | 20356301 | 28 | 0 | 0 | 0 | 0 | 1.09 | 0.94 | 1.27 | 0.240 | 0.896 | 18.15 | 0.90 | 283.68 | 1.09 | 0.94 | 1.27 | 0.241 | 0.859 | 1.03 | 0.83 | 1.28 | 0.779 | 0.99 | 0.77 | 1.27 | 0.948 | 0.03 | -0.04 | 0.10 | 0.350 |
| ALDH3A1 | OID2051 | Inflamma | ALDH3A1 | P30838 | 17 | 19737984 | 19748943 | 29 | 0 | 0 | 0 | 0 | 1.11 | 0.93 | 1.32 | 0.251 | 0.896 | 21.29 | 0.81 | 186.50 | 1.11 | 0.93 | 1.32 | 0.252 | 0.859 | 1.13 | 0.89 | 1.44 | 0.324 | 1.30 | 0.79 | 2.14 | 0.307 | -0.03 | -0.14 | 0.07 | 0.510 |
| CDC27 | OID2123 | Oncology | CDC27 | P30260 | 17 | 47117703 | 47189422 | 1 | 0 | 0 | 0 | 0 | 1.99 | 0.65 | 6.14 | 0.229 | 0.896 | - | - | 117.12 | 1.99 | 0.63 | 6.27 | 0.238 | 0.859 | - | - | - | - | - | - | - | - | - | - | - | - |
| TK1 | OID3129 | Oncology | TK1 | P04183 | 17 | 78174091 | 78187233 | 1 | 0 | 0 | 0 | 0 | 2.27 | 0.57 | 9.07 | 0.246 | 0.896 | - | - | 102.63 | 2.27 | 0.55 | 9.34 | 0.256 | 0.859 | - | - | - | - | - | - | - | - | - | - | - | - |
| TP53 | OID2123 | Oncology | TP53 | P04637 | 17 | 7661779 | 7687538 | 1 | 0 | 0 | 0 | 0 | 2.57 | 0.50 | 13.09 | 0.257 | 0.896 | - | - | 54.01 | 2.57 | 0.47 | 13.98 | 0.275 | 0.879 | - | - | - | - | - | - | - | - | - | - | - | - |
| BCL2 | OID3145 | Oncology | BCL2 | P10415 | 18 | 63123346 | 63320128 | 3 | 0 | 0 | 1 | 0 | 0.49 | 0.14 | 1.66 | 0.253 | 0.896 | 1.59 | 0.45 | 36.33 | 0.48 | 0.14 | 1.73 | 0.265 | 0.864 | 0.47 | 0.10 | 2.09 | 0.318 | 8.21 | 0.08 | 804.91 | 0.368 | -0.16 | -0.40 | 0.09 | 0.211 |
| GRP | OID3026 | Cardiom | GRP | P07492 | 18 | 59220158 | 59230774 | 9 | 0 | 0 | 0 | 0 | 0.87 | 0.69 | 1.10 | 0.238 | 0.896 | 6.26 | 0.62 | 333.54 | 0.87 | 0.69 | 1.10 | 0.239 | 0.859 | 0.88 | 0.68 | 1.15 | 0.356 | 1.02 | 0.72 | 1.43 | 0.932 | -0.04 | -0.09 | 0.02 | 0.243 |
| YES1 | OID2134 | Oncology | YES1 | P07947 | 18 | 721588 | 812546 | 2 | 0 | 0 | 0 | 0 | 1.89 | 0.66 | 5.41 | 0.238 | 0.896 | 3.27 | 0.07 | 72.20 | 1.91 | 0.66 | 5.57 | 0.235 | 0.859 | - | - | - | - | - | - | - | - | - | - | - | - |
| A1BG | OID3077 | Inflamma | A1BG | P04217 | 19 | 58345178 | 58353492 | 5 | 0 | 0 | 0 | 0 | 1.36 | 0.82 | 2.28 | 0.235 | 0.896 | 1.17 | 0.88 | 143.18 | 1.36 | 0.81 | 2.29 | 0.239 | 0.859 | 1.28 | 0.69 | 2.37 | 0.431 | 1.45 | 0.51 | 4.14 | 0.489 | -0.01 | -0.20 | 0.18 | 0.896 |
| IFI30 | OID3150 | Oncology | IFI30 | P13284 | 19 | 18173162 | 18178117 | 15 | 0 | 0 | 0 | 0 | 1.17 | 0.91 | 1.50 | 0.224 | 0.896 | 16.76 | 0.27 | 219.15 | 1.17 | 0.93 | 1.47 | 0.183 | 0.824 | 1.17 | 0.85 | 1.61 | 0.326 | 1.17 | 0.78 | 1.76 | 0.455 | 0.00 | -0.08 | 0.08 | 0.999 |

| | | | | | | | | | | | | | | |
|---|---|---|---|---|---|---|---|---|---|---|---|---|---|---|
| TGFA | OID2060 | Inflamma | TGFA | P01135 | 2 | 70447284 | 70554193 | 2 | 0 | 0 | 0 | 0 | 1.40 | 0.66 |
| TP53I3 | OID3063 | Inflamma | TP53I3 | Q53FA7 | 2 | 24077433 | 24085861 | 20 | 0 | 0 | 0 | 0 | 1.10 | 0.90 |
| ABHD14B | OID2092 | Neurolog | ABHD14B | Q96IU4 | 3 | 51968510 | 51983409 | 5 | 0 | 0 | 0 | 0 | 0.90 | 0.70 |
| CCDC50 | OID3100 | Neurolog | CCDC50 | Q8IVM0 | 3 | 191329085 | 191398659 | 3 | 0 | 0 | 0 | 0 | 0.83 | 0.52 |
| GP5 | OID3061 | Inflamma | GP5 | P40197 | 3 | 194394821 | 194399266 | 1 | 0 | 0 | 0 | 0 | 1.39 | 0.60 |
| LZTFL1 | OID3063 | Inflamma | LZTFL1 | Q9NQ48 | 3 | 45823316 | 45916042 | 2 | 0 | 0 | 0 | 0 | 1.32 | 0.70 |
| PODXL2 | OID2148 | Oncology | PODXL2 | Q9NZ53 | 3 | 127629185 | 127672802 | 4 | 0 | 0 | 0 | 0 | 1.26 | 0.71 |
| PTH1R | OID2057 | Inflamma | PTH1R | Q03431 | 3 | 46877721 | 46903799 | 7 | 0 | 0 | 0 | 0 | 1.20 | 0.70 |
| AFAP1 | OID3056 | Inflamma | AFAP1 | Q8N556 | 4 | 7758714 | 7939926 | 33 | 0 | 0 | 0 | 0 | 1.04 | 0.94 |
| DMP1 | OID3021 | Cardiome | DMP1 | Q13316 | 4 | 87650280 | 87664361 | 1 | 0 | 0 | 0 | 0 | 2.24 | 0.30 |
| FGFBP2 | OID3039 | Cardiome | FGFBP2 | Q9BYJ0 | 4 | 15960245 | 15969309 | 17 | 0 | 0 | 0 | 0 | 0.93 | 0.77 |
| PDE5A | OID3058 | Inflamma | PDE5A | O76074 | 4 | 119494397 | 119628804 | 7 | 0 | 0 | 0 | 0 | 0.83 | 0.52 |
| SNCA | OID3064 | Inflamma | SNCA | P37840 | 4 | 89700345 | 89838315 | 1 | 0 | 0 | 0 | 0 | 2.21 | 0.28 |
| TLR3 | OID2061 | Inflamma | TLR3 | O15455 | 4 | 186069155 | 186088073 | 40 | 0 | 0 | 0 | 0 | 0.96 | 0.80 |
| AP3B1 | OID3147 | Oncology | AP3B1 | O00203 | 5 | 78000522 | 78294762 | 1 | 0 | 0 | 0 | 0 | 0.46 | 0.04 |
| CSF1R | OID3068 | Inflamma | CSF1R | P07333 | 5 | 150053291 | 150113372 | 4 | 0 | 0 | 0 | 0 | 0.82 | 0.51 |
| DBN1 | OID3054 | Inflamma | DBN1 | Q16643 | 5 | 177456608 | 177474401 | 1 | 0 | 0 | 0 | 0 | 0.45 | 0.06 |
| DNAJC21 | OID3110 | Neurolog | DNAJC21 | Q5F1R6 | 5 | 34929559 | 34958964 | 2 | 0 | 0 | 0 | 0 | 0.72 | 0.32 |
| F12 | OID3073 | Inflamma | F12 | P00748 | 5 | 177402133 | 177416583 | 45 | 0 | 0 | 0 | 0 | 0.94 | 0.81 |
| FBN2 | OID3097 | Neurolog | FBN2 | P35556 | 5 | 128257909 | 128659185 | 9 | 0 | 0 | 0 | 0 | 1.18 | 0.82 |
| GFRA3 | OID2093 | Neurolog | GFRA3 | O60609 | 5 | 138252380 | 138274621 | 3 | 0 | 0 | 1 | 0 | 1.39 | 0.60 |
| GZMA | OID2066 | Inflamma | GZMA | P12544 | 5 | 55102646 | 55110252 | 2 | 0 | 0 | 0 | 0 | 1.76 | 0.50 |
| LMNB1 | OID3150 | Oncology | LMNB1 | P20700 | 5 | 126776623 | 126837020 | 1 | 0 | 0 | 0 | 0 | 2.43 | 0.28 |
| OXCT1 | OID3022 | Cardiome | OXCT1 | P55809 | 5 | 41730065 | 41870425 | 1 | 0 | 0 | 0 | 0 | 0.41 | 0.05 |
| PDLIM7 | OID2072 | Inflamma | PDLIM7 | Q9NR12 | 5 | 177483394 | 177497606 | 2 | 0 | 0 | 0 | 0 | 0.52 | 0.11 |
| SEPTIN8 | OID3107 | Neurolog | SEPTIN8 | Q92599 | 5 | 132750819 | 132807241 | 2 | 0 | 0 | 0 | 0 | 1.52 | 0.58 |
| SNX18 | OID3147 | Oncology | SNX18 | Q96RF0 | 5 | 54517759 | 54546586 | 1 | 0 | 0 | 0 | 0 | 0.60 | 0.17 |
| THBS4 | OID2036 | Cardiome | THBS4 | P35443 | 5 | 79991311 | 80083287 | 8 | 0 | 0 | 0 | 0 | 1.14 | 0.83 |
| AKAP12 | OID3057 | Inflamma | AKAP12 | Q02952 | 6 | 151239967 | 151358559 | 1 | 0 | 0 | 0 | 0 | 0.41 | 0.05 |
| ARG1 | OID2144 | Oncology | ARG1 | P05089 | 6 | 131470832 | 131584332 | 4 | 0 | 0 | 0 | 0 | 0.79 | 0.46 |
| ATP6V1C | OID3016 | Cardiome | ATP6V1C | O95670 | 6 | 31544444 | 31548427 | 8 | 0 | 0 | 0 | 0 | 1.22 | 0.76 |

| | | | | | | | | | | | | | | | | | | | | | | | | | | | | | | | | | | | | | | |
|---|---|---|---|---|---|---|---|---|---|---|---|---|---|---|---|---|---|---|---|---|---|---|---|---|---|---|---|---|---|---|---|---|---|---|---|---|---|---|
| CLEC4M | OID3040 | Cardiom | CLEC4M | Q9H2X3 | 19 | 7763210 | 7769605 | 13 | 0 | 0 | 0 | 0 | 1.08 | 0.86 | 1.34 | 0.516 | 0.945 | 13.05 | 0.37 | 393.37 | 1.08 | 1.08 | 0.87 | 1.33 | 0.498 | 0.920 | 1.07 | 0.82 | 1.38 | 0.624 | 1.09 | 0.67 | 1.79 | 0.722 | 0.00 | -0.11 | 0.10 | 0.941 |
| KIR2DL2 | OID3143 | Oncology | KIR2DL2 | P43627 | 19 | 54754693 | 54769187 | 18 | 0 | 0 | 0 | 0 | 1.07 | 0.88 | 1.29 | 0.510 | 0.945 | 18.73 | 0.34 | 328.48 | 1.07 | 1.07 | 0.89 | 1.28 | 0.489 | 0.912 | 1.07 | 0.87 | 1.33 | 0.513 | 0.97 | 0.72 | 1.31 | 0.830 | 0.03 | -0.04 | 0.09 | 0.412 |
| KLK4 | OID2139 | Oncology | KLK4 | Q9Y5K2 | 19 | 50906351 | 50911395 | 17 | 0 | 0 | 0 | 0 | 1.07 | 0.87 | 1.31 | 0.517 | 0.945 | 16.89 | 0.39 | 288.24 | 1.07 | 1.07 | 0.88 | 1.30 | 0.506 | 0.926 | 1.10 | 0.86 | 1.40 | 0.468 | 1.22 | 0.84 | 1.77 | 0.299 | -0.03 | -0.11 | 0.05 | 0.412 |
| CCN5 | OID2106 | Neurolog | CCN5 | O76076 | 20 | 44714844 | 44728509 | 8 | 0 | 0 | 0 | 0 | 1.14 | 0.77 | 1.67 | 0.515 | 0.945 | 8.32 | 0.31 | 187.23 | 1.14 | 1.14 | 0.80 | 1.62 | 0.478 | 0.910 | 1.30 | 0.83 | 2.03 | 0.251 | 1.00 | 0.35 | 2.85 | 0.998 | 0.02 | -0.13 | 0.17 | 0.796 |
| PCSK9 | OID2023 | Cardiom | PCSK9 | Q8NBP7 | 1 | 55039447 | 55064852 | 7 | 0 | 0 | 0 | 0 | 0.92 | 0.72 | 1.18 | 0.520 | 0.946 | 4.15 | 0.66 | 342.24 | 0.92 | 0.92 | 0.72 | 1.19 | 0.521 | 0.932 | 0.95 | 0.73 | 1.24 | 0.708 | 1.02 | 0.74 | 1.40 | 0.903 | -0.04 | -0.11 | 0.03 | 0.288 |
| TNFRSF | OID2098 | Neurolog | TNFRSF | Q07011 | 1 | 7915871 | 7943165 | 2 | 0 | 0 | 0 | 0 | 1.20 | 0.69 | 2.06 | 0.520 | 0.946 | 1.78 | 0.18 | 279.36 | 1.20 | 1.20 | 0.69 | 2.07 | 0.520 | 0.932 | - | - | - | - | - | - | - | - | - | - | - | - |
| VWC2L | OID3100 | Neurolog | VWC2L | B2RUY7 | 2 | 214411054 | 214578976 | 2 | 0 | 0 | 0 | 0 | 0.72 | 0.27 | 1.94 | 0.522 | 0.946 | 0.77 | 0.38 | 85.28 | 0.72 | 0.72 | 0.27 | 1.97 | 0.527 | 0.932 | - | - | - | - | - | - | - | - | - | - | - | - |
| SETMAR | OID2094 | Neurolog | SETMAR | Q53H47 | 3 | 4303332 | 4317265 | 8 | 0 | 0 | 0 | 0 | 1.12 | 0.79 | 1.60 | 0.522 | 0.946 | 8.32 | 0.30 | 197.21 | 1.12 | 1.12 | 0.81 | 1.56 | 0.485 | 0.911 | 1.37 | 0.92 | 2.04 | 0.121 | 1.16 | 0.54 | 2.49 | 0.712 | 0.00 | -0.11 | 0.10 | 0.931 |
| MRC1 | OID3069 | Inflamma | MRC1 | P22897 | 10 | 17809348 | 17911164 | 12 | 0 | 0 | 0 | 0 | 0.91 | 0.68 | 1.22 | 0.523 | 0.946 | 3.36 | 0.98 | 170.44 | 0.91 | 0.91 | 0.68 | 1.22 | 0.526 | 0.932 | 0.99 | 0.69 | 1.41 | 0.937 | 0.95 | 0.58 | 1.57 | 0.848 | -0.01 | -0.09 | 0.07 | 0.821 |
| IFNG | OID2049 | Inflamma | IFNG | P01579 | 12 | 68154768 | 68159740 | 1 | 0 | 0 | 0 | 0 | 2.08 | 0.22 | 19.52 | 0.521 | 0.946 | - | - | 30.68 | 2.08 | 2.08 | 0.19 | 22.25 | 0.544 | 0.932 | - | - | - | - | - | - | - | - | - | - | - | - |
| KITLG | OID2019 | Cardiom | KITLG | P21583 | 12 | 88492793 | 88580851 | 1 | 0 | 0 | 0 | 0 | 0.54 | 0.08 | 3.48 | 0.520 | 0.946 | - | - | 48.24 | 0.54 | 0.54 | 0.08 | 3.72 | 0.535 | 0.932 | - | - | - | - | - | - | - | - | - | - | - | - |
| ACP5 | OID2031 | Cardiom | ACP5 | P13686 | 19 | 11574660 | 11579008 | 14 | 0 | 0 | 0 | 0 | 1.07 | 0.87 | 1.33 | 0.522 | 0.946 | 7.79 | 0.86 | 266.75 | 1.07 | 1.07 | 0.86 | 1.33 | 0.523 | 0.932 | 1.03 | 0.79 | 1.35 | 0.812 | 1.00 | 0.67 | 1.49 | 0.991 | 0.02 | -0.07 | 0.11 | 0.692 |
| PVR | OID2101 | Neurolog | PVR | P15151 | 19 | 44643798 | 44666162 | 26 | 0 | 1 | 0 | 0 | 1.09 | 0.83 | 1.43 | 0.521 | 0.946 | 83.70 | 0.00 | 266.11 | 1.09 | 1.09 | 0.94 | 1.27 | 0.234 | 0.859 | 0.99 | 0.81 | 1.21 | 0.925 | 0.89 | 0.56 | 1.41 | 0.618 | 0.07 | -0.05 | 0.19 | 0.283 |
| ADAMTS | OID3027 | Cardiom | ADAMTS | Q6UY14 | 1 | 150549369 | 150560937 | 17 | 0 | 0 | 0 | 0 | 1.09 | 0.82 | 1.43 | 0.560 | 0.946 | 11.63 | 0.77 | 122.67 | 1.09 | 1.09 | 0.82 | 1.43 | 0.562 | 0.934 | 1.27 | 0.88 | 1.84 | 0.206 | 1.07 | 0.54 | 2.11 | 0.852 | 0.00 | -0.14 | 0.15 | 0.957 |
| AKR7L | OID3048 | Inflamma | AKR7L | Q8NHP1 | 1 | 19265982 | 19274194 | 7 | 0 | 0 | 0 | 0 | 1.15 | 0.71 | 1.89 | 0.567 | 0.946 | 8.91 | 0.18 | 135.61 | 1.16 | 1.16 | 0.77 | 1.74 | 0.484 | 0.911 | 1.28 | 0.80 | 2.05 | 0.306 | 2.93 | 1.23 | 6.99 | 0.016 | -0.19 | -0.34 | -0.03 | 0.018 |
| CEP85 | OID2124 | Oncology | CEP85 | Q6P2H3 | 1 | 26234200 | 26278808 | 2 | 0 | 0 | 0 | 0 | 0.75 | 0.29 | 1.96 | 0.564 | 0.946 | 0.44 | 0.51 | 83.74 | 0.75 | 0.75 | 0.29 | 2.00 | 0.570 | 0.934 | - | - | - | - | - | - | - | - | - | - | - | - |
| CREG1 | OID2151 | Oncology | CREG1 | O75629 | 1 | 167529117 | 167553805 | 3 | 0 | 0 | 0 | 0 | 1.16 | 0.72 | 1.87 | 0.546 | 0.946 | 2.84 | 0.24 | 244.84 | 1.16 | 1.16 | 0.72 | 1.87 | 0.546 | 0.932 | 1.10 | 0.68 | 1.79 | 0.696 | 0.69 | 0.29 | 1.61 | 0.386 | 0.10 | -0.04 | 0.24 | 0.145 |
| DDI2 | OID3060 | Inflamma | DDI2 | Q5TDH0 | 1 | 15617458 | 15669044 | 1 | 0 | 0 | 0 | 0 | 1.87 | 0.25 | 13.73 | 0.541 | 0.946 | - | - | 58.08 | 1.87 | 1.87 | 0.24 | 14.57 | 0.552 | 0.933 | - | - | - | - | - | - | - | - | - | - | - | - |
| DNAJC6 | OID3024 | Cardiom | DNAJC6 | O75061 | 1 | 65248219 | 65415871 | 1 | 0 | 0 | 0 | 0 | 0.72 | 0.22 | 2.38 | 0.586 | 0.946 | - | - | 109.52 | 0.72 | 0.72 | 0.21 | 2.42 | 0.592 | 0.934 | - | - | - | - | - | - | - | - | - | - | - | - |
| DRAXIN | OID2091 | Neurolog | DRAXIN | Q8NBI3 | 1 | 11691710 | 11725857 | 13 | 0 | 0 | 0 | 0 | 0.95 | 0.78 | 1.15 | 0.580 | 0.946 | 3.91 | 0.99 | 317.65 | 0.95 | 0.95 | 0.78 | 1.15 | 0.582 | 0.934 | 0.95 | 0.75 | 1.21 | 0.684 | 0.97 | 0.70 | 1.33 | 0.830 | 0.00 | -0.06 | 0.05 | 0.877 |
| EIF4G3 | OID3102 | Neurolog | EIF4G3 | O43432 | 1 | 20806292 | 21177285 | 1 | 0 | 0 | 0 | 0 | 1.48 | 0.40 | 5.56 | 0.559 | 0.946 | - | - | 92.73 | 1.48 | 1.48 | 0.39 | 5.70 | 0.566 | 0.934 | - | - | - | - | - | - | - | - | - | - | - | - |
| HDGF | OID2145 | Oncology | HDGF | P51858 | 1 | 156742109 | 156766925 | 39 | 0 | 0 | 0 | 0 | 1.03 | 0.93 | 1.14 | 0.588 | 0.946 | 42.53 | 0.28 | 413.12 | 1.03 | 1.03 | 0.93 | 1.13 | 0.566 | 0.934 | 0.99 | 0.87 | 1.14 | 0.896 | 1.01 | 0.86 | 1.19 | 0.866 | 0.01 | -0.05 | 0.06 | 0.818 |
| IGSF21 | OID3021 | Cardiom | IGSF21 | Q96ID5 | 1 | 18107798 | 18378483 | 7 | 0 | 0 | 0 | 0 | 0.87 | 0.55 | 1.39 | 0.570 | 0.946 | 3.33 | 0.77 | 107.69 | 0.87 | 0.87 | 0.55 | 1.40 | 0.572 | 0.934 | 0.62 | 0.35 | 1.10 | 0.103 | 0.61 | 0.18 | 2.06 | 0.430 | 0.04 | -0.10 | 0.19 | 0.535 |
| MECR | OID3027 | Cardiom | MECR | Q9BV79 | 1 | 29192657 | 29230942 | 2 | 0 | 0 | 0 | 0 | 1.38 | 0.51 | 3.79 | 0.527 | 0.946 | 1.47 | 0.23 | 68.43 | 1.39 | 1.39 | 0.50 | 3.87 | 0.530 | 0.932 | - | - | - | - | - | - | - | - | - | - | - | - |
| MYOC | OID2112 | Neurolog | MYOC | Q99972 | 1 | 171635417 | 171652688 | 13 | 0 | 0 | 0 | 0 | 1.11 | 0.77 | 1.59 | 0.586 | 0.946 | 24.09 | 0.02 | 202.57 | 1.11 | 1.11 | 0.86 | 1.43 | 0.437 | 0.909 | 0.95 | 0.67 | 1.34 | 0.765 | 0.50 | 0.19 | 1.32 | 0.161 | 0.15 | -0.02 | 0.33 | 0.086 |
| NEXN | OID3067 | Inflamma | NEXN | Q0ZGT2 | 1 | 77888513 | 77943895 | 1 | 0 | 0 | 0 | 0 | 1.48 | 0.43 | 5.15 | 0.538 | 0.946 | - | - | 112.55 | 1.48 | 1.48 | 0.42 | 5.25 | 0.544 | 0.932 | - | - | - | - | - | - | - | - | - | - | - | - |
| PFKFB2 | OID2135 | Oncology | PFKFB2 | O60825 | 1 | 207034366 | 207081024 | 1 | 0 | 0 | 0 | 0 | 1.15 | 0.70 | 1.90 | 0.582 | 0.946 | - | - | 644.17 | 1.15 | 1.15 | 0.70 | 1.90 | 0.583 | 0.934 | - | - | - | - | - | - | - | - | - | - | - | - |
| PGD | OID3034 | Cardiom | PGD | P52209 | 1 | 10398592 | 10420511 | 1 | 0 | 0 | 0 | 0 | 0.58 | 0.08 | 4.19 | 0.586 | 0.946 | - | - | 40.38 | 0.58 | 0.58 | 0.07 | 4.56 | 0.602 | 0.937 | - | - | - | - | - | - | - | - | - | - | - | - |
| SEMA6C | OID3053 | Inflamma | SEMA6C | Q9H3T2 | 1 | 151131685 | 151146631 | 1 | 0 | 0 | 0 | 0 | 1.38 | 0.48 | 3.91 | 0.549 | 0.946 | - | - | 143.19 | 1.38 | 1.38 | 0.48 | 3.96 | 0.554 | 0.933 | - | - | - | - | - | - | - | - | - | - | - | - |
| SORT1 | OID2021 | Cardiom | SORT1 | Q99523 | 1 | 109309568 | 109397918 | 3 | 0 | 0 | 0 | 0 | 0.86 | 0.54 | 1.39 | 0.542 | 0.946 | 2.02 | 0.36 | 229.90 | 0.86 | 0.86 | 0.54 | 1.39 | 0.543 | 0.932 | 0.79 | 0.49 | 1.30 | 0.356 | 0.71 | 0.00 | 132.05 | 0.896 | 0.03 | -0.84 | 0.91 | 0.940 |
| SPRR3 | OID3113 | Neurolog | SPRR3 | Q9UBC9 | 1 | 153001747 | 153003856 | 11 | 0 | 0 | 0 | 0 | 1.11 | 0.77 | 1.60 | 0.586 | 0.946 | 13.31 | 0.21 | 170.36 | 1.11 | 1.11 | 0.80 | 1.53 | 0.530 | 0.932 | 1.17 | 0.80 | 1.69 | 0.416 | 1.15 | 0.57 | 2.30 | 0.695 | -0.01 | -0.12 | 0.11 | 0.901 |
| TNR | OID2095 | Neurolog | TNR | Q92752 | 1 | 175315194 | 175743616 | 14 | 0 | 0 | 0 | 0 | 1.07 | 0.85 | 1.35 | 0.585 | 0.946 | 14.07 | 0.37 | 241.73 | 1.07 | 1.07 | 0.85 | 1.34 | 0.570 | 0.934 | 0.99 | 0.77 | 1.26 | 0.910 | 0.92 | 0.61 | 1.38 | 0.685 | 0.04 | -0.05 | 0.12 | 0.384 |
| TOR1AIP | OID3019 | Cardiom | TOR1AIP | Q5JTV8 | 1 | 179882042 | 179920077 | 34 | 0 | 0 | 0 | 0 | 0.96 | 0.84 | 1.10 | 0.537 | 0.946 | 20.95 | 0.95 | 247.89 | 0.96 | 0.96 | 0.84 | 1.10 | 0.538 | 0.932 | 1.02 | 0.84 | 1.23 | 0.876 | 1.03 | 0.78 | 1.37 | 0.835 | -0.02 | -0.11 | 0.06 | 0.571 |
| VTCN1 | OID2116 | Oncology | VTCN1 | Q7Z7D3 | 1 | 117143587 | 117210960 | 1 | 0 | 0 | 0 | 0 | 0.69 | 0.20 | 2.37 | 0.556 | 0.946 | - | - | 101.67 | 0.69 | 0.69 | 0.20 | 2.42 | 0.563 | 0.934 | - | - | - | - | - | - | - | - | - | - | - | - |
| VWA1 | OID2142 | Oncology | VWA1 | Q6PCB0 | 1 | 1434861 | 1442882 | 6 | 0 | 0 | 0 | 0 | 0.93 | 0.74 | 1.16 | 0.528 | 0.946 | 3.71 | 0.59 | 542.98 | 0.93 | 0.93 | 0.74 | 1.16 | 0.528 | 0.932 | 0.93 | 0.73 | 1.19 | 0.568 | 0.91 | 0.70 | 1.18 | 0.471 | 0.02 | -0.07 | 0.10 | 0.726 |
| ZBTB17 | OID2008 | Cardiom | ZBTB17 | Q13105 | 1 | 15941869 | 15976132 | 1 | 0 | 0 | 0 | 0 | 1.89 | 0.24 | 14.78 | 0.546 | 0.946 | - | - | 37.34 | 1.89 | 1.89 | 0.22 | 16.28 | 0.564 | 0.934 | - | - | - | - | - | - | - | - | - | - | - | - |
| AGXT | OID2013 | Cardiom | AGXT | P21549 | 2 | 240868824 | 240880502 | 6 | 0 | 0 | 0 | 0 | 1.11 | 0.78 | 1.57 | 0.566 | 0.946 | 6.40 | 0.27 | 290.24 | 1.11 | 1.11 | 0.81 | 1.51 | 0.515 | 0.930 | 1.15 | 0.83 | 1.59 | 0.400 | 0.95 | 0.45 | 1.98 | 0.884 | 0.03 | -0.10 | 0.16 | 0.628 |
| ATRAID | OID3145 | Oncology | ATRAID | Q6UW56 | 2 | 27212041 | 27217178 | 21 | 0 | 0 | 0 | 0 | 1.10 | 0.80 | 1.51 | 0.561 | 0.946 | 31.75 | 0.05 | 129.98 | 1.10 | 1.10 | 0.85 | 1.42 | 0.462 | 0.909 | 0.94 | 0.67 | 1.33 | 0.741 | 1.04 | 0.50 | 2.17 | 0.917 | 0.01 | -0.12 | 0.14 | 0.868 |
| ERBB4 | OID2151 | Oncology | ERBB4 | Q15303 | 2 | 211375717 | 212538841 | 12 | 0 | 0 | 0 | 0 | 1.10 | 0.81 | 1.50 | 0.548 | 0.946 | 12.28 | 0.34 | 155.91 | 1.10 | 1.10 | 0.82 | 1.48 | 0.525 | 0.932 | 1.45 | 0.97 | 2.18 | 0.072 | 1.09 | 0.54 | 2.23 | 0.808 | 0.00 | -0.09 | 0.09 | 0.982 |
| FKBP1B | OID2061 | Inflamma | FKBP1B | P68106 | 2 | 24049701 | 24063681 | 6 | 0 | 1 | 0 | 0 | 1.22 | 0.59 | 2.54 | 0.589 | 0.946 | 16.45 | 0.01 | 173.95 | 1.23 | 1.23 | 0.82 | 1.84 | 0.323 | 0.887 | 1.18 | 0.76 | 1.82 | 0.463 | 2.92 | 0.42 | 20.37 | 0.280 | -0.17 | -0.53 | 0.18 | 0.343 |
| GNLY | OID2113 | Neurolog | GNLY | P22749 | 2 | 85685175 | 85698852 | 26 | 0 | 0 | 0 | 0 | 0.95 | 0.81 | 1.12 | 0.575 | 0.946 | 23.00 | 0.58 | 225.11 | 0.95 | 0.95 | 0.81 | 1.12 | 0.575 | 0.934 | 0.93 | 0.74 | 1.17 | 0.546 | 0.97 | 0.75 | 1.25 | 0.791 | 0.00 | -0.07 | 0.06 | 0.909 |
| IL36G | OID3055 | Inflamma | IL36G | Q9NZH8 | 2 | 112973203 | 112985658 | 2 | 0 | 0 | 0 | 0 | 1.39 | 0.50 | 3.92 | 0.530 | 0.946 | 0.35 | 0.55 | 68.98 | 1.39 | 1.39 | 0.48 | 4.02 | 0.538 | 0.932 | - | - | - | - | - | - | - | - | - | - | - | - |
| POMC | OID3028 | Cardiom | POMC | P01189 | 2 | 25160853 | 25168903 | 2 | 0 | 0 | 0 | 0 | 0.72 | 0.25 | 2.07 | 0.537 | 0.946 | 1.52 | 0.22 | 78.41 | 0.71 | 0.71 | 0.24 | 2.09 | 0.540 | 0.932 | - | - | - | - | - | - | - | - | - | - | - | - |
| RBKS | OID2098 | Neurolog | RBKS | Q9H477 | 2 | 27781379 | 27890681 | 8 | 0 | 0 | 0 | 0 | 0.93 | 0.74 | 1.18 | 0.572 | 0.946 | 12.18 | 0.09 | 593.25 | 0.93 | 0.93 | 0.78 | 1.12 | 0.455 | 0.909 | 0.99 | 0.78 | 1.24 | 0.897 | 0.98 | 0.74 | 1.30 | 0.887 | -0.03 | -0.12 | 0.06 | 0.491 |
| REG1A | OID2041 | Cardiom | REG1A | P05451 | 2 | 79120362 | 79123409 | 9 | 0 | 0 | 0 | 0 | 1.07 | 0.84 | 1.37 | 0.566 | 0.946 | 3.58 | 0.89 | 307.33 | 1.07 | 1.07 | 0.84 | 1.37 | 0.567 | 0.934 | 1.14 | 0.86 | 1.51 | 0.375 | 1.32 | 0.88 | 1.98 | 0.178 | -0.05 | -0.13 | 0.03 | 0.210 |
| VIT | OID3033 | Cardiom | VIT | Q6UXI7 | 2 | 36696690 | 36814794 | 31 | 0 | 0 | 0 | 0 | 0.95 | 0.80 | 1.12 | 0.529 | 0.946 | 16.93 | 0.97 | 211.92 | 0.95 | 0.95 | 0.80 | 1.13 | 0.531 | 0.932 | 0.87 | 0.69 | 1.10 | 0.241 | 0.97 | 0.72 | 1.32 | 0.856 | -0.01 | -0.06 | 0.05 | 0.836 |
| ITGB5 | OID2152 | Oncology | ITGB5 | P18084 | 3 | 124761948 | 124901418 | 2 | 0 | 0 | 0 | 0 | 0.75 | 0.27 | 2.06 | 0.579 | 0.946 | 0.16 | 0.69 | 94.81 | 0.75 | 0.75 | 0.27 | 2.10 | 0.585 | 0.934 | - | - | - | - | - | - | - | - | - | - | - | - |
| NAA80 | OID3089 | Neurolog | NAA80 | Q93015 | 3 | 50296402 | 50299416 | 4 | 0 | 0 | 1 | 0 | 1.19 | 0.64 | 2.18 | 0.585 | 0.946 | 0.88 | 0.83 | 108.80 | 1.19 | 1.19 | 0.64 | 2.20 | 0.589 | 0.934 | 1.02 | 0.51 | 2.06 | 0.948 | 0.80 | 0.29 | 2.23 | 0.672 | 0.07 | -0.08 | 0.22 | 0.350 |
| RARRES | OID2117 | Oncology | RARRES | P49788 | 3 | 158696892 | 158732489 | 33 | 0 | 0 | 0 | 0 | 0.95 | 0.81 | 1.12 | 0.558 | 0.946 | 18.28 | 0.98 | 178.63 | 0.95 | 0.95 | 0.81 | 1.12 | 0.559 | 0.934 | 0.96 | 0.76 | 1.22 | 0.750 | 0.95 | 0.70 | 1.27 | 0.710 | 0.00 | -0.07 | 0.07 | 0.944 |
| SERPINI | OID3060 | Inflamma | SERPINI | Q99574 | 3 | 167735243 | 167825569 | 22 | 0 | 0 | 0 | 0 | 1.07 | 0.87 | 1.31 | 0.548 | 0.946 | 21.49 | 0.43 | 170.53 | 1.07 | 1.07 | 0.87 | 1.31 | 0.544 | 0.932 | 1.00 | 0.77 | 1.31 | 0.993 | 0.72 | 0.44 | 1.19 | 0.197 | 0.10 | -0.02 | 0.21 | 0.090 |
| ANXA5 | OID2081 | Neurolog | ANXA5 | P08758 | 4 | 121667946 | 121696995 | 1 | 0 | 0 | 0 | 0 | 1.29 | 0.53 | 3.15 | 0.577 | 0.946 | - | - | 190.76 | 1.29 | 1.29 | 0.52 | 3.18 | 0.580 | 0.934 | - | - | - | - | - | - | - | - | - | - | - | - |
| BANK1 | OID2059 | Inflamma | BANK1 | Q8NDB2 | 4 | 101411286 | 102074812 | 4 | 0 | 0 | 1 | 0 | 0.76 | 0.31 | 1.83 | 0.537 | 0.946 | 1.32 | 0.72 | 48.59 | 0.76 | 0.76 | 0.31 | 1.87 | 0.546 | 0.932 | 0.72 | 0.26 | 2.02 | 0.534 | 0.54 | 0.10 | 2.89 | 0.469 | 0.04 | -0.13 | 0.21 | 0.637 |
| EPGN | OID3137 | Oncology | EPGN | Q6UW88 | 4 | 74308470 | 74316789 | 1 | 0 | 0 | 0 | 0 | 0.52 | 0.07 | 3.95 | 0.525 | 0.946 | - | - | 36.55 | 0.52 | 0.52 | 0.06 | 4.36 | 0.544 | 0.932 | - | - | - | - | - | - | - | - | - | - | - | - |
| FAM13A | OID3146 | Oncology | FAM13A | O94988 | 4 | 88725955 | 89111398 | 4 | 0 | 0 | 0 | 0 | 1.15 | 0.71 | 1.85 | 0.575 | 0.946 | 2.75 | 0.43 | 168.58 | 1.15 | 1.15 | 0.71 | 1.86 | 0.576 | 0.934 | 1.15 | 0.65 | 2.05 | 0.622 | 2.43 | 0.31 | 19.31 | 0.402 | -0.14 | -0.50 | 0.23 | 0.465 |
| HPSE | OID3040 | Cardiom | HPSE | Q9Y251 | 4 | 83292461 | 83335153 | 2 | 0 | 0 | 0 | 0 | 1.20 | 0.67 | 2.17 | 0.541 | 0.946 | 0.02 | 0.89 | 231.40 | 1.20 | 1.20 | 0.66 | 2.18 | 0.544 | 0.932 | - | - | - | - | - | - | - | - | - | - | - | - |
| NFKB1 | OID3016 | Cardiom | NFKB1 | P19838 | 4 | 102501330 | 102617302 | 1 | 0 | 0 | 0 | 0 | 1.26 | 0.54 | 2.91 | 0.590 | 0.946 | - | - | 224.12 | 1.26 | 1.26 | 0.54 | 2.93 | 0.593 | 0.934 | - | - | - | - | - | - | - | - | - | - | - | - |
| SCRG1 | OID3033 | Cardiom | SCRG1 | O75711 | 4 | 173384701 | 173406380 | 3 | 0 | 0 | 0 | 0 | 1.26 | 0.61 | 2.60 | 0.525 | 0.946 | 0.18 | 0.91 | 130.65 | 1.26 | 1.26 | 0.61 | 2.62 | 0.530 | 0.932 | 1.29 | 0.60 | 2.74 | 0.514 | 0.92 | 0.13 | 6.43 | 0.932 | 0.03 | -0.16 | 0.22 | 0.729 |
| SORCS2 | OID2126 | Oncology | SORCS2 | Q96PQ0 | 4 | 7192538 | 7742836 | 27 | 0 | 0 | 0 | 0 | 0.94 | 0.77 | 1.16 | 0.585 | 0.946 | 14.38 | 0.97 | 146.93 | 0.94 | 0.94 | 0.77 | 1.16 | 0.587 | 0.934 | 0.81 | 0.61 | 1.07 | 0.142 | 0.79 | 0.57 | 1.11 | 0.176 | 0.03 | -0.01 | 0.07 | 0.199 |
| TACC3 | OID2128 | Oncology | TACC3 | Q9Y6A5 | 4 | 1712858 | 1745171 | 4 | 0 | 0 | 0 | 0 | 1.13 | 0.77 | 1.67 | 0.533 | 0.946 | 1.77 | 0.62 | 261.68 | 1.13 | 1.13 | 0.77 | 1.67 | 0.535 | 0.932 | 1.29 | 0.84 | 1.97 | 0.239 | 2.96 | 0.64 | 13.81 | 0.166 | -0.17 | -0.43 | 0.09 | 0.205 |
| EDIL3 | OID2008 | Cardiom | EDIL3 | O43854 | 5 | 83940554 | 84384880 | 2 | 0 | 0 | 0 | 0 | 0.75 | 0.27 | 2.07 | 0.585 | 0.946 | 0.56 | 0.45 | 77.40 | 0.75 | 0.75 | 0.27 | 2.11 | 0.590 | 0.934 | - | - | - | - | - | - | - | - | - | - | - | - |
| F2R | OID2069 | Inflamma | F2R | P25116 | 5 | 76716126 | 76735770 | 2 | 0 | 0 | 0 | 0 | 0.81 | 0.39 | 1.66 | 0.560 | 0.946 | 6.07 | 0.01 | 142.90 | 0.80 | 0.80 | 0.39 | 1.66 | 0.553 | 0.933 | - | - | - | - | - | - | - | - | - | - | - | - |
| OCLN | OID3106 | Neurolog | OCLN | Q16625 | 5 | 69492292 | 69558104 | 4 | 0 | 0 | 1 | 0 | 1.20 | 0.65 | 2.20 | 0.563 | 0.946 | 2.77 | 0.43 | 122.29 | 1.20 | 1.20 | 0.65 | 2.21 | 0.565 | 0.934 | 1.36 | 0.67 | 2.80 | 0.397 | 2.87 | 0.83 | 10.00 | 0.097 | -0.12 | -0.27 | 0.03 | 0.115 |
| SNX2 | OID3137 | Oncology | SNX2 | O60749 | 5 | 122775079 | 122834543 | 1 | 0 | 0 | 0 | 0 | 0.76 | 0.28 | 2.06 | 0.590 | 0.946 | - | - | 158.50 | 0.76 | 0.76 | 0.28 | 2.08 | 0.594 | 0.934 | - | - | - | - | - | - | - | - | - | - | - | - |
| C2 | OID2041 | Cardiom | C2 | P06681 | 6 | 31897785 | 31945672 | 21 | 0 | 0 | 0 | 0 | 0.94 | 0.78 | 1.14 | 0.545 | 0.946 | 14.79 | 0.79 | 207.12 | 0.94 | 0.94 | 0.78 | 1.14 | 0.546 | 0.932 | 0.85 | 0.66 | 1.11 | 0.246 | 0.81 | 0.54 | 1.22 | 0.315 | 0.03 | -0.05 | 0.12 | 0.414 |
| CRISP3 | OID3071 | Inflamma | CRISP3 | P54108 | 6 | 49727376 | 49744437 | 44 | 0 | 0 | 0 | 0 | 1.03 | 0.92 | 1.16 | 0.588 | 0.946 | 40.12 | 0.60 | 269.84 | 1.03 | 1.03 | 0.92 | 1.16 | 0.588 | 0.934 | 0.94 | 0.79 | 1.13 | 0.534 | 1.10 | 0.86 | 1.41 | 0.429 | -0.02 | -0.09 | 0.05 | 0.545 |
| LEG1 | OID3060 | Inflamma | C6orf58 | Q6P5S2 | 6 | 127519455 | 127591820 | 16 | 0 | 0 | 0 | 0 | 1.08 | 0.85 | 1.37 | 0.540 | 0.946 | 9.65 | 0.84 | 181.29 | 1.08 | 1.08 | 0.85 | 1.37 | 0.542 | 0.932 | 1.07 | 0.80 | 1.44 | 0.657 | 1.04 | 0.60 | 1.80 | 0.882 | 0.01 | -0.10 | 0.11 | 0.895 |
| CCL26 | OID2054 | Inflamma | CCL26 | Q9Y258 | 7 | 75769533 | 75789896 | 6 | 0 | 0 | 0 | 0 | 1.16 | 0.71 | 1.90 | 0.550 | 0.946 | 0.91 | 0.97 | 116.08 | 1.16 | 1.16 | 0.71 | 1.91 | 0.554 | 0.933 | 1.04 | 0.58 | 1.86 | 0.894 | 1.52 | 0.58 | 3.99 | 0.397 | -0.04 | -0.15 | 0.08 | 0.529 |
| DBNL | OID2068 | Inflamma | DBNL | Q9UJU6 | 7 | 44044640 | 44069456 | 1 | 0 | 0 | 0 | 0 | 1.61 | 0.29 | 9.01 | 0.590 | 0.946 | - | - | 52.77 | 1.61 | 1.61 | 0.27 | 9.52 | 0.602 | 0.937 | - | - | - | - | - | - | - | - | - | - | - | - |
| GPNMB | OID2017 | Cardiom | GPNMB | Q14956 | 7 | 23235967 | 23275108 | 15 | 0 | 0 | 0 | 0 | 1.07 | 0.87 | 1.31 | 0.546 | 0.946 | 10.54 | 0.72 | 243.72 | 1.07 | 1.07 | 0.87 | 1.31 | 0.546 | 0.932 | 1.02 | 0.80 | 1.30 | 0.873 | 1.02 | 0.78 | 1.34 | 0.885 | 0.01 | -0.04 | 0.07 | 0.635 |
| LAT2 | OID2129 | Oncology | LAT2 | Q9GZY6 | 7 | 74199652 | 74229834 | 1 | 0 | 0 | 0 | 0 | 1.93 | 0.20 | 18.70 | 0.572 | 0.946 | - | - | 31.51 | 1.93 | 1.93 | 0.17 | 21.21 | 0.592 | 0.934 | - | - | - | - | - | - | - | - | - | - | - | - |
| VGF | OID3106 | Neurolog | VGF | O15240 | 7 | 101162509 | 101165569 | 2 | 0 | 0 | 0 | 0 | 1.45 | 0.42 | 5.06 | 0.556 | 0.946 | 0.02 | 0.88 | 54.98 | 1.45 | 1.45 | 0.40 | 5.25 | 0.567 | 0.934 | - | - | - | - | - | - | - | - | - | - | - | - |
| CDH17 | OID2013 | Cardiom | CDH17 | Q12864 | 8 | 94127162 | 94217303 | 20 | 0 | 0 | 0 | 0 | 1.07 | 0.85 | 1.35 | 0.568 | 0.946 | 18.07 | 0.52 | 182.27 | 1.07 | 1.07 | 0.85 | 1.35 | 0.568 | 0.934 | 1.16 | 0.85 | 1.58 | 0.351 | 1.11 | 0.63 | 1.96 | 0.716 | -0.01 | -0.12 | 0.10 | 0.884 |
| CRH | OID2125 | Oncology | CRH | P06850 | 8 | 66176376 | 66178464 | 12 | 0 | 0 | 0 | 0 | 0.91 | 0.69 | 1.21 | 0.533 | 0.946 | 9.06 | 0.62 | 170.19 | 0.91 | 0.91 | 0.69 | 1.21 | 0.534 | 0.932 | 0.91 | 0.65 | 1.28 | 0.580 | 1.09 | 0.72 | 1.65 | 0.674 | -0.05 | -0.13 | 0.03 | 0.253 |
| FABP9 | OID2050 | Inflamma | FABP9 | Q0Z7S8 | 8 | 81458253 | 81461579 | 3 | 0 | 0 | 1 | 0 | 1.25 | 0.56 | 2.77 | 0.589 | 0.946 | 0.57 | 0.75 | 96.52 | 1.25 | 1.25 | 0.55 | 2.80 | 0.594 | 0.934 | 1.20 | 0.49 | 2.95 | 0.692 | 1.00 | 0.19 | 5.19 | 0.997 | 0.02 | -0.13 | 0.18 | 0.762 |
| IL7 | OID2053 | Inflamma | IL7 | P13232 | 8 | 78675743 | 78805523 | 2 | 0 | 0 | 0 | 0 | 1.30 | 0.56 | 3.04 | 0.541 | 0.946 | 1.20 | 0.27 | 116.22 | 1.30 | 1.30 | 0.55 | 3.08 | 0.544 | 0.932 | - | - | - | - | - | - | - | - | - | - | - | - |
| MSRA | OID2151 | Oncology | MSRA | Q9UJ68 | 8 | 10054292 | 10428891 | 1 | 0 | 0 | 0 | 0 | 0.62 | 0.12 | 3.31 | 0.575 | 0.946 | - | - | 56.24 | 0.62 | 0.62 | 0.11 | 3.48 | 0.586 | 0.934 | - | - | - | - | - | - | - | - | - | - | - | - |
| PLPBP | OID2014 | Cardiom | PLPBP | O94903 | 8 | 37762595 | 37779768 | 1 | 0 | 0 | 0 | 0 | 0.52 | 0.05 | 4.95 | 0.570 | 0.946 | - | - | 32.52 | 0.52 | 0.52 | 0.05 | 5.59 | 0.590 | 0.934 | - | - | - | - | - | - | - | - | - | - | - | - |
| RIDA | OID3063 | Inflamma | RIDA | P52758 | 8 | 98102344 | 98117171 | 5 | 0 | 0 | 0 | 0 | 1.11 | 0.77 | 1.60 | 0.569 | 0.946 | 5.21 | 0.27 | 328.88 | 1.11 | 1.11 | 0.81 | 1.53 | 0.516 | 0.930 | 1.12 | 0.80 | 1.56 | 0.500 | 1.11 | 0.55 | 2.22 | 0.774 | 0.00 | -0.13 | 0.13 | 0.990 |
| SCARA5 | OID2105 | Neurolog | SCARA5 | Q6ZMJ2 | 8 | 27869883 | 27992673 | 11 | 0 | 0 | 0 | 0 | 0.93 | 0.72 | 1.21 | 0.589 | 0.946 | 7.08 | 0.72 | 237.33 | 0.93 | 0.93 | 0.72 | 1.21 | 0.590 | 0.934 | 0.87 | 0.64 | 1.17 | 0.344 | 0.84 | 0.54 | 1.29 | 0.424 | 0.02 | -0.05 | 0.09 | 0.552 |
| TNFRSF1 | OID2073 | Inflamma | TNFRSF1 | O00300 | 8 | 118923557 | 118951885 | 13 | 0 | 0 | 0 | 0 | 0.89 | 0.63 | 1.27 | 0.536 | 0.946 | 11.93 | 0.45 | 120.50 | 0.89 | 0.89 | 0.63 | 1.28 | 0.537 | 0.932 | 0.95 | 0.61 | 1.47 | 0.813 | 1.35 | 0.63 | 2.86 | 0.438 | -0.06 | -0.16 | 0.04 | 0.229 |
| DPP7 | OID2014 | Cardiom | DPP7 | Q9UHL4 | 9 | 137110546 | 137115177 | 23 | 0 | 0 | 0 | 0 | 1.07 | 0.87 | 1.31 | 0.531 | 0.946 | 11.96 | 0.96 | 171.99 | 1.07 | 1.07 | 0.87 | 1.31 | 0.533 | 0.932 | 0.94 | 0.71 | 1.25 | 0.668 | 1.08 | 0.78 | 1.51 | 0.631 | 0.00 | -0.06 | 0.06 | 0.901 |
| MAMDC4 | OID3134 | Oncology | MAMDC4 | Q6UXC1 | 9 | 136850943 | 136860799 | 5 | 0 | 0 | 0 | 0 | 1.10 | 0.80 | 1.52 | 0.558 | 0.946 | 1.05 | 0.90 | 335.55 | 1.10 | 1.10 | 0.80 | 1.53 | 0.559 | 0.934 | 1.14 | 0.80 | 1.62 | 0.478 | 1.21 | 0.82 | 1.80 | 0.331 | -0.04 | -0.13 | 0.05 | 0.387 |
| PTGR1 | OID3029 | Cardiom | PTGR1 | Q14914 | 9 | 111549722 | 111599893 | 20 | 0 | 0 | 0 | 0 | 1.05 | 0.89 | 1.24 | 0.539 | 0.946 | 11.81 | 0.89 | 296.87 | 1.05 | 1.05 | 0.89 | 1.24 | 0.540 | 0.932 | 1.09 | 0.88 | 1.34 | 0.426 | 0.97 | 0.77 | 1.22 | 0.804 | 0.03 | -0.03 | 0.09 | 0.322 |
| SPINK4 | OID2050 | Inflamma | SPINK4 | O60575 | 9 | 33218365 | 33248567 | 28 | 0 | 0 | 0 | 0 | 0.96 | 0.85 | 1.09 | 0.548 | 0.946 | 24.74 | 0.59 | 362.30 | 0.96 | 0.96 | 0.85 | 1.09 | 0.549 | 0.932 | 0.97 | 0.81 | 1.15 | 0.687 | 1.02 | 0.84 | 1.23 | 0.834 | -0.02 | -0.07 | 0.03 | 0.410 |
| ASAH2 | OID2099 | Neurolog | ASAH2 | Q9NR71 | 10 | 50182778 | 50279720 | 43 | 0 | 0 | 0 | 0 | 1.03 | 0.93 | 1.15 | 0.530 | 0.946 | 28.76 | 0.94 | 328.62 | 1.03 | 1.03 | 0.93 | 1.15 | 0.531 | 0.932 | 0.96 | 0.83 | 1.11 | 0.560 | 0.96 | 0.80 | 1.14 | 0.621 | 0.03 | -0.03 | 0.09 | 0.292 |
| CXCL12 | OID2046 | Inflamma | CXCL12 | P48061 | 10 | 44370165 | 44386493 | 6 | 0 | 0 | 0 | 0 | 0.87 | 0.53 | 1.44 | 0.590 | 0.946 | 7.77 | 0.17 | 152.60 | 0.87 | 0.87 | 0.58 | 1.31 | 0.499 | 0.920 | 1.08 | 0.68 | 1.70 | 0.747 | 1.21 | 0.33 | 4.42 | 0.777 | -0.05 | -0.22 | 0.13 | 0.587 |
| PLAU | OID2112 | Neurolog | PLAU | P00749 | 10 | 73909177 | 73917496 | 6 | 0 | 0 | 1 | 0 | 1.20 | 0.65 | 2.21 | 0.552 | 0.946 | 3.06 | 0.69 | 75.62 | 1.20 | 1.20 | 0.65 | 2.24 | 0.556 | 0.934 | 1.59 | 0.74 | 3.45 | 0.236 | 0.38 | 0.06 | 2.49 | 0.313 | 0.17 | -0.09 | 0.42 | 0.204 |
| CASP1 | OID2085 | Neurolog | CASP1 | P29466 | 11 | 105025443 | 105035250 | 1 | 0 | 0 | 0 | 0 | 1.81 | 0.27 | 11.99 | 0.539 | 0.946 | - | - | 43.86 | 1.81 | 1.81 | 0.25 | 12.93 | 0.555 | 0.934 | - | - | - | - | - | - | - | - | - | - | - | - |
| CD248 | OID3039 | Cardiom | CD248 | Q9HCU0 | 11 | 66314494 | 66317044 | 4 | 0 | 0 | 1 | 0 | 0.77 | 0.32 | 1.87 | 0.570 | 0.946 | 3.03 | 0.39 | 67.85 | 0.77 | 0.77 | 0.31 | 1.89 | 0.572 | 0.934 | 1.01 | 0.33 | 3.10 | 0.984 | 0.35 | 0.08 | 1.41 | 0.139 | 0.11 | -0.04 | 0.25 | 0.151 |
| CTSD | OID2035 | Cardiom | CTSD | P07339 | 11 | 1752752 | 1764573 | 10 | 0 | 0 | 0 | 0 | 1.08 | 0.83 | 1.40 | 0.574 | 0.946 | 4.13 | 0.90 | 236.88 | 1.08 | 1.08 | 0.83 | 1.40 | 0.575 | 0.934 | 0.95 | 0.69 | 1.31 | 0.764 | 0.91 | 0.57 | 1.46 | 0.700 | 0.04 | -0.05 | 0.13 | 0.407 |
| JAM3 | OID3058 | Inflamma | JAM3 | Q9BX67 | 11 | 134069071 | 134152001 | 3 | 0 | 0 | 0 | 0 | 0.80 | 0.37 | 1.74 | 0.578 | 0.946 | 1.66 | 0.44 | 89.74 | 0.80 | 0.80 | 0.37 | 1.75 | 0.581 | 0.934 | 0.70 | 0.30 | 1.66 | 0.424 | 1.25 | 0.13 | 11.71 | 0.842 | -0.04 | -0.24 | 0.16 | 0.668 |
| LAYN | OID2097 | Neurolog | LAYN | Q6UX15 | 11 | 111540280 | 111561745 | 16 | 0 | 0 | 0 | 0 | 1.08 | 0.84 | 1.37 | 0.558 | 0.946 | 18.64 | 0.23 | 220.58 | 1.08 | 1.08 | 0.86 | 1.34 | 0.513 | 0.930 | 0.94 | 0.70 | 1.27 | 0.686 | 0.68 | 0.33 | 1.40 | 0.293 | 0.11 | -0.06 | 0.28 | 0.186 |
| MMP1 | OID2067 | Inflamma | MMP1 | P03956 | 11 | 102789401 | 102798160 | 36 | 0 | 0 | 0 | 0 | 0.96 | 0.83 | 1.11 | 0.588 | 0.946 | 29.35 | 0.74 | 210.21 | 0.96 | 0.96 | 0.83 | 1.11 | 0.588 | 0.934 | 1.02 | 0.82 | 1.28 | 0.842 | 0.90 | 0.67 | 1.21 | 0.476 | 0.02 | -0.05 | 0.09 | 0.605 |
| NCR3LG | OID3149 | Oncology | NCR3LG | Q68D85 | 11 | 17351800 | 17377341 | 12 | 0 | 0 | 0 | 0 | 0.93 | 0.71 | 1.21 | 0.587 | 0.946 | 8.04 | 0.71 | 217.29 | 0.93 | 0.93 | 0.71 | 1.21 | 0.588 | 0.934 | 0.99 | 0.71 | 1.37 | 0.944 | 1.20 | 0.70 | 2.05 | 0.503 | -0.05 | -0.15 | 0.04 | 0.276 |
| PAMR1 | OID2115 | Neurolog | PAMR1 | Q6UXH9 | 11 | 35431823 | 35530300 | 20 | 0 | 0 | 0 | 0 | 0.95 | 0.80 | 1.13 | 0.567 | 0.946 | 18.60 | 0.48 | 265.02 | 0.95 | 0.95 | 0.80 | 1.13 | 0.566 | 0.934 | 0.89 | 0.72 | 1.11 | 0.313 | 0.85 | 0.64 | 1.13 | 0.275 | 0.03 | -0.03 | 0.08 | 0.348 |
| PPME1 | OID2134 | Oncology | PPME1 | Q9Y570 | 11 | 74171267 | 74254703 | 1 | 0 | 0 | 0 | 0 | 1.96 | 0.24 | 16.22 | 0.533 | 0.946 | - | - | 39.19 | 1.96 | 1.96 | 0.22 | 17.83 | 0.551 | 0.933 | - | - | - | - | - | - | - | - | - | - | - | - |
| SAA4 | OID3072 | Inflamma | SAA4 | P35542 | 11 | 18231355 | 18236802 | 11 | 0 | 0 | 0 | 0 | 0.92 | 0.70 | 1.21 | 0.559 | 0.946 | 14.03 | 0.17 | 278.37 | 0.92 | 0.92 | 0.73 | 1.16 | 0.488 | 0.912 | 0.82 | 0.61 | 1.10 | 0.182 | 0.74 | 0.46 | 1.19 | 0.220 | 0.06 | -0.05 | 0.16 | 0.278 |
| SWAP70 | OID3147 | Oncology | SWAP70 | Q9UH65 | 11 | 9664077 | 9752993 | 6 | 0 | 0 | 0 | 0 | 1.16 | 0.70 | 1.92 | 0.573 | 0.946 | 0.59 | 0.99 | 100.87 | 1.16 | 1.16 | 0.69 | 1.94 | 0.578 | 0.934 | 1.09 | 0.61 | 1.95 | 0.769 | 1.10 | 0.39 | 3.15 | 0.852 | 0.01 | -0.16 | 0.18 | 0.921 |
| TMPRSS | OID2094 | Neurolog | TMPRSS | Q9H3S3 | 11 | 113687547 | 113706373 | 36 | 0 | 0 | 0 | 0 | 0.96 | 0.85 | 1.09 | 0.566 | 0.946 | 32.57 | 0.59 | 280.49 | 0.96 | 0.96 | 0.85 | 1.09 | 0.566 | 0.934 | 0.98 | 0.81 | 1.19 | 0.835 | 0.99 | 0.82 | 1.20 | 0.943 | -0.01 | -0.06 | 0.04 | 0.677 |
| TPP1 | OID2075 | Inflamma | TPP1 | O14773 | 11 | 6612768 | 6619448 | 4 | 0 | 0 | 0 | 0 | 1.18 | 0.69 | 2.03 | 0.544 | 0.946 | 0.40 | 0.94 | 142.06 | 1.18 | 1.18 | 0.69 | 2.04 | 0.548 | 0.932 | 1.21 | 0.67 | 2.18 | 0.521 | 1.12 | 0.40 | 3.12 | 0.830 | 0.01 | -0.12 | 0.14 | 0.902 |
| ZPR1 | OID3080 | Neurolog | ZPR1 | O75312 | 11 | 116773799 | 116788039 | 1 | 0 | 0 | 0 | 0 | 0.58 | 0.08 | 4.05 | 0.585 | 0.946 | - | - | 43.14 | 0.58 | 0.58 | 0.08 | 4.37 | 0.600 | 0.937 | - | - | - | - | - | - | - | - | - | - | - | - |
| ADGRD1 | OID3058 | Inflamma | ADGRD1 | Q6QNK2 | 12 | 130953907 | 131141469 | 19 | 0 | 0 | 0 | 0 | 0.93 | 0.74 | 1.17 | 0.560 | 0.946 | 19.73 | 0.35 | 206.37 | 0.93 | 0.93 | 0.75 | 1.16 | 0.541 | 0.932 | 0.91 | 0.67 | 1.23 | 0.549 | 1.01 | 0.65 | 1.57 | 0.947 | -0.02 | -0.08 | 0.05 | 0.662 |
| CD27 | OID2152 | Oncology | CD27 | P26842 | 12 | 6444955 | 6451718 | 12 | 0 | 0 | 0 | 0 | 1.09 | 0.80 | 1.50 | 0.584 | 0.946 | 13.19 | 0.28 | 159.28 | 1.09 | 1.09 | 0.82 | 1.46 | 0.549 | 0.932 | 1.15 | 0.81 | 1.63 | 0.427 | 1.04 | 0.59 | 1.82 | 0.904 | 0.01 | -0.08 | 0.10 | 0.817 |
| CIT | OID3097 | Neurolog | CIT | O14578 | 12 | 119685791 | 119877320 | 2 | 0 | 0 | 0 | 0 | 1.50 | 0.38 | 5.98 | 0.562 | 0.946 | 0.57 | 0.45 | 41.11 | 1.51 | 1.51 | 0.36 | 6.31 | 0.574 | 0.934 | - | - | - | - | - | - | - | - | - | - | - | - |
| MDM1 | OID3094 | Neurolog | MDM1 | Q8TC05 | 12 | 68272443 | 68332381 | 1 | 0 | 0 | 0 | 0 | 1.15 | 0.69 | 1.92 | 0.590 | 0.946 | - | - | 566.16 | 1.15 | 1.15 | 0.69 | 1.93 | 0.591 | 0.934 | - | - | - | - | - | - | - | - | - | - | - | - |
| PZP | OID3073 | Inflamma | PZP | P20742 | 12 | 9148840 | 9208395 | 28 | 0 | 0 | 0 | 0 | 1.06 | 0.85 | 1.32 | 0.584 | 0.946 | 24.06 | 0.63 | 219.03 | 1.06 | 1.06 | 0.85 | 1.32 | 0.584 | 0.934 | 1.05 | 0.79 | 1.40 | 0.730 | 0.95 | 0.65 | 1.39 | 0.799 | 0.02 | -0.04 | 0.09 | 0.484 |
| FLT3 | OID2127 | Oncology | FLT3 | P36888 | 13 | 28003274 | 28100592 | 1 | 0 | 0 | 0 | 0 | 0.71 | 0.22 | 2.24 | 0.554 | 0.946 | - | - | 114.88 | 0.71 | 0.71 | 0.22 | 2.27 | 0.560 | 0.934 | - | - | - | - | - | - | - | - | - | - | - | - |
| PIBF1 | OID3106 | Neurolog | PIBF1 | Q8WXW3 | 13 | 72782133 | 73016461 | 1 | 0 | 0 | 0 | 0 | 1.60 | 0.33 | 7.68 | 0.560 | 0.946 | - | - | 64.96 | 1.60 | 1.60 | 0.32 | 8.01 | 0.570 | 0.934 | - | - | - | - | - | - | - | - | - | - | - | - |
| SLITRK6 | OID2009 | Cardiom | SLITRK6 | Q9H5Y7 | 13 | 85792790 | 85806683 | 12 | 0 | 0 | 0 | 0 | 0.88 | 0.58 | 1.33 | 0.550 | 0.946 | 19.02 | 0.06 | 126.30 | 0.88 | 0.88 | 0.64 | 1.21 | 0.428 | 0.909 | 0.68 | 0.44 | 1.05 | 0.082 | 0.53 | 0.26 | 1.11 | 0.093 | 0.07 | -0.02 | 0.17 | 0.114 |
| SPRY2 | OID2047 | Inflamma | SPRY2 | O43597 | 13 | 80335976 | 80341126 | 2 | 0 | 0 | 0 | 0 | 1.36 | 0.51 | 3.66 | 0.542 | 0.946 | 0.80 | 0.37 | 80.50 | 1.36 | 1.36 | 0.50 | 3.73 | 0.547 | 0.932 | - | - | - | - | - | - | - | - | - | - | - | - |
| APEX1 | OID2143 | Oncology | APEX1 | P27695 | 14 | 20455191 | 20457772 | 6 | 0 | 0 | 0 | 0 | 1.07 | 0.85 | 1.34 | 0.580 | 0.946 | 1.97 | 0.85 | 521.76 | 1.07 | 1.07 | 0.85 | 1.34 | 0.581 | 0.934 | 1.05 | 0.83 | 1.34 | 0.666 | 1.00 | 0.70 | 1.42 | 0.984 | 0.02 | -0.06 | 0.10 | 0.613 |
| PNMA1 | OID3019 | Cardiom | PNMA1 | Q8ND90 | 14 | 73711783 | 73714384 | 2 | 0 | 0 | 0 | 0 | 1.38 | 0.46 | 4.14 | 0.569 | 0.946 | 0.39 | 0.53 | 70.52 | 1.38 | 1.38 | 0.45 | 4.25 | 0.576 | 0.934 | - | - | - | - | - | - | - | - | - | - | - | - |
| SERPINA | OID3074 | Inflamma | SERPINA | P08185 | 14 | 94304248 | 94323389 | 6 | 0 | 0 | 0 | 0 | 1.12 | 0.77 | 1.63 | 0.557 | 0.946 | 1.11 | 0.95 | 222.26 | 1.12 | 1.12 | 0.77 | 1.64 | 0.560 | 0.934 | 1.03 | 0.69 | 1.56 | 0.872 | 0.99 | 0.48 | 2.04 | 0.985 | 0.02 | -0.08 | 0.11 | 0.701 |
| PPCDC | OID2102 | Neurolog | PPCDC | Q96CD2 | 15 | 75023586 | 75117462 | 12 | 0 | 0 | 0 | 0 | 0.94 | 0.76 | 1.15 | 0.533 | 0.946 | 6.01 | 0.87 | 325.52 | 0.94 | 0.94 | 0.76 | 1.15 | 0.534 | 0.932 | 1.00 | 0.78 | 1.29 | 0.986 | 1.07 | 0.76 | 1.49 | 0.705 | -0.05 | -0.14 | 0.05 | 0.338 |
| AGRP | OID2065 | Inflamma | AGRP | O00253 | 16 | 67482571 | 67483547 | 6 | 0 | 0 | 1 | 0 | 0.81 | 0.42 | 1.59 | 0.544 | 0.946 | 1.73 | 0.89 | 60.04 | 0.81 | 0.81 | 0.41 | 1.61 | 0.551 | 0.933 | 0.73 | 0.32 | 1.66 | 0.452 | 0.90 | 0.27 | 2.98 | 0.858 | -0.02 | -0.23 | 0.19 | 0.848 |
| CA5A | OID2007 | Cardiom | CA5A | P35218 | 16 | 87881546 | 87936580 | 21 | 0 | 0 | 0 | 0 | 0.94 | 0.78 | 1.15 | 0.568 | 0.946 | 23.79 | 0.25 | 235.10 | 0.94 | 0.94 | 0.79 | 1.13 | 0.533 | 0.932 | 0.98 | 0.78 | 1.24 | 0.864 | 1.01 | 0.74 | 1.37 | 0.965 | -0.02 | -0.08 | 0.04 | 0.592 |
| CDH3 | OID2100 | Neurolog | CDH3 | P22223 | 16 | 68636189 | 68727468 | 3 | 0 | 0 | 1 | 0 | 1.34 | 0.53 | 3.42 | 0.539 | 0.946 | 2.38 | 0.30 | 61.48 | 1.34 | 1.34 | 0.52 | 3.49 | 0.543 | 0.932 | 1.77 | 0.63 | 5.02 | 0.279 | 0.78 | 0.02 | 29.12 | 0.895 | 0.05 | -0.25 | 0.34 | 0.753 |
| CNTNAP | OID3091 | Neurolog | CNTNAP | Q9C0A0 | 16 | 76277278 | 76560757 | 1 | 0 | 0 | 0 | 0 | 1.79 | 0.25 | 12.79 | 0.560 | 0.946 | - | - | 38.55 | 1.79 | 1.79 | 0.23 | 13.98 | 0.577 | 0.934 | - | - | - | - | - | - | - | - | - | - | - | - |
| CTF1 | OID2006 | Cardiom | CTF1 | Q16619 | 16 | 30896614 | 30903547 | 1 | 0 | 0 | 0 | 0 | 1.67 | 0.27 | 10.21 | 0.580 | 0.946 | - | - | 46.10 | 1.67 | 1.67 | 0.25 | 10.92 | 0.594 | 0.934 | - | - | - | - | - | - | - | - | - | - | - | - |
| HSBP1 | OID3023 | Cardiom | HSBP1 | O75506 | 16 | 83719311 | 83819737 | 24 | 0 | 0 | 0 | 0 | 0.95 | 0.80 | 1.13 | 0.549 | 0.946 | 27.48 | 0.24 | 264.83 | 0.95 | 0.95 | 0.81 | 1.11 | 0.512 | 0.930 | 0.95 | 0.77 | 1.16 | 0.592 | 1.21 | 0.89 | 1.64 | 0.230 | -0.06 | -0.13 | 0.01 | 0.071 |
| IL32 | OID2060 | Inflamma | IL32 | P24001 | 16 | 3065297 | 3082192 | 9 | 0 | 0 | 0 | 0 | 1.11 | 0.78 | 1.58 | 0.577 | 0.946 | 10.35 | 0.24 | 186.31 | 1.11 | 1.11 | 0.81 | 1.52 | 0.524 | 0.932 | 1.09 | 0.76 | 1.59 | 0.632 | 0.97 | 0.55 | 1.71 | 0.923 | 0.03 | -0.06 | 0.11 | 0.551 |
| JPT2 | OID3152 | Oncology | JPT2 | Q9H910 | 16 | 1678256 | 1702280 | 1 | 0 | 0 | 0 | 0 | 0.58 | 0.09 | 3.85 | 0.570 | 0.946 | - | - | 43.50 | 0.58 | 0.58 | 0.08 | 4.15 | 0.584 | 0.934 | - | - | - | - | - | - | - | - | - | - | - | - |
| PLA2G10 | OID2100 | Neurolog | PLA2G10 | O15496 | 16 | 14672548 | 14694308 | 5 | 0 | 0 | 0 | 0 | 0.87 | 0.53 | 1.43 | 0.571 | 0.946 | 2.63 | 0.62 | 147.61 | 0.87 | 0.87 | 0.52 | 1.43 | 0.573 | 0.934 | 1.07 | 0.58 | 2.00 | 0.822 | 1.37 | 0.51 | 3.68 | 0.530 | -0.07 | -0.20 | 0.06 | 0.289 |
| STX4 | OID2132 | Oncology | STX4 | Q12846 | 16 | 31032889 | 31042975 | 1 | 0 | 0 | 0 | 0 | 1.77 | 0.25 | 12.52 | 0.567 | 0.946 | - | - | 40.92 | 1.77 | 1.77 | 0.23 | 13.60 | 0.583 | 0.934 | - | - | - | - | - | - | - | - | - | - | - | - |
| ACOX1 | OID2012 | Cardiom | ACOX1 | Q15067 | 17 | 75941507 | 75979177 | 1 | 0 | 0 | 0 | 0 | 0.67 | 0.19 | 2.38 | 0.532 | 0.946 | - | - | 92.25 | 0.67 | 0.67 | 0.18 | 2.43 | 0.540 | 0.932 | - | - | - | - | - | - | - | - | - | - | - | - |
| CA4 | OID2024 | Cardiom | CA4 | P22748 | 17 | 60149942 | 60170899 | 10 | 0 | 0 | 0 | 0 | 1.07 | 0.84 | 1.36 | 0.579 | 0.946 | 7.79 | 0.56 | 280.53 | 1.07 | 1.07 | 0.84 | 1.36 | 0.580 | 0.934 | 1.02 | 0.76 | 1.36 | 0.902 | 1.07 | 0.75 | 1.54 | 0.704 | 0.00 | -0.12 | 0.12 | 0.992 |
| CCL15 | OID2032 | Cardiom | CCL15 | Q16663 | 17 | 35996440 | 36001553 | 28 | 0 | 1 | 0 | 0 | 1.06 | 0.87 | 1.29 | 0.536 | 0.946 | 48.86 | 0.01 | 282.25 | 1.06 | 1.06 | 0.92 | 1.23 | 0.403 | 0.895 | 0.90 | 0.72 | 1.13 | 0.365 | 1.04 | 0.76 | 1.42 | 0.827 | 0.01 | -0.07 | 0.08 | 0.826 |
| CCL7 | OID2052 | Inflamma | CCL7 | P80098 | 17 | 34270221 | 34272242 | 3 | 0 | 0 | 0 | 0 | 0.80 | 0.37 | 1.72 | 0.563 | 0.946 | 3.79 | 0.15 | 97.48 | 0.80 | 0.80 | 0.37 | 1.72 | 0.561 | 0.934 | 0.68 | 0.29 | 1.57 | 0.364 | 0.33 | 0.01 | 9.07 | 0.510 | 0.07 | -0.17 | 0.31 | 0.569 |
| CCL8 | OID2146 | Oncology | CCL8 | P80075 | 17 | 34319435 | 34321402 | 21 | 0 | 0 | 0 | 1 | 0.94 | 0.77 | 1.15 | 0.556 | 0.946 | 32.31 | 0.04 | 329.58 | 0.94 | 0.94 | 0.81 | 1.10 | 0.453 | 0.909 | 1.01 | 0.81 | 1.26 | 0.958 | 1.30 | 0.99 | 1.73 | 0.062 | -0.11 | -0.18 | -0.03 | 0.004 |
| GP1BA | OID2038 | Cardiom | GP1BA | P07359 | 17 | 4932277 | 4935023 | 2 | 0 | 0 | 0 | 0 | 0.79 | 0.37 | 1.71 | 0.554 | 0.946 | 3.29 | 0.07 | 155.93 | 0.79 | 0.79 | 0.37 | 1.71 | 0.552 | 0.933 | - | - | - | - | - | - | - | - | - | - | - | - |
| LGALS3 | OID3076 | Inflamma | LGALS3 | Q08380 | 17 | 78971238 | 78979947 | 6 | 0 | 0 | 0 | 0 | 1.17 | 0.72 | 1.89 | 0.534 | 0.946 | 3.82 | 0.58 | 133.90 | 1.17 | 1.17 | 0.72 | 1.90 | 0.536 | 0.932 | 1.28 | 0.71 | 2.29 | 0.408 | 1.31 | 0.65 | 2.64 | 0.452 | -0.02 | -0.12 | 0.08 | 0.656 |
| SKAP1 | OID2090 | Neurolog | SKAP1 | Q86WV1 | 17 | 48133442 | 48430275 | 1 | 0 | 0 | 0 | 0 | 1.63 | 0.29 | 9.11 | 0.577 | 0.946 | - | - | 53.55 | 1.63 | 1.63 | 0.28 | 9.63 | 0.589 | 0.934 | - | - | - | - | - | - | - | - | - | - | - | - |
| DSG2 | OID2111 | Neurolog | DSG2 | Q14126 | 18 | 31498177 | 31549008 | 7 | 0 | 0 | 0 | 0 | 0.87 | 0.56 | 1.36 | 0.549 | 0.946 | 8.20 | 0.22 | 174.66 | 0.87 | 0.87 | 0.59 | 1.28 | 0.484 | 0.911 | 0.81 | 0.53 | 1.24 | 0.327 | 0.80 | 0.27 | 2.43 | 0.699 | 0.01 | -0.17 | 0.20 | 0.872 |
| NFATC1 | OID2054 | Inflamma | NFATC1 | O95644 | 18 | 79395856 | 79529325 | 5 | 0 | 0 | 0 | 0 | 1.19 | 0.68 | 2.09 | 0.550 | 0.946 | 2.80 | 0.59 | 103.21 | 1.19 | 1.19 | 0.67 | 2.10 | 0.552 | 0.933 | 1.07 | 0.56 | 2.07 | 0.829 | 0.69 | 0.18 | 2.70 | 0.593 | 0.06 | -0.08 | 0.21 | 0.392 |
| ADGRE5 | OID2026 | Cardiom | ADGRE5 | P48960 | 19 | 14380501 | 14408725 | 8 | 0 | 0 | 0 | 0 | 1.13 | 0.76 | 1.67 | 0.542 | 0.946 | 5.18 | 0.64 | 155.76 | 1.13 | 1.13 | 0.76 | 1.68 | 0.544 | 0.932 | 1.39 | 0.84 | 2.29 | 0.201 | 1.47 | 0.82 | 2.64 | 0.190 | -0.05 | -0.13 | 0.03 | 0.223 |
| CLEC4G | OID2059 | Inflamma | CLEC4G | Q6UXB4 | 19 | 7728957 | 7733906 | 6 | 0 | 0 | 0 | 0 | 1.12 | 0.76 | 1.66 | 0.566 | 0.946 | 3.40 | 0.64 | 170.26 | 1.12 | 1.12 | 0.76 | 1.66 | 0.568 | 0.934 | 1.19 | 0.78 | 1.83 | 0.422 | 1.38 | 0.78 | 2.45 | 0.264 | -0.04 | -0.12 | 0.04 | 0.320 |
| FXYD5 | OID2046 | Inflamma | FXYD5 | Q96DB9 | 19 | 35154730 | 35169881 | 3 | 0 | 0 | 0 | 0 | 1.10 | 0.81 | 1.50 | 0.538 | 0.946 | 4.73 | 0.09 | 523.45 | 1.10 | 1.10 | 0.81 | 1.50 | 0.537 | 0.932 | 1.09 | 0.79 | 1.50 | 0.596 | 1.41 | 0.21 | 9.67 | 0.724 | -0.09 | -0.72 | 0.55 | 0.787 |
| HSD17B1 | OID3109 | Neurolog | HSD17B1 | Q9BPX1 | 19 | 48813018 | 48836510 | 25 | 0 | 0 | 0 | 0 | 1.04 | 0.91 | 1.20 | 0.569 | 0.946 | 18.70 | 0.77 | 320.25 | 1.04 | 1.04 | 0.91 | 1.20 | 0.569 | 0.934 | 1.08 | 0.89 | 1.31 | 0.439 | 1.07 | 0.87 | 1.31 | 0.509 | -0.01 | -0.06 | 0.04 | 0.710 |
| IL12RB1 | OID2048 | Inflamma | IL12RB1 | P42701 | 19 | 18058995 | 18098944 | 11 | 0 | 0 | 0 | 0 | 0.93 | 0.73 | 1.18 | 0.543 | 0.946 | 6.76 | 0.75 | 244.77 | 0.93 | 0.93 | 0.73 | 1.18 | 0.544 | 0.932 | 0.97 | 0.73 | 1.30 | 0.847 | 0.78 | 0.50 | 1.22 | 0.271 | 0.03 | -0.04 | 0.11 | 0.359 |
| KIR3DL2 | OID3119 | Oncology | KIR3DL2 | P43630 | 19 | 54850443 | 54867207 | 3 | 0 | 0 | 0 | 0 | 0.82 | 0.42 | 1.63 | 0.573 | 0.946 | 0.79 | 0.67 | 116.96 | 0.82 | 0.82 | 0.41 | 1.64 | 0.577 | 0.934 | 0.71 | 0.34 | 1.47 | 0.356 | 0.55 | 0.17 | 1.83 | 0.332 | 0.07 | -0.10 | 0.24 | 0.430 |
| LAIR1 | OID2073 | Inflamma | LAIR1 | Q6GTX8 | 19 | 54351384 | 54370558 | 18 | 0 | 0 | 0 | 0 | 0.95 | 0.79 | 1.14 | 0.577 | 0.946 | 16.28 | 0.50 | 283.89 | 0.95 | 0.95 | 0.79 | 1.14 | 0.577 | 0.934 | 0.94 | 0.74 | 1.19 | 0.595 | 0.89 | 0.67 | 1.17 | 0.396 | 0.02 | -0.04 | 0.08 | 0.523 |
| PTPRH | OID3143 | Oncology | PTPRH | Q9HD43 | 19 | 55181247 | 55209506 | 13 | 0 | 0 | 0 | 0 | 0.90 | 0.63 | 1.29 | 0.564 | 0.946 | 18.97 | 0.09 | 174.46 | 0.90 | 0.90 | 0.67 | 1.20 | 0.466 | 0.909 | 0.76 | 0.52 | 1.11 | 0.150 | 0.70 | 0.43 | 1.14 | 0.153 | 0.05 | -0.02 | 0.12 | 0.158 |
| RETN | OID2032 | Cardiom | RETN | Q9HD89 | 19 | 7669049 | 7670455 | 11 | 0 | 0 | 0 | 0 | 0.93 | 0.73 | 1.20 | 0.588 | 0.946 | 9.66 | 0.47 | 239.04 | 0.93 | 0.93 | 0.73 | 1.20 | 0.589 | 0.934 | 0.99 | 0.73 | 1.35 | 0.963 | 0.81 | 0.55 | 1.19 | 0.293 | 0.04 | -0.04 | 0.11 | 0.358 |
| CHGB | OID2103 | Neurolog | CHGB | P05060 | 20 | 5911510 | 5925353 | 24 | 0 | 0 | 0 | 0 | 0.95 | 0.80 | 1.13 | 0.570 | 0.946 | 18.34 | 0.74 | 229.57 | 0.95 | 0.95 | 0.80 | 1.13 | 0.571 | 0.934 | 0.90 | 0.72 | 1.12 | 0.344 | 0.99 | 0.74 | 1.33 | 0.944 | -0.01 | -0.07 | 0.05 | 0.751 |
| ENTPD6 | OID2010 | Cardiom | ENTPD6 | O75354 | 20 | 25195693 | 25228075 | 24 | 0 | 0 | 0 | 0 | 0.96 | 0.86 | 1.09 | 0.563 | 0.946 | 25.80 | 0.31 | 493.78 | 0.96 | 0.96 | 0.86 | 1.08 | 0.540 | 0.932 | 0.99 | 0.85 | 1.16 | 0.945 | 0.99 | 0.84 | 1.17 | 0.911 | -0.01 | -0.06 | 0.04 | 0.636 |
| RALY | OID3138 | Oncology | RALY | Q9UKM9 | 20 | 33993646 | 34108308 | 2 | 0 | 0 | 0 | 0 | 1.40 | 0.46 | 4.27 | 0.551 | 0.946 | 0.00 | 0.95 | 62.11 | 1.40 | 1.40 | 0.45 | 4.40 | 0.561 | 0.934 | - | - | - | - | - | - | - | - | - | - | - | - |
| CRYZL1 | OID3027 | Cardiom | CRYZL1 | O95825 | 21 | 33589341 | 33643926 | 1 | 0 | 0 | 0 | 0 | 0.81 | 0.41 | 1.61 | 0.548 | 0.946 | - | - | 296.85 | 0.81 | 0.81 | 0.41 | 1.61 | 0.551 | 0.933 | - | - | - | - | - | - | - | - | - | - | - | - |
| JAM2 | OID2106 | Neurolog | JAM2 | P57087 | 21 | 25639258 | 25717562 | 3 | 0 | 0 | 1 | 0 | 1.34 | 0.51 | 3.53 | 0.549 | 0.946 | 0.71 | 0.70 | 61.64 | 1.34 | 1.34 | 0.50 | 3.61 | 0.557 | 0.934 | 1.12 | 0.38 | 3.33 | 0.843 | 1.18 | 0.08 | 16.75 | 0.900 | 0.01 | -0.26 | 0.28 | 0.920 |
| TFF1 | OID2115 | Neurolog | TFF1 | P04155 | 21 | 42362282 | 42366535 | 4 | 0 | 0 | 0 | 0 | 1.11 | 0.79 | 1.58 | 0.542 | 0.946 | 0.77 | 0.86 | 354.14 | 1.11 | 1.11 | 0.78 | 1.58 | 0.544 | 0.932 | 1.16 | 0.81 | 1.66 | 0.430 | 1.44 | 0.69 | 3.00 | 0.336 | -0.07 | -0.23 | 0.10 | 0.445 |
| BCR | OID2055 | Inflamma | BCR | P11274 | 22 | 23179704 | 23318037 | 1 | 0 | 0 | 0 | 0 | 0.62 | 0.14 | 2.86 | 0.542 | 0.946 | - | - | 58.87 | 0.62 | 0.62 | 0.13 | 2.99 | 0.554 | 0.933 | - | - | - | - | - | - | - | - | - | - | - | - |
| COMT | OID2008 | Cardiom | COMT | P21964 | 22 | 19941371 | 19969975 | 18 | 0 | 0 | 0 | 0 | 0.93 | 0.74 | 1.18 | 0.557 | 0.946 | 13.66 | 0.69 | 157.36 | 0.93 | 0.93 | 0.74 | 1.18 | 0.557 | 0.934 | 0.86 | 0.65 | 1.14 | 0.297 | 0.76 | 0.48 | 1.21 | 0.244 | 0.05 | -0.05 | 0.14 | 0.315 |
| HMOX1 | OID2021 | Cardiom | HMOX1 | P09601 | 22 | 35380361 | 35394214 | 2 | 0 | 0 | 0 | 0 | 0.76 | 0.30 | 1.94 | 0.567 | 0.946 | 4.35 | 0.04 | 90.93 | 0.76 | 0.76 | 0.30 | 1.93 | 0.560 | 0.934 | - | - | - | - | - | - | - | - | - | - | - | - |
| CKMT1A | OID2072 | Inflamma | CKMT1B | P12532;P | 15;15 | 43593054;4 | 43604901;4 | 7 | 0 | 0 | 0 | 0 | 1.13 | 0.75 | 1.70 | 0.569 | 0.946 | 4.52 | 0.61 | 130.31 | 1.13 | 1.13 | 0.74 | 1.71 | 0.570 | 0.934 | 1.14 | 0.68 | 1.91 | 0.616 | 1.62 | 0.53 | 5.00 | 0.401 | -0.08 | -0.31 | 0.15 | 0.497 |
| CGB3_C | OID3067 | Inflamma | CGB5;C | P0DN86; | 19;19;19 | 49043848;4 | 49045311;4 | 1 | 0 | 0 | 0 | 0 | 1.57 | 0.33 | 7.41 | 0.572 | 0.946 | - | - | 75.67 | 1.57 | 1.57 | 0.32 | 7.68 | 0.580 | 0.934 | - | - | - | - | - | - | - | - | - | - | - | - |
| CETN3 | OID3152 | Oncology | CETN3 | O15182 | 5 | 90392257 | 90409766 | 2 | 0 | 0 | 0 | 0 | 1.17 | 0.66 | 2.09 | 0.593 | 0.947 | 0.08 | 0.78 | 233.73 | 1.17 | 1.17 | 0.65 | 2.10 | 0.596 | 0.934 | - | - | - | - | - | - | - | - | - | - | - | - |
| RARRES | OID2037 | Cardiom | RARRES | Q99969 | 7 | 150338317 | 150341662 | 2 | 0 | 0 | 0 | 0 | 0.88 | 0.54 | 1.43 | 0.594 | 0.947 | 0.04 | 0.84 | 346.25 | 0.88 | 0.88 | 0.54 | 1.43 | 0.596 | 0.934 | - | - | - | - | - | - | - | - | - | - | - | - |
| LIPF | OID3026 | Cardiom | LIPF | P07098 | 10 | 88664441 | 88678814 | 6 | 0 | 0 | 0 | 0 | 0.91 | 0.64 | 1.29 | 0.595 | 0.947 | 6.93 | 0.23 | 292.65 | 0.91 | 0.91 | 0.67 | 1.23 | 0.531 | 0.932 | 0.91 | 0.66 | 1.27 | 0.589 | 1.04 | 0.58 | 1.87 | 0.887 | -0.03 | -0.11 | 0.06 | 0.539 |
| PRTFDC | OID2086 | Neurolog | PRTFDC | Q9NRG1 | 10 | 24848614 | 24952606 | 3 | 0 | 0 | 0 | 0 | 1.11 | 0.75 | 1.66 | 0.594 | 0.947 | 1.00 | 0.61 | 328.87 | 1.11 | 1.11 | 0.75 | 1.66 | 0.595 | 0.934 | 1.06 | 0.69 | 1.61 | 0.804 | 0.97 | 0.59 | 1.59 | 0.892 | 0.05 | -0.06 | 0.17 | 0.346 |
| ADM | OID2146 | Oncology | ADM | P35318 | 11 | 10305073 | 10307397 | 4 | 0 | 0 | 0 | 0 | 1.13 | 0.72 | 1.76 | 0.595 | 0.947 | 0.65 | 0.88 | 223.97 | 1.13 | 1.13 | 0.72 | 1.77 | 0.597 | 0.934 | 1.16 | 0.71 | 1.90 | 0.563 | 0.54 | 0.06 | 4.44 | 0.563 | 0.10 | -0.17 | 0.37 | 0.480 |
| THY1 | OID2105 | Neurolog | THY1 | P04216 | 11 | 119415476 | 119424985 | 14 | 0 | 0 | 0 | 0 | 0.95 | 0.77 | 1.16 | 0.593 | 0.947 | 13.00 | 0.45 | 252.06 | 0.95 | 0.95 | 0.77 | 1.16 | 0.593 | 0.934 | 0.92 | 0.72 | 1.18 | 0.501 | 1.00 | 0.76 | 1.31 | 0.986 | -0.02 | -0.08 | 0.04 | 0.545 |
| TRIM21 | OID2053 | Inflamma | TRIM21 | P19474 | 11 | 4384897 | 4393702 | 2 | 0 | 0 | 0 | 0 | 0.67 | 0.15 | 2.91 | 0.594 | 0.947 | 2.02 | 0.15 | 40.22 | 0.66 | 0.66 | 0.15 | 3.00 | 0.595 | 0.934 | - | - | - | - | - | - | - | - | - | - | - | - |
| C1R | OID3075 | Inflamma | C1R | P00736 | 12 | 7080214 | 7092540 | 7 | 0 | 0 | 0 | 0 | 0.93 | 0.70 | 1.23 | 0.595 | 0.947 | 4.40 | 0.62 | 275.33 | 0.93 | 0.93 | 0.70 | 1.23 | 0.596 | 0.934 | 0.93 | 0.68 | 1.25 | 0.617 | 0.93 | 0.59 | 1.47 | 0.755 | 0.00 | -0.10 | 0.10 | 0.983 |
| DHRS4L | OID3107 | Neurolog | DHRS4L | Q6PKH6 | 14 | 23969874 | 24006408 | 9 | 0 | 0 | 0 | 0 | 1.10 | 0.77 | 1.59 | 0.595 | 0.947 | 12.12 | 0.15 | 221.41 | 1.10 | 1.10 | 0.82 | 1.48 | 0.512 | 0.930 | 1.23 | 0.88 | 1.72 | 0.217 | 1.37 | 0.84 | 2.22 | 0.207 | -0.07 | -0.17 | 0.04 | 0.211 |
| TPSAB1 | OID2064 | Inflamma | TPSAB1 | Q15661 | 16 | 1240379 | 1242554 | 39 | 0 | 0 | 0 | 0 | 1.03 | 0.93 | 1.13 | 0.591 | 0.947 | 40.58 | 0.36 | 413.26 | 1.03 | 1.03 | 0.93 | 1.13 | 0.579 | 0.934 | 1.02 | 0.88 | 1.17 | 0.811 | 1.03 | 0.89 | 1.20 | 0.665 | 0.00 | -0.05 | 0.05 | 0.923 |
| KLRB1 | OID2062 | Inflamma | KLRB1 | Q12918 | 12 | 9594551 | 9607916 | 9 | 0 | 0 | 0 | 0 | 0.89 | 0.58 | 1.37 | 0.597 | 0.949 | 7.35 | 0.50 | 97.67 | 0.89 | 0.89 | 0.58 | 1.37 | 0.598 | 0.935 | 0.86 | 0.49 | 1.49 | 0.588 | 1.29 | 0.34 | 4.95 | 0.707 | -0.06 | -0.26 | 0.14 | 0.566 |
| RLN2 | OID3060 | Inflamma | RLN2 | P04090 | 9 | 5299864 | 5304716 | 6 | 0 | 0 | 0 | 0 | 0.81 | 0.36 | 1.79 | 0.598 | 0.950 | 12.32 | 0.03 | 100.16 | 0.80 | 0.80 | 0.48 | 1.34 | 0.399 | 0.895 | 1.21 | 0.65 | 2.26 | 0.554 | 1.48 | 0.28 | 7.70 | 0.641 | -0.09 | -0.29 | 0.12 | 0.405 |
| MAX | OID2087 | Neurolog | MAX | P61244 | 14 | 65006174 | 65102695 | 1 | 0 | 0 | 0 | 0 | 1.39 | 0.41 | 4.70 | 0.599 | 0.950 | - | - | 102.12 | 1.39 | 1.39 | 0.40 | 4.80 | 0.605 | 0.939 | - | - | - | - | - | - | - | - | - | - | - | - |
| INSR | OID3056 | Inflamma | INSR | P06213 | 19 | 7112255 | 7294414 | 1 | 0 | 0 | 0 | 0 | 0.62 | 0.10 | 3.72 | 0.598 | 0.950 | - | - | 43.39 | 0.62 | 0.62 | 0.10 | 3.99 | 0.612 | 0.939 | - | - | - | - | - | - | - | - | - | - | - | - |
| CD164L2 | OID3092 | Neurolog | CD164L2 | Q6UWJ8 | 1 | 27379176 | 27383333 | 12 | 0 | 0 | 0 | 0 | 1.09 | 0.78 | 1.52 | 0.606 | 0.952 | 11.92 | 0.37 | 131.22 | 1.09 | 1.09 | 0.79 | 1.50 | 0.591 | 0.934 | 0.94 | 0.62 | 1.42 | 0.771 | 0.81 | 0.49 | 1.32 | 0.397 | 0.07 | -0.02 | 0.17 | 0.121 |
| CD55 | OID2037 | Cardiom | CD55 | P08174 | 1 | 207321532 | 207386804 | 15 | 0 | 0 | 0 | 0 | 0.95 | 0.77 | 1.18 | 0.664 | 0.952 | 8.86 | 0.84 | 226.66 | 0.95 | 0.95 | 0.77 | 1.18 | 0.665 | 0.947 | 0.90 | 0.69 | 1.17 | 0.428 | 0.82 | 0.56 | 1.19 | 0.294 | 0.04 | -0.04 | 0.13 | 0.330 |
| CRNN | OID2142 | Oncology | CRNN | Q9UBG3 | 1 | 152409243 | 152414263 | 24 | 0 | 0 | 0 | 0 | 0.96 | 0.80 | 1.15 | 0.646 | 0.952 | 16.47 | 0.83 | 201.28 | 0.96 | 0.96 | 0.79 | 1.15 | 0.647 | 0.945 | 0.96 | 0.74 | 1.25 | 0.754 | 0.84 | 0.63 | 1.13 | 0.253 | 0.04 | -0.03 | 0.10 | 0.270 |
| EFNA4 | OID2093 | Neurolog | EFNA4 | P52798 | 1 | 155063737 | 155069553 | 1 | 0 | 0 | 0 | 0 | 0.60 | 0.08 | 4.55 | 0.621 | 0.952 | - | - | 46.75 | 0.60 | 0.60 | 0.07 | 4.90 | 0.633 | 0.945 | - | - | - | - | - | - | - | - | - | - | - | - |
| FOXJ3 | OID3044 | Inflamma | FOXJ3 | Q9UPW6 | 1 | 42176539 | 42335877 | 1 | 0 | 0 | 0 | 0 | 1.39 | 0.36 | 5.30 | 0.633 | 0.952 | - | - | 86.53 | 1.39 | 1.39 | 0.35 | 5.44 | 0.640 | 0.945 | - | - | - | - | - | - | - | - | - | - | - | - |
| GALNT2 | OID2142 | Oncology | GALNT2 | Q10471 | 1 | 230057990 | 230282122 | 10 | 0 | 0 | 0 | 0 | 0.94 | 0.73 | 1.22 | 0.664 | 0.952 | 3.27 | 0.95 | 253.58 | 0.94 | 0.94 | 0.73 | 1.22 | 0.665 | 0.947 | 0.89 | 0.66 | 1.21 | 0.462 | 0.65 | 0.37 | 1.16 | 0.144 | 0.08 | -0.03 | 0.18 | 0.156 |
| HSPG2 | OID2038 | Cardiom | HSPG2 | P98160 | 1 | 21822244 | 21937310 | 7 | 0 | 0 | 0 | 0 | 1.17 | 0.61 | 2.24 | 0.645 | 0.952 | 13.18 | 0.04 | 121.41 | 1.17 | 1.17 | 0.75 | 1.82 | 0.489 | 0.912 | 1.17 | 0.69 | 1.97 | 0.558 | 0.42 | 0.13 | 1.37 | 0.149 | 0.16 | 0.00 | 0.32 | 0.056 |
| IL10 | OID2043 | Inflamma | IL10 | P22301 | 1 | 206767602 | 206774541 | 1 | 0 | 0 | 0 | 0 | 1.23 | 0.48 | 3.16 | 0.663 | 0.952 | - | - | 175.99 | 1.23 | 1.23 | 0.48 | 3.19 | 0.666 | 0.947 | - | - | - | - | - | - | - | - | - | - | - | - |
| LRRC38 | OID3092 | Neurolog | LRRC38 | Q5VT99 | 1 | 13474973 | 13514003 | 6 | 0 | 0 | 0 | 0 | 0.94 | 0.72 | 1.23 | 0.649 | 0.952 | 2.74 | 0.74 | 358.81 | 0.94 | 0.94 | 0.72 | 1.23 | 0.650 | 0.945 | 0.95 | 0.71 | 1.27 | 0.745 | 1.13 | 0.60 | 2.14 | 0.700 | -0.05 | -0.20 | 0.10 | 0.522 |
| NID1 | OID2036 | Cardiom | NID1 | P14543 | 1 | 235975830 | 236065109 | 3 | 0 | 0 | 0 | 0 | 1.13 | 0.69 | 1.85 | 0.636 | 0.952 | 0.44 | 0.80 | 221.74 | 1.13 | 1.13 | 0.68 | 1.86 | 0.638 | 0.945 | 1.12 | 0.67 | 1.88 | 0.668 | 0.99 | 0.46 | 2.14 | 0.976 | 0.03 | -0.11 | 0.18 | 0.663 |
| NOTCH2 | OID3040 | Cardiom | NOTCH2 | Q04721 | 1 | 119911553 | 120100779 | 4 | 0 | 0 | 0 | 0 | 0.85 | 0.44 | 1.62 | 0.615 | 0.952 | 3.00 | 0.39 | 95.53 | 0.85 | 0.85 | 0.44 | 1.63 | 0.617 | 0.942 | 0.92 | 0.43 | 1.97 | 0.836 | 3.32 | 0.31 | 34.99 | 0.319 | -0.19 | -0.50 | 0.12 | 0.238 |
| PRELP | OID2076 | Inflamma | PRELP | P51888 | 1 | 203475806 | 203491352 | 9 | 0 | 0 | 0 | 0 | 1.08 | 0.80 | 1.46 | 0.609 | 0.952 | 8.25 | 0.41 | 277.02 | 1.08 | 1.08 | 0.80 | 1.46 | 0.604 | 0.939 | 1.35 | 0.95 | 1.92 | 0.094 | 1.32 | 0.91 | 1.93 | 0.146 | -0.05 | -0.10 | 0.01 | 0.091 |
| PSRC1 | OID2116 | Oncology | PSRC1 | Q6PGN9 | 1 | 109279556 | 109283186 | 3 | 0 | 0 | 0 | 0 | 1.12 | 0.72 | 1.73 | 0.610 | 0.952 | 2.65 | 0.27 | 240.71 | 1.12 | 1.12 | 0.72 | 1.74 | 0.610 | 0.939 | 1.08 | 0.68 | 1.71 | 0.740 | 1.39 | 0.52 | 3.74 | 0.513 | -0.06 | -0.30 | 0.17 | 0.581 |
| S100A12 | OID2137 | Oncology | S100A12 | P80511 | 1 | 153373711 | 153375621 | 2 | 0 | 0 | 0 | 0 | 0.92 | 0.63 | 1.35 | 0.664 | 0.952 | 6.45 | 0.01 | 525.22 | 0.92 | 0.92 | 0.63 | 1.35 | 0.662 | 0.947 | - | - | - | - | - | - | - | - | - | - | - | - |
| TDRKH | OID2087 | Neurolog | TDRKH | Q9Y2W6 | 1 | 151770107 | 151791416 | 9 | 0 | 0 | 0 | 0 | 0.94 | 0.71 | 1.23 | 0.638 | 0.952 | 1.94 | 0.98 | 225.42 | 0.94 | 0.94 | 0.71 | 1.24 | 0.640 | 0.945 | 0.89 | 0.67 | 1.20 | 0.457 | 0.82 | 0.53 | 1.28 | 0.385 | 0.03 | -0.05 | 0.10 | 0.462 |
| CASP8 | OID2141 | Oncology | CASP8 | Q14790 | 2 | 201233443 | 201287711 | 8 | 0 | 0 | 0 | 0 | 1.10 | 0.72 | 1.68 | 0.644 | 0.952 | 8.60 | 0.28 | 143.75 | 1.11 | 1.11 | 0.75 | 1.62 | 0.608 | 0.939 | 0.92 | 0.60 | 1.42 | 0.701 | 0.58 | 0.28 | 1.19 | 0.138 | 0.14 | 0.01 | 0.28 | 0.040 |
| ITGB6 | OID2049 | Inflamma | ITGB6 | P18564 | 2 | 160099667 | 160200313 | 10 | 0 | 0 | 0 | 0 | 0.94 | 0.72 | 1.23 | 0.660 | 0.952 | 9.51 | 0.39 | 267.62 | 0.94 | 0.94 | 0.73 | 1.22 | 0.651 | 0.945 | 0.92 | 0.67 | 1.25 | 0.593 | 0.99 | 0.67 | 1.47 | 0.968 | -0.01 | -0.08 | 0.06 | 0.718 |
| LY75 | OID2058 | Inflamma | LY75 | O60449 | 2 | 159803355 | 159904756 | 54 | 0 | 0 | 0 | 0 | 0.98 | 0.90 | 1.07 | 0.615 | 0.952 | 39.83 | 0.91 | 372.07 | 0.98 | 0.98 | 0.90 | 1.07 | 0.615 | 0.941 | 0.97 | 0.85 | 1.11 | 0.648 | 0.95 | 0.79 | 1.15 | 0.612 | 0.01 | -0.06 | 0.08 | 0.759 |
| MERTK | OID2065 | Inflamma | MERTK | Q12866 | 2 | 111898607 | 112029561 | 10 | 0 | 0 | 0 | 0 | 0.93 | 0.68 | 1.27 | 0.631 | 0.952 | 2.49 | 0.98 | 176.54 | 0.93 | 0.93 | 0.68 | 1.27 | 0.633 | 0.945 | 0.95 | 0.67 | 1.35 | 0.778 | 0.83 | 0.48 | 1.42 | 0.485 | 0.02 | -0.07 | 0.11 | 0.607 |
| MTHFD2 | OID3107 | Neurolog | MTHFD2 | P13995 | 2 | 74186172 | 74217565 | 3 | 0 | 0 | 1 | 0 | 1.23 | 0.52 | 2.89 | 0.632 | 0.952 | 0.34 | 0.84 | 62.12 | 1.23 | 1.23 | 0.51 | 2.95 | 0.639 | 0.945 | 1.34 | 0.53 | 3.38 | 0.531 | 2.12 | 0.22 | 19.97 | 0.512 | -0.11 | -0.54 | 0.32 | 0.609 |
| RNF149 | OID3023 | Cardiom | RNF149 | Q8NC42 | 2 | 101271219 | 101308701 | 5 | 0 | 0 | 0 | 0 | 1.08 | 0.78 | 1.50 | 0.641 | 0.952 | 2.89 | 0.58 | 299.29 | 1.08 | 1.08 | 0.78 | 1.50 | 0.642 | 0.945 | 1.11 | 0.79 | 1.55 | 0.549 | 1.48 | 0.86 | 2.54 | 0.158 | -0.07 | -0.17 | 0.03 | 0.156 |
| SERPINE | OID3035 | Cardiom | SERPINE | P07093 | 2 | 223975045 | 224039318 | 21 | 0 | 0 | 0 | 0 | 0.93 | 0.72 | 1.22 | 0.614 | 0.952 | 30.59 | 0.06 | 164.32 | 0.93 | 0.93 | 0.75 | 1.16 | 0.530 | 0.932 | 1.01 | 0.76 | 1.33 | 0.965 | 1.13 | 0.68 | 1.88 | 0.624 | -0.04 | -0.14 | 0.05 | 0.377 |
| TGOLN2 | OID3069 | Inflamma | TGOLN2 | O43493 | 2 | 85318027 | 85328251 | 34 | 0 | 0 | 0 | 0 | 1.03 | 0.90 | 1.18 | 0.665 | 0.952 | 25.35 | 0.83 | 248.63 | 1.03 | 1.03 | 0.90 | 1.18 | 0.665 | 0.947 | 0.94 | 0.78 | 1.15 | 0.555 | 1.01 | 0.78 | 1.31 | 0.950 | 0.01 | -0.06 | 0.08 | 0.848 |
| BOC | OID2011 | Cardiom | BOC | Q9BWV1 | 3 | 113211003 | 113287459 | 7 | 0 | 0 | 0 | 0 | 0.91 | 0.61 | 1.35 | 0.639 | 0.952 | 4.22 | 0.65 | 150.03 | 0.91 | 0.91 | 0.61 | 1.35 | 0.641 | 0.945 | 0.95 | 0.58 | 1.55 | 0.837 | 1.51 | 0.34 | 6.76 | 0.593 | -0.09 | -0.37 | 0.18 | 0.495 |
| CD86 | OID3150 | Oncology | CD86 | P42081 | 3 | 122055362 | 122121139 | 10 | 0 | 0 | 0 | 0 | 0.90 | 0.58 | 1.39 | 0.640 | 0.952 | 8.06 | 0.53 | 90.52 | 0.90 | 0.90 | 0.58 | 1.40 | 0.642 | 0.945 | 0.87 | 0.49 | 1.55 | 0.637 | 1.75 | 0.35 | 8.68 | 0.491 | -0.10 | -0.35 | 0.14 | 0.397 |
| OSTN | OID3090 | Neurolog | OSTN | P61366 | 3 | 191199241 | 191265615 | 2 | 0 | 0 | 0 | 0 | 0.80 | 0.32 | 2.02 | 0.641 | 0.952 | 0.28 | 0.60 | 88.73 | 0.80 | 0.80 | 0.31 | 2.06 | 0.646 | 0.945 | - | - | - | - | - | - | - | - | - | - | - | - |
| RPL14 | OID3028 | Cardiom | RPL14 | P50914 | 3 | 40457292 | 40468587 | 1 | 0 | 0 | 0 | 0 | 0.67 | 0.12 | 3.84 | 0.653 | 0.952 | - | - | 46.68 | 0.67 | 0.67 | 0.11 | 4.08 | 0.664 | 0.947 | - | - | - | - | - | - | - | - | - | - | - | - |
| TDGF1 | OID2081 | Neurolog | TDGF1 | P13385 | 3 | 46574534 | 46582457 | 52 | 0 | 0 | 0 | 0 | 1.02 | 0.94 | 1.12 | 0.611 | 0.952 | 45.41 | 0.69 | 372.43 | 1.02 | 1.02 | 0.94 | 1.12 | 0.611 | 0.939 | 1.04 | 0.91 | 1.18 | 0.569 | 1.03 | 0.86 | 1.23 | 0.749 | 0.00 | -0.07 | 0.06 | 0.935 |
| FGFBP1 | OID2150 | Oncology | FGFBP1 | Q14512 | 4 | 15935577 | 15938740 | 6 | 0 | 0 | 1 | 0 | 0.87 | 0.50 | 1.52 | 0.629 | 0.952 | 1.23 | 0.94 | 94.13 | 0.87 | 0.87 | 0.49 | 1.53 | 0.633 | 0.945 | 0.86 | 0.44 | 1.67 | 0.656 | 0.59 | 0.09 | 3.85 | 0.585 | 0.05 | -0.20 | 0.30 | 0.673 |
| HGFAC | OID3071 | Inflamma | HGFAC | Q04756 | 4 | 3441968 | 3449486 | 24 | 0 | 0 | 0 | 0 | 1.04 | 0.88 | 1.23 | 0.651 | 0.952 | 14.72 | 0.90 | 234.49 | 1.04 | 1.04 | 0.88 | 1.23 | 0.652 | 0.945 | 0.99 | 0.80 | 1.24 | 0.956 | 0.94 | 0.70 | 1.25 | 0.654 | 0.03 | -0.04 | 0.11 | 0.377 |
| LAP3 | OID2043 | Inflamma | LAP3 | P28838 | 4 | 17577198 | 17607972 | 2 | 0 | 0 | 0 | 0 | 1.48 | 0.28 | 7.67 | 0.642 | 0.952 | 1.46 | 0.23 | 29.80 | 1.49 | 1.49 | 0.27 | 8.29 | 0.647 | 0.945 | - | - | - | - | - | - | - | - | - | - | - | - |
| PKD2 | OID3018 | Cardiom | PKD2 | Q13563 | 4 | 88007635 | 88077777 | 2 | 0 | 0 | 0 | 0 | 0.77 | 0.29 | 2.06 | 0.609 | 0.952 | 0.83 | 0.36 | 79.64 | 0.77 | 0.77 | 0.28 | 2.10 | 0.613 | 0.940 | - | - | - | - | - | - | - | - | - | - | - | - |
| C7 | OID3076 | Inflamma | C7 | P10643 | 5 | 40909492 | 40984643 | 25 | 0 | 0 | 0 | 0 | 0.96 | 0.82 | 1.12 | 0.603 | 0.952 | 25.45 | 0.38 | 272.22 | 0.96 | 0.96 | 0.82 | 1.12 | 0.592 | 0.934 | 1.26 | 0.97 | 1.62 | 0.079 | 1.09 | 0.75 | 1.59 | 0.651 | -0.03 | -0.12 | 0.05 | 0.458 |
| GALNT10 | OID2137 | Oncology | GALNT10 | Q86SR1 | 5 | 154190730 | 154420984 | 20 | 0 | 0 | 0 | 0 | 1.05 | 0.85 | 1.31 | 0.643 | 0.952 | 20.65 | 0.36 | 189.86 | 1.05 | 1.05 | 0.85 | 1.30 | 0.629 | 0.945 | 1.01 | 0.76 | 1.34 | 0.970 | 0.79 | 0.52 | 1.19 | 0.250 | 0.07 | -0.02 | 0.16 | 0.107 |
| HBEGF | OID2146 | Oncology | HBEGF | Q99075 | 5 | 140332843 | 140346603 | 2 | 0 | 1 | 0 | 0 | 0.72 | 0.17 | 3.05 | 0.658 | 0.952 | 8.42 | 0.00 | 38.28 | 0.69 | 0.69 | 0.17 | 2.88 | 0.614 | 0.940 | - | - | - | - | - | - | - | - | - | - | - | - |
| IL31RA | OID3047 | Inflamma | IL31RA | Q8NI17 | 5 | 55851357 | 55922854 | 33 | 0 | 0 | 0 | 0 | 0.97 | 0.85 | 1.10 | 0.615 | 0.952 | 32.77 | 0.43 | 284.24 | 0.97 | 0.97 | 0.85 | 1.10 | 0.610 | 0.939 | 0.94 | 0.78 | 1.12 | 0.480 | 1.00 | 0.82 | 1.24 | 0.964 | -0.01 | -0.07 | 0.04 | 0.645 |
| OSMR | OID2035 | Cardiom | OSMR | Q99650 | 5 | 38845858 | 38945596 | 22 | 0 | 0 | 0 | 0 | 1.04 | 0.87 | 1.25 | 0.663 | 0.952 | 19.45 | 0.56 | 244.42 | 1.04 | 1.04 | 0.87 | 1.25 | 0.663 | 0.947 | 1.04 | 0.80 | 1.35 | 0.755 | 1.05 | 0.77 | 1.43 | 0.750 | 0.00 | -0.07 | 0.06 | 0.940 |
| BTN2A1 | OID2071 | Inflamma | BTN2A1 | Q7KYR7 | 6 | 26457904 | 26476621 | 23 | 0 | 0 | 0 | 0 | 1.04 | 0.89 | 1.22 | 0.607 | 0.952 | 19.78 | 0.60 | 303.14 | 1.04 | 1.04 | 0.89 | 1.22 | 0.607 | 0.939 | 1.06 | 0.87 | 1.29 | 0.549 | 1.13 | 0.85 | 1.50 | 0.408 | -0.02 | -0.10 | 0.05 | 0.514 |
| GMPR | OID2064 | Inflamma | GMPR | P36959 | 6 | 16238587 | 16295549 | 5 | 0 | 0 | 0 | 0 | 0.92 | 0.64 | 1.32 | 0.639 | 0.952 | 0.14 | 1.00 | 252.66 | 0.92 | 0.92 | 0.64 | 1.32 | 0.641 | 0.945 | 0.90 | 0.62 | 1.31 | 0.581 | 0.85 | 0.44 | 1.61 | 0.610 | 0.02 | -0.09 | 0.12 | 0.766 |
| MLN | OID2054 | Inflamma | MLN | P12872 | 6 | 33794673 | 33804003 | 21 | 0 | 0 | 0 | 0 | 1.05 | 0.85 | 1.30 | 0.656 | 0.952 | 15.80 | 0.73 | 192.37 | 1.05 | 1.05 | 0.85 | 1.30 | 0.657 | 0.947 | 1.02 | 0.76 | 1.37 | 0.873 | 1.14 | 0.80 | 1.64 | 0.463 | -0.02 | -0.07 | 0.04 | 0.559 |
| SMPDL3 | OID2067 | Inflamma | SMPDL3 | Q92484 | 6 | 122789049 | 122809720 | 38 | 0 | 0 | 0 | 0 | 1.04 | 0.89 | 1.22 | 0.637 | 0.952 | 16.16 | 1.00 | 170.77 | 1.04 | 1.04 | 0.88 | 1.22 | 0.639 | 0.945 | 0.95 | 0.76 | 1.19 | 0.646 | 1.04 | 0.72 | 1.49 | 0.845 | 0.00 | -0.07 | 0.07 | 0.989 |
| WASF1 | OID2009 | Cardiom | WASF1 | Q92558 | 6 | 110099819 | 110180004 | 4 | 0 | 0 | 0 | 0 | 1.11 | 0.71 | 1.73 | 0.662 | 0.952 | 6.12 | 0.11 | 400.77 | 1.11 | 1.11 | 0.81 | 1.51 | 0.532 | 0.932 | 1.10 | 0.79 | 1.52 | 0.578 | 1.11 | 0.42 | 2.91 | 0.831 | 0.00 | -0.18 | 0.18 | 0.991 |
| PON2 | OID2008 | Cardiom | PON2 | Q15165 | 7 | 95404862 | 95435329 | 51 | 0 | 0 | 0 | 0 | 1.03 | 0.92 | 1.14 | 0.630 | 0.952 | 47.54 | 0.57 | 261.64 | 1.03 | 1.03 | 0.92 | 1.14 | 0.631 | 0.945 | 1.00 | 0.85 | 1.16 | 0.954 | 1.01 | 0.83 | 1.24 | 0.888 | 0.00 | -0.06 | 0.06 | 0.891 |
| SFRP4 | OID3064 | Inflamma | SFRP4 | Q6FHJ7 | 7 | 37905932 | 38025695 | 7 | 0 | 0 | 0 | 0 | 1.11 | 0.76 | 1.62 | 0.606 | 0.952 | 4.27 | 0.64 | 151.74 | 1.11 | 1.11 | 0.75 | 1.62 | 0.608 | 0.939 | 1.13 | 0.73 | 1.75 | 0.579 | 0.89 | 0.47 | 1.71 | 0.731 | 0.04 | -0.06 | 0.14 | 0.426 |
| SUMF2 | OID2103 | Neurolog | SUMF2 | Q8NBJ7 | 7 | 56064002 | 56080670 | 13 | 0 | 0 | 0 | 0 | 1.06 | 0.82 | 1.38 | 0.647 | 0.952 | 13.15 | 0.36 | 246.95 | 1.06 | 1.06 | 0.83 | 1.36 | 0.632 | 0.945 | 1.28 | 0.95 | 1.72 | 0.111 | 1.23 | 0.90 | 1.68 | 0.200 | -0.05 | -0.11 | 0.02 | 0.143 |
| VWC2 | OID2093 | Neurolog | VWC2 | Q2TAL6 | 7 | 49773638 | 49921950 | 23 | 0 | 0 | 0 | 0 | 1.05 | 0.87 | 1.27 | 0.623 | 0.952 | 24.82 | 0.31 | 233.85 | 1.05 | 1.05 | 0.88 | 1.26 | 0.601 | 0.937 | 1.20 | 0.93 | 1.55 | 0.167 | 1.09 | 0.78 | 1.52 | 0.634 | -0.01 | -0.08 | 0.06 | 0.809 |
| IMPA1 | OID2090 | Neurolog | IMPA1 | P29218 | 8 | 81656914 | 81686331 | 3 | 0 | 0 | 0 | 0 | 0.89 | 0.56 | 1.41 | 0.621 | 0.952 | 1.30 | 0.52 | 240.12 | 0.89 | 0.89 | 0.56 | 1.42 | 0.622 | 0.945 | 0.88 | 0.54 | 1.42 | 0.590 | 0.64 | 0.27 | 1.54 | 0.318 | 0.06 | -0.07 | 0.19 | 0.385 |
| MATN2 | OID2076 | Inflamma | MATN2 | O00339 | 8 | 97868840 | 98036724 | 8 | 0 | 0 | 0 | 0 | 0.93 | 0.68 | 1.27 | 0.634 | 0.952 | 9.67 | 0.21 | 301.83 | 0.93 | 0.93 | 0.71 | 1.21 | 0.576 | 0.934 | 1.06 | 0.78 | 1.44 | 0.712 | 1.07 | 0.57 | 1.99 | 0.833 | -0.03 | -0.14 | 0.08 | 0.596 |
| PENK | OID3066 | Inflamma | PENK | P01210 | 8 | 56436674 | 56446671 | 13 | 0 | 0 | 0 | 0 | 0.94 | 0.74 | 1.20 | 0.611 | 0.952 | 8.03 | 0.78 | 204.47 | 0.94 | 0.94 | 0.73 | 1.20 | 0.612 | 0.939 | 0.93 | 0.69 | 1.26 | 0.652 | 0.90 | 0.51 | 1.58 | 0.718 | 0.01 | -0.09 | 0.11 | 0.875 |
| PXDNL | OID3140 | Oncology | PXDNL | A1KZ92 | 8 | 51319577 | 51809445 | 8 | 0 | 0 | 0 | 0 | 0.88 | 0.54 | 1.44 | 0.607 | 0.952 | 14.31 | 0.05 | 161.05 | 0.88 | 0.88 | 0.62 | 1.24 | 0.459 | 0.909 | 1.07 | 0.72 | 1.58 | 0.736 | 1.28 | 0.55 | 2.96 | 0.568 | -0.08 | -0.23 | 0.07 | 0.283 |
| RRM2B | OID2117 | Oncology | RRM2B | Q7LG56 | 8 | 102204502 | 102238961 | 2 | 0 | 0 | 0 | 0 | 0.84 | 0.38 | 1.87 | 0.664 | 0.952 | 0.11 | 0.74 | 117.87 | 0.84 | 0.84 | 0.37 | 1.88 | 0.668 | 0.947 | - | - | - | - | - | - | - | - | - | - | - | - |
| SLURP1 | OID3040 | Cardiom | SLURP1 | P55000 | 8 | 142740949 | 142742406 | 16 | 0 | 0 | 0 | 0 | 1.06 | 0.85 | 1.33 | 0.608 | 0.952 | 10.61 | 0.78 | 196.51 | 1.06 | 1.06 | 0.85 | 1.33 | 0.609 | 0.939 | 1.06 | 0.81 | 1.39 | 0.686 | 1.06 | 0.72 | 1.58 | 0.756 | 0.00 | -0.07 | 0.07 | 0.981 |
| ABL1 | OID2128 | Oncology | ABL1 | P00519 | 9 | 130713016 | 130887675 | 1 | 0 | 0 | 0 | 0 | 0.62 | 0.07 | 5.33 | 0.664 | 0.952 | - | - | 31.50 | 0.62 | 0.62 | 0.06 | 5.98 | 0.681 | 0.947 | - | - | - | - | - | - | - | - | - | - | - | - |
| ANGPTL | OID2072 | Inflamma | ANGPTL | Q9UKU9 | 9 | 127087348 | 127122635 | 1 | 0 | 0 | 0 | 0 | 0.63 | 0.09 | 4.23 | 0.633 | 0.952 | - | - | 44.97 | 0.63 | 0.63 | 0.09 | 4.54 | 0.645 | 0.945 | - | - | - | - | - | - | - | - | - | - | - | - |
| EDF1 | OID3101 | Neurolog | EDF1 | O60869 | 9 | 136862119 | 136866308 | 1 | 0 | 0 | 0 | 0 | 1.81 | 0.19 | 16.91 | 0.602 | 0.952 | - | - | 30.85 | 1.81 | 1.81 | 0.17 | 19.17 | 0.621 | 0.944 | - | - | - | - | - | - | - | - | - | - | - | - |
| NPDC1 | OID2020 | Cardiom | NPDC1 | Q9NQX5 | 9 | 137039463 | 137046179 | 4 | 0 | 0 | 0 | 0 | 1.16 | 0.65 | 2.07 | 0.625 | 0.952 | 0.90 | 0.83 | 121.70 | 1.16 | 1.16 | 0.64 | 2.09 | 0.628 | 0.945 | 1.19 | 0.61 | 2.32 | 0.618 | 1.33 | 0.32 | 5.46 | 0.694 | -0.02 | -0.16 | 0.13 | 0.833 |
| OMD | OID2072 | Inflamma | OMD | Q99983 | 9 | 92412380 | 92424471 | 3 | 0 | 0 | 1 | 0 | 0.79 | 0.31 | 2.04 | 0.630 | 0.952 | 0.29 | 0.86 | 64.69 | 0.79 | 0.79 | 0.30 | 2.09 | 0.638 | 0.945 | 0.78 | 0.28 | 2.19 | 0.636 | 0.26 | 0.00 | 27.81 | 0.569 | 0.09 | -0.28 | 0.46 | 0.630 |
| PALM2 | OID3109 | Neurolog | PALM2AI | Q8IXS6 | 9 | 109498325 | 110172512 | 5 | 0 | 0 | 0 | 0 | 0.92 | 0.62 | 1.36 | 0.663 | 0.952 | 1.51 | 0.82 | 214.66 | 0.92 | 0.92 | 0.61 | 1.36 | 0.664 | 0.947 | 0.90 | 0.58 | 1.38 | 0.624 | 0.84 | 0.42 | 1.68 | 0.624 | 0.01 | -0.08 | 0.10 | 0.770 |
| PAXX | OID3059 | Inflamma | PAXX | Q9BUH6 | 9 | 136992422 | 136993984 | 2 | 0 | 0 | 0 | 0 | 1.16 | 0.65 | 2.08 | 0.617 | 0.952 | 0.24 | 0.62 | 229.30 | 1.16 | 1.16 | 0.65 | 2.09 | 0.619 | 0.943 | - | - | - | - | - | - | - | - | - | - | - | - |
| STXBP1 | OID3105 | Neurolog | STXBP1 | P61764 | 9 | 127579370 | 127696027 | 1 | 0 | 0 | 0 | 0 | 0.65 | 0.11 | 3.81 | 0.635 | 0.952 | - | - | 50.56 | 0.65 | 0.65 | 0.11 | 4.04 | 0.646 | 0.945 | - | - | - | - | - | - | - | - | - | - | - | - |
| TEK | OID2149 | Oncology | TEK | Q02763 | 9 | 27109141 | 27230174 | 16 | 0 | 0 | 0 | 0 | 1.04 | 0.88 | 1.23 | 0.620 | 0.952 | 18.22 | 0.25 | 421.17 | 1.04 | 1.04 | 0.90 | 1.22 | 0.585 | 0.934 | 0.93 | 0.74 | 1.16 | 0.500 | 1.10 | 0.88 | 1.37 | 0.411 | -0.02 | -0.08 | 0.04 | 0.483 |
| TGFBR1 | OID3050 | Inflamma | TGFBR1 | P36897 | 9 | 99104038 | 99154192 | 1 | 0 | 0 | 0 | 0 | 0.65 | 0.09 | 4.58 | 0.664 | 0.952 | - | - | 38.68 | 0.65 | 0.65 | 0.08 | 4.99 | 0.677 | 0.947 | - | - | - | - | - | - | - | - | - | - | - | - |
| ACTA2 | OID2007 | Cardiom | ACTA2 | P62736 | 10 | 88935074 | 88991339 | 1 | 0 | 0 | 0 | 0 | 1.52 | 0.32 | 7.28 | 0.603 | 0.952 | - | - | 77.84 | 1.52 | 1.52 | 0.31 | 7.53 | 0.611 | 0.939 | - | - | - | - | - | - | - | - | - | - | - | - |
| CDHR1 | OID2080 | Neurolog | CDHR1 | Q96JP9 | 10 | 84194537 | 84219621 | 9 | 0 | 0 | 0 | 0 | 0.91 | 0.65 | 1.29 | 0.613 | 0.952 | 9.94 | 0.27 | 165.06 | 0.91 | 0.91 | 0.67 | 1.25 | 0.573 | 0.934 | 0.81 | 0.55 | 1.18 | 0.266 | 0.61 | 0.33 | 1.13 | 0.118 | 0.08 | -0.02 | 0.18 | 0.133 |
| CDNF | OID2131 | Oncology | CDNF | Q49AH0 | 10 | 14819245 | 14838575 | 11 | 0 | 0 | 0 | 0 | 1.06 | 0.81 | 1.40 | 0.656 | 0.952 | 18.45 | 0.05 | 367.21 | 1.07 | 1.07 | 0.87 | 1.31 | 0.544 | 0.932 | 1.07 | 0.86 | 1.34 | 0.539 | 1.01 | 0.71 | 1.44 | 0.955 | 0.02 | -0.06 | 0.10 | 0.616 |
| DKK1 | OID2106 | Neurolog | DKK1 | O94907 | 10 | 52314281 | 52318042 | 4 | 0 | 0 | 0 | 0 | 0.79 | 0.30 | 2.09 | 0.639 | 0.952 | 6.76 | 0.08 | 100.45 | 0.79 | 0.79 | 0.41 | 1.51 | 0.477 | 0.910 | 0.52 | 0.23 | 1.18 | 0.118 | 0.27 | 0.00 | 27.31 | 0.575 | 0.11 | -0.35 | 0.57 | 0.633 |
| CAT | OID3071 | Inflamma | CAT | P04040 | 11 | 34438934 | 34472060 | 4 | 0 | 0 | 0 | 0 | 0.89 | 0.55 | 1.45 | 0.648 | 0.952 | 1.71 | 0.63 | 183.77 | 0.89 | 0.89 | 0.55 | 1.45 | 0.649 | 0.945 | 0.83 | 0.49 | 1.42 | 0.505 | 0.55 | 0.18 | 1.69 | 0.295 | 0.06 | -0.07 | 0.19 | 0.347 |
| F2 | OID3072 | Inflamma | F2 | P00734 | 11 | 46719196 | 46739506 | 8 | 0 | 0 | 0 | 0 | 0.90 | 0.59 | 1.39 | 0.646 | 0.952 | 5.88 | 0.55 | 97.40 | 0.90 | 0.90 | 0.59 | 1.39 | 0.647 | 0.945 | 1.00 | 0.59 | 1.72 | 0.990 | 1.03 | 0.56 | 1.90 | 0.918 | -0.02 | -0.10 | 0.05 | 0.553 |
| PRDX5 | OID2064 | Inflamma | PRDX5 | P30044 | 11 | 64318121 | 64321811 | 5 | 0 | 0 | 1 | 0 | 1.29 | 0.47 | 3.58 | 0.619 | 0.952 | 7.70 | 0.10 | 56.43 | 1.31 | 1.31 | 0.62 | 2.73 | 0.480 | 0.911 | 0.93 | 0.35 | 2.45 | 0.886 | 2.53 | 0.16 | 39.98 | 0.509 | -0.09 | -0.41 | 0.24 | 0.601 |
| USP28 | OID3108 | Neurolog | USP28 | Q96RU2 | 11 | 113797874 | 113875572 | 2 | 0 | 0 | 0 | 0 | 0.79 | 0.31 | 2.00 | 0.621 | 0.952 | 0.62 | 0.43 | 85.59 | 0.79 | 0.79 | 0.31 | 2.03 | 0.626 | 0.945 | - | - | - | - | - | - | - | - | - | - | - | - |
| CD4 | OID2058 | Inflamma | CD4 | P01730 | 12 | 6786858 | 6820799 | 5 | 0 | 0 | 0 | 0 | 0.93 | 0.68 | 1.27 | 0.655 | 0.952 | 2.39 | 0.66 | 329.64 | 0.93 | 0.93 | 0.68 | 1.27 | 0.655 | 0.947 | 0.92 | 0.67 | 1.27 | 0.621 | 0.75 | 0.41 | 1.40 | 0.369 | 0.06 | -0.10 | 0.22 | 0.436 |
| CLEC12A | OID3055 | Inflamma | CLEC12 | Q5QGZ9 | 12 | 9951316 | 9995694 | 18 | 0 | 0 | 0 | 0 | 1.05 | 0.86 | 1.28 | 0.630 | 0.952 | 14.28 | 0.65 | 196.04 | 1.05 | 1.05 | 0.86 | 1.29 | 0.631 | 0.945 | 1.21 | 0.94 | 1.56 | 0.138 | 1.23 | 0.88 | 1.73 | 0.222 | -0.04 | -0.10 | 0.03 | 0.245 |
| ENO2 | OID2104 | Neurolog | ENO2 | P09104 | 12 | 6913745 | 6923698 | 2 | 0 | 0 | 0 | 0 | 1.18 | 0.60 | 2.34 | 0.628 | 0.952 | 1.66 | 0.20 | 168.78 | 1.19 | 1.19 | 0.60 | 2.36 | 0.628 | 0.945 | - | - | - | - | - | - | - | - | - | - | - | - |
| GLT8D2 | OID2082 | Neurolog | GLT8D2 | Q9H1C3 | 12 | 103988987 | 104064183 | 2 | 0 | 0 | 0 | 0 | 1.13 | 0.69 | 1.84 | 0.632 | 0.952 | 0.16 | 0.69 | 275.18 | 1.13 | 1.13 | 0.69 | 1.84 | 0.634 | 0.945 | - | - | - | - | - | - | - | - | - | - | - | - |
| HEBP1 | OID2006 | Cardiom | HEBP1 | Q9NRV9 | 12 | 12974870 | 13000265 | 1 | 0 | 0 | 0 | 0 | 1.21 | 0.54 | 2.68 | 0.643 | 0.952 | - | - | 252.58 | 1.21 | 1.21 | 0.54 | 2.69 | 0.645 | 0.945 | - | - | - | - | - | - | - | - | - | - | - | - |
| LTA4H | OID2145 | Oncology | LTA4H | P09960 | 12 | 96000753 | 96043520 | 2 | 0 | 0 | 0 | 0 | 0.81 | 0.33 | 2.00 | 0.651 | 0.952 | 0.00 | 0.99 | 101.63 | 0.81 | 0.81 | 0.33 | 2.03 | 0.656 | 0.947 | - | - | - | - | - | - | - | - | - | - | - | - |
| METAP2 | OID2143 | Oncology | METAP2 | P50579 | 12 | 95473520 | 95515839 | 1 | 0 | 0 | 0 | 0 | 1.80 | 0.19 | 16.98 | 0.608 | 0.952 | - | - | 31.55 | 1.80 | 1.80 | 0.17 | 19.19 | 0.627 | 0.945 | - | - | - | - | - | - | - | - | - | - | - | - |
| PTPRB | OID3032 | Cardiom | PTPRB | P23467 | 12 | 70515870 | 70637440 | 19 | 0 | 0 | 0 | 0 | 0.96 | 0.79 | 1.16 | 0.662 | 0.952 | 19.57 | 0.36 | 256.18 | 0.96 | 0.96 | 0.80 | 1.15 | 0.648 | 0.945 | 0.97 | 0.76 | 1.23 | 0.790 | 0.86 | 0.59 | 1.25 | 0.430 | 0.03 | -0.05 | 0.10 | 0.509 |
| PTPRR | OID3109 | Neurolog | PTPRR | Q15256 | 12 | 70638073 | 70920738 | 1 | 0 | 0 | 0 | 0 | 1.81 | 0.17 | 19.07 | 0.621 | 0.952 | - | - | 30.05 | 1.81 | 1.81 | 0.15 | 21.81 | 0.640 | 0.945 | - | - | - | - | - | - | - | - | - | - | - | - |
| GGACT | OID3029 | Cardiom | GGACT | Q9BVM4 | 13 | 100530164 | 100589528 | 18 | 0 | 0 | 0 | 0 | 0.95 | 0.77 | 1.18 | 0.641 | 0.952 | 11.31 | 0.84 | 188.69 | 0.95 | 0.95 | 0.77 | 1.18 | 0.641 | 0.945 | 1.01 | 0.79 | 1.30 | 0.918 | 1.05 | 0.72 | 1.52 | 0.807 | -0.02 | -0.10 | 0.05 | 0.529 |
| TNFRSF1 | OID2144 | Oncology | TNFRSF1 | Q9NS68 | 13 | 23570370 | 23676104 | 12 | 0 | 0 | 0 | 0 | 0.93 | 0.70 | 1.25 | 0.648 | 0.952 | 11.01 | 0.44 | 170.16 | 0.93 | 0.93 | 0.70 | 1.25 | 0.648 | 0.945 | 0.96 | 0.67 | 1.38 | 0.845 | 1.27 | 0.74 | 2.18 | 0.390 | -0.06 | -0.15 | 0.03 | 0.189 |
| GMPR2 | OID3062 | Inflamma | GMPR2 | Q9P2T1 | 14 | 24232422 | 24239242 | 3 | 0 | 0 | 1 | 0 | 0.78 | 0.27 | 2.28 | 0.654 | 0.952 | 4.08 | 0.13 | 52.60 | 0.78 | 0.78 | 0.26 | 2.31 | 0.651 | 0.945 | 0.61 | 0.18 | 2.10 | 0.432 | 0.02 | 0.00 | 1.53 | 0.077 | 0.47 | -0.07 | 1.01 | 0.086 |
| MAP4K5 | OID2089 | Neurolog | MAP4K5 | Q9Y4K4 | 14 | 50418501 | 50561126 | 1 | 0 | 0 | 0 | 0 | 1.53 | 0.23 | 10.27 | 0.661 | 0.952 | - | - | 42.62 | 1.53 | 1.53 | 0.21 | 11.06 | 0.673 | 0.947 | - | - | - | - | - | - | - | - | - | - | - | - |
| SERPINA | OID3076 | Inflamma | SERPINA | P05154 | 14 | 94561442 | 94593118 | 7 | 0 | 0 | 0 | 0 | 0.91 | 0.63 | 1.31 | 0.604 | 0.952 | 2.92 | 0.82 | 171.65 | 0.91 | 0.91 | 0.63 | 1.31 | 0.606 | 0.939 | 0.81 | 0.52 | 1.26 | 0.352 | 0.71 | 0.30 | 1.70 | 0.444 | 0.04 | -0.09 | 0.17 | 0.546 |
| ISLR2 | OID2088 | Neurolog | ISLR2 | Q6UXK2 | 15 | 74100311 | 74138540 | 13 | 0 | 0 | 0 | 0 | 1.08 | 0.78 | 1.51 | 0.645 | 0.952 | 8.75 | 0.72 | 122.96 | 1.08 | 1.08 | 0.77 | 1.51 | 0.646 | 0.945 | 1.23 | 0.81 | 1.86 | 0.335 | 2.04 | 0.78 | 5.30 | 0.146 | -0.11 | -0.27 | 0.05 | 0.167 |
| SRP14 | OID2132 | Oncology | SRP14 | P37108 | 15 | 40035690 | 40039181 | 1 | 0 | 0 | 0 | 0 | 0.63 | 0.09 | 4.60 | 0.649 | 0.952 | - | - | 42.61 | 0.63 | 0.63 | 0.08 | 4.97 | 0.661 | 0.947 | - | - | - | - | - | - | - | - | - | - | - | - |
| CDH15 | OID2100 | Neurolog | CDH15 | P55291 | 16 | 89171748 | 89195492 | 20 | 0 | 0 | 0 | 0 | 1.06 | 0.84 | 1.35 | 0.625 | 0.952 | 13.07 | 0.83 | 186.85 | 1.06 | 1.06 | 0.84 | 1.35 | 0.626 | 0.945 | 1.12 | 0.82 | 1.53 | 0.468 | 0.97 | 0.60 | 1.58 | 0.916 | 0.02 | -0.08 | 0.12 | 0.689 |
| CES3 | OID2134 | Oncology | CES3 | Q6UWW8 | 16 | 66961245 | 66975149 | 11 | 0 | 0 | 0 | 0 | 1.08 | 0.78 | 1.49 | 0.659 | 0.952 | 7.94 | 0.63 | 142.16 | 1.08 | 1.08 | 0.78 | 1.49 | 0.659 | 0.947 | 1.04 | 0.68 | 1.59 | 0.866 | 0.95 | 0.59 | 1.53 | 0.819 | 0.04 | -0.06 | 0.13 | 0.472 |
| CRYM | OID3081 | Neurolog | CRYM | Q14894 | 16 | 21238874 | 21303083 | 20 | 0 | 0 | 0 | 0 | 1.05 | 0.84 | 1.32 | 0.654 | 0.952 | 19.70 | 0.41 | 150.08 | 1.05 | 1.05 | 0.84 | 1.32 | 0.648 | 0.945 | 1.04 | 0.80 | 1.36 | 0.775 | 1.04 | 0.70 | 1.52 | 0.856 | 0.00 | -0.07 | 0.07 | 0.919 |
| HAGH | OID2145 | Oncology | HAGH | Q16775 | 16 | 1795620 | 1827157 | 3 | 0 | 0 | 1 | 0 | 1.21 | 0.53 | 2.73 | 0.653 | 0.952 | 2.61 | 0.27 | 98.83 | 1.21 | 1.21 | 0.53 | 2.76 | 0.655 | 0.947 | 1.07 | 0.41 | 2.76 | 0.892 | 4.98 | 0.62 | 39.89 | 0.130 | -0.22 | -0.51 | 0.08 | 0.146 |
| MTHFSD | OID3130 | Oncology | MTHFSD | Q2M296 | 16 | 86530178 | 86555235 | 16 | 0 | 0 | 0 | 0 | 1.05 | 0.87 | 1.26 | 0.605 | 0.952 | 8.35 | 0.91 | 293.24 | 1.05 | 1.05 | 0.87 | 1.26 | 0.605 | 0.939 | 1.06 | 0.86 | 1.30 | 0.584 | 0.95 | 0.73 | 1.23 | 0.680 | 0.03 | -0.03 | 0.09 | 0.285 |
| OTOA | OID3082 | Neurolog | OTOA | Q7RTW8 | 16 | 21663968 | 21761935 | 1 | 0 | 0 | 0 | 0 | 1.55 | 0.28 | 8.43 | 0.614 | 0.952 | - | - | 52.59 | 1.55 | 1.55 | 0.27 | 8.90 | 0.625 | 0.945 | - | - | - | - | - | - | - | - | - | - | - | - |
| PYDC1 | OID3020 | Cardiom | PYDC1 | Q8WXC3 | 16 | 31215962 | 31217135 | 16 | 0 | 0 | 0 | 0 | 1.08 | 0.81 | 1.42 | 0.611 | 0.952 | 12.73 | 0.62 | 160.04 | 1.08 | 1.08 | 0.81 | 1.43 | 0.612 | 0.939 | 1.07 | 0.76 | 1.52 | 0.697 | 1.06 | 0.57 | 1.96 | 0.863 | 0.00 | -0.11 | 0.12 | 0.948 |
| AOC3 | OID2038 | Cardiom | AOC3 | Q16853 | 17 | 42851184 | 42858130 | 23 | 0 | 0 | 0 | 0 | 1.04 | 0.89 | 1.21 | 0.625 | 0.952 | 15.98 | 0.82 | 325.82 | 1.04 | 1.04 | 0.89 | 1.21 | 0.626 | 0.945 | 0.98 | 0.79 | 1.21 | 0.843 | 1.04 | 0.82 | 1.32 | 0.727 | 0.00 | -0.07 | 0.06 | 0.961 |
| CCL16 | OID2033 | Cardiom | CCL16 | O15467 | 17 | 35976493 | 35981497 | 21 | 0 | 0 | 0 | 0 | 0.97 | 0.87 | 1.09 | 0.610 | 0.952 | 15.26 | 0.76 | 566.17 | 0.97 | 0.97 | 0.87 | 1.09 | 0.610 | 0.939 | 0.94 | 0.81 | 1.08 | 0.369 | 0.96 | 0.84 | 1.10 | 0.589 | 0.00 | -0.04 | 0.05 | 0.839 |
| CHAD | OID3066 | Inflamma | CHAD | O15335 | 17 | 50464496 | 50468906 | 3 | 0 | 0 | 0 | 0 | 0.90 | 0.58 | 1.39 | 0.628 | 0.952 | 0.11 | 0.95 | 262.30 | 0.90 | 0.90 | 0.58 | 1.40 | 0.630 | 0.945 | 0.91 | 0.58 | 1.42 | 0.669 | 0.92 | 0.55 | 1.53 | 0.744 | -0.01 | -0.13 | 0.11 | 0.859 |
| CHMP6 | OID3032 | Cardiom | CHMP6 | Q96FZ7 | 17 | 80991598 | 81009517 | 2 | 0 | 0 | 0 | 0 | 1.14 | 0.65 | 2.01 | 0.656 | 0.952 | 0.21 | 0.64 | 243.81 | 1.14 | 1.14 | 0.64 | 2.01 | 0.657 | 0.947 | - | - | - | - | - | - | - | - | - | - | - | - |
| KIF1C | OID3016 | Cardiom | KIF1C | O43896 | 17 | 4997950 | 5028401 | 1 | 0 | 0 | 0 | 0 | 1.60 | 0.19 | 13.48 | 0.663 | 0.952 | - | - | 34.71 | 1.60 | 1.60 | 0.17 | 14.94 | 0.678 | 0.947 | - | - | - | - | - | - | - | - | - | - | - | - |
| SCARF1 | OID2029 | Cardiom | SCARF1 | Q14162 | 17 | 1633858 | 1645744 | 16 | 0 | 0 | 0 | 0 | 1.06 | 0.83 | 1.36 | 0.636 | 0.952 | 13.60 | 0.56 | 169.04 | 1.06 | 1.06 | 0.83 | 1.36 | 0.637 | 0.945 | 1.03 | 0.76 | 1.39 | 0.843 | 1.09 | 0.69 | 1.73 | 0.709 | -0.01 | -0.10 | 0.09 | 0.887 |
| SDK2 | OID3066 | Inflamma | SDK2 | Q58EX2 | 17 | 73334384 | 73644445 | 18 | 0 | 0 | 0 | 0 | 0.94 | 0.73 | 1.22 | 0.648 | 0.952 | 25.54 | 0.08 | 192.21 | 0.94 | 0.94 | 0.76 | 1.16 | 0.574 | 0.934 | 0.96 | 0.72 | 1.27 | 0.757 | 0.78 | 0.47 | 1.29 | 0.330 | 0.04 | -0.05 | 0.14 | 0.387 |
| SLC9A3R | OID3063 | Inflamma | SLC9A3R | O14745 | 17 | 74748628 | 74769353 | 1 | 0 | 0 | 0 | 0 | 1.29 | 0.44 | 3.75 | 0.640 | 0.952 | - | - | 133.81 | 1.29 | 1.29 | 0.44 | 3.79 | 0.644 | 0.945 | - | - | - | - | - | - | - | - | - | - | - | - |
| PSTPIP2 | OID3059 | Inflamma | PSTPIP2 | Q9H939 | 18 | 45983536 | 46072272 | 2 | 0 | 0 | 0 | 0 | 1.16 | 0.65 | 2.07 | 0.622 | 0.952 | 0.12 | 0.73 | 224.66 | 1.16 | 1.16 | 0.64 | 2.08 | 0.624 | 0.945 | - | - | - | - | - | - | - | - | - | - | - | - |
| PTPRM | OID2059 | Inflamma | PTPRM | P28827 | 18 | 7566782 | 8406861 | 6 | 0 | 0 | 1 | 0 | 0.81 | 0.33 | 1.98 | 0.637 | 0.952 | 6.96 | 0.22 | 63.14 | 0.80 | 0.80 | 0.37 | 1.74 | 0.576 | 0.934 | 1.26 | 0.46 | 3.49 | 0.654 | 0.37 | 0.04 | 3.76 | 0.398 | 0.07 | -0.11 | 0.24 | 0.468 |

| | | | | | | | | | | | | | | | | |
|---|---|---|---|---|---|---|---|---|---|---|---|---|---|---|---|---|
| APOC1 | OID3074 | Inflamma | APOC1 | P02654 | | 19 | 44914247 | 44919349 | 10 | 0 | 1 | 0 | | 0 | 0.88 | [illegible] |
| ARHGAP | OID3065 | Inflamma | ARHGAP | Q92619 | | 19 | 1065923 | 1086628 | 1 | 0 | 0 | 0 | | 0 | 0.72 | [illegible] |
| BLVRB | OID2114 | Neurolog | BLVRB | P30043 | | 19 | 40447765 | 40465764 | 4 | 0 | 0 | 0 | | 0 | 1.17 | [illegible] |
| CCL25 | OID2067 | Inflamma | CCL25 | O15444 | | 19 | 8052318 | 8062660 | 25 | 0 | 0 | 0 | | 0 | 0.97 | [illegible] |
| CD209 | OID2026 | Cardiom | CD209 | Q9NNX6 | | 19 | 7739993 | 7747564 | 15 | 0 | 0 | 0 | | 0 | 1.06 | [illegible] |
| CEACAM | OID3144 | Oncology | CEACAM | Q7Z692 | | 19 | 44662278 | 44684359 | 2 | 0 | 0 | 0 | | 0 | 0.79 | [illegible] |
| CEACAM | OID2117 | Oncology | CEACAM | P06731 | | 19 | 41708585 | 41730433 | 9 | 0 | 0 | 0 | | 0 | 1.06 | [illegible] |
| ICAM3 | OID2027 | Cardiom | ICAM3 | P32942 | | 19 | 10333776 | 10339661 | 12 | 0 | 0 | 0 | | 0 | 1.07 | [illegible] |
| ICAM4 | OID2057 | Inflamma | ICAM4 | Q14773 | | 19 | 10286955 | 10288522 | 2 | 0 | 0 | 0 | | 0 | 1.23 | [illegible] |
| LILRA6 | OID3034 | Cardiom | LILRA6 | Q6PI73 | | 19 | 54236592 | 54242791 | 32 | 0 | 0 | 0 | | 0 | 0.98 | [illegible] |
| MYBPC2 | OID3019 | Cardiom | MYBPC2 | Q14324 | | 19 | 50432892 | 50466321 | 1 | 0 | 0 | 0 | | 0 | 1.68 | [illegible] |
| NCAN | OID2105 | Neurolog | NCAN | O14594 | | 19 | 19211958 | 19252233 | 7 | 0 | 0 | 0 | | 0 | 1.13 | [illegible] |
| NTF4 | OID2120 | Oncology | NTF4 | P34130 | | 19 | 49061066 | 49065054 | 1 | 0 | 0 | 0 | | 0 | 0.61 | [illegible] |
| PEPD | OID3068 | Inflamma | PEPD | P12955 | | 19 | 33386950 | 33521823 | 12 | 0 | 0 | 0 | | 0 | 0.95 | [illegible] |
| PLAUR | OID2076 | Inflamma | PLAUR | Q03405 | | 19 | 43646095 | 43670547 | 4 | 0 | 0 | 0 | | 0 | 0.87 | [illegible] |
| PRTN3 | OID2032 | Cardiom | PRTN3 | P24158 | | 19 | 840999 | 848175 | 16 | 0 | 0 | 0 | | 0 | 0.94 | [illegible] |
| RTBDN | OID2134 | Oncology | RTBDN | Q9BSG5 | | 19 | 12825478 | 12835428 | 2 | 0 | 0 | 0 | | 0 | 0.86 | [illegible] |
| SIGLEC8 | OID3026 | Cardiom | SIGLEC8 | Q9NYZ4 | | 19 | 51450847 | 51458456 | 21 | 0 | 0 | 0 | | 0 | 1.07 | [illegible] |
| TBCB | OID2099 | Neurolog | TBCB | Q99426 | | 19 | 36114289 | 36125947 | 1 | 0 | 0 | 0 | | 0 | 1.59 | [illegible] |
| CEBPB | OID2005 | Cardiom | CEBPB | P17676 | | 20 | 50190830 | 50192668 | 1 | 0 | 0 | 0 | | 0 | 1.57 | [illegible] |
| CST3 | OID2040 | Cardiom | CST3 | P01034 | | 20 | 23626706 | 23638473 | 6 | 0 | 0 | 0 | | 0 | 0.93 | [illegible] |
| PRND | OID3112 | Neurolog | PRND | Q9UKY0 | | 20 | 4721909 | 4728460 | 8 | 0 | 0 | 0 | | 0 | 0.94 | [illegible] |
| TNFRSF6 | OID2096 | Neurolog | TNFRSF6 | O95407 | | 20 | 63696652 | 63698684 | 18 | 0 | 0 | 0 | | 0 | 0.95 | [illegible] |
| ADAMTS | OID3052 | Inflamma | ADAMTS | Q9UHI8 | | 21 | 26835755 | 26845409 | 1 | 0 | 0 | 0 | | 0 | 1.66 | [illegible] |
| COL18A1 | OID2033 | Cardiom | COL18A1 | P39060 | | 21 | 45405165 | 45513720 | 6 | 0 | 0 | 0 | | 0 | 1.15 | [illegible] |
| CRYBB1 | OID3031 | Cardiom | CRYBB1 | P53674 | | 22 | 26599278 | 26618027 | 3 | 0 | 0 | 0 | | 0 | 0.86 | [illegible] |
| DDT | OID3037 | Cardiom | DDT | P30046 | | 22 | 23970542 | 23980525 | 3 | 0 | 0 | 0 | | 0 | 1.11 | [illegible] |
| SUSD2 | OID2109 | Neurolog | SUSD2 | Q9UGT4 | | 22 | 24181487 | 24189106 | 19 | 0 | 0 | 0 | | 0 | 1.04 | [illegible] |
| AMY1A | OID3070 | Inflamma | AMY1A;A | P0DUB6 | 1;1;1 | | 103655760 | 103664554 | 30 | 0 | 0 | 0 | | 0 | 0.96 | [illegible] |
| FUT3_FU | OID2101 | Neurolog | FUT3;FU | P21217;( | 19;19 | | 5842888;58 | 5851471;58 | 23 | 0 | 0 | 0 | | 0 | 0.96 | [illegible] |
| LGALS7 | OID2140 | Oncology | LGALS7B | P47929;F | 19;19 | | 38789200;3 | 38791754;3 | 8 | 0 | 0 | 0 | | 0 | 1.09 | [illegible] |
| PTX3 | OID2057 | Inflamma | PTX3 | P26022 | | 3 | 157436850 | 157443633 | 3 | 0 | 0 | 1 | | 0 | 0.80 | [illegible] |
| AHNAK2 | OID3024 | Cardiom | AHNAK2 | Q8IVF2 | | 14 | 104937244 | 104978374 | 2 | 0 | 0 | 0 | | 0 | 1.10 | [illegible] |
| CR2 | OID2039 | Cardiom | CR2 | P20023 | | 1 | 207454230 | 207489895 | 6 | 0 | 0 | 0 | | 0 | 0.91 | [illegible] |
| PACS2 | OID3018 | Cardiom | PACS2 | Q86VP3 | | 14 | 105300563 | 105398147 | 1 | 0 | 0 | 0 | | 0 | 1.13 | [illegible] |
| GFAP | OID2124 | Oncology | GFAP | P14136 | | 17 | 44903159 | 44916937 | 4 | 0 | 0 | 1 | | 0 | 1.26 | [illegible] |
| NOS2 | OID3008 | Cardiom | NOS2 | P35228 | | 17 | 27756766 | 27800529 | 2 | 0 | 0 | 0 | | 0 | 1.23 | [illegible] |
| MSMB | OID2027 | Cardiom | MSMB | P08118 | | 10 | 46033307 | 46048180 | 52 | 0 | 0 | 0 | | 0 | 0.98 | [illegible] |
| CLUL1 | OID2019 | Cardiom | CLUL1 | Q15846 | | 18 | 596988 | 650334 | 20 | 0 | 0 | 0 | | 0 | 1.03 | [illegible] |
| IL36A | OID3047 | Inflamma | IL36A | Q9UHA7 | | 2 | 113005459 | 113008044 | 1 | 0 | 0 | 0 | | 0 | 0.70 | [illegible] |
| HEG1 | OID3057 | Inflamma | HEG1 | Q9ULI3 | | 3 | 124965710 | 125055997 | 3 | 0 | 0 | 0 | | 0 | 0.87 | [illegible] |
| CGA | OID2106 | Neurolog | CGA | P01215 | | 6 | 87085498 | 87095106 | 1 | 0 | 0 | 0 | | 0 | 1.81 | [illegible] |
| EPHB6 | OID2098 | Neurolog | EPHB6 | O15197 | | 7 | 142855061 | 142871094 | 5 | 0 | 0 | 0 | | 0 | 0.93 | [illegible] |
| PIK3AP1 | OID2058 | Inflamma | PIK3AP1 | Q6ZUJ8 | | 10 | 96593315 | 96720514 | 6 | 0 | 0 | 0 | | 0 | 0.93 | [illegible] |
| MANSC4 | OID3014 | Cardiom | MANSC4 | A6NHS7 | | 12 | 27762427 | 27780236 | 27 | 0 | 0 | 0 | | 0 | 1.04 | [illegible] |
| MEP1B | OID2016 | Cardiom | MEP1B | Q16820 | | 18 | 32185069 | 32220404 | 37 | 0 | 0 | 0 | | 0 | 1.02 | [illegible] |
| ANKMY2 | OID3051 | Inflamma | ANKMY2 | Q8IV38 | | 7 | 16599779 | 16645817 | 1 | 0 | 0 | 0 | | 0 | 0.77 | [illegible] |
| IL22RA1 | OID2044 | Inflamma | IL22RA1 | Q8N6P7 | | 1 | 24119771 | 24143140 | 5 | 0 | 0 | 0 | | 0 | 1.08 | [illegible] |
| FGF2 | OID2050 | Inflamma | FGF2 | P09038 | | 4 | 122826708 | 122898236 | 44 | 0 | 0 | 0 | | 0 | 0.97 | [illegible] |
| PDGFRB | OID2026 | Cardiom | PDGFRB | P09619 | | 5 | 150113839 | 150155872 | 36 | 0 | 0 | 0 | | 0 | 0.98 | [illegible] |
| ADAMTS | OID2024 | Cardiom | ADAMTS | Q76LX8 | | 9 | 133414358 | 133459402 | 12 | 0 | 0 | 0 | | 0 | 1.05 | [illegible] |
| CD72 | OID3064 | Inflamma | CD72 | P21854 | | 9 | 35609982 | 35646810 | 3 | 0 | 0 | 0 | | 0 | 1.13 | [illegible] |
| CLMP | OID2141 | Oncology | CLMP | Q9H6B4 | | 11 | 123069872 | 123195248 | 10 | 0 | 0 | 0 | | 0 | 1.05 | [illegible] |
| HYOU1 | OID2030 | Cardiom | HYOU1 | Q9Y4L1 | | 11 | 119044188 | 119057227 | 3 | 0 | 0 | 0 | | 0 | 0.90 | [illegible] |
| BAIAP2 | OID2125 | Oncology | BAIAP2 | Q9UQB8 | | 17 | 81035122 | 81117434 | 1 | 0 | 0 | 0 | | 0 | 1.31 | [illegible] |
| ENO3 | OID3020 | Cardiom | ENO3 | P13929 | | 17 | 4948092 | 4957131 | 1 | 0 | 0 | 0 | | 0 | 1.26 | [illegible] |
| GIP | OID3028 | Cardiom | GIP | P09681 | | 17 | 48958554 | 48968596 | 1 | 0 | 0 | 0 | | 0 | 1.61 | [illegible] |
| LILRA3 | OID3037 | Cardiom | LILRA3 | Q8N6C8 | | 19 | 54294163 | 54306461 | 25 | 0 | 0 | 0 | | 0 | 0.97 | [illegible] |
| PROCR | OID3014 | Cardiom | PROCR | Q9UNN8 | | 20 | 35172072 | 35216240 | 9 | 0 | 0 | 0 | | 0 | 1.06 | [illegible] |
| CXADR | OID2056 | Inflamma | CXADR | P78310 | | 21 | 17513043 | 17593579 | 2 | 0 | 0 | 0 | | 0 | 0.85 | [illegible] |
| NUMB | OID3056 | Inflamma | NUMB | P49757 | | 14 | 73275107 | 73458617 | 3 | 0 | 0 | 1 | | 0 | 1.18 | [illegible] |
| BCL2L15 | OID3049 | Inflamma | BCL2L15 | Q5TBC7 | | 1 | 113876816 | 113887581 | 8 | 0 | 0 | 0 | | 0 | 1.10 | [illegible] |
| SCG2 | OID2140 | Oncology | SCG2 | P13521 | | 2 | 223596940 | 223602361 | 1 | 0 | 0 | 0 | | 0 | 1.55 | [illegible] |
| FHIT | OID2087 | Neurolog | FHIT | P49789 | | 3 | 59747277 | 61251459 | 3 | 0 | 0 | 1 | | 0 | 0.86 | [illegible] |
| ADD1 | OID3065 | Inflamma | ADD1 | P35611 | | 4 | 2843844 | 2930076 | 2 | 0 | 0 | 0 | | 0 | 1.14 | [illegible] |
| ANXA3 | OID2103 | Neurolog | ANXA3 | P12429 | | 4 | 78551747 | 78610451 | 3 | 0 | 0 | 1 | | 0 | 1.19 | [illegible] |
| [illegible] | | | | | | | | | | | | | | | | |

| | | | | | | | | | | | | | | | |
|---|---|---|---|---|---|---|---|---|---|---|---|---|---|---|---|
| MCFD2 | OID2017 | Cardiom | MCFD2 | Q8NI22 | 2 | 46901870 | 46941855 | 2 | 0 | 0 | 0 | 0 | 0.92 | 0.47 | 1.81 |
| DCBLD2 | OID2147 | Oncology | DCBLD2 | Q96PD2 | 3 | 98795941 | 98901695 | 7 | 0 | 0 | 0 | 0 | 1.06 | 0.65 | 1.73 |
| FBLN2 | OID3037 | Cardiom | FBLN2 | P98095 | 3 | 13549125 | 13638422 | 8 | 0 | 0 | 0 | 0 | 0.96 | 0.67 | 1.37 |
| LRPAP1 | OID2105 | Neurolog | LRPAP1 | P30533 | 4 | 3503612 | 3532446 | 9 | 0 | 0 | 0 | 0 | 1.04 | 0.72 | 1.51 |
| PCDH7 | OID3145 | Oncology | PCDH7 | O60245 | 4 | 30720369 | 31146805 | 2 | 0 | 0 | 0 | 0 | 0.91 | 0.41 | 2.00 |
| IQGAP2 | OID2136 | Oncology | IQGAP2 | Q13576 | 5 | 76403285 | 76708132 | 1 | 0 | 0 | 0 | 0 | 1.09 | 0.56 | 2.14 |
| RGMB | OID2104 | Neurolog | RGMB | Q6NW40 | 5 | 98768650 | 98798643 | 9 | 0 | 0 | 0 | 0 | 0.95 | 0.60 | 1.49 |
| SIL1 | OID3020 | Cardiom | SIL1 | Q9H173 | 5 | 138946724 | 139293557 | 1 | 0 | 0 | 0 | 0 | 0.83 | 0.16 | 4.23 |
| LTB | OID3138 | Oncology | LTB | Q06643 | 6 | 31580525 | 31582522 | 10 | 0 | 0 | 0 | 0 | 0.96 | 0.71 | 1.31 |
| TNF | OID2007 | Cardiom | TNF | P01375 | 6 | 31575565 | 31578336 | 14 | 0 | 0 | 1 | 0 | 1.06 | 0.68 | 1.65 |
| TXNDC5 | OID2101 | Neurolog | TXNDC5 | Q8NBS9 | 6 | 7881517 | 7910788 | 3 | 0 | 0 | 1 | 0 | 1.09 | 0.56 | 2.12 |
| ARHGEF | OID3087 | Neurolog | ARHGEF | Q12774 | 7 | 144355288 | 144380632 | 27 | 0 | 0 | 0 | 1 | 0.98 | 0.84 | 1.15 |
| NT5C3A | OID2055 | Inflamma | NT5C3A | Q9H0P0 | 7 | 33014130 | 33062796 | 1 | 0 | 0 | 0 | 0 | 0.89 | 0.35 | 2.24 |
| SSC4D | OID2018 | Cardiom | SSC4D | Q8WTU2 | 7 | 76389334 | 76409697 | 13 | 0 | 0 | 0 | 0 | 1.03 | 0.79 | 1.34 |
| CPQ | OID3040 | Cardiom | CPQ | Q9Y646 | 8 | 96645242 | 97149654 | 20 | 0 | 0 | 0 | 0 | 0.97 | 0.73 | 1.28 |
| DKK4 | OID2093 | Neurolog | DKK4 | Q9UBT3 | 8 | 42374063 | 42377229 | 5 | 0 | 0 | 0 | 0 | 0.96 | 0.68 | 1.36 |
| PTGES2 | OID3056 | Inflamma | PTGES2 | Q9H7Z7 | 9 | 128120693 | 128128462 | 1 | 0 | 0 | 0 | 0 | 0.86 | 0.26 | 2.83 |
| SLC27A4 | OID2081 | Neurolog | SLC27A4 | Q6P1M0 | 9 | 128340527 | 128361470 | 1 | 0 | 0 | 0 | 0 | 0.81 | 0.13 | 4.93 |
| TYRP1 | OID3051 | Inflamma | TYRP1 | P17643 | 9 | 12685439 | 12710285 | 1 | 0 | 0 | 0 | 0 | 0.87 | 0.26 | 2.94 |
| BLNK | OID3054 | Inflamma | BLNK | Q8WV28 | 10 | 96189171 | 96271587 | 1 | 0 | 0 | 0 | 0 | 1.20 | 0.23 | 6.31 |
| ECHS1 | OID3023 | Cardiom | ECHS1 | P30084 | 10 | 133362485 | 133373354 | 5 | 0 | 0 | 0 | 0 | 0.95 | 0.61 | 1.48 |
| NUDT5 | OID2111 | Neurolog | NUDT5 | Q9UKK9 | 10 | 12165330 | 12196144 | 1 | 0 | 0 | 0 | 0 | 1.22 | 0.24 | 6.24 |
| ACRV1 | OID3051 | Inflamma | ACRV1 | P26436 | 11 | 125671522 | 125680874 | 1 | 0 | 0 | 0 | 0 | 1.27 | 0.20 | 7.94 |
| MMP10 | OID2068 | Inflamma | MMP10 | P09238 | 11 | 102770502 | 102780628 | 21 | 0 | 0 | 0 | 0 | 1.02 | 0.85 | 1.22 |
| TIMM10 | OID3035 | Cardiom | TIMM10 | P62072 | 11 | 57528464 | 57530803 | 4 | 0 | 0 | 0 | 0 | 0.95 | 0.60 | 1.49 |
| APOF | OID3070 | Inflamma | APOF | Q13790 | 12 | 56360568 | 56362857 | 5 | 0 | 0 | 0 | 0 | 0.95 | 0.61 | 1.48 |
| BCAT1 | OID3031 | Cardiom | BCAT1 | P54687 | 12 | 24810024 | 24949101 | 12 | 0 | 0 | 0 | 0 | 1.03 | 0.79 | 1.35 |
| CD69 | OID2020 | Cardiom | CD69 | Q07108 | 12 | 9752486 | 9760901 | 1 | 0 | 0 | 0 | 0 | 1.24 | 0.22 | 6.97 |
| CRADD | OID2092 | Neurolog | CRADD | P78560 | 12 | 93677375 | 93894840 | 1 | 0 | 0 | 0 | 0 | 1.18 | 0.32 | 4.37 |
| IL22 | OID3140 | Oncology | IL22 | Q9GZX6 | 12 | 68248242 | 68253604 | 2 | 0 | 0 | 0 | 0 | 1.11 | 0.45 | 2.74 |
| KLRK1 | OID3108 | Neurolog | KLRK1 | P26718 | 12 | 10372353 | 10391874 | 16 | 0 | 0 | 0 | 0 | 0.98 | 0.80 | 1.20 |
| LGALS3 | OID2028 | Cardiom | LGALS3 | P17931 | 14 | 55124110 | 55145423 | 34 | 0 | 0 | 0 | 0 | 0.99 | 0.87 | 1.12 |
| SMOC1 | OID2146 | Oncology | SMOC1 | Q9H4F8 | 14 | 69854131 | 70032366 | 14 | 0 | 0 | 0 | 0 | 1.03 | 0.80 | 1.33 |
| TCL1A | OID2098 | Neurolog | TCL1A | P56279 | 14 | 95709947 | 95714196 | 8 | 0 | 0 | 1 | 0 | 0.92 | 0.44 | 1.91 |
| CTSH | OID2011 | Cardiom | CTSH | P09668 | 15 | 78921058 | 78949574 | 27 | 0 | 0 | 0 | 0 | 1.01 | 0.91 | 1.13 |
| NTRK3 | OID2105 | Neurolog | NTRK3 | Q16288 | 15 | 87859751 | 88256768 | 7 | 0 | 0 | 0 | 0 | 1.04 | 0.74 | 1.48 |
| MAP2K6 | OID2055 | Inflamma | MAP2K6 | P52564 | 17 | 69414697 | 69553865 | 1 | 0 | 0 | 0 | 0 | 0.75 | 0.08 | 7.24 |
| AZU1 | OID2030 | Cardiom | AZU1 | P20160 | 19 | 825097 | 832018 | 3 | 0 | 0 | 0 | 0 | 0.91 | 0.41 | 2.03 |
| MUC16 | OID2137 | Oncology | MUC16 | Q8WXI7 | 19 | 8848844 | 8981342 | 3 | 0 | 0 | 1 | 0 | 0.90 | 0.38 | 2.16 |
| PTPRS | OID2030 | Cardiom | PTPRS | Q13332 | 19 | 5158495 | 5340812 | 20 | 0 | 0 | 0 | 0 | 1.03 | 0.82 | 1.29 |
| SPINT2 | OID2072 | Inflamma | SPINT2 | O43291 | 19 | 38244035 | 38292615 | 6 | 0 | 0 | 0 | 0 | 1.06 | 0.65 | 1.73 |
| SPINT3 | OID3150 | Oncology | SPINT3 | P49223 | 20 | 45512461 | 45515622 | 1 | 0 | 0 | 0 | 0 | 1.12 | 0.40 | 3.18 |
| PVALB | OID2142 | Oncology | PVALB | P20472 | 22 | 36800684 | 36819479 | 21 | 0 | 0 | 0 | 0 | 0.98 | 0.83 | 1.15 |
| CCL22 | OID2076 | Inflamma | CCL22 | O00626 | 16 | 57358783 | 57366189 | 3 | 0 | 0 | 1 | 0 | 0.95 | 0.60 | 1.50 |
| NPPC | OID2056 | Inflamma | NPPC | P23582 | 2 | 231921809 | 231926396 | 2 | 0 | 0 | 0 | 0 | 1.19 | 0.24 | 5.83 |
| STX8 | OID2053 | Inflamma | STX8 | Q9UNK0 | 17 | 9250471 | 9576591 | 1 | 0 | 0 | 0 | 0 | 0.89 | 0.30 | 2.62 |
| DNAJB1 | OID2136 | Oncology | DNAJB1 | P25685 | 19 | 14514769 | 14560391 | 1 | 0 | 0 | 0 | 0 | 1.21 | 0.21 | 7.02 |
| SEZ6L | OID2148 | Oncology | SEZ6L | Q9BYH1 | 22 | 26169462 | 26383597 | 8 | 0 | 0 | 1 | 0 | 1.06 | 0.64 | 1.74 |
| PSCA | OID3149 | Oncology | PSCA | O43653 | 8 | 142670308 | 142682725 | 45 | 0 | 0 | 0 | 0 | 1.01 | 0.92 | 1.11 |
| DLK1 | OID2029 | Cardiom | DLK1 | P80370 | 14 | 100725705 | 100738224 | 22 | 0 | 0 | 0 | 0 | 1.02 | 0.85 | 1.22 |
| ACP6 | OID2143 | Oncology | ACP6 | Q9NPH0 | 1 | 147629652 | 147670524 | 38 | 0 | 0 | 0 | 0 | 1.01 | 0.91 | 1.13 |
| DAG1 | OID2077 | Inflamma | DAG1 | Q14118 | 3 | 49468703 | 49535618 | 2 | 0 | 0 | 0 | 0 | 1.15 | 0.30 | 4.48 |
| EPHA1 | OID2067 | Inflamma | EPHA1 | P21709 | 7 | 143390289 | 143408856 | 11 | 0 | 0 | 0 | 0 | 0.96 | 0.69 | 1.36 |
| TPK1 | OID2033 | Cardiom | TPK1 | Q9H3S4 | 7 | 144451941 | 144836395 | 17 | 0 | 0 | 0 | 0 | 0.98 | 0.79 | 1.21 |
| SNCG | OID2097 | Neurolog | SNCG | O76070 | 10 | 86958599 | 86963258 | 46 | 0 | 0 | 0 | 0 | 0.99 | 0.88 | 1.11 |
| PPP1R14 | OID3099 | Neurolog | PPP1R14 | Q9NXH3 | 15 | 40815451 | 40828709 | 1 | 0 | 0 | 0 | 0 | 0.84 | 0.16 | 4.33 |
| CCL4 | OID2069 | Inflamma | CCL4 | P13236 | 17 | 36103827 | 36105621 | 20 | 0 | 0 | 0 | 0 | 0.98 | 0.79 | 1.21 |
| TIMP3 | OID2068 | Inflamma | TIMP3 | P35625 | 22 | 32801705 | 32863041 | 21 | 0 | 0 | 0 | 0 | 1.02 | 0.87 | 1.19 |
| MNDA | OID2015 | Cardiom | MNDA | P41218 | 1 | 158831351 | 158849506 | 1 | 0 | 0 | 0 | 0 | 1.27 | 0.13 | 12.95 |
| OLFM4 | OID3066 | Inflamma | OLFM4 | Q6UX06 | 13 | 53028813 | 53052057 | 30 | 0 | 0 | 0 | 0 | 1.02 | 0.87 | 1.19 |
| TNFRSF1 | OID2026 | Cardiom | TNFRSF1 | O14798 | 8 | 23102921 | 23117445 | 9 | 0 | 0 | 0 | 0 | 0.96 | 0.68 | 1.36 |
| PLXNB2 | OID2031 | Cardiom | PLXNB2 | O15031 | 22 | 50274979 | 50307646 | 29 | 0 | 0 | 0 | 0 | 0.99 | 0.87 | 1.12 |
| TNFSF10 | OID2061 | Inflamma | TNFSF10 | P50591 | 3 | 172505508 | 172523475 | 2 | 0 | 0 | 0 | 0 | 1.05 | 0.65 | 1.70 |
| BOLA2_E | OID3026 | Cardiom | BOLA2B | Q9H3K6 | 16;16 | 30192932;2 | 30194306;2 | 4 | 0 | 0 | 1 | 0 | 1.09 | 0.46 | 2.61 |
| HSPA1A | OID2071 | Inflamma | HSPA1A | P0DMV8 | 6 | 31815543 | 31817946 | 4 | 0 | 0 | 1 | 0 | 0.92 | 0.40 | 2.09 |
| CTSZ | OID2036 | Cardiom | CTSZ | Q9UBR2 | 20 | 58985686 | 59008238 | 4 | 0 | 0 | 0 | 0 | 1.07 | 0.56 | 2.02 |
| ADAMTS | OID3047 | Inflamma | ADAMTS | O75173 | 1 | 161184302 | 161199054 | 1 | 0 | 0 | 0 | 0 | 0.92 | 0.41 | 2.07 |
| IGFBP2 | OID2032 | Cardiom | IGFBP2 | P18065 | 2 | 216632828 | 216664436 | 1 | 0 | 0 | 0 | 0 | 1.17 | 0.24 | 5.61 |
| DLL1 | OID2152 | Oncology | DLL1 | O00548 | 6 | 170282206 | 170306565 | 6 | 0 | 0 | 0 | 0 | 0.95 | 0.55 | 1.64 |
| HLA-E | OID2053 | Inflamma | HLA-E | P13747 | 6 | 30489509 | 30494194 | 25 | 0 | 0 | 0 | 0 | 1.01 | 0.88 | 1.16 |
| MELTF | OID3034 | Cardiom | MELTF | P08582 | 3 | 196980590 | 197029835 | 25 | 0 | 0 | 0 | 0 | 0.99 | 0.85 | 1.14 |
| PDIA5 | OID3120 | Oncology | PDIA5 | Q14554 | 3 | 123067025 | 123225227 | 3 | 0 | 0 | 0 | 0 | 0.96 | 0.66 | 1.40 |
| SEC31A | OID3026 | Cardiom | SEC31A | O94979 | 4 | 82818509 | 82901166 | 1 | 0 | 0 | 0 | 0 | 0.91 | 0.35 | 2.39 |
| IGHMBP2 | OID3012 | Cardiom | IGHMBP2 | P38935 | 11 | 68903863 | 68940602 | 1 | 0 | 0 | 0 | 0 | 1.13 | 0.34 | 3.78 |
| SUOX | OID3103 | Neurolog | SUOX | P51687 | 12 | 55997180 | 56006641 | 1 | 0 | 0 | 0 | 0 | 1.18 | 0.22 | 6.23 |
| NPHS1 | OID3054 | Inflamma | NPHS1 | O60500 | 19 | 35825372 | 35869287 | 22 | 0 | 0 | 0 | 0 | 0.98 | 0.83 | 1.17 |
| WFDC12 | OID2146 | Oncology | WFDC12 | Q8WWY | 20 | 45123425 | 45124465 | 23 | 0 | 0 | 0 | 0 | 1.02 | 0.85 | 1.22 |
| EPPK1 | OID3025 | Cardiom | EPPK1 | P58107 | 8 | 143857324 | 143878467 | 35 | 0 | 0 | 0 | 1 | 0.98 | 0.84 | 1.16 |
| LCP1 | OID3036 | Cardiom | LCP1 | P13796 | 13 | 46125920 | 46211871 | 18 | 0 | 0 | 0 | 0 | 1.02 | 0.85 | 1.22 |
| KHK | OID3024 | Cardiom | KHK | P50053 | 2 | 27086747 | 27100762 | 31 | 0 | 0 | 0 | 0 | 0.98 | 0.80 | 1.21 |
| PDGFRA | OID2013 | Cardiom | PDGFRA | P16234 | 4 | 54229280 | 54298245 | 9 | 0 | 1 | 0 | 0 | 0.95 | 0.57 | 1.59 |
| NPC2 | OID3041 | Cardiom | NPC2 | P61916 | 14 | 74476192 | 74494177 | 12 | 0 | 0 | 0 | 0 | 0.96 | 0.63 | 1.47 |
| GP2 | OID2009 | Cardiom | GP2 | P55259 | 16 | 20309574 | 20327808 | 6 | 0 | 0 | 0 | 0 | 1.04 | 0.71 | 1.51 |
| CELA3A | OID2029 | Cardiom | CELA3A | P09093 | 1 | 22001657 | 22012542 | 7 | 0 | 0 | 0 | 0 | 0.97 | 0.65 | 1.44 |
| DPT | OID2027 | Cardiom | DPT | Q07507 | 1 | 168695468 | 168729206 | 18 | 0 | 0 | 0 | 0 | 1.02 | 0.83 | 1.25 |
| FECAB1 | OID3146 | Oncology | FECAB1 | O75071 | 1 | 46674659 | 46719146 | 2 | 0 | 0 | 0 | 0 | 1.09 | 0.40 | 2.99 |
| EGLN1 | OID2057 | Inflamma | EGLN1 | Q9GZT9 | 1 | 231363751 | 231422287 | 3 | 0 | 0 | 0 | 0 | 0.95 | 0.51 | 1.78 |
| F11R | OID2115 | Neurolog | F11R | Q9Y624 | 1 | 160995211 | 161021343 | 1 | 0 | 0 | 0 | 0 | 1.14 | 0.22 | 5.94 |
| IGSF9 | OID3033 | Cardiom | IGSF9 | Q9P2J2 | 1 | 159927039 | 159945613 | 5 | 0 | 0 | 0 | 0 | 0.97 | 0.69 | 1.37 |
| PIGR | OID2110 | Neurolog | PIGR | P01833 | 1 | 206928522 | 206946466 | 2 | 0 | 0 | 0 | 0 | 1.10 | 0.37 | 3.27 |
| ARHGAP | OID2125 | Oncology | ARHGAP | P42331 | 2 | 68679601 | 68826833 | 2 | 0 | 0 | 0 | 0 | 0.94 | 0.39 | 2.24 |
| DTYMK | OID3030 | Cardiom | DTYMK | P23919 | 2 | 241675747 | 241686944 | 4 | 0 | 0 | 1 | 0 | 0.93 | 0.34 | 2.49 |
| MCEE | OID3103 | Neurolog | MCEE | Q96PE7 | 2 | 71109684 | 71130239 | 1 | 0 | 0 | 0 | 0 | 1.04 | 0.63 | 1.72 |
| MITD1 | OID2095 | Neurolog | MITD1 | Q8WV92 | 2 | 99161427 | 99181058 | 1 | 0 | 0 | 0 | 0 | 0.90 | 0.24 | 3.39 |
| NAGK | OID3027 | Cardiom | NAGK | Q9UJ70 | 2 | 71064344 | 71079808 | 5 | 0 | 0 | 0 | 0 | 0.97 | 0.66 | 1.43 |
| MANF | OID2070 | Inflamma | MANF | P55145 | 3 | 51385291 | 51389397 | 1 | 0 | 0 | 0 | 0 | 0.89 | 0.22 | 3.57 |
| MASP1 | OID2095 | Neurolog | MASP1 | P48740 | 3 | 187217282 | 187291980 | 9 | 0 | 0 | 0 | 0 | 1.03 | 0.72 | 1.47 |
| ANXA10 | OID2088 | Neurolog | ANXA10 | Q9UJ72 | 4 | 168092537 | 168187736 | 1 | 0 | 0 | 0 | 0 | 0.86 | 0.13 | 5.72 |
| CXCL8 | OID2015 | Cardiom | CXCL8 | P10145 | 4 | 73740541 | 73743716 | 6 | 0 | 0 | 1 | 0 | 1.05 | 0.59 | 1.90 |
| ACOT13 | OID3152 | Oncology | ACOT13 | Q9NPJ3 | 6 | 24667035 | 24705065 | 1 | 0 | 0 | 0 | 0 | 1.14 | 0.23 | 5.68 |
| CPA4 | OID3060 | Inflamma | CPA4 | Q9UI42 | 7 | 130293134 | 130324180 | 22 | 0 | 0 | 0 | 0 | 0.98 | 0.82 | 1.18 |
| IGFBP1 | OID2024 | Cardiom | IGFBP1 | P08833 | 7 | 45888360 | 45893660 | 1 | 0 | 0 | 0 | 0 | 0.86 | 0.11 | 6.87 |
| MAD1L1 | OID2090 | Neurolog | MAD1L1 | Q9Y6D9 | 7 | 1815793 | 2233243 | 2 | 0 | 0 | 0 | 0 | 1.07 | 0.48 | 2.38 |
| TNFRSF1 | OID2098 | Neurolog | TNFRSF1 | O14763 | 8 | 23020133 | 23069031 | 12 | 0 | 0 | 0 | 0 | 1.02 | 0.79 | 1.32 |
| EGFL7 | OID2125 | Oncology | EGFL7 | Q9UHF1 | 9 | 136658856 | 136672678 | 7 | 0 | 0 | 0 | 0 | 1.04 | 0.68 | 1.61 |
| PAEP | OID2092 | Neurolog | PAEP | P09466 | 9 | 135561756 | 135566955 | 28 | 0 | 0 | 0 | 0 | 0.99 | 0.85 | 1.15 |
| GFRA1 | OID2145 | Oncology | GFRA1 | P56159 | 10 | 116056925 | 116276803 | 10 | 0 | 0 | 0 | 0 | 1.04 | 0.66 | 1.62 |
| KAZALD1 | OID2133 | Oncology | KAZALD1 | Q96I82 | 10 | 101061989 | 101068131 | 26 | 0 | 0 | 0 | 0 | 1.01 | 0.86 | 1.20 |
| CD5 | OID2144 | Oncology | CD5 | P06127 | 11 | 61102489 | 61127852 | 6 | 0 | 0 | 0 | 0 | 1.04 | 0.64 | 1.70 |
| FSHB | OID3023 | Cardiom | FSHB | P01225 | 11 | 30231014 | 30235261 | 1 | 0 | 0 | 0 | 0 | 1.08 | 0.43 | 2.71 |
| BIN2 | OID2106 | Neurolog | BIN2 | Q9UBW5 | 12 | 51281038 | 51324668 | 1 | 0 | 0 | 0 | 0 | 1.07 | 0.45 | 2.50 |
| C1S | OID3073 | Inflamma | C1S | P09871 | 12 | 6988259 | 7071032 | 13 | 0 | 0 | 0 | 0 | 0.98 | 0.78 | 1.24 |
| IGFBP6 | OID2041 | Cardiom | IGFBP6 | P24592 | 12 | 53097436 | 53102345 | 3 | 0 | 0 | 0 | 0 | 1.09 | 0.41 | 2.86 |
| KLRD1 | OID2063 | Inflamma | KLRD1 | Q13241 | 12 | 10226058 | 10329608 | 18 | 0 | 0 | 0 | 0 | 0.98 | 0.79 | 1.22 |
| LRIG3 | OID3045 | Inflamma | LRIG3 | Q6UXM1 | 12 | 58872149 | 58920504 | 1 | 0 | 0 | 0 | 0 | 0.90 | 0.24 | 3.31 |
| NTF3 | OID2059 | Inflamma | NTF3 | P20783 | 12 | 5432108 | 5521536 | 6 | 0 | 0 | 0 | 0 | 1.03 | 0.67 | 1.58 |
| THSD1 | OID3047 | Inflamma | THSD1 | Q9NS62 | 13 | 52377167 | 52416373 | 4 | 0 | 0 | 1 | 0 | 0.95 | 0.46 | 1.94 |
| WASF3 | OID2088 | Neurolog | WASF3 | Q9UPY6 | 13 | 26557683 | 26688948 | 2 | 0 | 0 | 0 | 0 | 0.94 | 0.50 | 1.79 |
| RNASE1 | OID3067 | Inflamma | RNASE1 | P07998 | 14 | 20801228 | 20802855 | 4 | 0 | 0 | 0 | 0 | 0.97 | 0.69 | 1.38 |
| FES | OID2120 | Oncology | FES | P07332 | 15 | 90883695 | 90895776 | 1 | 0 | 0 | 0 | 0 | 0.94 | 0.46 | 1.92 |
| HBQ1 | OID2143 | Oncology | HBQ1 | P09105 | 16 | 180459 | 181179 | 11 | 0 | 0 | 0 | 0 | 1.01 | 0.85 | 1.21 |
| LCAT | OID3067 | Inflamma | LCAT | P04180 | 16 | 67939750 | 67944131 | 7 | 0 | 0 | 0 | 0 | 1.05 | 0.58 | 1.92 |
| ACE | OID3069 | Inflamma | ACE | P12821 | 17 | 63477061 | 63498380 | 30 | 0 | 0 | 0 | 0 | 1.02 | 0.83 | 1.25 |
| ASGR1 | OID2099 | Neurolog | ASGR1 | P07306 | 17 | 7173431 | 7179564 | 3 | 0 | 0 | 0 | 0 | 0.96 | 0.56 | 1.65 |
| BLMH | OID2033 | Cardiom | BLMH | Q13867 | 17 | 30248203 | 30292056 | 49 | 0 | 0 | 0 | 0 | 1.01 | 0.88 | 1.17 |
| CCL11 | OID2066 | Inflamma | CCL11 | P51671 | 17 | 34285742 | 34288334 | 1 | 0 | 0 | 0 | 0 | 1.08 | 0.41 | 2.82 |
| CD7 | OID3054 | Inflamma | CD7 | P09564 | 17 | 82314868 | 82317608 | 8 | 0 | 0 | 1 | 0 | 1.06 | 0.55 | 2.03 |
| IGFBP4 | OID2115 | Neurolog | IGFBP4 | P22692 | 17 | 40443450 | 40457725 | 1 | 0 | 0 | 0 | 0 | 0.82 | 0.08 | 8.39 |
| NT5C | OID3029 | Cardiom | NT5C | Q8TCD5 | 17 | 75130225 | 75131757 | 12 | 0 | 0 | 0 | 0 | 0.99 | 0.81 | 1.19 |
| SCPEP1 | OID3027 | Cardiom | SCPEP1 | Q9HB40 | 17 | 56978129 | 57006768 | 7 | 0 | 0 | 0 | 0 | 0.98 | 0.74 | 1.29 |
| CD177 | OID2111 | Neurolog | CD177 | Q8N6Q3 | 19 | 43353686 | 43363172 | 40 | 0 | 0 | 0 | 0 | 1.01 | 0.90 | 1.13 |
| CEACAM | OID3036 | Cardiom | CEACAM | P40199 | 19 | 41750977 | 41772211 | 1 | 0 | 0 | 0 | 0 | 0.92 | 0.39 | 2.19 |
| CFD | OID3073 | Inflamma | CFD | P00746 | 19 | 859453 | 867884 | 9 | 0 | 0 | 0 | 0 | 0.96 | 0.65 | 1.44 |
| ICAM1 | OID2041 | Cardiom | ICAM1 | P05362 | 19 | 10271093 | 10286615 | 23 | 0 | 0 | 0 | 0 | 1.02 | 0.85 | 1.22 |
| KIR2DL3 | OID2119 | Oncology | KIR2DL3 | P43628 | 19 | 54738513 | 54753052 | 15 | 0 | 0 | 0 | 0 | 0.98 | 0.80 | 1.21 |
| KIR2DS4 | OID3143 | Oncology | KIR2DS4 | P43632 | 19 | 54832676 | 54848569 | 23 | 0 | 1 | 0 | 0 | 1.01 | 0.90 | 1.13 |
| MYO9B | OID2045 | Inflamma | MYO9B | Q13459 | 19 | 17075777 | 17214537 | 1 | 0 | 0 | 0 | 0 | 0.87 | 0.13 | 5.64 |
| NOTCH3 | OID2025 | Cardiom | NOTCH3 | Q9UM47 | 19 | 15159038 | 15200995 | 4 | 0 | 0 | 1 | 0 | 0.94 | 0.45 | 1.97 |
| PRKD2 | OID3014 | Cardiom | PRKD2 | Q9BZL6 | 19 | 46674275 | 46717127 | 1 | 0 | 0 | 0 | 0 | 1.18 | 0.14 | 10.09 |
| YJU2 | OID3140 | Oncology | YJU2 | Q9BW85 | 19 | 4247080 | 4269088 | 1 | 0 | 0 | 0 | 0 | 0.85 | 0.13 | 5.61 |
| RASSF2 | OID2119 | Oncology | RASSF2 | P50749 | 20 | 4780023 | 4823608 | 3 | 0 | 0 | 1 | 0 | 0.94 | 0.45 | 1.98 |
| CRELD2 | OID2075 | Inflamma | CRELD2 | Q6UXH1 | 22 | 49918167 | 49927540 | 12 | 0 | 0 | 0 | 0 | 0.98 | 0.78 | 1.23 |
| TYMP | OID2023 | Cardiom | TYMP | P19971 | 22 | 50525752 | 50530032 | 3 | 0 | 0 | 0 | 0 | 1.04 | 0.65 | 1.65 |
| DEFB4A | OID2137 | Oncology | DEFB4A | O15263 | 8;8 | 7894677;74 | 7896716;74 | 6 | 0 | 0 | 1 | 0 | 0.94 | 0.45 | 1.93 |
| APP | OID2107 | Neurolog | APP | P05067 | 21 | 25880550 | 26171128 | 1 | 0 | 0 | 0 | 0 | 0.90 | 0.23 | 3.52 |
| AGT | OID3070 | Inflamma | AGT | P01019 | 1 | 230690776 | 230745576 | 18 | 0 | 0 | 0 | 0 | 1.02 | 0.81 | 1.28 |
| C1QA | OID2065 | Inflamma | C1QA | P02745 | 1 | 22636628 | 22639678 | 11 | 0 | 0 | 0 | 0 | 1.02 | 0.80 | 1.29 |
| C8B | OID3071 | Inflamma | C8B | P07358 | 1 | 56929207 | 56974383 | 16 | 0 | 0 | 0 | 0 | 1.01 | 0.85 | 1.20 |
| CA14 | OID2140 | Oncology | CA14 | Q9ULX7 | 1 | 150257251 | 150265078 | 15 | 0 | 0 | 0 | 0 | 0.99 | 0.75 | 1.30 |
| CNTN2 | OID2142 | Oncology | CNTN2 | Q02246 | 1 | 205042937 | 205078289 | 31 | 0 | 0 | 0 | 0 | 1.01 | 0.88 | 1.14 |
| CR1 | OID3069 | Inflamma | CR1 | P17927 | 1 | 207496147 | 207641765 | 18 | 0 | 0 | 0 | 0 | 0.99 | 0.83 | 1.18 |
| DDAH1 | OID2129 | Oncology | DDAH1 | O94760 | 1 | 85318481 | 85578363 | 8 | 0 | 0 | 0 | 0 | 0.98 | 0.59 | 1.61 |
| FCGR3B | OID2038 | Cardiom | FCGR3B | O75015 | 1 | 161623196 | 161631963 | 32 | 0 | 0 | 0 | 0 | 0.99 | 0.86 | 1.14 |
| GPA33 | OID2137 | Oncology | GPA33 | Q99795 | 1 | 167052836 | 167166479 | 3 | 0 | 0 | 0 | 0 | 1.02 | 0.63 | 1.66 |
| GUCA2A | OID2110 | Neurolog | GUCA2A | Q02747 | 1 | 42162690 | 42164745 | 13 | 0 | 0 | 0 | 0 | 1.02 | 0.71 | 1.46 |
| LMOD1 | OID3021 | Cardiom | LMOD1 | P29536 | 1 | 201896456 | 201946588 | 7 | 0 | 0 | 0 | 0 | 0.99 | 0.65 | 1.49 |
| MYOM3 | OID3063 | Inflamma | MYOM3 | Q5VTT5 | 1 | 24056035 | 24112135 | 1 | 0 | 0 | 0 | 0 | 0.94 | 0.27 | 3.31 |
| NCF2 | OID2056 | Inflamma | NCF2 | P19878 | 1 | 183555562 | 183590876 | 1 | 0 | 0 | 0 | 0 | 0.93 | 0.10 | 8.38 |
| NMNAT1 | OID2091 | Neurolog | NMNAT1 | Q9HAN9 | 1 | 9943428 | 9985501 | 3 | 0 | 0 | 0 | 0 | 1.03 | 0.56 | 1.92 |
| PBXIP1 | OID3143 | Oncology | PBXIP1 | Q96AQ6 | 1 | 154944076 | 154956123 | 1 | 0 | 0 | 0 | 0 | 1.12 | 0.14 | 8.77 |
| RBP7 | OID3152 | Oncology | RBP7 | Q96R05 | 1 | 9997206 | 10016021 | 7 | 0 | 0 | 0 | 0 | 0.98 | 0.70 | 1.36 |
| REG4 | OID2078 | Inflamma | REG4 | Q9BYZ8 | 1 | 119794017 | 119811580 | 5 | 0 | 0 | 0 | 0 | 0.99 | 0.67 | 1.46 |
| S100A11 | OID2013 | Cardiom | S100A11 | P31949 | 1 | 152032506 | 152047907 | 2 | 0 | 0 | 0 | 0 | 0.97 | 0.48 | 1.97 |
| S100A13 | OID3052 | Inflamma | S100A13 | Q99584 | 1 | 153618787 | 153631360 | 8 | 0 | 0 | 0 | 0 | 1.01 | 0.73 | 1.39 |
| SELE | OID2029 | Cardiom | SELE | P16581 | 1 | 169722640 | 169764705 | 2 | 0 | 0 | 0 | 0 | 1.02 | 0.40 | 2.62 |
| SUSD4 | OID3139 | Oncology | SUSD4 | Q5VX71 | 1 | 223220819 | 223364233 | 11 | 0 | 0 | 0 | 0 | 0.99 | 0.72 | 1.35 |
| TGFBR3 | OID2025 | Cardiom | TGFBR3 | Q03167 | 1 | 91680343 | 91906335 | 7 | 0 | 0 | 0 | 0 | 1.02 | 0.61 | 1.72 |
| TNFRSF1 | OID2078 | Inflamma | TNFRSF1 | Q92956 | 1 | 2555639 | 2565382 | 1 | 0 | 0 | 0 | 0 | 1.02 | 0.45 | 2.32 |
| BMP10 | OID3057 | Inflamma | BMP10 | O95393 | 2 | 68860909 | 68871397 | 6 | 0 | 0 | 1 | 0 | 1.05 | 0.53 | 2.07 |
| EDAR | OID2052 | Inflamma | EDAR | Q9UNE0 | 2 | 108894471 | 108989372 | 19 | 0 | 0 | 0 | 0 | 1.01 | 0.80 | 1.29 |
| GORASP | OID3143 | Oncology | GORASP | Q9H8Y8 | 2 | 170928464 | 170967130 | 1 | 0 | 0 | 0 | 0 | 1.07 | 0.18 | 6.52 |
| IL1RN | OID2070 | Inflamma | IL1RN | P18510 | 2 | 113107214 | 113134016 | 8 | 0 | 0 | 0 | 0 | 0.99 | 0.71 | 1.38 |
| INPP1 | OID2134 | Oncology | INPP1 | P49441 | 2 | 190343570 | 190371665 | 1 | 0 | 0 | 0 | 0 | 1.09 | 0.30 | 4.05 |
| PLB1 | OID3111 | Neurolog | PLB1 | Q6P1J6 | 2 | 28457145 | 28644142 | 32 | 0 | 0 | 0 | 0 | 0.99 | 0.88 | 1.12 |
| TFPI | OID2038 | Cardiom | TFPI | P10646 | 2 | 187464230 | 187565760 | 24 | 0 | 0 | 0 | 0 | 0.98 | 0.79 | 1.23 |
| TMSB10 | OID2111 | Neurolog | TMSB10 | P63313 | 2 | 84905656 | 84906671 | 1 | 0 | 0 | 0 | 0 | 1.06 | 0.12 | 9.43 |
| CLEC3B | OID3075 | Inflamma | CLEC3B | P05452 | 3 | 45001548 | 45036071 | 8 | 0 | 0 | 0 | 0 | 0.99 | 0.70 | 1.38 |
| CPOX | OID3049 | Inflamma | CPOX | P36551 | 3 | 98579446 | 98593648 | 3 | 0 | 0 | 0 | 0 | 1.03 | 0.55 | 1.95 |
| IL17RB | OID2058 | Inflamma | IL17RB | Q9NRM6 | 3 | 53846568 | 53865794 | 32 | 0 | 0 | 0 | 0 | 1.00 | 0.86 | 1.17 |
| LAMP3 | OID2063 | Inflamma | LAMP3 | Q9UQV4 | 3 | 183122215 | 183163839 | 4 | 0 | 0 | 0 | 0 | 0.97 | 0.65 | 1.45 |
| ROBO1 | OID2073 | Inflamma | ROBO1 | Q9Y6N7 | 3 | 78597239 | 79767998 | 12 | 0 | 0 | 0 | 0 | 1.01 | 0.71 | 1.43 |
| SUSD5 | OID3065 | Inflamma | SUSD5 | O60279 | 3 | 33150043 | 33218810 | 26 | 0 | 0 | 0 | 0 | 1.00 | 0.87 | 1.16 |
| AREG | OID2135 | Oncology | AREG | P15514 | 4 | 74445136 | 74455005 | 5 | 0 | 0 | 0 | 0 | 0.99 | 0.60 | 1.63 |
| BST1 | OID2105 | Neurolog | BST1 | Q10588 | 4 | 15703065 | 15738313 | 41 | 0 | 0 | 0 | 0 | 1.00 | 0.90 | 1.12 |
| BTC | OID2119 | Oncology | BTC | P35070 | 4 | 74744759 | 74794523 | 20 | 0 | 0 | 0 | 0 | 0.99 | 0.83 | 1.18 |
| CTSO | OID2066 | Inflamma | CTSO | P43234 | 4 | 155921580 | 155953912 | 7 | 0 | 0 | 0 | 0 | 0.97 | 0.67 | 1.42 |
| CXCL11 | OID2104 | Neurolog | CXCL11 | O14625 | 4 | 76033682 | 76041415 | 6 | 0 | 0 | 0 | 0 | 1.01 | 0.67 | 1.53 |
| CXCL6 | OID2061 | Inflamma | CXCL6 | P80162 | 4 | 73836640 | 73849064 | 25 | 0 | 0 | 0 | 0 | 1.00 | 0.85 | 1.19 |
| DAPP1 | OID2052 | Inflamma | DAPP1 | Q9UN19 | 4 | 99816827 | 99870190 | 1 | 0 | 0 | 0 | 0 | 0.95 | 0.37 | 2.45 |
| EREG | OID2090 | Neurolog | EREG | O14944 | 4 | 74365145 | 74388749 | 1 | 0 | 0 | 0 | 0 | 1.04 | 0.36 | 3.04 |
| GC | OID3078 | Inflamma | GC | P02774 | 4 | 71741696 | 71804041 | 49 | 0 | 0 | 0 | 1 | 1.00 | 0.89 | 1.11 |
| KDR | OID2149 | Oncology | KDR | P35968 | 4 | 55078481 | 55125595 | 13 | 0 | 0 | 0 | 0 | 1.01 | 0.78 | 1.32 |
| KIT | OID2037 | Cardiom | KIT | P10721 | 4 | 54657267 | 54740783 | 2 | 0 | 0 | 0 | 0 | 1.09 | 0.27 | 4.31 |
| ODAM | OID2079 | Neurolog | ODAM | A1E959 | 4 | 70195725 | 70204576 | 9 | 0 | 0 | 0 | 0 | 1.02 | 0.79 | 1.30 |
| QDPR | OID2017 | Cardiom | QDPR | P09417 | 4 | 17460261 | 17512206 | 9 | 0 | 0 | 0 | 0 | 1.02 | 0.80 | 1.28 |
| CERT | OID2085 | Neurolog | CERT1 | Q9Y5P4 | 5 | 75356345 | 75512138 | 1 | 0 | 0 | 0 | 0 | 0.96 | 0.17 | 5.36 |
| FST | OID2077 | Inflamma | FST | P19883 | 5 | 53480626 | 53487134 | 4 | 0 | 0 | 0 | 0 | 1.01 | 0.59 | 1.73 |
| IL7R | OID2113 | Neurolog | IL7R | P16871 | 5 | 35852695 | 35879603 | 40 | 0 | 0 | 0 | 0 | 1.01 | 0.89 | 1.13 |
| MEGF10 | OID2074 | Inflamma | MEGF10 | Q96KG7 | 5 | 127290796 | 127465737 | 22 | 0 | 0 | 0 | 0 | 1.00 | 0.83 | 1.20 |
| MZB1 | OID2073 | Inflamma | MZB1 | Q8WU39 | 5 | 139387467 | 139390081 | 3 | 0 | 0 | 1 | 0 | 1.03 | 0.36 | 3.01 |
| TBCA | OID3061 | Inflamma | TBCA | O75347 | 5 | 77691166 | 77868780 | 1 | 0 | 0 | 0 | 0 | 0.97 | 0.47 | 2.01 |
| CD83 | OID2056 | Inflamma | CD83 | Q01151 | 6 | 14117256 | 14136918 | 5 | 0 | 0 | 0 | 0 | 0.98 | 0.61 | 1.57 |
| IGF2R | OID2114 | Neurolog | IGF2R | P11717 | 6 | 159969082 | 160113507 | 27 | 0 | 0 | 0 | 0 | 1.01 | 0.86 | 1.18 |
| IL17F | OID2045 | Inflamma | IL17F | Q96PD4 | 6 | 52236681 | 52244500 | 2 | 0 | 0 | 0 | 0 | 1.01 | 0.60 | 1.71 |
| LTA | OID2058 | Inflamma | LTA | P01374 | 6 | 31572054 | 31574324 | 27 | 0 | 0 | 0 | 0 | 1.00 | 0.87 | 1.15 |
| NT5E | OID2149 | Oncology | NT5E | P21589 | 6 | 85449584 | 85495791 | 30 | 0 | 0 | 0 | 0 | 1.00 | 0.88 | 1.13 |
| PI16 | OID3067 | Inflamma | PI16 | Q6UXB8 | 6 | 36948263 | 36964837 | 12 | 0 | 0 | 0 | 0 | 1.01 | 0.75 | 1.34 |
| SERPINF | OID2108 | Neurolog | SERPINF | P30740 | 6 | 2832332 | 2841959 | 3 | 0 | 0 | 1 | 0 | 1.04 | 0.47 | 2.28 |
| TPMT | OID2127 | Oncology | TPMT | P51580 | 6 | 18128311 | 18155077 | 15 | 0 | 0 | 0 | 0 | 1.01 | 0.77 | 1.32 |
| TREM2 | OID2073 | Inflamma | TREM2 | Q9NZC2 | 6 | 41158506 | 41163186 | 3 | 0 | 0 | 1 | 0 | 1.01 | 0.43 | 2.37 |
| CD36 | OID3048 | Inflamma | CD36 | P16671 | 7 | 80369575 | 80679277 | 6 | 0 | 0 | 0 | 0 | 1.01 | 0.74 | 1.39 |
| CLIP2 | OID2055 | Inflamma | CLIP2 | Q9UDT6 | 7 | 74289407 | 74405935 | 3 | 0 | 0 | 0 | 0 | 1.05 | 0.53 | 2.08 |
| CPA1 | OID2028 | Cardiom | CPA1 | P15085 | 7 | 130380339 | 130388114 | 3 | 0 | 0 | 0 | 0 | 1.02 | 0.61 | 1.69 |
| CPA2 | OID2106 | Neurolog | CPA2 | P48052 | 7 | 130266863 | 130289798 | 15 | 0 | 0 | 0 | 0 | 0.99 | 0.78 | 1.25 |
| EPO | OID2052 | Inflamma | EPO | P01588 | 7 | 100720468 | 100723700 | 4 | 0 | 0 | 1 | 0 | 1.03 | 0.36 | 2.95 |
| GIMAP8 | OID3119 | Oncology | GIMAP8 | Q8ND71 | 7 | 150450630 | 150479393 | 2 | 0 | 0 | 0 | 0 | 1.04 | 0.46 | 2.34 |
| HGF | OID2065 | Inflamma | HGF | P14210 | 7 | 81699010 | 81770438 | 2 | 0 | 0 | 0 | 0 | 1.02 | 0.40 | 2.59 |
| PON1 | OID3070 | Inflamma | PON1 | P27169 | 7 | 95297676 | 95324532 | 48 | 0 | 0 | 0 | 0 | 1.00 | 0.91 | 1.10 |
| SERPINE | OID2034 | Cardiom | SERPINE | P05121 | 7 | 101127104 | 101139247 | 4 | 0 | 0 | 0 | 0 | 0.98 | 0.45 | 2.11 |
| CA13 | OID2022 | Cardiom | CA13 | Q8N1Q1 | 8 | 85220587 | 85284073 | 12 | 0 | 0 | 0 | 0 | 1.00 | 0.80 | 1.26 |
| CCN4 | OID2144 | Oncology | CCN4 | O95388 | 8 | 133191039 | 133231690 | 19 | 0 | 0 | 0 | 0 | 1.00 | 0.84 | 1.18 |
| EPHX2 | OID2011 | Cardiom | EPHX2 | P34913 | 8 | 27490781 | 27545564 | 3 | 0 | 0 | 0 | 0 | 1.03 | 0.59 | 1.79 |
| FNTA | OID3138 | Oncology | FNTA | P49354 | 8 | 43034194 | 43085788 | 1 | 0 | 0 | 0 | 0 | 0.90 | 0.13 | 6.16 |
| GPIHBP1 | OID3146 | Oncology | GPIHBP1 | Q8IV16 | 8 | 143213218 | 143217170 | 13 | 0 | 0 | 0 | 0 | 1.01 | 0.76 | 1.34 |
| LY96 | OID2094 | Neurolog | LY96 | Q9Y6Y9 | 8 | 73991392 | 74029079 | 3 | 0 | 0 | 0 | 0 | 0.99 | 0.42 | 2.31 |
| OPLAH | OID3020 | Cardiom | OPLAH | O14841 | 8 | 144051266 | 144063965 | 3 | 0 | 0 | 0 | 0 | 0.97 | 0.60 | 1.57 |
| ST3GAL1 | OID2132 | Oncology | ST3GAL1 | Q11201 | 8 | 133454848 | 133571940 | 4 | 0 | 0 | 0 | 0 | 0.99 | 0.69 | 1.41 |
| ABCA2 | OID3014 | Cardiom | ABCA2 | Q9BZC7 | 9 | 137007234 | 137028915 | 1 | 0 | 0 | 0 | 0 | 1.05 | 0.23 | 4.87 |
| ASPN | OID3031 | Cardiom | ASPN | Q9BXN1 | 9 | 92456205 | 92482506 | 12 | 0 | 0 | 0 | 0 | 0.99 | 0.76 | 1.28 |
| COL15A1 | OID3039 | Cardiom | COL15A1 | P39059 | 9 | 98943179 | 99070792 | 9 | 0 | 0 | 0 | 0 | 1.01 | 0.72 | 1.40 |
| KYAT1 | OID2023 | Cardiom | KYAT1 | Q16773 | 9 | 128832942 | 128882494 | 1 | 0 | 0 | 0 | 0 | 0.97 | 0.45 | 2.07 |
| SEMA4D | OID2102 | Neurolog | SEMA4D | Q92854 | 9 | 89360787 | 89498130 | 1 | 0 | 0 | 0 | 0 | 0.93 | 0.29 | 2.93 |
| SPTLC1 | OID3108 | Neurolog | SPTLC1 | O15269 | 9 | 92000087 | 92115413 | 5 | 0 | 0 | 0 | 0 | 0.99 | 0.64 | 1.54 |
| SSNA1 | OID3145 | Oncology | SSNA1 | O43805 | 9 | 137188660 | 137190370 | 1 | 0 | 0 | 0 | 0 | 1.09 | 0.16 | 7.43 |
| TNC | OID2035 | Cardiom | TNC | P24821 | 9 | 115019575 | 115118257 | 11 | 0 | 0 | 0 | 0 | 0.99 | 0.73 | 1.34 |
| APBB1IP | OID2135 | Oncology | APBB1IF | Q7Z5R6 | 10 | 26438341 | 26567803 | 4 | 0 | 0 | 0 | 0 | 0.98 | 0.64 | 1.49 |
| FAS | OID2031 | Cardiom | FAS | P25445 | 10 | 88953813 | 89029605 | 10 | 0 | 0 | 0 | 0 | 1.02 | 0.72 | 1.45 |
| MBL2 | OID3075 | Inflamma | MBL2 | P11226 | 10 | 52765380 | 52772784 | 40 | 0 | 0 | 0 | 0 | 1.01 | 0.89 | 1.15 |
| MGMT | OID2058 | Inflamma | MGMT | P16455 | 10 | 129467190 | 129770983 | 19 | 0 | 0 | 0 | 0 | 1.01 | 0.83 | 1.22 |
| NRP1 | OID2039 | Cardiom | NRP1 | O14786 | 10 | 33177492 | 33336262 | 17 | 0 | 0 | 0 | 0 | 1.02 | 0.76 | 1.38 |
| PCBD1 | OID3061 | Inflamma | PCBD1 | P61457 | 10 | 70882280 | 70888565 | 4 | 0 | 0 | 0 | 0 | 1.02 | 0.52 | 1.99 |
| PSAP | OID3041 | Cardiom | PSAP | P07602 | 10 | 71816298 | 71851251 | 7 | 0 | 0 | 0 | 0 | 0.97 | 0.66 | 1.45 |
| VSIR | OID2013 | Cardiom | VSIR | Q9H7M9 | 10 | 71747556 | 71773520 | 12 | 0 | 0 | 0 | 0 | 0.99 | 0.76 | 1.28 |
| ACY3 | OID3008 | Cardiom | ACY3 | Q96HD9 | 11 | 67642555 | 67650730 | 22 | 0 | 0 | 0 | 0 | 0.98 | 0.72 | 1.33 |
| ADAMTS | OID2127 | Oncology | ADAMTS | Q8TE58 | 11 | 130448645 | 130476645 | 2 | 0 | 0 | 0 | 0 | 0.97 | 0.38 | 2.52 |
| CCS | OID2097 | Neurolog | CCS | O14618 | 11 | 66593153 | 66606019 | 35 | 0 | 0 | 0 | 0 | 0.99 | 0.80 | 1.23 |
| IL18 | OID2069 | Inflamma | IL18 | Q14116 | 11 | 112143253 | 112164096 | 9 | 0 | 0 | 0 | 0 | 0.98 | 0.70 | 1.37 |
| PHLDB1 | OID3146 | Oncology | PHLDB1 | Q86UU1 | 11 | 118606440 | 118658031 | 1 | 0 | 0 | 0 | 0 | 1.11 | 0.12 | 9.96 |
| PRCP | OID2033 | Cardiom | PRCP | P42785 | 11 | 82822936 | 82970584 | 1 | 0 | 0 | 0 | 0 | 0.96 | 0.24 | 3.87 |
| PTS | OID2084 | Neurolog | PTS | Q03393 | 11 | 112226367 | 112269955 | 1 | 0 | 0 | 0 | 0 | 0.98 | 0.61 | 1.58 |
| SCGB1A | OID2068 | Inflamma | SCGB1A | P11684 | 11 | 62405103 | 62423195 | 11 | 0 | 0 | 0 | 0 | 1.00 | 0.78 | 1.30 |
| ACRBP | OID3138 | Oncology | ACRBP | Q8NEB7 | 12 | 6638075 | 6647433 | 2 | 0 | 0 | 0 | 0 | 1.03 | 0.59 | 1.79 |
| ITGA5 | OID2113 | Neurolog | ITGA5 | P08648 | 12 | 54395261 | 54419266 | 1 | 0 | 0 | 0 | 0 | 0.96 | 0.13 | 6.90 |
| KRT5 | OID2086 | Neurolog | KRT5 | P13647 | 12 | 52514575 | 52520530 | 2 | 0 | 0 | 0 | 0 | 1.03 | 0.44 | 2.39 |
| LAG3 | OID2131 | Oncology | LAG3 | P18627 | 12 | 6772512 | 6778455 | 4 | 0 | 0 | 0 | 0 | 1.02 | 0.62 | 1.67 |
| LRTM2 | OID3102 | Neurolog | LRTM2 | Q8N967 | 12 | 1820267 | 1836753 | 2 | 0 | 0 | 0 | 0 | 0.97 | 0.47 | 2.00 |
| M6PR | OID3020 | Cardiom | M6PR | P20645 | 12 | 8940361 | 8949761 | 3 | 0 | 0 | 0 | 0 | 1.02 | 0.61 | 1.68 |
| NFE2 | OID3026 | Cardiom | NFE2 | Q16621 | 12 | 54292111 | 54301015 | 1 | 0 | 0 | 0 | 0 | 1.06 | 0.25 | 4.39 |
| PCBP2 | OID3100 | Neurolog | PCBP2 | Q15366 | 12 | 53452102 | 53481162 | 1 | 0 | 0 | 0 | 0 | 1.05 | 0.14 | 7.66 |
| PEBP1 | OID2108 | Neurolog | PEBP1 | P30086 | 12 | 118136124 | 118145584 | 1 | 0 | 0 | 0 | 0 | 1.06 | 0.33 | 3.37 |
| PLA2G1E | OID2025 | Cardiom | PLA2G1E | P04054 | 12 | 120322115 | 120327779 | 1 | 0 | 0 | 0 | 0 | 0.92 | 0.17 | 5.10 |
| TARBP2 | OID2087 | Neurolog | TARBP2 | Q15633 | 12 | 53500921 | 53506431 | 1 | 0 | 0 | 0 | 0 | 1.04 | 0.16 | 6.88 |
| TIGAR | OID2089 | Neurolog | TIGAR | Q9NQ88 | 12 | 4307763 | 4360028 | 1 | 0 | 0 | 0 | 0 | 1.12 | 0.21 | 5.89 |
| VSIG10 | OID3140 | Oncology | VSIG10 | Q8N0Z9 | 12 | 118063593 | 118136026 | 27 | 0 | 0 | 0 | 0 | 1.01 | 0.86 | 1.18 |
| CPB2 | OID3073 | Inflamma | CPB2 | Q96IY4 | 13 | 46053186 | 46105033 | 21 | 0 | 0 | 0 | 0 | 0.99 | 0.83 | 1.19 |
| LAMP1 | OID3041 | Cardiom | LAMP1 | P11279 | 13 | 113297239 | 113323672 | 2 | 0 | 0 | 0 | 0 | 1.03 | 0.32 | 3.33 |
| NHLRC3 | OID3061 | Inflamma | NHLRC3 | Q5JS37 | 13 | 39038306 | 39050109 | 8 | 0 | 0 | 1 | 0 | 0.98 | 0.54 | 1.79 |
| POSTN | OID3041 | Cardiom | POSTN | Q15063 | 13 | 37562583 | 37598844 | 7 | 0 | 0 | 0 | 0 | 1.03 | 0.66 | 1.60 |
| SLITRK1 | OID3051 | Inflamma | SLITRK1 | Q96PX8 | 13 | 83877205 | 83882474 | 1 | 0 | 0 | 0 | 0 | 1.03 | 0.46 | 2.31 |
| GZMH | OID2009 | Cardiom | GZMH | P20718 | 14 | 24606480 | 24609699 | 5 | 0 | 0 | 0 | 0 | 1.01 | 0.68 | 1.51 |
| HSPA2 | OID3142 | Oncology | HSPA2 | P54652 | 14 | 64535905 | 64546173 | 4 | 0 | 0 | 0 | 0 | 0.97 | 0.45 | 2.11 |

| | | | | | | | | | | | | | | | | | | | | | | | | | | | | | | | | | | | | | |
|---|---|---|---|---|---|---|---|---|---|---|---|---|---|---|---|---|---|---|---|---|---|---|---|---|---|---|---|---|---|---|---|---|---|---|---|---|---|
| LTBP2 | OID2025 | Cardiom | LTBP2 | Q14767 | 14 | 74498183 | 74612378 | 8 | 0 | 0 | 0 | 0 | 0.99 | 0.52 | 1.87 | 0.977 | 0.995 | 9.64 | 0.21 | 92.81 | 0.99 | 0.57 | 1.71 | 0.972 | 0.994 | 0.84 | 0.40 | 1.74 | 0.632 | 0.69 | 0.15 | 3.28 | 0.641 | 0.03 | -0.10 | 0.17 | 0.615 |
| TDP1 | OID3140 | Oncology | TDP1 | Q9NUW8 | 14 | 89954939 | 90044764 | 1 | 0 | 0 | 0 | 0 | 0.98 | 0.30 | 3.20 | 0.968 | 0.995 | - | - | 113.06 | 0.98 | 0.29 | 3.25 | 0.969 | 0.994 | - | - | - | - | - | - | - | - | - | - | - | - |
| WARS | OID2108 | Neurolog | WARS1 | P23381 | 14 | 100333790 | 100376805 | 2 | 0 | 0 | 0 | 0 | 0.98 | 0.66 | 1.45 | 0.917 | 0.995 | 0.15 | 0.70 | 496.66 | 0.98 | 0.66 | 1.45 | 0.917 | 0.994 | - | - | - | - | - | - | - | - | - | - | - | - |
| CAPN3 | OID3138 | Oncology | CAPN3 | P20807 | 15 | 42359498 | 42412949 | 1 | 0 | 0 | 0 | 0 | 1.11 | 0.12 | 10.59 | 0.927 | 0.995 | - | - | 30.62 | 1.11 | 0.10 | 11.92 | 0.930 | 0.994 | - | - | - | - | - | - | - | - | - | - | - | - |
| DNAJA4 | OID3113 | Neurolog | DNAJA4 | Q8WW22 | 15 | 78264086 | 78282196 | 4 | 0 | 0 | 0 | 0 | 1.03 | 0.49 | 2.17 | 0.935 | 0.995 | 4.49 | 0.21 | 115.84 | 1.03 | 0.56 | 1.91 | 0.920 | 0.994 | 0.84 | 0.42 | 1.66 | 0.608 | 0.39 | 0.12 | 1.24 | 0.111 | 0.12 | 0.00 | 0.24 | 0.054 |
| MEGF11 | OID3009 | Cardiom | MEGF11 | A6BM72 | 15 | 65895079 | 66253756 | 3 | 0 | 0 | 0 | 0 | 0.97 | 0.53 | 1.77 | 0.932 | 0.995 | 1.31 | 0.52 | 142.64 | 0.97 | 0.53 | 1.78 | 0.933 | 0.994 | 0.97 | 0.52 | 1.83 | 0.935 | 0.38 | 0.07 | 2.13 | 0.270 | 0.12 | -0.09 | 0.32 | 0.252 |
| MFGE8 | OID2113 | Neurolog | MFGE8 | Q08431 | 15 | 88898683 | 88913381 | 8 | 0 | 0 | 0 | 0 | 1.01 | 0.76 | 1.34 | 0.945 | 0.995 | 7.35 | 0.39 | 270.48 | 1.01 | 0.77 | 1.33 | 0.944 | 0.994 | 0.97 | 0.72 | 1.32 | 0.858 | 1.13 | 0.70 | 1.83 | 0.625 | -0.03 | -0.12 | 0.07 | 0.569 |
| SMAD3 | OID3141 | Oncology | SMAD3 | P84022 | 15 | 67063763 | 67195173 | 5 | 0 | 0 | 0 | 0 | 0.97 | 0.50 | 1.88 | 0.921 | 0.995 | 12.84 | 0.01 | 224.27 | 0.97 | 0.67 | 1.40 | 0.857 | 0.987 | 1.14 | 0.76 | 1.72 | 0.532 | 3.38 | 1.11 | 10.24 | 0.032 | -0.24 | -0.44 | -0.05 | 0.016 |
| SPESP1 | OID3029 | Cardiom | SPESP1 | Q6UW49 | 15 | 68818221 | 68946811 | 7 | 0 | 0 | 0 | 0 | 0.98 | 0.49 | 1.95 | 0.961 | 0.995 | 9.63 | 0.14 | 83.60 | 0.98 | 0.57 | 1.70 | 0.950 | 0.994 | 1.01 | 0.48 | 2.14 | 0.975 | 0.58 | 0.14 | 2.50 | 0.467 | 0.09 | -0.13 | 0.32 | 0.420 |
| ZFYVE1 | OID3145 | Oncology | ZFYVE1 | Q96K21 | 15 | 40807086 | 40815084 | 16 | 0 | 0 | 1 | 0 | 1.01 | 0.68 | 1.52 | 0.948 | 0.995 | 8.55 | 0.90 | 61.39 | 1.01 | 0.67 | 1.53 | 0.948 | 0.994 | 1.15 | 0.66 | 2.00 | 0.628 | 1.06 | 0.23 | 4.94 | 0.938 | -0.01 | -0.24 | 0.22 | 0.950 |
| CALB2 | OID2080 | Neurolog | CALB2 | P22676 | 16 | 71358713 | 71390436 | 1 | 0 | 0 | 0 | 0 | 0.91 | 0.20 | 4.11 | 0.904 | 0.995 | - | - | 67.90 | 0.91 | 0.19 | 4.26 | 0.906 | 0.994 | - | - | - | - | - | - | - | - | - | - | - | - |
| CES1 | OID2024 | Cardiom | CES1 | P23141 | 16 | 55802851 | 55833337 | 9 | 0 | 0 | 0 | 0 | 1.02 | 0.77 | 1.35 | 0.917 | 0.995 | 3.93 | 0.86 | 253.25 | 1.02 | 0.76 | 1.35 | 0.917 | 0.994 | 1.06 | 0.76 | 1.46 | 0.745 | 0.96 | 0.67 | 1.38 | 0.836 | 0.01 | -0.04 | 0.07 | 0.638 |
| GFER | OID2125 | Oncology | GFER | P55789 | 16 | 1984193 | 1987749 | 1 | 0 | 0 | 0 | 0 | 0.97 | 0.24 | 3.92 | 0.969 | 0.995 | - | - | 82.24 | 0.97 | 0.23 | 4.03 | 0.969 | 0.994 | - | - | - | - | - | - | - | - | - | - | - | - |
| ITGAM | OID2107 | Neurolog | ITGAM | P11215 | 16 | 31259967 | 31332892 | 4 | 0 | 0 | 0 | 0 | 1.03 | 0.59 | 1.77 | 0.928 | 0.995 | 0.42 | 0.94 | 136.28 | 1.03 | 0.59 | 1.78 | 0.928 | 0.994 | 1.00 | 0.56 | 1.80 | 0.988 | 0.78 | 0.27 | 2.27 | 0.646 | 0.03 | -0.08 | 0.14 | 0.557 |
| NAGPA | OID3064 | Inflamma | NAGPA | Q9UK23 | 16 | 5024844 | 5034141 | 9 | 0 | 0 | 0 | 0 | 0.98 | 0.72 | 1.34 | 0.912 | 0.995 | 11.59 | 0.17 | 262.08 | 0.98 | 0.76 | 1.27 | 0.895 | 0.993 | 1.00 | 0.75 | 1.33 | 0.998 | 1.17 | 0.72 | 1.90 | 0.536 | -0.04 | -0.14 | 0.05 | 0.365 |
| PLA2G15 | OID2147 | Oncology | PLA2G15 | Q8NCC3 | 16 | 68245304 | 68261058 | 15 | 0 | 0 | 0 | 0 | 1.01 | 0.79 | 1.30 | 0.922 | 0.995 | 11.48 | 0.65 | 180.32 | 1.01 | 0.79 | 1.30 | 0.922 | 0.994 | 0.97 | 0.69 | 1.36 | 0.838 | 0.63 | 0.33 | 1.18 | 0.145 | 0.10 | -0.02 | 0.21 | 0.103 |
| SLC9A3R | OID3144 | Oncology | SLC9A3R | Q15599 | 16 | 2025356 | 2039026 | 1 | 0 | 0 | 0 | 0 | 1.06 | 0.44 | 2.56 | 0.895 | 0.995 | - | - | 204.81 | 1.06 | 0.44 | 2.58 | 0.896 | 0.993 | - | - | - | - | - | - | - | - | - | - | - | - |
| TPSD1 | OID3098 | Neurolog | TPSD1 | Q9BZJ3 | 16 | 1256059 | 1259008 | 13 | 0 | 0 | 0 | 0 | 1.00 | 0.79 | 1.26 | 0.969 | 0.995 | 14.80 | 0.25 | 238.31 | 1.00 | 0.81 | 1.23 | 0.966 | 0.994 | 0.93 | 0.67 | 1.30 | 0.689 | 1.07 | 0.75 | 1.54 | 0.702 | -0.02 | -0.10 | 0.06 | 0.583 |
| BAP18 | OID3115 | Neurolog | C17orf49 | Q8IXM2 | 17 | 7014495 | 7017525 | 1 | 0 | 0 | 0 | 0 | 1.04 | 0.21 | 5.03 | 0.965 | 0.995 | - | - | 66.67 | 1.04 | 0.21 | 5.22 | 0.965 | 0.994 | - | - | - | - | - | - | - | - | - | - | - | - |
| C1QTNF1 | OID2036 | Cardiom | C1QTNF1 | Q9BXJ1 | 17 | 79022814 | 79049788 | 4 | 0 | 0 | 0 | 0 | 1.01 | 0.65 | 1.57 | 0.964 | 0.995 | 4.09 | 0.25 | 286.83 | 1.01 | 0.69 | 1.48 | 0.958 | 0.994 | 0.95 | 0.64 | 1.40 | 0.787 | 0.69 | 0.39 | 1.19 | 0.181 | 0.12 | 0.00 | 0.25 | 0.059 |
| CD300A | OID3057 | Inflamma | CD300A | Q9UGN4 | 17 | 74466399 | 74484794 | 5 | 0 | 0 | 0 | 0 | 0.98 | 0.68 | 1.43 | 0.927 | 0.995 | 6.79 | 0.15 | 387.62 | 0.98 | 0.74 | 1.31 | 0.905 | 0.994 | 1.04 | 0.76 | 1.42 | 0.815 | 0.85 | 0.47 | 1.54 | 0.586 | 0.05 | -0.10 | 0.19 | 0.507 |
| CD300C | OID2101 | Neurolog | CD300C | Q08708 | 17 | 74541073 | 74546115 | 7 | 0 | 0 | 0 | 0 | 1.01 | 0.82 | 1.24 | 0.939 | 0.995 | 2.20 | 0.90 | 551.02 | 1.01 | 0.82 | 1.24 | 0.939 | 0.994 | 0.98 | 0.77 | 1.23 | 0.830 | 0.94 | 0.68 | 1.29 | 0.699 | 0.02 | -0.06 | 0.10 | 0.561 |
| CD300LG | OID2113 | Neurolog | CD300LG | Q6UXG3 | 17 | 43847148 | 43863639 | 9 | 0 | 0 | 0 | 0 | 1.01 | 0.77 | 1.31 | 0.963 | 0.995 | 5.93 | 0.65 | 264.83 | 1.01 | 0.77 | 1.32 | 0.963 | 0.994 | 0.98 | 0.73 | 1.33 | 0.919 | 0.89 | 0.64 | 1.24 | 0.481 | 0.04 | -0.02 | 0.10 | 0.211 |
| DCXR | OID2148 | Oncology | DCXR | Q7Z4W1 | 17 | 82035136 | 82037709 | 1 | 0 | 0 | 0 | 0 | 1.04 | 0.32 | 3.33 | 0.947 | 0.995 | - | - | 129.18 | 1.04 | 0.32 | 3.37 | 0.948 | 0.994 | - | - | - | - | - | - | - | - | - | - | - | - |
| MINK1 | OID3115 | Neurolog | MINK1 | Q8N4C8 | 17 | 4833340 | 4898061 | 1 | 0 | 0 | 0 | 0 | 0.93 | 0.16 | 5.36 | 0.940 | 0.995 | - | - | 51.54 | 0.93 | 0.15 | 5.65 | 0.941 | 0.994 | - | - | - | - | - | - | - | - | - | - | - | - |
| RABEP1 | OID3064 | Inflamma | RABEP1 | Q15276 | 17 | 5282265 | 5386340 | 1 | 0 | 0 | 0 | 0 | 0.95 | 0.25 | 3.64 | 0.943 | 0.995 | - | - | 89.38 | 0.95 | 0.24 | 3.73 | 0.944 | 0.994 | - | - | - | - | - | - | - | - | - | - | - | - |
| TNFSF12 | OID2062 | Inflamma | TNFSF12 | O43508 | 17 | 7549058 | 7557890 | 12 | 0 | 0 | 0 | 0 | 1.01 | 0.79 | 1.29 | 0.945 | 0.995 | 7.85 | 0.73 | 222.88 | 1.01 | 0.79 | 1.29 | 0.945 | 0.994 | 0.91 | 0.69 | 1.22 | 0.538 | 1.00 | 0.65 | 1.54 | 0.986 | 0.00 | -0.08 | 0.08 | 0.978 |
| CDH2 | OID2017 | Cardiom | CDH2 | P19022 | 18 | 27932879 | 28177946 | 3 | 0 | 0 | 1 | 1 | 0.96 | 0.48 | 1.92 | 0.916 | 0.995 | 8.84 | 0.01 | 133.22 | 0.96 | 0.48 | 1.92 | 0.914 | 0.994 | 0.91 | 0.38 | 2.21 | 0.838 | 0.07 | 0.01 | 0.60 | 0.015 | 0.37 | 0.09 | 0.65 | 0.010 |
| AXL | OID2036 | Cardiom | AXL | P30530 | 19 | 41219223 | 41261766 | 5 | 0 | 1 | 0 | 0 | 1.03 | 0.45 | 2.34 | 0.942 | 0.995 | 15.32 | 0.00 | 177.49 | 1.03 | 0.68 | 1.57 | 0.883 | 0.990 | 1.00 | 0.62 | 1.60 | 0.986 | 8.13 | 0.01 | 4796.20 | 0.520 | -0.38 | -1.54 | 0.78 | 0.522 |
| CC2D1A | OID2095 | Neurolog | CC2D1A | Q6P1N0 | 19 | 13906201 | 13930879 | 2 | 0 | 0 | 0 | 0 | 0.99 | 0.37 | 2.60 | 0.977 | 0.995 | 0.65 | 0.42 | 82.58 | 0.99 | 0.37 | 2.64 | 0.977 | 0.994 | - | - | - | - | - | - | - | - | - | - | - | - |
| CD22 | OID2063 | Inflamma | CD22 | P20273 | 19 | 35319261 | 35347361 | 6 | 0 | 0 | 0 | 0 | 1.02 | 0.71 | 1.46 | 0.923 | 0.995 | 1.98 | 0.85 | 225.28 | 1.02 | 0.71 | 1.46 | 0.923 | 0.994 | 0.98 | 0.67 | 1.43 | 0.905 | 0.88 | 0.48 | 1.60 | 0.667 | 0.03 | -0.07 | 0.14 | 0.542 |
| FGF21 | OID2140 | Oncology | FGF21 | Q9NSA1 | 19 | 48755524 | 48758333 | 2 | 0 | 0 | 0 | 0 | 1.03 | 0.50 | 2.13 | 0.931 | 0.995 | 0.00 | 0.95 | 152.43 | 1.03 | 0.50 | 2.15 | 0.932 | 0.994 | - | - | - | - | - | - | - | - | - | - | - | - |
| ICAM5 | OID2014 | Cardiom | ICAM5 | Q9UMF0 | 19 | 10289952 | 10296778 | 34 | 0 | 0 | 0 | 0 | 1.00 | 0.89 | 1.11 | 0.945 | 0.995 | 24.69 | 0.85 | 377.93 | 1.00 | 0.89 | 1.11 | 0.945 | 0.994 | 0.96 | 0.82 | 1.12 | 0.584 | 0.98 | 0.84 | 1.15 | 0.829 | 0.01 | -0.04 | 0.05 | 0.815 |
| IFNL1 | OID2079 | Neurolog | IFNL1 | Q8IU54 | 19 | 39296407 | 39298673 | 1 | 0 | 0 | 0 | 0 | 0.97 | 0.16 | 5.94 | 0.974 | 0.995 | - | - | 45.07 | 0.97 | 0.15 | 6.33 | 0.975 | 0.994 | - | - | - | - | - | - | - | - | - | - | - | - |
| MCEMP1 | OID3050 | Inflamma | MCEMP1 | Q8IX19 | 19 | 7677088 | 7679829 | 7 | 0 | 0 | 0 | 0 | 0.98 | 0.68 | 1.41 | 0.895 | 0.995 | 2.39 | 0.88 | 163.33 | 0.98 | 0.67 | 1.41 | 0.896 | 0.993 | 1.00 | 0.63 | 1.60 | 0.989 | 1.08 | 0.23 | 5.06 | 0.923 | -0.02 | -0.30 | 0.26 | 0.895 |
| PALM | OID3151 | Oncology | PALM | O75781 | 19 | 708935 | 748329 | 5 | 0 | 0 | 0 | 0 | 0.98 | 0.63 | 1.52 | 0.920 | 0.995 | 0.49 | 0.97 | 167.18 | 0.98 | 0.63 | 1.53 | 0.920 | 0.994 | 1.03 | 0.62 | 1.72 | 0.915 | 1.13 | 0.54 | 2.38 | 0.743 | -0.02 | -0.11 | 0.07 | 0.630 |
| PRDX2 | OID3066 | Inflamma | PRDX2 | P32119 | 19 | 12796820 | 12801800 | 3 | 0 | 0 | 1 | 0 | 0.95 | 0.35 | 2.59 | 0.928 | 0.995 | 0.34 | 0.84 | 55.15 | 0.95 | 0.34 | 2.66 | 0.929 | 0.994 | 0.88 | 0.28 | 2.72 | 0.823 | 0.71 | 0.17 | 3.01 | 0.645 | 0.04 | -0.10 | 0.17 | 0.582 |
| SIGLEC7 | OID2019 | Cardiom | SIGLEC7 | Q9Y286 | 19 | 51142299 | 51153526 | 11 | 0 | 0 | 0 | 0 | 0.99 | 0.76 | 1.28 | 0.931 | 0.995 | 12.79 | 0.24 | 312.44 | 0.99 | 0.79 | 1.24 | 0.922 | 0.994 | 1.01 | 0.75 | 1.34 | 0.970 | 1.04 | 0.70 | 1.53 | 0.845 | -0.01 | -0.09 | 0.06 | 0.724 |
| TGFB1 | OID2062 | Inflamma | TGFB1 | P01137 | 19 | 41301587 | 41353922 | 4 | 0 | 0 | 0 | 0 | 1.01 | 0.71 | 1.45 | 0.939 | 0.995 | 1.76 | 0.62 | 306.57 | 1.01 | 0.71 | 1.46 | 0.939 | 0.994 | 1.05 | 0.71 | 1.56 | 0.801 | 1.23 | 0.62 | 2.44 | 0.545 | -0.04 | -0.15 | 0.07 | 0.506 |
| CST1 | OID3058 | Inflamma | CST1 | P01037 | 20 | 23747562 | 23751268 | 25 | 0 | 0 | 0 | 0 | 0.99 | 0.84 | 1.17 | 0.913 | 0.995 | 19.94 | 0.70 | 214.56 | 0.99 | 0.84 | 1.17 | 0.913 | 0.994 | 1.12 | 0.89 | 1.41 | 0.315 | 1.09 | 0.81 | 1.47 | 0.575 | -0.03 | -0.11 | 0.05 | 0.456 |
| MAVS | OID2120 | Oncology | MAVS | Q7Z434 | 20 | 3846799 | 3876123 | 1 | 0 | 0 | 0 | 0 | 0.93 | 0.33 | 2.63 | 0.892 | 0.995 | - | - | 147.08 | 0.93 | 0.33 | 2.66 | 0.893 | 0.993 | - | - | - | - | - | - | - | - | - | - | - | - |
| THBD | OID2023 | Cardiom | THBD | P07204 | 20 | 23045633 | 23049672 | 8 | 0 | 0 | 0 | 0 | 1.03 | 0.66 | 1.61 | 0.899 | 0.995 | 3.01 | 0.88 | 106.46 | 1.03 | 0.66 | 1.62 | 0.900 | 0.993 | 1.00 | 0.58 | 1.70 | 0.987 | 0.55 | 0.19 | 1.58 | 0.265 | 0.08 | -0.04 | 0.20 | 0.198 |
| VSTM2L | OID2006 | Cardiom | VSTM2L | Q96N03 | 20 | 37903111 | 37945350 | 2 | 0 | 0 | 0 | 0 | 1.03 | 0.55 | 1.93 | 0.926 | 0.995 | 0.34 | 0.56 | 190.50 | 1.03 | 0.55 | 1.93 | 0.926 | 0.994 | - | - | - | - | - | - | - | - | - | - | - | - |
| BACH1 | OID2050 | Inflamma | BACH1 | O14867 | 21 | 29194071 | 29630751 | 1 | 0 | 0 | 0 | 0 | 1.04 | 0.10 | 10.67 | 0.973 | 0.995 | - | - | 30.11 | 1.04 | 0.09 | 12.07 | 0.974 | 0.994 | - | - | - | - | - | - | - | - | - | - | - | - |
| CSTB | OID2029 | Cardiom | CSTB | P04080 | 21 | 43772511 | 43776330 | 35 | 0 | 0 | 0 | 0 | 1.00 | 0.85 | 1.18 | 0.972 | 0.995 | 48.63 | 0.05 | 233.39 | 1.00 | 0.87 | 1.15 | 0.966 | 0.994 | 0.95 | 0.78 | 1.15 | 0.604 | 1.00 | 0.76 | 1.31 | 0.980 | 0.00 | -0.06 | 0.06 | 0.954 |
| ARSA | OID2113 | Neurolog | ARSA | P15289 | 22 | 50622754 | 50628173 | 15 | 0 | 0 | 0 | 0 | 1.01 | 0.74 | 1.37 | 0.958 | 0.995 | 20.09 | 0.13 | 178.17 | 1.01 | 0.78 | 1.30 | 0.949 | 0.994 | 0.99 | 0.71 | 1.39 | 0.975 | 0.98 | 0.61 | 1.57 | 0.934 | 0.01 | -0.07 | 0.09 | 0.875 |
| GP1BB | OID3017 | Cardiom | GP1BB | P13224 | 22 | 19723539 | 19724771 | 1 | 0 | 0 | 0 | 0 | 0.93 | 0.10 | 8.63 | 0.950 | 0.995 | - | - | 31.59 | 0.93 | 0.09 | 9.66 | 0.952 | 0.994 | - | - | - | - | - | - | - | - | - | - | - | - |
| RTN4R | OID2142 | Oncology | RTN4R | Q9BZR6 | 22 | 20241415 | 20283246 | 13 | 0 | 0 | 0 | 0 | 0.99 | 0.77 | 1.27 | 0.937 | 0.995 | 15.12 | 0.23 | 263.58 | 0.99 | 0.79 | 1.23 | 0.929 | 0.994 | 0.99 | 0.73 | 1.33 | 0.938 | 0.78 | 0.50 | 1.22 | 0.273 | 0.05 | -0.03 | 0.13 | 0.212 |
| SCARF2 | OID2104 | Neurolog | SCARF2 | Q96GP6 | 22 | 20424584 | 20437826 | 8 | 0 | 0 | 0 | 0 | 0.99 | 0.72 | 1.36 | 0.929 | 0.995 | 1.48 | 0.98 | 253.26 | 0.99 | 0.71 | 1.36 | 0.930 | 0.994 | 0.96 | 0.68 | 1.37 | 0.841 | 0.79 | 0.27 | 2.31 | 0.664 | 0.05 | -0.19 | 0.29 | 0.669 |
| UPB1 | OID3035 | Cardiom | UPB1 | Q9UBR1 | 22 | 24494107 | 24528390 | 1 | 0 | 0 | 0 | 0 | 0.94 | 0.10 | 9.17 | 0.957 | 0.995 | - | - | 31.06 | 0.94 | 0.09 | 10.32 | 0.959 | 0.994 | - | - | - | - | - | - | - | - | - | - | - | - |
| IL12A_IL | OID2132 | Oncology | IL12A;IL1 | P29459;F | 3;5 | 159988835 | 159996019 | 22 | 0 | 0 | 0 | 0 | 0.99 | 0.79 | 1.24 | 0.938 | 0.995 | 24.34 | 0.28 | 174.42 | 0.99 | 0.80 | 1.22 | 0.933 | 0.994 | 1.19 | 0.89 | 1.61 | 0.247 | 1.06 | 0.71 | 1.57 | 0.781 | -0.01 | -0.09 | 0.06 | 0.695 |
| VMO1 | OID2138 | Oncology | VMO1 | Q7Z5L0 | 17 | 4785285 | 4786433 | 39 | 0 | 0 | 0 | 0 | 1.00 | 0.88 | 1.13 | 0.978 | 0.996 | 40.23 | 0.37 | 281.68 | 1.00 | 0.89 | 1.12 | 0.978 | 0.994 | 0.89 | 0.74 | 1.08 | 0.237 | 0.92 | 0.74 | 1.13 | 0.413 | 0.03 | -0.03 | 0.08 | 0.329 |
| MDGA1 | OID2095 | Neurolog | MDGA1 | Q8NFP4 | 6 | 37630679 | 37699306 | 51 | 0 | 0 | 0 | 0 | 1.00 | 0.90 | 1.10 | 0.979 | 0.996 | 39.54 | 0.86 | 301.12 | 1.00 | 0.90 | 1.11 | 0.979 | 0.995 | 0.98 | 0.84 | 1.13 | 0.748 | 1.09 | 0.92 | 1.29 | 0.311 | -0.03 | -0.08 | 0.02 | 0.195 |
| BCHE | OID3071 | Inflamma | BCHE | P06276 | 3 | 165772904 | 165837462 | 37 | 0 | 0 | 0 | 0 | 1.00 | 0.83 | 1.21 | 0.980 | 0.996 | 42.29 | 0.22 | 150.84 | 1.00 | 0.84 | 1.19 | 0.978 | 0.994 | 1.03 | 0.79 | 1.34 | 0.831 | 0.80 | 0.54 | 1.18 | 0.260 | 0.06 | -0.03 | 0.15 | 0.199 |
| CACYBP | OID3064 | Inflamma | CACYBP | Q9HB71 | 1 | 174999163 | 175012027 | 3 | 0 | 0 | 0 | 0 | 0.99 | 0.55 | 1.78 | 0.982 | 0.997 | 0.02 | 0.99 | 145.43 | 0.99 | 0.55 | 1.79 | 0.982 | 0.997 | 1.00 | 0.54 | 1.87 | 0.995 | 2.39 | 0.00 | 301908.69 | 0.885 | -0.15 | -2.15 | 1.85 | 0.884 |
| CDA | OID3034 | Cardiom | CDA | P32320 | 1 | 20589086 | 20618903 | 23 | 0 | 1 | 0 | 0 | 1.00 | 0.78 | 1.29 | 0.987 | 0.997 | 40.57 | 0.01 | 222.80 | 1.00 | 0.83 | 1.21 | 0.982 | 0.997 | 1.00 | 0.80 | 1.25 | 0.988 | 1.13 | 0.76 | 1.68 | 0.535 | -0.03 | -0.11 | 0.05 | 0.425 |
| CTRC | OID2075 | Inflamma | CTRC | Q99895 | 1 | 15438442 | 15449242 | 13 | 0 | 0 | 0 | 0 | 1.00 | 0.78 | 1.28 | 0.982 | 0.997 | 5.96 | 0.92 | 224.65 | 1.00 | 0.78 | 1.29 | 0.982 | 0.997 | 0.99 | 0.75 | 1.30 | 0.946 | 1.03 | 0.73 | 1.46 | 0.873 | -0.01 | -0.09 | 0.07 | 0.839 |
| GBP1 | OID3112 | Neurolog | GBP1 | P32455 | 1 | 89051882 | 89065360 | 11 | 0 | 0 | 1 | 0 | 1.00 | 0.58 | 1.70 | 0.988 | 0.997 | 7.80 | 0.65 | 53.07 | 1.00 | 0.58 | 1.72 | 0.988 | 0.997 | 1.15 | 0.55 | 2.41 | 0.709 | 2.16 | 0.60 | 7.70 | 0.237 | -0.08 | -0.19 | 0.04 | 0.191 |
| ADIPOQ | OID3074 | Inflamma | ADIPOQ | Q15848 | 3 | 186842704 | 186858463 | 8 | 0 | 0 | 0 | 0 | 1.00 | 0.64 | 1.58 | 0.986 | 0.997 | 7.43 | 0.39 | 138.03 | 1.00 | 0.64 | 1.56 | 0.986 | 0.997 | 0.87 | 0.53 | 1.43 | 0.580 | 0.72 | 0.39 | 1.35 | 0.307 | 0.06 | -0.02 | 0.13 | 0.147 |
| S100P | OID2010 | Cardiom | S100P | P25815 | 4 | 6693878 | 6697170 | 4 | 0 | 0 | 0 | 0 | 1.00 | 0.70 | 1.43 | 0.989 | 0.997 | 1.33 | 0.72 | 309.03 | 1.00 | 0.69 | 1.43 | 0.989 | 0.997 | 1.01 | 0.69 | 1.47 | 0.962 | 1.01 | 0.65 | 1.57 | 0.955 | 0.00 | -0.08 | 0.07 | 0.903 |
| TSPYL1 | OID3051 | Inflamma | TSPYL1 | Q9H0U9 | 6 | 116267760 | 116279930 | 2 | 0 | 0 | 0 | 0 | 1.01 | 0.47 | 2.18 | 0.983 | 0.997 | 1.13 | 0.29 | 130.92 | 1.01 | 0.46 | 2.20 | 0.983 | 0.997 | - | - | - | - | - | - | - | - | - | - | - | - |
| PDIA4 | OID3037 | Cardiom | PDIA4 | P13667 | 7 | 149003062 | 149028662 | 1 | 0 | 0 | 0 | 0 | 0.99 | 0.35 | 2.84 | 0.989 | 0.997 | - | - | 139.89 | 0.99 | 0.34 | 2.88 | 0.989 | 0.997 | - | - | - | - | - | - | - | - | - | - | - | - |
| TXN | OID3067 | Inflamma | TXN | P10599 | 9 | 110243810 | 110256507 | 2 | 0 | 0 | 0 | 0 | 0.99 | 0.29 | 3.42 | 0.986 | 0.997 | 1.03 | 0.31 | 51.67 | 0.99 | 0.28 | 3.52 | 0.986 | 0.997 | - | - | - | - | - | - | - | - | - | - | - | - |
| PBLD | OID2083 | Neurolog | PBLD | P30039 | 10 | 68282660 | 68333049 | 30 | 0 | 0 | 0 | 0 | 1.00 | 0.83 | 1.19 | 0.987 | 0.997 | 27.23 | 0.56 | 159.68 | 1.00 | 0.83 | 1.20 | 0.987 | 0.997 | 1.00 | 0.77 | 1.31 | 0.996 | 0.93 | 0.67 | 1.30 | 0.685 | 0.02 | -0.05 | 0.08 | 0.637 |
| PRDX3 | OID2048 | Inflamma | PRDX3 | P30048 | 10 | 119167720 | 119178812 | 1 | 0 | 0 | 0 | 0 | 0.99 | 0.21 | 4.72 | 0.985 | 0.997 | - | - | 62.50 | 0.99 | 0.20 | 4.91 | 0.985 | 0.997 | - | - | - | - | - | - | - | - | - | - | - | - |
| TCN1 | OID3068 | Inflamma | TCN1 | P20061 | 11 | 59852800 | 59866489 | 23 | 0 | 0 | 0 | 0 | 1.00 | 0.85 | 1.18 | 0.987 | 0.997 | 20.57 | 0.55 | 270.80 | 1.00 | 0.85 | 1.18 | 0.987 | 0.997 | 1.03 | 0.85 | 1.25 | 0.768 | 0.95 | 0.76 | 1.19 | 0.669 | 0.02 | -0.04 | 0.07 | 0.549 |
| ENTPD5 | OID2010 | Cardiom | ENTPD5 | O75356 | 14 | 73958010 | 74019399 | 16 | 0 | 0 | 0 | 0 | 1.00 | 0.76 | 1.31 | 0.983 | 0.997 | 25.45 | 0.04 | 224.34 | 1.00 | 0.81 | 1.23 | 0.978 | 0.994 | 0.96 | 0.74 | 1.24 | 0.768 | 0.93 | 0.56 | 1.55 | 0.789 | 0.02 | -0.10 | 0.14 | 0.760 |
| GOLM2 | OID2104 | Neurolog | GOLM2 | Q6P4E1 | 15 | 44288719 | 44415758 | 1 | 0 | 0 | 0 | 0 | 1.02 | 0.11 | 9.48 | 0.987 | 0.997 | - | - | 32.01 | 1.02 | 0.10 | 10.60 | 0.988 | 0.997 | - | - | - | - | - | - | - | - | - | - | - | - |
| PRSS27 | OID2021 | Cardiom | PRSS27 | Q9BQR3 | 16 | 2712419 | 2720551 | 6 | 0 | 0 | 0 | 0 | 1.00 | 0.68 | 1.47 | 0.989 | 0.997 | 2.84 | 0.72 | 218.35 | 1.00 | 0.68 | 1.47 | 0.989 | 0.997 | 1.05 | 0.68 | 1.61 | 0.825 | 1.29 | 0.72 | 2.32 | 0.395 | -0.05 | -0.15 | 0.04 | 0.253 |
| SIRPB1 | OID2073 | Inflamma | SIRPB1 | O00241 | 20 | 1561385 | 1620061 | 20 | 0 | 0 | 0 | 0 | 1.00 | 0.84 | 1.18 | 0.987 | 0.997 | 15.85 | 0.67 | 267.95 | 1.00 | 0.84 | 1.19 | 0.987 | 0.997 | 0.90 | 0.72 | 1.13 | 0.353 | 0.96 | 0.74 | 1.25 | 0.756 | 0.01 | -0.04 | 0.05 | 0.692 |
| ST13 | OID3062 | Inflamma | ST13 | P50502 | 22 | 40824535 | 40856639 | 1 | 0 | 0 | 0 | 0 | 1.02 | 0.18 | 5.84 | 0.985 | 0.997 | - | - | 53.17 | 1.02 | 0.17 | 6.14 | 0.986 | 0.997 | - | - | - | - | - | - | - | - | - | - | - | - |
| MUC2 | OID3037 | Cardiom | MUC2 | Q02817 | 11 | 1074875 | 1110511 | 5 | 0 | 0 | 0 | 0 | 1.00 | 0.67 | 1.49 | 0.989 | 0.997 | 1.31 | 0.86 | 286.24 | 1.00 | 0.67 | 1.49 | 0.989 | 0.997 | 1.02 | 0.67 | 1.55 | 0.924 | 1.14 | 0.60 | 2.15 | 0.688 | -0.02 | -0.11 | 0.06 | 0.597 |
| CD164 | OID2108 | Neurolog | CD164 | Q04900 | 6 | 109366514 | 109382467 | 5 | 0 | 0 | 0 | 0 | 1.00 | 0.58 | 1.73 | 0.990 | 0.997 | 1.49 | 0.83 | 104.33 | 1.00 | 0.58 | 1.74 | 0.990 | 0.997 | 0.95 | 0.52 | 1.74 | 0.867 | 0.95 | 0.28 | 3.15 | 0.927 | 0.01 | -0.12 | 0.13 | 0.914 |
| SCRN1 | OID2054 | Inflamma | SCRN1 | Q12765 | 7 | 29920104 | 29990289 | 2 | 0 | 0 | 0 | 0 | 1.00 | 0.52 | 1.93 | 0.991 | 0.998 | 1.03 | 0.31 | 194.64 | 1.00 | 0.52 | 1.94 | 0.991 | 0.997 | - | - | - | - | - | - | - | - | - | - | - | - |
| HMCN2 | OID3059 | Inflamma | HMCN2 | Q8NDA2 | 9 | 130265760 | 130434123 | 33 | 0 | 0 | 0 | 0 | 1.00 | 0.85 | 1.18 | 0.992 | 0.998 | 42.90 | 0.09 | 236.03 | 1.00 | 0.87 | 1.15 | 0.991 | 0.997 | 0.95 | 0.77 | 1.17 | 0.640 | 1.10 | 0.83 | 1.45 | 0.508 | -0.02 | -0.08 | 0.03 | 0.408 |
| EXTL1 | OID3011 | Cardiom | EXTL1 | Q92935 | 1 | 26019884 | 26036464 | 15 | 0 | 0 | 0 | 0 | 1.00 | 0.79 | 1.27 | 0.993 | 0.998 | 11.87 | 0.62 | 173.34 | 1.00 | 0.79 | 1.27 | 0.993 | 0.998 | 1.09 | 0.81 | 1.47 | 0.554 | 1.18 | 0.70 | 1.98 | 0.536 | -0.03 | -0.13 | 0.06 | 0.483 |
| PTPRF | OID2031 | Cardiom | PTPRF | P10586 | 1 | 43525187 | 43623666 | 2 | 0 | 0 | 0 | 0 | 1.00 | 0.32 | 3.09 | 0.994 | 0.999 | 1.21 | 0.27 | 69.76 | 1.00 | 0.31 | 3.15 | 0.994 | 0.999 | - | - | - | - | - | - | - | - | - | - | - | - |
| RHOC | OID2095 | Neurolog | RHOC | P08134 | 1 | 112701127 | 112707434 | 1 | 0 | 0 | 0 | 0 | 1.00 | 0.31 | 3.19 | 0.995 | 0.999 | - | - | 113.29 | 1.00 | 0.31 | 3.24 | 0.995 | 0.999 | - | - | - | - | - | - | - | - | - | - | - | - |
| FABP2 | OID2020 | Cardiom | FABP2 | P12104 | 4 | 119317250 | 119322138 | 5 | 0 | 0 | 0 | 0 | 1.00 | 0.67 | 1.49 | 0.994 | 0.999 | 1.44 | 0.84 | 202.97 | 1.00 | 0.67 | 1.49 | 0.994 | 0.999 | 1.02 | 0.66 | 1.56 | 0.938 | 1.23 | 0.53 | 2.83 | 0.629 | -0.03 | -0.14 | 0.08 | 0.586 |
| MAMDC2 | OID3030 | Cardiom | MAMDC2 | Q7Z304 | 9 | 70043848 | 70226972 | 7 | 0 | 0 | 0 | 0 | 1.00 | 0.66 | 1.50 | 0.995 | 0.999 | 4.79 | 0.57 | 144.97 | 1.00 | 0.66 | 1.50 | 0.995 | 0.999 | 1.11 | 0.71 | 1.73 | 0.640 | 1.68 | 0.38 | 7.34 | 0.494 | -0.10 | -0.37 | 0.17 | 0.475 |
| ATP5IF1 | OID2076 | Inflamma | ATP5IF1 | Q9UII2 | 1 | 28236109 | 28246906 | 3 | 0 | 0 | 0 | 0 | 1.00 | 0.48 | 2.10 | 0.997 | 0.999 | 0.24 | 0.89 | 95.97 | 1.00 | 0.47 | 2.12 | 0.997 | 0.999 | 0.97 | 0.44 | 2.11 | 0.936 | 0.65 | 0.08 | 5.67 | 0.700 | 0.04 | -0.14 | 0.21 | 0.681 |
| PHYKPL | OID3063 | Inflamma | PHYKPL | Q8IUZ5 | 5 | 178208471 | 178232802 | 8 | 0 | 0 | 0 | 0 | 1.00 | 0.60 | 1.66 | 0.997 | 0.999 | 8.95 | 0.26 | 94.51 | 1.00 | 0.63 | 1.57 | 0.997 | 0.999 | 0.66 | 0.37 | 1.20 | 0.173 | 0.32 | 0.12 | 0.87 | 0.026 | 0.16 | 0.03 | 0.29 | 0.013 |
| EGFR | OID2031 | Cardiom | EGFR | P00533 | 7 | 55019017 | 55211628 | 8 | 0 | 0 | 0 | 0 | 1.00 | 0.65 | 1.54 | 0.997 | 0.999 | 7.87 | 0.34 | 128.48 | 1.00 | 0.66 | 1.51 | 0.997 | 0.999 | 0.84 | 0.50 | 1.39 | 0.489 | 0.53 | 0.22 | 1.29 | 0.164 | 0.09 | -0.02 | 0.19 | 0.117 |
| ADAMTS | OID3035 | Cardiom | ADAMTS | Q6ZMM2 | 19 | 1505022 | 1513604 | 10 | 0 | 0 | 0 | 0 | 1.00 | 0.78 | 1.28 | 0.996 | 0.999 | 6.21 | 0.72 | 262.05 | 1.00 | 0.78 | 1.28 | 0.996 | 0.999 | 0.98 | 0.74 | 1.29 | 0.867 | 1.03 | 0.67 | 1.58 | 0.903 | -0.01 | -0.09 | 0.08 | 0.879 |
| HAVCR2 | OID2113 | Neurolog | HAVCR2 | Q8TDQ0 | 5 | 157085832 | 157142869 | 5 | 0 | 0 | 1 | 0 | 1.00 | 0.55 | 1.83 | 0.999 | 0.999 | 1.04 | 0.90 | 94.17 | 1.00 | 0.54 | 1.85 | 0.999 | 0.999 | 0.95 | 0.47 | 1.93 | 0.890 | 1.30 | 0.45 | 3.80 | 0.630 | -0.05 | -0.20 | 0.11 | 0.558 |
| OSCAR | OID2077 | Inflamma | OSCAR | Q8IYS5 | 19 | 54094668 | 54102692 | 22 | 0 | 0 | 0 | 0 | 1.00 | 0.85 | 1.18 | 0.999 | 0.999 | 30.09 | 0.09 | 377.52 | 1.00 | 0.87 | 1.15 | 0.998 | 0.999 | 0.90 | 0.75 | 1.08 | 0.259 | 0.88 | 0.67 | 1.16 | 0.362 | 0.04 | -0.03 | 0.10 | 0.261 |
| PADI2 | OID2046 | Inflamma | PADI2 | Q9Y2J8 | 1 | 17066761 | 17119451 | 1 | 0 | 0 | 0 | 0 | 1.00 | 0.45 | 2.23 | 1.000 | 1.000 | - | - | 238.35 | 1.00 | 0.45 | 2.25 | 1.000 | 1.000 | - | - | - | - | - | - | - | - | - | - | - | - |
| ACTN2 | OID3019 | Cardiom | ACTN2 | P35609 | 1 | 236664141 | 236764631 | 0 | - | - | - | - | - | - | - | - | - | - | - | - | - | - | - | - | - | - | - | - | - | - | - | - | - | - | - | - | - |
| AIDA | OID3095 | Neurolog | AIDA | Q96BJ3 | 1 | 222668013 | 222713210 | 0 | - | - | - | - | - | - | - | - | - | - | - | - | - | - | - | - | - | - | - | - | - | - | - | - | - | - | - | - | - |
| AK2 | OID3114 | Neurolog | AK2 | P54819 | 1 | 33007986 | 33080996 | 0 | - | - | - | - | - | - | - | - | - | - | - | - | - | - | - | - | - | - | - | - | - | - | - | - | - | - | - | - | - |
| ARHGAP | OID3133 | Oncology | ARHGAP | Q7Z6I6 | 1 | 161046946 | 161069970 | 0 | - | - | - | - | - | - | - | - | - | - | - | - | - | - | - | - | - | - | - | - | - | - | - | - | - | - | - | - | - |
| ARID4B | OID2080 | Neurolog | ARID4B | Q4LE39 | 1 | 235131634 | 235328219 | 0 | - | - | - | - | - | - | - | - | - | - | - | - | - | - | - | - | - | - | - | - | - | - | - | - | - | - | - | - | - |
| ARNT | OID2043 | Inflamma | ARNT | P27540 | 1 | 150809713 | 150876708 | 0 | - | - | - | - | - | - | - | - | - | - | - | - | - | - | - | - | - | - | - | - | - | - | - | - | - | - | - | - | - |
| ARTN | OID2044 | Inflamma | ARTN | Q5T4W7 | 1 | 43933320 | 43937240 | 0 | - | - | - | - | - | - | - | - | - | - | - | - | - | - | - | - | - | - | - | - | - | - | - | - | - | - | - | - | - |
| ATP1B1 | OID3012 | Cardiom | ATP1B1 | P05026 | 1 | 169105697 | 169310992 | 0 | - | - | - | - | - | - | - | - | - | - | - | - | - | - | - | - | - | - | - | - | - | - | - | - | - | - | - | - | - |
| ATP2B4 | OID3006 | Cardiom | ATP2B4 | P23634 | 1 | 203626832 | 203744081 | 0 | - | - | - | - | - | - | - | - | - | - | - | - | - | - | - | - | - | - | - | - | - | - | - | - | - | - | - | - | - |
| BRDT | OID3017 | Cardiom | BRDT | Q58F21 | 1 | 91949343 | 92014426 | 0 | - | - | - | - | - | - | - | - | - | - | - | - | - | - | - | - | - | - | - | - | - | - | - | - | - | - | - | - | - |
| BSND | OID3118 | Oncology | BSND | Q8WZ55 | 1 | 54998933 | 55017172 | 0 | - | - | - | - | - | - | - | - | - | - | - | - | - | - | - | - | - | - | - | - | - | - | - | - | - | - | - | - | - |
| BTNL10 | OID3119 | Oncology | BTNL10 | A8MVZ5 | 1 | 228510425 | 228512305 | 0 | - | - | - | - | - | - | - | - | - | - | - | - | - | - | - | - | - | - | - | - | - | - | - | - | - | - | - | - | - |
| CASQ2 | OID3006 | Cardiom | CASQ2 | O14958 | 1 | 115700021 | 115768714 | 0 | - | - | - | - | - | - | - | - | - | - | - | - | - | - | - | - | - | - | - | - | - | - | - | - | - | - | - | - | - |
| CEP350 | OID3124 | Oncology | CEP350 | Q5VT06 | 1 | 179954674 | 180114880 | 0 | - | - | - | - | - | - | - | - | - | - | - | - | - | - | - | - | - | - | - | - | - | - | - | - | - | - | - | - | - |
| CGN | OID3089 | Neurolog | CGN | Q9P2M7 | 1 | 151510510 | 151538692 | 0 | - | - | - | - | - | - | - | - | - | - | - | - | - | - | - | - | - | - | - | - | - | - | - | - | - | - | - | - | - |
| CLSPN | OID2083 | Neurolog | CLSPN | Q9HAW4 | 1 | 35720218 | 35769978 | 0 | - | - | - | - | - | - | - | - | - | - | - | - | - | - | - | - | - | - | - | - | - | - | - | - | - | - | - | - | - |
| CLSTN1 | OID2087 | Neurolog | CLSTN1 | O94985 | 1 | 9728926 | 9823984 | 0 | - | - | - | - | - | - | - | - | - | - | - | - | - | - | - | - | - | - | - | - | - | - | - | - | - | - | - | - | - |
| CNST | OID2012 | Cardiom | CNST | Q6PJW8 | 1 | 246566444 | 246668595 | 0 | - | - | - | - | - | - | - | - | - | - | - | - | - | - | - | - | - | - | - | - | - | - | - | - | - | - | - | - | - |
| COL9A2 | OID3125 | Oncology | COL9A2 | Q14055 | 1 | 40300489 | 40317813 | 0 | - | - | - | - | - | - | - | - | - | - | - | - | - | - | - | - | - | - | - | - | - | - | - | - | - | - | - | - | - |
| CSDE1 | OID3018 | Cardiom | CSDE1 | O75534 | 1 | 114716913 | 114758676 | 0 | - | - | - | - | - | - | - | - | - | - | - | - | - | - | - | - | - | - | - | - | - | - | - | - | - | - | - | - | - |
| DFFA | OID2062 | Inflamma | DFFA | O00273 | 1 | 10456522 | 10472529 | 0 | - | - | - | - | - | - | - | - | - | - | - | - | - | - | - | - | - | - | - | - | - | - | - | - | - | - | - | - | - |
| DNM3 | OID3100 | Neurolog | DNM3 | Q9UQ16 | 1 | 171817887 | 172418466 | 0 | - | - | - | - | - | - | - | - | - | - | - | - | - | - | - | - | - | - | - | - | - | - | - | - | - | - | - | - | - |
| ELAVL4 | OID3085 | Neurolog | ELAVL4 | P26378 | 1 | 50024029 | 50203772 | 0 | - | - | - | - | - | - | - | - | - | - | - | - | - | - | - | - | - | - | - | - | - | - | - | - | - | - | - | - | - |
| ENSA | OID3135 | Oncology | ENSA | O43768 | 1 | 150600851 | 150629612 | 0 | - | - | - | - | - | - | - | - | - | - | - | - | - | - | - | - | - | - | - | - | - | - | - | - | - | - | - | - | - |
| EPHA10 | OID2084 | Neurolog | EPHA10 | Q5JZY3 | 1 | 37713880 | 37765133 | 0 | - | - | - | - | - | - | - | - | - | - | - | - | - | - | - | - | - | - | - | - | - | - | - | - | - | - | - | - | - |
| EXOSC1 | OID3008 | Cardiom | EXOSC1 | Q01780 | 1 | 11066618 | 11099869 | 0 | - | - | - | - | - | - | - | - | - | - | - | - | - | - | - | - | - | - | - | - | - | - | - | - | - | - | - | - | - |
| FH | OID3083 | Neurolog | FH | P07954 | 1 | 241497511 | 241519799 | 0 | - | - | - | - | - | - | - | - | - | - | - | - | - | - | - | - | - | - | - | - | - | - | - | - | - | - | - | - | - |
| GBA | OID3096 | Neurolog | GBA | P04062 | 1 | 155234452 | 155244699 | 0 | - | - | - | - | - | - | - | - | - | - | - | - | - | - | - | - | - | - | - | - | - | - | - | - | - | - | - | - | - |
| GBP6 | OID3013 | Cardiom | GBP6 | Q6ZN66 | 1 | 89364059 | 89388160 | 0 | - | - | - | - | - | - | - | - | - | - | - | - | - | - | - | - | - | - | - | - | - | - | - | - | - | - | - | - | - |
| GJA8 | OID3118 | Oncology | GJA8 | P48165 | 1 | 147902795 | 147909269 | 0 | - | - | - | - | - | - | - | - | - | - | - | - | - | - | - | - | - | - | - | - | - | - | - | - | - | - | - | - | - |
| GUK1 | OID3014 | Cardiom | GUK1 | Q16774 | 1 | 228139962 | 228148984 | 0 | - | - | - | - | - | - | - | - | - | - | - | - | - | - | - | - | - | - | - | - | - | - | - | - | - | - | - | - | - |
| HMGCL | OID3021 | Cardiom | HMGCL | P35914 | 1 | 23801885 | 23838620 | 0 | - | - | - | - | - | - | - | - | - | - | - | - | - | - | - | - | - | - | - | - | - | - | - | - | - | - | - | - | - |
| HSD11B1 | OID2057 | Inflamma | HSD11B1 | P28845 | 1 | 209686178 | 209734949 | 0 | - | - | - | - | - | - | - | - | - | - | - | - | - | - | - | - | - | - | - | - | - | - | - | - | - | - | - | - | - |
| IL12RB2 | OID3042 | Inflamma | IL12RB2 | Q99665 | 1 | 67307364 | 67398724 | 0 | - | - | - | - | - | - | - | - | - | - | - | - | - | - | - | - | - | - | - | - | - | - | - | - | - | - | - | - | - |
| IL20 | OID2045 | Inflamma | IL20 | Q9NYY1 | 1 | 206865623 | 206869223 | 0 | - | - | - | - | - | - | - | - | - | - | - | - | - | - | - | - | - | - | - | - | - | - | - | - | - | - | - | - | - |
| IL24 | OID2044 | Inflamma | IL24 | Q13007 | 1 | 206897443 | 206904139 | 0 | - | - | - | - | - | - | - | - | - | - | - | - | - | - | - | - | - | - | - | - | - | - | - | - | - | - | - | - | - |
| JUN | OID2042 | Inflamma | JUN | P05412 | 1 | 58776845 | 58784048 | 0 | - | - | - | - | - | - | - | - | - | - | - | - | - | - | - | - | - | - | - | - | - | - | - | - | - | - | - | - | - |
| KAZN | OID3148 | Oncology | KAZN | Q674X7 | 1 | 13892792 | 15118043 | 0 | - | - | - | - | - | - | - | - | - | - | - | - | - | - | - | - | - | - | - | - | - | - | - | - | - | - | - | - | - |
| KCNC4 | OID3081 | Neurolog | KCNC4 | Q03721 | 1 | 110210314 | 110283100 | 0 | - | - | - | - | - | - | - | - | - | - | - | - | - | - | - | - | - | - | - | - | - | - | - | - | - | - | - | - | - |
| KIAA201 | OID3139 | Oncology | KIAA201 | Q8IYS2 | 1 | 11919591 | 11926428 | 0 | - | - | - | - | - | - | - | - | - | - | - | - | - | - | - | - | - | - | - | - | - | - | - | - | - | - | - | - | - |
| KIRREL1 | OID3080 | Neurolog | KIRREL1 | Q96J84 | 1 | 157993273 | 158100262 | 0 | - | - | - | - | - | - | - | - | - | - | - | - | - | - | - | - | - | - | - | - | - | - | - | - | - | - | - | - | - |
| LAMTOR | OID3139 | Oncology | LAMTOR | O43504 | 1 | 110401249 | 110407942 | 0 | - | - | - | - | - | - | - | - | - | - | - | - | - | - | - | - | - | - | - | - | - | - | - | - | - | - | - | - | - |
| LBR | OID2103 | Neurolog | LBR | Q14739 | 1 | 225401502 | 225428925 | 0 | - | - | - | - | - | - | - | - | - | - | - | - | - | - | - | - | - | - | - | - | - | - | - | - | - | - | - | - | - |
| LELP1 | OID3084 | Neurolog | LELP1 | Q5T871 | 1 | 153203430 | 153205120 | 0 | - | - | - | - | - | - | - | - | - | - | - | - | - | - | - | - | - | - | - | - | - | - | - | - | - | - | - | - | - |
| LYPLA2 | OID3081 | Neurolog | LYPLA2 | O95372 | 1 | 23791145 | 23795539 | 0 | - | - | - | - | - | - | - | - | - | - | - | - | - | - | - | - | - | - | - | - | - | - | - | - | - | - | - | - | - |
| MANEAL | OID3124 | Oncology | MANEAL | Q5VSG8 | 1 | 37793847 | 37801137 | 0 | - | - | - | - | - | - | - | - | - | - | - | - | - | - | - | - | - | - | - | - | - | - | - | - | - | - | - | - | - |
| MED18 | OID2123 | Oncology | MED18 | Q9BUE0 | 1 | 28329002 | 28335965 | 0 | - | - | - | - | - | - | - | - | - | - | - | - | - | - | - | - | - | - | - | - | - | - | - | - | - | - | - | - | - |
| MRPL24 | OID3017 | Cardiom | MRPL24 | Q96A35 | 1 | 156737303 | 156741590 | 0 | - | - | - | - | - | - | - | - | - | - | - | - | - | - | - | - | - | - | - | - | - | - | - | - | - | - | - | - | - |
| NECAP2 | OID3008 | Cardiom | NECAP2 | Q9NVZ3 | 1 | 16440721 | 16460078 | 0 | - | - | - | - | - | - | - | - | - | - | - | - | - | - | - | - | - | - | - | - | - | - | - | - | - | - | - | - | - |
| NEK7 | OID3125 | Oncology | NEK7 | Q8TDX7 | 1 | 198156994 | 198322420 | 0 | - | - | - | - | - | - | - | - | - | - | - | - | - | - | - | - | - | - | - | - | - | - | - | - | - | - | - | - | - |
| NENF | OID3135 | Oncology | NENF | Q9UMX5 | 1 | 212432920 | 212446379 | 0 | - | - | - | - | - | - | - | - | - | - | - | - | - | - | - | - | - | - | - | - | - | - | - | - | - | - | - | - | - |
| NGF | OID2092 | Neurolog | NGF | P01138 | 1 | 115285904 | 115338770 | 0 | - | - | - | - | - | - | - | - | - | - | - | - | - | - | - | - | - | - | - | - | - | - | - | - | - | - | - | - | - |
| NIT1 | OID3025 | Cardiom | NIT1 | Q86X76 | 1 | 161118086 | 161125445 | 0 | - | - | - | - | - | - | - | - | - | - | - | - | - | - | - | - | - | - | - | - | - | - | - | - | - | - | - | - | - |
| NPHS2 | OID3046 | Inflamma | NPHS2 | Q9NP85 | 1 | 179550539 | 179575952 | 0 | - | - | - | - | - | - | - | - | - | - | - | - | - | - | - | - | - | - | - | - | - | - | - | - | - | - | - | - | - |
| NPR1 | OID3011 | Cardiom | NPR1 | P16066 | 1 | 153678688 | 153693992 | 0 | - | - | - | - | - | - | - | - | - | - | - | - | - | - | - | - | - | - | - | - | - | - | - | - | - | - | - | - | - |
| NT5C1A | OID3110 | Neurolog | NT5C1A | Q9BXI3 | 1 | 39651229 | 39672107 | 0 | - | - | - | - | - | - | - | - | - | - | - | - | - | - | - | - | - | - | - | - | - | - | - | - | - | - | - | - | - |
| NUDC | OID2062 | Inflamma | NUDC | Q9Y266 | 1 | 26900238 | 26946871 | 0 | - | - | - | - | - | - | - | - | - | - | - | - | - | - | - | - | - | - | - | - | - | - | - | - | - | - | - | - | - |
| OTUD7B | OID3106 | Neurolog | OTUD7B | Q6GQQ9 | 1 | 149937812 | 150010726 | 0 | - | - | - | - | - | - | - | - | - | - | - | - | - | - | - | - | - | - | - | - | - | - | - | - | - | - | - | - | - |
| PAFAH2 | OID3128 | Oncology | PAFAH2 | Q99487 | 1 | 25959767 | 25998117 | 0 | - | - | - | - | - | - | - | - | - | - | - | - | - | - | - | - | - | - | - | - | - | - | - | - | - | - | - | - | - |
| PFDN2 | OID2089 | Neurolog | PFDN2 | Q9UHV9 | 1 | 161100556 | 161118055 | 0 | - | - | - | - | - | - | - | - | - | - | - | - | - | - | - | - | - | - | - | - | - | - | - | - | - | - | - | - | - |
| PITHD1 | OID3121 | Oncology | PITHD1 | Q9GZP4 | 1 | 23778418 | 23788232 | 0 | - | - | - | - | - | - | - | - | - | - | - | - | - | - | - | - | - | - | - | - | - | - | - | - | - | - | - | - | - |
| PPP1R12 | OID3130 | Oncology | PPP1R12 | O60237 | 1 | 202348699 | 202592706 | 0 | - | - | - | - | - | - | - | - | - | - | - | - | - | - | - | - | - | - | - | - | - | - | - | - | - | - | - | - | - |
| PPT1 | OID3094 | Neurolog | PPT1 | P50897 | 1 | 40072710 | 40097260 | 0 | - | - | - | - | - | - | - | - | - | - | - | - | - | - | - | - | - | - | - | - | - | - | - | - | - | - | - | - | - |
| RAP1A | OID3045 | Inflamma | RAP1A | P62834 | 1 | 111542218 | 111716691 | 0 | - | - | - | - | - | - | - | - | - | - | - | - | - | - | - | - | - | - | - | - | - | - | - | - | - | - | - | - | - |
| RCC1 | OID3135 | Oncology | RCC1 | P18754 | 1 | 28505943 | 28539300 | 0 | - | - | - | - | - | - | - | - | - | - | - | - | - | - | - | - | - | - | - | - | - | - | - | - | - | - | - | - | - |
| RGS8 | OID2045 | Inflamma | RGS8 | P57771 | 1 | 182641816 | 182684576 | 0 | - | - | - | - | - | - | - | - | - | - | - | - | - | - | - | - | - | - | - | - | - | - | - | - | - | - | - | - | - |
| RRP15 | OID3086 | Neurolog | RRP15 | Q9Y3B9 | 1 | 218285293 | 218337983 | 0 | - | - | - | - | - | - | - | - | - | - | - | - | - | - | - | - | - | - | - | - | - | - | - | - | - | - | - | - | - |
| SARG | OID3033 | Cardiom | C1orf116 | Q9BW04 | 1 | 207018522 | 207032756 | 0 | - | - | - | - | - | - | - | - | - | - | - | - | - | - | - | - | - | - | - | - | - | - | - | - | - | - | - | - | - |
| SHC1 | OID3122 | Oncology | SHC1 | P29353 | 1 | 154962298 | 154974395 | 0 | - | - | - | - | - | - | - | - | - | - | - | - | - | - | - | - | - | - | - | - | - | - | - | - | - | - | - | - | - |
| SMPDL3 | OID3138 | Oncology | SMPDL3 | Q92485 | 1 | 27935000 | 27959157 | 0 | - | - | - | - | - | - | - | - | - | - | - | - | - | - | - | - | - | - | - | - | - | - | - | - | - | - | - | - | - |
| SNAPIN | OID3101 | Neurolog | SNAPIN | O95295 | 1 | 153658703 | 153661852 | 0 | - | - | - | - | - | - | - | - | - | - | - | - | - | - | - | - | - | - | - | - | - | - | - | - | - | - | - | - | - |
| SPRR1B | OID3129 | Oncology | SPRR1B | P22528 | 1 | 153031203 | 153032900 | 0 | - | - | - | - | - | - | - | - | - | - | - | - | - | - | - | - | - | - | - | - | - | - | - | - | - | - | - | - | - |
| STX6 | OID2121 | Oncology | STX6 | O43752 | 1 | 180972712 | 181023121 | 0 | - | - | - | - | - | - | - | - | - | - | - | - | - | - | - | - | - | - | - | - | - | - | - | - | - | - | - | - | - |
| STXBP3 | OID2124 | Oncology | STXBP3 | O00186 | 1 | 108746674 | 108809523 | 0 | - | - | - | - | - | - | - | - | - | - | - | - | - | - | - | - | - | - | - | - | - | - | - | - | - | - | - | - | - |
| THRAP3 | OID3005 | Cardiom | THRAP3 | Q9Y2W1 | 1 | 36224432 | 36305357 | 0 | - | - | - | - | - | - | - | - | - | - | - | - | - | - | - | - | - | - | - | - | - | - | - | - | - | - | - | - | - |
| TMOD4 | OID3124 | Oncology | TMOD4 | Q9NZQ9 | 1 | 151169986 | 151176284 | 0 | - | - | - | - | - | - | - | - | - | - | - | - | - | - | - | - | - | - | - | - | - | - | - | - | - | - | - | - | - |
| TOMM20 | OID3035 | Cardiom | TOMM20 | Q15388 | 1 | 235109341 | 235128837 | 0 | - | - | - | - | - | - | - | - | - | - | - | - | - | - | - | - | - | - | - | - | - | - | - | - | - | - | - | - | - |
| TP73 | OID3087 | Neurolog | TP73 | O15350 | 1 | 3652516 | 3736201 | 0 | - | - | - | - | - | - | - | - | - | - | - | - | - | - | - | - | - | - | - | - | - | - | - | - | - | - | - | - | - |
| TPM3 | OID3005 | Cardiom | TPM3 | P06753 | 1 | 154155308 | 154194648 | 0 | - | - | - | - | - | - | - | - | - | - | - | - | - | - | - | - | - | - | - | - | - | - | - | - | - | - | - | - | - |
| TPR | OID3014 | Cardiom | TPR | P12270 | 1 | 186311652 | 186375693 | 0 | - | - | - | - | - | - | - | - | - | - | - | - | - | - | - | - | - | - | - | - | - | - | - | - | - | - | - | - | - |
| TSHB | OID2012 | Cardiom | TSHB | P01222 | 1 | 115029826 | 115034302 | 0 | - | - | - | - | - | - | - | - | - | - | - | - | - | - | - | - | - | - | - | - | - | - | - | - | - | - | - | - | - |
| TSNAX | OID3018 | Cardiom | TSNAX | Q99598 | 1 | 231528541 | 231566524 | 0 | - | - | - | - | - | - | - | - | - | - | - | - | - | - | - | - | - | - | - | - | - | - | - | - | - | - | - | - | - |
| TXLNA | OID2095 | Neurolog | TXLNA | P40222 | 1 | 32179675 | 32198285 | 0 | - | - | - | - | - | - | - | - | - | - | - | - | - | - | - | - | - | - | - | - | - | - | - | - | - | - | - | - | - |
| YARS1 | OID3149 | Oncology | YARS1 | P54577 | 1 | 32775237 | 32818031 | 0 | - | - | - | - | - | - | - | - | - | - | - | - | - | - | - | - | - | - | - | - | - | - | - | - | - | - | - | - | - |
| ZP4 | OID3118 | Oncology | ZP4 | Q12836 | 1 | 237877864 | 237890922 | 0 | - | - | - | - | - | - | - | - | - | - | - | - | - | - | - | - | - | - | - | - | - | - | - | - | - | - | - | - | - |
| ALMS1 | OID3127 | Oncology | ALMS1 | Q8TCU4 | 2 | 73385758 | 73625166 | 0 | - | - | - | - | - | - | - | - | - | - | - | - | - | - | - | - | - | - | - | - | - | - | - | - | - | - | - | - | - |
| ALPI | OID3049 | Inflamma | ALPI | P09923 | 2 | 232456125 | 232460753 | 0 | - | - | - | - | - | - | - | - | - | - | - | - | - | - | - | - | - | - | - | - | - | - | - | - | - | - | - | - | - |
| APOB | OID3067 | Inflamma | APOB | P04114 | 2 | 21001429 | 21044073 | 0 | - | - | - | - | - | - | - | - | - | - | - | - | - | - | - | - | - | - | - | - | - | - | - | - | - | - | - | - | - |
| ATF2 | OID2079 | Neurolog | ATF2 | P15336 | 2 | 175072250 | 175168382 | 0 | - | - | - | - | - | - | - | - | - | - | - | - | - | - | - | - | - | - | - | - | - | - | - | - | - | - | - | - | - |
| ATG16L1 | OID3146 | Oncology | ATG16L1 | Q676U5 | 2 | 233210051 | 233295674 | 0 | - | - | - | - | - | - | - | - | - | - | - | - | - | - | - | - | - | - | - | - | - | - | - | - | - | - | - | - | - |
| BCL2L11 | OID2051 | Inflamma | BCL2L11 | O43521 | 2 | 111119378 | 111168445 | 0 | - | - | - | - | - | - | - | - | - | - | - | - | - | - | - | - | - | - | - | - | - | - | - | - | - | - | - | - | - |
| CFC1 | OID2126 | Oncology | CFC1 | P0CG37 | 2 | 130592165 | 130599575 | 0 | - | - | - | - | - | - | - | - | - | - | - | - | - | - | - | - | - | - | - | - | - | - | - | - | - | - | - | - | - |
| CLASP1 | OID3119 | Oncology | CLASP1 | Q7Z460 | 2 | 121337776 | 121649476 | 0 | - | - | - | - | - | - | - | - | - | - | - | - | - | - | - | - | - | - | - | - | - | - | - | - | - | - | - | - | - |
| COL4A4 | OID3133 | Oncology | COL4A4 | P53420 | 2 | 227002714 | 227164453 | 0 | - | - | - | - | - | - | - | - | - | - | - | - | - | - | - | - | - | - | - | - | - | - | - | - | - | - | - | - | - |
| COMMD1 | OID3022 | Cardiom | COMMD1 | Q8N668 | 2 | 61888724 | 62147247 | 0 | - | - | - | - | - | - | - | - | - | - | - | - | - | - | - | - | - | - | - | - | - | - | - | - | - | - | - | - | - |
| COX5B | OID2129 | Oncology | COX5B | P10606 | 2 | 97646062 | 97648383 | 0 | - | - | - | - | - | - | - | - | - | - | - | - | - | - | - | - | - | - | - | - | - | - | - | - | - | - | - | - | - |
| CTLA4 | OID3036 | Cardiom | CTLA4 | P16410 | 2 | 203853888 | 203873965 | 0 | - | - | - | - | - | - | - | - | - | - | - | - | - | - | - | - | - | - | - | - | - | - | - | - | - | - | - | - | - |
| DCDC2C | OID3123 | Oncology | DCDC2C | A8MYV0 | 2 | 3703575 | 3848008 | 0 | - | - | - | - | - | - | - | - | - | - | - | - | - | - | - | - | - | - | - | - | - | - | - | - | - | - | - | - | - |
| DCTN1 | OID2139 | Oncology | DCTN1 | Q14203 | 2 | 74361154 | 74392087 | 0 | - | - | - | - | - | - | - | - | - | - | - | - | - | - | - | - | - | - | - | - | - | - | - | - | - | - | - | - | - |
| DDX1 | OID3151 | Oncology | DDX1 | Q92499 | 2 | 15591178 | 15634346 | 0 | - | - | - | - | - | - | - | - | - | - | - | - | - | - | - | - | - | - | - | - | - | - | - | - | - | - | - | - | - |
| DNAJB2 | OID3066 | Inflamma | DNAJB2 | P25686 | 2 | 219279342 | 219286898 | 0 | - | - | - | - | - | - | - | - | - | - | - | - | - | - | - | - | - | - | - | - | - | - | - | - | - | - | - | - | - |
| DOK1 | OID3111 | Neurolog | DOK1 | Q99704 | 2 | 74549026 | 74557551 | 0 | - | - | - | - | - | - | - | - | - | - | - | - | - | - | - | - | - | - | - | - | - | - | - | - | - | - | - | - | - |
| EPB41L5 | OID3088 | Neurolog | EPB41L5 | Q9HCM4 | 2 | 120013077 | 120179119 | 0 | - | - | - | - | - | - | - | - | - | - | - | - | - | - | - | - | - | - | - | - | - | - | - | - | - | - | - | - | - |
| EPCAM | OID2074 | Inflamma | EPCAM | P16422 | 2 | 47345158 | 47387601 | 0 | - | - | - | - | - | - | - | - | - | - | - | - | - | - | - | - | - | - | - | - | - | - | - | - | - | - | - | - | - |
| GAD1 | OID3116 | Oncology | GAD1 | Q99259 | 2 | 170813213 | 170861151 | 0 | - | - | - | - | - | - | - | - | - | - | - | - | - | - | - | - | - | - | - | - | - | - | - | - | - | - | - | - | - |
| GCG | OID2126 | Oncology | GCG | P01275 | 2 | 162142882 | 162152404 | 0 | - | - | - | - | - | - | - | - | - | - | - | - | - | - | - | - | - | - | - | - | - | - | - | - | - | - | - | - | - |
| GIGYF2 | OID3106 | Neurolog | GIGYF2 | Q6Y7W6 | 2 | 232697299 | 232860605 | 0 | - | - | - | - | - | - | - | - | - | - | - | - | - | - | - | - | - | - | - | - | - | - | - | - | - | - | - | - | - |
| GKN1 | OID2088 | Neurolog | GKN1 | Q9NS71 | 2 | 68974573 | 68980980 | 0 | - | - | - | - | - | - | - | - | - | - | - | - | - | - | - | - | - | - | - | - | - | - | - | - | - | - | - | - | - |
| GLI2 | OID3042 | Inflamma | GLI2 | P10070 | 2 | 120735623 | 120992653 | 0 | - | - | - | - | - | - | - | - | - | - | - | - | - | - | - | - | - | - | - | - | - | - | - | - | - | - | - | - | - |
| HK2 | OID2006 | Cardiom | HK2 | P52789 | 2 | 74834127 | 74893359 | 0 | - | - | - | - | - | - | - | - | - | - | - | - | - | - | - | - | - | - | - | - | - | - | - | - | - | - | - | - | - |
| HTRA2 | OID2147 | Oncology | HTRA2 | O43464 | 2 | 74529725 | 74533332 | 0 | - | - | - | - | - | - | - | - | - | - | - | - | - | - | - | - | - | - | - | - | - | - | - | - | - | - | - | - | - |
| IKZF2 | OID2121 | Oncology | IKZF2 | Q9UKS7 | 2 | 212999691 | 213152427 | 0 | - | - | - | - | - | - | - | - | - | - | - | - | - | - | - | - | - | - | - | - | - | - | - | - | - | - | - | - | - |
| IL18RAP | OID2090 | Neurolog | IL18RAP | O95256 | 2 | 102418689 | 102452565 | 0 | - | - | - | - | - | - | - | - | - | - | - | - | - | - | - | - | - | - | - | - | - | - | - | - | - | - | - | - | - |
| IL1A | OID2048 | Inflamma | IL1A | P01583 | 2 | 112773925 | 112784493 | 0 | - | - | - | - | - | - | - | - | - | - | - | - | - | - | - | - | - | - | - | - | - | - | - | - | - | - | - | - | - |
| IL1B | OID2042 | Inflamma | IL1B | P01584 | 2 | 112829751 | 112836816 | 0 | - | - | - | - | - | - | - | - | - | - | - | - | - | - | - | - | - | - | - | - | - | - | - | - | - | - | - | - | - |
| ILKAP | OID2085 | Neurolog | ILKAP | Q9H0C8 | 2 | 238170402 | 238203708 | 0 | - | - | - | - | - | - | - | - | - | - | - | - | - | - | - | - | - | - | - | - | - | - | - | - | - | - | - | - | - |
| ITGB1BP | OID2126 | Oncology | ITGB1BP | O14713 | 2 | 9403475 | 9423528 | 0 | - | - | - | - | - | - | - | - | - | - | - | - | - | - | - | - | - | - | - | - | - | - | - | - | - | - | - | - | - |
| KDM3A | OID3042 | Inflamma | KDM3A | Q9Y4C1 | 2 | 86440647 | 86492716 | 0 | - | - | - | - | - | - | - | - | - | - | - | - | - | - | - | - | - | - | - | - | - | - | - | - | - | - | - | - | - |
| KLHL41 | OID3098 | Neurolog | KLHL41 | O60662 | 2 | 169509702 | 169526258 | 0 | - | - | - | - | - | - | - | - | - | - | - | - | - | - | - | - | - | - | - | - | - | - | - | - | - | - | - | - | - |
| LRP2 | OID3096 | Neurolog | LRP2 | P98164 | 2 | 169127109 | 169362534 | 0 | - | - | - | - | - | - | - | - | - | - | - | - | - | - | - | - | - | - | - | - | - | - | - | - | - | - | - | - | - |
| LYPD1 | OID2082 | Neurolog | LYPD1 | Q8N2G4 | 2 | 132643286 | 132671579 | 0 | - | - | - | - | - | - | - | - | - | - | - | - | - | - | - | - | - | - | - | - | - | - | - | - | - | - | - | - | - |
| MAPRE3 | OID3117 | Oncology | MAPRE3 | Q9UPY8 | 2 | 26970637 | 27027219 | 0 | - | - | - | - | - | - | - | - | - | - | - | - | - | - | - | - | - | - | - | - | - | - | - | - | - | - | - | - | - |
| MYL1 | OID3012 | Cardiom | MYL1 | P05976 | 2 | 210290150 | 210315174 | 0 | - | - | - | - | - | - | - | - | - | - | - | - | - | - | - | - | - | - | - | - | - | - | - | - | - | - | - | - | - |
| NCK2 | OID2068 | Inflamma | NCK2 | O43639 | 2 | 105744912 | 105894274 | 0 | - | - | - | - | - | - | - | - | - | - | - | - | - | - | - | - | - | - | - | - | - | - | - | - | - | - | - | - | - |
| NEB | OID3010 | Cardiom | NEB | P20929 | 2 | 151485336 | 151734487 | 0 | - | - | - | - | - | - | - | - | - | - | - | - | - | - | - | - | - | - | - | - | - | - | - | - | - | - | - | - | - |
| PAIP2B | OID3122 | Oncology | PAIP2B | Q9ULR5 | 2 | 71182738 | 71227103 | 0 | - | - | - | - | - | - | - | - | - | - | - | - | - | - | - | - | - | - | - | - | - | - | - | - | - | - | - | - | - |
| PCARE | OID3083 | Neurolog | PCARE | A6NGG8 | 2 | 29060976 | 29074523 | 0 | - | - | - | - | - | - | - | - | - | - | - | - | - | - | - | - | - | - | - | - | - | - | - | - | - | - | - | - | - |
| PIKFYVE | OID3046 | Inflamma | PIKFYVE | Q9Y2I7 | 2 | 208266255 | 208358746 | 0 | - | - | - | - | - | - | - | - | - | - | - | - | - | - | - | - | - | - | - | - | - | - | - | - | - | - | - | - | - |
| PNPT1 | OID2044 | Inflamma | PNPT1 | Q8TCS8 | 2 | 55634061 | 55693863 | 0 | - | - | - | - | - | - | - | - | - | - | - | - | - | - | - | - | - | - | - | - | - | - | - | - | - | - | - | - | - |
| PPM1B | OID3054 | Inflamma | PPM1B | O75688 | 2 | 44167969 | 44244384 | 0 | - | - | - | - | - | - | - | - | - | - | - | - | - | - | - | - | - | - | - | - | - | - | - | - | - | - | - | - | - |
| PPP3R1 | OID2090 | Neurolog | PPP3R1 | P63098 | 2 | 68178857 | 68256237 | 0 | - | - | - | - | - | - | - | - | - | - | - | - | - | - | - | - | - | - | - | - | - | - | - | - | - | - | - | - | - |
| PREB | OID2043 | Inflamma | PREB | Q9HCU5 | 2 | 27130756 | 27134666 | 0 | - | - | - | - | - | - | - | - | - | - | - | - | - | - | - | - | - | - | - | - | - | - | - | - | - | - | - | - | - |
| PRKAG3 | OID3094 | Neurolog | PRKAG3 | Q9UGI9 | 2 | 218822308 | 218832086 | 0 | - | - | - | - | - | - | - | - | - | - | - | - | - | - | - | - | - | - | - | - | - | - | - | - | - | - | - | - | - |
| PRKRA | OID2122 | Oncology | PRKRA | O75569 | 2 | 178431292 | 178451512 | 0 | - | - | - | - | - | - | - | - | - | - | - | - | - | - | - | - | - | - | - | - | - | - | - | - | - | - | - | - | - |
| PSMD1 | OID3105 | Neurolog | PSMD1 | Q99460 | 2 | 231056845 | 231173116 | 0 | - | - | - | - | - | - | - | - | - | - | - | - | - | - | - | - | - | - | - | - | - | - | - | - | - | - | - | - | - |
| PTRHD1 | OID3065 | Inflamma | PTRHD1 | Q6GMV3 | 2 | 24789728 | 24793391 | 0 | - | - | - | - | - | - | - | - | - | - | - | - | - | - | - | - | - | - | - | - | - | - | - | - | - | - | - | - | - |
| RAB10 | OID3035 | Cardiom | RAB10 | P61026 | 2 | 26034084 | 26137454 | 0 | - | - | - | - | - | - | - | - | - | - | - | - | - | - | - | - | - | - | - | - | - | - | - | - | - | - | - | - | - |
| RAB3GAI | OID3082 | Neurolog | RAB3GAI | Q15042 | 2 | 135052289 | 135176667 | 0 | - | - | - | - | - | - | - | - | - | - | - | - | - | - | - | - | - | - | - | - | - | - | - | - | - | - | - | - | - |
| RANBP2 | OID3081 | Neurolog | RANBP2 | P49792 | 2 | 108719482 | 108785810 | 0 | - | - | - | - | - | - | - | - | - | - | - | - | - | - | - | - | - | - | - | - | - | - | - | - | - | - | - | - | - |
| SAG | OID3087 | Neurolog | SAG | P10523 | 2 | 233307816 | 233347055 | 0 | - | - | - | - | - | - | - | - | - | - | - | - | - | - | - | - | - | - | - | - | - | - | - | - | - | - | - | - | - |
| SCN2A | OID3083 | Neurolog | SCN2A | Q99250 | 2 | 165194993 | 165392310 | 0 | - | - | - | - | - | - | - | - | - | - | - | - | - | - | - | - | - | - | - | - | - | - | - | - | - | - | - | - | - |
| SCN3A | OID3090 | Neurolog | SCN3A | Q9NY46 | 2 | 165087526 | 165204050 | 0 | - | - | - | - | - | - | - | - | - | - | - | - | - | - | - | - | - | - | - | - | - | - | - | - | - | - | - | - | - |
| SEMA4C | OID2132 | Oncology | SEMA4C | Q9C0C4 | 2 | 96859718 | 96870757 | 0 | - | - | - | - | - | - | - | - | - | - | - | - | - | - | - | - | - | - | - | - | - | - | - | - | - | - | - | - | - |
| SLC1A4 | OID3092 | Neurolog | SLC1A4 | P43007 | 2 | 64988477 | 65023865 | 0 | - | - | - | - | - | - | - | - | - | - | - | - | - | - | - | - | - | - | - | - | - | - | - | - | - | - | - | - | - |
| SSB | OID2081 | Neurolog | SSB | P05455 | 2 | 169791933 | 169812064 | 0 | - | - | - | - | - | - | - | - | - | - | - | - | - | - | - | - | - | - | - | - | - | - | - | - | - | - | - | - | - |
| TBR1 | OID3092 | Neurolog | TBR1 | Q16650 | 2 | 161416297 | 161425870 | 0 | - | - | - | - | - | - | - | - | - | - | - | - | - | - | - | - | - | - | - | - | - | - | - | - | - | - | - | - | - |
| TIA1 | OID2006 | Cardiom | TIA1 | P31483 | 2 | 70209444 | 70248660 | 0 | - | - | - | - | - | - | - | - | - | - | - | - | - | - | - | - | - | - | - | - | - | - | - | - | - | - | - | - | - |
| TPRKB | OID3123 | Oncology | TPRKB | Q9Y3C4 | 2 | 73729104 | 73737400 | 0 | - | - | - | - | - | - | - | - | - | - | - | - | - | - | - | - | - | - | - | - | - | - | - | - | - | - | - | - | - |
| TTN | OID3010 | Cardiom | TTN | Q8WZ42 | 2 | 178525989 | 178830802 | 0 | - | - | - | - | - | - | - | - | - | - | - | - | - | - | - | - | - | - | - | - | - | - | - | - | - | - | - | - | - |
| YWHAQ | OID3060 | Inflamma | YWHAQ | P27348 | 2 | 9583967 | 9630997 | 0 | - | - | - | - | - | - | - | - | - | - | - | - | - | - | - | - | - | - | - | - | - | - | - | - | - | - | - | - | - |
| ARL13B | OID3016 | Cardiom | ARL13B | Q3SXY8 | 3 | 93980139 | 94055678 | 0 | - | - | - | - | - | - | - | - | - | - | - | - | - | - | - | - | - | - | - | - | - | - | - | - | - | - | - | - | - |
| ATP1B3 | OID3006 | Cardiom | ATP1B3 | P54709 | 3 | 141876124 | 141926549 | 0 | - | - | - | - | - | - | - | - | - | - | - | - | - | - | - | - | - | - | - | - | - | - | - | - | - | - | - | - | - |
| AZI2 | OID3105 | Neurolog | AZI2 | Q9H6S1 | 3 | 28315003 | 28349050 | 0 | - | - | - | - | - | - | - | - | - | - | - | - | - | - | - | - | - | - | - | - | - | - | - | - | - | - | - | - | - |
| BHLHE4 | OID3087 | Neurolog | BHLHE4 | O14503 | 3 | 4979437 | 4985323 | 0 | - | - | - | - | - | - | - | - | - | - | - | - | - | - | - | - | - | - | - | - | - | - | - | - | - | - | - | - | - |
| BRK1 | OID2088 | Neurolog | BRK1 | Q8WUW1 | 3 | 10115675 | 10127190 | 0 | - | - | - | - | - | - | - | - | - | - | - | - | - | - | - | - | - | - | - | - | - | - | - | - | - | - | - | - | - |
| BTLA | OID3123 | Oncology | BTLA | Q7Z6A9 | 3 | 112463966 | 112499472 | 0 | - | - | - | - | - | - | - | - | - | - | - | - | - | - | - | - | - | - | - | - | - | - | - | - | - | - | - | - | - |
| CADPS | OID3091 | Neurolog | CADPS | Q9ULU8 | 3 | 62398346 | 62875416 | 0 | - | - | - | - | - | - | - | - | - | - | - | - | - | - | - | - | - | - | - | - | - | - | - | - | - | - | - | - | - |
| CDC25A | OID3118 | Oncology | CDC25A | P30304 | 3 | 48157146 | 48188417 | 0 | - | - | - | - | - | - | - | - | - | - | - | - | - | - | - | - | - | - | - | - | - | - | - | - | - | - | - | - | - |
| CMC1 | OID3036 | Cardiom | CMC1 | Q7Z7K0 | 3 | 28241584 | 28325142 | 0 | - | - | - | - | - | - | - | - | - | - | - | - | - | - | - | - | - | - | - | - | - | - | - | - | - | - | - | - | - |
| COPB2 | OID3017 | Cardiom | COPB2 | P35606 | 3 | 139353946 | 139389736 | 0 | - | - | - | - | - | - | - | - | - | - | - | - | - | - | - | - | - | - | - | - | - | - | - | - | - | - | - | - | - |
| CRTAP | OID3090 | Neurolog | CRTAP | O75718 | 3 | 33114014 | 33147773 | 0 | - | - | - | - | - | - | - | - | - | - | - | - | - | - | - | - | - | - | - | - | - | - | - | - | - | - | - | - | - |
| CSPG5 | OID3084 | Neurolog | CSPG5 | O95196 | 3 | 47562238 | 47580792 | 0 | - | - | - | - | - | - | - | - | - | - | - | - | - | - | - | - | - | - | - | - | - | - | - | - | - | - | - | - | - |
| DCUN1D | OID3083 | Neurolog | DCUN1D | Q96GG9 | 3 | 182938074 | 182985953 | 0 | - | - | - | - | - | - | - | - | - | - | - | - | - | - | - | - | - | - | - | - | - | - | - | - | - | - | - | - | - |
| DNAJB8 | OID2009 | Cardiom | DNAJB8 | Q8NHS0 | 3 | 128462437 | 128466968 | 0 | - | - | - | - | - | - | - | - | - | - | - | - | - | - | - | - | - | - | - | - | - | - | - | - | - | - | - | - | - |
| EIF4G1 | OID2062 | Inflamma | EIF4G1 | Q04637 | 3 | 184314495 | 184335358 | 0 | - | - | - | - | - | - | - | - | - | - | - | - | - | - | - | - | - | - | - | - | - | - | - | - | - | - | - | - | - |
| ERC2 | OID3088 | Neurolog | ERC2 | O15083 | 3 | 55508311 | 56469095 | 0 | - | - | - | - | - | - | - | - | - | - | - | - | - | - | - | - | - | - | - | - | - | - | - | - | - | - | - | - | - |
| FGF12 | OID3045 | Inflamma | FGF12 | P61328 | 3 | 192139390 | 192767764 | 0 | - | - | - | - | - | - | - | - | - | - | - | - | - | - | - | - | - | - | - | - | - | - | - | - | - | - | - | - | - |
| FRMD4B | OID3015 | Cardiom | FRMD4B | Q9Y2L6 | 3 | 69168782 | 69542583 | 0 | - | - | - | - | - | - | - | - | - | - | - | - | - | - | - | - | - | - | - | - | - | - | - | - | - | - | - | - | - |
| LPP | OID3019 | Cardiom | LPP | Q93052 | 3 | 188153284 | 188890671 | 0 | - | - | - | - | - | - | - | - | - | - | - | - | - | - | - | - | - | - | - | - | - | - | - | - | - | - | - | - | - |
| LRTM1 | OID3090 | Neurolog | LRTM1 | Q9HBL6 | 3 | 54918231 | 54967088 | 0 | - | - | - | - | - | - | - | - | - | - | - | - | - | - | - | - | - | - | - | - | - | - | - | - | - | - | - | - | - |
| MYL3 | OID3025 | Cardiom | MYL3 | P08590 | 3 | 46835111 | 46882172 | 0 | - | - | - | - | - | - | - | - | - | - | - | - | - | - | - | - | - | - | - | - | - | - | - | - | - | - | - | - | - |
| NLGN1 | OID3090 | Neurolog | NLGN1 | Q8N2Q7 | 3 | 173396284 | 174294372 | 0 | - | - | - | - | - | - | - | - | - | - | - | - | - | - | - | - | - | - | - | - | - | - | - | - | - | - | - | - | - |
| NUDT16 | OID3148 | Oncology | NUDT16 | Q96DE0 | 3 | 131381671 | 131388830 | 0 | - | - | - | - | - | - | - | - | - | - | - | - | - | - | - | - | - | - | - | - | - | - | - | - | - | - | - | - | - |
| PHLDB2 | OID3116 | Oncology | PHLDB2 | Q86SQ0 | 3 | 111732497 | 111976517 | 0 | - | - | - | - | - | - | - | - | - | - | - | - | - | - | - | - | - | - | - | - | - | - | - | - | - | - | - | - | - |
| PPP1R2 | OID2017 | Cardiom | PPP1R2 | P41236 | 3 | 195514428 | 195543386 | 0 | - | - | - | - | - | - | - | - | - | - | - | - | - | - | - | - | - | - | - | - | - | - | - | - | - | - | - | - | - |
| RAB6B | OID2081 | Neurolog | RAB6B | Q9NRW1 | 3 | 133824235 | 133895882 | 0 | - | - | - | - | - | - | - | - | - | - | - | - | - | - | - | - | - | - | - | - | - | - | - | - | - | - | - | - | - |
| RFC4 | OID3137 | Oncology | RFC4 | P35249 | 3 | 186789880 | 186807058 | 0 | - | - | - | - | - | - | - | - | - | - | - | - | - | - | - | - | - | - | - | - | - | - | - | - | - | - | - | - | - |
| RNF168 | OID3050 | Inflamma | RNF168 | Q8IYW5 | 3 | 196468783 | 196503768 | 0 | - | - | - | - | - | - | - | - | - | - | - | - | - | - | - | - | - | - | - | - | - | - | - | - | - | - | - | - | - |
| RUVBL1 | OID2121 | Oncology | RUVBL1 | Q9Y265 | 3 | 128064778 | 128153914 | 0 | - | - | - | - | - | - | - | - | - | - | - | - | - | - | - | - | - | - | - | - | - | - | - | - | - | - | - | - | - |
| SATB1 | OID3103 | Neurolog | SATB1 | Q01826 | 3 | 18345377 | 18445621 | 0 | - | - | - | - | - | - | - | - | - | - | - | - | - | - | - | - | - | - | - | - | - | - | - | - | - | - | - | - | - |
| SEMA3G | OID3059 | Inflamma | SEMA3G | Q9NS98 | 3 | 52433035 | 52445103 | 0 | - | - | - | - | - | - | - | - | - | - | - | - | - | - | - | - | - | - | - | - | - | - | - | - | - | - | - | - | - |
| SOX2 | OID3098 | Neurolog | SOX2 | P48431 | 3 | 181711925 | 181714436 | 0 | - | - | - | - | - | - | - | - | - | - | - | - | - | - | - | - | - | - | - | - | - | - | - | - | - | - | - | - | - |
| TADA3 | OID3142 | Oncology | TADA3 | O75528 | 3 | 9779967 | 9793011 | 0 | - | - | - | - | - | - | - | - | - | - | - | - | - | - | - | - | - | - | - | - | - | - | - | - | - | - | - | - | - |
| TAGLN3 | OID3122 | Oncology | TAGLN3 | Q9UI15 | 3 | 111998739 | 112013887 | 0 | - | - | - | - | - | - | - | - | - | - | - | - | - | - | - | - | - | - | - | - | - | - | - | - | - | - | - | - | - |

| | | | | | | | | | | | | | | | | | | | | | | | | | | | | | | | | | | | | | |
|---|---|---|---|---|---|---|---|---|---|---|---|---|---|---|---|---|---|---|---|---|---|---|---|---|---|---|---|---|---|---|---|---|---|---|---|---|---|
| TBC1D5 | OID2052 | Inflamma | TBC1D5 | Q92609 | 3 | 17157162 | 18444817 | 0 | - | - | - | - | - | - | - | - | - | - | - | - | - | - | - | - | - | - | - | - | - | - | - | - | - | - | - | - | - |
| VIPR1 | OID3019 | Cardiom | VIPR1 | P32241 | 3 | 42489299 | 42537573 | 0 | - | - | - | - | - | - | - | - | - | - | - | - | - | - | - | - | - | - | - | - | - | - | - | - | - | - | - | - | - |
| ACSL1 | OID3008 | Cardiom | ACSL1 | P33121 | 4 | 184755595 | 184826818 | 0 | - | - | - | - | - | - | - | - | - | - | - | - | - | - | - | - | - | - | - | - | - | - | - | - | - | - | - | - | - |
| ANK2 | OID3016 | Cardiom | ANK2 | Q01484 | 4 | 112818032 | 113384221 | 0 | - | - | - | - | - | - | - | - | - | - | - | - | - | - | - | - | - | - | - | - | - | - | - | - | - | - | - | - | - |
| ANP32C | OID3020 | Cardiom | ANP32C | O43423 | 4 | 164197007 | 16419771 | 0 | - | - | - | - | - | - | - | - | - | - | - | - | - | - | - | - | - | - | - | - | - | - | - | - | - | - | - | - | - |
| CXCL3 | OID2078 | Inflamma | CXCL3 | P19876 | 4 | 74036589 | 74038807 | 0 | - | - | - | - | - | - | - | - | - | - | - | - | - | - | - | - | - | - | - | - | - | - | - | - | - | - | - | - | - |
| EIF4E | OID3055 | Inflamma | EIF4E | P06730 | 4 | 98879276 | 98929133 | 0 | - | - | - | - | - | - | - | - | - | - | - | - | - | - | - | - | - | - | - | - | - | - | - | - | - | - | - | - | - |
| ENOPH1 | OID3134 | Oncology | ENOPH1 | Q9UHY7 | 4 | 82430590 | 82461177 | 0 | - | - | - | - | - | - | - | - | - | - | - | - | - | - | - | - | - | - | - | - | - | - | - | - | - | - | - | - | - |
| GABRA4 | OID3080 | Neurolog | GABRA4 | P48169 | 4 | 46918900 | 46993581 | 0 | - | - | - | - | - | - | - | - | - | - | - | - | - | - | - | - | - | - | - | - | - | - | - | - | - | - | - | - | - |
| GRSF1 | OID3122 | Oncology | GRSF1 | Q12849 | 4 | 70815783 | 70839897 | 0 | - | - | - | - | - | - | - | - | - | - | - | - | - | - | - | - | - | - | - | - | - | - | - | - | - | - | - | - | - |
| IL15 | OID2056 | Inflamma | IL15 | P40933 | 4 | 141636583 | 141733987 | 0 | - | - | - | - | - | - | - | - | - | - | - | - | - | - | - | - | - | - | - | - | - | - | - | - | - | - | - | - | - |
| IL2 | OID2041 | Inflamma | IL2 | P60568 | 4 | 122451470 | 122456725 | 0 | - | - | - | - | - | - | - | - | - | - | - | - | - | - | - | - | - | - | - | - | - | - | - | - | - | - | - | - | - |
| JCHAIN | OID2049 | Inflamma | JCHAIN | P01591 | 4 | 70655541 | 70681817 | 0 | - | - | - | - | - | - | - | - | - | - | - | - | - | - | - | - | - | - | - | - | - | - | - | - | - | - | - | - | - |
| KCNIP4 | OID2081 | Neurolog | KCNIP4 | Q6PIL6 | 4 | 20728606 | 21948772 | 0 | - | - | - | - | - | - | - | - | - | - | - | - | - | - | - | - | - | - | - | - | - | - | - | - | - | - | - | - | - |
| LETM1 | OID3101 | Neurolog | LETM1 | O95202 | 4 | 1811479 | 1856156 | 0 | - | - | - | - | - | - | - | - | - | - | - | - | - | - | - | - | - | - | - | - | - | - | - | - | - | - | - | - | - |
| LRP2BP | OID3090 | Neurolog | LRP2BP | Q9P2M1 | 4 | 185363872 | 185395924 | 0 | - | - | - | - | - | - | - | - | - | - | - | - | - | - | - | - | - | - | - | - | - | - | - | - | - | - | - | - | - |
| MAEA | OID2120 | Oncology | MAEA | Q7L5Y9 | 4 | 1289887 | 1340147 | 0 | - | - | - | - | - | - | - | - | - | - | - | - | - | - | - | - | - | - | - | - | - | - | - | - | - | - | - | - | - |
| METAP1 | OID2081 | Neurolog | METAP1 | P53582 | 4 | 98995659 | 99062809 | 0 | - | - | - | - | - | - | - | - | - | - | - | - | - | - | - | - | - | - | - | - | - | - | - | - | - | - | - | - | - |
| MFAP3L | OID3093 | Neurolog | MFAP3L | O75121 | 4 | 169986597 | 170033031 | 0 | - | - | - | - | - | - | - | - | - | - | - | - | - | - | - | - | - | - | - | - | - | - | - | - | - | - | - | - | - |
| PALLD | OID3044 | Inflamma | PALLD | Q8WX93 | 4 | 168497052 | 168928457 | 0 | - | - | - | - | - | - | - | - | - | - | - | - | - | - | - | - | - | - | - | - | - | - | - | - | - | - | - | - | - |
| PDCL2 | OID3116 | Oncology | PDCL2 | Q8N4E4 | 4 | 55556519 | 55592245 | 0 | - | - | - | - | - | - | - | - | - | - | - | - | - | - | - | - | - | - | - | - | - | - | - | - | - | - | - | - | - |
| PDLIM5 | OID3106 | Neurolog | PDLIM5 | Q96HC4 | 4 | 94451857 | 94668227 | 0 | - | - | - | - | - | - | - | - | - | - | - | - | - | - | - | - | - | - | - | - | - | - | - | - | - | - | - | - | - |
| PGM2 | OID3090 | Neurolog | PGM2 | Q96G03 | 4 | 37826660 | 37862937 | 0 | - | - | - | - | - | - | - | - | - | - | - | - | - | - | - | - | - | - | - | - | - | - | - | - | - | - | - | - | - |
| PPBP | OID3076 | Inflamma | PPBP | P02775 | 4 | 73986439 | 73988190 | 0 | - | - | - | - | - | - | - | - | - | - | - | - | - | - | - | - | - | - | - | - | - | - | - | - | - | - | - | - | - |
| REST | OID3124 | Oncology | REST | Q13127 | 4 | 56907876 | 56966808 | 0 | - | - | - | - | - | - | - | - | - | - | - | - | - | - | - | - | - | - | - | - | - | - | - | - | - | - | - | - | - |
| RNF4 | OID3132 | Oncology | RNF4 | P78317 | 4 | 2462220 | 2515857 | 0 | - | - | - | - | - | - | - | - | - | - | - | - | - | - | - | - | - | - | - | - | - | - | - | - | - | - | - | - | - |
| SMAD1 | OID2118 | Oncology | SMAD1 | Q15797 | 4 | 145481194 | 145559176 | 0 | - | - | - | - | - | - | - | - | - | - | - | - | - | - | - | - | - | - | - | - | - | - | - | - | - | - | - | - | - |
| TDO2 | OID3091 | Neurolog | TDO2 | P48775 | 4 | 155854738 | 155920406 | 0 | - | - | - | - | - | - | - | - | - | - | - | - | - | - | - | - | - | - | - | - | - | - | - | - | - | - | - | - | - |
| TET2 | OID3126 | Oncology | TET2 | Q6N021 | 4 | 105145875 | 105279816 | 0 | - | - | - | - | - | - | - | - | - | - | - | - | - | - | - | - | - | - | - | - | - | - | - | - | - | - | - | - | - |
| TLR2 | OID3129 | Oncology | TLR2 | O60603 | 4 | 153684070 | 153706260 | 0 | - | - | - | - | - | - | - | - | - | - | - | - | - | - | - | - | - | - | - | - | - | - | - | - | - | - | - | - | - |
| TMPRSS | OID3085 | Neurolog | TMPRSS | Q86T26 | 4 | 68226653 | 68245694 | 0 | - | - | - | - | - | - | - | - | - | - | - | - | - | - | - | - | - | - | - | - | - | - | - | - | - | - | - | - | - |
| TXK | OID3126 | Oncology | TXK | P42681 | 4 | 48066393 | 48134250 | 0 | - | - | - | - | - | - | - | - | - | - | - | - | - | - | - | - | - | - | - | - | - | - | - | - | - | - | - | - | - |
| UGDH | OID3008 | Cardiom | UGDH | O60701 | 4 | 39498755 | 39528311 | 0 | - | - | - | - | - | - | - | - | - | - | - | - | - | - | - | - | - | - | - | - | - | - | - | - | - | - | - | - | - |
| USO1 | OID2136 | Oncology | USO1 | O60763 | 4 | 75724577 | 75814286 | 0 | - | - | - | - | - | - | - | - | - | - | - | - | - | - | - | - | - | - | - | - | - | - | - | - | - | - | - | - | - |
| ADGRV1 | OID3087 | Neurolog | ADGRV1 | Q8WXG9 | 5 | 90529344 | 91164437 | 0 | - | - | - | - | - | - | - | - | - | - | - | - | - | - | - | - | - | - | - | - | - | - | - | - | - | - | - | - | - |
| ANKRA2 | OID3117 | Oncology | ANKRA2 | Q9H9E1 | 5 | 73552190 | 73565667 | 0 | - | - | - | - | - | - | - | - | - | - | - | - | - | - | - | - | - | - | - | - | - | - | - | - | - | - | - | - | - |
| BHMT2 | OID3005 | Cardiom | BHMT2 | Q9H2M3 | 5 | 79069767 | 79090069 | 0 | - | - | - | - | - | - | - | - | - | - | - | - | - | - | - | - | - | - | - | - | - | - | - | - | - | - | - | - | - |
| CAMLG | OID3095 | Neurolog | CAMLG | P49069 | 5 | 134738495 | 134752157 | 0 | - | - | - | - | - | - | - | - | - | - | - | - | - | - | - | - | - | - | - | - | - | - | - | - | - | - | - | - | - |
| CCT5 | OID2120 | Oncology | CCT5 | P48643 | 5 | 10249929 | 10266389 | 0 | - | - | - | - | - | - | - | - | - | - | - | - | - | - | - | - | - | - | - | - | - | - | - | - | - | - | - | - | - |
| CLINT1 | OID3118 | Oncology | CLINT1 | Q14677 | 5 | 157785743 | 157859145 | 0 | - | - | - | - | - | - | - | - | - | - | - | - | - | - | - | - | - | - | - | - | - | - | - | - | - | - | - | - | - |
| CPLX2 | OID3118 | Oncology | CPLX2 | Q6PUV4 | 5 | 175796310 | 175884021 | 0 | - | - | - | - | - | - | - | - | - | - | - | - | - | - | - | - | - | - | - | - | - | - | - | - | - | - | - | - | - |
| CTNNA1 | OID3080 | Neurolog | CTNNA1 | P35221 | 5 | 138610967 | 138935034 | 0 | - | - | - | - | - | - | - | - | - | - | - | - | - | - | - | - | - | - | - | - | - | - | - | - | - | - | - | - | - |
| DAB2 | OID2142 | Oncology | DAB2 | P98082 | 5 | 39371675 | 39462300 | 0 | - | - | - | - | - | - | - | - | - | - | - | - | - | - | - | - | - | - | - | - | - | - | - | - | - | - | - | - | - |
| DDX4 | OID3043 | Inflamma | DDX4 | Q9NQI0 | 5 | 55738017 | 55817157 | 0 | - | - | - | - | - | - | - | - | - | - | - | - | - | - | - | - | - | - | - | - | - | - | - | - | - | - | - | - | - |
| ECSCR | OID3083 | Neurolog | ECSCR | Q19T08 | 5 | 139448560 | 139462743 | 0 | - | - | - | - | - | - | - | - | - | - | - | - | - | - | - | - | - | - | - | - | - | - | - | - | - | - | - | - | - |
| ERBIN | OID2132 | Oncology | ERBIN | Q96RT1 | 5 | 65926556 | 66082546 | 0 | - | - | - | - | - | - | - | - | - | - | - | - | - | - | - | - | - | - | - | - | - | - | - | - | - | - | - | - | - |
| FYB1 | OID2114 | Neurolog | FYB1 | O15117 | 5 | 39105252 | 39274528 | 0 | - | - | - | - | - | - | - | - | - | - | - | - | - | - | - | - | - | - | - | - | - | - | - | - | - | - | - | - | - |
| HARS1 | OID2108 | Neurolog | HARS1 | P12081 | 5 | 140673035 | 140691537 | 0 | - | - | - | - | - | - | - | - | - | - | - | - | - | - | - | - | - | - | - | - | - | - | - | - | - | - | - | - | - |
| HMGCS1 | OID3015 | Cardiom | HMGCS1 | Q01581 | 5 | 43287470 | 43313512 | 0 | - | - | - | - | - | - | - | - | - | - | - | - | - | - | - | - | - | - | - | - | - | - | - | - | - | - | - | - | - |
| HMMR | OID3117 | Oncology | HMMR | O75330 | 5 | 163460203 | 163491941 | 0 | - | - | - | - | - | - | - | - | - | - | - | - | - | - | - | - | - | - | - | - | - | - | - | - | - | - | - | - | - |
| HTR1A | OID3082 | Neurolog | HTR1A | P08908 | 5 | 63957874 | 63962507 | 0 | - | - | - | - | - | - | - | - | - | - | - | - | - | - | - | - | - | - | - | - | - | - | - | - | - | - | - | - | - |
| IL13 | OID2043 | Inflamma | IL13 | P35225 | 5 | 132656263 | 132661110 | 0 | - | - | - | - | - | - | - | - | - | - | - | - | - | - | - | - | - | - | - | - | - | - | - | - | - | - | - | - | - |
| IL3 | OID3122 | Oncology | IL3 | P08700 | 5 | 132060655 | 132063204 | 0 | - | - | - | - | - | - | - | - | - | - | - | - | - | - | - | - | - | - | - | - | - | - | - | - | - | - | - | - | - |
| IL4 | OID2042 | Inflamma | IL4 | P05112 | 5 | 132673986 | 132682678 | 0 | - | - | - | - | - | - | - | - | - | - | - | - | - | - | - | - | - | - | - | - | - | - | - | - | - | - | - | - | - |
| IL5 | OID2047 | Inflamma | IL5 | P05113 | 5 | 132541445 | 132556838 | 0 | - | - | - | - | - | - | - | - | - | - | - | - | - | - | - | - | - | - | - | - | - | - | - | - | - | - | - | - | - |
| IL9 | OID3129 | Oncology | IL9 | P15248 | 5 | 135892246 | 135895841 | 0 | - | - | - | - | - | - | - | - | - | - | - | - | - | - | - | - | - | - | - | - | - | - | - | - | - | - | - | - | - |
| LARP1 | OID3137 | Oncology | LARP1 | Q6PKG0 | 5 | 154682986 | 154817605 | 0 | - | - | - | - | - | - | - | - | - | - | - | - | - | - | - | - | - | - | - | - | - | - | - | - | - | - | - | - | - |
| MFAP3 | OID2005 | Cardiom | MFAP3 | P55082 | 5 | 154038959 | 154220478 | 0 | - | - | - | - | - | - | - | - | - | - | - | - | - | - | - | - | - | - | - | - | - | - | - | - | - | - | - | - | - |
| NDST1 | OID3136 | Oncology | NDST1 | P52848 | 5 | 150485818 | 150558211 | 0 | - | - | - | - | - | - | - | - | - | - | - | - | - | - | - | - | - | - | - | - | - | - | - | - | - | - | - | - | - |
| NDUFS6 | OID2122 | Oncology | NDUFS6 | O75380 | 5 | 1801407 | 1816048 | 0 | - | - | - | - | - | - | - | - | - | - | - | - | - | - | - | - | - | - | - | - | - | - | - | - | - | - | - | - | - |
| PDE4D | OID3008 | Cardiom | PDE4D | Q08499 | 5 | 58969038 | 60522120 | 0 | - | - | - | - | - | - | - | - | - | - | - | - | - | - | - | - | - | - | - | - | - | - | - | - | - | - | - | - | - |
| PTTG1 | OID3132 | Oncology | PTTG1 | O95997 | 5 | 160421855 | 160428739 | 0 | - | - | - | - | - | - | - | - | - | - | - | - | - | - | - | - | - | - | - | - | - | - | - | - | - | - | - | - | - |
| RASA1 | OID2082 | Neurolog | RASA1 | P20936 | 5 | 87267883 | 87391931 | 0 | - | - | - | - | - | - | - | - | - | - | - | - | - | - | - | - | - | - | - | - | - | - | - | - | - | - | - | - | - |
| RICTOR | OID3043 | Inflamma | RICTOR | Q6R327 | 5 | 38937920 | 39074399 | 0 | - | - | - | - | - | - | - | - | - | - | - | - | - | - | - | - | - | - | - | - | - | - | - | - | - | - | - | - | - |
| SLC12A2 | OID3007 | Cardiom | SLC12A2 | P55011 | 5 | 128083766 | 128189677 | 0 | - | - | - | - | - | - | - | - | - | - | - | - | - | - | - | - | - | - | - | - | - | - | - | - | - | - | - | - | - |
| SMAD5 | OID2135 | Oncology | SMAD5 | Q99717 | 5 | 136132845 | 136188747 | 0 | - | - | - | - | - | - | - | - | - | - | - | - | - | - | - | - | - | - | - | - | - | - | - | - | - | - | - | - | - |
| SOWAHA | OID3088 | Neurolog | SOWAHA | Q2M3V2 | 5 | 132813302 | 132816786 | 0 | - | - | - | - | - | - | - | - | - | - | - | - | - | - | - | - | - | - | - | - | - | - | - | - | - | - | - | - | - |
| TARS1 | OID3134 | Oncology | TARS1 | P26639 | 5 | 33440696 | 33468091 | 0 | - | - | - | - | - | - | - | - | - | - | - | - | - | - | - | - | - | - | - | - | - | - | - | - | - | - | - | - | - |
| TCOF1 | OID3031 | Cardiom | TCOF1 | Q13428 | 5 | 150357629 | 150400308 | 0 | - | - | - | - | - | - | - | - | - | - | - | - | - | - | - | - | - | - | - | - | - | - | - | - | - | - | - | - | - |
| TNPO1 | OID3133 | Oncology | TNPO1 | Q92973 | 5 | 72816312 | 72916733 | 0 | - | - | - | - | - | - | - | - | - | - | - | - | - | - | - | - | - | - | - | - | - | - | - | - | - | - | - | - | - |
| TSLP | OID2005 | Cardiom | TSLP | Q969D9 | 5 | 111070062 | 111078026 | 0 | - | - | - | - | - | - | - | - | - | - | - | - | - | - | - | - | - | - | - | - | - | - | - | - | - | - | - | - | - |
| UBE2B | OID3136 | Oncology | UBE2B | P63146 | 5 | 134371184 | 134392108 | 0 | - | - | - | - | - | - | - | - | - | - | - | - | - | - | - | - | - | - | - | - | - | - | - | - | - | - | - | - | - |
| ADGRF5 | OID3034 | Cardiom | ADGRF5 | Q8IZF2 | 6 | 46852522 | 46954943 | 0 | - | - | - | - | - | - | - | - | - | - | - | - | - | - | - | - | - | - | - | - | - | - | - | - | - | - | - | - | - |
| BAG6 | OID2011 | Cardiom | BAG6 | P46379 | 6 | 31639028 | 31652705 | 0 | - | - | - | - | - | - | - | - | - | - | - | - | - | - | - | - | - | - | - | - | - | - | - | - | - | - | - | - | - |
| BRD2 | OID3082 | Neurolog | BRD2 | P25440 | 6 | 32968594 | 32981505 | 0 | - | - | - | - | - | - | - | - | - | - | - | - | - | - | - | - | - | - | - | - | - | - | - | - | - | - | - | - | - |
| CCDC28 | OID3097 | Neurolog | CCDC28 | Q8IWP9 | 6 | 138773769 | 138793319 | 0 | - | - | - | - | - | - | - | - | - | - | - | - | - | - | - | - | - | - | - | - | - | - | - | - | - | - | - | - | - |
| ESR1 | OID3043 | Inflamma | ESR1 | P03372 | 6 | 151656691 | 152129619 | 0 | - | - | - | - | - | - | - | - | - | - | - | - | - | - | - | - | - | - | - | - | - | - | - | - | - | - | - | - | - |
| FOXO3 | OID2127 | Oncology | FOXO3 | O43524 | 6 | 108559835 | 108684774 | 0 | - | - | - | - | - | - | - | - | - | - | - | - | - | - | - | - | - | - | - | - | - | - | - | - | - | - | - | - | - |
| GLP1R | OID3080 | Neurolog | GLP1R | P43220 | 6 | 39048781 | 39091303 | 0 | - | - | - | - | - | - | - | - | - | - | - | - | - | - | - | - | - | - | - | - | - | - | - | - | - | - | - | - | - |
| GOPC | OID2055 | Inflamma | GOPC | Q9HD26 | 6 | 117560269 | 117602542 | 0 | - | - | - | - | - | - | - | - | - | - | - | - | - | - | - | - | - | - | - | - | - | - | - | - | - | - | - | - | - |
| GTPBP2 | OID3094 | Neurolog | GTPBP2 | Q9BX10 | 6 | 43605316 | 43629264 | 0 | - | - | - | - | - | - | - | - | - | - | - | - | - | - | - | - | - | - | - | - | - | - | - | - | - | - | - | - | - |
| HTR1B | OID3086 | Neurolog | HTR1B | P28222 | 6 | 77460924 | 77463491 | 0 | - | - | - | - | - | - | - | - | - | - | - | - | - | - | - | - | - | - | - | - | - | - | - | - | - | - | - | - | - |
| ID4 | OID3128 | Oncology | ID4 | P47928 | 6 | 19837370 | 19842197 | 0 | - | - | - | - | - | - | - | - | - | - | - | - | - | - | - | - | - | - | - | - | - | - | - | - | - | - | - | - | - |
| IL17A | OID2046 | Inflamma | IL17A | Q16552 | 6 | 52186375 | 52190638 | 0 | - | - | - | - | - | - | - | - | - | - | - | - | - | - | - | - | - | - | - | - | - | - | - | - | - | - | - | - | - |
| IL20RA | OID2042 | Inflamma | IL20RA | Q9UHF4 | 6 | 136999971 | 137045180 | 0 | - | - | - | - | - | - | - | - | - | - | - | - | - | - | - | - | - | - | - | - | - | - | - | - | - | - | - | - | - |
| IMPG1 | OID3081 | Neurolog | IMPG1 | Q17R60 | 6 | 75921114 | 76072678 | 0 | - | - | - | - | - | - | - | - | - | - | - | - | - | - | - | - | - | - | - | - | - | - | - | - | - | - | - | - | - |
| KHDC3L | OID3124 | Oncology | KHDC3L | Q587J8 | 6 | 73362658 | 73364171 | 0 | - | - | - | - | - | - | - | - | - | - | - | - | - | - | - | - | - | - | - | - | - | - | - | - | - | - | - | - | - |
| LRFN2 | OID3089 | Neurolog | LRFN2 | Q9ULH4 | 6 | 40391591 | 40587364 | 0 | - | - | - | - | - | - | - | - | - | - | - | - | - | - | - | - | - | - | - | - | - | - | - | - | - | - | - | - | - |
| MAP3K5 | OID2125 | Oncology | MAP3K5 | Q99683 | 6 | 136557046 | 136792477 | 0 | - | - | - | - | - | - | - | - | - | - | - | - | - | - | - | - | - | - | - | - | - | - | - | - | - | - | - | - | - |
| MPIG6B | OID2077 | Inflamma | MPIG6B | O95866 | 6 | 31718594 | 31726714 | 0 | - | - | - | - | - | - | - | - | - | - | - | - | - | - | - | - | - | - | - | - | - | - | - | - | - | - | - | - | - |
| MUCL3 | OID3117 | Oncology | MUCL3 | Q3MIW9 | 6 | 30934523 | 30954221 | 0 | - | - | - | - | - | - | - | - | - | - | - | - | - | - | - | - | - | - | - | - | - | - | - | - | - | - | - | - | - |
| MYO6 | OID3098 | Neurolog | MYO6 | Q9UM54 | 6 | 75749201 | 75919537 | 0 | - | - | - | - | - | - | - | - | - | - | - | - | - | - | - | - | - | - | - | - | - | - | - | - | - | - | - | - | - |
| NEDD9 | OID3050 | Inflamma | NEDD9 | Q14511 | 6 | 11183298 | 11382348 | 0 | - | - | - | - | - | - | - | - | - | - | - | - | - | - | - | - | - | - | - | - | - | - | - | - | - | - | - | - | - |
| NFYA | OID3013 | Cardiom | NFYA | P23511 | 6 | 41072974 | 41102403 | 0 | - | - | - | - | - | - | - | - | - | - | - | - | - | - | - | - | - | - | - | - | - | - | - | - | - | - | - | - | - |
| NRN1 | OID3098 | Neurolog | NRN1 | Q9NPD7 | 6 | 5997999 | 6007605 | 0 | - | - | - | - | - | - | - | - | - | - | - | - | - | - | - | - | - | - | - | - | - | - | - | - | - | - | - | - | - |
| PFDN6 | OID3093 | Neurolog | PFDN6 | O15212 | 6 | 33289302 | 33298401 | 0 | - | - | - | - | - | - | - | - | - | - | - | - | - | - | - | - | - | - | - | - | - | - | - | - | - | - | - | - | - |
| PRL | OID2103 | Neurolog | PRL | P01236 | 6 | 22287244 | 22302826 | 0 | - | - | - | - | - | - | - | - | - | - | - | - | - | - | - | - | - | - | - | - | - | - | - | - | - | - | - | - | - |
| REPS1 | OID3049 | Inflamma | REPS1 | Q96D71 | 6 | 138903493 | 138988261 | 0 | - | - | - | - | - | - | - | - | - | - | - | - | - | - | - | - | - | - | - | - | - | - | - | - | - | - | - | - | - |
| RGL2 | OID3129 | Oncology | RGL2 | O15211 | 6 | 33291654 | 33298942 | 0 | - | - | - | - | - | - | - | - | - | - | - | - | - | - | - | - | - | - | - | - | - | - | - | - | - | - | - | - | - |
| RNF5 | OID3021 | Cardiom | RNF5 | Q99942 | 6 | 32178405 | 32180793 | 0 | - | - | - | - | - | - | - | - | - | - | - | - | - | - | - | - | - | - | - | - | - | - | - | - | - | - | - | - | - |
| RPS10 | OID3007 | Cardiom | RPS10 | P46783 | 6 | 34417454 | 34426069 | 0 | - | - | - | - | - | - | - | - | - | - | - | - | - | - | - | - | - | - | - | - | - | - | - | - | - | - | - | - | - |
| SH3BGRL | OID3086 | Neurolog | SH3BGR | Q9UJC5 | 6 | 79631329 | 79703655 | 0 | - | - | - | - | - | - | - | - | - | - | - | - | - | - | - | - | - | - | - | - | - | - | - | - | - | - | - | - | - |
| SIRT5 | OID2082 | Neurolog | SIRT5 | Q9NXA8 | 6 | 13574227 | 13615158 | 0 | - | - | - | - | - | - | - | - | - | - | - | - | - | - | - | - | - | - | - | - | - | - | - | - | - | - | - | - | - |
| SKIV2L | OID3005 | Cardiom | SKIV2L | Q15477 | 6 | 31959116 | 31969818 | 0 | - | - | - | - | - | - | - | - | - | - | - | - | - | - | - | - | - | - | - | - | - | - | - | - | - | - | - | - | - |
| SLC44A4 | OID3089 | Neurolog | SLC44A4 | Q53GD3 | 6 | 31863192 | 31879046 | 0 | - | - | - | - | - | - | - | - | - | - | - | - | - | - | - | - | - | - | - | - | - | - | - | - | - | - | - | - | - |
| SYNGAP | OID3079 | Neurolog | SYNGAP | Q96PV0 | 6 | 33419661 | 33453689 | 0 | - | - | - | - | - | - | - | - | - | - | - | - | - | - | - | - | - | - | - | - | - | - | - | - | - | - | - | - | - |
| TAP1 | OID3128 | Oncology | TAP1 | Q03518 | 6 | 32845209 | 32853816 | 0 | - | - | - | - | - | - | - | - | - | - | - | - | - | - | - | - | - | - | - | - | - | - | - | - | - | - | - | - | - |
| TCP11 | OID3126 | Oncology | TCP11 | Q8WWU | 6 | 35118071 | 35148610 | 0 | - | - | - | - | - | - | - | - | - | - | - | - | - | - | - | - | - | - | - | - | - | - | - | - | - | - | - | - | - |
| TFAP2A | OID3084 | Neurolog | TFAP2A | P05549 | 6 | 10393186 | 10419659 | 0 | - | - | - | - | - | - | - | - | - | - | - | - | - | - | - | - | - | - | - | - | - | - | - | - | - | - | - | - | - |
| TRAF3IP | OID3005 | Cardiom | TRAF3IP | O43734 | 6 | 111555381 | 111606906 | 0 | - | - | - | - | - | - | - | - | - | - | - | - | - | - | - | - | - | - | - | - | - | - | - | - | - | - | - | - | - |
| TREML1 | OID3068 | Inflamma | TREML1 | Q86YW5 | 6 | 41149337 | 41154347 | 0 | - | - | - | - | - | - | - | - | - | - | - | - | - | - | - | - | - | - | - | - | - | - | - | - | - | - | - | - | - |
| TRIM26 | OID3120 | Oncology | TRIM26 | Q12899 | 6 | 30184455 | 30213427 | 0 | - | - | - | - | - | - | - | - | - | - | - | - | - | - | - | - | - | - | - | - | - | - | - | - | - | - | - | - | - |
| VTA1 | OID2101 | Neurolog | VTA1 | Q9NP79 | 6 | 142147162 | 142224685 | 0 | - | - | - | - | - | - | - | - | - | - | - | - | - | - | - | - | - | - | - | - | - | - | - | - | - | - | - | - | - |
| WDR46 | OID3138 | Oncology | WDR46 | O15213 | 6 | 33279108 | 33289247 | 0 | - | - | - | - | - | - | - | - | - | - | - | - | - | - | - | - | - | - | - | - | - | - | - | - | - | - | - | - | - |
| ADCYAP | OID2118 | Oncology | ADCYAP | P41586 | 7 | 31052308 | 31111479 | 0 | - | - | - | - | - | - | - | - | - | - | - | - | - | - | - | - | - | - | - | - | - | - | - | - | - | - | - | - | - |
| AGR3 | OID2124 | Oncology | AGR3 | Q8TD06 | 7 | 16859412 | 16881987 | 0 | - | - | - | - | - | - | - | - | - | - | - | - | - | - | - | - | - | - | - | - | - | - | - | - | - | - | - | - | - |
| AKR1B10 | OID3095 | Neurolog | AKR1B10 | O60218 | 7 | 134527567 | 134541412 | 0 | - | - | - | - | - | - | - | - | - | - | - | - | - | - | - | - | - | - | - | - | - | - | - | - | - | - | - | - | - |
| ATP6V1F | OID2084 | Neurolog | ATP6V1F | Q16864 | 7 | 128862856 | 128865847 | 0 | - | - | - | - | - | - | - | - | - | - | - | - | - | - | - | - | - | - | - | - | - | - | - | - | - | - | - | - | - |
| BCL7B | OID3142 | Oncology | BCL7B | Q9BQE9 | 7 | 73536356 | 73557690 | 0 | - | - | - | - | - | - | - | - | - | - | - | - | - | - | - | - | - | - | - | - | - | - | - | - | - | - | - | - | - |
| CASP2 | OID2055 | Inflamma | CASP2 | P42575 | 7 | 143288215 | 143307696 | 0 | - | - | - | - | - | - | - | - | - | - | - | - | - | - | - | - | - | - | - | - | - | - | - | - | - | - | - | - | - |
| CLEC2L | OID3088 | Neurolog | CLEC2L | P0C7M8 | 7 | 139523685 | 139544985 | 0 | - | - | - | - | - | - | - | - | - | - | - | - | - | - | - | - | - | - | - | - | - | - | - | - | - | - | - | - | - |
| CYTH3 | OID3120 | Oncology | CYTH3 | O43739 | 7 | 6161776 | 6272644 | 0 | - | - | - | - | - | - | - | - | - | - | - | - | - | - | - | - | - | - | - | - | - | - | - | - | - | - | - | - | - |
| DTX2 | OID3088 | Neurolog | DTX2 | Q86UW9 | 7 | 76461676 | 76505995 | 0 | - | - | - | - | - | - | - | - | - | - | - | - | - | - | - | - | - | - | - | - | - | - | - | - | - | - | - | - | - |
| FKBP14 | OID3106 | Neurolog | FKBP14 | Q9NWM8 | 7 | 30010587 | 30026702 | 0 | - | - | - | - | - | - | - | - | - | - | - | - | - | - | - | - | - | - | - | - | - | - | - | - | - | - | - | - | - |
| GCC1 | OID3114 | Neurolog | GCC1 | Q96CN9 | 7 | 127580628 | 127593611 | 0 | - | - | - | - | - | - | - | - | - | - | - | - | - | - | - | - | - | - | - | - | - | - | - | - | - | - | - | - | - |
| GHRHR | OID2084 | Neurolog | GHRHR | Q02643 | 7 | 30938669 | 30993254 | 0 | - | - | - | - | - | - | - | - | - | - | - | - | - | - | - | - | - | - | - | - | - | - | - | - | - | - | - | - | - |
| GNGT1 | OID3092 | Neurolog | GNGT1 | P63211 | 7 | 93591573 | 93911265 | 0 | - | - | - | - | - | - | - | - | - | - | - | - | - | - | - | - | - | - | - | - | - | - | - | - | - | - | - | - | - |
| GSAP | OID2117 | Oncology | GSAP | A4D1B5 | 7 | 77310751 | 77416349 | 0 | - | - | - | - | - | - | - | - | - | - | - | - | - | - | - | - | - | - | - | - | - | - | - | - | - | - | - | - | - |
| GTF2IRD | OID3126 | Oncology | GTF2IRD | Q9UHL9 | 7 | 74453790 | 74602605 | 0 | - | - | - | - | - | - | - | - | - | - | - | - | - | - | - | - | - | - | - | - | - | - | - | - | - | - | - | - | - |
| HDAC9 | OID3083 | Neurolog | HDAC9 | Q9UKV0 | 7 | 18086949 | 19002416 | 0 | - | - | - | - | - | - | - | - | - | - | - | - | - | - | - | - | - | - | - | - | - | - | - | - | - | - | - | - | - |
| HEPACA | OID3115 | Neurolog | HEPACA | A8MVW5 | 7 | 93188534 | 93226469 | 0 | - | - | - | - | - | - | - | - | - | - | - | - | - | - | - | - | - | - | - | - | - | - | - | - | - | - | - | - | - |
| ICA1 | OID2048 | Inflamma | ICA1 | Q05084 | 7 | 8113184 | 8262687 | 0 | - | - | - | - | - | - | - | - | - | - | - | - | - | - | - | - | - | - | - | - | - | - | - | - | - | - | - | - | - |
| IGF2BP3 | OID3011 | Cardiom | IGF2BP3 | O00425 | 7 | 23310209 | 23470491 | 0 | - | - | - | - | - | - | - | - | - | - | - | - | - | - | - | - | - | - | - | - | - | - | - | - | - | - | - | - | - |
| IL6 | OID2010 | Cardiom | IL6 | P05231 | 7 | 22725884 | 22732002 | 0 | - | - | - | - | - | - | - | - | - | - | - | - | - | - | - | - | - | - | - | - | - | - | - | - | - | - | - | - | - |
| KCNH2 | OID3089 | Neurolog | KCNH2 | Q12809 | 7 | 150944961 | 150978321 | 0 | - | - | - | - | - | - | - | - | - | - | - | - | - | - | - | - | - | - | - | - | - | - | - | - | - | - | - | - | - |
| KIAA154 | OID3130 | Oncology | KIAA154 | Q9HCM3 | 7 | 138831381 | 138981389 | 0 | - | - | - | - | - | - | - | - | - | - | - | - | - | - | - | - | - | - | - | - | - | - | - | - | - | - | - | - | - |
| LEP | OID2018 | Cardiom | LEP | P41159 | 7 | 128241278 | 128257629 | 0 | - | - | - | - | - | - | - | - | - | - | - | - | - | - | - | - | - | - | - | - | - | - | - | - | - | - | - | - | - |
| LMOD2 | OID3086 | Neurolog | LMOD2 | Q6P5Q4 | 7 | 123655866 | 123664290 | 0 | - | - | - | - | - | - | - | - | - | - | - | - | - | - | - | - | - | - | - | - | - | - | - | - | - | - | - | - | - |
| LSM8 | OID3126 | Oncology | LSM8 | O95777 | 7 | 118184144 | 118204035 | 0 | - | - | - | - | - | - | - | - | - | - | - | - | - | - | - | - | - | - | - | - | - | - | - | - | - | - | - | - | - |
| MTPN | OID2008 | Cardiom | MTPN | P58546 | 7 | 135926760 | 135977359 | 0 | - | - | - | - | - | - | - | - | - | - | - | - | - | - | - | - | - | - | - | - | - | - | - | - | - | - | - | - | - |
| NDUFA5 | OID3052 | Inflamma | NDUFA5 | Q16718 | 7 | 123536997 | 123557904 | 0 | - | - | - | - | - | - | - | - | - | - | - | - | - | - | - | - | - | - | - | - | - | - | - | - | - | - | - | - | - |
| NXPH1 | OID2084 | Neurolog | NXPH1 | P58417 | 7 | 8433609 | 8752961 | 0 | - | - | - | - | - | - | - | - | - | - | - | - | - | - | - | - | - | - | - | - | - | - | - | - | - | - | - | - | - |
| PDAP1 | OID3032 | Cardiom | PDAP1 | Q13442 | 7 | 99392048 | 99408597 | 0 | - | - | - | - | - | - | - | - | - | - | - | - | - | - | - | - | - | - | - | - | - | - | - | - | - | - | - | - | - |
| PDE1C | OID3120 | Oncology | PDE1C | Q14123 | 7 | 31751179 | 32428131 | 0 | - | - | - | - | - | - | - | - | - | - | - | - | - | - | - | - | - | - | - | - | - | - | - | - | - | - | - | - | - |
| PSMG3 | OID2054 | Inflamma | PSMG3 | Q9BT73 | 7 | 1567332 | 1571005 | 0 | - | - | - | - | - | - | - | - | - | - | - | - | - | - | - | - | - | - | - | - | - | - | - | - | - | - | - | - | - |
| SCIN | OID3085 | Neurolog | SCIN | Q9Y6U3 | 7 | 12570577 | 12660182 | 0 | - | - | - | - | - | - | - | - | - | - | - | - | - | - | - | - | - | - | - | - | - | - | - | - | - | - | - | - | - |
| SEPTIN7 | OID3128 | Oncology | SEPTIN7 | Q16181 | 7 | 35800932 | 35907110 | 0 | - | - | - | - | - | - | - | - | - | - | - | - | - | - | - | - | - | - | - | - | - | - | - | - | - | - | - | - | - |
| SKAP2 | OID2076 | Inflamma | SKAP2 | O75563 | 7 | 26667068 | 26995239 | 0 | - | - | - | - | - | - | - | - | - | - | - | - | - | - | - | - | - | - | - | - | - | - | - | - | - | - | - | - | - |
| SLC13A1 | OID3138 | Oncology | SLC13A1 | Q9BZW2 | 7 | 123113531 | 123199972 | 0 | - | - | - | - | - | - | - | - | - | - | - | - | - | - | - | - | - | - | - | - | - | - | - | - | - | - | - | - | - |
| SRPK2 | OID2050 | Inflamma | SRPK2 | P78362 | 7 | 105110704 | 105399308 | 0 | - | - | - | - | - | - | - | - | - | - | - | - | - | - | - | - | - | - | - | - | - | - | - | - | - | - | - | - | - |
| SSBP1 | OID3046 | Inflamma | SSBP1 | Q04837 | 7 | 141738334 | 141787922 | 0 | - | - | - | - | - | - | - | - | - | - | - | - | - | - | - | - | - | - | - | - | - | - | - | - | - | - | - | - | - |
| STEAP4 | OID3084 | Neurolog | STEAP4 | Q687X5 | 7 | 88270892 | 88306894 | 0 | - | - | - | - | - | - | - | - | - | - | - | - | - | - | - | - | - | - | - | - | - | - | - | - | - | - | - | - | - |
| TAX1BP1 | OID3107 | Neurolog | TAX1BP1 | Q86VP1 | 7 | 27739331 | 27844564 | 0 | - | - | - | - | - | - | - | - | - | - | - | - | - | - | - | - | - | - | - | - | - | - | - | - | - | - | - | - | - |
| TMED4 | OID3015 | Cardiom | TMED4 | Q7Z7H5 | 7 | 44577894 | 44582287 | 0 | - | - | - | - | - | - | - | - | - | - | - | - | - | - | - | - | - | - | - | - | - | - | - | - | - | - | - | - | - |
| TRIM24 | OID3127 | Oncology | TRIM24 | O15164 | 7 | 138460259 | 138589996 | 0 | - | - | - | - | - | - | - | - | - | - | - | - | - | - | - | - | - | - | - | - | - | - | - | - | - | - | - | - | - |
| UPK3BL1 | OID3128 | Oncology | UPK3BL1 | B0FP48 | 7 | 102637025 | 102642791 | 0 | - | - | - | - | - | - | - | - | - | - | - | - | - | - | - | - | - | - | - | - | - | - | - | - | - | - | - | - | - |
| WASL | OID3046 | Inflamma | WASL | O00401 | 7 | 123681943 | 123749003 | 0 | - | - | - | - | - | - | - | - | - | - | - | - | - | - | - | - | - | - | - | - | - | - | - | - | - | - | - | - | - |
| BAG4 | OID3047 | Inflamma | BAG4 | O95429 | 8 | 38176533 | 38213301 | 0 | - | - | - | - | - | - | - | - | - | - | - | - | - | - | - | - | - | - | - | - | - | - | - | - | - | - | - | - | - |
| CA8 | OID3048 | Inflamma | CA8 | P35219 | 8 | 60185412 | 60281400 | 0 | - | - | - | - | - | - | - | - | - | - | - | - | - | - | - | - | - | - | - | - | - | - | - | - | - | - | - | - | - |
| CCAR2 | OID3107 | Neurolog | CCAR2 | Q8N163 | 8 | 22604757 | 22620964 | 0 | - | - | - | - | - | - | - | - | - | - | - | - | - | - | - | - | - | - | - | - | - | - | - | - | - | - | - | - | - |
| CNGB3 | OID3080 | Neurolog | CNGB3 | Q9NQW8 | 8 | 86553977 | 86743675 | 0 | - | - | - | - | - | - | - | - | - | - | - | - | - | - | - | - | - | - | - | - | - | - | - | - | - | - | - | - | - |
| DCTN6 | OID2087 | Neurolog | DCTN6 | O00399 | 8 | 30156319 | 30183639 | 0 | - | - | - | - | - | - | - | - | - | - | - | - | - | - | - | - | - | - | - | - | - | - | - | - | - | - | - | - | - |
| EEF1D | OID3033 | Cardiom | EEF1D | P29692 | 8 | 143579697 | 143599541 | 0 | - | - | - | - | - | - | - | - | - | - | - | - | - | - | - | - | - | - | - | - | - | - | - | - | - | - | - | - | - |
| EIF4EBP | OID2016 | Cardiom | EIF4EBP | Q13541 | 8 | 38030534 | 38060365 | 0 | - | - | - | - | - | - | - | - | - | - | - | - | - | - | - | - | - | - | - | - | - | - | - | - | - | - | - | - | - |
| FGF20 | OID3045 | Inflamma | FGF20 | Q9NP95 | 8 | 16992181 | 17002345 | 0 | - | - | - | - | - | - | - | - | - | - | - | - | - | - | - | - | - | - | - | - | - | - | - | - | - | - | - | - | - |
| LSM1 | OID2121 | Oncology | LSM1 | O15116 | 8 | 38163335 | 38176730 | 0 | - | - | - | - | - | - | - | - | - | - | - | - | - | - | - | - | - | - | - | - | - | - | - | - | - | - | - | - | - |
| LYN | OID2135 | Oncology | LYN | P07948 | 8 | 55879835 | 56014169 | 0 | - | - | - | - | - | - | - | - | - | - | - | - | - | - | - | - | - | - | - | - | - | - | - | - | - | - | - | - | - |
| MTDH | OID3053 | Inflamma | MTDH | Q86UE4 | 8 | 97644184 | 97730260 | 0 | - | - | - | - | - | - | - | - | - | - | - | - | - | - | - | - | - | - | - | - | - | - | - | - | - | - | - | - | - |
| NDRG1 | OID2079 | Neurolog | NDRG1 | Q92597 | 8 | 133237175 | 133302022 | 0 | - | - | - | - | - | - | - | - | - | - | - | - | - | - | - | - | - | - | - | - | - | - | - | - | - | - | - | - | - |
| NEFL | OID2087 | Neurolog | NEFL | P07196 | 8 | 24950955 | 24956721 | 0 | - | - | - | - | - | - | - | - | - | - | - | - | - | - | - | - | - | - | - | - | - | - | - | - | - | - | - | - | - |
| OTUD6B | OID3148 | Oncology | OTUD6B | Q8N6M0 | 8 | 91070196 | 91087095 | 0 | - | - | - | - | - | - | - | - | - | - | - | - | - | - | - | - | - | - | - | - | - | - | - | - | - | - | - | - | - |
| PBK | OID3115 | Oncology | PBK | Q96KB5 | 8 | 27809624 | 27838082 | 0 | - | - | - | - | - | - | - | - | - | - | - | - | - | - | - | - | - | - | - | - | - | - | - | - | - | - | - | - | - |
| PDP1 | OID2117 | Oncology | PDP1 | Q9P0J1 | 8 | 93857807 | 93926068 | 0 | - | - | - | - | - | - | - | - | - | - | - | - | - | - | - | - | - | - | - | - | - | - | - | - | - | - | - | - | - |
| PNMA2 | OID3084 | Neurolog | PNMA2 | Q9UL42 | 8 | 26504701 | 26514092 | 0 | - | - | - | - | - | - | - | - | - | - | - | - | - | - | - | - | - | - | - | - | - | - | - | - | - | - | - | - | - |
| PTP4A3 | OID3046 | Inflamma | PTP4A3 | O75365 | 8 | 141391995 | 141432454 | 0 | - | - | - | - | - | - | - | - | - | - | - | - | - | - | - | - | - | - | - | - | - | - | - | - | - | - | - | - | - |
| RBPMS | OID3050 | Inflamma | RBPMS | Q93062 | 8 | 30384511 | 30572256 | 0 | - | - | - | - | - | - | - | - | - | - | - | - | - | - | - | - | - | - | - | - | - | - | - | - | - | - | - | - | - |
| SCRIB | OID3022 | Cardiom | SCRIB | Q14160 | 8 | 143790920 | 143815773 | 0 | - | - | - | - | - | - | - | - | - | - | - | - | - | - | - | - | - | - | - | - | - | - | - | - | - | - | - | - | - |
| TERF1 | OID3048 | Inflamma | TERF1 | P54274 | 8 | 73008856 | 73048123 | 0 | - | - | - | - | - | - | - | - | - | - | - | - | - | - | - | - | - | - | - | - | - | - | - | - | - | - | - | - | - |
| TOP1MT | OID3123 | Oncology | TOP1MT | Q969P6 | 8 | 143304384 | 143359979 | 0 | - | - | - | - | - | - | - | - | - | - | - | - | - | - | - | - | - | - | - | - | - | - | - | - | - | - | - | - | - |
| TP53INP1 | OID2012 | Cardiom | TP53INP | Q96A56 | 8 | 94925972 | 94949378 | 0 | - | - | - | - | - | - | - | - | - | - | - | - | - | - | - | - | - | - | - | - | - | - | - | - | - | - | - | - | - |
| VPS28 | OID3127 | Oncology | VPS28 | Q9UK41 | 8 | 144423601 | 144428563 | 0 | - | - | - | - | - | - | - | - | - | - | - | - | - | - | - | - | - | - | - | - | - | - | - | - | - | - | - | - | - |
| VPS37A | OID2135 | Oncology | VPS37A | Q8NEZ2 | 8 | 17246931 | 17302427 | 0 | - | - | - | - | - | - | - | - | - | - | - | - | - | - | - | - | - | - | - | - | - | - | - | - | - | - | - | - | - |
| YTHDF3 | OID2047 | Inflamma | YTHDF3 | Q7Z739 | 8 | 63168553 | 63212786 | 0 | - | - | - | - | - | - | - | - | - | - | - | - | - | - | - | - | - | - | - | - | - | - | - | - | - | - | - | - | - |
| ZHX2 | OID3114 | Neurolog | ZHX2 | Q9Y6X8 | 8 | 122781655 | 122974510 | 0 | - | - | - | - | - | - | - | - | - | - | - | - | - | - | - | - | - | - | - | - | - | - | - | - | - | - | - | - | - |
| AK1 | OID2012 | Cardiom | AK1 | P00568 | 9 | 127866486 | 127877675 | 0 | - | - | - | - | - | - | - | - | - | - | - | - | - | - | - | - | - | - | - | - | - | - | - | - | - | - | - | - | - |
| ANXA1 | OID3061 | Inflamma | ANXA1 | P04083 | 9 | 73151865 | 73170393 | 0 | - | - | - | - | - | - | - | - | - | - | - | - | - | - | - | - | - | - | - | - | - | - | - | - | - | - | - | - | - |
| ASS1 | OID3017 | Cardiom | ASS1 | P00966 | 9 | 130444961 | 130501274 | 0 | - | - | - | - | - | - | - | - | - | - | - | - | - | - | - | - | - | - | - | - | - | - | - | - | - | - | - | - | - |
| ATP6V1G | OID3024 | Cardiom | ATP6V1G | O75348 | 9 | 114587706 | 114598915 | 0 | - | - | - | - | - | - | - | - | - | - | - | - | - | - | - | - | - | - | - | - | - | - | - | - | - | - | - | - | - |
| BRD3 | OID3009 | Cardiom | BRD3 | Q15059 | 9 | 134030305 | 134068535 | 0 | - | - | - | - | - | - | - | - | - | - | - | - | - | - | - | - | - | - | - | - | - | - | - | - | - | - | - | - | - |
| C9orf40 | OID3148 | Oncology | C9orf40 | Q8IXQ3 | 9 | 74946583 | 74952912 | 0 | - | - | - | - | - | - | - | - | - | - | - | - | - | - | - | - | - | - | - | - | - | - | - | - | - | - | - | - | - |
| CAMSAP | OID3107 | Neurolog | CAMSAP | Q5T5Y3 | 9 | 135808487 | 135907546 | 0 | - | - | - | - | - | - | - | - | - | - | - | - | - | - | - | - | - | - | - | - | - | - | - | - | - | - | - | - | - |
| CDC26 | OID3150 | Oncology | CDC26 | Q8NHZ8 | 9 | 113255835 | 113275572 | 0 | - | - | - | - | - | - | - | - | - | - | - | - | - | - | - | - | - | - | - | - | - | - | - | - | - | - | - | - | - |
| CLTA | OID2011 | Cardiom | CLTA | P09496 | 9 | 36190856 | 36304781 | 0 | - | - | - | - | - | - | - | - | - | - | - | - | - | - | - | - | - | - | - | - | - | - | - | - | - | - | - | - | - |
| CREB3 | OID3118 | Oncology | CREB3 | O43889 | 9 | 35732598 | 35736999 | 0 | - | - | - | - | - | - | - | - | - | - | - | - | - | - | - | - | - | - | - | - | - | - | - | - | - | - | - | - | - |
| DNAJA1 | OID3120 | Oncology | DNAJA1 | P31689 | 9 | 33025273 | 33039907 | 0 | - | - | - | - | - | - | - | - | - | - | - | - | - | - | - | - | - | - | - | - | - | - | - | - | - | - | - | - | - |
| DNLZ | OID3082 | Neurolog | DNLZ | Q5SXM8 | 9 | 136359480 | 136363744 | 0 | - | - | - | - | - | - | - | - | - | - | - | - | - | - | - | - | - | - | - | - | - | - | - | - | - | - | - | - | - |
| FGD3 | OID3104 | Neurolog | FGD3 | Q5JSP0 | 9 | 92947523 | 93036236 | 0 | - | - | - | - | - | - | - | - | - | - | - | - | - | - | - | - | - | - | - | - | - | - | - | - | - | - | - | - | - |
| GNE | OID2123 | Oncology | GNE | Q9Y223 | 9 | 36214441 | 36277042 | 0 | - | - | - | - | - | - | - | - | - | - | - | - | - | - | - | - | - | - | - | - | - | - | - | - | - | - | - | - | - |
| HNRNPK | OID2005 | Cardiom | HNRNPK | P61978 | 9 | 83968083 | 83980616 | 0 | - | - | - | - | - | - | - | - | - | - | - | - | - | - | - | - | - | - | - | - | - | - | - | - | - | - | - | - | - |
| HSD17B | OID3014 | Cardiom | HSD17B | P37058 | 9 | 96235306 | 96302176 | 0 | - | - | - | - | - | - | - | - | - | - | - | - | - | - | - | - | - | - | - | - | - | - | - | - | - | - | - | - | - |
| IFNW1 | OID3012 | Cardiom | IFNW1 | P05000 | 9 | 21140632 | 21141832 | 0 | - | - | - | - | - | - | - | - | - | - | - | - | - | - | - | - | - | - | - | - | - | - | - | - | - | - | - | - | - |
| IL33 | OID2042 | Inflamma | IL33 | O95760 | 9 | 6215786 | 6257983 | 0 | - | - | - | - | - | - | - | - | - | - | - | - | - | - | - | - | - | - | - | - | - | - | - | - | - | - | - | - | - |
| KLF4 | OID3080 | Neurolog | KLF4 | O43474 | 9 | 107484852 | 107490482 | 0 | - | - | - | - | - | - | - | - | - | - | - | - | - | - | - | - | - | - | - | - | - | - | - | - | - | - | - | - | - |
| NFX1 | OID3015 | Cardiom | NFX1 | Q12986 | 9 | 33290512 | 33371157 | 0 | - | - | - | - | - | - | - | - | - | - | - | - | - | - | - | - | - | - | - | - | - | - | - | - | - | - | - | - | - |
| PRUNE2 | OID3127 | Oncology | PRUNE2 | Q8WUY3 | 9 | 76611376 | 76906114 | 0 | - | - | - | - | - | - | - | - | - | - | - | - | - | - | - | - | - | - | - | - | - | - | - | - | - | - | - | - | - |
| PSIP1 | OID2053 | Inflamma | PSIP1 | O75475 | 9 | 15464066 | 15510995 | 0 | - | - | - | - | - | - | - | - | - | - | - | - | - | - | - | - | - | - | - | - | - | - | - | - | - | - | - | - | - |
| PSMD5 | OID3091 | Neurolog | PSMD5 | Q16401 | 9 | 120815496 | 120842951 | 0 | - | - | - | - | - | - | - | - | - | - | - | - | - | - | - | - | - | - | - | - | - | - | - | - | - | - | - | - | - |
| RLN1 | OID3099 | Neurolog | RLN1 | P04808 | 9 | 5334930 | 5339876 | 0 | - | - | - | - | - | - | - | - | - | - | - | - | - | - | - | - | - | - | - | - | - | - | - | - | - | - | - | - | - |
| SIT1 | OID2046 | Inflamma | SIT1 | Q9Y3P8 | 9 | 35649295 | 35650931 | 0 | - | - | - | - | - | - | - | - | - | - | - | - | - | - | - | - | - | - | - | - | - | - | - | - | - | - | - | - | - |
| SLC34A3 | OID3120 | Oncology | SLC34A3 | Q8N130 | 9 | 137230757 | 137236555 | 0 | - | - | - | - | - | - | - | - | - | - | - | - | - | - | - | - | - | - | - | - | - | - | - | - | - | - | - | - | - |
| STOML2 | OID3118 | Oncology | STOML2 | Q9UJZ1 | 9 | 35099776 | 35103195 | 0 | - | - | - | - | - | - | - | - | - | - | - | - | - | - | - | - | - | - | - | - | - | - | - | - | - | - | - | - | - |
| TSC1 | OID3047 | Inflamma | TSC1 | Q92574 | 9 | 132891348 | 132946874 | 0 | - | - | - | - | - | - | - | - | - | - | - | - | - | - | - | - | - | - | - | - | - | - | - | - | - | - | - | - | - |
| UHRF2 | OID3017 | Cardiom | UHRF2 | Q96PU4 | 9 | 6413151 | 6507054 | 0 | - | - | - | - | - | - | - | - | - | - | - | - | - | - | - | - | - | - | - | - | - | - | - | - | - | - | - | - | - |
| ABRAXAS | OID3013 | Cardiom | ABRAXA | Q15018 | 10 | 124801819 | 124836667 | 0 | - | - | - | - | - | - | - | - | - | - | - | - | - | - | - | - | - | - | - | - | - | - | - | - | - | - | - | - | - |
| ADRA2A | OID3006 | Cardiom | ADRA2A | P08913 | 10 | 111077029 | 111080907 | 0 | - | - | - | - | - | - | - | - | - | - | - | - | - | - | - | - | - | - | - | - | - | - | - | - | - | - | - | - | - |
| BLOC1S | OID3085 | Neurolog | BLOC1S | Q6QNY1 | 10 | 100273280 | 100286680 | 0 | - | - | - | - | - | - | - | - | - | - | - | - | - | - | - | - | - | - | - | - | - | - | - | - | - | - | - | - | - |
| CALY | OID3014 | Cardiom | CALY | Q9NYX4 | 10 | 133324072 | 133336935 | 0 | - | - | - | - | - | - | - | - | - | - | - | - | - | - | - | - | - | - | - | - | - | - | - | - | - | - | - | - | - |
| CDC123 | OID3124 | Oncology | CDC123 | O75794 | 10 | 12195965 | 12250589 | 0 | - | - | - | - | - | - | - | - | - | - | - | - | - | - | - | - | - | - | - | - | - | - | - | - | - | - | - | - | - |
| CDK1 | OID3116 | Oncology | CDK1 | P06493 | 10 | 60778331 | 60794852 | 0 | - | - | - | - | - | - | - | - | - | - | - | - | - | - | - | - | - | - | - | - | - | - | - | - | - | - | - | - | - |
| CUZD1 | OID3091 | Neurolog | CUZD1 | Q86UP6 | 10 | 122832158 | 122846175 | 0 | - | - | - | - | - | - | - | - | - | - | - | - | - | - | - | - | - | - | - | - | - | - | - | - | - | - | - | - | - |
| DCLRE1 | OID3124 | Oncology | DCLRE1 | Q96SD1 | 10 | 14897359 | 14954432 | 0 | - | - | - | - | - | - | - | - | - | - | - | - | - | - | - | - | - | - | - | - | - | - | - | - | - | - | - | - | - |
| DNAJC9 | OID3147 | Oncology | DNAJC9 | Q8WXX5 | 10 | 73183362 | 73247255 | 0 | - | - | - | - | - | - | - | - | - | - | - | - | - | - | - | - | - | - | - | - | - | - | - | - | - | - | - | - | - |
| FHIP2A | OID3018 | Cardiom | FHIP2A | Q5W0V3 | 10 | 114821744 | 114899832 | 0 | - | - | - | - | - | - | - | - | - | - | - | - | - | - | - | - | - | - | - | - | - | - | - | - | - | - | - | - | - |
| FZD8 | OID3081 | Neurolog | FZD8 | Q9H461 | 10 | 35638247 | 35642296 | 0 | - | - | - | - | - | - | - | - | - | - | - | - | - | - | - | - | - | - | - | - | - | - | - | - | - | - | - | - | - |
| GAD2 | OID3044 | Inflamma | GAD2 | Q05329 | 10 | 26216665 | 26304558 | 0 | - | - | - | - | - | - | - | - | - | - | - | - | - | - | - | - | - | - | - | - | - | - | - | - | - | - | - | - | - |
| GATA3 | OID3121 | Oncology | GATA3 | P23771 | 10 | 8045378 | 8075198 | 0 | - | - | - | - | - | - | - | - | - | - | - | - | - | - | - | - | - | - | - | - | - | - | - | - | - | - | - | - | - |
| GDF2 | OID2020 | Cardiom | GDF2 | Q9UK05 | 10 | 47322454 | 47327588 | 0 | - | - | - | - | - | - | - | - | - | - | - | - | - | - | - | - | - | - | - | - | - | - | - | - | - | - | - | - | - |
| HHEX | OID3110 | Neurolog | HHEX | Q03014 | 10 | 92689955 | 92695647 | 0 | - | - | - | - | - | - | - | - | - | - | - | - | - | - | - | - | - | - | - | - | - | - | - | - | - | - | - | - | - |
| IFIT1 | OID3131 | Oncology | IFIT1 | P09914 | 10 | 89392546 | 89406487 | 0 | - | - | - | - | - | - | - | - | - | - | - | - | - | - | - | - | - | - | - | - | - | - | - | - | - | - | - | - | - |
| ITGB1 | OID2037 | Cardiom | ITGB1 | P05556 | 10 | 32887273 | 33005792 | 0 | - | - | - | - | - | - | - | - | - | - | - | - | - | - | - | - | - | - | - | - | - | - | - | - | - | - | - | - | - |
| ITPRIP | OID3086 | Neurolog | ITPRIP | Q8IWB1 | 10 | 104309698 | 104338465 | 0 | - | - | - | - | - | - | - | - | - | - | - | - | - | - | - | - | - | - | - | - | - | - | - | - | - | - | - | - | - |
| JMJD1C | OID3117 | Oncology | JMJD1C | Q15652 | 10 | 63167221 | 63521850 | 0 | - | - | - | - | - | - | - | - | - | - | - | - | - | - | - | - | - | - | - | - | - | - | - | - | - | - | - | - | - |
| KIF20B | OID3081 | Neurolog | KIF20B | Q96Q89 | 10 | 89701590 | 89774939 | 0 | - | - | - | - | - | - | - | - | - | - | - | - | - | - | - | - | - | - | - | - | - | - | - | - | - | - | - | - | - |
| MKI67 | OID3045 | Inflamma | MKI67 | P46013 | 10 | 128096659 | 128126423 | 0 | - | - | - | - | - | - | - | - | - | - | - | - | - | - | - | - | - | - | - | - | - | - | - | - | - | - | - | - | - |
| MORN4 | OID3121 | Oncology | MORN4 | Q8NDC4 | 10 | 97614553 | 97633500 | 0 | - | - | - | - | - | - | - | - | - | - | - | - | - | - | - | - | - | - | - | - | - | - | - | - | - | - | - | - | - |
| MRPS16 | OID3048 | Inflamma | MRPS16 | Q9Y3D3 | 10 | 73248843 | 73252693 | 0 | - | - | - | - | - | - | - | - | - | - | - | - | - | - | - | - | - | - | - | - | - | - | - | - | - | - | - | - | - |
| NFKB2 | OID3125 | Oncology | NFKB2 | Q00653 | 10 | 102394110 | 102402524 | 0 | - | - | - | - | - | - | - | - | - | - | - | - | - | - | - | - | - | - | - | - | - | - | - | - | - | - | - | - | - |
| PNLIP | OID3069 | Inflamma | PNLIP | P16233 | 10 | 116545931 | 116567855 | 0 | - | - | - | - | - | - | - | - | - | - | - | - | - | - | - | - | - | - | - | - | - | - | - | - | - | - | - | - | - |
| PPIF | OID3038 | Cardiom | PPIF | P30405 | 10 | 79347469 | 79355334 | 0 | - | - | - | - | - | - | - | - | - | - | - | - | - | - | - | - | - | - | - | - | - | - | - | - | - | - | - | - | - |
| PRKCQ | OID2046 | Inflamma | PRKCQ | Q04759 | 10 | 6427143 | 6580301 | 0 | - | - | - | - | - | - | - | - | - | - | - | - | - | - | - | - | - | - | - | - | - | - | - | - | - | - | - | - | - |
| PTEN | OID2079 | Neurolog | PTEN | P60484 | 10 | 87862638 | 87971930 | 0 | - | - | - | - | - | - | - | - | - | - | - | - | - | - | - | - | - | - | - | - | - | - | - | - | - | - | - | - | - |
| RBM17 | OID3103 | Neurolog | RBM17 | Q96I25 | 10 | 6089034 | 6117457 | 0 | - | - | - | - | - | - | - | - | - | - | - | - | - | - | - | - | - | - | - | - | - | - | - | - | - | - | - | - | - |
| RGS10 | OID3099 | Neurolog | RGS10 | O43665 | 10 | 119499817 | 119542719 | 0 | - | - | - | - | - | - | - | - | - | - | - | - | - | - | - | - | - | - | - | - | - | - | - | - | - | - | - | - | - |
| RTKN2 | OID3004 | Cardiom | RTKN2 | Q8IZC4 | 10 | 62183035 | 62268844 | 0 | - | - | - | - | - | - | - | - | - | - | - | - | - | - | - | - | - | - | - | - | - | - | - | - | - | - | - | - | - |
| SIRT1 | OID3043 | Inflamma | SIRT1 | Q96EB6 | 10 | 67884656 | 67918390 | 0 | - | - | - | - | - | - | - | - | - | - | - | - | - | - | - | - | - | - | - | - | - | - | - | - | - | - | - | - | - |
| SLK | OID3128 | Oncology | SLK | Q9H2G2 | 10 | 103967140 | 104029233 | 0 | - | - | - | - | - | - | - | - | - | - | - | - | - | - | - | - | - | - | - | - | - | - | - | - | - | - | - | - | - |
| SMC3 | OID3119 | Oncology | SMC3 | Q9UQE7 | 10 | 110567684 | 110606048 | 0 | - | - | - | - | - | - | - | - | - | - | - | - | - | - | - | - | - | - | - | - | - | - | - | - | - | - | - | - | - |
| SMNDC1 | OID3147 | Oncology | SMNDC1 | O75940 | 10 | 110290730 | 110304938 | 0 | - | - | - | - | - | - | - | - | - | - | - | - | - | - | - | - | - | - | - | - | - | - | - | - | - | - | - | - | - |
| STAM | OID3142 | Oncology | STAM | Q92783 | 10 | 17644151 | 17716824 | 0 | - | - | - | - | - | - | - | - | - | - | - | - | - | - | - | - | - | - | - | - | - | - | - | - | - | - | - | - | - |
| VIM | OID2005 | Cardiom | VIM | P08670 | 10 | 17228241 | 17237593 | 0 | - | - | - | - | - | - | - | - | - | - | - | - | - | - | - | - | - | - | - | - | - | - | - | - | - | - | - | - | - |
| AGBL2 | OID3079 | Neurolog | AGBL2 | Q5U5Z8 | 11 | 47659591 | 47715389 | 0 | - | - | - | - | - | - | - | - | - | - | - | - | - | - | - | - | - | - | - | - | - | - | - | - | - | - | - | - | - |
| ARHGAP | OID2120 | Oncology | ARHGAP | Q07960 | 11 | 46677080 | 46700619 | 0 | - | - | - | - | - | - | - | - | - | - | - | - | - | - | - | - | - | - | - | - | - | - | - | - | - | - | - | - | - |
| ARHGEF | OID2055 | Inflamma | ARHGEF | Q9NZN5 | 11 | 120336413 | 120489937 | 0 | - | - | - | - | - | - | - | - | - | - | - | - | - | - | - | - | - | - | - | - | - | - | - | - | - | - | - | - | - |
| ARNTL | OID3016 | Cardiom | ARNTL | O00327 | 11 | 13276652 | 13387266 | 0 | - | - | - | - | - | - | - | - | - | - | - | - | - | - | - | - | - | - | - | - | - | - | - | - | - | - | - | - | - |
| B3GAT3 | OID3016 | Cardiom | B3GAT3 | O94766 | 11 | 62615296 | 62622154 | 0 | - | - | - | - | - | - | - | - | - | - | - | - | - | - | - | - | - | - | - | - | - | - | - | - | - | - | - | - | - |
| BIRC2 | OID2121 | Oncology | BIRC2 | Q13490 | 11 | 102347211 | 102378670 | 0 | - | - | - | - | - | - | - | - | - | - | - | - | - | - | - | - | - | - | - | - | - | - | - | - | - | - | - | - | - |
| CABP2 | OID3091 | Neurolog | CABP2 | Q9NPB3 | 11 | 67518912 | 67524517 | 0 | - | - | - | - | - | - | - | - | - | - | - | - | - | - | - | - | - | - | - | - | - | - | - | - | - | - | - | - | - |
| CASP4 | OID3127 | Oncology | CASP4 | P49662 | 11 | 104942866 | 104969366 | 0 | - | - | - | - | - | - | - | - | - | - | - | - | - | - | - | - | - | - | - | - | - | - | - | - | - | - | - | - | - |
| CD3D | OID3082 | Neurolog | CD3D | P04234 | 11 | 118339075 | 118342705 | 0 | - | - | - | - | - | - | - | - | - | - | - | - | - | - | - | - | - | - | - | - | - | - | - | - | - | - | - | - | - |
| CD3E | OID3130 | Oncology | CD3E | P07766 | 11 | 118304730 | 118316175 | 0 | - | - | - | - | - | - | - | - | - | - | - | - | - | - | - | - | - | - | - | - | - | - | - | - | - | - | - | - | - |
| CD3G | OID3044 | Inflamma | CD3G | P09693 | 11 | 118344344 | 118355161 | 0 | - | - | - | - | - | - | - | - | - | - | - | - | - | - | - | - | - | - | - | - | - | - | - | - | - | - | - | - | - |
| CD82 | OID3125 | Oncology | CD82 | P27701 | 11 | 44564427 | 44620358 | 0 | - | - | - | - | - | - | - | - | - | - | - | - | - | - | - | - | - | - | - | - | - | - | - | - | - | - | - | - | - |
| CEND1 | OID3082 | Neurolog | CEND1 | Q8N111 | 11 | 787115 | 790113 | 0 | - | - | - | - | - | - | - | - | - | - | - | - | - | - | - | - | - | - | - | - | - | - | - | - | - | - | - | - | - |
| CEP164 | OID2044 | Inflamma | CEP164 | Q9UPV0 | 11 | 117314557 | 117413266 | 0 | - | - | - | - | - | - | - | - | - | - | - | - | - | - | - | - | - | - | - | - | - | - | - | - | - | - | - | - | - |
| CHRM1 | OID3093 | Neurolog | CHRM1 | P11229 | 11 | 62908679 | 62921807 | 0 | - | - | - | - | - | - | - | - | - | - | - | - | - | - | - | - | - | - | - | - | - | - | - | - | - | - | - | - | - |
| CLNS1A | OID3092 | Neurolog | CLNS1A | P54105 | 11 | 77514936 | 77637794 | 0 | - | - | - | - | - | - | - | - | - | - | - | - | - | - | - | - | - | - | - | - | - | - | - | - | - | - | - | - | - |
| CNTF | OID3080 | Neurolog | CNTF | P26441 | 11 | 58622665 | 58625733 | 0 | - | - | - | - | - | - | - | - | - | - | - | - | - | - | - | - | - | - | - | - | - | - | - | - | - | - | - | - | - |
| COMMD9 | OID3139 | Oncology | COMMD9 | Q9P000 | 11 | 36269284 | 36289449 | 0 | - | - | - | - | - | - | - | - | - | - | - | - | - | - | - | - | - | - | - | - | - | - | - | - | - | - | - | - | - |
| CREBZF | OID3120 | Oncology | CREBZF | Q9NS37 | 11 | 85657742 | 85682908 | 0 | - | - | - | - | - | - | - | - | - | - | - | - | - | - | - | - | - | - | - | - | - | - | - | - | - | - | - | - | - |
| CSRP3 | OID3017 | Cardiom | CSRP3 | P50461 | 11 | 19182030 | 19210571 | 0 | - | - | - | - | - | - | - | - | - | - | - | - | - | - | - | - | - | - | - | - | - | - | - | - | - | - | - | - | - |
| DDX25 | OID3089 | Neurolog | DDX25 | Q9UHL0 | 11 | 125903348 | 125943702 | 0 | - | - | - | - | - | - | - | - | - | - | - | - | - | - | - | - | - | - | - | - | - | - | - | - | - | - | - | - | - |

| | | | | | | | | | | | | | | | | | | | | | | | | | | | | | | | | | | | | | |
|---|---|---|---|---|---|---|---|---|---|---|---|---|---|---|---|---|---|---|---|---|---|---|---|---|---|---|---|---|---|---|---|---|---|---|---|---|---|
| DENND2 | OID3005 | Cardiom | DENND2 | P78524 | 11 | 8693351 | 8910951 | 0 | - | - | - | - | - | - | - | - | - | - | - | - | - | - | - | - | - | - | - | - | - | - | - | - | - | - | - | - | - |
| DGKZ | OID2046 | Inflamma | DGKZ | Q13574 | 11 | 46332905 | 46380554 | 0 | - | - | - | - | - | - | - | - | - | - | - | - | - | - | - | - | - | - | - | - | - | - | - | - | - | - | - | - | - |
| FEN1 | OID2119 | Oncology | FEN1 | P39748 | 11 | 61792911 | 61797238 | 0 | - | - | - | - | - | - | - | - | - | - | - | - | - | - | - | - | - | - | - | - | - | - | - | - | - | - | - | - | - |
| FGF19 | OID2071 | Inflamma | FGF19 | O95750 | 11 | 69698238 | 69704022 | 0 | - | - | - | - | - | - | - | - | - | - | - | - | - | - | - | - | - | - | - | - | - | - | - | - | - | - | - | - | - |
| FGF3 | OID3054 | Inflamma | FGF3 | P11487 | 11 | 69809968 | 69819416 | 0 | - | - | - | - | - | - | - | - | - | - | - | - | - | - | - | - | - | - | - | - | - | - | - | - | - | - | - | - | - |
| FLI1 | OID2123 | Oncology | FLI1 | Q01543 | 11 | 128686535 | 128813267 | 0 | - | - | - | - | - | - | - | - | - | - | - | - | - | - | - | - | - | - | - | - | - | - | - | - | - | - | - | - | - |
| HRAS | OID3126 | Oncology | HRAS | P01112 | 11 | 532242 | 537321 | 0 | - | - | - | - | - | - | - | - | - | - | - | - | - | - | - | - | - | - | - | - | - | - | - | - | - | - | - | - | - |
| INPPL1 | OID2131 | Oncology | INPPL1 | O15357 | 11 | 72223701 | 72239147 | 0 | - | - | - | - | - | - | - | - | - | - | - | - | - | - | - | - | - | - | - | - | - | - | - | - | - | - | - | - | - |
| KIAA154 | OID3131 | Oncology | KIAA154 | Q6ZVL6 | 11 | 33376108 | 33674102 | 0 | - | - | - | - | - | - | - | - | - | - | - | - | - | - | - | - | - | - | - | - | - | - | - | - | - | - | - | - | - |
| LTO1 | OID2043 | Inflamma | LTO1 | Q8WV07 | 11 | 69653076 | 69675416 | 0 | - | - | - | - | - | - | - | - | - | - | - | - | - | - | - | - | - | - | - | - | - | - | - | - | - | - | - | - | - |
| NRGN | OID3067 | Inflamma | NRGN | Q92686 | 11 | 124739942 | 124747210 | 0 | - | - | - | - | - | - | - | - | - | - | - | - | - | - | - | - | - | - | - | - | - | - | - | - | - | - | - | - | - |
| NXPE4 | OID3007 | Cardiom | NXPE4 | Q6UWF7 | 11 | 114570591 | 114595786 | 0 | - | - | - | - | - | - | - | - | - | - | - | - | - | - | - | - | - | - | - | - | - | - | - | - | - | - | - | - | - |
| PGR | OID3046 | Inflamma | PGR | P06401 | 11 | 101029624 | 101129813 | 0 | - | - | - | - | - | - | - | - | - | - | - | - | - | - | - | - | - | - | - | - | - | - | - | - | - | - | - | - | - |
| PSMA1 | OID2122 | Oncology | PSMA1 | P25786 | 11 | 14504874 | 14643635 | 0 | - | - | - | - | - | - | - | - | - | - | - | - | - | - | - | - | - | - | - | - | - | - | - | - | - | - | - | - | - |
| PSMC3 | OID3120 | Oncology | PSMC3 | P17980 | 11 | 47418769 | 47426473 | 0 | - | - | - | - | - | - | - | - | - | - | - | - | - | - | - | - | - | - | - | - | - | - | - | - | - | - | - | - | - |
| PTH | OID3096 | Neurolog | PTH | P01270 | 11 | 13492054 | 13496181 | 0 | - | - | - | - | - | - | - | - | - | - | - | - | - | - | - | - | - | - | - | - | - | - | - | - | - | - | - | - | - |
| REXO2 | OID3007 | Cardiom | REXO2 | Q9Y3B8 | 11 | 114439435 | 114450279 | 0 | - | - | - | - | - | - | - | - | - | - | - | - | - | - | - | - | - | - | - | - | - | - | - | - | - | - | - | - | - |
| SART1 | OID3011 | Cardiom | SART1 | O43290 | 11 | 65961728 | 65980137 | 0 | - | - | - | - | - | - | - | - | - | - | - | - | - | - | - | - | - | - | - | - | - | - | - | - | - | - | - | - | - |
| SCGB2A | OID3010 | Cardiom | SCGB2A | Q13296 | 11 | 62270158 | 62273160 | 0 | - | - | - | - | - | - | - | - | - | - | - | - | - | - | - | - | - | - | - | - | - | - | - | - | - | - | - | - | - |
| SCN2B | OID3094 | Neurolog | SCN2B | O60939 | 11 | 118162806 | 118176639 | 0 | - | - | - | - | - | - | - | - | - | - | - | - | - | - | - | - | - | - | - | - | - | - | - | - | - | - | - | - | - |
| SCN3B | OID3080 | Neurolog | SCN3B | Q9NY72 | 11 | 123629187 | 123655244 | 0 | - | - | - | - | - | - | - | - | - | - | - | - | - | - | - | - | - | - | - | - | - | - | - | - | - | - | - | - | - |
| SCT | OID3095 | Neurolog | SCT | P09683 | 11 | 626309 | 627181 | 0 | - | - | - | - | - | - | - | - | - | - | - | - | - | - | - | - | - | - | - | - | - | - | - | - | - | - | - | - | - |
| SPTBN2 | OID3087 | Neurolog | SPTBN2 | O15020 | 11 | 66682497 | 66744670 | 0 | - | - | - | - | - | - | - | - | - | - | - | - | - | - | - | - | - | - | - | - | - | - | - | - | - | - | - | - | - |
| SSH3 | OID3121 | Oncology | SSH3 | Q8TE77 | 11 | 67303478 | 67312607 | 0 | - | - | - | - | - | - | - | - | - | - | - | - | - | - | - | - | - | - | - | - | - | - | - | - | - | - | - | - | - |
| STIP1 | OID2107 | Neurolog | STIP1 | P31948 | 11 | 64185272 | 64204543 | 0 | - | - | - | - | - | - | - | - | - | - | - | - | - | - | - | - | - | - | - | - | - | - | - | - | - | - | - | - | - |
| STX3 | OID3109 | Neurolog | STX3 | Q13277 | 11 | 59713456 | 59805882 | 0 | - | - | - | - | - | - | - | - | - | - | - | - | - | - | - | - | - | - | - | - | - | - | - | - | - | - | - | - | - |
| STX5 | OID3043 | Inflamma | STX5 | Q13190 | 11 | 62806860 | 62832051 | 0 | - | - | - | - | - | - | - | - | - | - | - | - | - | - | - | - | - | - | - | - | - | - | - | - | - | - | - | - | - |
| THAP12 | OID3123 | Oncology | THAP12 | O43422 | 11 | 76349898 | 76381132 | 0 | - | - | - | - | - | - | - | - | - | - | - | - | - | - | - | - | - | - | - | - | - | - | - | - | - | - | - | - | - |
| TPBGL | OID3092 | Neurolog | TPBGL | P0DKB5 | 11 | 75240774 | 75243704 | 0 | - | - | - | - | - | - | - | - | - | - | - | - | - | - | - | - | - | - | - | - | - | - | - | - | - | - | - | - | - |
| UBQLN3 | OID3009 | Cardiom | UBQLN3 | Q9H347 | 11 | 5507300 | 5509958 | 0 | - | - | - | - | - | - | - | - | - | - | - | - | - | - | - | - | - | - | - | - | - | - | - | - | - | - | - | - | - |
| UBXN1 | OID3065 | Inflamma | UBXN1 | Q04323 | 11 | 62676498 | 62679117 | 0 | - | - | - | - | - | - | - | - | - | - | - | - | - | - | - | - | - | - | - | - | - | - | - | - | - | - | - | - | - |
| USP47 | OID3011 | Cardiom | USP47 | Q96K76 | 11 | 11841423 | 11961887 | 0 | - | - | - | - | - | - | - | - | - | - | - | - | - | - | - | - | - | - | - | - | - | - | - | - | - | - | - | - | - |
| VWA5A | OID3119 | Oncology | VWA5A | O00534 | 11 | 124115404 | 124147721 | 0 | - | - | - | - | - | - | - | - | - | - | - | - | - | - | - | - | - | - | - | - | - | - | - | - | - | - | - | - | - |
| ZNRD2 | OID3145 | Oncology | ZNRD2 | O60232 | 11 | 65570460 | 65573942 | 0 | - | - | - | - | - | - | - | - | - | - | - | - | - | - | - | - | - | - | - | - | - | - | - | - | - | - | - | - | - |
| ALDH2 | OID3138 | Oncology | ALDH2 | P05091 | 12 | 111766887 | 111817532 | 0 | - | - | - | - | - | - | - | - | - | - | - | - | - | - | - | - | - | - | - | - | - | - | - | - | - | - | - | - | - |
| ATXN2 | OID3085 | Neurolog | ATXN2 | Q99700 | 12 | 111443485 | 111599676 | 0 | - | - | - | - | - | - | - | - | - | - | - | - | - | - | - | - | - | - | - | - | - | - | - | - | - | - | - | - | - |
| BCL7A | OID3121 | Oncology | BCL7A | Q4VC05 | 12 | 122019422 | 122062044 | 0 | - | - | - | - | - | - | - | - | - | - | - | - | - | - | - | - | - | - | - | - | - | - | - | - | - | - | - | - | - |
| BRAP | OID3150 | Oncology | BRAP | Q7Z569 | 12 | 111642146 | 111685956 | 0 | - | - | - | - | - | - | - | - | - | - | - | - | - | - | - | - | - | - | - | - | - | - | - | - | - | - | - | - | - |
| CACNA1 | OID3084 | Neurolog | CACNA1 | Q13936 | 12 | 1970772 | 2697950 | 0 | - | - | - | - | - | - | - | - | - | - | - | - | - | - | - | - | - | - | - | - | - | - | - | - | - | - | - | - | - |
| CCND2 | OID3131 | Oncology | CCND2 | P30279 | 12 | 4269771 | 4305353 | 0 | - | - | - | - | - | - | - | - | - | - | - | - | - | - | - | - | - | - | - | - | - | - | - | - | - | - | - | - | - |
| CD63 | OID2098 | Neurolog | CD63 | P08962 | 12 | 55725323 | 55729707 | 0 | - | - | - | - | - | - | - | - | - | - | - | - | - | - | - | - | - | - | - | - | - | - | - | - | - | - | - | - | - |
| CEP290 | OID3128 | Oncology | CEP290 | O15078 | 12 | 88049016 | 88142099 | 0 | - | - | - | - | - | - | - | - | - | - | - | - | - | - | - | - | - | - | - | - | - | - | - | - | - | - | - | - | - |
| CNPY2 | OID2006 | Cardiom | CNPY2 | Q9Y2B0 | 12 | 56309842 | 56316119 | 0 | - | - | - | - | - | - | - | - | - | - | - | - | - | - | - | - | - | - | - | - | - | - | - | - | - | - | - | - | - |
| DCTN2 | OID2133 | Oncology | DCTN2 | Q13561 | 12 | 57529633 | 57547224 | 0 | - | - | - | - | - | - | - | - | - | - | - | - | - | - | - | - | - | - | - | - | - | - | - | - | - | - | - | - | - |
| DENR | OID3049 | Inflamma | DENR | O43583 | 12 | 122752824 | 122771064 | 0 | - | - | - | - | - | - | - | - | - | - | - | - | - | - | - | - | - | - | - | - | - | - | - | - | - | - | - | - | - |
| DGKA | OID3054 | Inflamma | DGKA | P23743 | 12 | 55927319 | 55954027 | 0 | - | - | - | - | - | - | - | - | - | - | - | - | - | - | - | - | - | - | - | - | - | - | - | - | - | - | - | - | - |
| DIABLO | OID2018 | Cardiom | DIABLO | Q9NR28 | 12 | 122207663 | 122227456 | 0 | - | - | - | - | - | - | - | - | - | - | - | - | - | - | - | - | - | - | - | - | - | - | - | - | - | - | - | - | - |
| EIF4B | OID2100 | Neurolog | EIF4B | P23588 | 12 | 53006282 | 53042215 | 0 | - | - | - | - | - | - | - | - | - | - | - | - | - | - | - | - | - | - | - | - | - | - | - | - | - | - | - | - | - |
| ERP29 | OID3057 | Inflamma | ERP29 | P30040 | 12 | 112013348 | 112023449 | 0 | - | - | - | - | - | - | - | - | - | - | - | - | - | - | - | - | - | - | - | - | - | - | - | - | - | - | - | - | - |
| ESPL1 | OID3117 | Oncology | ESPL1 | Q14674 | 12 | 53268299 | 53293638 | 0 | - | - | - | - | - | - | - | - | - | - | - | - | - | - | - | - | - | - | - | - | - | - | - | - | - | - | - | - | - |
| FGF6 | OID3044 | Inflamma | FGF6 | P10767 | 12 | 4428155 | 4445815 | 0 | - | - | - | - | - | - | - | - | - | - | - | - | - | - | - | - | - | - | - | - | - | - | - | - | - | - | - | - | - |
| FZD10 | OID3093 | Neurolog | FZD10 | Q9ULW2 | 12 | 130162459 | 130165740 | 0 | - | - | - | - | - | - | - | - | - | - | - | - | - | - | - | - | - | - | - | - | - | - | - | - | - | - | - | - | - |
| GABARA | OID3125 | Oncology | GABARA | Q9H0R8 | 12 | 10212458 | 10223128 | 0 | - | - | - | - | - | - | - | - | - | - | - | - | - | - | - | - | - | - | - | - | - | - | - | - | - | - | - | - | - |
| GAPDH | OID3060 | Inflamma | GAPDH | P04406 | 12 | 6534512 | 6538374 | 0 | - | - | - | - | - | - | - | - | - | - | - | - | - | - | - | - | - | - | - | - | - | - | - | - | - | - | - | - | - |
| GRIN2B | OID3080 | Neurolog | GRIN2B | Q13224 | 12 | 13437942 | 13982002 | 0 | - | - | - | - | - | - | - | - | - | - | - | - | - | - | - | - | - | - | - | - | - | - | - | - | - | - | - | - | - |
| GUCY2C | OID3131 | Oncology | GUCY2C | P25092 | 12 | 14612632 | 14696599 | 0 | - | - | - | - | - | - | - | - | - | - | - | - | - | - | - | - | - | - | - | - | - | - | - | - | - | - | - | - | - |
| HNF1A | OID3091 | Neurolog | HNF1A | P20823 | 12 | 120978543 | 121002512 | 0 | - | - | - | - | - | - | - | - | - | - | - | - | - | - | - | - | - | - | - | - | - | - | - | - | - | - | - | - | - |
| HSP90B1 | OID2082 | Neurolog | HSP90B1 | P14625 | 12 | 103930107 | 103953931 | 0 | - | - | - | - | - | - | - | - | - | - | - | - | - | - | - | - | - | - | - | - | - | - | - | - | - | - | - | - | - |
| IL31 | OID3048 | Inflamma | IL31 | Q6EBC2 | 12 | 122172029 | 122174221 | 0 | - | - | - | - | - | - | - | - | - | - | - | - | - | - | - | - | - | - | - | - | - | - | - | - | - | - | - | - | - |
| IRAG2 | OID2008 | Cardiom | IRAG2 | Q12912 | 12 | 25004342 | 25108335 | 0 | - | - | - | - | - | - | - | - | - | - | - | - | - | - | - | - | - | - | - | - | - | - | - | - | - | - | - | - | - |
| IRAK4 | OID2057 | Inflamma | IRAK4 | Q9NWZ3 | 12 | 43758944 | 43789543 | 0 | - | - | - | - | - | - | - | - | - | - | - | - | - | - | - | - | - | - | - | - | - | - | - | - | - | - | - | - | - |
| KLRC1 | OID3085 | Neurolog | KLRC1 | P26715 | 12 | 10442264 | 10454685 | 0 | - | - | - | - | - | - | - | - | - | - | - | - | - | - | - | - | - | - | - | - | - | - | - | - | - | - | - | - | - |
| KRT6C | OID3016 | Cardiom | KRT6C | P48668 | 12 | 52468516 | 52473805 | 0 | - | - | - | - | - | - | - | - | - | - | - | - | - | - | - | - | - | - | - | - | - | - | - | - | - | - | - | - | - |
| KRT8 | OID3086 | Neurolog | KRT8 | P05787 | 12 | 52897187 | 52949954 | 0 | - | - | - | - | - | - | - | - | - | - | - | - | - | - | - | - | - | - | - | - | - | - | - | - | - | - | - | - | - |
| LRP1 | OID2129 | Oncology | LRP1 | Q07954 | 12 | 57128483 | 57213361 | 0 | - | - | - | - | - | - | - | - | - | - | - | - | - | - | - | - | - | - | - | - | - | - | - | - | - | - | - | - | - |
| MAP1LC | OID3007 | Cardiom | MAP1LC | A6NCE7 | 12 | 116548105 | 116576606 | 0 | - | - | - | - | - | - | - | - | - | - | - | - | - | - | - | - | - | - | - | - | - | - | - | - | - | - | - | - | - |
| MARS1 | OID3065 | Inflamma | MARS1 | P56192 | 12 | 57475445 | 57517569 | 0 | - | - | - | - | - | - | - | - | - | - | - | - | - | - | - | - | - | - | - | - | - | - | - | - | - | - | - | - | - |
| MED21 | OID3006 | Cardiom | MED21 | Q13503 | 12 | 27022546 | 27066343 | 0 | - | - | - | - | - | - | - | - | - | - | - | - | - | - | - | - | - | - | - | - | - | - | - | - | - | - | - | - | - |
| MYBPC1 | OID3029 | Cardiom | MYBPC1 | Q00872 | 12 | 101568353 | 101686028 | 0 | - | - | - | - | - | - | - | - | - | - | - | - | - | - | - | - | - | - | - | - | - | - | - | - | - | - | - | - | - |
| MYL6B | OID3093 | Neurolog | MYL6B | P14649 | 12 | 56152256 | 56159647 | 0 | - | - | - | - | - | - | - | - | - | - | - | - | - | - | - | - | - | - | - | - | - | - | - | - | - | - | - | - | - |
| PMCH | OID3017 | Cardiom | PMCH | P20382 | 12 | 102196459 | 102197833 | 0 | - | - | - | - | - | - | - | - | - | - | - | - | - | - | - | - | - | - | - | - | - | - | - | - | - | - | - | - | - |
| PPP1CC | OID3136 | Oncology | PPP1CC | P36873 | 12 | 110719680 | 110742939 | 0 | - | - | - | - | - | - | - | - | - | - | - | - | - | - | - | - | - | - | - | - | - | - | - | - | - | - | - | - | - |
| PPP1R12 | OID2147 | Oncology | PPP1R12 | O14974 | 12 | 79773563 | 79935460 | 0 | - | - | - | - | - | - | - | - | - | - | - | - | - | - | - | - | - | - | - | - | - | - | - | - | - | - | - | - | - |
| PTPN6 | OID2071 | Inflamma | PTPN6 | P29350 | 12 | 6946468 | 6961316 | 0 | - | - | - | - | - | - | - | - | - | - | - | - | - | - | - | - | - | - | - | - | - | - | - | - | - | - | - | - | - |
| RBM19 | OID3009 | Cardiom | RBM19 | Q9Y4C8 | 12 | 113816738 | 113966325 | 0 | - | - | - | - | - | - | - | - | - | - | - | - | - | - | - | - | - | - | - | - | - | - | - | - | - | - | - | - | - |
| RNF41 | OID2117 | Oncology | RNF41 | Q9H4P4 | 12 | 56202179 | 56221933 | 0 | - | - | - | - | - | - | - | - | - | - | - | - | - | - | - | - | - | - | - | - | - | - | - | - | - | - | - | - | - |
| SCARB1 | OID2080 | Neurolog | SCARB1 | Q8WTV0 | 12 | 124776856 | 124882668 | 0 | - | - | - | - | - | - | - | - | - | - | - | - | - | - | - | - | - | - | - | - | - | - | - | - | - | - | - | - | - |
| SPRING1 | OID3119 | Oncology | SPRING1 | Q9H741 | 12 | 116710171 | 116738070 | 0 | - | - | - | - | - | - | - | - | - | - | - | - | - | - | - | - | - | - | - | - | - | - | - | - | - | - | - | - | - |
| ST8SIA1 | OID3044 | Inflamma | ST8SIA1 | Q92185 | 12 | 22063773 | 22437041 | 0 | - | - | - | - | - | - | - | - | - | - | - | - | - | - | - | - | - | - | - | - | - | - | - | - | - | - | - | - | - |
| SYT1 | OID3108 | Neurolog | SYT1 | P21579 | 12 | 78863993 | 79452008 | 0 | - | - | - | - | - | - | - | - | - | - | - | - | - | - | - | - | - | - | - | - | - | - | - | - | - | - | - | - | - |
| TRIAP1 | OID2140 | Oncology | TRIAP1 | O43715 | 12 | 120443964 | 120446384 | 0 | - | - | - | - | - | - | - | - | - | - | - | - | - | - | - | - | - | - | - | - | - | - | - | - | - | - | - | - | - |
| UNG | OID3141 | Oncology | UNG | P13051 | 12 | 109097597 | 109126725 | 0 | - | - | - | - | - | - | - | - | - | - | - | - | - | - | - | - | - | - | - | - | - | - | - | - | - | - | - | - | - |
| ZCCHC8 | OID3011 | Cardiom | ZCCHC8 | Q6NZY4 | 12 | 122471599 | 122501073 | 0 | - | - | - | - | - | - | - | - | - | - | - | - | - | - | - | - | - | - | - | - | - | - | - | - | - | - | - | - | - |
| CENPJ | OID3131 | Oncology | CENPJ | Q9HC77 | 13 | 24882279 | 24922889 | 0 | - | - | - | - | - | - | - | - | - | - | - | - | - | - | - | - | - | - | - | - | - | - | - | - | - | - | - | - | - |
| DCUN1D | OID3123 | Oncology | DCUN1D | Q6PH85 | 13 | 113455819 | 113490951 | 0 | - | - | - | - | - | - | - | - | - | - | - | - | - | - | - | - | - | - | - | - | - | - | - | - | - | - | - | - | - |
| DOCK9 | OID3083 | Neurolog | DOCK9 | Q9BZ29 | 13 | 98793429 | 99086625 | 0 | - | - | - | - | - | - | - | - | - | - | - | - | - | - | - | - | - | - | - | - | - | - | - | - | - | - | - | - | - |
| EDNRB | OID3042 | Inflamma | EDNRB | P24530 | 13 | 77895481 | 77975529 | 0 | - | - | - | - | - | - | - | - | - | - | - | - | - | - | - | - | - | - | - | - | - | - | - | - | - | - | - | - | - |
| EFNB2 | OID3095 | Neurolog | EFNB2 | P52799 | 13 | 106489745 | 106535662 | 0 | - | - | - | - | - | - | - | - | - | - | - | - | - | - | - | - | - | - | - | - | - | - | - | - | - | - | - | - | - |
| FGF9 | OID3119 | Oncology | FGF9 | P31371 | 13 | 21671073 | 21704498 | 0 | - | - | - | - | - | - | - | - | - | - | - | - | - | - | - | - | - | - | - | - | - | - | - | - | - | - | - | - | - |
| FLT1 | OID2130 | Oncology | FLT1 | P17948 | 13 | 28300346 | 28495145 | 0 | - | - | - | - | - | - | - | - | - | - | - | - | - | - | - | - | - | - | - | - | - | - | - | - | - | - | - | - | - |
| MYCBP2 | OID3099 | Neurolog | MYCBP2 | O75592 | 13 | 77042474 | 77327094 | 0 | - | - | - | - | - | - | - | - | - | - | - | - | - | - | - | - | - | - | - | - | - | - | - | - | - | - | - | - | - |
| MZT1 | OID2118 | Oncology | MZT1 | Q08AG7 | 13 | 72708367 | 72727629 | 0 | - | - | - | - | - | - | - | - | - | - | - | - | - | - | - | - | - | - | - | - | - | - | - | - | - | - | - | - | - |
| NUDT15 | OID3010 | Cardiom | NUDT15 | Q9NV35 | 13 | 48037726 | 48047221 | 0 | - | - | - | - | - | - | - | - | - | - | - | - | - | - | - | - | - | - | - | - | - | - | - | - | - | - | - | - | - |
| RGCC | OID3106 | Neurolog | RGCC | Q9H4X1 | 13 | 41457550 | 41470871 | 0 | - | - | - | - | - | - | - | - | - | - | - | - | - | - | - | - | - | - | - | - | - | - | - | - | - | - | - | - | - |
| SAP18 | OID3132 | Oncology | SAP18 | O00422 | 13 | 21140119 | 21149097 | 0 | - | - | - | - | - | - | - | - | - | - | - | - | - | - | - | - | - | - | - | - | - | - | - | - | - | - | - | - | - |
| SPART | OID3065 | Inflamma | SPART | Q8N0X7 | 13 | 36301638 | 36370180 | 0 | - | - | - | - | - | - | - | - | - | - | - | - | - | - | - | - | - | - | - | - | - | - | - | - | - | - | - | - | - |
| STK24 | OID2083 | Neurolog | STK24 | Q9Y6E0 | 13 | 98445185 | 98577940 | 0 | - | - | - | - | - | - | - | - | - | - | - | - | - | - | - | - | - | - | - | - | - | - | - | - | - | - | - | - | - |
| SUGT1 | OID2122 | Oncology | SUGT1 | Q9Y2Z0 | 13 | 52652709 | 52700909 | 0 | - | - | - | - | - | - | - | - | - | - | - | - | - | - | - | - | - | - | - | - | - | - | - | - | - | - | - | - | - |
| TPT1 | OID2047 | Inflamma | TPT1 | P13693 | 13 | 45333471 | 45341284 | 0 | - | - | - | - | - | - | - | - | - | - | - | - | - | - | - | - | - | - | - | - | - | - | - | - | - | - | - | - | - |
| AHSA1 | OID3137 | Oncology | AHSA1 | O95433 | 14 | 77457870 | 77469472 | 0 | - | - | - | - | - | - | - | - | - | - | - | - | - | - | - | - | - | - | - | - | - | - | - | - | - | - | - | - | - |
| ARF6 | OID3152 | Oncology | ARF6 | P62330 | 14 | 49893079 | 49897054 | 0 | - | - | - | - | - | - | - | - | - | - | - | - | - | - | - | - | - | - | - | - | - | - | - | - | - | - | - | - | - |
| ARHGAP | OID3119 | Oncology | ARHGAP | Q13017 | 14 | 32076114 | 32159728 | 0 | - | - | - | - | - | - | - | - | - | - | - | - | - | - | - | - | - | - | - | - | - | - | - | - | - | - | - | - | - |
| ATP6V1D | OID2134 | Oncology | ATP6V1D | Q9Y5K8 | 14 | 67294371 | 67360265 | 0 | - | - | - | - | - | - | - | - | - | - | - | - | - | - | - | - | - | - | - | - | - | - | - | - | - | - | - | - | - |
| BATF | OID3089 | Neurolog | BATF | Q16520 | 14 | 75522455 | 75546993 | 0 | - | - | - | - | - | - | - | - | - | - | - | - | - | - | - | - | - | - | - | - | - | - | - | - | - | - | - | - | - |
| BMP4 | OID2099 | Neurolog | BMP4 | P12644 | 14 | 53949736 | 53958761 | 0 | - | - | - | - | - | - | - | - | - | - | - | - | - | - | - | - | - | - | - | - | - | - | - | - | - | - | - | - | - |
| CDC42BI | OID3148 | Oncology | CDC42BI | Q9Y5S2 | 14 | 102932380 | 103057549 | 0 | - | - | - | - | - | - | - | - | - | - | - | - | - | - | - | - | - | - | - | - | - | - | - | - | - | - | - | - | - |
| CINP | OID3132 | Oncology | CINP | Q9BW66 | 14 | 102341102 | 102362916 | 0 | - | - | - | - | - | - | - | - | - | - | - | - | - | - | - | - | - | - | - | - | - | - | - | - | - | - | - | - | - |
| CKB | OID3040 | Cardiom | CKB | P12277 | 14 | 103519667 | 103522833 | 0 | - | - | - | - | - | - | - | - | - | - | - | - | - | - | - | - | - | - | - | - | - | - | - | - | - | - | - | - | - |
| DAAM1 | OID3066 | Inflamma | DAAM1 | Q9Y4D1 | 14 | 59188646 | 59371405 | 0 | - | - | - | - | - | - | - | - | - | - | - | - | - | - | - | - | - | - | - | - | - | - | - | - | - | - | - | - | - |
| DLGAP5 | OID3097 | Neurolog | DLGAP5 | Q15398 | 14 | 55148112 | 55191608 | 0 | - | - | - | - | - | - | - | - | - | - | - | - | - | - | - | - | - | - | - | - | - | - | - | - | - | - | - | - | - |
| DYNC1H | OID3134 | Oncology | DYNC1H | Q14204 | 14 | 101964573 | 102056443 | 0 | - | - | - | - | - | - | - | - | - | - | - | - | - | - | - | - | - | - | - | - | - | - | - | - | - | - | - | - | - |
| EIF5 | OID3015 | Cardiom | EIF5 | P55010 | 14 | 103333544 | 103345025 | 0 | - | - | - | - | - | - | - | - | - | - | - | - | - | - | - | - | - | - | - | - | - | - | - | - | - | - | - | - | - |
| FOS | OID3122 | Oncology | FOS | P01100 | 14 | 75278826 | 75282230 | 0 | - | - | - | - | - | - | - | - | - | - | - | - | - | - | - | - | - | - | - | - | - | - | - | - | - | - | - | - | - |
| HIF1A | OID3042 | Inflamma | HIF1A | Q16665 | 14 | 61695513 | 61748259 | 0 | - | - | - | - | - | - | - | - | - | - | - | - | - | - | - | - | - | - | - | - | - | - | - | - | - | - | - | - | - |
| IL25 | OID3124 | Oncology | IL25 | Q9H293 | 14 | 23372809 | 23376403 | 0 | - | - | - | - | - | - | - | - | - | - | - | - | - | - | - | - | - | - | - | - | - | - | - | - | - | - | - | - | - |
| ISM2 | OID3036 | Cardiom | ISM2 | Q6H9L7 | 14 | 77474394 | 77498816 | 0 | - | - | - | - | - | - | - | - | - | - | - | - | - | - | - | - | - | - | - | - | - | - | - | - | - | - | - | - | - |
| L3HYPDH | OID3031 | Cardiom | L3HYPDH | Q96EM0 | 14 | 59460363 | 59484408 | 0 | - | - | - | - | - | - | - | - | - | - | - | - | - | - | - | - | - | - | - | - | - | - | - | - | - | - | - | - | - |
| MNAT1 | OID3132 | Oncology | MNAT1 | P51948 | 14 | 60734742 | 60969965 | 0 | - | - | - | - | - | - | - | - | - | - | - | - | - | - | - | - | - | - | - | - | - | - | - | - | - | - | - | - | - |
| MRPL52 | OID3123 | Oncology | MRPL52 | Q86TS9 | 14 | 22829879 | 22835037 | 0 | - | - | - | - | - | - | - | - | - | - | - | - | - | - | - | - | - | - | - | - | - | - | - | - | - | - | - | - | - |
| NRXN3 | OID3090 | Neurolog | NRXN3 | Q9Y4C0 | 14 | 78170373 | 79868291 | 0 | - | - | - | - | - | - | - | - | - | - | - | - | - | - | - | - | - | - | - | - | - | - | - | - | - | - | - | - | - |
| PPM1A | OID2118 | Oncology | PPM1A | P35813 | 14 | 60245752 | 60299087 | 0 | - | - | - | - | - | - | - | - | - | - | - | - | - | - | - | - | - | - | - | - | - | - | - | - | - | - | - | - | - |
| RBM25 | OID3123 | Oncology | RBM25 | P49756 | 14 | 73058532 | 73123899 | 0 | - | - | - | - | - | - | - | - | - | - | - | - | - | - | - | - | - | - | - | - | - | - | - | - | - | - | - | - | - |
| RCOR1 | OID2007 | Cardiom | RCOR1 | Q9UKL0 | 14 | 102592649 | 102730561 | 0 | - | - | - | - | - | - | - | - | - | - | - | - | - | - | - | - | - | - | - | - | - | - | - | - | - | - | - | - | - |
| RNF31 | OID3047 | Inflamma | RNF31 | Q96EP0 | 14 | 24146683 | 24160660 | 0 | - | - | - | - | - | - | - | - | - | - | - | - | - | - | - | - | - | - | - | - | - | - | - | - | - | - | - | - | - |
| SEL1L | OID3019 | Cardiom | SEL1L | Q9UBV2 | 14 | 81471547 | 81533853 | 0 | - | - | - | - | - | - | - | - | - | - | - | - | - | - | - | - | - | - | - | - | - | - | - | - | - | - | - | - | - |
| SLIRP | OID3117 | Oncology | SLIRP | Q9GZT3 | 14 | 77708071 | 77761104 | 0 | - | - | - | - | - | - | - | - | - | - | - | - | - | - | - | - | - | - | - | - | - | - | - | - | - | - | - | - | - |
| TCL1B | OID2005 | Cardiom | TCL1B | O95988 | 14 | 95686426 | 95692628 | 0 | - | - | - | - | - | - | - | - | - | - | - | - | - | - | - | - | - | - | - | - | - | - | - | - | - | - | - | - | - |
| TMED10 | OID3006 | Cardiom | TMED10 | P49755 | 14 | 75131469 | 75176612 | 0 | - | - | - | - | - | - | - | - | - | - | - | - | - | - | - | - | - | - | - | - | - | - | - | - | - | - | - | - | - |
| TPPP2 | OID3127 | Oncology | TPPP2 | P59282 | 14 | 21024109 | 21036276 | 0 | - | - | - | - | - | - | - | - | - | - | - | - | - | - | - | - | - | - | - | - | - | - | - | - | - | - | - | - | - |
| TRAF3 | OID3047 | Inflamma | TRAF3 | Q13114 | 14 | 102777449 | 102911500 | 0 | - | - | - | - | - | - | - | - | - | - | - | - | - | - | - | - | - | - | - | - | - | - | - | - | - | - | - | - | - |
| UNC79 | OID3118 | Oncology | UNC79 | Q9P2D8 | 14 | 93333219 | 93707876 | 0 | - | - | - | - | - | - | - | - | - | - | - | - | - | - | - | - | - | - | - | - | - | - | - | - | - | - | - | - | - |
| YY1 | OID3044 | Inflamma | YY1 | P25490 | 14 | 100238298 | 100282788 | 0 | - | - | - | - | - | - | - | - | - | - | - | - | - | - | - | - | - | - | - | - | - | - | - | - | - | - | - | - | - |
| AP3S2 | OID3111 | Neurolog | AP3S2 | P59780 | 15 | 89830599 | 89894638 | 0 | - | - | - | - | - | - | - | - | - | - | - | - | - | - | - | - | - | - | - | - | - | - | - | - | - | - | - | - | - |
| B2M | OID3072 | Inflamma | B2M | P61769 | 15 | 44711358 | 44718851 | 0 | - | - | - | - | - | - | - | - | - | - | - | - | - | - | - | - | - | - | - | - | - | - | - | - | - | - | - | - | - |
| BNIP2 | OID3029 | Cardiom | BNIP2 | Q12982 | 15 | 59659146 | 59689339 | 0 | - | - | - | - | - | - | - | - | - | - | - | - | - | - | - | - | - | - | - | - | - | - | - | - | - | - | - | - | - |
| CDAN1 | OID3107 | Neurolog | CDAN1 | Q8IWY9 | 15 | 42723544 | 42737128 | 0 | - | - | - | - | - | - | - | - | - | - | - | - | - | - | - | - | - | - | - | - | - | - | - | - | - | - | - | - | - |
| CHP1 | OID3086 | Neurolog | CHP1 | Q99653 | 15 | 41230839 | 41281887 | 0 | - | - | - | - | - | - | - | - | - | - | - | - | - | - | - | - | - | - | - | - | - | - | - | - | - | - | - | - | - |
| DLL4 | OID3025 | Cardiom | DLL4 | Q9NR61 | 15 | 40929340 | 40939073 | 0 | - | - | - | - | - | - | - | - | - | - | - | - | - | - | - | - | - | - | - | - | - | - | - | - | - | - | - | - | - |
| DUOX2 | OID2005 | Cardiom | DUOX2 | Q9NRD8 | 15 | 45092650 | 45114172 | 0 | - | - | - | - | - | - | - | - | - | - | - | - | - | - | - | - | - | - | - | - | - | - | - | - | - | - | - | - | - |
| DUT | OID3145 | Oncology | DUT | P33316 | 15 | 48331011 | 48343373 | 0 | - | - | - | - | - | - | - | - | - | - | - | - | - | - | - | - | - | - | - | - | - | - | - | - | - | - | - | - | - |
| FGF7 | OID3124 | Oncology | FGF7 | P21781 | 15 | 49423237 | 49488775 | 0 | - | - | - | - | - | - | - | - | - | - | - | - | - | - | - | - | - | - | - | - | - | - | - | - | - | - | - | - | - |
| IGDCC3 | OID3086 | Neurolog | IGDCC3 | Q8IVU1 | 15 | 65327127 | 65378002 | 0 | - | - | - | - | - | - | - | - | - | - | - | - | - | - | - | - | - | - | - | - | - | - | - | - | - | - | - | - | - |
| IVD | OID2083 | Neurolog | IVD | P26440 | 15 | 40405485 | 40435947 | 0 | - | - | - | - | - | - | - | - | - | - | - | - | - | - | - | - | - | - | - | - | - | - | - | - | - | - | - | - | - |
| LEO1 | OID3099 | Neurolog | LEO1 | Q8WVC0 | 15 | 51938025 | 51971778 | 0 | - | - | - | - | - | - | - | - | - | - | - | - | - | - | - | - | - | - | - | - | - | - | - | - | - | - | - | - | - |
| MORF4L1 | OID3125 | Oncology | MORF4L1 | Q9UBU8 | 15 | 78810487 | 78898139 | 0 | - | - | - | - | - | - | - | - | - | - | - | - | - | - | - | - | - | - | - | - | - | - | - | - | - | - | - | - | - |
| MRPL46 | OID2085 | Neurolog | MRPL46 | Q9H2W6 | 15 | 88459477 | 88467390 | 0 | - | - | - | - | - | - | - | - | - | - | - | - | - | - | - | - | - | - | - | - | - | - | - | - | - | - | - | - | - |
| NGRN | OID3111 | Neurolog | NGRN | Q9NPE2 | 15 | 90265659 | 90275778 | 0 | - | - | - | - | - | - | - | - | - | - | - | - | - | - | - | - | - | - | - | - | - | - | - | - | - | - | - | - | - |
| NPTN | OID2123 | Oncology | NPTN | Q9Y639 | 15 | 73560014 | 73634134 | 0 | - | - | - | - | - | - | - | - | - | - | - | - | - | - | - | - | - | - | - | - | - | - | - | - | - | - | - | - | - |
| PLIN1 | OID2083 | Neurolog | PLIN1 | O60240 | 15 | 89664367 | 89679427 | 0 | - | - | - | - | - | - | - | - | - | - | - | - | - | - | - | - | - | - | - | - | - | - | - | - | - | - | - | - | - |
| PPIB | OID2033 | Cardiom | PPIB | P23284 | 15 | 64155740 | 64163134 | 0 | - | - | - | - | - | - | - | - | - | - | - | - | - | - | - | - | - | - | - | - | - | - | - | - | - | - | - | - | - |
| PRC1 | OID3134 | Oncology | PRC1 | O43663 | 15 | 90966040 | 90995629 | 0 | - | - | - | - | - | - | - | - | - | - | - | - | - | - | - | - | - | - | - | - | - | - | - | - | - | - | - | - | - |
| PTPN9 | OID3043 | Inflamma | PTPN9 | P43378 | 15 | 75463251 | 75579315 | 0 | - | - | - | - | - | - | - | - | - | - | - | - | - | - | - | - | - | - | - | - | - | - | - | - | - | - | - | - | - |
| RAD51 | OID3125 | Oncology | RAD51 | Q06609 | 15 | 40694774 | 40732340 | 0 | - | - | - | - | - | - | - | - | - | - | - | - | - | - | - | - | - | - | - | - | - | - | - | - | - | - | - | - | - |
| RASGRF | OID3121 | Oncology | RASGRF | Q13972 | 15 | 78959906 | 79090780 | 0 | - | - | - | - | - | - | - | - | - | - | - | - | - | - | - | - | - | - | - | - | - | - | - | - | - | - | - | - | - |
| SH3GL3 | OID3133 | Oncology | SH3GL3 | Q99963 | 15 | 83447228 | 83618743 | 0 | - | - | - | - | - | - | - | - | - | - | - | - | - | - | - | - | - | - | - | - | - | - | - | - | - | - | - | - | - |
| SLC28A1 | OID3122 | Oncology | SLC28A1 | O00337 | 15 | 84884654 | 84945798 | 0 | - | - | - | - | - | - | - | - | - | - | - | - | - | - | - | - | - | - | - | - | - | - | - | - | - | - | - | - | - |
| SLC51B | OID3010 | Cardiom | SLC51B | Q86UW2 | 15 | 65045387 | 65053397 | 0 | - | - | - | - | - | - | - | - | - | - | - | - | - | - | - | - | - | - | - | - | - | - | - | - | - | - | - | - | - |
| SNAP23 | OID2021 | Cardiom | SNAP23 | O00161 | 15 | 42491233 | 42545356 | 0 | - | - | - | - | - | - | - | - | - | - | - | - | - | - | - | - | - | - | - | - | - | - | - | - | - | - | - | - | - |
| TMCO5A | OID3094 | Neurolog | TMCO5A | Q8N6Q1 | 15 | 37921939 | 37967724 | 0 | - | - | - | - | - | - | - | - | - | - | - | - | - | - | - | - | - | - | - | - | - | - | - | - | - | - | - | - | - |
| TP53BP1 | OID3047 | Inflamma | TP53BP1 | Q12888 | 15 | 43403061 | 43510728 | 0 | - | - | - | - | - | - | - | - | - | - | - | - | - | - | - | - | - | - | - | - | - | - | - | - | - | - | - | - | - |
| AMDHD2 | OID3036 | Cardiom | AMDHD2 | Q9Y303 | 16 | 2520357 | 2531422 | 0 | - | - | - | - | - | - | - | - | - | - | - | - | - | - | - | - | - | - | - | - | - | - | - | - | - | - | - | - | - |
| CA7 | OID3126 | Oncology | CA7 | P43166 | 16 | 66844414 | 66854153 | 0 | - | - | - | - | - | - | - | - | - | - | - | - | - | - | - | - | - | - | - | - | - | - | - | - | - | - | - | - | - |
| CACNA1 | OID3006 | Cardiom | CACNA1 | O95180 | 16 | 1153106 | 1221771 | 0 | - | - | - | - | - | - | - | - | - | - | - | - | - | - | - | - | - | - | - | - | - | - | - | - | - | - | - | - | - |
| CBLN1 | OID3085 | Neurolog | CBLN1 | P23435 | 16 | 49277917 | 49281838 | 0 | - | - | - | - | - | - | - | - | - | - | - | - | - | - | - | - | - | - | - | - | - | - | - | - | - | - | - | - | - |
| CMIP | OID3140 | Oncology | CMIP | Q8IY22 | 16 | 81444808 | 81711762 | 0 | - | - | - | - | - | - | - | - | - | - | - | - | - | - | - | - | - | - | - | - | - | - | - | - | - | - | - | - | - |
| CORO1A | OID2028 | Cardiom | CORO1A | P31146 | 16 | 30182827 | 30189076 | 0 | - | - | - | - | - | - | - | - | - | - | - | - | - | - | - | - | - | - | - | - | - | - | - | - | - | - | - | - | - |
| DHODH | OID3013 | Cardiom | DHODH | Q02127 | 16 | 72008588 | 72027664 | 0 | - | - | - | - | - | - | - | - | - | - | - | - | - | - | - | - | - | - | - | - | - | - | - | - | - | - | - | - | - |
| DNAJA2 | OID2062 | Inflamma | DNAJA2 | O60884 | 16 | 46955362 | 46973674 | 0 | - | - | - | - | - | - | - | - | - | - | - | - | - | - | - | - | - | - | - | - | - | - | - | - | - | - | - | - | - |
| ELOB | OID3012 | Cardiom | ELOB | Q15370 | 16 | 2771414 | 2777289 | 0 | - | - | - | - | - | - | - | - | - | - | - | - | - | - | - | - | - | - | - | - | - | - | - | - | - | - | - | - | - |
| FUS | OID2125 | Oncology | FUS | P35637 | 16 | 31180138 | 31191605 | 0 | - | - | - | - | - | - | - | - | - | - | - | - | - | - | - | - | - | - | - | - | - | - | - | - | - | - | - | - | - |
| GFOD2 | OID2121 | Oncology | GFOD2 | Q3B7J2 | 16 | 67674531 | 67719339 | 0 | - | - | - | - | - | - | - | - | - | - | - | - | - | - | - | - | - | - | - | - | - | - | - | - | - | - | - | - | - |
| GLYR1 | OID3136 | Oncology | GLYR1 | Q49A26 | 16 | 4803203 | 4847288 | 0 | - | - | - | - | - | - | - | - | - | - | - | - | - | - | - | - | - | - | - | - | - | - | - | - | - | - | - | - | - |
| IL21R | OID3044 | Inflamma | IL21R | Q9HBE5 | 16 | 27402174 | 27452042 | 0 | - | - | - | - | - | - | - | - | - | - | - | - | - | - | - | - | - | - | - | - | - | - | - | - | - | - | - | - | - |
| IST1 | OID3149 | Oncology | IST1 | P53990 | 16 | 71845996 | 71931199 | 0 | - | - | - | - | - | - | - | - | - | - | - | - | - | - | - | - | - | - | - | - | - | - | - | - | - | - | - | - | - |
| ITGAX | OID3132 | Oncology | ITGAX | P20702 | 16 | 31355134 | 31382999 | 0 | - | - | - | - | - | - | - | - | - | - | - | - | - | - | - | - | - | - | - | - | - | - | - | - | - | - | - | - | - |
| KCTD5 | OID3114 | Neurolog | KCTD5 | Q9NXV2 | 16 | 2682523 | 2709030 | 0 | - | - | - | - | - | - | - | - | - | - | - | - | - | - | - | - | - | - | - | - | - | - | - | - | - | - | - | - | - |
| KIF22 | OID3017 | Cardiom | KIF22 | Q14807 | 16 | 29790727 | 29805385 | 0 | - | - | - | - | - | - | - | - | - | - | - | - | - | - | - | - | - | - | - | - | - | - | - | - | - | - | - | - | - |
| MMP15 | OID3029 | Cardiom | MMP15 | P51511 | 16 | 58025754 | 58046901 | 0 | - | - | - | - | - | - | - | - | - | - | - | - | - | - | - | - | - | - | - | - | - | - | - | - | - | - | - | - | - |
| MRPL28 | OID3102 | Neurolog | MRPL28 | Q13084 | 16 | 366969 | 371289 | 0 | - | - | - | - | - | - | - | - | - | - | - | - | - | - | - | - | - | - | - | - | - | - | - | - | - | - | - | - | - |
| MSLNL | OID3093 | Neurolog | MSLNL | Q96KJ4 | 16 | 769428 | 783370 | 0 | - | - | - | - | - | - | - | - | - | - | - | - | - | - | - | - | - | - | - | - | - | - | - | - | - | - | - | - | - |
| MYLPF | OID3091 | Neurolog | MYL11 | Q96A32 | 16 | 30370934 | 30377991 | 0 | - | - | - | - | - | - | - | - | - | - | - | - | - | - | - | - | - | - | - | - | - | - | - | - | - | - | - | - | - |
| NFAT5 | OID3052 | Inflamma | NFAT5 | O94916 | 16 | 69565094 | 69704666 | 0 | - | - | - | - | - | - | - | - | - | - | - | - | - | - | - | - | - | - | - | - | - | - | - | - | - | - | - | - | - |
| NFATC3 | OID2047 | Inflamma | NFATC3 | Q12968 | 16 | 68084751 | 68229259 | 0 | - | - | - | - | - | - | - | - | - | - | - | - | - | - | - | - | - | - | - | - | - | - | - | - | - | - | - | - | - |
| NUBP1 | OID3141 | Oncology | NUBP1 | P53384 | 16 | 10743786 | 10769351 | 0 | - | - | - | - | - | - | - | - | - | - | - | - | - | - | - | - | - | - | - | - | - | - | - | - | - | - | - | - | - |
| PAGR1 | OID3020 | Cardiom | PAGR1 | Q9BTK6 | 16 | 29816152 | 29822489 | 0 | - | - | - | - | - | - | - | - | - | - | - | - | - | - | - | - | - | - | - | - | - | - | - | - | - | - | - | - | - |
| PDIA2 | OID3121 | Oncology | PDIA2 | Q13087 | 16 | 283164 | 287215 | 0 | - | - | - | - | - | - | - | - | - | - | - | - | - | - | - | - | - | - | - | - | - | - | - | - | - | - | - | - | - |
| PDXDC1 | OID3122 | Oncology | PDXDC1 | Q6P996 | 16 | 14974591 | 15139339 | 0 | - | - | - | - | - | - | - | - | - | - | - | - | - | - | - | - | - | - | - | - | - | - | - | - | - | - | - | - | - |
| STX1B | OID3093 | Neurolog | STX1B | P61266 | 16 | 30989256 | 31010638 | 0 | - | - | - | - | - | - | - | - | - | - | - | - | - | - | - | - | - | - | - | - | - | - | - | - | - | - | - | - | - |
| TPSG1 | OID3042 | Inflamma | TPSG1 | Q9NRR2 | 16 | 1221651 | 1225268 | 0 | - | - | - | - | - | - | - | - | - | - | - | - | - | - | - | - | - | - | - | - | - | - | - | - | - | - | - | - | - |
| TUBB3 | OID3079 | Neurolog | TUBB3 | Q13509 | 16 | 89921392 | 89938761 | 0 | - | - | - | - | - | - | - | - | - | - | - | - | - | - | - | - | - | - | - | - | - | - | - | - | - | - | - | - | - |
| ZNF174 | OID3050 | Inflamma | ZNF174 | Q15697 | 16 | 3401215 | 3409364 | 0 | - | - | - | - | - | - | - | - | - | - | - | - | - | - | - | - | - | - | - | - | - | - | - | - | - | - | - | - | - |
| AP2B1 | OID3094 | Neurolog | AP2B1 | P63010 | 17 | 35578046 | 35726413 | 0 | - | - | - | - | - | - | - | - | - | - | - | - | - | - | - | - | - | - | - | - | - | - | - | - | - | - | - | - | - |
| ATP1B2 | OID3011 | Cardiom | ATP1B2 | P14415 | 17 | 7646627 | 7657770 | 0 | - | - | - | - | - | - | - | - | - | - | - | - | - | - | - | - | - | - | - | - | - | - | - | - | - | - | - | - | - |
| BECN1 | OID3022 | Cardiom | BECN1 | Q14457 | 17 | 42810134 | 42833350 | 0 | - | - | - | - | - | - | - | - | - | - | - | - | - | - | - | - | - | - | - | - | - | - | - | - | - | - | - | - | - |
| C1QBP | OID3100 | Neurolog | C1QBP | Q07021 | 17 | 5432777 | 5448830 | 0 | - | - | - | - | - | - | - | - | - | - | - | - | - | - | - | - | - | - | - | - | - | - | - | - | - | - | - | - | - |
| CACNB1 | OID3082 | Neurolog | CACNB1 | Q02641 | 17 | 39173453 | 39197702 | 0 | - | - | - | - | - | - | - | - | - | - | - | - | - | - | - | - | - | - | - | - | - | - | - | - | - | - | - | - | - |
| CASC3 | OID3092 | Neurolog | CASC3 | O15234 | 17 | 40140318 | 40172171 | 0 | - | - | - | - | - | - | - | - | - | - | - | - | - | - | - | - | - | - | - | - | - | - | - | - | - | - | - | - | - |
| CBX2 | OID3017 | Cardiom | CBX2 | Q14781 | 17 | 79778148 | 79787983 | 0 | - | - | - | - | - | - | - | - | - | - | - | - | - | - | - | - | - | - | - | - | - | - | - | - | - | - | - | - | - |
| CDK5RA | OID3098 | Neurolog | CDK5RA | Q96JB5 | 17 | 47967810 | 47981781 | 0 | - | - | - | - | - | - | - | - | - | - | - | - | - | - | - | - | - | - | - | - | - | - | - | - | - | - | - | - | - |
| CORO6 | OID3087 | Neurolog | CORO6 | Q6QEF8 | 17 | 29614756 | 29622912 | 0 | - | - | - | - | - | - | - | - | - | - | - | - | - | - | - | - | - | - | - | - | - | - | - | - | - | - | - | - | - |
| CRHR1 | OID2005 | Cardiom | CRHR1 | P34998 | 17 | 45784280 | 45835828 | 0 | - | - | - | - | - | - | - | - | - | - | - | - | - | - | - | - | - | - | - | - | - | - | - | - | - | - | - | - | - |
| CSH1 | OID3066 | Inflamma | CSH1 | P0DML2 | 17 | 63894909 | 63896661 | 0 | - | - | - | - | - | - | - | - | - | - | - | - | - | - | - | - | - | - | - | - | - | - | - | - | - | - | - | - | - |
| CSNK1D | OID3049 | Inflamma | CSNK1D | P48730 | 17 | 82239023 | 82273700 | 0 | - | - | - | - | - | - | - | - | - | - | - | - | - | - | - | - | - | - | - | - | - | - | - | - | - | - | - | - | - |
| DLG4 | OID3107 | Neurolog | DLG4 | P78352 | 17 | 7187187 | 7219841 | 0 | - | - | - | - | - | - | - | - | - | - | - | - | - | - | - | - | - | - | - | - | - | - | - | - | - | - | - | - | - |
| DRG2 | OID2133 | Oncology | DRG2 | P55039 | 17 | 18087892 | 18107970 | 0 | - | - | - | - | - | - | - | - | - | - | - | - | - | - | - | - | - | - | - | - | - | - | - | - | - | - | - | - | - |
| DUSP3 | OID2082 | Neurolog | DUSP3 | P51452 | 17 | 43766125 | 43778977 | 0 | - | - | - | - | - | - | - | - | - | - | - | - | - | - | - | - | - | - | - | - | - | - | - | - | - | - | - | - | - |
| EIF5A | OID2042 | Inflamma | EIF5A | P63241 | 17 | 7306999 | 7312463 | 0 | - | - | - | - | - | - | - | - | - | - | - | - | - | - | - | - | - | - | - | - | - | - | - | - | - | - | - | - | - |
| EVI2B | OID3122 | Oncology | EVI2B | P34910 | 17 | 31303770 | 31314105 | 0 | - | - | - | - | - | - | - | - | - | - | - | - | - | - | - | - | - | - | - | - | - | - | - | - | - | - | - | - | - |
| EVPL | OID3130 | Oncology | EVPL | Q92817 | 17 | 76004502 | 76027306 | 0 | - | - | - | - | - | - | - | - | - | - | - | - | - | - | - | - | - | - | - | - | - | - | - | - | - | - | - | - | - |
| FAM171A | OID3100 | Neurolog | FAM171A | A8MVW0 | 17 | 44353215 | 44363853 | 0 | - | - | - | - | - | - | - | - | - | - | - | - | - | - | - | - | - | - | - | - | - | - | - | - | - | - | - | - | - |
| FMNL1 | OID2088 | Neurolog | FMNL1 | O95466 | 17 | 45221444 | 45247319 | 0 | - | - | - | - | - | - | - | - | - | - | - | - | - | - | - | - | - | - | - | - | - | - | - | - | - | - | - | - | - |
| GABARA | OID3141 | Oncology | GABARA | O95166 | 17 | 7240008 | 7242449 | 0 | - | - | - | - | - | - | - | - | - | - | - | - | - | - | - | - | - | - | - | - | - | - | - | - | - | - | - | - | - |
| GAST | OID3108 | Neurolog | GAST | P01350 | 17 | 41712331 | 41715969 | 0 | - | - | - | - | - | - | - | - | - | - | - | - | - | - | - | - | - | - | - | - | - | - | - | - | - | - | - | - | - |
| GH1 | OID2022 | Cardiom | GH1 | P01241 | 17 | 63917200 | 63918839 | 0 | - | - | - | - | - | - | - | - | - | - | - | - | - | - | - | - | - | - | - | - | - | - | - | - | - | - | - | - | - |
| GH2 | OID2116 | Oncology | GH2 | P01242 | 17 | 63880215 | 63881944 | 0 | - | - | - | - | - | - | - | - | - | - | - | - | - | - | - | - | - | - | - | - | - | - | - | - | - | - | - | - | - |
| GLOD4 | OID2071 | Inflamma | GLOD4 | Q9HC38 | 17 | 757097 | 782294 | 0 | - | - | - | - | - | - | - | - | - | - | - | - | - | - | - | - | - | - | - | - | - | - | - | - | - | - | - | - | - |
| HGS | OID2117 | Oncology | HGS | O14964 | 17 | 81683326 | 81703138 | 0 | - | - | - | - | - | - | - | - | - | - | - | - | - | - | - | - | - | - | - | - | - | - | - | - | - | - | - | - | - |
| IFT20 | OID3096 | Neurolog | IFT20 | Q8IY31 | 17 | 28328325 | 28335489 | 0 | - | - | - | - | - | - | - | - | - | - | - | - | - | - | - | - | - | - | - | - | - | - | - | - | - | - | - | - | - |
| KRT14 | OID2080 | Neurolog | KRT14 | P02533 | 17 | 41582279 | 41586895 | 0 | - | - | - | - | - | - | - | - | - | - | - | - | - | - | - | - | - | - | - | - | - | - | - | - | - | - | - | - | - |
| KRT17 | OID3009 | Cardiom | KRT17 | Q04695 | 17 | 41619442 | 41624842 | 0 | - | - | - | - | - | - | - | - | - | - | - | - | - | - | - | - | - | - | - | - | - | - | - | - | - | - | - | - | - |
| LRRC59 | OID3036 | Cardiom | LRRC59 | Q96AG4 | 17 | 50375059 | 50397523 | 0 | - | - | - | - | - | - | - | - | - | - | - | - | - | - | - | - | - | - | - | - | - | - | - | - | - | - | - | - | - |
| MAPT | OID2083 | Neurolog | MAPT | P10636 | 17 | 45894527 | 46028334 | 0 | - | - | - | - | - | - | - | - | - | - | - | - | - | - | - | - | - | - | - | - | - | - | - | - | - | - | - | - | - |
| MPRIP | OID3142 | Oncology | MPRIP | Q6WCQ1 | 17 | 17042457 | 17217679 | 0 | - | - | - | - | - | - | - | - | - | - | - | - | - | - | - | - | - | - | - | - | - | - | - | - | - | - | - | - | - |
| MRPL58 | OID3103 | Neurolog | MRPL58 | Q14197 | 17 | 75012670 | 75021261 | 0 | - | - | - | - | - | - | - | - | - | - | - | - | - | - | - | - | - | - | - | - | - | - | - | - | - | - | - | - | - |
| MYH4 | OID3006 | Cardiom | MYH4 | Q9Y623 | 17 | 10443290 | 10469559 | 0 | - | - | - | - | - | - | - | - | - | - | - | - | - | - | - | - | - | - | - | - | - | - | - | - | - | - | - | - | - |
| MYL4 | OID3024 | Cardiom | MYL4 | P12829 | 17 | 47200446 | 47223679 | 0 | - | - | - | - | - | - | - | - | - | - | - | - | - | - | - | - | - | - | - | - | - | - | - | - | - | - | - | - | - |
| NGFR | OID3030 | Cardiom | NGFR | P08138 | 17 | 49495293 | 49515008 | 0 | - | - | - | - | - | - | - | - | - | - | - | - | - | - | - | - | - | - | - | - | - | - | - | - | - | - | - | - | - |
| NLGN2 | OID3087 | Neurolog | NLGN2 | Q8NFZ4 | 17 | 7404874 | 7419860 | 0 | - | - | - | - | - | - | - | - | - | - | - | - | - | - | - | - | - | - | - | - | - | - | - | - | - | - | - | - | - |
| NME1 | OID3045 | Inflamma | NME1 | P15531 | 17 | 51153559 | 51162428 | 0 | - | - | - | - | - | - | - | - | - | - | - | - | - | - | - | - | - | - | - | - | - | - | - | - | - | - | - | - | - |
| NMT1 | OID3102 | Neurolog | NMT1 | P30419 | 17 | 44957992 | 45109016 | 0 | - | - | - | - | - | - | - | - | - | - | - | - | - | - | - | - | - | - | - | - | - | - | - | - | - | - | - | - | - |
| PCYT2 | OID3151 | Oncology | PCYT2 | Q99447 | 17 | 81900958 | 81911432 | 0 | - | - | - | - | - | - | - | - | - | - | - | - | - | - | - | - | - | - | - | - | - | - | - | - | - | - | - | - | - |
| PHOSPH | OID2092 | Neurolog | PHOSPH | Q8TCT1 | 17 | 49223366 | 49230787 | 0 | - | - | - | - | - | - | - | - | - | - | - | - | - | - | - | - | - | - | - | - | - | - | - | - | - | - | - | - | - |
| POLR2A | OID3044 | Inflamma | POLR2A | P24928 | 17 | 7484366 | 7514616 | 0 | - | - | - | - | - | - | - | - | - | - | - | - | - | - | - | - | - | - | - | - | - | - | - | - | - | - | - | - | - |
| PPP1R9B | OID2064 | Inflamma | PPP1R9B | Q96SB3 | 17 | 50133737 | 50150677 | 0 | - | - | - | - | - | - | - | - | - | - | - | - | - | - | - | - | - | - | - | - | - | - | - | - | - | - | - | - | - |
| PPY | OID2151 | Oncology | PPY | P01298 | 17 | 43940804 | 43942476 | 0 | - | - | - | - | - | - | - | - | - | - | - | - | - | - | - | - | - | - | - | - | - | - | - | - | - | - | - | - | - |
| PRKAR1A | OID2014 | Cardiom | PRKAR1A | P10644 | 17 | 68511780 | 68551319 | 0 | - | - | - | - | - | - | - | - | - | - | - | - | - | - | - | - | - | - | - | - | - | - | - | - | - | - | - | - | - |
| RAB37 | OID2042 | Inflamma | RAB37 | Q96AX2 | 17 | 74670578 | 74747335 | 0 | - | - | - | - | - | - | - | - | - | - | - | - | - | - | - | - | - | - | - | - | - | - | - | - | - | - | - | - | - |
| RAC3 | OID3095 | Neurolog | RAC3 | P60763 | 17 | 82031678 | 82034204 | 0 | - | - | - | - | - | - | - | - | - | - | - | - | - | - | - | - | - | - | - | - | - | - | - | - | - | - | - | - | - |
| RBFOX3 | OID3098 | Neurolog | RBFOX3 | A6NFN3 | 17 | 79089345 | 79611051 | 0 | - | - | - | - | - | - | - | - | - | - | - | - | - | - | - | - | - | - | - | - | - | - | - | - | - | - | - | - | - |
| RILP | OID2143 | Oncology | RILP | Q96NA2 | 17 | 1646145 | 1649866 | 0 | - | - | - | - | - | - | - | - | - | - | - | - | - | - | - | - | - | - | - | - | - | - | - | - | - | - | - | - | - |
| SOX9 | OID3090 | Neurolog | SOX9 | P48436 | 17 | 72121020 | 72126416 | 0 | - | - | - | - | - | - | - | - | - | - | - | - | - | - | - | - | - | - | - | - | - | - | - | - | - | - | - | - | - |
| STAT5B | OID2127 | Oncology | STAT5B | P51692 | 17 | 42199177 | 42276707 | 0 | - | - | - | - | - | - | - | - | - | - | - | - | - | - | - | - | - | - | - | - | - | - | - | - | - | - | - | - | - |
| TRPV3 | OID3012 | Cardiom | TRPV3 | Q8NET8 | 17 | 3510502 | 3557812 | 0 | - | - | - | - | - | - | - | - | - | - | - | - | - | - | - | - | - | - | - | - | - | - | - | - | - | - | - | - | - |
| UBE2Z | OID3042 | Inflamma | UBE2Z | Q9H832 | 17 | 48908407 | 48929056 | 0 | - | - | - | - | - | - | - | - | - | - | - | - | - | - | - | - | - | - | - | - | - | - | - | - | - | - | - | - | - |
| VPS53 | OID2128 | Oncology | VPS53 | Q5VIR6 | 17 | 508503 | 721717 | 0 | - | - | - | - | - | - | - | - | - | - | - | - | - | - | - | - | - | - | - | - | - | - | - | - | - | - | - | - | - |
| ZNF830 | OID3135 | Oncology | ZNF830 | Q96NB3 | 17 | 34961540 | 34963777 | 0 | - | - | - | - | - | - | - | - | - | - | - | - | - | - | - | - | - | - | - | - | - | - | - | - | - | - | - | - | - |
| CYB5A | OID3117 | Oncology | CYB5A | P00167 | 18 | 74250846 | 74291973 | 0 | - | - | - | - | - | - | - | - | - | - | - | - | - | - | - | - | - | - | - | - | - | - | - | - | - | - | - | - | - |
| DIPK1C | OID3093 | Neurolog | DIPK1C | Q0P6D2 | 18 | 74434775 | 74457944 | 0 | - | - | - | - | - | - | - | - | - | - | - | - | - | - | - | - | - | - | - | - | - | - | - | - | - | - | - | - | - |
| IMPACT | OID3111 | Neurolog | IMPACT | Q9P2X3 | 18 | 24426634 | 24453531 | 0 | - | - | - | - | - | - | - | - | - | - | - | - | - | - | - | - | - | - | - | - | - | - | - | - | - | - | - | - | - |
| LAMA1 | OID3005 | Cardiom | LAMA1 | P25391 | 18 | 6941742 | 7117797 | 0 | - | - | - | - | - | - | - | - | - | - | - | - | - | - | - | - | - | - | - | - | - | - | - | - | - | - | - | - | - |
| MYOM1 | OID3083 | Neurolog | MYOM1 | P52179 | 18 | 3066807 | 3219968 | 0 | - | - | - | - | - | - | - | - | - | - | - | - | - | - | - | - | - | - | - | - | - | - | - | - | - | - | - | - | - |
| NARS1 | OID3103 | Neurolog | NARS1 | O43776 | 18 | 57600656 | 57622213 | 0 | - | - | - | - | - | - | - | - | - | - | - | - | - | - | - | - | - | - | - | - | - | - | - | - | - | - | - | - | - |
| NEDD4L | OID3052 | Inflamma | NEDD4L | Q96PU5 | 18 | 58044226 | 58401540 | 0 | - | - | - | - | - | - | - | - | - | - | - | - | - | - | - | - | - | - | - | - | - | - | - | - | - | - | - | - | - |
| RAB27B | OID3035 | Cardiom | RAB27B | O00194 | 18 | 54717860 | 54895516 | 0 | - | - | - | - | - | - | - | - | - | - | - | - | - | - | - | - | - | - | - | - | - | - | - | - | - | - | - | - | - |
| SIGLEC1 | OID2081 | Neurolog | SIGLEC1 | Q6ZMC9 | 18 | 45825675 | 45844094 | 0 | - | - | - | - | - | - | - | - | - | - | - | - | - | - | - | - | - | - | - | - | - | - | - | - | - | - | - | - | - |
| SMAD2 | OID3135 | Oncology | SMAD2 | Q15796 | 18 | 47808957 | 47931146 | 0 | - | - | - | - | - | - | - | - | - | - | - | - | - | - | - | - | - | - | - | - | - | - | - | - | - | - | - | - | - |
| TXNL1 | OID3087 | Neurolog | TXNL1 | O43396 | 18 | 56597209 | 56651600 | 0 | - | - | - | - | - | - | - | - | - | - | - | - | - | - | - | - | - | - | - | - | - | - | - | - | - | - | - | - | - |
| VPS4B | OID3138 | Oncology | VPS4B | O75351 | 18 | 63389190 | 63422483 | 0 | - | - | - | - | - | - | - | - | - | - | - | - | - | - | - | - | - | - | - | - | - | - | - | - | - | - | - | - | - |
| ACTN4 | OID2043 | Inflamma | ACTN4 | O43707 | 19 | 38647649 | 38731589 | 0 | - | - | - | - | - | - | - | - | - | - | - | - | - | - | - | - | - | - | - | - | - | - | - | - | - | - | - | - | - |
| AKT1S1 | OID2089 | Neurolog | AKT1S1 | Q96B36 | 19 | 49869033 | 49878459 | 0 | - | - | - | - | - | - | - | - | - | - | - | - | - | - | - | - | - | - | - | - | - | - | - | - | - | - | - | - | - |
| AKT2 | OID3105 | Neurolog | AKT2 | P31751 | 19 | 40230317 | 40285536 | 0 | - | - | - | - | - | - | - | - | - | - | - | - | - | - | - | - | - | - | - | - | - | - | - | - | - | - | - | - | - |
| APLP1 | OID2016 | Cardiom | APLP1 | P51693 | 19 | 35867899 | 35879792 | 0 | - | - | - | - | - | - | - | - | - | - | - | - | - | - | - | - | - | - | - | - | - | - | - | - | - | - | - | - | - |
| ARHGEF | OID3113 | Neurolog | ARHGEF | Q92888 | 19 | 41883173 | 41930150 | 0 | - | - | - | - | - | - | - | - | - | - | - | - | - | - | - | - | - | - | - | - | - | - | - | - | - | - | - | - | - |
| ARID3A | OID3086 | Neurolog | ARID3A | Q99856 | 19 | 925781 | 975939 | 0 | - | - | - | - | - | - | - | - | - | - | - | - | - | - | - | - | - | - | - | - | - | - | - | - | - | - | - | - | - |
| ATP5F1D | OID3007 | Cardiom | ATP5F1D | P30049 | 19 | 1241746 | 1244825 | 0 | - | - | - | - | - | - | - | - | - | - | - | - | - | - | - | - | - | - | - | - | - | - | - | - | - | - | - | - | - |
| BABAM1 | OID3046 | Inflamma | BABAM1 | Q9NWV8 | 19 | 17267376 | 17281249 | 0 | - | - | - | - | - | - | - | - | - | - | - | - | - | - | - | - | - | - | - | - | - | - | - | - | - | - | - | - | - |
| BAX | OID2085 | Neurolog | BAX | Q07812 | 19 | 48954815 | 48961798 | 0 | - | - | - | - | - | - | - | - | - | - | - | - | - | - | - | - | - | - | - | - | - | - | - | - | - | - | - | - | - |
| BCAT2 | OID3084 | Neurolog | BCAT2 | O15382 | 19 | 48795062 | 48811029 | 0 | - | - | - | - | - | - | - | - | - | - | - | - | - | - | - | - | - | - | - | - | - | - | - | - | - | - | - | - | - |
| BLOC1S3 | OID3110 | Neurolog | BLOC1S3 | Q6QNY0 | 19 | 45178784 | 45216933 | 0 | - | - | - | - | - | - | - | - | - | - | - | - | - | - | - | - | - | - | - | - | - | - | - | - | - | - | - | - | - |
| BRME1 | OID3084 | Neurolog | BRME1 | Q0VDD7 | 19 | 13882348 | 13906452 | 0 | - | - | - | - | - | - | - | - | - | - | - | - | - | - | - | - | - | - | - | - | - | - | - | - | - | - | - | - | - |
| CCER2 | OID3022 | Cardiom | CCER2 | I3L3R5 | 19 | 38908980 | 38912186 | 0 | - | - | - | - | - | - | - | - | - | - | - | - | - | - | - | - | - | - | - | - | - | - | - | - | - | - | - | - | - |

| | | | | | | | | | | | | | | | | | | | | | | | | | | | | | | | | | | | | | |
|---|---|---|---|---|---|---|---|---|---|---|---|---|---|---|---|---|---|---|---|---|---|---|---|---|---|---|---|---|---|---|---|---|---|---|---|---|---|
| CCNE1 | OID3045 | Inflamma | CCNE1 | P24864 | 19 | 29811991 | 29824312 | 0 | - | - | - | - | - | - | - | - | - | - | - | - | - | - | - | - | - | - | - | - | - | - | - | - | - | - | - | - | - |
| CDC37 | OID2147 | Oncology | CDC37 | Q16543 | 19 | 10391133 | 10420121 | 0 | - | - | - | - | - | - | - | - | - | - | - | - | - | - | - | - | - | - | - | - | - | - | - | - | - | - | - | - | - |
| CDKN2D | OID2129 | Oncology | CDKN2D | P55273 | 19 | 10566460 | 10569059 | 0 | - | - | - | - | - | - | - | - | - | - | - | - | - | - | - | - | - | - | - | - | - | - | - | - | - | - | - | - | - |
| CEACAM | OID3125 | Oncology | CEACAM | A8MTB9 | 19 | 51478539 | 51491301 | 0 | - | - | - | - | - | - | - | - | - | - | - | - | - | - | - | - | - | - | - | - | - | - | - | - | - | - | - | - | - |
| CEACAM | OID2124 | Oncology | CEACAM | P40198 | 19 | 41796587 | 41811554 | 0 | - | - | - | - | - | - | - | - | - | - | - | - | - | - | - | - | - | - | - | - | - | - | - | - | - | - | - | - | - |
| CEBPA | OID3045 | Inflamma | CEBPA | P49715 | 19 | 33299934 | 33302534 | 0 | - | - | - | - | - | - | - | - | - | - | - | - | - | - | - | - | - | - | - | - | - | - | - | - | - | - | - | - | - |
| CIRBP | OID3150 | Oncology | CIRBP | Q14011 | 19 | 1259384 | 1274880 | 0 | - | - | - | - | - | - | - | - | - | - | - | - | - | - | - | - | - | - | - | - | - | - | - | - | - | - | - | - | - |
| CLC | OID2012 | Cardiome | CLC | Q05315 | 19 | 39731255 | 39738029 | 0 | - | - | - | - | - | - | - | - | - | - | - | - | - | - | - | - | - | - | - | - | - | - | - | - | - | - | - | - | - |
| CLPP | OID2091 | Neurolog | CLPP | Q16740 | 19 | 6361531 | 6370242 | 0 | - | - | - | - | - | - | - | - | - | - | - | - | - | - | - | - | - | - | - | - | - | - | - | - | - | - | - | - | - |
| COPE | OID2079 | Neurolog | COPE | O14579 | 19 | 18899514 | 18919387 | 0 | - | - | - | - | - | - | - | - | - | - | - | - | - | - | - | - | - | - | - | - | - | - | - | - | - | - | - | - | - |
| COX6B1 | OID3031 | Cardiome | COX6B1 | P14854 | 19 | 35648323 | 35658782 | 0 | - | - | - | - | - | - | - | - | - | - | - | - | - | - | - | - | - | - | - | - | - | - | - | - | - | - | - | - | - |
| CRLF1 | OID2069 | Inflamma | CRLF1 | O75462 | 19 | 18572220 | 18607741 | 0 | - | - | - | - | - | - | - | - | - | - | - | - | - | - | - | - | - | - | - | - | - | - | - | - | - | - | - | - | - |
| CRX | OID2007 | Cardiome | CRX | O43186 | 19 | 47819779 | 47843330 | 0 | - | - | - | - | - | - | - | - | - | - | - | - | - | - | - | - | - | - | - | - | - | - | - | - | - | - | - | - | - |
| DAND5 | OID3042 | Inflamma | DAND5 | Q8N907 | 19 | 12965159 | 12974760 | 0 | - | - | - | - | - | - | - | - | - | - | - | - | - | - | - | - | - | - | - | - | - | - | - | - | - | - | - | - | - |
| DDA1 | OID3012 | Cardiome | DDA1 | Q9BW61 | 19 | 17309518 | 17323298 | 0 | - | - | - | - | - | - | - | - | - | - | - | - | - | - | - | - | - | - | - | - | - | - | - | - | - | - | - | - | - |
| DDX39A | OID3049 | Inflamma | DDX39A | O00148 | 19 | 14408798 | 14419383 | 0 | - | - | - | - | - | - | - | - | - | - | - | - | - | - | - | - | - | - | - | - | - | - | - | - | - | - | - | - | - |
| DHPS | OID3135 | Oncology | DHPS | P49366 | 19 | 12675717 | 12681880 | 0 | - | - | - | - | - | - | - | - | - | - | - | - | - | - | - | - | - | - | - | - | - | - | - | - | - | - | - | - | - |
| EPN1 | OID3125 | Oncology | EPN1 | Q9Y6I3 | 19 | 55675226 | 55709858 | 0 | - | - | - | - | - | - | - | - | - | - | - | - | - | - | - | - | - | - | - | - | - | - | - | - | - | - | - | - | - |
| ERCC1 | OID3122 | Oncology | ERCC1 | P07992 | 19 | 45407334 | 45478828 | 0 | - | - | - | - | - | - | - | - | - | - | - | - | - | - | - | - | - | - | - | - | - | - | - | - | - | - | - | - | - |
| ERVV-1 | OID3009 | Cardiome | ERVV-1 | B6SEH8 | 19 | 53013921 | 53016123 | 0 | - | - | - | - | - | - | - | - | - | - | - | - | - | - | - | - | - | - | - | - | - | - | - | - | - | - | - | - | - |
| FARSA | OID3088 | Neurolog | FARSA | Q9Y285 | 19 | 12922479 | 12934037 | 0 | - | - | - | - | - | - | - | - | - | - | - | - | - | - | - | - | - | - | - | - | - | - | - | - | - | - | - | - | - |
| FDX2 | OID3014 | Cardiome | FDX2 | Q6P4F2 | 19 | 10310045 | 10316015 | 0 | - | - | - | - | - | - | - | - | - | - | - | - | - | - | - | - | - | - | - | - | - | - | - | - | - | - | - | - | - |
| FOSB | OID2079 | Neurolog | FOSB | P53539 | 19 | 45467995 | 45475179 | 0 | - | - | - | - | - | - | - | - | - | - | - | - | - | - | - | - | - | - | - | - | - | - | - | - | - | - | - | - | - |
| FUT1 | OID3129 | Oncology | FUT1 | P19526 | 19 | 48748011 | 48755390 | 0 | - | - | - | - | - | - | - | - | - | - | - | - | - | - | - | - | - | - | - | - | - | - | - | - | - | - | - | - | - |
| GADD45 | OID3126 | Oncology | GADD45 | O75293 | 19 | 2476122 | 2478259 | 0 | - | - | - | - | - | - | - | - | - | - | - | - | - | - | - | - | - | - | - | - | - | - | - | - | - | - | - | - | - |
| GADD45 | OID3023 | Cardiome | GADD45 | Q8TAE8 | 19 | 12953119 | 12957223 | 0 | - | - | - | - | - | - | - | - | - | - | - | - | - | - | - | - | - | - | - | - | - | - | - | - | - | - | - | - | - |
| GAMT | OID3013 | Cardiome | GAMT | Q14353 | 19 | 1397026 | 1401570 | 0 | - | - | - | - | - | - | - | - | - | - | - | - | - | - | - | - | - | - | - | - | - | - | - | - | - | - | - | - | - |
| GET3 | OID3030 | Cardiome | GET3 | O43681 | 19 | 12737139 | 12748323 | 0 | - | - | - | - | - | - | - | - | - | - | - | - | - | - | - | - | - | - | - | - | - | - | - | - | - | - | - | - | - |
| GIPR | OID3054 | Inflamma | GIPR | P48546 | 19 | 45668221 | 45683722 | 0 | - | - | - | - | - | - | - | - | - | - | - | - | - | - | - | - | - | - | - | - | - | - | - | - | - | - | - | - | - |
| GPI | OID3068 | Inflamma | GPI | P06744 | 19 | 34359480 | 34402413 | 0 | - | - | - | - | - | - | - | - | - | - | - | - | - | - | - | - | - | - | - | - | - | - | - | - | - | - | - | - | - |
| GYS1 | OID2009 | Cardiome | GYS1 | P13807 | 19 | 48968130 | 48993310 | 0 | - | - | - | - | - | - | - | - | - | - | - | - | - | - | - | - | - | - | - | - | - | - | - | - | - | - | - | - | - |
| HNRNPU | OID3103 | Neurolog | HNRNPU | Q9BUJ2 | 19 | 41262496 | 41307787 | 0 | - | - | - | - | - | - | - | - | - | - | - | - | - | - | - | - | - | - | - | - | - | - | - | - | - | - | - | - | - |
| IFNL2 | OID3079 | Neurolog | IFNL2 | Q8IZJ0 | 19 | 39268396 | 39270188 | 0 | - | - | - | - | - | - | - | - | - | - | - | - | - | - | - | - | - | - | - | - | - | - | - | - | - | - | - | - | - |
| IGFL4 | OID3050 | Inflamma | IGFL4 | Q6B9Z1 | 19 | 46039182 | 46077118 | 0 | - | - | - | - | - | - | - | - | - | - | - | - | - | - | - | - | - | - | - | - | - | - | - | - | - | - | - | - | - |
| IGLON5 | OID3116 | Oncology | IGLON5 | A6NGN9 | 19 | 51311644 | 51330891 | 0 | - | - | - | - | - | - | - | - | - | - | - | - | - | - | - | - | - | - | - | - | - | - | - | - | - | - | - | - | - |
| IL11 | OID2045 | Inflamma | IL11 | P20809 | 19 | 55364382 | 55370463 | 0 | - | - | - | - | - | - | - | - | - | - | - | - | - | - | - | - | - | - | - | - | - | - | - | - | - | - | - | - | - |
| INSL3 | OID3112 | Neurolog | INSL3 | P51460 | 19 | 17816512 | 17821574 | 0 | - | - | - | - | - | - | - | - | - | - | - | - | - | - | - | - | - | - | - | - | - | - | - | - | - | - | - | - | - |
| IZUMO1 | OID3115 | Oncology | IZUMO1 | Q8IYV9 | 19 | 48740852 | 48746909 | 0 | - | - | - | - | - | - | - | - | - | - | - | - | - | - | - | - | - | - | - | - | - | - | - | - | - | - | - | - | - |
| LILRA4 | OID3016 | Cardiome | LILRA4 | P59901 | 19 | 54333185 | 54339162 | 0 | - | - | - | - | - | - | - | - | - | - | - | - | - | - | - | - | - | - | - | - | - | - | - | - | - | - | - | - | - |
| MAG | OID3096 | Neurolog | MAG | P20916 | 19 | 35292125 | 35313807 | 0 | - | - | - | - | - | - | - | - | - | - | - | - | - | - | - | - | - | - | - | - | - | - | - | - | - | - | - | - | - |
| MLLT1 | OID3121 | Oncology | MLLT1 | Q03111 | 19 | 6210381 | 6279975 | 0 | - | - | - | - | - | - | - | - | - | - | - | - | - | - | - | - | - | - | - | - | - | - | - | - | - | - | - | - | - |
| MYDGF | OID3031 | Cardiome | MYDGF | Q969H8 | 19 | 4641374 | 4670362 | 0 | - | - | - | - | - | - | - | - | - | - | - | - | - | - | - | - | - | - | - | - | - | - | - | - | - | - | - | - | - |
| NACC1 | OID3139 | Oncology | NACC1 | Q96RE7 | 19 | 13116862 | 13141147 | 0 | - | - | - | - | - | - | - | - | - | - | - | - | - | - | - | - | - | - | - | - | - | - | - | - | - | - | - | - | - |
| NCLN | OID2044 | Inflamma | NCLN | Q969V3 | 19 | 3185563 | 3209575 | 0 | - | - | - | - | - | - | - | - | - | - | - | - | - | - | - | - | - | - | - | - | - | - | - | - | - | - | - | - | - |
| NDUFB7 | OID3152 | Oncology | NDUFB7 | P17568 | 19 | 14566078 | 14572066 | 0 | - | - | - | - | - | - | - | - | - | - | - | - | - | - | - | - | - | - | - | - | - | - | - | - | - | - | - | - | - |
| NFIC | OID3098 | Neurolog | NFIC | P08651 | 19 | 3314403 | 3469217 | 0 | - | - | - | - | - | - | - | - | - | - | - | - | - | - | - | - | - | - | - | - | - | - | - | - | - | - | - | - | - |
| NMRK2 | OID3116 | Oncology | NMRK2 | Q9NPI5 | 19 | 3933069 | 3942416 | 0 | - | - | - | - | - | - | - | - | - | - | - | - | - | - | - | - | - | - | - | - | - | - | - | - | - | - | - | - | - |
| PAFAH1 | OID3099 | Neurolog | PAFAH1 | Q15102 | 19 | 42297033 | 42303546 | 0 | - | - | - | - | - | - | - | - | - | - | - | - | - | - | - | - | - | - | - | - | - | - | - | - | - | - | - | - | - |
| PAK4 | OID2081 | Neurolog | PAK4 | O96013 | 19 | 39125770 | 39182816 | 0 | - | - | - | - | - | - | - | - | - | - | - | - | - | - | - | - | - | - | - | - | - | - | - | - | - | - | - | - | - |
| PALM3 | OID3018 | Cardiome | PALM3 | A6NDB9 | 19 | 14053363 | 14062076 | 0 | - | - | - | - | - | - | - | - | - | - | - | - | - | - | - | - | - | - | - | - | - | - | - | - | - | - | - | - | - |
| PSPN | OID2051 | Inflamma | PSPN | O60542 | 19 | 6375148 | 6379058 | 0 | - | - | - | - | - | - | - | - | - | - | - | - | - | - | - | - | - | - | - | - | - | - | - | - | - | - | - | - | - |
| RELB | OID3046 | Inflamma | RELB | Q01201 | 19 | 45001461 | 45038192 | 0 | - | - | - | - | - | - | - | - | - | - | - | - | - | - | - | - | - | - | - | - | - | - | - | - | - | - | - | - | - |
| RRAS | OID3096 | Neurolog | RRAS | P10301 | 19 | 49635292 | 49640143 | 0 | - | - | - | - | - | - | - | - | - | - | - | - | - | - | - | - | - | - | - | - | - | - | - | - | - | - | - | - | - |
| RYR1 | OID3008 | Cardiome | RYR1 | P21817 | 19 | 38433691 | 38595273 | 0 | - | - | - | - | - | - | - | - | - | - | - | - | - | - | - | - | - | - | - | - | - | - | - | - | - | - | - | - | - |
| SAFB2 | OID3101 | Neurolog | SAFB2 | Q14151 | 19 | 5586999 | 5624046 | 0 | - | - | - | - | - | - | - | - | - | - | - | - | - | - | - | - | - | - | - | - | - | - | - | - | - | - | - | - | - |
| SHD | OID3009 | Cardiome | SHD | Q96IW2 | 19 | 4279266 | 4290722 | 0 | - | - | - | - | - | - | - | - | - | - | - | - | - | - | - | - | - | - | - | - | - | - | - | - | - | - | - | - | - |
| STK11 | OID2007 | Cardiome | STK11 | Q15831 | 19 | 1177558 | 1228431 | 0 | - | - | - | - | - | - | - | - | - | - | - | - | - | - | - | - | - | - | - | - | - | - | - | - | - | - | - | - | - |
| TARM1 | OID3093 | Neurolog | TARM1 | B6A8C7 | 19 | 54069895 | 54081365 | 0 | - | - | - | - | - | - | - | - | - | - | - | - | - | - | - | - | - | - | - | - | - | - | - | - | - | - | - | - | - |
| TJP3 | OID3079 | Neurolog | TJP3 | O95049 | 19 | 3708362 | 3750813 | 0 | - | - | - | - | - | - | - | - | - | - | - | - | - | - | - | - | - | - | - | - | - | - | - | - | - | - | - | - | - |
| TMED1 | OID3127 | Oncology | TMED1 | Q13445 | 19 | 10832067 | 10836318 | 0 | - | - | - | - | - | - | - | - | - | - | - | - | - | - | - | - | - | - | - | - | - | - | - | - | - | - | - | - | - |
| TNNI3 | OID2006 | Cardiome | TNNI3 | P19429 | 19 | 55151767 | 55157773 | 0 | - | - | - | - | - | - | - | - | - | - | - | - | - | - | - | - | - | - | - | - | - | - | - | - | - | - | - | - | - |
| VASP | OID3069 | Inflamma | VASP | P50552 | 19 | 45506579 | 45526989 | 0 | - | - | - | - | - | - | - | - | - | - | - | - | - | - | - | - | - | - | - | - | - | - | - | - | - | - | - | - | - |
| VSTM2B | OID3127 | Oncology | VSTM2B | A6NLU5 | 19 | 29525994 | 29564551 | 0 | - | - | - | - | - | - | - | - | - | - | - | - | - | - | - | - | - | - | - | - | - | - | - | - | - | - | - | - | - |
| ZNRF4 | OID3134 | Oncology | ZNRF4 | Q8WWF | 19 | 5455417 | 5456856 | 0 | - | - | - | - | - | - | - | - | - | - | - | - | - | - | - | - | - | - | - | - | - | - | - | - | - | - | - | - | - |
| BCL2L1 | OID3115 | Neurolog | BCL2L1 | Q07817 | 20 | 31664452 | 31723989 | 0 | - | - | - | - | - | - | - | - | - | - | - | - | - | - | - | - | - | - | - | - | - | - | - | - | - | - | - | - | - |
| CD93 | OID2039 | Cardiome | CD93 | Q9NPY3 | 20 | 23079360 | 23086324 | 0 | - | - | - | - | - | - | - | - | - | - | - | - | - | - | - | - | - | - | - | - | - | - | - | - | - | - | - | - | - |
| CDH22 | OID3133 | Oncology | CDH22 | Q9UJ99 | 20 | 46173739 | 46308498 | 0 | - | - | - | - | - | - | - | - | - | - | - | - | - | - | - | - | - | - | - | - | - | - | - | - | - | - | - | - | - |
| CDH4 | OID3087 | Neurolog | CDH4 | P55283 | 20 | 61252261 | 61940617 | 0 | - | - | - | - | - | - | - | - | - | - | - | - | - | - | - | - | - | - | - | - | - | - | - | - | - | - | - | - | - |
| CSNK2A | OID3105 | Neurolog | CSNK2A | P68400 | 20 | 472498 | 543835 | 0 | - | - | - | - | - | - | - | - | - | - | - | - | - | - | - | - | - | - | - | - | - | - | - | - | - | - | - | - | - |
| CYP24A1 | OID3009 | Cardiome | CYP24A1 | Q07973 | 20 | 54153446 | 54173986 | 0 | - | - | - | - | - | - | - | - | - | - | - | - | - | - | - | - | - | - | - | - | - | - | - | - | - | - | - | - | - |
| DEFB116 | OID3082 | Neurolog | DEFB116 | Q30KQ4 | 20 | 31303212 | 31308585 | 0 | - | - | - | - | - | - | - | - | - | - | - | - | - | - | - | - | - | - | - | - | - | - | - | - | - | - | - | - | - |
| DEFB118 | OID3085 | Neurolog | DEFB118 | Q96PH6 | 20 | 31368601 | 31373923 | 0 | - | - | - | - | - | - | - | - | - | - | - | - | - | - | - | - | - | - | - | - | - | - | - | - | - | - | - | - | - |
| EDEM2 | OID3013 | Cardiome | EDEM2 | Q9BV94 | 20 | 35115364 | 35147336 | 0 | - | - | - | - | - | - | - | - | - | - | - | - | - | - | - | - | - | - | - | - | - | - | - | - | - | - | - | - | - |
| EIF2S2 | OID3027 | Cardiome | EIF2S2 | P20042 | 20 | 34088309 | 34112243 | 0 | - | - | - | - | - | - | - | - | - | - | - | - | - | - | - | - | - | - | - | - | - | - | - | - | - | - | - | - | - |
| GID8 | OID3104 | Neurolog | GID8 | Q9NWU2 | 20 | 62938147 | 62948475 | 0 | - | - | - | - | - | - | - | - | - | - | - | - | - | - | - | - | - | - | - | - | - | - | - | - | - | - | - | - | - |
| HAO1 | OID2139 | Oncology | HAO1 | Q9UJM8 | 20 | 7882985 | 7940458 | 0 | - | - | - | - | - | - | - | - | - | - | - | - | - | - | - | - | - | - | - | - | - | - | - | - | - | - | - | - | - |
| MAP1LC | OID3086 | Neurolog | MAP1LC | Q9H492 | 20 | 34546854 | 34560345 | 0 | - | - | - | - | - | - | - | - | - | - | - | - | - | - | - | - | - | - | - | - | - | - | - | - | - | - | - | - | - |
| MYH7B | OID3117 | Oncology | MYH7B | A7E2Y1 | 20 | 34955810 | 35002437 | 0 | - | - | - | - | - | - | - | - | - | - | - | - | - | - | - | - | - | - | - | - | - | - | - | - | - | - | - | - | - |
| NOP56 | OID3005 | Cardiome | NOP56 | O00567 | 20 | 2652593 | 2658393 | 0 | - | - | - | - | - | - | - | - | - | - | - | - | - | - | - | - | - | - | - | - | - | - | - | - | - | - | - | - | - |
| OSBPL2 | OID3108 | Neurolog | OSBPL2 | Q9H1P3 | 20 | 62231922 | 62296213 | 0 | - | - | - | - | - | - | - | - | - | - | - | - | - | - | - | - | - | - | - | - | - | - | - | - | - | - | - | - | - |
| PCNA | OID3116 | Oncology | PCNA | P12004 | 20 | 5114953 | 5126626 | 0 | - | - | - | - | - | - | - | - | - | - | - | - | - | - | - | - | - | - | - | - | - | - | - | - | - | - | - | - | - |
| PDRG1 | OID3091 | Neurolog | PDRG1 | Q9NUG6 | 20 | 31944337 | 31952046 | 0 | - | - | - | - | - | - | - | - | - | - | - | - | - | - | - | - | - | - | - | - | - | - | - | - | - | - | - | - | - |
| PFDN4 | OID3121 | Oncology | PFDN4 | Q9NQP4 | 20 | 54208087 | 54228052 | 0 | - | - | - | - | - | - | - | - | - | - | - | - | - | - | - | - | - | - | - | - | - | - | - | - | - | - | - | - | - |
| PLCB1 | OID3048 | Inflamma | PLCB1 | Q9NQ66 | 20 | 8077251 | 8968360 | 0 | - | - | - | - | - | - | - | - | - | - | - | - | - | - | - | - | - | - | - | - | - | - | - | - | - | - | - | - | - |
| PTPN1 | OID2090 | Neurolog | PTPN1 | P18031 | 20 | 50510321 | 50585241 | 0 | - | - | - | - | - | - | - | - | - | - | - | - | - | - | - | - | - | - | - | - | - | - | - | - | - | - | - | - | - |
| SNRPB2 | OID3113 | Neurolog | SNRPB2 | P08579 | 20 | 16729961 | 16742564 | 0 | - | - | - | - | - | - | - | - | - | - | - | - | - | - | - | - | - | - | - | - | - | - | - | - | - | - | - | - | - |
| SNX5 | OID3021 | Cardiome | SNX5 | Q9Y5X3 | 20 | 17941597 | 17968980 | 0 | - | - | - | - | - | - | - | - | - | - | - | - | - | - | - | - | - | - | - | - | - | - | - | - | - | - | - | - | - |
| SRC | OID2152 | Oncology | SRC | P12931 | 20 | 37344685 | 37406050 | 0 | - | - | - | - | - | - | - | - | - | - | - | - | - | - | - | - | - | - | - | - | - | - | - | - | - | - | - | - | - |
| STAU1 | OID3121 | Oncology | STAU1 | O95793 | 20 | 49113339 | 49188367 | 0 | - | - | - | - | - | - | - | - | - | - | - | - | - | - | - | - | - | - | - | - | - | - | - | - | - | - | - | - | - |
| TOP1 | OID3144 | Oncology | TOP1 | P11387 | 20 | 41028822 | 41124487 | 0 | - | - | - | - | - | - | - | - | - | - | - | - | - | - | - | - | - | - | - | - | - | - | - | - | - | - | - | - | - |
| TPD52L2 | OID3060 | Inflamma | TPD52L2 | O43399 | 20 | 63865228 | 63891545 | 0 | - | - | - | - | - | - | - | - | - | - | - | - | - | - | - | - | - | - | - | - | - | - | - | - | - | - | - | - | - |
| ATP5PO | OID2081 | Neurolog | ATP5PO | P48047 | 21 | 33903453 | 33915814 | 0 | - | - | - | - | - | - | - | - | - | - | - | - | - | - | - | - | - | - | - | - | - | - | - | - | - | - | - | - | - |
| MORC3 | OID3107 | Neurolog | MORC3 | Q14149 | 21 | 36320189 | 36386148 | 0 | - | - | - | - | - | - | - | - | - | - | - | - | - | - | - | - | - | - | - | - | - | - | - | - | - | - | - | - | - |
| RIPK4 | OID3013 | Cardiome | RIPK4 | P57078 | 21 | 41739369 | 41767089 | 0 | - | - | - | - | - | - | - | - | - | - | - | - | - | - | - | - | - | - | - | - | - | - | - | - | - | - | - | - | - |
| SOD1 | OID2038 | Cardiome | SOD1 | P00441 | 21 | 31659666 | 31668931 | 0 | - | - | - | - | - | - | - | - | - | - | - | - | - | - | - | - | - | - | - | - | - | - | - | - | - | - | - | - | - |
| USP25 | OID3142 | Oncology | USP25 | Q9UHP3 | 21 | 15729982 | 15880064 | 0 | - | - | - | - | - | - | - | - | - | - | - | - | - | - | - | - | - | - | - | - | - | - | - | - | - | - | - | - | - |
| ATF4 | OID3116 | Oncology | ATF4 | P18848 | 22 | 39519695 | 39522683 | 0 | - | - | - | - | - | - | - | - | - | - | - | - | - | - | - | - | - | - | - | - | - | - | - | - | - | - | - | - | - |
| BRD1 | OID3125 | Oncology | BRD1 | O95696 | 22 | 49773283 | 49827873 | 0 | - | - | - | - | - | - | - | - | - | - | - | - | - | - | - | - | - | - | - | - | - | - | - | - | - | - | - | - | - |
| CHEK2 | OID2005 | Cardiome | CHEK2 | O96017 | 22 | 28687743 | 28742422 | 0 | - | - | - | - | - | - | - | - | - | - | - | - | - | - | - | - | - | - | - | - | - | - | - | - | - | - | - | - | - |
| CRKL | OID2074 | Inflamma | CRKL | P46109 | 22 | 20917407 | 20953747 | 0 | - | - | - | - | - | - | - | - | - | - | - | - | - | - | - | - | - | - | - | - | - | - | - | - | - | - | - | - | - |
| CRYBB2 | OID3108 | Neurolog | CRYBB2 | P43320 | 22 | 25212564 | 25231870 | 0 | - | - | - | - | - | - | - | - | - | - | - | - | - | - | - | - | - | - | - | - | - | - | - | - | - | - | - | - | - |
| EP300 | OID3043 | Inflamma | EP300 | Q09472 | 22 | 41092592 | 41180077 | 0 | - | - | - | - | - | - | - | - | - | - | - | - | - | - | - | - | - | - | - | - | - | - | - | - | - | - | - | - | - |
| GGA1 | OID2083 | Neurolog | GGA1 | Q9UJY5 | 22 | 37608725 | 37633564 | 0 | - | - | - | - | - | - | - | - | - | - | - | - | - | - | - | - | - | - | - | - | - | - | - | - | - | - | - | - | - |
| IL2RB | OID2045 | Inflamma | IL2RB | P14784 | 22 | 37125843 | 37175054 | 0 | - | - | - | - | - | - | - | - | - | - | - | - | - | - | - | - | - | - | - | - | - | - | - | - | - | - | - | - | - |
| INPP5J | OID3097 | Neurolog | INPP5J | Q15735 | 22 | 31122731 | 31134697 | 0 | - | - | - | - | - | - | - | - | - | - | - | - | - | - | - | - | - | - | - | - | - | - | - | - | - | - | - | - | - |
| MN1 | OID3090 | Neurolog | MN1 | Q10571 | 22 | 27748277 | 27801756 | 0 | - | - | - | - | - | - | - | - | - | - | - | - | - | - | - | - | - | - | - | - | - | - | - | - | - | - | - | - | - |
| MYH9 | OID3038 | Cardiome | MYH9 | P35579 | 22 | 36281280 | 36388010 | 0 | - | - | - | - | - | - | - | - | - | - | - | - | - | - | - | - | - | - | - | - | - | - | - | - | - | - | - | - | - |
| NUP50 | OID3010 | Cardiome | NUP50 | Q9UKX7 | 22 | 45163925 | 45188017 | 0 | - | - | - | - | - | - | - | - | - | - | - | - | - | - | - | - | - | - | - | - | - | - | - | - | - | - | - | - | - |
| OSM | OID2057 | Inflamma | OSM | P13725 | 22 | 30262829 | 30266851 | 0 | - | - | - | - | - | - | - | - | - | - | - | - | - | - | - | - | - | - | - | - | - | - | - | - | - | - | - | - | - |
| PDGFB | OID2074 | Inflamma | PDGFB | P01127 | 22 | 39223359 | 39244982 | 0 | - | - | - | - | - | - | - | - | - | - | - | - | - | - | - | - | - | - | - | - | - | - | - | - | - | - | - | - | - |
| POLR2F | OID2128 | Oncology | POLR2F | P61218 | 22 | 37952607 | 38041915 | 0 | - | - | - | - | - | - | - | - | - | - | - | - | - | - | - | - | - | - | - | - | - | - | - | - | - | - | - | - | - |
| PPM1F | OID3035 | Cardiome | PPM1F | P49593 | 22 | 21919425 | 21952848 | 0 | - | - | - | - | - | - | - | - | - | - | - | - | - | - | - | - | - | - | - | - | - | - | - | - | - | - | - | - | - |
| PRAME | OID3116 | Oncology | PRAME | P78395 | 22 | 22547701 | 22559361 | 0 | - | - | - | - | - | - | - | - | - | - | - | - | - | - | - | - | - | - | - | - | - | - | - | - | - | - | - | - | - |
| PRR5 | OID3045 | Inflamma | PRR5 | P85299 | 22 | 44668547 | 44737681 | 0 | - | - | - | - | - | - | - | - | - | - | - | - | - | - | - | - | - | - | - | - | - | - | - | - | - | - | - | - | - |
| RANGAP | OID2120 | Oncology | RANGAP | P46060 | 22 | 41244779 | 41286187 | 0 | - | - | - | - | - | - | - | - | - | - | - | - | - | - | - | - | - | - | - | - | - | - | - | - | - | - | - | - | - |
| SEPTIN3 | OID3102 | Neurolog | SEPTIN3 | Q9UH03 | 22 | 41969443 | 41998221 | 0 | - | - | - | - | - | - | - | - | - | - | - | - | - | - | - | - | - | - | - | - | - | - | - | - | - | - | - | - | - |
| SMTN | OID3115 | Neurolog | SMTN | P53814 | 22 | 31064105 | 31104757 | 0 | - | - | - | - | - | - | - | - | - | - | - | - | - | - | - | - | - | - | - | - | - | - | - | - | - | - | - | - | - |
| SNU13 | OID3005 | Cardiome | SNU13 | P55769 | 22 | 41673933 | 41690504 | 0 | - | - | - | - | - | - | - | - | - | - | - | - | - | - | - | - | - | - | - | - | - | - | - | - | - | - | - | - | - |
| TEX33 | OID3128 | Oncology | TEX33 | O43247 | 22 | 36991120 | 37007851 | 0 | - | - | - | - | - | - | - | - | - | - | - | - | - | - | - | - | - | - | - | - | - | - | - | - | - | - | - | - | - |
| UFD1 | OID3148 | Oncology | UFD1 | Q92890 | 22 | 19449911 | 19479202 | 0 | - | - | - | - | - | - | - | - | - | - | - | - | - | - | - | - | - | - | - | - | - | - | - | - | - | - | - | - | - |
| UPK3A | OID3079 | Neurolog | UPK3A | O75631 | 22 | 45284949 | 45295874 | 0 | - | - | - | - | - | - | - | - | - | - | - | - | - | - | - | - | - | - | - | - | - | - | - | - | - | - | - | - | - |
| DEFB103 | OID3053 | Inflamma | DEFB103 | P81534;P | 8;8 | 7428888;78 | 7430348;78 | 0 | - | - | - | - | - | - | - | - | - | - | - | - | - | - | - | - | - | - | - | - | - | - | - | - | - | - | - | - | - |
| ACE2 | OID2010 | Cardiome | ACE2 | Q9BYF1 | X | 15494566 | 15607236 | 0 | - | - | - | - | - | - | - | - | - | - | - | - | - | - | - | - | - | - | - | - | - | - | - | - | - | - | - | - | - |
| ADGRG2 | OID2018 | Cardiome | ADGRG2 | Q8IZP9 | X | 18989307 | 19122637 | 0 | - | - | - | - | - | - | - | - | - | - | - | - | - | - | - | - | - | - | - | - | - | - | - | - | - | - | - | - | - |
| AIFM1 | OID2128 | Oncology | AIFM1 | O95831 | X | 130124666 | 130165879 | 0 | - | - | - | - | - | - | - | - | - | - | - | - | - | - | - | - | - | - | - | - | - | - | - | - | - | - | - | - | - |
| AMOT | OID3057 | Inflamma | AMOT | Q4VCS5 | X | 112774503 | 112840831 | 0 | - | - | - | - | - | - | - | - | - | - | - | - | - | - | - | - | - | - | - | - | - | - | - | - | - | - | - | - | - |
| ARAF | OID3126 | Oncology | ARAF | P10398 | X | 47561205 | 47571908 | 0 | - | - | - | - | - | - | - | - | - | - | - | - | - | - | - | - | - | - | - | - | - | - | - | - | - | - | - | - | - |
| ARMCX2 | OID3079 | Neurolog | ARMCX2 | Q7L311 | X | 101655281 | 101659850 | 0 | - | - | - | - | - | - | - | - | - | - | - | - | - | - | - | - | - | - | - | - | - | - | - | - | - | - | - | - | - |
| ATG4A | OID2124 | Oncology | ATG4A | Q8WYN0 | X | 108091668 | 108154671 | 0 | - | - | - | - | - | - | - | - | - | - | - | - | - | - | - | - | - | - | - | - | - | - | - | - | - | - | - | - | - |
| ATP1B4 | OID3123 | Oncology | ATP1B4 | Q9UN42 | X | 120362085 | 120383249 | 0 | - | - | - | - | - | - | - | - | - | - | - | - | - | - | - | - | - | - | - | - | - | - | - | - | - | - | - | - | - |
| ATP6AP | OID2117 | Oncology | ATP6AP | O75787 | X | 40579372 | 40606848 | 0 | - | - | - | - | - | - | - | - | - | - | - | - | - | - | - | - | - | - | - | - | - | - | - | - | - | - | - | - | - |
| BEX3 | OID3120 | Oncology | BEX3 | Q00994 | X | 103376395 | 103378164 | 0 | - | - | - | - | - | - | - | - | - | - | - | - | - | - | - | - | - | - | - | - | - | - | - | - | - | - | - | - | - |
| BGN | OID2119 | Oncology | BGN | P21810 | X | 153494980 | 153509546 | 0 | - | - | - | - | - | - | - | - | - | - | - | - | - | - | - | - | - | - | - | - | - | - | - | - | - | - | - | - | - |
| C1GALT1 | OID3018 | Cardiome | C1GALT1 | Q96EU7 | X | 120625674 | 120630054 | 0 | - | - | - | - | - | - | - | - | - | - | - | - | - | - | - | - | - | - | - | - | - | - | - | - | - | - | - | - | - |
| CD40LG | OID2061 | Inflamma | CD40LG | P29965 | X | 136648158 | 136660390 | 0 | - | - | - | - | - | - | - | - | - | - | - | - | - | - | - | - | - | - | - | - | - | - | - | - | - | - | - | - | - |
| CD99 | OID2111 | Neurolog | CD99 | P14209 | X | 2691187 | 2741309 | 0 | - | - | - | - | - | - | - | - | - | - | - | - | - | - | - | - | - | - | - | - | - | - | - | - | - | - | - | - | - |
| CD99L2 | OID2112 | Neurolog | CD99L2 | Q8TCZ2 | X | 150766336 | 150898816 | 0 | - | - | - | - | - | - | - | - | - | - | - | - | - | - | - | - | - | - | - | - | - | - | - | - | - | - | - | - | - |
| CDKL5 | OID3079 | Neurolog | CDKL5 | O76039 | X | 18425583 | 18653629 | 0 | - | - | - | - | - | - | - | - | - | - | - | - | - | - | - | - | - | - | - | - | - | - | - | - | - | - | - | - | - |
| CETN2 | OID2083 | Neurolog | CETN2 | P41208 | X | 152826994 | 152830757 | 0 | - | - | - | - | - | - | - | - | - | - | - | - | - | - | - | - | - | - | - | - | - | - | - | - | - | - | - | - | - |
| CFP | OID3078 | Inflamma | CFP | P27918 | X | 47623172 | 47630305 | 0 | - | - | - | - | - | - | - | - | - | - | - | - | - | - | - | - | - | - | - | - | - | - | - | - | - | - | - | - | - |
| CHM | OID3097 | Neurolog | CHM | P24386 | X | 85861180 | 86047561 | 0 | - | - | - | - | - | - | - | - | - | - | - | - | - | - | - | - | - | - | - | - | - | - | - | - | - | - | - | - | - |
| CHRDL1 | OID2077 | Inflamma | CHRDL1 | Q9BU40 | X | 110673856 | 110795819 | 0 | - | - | - | - | - | - | - | - | - | - | - | - | - | - | - | - | - | - | - | - | - | - | - | - | - | - | - | - | - |
| CSF2RA | OID2091 | Neurolog | CSF2RA | P15509 | X | 1268800 | 1310381 | 0 | - | - | - | - | - | - | - | - | - | - | - | - | - | - | - | - | - | - | - | - | - | - | - | - | - | - | - | - | - |
| DDX53 | OID3094 | Neurolog | DDX53 | Q86TM3 | X | 22999960 | 23003589 | 0 | - | - | - | - | - | - | - | - | - | - | - | - | - | - | - | - | - | - | - | - | - | - | - | - | - | - | - | - | - |
| DIPK2B | OID3058 | Inflamma | DIPK2B | Q9H7Y0 | X | 45148373 | 45200901 | 0 | - | - | - | - | - | - | - | - | - | - | - | - | - | - | - | - | - | - | - | - | - | - | - | - | - | - | - | - | - |
| DMD | OID3022 | Cardiome | DMD | P11532 | X | 31097677 | 33339609 | 0 | - | - | - | - | - | - | - | - | - | - | - | - | - | - | - | - | - | - | - | - | - | - | - | - | - | - | - | - | - |
| DYNLT3 | OID3141 | Oncology | DYNLT3 | P51808 | X | 37836757 | 37847571 | 0 | - | - | - | - | - | - | - | - | - | - | - | - | - | - | - | - | - | - | - | - | - | - | - | - | - | - | - | - | - |
| EDA2R | OID2145 | Oncology | EDA2R | Q9HAV5 | X | 66595637 | 66639298 | 0 | - | - | - | - | - | - | - | - | - | - | - | - | - | - | - | - | - | - | - | - | - | - | - | - | - | - | - | - | - |
| EIF1AX | OID3112 | Neurolog | EIF1AX | P47813 | X | 20124525 | 20141838 | 0 | - | - | - | - | - | - | - | - | - | - | - | - | - | - | - | - | - | - | - | - | - | - | - | - | - | - | - | - | - |
| ENOX2 | OID3015 | Cardiome | ENOX2 | Q16206 | X | 130622325 | 130903234 | 0 | - | - | - | - | - | - | - | - | - | - | - | - | - | - | - | - | - | - | - | - | - | - | - | - | - | - | - | - | - |
| F9 | OID2034 | Cardiome | F9 | P00740 | X | 139530739 | 139563459 | 0 | - | - | - | - | - | - | - | - | - | - | - | - | - | - | - | - | - | - | - | - | - | - | - | - | - | - | - | - | - |
| FGF16 | OID3053 | Inflamma | FGF16 | O43320 | X | 77447389 | 77457278 | 0 | - | - | - | - | - | - | - | - | - | - | - | - | - | - | - | - | - | - | - | - | - | - | - | - | - | - | - | - | - |
| FMR1 | OID2123 | Oncology | FMR1 | Q06787 | X | 147911919 | 147951125 | 0 | - | - | - | - | - | - | - | - | - | - | - | - | - | - | - | - | - | - | - | - | - | - | - | - | - | - | - | - | - |
| FRMD7 | OID3089 | Neurolog | FRMD7 | Q6ZUT3 | X | 132076990 | 132128020 | 0 | - | - | - | - | - | - | - | - | - | - | - | - | - | - | - | - | - | - | - | - | - | - | - | - | - | - | - | - | - |
| GAGE2A | OID3119 | Oncology | GAGE2A | Q6NT46 | X | 49589496 | 49596824 | 0 | - | - | - | - | - | - | - | - | - | - | - | - | - | - | - | - | - | - | - | - | - | - | - | - | - | - | - | - | - |
| GLA | OID3059 | Inflamma | GLA | P06280 | X | 101393273 | 101408012 | 0 | - | - | - | - | - | - | - | - | - | - | - | - | - | - | - | - | - | - | - | - | - | - | - | - | - | - | - | - | - |
| GPKOW | OID2089 | Neurolog | GPKOW | Q92917 | X | 49113407 | 49123735 | 0 | - | - | - | - | - | - | - | - | - | - | - | - | - | - | - | - | - | - | - | - | - | - | - | - | - | - | - | - | - |
| GPR101 | OID3081 | Neurolog | GPR101 | Q96P66 | X | 137023929 | 137033995 | 0 | - | - | - | - | - | - | - | - | - | - | - | - | - | - | - | - | - | - | - | - | - | - | - | - | - | - | - | - | - |
| H2AP | OID3123 | Oncology | H2AP | O75409 | X | 37990779 | 37991314 | 0 | - | - | - | - | - | - | - | - | - | - | - | - | - | - | - | - | - | - | - | - | - | - | - | - | - | - | - | - | - |
| HDAC8 | OID3043 | Inflamma | HDAC8 | Q9BY41 | X | 72329516 | 72573101 | 0 | - | - | - | - | - | - | - | - | - | - | - | - | - | - | - | - | - | - | - | - | - | - | - | - | - | - | - | - | - |
| HEPH | OID3033 | Cardiome | HEPH | Q9BQS7 | X | 66162671 | 66268867 | 0 | - | - | - | - | - | - | - | - | - | - | - | - | - | - | - | - | - | - | - | - | - | - | - | - | - | - | - | - | - |
| HS6ST2 | OID3048 | Inflamma | HS6ST2 | Q96MM7 | X | 132626015 | 132961395 | 0 | - | - | - | - | - | - | - | - | - | - | - | - | - | - | - | - | - | - | - | - | - | - | - | - | - | - | - | - | - |
| IDS | OID2061 | Inflamma | IDS | P22304 | X | 149476988 | 149521096 | 0 | - | - | - | - | - | - | - | - | - | - | - | - | - | - | - | - | - | - | - | - | - | - | - | - | - | - | - | - | - |
| IGBP1 | OID3105 | Neurolog | IGBP1 | P78318 | X | 70133447 | 70166324 | 0 | - | - | - | - | - | - | - | - | - | - | - | - | - | - | - | - | - | - | - | - | - | - | - | - | - | - | - | - | - |
| IKBKG | OID2054 | Inflamma | IKBKG | Q9Y6K9 | X | 154541199 | 154565046 | 0 | - | - | - | - | - | - | - | - | - | - | - | - | - | - | - | - | - | - | - | - | - | - | - | - | - | - | - | - | - |
| IL13RA1 | OID2150 | Oncology | IL13RA1 | P78552 | X | 118727133 | 118794535 | 0 | - | - | - | - | - | - | - | - | - | - | - | - | - | - | - | - | - | - | - | - | - | - | - | - | - | - | - | - | - |
| IL13RA2 | OID3084 | Neurolog | IL13RA2 | Q14627 | X | 115003975 | 115019977 | 0 | - | - | - | - | - | - | - | - | - | - | - | - | - | - | - | - | - | - | - | - | - | - | - | - | - | - | - | - | - |
| IL2RG | OID3136 | Oncology | IL2RG | P31785 | X | 71107404 | 71112108 | 0 | - | - | - | - | - | - | - | - | - | - | - | - | - | - | - | - | - | - | - | - | - | - | - | - | - | - | - | - | - |
| IL3RA | OID2046 | Inflamma | IL3RA | P26951 | X | 1336616 | 1382689 | 0 | - | - | - | - | - | - | - | - | - | - | - | - | - | - | - | - | - | - | - | - | - | - | - | - | - | - | - | - | - |
| IRAK1 | OID2048 | Inflamma | IRAK1 | P51617 | X | 154010506 | 154019902 | 0 | - | - | - | - | - | - | - | - | - | - | - | - | - | - | - | - | - | - | - | - | - | - | - | - | - | - | - | - | - |
| ITGB1BP | OID2011 | Cardiome | ITGB1BP | Q9UKP3 | X | 71301750 | 71305371 | 0 | - | - | - | - | - | - | - | - | - | - | - | - | - | - | - | - | - | - | - | - | - | - | - | - | - | - | - | - | - |
| ITM2A | OID2050 | Inflamma | ITM2A | O43736 | X | 79360384 | 79367667 | 0 | - | - | - | - | - | - | - | - | - | - | - | - | - | - | - | - | - | - | - | - | - | - | - | - | - | - | - | - | - |
| L1CAM | OID2148 | Oncology | L1CAM | P32004 | X | 153861514 | 153886173 | 0 | - | - | - | - | - | - | - | - | - | - | - | - | - | - | - | - | - | - | - | - | - | - | - | - | - | - | - | - | - |
| LAMP2 | OID2103 | Neurolog | LAMP2 | P13473 | X | 120426148 | 120469365 | 0 | - | - | - | - | - | - | - | - | - | - | - | - | - | - | - | - | - | - | - | - | - | - | - | - | - | - | - | - | - |
| MAGEA3 | OID3133 | Oncology | MAGEA3 | P43357 | X | 152698767 | 152702347 | 0 | - | - | - | - | - | - | - | - | - | - | - | - | - | - | - | - | - | - | - | - | - | - | - | - | - | - | - | - | - |
| MAGED1 | OID2118 | Oncology | MAGED1 | Q9Y5V3 | X | 51803007 | 51902354 | 0 | - | - | - | - | - | - | - | - | - | - | - | - | - | - | - | - | - | - | - | - | - | - | - | - | - | - | - | - | - |
| MCTS1 | OID3144 | Oncology | MCTS1 | Q9ULC4 | X | 120594010 | 120621159 | 0 | - | - | - | - | - | - | - | - | - | - | - | - | - | - | - | - | - | - | - | - | - | - | - | - | - | - | - | - | - |
| MORF4L | OID3116 | Oncology | MORF4L | Q15014 | X | 103675496 | 103688158 | 0 | - | - | - | - | - | - | - | - | - | - | - | - | - | - | - | - | - | - | - | - | - | - | - | - | - | - | - | - | - |
| NAA10 | OID3018 | Cardiome | NAA10 | P41227 | X | 153929225 | 153935080 | 0 | - | - | - | - | - | - | - | - | - | - | - | - | - | - | - | - | - | - | - | - | - | - | - | - | - | - | - | - | - |
| NUDT10 | OID3019 | Cardiome | NUDT10 | Q8NFP7 | X | 51332231 | 51337525 | 0 | - | - | - | - | - | - | - | - | - | - | - | - | - | - | - | - | - | - | - | - | - | - | - | - | - | - | - | - | - |
| OFD1 | OID3126 | Oncology | OFD1 | O75665 | X | 13734743 | 13777955 | 0 | - | - | - | - | - | - | - | - | - | - | - | - | - | - | - | - | - | - | - | - | - | - | - | - | - | - | - | - | - |
| OGT | OID3130 | Oncology | OGT | O15294 | X | 71533087 | 71575892 | 0 | - | - | - | - | - | - | - | - | - | - | - | - | - | - | - | - | - | - | - | - | - | - | - | - | - | - | - | - | - |
| OPHN1 | OID3108 | Neurolog | OPHN1 | O60890 | X | 67949349 | 68433913 | 0 | - | - | - | - | - | - | - | - | - | - | - | - | - | - | - | - | - | - | - | - | - | - | - | - | - | - | - | - | - |
| PLXNB3 | OID2016 | Cardiome | PLXNB3 | Q9ULL4 | X | 153764196 | 153779346 | 0 | - | - | - | - | - | - | - | - | - | - | - | - | - | - | - | - | - | - | - | - | - | - | - | - | - | - | - | - | - |
| POF1B | OID3050 | Inflamma | POF1B | Q8WVV4 | X | 85277396 | 85379717 | 0 | - | - | - | - | - | - | - | - | - | - | - | - | - | - | - | - | - | - | - | - | - | - | - | - | - | - | - | - | - |
| PQBP1 | OID2117 | Oncology | PQBP1 | O60828 | X | 48890197 | 48903402 | 0 | - | - | - | - | - | - | - | - | - | - | - | - | - | - | - | - | - | - | - | - | - | - | - | - | - | - | - | - | - |
| RAB33A | OID3012 | Cardiome | RAB33A | Q14088 | X | 130171962 | 130184870 | 0 | - | - | - | - | - | - | - | - | - | - | - | - | - | - | - | - | - | - | - | - | - | - | - | - | - | - | - | - | - |
| RAB39B | OID3006 | Cardiome | RAB39B | Q96DA2 | X | 155258235 | 155264491 | 0 | - | - | - | - | - | - | - | - | - | - | - | - | - | - | - | - | - | - | - | - | - | - | - | - | - | - | - | - | - |
| RP2 | OID2130 | Oncology | RP2 | O75695 | X | 46837043 | 46882358 | 0 | - | - | - | - | - | - | - | - | - | - | - | - | - | - | - | - | - | - | - | - | - | - | - | - | - | - | - | - | - |
| RPGR | OID3095 | Neurolog | RPGR | Q92834 | X | 38269163 | 38327544 | 0 | - | - | - | - | - | - | - | - | - | - | - | - | - | - | - | - | - | - | - | - | - | - | - | - | - | - | - | - | - |
| S100G | OID3105 | Neurolog | S100G | P29377 | X | 16650158 | 16654670 | 0 | - | - | - | - | - | - | - | - | - | - | - | - | - | - | - | - | - | - | - | - | - | - | - | - | - | - | - | - | - |
| SAT1 | OID3011 | Cardiome | SAT1 | P21673 | X | 23783173 | 23786210 | 0 | - | - | - | - | - | - | - | - | - | - | - | - | - | - | - | - | - | - | - | - | - | - | - | - | - | - | - | - | - |
| SERPINA | OID3072 | Inflamma | SERPINA | P05543 | X | 106032435 | 106038727 | 0 | - | - | - | - | - | - | - | - | - | - | - | - | - | - | - | - | - | - | - | - | - | - | - | - | - | - | - | - | - |
| SH2D1A | OID2051 | Inflamma | SH2D1A | O60880 | X | 124227868 | 124373197 | 0 | - | - | - | - | - | - | - | - | - | - | - | - | - | - | - | - | - | - | - | - | - | - | - | - | - | - | - | - | - |
| SLITRK2 | OID2140 | Oncology | SLITRK2 | Q9H156 | X | 145817829 | 145829856 | 0 | - | - | - | - | - | - | - | - | - | - | - | - | - | - | - | - | - | - | - | - | - | - | - | - | - | - | - | - | - |
| SMS | OID3085 | Neurolog | SMS | P52788 | X | 21940709 | 21994837 | 0 | - | - | - | - | - | - | - | - | - | - | - | - | - | - | - | - | - | - | - | - | - | - | - | - | - | - | - | - | - |
| SRPX | OID3031 | Cardiome | SRPX | P78539 | X | 38149336 | 38220924 | 0 | - | - | - | - | - | - | - | - | - | - | - | - | - | - | - | - | - | - | - | - | - | - | - | - | - | - | - | - | - |
| SYAP1 | OID3063 | Inflamma | SYAP1 | Q96A49 | X | 16719612 | 16765340 | 0 | - | - | - | - | - | - | - | - | - | - | - | - | - | - | - | - | - | - | - | - | - | - | - | - | - | - | - | - | - |
| SYTL4 | OID3033 | Cardiome | SYTL4 | Q96C24 | X | 100671783 | 100732123 | 0 | - | - | - | - | - | - | - | - | - | - | - | - | - | - | - | - | - | - | - | - | - | - | - | - | - | - | - | - | - |
| TBL1X | OID2122 | Oncology | TBL1X | O60907 | X | 9463320 | 9741037 | 0 | - | - | - | - | - | - | - | - | - | - | - | - | - | - | - | - | - | - | - | - | - | - | - | - | - | - | - | - | - |
| TIMM8A | OID3114 | Neurolog | TIMM8A | O60220 | X | 101345661 | 101348742 | 0 | - | - | - | - | - | - | - | - | - | - | - | - | - | - | - | - | - | - | - | - | - | - | - | - | - | - | - | - | - |
| TIMP1 | OID2041 | Cardiome | TIMP1 | P01033 | X | 47582408 | 47586789 | 0 | - | - | - | - | - | - | - | - | - | - | - | - | - | - | - | - | - | - | - | - | - | - | - | - | - | - | - | - | - |
| TSPAN7 | OID3081 | Neurolog | TSPAN7 | P41732 | X | 38561542 | 38688920 | 0 | - | - | - | - | - | - | - | - | - | - | - | - | - | - | - | - | - | - | - | - | - | - | - | - | - | - | - | - | - |
| VEGFD | OID2066 | Inflamma | VEGFD | O43915 | X | 15345596 | 15384413 | 0 | - | - | - | - | - | - | - | - | - | - | - | - | - | - | - | - | - | - | - | - | - | - | - | - | - | - | - | - | - |
| VSIG4 | OID2114 | Neurolog | VSIG4 | Q9Y279 | X | 66021738 | 66040125 | 0 | - | - | - | - | - | - | - | - | - | - | - | - | - | - | - | - | - | - | - | - | - | - | - | - | - | - | - | - | - |
| WAS | OID2047 | Inflamma | WAS | P42768 | X | 48676596 | 48691427 | 0 | - | - | - | - | - | - | - | - | - | - | - | - | - | - | - | - | - | - | - | - | - | - | - | - | - | - | - | - | - |
| XG | OID2025 | Cardiome | XG | P55808 | X | 2752040 | 2816500 | 0 | - | - | - | - | - | - | - | - | - | - | - | - | - | - | - | - | - | - | - | - | - | - | - | - | - | - | - | - | - |
| XIAP | OID3049 | Inflamma | XIAP | P98170 | X | 123859712 | 123913972 | 0 | - | - | - | - | - | - | - | - | - | - | - | - | - | - | - | - | - | - | - | - | - | - | - | - | - | - | - | - | - |
| XPNPEP | OID2150 | Oncology | XPNPEP | O43895 | X | 129738949 | 129769536 | 0 | - | - | - | - | - | - | - | - | - | - | - | - | - | - | - | - | - | - | - | - | - | - | - | - | - | - | - | - | - |
| ZNF75D | OID3130 | Oncology | ZNF75D | P51815 | X | 135248920 | 135344109 | 0 | - | - | - | - | - | - | - | - | - | - | - | - | - | - | - | - | - | - | - | - | - | - | - | - | - | - | - | - | - |
| CTAG1A | OID3129 | Oncology | CTAG1A | P78358;P | X;X | 154585133; | 154586816; | 0 | - | - | - | - | - | - | - | - | - | - | - | - | - | - | - | - | - | - | - | - | - | - | - | - | - | - | - | - | - |
| SPACA5 | OID3099 | Neurolog | SPACA5 | Q96QH8 | X;X | 48130626;4 | 48132620;4 | 0 | - | - | - | - | - | - | - | - | - | - | - | - | - | - | - | - | - | - | - | - | - | - | - | - | - | - | - | - | - |

**Table S7 Two-sample Mendelian randomization analysis results using cis-pQTL reported by Sun et al. to test the causal effects of proteins on incident ADRD in participants with a history of MDD at baseline**

| Assay Target | Olink ID | Protein pa | Gene sym | UniProt | Gene CHF | Gene start | Gene end | SNPs | Fstat | IVW HR | VW Lower | VW Upper | IVW P | IVW Padj |
|---|---|---|---|---|---|---|---|---|---|---|---|---|---|---|
| APOE | OID30727 | Inflammat | APOE | P02649 | 19 | 44905791 | 44909393 | 1 | 10241.44 | 0.48 | 0.43 | 0.53 | 3.67E-42 | 6.37E-39 |
| BAG3 | OID20972 | Neurology | BAG3 | O95817 | 10 | 119651380 | 119677819 | 1 | 65.46 | 6.50 | 1.79 | 23.62 | 0.004 | 0.813 |
| CD70 | OID20599 | Inflammat | CD70 | P32970 | 19 | 6583183 | 6604103 | 1 | 11479.59 | 0.84 | 0.74 | 0.95 | 0.005 | 0.813 |
| CRISP2 | OID21456 | Oncology | CRISP2 | P16562 | 6 | 49692358 | 49713590 | 1 | 4880.02 | 0.75 | 0.61 | 0.91 | 0.003 | 0.813 |
| CXCL16 | OID20282 | Cardiome | CXCL16 | Q9H2A7 | 17 | 4733533 | 4739928 | 1 | 729.00 | 2.08 | 1.25 | 3.44 | 0.005 | 0.813 |
| DDC | OID20215 | Cardiome | DDC | P20711 | 7 | 50458436 | 50565405 | 1 | 2288.03 | 0.71 | 0.57 | 0.88 | 0.002 | 0.813 |
| GRAP2 | OID20192 | Cardiome | GRAP2 | O75791 | 22 | 39901084 | 39973721 | 1 | 89.69 | 0.04 | 0.00 | 0.39 | 0.005 | 0.813 |
| HSDL2 | OID30256 | Cardiome | HSDL2 | Q6YN16 | 9 | 112380080 | 112472405 | 1 | 5643.77 | 0.77 | 0.66 | 0.91 | 0.002 | 0.813 |
| IL17RA | OID21089 | Neurology | IL17RA | Q96F46 | 22 | 17084954 | 17115694 | 1 | 11475.77 | 0.84 | 0.75 | 0.94 | 0.003 | 0.813 |
| SCN4B | OID30277 | Cardiome | SCN4B | Q8IWT1 | 11 | 118133377 | 118152888 | 1 | 6806.25 | 1.27 | 1.09 | 1.47 | 0.002 | 0.813 |
| VEGFA | OID20650 | Inflammat | VEGFA | P15692 | 6 | 43770184 | 43786487 | 1 | 9660.08 | 1.22 | 1.07 | 1.41 | 0.004 | 0.813 |
| ANGPT2 | OID21463 | Oncology | ANGPT2 | O15123 | 8 | 6499632 | 6563409 | 1 | 385.14 | 2.45 | 1.28 | 4.69 | 0.007 | 0.869 |
| APOA4 | OID30733 | Inflammat | APOA4 | P06727 | 11 | 116820700 | 116823304 | 1 | 310.64 | 2.39 | 1.21 | 4.74 | 0.012 | 0.869 |
| ART5 | OID30828 | Neurology | ART5 | Q96L15 | 11 | 3638512 | 3642316 | 1 | 544.44 | 1.95 | 1.19 | 3.20 | 0.008 | 0.869 |
| CBLN4 | OID21386 | Oncology | CBLN4 | Q9NTU7 | 20 | 55997357 | 56005519 | 1 | 552.25 | 0.45 | 0.26 | 0.81 | 0.007 | 0.869 |
| CD101 | OID31480 | Oncology | CD101 | Q93033 | 1 | 117001750 | 117036552 | 1 | 8333.08 | 0.85 | 0.75 | 0.96 | 0.010 | 0.869 |
| DNPH1 | OID20744 | Inflammat | DNPH1 | O43598 | 6 | 43225629 | 43229481 | 1 | 1469.44 | 0.67 | 0.49 | 0.91 | 0.011 | 0.869 |
| EPS8L2 | OID21397 | Oncology | EPS8L2 | Q9H6S3 | 11 | 694438 | 727727 | 1 | 1552.36 | 0.67 | 0.49 | 0.91 | 0.011 | 0.869 |
| KLK7 | OID30652 | Inflammat | KLK7 | P49862 | 19 | 50976468 | 50984099 | 1 | 2841.71 | 0.75 | 0.60 | 0.94 | 0.013 | 0.869 |
| LYPD8 | OID21251 | Oncology | LYPD8 | Q6UX82 | 1 | 248739415 | 248755759 | 1 | 4235.01 | 1.26 | 1.05 | 1.51 | 0.011 | 0.869 |
| PVR | OID21011 | Neurology | PVR | P15151 | 19 | 44643798 | 44666162 | 1 | 7627.11 | 1.21 | 1.04 | 1.39 | 0.012 | 0.869 |
| SLMAP | OID31441 | Oncology | SLMAP | Q14BN4 | 3 | 57755450 | 57930003 | 1 | 78.77 | 6.57 | 1.66 | 25.90 | 0.007 | 0.869 |
| TNFSF11 | OID20592 | Inflammat | TNFSF11 | O14788 | 13 | 42562736 | 42608013 | 1 | 256.00 | 0.33 | 0.14 | 0.78 | 0.011 | 0.869 |
| TREML2 | OID21120 | Neurology | TREML2 | Q5T2D2 | 6 | 41189749 | 41201149 | 1 | 1163.57 | 1.58 | 1.11 | 2.24 | 0.010 | 0.869 |
| WFIKKN2 | OID20785 | Inflammat | WFIKKN2 | Q8TEU8 | 17 | 50834650 | 50842353 | 1 | 7225.00 | 0.82 | 0.70 | 0.95 | 0.008 | 0.869 |
| AHSG | OID30706 | Inflammat | AHSG | P02765 | 3 | 186613060 | 186621318 | 1 | 7140.25 | 1.20 | 1.03 | 1.39 | 0.017 | 0.876 |
| AMN | OID20492 | Inflammat | AMN | Q9BXJ7 | 14 | 102922663 | 102933596 | 1 | 7319.75 | 0.86 | 0.75 | 0.99 | 0.031 | 0.876 |
| BCHE | OID30717 | Inflammat | BCHE | P06276 | 3 | 165772904 | 165837462 | 1 | 4225.00 | 1.24 | 1.01 | 1.52 | 0.038 | 0.876 |
| BPIFB2 | OID30311 | Cardiome | BPIFB2 | Q8N4F0 | 20 | 33007704 | 33023703 | 1 | 1097.27 | 1.46 | 1.02 | 2.10 | 0.040 | 0.876 |
| CDH2 | OID20170 | Cardiome | CDH2 | P19022 | 18 | 27932879 | 28177946 | 1 | 208.18 | 2.92 | 1.05 | 8.17 | 0.041 | 0.876 |
| CEACAM1 | OID21528 | Oncology | CEACAM | P13688 | 19 | 42507304 | 42561234 | 1 | 1380.41 | 1.58 | 1.08 | 2.31 | 0.019 | 0.876 |
| CFHR4 | OID30758 | Inflammat | CFHR4 | Q92496 | 1 | 196888014 | 196918795 | 1 | 7763.57 | 1.16 | 1.01 | 1.34 | 0.041 | 0.876 |
| CLEC14A | OID21090 | Neurology | CLEC14A | Q86T13 | 14 | 38254000 | 38256093 | 1 | 144.00 | 0.29 | 0.09 | 0.93 | 0.037 | 0.876 |
| CLEC5A | OID20165 | Cardiome | CLEC5A | Q9NY25 | 7 | 141927357 | 141947007 | 1 | 1278.06 | 0.67 | 0.46 | 0.96 | 0.030 | 0.876 |
| CNPY4 | OID21317 | Oncology | CNPY4 | Q8N129 | 7 | 100119634 | 100125508 | 1 | 498.78 | 0.58 | 0.35 | 0.97 | 0.038 | 0.876 |
| CNTN4 | OID21117 | Neurology | CNTN4 | Q8IWV2 | 3 | 2098813 | 3057959 | 1 | 998.56 | 0.62 | 0.42 | 0.93 | 0.021 | 0.876 |
| COL6A3 | OID20292 | Cardiome | COL6A3 | P12111 | 2 | 237324003 | 237414328 | 1 | 132.25 | 3.37 | 1.15 | 9.87 | 0.027 | 0.876 |
| COQ7 | OID30217 | Cardiome | COQ7 | Q99807 | 16 | 19067614 | 19080095 | 1 | 228.77 | 2.51 | 1.13 | 5.60 | 0.024 | 0.876 |
| CPE | OID21392 | Oncology | CPE | P16870 | 4 | 165361194 | 165498547 | 1 | 1024.00 | 1.60 | 1.08 | 2.36 | 0.020 | 0.876 |
| CRIM1 | OID20701 | Inflammat | CRIM1 | Q9NZV1 | 2 | 36355778 | 36551135 | 1 | 289.00 | 0.39 | 0.18 | 0.83 | 0.015 | 0.876 |
| CTHRC1 | OID30393 | Cardiome | CTHRC1 | Q96CG8 | 8 | 103371538 | 103382989 | 1 | 535.59 | 1.92 | 1.07 | 3.45 | 0.028 | 0.876 |
| CTRB1 | OID21150 | Neurology | CTRB1 | P17538 | 16 | 75218988 | 75226338 | 1 | 5148.06 | 0.80 | 0.67 | 0.96 | 0.016 | 0.876 |
| CX3CL1 | OID20976 | Neurology | CX3CL1 | P78423 | 16 | 57372477 | 57385044 | 1 | 841.00 | 0.62 | 0.40 | 0.95 | 0.028 | 0.876 |
| CXCL17 | OID20622 | Inflammat | CXCL17 | Q6UXB2 | 19 | 42428278 | 42442946 | 1 | 96.69 | 0.14 | 0.03 | 0.72 | 0.019 | 0.876 |
| DPP6 | OID21354 | Oncology | DPP6 | P42658 | 7 | 153887097 | 154894285 | 1 | 1064.39 | 1.55 | 1.03 | 2.31 | 0.034 | 0.876 |
| DSCAM | OID30993 | Neurology | DSCAM | O60469 | 21 | 40010999 | 40847158 | 1 | 2256.25 | 1.31 | 1.03 | 1.66 | 0.027 | 0.876 |
| FLT4 | OID21491 | Oncology | FLT4 | P35916 | 5 | 180601506 | 180649624 | 1 | 1962.49 | 1.50 | 1.05 | 2.16 | 0.027 | 0.876 |
| GAL | OID20727 | Inflammat | GAL | P22466 | 11 | 68683779 | 68691175 | 1 | 136.71 | 3.81 | 1.32 | 10.95 | 0.013 | 0.876 |
| GRN | OID21159 | Neurology | GRN | P28799 | 17 | 44345246 | 44353106 | 1 | 976.56 | 0.62 | 0.41 | 0.94 | 0.024 | 0.876 |
| HMBS | OID21385 | Oncology | HMBS | P08397 | 11 | 119084866 | 119093834 | 1 | 327.44 | 1.95 | 1.04 | 3.67 | 0.039 | 0.876 |
| IFNGR1 | OID20678 | Inflammat | IFNGR1 | P15260 | 6 | 137197485 | 137219449 | 1 | 503.75 | 0.56 | 0.33 | 0.95 | 0.032 | 0.876 |
| IL10RB | OID20696 | Inflammat | IL10RB | Q08334 | 21 | 33266367 | 33310187 | 1 | 5776.00 | 1.22 | 1.02 | 1.46 | 0.028 | 0.876 |
| IST1 | OID31497 | Oncology | IST1 | P53990 | 16 | 71845996 | 71931199 | 1 | 105.68 | 0.32 | 0.11 | 0.92 | 0.034 | 0.876 |
| ITGAL | OID30537 | Inflammat | ITGAL | P20701 | 16 | 30472658 | 30523567 | 1 | 380.25 | 0.53 | 0.29 | 0.98 | 0.042 | 0.876 |
| KLK11 | OID21518 | Oncology | KLK11 | Q9UBX7 | 19 | 51022216 | 51028039 | 1 | 2983.24 | 0.79 | 0.64 | 0.98 | 0.034 | 0.876 |
| LGALS9 | OID20781 | Inflammat | LGALS9 | O00182 | 17 | 27629798 | 27649560 | 1 | 4128.06 | 0.78 | 0.63 | 0.96 | 0.021 | 0.876 |
| LPO | OID20963 | Neurology | LPO | P22079 | 17 | 58218548 | 58268518 | 1 | 1163.57 | 1.51 | 1.03 | 2.21 | 0.035 | 0.876 |
| LRG1 | OID30751 | Inflammat | LRG1 | P02750 | 19 | 4536402 | 4540036 | 1 | 6400.00 | 1.24 | 1.04 | 1.47 | 0.014 | 0.876 |
| MEP1A | OID31459 | Oncology | MEP1A | Q16819 | 6 | 46793389 | 46839778 | 1 | 768.08 | 0.56 | 0.34 | 0.92 | 0.022 | 0.876 |
| MET | OID20269 | Cardiome | MET | P08581 | 7 | 116672196 | 116798377 | 1 | 400.00 | 2.01 | 1.08 | 3.75 | 0.028 | 0.876 |
| MFAP4 | OID30598 | Inflammat | MFAP4 | P55083 | 17 | 19383442 | 19387190 | 1 | 834.05 | 1.99 | 1.13 | 3.48 | 0.016 | 0.876 |
| MPHOSPH8 | OID20092 | Cardiome | MPHOSP | Q99549 | 13 | 19633659 | 19673441 | 1 | 53.78 | 5.09 | 1.08 | 23.99 | 0.040 | 0.876 |
| MRI1 | OID30891 | Neurology | MRI1 | Q9BV20 | 19 | 13764522 | 13774282 | 1 | 1881.39 | 1.34 | 1.01 | 1.77 | 0.040 | 0.876 |
| NRP2 | OID21081 | Neurology | NRP2 | O60462 | 2 | 205681990 | 205798133 | 1 | 503.75 | 1.90 | 1.07 | 3.36 | 0.029 | 0.876 |
| NUDT2 | OID21308 | Oncology | NUDT2 | P50583 | 9 | 34329506 | 34343711 | 1 | 1102.24 | 0.65 | 0.43 | 0.98 | 0.038 | 0.876 |
| PLA2G4A | OID20685 | Inflammat | PLA2G4A | P47712 | 1 | 186828949 | 186988981 | 1 | 86.22 | 0.21 | 0.05 | 0.90 | 0.035 | 0.876 |
| PRDX1 | OID20962 | Neurology | PRDX1 | Q06830 | 1 | 45510914 | 45542732 | 1 | 82.51 | 0.22 | 0.06 | 0.86 | 0.029 | 0.876 |
| PROC | OID20402 | Cardiome | PROC | P04070 | 2 | 127418427 | 127429242 | 1 | 808.18 | 1.71 | 1.05 | 2.77 | 0.029 | 0.876 |
| RNASE4 | OID30705 | Inflammat | RNASE4 | P34096 | 14 | 20684560 | 20701216 | 1 | 1644.75 | 1.46 | 1.05 | 2.02 | 0.023 | 0.876 |
| SAA4 | OID30726 | Inflammat | SAA4 | P35542 | 11 | 18231355 | 18236802 | 1 | 2070.25 | 0.76 | 0.58 | 0.98 | 0.037 | 0.876 |
| SCLY | OID21425 | Oncology | SCLY | Q96I15 | 2 | 238060924 | 238099413 | 1 | 588.06 | 1.77 | 1.04 | 3.00 | 0.036 | 0.876 |
| SDCCAG8 | OID31423 | Oncology | SDCCAG | Q86SQ7 | 1 | 243256034 | 243500091 | 1 | 841.00 | 0.59 | 0.37 | 0.94 | 0.026 | 0.876 |
| SELENOP | OID30745 | Inflammat | SELENOF | P49908 | 5 | 42799880 | 42887392 | 1 | 130.98 | 3.52 | 1.21 | 10.24 | 0.021 | 0.876 |
| SERPINB8 | OID20630 | Inflammat | SERPINB | P50452 | 18 | 63970029 | 64019779 | 1 | 8258.27 | 1.15 | 1.01 | 1.31 | 0.042 | 0.876 |
| SLITRK6 | OID20094 | Cardiome | SLITRK6 | Q9H5Y7 | 13 | 85792790 | 85806683 | 1 | 2367.33 | 0.77 | 0.62 | 0.96 | 0.019 | 0.876 |
| SPINK5 | OID21148 | Neurology | SPINK5 | Q9NQ38 | 5 | 148025683 | 148137382 | 1 | 2626.56 | 1.32 | 1.04 | 1.68 | 0.023 | 0.876 |
| SPINT1 | OID21109 | Neurology | SPINT1 | O43278 | 15 | 40844018 | 40858207 | 1 | 1785.06 | 0.74 | 0.56 | 0.98 | 0.036 | 0.876 |
| TGFBR2 | OID21493 | Oncology | TGFBR2 | P37173 | 3 | 30606601 | 30694142 | 1 | 40.64 | 8.93 | 1.26 | 63.52 | 0.029 | 0.876 |
| TNFRSF12A | OID21482 | Oncology | TNFRSF1 | Q9NP84 | 16 | 3018445 | 3022383 | 1 | 78.03 | 4.95 | 1.30 | 18.86 | 0.019 | 0.876 |
| TPSD1 | OID30982 | Neurology | TPSD1 | Q9BZJ3 | 16 | 1256059 | 1259008 | 1 | 1113.89 | 0.69 | 0.48 | 0.99 | 0.042 | 0.876 |
| TTR | OID30782 | Inflammat | TTR | P02766 | 18 | 31557009 | 31598833 | 1 | 284.16 | 2.32 | 1.03 | 5.22 | 0.042 | 0.876 |
| TWF2 | OID30386 | Cardiome | TWF2 | Q6IBS0 | 3 | 52228612 | 52246788 | 1 | 66.31 | 6.01 | 1.16 | 31.22 | 0.033 | 0.876 |
| USP8 | OID20110 | Cardiome | USP8 | P40818 | 15 | 50424380 | 50514421 | 1 | 102.23 | 0.23 | 0.06 | 0.89 | 0.033 | 0.876 |
| VSIG10L | OID30189 | Cardiome | VSIG10L | Q86VR7 | 19 | 51331536 | 51342139 | 1 | 676.00 | 1.69 | 1.06 | 2.70 | 0.026 | 0.876 |
| GRK5 | OID20066 | Cardiome | GRK5 | P34947 | 10 | 119207571 | 119459745 | 1 | 1150.34 | 1.47 | 1.01 | 2.15 | 0.043 | 0.878 |
| FUOM | OID30626 | Inflammat | FUOM | A2VDF0 | 10 | 133355158 | 133358025 | 1 | 573.72 | 0.54 | 0.30 | 0.98 | 0.044 | 0.882 |
| GP6 | OID21091 | Neurology | GP6 | Q9HCN6 | 19 | 55013705 | 55038264 | 1 | 1705.69 | 1.38 | 1.01 | 1.90 | 0.046 | 0.882 |
| IFI30 | OID31505 | Oncology | IFI30 | P13284 | 19 | 18173162 | 18178117 | 1 | 3173.44 | 1.27 | 1.00 | 1.60 | 0.046 | 0.882 |
| IL1R1 | OID21116 | Neurology | IL1R1 | P14778 | 2 | 102064544 | 102179874 | 1 | 529.00 | 0.57 | 0.33 | 0.99 | 0.046 | 0.882 |
| PDCD1LG2 | OID21273 | Oncology | PDCD1LC | Q9BQ51 | 9 | 5510531 | 5571282 | 1 | 7371.45 | 1.17 | 1.00 | 1.37 | 0.047 | 0.882 |
| VSIG2 | OID31371 | Oncology | VSIG2 | Q96IQ7 | 11 | 124747474 | 124752255 | 1 | 708.89 | 0.58 | 0.34 | 0.99 | 0.045 | 0.882 |
| YES1 | OID21348 | Oncology | YES1 | P07947 | 18 | 721588 | 812546 | 1 | 89.20 | 3.76 | 1.03 | 13.70 | 0.045 | 0.882 |
| CD59 | OID20339 | Cardiome | CD59 | P13987 | 11 | 33703010 | 33736479 | 1 | 945.56 | 1.52 | 1.00 | 2.32 | 0.049 | 0.900 |
| HPCAL1 | OID20615 | Inflammat | HPCAL1 | P37235 | 2 | 10302889 | 10427617 | 1 | 396.37 | 1.79 | 1.00 | 3.19 | 0.051 | 0.900 |
| IL6ST | OID20147 | Cardiome | IL6ST | P40189 | 5 | 55935095 | 55995022 | 1 | 562.98 | 1.70 | 1.00 | 2.89 | 0.050 | 0.900 |
| LGALS4 | OID20748 | Inflammat | LGALS4 | P56470 | 19 | 38801671 | 38812945 | 1 | 217.17 | 2.43 | 1.00 | 5.92 | 0.051 | 0.900 |
| NADK | OID20178 | Cardiome | NADK | O95544 | 1 | 1751232 | 1780457 | 1 | 1296.00 | 0.69 | 0.47 | 1.00 | 0.051 | 0.900 |
| WFIKKN1 | OID20939 | Neurology | WFIKKN1 | Q96NZ8 | 16 | 629239 | 634117 | 1 | 1207.56 | 0.68 | 0.46 | 1.00 | 0.050 | 0.900 |
| CFHR2 | OID30720 | Inflammat | CFHR2 | P36980 | 1 | 196943738 | 196959622 | 1 | 13276.16 | 1.12 | 1.00 | 1.25 | 0.054 | 0.910 |
| NSFL1C | OID21022 | Neurology | NSFL1C | Q9UNZ2 | 20 | 1442162 | 1473842 | 1 | 100.00 | 3.35 | 0.98 | 11.46 | 0.054 | 0.910 |
| PILRA | OID21129 | Neurology | PILRA | Q9UKJ1 | 7 | 100367530 | 100400096 | 1 | 15625.00 | 1.11 | 1.00 | 1.22 | 0.054 | 0.910 |
| PILRB | OID20197 | Cardiome | PILRB | Q9UKJ0 | 7 | 100352176 | 100367831 | 1 | 20556.39 | 1.09 | 1.00 | 1.19 | 0.054 | 0.910 |
| SERPINA1 | OID30750 | Inflammat | SERPINA | P01009 | 14 | 94376747 | 94390693 | 1 | 431.72 | 0.59 | 0.35 | 1.01 | 0.054 | 0.910 |
| TMPRSS5 | OID20947 | Neurology | TMPRSS5 | Q9H3S3 | 11 | 113687547 | 113706373 | 1 | 9628.52 | 0.89 | 0.78 | 1.00 | 0.055 | 0.910 |
| ACP1 | OID30524 | Inflammat | ACP1 | P24666 | 2 | 264140 | 278283 | 1 | 2244.39 | 0.78 | 0.60 | 1.01 | 0.059 | 0.914 |

| | | | | | | | | | | | | | | |
|---|---|---|---|---|---|---|---|---|---|---|---|---|---|---|
| AMY2A | OID20333 | Cardiome | AMY2A | P04746 | 1 | 103617427 | 103625780 | 1 | 2107.25 | 1.34 | 0.99 | 1.82 | 0.058 | 0.914 |
| CEMIP2 | OID30499 | Inflammat | CEMIP2 | Q9UHN6 | 9 | 71683366 | 71816690 | 1 | 329.16 | 0.50 | 0.24 | 1.02 | 0.058 | 0.914 |
| DEFA1_DEFA | OID20344 | Cardiome | DEFA1;DI | P59665;P | 8;8 | 6977649;69 | 6980092;69 | 1 | 926.86 | 0.69 | 0.46 | 1.01 | 0.058 | 0.914 |
| ENAH | OID20497 | Inflammat | ENAH | Q8N8S7 | 1 | 225486829 | 225653142 | 1 | 189.83 | 2.40 | 0.97 | 5.94 | 0.059 | 0.914 |
| FSTL3 | OID20782 | Inflammat | FSTL3 | O95633 | 19 | 676392 | 683392 | 1 | 118.27 | 3.01 | 0.96 | 9.44 | 0.059 | 0.914 |
| GHR | OID30574 | Inflammat | GHR | P10912 | 5 | 42423439 | 42721878 | 1 | 5792.90 | 1.17 | 1.00 | 1.37 | 0.056 | 0.914 |
| SPINK6 | OID21450 | Oncology | SPINK6 | Q6UWN8 | 5 | 148202794 | 148215137 | 1 | 2630.22 | 1.29 | 0.99 | 1.68 | 0.056 | 0.914 |
| MMP7 | OID20087 | Cardiome | MMP7 | P09237 | 11 | 102520508 | 102530750 | 1 | 1428.84 | 1.36 | 0.99 | 1.87 | 0.060 | 0.914 |
| PCSK7 | OID31410 | Oncology | PCSK7 | Q16549 | 11 | 117204337 | 117232525 | 1 | 1773.35 | 1.35 | 0.99 | 1.83 | 0.060 | 0.914 |
| SULT1A1 | OID21031 | Neurology | SULT1A1 | P50225 | 16 | 28603944 | 28623625 | 1 | 156.25 | 0.38 | 0.14 | 1.05 | 0.063 | 0.953 |
| AARSD1 | OID21311 | Oncology | AARSD1 | Q9BTE6 | 17 | 42950526 | 42964498 | 1 | 272.25 | 0.63 | 0.29 | 1.40 | 0.261 | 0.955 |
| ACADM | OID31277 | Oncology | ACADM | P11310 | 1 | 75724431 | 75787575 | 1 | 315.06 | 1.49 | 0.73 | 3.03 | 0.273 | 0.955 |
| ACADSB | OID30356 | Cardiome | ACADSB | P45954 | 10 | 123008979 | 123058290 | 1 | 713.65 | 1.31 | 0.84 | 2.04 | 0.239 | 0.955 |
| ACE | OID30696 | Inflammat | ACE | P12821 | 17 | 63477061 | 63498380 | 1 | 3136.00 | 0.90 | 0.73 | 1.11 | 0.340 | 0.955 |
| ADA | OID20645 | Inflammat | ADA | P00813 | 20 | 44619522 | 44652233 | 1 | 7281.78 | 1.12 | 0.97 | 1.30 | 0.119 | 0.955 |
| ADA2 | OID20394 | Cardiome | ADA2 | Q9NZK5 | 22 | 17178790 | 17258235 | 1 | 669.52 | 1.50 | 0.93 | 2.41 | 0.097 | 0.955 |
| ADAM23 | OID20651 | Inflammat | ADAM23 | O75077 | 2 | 206443532 | 206621127 | 1 | 5353.36 | 1.10 | 0.93 | 1.31 | 0.268 | 0.955 |
| ADAMTS16 | OID20089 | Cardiome | ADAMTS1 | Q8TE57 | 5 | 5140330 | 5320304 | 1 | 308.76 | 1.98 | 0.86 | 4.52 | 0.107 | 0.955 |
| ADAMTSL2 | OID30370 | Cardiome | ADAMTSI | Q86TH1 | 9 | 133532164 | 133575519 | 1 | 138.72 | 1.67 | 0.61 | 4.57 | 0.314 | 0.955 |
| ADGRD1 | OID30588 | Inflammat | ADGRD1 | Q6QNK2 | 12 | 130953907 | 131141469 | 1 | 3017.67 | 0.88 | 0.69 | 1.12 | 0.310 | 0.955 |
| ADGRE5 | OID20260 | Cardiome | ADGRE5 | P48960 | 19 | 14380501 | 14408725 | 1 | 652.07 | 1.40 | 0.79 | 2.50 | 0.250 | 0.955 |
| ADGRG1 | OID21294 | Oncology | ADGRG1 | Q9Y653 | 16 | 57610652 | 57665580 | 1 | 102.88 | 0.40 | 0.10 | 1.59 | 0.194 | 0.955 |
| AFP | OID20978 | Neurology | AFP | P02771 | 4 | 73431138 | 73456174 | 1 | 706.04 | 0.75 | 0.45 | 1.25 | 0.263 | 0.955 |
| AGER | OID20756 | Inflammat | AGER | Q15109 | 6 | 32180968 | 32184322 | 1 | 451.56 | 1.51 | 0.83 | 2.75 | 0.175 | 0.955 |
| AGRN | OID20786 | Inflammat | AGRN | O00468 | 1 | 1020120 | 1056118 | 1 | 2232.56 | 0.88 | 0.68 | 1.14 | 0.319 | 0.955 |
| AHCY | OID20199 | Cardiome | AHCY | P23526 | 20 | 34280268 | 34311802 | 1 | 2100.69 | 0.87 | 0.67 | 1.13 | 0.292 | 0.955 |
| AHNAK | OID30260 | Cardiome | AHNAK | Q09666 | 11 | 62433542 | 62556235 | 1 | 462.25 | 1.67 | 0.87 | 3.24 | 0.126 | 0.955 |
| AKR1C4 | OID20062 | Cardiome | AKR1C4 | P17516 | 10 | 5195462 | 5218949 | 1 | 46.24 | 0.39 | 0.07 | 2.29 | 0.298 | 0.955 |
| AKR7L | OID30488 | Inflammat | AKR7L | Q8NHP1 | 1 | 19265982 | 19274194 | 1 | 594.61 | 1.44 | 0.86 | 2.43 | 0.168 | 0.955 |
| AKT3 | OID21197 | Oncology | AKT3 | Q9Y243 | 1 | 243488233 | 243851079 | 1 | 52.16 | 4.68 | 0.83 | 26.25 | 0.080 | 0.955 |
| AMBN | OID20437 | Inflammat | AMBN | Q9NP70 | 4 | 70592256 | 70607288 | 1 | 97.79 | 0.47 | 0.14 | 1.59 | 0.224 | 0.955 |
| AMY1A_AMY | OID30707 | Inflammat | AMY1A;A | P0DUB6;F | 1;1;1 | 103655760;1 | 103664554;1 | 1 | 1624.09 | 1.16 | 0.87 | 1.54 | 0.308 | 0.955 |
| ANGPTL1 | OID20211 | Cardiome | ANGPTL1 | O95841 | 1 | 178849535 | 178871077 | 1 | 1260.25 | 0.77 | 0.55 | 1.08 | 0.127 | 0.955 |
| ANGPTL4 | OID20703 | Inflammat | ANGPTL4 | Q9BY76 | 19 | 8363289 | 8374370 | 1 | 1225.00 | 0.78 | 0.56 | 1.09 | 0.151 | 0.955 |
| ANKRD54 | OID21162 | Oncology | ANKRD54 | Q6NXT1 | 22 | 37830855 | 37849327 | 1 | 79.88 | 0.30 | 0.06 | 1.36 | 0.118 | 0.955 |
| AP1G2 | OID31348 | Oncology | AP1G2 | O75843 | 14 | 23559565 | 23568070 | 1 | 218.13 | 1.83 | 0.74 | 4.51 | 0.190 | 0.955 |
| APOA1 | OID30769 | Inflammat | APOA1 | P02647 | 11 | 116835751 | 116837622 | 1 | 83.59 | 0.31 | 0.06 | 1.48 | 0.141 | 0.955 |
| APOA2 | OID30686 | Inflammat | APOA2 | P02652 | 1 | 161222292 | 161223631 | 1 | 70.14 | 0.46 | 0.11 | 1.97 | 0.297 | 0.955 |
| APOC1 | OID30749 | Inflammat | APOC1 | P02654 | 19 | 44914247 | 44919349 | 1 | 1859.77 | 0.83 | 0.62 | 1.11 | 0.216 | 0.955 |
| APOL1 | OID30708 | Inflammat | APOL1 | O14791 | 22 | 36253071 | 36267530 | 1 | 1248.44 | 0.83 | 0.56 | 1.22 | 0.338 | 0.955 |
| APOM | OID20374 | Cardiome | APOM | O95445 | 6 | 31652416 | 31658210 | 1 | 1269.95 | 0.78 | 0.57 | 1.07 | 0.119 | 0.955 |
| APPL2 | OID30590 | Inflammat | APPL2 | Q8NEU8 | 12 | 105173297 | 105236203 | 1 | 260.02 | 2.02 | 0.95 | 4.29 | 0.067 | 0.955 |
| ARG1 | OID21445 | Oncology | ARG1 | P05089 | 6 | 131470832 | 131584332 | 1 | 561.69 | 0.62 | 0.37 | 1.05 | 0.073 | 0.955 |
| ARHGEF10 | OID30994 | Neurology | ARHGEF1 | O15013 | 8 | 1823926 | 1958641 | 1 | 1240.60 | 0.78 | 0.54 | 1.11 | 0.167 | 0.955 |
| ARL2BP | OID30121 | Cardiome | ARL2BP | Q9Y2Y0 | 16 | 57245259 | 57253635 | 1 | 1261.94 | 1.24 | 0.87 | 1.79 | 0.239 | 0.955 |
| ARSB | OID21331 | Oncology | ARSB | P15848 | 5 | 78777209 | 78986087 | 1 | 1350.56 | 1.24 | 0.90 | 1.72 | 0.189 | 0.955 |
| ASGR2 | OID30679 | Inflammat | ASGR2 | P07307 | 17 | 7101322 | 7115700 | 1 | 1650.39 | 1.31 | 0.96 | 1.80 | 0.088 | 0.955 |
| ASPSCR1 | OID31055 | Neurology | ASPSCR1 | Q9BZE9 | 17 | 81976807 | 82017406 | 1 | 708.89 | 1.48 | 0.94 | 2.32 | 0.091 | 0.955 |
| ATXN3 | OID31149 | Neurology | ATXN3 | P54252 | 14 | 92044496 | 92106621 | 1 | 1472.64 | 1.33 | 0.93 | 1.91 | 0.123 | 0.955 |
| B3GNT7 | OID30281 | Cardiome | B3GNT7 | Q8NFL0 | 2 | 231395710 | 231401164 | 1 | 6889.00 | 0.93 | 0.80 | 1.08 | 0.337 | 0.955 |
| B4GAT1 | OID21127 | Neurology | B4GAT1 | O43505 | 11 | 66345374 | 66347629 | 1 | 1232.79 | 0.81 | 0.57 | 1.16 | 0.250 | 0.955 |
| BCAM | OID21076 | Neurology | BCAM | P50895 | 19 | 44809071 | 44821421 | 1 | 1069.29 | 1.31 | 0.85 | 2.01 | 0.220 | 0.955 |
| BCAN | OID20998 | Neurology | BCAN | Q96GW7 | 1 | 156641390 | 156659532 | 1 | 833.77 | 0.80 | 0.52 | 1.23 | 0.302 | 0.955 |
| BCAT1 | OID30312 | Cardiome | BCAT1 | P54687 | 12 | 24810024 | 24949101 | 1 | 1242.56 | 1.31 | 0.91 | 1.88 | 0.150 | 0.955 |
| BGLAP | OID30389 | Cardiome | BGLAP | P02818 | 1 | 156242184 | 156243317 | 1 | 289.00 | 0.69 | 0.34 | 1.39 | 0.298 | 0.955 |
| BMP10 | OID30575 | Inflammat | BMP10 | O95393 | 2 | 68860909 | 68871397 | 1 | 3980.82 | 1.15 | 0.91 | 1.46 | 0.248 | 0.955 |
| BMP6 | OID20111 | Cardiome | BMP6 | P22004 | 6 | 7726099 | 7881728 | 1 | 284.77 | 0.70 | 0.35 | 1.42 | 0.322 | 0.955 |
| BOLA1 | OID30837 | Neurology | BOLA1 | Q9Y3E2 | 1 | 149887890 | 149900798 | 1 | 188.08 | 1.57 | 0.63 | 3.92 | 0.330 | 0.955 |
| BPIFB1 | OID20308 | Cardiome | BPIFB1 | Q8TDL5 | 20 | 33273480 | 33309871 | 1 | 3634.37 | 0.84 | 0.66 | 1.05 | 0.132 | 0.955 |
| BTC | OID21199 | Oncology | BTC | P35070 | 4 | 74744759 | 74794523 | 1 | 2955.40 | 0.89 | 0.71 | 1.12 | 0.321 | 0.955 |
| BTD | OID30754 | Inflammat | BTD | P43251 | 3 | 15601341 | 15722311 | 1 | 8080.01 | 1.11 | 0.96 | 1.30 | 0.167 | 0.955 |
| C1QA | OID20654 | Inflammat | C1QA | P02745 | 1 | 22636628 | 22639678 | 1 | 2355.98 | 1.22 | 0.92 | 1.62 | 0.176 | 0.955 |
| C1QL2 | OID30515 | Inflammat | C1QL2 | Q7Z5L3 | 2 | 119156243 | 119158751 | 1 | 961.00 | 1.38 | 0.89 | 2.15 | 0.149 | 0.955 |
| C1QTNF6 | OID31049 | Neurology | C1QTNF6 | Q9BXI9 | 22 | 37180166 | 37199385 | 1 | 74.77 | 3.13 | 0.66 | 14.81 | 0.149 | 0.955 |
| C1QTNF9 | OID30620 | Inflammat | C1QTNF9 | P0C862 | 13 | 24307166 | 24322535 | 1 | 13248.01 | 0.94 | 0.85 | 1.05 | 0.269 | 0.955 |
| C2orf69 | OID30883 | Neurology | C2orf69 | Q8N8R5 | 2 | 199911293 | 199955935 | 1 | 132.25 | 0.58 | 0.19 | 1.81 | 0.349 | 0.955 |
| C4BPB | OID21481 | Oncology | C4BPB | P20851 | 1 | 207088860 | 207099993 | 1 | 490.31 | 0.73 | 0.41 | 1.29 | 0.272 | 0.955 |
| C5 | OID30744 | Inflammat | C5 | P01031 | 9 | 120932987 | 121075195 | 1 | 319.52 | 0.67 | 0.34 | 1.31 | 0.243 | 0.955 |
| CA11 | OID21195 | Oncology | CA11 | O75493 | 19 | 48637946 | 48646187 | 1 | 59.29 | 2.17 | 0.49 | 9.60 | 0.308 | 0.955 |
| CA12 | OID21344 | Oncology | CA12 | O43570 | 15 | 63321378 | 63381846 | 1 | 1350.56 | 1.20 | 0.84 | 1.73 | 0.318 | 0.955 |
| CA14 | OID21401 | Oncology | CA14 | Q9ULX7 | 1 | 150257251 | 150265078 | 1 | 617.88 | 1.32 | 0.77 | 2.26 | 0.319 | 0.955 |
| CACNB3 | OID31039 | Neurology | CACNB3 | P54284 | 12 | 48813794 | 48828941 | 1 | 174.83 | 2.36 | 0.95 | 5.87 | 0.065 | 0.955 |
| CALCOCO1 | OID21387 | Oncology | CALCOC( | Q9P1Z2 | 12 | 53708517 | 53727745 | 1 | 193.21 | 0.49 | 0.18 | 1.29 | 0.148 | 0.955 |
| CAMKK1 | OID21182 | Oncology | CAMKK1 | Q8N5S9 | 17 | 3860315 | 3894891 | 1 | 1378.27 | 1.31 | 0.92 | 1.87 | 0.135 | 0.955 |
| CASP10 | OID20893 | Neurology | CASP10 | Q92851 | 2 | 201182898 | 201229406 | 1 | 517.56 | 1.44 | 0.86 | 2.42 | 0.166 | 0.955 |
| CCL13 | OID20655 | Inflammat | CCL13 | Q99616 | 17 | 34356480 | 34358610 | 1 | 833.77 | 0.79 | 0.51 | 1.22 | 0.292 | 0.955 |
| CCL14 | OID20401 | Cardiome | CCL14 | Q16627 | 17 | 35983288 | 35987004 | 1 | 3550.76 | 0.89 | 0.74 | 1.09 | 0.261 | 0.955 |
| CCL17 | OID20745 | Inflammat | CCL17 | Q92583 | 16 | 57404767 | 57416063 | 1 | 858.94 | 0.77 | 0.47 | 1.25 | 0.288 | 0.955 |
| CCL2 | OID21004 | Neurology | CCL2 | P13500 | 17 | 34255218 | 34257203 | 1 | 72.25 | 0.34 | 0.07 | 1.72 | 0.192 | 0.955 |
| CCL20 | OID20671 | Inflammat | CCL20 | P78556 | 2 | 227805739 | 227817564 | 1 | 135.14 | 2.45 | 0.86 | 7.01 | 0.094 | 0.955 |
| CCL23 | OID20693 | Inflammat | CCL23 | P55773 | 17 | 36013056 | 36017972 | 1 | 2675.71 | 1.23 | 0.95 | 1.59 | 0.118 | 0.955 |
| CCL25 | OID20674 | Inflammat | CCL25 | O15444 | 19 | 8052318 | 8062660 | 1 | 7319.75 | 1.10 | 0.95 | 1.27 | 0.211 | 0.955 |
| CCL28 | OID20569 | Inflammat | CCL28 | Q9NRJ3 | 5 | 43376645 | 43412391 | 1 | 66.31 | 0.45 | 0.08 | 2.35 | 0.341 | 0.955 |
| CCL4 | OID20695 | Inflammat | CCL4 | P13236 | 17 | 36103827 | 36105621 | 1 | 1540.56 | 0.80 | 0.57 | 1.14 | 0.215 | 0.955 |
| CCN3 | OID20299 | Cardiome | CCN3 | P48745 | 8 | 119416446 | 119424434 | 1 | 1173.06 | 0.71 | 0.49 | 1.04 | 0.082 | 0.955 |
| CCN4 | OID21444 | Oncology | CCN4 | O95388 | 8 | 133191039 | 133231690 | 1 | 3011.27 | 0.86 | 0.69 | 1.08 | 0.194 | 0.955 |
| CCN5 | OID21068 | Neurology | CCN5 | O76076 | 20 | 44714844 | 44728509 | 1 | 816.33 | 1.38 | 0.85 | 2.24 | 0.189 | 0.955 |
| CCS | OID20973 | Neurology | CCS | O14618 | 11 | 66593153 | 66606019 | 1 | 4721.65 | 0.89 | 0.73 | 1.08 | 0.248 | 0.955 |
| CD14 | OID20378 | Cardiome | CD14 | P08571 | 5 | 140631728 | 140633701 | 1 | 1272.11 | 1.21 | 0.85 | 1.72 | 0.297 | 0.955 |
| CD160 | OID20647 | Inflammat | CD160 | O95971 | 1 | 145719471 | 145739288 | 1 | 2425.56 | 0.82 | 0.64 | 1.04 | 0.098 | 0.955 |
| CD164L2 | OID30922 | Neurology | CD164L2 | Q6UWJ8 | 1 | 27379176 | 27383333 | 1 | 3200.33 | 0.90 | 0.72 | 1.12 | 0.341 | 0.955 |
| CD200 | OID20603 | Inflammat | CD200 | P41217 | 3 | 112332347 | 112362812 | 1 | 521.36 | 1.38 | 0.76 | 2.50 | 0.284 | 0.955 |
| CD207 | OID21298 | Oncology | CD207 | Q9UJ71 | 2 | 70830211 | 70835816 | 1 | 3495.77 | 0.83 | 0.67 | 1.03 | 0.089 | 0.955 |
| CD226 | OID30556 | Inflammat | CD226 | Q15762 | 18 | 69831158 | 69961803 | 1 | 1089.00 | 1.19 | 0.83 | 1.70 | 0.345 | 0.955 |
| CD274 | OID20966 | Neurology | CD274 | Q9NZQ7 | 9 | 5450503 | 5470566 | 1 | 1435.57 | 0.78 | 0.57 | 1.06 | 0.106 | 0.955 |
| CD28 | OID21265 | Oncology | CD28 | P10747 | 2 | 203706475 | 203738912 | 1 | 681.21 | 0.79 | 0.50 | 1.27 | 0.333 | 0.955 |
| CD300LG | OID21130 | Neurology | CD300LG | Q6UXG3 | 17 | 43847148 | 43863639 | 1 | 2872.96 | 0.87 | 0.69 | 1.10 | 0.251 | 0.955 |
| CD34 | OID21025 | Neurology | CD34 | P28906 | 1 | 207880972 | 207911402 | 1 | 67.24 | 0.43 | 0.08 | 2.24 | 0.319 | 0.955 |
| CD48 | OID20692 | Inflammat | CD48 | P09326 | 1 | 160678746 | 160711831 | 1 | 7194.12 | 1.10 | 0.95 | 1.28 | 0.203 | 0.955 |
| CD58 | OID20716 | Inflammat | CD58 | P19256 | 1 | 116514534 | 116571039 | 1 | 550.12 | 1.70 | 0.94 | 3.09 | 0.080 | 0.955 |
| CD6 | OID20649 | Inflammat | CD6 | P30203 | 11 | 60971680 | 61020377 | 1 | 10251.56 | 0.93 | 0.83 | 1.05 | 0.270 | 0.955 |
| CD79B | OID20635 | Inflammat | CD79B | P40259 | 17 | 63928740 | 63932336 | 1 | 542.22 | 1.60 | 0.88 | 2.91 | 0.124 | 0.955 |
| CD84 | OID20607 | Inflammat | CD84 | Q9UIB8 | 1 | 160541095 | 160579516 | 1 | 216.51 | 1.51 | 0.67 | 3.39 | 0.321 | 0.955 |
| CDC27 | OID21238 | Oncology | CDC27 | P30260 | 17 | 47117703 | 47189422 | 1 | 109.44 | 2.00 | 0.65 | 6.14 | 0.229 | 0.955 |
| CDH23 | OID31074 | Neurology | CDH23 | Q9H251 | 10 | 71396920 | 71815947 | 1 | 348.44 | 1.41 | 0.70 | 2.82 | 0.336 | 0.955 |
| CDHR1 | OID20808 | Neurology | CDHR1 | Q96JP9 | 10 | 84194537 | 84219621 | 1 | 798.06 | 0.79 | 0.52 | 1.20 | 0.270 | 0.955 |
| CDHR2 | OID21282 | Oncology | CDHR2 | Q9BYE9 | 5 | 176542511 | 176595974 | 1 | 289.00 | 1.56 | 0.62 | 3.93 | 0.340 | 0.955 |

| | | | | | | | | | | | | | | |
|---|---|---|---|---|---|---|---|---|---|---|---|---|---|---|
| CDHR5 | OID20193 | Cardiome | CDHR5 | Q9HBB8 | 11 | 616577 | 626078 | 1 | 4761.00 | 1.14 | 0.94 | 1.37 | 0.185 | 0.955 |
| CDKN1A | OID21319 | Oncology | CDKN1A | P38936 | 6 | 36676460 | 36687337 | 1 | 319.22 | 0.71 | 0.36 | 1.42 | 0.338 | 0.955 |
| CEACAM16 | OID31495 | Oncology | CEACAM | Q2WEN9 | 19 | 44699151 | 44710718 | 1 | 1753.52 | 0.79 | 0.59 | 1.05 | 0.108 | 0.955 |
| CEACAM19 | OID31443 | Oncology | CEACAM | Q7Z692 | 19 | 44662278 | 44684359 | 1 | 801.92 | 0.75 | 0.46 | 1.21 | 0.238 | 0.955 |
| CEACAM20 | OID31308 | Oncology | CEACAM: | Q6UY09 | 19 | 44501677 | 44529788 | 1 | 408.62 | 1.55 | 0.82 | 2.95 | 0.176 | 0.955 |
| CEACAM5 | OID21171 | Oncology | CEACAM! | P06731 | 19 | 41708585 | 41730433 | 1 | 2568.89 | 1.13 | 0.90 | 1.42 | 0.296 | 0.955 |
| CELA3A | OID20298 | Cardiome | CELA3A | P09093 | 1 | 22001657 | 22012542 | 1 | 693.44 | 0.73 | 0.45 | 1.18 | 0.197 | 0.955 |
| CEP152 | OID31249 | Oncology | CEP152 | O94986 | 15 | 48712928 | 48811146 | 1 | 50.93 | 0.21 | 0.03 | 1.52 | 0.123 | 0.955 |
| CEP170 | OID30240 | Cardiome | CEP170 | Q5SW79 | 1 | 243124428 | 243255348 | 1 | 76.56 | 3.61 | 0.90 | 14.48 | 0.071 | 0.955 |
| CFB | OID30755 | Inflammat | CFB | P00751 | 6 | 31945650 | 31952086 | 1 | 2348.52 | 0.86 | 0.66 | 1.12 | 0.251 | 0.955 |
| CFHR5 | OID30716 | Inflammat | CFHR5 | Q9BXR6 | 1 | 196975010 | 197009678 | 1 | 2805.47 | 0.90 | 0.73 | 1.12 | 0.349 | 0.955 |
| CFI | OID30772 | Inflammat | CFI | P05156 | 4 | 109731008 | 109802150 | 1 | 952.16 | 1.28 | 0.83 | 1.98 | 0.267 | 0.955 |
| CHIT1 | OID20312 | Cardiome | CHIT1 | Q13231 | 1 | 203212827 | 203273641 | 1 | 14721.78 | 0.92 | 0.83 | 1.02 | 0.114 | 0.955 |
| CHL1 | OID20366 | Cardiome | CHL1 | O00533 | 3 | 196763 | 409417 | 1 | 1699.98 | 1.25 | 0.93 | 1.69 | 0.143 | 0.955 |
| CHMP1A | OID20930 | Neurology | CHMP1A | Q9HD42 | 16 | 89640816 | 89657738 | 1 | 141.02 | 1.73 | 0.63 | 4.72 | 0.284 | 0.955 |
| CIAPIN1 | OID21501 | Oncology | CIAPIN1 | Q6FI81 | 16 | 57428187 | 57447420 | 1 | 49.00 | 2.77 | 0.51 | 14.91 | 0.236 | 0.955 |
| CKAP4 | OID20617 | Inflammat | CKAP4 | Q07065 | 12 | 106237881 | 106304279 | 1 | 613.94 | 0.72 | 0.43 | 1.22 | 0.222 | 0.955 |
| CLEC12A | OID30552 | Inflammat | CLEC12A | Q5QGZ9 | 12 | 9951316 | 9995694 | 1 | 3012.79 | 1.13 | 0.92 | 1.40 | 0.255 | 0.955 |
| CLEC4D | OID20609 | Inflammat | CLEC4D | Q8WXI8 | 12 | 8509475 | 8522366 | 1 | 4405.64 | 0.85 | 0.71 | 1.02 | 0.077 | 0.955 |
| CLGN | OID30957 | Neurology | CLGN | O14967 | 4 | 140388453 | 140427661 | 1 | 8169.38 | 1.11 | 0.97 | 1.27 | 0.112 | 0.955 |
| CLIC5 | OID30890 | Neurology | CLIC5 | Q9NZA1 | 6 | 45880827 | 46080348 | 1 | 228.77 | 0.48 | 0.22 | 1.08 | 0.075 | 0.955 |
| CLPS | OID21156 | Neurology | CLPS | P04118 | 6 | 35794982 | 35797344 | 1 | 4726.56 | 1.10 | 0.92 | 1.32 | 0.280 | 0.955 |
| CNDP1 | OID20347 | Cardiome | CNDP1 | Q96KN2 | 18 | 74534500 | 74587212 | 1 | 1350.56 | 0.85 | 0.61 | 1.19 | 0.348 | 0.955 |
| CNP | OID30285 | Cardiome | CNP | P09543 | 17 | 41966763 | 41977740 | 1 | 115.56 | 0.52 | 0.15 | 1.76 | 0.293 | 0.955 |
| CNTN5 | OID21006 | Neurology | CNTN5 | O94779 | 11 | 99020949 | 100358885 | 1 | 3711.62 | 1.18 | 0.96 | 1.45 | 0.122 | 0.955 |
| COCH | OID30305 | Cardiome | COCH | O43405 | 14 | 30874514 | 30895065 | 1 | 1806.25 | 0.79 | 0.57 | 1.09 | 0.146 | 0.955 |
| COL24A1 | OID31464 | Oncology | COL24A1 | Q17RW2 | 1 | 85729233 | 86156943 | 1 | 390.82 | 0.59 | 0.33 | 1.08 | 0.089 | 0.955 |
| COL28A1 | OID30969 | Neurology | COL28A1 | Q2UY09 | 7 | 7356203 | 7535873 | 1 | 10111.42 | 0.90 | 0.79 | 1.02 | 0.090 | 0.955 |
| CPA2 | OID21060 | Neurology | CPA2 | P48052 | 7 | 130266863 | 130289798 | 1 | 2127.52 | 1.16 | 0.89 | 1.53 | 0.279 | 0.955 |
| CPB1 | OID20289 | Cardiome | CPB1 | P15086 | 3 | 148791102 | 148860187 | 1 | 205.44 | 1.88 | 0.81 | 4.40 | 0.144 | 0.955 |
| CPB2 | OID30734 | Inflammat | CPB2 | Q96IY4 | 13 | 46053186 | 46105033 | 1 | 7098.06 | 0.87 | 0.74 | 1.02 | 0.078 | 0.955 |
| CPM | OID21054 | Neurology | CPM | P14384 | 12 | 68842197 | 68971570 | 1 | 234.78 | 1.83 | 0.73 | 4.61 | 0.200 | 0.955 |
| CPXM2 | OID30322 | Cardiome | CPXM2 | Q8N436 | 10 | 123706207 | 123940267 | 1 | 3555.14 | 1.13 | 0.92 | 1.38 | 0.250 | 0.955 |
| CRIP2 | OID20901 | Neurology | CRIP2 | P52943 | 14 | 105472962 | 105480162 | 1 | 334.89 | 0.69 | 0.33 | 1.43 | 0.317 | 0.955 |
| CRYGD | OID31144 | Neurology | CRYGD | P07320 | 2 | 208121607 | 208124524 | 1 | 7876.56 | 1.09 | 0.95 | 1.27 | 0.223 | 0.955 |
| CSF3 | OID20491 | Inflammat | CSF3 | P09919 | 17 | 40015361 | 40017813 | 1 | 90.25 | 2.87 | 0.83 | 9.92 | 0.096 | 0.955 |
| CST3 | OID20400 | Cardiome | CST3 | P01034 | 20 | 23626706 | 23638473 | 1 | 1681.00 | 0.82 | 0.58 | 1.16 | 0.270 | 0.955 |
| CTSB | OID20276 | Cardiome | CTSB | P07858 | 8 | 11842524 | 11869533 | 1 | 4312.11 | 1.10 | 0.92 | 1.31 | 0.302 | 0.955 |
| CTSC | OID20679 | Inflammat | CTSC | P53634 | 11 | 88265069 | 88359684 | 1 | 5787.70 | 0.91 | 0.77 | 1.07 | 0.254 | 0.955 |
| CTSS | OID21056 | Neurology | CTSS | P25774 | 1 | 150730079 | 150765957 | 1 | 3038.77 | 0.83 | 0.67 | 1.03 | 0.084 | 0.955 |
| CXCL13 | OID21024 | Neurology | CXCL13 | O43927 | 4 | 77511753 | 77611834 | 1 | 47.27 | 0.41 | 0.07 | 2.35 | 0.314 | 0.955 |
| CXCL9 | OID20675 | Inflammat | CXCL9 | Q07325 | 4 | 76001275 | 76007509 | 1 | 346.89 | 0.67 | 0.33 | 1.39 | 0.285 | 0.955 |
| CYTL1 | OID30374 | Cardiome | CYTL1 | Q9NRR1 | 4 | 5014586 | 5019458 | 1 | 4804.68 | 1.18 | 0.95 | 1.46 | 0.126 | 0.955 |
| DBI | OID21143 | Neurology | DBI | P07108 | 2 | 119366924 | 119372550 | 1 | 1312.05 | 0.82 | 0.58 | 1.16 | 0.253 | 0.955 |
| DCC | OID30953 | Neurology | DCC | P43146 | 18 | 52340197 | 53535903 | 1 | 953.27 | 0.72 | 0.48 | 1.10 | 0.132 | 0.955 |
| DDHD2 | OID31043 | Neurology | DDHD2 | O94830 | 8 | 38225218 | 38275558 | 1 | 146.68 | 1.73 | 0.62 | 4.81 | 0.293 | 0.955 |
| DDX58 | OID21226 | Oncology | DDX58 | O95786 | 9 | 32455302 | 32526208 | 1 | 1305.02 | 1.19 | 0.84 | 1.70 | 0.329 | 0.955 |
| DEFB4A_DEF | OID21373 | Oncology | DEFB4A;I | O15263;C | 8;8 | 7894677;741 | 7896716;741 | 1 | 80.31 | 1.93 | 0.51 | 7.30 | 0.332 | 0.955 |
| DKK1 | OID21066 | Neurology | DKK1 | O94907 | 10 | 52314281 | 52318042 | 1 | 165.77 | 0.51 | 0.20 | 1.28 | 0.152 | 0.955 |
| DLK1 | OID20295 | Cardiome | DLK1 | P80370 | 14 | 100725705 | 100738224 | 1 | 3782.25 | 1.16 | 0.94 | 1.43 | 0.158 | 0.955 |
| DOC2B | OID30927 | Neurology | DOC2B | Q14184 | 17 | 142789 | 181650 | 1 | 70.14 | 2.08 | 0.45 | 9.59 | 0.348 | 0.955 |
| DPP10 | OID20527 | Inflammat | DPP10 | Q8N608 | 2 | 114442299 | 115845780 | 1 | 1390.22 | 0.81 | 0.56 | 1.16 | 0.246 | 0.955 |
| DPP4 | OID20406 | Cardiome | DPP4 | P27487 | 2 | 161992245 | 162074394 | 1 | 819.39 | 1.37 | 0.90 | 2.10 | 0.146 | 0.955 |
| DPY30 | OID21410 | Oncology | DPY30 | Q9C005 | 2 | 31867809 | 32039805 | 1 | 54.60 | 3.30 | 0.70 | 15.62 | 0.133 | 0.955 |
| DSG2 | OID21113 | Neurology | DSG2 | Q14126 | 18 | 31498177 | 31549008 | 1 | 969.88 | 0.78 | 0.50 | 1.20 | 0.257 | 0.955 |
| DTD1 | OID30580 | Inflammat | DTD1 | Q8TEA8 | 20 | 18587942 | 18766644 | 1 | 430.56 | 1.42 | 0.79 | 2.55 | 0.237 | 0.955 |
| DTX3 | OID21349 | Oncology | DTX3 | Q8N9I9 | 12 | 57604622 | 57609804 | 1 | 185.64 | 0.48 | 0.18 | 1.29 | 0.145 | 0.955 |
| DUSP13 | OID31394 | Oncology | DUSP13 | Q6B8I1 | 10 | 75094432 | 75109221 | 1 | 1097.27 | 0.83 | 0.58 | 1.19 | 0.312 | 0.955 |
| ECHDC3 | OID30293 | Cardiome | ECHDC3 | Q96DC8 | 10 | 11742366 | 11764070 | 1 | 5396.60 | 0.91 | 0.77 | 1.07 | 0.264 | 0.955 |
| ECI2 | OID30142 | Cardiome | ECI2 | O75521 | 6 | 4115693 | 4135597 | 1 | 1207.56 | 1.26 | 0.88 | 1.82 | 0.210 | 0.955 |
| EDDM3B | OID31365 | Oncology | EDDM3B | P56851 | 14 | 20768404 | 20770948 | 1 | 1024.00 | 0.73 | 0.39 | 1.37 | 0.325 | 0.955 |
| EFHD1 | OID31044 | Neurology | EFHD1 | Q9BUP0 | 2 | 232606057 | 232682776 | 1 | 625.00 | 1.42 | 0.85 | 2.35 | 0.179 | 0.955 |
| EHBP1 | OID30302 | Cardiome | EHBP1 | Q8NDI1 | 2 | 62673851 | 63046487 | 1 | 469.44 | 0.75 | 0.41 | 1.35 | 0.336 | 0.955 |
| EHD3 | OID30229 | Cardiome | EHD3 | Q9NZN3 | 2 | 31234152 | 31269451 | 1 | 459.86 | 0.63 | 0.36 | 1.09 | 0.099 | 0.955 |
| EIF2AK3 | OID30060 | Cardiome | EIF2AK3 | Q9NZJ5 | 2 | 88556741 | 88691518 | 1 | 228.77 | 0.66 | 0.29 | 1.51 | 0.327 | 0.955 |
| ELN | OID30301 | Cardiome | ELN | P15502 | 7 | 74027789 | 74069907 | 1 | 256.00 | 0.38 | 0.12 | 1.21 | 0.102 | 0.955 |
| ENPP2 | OID20288 | Cardiome | ENPP2 | Q13822 | 8 | 119557086 | 119673453 | 1 | 1400.90 | 1.27 | 0.86 | 1.88 | 0.235 | 0.955 |
| ENPP5 | OID20736 | Inflammat | ENPP5 | Q9UJA9 | 6 | 46159185 | 46170980 | 1 | 12352.73 | 0.93 | 0.82 | 1.06 | 0.287 | 0.955 |
| ENSA | OID31355 | Oncology | ENSA | O43768 | 1 | 150600851 | 150629612 | 1 | 47.54 | 2.43 | 0.45 | 13.30 | 0.305 | 0.955 |
| ENTR1 | OID30231 | Cardiome | ENTR1 | Q96C92 | 9 | 136401922 | 136410614 | 1 | 882.09 | 1.25 | 0.81 | 1.92 | 0.309 | 0.955 |
| EPHA1 | OID20677 | Inflammat | EPHA1 | P21709 | 7 | 143390289 | 143408856 | 1 | 796.49 | 0.75 | 0.46 | 1.24 | 0.259 | 0.955 |
| EPHB4 | OID20242 | Cardiome | EPHB4 | P54760 | 7 | 100802565 | 100827523 | 1 | 484.00 | 0.61 | 0.33 | 1.12 | 0.109 | 0.955 |
| EPPK1 | OID30250 | Cardiome | EPPK1 | P58107 | 8 | 143857324 | 143878467 | 1 | 2445.75 | 1.19 | 0.91 | 1.55 | 0.201 | 0.955 |
| ERBB4 | OID21510 | Oncology | ERBB4 | Q15303 | 2 | 211375717 | 212538841 | 1 | 749.39 | 1.52 | 0.94 | 2.46 | 0.086 | 0.955 |
| ERI1 | OID31317 | Oncology | ERI1 | Q8IV48 | 8 | 9002147 | 9116746 | 1 | 78.77 | 1.89 | 0.50 | 7.15 | 0.345 | 0.955 |
| ESM1 | OID20758 | Inflammat | ESM1 | Q9NQ30 | 5 | 54977867 | 55022671 | 1 | 455.11 | 1.72 | 0.97 | 3.06 | 0.066 | 0.955 |
| ESYT2 | OID31134 | Neurology | ESYT2 | A0FGR8 | 7 | 158730995 | 158830253 | 1 | 330.79 | 0.63 | 0.31 | 1.32 | 0.222 | 0.955 |
| EVI5 | OID30519 | Inflammat | EVI5 | O60447 | 1 | 92508696 | 92792404 | 1 | 259.57 | 1.71 | 0.81 | 3.63 | 0.159 | 0.955 |
| F10 | OID30684 | Inflammat | F10 | P00742 | 13 | 113122799 | 113149529 | 1 | 500.64 | 0.69 | 0.40 | 1.21 | 0.195 | 0.955 |
| F13B | OID30781 | Inflammat | F13B | P05160 | 1 | 197038741 | 197067260 | 1 | 3124.81 | 1.24 | 0.98 | 1.57 | 0.078 | 0.955 |
| F2R | OID20691 | Inflammat | F2R | P25116 | 5 | 76716126 | 76735770 | 1 | 231.04 | 0.53 | 0.24 | 1.17 | 0.115 | 0.955 |
| F3 | OID21440 | Oncology | F3 | P13726 | 1 | 94529173 | 94541759 | 1 | 698.47 | 1.29 | 0.77 | 2.18 | 0.338 | 0.955 |
| FABP3 | OID30351 | Cardiome | FABP3 | P05413 | 1 | 31365253 | 31376850 | 1 | 144.00 | 0.53 | 0.17 | 1.66 | 0.276 | 0.955 |
| FABP5 | OID21043 | Neurology | FABP5 | Q01469 | 8 | 81280536 | 81284777 | 1 | 204.08 | 2.21 | 0.84 | 5.81 | 0.109 | 0.955 |
| FADD | OID20151 | Cardiome | FADD | Q13158 | 11 | 70203296 | 70207390 | 1 | 100.00 | 0.56 | 0.17 | 1.88 | 0.350 | 0.955 |
| FAM172A | OID30248 | Cardiome | FAM172A | Q8WUF8 | 5 | 93617725 | 94111699 | 1 | 301.50 | 0.66 | 0.31 | 1.40 | 0.281 | 0.955 |
| FAM3C | OID20233 | Cardiome | FAM3C | Q92520 | 7 | 121348878 | 121396364 | 1 | 203.06 | 0.58 | 0.23 | 1.50 | 0.265 | 0.955 |
| FCAMR | OID30332 | Cardiome | FCAMR | Q8WWV6 | 1 | 206957965 | 206970625 | 1 | 1998.09 | 1.21 | 0.92 | 1.58 | 0.169 | 0.955 |
| FCER1A | OID30992 | Neurology | FCER1A | P12319 | 1 | 159289714 | 159308224 | 1 | 302.76 | 0.67 | 0.32 | 1.43 | 0.303 | 0.955 |
| FCGR2B | OID21215 | Oncology | FCGR2B | P31994 | 1 | 161663143 | 161678654 | 1 | 3178.14 | 0.90 | 0.73 | 1.12 | 0.346 | 0.955 |
| FCRL6 | OID20529 | Inflammat | FCRL6 | Q6DN72 | 1 | 159800511 | 159816257 | 1 | 9801.00 | 0.92 | 0.81 | 1.05 | 0.195 | 0.955 |
| FCRLB | OID21193 | Oncology | FCRLB | Q6BAA4 | 1 | 161721563 | 161728143 | 1 | 7256.91 | 0.91 | 0.78 | 1.06 | 0.214 | 0.955 |
| FGF23 | OID21411 | Oncology | FGF23 | Q9GZV9 | 12 | 4368227 | 4379712 | 1 | 182.25 | 2.08 | 0.83 | 5.19 | 0.116 | 0.955 |
| FGR | OID20828 | Neurology | FGR | P09769 | 1 | 27612064 | 27635185 | 1 | 163.47 | 2.24 | 0.75 | 6.64 | 0.147 | 0.955 |
| FIS1 | OID20722 | Inflammat | FIS1 | Q9Y3D6 | 7 | 101239458 | 101252316 | 1 | 414.24 | 0.63 | 0.36 | 1.11 | 0.108 | 0.955 |
| FKBP1B | OID20618 | Inflammat | FKBP1B | P68106 | 2 | 24049701 | 24063681 | 1 | 802.78 | 1.38 | 0.86 | 2.20 | 0.182 | 0.955 |
| FLRT2 | OID20946 | Neurology | FLRT2 | O43155 | 14 | 85530144 | 85654428 | 1 | 4886.01 | 0.86 | 0.71 | 1.03 | 0.099 | 0.955 |
| FN1 | OID30787 | Inflammat | FN1 | P02751 | 2 | 215360440 | 215436073 | 1 | 3136.00 | 1.22 | 0.95 | 1.55 | 0.114 | 0.955 |
| FOLH1 | OID30430 | Inflammat | FOLH1 | Q04609 | 11 | 49145092 | 49208638 | 1 | 126.23 | 1.71 | 0.59 | 4.94 | 0.323 | 0.955 |
| FOLR2 | OID21062 | Neurology | FOLR2 | P14207 | 11 | 72216601 | 72221950 | 1 | 233.65 | 1.51 | 0.67 | 3.42 | 0.321 | 0.955 |
| FOLR3 | OID21485 | Oncology | FOLR3 | P41439 | 11 | 72114869 | 72139892 | 1 | 14744.90 | 0.95 | 0.85 | 1.06 | 0.329 | 0.955 |
| FSTL1 | OID30254 | Cardiome | FSTL1 | Q12841 | 3 | 120392293 | 120450993 | 1 | 346.89 | 0.58 | 0.30 | 1.11 | 0.102 | 0.955 |
| FURIN | OID21514 | Oncology | FURIN | P09958 | 15 | 90868588 | 90883564 | 1 | 588.06 | 1.34 | 0.80 | 2.24 | 0.269 | 0.955 |
| FUT8 | OID20992 | Neurology | FUT8 | Q9BYC5 | 14 | 65410592 | 65744121 | 1 | 10336.11 | 0.91 | 0.80 | 1.03 | 0.147 | 0.955 |
| GALNT5 | OID30295 | Cardiome | GALNT5 | Q7Z7M9 | 2 | 157257705 | 157318491 | 1 | 1129.44 | 0.78 | 0.55 | 1.10 | 0.156 | 0.955 |
| GAS2 | OID31285 | Oncology | GAS2 | O43903 | 11 | 22625509 | 22813001 | 1 | 203.06 | 1.60 | 0.64 | 3.98 | 0.315 | 0.955 |
| GAS6 | OID20318 | Cardiome | GAS6 | Q14393 | 13 | 113820549 | 113864076 | 1 | 1785.06 | 0.80 | 0.60 | 1.08 | 0.144 | 0.955 |

| | | | | | | | | | | | | | | |
|---|---|---|---|---|---|---|---|---|---|---|---|---|---|---|
| GBP1 | OID31128 | Neurology | GBP1 | P32455 | 1 | 89051882 | 89065360 | 1 | 3263.89 | 0.85 | 0.69 | 1.03 | 0.103 | 0.955 |
| GDF15 | OID20251 | Cardiome | GDF15 | Q99988 | 19 | 18374731 | 18389176 | 1 | 1897.09 | 0.78 | 0.55 | 1.10 | 0.161 | 0.955 |
| GDNF | OID20861 | Neurology | GDNF | P39905 | 5 | 37812677 | 37840041 | 1 | 913.90 | 0.73 | 0.49 | 1.09 | 0.128 | 0.955 |
| GFRAL | OID31091 | Neurology | GFRAL | Q6UXV0 | 6 | 55327469 | 55402493 | 1 | 1369.00 | 0.81 | 0.57 | 1.17 | 0.267 | 0.955 |
| GGCT | OID30304 | Cardiome | GGCT | O75223 | 7 | 30496621 | 30504841 | 1 | 221.27 | 0.53 | 0.21 | 1.30 | 0.164 | 0.955 |
| GGT1 | OID21075 | Neurology | GGT1 | P19440 | 22 | 24594811 | 24629005 | 1 | 1316.65 | 1.40 | 0.95 | 2.05 | 0.088 | 0.955 |
| GIPC2 | OID31153 | Neurology | GIPC2 | Q8TF65 | 1 | 77979542 | 78138444 | 1 | 1217.23 | 0.74 | 0.52 | 1.06 | 0.104 | 0.955 |
| GLB1 | OID20949 | Neurology | GLB1 | P16278 | 3 | 32996609 | 33097202 | 1 | 1060.90 | 0.74 | 0.49 | 1.12 | 0.154 | 0.955 |
| GLRX5 | OID30663 | Inflammat | GLRX5 | Q86SX6 | 14 | 95533503 | 95544724 | 1 | 243.36 | 2.04 | 0.94 | 4.42 | 0.070 | 0.955 |
| GNAS | OID31008 | Neurology | GNAS | O95467 | 20 | 58839718 | 58911192 | 1 | 87.89 | 0.47 | 0.13 | 1.73 | 0.258 | 0.955 |
| GOLGA3 | OID31026 | Neurology | GOLGA3 | Q08378 | 12 | 132768902 | 132829143 | 1 | 171.90 | 2.15 | 0.83 | 5.57 | 0.117 | 0.955 |
| GPC5 | OID20944 | Neurology | GPC5 | P78333 | 13 | 91398621 | 92873682 | 1 | 9517.09 | 0.94 | 0.83 | 1.06 | 0.341 | 0.955 |
| GPR37 | OID20095 | Cardiome | GPR37 | O15354 | 7 | 124743885 | 124765792 | 1 | 7607.72 | 1.08 | 0.94 | 1.24 | 0.299 | 0.955 |
| GRHPR | OID30289 | Cardiome | GRHPR | Q9UBQ7 | 9 | 37422666 | 37436990 | 1 | 1130.64 | 0.81 | 0.56 | 1.19 | 0.282 | 0.955 |
| GRIK2 | OID30977 | Neurology | GRIK2 | Q13002 | 6 | 100962701 | 102081622 | 1 | 1029.57 | 0.72 | 0.46 | 1.11 | 0.137 | 0.955 |
| GSR | OID30700 | Inflammat | GSR | P00390 | 8 | 30678066 | 30727846 | 1 | 244.14 | 0.61 | 0.27 | 1.39 | 0.241 | 0.955 |
| GSTM4 | OID30164 | Cardiome | GSTM4 | Q03013 | 1 | 109656099 | 109665496 | 1 | 2282.72 | 0.89 | 0.69 | 1.14 | 0.340 | 0.955 |
| GSTT2B | OID31097 | Neurology | GSTT2B | P0CG30 | 22 | 23957414 | 23961195 | 1 | 16802.64 | 1.05 | 0.96 | 1.15 | 0.324 | 0.955 |
| GZMA | OID20663 | Inflammat | GZMA | P12544 | 5 | 55102646 | 55110252 | 1 | 60.49 | 3.07 | 0.61 | 15.38 | 0.173 | 0.955 |
| HBQ1 | OID21433 | Oncology | HBQ1 | P09105 | 16 | 180459 | 181179 | 1 | 2750.42 | 0.82 | 0.64 | 1.06 | 0.134 | 0.955 |
| HCLS1 | OID20648 | Inflammat | HCLS1 | P14317 | 3 | 121631399 | 121660927 | 1 | 111.42 | 1.76 | 0.55 | 5.69 | 0.342 | 0.955 |
| HDDC2 | OID31447 | Oncology | HDDC2 | Q7Z4H3 | 6 | 125219962 | 125302078 | 1 | 7638.76 | 0.89 | 0.78 | 1.03 | 0.108 | 0.955 |
| HDGF | OID21455 | Oncology | HDGF | P51858 | 1 | 156742109 | 156766925 | 1 | 20520.56 | 1.04 | 0.96 | 1.14 | 0.337 | 0.955 |
| HEXIM1 | OID20589 | Inflammat | HEXIM1 | O94992 | 17 | 45148475 | 45152099 | 1 | 87.11 | 0.29 | 0.07 | 1.27 | 0.101 | 0.955 |
| HJV | OID31006 | Neurology | HJV | Q6ZVN8 | 1 | 146017468 | 146036746 | 1 | 388.47 | 0.70 | 0.40 | 1.24 | 0.223 | 0.955 |
| HLA-DRA | OID20520 | Inflammat | HLA-DRA | P01903 | 6 | 32439878 | 32445046 | 1 | 4637.61 | 0.90 | 0.75 | 1.07 | 0.229 | 0.955 |
| HMOX1 | OID20217 | Cardiome | HMOX1 | P09601 | 22 | 35380361 | 35394214 | 1 | 93.85 | 2.24 | 0.57 | 8.90 | 0.250 | 0.955 |
| HNMT | OID20975 | Neurology | HNMT | P50135 | 2 | 137964020 | 138016364 | 1 | 8664.51 | 0.93 | 0.82 | 1.06 | 0.281 | 0.955 |
| HRG | OID30770 | Inflammat | HRG | P04196 | 3 | 186660216 | 186678234 | 1 | 3921.89 | 1.14 | 0.94 | 1.39 | 0.186 | 0.955 |
| HS6ST1 | OID21441 | Oncology | HS6ST1 | O60243 | 2 | 128236716 | 128318868 | 1 | 689.06 | 1.27 | 0.81 | 2.01 | 0.301 | 0.955 |
| HSBP1 | OID30232 | Cardiome | HSBP1 | O75506 | 16 | 83719311 | 83819737 | 1 | 5041.00 | 0.87 | 0.73 | 1.04 | 0.131 | 0.955 |
| HYAL1 | OID20357 | Cardiome | HYAL1 | Q12794 | 3 | 50299890 | 50312381 | 1 | 1177.35 | 1.26 | 0.87 | 1.83 | 0.217 | 0.955 |
| ICAM2 | OID20248 | Cardiome | ICAM2 | P13598 | 17 | 64002594 | 64020634 | 1 | 208.18 | 1.84 | 0.54 | 6.24 | 0.329 | 0.955 |
| ICOSLG | OID21351 | Oncology | ICOSLG | O75144 | 21 | 44217014 | 44240966 | 1 | 1995.11 | 1.16 | 0.90 | 1.50 | 0.254 | 0.955 |
| IDUA | OID21468 | Oncology | IDUA | P35475 | 4 | 986997 | 1004564 | 1 | 4607.02 | 1.11 | 0.93 | 1.33 | 0.261 | 0.955 |
| IFNAR1 | OID31491 | Oncology | IFNAR1 | P17181 | 21 | 33324387 | 33359864 | 1 | 3164.06 | 0.87 | 0.68 | 1.10 | 0.244 | 0.955 |
| IFNGR2 | OID20941 | Neurology | IFNGR2 | P38484 | 21 | 33403413 | 33479348 | 1 | 16970.98 | 0.91 | 0.83 | 1.01 | 0.073 | 0.955 |
| IGDCC4 | OID30402 | Cardiome | IGDCC4 | Q8TDY8 | 15 | 65381484 | 65422947 | 1 | 1374.70 | 1.30 | 0.94 | 1.78 | 0.110 | 0.955 |
| IGF2R | OID21146 | Neurology | IGF2R | P11717 | 6 | 159969082 | 160113507 | 1 | 2554.84 | 1.14 | 0.89 | 1.45 | 0.298 | 0.955 |
| IGFBP3 | OID20398 | Cardiome | IGFBP3 | P17936 | 7 | 45912245 | 45921874 | 1 | 4337.16 | 1.12 | 0.91 | 1.38 | 0.276 | 0.955 |
| IGLC2 | OID30740 | Inflammat | IGLC2 | P0DOY2 | 22 | 22900976 | 22901437 | 1 | 364.17 | 0.62 | 0.32 | 1.20 | 0.157 | 0.955 |
| IGSF21 | OID30210 | Cardiome | IGSF21 | Q96ID5 | 1 | 18107798 | 18378483 | 1 | 361.00 | 0.61 | 0.32 | 1.19 | 0.146 | 0.955 |
| IGSF8 | OID20167 | Cardiome | IGSF8 | Q969P0 | 1 | 160091340 | 160098943 | 1 | 300.44 | 0.64 | 0.30 | 1.35 | 0.244 | 0.955 |
| IL10RA | OID20420 | Inflammat | IL10RA | Q13651 | 11 | 117986370 | 118003037 | 1 | 357.21 | 1.46 | 0.76 | 2.82 | 0.253 | 0.955 |
| IL15RA | OID20498 | Inflammat | IL15RA | Q13261 | 10 | 5943639 | 5978187 | 1 | 1048.14 | 0.73 | 0.49 | 1.08 | 0.110 | 0.955 |
| IL17C | OID20477 | Inflammat | IL17C | Q9P0M4 | 16 | 88638572 | 88640468 | 1 | 225.00 | 0.56 | 0.25 | 1.28 | 0.170 | 0.955 |
| IL18R1 | OID20652 | Inflammat | IL18R1 | Q13478 | 2 | 102311529 | 102398777 | 1 | 9955.60 | 0.90 | 0.79 | 1.03 | 0.130 | 0.955 |
| IL19 | OID20158 | Cardiome | IL19 | Q9UHD0 | 1 | 206770764 | 206842981 | 1 | 1564.64 | 0.83 | 0.61 | 1.14 | 0.248 | 0.955 |
| IL2RA | OID20267 | Cardiome | IL2RA | P01589 | 10 | 6010689 | 6062370 | 1 | 5407.91 | 0.88 | 0.73 | 1.06 | 0.172 | 0.955 |
| IL6R | OID20385 | Cardiome | IL6R | P08887 | 1 | 154405193 | 154469450 | 1 | 19078.52 | 0.95 | 0.87 | 1.04 | 0.238 | 0.955 |
| IL7 | OID20535 | Inflammat | IL7 | P13232 | 8 | 78675743 | 78805523 | 1 | 198.16 | 1.59 | 0.63 | 4.01 | 0.322 | 0.955 |
| IL7R | OID21136 | Neurology | IL7R | P16871 | 5 | 35852695 | 35879603 | 1 | 12348.77 | 1.09 | 0.96 | 1.23 | 0.176 | 0.955 |
| INSL4 | OID31370 | Oncology | INSL4 | Q14641 | 9 | 5231419 | 5235304 | 1 | 472.34 | 0.76 | 0.45 | 1.28 | 0.302 | 0.955 |
| ISLR2 | OID20889 | Neurology | ISLR2 | Q6UXK2 | 15 | 74100311 | 74138540 | 1 | 945.56 | 1.24 | 0.79 | 1.94 | 0.349 | 0.955 |
| ITGA11 | OID20581 | Inflammat | ITGA11 | Q9UKX5 | 15 | 68296532 | 68432163 | 1 | 762.61 | 1.25 | 0.78 | 2.00 | 0.349 | 0.955 |
| ITGA6 | OID20528 | Inflammat | ITGA6 | P23229 | 2 | 172427354 | 172506459 | 1 | 833.77 | 0.71 | 0.46 | 1.11 | 0.136 | 0.955 |
| ITGB2 | OID20315 | Cardiome | ITGB2 | P05107 | 21 | 44885953 | 44931989 | 1 | 274.09 | 0.67 | 0.30 | 1.49 | 0.325 | 0.955 |
| ITGB6 | OID20493 | Inflammat | ITGB6 | P18564 | 2 | 160099667 | 160200313 | 1 | 1305.02 | 0.81 | 0.57 | 1.15 | 0.237 | 0.955 |
| ITGB7 | OID21393 | Oncology | ITGB7 | P26010 | 12 | 53191323 | 53207282 | 1 | 193.85 | 1.91 | 0.73 | 4.95 | 0.186 | 0.955 |
| ITIH1 | OID30722 | Inflammat | ITIH1 | P19827 | 3 | 52777595 | 52792068 | 1 | 306.25 | 0.60 | 0.30 | 1.21 | 0.152 | 0.955 |
| ITIH3 | OID20352 | Cardiome | ITIH3 | Q06033 | 3 | 52794768 | 52809009 | 1 | 1961.22 | 1.26 | 0.91 | 1.73 | 0.162 | 0.955 |
| ITPA | OID30082 | Cardiome | ITPA | Q9BY32 | 20 | 3208868 | 3223870 | 1 | 6241.00 | 0.92 | 0.80 | 1.07 | 0.270 | 0.955 |
| ITPR1 | OID30216 | Cardiome | ITPR1 | Q14643 | 3 | 4493345 | 4847506 | 1 | 132.25 | 0.59 | 0.21 | 1.63 | 0.309 | 0.955 |
| KDR | OID21497 | Oncology | KDR | P35968 | 4 | 55078481 | 55125595 | 1 | 4389.16 | 1.16 | 0.94 | 1.44 | 0.170 | 0.955 |
| KIR2DL2 | OID31431 | Oncology | KIR2DL2 | P43627 | 19 | 54754693 | 54769187 | 1 | 7844.90 | 1.13 | 0.96 | 1.33 | 0.154 | 0.955 |
| KIT | OID20370 | Cardiome | KIT | P10721 | 4 | 54657267 | 54740783 | 1 | 52.89 | 5.87 | 0.89 | 38.63 | 0.066 | 0.955 |
| KLK14 | OID21407 | Oncology | KLK14 | Q9P0G3 | 19 | 51077495 | 51084245 | 1 | 2704.00 | 1.22 | 0.96 | 1.56 | 0.106 | 0.955 |
| KLK3 | OID30364 | Cardiome | KLK3 | P07288 | 19 | 50854915 | 50860764 | 1 | 136.11 | 3.37 | 0.54 | 21.12 | 0.194 | 0.955 |
| KLK6 | OID21526 | Oncology | KLK6 | Q92876 | 19 | 50958631 | 50969673 | 1 | 576.00 | 1.55 | 0.93 | 2.59 | 0.093 | 0.955 |
| KLRK1 | OID31082 | Neurology | KLRK1 | P26718 | 12 | 10372353 | 10391874 | 1 | 4438.89 | 0.90 | 0.74 | 1.10 | 0.308 | 0.955 |
| KYNU | OID20669 | Inflammat | KYNU | Q16719 | 2 | 142877657 | 143055833 | 1 | 1124.22 | 0.71 | 0.46 | 1.09 | 0.116 | 0.955 |
| LAMB1 | OID30280 | Cardiome | LAMB1 | P07942 | 7 | 107923799 | 108003213 | 1 | 6340.14 | 1.12 | 0.96 | 1.30 | 0.152 | 0.955 |
| LATS1 | OID30523 | Inflammat | LATS1 | O95835 | 6 | 149658153 | 149718105 | 1 | 66.02 | 0.33 | 0.07 | 1.48 | 0.148 | 0.955 |
| LCAT | OID30677 | Inflammat | LCAT | P04180 | 16 | 67939750 | 67944131 | 1 | 252.81 | 1.70 | 0.71 | 4.06 | 0.231 | 0.955 |
| LCN15 | OID31032 | Neurology | LCN15 | Q6UWW( | 9 | 136759634 | 136766255 | 1 | 5329.00 | 0.86 | 0.73 | 1.02 | 0.089 | 0.955 |
| LCN2 | OID20392 | Cardiome | LCN2 | P80188 | 9 | 128149071 | 128153453 | 1 | 89.11 | 0.44 | 0.12 | 1.54 | 0.199 | 0.955 |
| LEPR | OID20128 | Cardiome | LEPR | P48357 | 1 | 65420652 | 65641559 | 1 | 761.76 | 1.56 | 0.97 | 2.50 | 0.067 | 0.955 |
| LGALS1 | OID20154 | Cardiome | LGALS1 | P09382 | 22 | 37675636 | 37679802 | 1 | 1251.39 | 1.20 | 0.84 | 1.71 | 0.317 | 0.955 |
| LGALS3 | OID20284 | Cardiome | LGALS3 | P17931 | 14 | 55124110 | 55145423 | 1 | 5819.51 | 0.90 | 0.75 | 1.07 | 0.227 | 0.955 |
| LGALS8 | OID21041 | Neurology | LGALS8 | O00214 | 1 | 236518000 | 236552981 | 1 | 1156.00 | 0.76 | 0.53 | 1.08 | 0.125 | 0.955 |
| LIFR | OID20606 | Inflammat | LIFR | P42702 | 5 | 38474668 | 38608354 | 1 | 400.00 | 0.68 | 0.38 | 1.24 | 0.209 | 0.955 |
| LILRA5 | OID20209 | Cardiome | LILRA5 | A6NI73 | 19 | 54307070 | 54313166 | 1 | 2025.00 | 0.81 | 0.61 | 1.06 | 0.125 | 0.955 |
| LILRA6 | OID30346 | Cardiome | LILRA6 | Q6PI73 | 19 | 54236592 | 54242791 | 1 | 9900.25 | 1.07 | 0.95 | 1.21 | 0.272 | 0.955 |
| LONP1 | OID30219 | Cardiome | LONP1 | P36776 | 19 | 5691834 | 5720572 | 1 | 467.22 | 0.66 | 0.39 | 1.12 | 0.126 | 0.955 |
| LPCAT2 | OID21290 | Oncology | LPCAT2 | Q7L5N7 | 16 | 55509072 | 55586666 | 1 | 197.56 | 1.81 | 0.71 | 4.60 | 0.213 | 0.955 |
| LRCH4 | OID30191 | Cardiome | LRCH4 | O75427 | 7 | 100574011 | 100586129 | 1 | 73.20 | 2.35 | 0.55 | 10.01 | 0.248 | 0.955 |
| LRP11 | OID20185 | Cardiome | LRP11 | Q86VZ4 | 6 | 149818757 | 149864359 | 1 | 5719.14 | 1.13 | 0.96 | 1.33 | 0.135 | 0.955 |
| LRPAP1 | OID21059 | Neurology | LRPAP1 | P30533 | 4 | 3503612 | 3532446 | 1 | 933.64 | 1.31 | 0.78 | 2.21 | 0.307 | 0.955 |
| LRRN1 | OID20438 | Inflammat | LRRN1 | Q6UXK5 | 3 | 3799431 | 3849834 | 1 | 10276.89 | 0.92 | 0.81 | 1.04 | 0.194 | 0.955 |
| LSP1 | OID20530 | Inflammat | LSP1 | P33241 | 11 | 1850904 | 1892267 | 1 | 346.89 | 1.58 | 0.82 | 3.03 | 0.169 | 0.955 |
| LTBP3 | OID21191 | Oncology | LTBP3 | Q9NS15 | 11 | 65538559 | 65558930 | 1 | 3136.00 | 1.15 | 0.92 | 1.43 | 0.219 | 0.955 |
| LTBR | OID20768 | Inflammat | LTBR | P36941 | 12 | 6375045 | 6391571 | 1 | 1511.27 | 0.84 | 0.60 | 1.15 | 0.274 | 0.955 |
| LY75 | OID20580 | Inflammat | LY75 | O60449 | 2 | 159803355 | 159904756 | 1 | 13630.56 | 0.95 | 0.86 | 1.05 | 0.322 | 0.955 |
| LYAR | OID21264 | Oncology | LYAR | Q9NX58 | 4 | 4267701 | 4290154 | 1 | 47.27 | 3.10 | 0.53 | 17.99 | 0.207 | 0.955 |
| LYSMD3 | OID30950 | Neurology | LYSMD3 | Q7Z3D4 | 5 | 90515611 | 90529584 | 1 | 47.27 | 2.65 | 0.45 | 15.55 | 0.280 | 0.955 |
| LYZL2 | OID31180 | Oncology | LYZL2 | Q7Z4W2 | 10 | 30611779 | 30629762 | 1 | 756.25 | 1.47 | 0.92 | 2.34 | 0.104 | 0.955 |
| LZTFL1 | OID30637 | Inflammat | LZTFL1 | Q9NQ48 | 3 | 45823316 | 45916042 | 1 | 361.00 | 1.53 | 0.76 | 3.08 | 0.236 | 0.955 |
| MAN1A2 | OID30262 | Cardiome | MAN1A2 | O60476 | 1 | 117367449 | 117528872 | 1 | 760.18 | 0.72 | 0.44 | 1.18 | 0.195 | 0.955 |
| MAP2K1 | OID31468 | Oncology | MAP2K1 | Q02750 | 15 | 66386837 | 66491656 | 1 | 43.76 | 0.27 | 0.04 | 1.96 | 0.196 | 0.955 |
| MAPK13 | OID31205 | Oncology | MAPK13 | O15264 | 6 | 36127809 | 36144524 | 1 | 53.78 | 0.28 | 0.06 | 1.37 | 0.115 | 0.955 |
| MAPKAPK2 | OID31293 | Oncology | MAPKAPI | P49137 | 1 | 206684905 | 206734281 | 1 | 337.64 | 1.41 | 0.70 | 2.82 | 0.332 | 0.955 |
| MATN2 | OID20767 | Inflammat | MATN2 | O00339 | 8 | 97868840 | 98036724 | 1 | 2450.25 | 0.88 | 0.69 | 1.13 | 0.328 | 0.955 |
| MCAM | OID20343 | Cardiome | MCAM | P43121 | 11 | 119308529 | 119321521 | 1 | 410.06 | 1.59 | 0.80 | 3.17 | 0.189 | 0.955 |
| MDH1 | OID30669 | Inflammat | MDH1 | P40925 | 2 | 63588609 | 63607197 | 1 | 132.25 | 1.94 | 0.61 | 6.17 | 0.262 | 0.955 |
| MECR | OID30274 | Cardiome | MECR | Q9BV79 | 1 | 29192657 | 29230942 | 1 | 94.62 | 2.22 | 0.63 | 7.91 | 0.217 | 0.955 |
| MGLL | OID20711 | Inflammat | MGLL | Q99685 | 3 | 127689062 | 128052190 | 1 | 56.25 | 0.46 | 0.10 | 2.24 | 0.339 | 0.955 |
| MGMT | OID20588 | Inflammat | MGMT | P16455 | 10 | 129467190 | 129770983 | 1 | 2678.06 | 1.24 | 0.98 | 1.57 | 0.080 | 0.955 |

| | | | | | | | | | | | | | | |
|---|---|---|---|---|---|---|---|---|---|---|---|---|---|---|
| MIA | OID21531 | Oncology | MIA | Q16674 | 19 | 40771648 | 40777490 | 1 | 9758.62 | 1.07 | 0.94 | 1.22 | 0.284 | 0.955 |
| MIF | OID21073 | Neurology | MIF | P14174 | 22 | 23894383 | 23895227 | 1 | 189.06 | 1.98 | 0.83 | 4.71 | 0.124 | 0.955 |
| MMP3 | OID21083 | Neurology | MMP3 | P08254 | 11 | 102835801 | 102843609 | 1 | 6373.36 | 0.90 | 0.74 | 1.10 | 0.306 | 0.955 |
| MMP9 | OID21103 | Neurology | MMP9 | P14780 | 20 | 46008908 | 46016561 | 1 | 328.52 | 1.44 | 0.74 | 2.83 | 0.283 | 0.955 |
| MMUT | OID30159 | Cardiome | MMUT | P22033 | 6 | 49430360 | 49463253 | 1 | 165.77 | 0.44 | 0.17 | 1.16 | 0.096 | 0.955 |
| MPI | OID21288 | Oncology | MPI | P34949 | 15 | 74890005 | 74902219 | 1 | 625.00 | 1.42 | 0.88 | 2.31 | 0.153 | 0.955 |
| MSR1 | OID21063 | Neurology | MSR1 | P21757 | 8 | 16107878 | 16567490 | 1 | 2197.62 | 1.29 | 0.92 | 1.80 | 0.145 | 0.955 |
| MUC13 | OID20866 | Neurology | MUC13 | Q9H3R2 | 3 | 124905442 | 124953819 | 1 | 735.01 | 0.75 | 0.48 | 1.17 | 0.203 | 0.955 |
| MYDGF | OID30313 | Cardiome | MYDGF | Q969H8 | 19 | 4641374 | 4670362 | 1 | 139.42 | 0.55 | 0.20 | 1.48 | 0.235 | 0.955 |
| MYOC | OID21121 | Neurology | MYOC | Q99972 | 1 | 171635417 | 171652688 | 1 | 1454.88 | 0.84 | 0.59 | 1.20 | 0.349 | 0.955 |
| MZB1 | OID20732 | Inflammat | MZB1 | Q8WU39 | 5 | 139387467 | 139390081 | 1 | 212.82 | 0.62 | 0.25 | 1.52 | 0.295 | 0.955 |
| NCAM1 | OID20354 | Cardiome | NCAM1 | P13591 | 11 | 112961247 | 113278436 | 1 | 1369.00 | 1.31 | 0.90 | 1.89 | 0.159 | 0.955 |
| NCR1 | OID20566 | Inflammat | NCR1 | O76036 | 19 | 54906148 | 54916140 | 1 | 1073.82 | 1.30 | 0.87 | 1.94 | 0.198 | 0.955 |
| NCS1 | OID21458 | Oncology | NCS1 | P62166 | 9 | 130172404 | 130237303 | 1 | 284.16 | 0.57 | 0.26 | 1.27 | 0.172 | 0.955 |
| NELL2 | OID20706 | Inflammat | NELL2 | Q99435 | 12 | 44508275 | 44921848 | 1 | 617.88 | 1.46 | 0.85 | 2.51 | 0.174 | 0.955 |
| NEO1 | OID31066 | Neurology | NEO1 | Q92859 | 15 | 73051710 | 73305205 | 1 | 76.90 | 0.39 | 0.09 | 1.68 | 0.206 | 0.955 |
| NFKBIE | OID21212 | Oncology | NFKBIE | O00221 | 6 | 44258166 | 44265788 | 1 | 757.70 | 1.33 | 0.83 | 2.14 | 0.236 | 0.955 |
| NFU1 | OID31510 | Oncology | NFU1 | Q9UMS0 | 2 | 69396113 | 69437628 | 1 | 243.95 | 0.66 | 0.31 | 1.38 | 0.267 | 0.955 |
| NID2 | OID21085 | Neurology | NID2 | Q14112 | 14 | 52004809 | 52069059 | 1 | 5184.00 | 0.90 | 0.76 | 1.06 | 0.197 | 0.955 |
| NIT1 | OID30252 | Cardiome | NIT1 | Q86X76 | 1 | 161118086 | 161125445 | 1 | 996.36 | 0.83 | 0.56 | 1.21 | 0.325 | 0.955 |
| NOMO1 | OID21106 | Neurology | NOMO1 | Q15155 | 16 | 14833721 | 14896157 | 1 | 3223.72 | 0.88 | 0.71 | 1.09 | 0.253 | 0.955 |
| NOS1 | OID20863 | Neurology | NOS1 | P29475 | 12 | 117208142 | 117452170 | 1 | 248.94 | 1.59 | 0.69 | 3.70 | 0.278 | 0.955 |
| NPPB | OID20049 | Cardiome | NPPB | P16860 | 1 | 11857464 | 11858945 | 1 | 523.27 | 1.31 | 0.77 | 2.22 | 0.317 | 0.955 |
| NPTX2 | OID30390 | Cardiome | NPTX2 | P47972 | 7 | 98617285 | 98629869 | 1 | 2415.02 | 0.84 | 0.62 | 1.13 | 0.255 | 0.955 |
| NPY | OID21388 | Oncology | NPY | P01303 | 7 | 24284188 | 24291862 | 1 | 509.47 | 0.74 | 0.40 | 1.34 | 0.320 | 0.955 |
| NRP1 | OID20390 | Cardiome | NRP1 | O14786 | 10 | 33177492 | 33336262 | 1 | 1453.52 | 1.18 | 0.85 | 1.64 | 0.313 | 0.955 |
| NRTN | OID20465 | Inflammat | NRTN | Q99748 | 19 | 5805067 | 5828324 | 1 | 47.74 | 0.24 | 0.04 | 1.25 | 0.090 | 0.955 |
| NT5E | OID21498 | Oncology | NT5E | P21589 | 6 | 85449584 | 85495791 | 1 | 5383.89 | 1.09 | 0.92 | 1.28 | 0.331 | 0.955 |
| NTproBNP | OID20125 | Cardiome | NPPB | P16860 | 1 | 11857464 | 11858945 | 1 | 1390.22 | 1.24 | 0.86 | 1.79 | 0.251 | 0.955 |
| NUCB2 | OID21448 | Oncology | NUCB2 | P80303 | 11 | 17208153 | 17349980 | 1 | 2575.56 | 1.17 | 0.92 | 1.48 | 0.192 | 0.955 |
| NUDT16 | OID31484 | Oncology | NUDT16 | Q96DE0 | 3 | 131381671 | 131388830 | 1 | 1131.57 | 0.82 | 0.60 | 1.13 | 0.223 | 0.955 |
| NUMB | OID30568 | Inflammat | NUMB | P49757 | 14 | 73275107 | 73458617 | 1 | 110.25 | 3.15 | 0.88 | 11.21 | 0.077 | 0.955 |
| OGN | OID21123 | Neurology | OGN | P20774 | 9 | 92383268 | 92404696 | 1 | 7273.65 | 0.89 | 0.76 | 1.05 | 0.177 | 0.955 |
| OLFM4 | OID30665 | Inflammat | OLFM4 | Q6UX06 | 13 | 53028813 | 53052057 | 1 | 9109.64 | 0.91 | 0.80 | 1.03 | 0.144 | 0.955 |
| OMG | OID21352 | Oncology | OMG | P23515 | 17 | 31272013 | 31297539 | 1 | 129.13 | 0.54 | 0.19 | 1.58 | 0.262 | 0.955 |
| OMP | OID30835 | Neurology | OMP | P47874 | 11 | 77102840 | 77103331 | 1 | 518.23 | 1.49 | 0.84 | 2.64 | 0.168 | 0.955 |
| OSMR | OID20356 | Cardiome | OSMR | Q99650 | 5 | 38845858 | 38945596 | 1 | 5550.25 | 1.18 | 0.98 | 1.41 | 0.075 | 0.955 |
| PAG1 | OID20108 | Cardiome | PAG1 | Q9NWQ8 | 8 | 80967810 | 81112068 | 1 | 95.06 | 0.36 | 0.10 | 1.25 | 0.106 | 0.955 |
| PAPPA | OID20421 | Inflammat | PAPPA | Q13219 | 9 | 116153791 | 116402321 | 1 | 51.73 | 2.36 | 0.44 | 12.67 | 0.315 | 0.955 |
| PARD3 | OID31301 | Oncology | PARD3 | Q8TEW0 | 10 | 34109560 | 34815325 | 1 | 71.31 | 3.30 | 0.83 | 13.16 | 0.091 | 0.955 |
| PARP1 | OID20500 | Inflammat | PARP1 | P09874 | 1 | 226360210 | 226408154 | 1 | 252.81 | 1.78 | 0.78 | 4.06 | 0.173 | 0.955 |
| PCDH1 | OID20614 | Inflammat | PCDH1 | Q08174 | 5 | 141853090 | 141879246 | 1 | 133.90 | 2.91 | 0.89 | 9.56 | 0.078 | 0.955 |
| PCDH12 | OID31045 | Neurology | PCDH12 | Q9NPG4 | 5 | 141943581 | 141969741 | 1 | 1670.77 | 1.19 | 0.89 | 1.58 | 0.241 | 0.955 |
| PCDH17 | OID20132 | Cardiome | PCDH17 | O14917 | 13 | 57631744 | 57729311 | 1 | 1947.02 | 0.80 | 0.60 | 1.05 | 0.104 | 0.955 |
| PCDHB15 | OID31377 | Oncology | PCDHB15 | Q9Y5E8 | 5 | 141245395 | 141249365 | 1 | 2065.20 | 1.22 | 0.92 | 1.60 | 0.169 | 0.955 |
| PDCD1 | OID21396 | Oncology | PDCD1 | Q15116 | 2 | 241849884 | 241858894 | 1 | 1081.68 | 0.76 | 0.52 | 1.12 | 0.173 | 0.955 |
| PDE5A | OID30584 | Inflammat | PDE5A | O76074 | 4 | 119494397 | 119628804 | 1 | 390.06 | 0.73 | 0.39 | 1.36 | 0.319 | 0.955 |
| PDGFRA | OID20133 | Cardiome | PDGFRA | P16234 | 4 | 54229280 | 54298245 | 1 | 1254.34 | 1.29 | 0.89 | 1.88 | 0.185 | 0.955 |
| PDZK1 | OID30550 | Inflammat | PDZK1 | Q5T2W1 | 1 | 145670852 | 145708148 | 1 | 192.52 | 2.00 | 0.86 | 4.65 | 0.106 | 0.955 |
| PF4 | OID30764 | Inflammat | PF4 | P02776 | 4 | 73980811 | 73982027 | 1 | 124.69 | 1.97 | 0.57 | 6.76 | 0.280 | 0.955 |
| PGA4 | OID30699 | Inflammat | PGA4 | P0DJD7 | 11 | 61222347 | 61231694 | 1 | 60.84 | 0.40 | 0.08 | 1.97 | 0.258 | 0.955 |
| PGD | OID30344 | Cardiome | PGD | P52209 | 1 | 10398592 | 10420511 | 1 | 893.80 | 1.26 | 0.86 | 1.85 | 0.232 | 0.955 |
| PGF | OID20673 | Inflammat | PGF | P49763 | 14 | 74941834 | 74955626 | 1 | 490.31 | 1.74 | 0.95 | 3.19 | 0.075 | 0.955 |
| PHACTR2 | OID31009 | Neurology | PHACTR2 | O75167 | 6 | 143536845 | 143831185 | 1 | 759.31 | 0.79 | 0.49 | 1.28 | 0.345 | 0.955 |
| PIGR | OID21102 | Neurology | PIGR | P01833 | 1 | 206928522 | 206946466 | 1 | 412.90 | 0.63 | 0.37 | 1.09 | 0.099 | 0.955 |
| PKN3 | OID31311 | Oncology | PKN3 | Q6P5Z2 | 9 | 128702503 | 128720916 | 1 | 382.02 | 1.34 | 0.73 | 2.46 | 0.345 | 0.955 |
| PLA2G2A | OID20274 | Cardiome | PLA2G2A | P14555 | 1 | 19975431 | 19980416 | 1 | 6597.05 | 0.91 | 0.78 | 1.06 | 0.235 | 0.955 |
| PLEKHO1 | OID31088 | Neurology | PLEKHO1 | Q53GL0 | 1 | 150149183 | 150164720 | 1 | 60.84 | 0.22 | 0.04 | 1.17 | 0.076 | 0.955 |
| PLIN3 | OID20143 | Cardiome | PLIN3 | O60664 | 19 | 4838341 | 4867694 | 1 | 67.60 | 0.42 | 0.10 | 1.87 | 0.256 | 0.955 |
| PLSCR3 | OID31146 | Neurology | PLSCR3 | Q9NRY6 | 17 | 7389727 | 7394842 | 1 | 74.22 | 0.50 | 0.13 | 1.95 | 0.316 | 0.955 |
| PLTP | OID20238 | Cardiome | PLTP | P55058 | 20 | 45898621 | 45912155 | 1 | 3111.16 | 0.89 | 0.70 | 1.11 | 0.301 | 0.955 |
| PMM2 | OID30125 | Cardiome | PMM2 | O15305 | 16 | 8788823 | 8862534 | 1 | 802.78 | 1.40 | 0.86 | 2.30 | 0.178 | 0.955 |
| PODXL | OID21358 | Oncology | PODXL | O00592 | 7 | 131500262 | 131558217 | 1 | 930.25 | 0.76 | 0.51 | 1.11 | 0.154 | 0.955 |
| POMC | OID30282 | Cardiome | POMC | P01189 | 2 | 25160853 | 25168903 | 1 | 107.27 | 0.43 | 0.11 | 1.63 | 0.215 | 0.955 |
| PON1 | OID30704 | Inflammat | PON1 | P27169 | 7 | 95297676 | 95324532 | 1 | 8953.89 | 1.06 | 0.94 | 1.20 | 0.339 | 0.955 |
| PPL | OID30564 | Inflammat | PPL | O60437 | 16 | 4882507 | 4960741 | 1 | 130.34 | 1.75 | 0.59 | 5.16 | 0.313 | 0.955 |
| PRDX6 | OID21404 | Oncology | PRDX6 | P30041 | 1 | 173477330 | 173488815 | 1 | 302.76 | 0.54 | 0.26 | 1.11 | 0.095 | 0.955 |
| PRG2 | OID30381 | Cardiome | PRG2 | P13727 | 11 | 57386780 | 57390650 | 1 | 176.17 | 1.85 | 0.71 | 4.80 | 0.205 | 0.955 |
| PRKAB1 | OID20444 | Inflammat | PRKAB1 | Q9Y478 | 12 | 119667864 | 119681624 | 1 | 600.25 | 1.30 | 0.80 | 2.11 | 0.292 | 0.955 |
| PRKG1 | OID30621 | Inflammat | PRKG1 | Q13976 | 10 | 50990888 | 52298423 | 1 | 586.72 | 1.47 | 0.88 | 2.47 | 0.142 | 0.955 |
| PRND | OID31124 | Neurology | PRND | Q9UKY0 | 20 | 4721909 | 4728460 | 1 | 1008.06 | 0.83 | 0.56 | 1.22 | 0.335 | 0.955 |
| PSMD9 | OID21304 | Oncology | PSMD9 | O00233 | 12 | 121888732 | 121918297 | 1 | 588.06 | 0.73 | 0.43 | 1.24 | 0.248 | 0.955 |
| PTGDS | OID20408 | Cardiome | PTGDS | P41222 | 9 | 136975092 | 136981742 | 1 | 792.02 | 1.30 | 0.80 | 2.11 | 0.283 | 0.955 |
| PTK7 | OID20880 | Neurology | PTK7 | Q13308 | 6 | 43076307 | 43161719 | 1 | 55.18 | 2.81 | 0.65 | 12.21 | 0.167 | 0.955 |
| PTPRN2 | OID20912 | Neurology | PTPRN2 | Q92932 | 7 | 157539056 | 158587823 | 1 | 375.39 | 1.77 | 0.93 | 3.36 | 0.080 | 0.955 |
| PTPRZ1 | OID30343 | Cardiome | PTPRZ1 | P23471 | 7 | 121873089 | 122062036 | 1 | 582.88 | 0.67 | 0.38 | 1.17 | 0.155 | 0.955 |
| PYY | OID30150 | Cardiome | PYY | P10082 | 17 | 43952733 | 44004469 | 1 | 937.89 | 0.79 | 0.53 | 1.19 | 0.261 | 0.955 |
| QSOX1 | OID30729 | Inflammat | QSOX1 | O00391 | 1 | 180154869 | 180204030 | 1 | 6978.67 | 0.93 | 0.80 | 1.07 | 0.315 | 0.955 |
| RAB11FIP3 | OID30325 | Cardiome | RAB11FIF | O75154 | 16 | 425649 | 523011 | 1 | 56.25 | 0.40 | 0.08 | 2.00 | 0.266 | 0.955 |
| RASSF2 | OID21194 | Oncology | RASSF2 | P50749 | 20 | 4780023 | 4823608 | 1 | 141.02 | 0.53 | 0.19 | 1.44 | 0.211 | 0.955 |
| RBM17 | OID31034 | Neurology | RBM17 | Q96I25 | 10 | 6089034 | 6117457 | 1 | 357.21 | 1.46 | 0.72 | 2.96 | 0.296 | 0.955 |
| RBP5 | OID21369 | Oncology | RBP5 | P82980 | 12 | 7115736 | 7128889 | 1 | 1545.09 | 1.24 | 0.88 | 1.74 | 0.216 | 0.955 |
| RECK | OID30234 | Cardiome | RECK | O95980 | 9 | 36036913 | 36124455 | 1 | 865.17 | 0.79 | 0.52 | 1.18 | 0.248 | 0.955 |
| REG3A | OID20258 | Cardiome | REG3A | Q06141 | 2 | 79157003 | 79159753 | 1 | 736.22 | 1.53 | 0.97 | 2.42 | 0.067 | 0.955 |
| RELT | OID21142 | Neurology | RELT | Q969Z4 | 11 | 73376399 | 73397474 | 1 | 2487.52 | 1.21 | 0.91 | 1.62 | 0.188 | 0.955 |
| RET | OID21346 | Oncology | RET | P07949 | 10 | 43077064 | 43130351 | 1 | 2218.41 | 1.20 | 0.93 | 1.55 | 0.157 | 0.955 |
| RNASE6 | OID30689 | Inflammat | RNASE6 | Q93091 | 14 | 20781268 | 20782467 | 1 | 2444.75 | 0.89 | 0.69 | 1.14 | 0.348 | 0.955 |
| RNF43 | OID31291 | Oncology | RNF43 | Q68DV7 | 17 | 58353676 | 58417595 | 1 | 159.68 | 2.18 | 0.89 | 5.36 | 0.090 | 0.955 |
| ROBO1 | OID20734 | Inflammat | ROBO1 | Q9Y6N7 | 3 | 78597239 | 79767998 | 1 | 687.60 | 1.32 | 0.81 | 2.17 | 0.266 | 0.955 |
| ROBO2 | OID21007 | Neurology | ROBO2 | Q9HCK4 | 3 | 75906695 | 77649964 | 1 | 216.51 | 1.61 | 0.64 | 4.05 | 0.313 | 0.955 |
| ROBO4 | OID30226 | Cardiome | ROBO4 | Q8WZ75 | 11 | 124883691 | 124898500 | 1 | 133.53 | 0.60 | 0.21 | 1.71 | 0.337 | 0.955 |
| RPE | OID31324 | Oncology | RPE | Q96AT9 | 2 | 210002565 | 210022260 | 1 | 50.77 | 3.61 | 0.69 | 19.00 | 0.129 | 0.955 |
| RSPO1 | OID20938 | Neurology | RSPO1 | Q2MKA7 | 1 | 37611350 | 37634892 | 1 | 892.52 | 1.29 | 0.86 | 1.94 | 0.217 | 0.955 |
| RSPO3 | OID21415 | Oncology | RSPO3 | Q9BXY4 | 6 | 127118671 | 127199481 | 1 | 503.04 | 1.36 | 0.74 | 2.49 | 0.326 | 0.955 |
| RTN4R | OID21424 | Oncology | RTN4R | Q9BZR6 | 22 | 20241415 | 20283246 | 1 | 1096.35 | 0.73 | 0.50 | 1.05 | 0.093 | 0.955 |
| RWDD1 | OID21015 | Neurology | RWDD1 | Q9H446 | 6 | 116571409 | 116597675 | 1 | 290.89 | 0.53 | 0.23 | 1.22 | 0.137 | 0.955 |
| S100A12 | OID21374 | Oncology | S100A12 | P80511 | 1 | 153373711 | 153375621 | 1 | 877.64 | 0.69 | 0.45 | 1.07 | 0.100 | 0.955 |
| S100A16 | OID20878 | Neurology | S100A16 | Q96FQ6 | 1 | 153606886 | 153613145 | 1 | 169.00 | 1.58 | 0.63 | 3.96 | 0.334 | 0.955 |
| SCAMP3 | OID21332 | Oncology | SCAMP3 | O14828 | 1 | 155255979 | 155262430 | 1 | 89.20 | 2.26 | 0.63 | 8.14 | 0.213 | 0.955 |
| SCARA5 | OID21058 | Neurology | SCARA5 | Q6ZMJ2 | 8 | 27869883 | 27992673 | 1 | 1774.52 | 0.81 | 0.59 | 1.10 | 0.173 | 0.955 |
| SCARB2 | OID20943 | Neurology | SCARB2 | Q14108 | 4 | 76158737 | 76234536 | 1 | 992.25 | 0.81 | 0.54 | 1.22 | 0.317 | 0.955 |
| SCARF1 | OID20297 | Cardiome | SCARF1 | Q14162 | 17 | 1633858 | 1645744 | 1 | 2244.39 | 0.86 | 0.67 | 1.12 | 0.264 | 0.955 |
| SCGB3A1 | OID30577 | Inflammat | SCGB3A1 | Q96QR1 | 5 | 180590105 | 180591499 | 1 | 284.16 | 2.10 | 0.93 | 4.70 | 0.073 | 0.955 |
| SCGB3A2 | OID20723 | Inflammat | SCGB3A2 | Q96PL1 | 5 | 147870682 | 147882191 | 1 | 759.31 | 0.70 | 0.44 | 1.12 | 0.142 | 0.955 |
| SDC1 | OID20169 | Cardiome | SDC1 | P18827 | 2 | 20200797 | 20225433 | 1 | 350.22 | 0.67 | 0.32 | 1.38 | 0.273 | 0.955 |
| SDC4 | OID20216 | Cardiome | SDC4 | P31431 | 20 | 45325288 | 45348424 | 1 | 1014.88 | 0.77 | 0.51 | 1.18 | 0.233 | 0.955 |
| SDHB | OID31436 | Oncology | SDHB | P21912 | 1 | 17018722 | 17054032 | 1 | 56.25 | 2.39 | 0.45 | 12.55 | 0.304 | 0.955 |

| | | | | | | | | | | | | | | |
|---|---|---|---|---|---|---|---|---|---|---|---|---|---|---|
| SEL1L | OID30196 | Cardiome | SEL1L | Q9UBV2 | 14 | 81471547 | 81533853 | 1 | 1538.11 | 0.79 | 0.60 | 1.04 | 0.096 | 0.955 |
| SEMA7A | OID20283 | Cardiome | SEMA7A | O75326 | 15 | 74409289 | 74433958 | 1 | 370.56 | 1.64 | 0.81 | 3.32 | 0.165 | 0.955 |
| SEPTIN8 | OID31079 | Neurology | SEPTIN8 | Q92599 | 5 | 132750819 | 132807241 | 1 | 173.76 | 2.11 | 0.72 | 6.22 | 0.176 | 0.955 |
| SERPINB5 | OID20064 | Cardiome | SERPINB | P36952 | 18 | 63476958 | 63505085 | 1 | 49.00 | 0.36 | 0.06 | 2.17 | 0.266 | 0.955 |
| SERPINB6 | OID21023 | Neurology | SERPINB | P35237 | 6 | 2948159 | 2972165 | 1 | 405.02 | 0.61 | 0.34 | 1.10 | 0.102 | 0.955 |
| SERPINF2 | OID30779 | Inflammat | SERPINF | P08697 | 17 | 1742836 | 1755265 | 1 | 529.00 | 0.61 | 0.34 | 1.09 | 0.093 | 0.955 |
| SERPINI1 | OID30604 | Inflammat | SERPINI1 | Q99574 | 3 | 167735243 | 167825569 | 1 | 2635.11 | 1.12 | 0.88 | 1.43 | 0.346 | 0.955 |
| SETMAR | OID20948 | Neurology | SETMAR | Q53H47 | 3 | 4303332 | 4317265 | 1 | 952.16 | 1.42 | 0.91 | 2.19 | 0.119 | 0.955 |
| SEZ6L2 | OID21289 | Oncology | SEZ6L2 | Q6UXD5 | 16 | 29871159 | 29899550 | 1 | 535.59 | 1.60 | 0.85 | 2.98 | 0.142 | 0.955 |
| SFTPA1 | OID21165 | Oncology | SFTPA1 | Q8IWL2 | 10 | 79610939 | 79615455 | 1 | 806.56 | 0.77 | 0.51 | 1.17 | 0.217 | 0.955 |
| SFTPA2 | OID21218 | Oncology | SFTPA2 | Q8IWL1 | 10 | 79555852 | 79560407 | 1 | 7302.48 | 1.08 | 0.94 | 1.25 | 0.274 | 0.955 |
| SH3GLB2 | OID31145 | Neurology | SH3GLB2 | Q9NR46 | 9 | 129007036 | 129028331 | 1 | 1521.00 | 0.82 | 0.58 | 1.16 | 0.259 | 0.955 |
| SHBG | OID30685 | Inflammat | SHBG | P04278 | 17 | 7613946 | 7633382 | 1 | 1532.16 | 0.76 | 0.54 | 1.07 | 0.119 | 0.955 |
| SHH | OID30516 | Inflammat | SHH | Q15465 | 7 | 155799980 | 155812463 | 1 | 50.77 | 2.37 | 0.44 | 12.82 | 0.317 | 0.955 |
| SHISA5 | OID30387 | Cardiome | SHISA5 | Q8N114 | 3 | 48467798 | 48504826 | 1 | 290.48 | 1.86 | 0.75 | 4.58 | 0.178 | 0.955 |
| SHPK | OID31292 | Oncology | SHPK | Q9UHJ6 | 17 | 3608240 | 3636250 | 1 | 103.10 | 0.45 | 0.15 | 1.41 | 0.174 | 0.955 |
| SIGLEC5 | OID21082 | Neurology | SIGLEC5 | O15389 | 19 | 51611927 | 51645545 | 1 | 10319.17 | 0.92 | 0.82 | 1.04 | 0.172 | 0.955 |
| SIGLEC7 | OID20190 | Cardiome | SIGLEC7 | Q9Y286 | 19 | 51142299 | 51153526 | 1 | 1792.11 | 1.18 | 0.85 | 1.64 | 0.318 | 0.955 |
| SLAMF1 | OID20496 | Inflammat | SLAMF1 | Q13291 | 1 | 160608106 | 160647044 | 1 | 173.76 | 0.56 | 0.22 | 1.39 | 0.211 | 0.955 |
| SLAMF7 | OID20602 | Inflammat | SLAMF7 | Q9NQ25 | 1 | 160739057 | 160754821 | 1 | 7349.17 | 1.09 | 0.93 | 1.27 | 0.286 | 0.955 |
| SLAMF8 | OID21214 | Oncology | SLAMF8 | Q9P0V8 | 1 | 159826811 | 159837492 | 1 | 12525.34 | 1.09 | 0.97 | 1.22 | 0.132 | 0.955 |
| SLC39A14 | OID20852 | Neurology | SLC39A1 | Q15043 | 8 | 22367278 | 22434129 | 1 | 141.35 | 0.53 | 0.19 | 1.42 | 0.205 | 0.955 |
| SLC4A1 | OID30369 | Cardiome | SLC4A1 | P02730 | 17 | 44248390 | 44268141 | 1 | 65.90 | 0.45 | 0.10 | 2.03 | 0.301 | 0.955 |
| SLIT2 | OID20862 | Neurology | SLIT2 | O94813 | 4 | 20251905 | 20620561 | 1 | 58.14 | 2.79 | 0.53 | 14.69 | 0.227 | 0.955 |
| SMPDL3A | OID20676 | Inflammat | SMPDL3A | Q92484 | 6 | 122789049 | 122809720 | 1 | 8547.84 | 1.10 | 0.96 | 1.27 | 0.158 | 0.955 |
| SOST | OID20204 | Cardiome | SOST | Q9BQB4 | 17 | 43753738 | 43758791 | 1 | 75.94 | 2.40 | 0.49 | 11.80 | 0.282 | 0.955 |
| SPAG1 | OID30979 | Neurology | SPAG1 | Q07617 | 8 | 100157906 | 100259278 | 1 | 7987.89 | 1.13 | 0.99 | 1.29 | 0.065 | 0.955 |
| SPOCK1 | OID20924 | Neurology | SPOCK1 | Q08629 | 5 | 136975298 | 137598379 | 1 | 268.14 | 0.68 | 0.32 | 1.43 | 0.306 | 0.955 |
| SPON2 | OID20221 | Cardiome | SPON2 | Q9BUD6 | 4 | 1166932 | 1208962 | 1 | 689.06 | 0.78 | 0.48 | 1.25 | 0.300 | 0.955 |
| SSC4D | OID20182 | Cardiome | SSC4D | Q8WTU2 | 7 | 76389334 | 76409697 | 1 | 2097.64 | 1.17 | 0.89 | 1.53 | 0.269 | 0.955 |
| SSC5D | OID20351 | Cardiome | SSC5D | A1L4H1 | 19 | 55488404 | 55519099 | 1 | 4539.39 | 0.86 | 0.70 | 1.05 | 0.141 | 0.955 |
| ST6GAL1 | OID20302 | Cardiome | ST6GAL1 | P15907 | 3 | 186930325 | 187078553 | 1 | 506.25 | 1.31 | 0.74 | 2.31 | 0.349 | 0.955 |
| STAB2 | OID30401 | Cardiome | STAB2 | Q8WWQ | 12 | 103587273 | 103766719 | 1 | 3265.31 | 1.17 | 0.94 | 1.46 | 0.161 | 0.955 |
| STC1 | OID21014 | Neurology | STC1 | P52823 | 8 | 23841929 | 23854806 | 1 | 74.70 | 2.35 | 0.59 | 9.39 | 0.228 | 0.955 |
| STC2 | OID21157 | Neurology | STC2 | O76061 | 5 | 173314723 | 173328447 | 1 | 105.06 | 0.55 | 0.16 | 1.90 | 0.343 | 0.955 |
| STK4 | OID20140 | Cardiome | STK4 | Q13043 | 20 | 44966479 | 45080021 | 1 | 43.89 | 2.44 | 0.41 | 14.40 | 0.326 | 0.955 |
| STX16 | OID21170 | Oncology | STX16 | O14662 | 20 | 58651253 | 58679526 | 1 | 60.06 | 3.91 | 0.82 | 18.57 | 0.086 | 0.955 |
| SUMF1 | OID30486 | Inflammat | SUMF1 | Q8NBK3 | 3 | 3700814 | 4467273 | 1 | 189.06 | 1.80 | 0.76 | 4.23 | 0.180 | 0.955 |
| SUMF2 | OID21032 | Neurology | SUMF2 | Q8NBJ7 | 7 | 56064002 | 56080670 | 1 | 3223.72 | 1.16 | 0.90 | 1.48 | 0.252 | 0.955 |
| SUSD2 | OID21098 | Neurology | SUSD2 | Q9UGT4 | 22 | 24181487 | 24189106 | 1 | 4022.23 | 1.14 | 0.94 | 1.38 | 0.180 | 0.955 |
| SUSD5 | OID30650 | Inflammat | SUSD5 | O60279 | 3 | 33150043 | 33218810 | 1 | 2976.31 | 0.85 | 0.66 | 1.10 | 0.214 | 0.955 |
| SV2A | OID30965 | Neurology | SV2A | Q7L0J3 | 1 | 149903318 | 149917844 | 1 | 447.02 | 1.34 | 0.74 | 2.43 | 0.342 | 0.955 |
| TACC3 | OID21285 | Oncology | TACC3 | Q9Y6A5 | 4 | 1712858 | 1745171 | 1 | 656.64 | 1.32 | 0.83 | 2.09 | 0.235 | 0.955 |
| TBCC | OID20942 | Neurology | TBCC | Q15814 | 6 | 42744498 | 42746103 | 1 | 305.72 | 1.79 | 0.78 | 4.10 | 0.168 | 0.955 |
| TDGF1 | OID20816 | Neurology | TDGF1 | P13385 | 3 | 46574534 | 46582457 | 1 | 21478.53 | 1.06 | 0.97 | 1.15 | 0.197 | 0.955 |
| TEX101 | OID30971 | Neurology | TEX101 | Q9BY14 | 19 | 43401496 | 43418597 | 1 | 1722.25 | 1.29 | 0.84 | 1.98 | 0.242 | 0.955 |
| TFPI2 | OID21512 | Oncology | TFPI2 | P48307 | 7 | 93885396 | 93890753 | 1 | 260.28 | 1.62 | 0.74 | 3.56 | 0.226 | 0.955 |
| TFRC | OID20369 | Cardiome | TFRC | P02786 | 3 | 196027183 | 196082096 | 1 | 4290.25 | 1.10 | 0.91 | 1.31 | 0.324 | 0.955 |
| TG | OID31359 | Oncology | TG | P01266 | 8 | 132866958 | 133134903 | 1 | 125.03 | 1.76 | 0.58 | 5.33 | 0.321 | 0.955 |
| TGFB2 | OID31417 | Oncology | TGFB2 | P61812 | 1 | 218345336 | 218444619 | 1 | 269.51 | 1.80 | 0.88 | 3.70 | 0.107 | 0.955 |
| TGFBI | OID20417 | Cardiome | TGFBI | Q15582 | 5 | 136028988 | 136063818 | 1 | 6084.00 | 1.12 | 0.95 | 1.34 | 0.186 | 0.955 |
| THBS2 | OID21104 | Neurology | THBS2 | P35442 | 6 | 169215780 | 169254050 | 1 | 2585.33 | 1.15 | 0.91 | 1.44 | 0.248 | 0.955 |
| THBS4 | OID20368 | Cardiome | THBS4 | P35443 | 5 | 79991311 | 80083287 | 1 | 930.25 | 1.30 | 0.88 | 1.93 | 0.195 | 0.955 |
| THPO | OID20098 | Cardiome | THPO | P40225 | 3 | 184371935 | 184381968 | 1 | 60.06 | 2.96 | 0.64 | 13.69 | 0.164 | 0.955 |
| THTPA | OID31375 | Oncology | THTPA | Q9BU02 | 14 | 23555988 | 23560271 | 1 | 3328.40 | 1.17 | 0.93 | 1.47 | 0.190 | 0.955 |
| TIE1 | OID20303 | Cardiome | TIE1 | P35590 | 1 | 43300982 | 43323108 | 1 | 744.51 | 0.78 | 0.47 | 1.29 | 0.329 | 0.955 |
| TK1 | OID31294 | Oncology | TK1 | P04183 | 17 | 78174091 | 78187233 | 1 | 100.00 | 2.27 | 0.57 | 9.07 | 0.246 | 0.955 |
| TLR4 | OID30428 | Inflammat | TLR4 | O00206 | 9 | 117704175 | 117724735 | 1 | 417.69 | 0.74 | 0.41 | 1.32 | 0.304 | 0.955 |
| TMED8 | OID31462 | Oncology | TMED8 | Q6PL24 | 14 | 77335029 | 77377094 | 1 | 68.06 | 0.43 | 0.10 | 1.89 | 0.266 | 0.955 |
| TMEM25 | OID31015 | Neurology | TMEM25 | Q86YD3 | 11 | 118531041 | 118547280 | 1 | 2359.18 | 0.88 | 0.69 | 1.13 | 0.314 | 0.955 |
| TNC | OID20359 | Cardiome | TNC | P24821 | 9 | 115019575 | 115118257 | 1 | 930.25 | 1.33 | 0.87 | 2.05 | 0.192 | 0.955 |
| TNFAIP8 | OID20433 | Inflammat | TNFAIP8 | O95379 | 5 | 119268692 | 119399688 | 1 | 105.06 | 2.17 | 0.62 | 7.62 | 0.226 | 0.955 |
| TNFRSF10A | OID20967 | Neurology | TNFRSF1 | O00220 | 8 | 23190452 | 23225102 | 1 | 4356.00 | 0.90 | 0.74 | 1.11 | 0.338 | 0.955 |
| TNFRSF13C | OID20480 | Inflammat | TNFRSF1 | Q96RJ3 | 22 | 41922032 | 41926806 | 1 | 3721.00 | 1.13 | 0.90 | 1.40 | 0.287 | 0.955 |
| TNFRSF1B | OID21145 | Neurology | TNFRSF1 | P20333 | 1 | 12166991 | 12209228 | 1 | 425.66 | 1.75 | 0.90 | 3.40 | 0.101 | 0.955 |
| TNFRSF4 | OID20653 | Inflammat | TNFRSF4 | P43489 | 1 | 1211340 | 1214153 | 1 | 298.52 | 1.50 | 0.68 | 3.31 | 0.314 | 0.955 |
| TNFRSF8 | OID21049 | Neurology | TNFRSF8 | P28908 | 1 | 12063303 | 12144207 | 1 | 716.59 | 1.25 | 0.80 | 1.96 | 0.321 | 0.955 |
| TNFSF13 | OID20733 | Inflammat | TNFSF13 | O75888 | 17 | 7558292 | 7561608 | 1 | 1304.01 | 1.25 | 0.88 | 1.78 | 0.204 | 0.955 |
| TNFSF14 | OID20953 | Neurology | TNFSF14 | O43557 | 19 | 6661253 | 6670588 | 1 | 1061.99 | 0.81 | 0.57 | 1.16 | 0.258 | 0.955 |
| TNFSF8 | OID30384 | Cardiome | TNFSF8 | P32971 | 9 | 114893343 | 114930595 | 1 | 870.25 | 0.71 | 0.47 | 1.06 | 0.089 | 0.955 |
| TNXB | OID21107 | Neurology | TNXB | P22105 | 6 | 32041153 | 32115334 | 1 | 2870.89 | 1.21 | 0.98 | 1.49 | 0.074 | 0.955 |
| TOP2B | OID30559 | Inflammat | TOP2B | Q02880 | 3 | 25597978 | 25664909 | 1 | 118.57 | 2.32 | 0.71 | 7.62 | 0.165 | 0.955 |
| TOR1AIP1 | OID30195 | Cardiome | TOR1AIP | Q5JTV8 | 1 | 179882042 | 179920077 | 1 | 7854.39 | 1.07 | 0.93 | 1.23 | 0.348 | 0.955 |
| TP53 | OID21232 | Oncology | TP53 | P04637 | 17 | 7661779 | 7687538 | 1 | 53.78 | 2.55 | 0.51 | 12.87 | 0.257 | 0.955 |
| TP53I3 | OID30632 | Inflammat | TP53I3 | Q53FA7 | 2 | 24077433 | 24085861 | 1 | 3552.16 | 1.11 | 0.89 | 1.39 | 0.345 | 0.955 |
| TPMT | OID21270 | Oncology | TPMT | P51580 | 6 | 18128311 | 18155077 | 1 | 4148.88 | 0.91 | 0.75 | 1.09 | 0.310 | 0.955 |
| TPPP3 | OID20842 | Neurology | TPPP3 | Q9BW30 | 16 | 67389809 | 67393518 | 1 | 60.84 | 2.96 | 0.60 | 14.75 | 0.184 | 0.955 |
| TRAF2 | OID20507 | Inflammat | TRAF2 | Q12933 | 9 | 136881912 | 136926607 | 1 | 66.31 | 2.82 | 0.52 | 15.19 | 0.228 | 0.955 |
| TRDMT1 | OID31335 | Oncology | TRDMT1 | O14717 | 10 | 17137336 | 17202054 | 1 | 337.64 | 0.60 | 0.32 | 1.15 | 0.123 | 0.955 |
| TREH | OID30388 | Cardiome | TREH | O43280 | 11 | 118657316 | 118679690 | 1 | 12210.25 | 0.92 | 0.82 | 1.03 | 0.145 | 0.955 |
| TRIM25 | OID31482 | Oncology | TRIM25 | Q14258 | 17 | 56836387 | 56914080 | 1 | 156.25 | 2.19 | 0.76 | 6.29 | 0.146 | 0.955 |
| TRIM40 | OID30885 | Neurology | TRIM40 | Q6P9F5 | 6 | 30136124 | 30148735 | 1 | 933.64 | 1.24 | 0.82 | 1.87 | 0.304 | 0.955 |
| TRIM58 | OID31429 | Oncology | TRIM58 | Q8NG06 | 1 | 247857187 | 247880138 | 1 | 630.57 | 1.35 | 0.82 | 2.24 | 0.240 | 0.955 |
| TXNDC15 | OID21495 | Oncology | TXNDC1 | Q96J42 | 5 | 134874371 | 134901635 | 1 | 11313.22 | 0.91 | 0.80 | 1.02 | 0.116 | 0.955 |
| ULBP2 | OID20919 | Neurology | ULBP2 | Q9BZM5 | 6 | 149942014 | 149949235 | 1 | 3776.66 | 0.89 | 0.73 | 1.09 | 0.268 | 0.955 |
| UMOD | OID20237 | Cardiome | UMOD | P07911 | 16 | 20333051 | 20356301 | 1 | 11592.11 | 1.10 | 0.97 | 1.25 | 0.153 | 0.955 |
| UROD | OID30603 | Inflammat | UROD | P06132 | 1 | 45010950 | 45015575 | 1 | 3555.14 | 0.88 | 0.70 | 1.10 | 0.246 | 0.955 |
| UXS1 | OID21189 | Oncology | UXS1 | Q8NBZ7 | 2 | 106093308 | 106194301 | 1 | 156.25 | 1.87 | 0.71 | 4.94 | 0.204 | 0.955 |
| VAMP8 | OID30555 | Inflammat | VAMP8 | Q9BV40 | 2 | 85561562 | 85582031 | 1 | 467.64 | 1.48 | 0.80 | 2.74 | 0.206 | 0.955 |
| VAT1 | OID21382 | Oncology | VAT1 | Q99536 | 17 | 43014607 | 43025123 | 1 | 310.64 | 1.58 | 0.77 | 3.23 | 0.212 | 0.955 |
| VCAN | OID21026 | Neurology | VCAN | P13611 | 5 | 83471618 | 83582303 | 1 | 8595.94 | 0.92 | 0.80 | 1.07 | 0.284 | 0.955 |
| VMO1 | OID21381 | Oncology | VMO1 | Q7Z5L0 | 17 | 4785285 | 4786433 | 1 | 8906.64 | 1.10 | 0.97 | 1.25 | 0.125 | 0.955 |
| VSNL1 | OID31105 | Neurology | VSNL1 | P62760 | 2 | 17539126 | 17657018 | 1 | 1530.77 | 0.80 | 0.57 | 1.12 | 0.191 | 0.955 |
| VSTM1 | OID20851 | Neurology | VSTM1 | Q6UX27 | 19 | 54040825 | 54063953 | 1 | 9216.00 | 0.91 | 0.81 | 1.03 | 0.154 | 0.955 |
| VTI1A | OID30640 | Inflammat | VTI1A | Q96AJ9 | 10 | 112446998 | 112818744 | 1 | 51.36 | 0.33 | 0.07 | 1.70 | 0.186 | 0.955 |
| VWC2L | OID31002 | Neurology | VWC2L | B2RUY7 | 2 | 214411054 | 214578976 | 1 | 121.00 | 0.57 | 0.18 | 1.75 | 0.324 | 0.955 |
| WIF1 | OID21464 | Oncology | WIF1 | Q9Y5W5 | 12 | 65050626 | 65121305 | 1 | 752.33 | 0.75 | 0.46 | 1.23 | 0.254 | 0.955 |
| XCL1 | OID21398 | Oncology | XCL1 | P47992 | 1 | 168576605 | 168582069 | 1 | 6938.89 | 0.92 | 0.80 | 1.07 | 0.278 | 0.955 |
| YAP1 | OID31113 | Neurology | YAP1 | P46937 | 11 | 102110447 | 102233424 | 1 | 78.45 | 2.51 | 0.54 | 11.58 | 0.238 | 0.955 |
| ZBTB16 | OID21205 | Oncology | ZBTB16 | Q05516 | 11 | 114059041 | 114256765 | 1 | 1814.76 | 1.16 | 0.88 | 1.53 | 0.281 | 0.955 |
| ZP3 | OID30265 | Cardiome | ZP3 | P21754 | 7 | 76397518 | 76442071 | 1 | 19078.52 | 0.95 | 0.87 | 1.03 | 0.231 | 0.955 |
| DAPK2 | OID30505 | Inflammat | DAPK2 | Q9UIK4 | 15 | 63907036 | 64072033 | 1 | 2340.14 | 0.89 | 0.69 | 1.14 | 0.350 | 0.956 |
| OCLN | OID31069 | Neurology | OCLN | Q16625 | 5 | 69492292 | 69558104 | 1 | 388.42 | 1.39 | 0.69 | 2.79 | 0.352 | 0.958 |
| ABHD14B | OID20921 | Neurology | ABHD14B | Q96IU4 | 3 | 51968510 | 51983409 | 1 | 1622.30 | 0.88 | 0.67 | 1.16 | 0.364 | 0.961 |
| ACAN | OID20159 | Cardiome | ACAN | P16112 | 15 | 88803436 | 88875353 | 1 | 221.68 | 0.67 | 0.27 | 1.66 | 0.387 | 0.961 |
| ANXA11 | OID20583 | Inflammat | ANXA11 | P50995 | 10 | 80150889 | 80205572 | 1 | 68.06 | 0.51 | 0.11 | 2.27 | 0.375 | 0.961 |
| ASRGL1 | OID30283 | Cardiome | ASRGL1 | Q7L266 | 11 | 62337448 | 62393412 | 1 | 2162.25 | 0.88 | 0.67 | 1.17 | 0.380 | 0.961 |

| | | | | | | | | | | | | | | |
|---|---|---|---|---|---|---|---|---|---|---|---|---|---|---|
| ATP6V1G2 | OID30167 | Cardiome | ATP6V1G | O95670 | 6 | 31544444 | 31548427 | 1 | 343.93 | 1.32 | 0.72 | 2.44 | 0.370 | 0.961 |
| BCL2L15 | OID30498 | Inflammat | BCL2L15 | Q5TBC7 | 1 | 113876816 | 113887581 | 1 | 489.52 | 1.29 | 0.73 | 2.29 | 0.387 | 0.961 |
| BNIP3L | OID30525 | Inflammat | BNIP3L | O60238 | 8 | 26383054 | 26505636 | 1 | 45.56 | 0.43 | 0.07 | 2.72 | 0.371 | 0.961 |
| C1RL | OID30721 | Inflammat | C1RL | Q9NZP8 | 12 | 7089587 | 7109238 | 1 | 4006.15 | 1.11 | 0.88 | 1.39 | 0.371 | 0.961 |
| CA13 | OID20225 | Cardiome | CA13 | Q8N1Q1 | 8 | 85220587 | 85284073 | 1 | 1229.67 | 0.86 | 0.60 | 1.22 | 0.384 | 0.961 |
| CA3 | OID20271 | Cardiome | CA3 | P07451 | 8 | 85373436 | 85449040 | 1 | 473.06 | 0.76 | 0.40 | 1.42 | 0.387 | 0.961 |
| CA6 | OID21096 | Neurology | CA6 | P23280 | 1 | 8945867 | 8975092 | 1 | 9520.18 | 0.94 | 0.81 | 1.08 | 0.384 | 0.961 |
| CALCB | OID31100 | Neurology | CALCB | P10092 | 11 | 14904997 | 15082342 | 1 | 199.12 | 1.46 | 0.62 | 3.45 | 0.382 | 0.961 |
| CAT | OID30712 | Inflammat | CAT | P04040 | 11 | 34438934 | 34472060 | 1 | 417.98 | 0.76 | 0.41 | 1.40 | 0.376 | 0.961 |
| CBLIF | OID20179 | Cardiome | CBLIF | P27352 | 11 | 59829273 | 59845499 | 1 | 987.76 | 0.82 | 0.52 | 1.28 | 0.380 | 0.961 |
| CCN1 | OID21368 | Oncology | CCN1 | O00622 | 1 | 85580761 | 85584589 | 1 | 217.56 | 1.53 | 0.60 | 3.88 | 0.370 | 0.961 |
| CD8A | OID21018 | Neurology | CD8A | P01732 | 2 | 86784610 | 86808396 | 1 | 1310.44 | 1.17 | 0.83 | 1.65 | 0.378 | 0.961 |
| CDH17 | OID20134 | Cardiome | CDH17 | Q12864 | 8 | 94127162 | 94217303 | 1 | 1532.16 | 1.17 | 0.83 | 1.66 | 0.367 | 0.961 |
| CEP43 | OID20084 | Cardiome | CEP43 | O95684 | 6 | 166999317 | 167094789 | 1 | 66.02 | 1.98 | 0.46 | 8.43 | 0.356 | 0.961 |
| CEP85 | OID21241 | Oncology | CEP85 | Q6P2H3 | 1 | 26234200 | 26278808 | 1 | 103.10 | 0.57 | 0.16 | 2.01 | 0.384 | 0.961 |
| CHAC2 | OID21230 | Oncology | CHAC2 | Q8WUX2 | 2 | 53767804 | 53775196 | 1 | 274.09 | 0.72 | 0.36 | 1.46 | 0.365 | 0.961 |
| CHGB | OID21038 | Neurology | CHGB | P05060 | 20 | 5911510 | 5925353 | 1 | 5833.14 | 0.93 | 0.80 | 1.09 | 0.386 | 0.961 |
| CLEC1B | OID21061 | Neurology | CLEC1B | Q9P126 | 12 | 9985642 | 10013424 | 1 | 344.31 | 0.73 | 0.38 | 1.42 | 0.357 | 0.961 |
| CNTN3 | OID21092 | Neurology | CNTN3 | Q9P232 | 3 | 74262568 | 74614659 | 1 | 1337.47 | 1.17 | 0.83 | 1.65 | 0.379 | 0.961 |
| CSPG4 | OID30397 | Cardiome | CSPG4 | Q6UVK1 | 15 | 75674322 | 75712848 | 1 | 280.56 | 0.69 | 0.31 | 1.54 | 0.365 | 0.961 |
| CTSO | OID20660 | Inflammat | CTSO | P43234 | 4 | 155921580 | 155953912 | 1 | 586.72 | 0.78 | 0.46 | 1.33 | 0.367 | 0.961 |
| DCBLD2 | OID21474 | Oncology | DCBLD2 | Q96PD2 | 3 | 98795941 | 98901695 | 1 | 1162.38 | 0.84 | 0.58 | 1.22 | 0.366 | 0.961 |
| EFEMP1 | OID20414 | Cardiome | EFEMP1 | Q12805 | 2 | 55865967 | 55924139 | 1 | 798.06 | 1.25 | 0.76 | 2.03 | 0.378 | 0.961 |
| EIF2AK2 | OID31442 | Oncology | EIF2AK2 | P19525 | 2 | 37099210 | 37157522 | 1 | 351.56 | 1.32 | 0.70 | 2.48 | 0.387 | 0.961 |
| F11 | OID30773 | Inflammat | F11 | P03951 | 4 | 186266189 | 186289681 | 1 | 3025.00 | 1.12 | 0.87 | 1.43 | 0.379 | 0.961 |
| FGF5 | OID20490 | Inflammat | FGF5 | P12034 | 4 | 80266639 | 80336680 | 1 | 7267.56 | 1.07 | 0.92 | 1.24 | 0.382 | 0.961 |
| FKBP5 | OID20937 | Neurology | FKBP5 | Q13451 | 6 | 35573585 | 35728583 | 1 | 682.52 | 1.25 | 0.77 | 2.04 | 0.371 | 0.961 |
| GART | OID31004 | Neurology | GART | P22102 | 21 | 33503931 | 33543491 | 1 | 205.44 | 1.46 | 0.62 | 3.43 | 0.386 | 0.961 |
| GASK1A | OID30109 | Cardiome | GASK1A | Q9UFP1 | 3 | 42979087 | 43067898 | 1 | 8326.56 | 1.07 | 0.93 | 1.22 | 0.357 | 0.961 |
| GBP2 | OID20489 | Inflammat | GBP2 | P32456 | 1 | 89106132 | 89150456 | 1 | 189.83 | 0.65 | 0.26 | 1.63 | 0.354 | 0.961 |
| HPGDS | OID21266 | Oncology | HPGDS | O60760 | 4 | 94298535 | 94342876 | 1 | 3349.52 | 1.10 | 0.89 | 1.36 | 0.355 | 0.961 |
| ICAM1 | OID20411 | Cardiome | ICAM1 | P05362 | 19 | 10271093 | 10286615 | 1 | 2691.02 | 0.90 | 0.71 | 1.14 | 0.372 | 0.961 |
| ICAM4 | OID20578 | Inflammat | ICAM4 | Q14773 | 19 | 10286955 | 10288522 | 1 | 269.90 | 1.47 | 0.63 | 3.43 | 0.372 | 0.961 |
| IDI2 | OID20823 | Neurology | IDI2 | Q9BXS1 | 10 | 1018910 | 1025859 | 1 | 1936.00 | 0.88 | 0.67 | 1.16 | 0.368 | 0.961 |
| IL1RN | OID20700 | Inflammat | IL1RN | P18510 | 2 | 113107214 | 113134016 | 1 | 708.89 | 1.22 | 0.78 | 1.91 | 0.373 | 0.961 |
| KIR3DL2 | OID31191 | Oncology | KIR3DL2 | P43630 | 19 | 54850443 | 54867207 | 1 | 240.25 | 0.69 | 0.30 | 1.60 | 0.386 | 0.961 |
| KLB | OID20888 | Neurology | KLB | Q86Z14 | 4 | 39406930 | 39451533 | 1 | 9072.56 | 1.06 | 0.93 | 1.21 | 0.388 | 0.961 |
| KLK1 | OID21499 | Oncology | KLK1 | P06870 | 19 | 50819146 | 50823787 | 1 | 12348.77 | 0.95 | 0.86 | 1.06 | 0.358 | 0.961 |
| KLK10 | OID21523 | Oncology | KLK10 | O43240 | 19 | 51012739 | 51020175 | 1 | 5494.52 | 0.93 | 0.79 | 1.10 | 0.382 | 0.961 |
| LY96 | OID20945 | Neurology | LY96 | Q9Y6Y9 | 8 | 73991392 | 74029079 | 1 | 125.94 | 1.61 | 0.56 | 4.65 | 0.377 | 0.961 |
| MMP10 | OID20687 | Inflammat | MMP10 | P09238 | 11 | 102770502 | 102780628 | 1 | 1568.16 | 1.16 | 0.84 | 1.61 | 0.373 | 0.961 |
| NELL1 | OID21494 | Oncology | NELL1 | Q92832 | 11 | 20669551 | 21575686 | 1 | 2181.44 | 0.88 | 0.66 | 1.17 | 0.369 | 0.961 |
| OLR1 | OID20183 | Cardiome | OLR1 | P78380 | 12 | 10158301 | 10172138 | 1 | 144.00 | 0.63 | 0.23 | 1.75 | 0.378 | 0.961 |
| PBLD | OID20839 | Neurology | PBLD | P30039 | 10 | 68282660 | 68333049 | 1 | 4096.00 | 0.92 | 0.76 | 1.11 | 0.380 | 0.961 |
| PTPRS | OID20306 | Cardiome | PTPRS | Q13332 | 19 | 5158495 | 5340812 | 1 | 1340.37 | 0.84 | 0.58 | 1.22 | 0.364 | 0.961 |
| RAB2B | OID31362 | Oncology | RAB2B | Q8WUD1 | 14 | 21459020 | 21476960 | 1 | 448.67 | 0.76 | 0.43 | 1.36 | 0.357 | 0.961 |
| RALB | OID30567 | Inflammat | RALB | P11234 | 2 | 120240064 | 120294710 | 1 | 213.89 | 0.67 | 0.28 | 1.63 | 0.381 | 0.961 |
| REEP4 | OID31028 | Neurology | REEP4 | Q9H6H4 | 8 | 22138020 | 22141951 | 1 | 454.60 | 1.34 | 0.72 | 2.52 | 0.359 | 0.961 |
| S100A14 | OID30987 | Neurology | S100A14 | Q9HCY8 | 1 | 153614255 | 153616986 | 1 | 625.00 | 1.28 | 0.74 | 2.21 | 0.369 | 0.961 |
| SAT2 | OID31306 | Oncology | SAT2 | Q96F10 | 17 | 7626234 | 7627876 | 1 | 4569.76 | 0.92 | 0.78 | 1.10 | 0.369 | 0.961 |
| SERPINA5 | OID30763 | Inflammat | SERPINA | P05154 | 14 | 94561442 | 94593118 | 1 | 631.27 | 0.81 | 0.50 | 1.30 | 0.379 | 0.961 |
| SERPINI2 | OID31504 | Oncology | SERPINI2 | O75830 | 3 | 167441914 | 167478972 | 1 | 2590.81 | 1.13 | 0.87 | 1.49 | 0.359 | 0.961 |
| SEZ6 | OID30968 | Neurology | SEZ6 | Q53EL9 | 17 | 28954901 | 29006440 | 1 | 64.00 | 0.52 | 0.12 | 2.25 | 0.379 | 0.961 |
| SOD1 | OID20386 | Cardiome | SOD1 | P00441 | 21 | 31659666 | 31668931 | 1 | 66.69 | 2.06 | 0.43 | 9.83 | 0.363 | 0.961 |
| STX7 | OID30560 | Inflammat | STX7 | O15400 | 6 | 132445867 | 132513198 | 1 | 5439.06 | 1.08 | 0.91 | 1.27 | 0.378 | 0.961 |
| TGM2 | OID20141 | Cardiome | TGM2 | P21980 | 20 | 38127385 | 38166578 | 1 | 582.88 | 0.76 | 0.43 | 1.35 | 0.355 | 0.961 |
| VASH1 | OID20516 | Inflammat | VASH1 | Q7L8A9 | 14 | 76761468 | 76783015 | 1 | 172.27 | 0.63 | 0.23 | 1.71 | 0.362 | 0.961 |
| VWF | OID20256 | Cardiome | VWF | P04275 | 12 | 5948877 | 6124770 | 1 | 150.06 | 1.59 | 0.58 | 4.36 | 0.368 | 0.961 |
| WFDC1 | OID31366 | Oncology | WFDC1 | Q9HC57 | 16 | 84294846 | 84329844 | 1 | 1274.49 | 1.17 | 0.82 | 1.66 | 0.388 | 0.961 |
| MTIF3 | OID31513 | Oncology | MTIF3 | Q9H2K0 | 13 | 27435643 | 27450591 | 1 | 131.65 | 0.64 | 0.23 | 1.76 | 0.390 | 0.964 |
| HADH | OID30132 | Cardiome | HADH | Q16836 | 4 | 107989714 | 108035241 | 1 | 57.33 | 1.98 | 0.42 | 9.36 | 0.391 | 0.966 |
| SF3B4 | OID21278 | Oncology | SF3B4 | Q15427 | 1 | 149923317 | 149927803 | 1 | 48.01 | 2.23 | 0.36 | 13.98 | 0.392 | 0.966 |
| ADAM12 | OID30491 | Inflammat | ADAM12 | O43184 | 10 | 126012381 | 126388477 | 1 | 260.02 | 0.73 | 0.34 | 1.54 | 0.403 | 0.969 |
| ALCAM | OID20273 | Cardiome | ALCAM | Q13740 | 3 | 105366909 | 105576900 | 1 | 358.63 | 1.39 | 0.65 | 2.98 | 0.399 | 0.969 |
| BANK1 | OID20594 | Inflammat | BANK1 | Q8NDB2 | 4 | 101411286 | 102074812 | 1 | 253.87 | 0.72 | 0.33 | 1.55 | 0.400 | 0.969 |
| CCL19 | OID21030 | Neurology | CCL19 | Q99731 | 9 | 34689570 | 34691276 | 1 | 91.12 | 1.81 | 0.46 | 7.06 | 0.395 | 0.969 |
| CLEC11A | OID21112 | Neurology | CLEC11A | Q9Y240 | 19 | 50723364 | 50725708 | 1 | 1271.08 | 0.85 | 0.59 | 1.23 | 0.396 | 0.969 |
| COL5A1 | OID30587 | Inflammat | COL5A1 | P20908 | 9 | 134641803 | 134844843 | 1 | 687.60 | 0.82 | 0.52 | 1.30 | 0.404 | 0.969 |
| CPOX | OID30495 | Inflammat | CPOX | P36551 | 3 | 98579446 | 98593648 | 1 | 203.71 | 0.69 | 0.29 | 1.65 | 0.404 | 0.969 |
| CSF2RB | OID30553 | Inflammat | CSF2RB | P32927 | 22 | 36913628 | 36940439 | 1 | 19670.06 | 1.04 | 0.95 | 1.13 | 0.401 | 0.969 |
| CTBS | OID30710 | Inflammat | CTBS | Q01459 | 1 | 84549611 | 84574480 | 1 | 4305.86 | 1.08 | 0.90 | 1.30 | 0.408 | 0.969 |
| CTRL | OID31157 | Neurology | CTRL | P40313 | 16 | 67927640 | 67932365 | 1 | 79.21 | 0.54 | 0.13 | 2.29 | 0.406 | 0.969 |
| CTSL | OID20228 | Cardiome | CTSL | P07711 | 9 | 87724051 | 87731469 | 1 | 447.02 | 1.31 | 0.69 | 2.46 | 0.407 | 0.969 |
| DNM1 | OID31458 | Oncology | DNM1 | Q05193 | 9 | 128191655 | 128255248 | 1 | 1272.11 | 0.86 | 0.60 | 1.23 | 0.398 | 0.969 |
| EZR | OID21017 | Neurology | EZR | P15311 | 6 | 158765741 | 158819368 | 1 | 147.02 | 0.63 | 0.22 | 1.82 | 0.395 | 0.969 |
| GBP4 | OID20845 | Neurology | GBP4 | Q96PP9 | 1 | 89181144 | 89198942 | 1 | 10125.39 | 0.95 | 0.85 | 1.07 | 0.400 | 0.969 |
| GCHFR | OID30631 | Inflammat | GCHFR | P30047 | 15 | 40764068 | 40767708 | 1 | 251.45 | 0.69 | 0.30 | 1.62 | 0.400 | 0.969 |
| HSPB1 | OID20230 | Cardiome | HSPB1 | P04792 | 7 | 76302673 | 76304295 | 1 | 3775.42 | 0.92 | 0.75 | 1.12 | 0.402 | 0.969 |
| HSPG2 | OID20383 | Cardiome | HSPG2 | P98160 | 1 | 21822244 | 21937310 | 1 | 478.52 | 1.29 | 0.71 | 2.35 | 0.395 | 0.969 |
| IL34 | OID20809 | Neurology | IL34 | Q6ZMJ4 | 16 | 70579895 | 70660682 | 1 | 7950.69 | 1.06 | 0.92 | 1.22 | 0.407 | 0.969 |
| KIAA0319 | OID31094 | Neurology | KIAA0319 | Q5VV43 | 6 | 24544104 | 24646191 | 1 | 1482.25 | 0.87 | 0.63 | 1.20 | 0.403 | 0.969 |
| NCAM2 | OID21095 | Neurology | NCAM2 | O15394 | 21 | 20998409 | 21543329 | 1 | 4624.00 | 1.08 | 0.90 | 1.30 | 0.405 | 0.969 |
| PNLIPRP1 | OID31122 | Neurology | PNLIPRP | P54315 | 10 | 116590385 | 116609175 | 1 | 1098.45 | 1.19 | 0.79 | 1.80 | 0.403 | 0.969 |
| RNASEH2A | OID31027 | Neurology | RNASEH2 | O75792 | 19 | 12806584 | 12813640 | 1 | 185.48 | 0.68 | 0.28 | 1.67 | 0.402 | 0.969 |
| SERPINA11 | OID20281 | Cardiome | SERPINA | Q86U17 | 14 | 94442464 | 94452800 | 1 | 7870.22 | 1.07 | 0.91 | 1.25 | 0.406 | 0.969 |
| SNAP29 | OID21506 | Oncology | SNAP29 | O95721 | 22 | 20859007 | 20891214 | 1 | 86.22 | 0.58 | 0.16 | 2.06 | 0.398 | 0.969 |
| SORCS2 | OID21262 | Oncology | SORCS2 | Q96PQ0 | 4 | 7192538 | 7742836 | 1 | 3263.27 | 0.91 | 0.74 | 1.13 | 0.395 | 0.969 |
| TGFA | OID20600 | Inflammat | TGFA | P01135 | 2 | 70447284 | 70554193 | 1 | 212.67 | 1.42 | 0.63 | 3.20 | 0.404 | 0.969 |
| USP28 | OID31084 | Neurology | USP28 | Q96RU2 | 11 | 113797874 | 113875572 | 1 | 119.43 | 0.62 | 0.20 | 1.89 | 0.396 | 0.969 |
| VNN2 | OID21310 | Oncology | VNN2 | O95498 | 6 | 132743870 | 132763459 | 1 | 7590.77 | 0.94 | 0.82 | 1.09 | 0.409 | 0.971 |
| MASP1 | OID20954 | Neurology | MASP1 | P48740 | 3 | 187217282 | 187291980 | 1 | 708.89 | 0.82 | 0.52 | 1.31 | 0.411 | 0.973 |
| DSG3 | OID21460 | Oncology | DSG3 | P32926 | 18 | 31447741 | 31478702 | 1 | 1171.16 | 1.19 | 0.79 | 1.78 | 0.412 | 0.974 |
| FRZB | OID20968 | Neurology | FRZB | Q92765 | 2 | 182833275 | 182866637 | 1 | 3405.56 | 0.92 | 0.74 | 1.13 | 0.412 | 0.974 |
| ATXN10 | OID20865 | Neurology | ATXN10 | Q9UBB4 | 22 | 45671798 | 45845307 | 1 | 175.56 | 1.49 | 0.56 | 3.96 | 0.422 | 0.975 |
| BTN1A1 | OID31363 | Oncology | BTN1A1 | Q13410 | 6 | 26500303 | 26510425 | 1 | 625.00 | 1.23 | 0.74 | 2.03 | 0.427 | 0.975 |
| CD27 | OID21527 | Oncology | CD27 | P26842 | 12 | 6444955 | 6451718 | 1 | 1067.11 | 1.16 | 0.80 | 1.69 | 0.422 | 0.975 |
| CR1 | OID30697 | Inflammat | CR1 | P17927 | 1 | 207496147 | 207641765 | 1 | 2916.00 | 0.91 | 0.72 | 1.15 | 0.425 | 0.975 |
| CXCL14 | OID20458 | Inflammat | CXCL14 | O95715 | 5 | 135570679 | 135579279 | 1 | 105.80 | 1.74 | 0.44 | 6.79 | 0.427 | 0.975 |
| DEFB104A_D | OID30849 | Neurology | DEFB104 | Q8WTQ1 | 8;8 | 7470308;783 | 7475082;784 | 1 | 362.81 | 1.31 | 0.68 | 2.52 | 0.416 | 0.975 |
| DOK2 | OID20138 | Cardiome | DOK2 | O60496 | 8 | 21908873 | 21913690 | 1 | 60.06 | 1.88 | 0.41 | 8.63 | 0.417 | 0.975 |
| DYNLT1 | OID30925 | Neurology | DYNLT1 | P63172 | 6 | 158636474 | 158644743 | 1 | 133.90 | 0.62 | 0.19 | 2.01 | 0.425 | 0.975 |
| EFNA1 | OID21125 | Neurology | EFNA1 | P20827 | 1 | 155127876 | 155134899 | 1 | 1570.14 | 1.13 | 0.84 | 1.53 | 0.413 | 0.975 |
| EPHX2 | OID20119 | Cardiome | EPHX2 | P34913 | 8 | 27490781 | 27545564 | 1 | 233.91 | 1.36 | 0.64 | 2.90 | 0.419 | 0.975 |
| FAM171B | OID31338 | Oncology | FAM171B | Q6P995 | 2 | 186694060 | 186765959 | 1 | 2220.77 | 1.12 | 0.85 | 1.49 | 0.420 | 0.975 |
| FXN | OID21442 | Oncology | FXN | Q16595 | 9 | 69035751 | 69079076 | 1 | 97.52 | 1.65 | 0.48 | 5.61 | 0.425 | 0.975 |
| GNLY | OID21132 | Neurology | GNLY | P22749 | 2 | 85685175 | 85698852 | 1 | 6188.44 | 0.94 | 0.80 | 1.10 | 0.425 | 0.975 |
| NCF2 | OID20567 | Inflammat | NCF2 | P19878 | 1 | 183555562 | 183590876 | 1 | 693.44 | 1.27 | 0.71 | 2.25 | 0.422 | 0.975 |

| | | | | | | | | | | | | | | |
|---|---|---|---|---|---|---|---|---|---|---|---|---|---|---|
| ODAM | OID20793 | Neurology | ODAM | A1E959 | 4 | 70195725 | 70204576 | 1 | 1192.28 | 0.86 | 0.61 | 1.23 | 0.416 | 0.975 |
| PKD1 | OID30328 | Cardiome | PKD1 | P98161 | 16 | 2088708 | 2135898 | 1 | 1012.24 | 0.85 | 0.56 | 1.27 | 0.418 | 0.975 |
| PRAP1 | OID30391 | Cardiome | PRAP1 | Q96NZ9 | 10 | 133347368 | 133352683 | 1 | 2148.98 | 1.13 | 0.83 | 1.54 | 0.423 | 0.975 |
| PRR4 | OID30607 | Inflammat | PRR4 | Q16378 | 12 | 10845849 | 10849475 | 1 | 9579.52 | 1.05 | 0.93 | 1.19 | 0.426 | 0.975 |
| RLN2 | OID30601 | Inflammat | RLN2 | P04090 | 9 | 5299864 | 5304716 | 1 | 308.20 | 1.34 | 0.66 | 2.71 | 0.418 | 0.975 |
| SESTD1 | OID20790 | Neurology | SESTD1 | Q86VW0 | 2 | 179101678 | 179264832 | 1 | 169.00 | 1.49 | 0.57 | 3.89 | 0.419 | 0.975 |
| SPINK1 | OID21097 | Neurology | SPINK1 | P00995 | 5 | 147824572 | 147831671 | 1 | 2562.89 | 0.91 | 0.71 | 1.15 | 0.427 | 0.975 |
| TFF1 | OID21154 | Neurology | TFF1 | P04155 | 21 | 42362282 | 42366535 | 1 | 1140.94 | 1.16 | 0.81 | 1.68 | 0.421 | 0.975 |
| TIMP3 | OID20684 | Inflammat | TIMP3 | P35625 | 22 | 32801705 | 32863041 | 1 | 6642.25 | 0.94 | 0.79 | 1.10 | 0.422 | 0.975 |
| TNFRSF1A | OID21155 | Neurology | TNFRSF1 | P19438 | 12 | 6328757 | 6342114 | 1 | 117.10 | 1.73 | 0.45 | 6.58 | 0.423 | 0.975 |
| TSC22D1 | OID31016 | Neurology | TSC22D1 | Q15714 | 13 | 44432143 | 44577147 | 1 | 58.78 | 0.50 | 0.09 | 2.75 | 0.427 | 0.975 |
| VAMP5 | OID20071 | Cardiome | VAMP5 | O95183 | 2 | 85584431 | 85593406 | 1 | 110.25 | 0.60 | 0.18 | 2.09 | 0.425 | 0.975 |
| CCDC134 | OID31413 | Oncology | CCDC134 | Q9H6E4 | 22 | 41800679 | 41832164 | 1 | 56.25 | 1.95 | 0.37 | 10.30 | 0.430 | 0.975 |
| CCL15 | OID20328 | Cardiome | CCL15 | Q16663 | 17 | 35996440 | 36001553 | 1 | 12720.62 | 0.95 | 0.85 | 1.07 | 0.428 | 0.975 |
| CLEC4G | OID20590 | Inflammat | CLEC4G | Q6UXB4 | 19 | 7728957 | 7733906 | 1 | 713.65 | 1.20 | 0.76 | 1.92 | 0.435 | 0.975 |
| ELOA | OID21337 | Oncology | ELOA | Q14241 | 1 | 23743448 | 23762059 | 1 | 115.56 | 1.57 | 0.51 | 4.85 | 0.429 | 0.975 |
| ENDOU | OID31498 | Oncology | ENDOU | P21128 | 12 | 47709734 | 47725567 | 1 | 2182.22 | 1.13 | 0.83 | 1.55 | 0.433 | 0.975 |
| GSTA3 | OID21242 | Oncology | GSTA3 | Q16772 | 6 | 52896639 | 52909698 | 1 | 1650.39 | 1.13 | 0.83 | 1.54 | 0.435 | 0.975 |
| HGFAC | OID30718 | Inflammat | HGFAC | Q04756 | 4 | 3441968 | 3449486 | 1 | 3645.14 | 1.09 | 0.88 | 1.34 | 0.439 | 0.975 |
| ITIH4 | OID30762 | Inflammat | ITIH4 | Q14624 | 3 | 52812962 | 52830688 | 1 | 961.00 | 1.17 | 0.79 | 1.75 | 0.436 | 0.975 |
| ITIH5 | OID31430 | Oncology | ITIH5 | Q86UX2 | 10 | 7559270 | 7666998 | 1 | 169.00 | 0.68 | 0.26 | 1.79 | 0.433 | 0.975 |
| LIPF | OID30266 | Cardiome | LIPF | P07098 | 10 | 88664441 | 88678814 | 1 | 1501.56 | 0.87 | 0.62 | 1.23 | 0.438 | 0.975 |
| NAAA | OID20931 | Neurology | NAAA | Q02083 | 4 | 75913660 | 75941013 | 1 | 5525.44 | 0.94 | 0.79 | 1.10 | 0.429 | 0.975 |
| NPTX1 | OID21074 | Neurology | NPTX1 | Q15818 | 17 | 80466834 | 80477843 | 1 | 3959.31 | 0.92 | 0.75 | 1.13 | 0.439 | 0.975 |
| NUB1 | OID20510 | Inflammat | NUB1 | Q9Y5A7 | 7 | 151341772 | 151378449 | 1 | 491.36 | 1.25 | 0.72 | 2.16 | 0.433 | 0.975 |
| PLXNA4 | OID20597 | Inflammat | PLXNA4 | Q9HCM2 | 7 | 132123340 | 132648688 | 1 | 133.53 | 0.64 | 0.21 | 1.98 | 0.439 | 0.975 |
| PSRC1 | OID21169 | Oncology | PSRC1 | Q6PGN9 | 1 | 109279556 | 109283186 | 1 | 454.69 | 1.24 | 0.72 | 2.13 | 0.434 | 0.975 |
| PTGR1 | OID30297 | Cardiome | PTGR1 | Q14914 | 9 | 111549722 | 111599893 | 1 | 1988.11 | 1.11 | 0.85 | 1.46 | 0.430 | 0.975 |
| RILPL2 | OID30342 | Cardiome | RILPL2 | Q969X0 | 12 | 123415039 | 123436684 | 1 | 114.10 | 1.57 | 0.51 | 4.81 | 0.433 | 0.975 |
| SNX18 | OID31476 | Oncology | SNX18 | Q96RF0 | 5 | 54517759 | 54546586 | 1 | 92.16 | 0.60 | 0.17 | 2.17 | 0.437 | 0.975 |
| SORT1 | OID20213 | Cardiome | SORT1 | Q99523 | 1 | 109309568 | 109397918 | 1 | 431.72 | 0.79 | 0.44 | 1.43 | 0.437 | 0.975 |
| THOP1 | OID20194 | Cardiome | THOP1 | P52888 | 19 | 2785503 | 2815807 | 1 | 1122.25 | 0.86 | 0.58 | 1.26 | 0.434 | 0.975 |
| TNFRSF17 | OID30589 | Inflammat | TNFRSF1 | Q02223 | 16 | 11965210 | 11968068 | 1 | 324.00 | 0.76 | 0.38 | 1.52 | 0.436 | 0.975 |
| TFPI | OID20388 | Cardiome | TFPI | P10646 | 2 | 187464230 | 187565760 | 1 | 1881.39 | 1.12 | 0.84 | 1.51 | 0.440 | 0.976 |
| C2CD2L | OID20864 | Neurology | C2CD2L | O14523 | 11 | 119102198 | 119118544 | 1 | 49.00 | 2.02 | 0.34 | 12.11 | 0.442 | 0.977 |
| GP5 | OID30611 | Inflammat | GP5 | P40197 | 3 | 194394821 | 194399266 | 1 | 231.72 | 1.39 | 0.60 | 3.20 | 0.442 | 0.977 |
| VAV3 | OID30795 | Neurology | VAV3 | Q9UKW4 | 1 | 107571161 | 107965180 | 1 | 52.16 | 1.98 | 0.35 | 11.30 | 0.441 | 0.977 |
| NINJ1 | OID21167 | Oncology | NINJ1 | Q92982 | 9 | 93121496 | 93134251 | 1 | 922.64 | 1.17 | 0.78 | 1.76 | 0.444 | 0.978 |
| RNASET2 | OID20321 | Cardiome | RNASET2 | O00584 | 6 | 166922113 | 166957191 | 1 | 2657.98 | 0.91 | 0.71 | 1.16 | 0.443 | 0.978 |
| COL4A1 | OID20208 | Cardiome | COL4A1 | P02462 | 13 | 110148963 | 110307157 | 1 | 529.00 | 0.81 | 0.47 | 1.39 | 0.446 | 0.982 |
| C1S | OID30731 | Inflammat | C1S | P09871 | 12 | 6988259 | 7071032 | 1 | 3687.80 | 0.92 | 0.75 | 1.13 | 0.447 | 0.982 |
| PLXDC2 | OID31101 | Neurology | PLXDC2 | Q6UX71 | 10 | 19816239 | 20289856 | 1 | 91.31 | 0.59 | 0.15 | 2.31 | 0.448 | 0.983 |
| ADAM15 | OID20109 | Cardiome | ADAM15 | Q13444 | 1 | 155050566 | 155062775 | 1 | 10485.76 | 1.05 | 0.93 | 1.18 | 0.450 | 0.984 |
| CTSH | OID20113 | Cardiome | CTSH | P09668 | 15 | 78921058 | 78949574 | 1 | 13409.64 | 0.96 | 0.85 | 1.07 | 0.452 | 0.984 |
| ENPEP | OID30135 | Cardiome | ENPEP | Q07075 | 4 | 110365733 | 110565285 | 1 | 90.25 | 1.76 | 0.40 | 7.70 | 0.451 | 0.984 |
| FCRL5 | OID21111 | Neurology | FCRL5 | Q96RD9 | 1 | 157513377 | 157552515 | 1 | 3000.60 | 1.10 | 0.86 | 1.39 | 0.451 | 0.984 |
| IL16 | OID20633 | Inflammat | IL16 | Q14005 | 15 | 81159575 | 81314058 | 1 | 5544.52 | 1.07 | 0.90 | 1.27 | 0.452 | 0.984 |
| MMP13 | OID20807 | Neurology | MMP13 | P45452 | 11 | 102942995 | 102955732 | 1 | 297.56 | 0.77 | 0.39 | 1.53 | 0.452 | 0.984 |
| PI3 | OID20326 | Cardiome | PI3 | P19957 | 20 | 45174902 | 45176544 | 1 | 1538.38 | 1.14 | 0.81 | 1.60 | 0.451 | 0.984 |
| C1R | OID30753 | Inflammat | C1R | P00736 | 12 | 7080214 | 7092540 | 1 | 1741.17 | 0.89 | 0.66 | 1.20 | 0.456 | 0.985 |
| CCL27 | OID20121 | Cardiome | CCL27 | Q9Y4X3 | 9 | 34661880 | 34664048 | 1 | 747.11 | 0.83 | 0.51 | 1.35 | 0.456 | 0.985 |
| CHGA | OID30403 | Cardiome | CHGA | P10645 | 14 | 92923150 | 92935285 | 1 | 605.16 | 1.23 | 0.72 | 2.10 | 0.457 | 0.985 |
| GPR158 | OID31013 | Neurology | GPR158 | Q5T848 | 10 | 25174802 | 25602229 | 1 | 64.00 | 0.54 | 0.11 | 2.72 | 0.456 | 0.985 |
| IL5RA | OID20601 | Inflammat | IL5RA | Q01344 | 3 | 3066324 | 3126613 | 1 | 2209.00 | 0.90 | 0.68 | 1.19 | 0.454 | 0.985 |
| JAM3 | OID30586 | Inflammat | JAM3 | Q9BX67 | 11 | 134069071 | 134152001 | 1 | 186.78 | 0.70 | 0.28 | 1.78 | 0.453 | 0.985 |
| RTN4IP1 | OID31030 | Neurology | RTN4IP1 | Q8WWV3 | 6 | 106570771 | 106629498 | 1 | 135.14 | 1.47 | 0.53 | 4.05 | 0.455 | 0.985 |
| SPRR3 | OID31131 | Neurology | SPRR3 | Q9UBC9 | 1 | 153001747 | 153003856 | 1 | 1175.51 | 1.16 | 0.78 | 1.72 | 0.455 | 0.985 |
| TNFAIP8L2 | OID30629 | Inflammat | TNFAIP8L | Q6P589 | 1 | 151156649 | 151159749 | 1 | 53.78 | 1.98 | 0.33 | 12.03 | 0.458 | 0.985 |
| CA5A | OID20075 | Cardiome | CA5A | P35218 | 16 | 87881546 | 87936580 | 1 | 8140.05 | 0.94 | 0.81 | 1.10 | 0.460 | 0.985 |
| CD300C | OID21010 | Neurology | CD300C | Q08708 | 17 | 74541073 | 74546115 | 1 | 2752.21 | 0.91 | 0.71 | 1.16 | 0.459 | 0.985 |
| MMP12 | OID21439 | Oncology | MMP12 | P39900 | 11 | 102862736 | 102874982 | 1 | 4636.37 | 0.93 | 0.77 | 1.12 | 0.460 | 0.985 |
| RNASE3 | OID20203 | Cardiome | RNASE3 | P12724 | 14 | 20891385 | 20892348 | 1 | 1120.28 | 1.15 | 0.80 | 1.66 | 0.459 | 0.985 |
| C1QTNF5 | OID30618 | Inflammat | C1QTNF5 | Q9BXJ0 | 11 | 119338942 | 119340940 | 1 | 265.69 | 0.73 | 0.31 | 1.70 | 0.465 | 0.986 |
| CD109 | OID21108 | Neurology | CD109 | Q6YHK3 | 6 | 73695785 | 73828316 | 1 | 11502.56 | 0.96 | 0.85 | 1.07 | 0.463 | 0.986 |
| EGFR | OID20319 | Cardiome | EGFR | P00533 | 7 | 55019017 | 55211628 | 1 | 596.76 | 0.81 | 0.46 | 1.42 | 0.465 | 0.986 |
| ENTPD5 | OID20104 | Cardiome | ENTPD5 | O75356 | 14 | 73958010 | 74019399 | 1 | 3275.88 | 0.92 | 0.75 | 1.14 | 0.467 | 0.986 |
| ERMAP | OID30539 | Inflammat | ERMAP | Q96PL5 | 1 | 42817122 | 42844991 | 1 | 169.00 | 1.47 | 0.52 | 4.18 | 0.467 | 0.986 |
| FOXO1 | OID20515 | Inflammat | FOXO1 | Q12778 | 13 | 40555667 | 40666641 | 1 | 241.98 | 0.71 | 0.29 | 1.77 | 0.466 | 0.986 |
| LEG1 | OID30608 | Inflammat | C6orf58 | Q6P5S2 | 6 | 127519455 | 127591820 | 1 | 2824.16 | 1.10 | 0.85 | 1.42 | 0.463 | 0.986 |
| LPA | OID30747 | Inflammat | LPA | P08519 | 6 | 160531482 | 160664275 | 1 | 5791.21 | 1.06 | 0.90 | 1.25 | 0.464 | 0.986 |
| RGMB | OID21045 | Neurology | RGMB | Q6NW40 | 5 | 98768650 | 98798643 | 1 | 596.76 | 0.81 | 0.47 | 1.42 | 0.465 | 0.986 |
| SEZ6L | OID21488 | Oncology | SEZ6L | Q9BYH1 | 22 | 26169462 | 26383597 | 1 | 482.31 | 0.81 | 0.47 | 1.41 | 0.462 | 0.986 |
| VASN | OID20345 | Cardiome | VASN | Q6EMK4 | 16 | 4371848 | 4383538 | 1 | 3192.25 | 0.92 | 0.75 | 1.14 | 0.464 | 0.986 |
| CFH | OID30790 | Inflammat | CFH | P08603 | 1 | 196651754 | 196752476 | 1 | 1772.41 | 0.90 | 0.68 | 1.20 | 0.468 | 0.987 |
| MMP1 | OID20672 | Inflammat | MMP1 | P03956 | 11 | 102789401 | 102798160 | 1 | 3136.00 | 0.92 | 0.74 | 1.15 | 0.468 | 0.987 |
| LRRC37A2 | OID31440 | Oncology | LRRC37A | A6NM11 | 17 | 46511508 | 46556910 | 1 | 15348.46 | 1.04 | 0.94 | 1.15 | 0.472 | 0.993 |
| SELE | OID20290 | Cardiome | SELE | P16581 | 1 | 169722640 | 169764705 | 1 | 184.18 | 1.47 | 0.52 | 4.16 | 0.472 | 0.993 |
| ACHE | OID30585 | Inflammat | ACHE | P22303 | 7 | 100889994 | 100896974 | 1 | 1999.37 | 0.90 | 0.66 | 1.21 | 0.483 | 0.994 |
| AGXT | OID20139 | Cardiome | AGXT | P21549 | 2 | 240868824 | 240880502 | 1 | 1296.00 | 1.14 | 0.79 | 1.63 | 0.485 | 0.994 |
| CCDC50 | OID31001 | Neurology | CCDC50 | Q8IVM0 | 3 | 191329085 | 191398659 | 1 | 541.85 | 0.83 | 0.49 | 1.40 | 0.483 | 0.994 |
| CCL21 | OID20686 | Inflammat | CCL21 | O00585 | 9 | 34709005 | 34710136 | 1 | 165.77 | 0.71 | 0.27 | 1.88 | 0.490 | 0.994 |
| CCL3 | OID20610 | Inflammat | CCL3 | P10147 | 17 | 36088256 | 36090169 | 1 | 2177.78 | 0.91 | 0.69 | 1.19 | 0.489 | 0.994 |
| CCL7 | OID20523 | Inflammat | CCL7 | P80098 | 17 | 34270221 | 34272242 | 1 | 225.00 | 0.72 | 0.29 | 1.81 | 0.491 | 0.994 |
| CCN2 | OID20709 | Inflammat | CCN2 | P29279 | 6 | 131948176 | 131951372 | 1 | 1555.86 | 0.89 | 0.65 | 1.23 | 0.482 | 0.994 |
| CD55 | OID20377 | Cardiome | CD55 | P08174 | 1 | 207321532 | 207386804 | 1 | 2928.01 | 0.92 | 0.74 | 1.16 | 0.486 | 0.994 |
| CD74 | OID21040 | Neurology | CD74 | P04233 | 5 | 150401637 | 150412969 | 1 | 62.55 | 1.87 | 0.32 | 10.86 | 0.486 | 0.994 |
| DUSP29 | OID30935 | Neurology | DUSP29 | Q68J44 | 10 | 75037472 | 75073642 | 1 | 306.25 | 0.78 | 0.38 | 1.58 | 0.484 | 0.994 |
| EDAR | OID20525 | Inflammat | EDAR | Q9UNE0 | 2 | 108894471 | 108989372 | 1 | 2691.02 | 1.09 | 0.86 | 1.37 | 0.488 | 0.994 |
| EDN1 | OID30224 | Cardiome | EDN1 | P05305 | 6 | 12290361 | 12297194 | 1 | 913.38 | 1.16 | 0.77 | 1.75 | 0.484 | 0.994 |
| FASLG | OID20665 | Inflammat | FASLG | P48023 | 1 | 172659103 | 172666876 | 1 | 121.00 | 0.63 | 0.18 | 2.25 | 0.478 | 0.994 |
| IFIT3 | OID31283 | Oncology | IFIT3 | O14879 | 10 | 89327307 | 89377473 | 1 | 144.00 | 0.70 | 0.26 | 1.91 | 0.487 | 0.994 |
| IFNLR1 | OID20506 | Inflammat | IFNLR1 | Q8IU57 | 1 | 24154168 | 24187959 | 1 | 2012.16 | 1.12 | 0.82 | 1.51 | 0.480 | 0.994 |
| LACRT | OID31367 | Oncology | LACRT | Q9GZZ8 | 12 | 54630811 | 54634895 | 1 | 76.56 | 1.68 | 0.38 | 7.36 | 0.491 | 0.994 |
| LGALS3BP | OID30765 | Inflammat | LGALS3B | Q08380 | 17 | 78971238 | 78979947 | 1 | 293.88 | 1.37 | 0.58 | 3.27 | 0.475 | 0.994 |
| LILRA3 | OID30376 | Cardiome | LILRA3 | Q8N6C8 | 19 | 54294163 | 54306461 | 1 | 18345.20 | 0.97 | 0.88 | 1.06 | 0.476 | 0.994 |
| LILRB1 | OID20323 | Cardiome | LILRB1 | Q8NHL6 | 19 | 54617158 | 54638022 | 1 | 5558.53 | 0.94 | 0.80 | 1.11 | 0.476 | 0.994 |
| PALM2 | OID31096 | Neurology | PALM2AK | Q8IXS6 | 9 | 109498325 | 110172512 | 1 | 676.00 | 0.84 | 0.52 | 1.36 | 0.481 | 0.994 |
| PDGFC | OID21183 | Oncology | PDGFC | Q9NRA1 | 4 | 156760454 | 156971799 | 1 | 102.23 | 1.51 | 0.47 | 4.84 | 0.491 | 0.994 |
| RABGAP1L | OID20447 | Inflammat | RABGAP1 | Q5R372 | 1 | 174159410 | 174995308 | 1 | 104.49 | 0.63 | 0.17 | 2.29 | 0.485 | 0.994 |
| REG4 | OID20784 | Inflammat | REG4 | Q9BYZ8 | 1 | 119794017 | 119811580 | 1 | 511.08 | 0.81 | 0.45 | 1.45 | 0.477 | 0.994 |
| RNASE1 | OID30672 | Inflammat | RNASE1 | P07998 | 14 | 20801228 | 20802855 | 1 | 735.77 | 1.19 | 0.74 | 1.92 | 0.481 | 0.994 |
| SCRN1 | OID20542 | Inflammat | SCRN1 | Q12765 | 7 | 29920104 | 29990289 | 1 | 197.48 | 0.73 | 0.29 | 1.80 | 0.489 | 0.994 |
| SERPINE2 | OID30359 | Cardiome | SERPINE | P07093 | 2 | 223975045 | 224039318 | 1 | 4931.16 | 0.94 | 0.79 | 1.12 | 0.491 | 0.994 |
| SLC16A1 | OID20799 | Neurology | SLC16A1 | P53985 | 1 | 112911847 | 112957593 | 1 | 60.06 | 0.58 | 0.12 | 2.72 | 0.491 | 0.994 |
| SOD2 | OID21114 | Neurology | SOD2 | P04179 | 6 | 159669069 | 159762529 | 1 | 390.06 | 0.79 | 0.41 | 1.53 | 0.491 | 0.994 |
| TDRKH | OID20875 | Neurology | TDRKH | Q9Y2W6 | 1 | 151770107 | 151791416 | 1 | 1540.56 | 0.90 | 0.67 | 1.22 | 0.490 | 0.994 |
| TEK | OID21496 | Oncology | TEK | Q02763 | 9 | 27109141 | 27230174 | 1 | 3025.00 | 0.92 | 0.72 | 1.16 | 0.474 | 0.994 |

| | | | | | | | | | | | | | | |
|---|---|---|---|---|---|---|---|---|---|---|---|---|---|---|
| THY1 | OID21050 | Neurology | THY1 | P04216 | 11 | 119415476 | 119424985 | 1 | 1914.94 | 0.91 | 0.69 | 1.19 | 0.489 | 0.994 |
| TIGIT | OID31320 | Oncology | TIGIT | Q495A1 | 3 | 114276913 | 114310288 | 1 | 141.02 | 0.67 | 0.22 | 2.03 | 0.477 | 0.994 |
| LGMN | OID20773 | Inflammat | LGMN | Q99538 | 14 | 92703807 | 92748679 | 1 | 962.82 | 1.18 | 0.74 | 1.88 | 0.493 | 0.995 |
| PPP1R14A | OID31011 | Neurology | PPP1R14 | Q96A00 | 19 | 38251237 | 38256532 | 1 | 630.57 | 1.19 | 0.73 | 1.93 | 0.493 | 0.995 |
| APOD | OID30746 | Inflammat | APOD | P05090 | 3 | 195568705 | 195584033 | 1 | 595.80 | 0.85 | 0.52 | 1.38 | 0.506 | 0.996 |
| BSG | OID20626 | Inflammat | BSG | P35613 | 19 | 571277 | 583494 | 1 | 343.67 | 1.26 | 0.65 | 2.43 | 0.498 | 0.996 |
| CARHSP1 | OID20859 | Neurology | CARHSP | Q9Y2V2 | 16 | 8852942 | 8869012 | 1 | 124.16 | 1.49 | 0.45 | 4.97 | 0.513 | 0.996 |
| CD5L | OID30775 | Inflammat | CD5L | O43866 | 1 | 157830911 | 157898256 | 1 | 625.00 | 0.85 | 0.52 | 1.38 | 0.507 | 0.996 |
| CEACAM21 | OID20548 | Inflammat | CEACAM | Q3KPI0 | 19 | 41549518 | 41586844 | 1 | 19818.38 | 0.97 | 0.89 | 1.06 | 0.498 | 0.996 |
| CELSR2 | OID30593 | Inflammat | CELSR2 | Q9HCU4 | 1 | 109249539 | 109275751 | 1 | 2612.35 | 0.92 | 0.72 | 1.18 | 0.504 | 0.996 |
| CLIP2 | OID20559 | Inflammat | CLIP2 | Q9UDT6 | 7 | 74289407 | 74405935 | 1 | 277.78 | 1.30 | 0.61 | 2.80 | 0.499 | 0.996 |
| CLSTN3 | OID31092 | Neurology | CLSTN3 | Q9BQT9 | 12 | 7129698 | 7158945 | 1 | 3422.25 | 0.93 | 0.76 | 1.14 | 0.509 | 0.996 |
| DDT | OID30372 | Cardiome | DDT | P30046 | 22 | 23970542 | 23980525 | 1 | 663.06 | 1.17 | 0.73 | 1.88 | 0.504 | 0.996 |
| DKK4 | OID20934 | Neurology | DKK4 | Q9UBT3 | 8 | 42374063 | 42377229 | 1 | 1046.71 | 0.88 | 0.60 | 1.28 | 0.496 | 0.996 |
| DPP7 | OID20142 | Cardiome | DPP7 | Q9UHL4 | 9 | 137110546 | 137115177 | 1 | 3299.86 | 0.93 | 0.76 | 1.15 | 0.510 | 0.996 |
| ENG | OID20287 | Cardiome | ENG | P17813 | 9 | 127815013 | 127854658 | 1 | 1048.14 | 0.88 | 0.61 | 1.28 | 0.499 | 0.996 |
| FABP6 | OID20076 | Cardiome | FABP6 | P51161 | 5 | 160187367 | 160238735 | 1 | 268.14 | 0.77 | 0.37 | 1.64 | 0.502 | 0.996 |
| FCRL3 | OID20443 | Inflammat | FCRL3 | Q96P31 | 1 | 157674321 | 157700769 | 1 | 14550.39 | 1.03 | 0.94 | 1.14 | 0.515 | 0.996 |
| GZMB | OID20604 | Inflammat | GZMB | P10144 | 14 | 24630954 | 24634267 | 1 | 1547.11 | 0.90 | 0.65 | 1.23 | 0.507 | 0.996 |
| HMCN2 | OID30592 | Inflammat | HMCN2 | Q8NDA2 | 9 | 130265760 | 130434123 | 1 | 6084.00 | 1.05 | 0.91 | 1.22 | 0.503 | 0.996 |
| IGFBPL1 | OID20155 | Cardiome | IGFBPL1 | Q8WX77 | 9 | 38406528 | 38424454 | 1 | 3025.00 | 0.92 | 0.73 | 1.17 | 0.516 | 0.996 |
| IL17RB | OID20585 | Inflammat | IL17RB | Q9NRM6 | 3 | 53846568 | 53865794 | 1 | 10175.77 | 1.04 | 0.92 | 1.18 | 0.506 | 0.996 |
| IL1RAP | OID21126 | Neurology | IL1RAP | Q9NPH3 | 3 | 190514051 | 190659750 | 1 | 19628.01 | 1.03 | 0.94 | 1.13 | 0.516 | 0.996 |
| ITGB5 | OID21522 | Oncology | ITGB5 | P18084 | 3 | 124761948 | 124901418 | 1 | 121.00 | 0.63 | 0.17 | 2.39 | 0.497 | 0.996 |
| KLRD1 | OID20632 | Inflammat | KLRD1 | Q13241 | 12 | 10226058 | 10329608 | 1 | 3565.80 | 1.08 | 0.86 | 1.36 | 0.514 | 0.996 |
| MEGF11 | OID30099 | Cardiome | MEGF11 | A6BM72 | 15 | 65895079 | 66253756 | 1 | 332.05 | 0.78 | 0.38 | 1.59 | 0.494 | 0.996 |
| MEP1B | OID20168 | Cardiome | MEP1B | Q16820 | 18 | 32185069 | 32220404 | 1 | 18023.06 | 1.03 | 0.94 | 1.13 | 0.505 | 0.996 |
| NAP1L4 | OID31475 | Oncology | NAP1L4 | Q99733 | 11 | 2944431 | 2992377 | 1 | 853.94 | 1.15 | 0.75 | 1.77 | 0.515 | 0.996 |
| PKLR | OID20682 | Inflammat | PKLR | P30613 | 1 | 155289293 | 155301438 | 1 | 588.87 | 1.20 | 0.71 | 2.03 | 0.495 | 0.996 |
| PROCR | OID30149 | Cardiome | PROCR | Q9UNN8 | 20 | 35172072 | 35216240 | 1 | 2718.88 | 0.92 | 0.73 | 1.17 | 0.516 | 0.996 |
| PSAP | OID30419 | Cardiome | PSAP | P07602 | 10 | 71816298 | 71851251 | 1 | 765.44 | 0.86 | 0.54 | 1.35 | 0.501 | 0.996 |
| PYDC1 | OID30202 | Cardiome | PYDC1 | Q8WXC3 | 16 | 31215962 | 31217135 | 1 | 1910.94 | 0.90 | 0.65 | 1.25 | 0.515 | 0.996 |
| RAD23B | OID21166 | Oncology | RAD23B | P54727 | 9 | 107283137 | 107332192 | 1 | 64.00 | 1.64 | 0.37 | 7.24 | 0.516 | 0.996 |
| RANBP1 | OID30258 | Cardiome | RANBP1 | P43487 | 22 | 20115938 | 20127355 | 1 | 52.35 | 0.55 | 0.10 | 3.14 | 0.500 | 0.996 |
| RBP1 | OID30913 | Neurology | RBP1 | P09455 | 3 | 139517434 | 139539829 | 1 | 187.69 | 0.74 | 0.30 | 1.83 | 0.511 | 0.996 |
| SFRP1 | OID20984 | Neurology | SFRP1 | Q8N474 | 8 | 41261962 | 41309473 | 1 | 729.00 | 1.19 | 0.71 | 1.98 | 0.514 | 0.996 |
| SIRPA | OID20304 | Cardiome | SIRPA | P78324 | 20 | 1894167 | 1940592 | 1 | 23793.06 | 1.03 | 0.95 | 1.11 | 0.507 | 0.996 |
| SNX15 | OID30644 | Inflammat | SNX15 | Q9NRS6 | 11 | 65027439 | 65040572 | 1 | 3609.24 | 0.93 | 0.75 | 1.15 | 0.509 | 0.996 |
| TCN2 | OID20341 | Cardiome | TCN2 | P20062 | 22 | 30607003 | 30627271 | 1 | 4946.78 | 1.06 | 0.89 | 1.28 | 0.502 | 0.996 |
| TMEM106A | OID31411 | Oncology | TMEM106 | Q96A25 | 17 | 43211835 | 43220041 | 1 | 178.89 | 1.38 | 0.53 | 3.55 | 0.507 | 0.996 |
| TYRO3 | OID20161 | Cardiome | TYRO3 | Q06418 | 15 | 41557675 | 41583589 | 1 | 6520.56 | 0.95 | 0.81 | 1.11 | 0.508 | 0.996 |
| VIT | OID30334 | Cardiome | VIT | Q6UXI7 | 2 | 36696690 | 36814794 | 1 | 3808.65 | 0.93 | 0.74 | 1.16 | 0.509 | 0.996 |
| XRCC4 | OID20869 | Neurology | XRCC4 | Q13426 | 5 | 83077498 | 83353787 | 1 | 75.11 | 1.62 | 0.39 | 6.72 | 0.506 | 0.996 |
| YOD1 | OID30181 | Cardiome | YOD1 | Q5VVQ6 | 1 | 207043849 | 207052980 | 1 | 229.31 | 1.35 | 0.55 | 3.30 | 0.515 | 0.996 |
| GNPDA1 | OID30197 | Cardiome | GNPDA1 | P46926 | 5 | 141991749 | 142013041 | 1 | 135.40 | 0.70 | 0.24 | 2.05 | 0.519 | 0.997 |
| LRIG1 | OID21429 | Oncology | LRIG1 | Q96JA1 | 3 | 66378797 | 66501263 | 1 | 11502.56 | 1.04 | 0.92 | 1.18 | 0.519 | 0.997 |
| OSTN | OID30909 | Neurology | OSTN | P61366 | 3 | 191199241 | 191265615 | 1 | 138.72 | 0.71 | 0.26 | 1.98 | 0.518 | 0.997 |
| SARG | OID30331 | Cardiome | C1orf116 | Q9BW04 | 1 | 207018522 | 207032756 | 1 | 88.23 | 0.62 | 0.14 | 2.67 | 0.519 | 0.997 |
| ACOX1 | OID20124 | Cardiome | ACOX1 | Q15067 | 17 | 75941507 | 75979177 | 1 | 95.06 | 0.67 | 0.19 | 2.37 | 0.532 | 0.998 |
| ADAMTS13 | OID20249 | Cardiome | ADAMTS | Q76LX8 | 9 | 133414358 | 133459402 | 1 | 2531.13 | 1.09 | 0.83 | 1.42 | 0.528 | 0.998 |
| CDCP1 | OID20940 | Neurology | CDCP1 | Q9H5V8 | 3 | 45082277 | 45146422 | 1 | 1133.44 | 0.88 | 0.59 | 1.31 | 0.525 | 0.998 |
| CST1 | OID30581 | Inflammat | CST1 | P01037 | 20 | 23747562 | 23751268 | 1 | 4196.16 | 1.06 | 0.89 | 1.26 | 0.531 | 0.998 |
| DDAH1 | OID21299 | Oncology | DDAH1 | O94760 | 1 | 85318481 | 85578363 | 1 | 427.11 | 1.22 | 0.65 | 2.30 | 0.534 | 0.998 |
| FCER2 | OID21133 | Neurology | FCER2 | P06734 | 19 | 7688758 | 7702146 | 1 | 1740.08 | 0.90 | 0.65 | 1.25 | 0.529 | 0.998 |
| GNPDA2 | OID30597 | Inflammat | GNPDA2 | Q8TDQ7 | 4 | 44682200 | 44726588 | 1 | 870.25 | 0.87 | 0.58 | 1.32 | 0.528 | 0.998 |
| HLA-A | OID31048 | Neurology | HLA-A | P04439 | 6 | 29941260 | 29945884 | 1 | 15907.52 | 1.03 | 0.94 | 1.14 | 0.525 | 0.998 |
| KITLG | OID20196 | Cardiome | KITLG | P21583 | 12 | 88492793 | 88580851 | 1 | 45.56 | 0.54 | 0.09 | 3.47 | 0.520 | 0.998 |
| KLK4 | OID21391 | Oncology | KLK4 | Q9Y5K2 | 19 | 50906351 | 50911395 | 1 | 3565.80 | 1.08 | 0.85 | 1.37 | 0.526 | 0.998 |
| MLN | OID20541 | Inflammat | MLN | P12872 | 6 | 33794673 | 33804003 | 1 | 3497.88 | 0.93 | 0.74 | 1.17 | 0.531 | 0.998 |
| PAXX | OID30591 | Inflammat | PAXX | Q9BUH6 | 9 | 136992422 | 136993984 | 1 | 425.39 | 1.22 | 0.66 | 2.26 | 0.529 | 0.998 |
| PENK | OID30666 | Inflammat | PENK | P01210 | 8 | 56436674 | 56446671 | 1 | 1849.00 | 0.90 | 0.66 | 1.24 | 0.532 | 0.998 |
| PGLYRP1 | OID20338 | Cardiome | PGLYRP1 | O75594 | 19 | 46019153 | 46023053 | 1 | 1378.27 | 1.11 | 0.79 | 1.56 | 0.534 | 0.998 |
| PNLIPRP2 | OID20775 | Inflammat | PNLIPRP | P54317 | 10 | 116620953 | 116645143 | 1 | 20164.00 | 0.97 | 0.90 | 1.06 | 0.522 | 0.998 |
| PRSS22 | OID30483 | Inflammat | PRSS22 | Q9GZN4 | 16 | 2852730 | 2858170 | 1 | 1056.25 | 0.88 | 0.60 | 1.30 | 0.525 | 0.998 |
| PRSS53 | OID31093 | Neurology | PRSS53 | Q2L4Q9 | 16 | 31083437 | 31089628 | 1 | 12237.89 | 0.97 | 0.87 | 1.08 | 0.525 | 0.998 |
| PRTN3 | OID20329 | Cardiome | PRTN3 | P24158 | 19 | 840999 | 848175 | 1 | 3074.09 | 0.93 | 0.75 | 1.16 | 0.521 | 0.998 |
| PSAPL1 | OID31473 | Oncology | PSAPL1 | Q6NUJ1 | 4 | 7430285 | 7434930 | 1 | 4900.00 | 0.94 | 0.77 | 1.14 | 0.531 | 0.998 |
| REG1A | OID20415 | Cardiome | REG1A | P05451 | 2 | 79120362 | 79123409 | 1 | 984.39 | 1.13 | 0.77 | 1.67 | 0.529 | 0.998 |
| REG1B | OID20266 | Cardiome | REG1B | P48304 | 2 | 79085023 | 79088019 | 1 | 1859.77 | 1.10 | 0.82 | 1.45 | 0.529 | 0.998 |
| RNF149 | OID30235 | Cardiome | RNF149 | Q8NC42 | 2 | 101271219 | 101308701 | 1 | 1233.77 | 1.12 | 0.79 | 1.57 | 0.533 | 0.998 |
| SIGLEC10 | OID20537 | Inflammat | SIGLEC1 | Q96LC7 | 19 | 51410020 | 51417803 | 1 | 2957.11 | 1.08 | 0.84 | 1.39 | 0.534 | 0.998 |
| SUSD4 | OID31398 | Oncology | SUSD4 | Q5VX71 | 1 | 223220819 | 223364233 | 1 | 2256.25 | 0.92 | 0.72 | 1.19 | 0.525 | 0.998 |
| TGFB1 | OID20621 | Inflammat | TGFB1 | P01137 | 19 | 41301587 | 41353922 | 1 | 829.44 | 1.15 | 0.75 | 1.77 | 0.532 | 0.998 |
| TPK1 | OID30338 | Cardiome | TPK1 | Q9H3S4 | 7 | 144451941 | 144836395 | 1 | 1743.06 | 0.91 | 0.69 | 1.21 | 0.525 | 0.998 |
| AAMDC | OID30236 | Cardiome | AAMDC | Q9H7C9 | 11 | 77821109 | 77918432 | 1 | 7861.78 | 0.97 | 0.84 | 1.12 | 0.678 | 0.998 |
| ABCA2 | OID30146 | Cardiome | ABCA2 | Q9BZC7 | 9 | 137007234 | 137028915 | 1 | 58.41 | 1.05 | 0.23 | 4.87 | 0.953 | 0.998 |
| ABO | OID30675 | Inflammat | ABO | P16442 | 9 | 133233278 | 133276024 | 1 | 21425.64 | 1.02 | 0.94 | 1.11 | 0.655 | 0.998 |
| ACAA1 | OID21269 | Oncology | ACAA1 | P09110 | 3 | 38103129 | 38137242 | 1 | 129.13 | 0.88 | 0.31 | 2.49 | 0.803 | 0.998 |
| ACP5 | OID20314 | Cardiome | ACP5 | P13686 | 19 | 11574660 | 11579008 | 1 | 2099.31 | 1.04 | 0.78 | 1.38 | 0.799 | 0.998 |
| ACP6 | OID21432 | Oncology | ACP6 | Q9NPH0 | 1 | 147629652 | 147670524 | 1 | 16065.56 | 0.98 | 0.89 | 1.09 | 0.735 | 0.998 |
| ACRBP | OID31385 | Oncology | ACRBP | Q8NEB7 | 12 | 6638075 | 6647433 | 1 | 405.02 | 1.17 | 0.64 | 2.14 | 0.607 | 0.998 |
| ACRV1 | OID30513 | Inflammat | ACRV1 | P26436 | 11 | 125671522 | 125680874 | 1 | 121.00 | 1.26 | 0.20 | 7.83 | 0.802 | 0.998 |
| ACTA2 | OID20079 | Cardiome | ACTA2 | P62736 | 10 | 88935074 | 88991339 | 1 | 73.47 | 1.52 | 0.31 | 7.33 | 0.603 | 0.998 |
| ACVRL1 | OID20999 | Neurology | ACVRL1 | P37023 | 12 | 51906908 | 51923361 | 1 | 1909.02 | 1.01 | 0.75 | 1.38 | 0.925 | 0.998 |
| ACY3 | OID30086 | Cardiome | ACY3 | Q96HD9 | 11 | 67642555 | 67650730 | 1 | 1173.06 | 1.07 | 0.74 | 1.56 | 0.717 | 0.998 |
| ACYP1 | OID30642 | Inflammat | ACYP1 | P07311 | 14 | 75053237 | 75069483 | 1 | 1230.84 | 0.96 | 0.68 | 1.35 | 0.806 | 0.998 |
| ADAM22 | OID21001 | Neurology | ADAM22 | Q9P0K1 | 7 | 87934143 | 88202889 | 1 | 4987.89 | 0.98 | 0.83 | 1.15 | 0.769 | 0.998 |
| ADAM8 | OID21039 | Neurology | ADAM8 | P78325 | 10 | 133262420 | 133276868 | 1 | 3490.84 | 1.01 | 0.82 | 1.25 | 0.915 | 0.998 |
| ADAMTS15 | OID21275 | Oncology | ADAMTS | Q8TE58 | 11 | 130448645 | 130476645 | 1 | 128.89 | 0.96 | 0.31 | 2.96 | 0.950 | 0.998 |
| ADAMTS4 | OID30471 | Inflammat | ADAMTS | O75173 | 1 | 161184302 | 161199054 | 1 | 217.56 | 0.92 | 0.41 | 2.08 | 0.844 | 0.998 |
| ADAMTS8 | OID21502 | Oncology | ADAMTS | Q9UP79 | 11 | 130404923 | 130428609 | 1 | 3797.64 | 1.04 | 0.84 | 1.29 | 0.707 | 0.998 |
| ADAMTSL5 | OID30355 | Cardiome | ADAMTSI | Q6ZMM2 | 19 | 1505022 | 1513604 | 1 | 2575.56 | 1.07 | 0.83 | 1.38 | 0.598 | 0.998 |
| ADD1 | OID30655 | Inflammat | ADD1 | P35611 | 4 | 2843844 | 2930076 | 1 | 386.78 | 1.10 | 0.55 | 2.20 | 0.785 | 0.998 |
| ADGRB3 | OID20974 | Neurology | ADGRB3 | O60242 | 6 | 68635282 | 69390571 | 1 | 13835.64 | 1.00 | 0.90 | 1.11 | 0.953 | 0.998 |
| ADGRE1 | OID31483 | Oncology | ADGRE1 | Q14246 | 19 | 6887566 | 6940459 | 1 | 1369.00 | 0.98 | 0.70 | 1.37 | 0.904 | 0.998 |
| ADGRE2 | OID20755 | Inflammat | ADGRE2 | Q9UHX3 | 19 | 14732392 | 14778560 | 1 | 1287.02 | 0.97 | 0.66 | 1.42 | 0.867 | 0.998 |
| ADH4 | OID20117 | Cardiome | ADH4 | P08319 | 4 | 99123657 | 99157792 | 1 | 344.90 | 0.90 | 0.43 | 1.91 | 0.789 | 0.998 |
| ADIPOQ | OID30748 | Inflammat | ADIPOQ | Q15848 | 3 | 186842704 | 186858463 | 1 | 945.56 | 1.01 | 0.62 | 1.65 | 0.956 | 0.998 |
| ADM | OID21467 | Oncology | ADM | P35318 | 11 | 10305073 | 10307397 | 1 | 522.45 | 1.02 | 0.57 | 1.84 | 0.949 | 0.998 |
| AFAP1 | OID30569 | Inflammat | AFAP1 | Q8N556 | 4 | 7758714 | 7939926 | 1 | 18326.39 | 0.97 | 0.89 | 1.07 | 0.593 | 0.998 |
| AGR2 | OID20896 | Neurology | AGR2 | O95994 | 7 | 16791811 | 16833433 | 1 | 178.89 | 0.87 | 0.36 | 2.11 | 0.757 | 0.998 |
| AGT | OID30709 | Inflammat | AGT | P01019 | 1 | 230690776 | 230745576 | 1 | 3844.00 | 0.96 | 0.79 | 1.17 | 0.675 | 0.998 |
| AHNAK2 | OID30244 | Cardiome | AHNAK2 | Q8IVF2 | 14 | 104937244 | 104978374 | 1 | 729.00 | 0.99 | 0.63 | 1.56 | 0.960 | 0.998 |
| AIF1 | OID21234 | Oncology | AIF1 | P55008 | 6 | 31615217 | 31617021 | 1 | 192.52 | 0.94 | 0.40 | 2.20 | 0.892 | 0.998 |
| AIF1L | OID30204 | Cardiome | AIF1L | Q9BQI0 | 9 | 131096476 | 131123152 | 1 | 1511.27 | 0.93 | 0.67 | 1.29 | 0.674 | 0.998 |
| AKR1B1 | OID21244 | Oncology | AKR1B1 | P15121 | 7 | 134442356 | 134459284 | 1 | 316.41 | 1.02 | 0.51 | 2.04 | 0.952 | 0.998 |
| ALDH1A1 | OID21128 | Neurology | ALDH1A1 | P00352 | 9 | 72900671 | 73080442 | 1 | 129.39 | 0.81 | 0.25 | 2.66 | 0.732 | 0.998 |

| | | | | | | | | | | | | | | |
|---|---|---|---|---|---|---|---|---|---|---|---|---|---|---|
| ALDH3A1 | OID20512 | Inflammat | ALDH3A1 | P30838 | 17 | 19737984 | 19748943 | 1 | 3660.25 | 0.96 | 0.78 | 1.17 | 0.670 | 0.998 |
| ALDH5A1 | OID31047 | Neurology | ALDH5A1 | P51649 | 6 | 24494867 | 24537207 | 1 | 310.64 | 0.92 | 0.47 | 1.80 | 0.814 | 0.998 |
| ALPP | OID21383 | Oncology | ALPP | P05187 | 2 | 232378724 | 232382889 | 1 | 3160.94 | 1.04 | 0.84 | 1.30 | 0.705 | 0.998 |
| AMIGO2 | OID21370 | Oncology | AMIGO2 | Q86SJ2 | 12 | 47075707 | 47079959 | 1 | 385.14 | 0.91 | 0.49 | 1.69 | 0.769 | 0.998 |
| AMOTL2 | OID31351 | Oncology | AMOTL2 | Q9Y2J4 | 3 | 134355347 | 134375479 | 1 | 58.14 | 1.32 | 0.27 | 6.47 | 0.735 | 0.998 |
| AMY2B | OID20340 | Cardiome | AMY2B | P19961 | 1 | 103553815 | 103579534 | 1 | 2215.72 | 1.03 | 0.81 | 1.32 | 0.804 | 0.998 |
| ANG | OID20403 | Cardiome | ANG | P03950 | 14 | 20684177 | 20698971 | 1 | 3676.77 | 1.00 | 0.81 | 1.25 | 0.966 | 0.998 |
| ANGPT1 | OID20740 | Inflammat | ANGPT1 | Q15389 | 8 | 107249482 | 107498055 | 1 | 594.14 | 1.17 | 0.68 | 2.01 | 0.572 | 0.998 |
| ANKMY2 | OID30518 | Inflammat | ANKMY2 | Q8IV38 | 7 | 16599779 | 16645817 | 1 | 105.06 | 0.77 | 0.23 | 2.60 | 0.676 | 0.998 |
| ANPEP | OID20350 | Cardiome | ANPEP | P15144 | 15 | 89784895 | 89815401 | 1 | 2620.16 | 0.93 | 0.72 | 1.20 | 0.594 | 0.998 |
| ANXA2 | OID30263 | Cardiome | ANXA2 | P07355 | 15 | 60347134 | 60402883 | 1 | 3320.64 | 1.03 | 0.84 | 1.27 | 0.751 | 0.998 |
| ANXA5 | OID20810 | Neurology | ANXA5 | P08758 | 4 | 121667946 | 121696995 | 1 | 213.89 | 1.29 | 0.53 | 3.14 | 0.577 | 0.998 |
| AOC1 | OID20452 | Inflammat | AOC1 | P19801 | 7 | 150824627 | 150861504 | 1 | 1250.13 | 1.06 | 0.75 | 1.50 | 0.727 | 0.998 |
| AOC3 | OID20389 | Cardiome | AOC3 | Q16853 | 17 | 42851184 | 42858130 | 1 | 5220.06 | 0.98 | 0.82 | 1.16 | 0.804 | 0.998 |
| APBB1IP | OID21359 | Oncology | APBB1IP | Q7Z5R6 | 10 | 26438341 | 26567803 | 1 | 735.77 | 0.97 | 0.62 | 1.51 | 0.879 | 0.998 |
| APCS | OID30778 | Inflammat | APCS | P02743 | 1 | 159587826 | 159588865 | 1 | 496.92 | 1.06 | 0.58 | 1.96 | 0.844 | 0.998 |
| APEX1 | OID21434 | Oncology | APEX1 | P27695 | 14 | 20455191 | 20457772 | 1 | 3088.18 | 1.05 | 0.83 | 1.34 | 0.678 | 0.998 |
| APOBR | OID30246 | Cardiome | APOBR | Q0VD83 | 16 | 28494643 | 28498964 | 1 | 19705.14 | 1.02 | 0.93 | 1.11 | 0.722 | 0.998 |
| APOF | OID30701 | Inflammat | APOF | Q13790 | 12 | 56360568 | 56362857 | 1 | 628.58 | 0.98 | 0.57 | 1.69 | 0.953 | 0.998 |
| APOH | OID21072 | Neurology | APOH | P02749 | 17 | 66212033 | 66256525 | 1 | 4416.21 | 1.02 | 0.84 | 1.23 | 0.863 | 0.998 |
| APP | OID21070 | Neurology | APP | P05067 | 21 | 25880550 | 26171128 | 1 | 83.27 | 0.91 | 0.23 | 3.50 | 0.885 | 0.998 |
| AREG | OID21357 | Oncology | AREG | P15514 | 4 | 74445136 | 74455005 | 1 | 489.52 | 1.02 | 0.57 | 1.82 | 0.951 | 0.998 |
| ARFIP1 | OID31024 | Neurology | ARFIP1 | P53367 | 4 | 152779937 | 152918463 | 1 | 395.02 | 1.13 | 0.62 | 2.08 | 0.686 | 0.998 |
| ARHGAP25 | OID21255 | Oncology | ARHGAP2 | P42331 | 2 | 68679601 | 68826833 | 1 | 138.72 | 1.03 | 0.36 | 2.94 | 0.956 | 0.998 |
| ARHGAP45 | OID30653 | Inflammat | ARHGAP4 | Q92619 | 19 | 1065923 | 1086628 | 1 | 78.77 | 0.72 | 0.19 | 2.74 | 0.632 | 0.998 |
| ARHGEF5 | OID30871 | Neurology | ARHGEF5 | Q12774 | 7 | 144355288 | 144380632 | 1 | 2724.84 | 0.96 | 0.77 | 1.20 | 0.703 | 0.998 |
| ARSA | OID21138 | Neurology | ARSA | P15289 | 22 | 50622754 | 50628173 | 1 | 6935.19 | 0.99 | 0.85 | 1.15 | 0.896 | 0.998 |
| ASAH1 | OID30415 | Cardiome | ASAH1 | Q13510 | 8 | 18055992 | 18084998 | 1 | 3345.71 | 0.95 | 0.77 | 1.17 | 0.642 | 0.998 |
| ASAH2 | OID20996 | Neurology | ASAH2 | Q9NR71 | 10 | 50182778 | 50279720 | 1 | 5894.83 | 0.99 | 0.83 | 1.17 | 0.907 | 0.998 |
| ASGR1 | OID20990 | Neurology | ASGR1 | P07306 | 17 | 7173431 | 7179564 | 1 | 459.86 | 0.94 | 0.51 | 1.74 | 0.838 | 0.998 |
| ASPN | OID30315 | Cardiome | ASPN | Q9BXN1 | 9 | 92456205 | 92482506 | 1 | 1032.02 | 1.04 | 0.70 | 1.54 | 0.842 | 0.998 |
| ATP5IF1 | OID20760 | Inflammat | ATP5IF1 | Q9UII2 | 1 | 28236109 | 28246906 | 1 | 196.00 | 0.90 | 0.37 | 2.17 | 0.816 | 0.998 |
| ATRAID | OID31451 | Oncology | ATRAID | Q6UW56 | 2 | 27212041 | 27217178 | 1 | 1216.27 | 0.93 | 0.64 | 1.35 | 0.714 | 0.998 |
| ATRN | OID30735 | Inflammat | ATRN | O75882 | 20 | 3471018 | 3651118 | 1 | 6530.66 | 0.98 | 0.84 | 1.15 | 0.845 | 0.998 |
| AXL | OID20363 | Cardiome | AXL | P30530 | 19 | 41219223 | 41261766 | 1 | 473.06 | 0.93 | 0.53 | 1.62 | 0.796 | 0.998 |
| AZU1 | OID20309 | Cardiome | AZU1 | P20160 | 19 | 825097 | 832018 | 1 | 1061.67 | 1.03 | 0.71 | 1.48 | 0.894 | 0.998 |
| B4GALT1 | OID20780 | Inflammat | B4GALT1 | P15291 | 9 | 33104082 | 33167356 | 1 | 765.44 | 1.01 | 0.64 | 1.62 | 0.953 | 0.998 |
| BAIAP2 | OID21250 | Oncology | BAIAP2 | Q9UQB8 | 17 | 81035122 | 81117434 | 1 | 90.25 | 1.31 | 0.36 | 4.80 | 0.681 | 0.998 |
| BAP18 | OID31150 | Neurology | C17orf49 | Q8IXM2 | 17 | 7014495 | 7017525 | 1 | 64.00 | 1.04 | 0.21 | 5.05 | 0.965 | 0.998 |
| BIN2 | OID21067 | Neurology | BIN2 | Q9UBW5 | 12 | 51281038 | 51324668 | 1 | 235.11 | 1.06 | 0.45 | 2.49 | 0.885 | 0.998 |
| BLMH | OID20336 | Cardiome | BLMH | Q13867 | 17 | 30248203 | 30292056 | 1 | 5238.14 | 0.99 | 0.83 | 1.18 | 0.920 | 0.998 |
| BLNK | OID30541 | Inflammat | BLNK | Q8WV28 | 10 | 96189171 | 96271587 | 1 | 55.50 | 1.20 | 0.23 | 6.34 | 0.827 | 0.998 |
| BMPER | OID30615 | Inflammat | BMPER | Q8N8U9 | 7 | 33904308 | 34156427 | 1 | 535.59 | 0.95 | 0.56 | 1.63 | 0.862 | 0.998 |
| BPIFA2 | OID30279 | Cardiome | BPIFA2 | Q96DR5 | 20 | 33161768 | 33181412 | 1 | 339.34 | 0.94 | 0.46 | 1.93 | 0.872 | 0.998 |
| BRSK2 | OID31042 | Neurology | BRSK2 | Q8IWQ3 | 11 | 1389899 | 1462689 | 1 | 4533.78 | 1.02 | 0.85 | 1.21 | 0.853 | 0.998 |
| BST1 | OID21051 | Neurology | BST1 | Q10588 | 4 | 15703065 | 15738313 | 1 | 18397.22 | 1.01 | 0.92 | 1.10 | 0.901 | 0.998 |
| BST2 | OID21029 | Neurology | BST2 | Q10589 | 19 | 17402939 | 17405630 | 1 | 365.23 | 0.97 | 0.48 | 1.97 | 0.940 | 0.998 |
| BTN2A1 | OID20713 | Inflammat | BTN2A1 | Q7KYR7 | 6 | 26457904 | 26476621 | 1 | 1547.11 | 0.97 | 0.70 | 1.34 | 0.842 | 0.998 |
| BTN3A2 | OID20540 | Inflammat | BTN3A2 | P78410 | 6 | 26365159 | 26378320 | 1 | 7553.19 | 1.00 | 0.87 | 1.16 | 0.965 | 0.998 |
| C1QTNF1 | OID20367 | Cardiome | C1QTNF1 | Q9BXJ1 | 17 | 79022814 | 79049788 | 1 | 937.30 | 0.89 | 0.59 | 1.34 | 0.586 | 0.998 |
| C3 | OID30776 | Inflammat | C3 | P01024 | 19 | 6677704 | 6730562 | 1 | 230.49 | 0.86 | 0.38 | 1.94 | 0.718 | 0.998 |
| C7 | OID30766 | Inflammat | C7 | P10643 | 5 | 40909492 | 40984643 | 1 | 4241.97 | 0.99 | 0.82 | 1.20 | 0.918 | 0.998 |
| C7orf50 | OID31095 | Neurology | C7orf50 | Q9BRJ6 | 7 | 996986 | 1138260 | 1 | 1698.87 | 1.05 | 0.78 | 1.42 | 0.760 | 0.998 |
| C8B | OID30719 | Inflammat | C8B | P07358 | 1 | 56929207 | 56974383 | 1 | 4208.77 | 1.03 | 0.85 | 1.25 | 0.778 | 0.998 |
| CA1 | OID20409 | Cardiome | CA1 | P00915 | 8 | 85327608 | 85379014 | 1 | 161.65 | 0.83 | 0.28 | 2.45 | 0.734 | 0.998 |
| CA2 | OID21149 | Neurology | CA2 | P00918 | 8 | 85463968 | 85481493 | 1 | 212.33 | 1.04 | 0.41 | 2.62 | 0.929 | 0.998 |
| CA4 | OID20241 | Cardiome | CA4 | P22748 | 17 | 60149942 | 60170899 | 1 | 1701.56 | 0.99 | 0.73 | 1.36 | 0.964 | 0.998 |
| CACYBP | OID30649 | Inflammat | CACYBP | Q9HB71 | 1 | 174999163 | 175012027 | 1 | 210.25 | 1.02 | 0.43 | 2.39 | 0.967 | 0.998 |
| CALB2 | OID20801 | Neurology | CALB2 | P22676 | 16 | 71358713 | 71390436 | 1 | 74.39 | 0.91 | 0.20 | 4.14 | 0.904 | 0.998 |
| CALCA | OID20983 | Neurology | CALCA | P01258 | 11 | 14966668 | 14972351 | 1 | 289.00 | 0.89 | 0.35 | 2.26 | 0.812 | 0.998 |
| CALCOCO2 | OID30249 | Cardiome | CALCOC( | Q13137 | 17 | 48831018 | 48866522 | 1 | 56.25 | 1.13 | 0.21 | 6.24 | 0.887 | 0.998 |
| CANT1 | OID20181 | Cardiome | CANT1 | Q8WVQ1 | 17 | 78991716 | 79009867 | 1 | 321.01 | 1.01 | 0.51 | 2.00 | 0.967 | 0.998 |
| CAPG | OID21431 | Oncology | CAPG | P40121 | 2 | 85394753 | 85418432 | 1 | 11881.00 | 1.02 | 0.91 | 1.14 | 0.726 | 0.998 |
| CASP3 | OID20305 | Cardiome | CASP3 | P42574 | 4 | 184627696 | 184649509 | 1 | 256.00 | 0.83 | 0.36 | 1.90 | 0.656 | 0.998 |
| CASP8 | OID21414 | Oncology | CASP8 | Q14790 | 2 | 201233443 | 201287711 | 1 | 794.21 | 0.89 | 0.57 | 1.40 | 0.615 | 0.998 |
| CASP9 | OID30530 | Inflammat | CASP9 | P55211 | 1 | 15490832 | 15526534 | 1 | 102.52 | 1.22 | 0.38 | 3.88 | 0.738 | 0.998 |
| CBS | OID30200 | Cardiome | CBS | P35520 | 21 | 43053191 | 43076943 | 1 | 243.36 | 1.23 | 0.56 | 2.68 | 0.608 | 0.998 |
| CC2D1A | OID20958 | Neurology | CC2D1A | Q6P1N0 | 19 | 13906201 | 13930879 | 1 | 103.15 | 0.74 | 0.22 | 2.44 | 0.619 | 0.998 |
| CCDC80 | OID20226 | Cardiome | CCDC80 | Q76M96 | 3 | 112596797 | 112649530 | 1 | 100.00 | 1.34 | 0.33 | 5.42 | 0.680 | 0.998 |
| CCL11 | OID20668 | Inflammat | CCL11 | P51671 | 17 | 34285742 | 34288334 | 1 | 180.75 | 1.08 | 0.41 | 2.81 | 0.880 | 0.998 |
| CCL16 | OID20334 | Cardiome | CCL16 | O15467 | 17 | 35976493 | 35981497 | 1 | 12321.00 | 0.97 | 0.87 | 1.09 | 0.635 | 0.998 |
| CCL18 | OID20395 | Cardiome | CCL18 | P55774 | 17 | 36064272 | 36072032 | 1 | 4119.31 | 1.04 | 0.86 | 1.27 | 0.665 | 0.998 |
| CCL24 | OID20772 | Inflammat | CCL24 | O00175 | 7 | 75810825 | 75823356 | 1 | 13129.34 | 0.98 | 0.87 | 1.09 | 0.675 | 0.998 |
| CCL5 | OID20412 | Cardiome | CCL5 | P13501 | 17 | 35871491 | 35880793 | 1 | 484.00 | 0.91 | 0.52 | 1.58 | 0.736 | 0.998 |
| CCL8 | OID21466 | Oncology | CCL8 | P80075 | 17 | 34319435 | 34321402 | 1 | 11534.76 | 0.98 | 0.87 | 1.11 | 0.795 | 0.998 |
| CD163 | OID20360 | Cardiome | CD163 | Q86VB7 | 12 | 7470811 | 7503893 | 1 | 240.25 | 0.88 | 0.38 | 2.05 | 0.773 | 0.998 |
| CD164 | OID21080 | Neurology | CD164 | Q04900 | 6 | 109366514 | 109382467 | 1 | 346.89 | 0.92 | 0.47 | 1.79 | 0.801 | 0.998 |
| CD177 | OID21119 | Neurology | CD177 | Q8N6Q3 | 19 | 43353686 | 43363172 | 1 | 3802.78 | 1.05 | 0.87 | 1.28 | 0.605 | 0.998 |
| CD200R1 | OID20595 | Inflammat | CD200R1 | Q8TD46 | 3 | 112921205 | 112975103 | 1 | 8326.56 | 1.03 | 0.90 | 1.18 | 0.626 | 0.998 |
| CD209 | OID20262 | Cardiome | CD209 | Q9NNX6 | 19 | 7739993 | 7747564 | 1 | 2678.06 | 0.99 | 0.75 | 1.30 | 0.944 | 0.998 |
| CD22 | OID20637 | Inflammat | CD22 | P20273 | 19 | 35319261 | 35347361 | 1 | 1074.38 | 0.98 | 0.66 | 1.44 | 0.904 | 0.998 |
| CD276 | OID20680 | Inflammat | CD276 | Q5ZPR3 | 15 | 73683966 | 73714514 | 1 | 12413.67 | 1.03 | 0.91 | 1.16 | 0.620 | 0.998 |
| CD2AP | OID20177 | Cardiome | CD2AP | Q9Y5K6 | 6 | 47477789 | 47627263 | 1 | 1278.06 | 1.01 | 0.72 | 1.43 | 0.944 | 0.998 |
| CD300A | OID30576 | Inflammat | CD300A | Q9UGN4 | 17 | 74466399 | 74484794 | 1 | 1343.01 | 0.92 | 0.66 | 1.29 | 0.629 | 0.998 |
| CD300E | OID21418 | Oncology | CD300E | Q496F6 | 17 | 74609885 | 74623738 | 1 | 2036.27 | 1.05 | 0.79 | 1.38 | 0.751 | 0.998 |
| CD300LF | OID21469 | Oncology | CD300LF | Q8TDQ1 | 17 | 74694311 | 74712978 | 1 | 16770.25 | 1.02 | 0.93 | 1.12 | 0.633 | 0.998 |
| CD302 | OID21328 | Oncology | CD302 | Q8IX05 | 2 | 159768628 | 159798255 | 1 | 1450.92 | 0.90 | 0.64 | 1.26 | 0.548 | 0.998 |
| CD33 | OID21472 | Oncology | CD33 | P20138 | 19 | 51225064 | 51243860 | 1 | 21645.77 | 1.02 | 0.94 | 1.11 | 0.567 | 0.998 |
| CD36 | OID30485 | Inflammat | CD36 | P16671 | 7 | 80369575 | 80679277 | 1 | 1164.52 | 1.06 | 0.74 | 1.50 | 0.755 | 0.998 |
| CD38 | OID21316 | Oncology | CD38 | P28907 | 4 | 15778275 | 15853232 | 1 | 3651.61 | 1.01 | 0.77 | 1.33 | 0.924 | 0.998 |
| CD4 | OID20584 | Inflammat | CD4 | P01730 | 12 | 6786858 | 6820799 | 1 | 2013.77 | 0.99 | 0.75 | 1.32 | 0.969 | 0.998 |
| CD40 | OID20724 | Inflammat | CD40 | P25942 | 20 | 46118278 | 46129863 | 1 | 2219.46 | 1.07 | 0.83 | 1.39 | 0.596 | 0.998 |
| CD46 | OID20380 | Cardiome | CD46 | P15529 | 1 | 207752054 | 207795513 | 1 | 123.77 | 1.04 | 0.36 | 3.06 | 0.936 | 0.998 |
| CD5 | OID21449 | Oncology | CD5 | P06127 | 11 | 61102489 | 61127852 | 1 | 289.00 | 0.91 | 0.45 | 1.85 | 0.791 | 0.998 |
| CD69 | OID20206 | Cardiome | CD69 | Q07108 | 12 | 9752486 | 9760901 | 1 | 47.27 | 1.24 | 0.22 | 7.09 | 0.811 | 0.998 |
| CD72 | OID30641 | Inflammat | CD72 | P21854 | 9 | 35609982 | 35646810 | 1 | 340.20 | 1.06 | 0.53 | 2.11 | 0.875 | 0.998 |
| CD80 | OID30291 | Cardiome | CD80 | P33681 | 3 | 119524293 | 119559614 | 1 | 2541.84 | 1.05 | 0.81 | 1.35 | 0.711 | 0.998 |
| CDA | OID30345 | Cardiome | CDA | P32320 | 1 | 20589086 | 20618903 | 1 | 3736.27 | 1.04 | 0.85 | 1.27 | 0.719 | 0.998 |
| CDH15 | OID21005 | Neurology | CDH15 | P55291 | 16 | 89171748 | 89195492 | 1 | 2368.44 | 0.97 | 0.71 | 1.31 | 0.824 | 0.998 |
| CDH5 | OID20243 | Cardiome | CDH5 | P33151 | 16 | 66366622 | 66404784 | 1 | 784.00 | 1.05 | 0.64 | 1.72 | 0.857 | 0.998 |
| CDH6 | OID20157 | Cardiome | CDH6 | P55285 | 5 | 31193686 | 31329146 | 1 | 4064.06 | 0.94 | 0.78 | 1.14 | 0.545 | 0.998 |
| CDNF | OID21318 | Oncology | CDNF | Q49AH0 | 10 | 14819245 | 14838575 | 1 | 2890.34 | 1.08 | 0.84 | 1.38 | 0.553 | 0.998 |
| CDON | OID20754 | Inflammat | CDON | Q4KMG0 | 11 | 125955796 | 126063335 | 1 | 193.85 | 0.82 | 0.33 | 2.02 | 0.668 | 0.998 |
| CDSN | OID20667 | Inflammat | CDSN | Q15517 | 6 | 31115087 | 31120446 | 1 | 6480.25 | 1.03 | 0.89 | 1.20 | 0.671 | 0.998 |
| CEACAM6 | OID30362 | Cardiome | CEACAM( | P40199 | 19 | 41750977 | 41772211 | 1 | 225.00 | 0.92 | 0.39 | 2.19 | 0.856 | 0.998 |
| CEACAM8 | OID20102 | Cardiome | CEACAM | P31997 | 19 | 42580243 | 42595055 | 1 | 268.14 | 0.93 | 0.42 | 2.05 | 0.856 | 0.998 |
| CELA2A | OID30411 | Cardiome | CELA2A | P08217 | 1 | 15456728 | 15472091 | 1 | 233.65 | 0.91 | 0.40 | 2.07 | 0.815 | 0.998 |

| | | | | | | | | | | | | | | |
|---|---|---|---|---|---|---|---|---|---|---|---|---|---|---|
| CEP112 | OID30115 | Cardiome | CEP112 | Q8N8E3 | 17 | 65635537 | 66192133 | 1 | 819.39 | 0.92 | 0.61 | 1.40 | 0.698 | 0.998 |
| CEP20 | OID21209 | Oncology | CEP20 | Q96NB1 | 16 | 15865719 | 15888625 | 1 | 161.44 | 1.07 | 0.42 | 2.75 | 0.883 | 0.998 |
| CERT | OID20857 | Neurology | CERT1 | Q9Y5P4 | 5 | 75356345 | 75512138 | 1 | 52.05 | 0.96 | 0.17 | 5.33 | 0.962 | 0.998 |
| CES1 | OID20244 | Cardiome | CES1 | P23141 | 16 | 55802851 | 55833337 | 1 | 5184.00 | 0.99 | 0.82 | 1.20 | 0.935 | 0.998 |
| CES3 | OID21340 | Oncology | CES3 | Q6UWW8 | 16 | 66961245 | 66975149 | 1 | 669.71 | 0.91 | 0.56 | 1.49 | 0.707 | 0.998 |
| CETN3 | OID31525 | Oncology | CETN3 | O15182 | 5 | 90392257 | 90409766 | 1 | 467.64 | 1.14 | 0.62 | 2.10 | 0.677 | 0.998 |
| CFD | OID30738 | Inflammat | CFD | P00746 | 19 | 859453 | 867884 | 1 | 422.53 | 1.02 | 0.54 | 1.91 | 0.960 | 0.998 |
| CGB3_CGB5 | OID30671 | Inflammat | CGB5;CG | P0DN86;F | 19;19;19 | 49043848;4 | 49045311;4 | 1 | 75.94 | 1.57 | 0.33 | 7.49 | 0.572 | 0.998 |
| CGREF1 | OID20152 | Cardiome | CGREF1 | Q99674 | 2 | 27098889 | 27119128 | 1 | 4917.52 | 1.04 | 0.87 | 1.24 | 0.665 | 0.998 |
| CHCHD10 | OID31116 | Neurology | CHCHD1( | Q8WYQ3 | 22 | 23765834 | 23767972 | 1 | 115.07 | 1.14 | 0.31 | 4.18 | 0.838 | 0.998 |
| CHCHD6 | OID31426 | Oncology | CHCHD6 | Q9BRQ6 | 3 | 126704240 | 126960420 | 1 | 936.36 | 0.91 | 0.61 | 1.38 | 0.666 | 0.998 |
| CHI3L1 | OID20399 | Cardiome | CHI3L1 | P36222 | 1 | 203178931 | 203186704 | 1 | 5575.11 | 0.96 | 0.81 | 1.14 | 0.612 | 0.998 |
| CHMP6 | OID30327 | Cardiome | CHMP6 | Q96FZ7 | 17 | 80991598 | 81009517 | 1 | 412.09 | 1.08 | 0.59 | 1.99 | 0.810 | 0.998 |
| CHRDL2 | OID20223 | Cardiome | CHRDL2 | Q6WN34 | 11 | 74696429 | 74731426 | 1 | 1373.11 | 1.06 | 0.74 | 1.53 | 0.756 | 0.998 |
| CIT | OID30970 | Neurology | CIT | O14578 | 12 | 119685791 | 119877320 | 1 | 49.48 | 0.95 | 0.15 | 5.90 | 0.953 | 0.998 |
| CKMT1A_CKI | OID20721 | Inflammat | CKMT1B; | P12532;P | 15;15 | 43593054;4 | 43604901;4 | 1 | 432.05 | 1.04 | 0.57 | 1.92 | 0.888 | 0.998 |
| CLEC10A | OID20928 | Neurology | CLEC10A | Q8IUN9 | 17 | 7074537 | 7080307 | 1 | 4624.00 | 1.03 | 0.85 | 1.23 | 0.782 | 0.998 |
| CLEC1A | OID20099 | Cardiome | CLEC1A | Q8NC01 | 12 | 10069554 | 10111627 | 1 | 1570.14 | 0.92 | 0.67 | 1.25 | 0.581 | 0.998 |
| CLEC3B | OID30756 | Inflammat | CLEC3B | P05452 | 3 | 45001548 | 45036071 | 1 | 885.06 | 0.92 | 0.59 | 1.43 | 0.713 | 0.998 |
| CLEC4A | OID20573 | Inflammat | CLEC4A | Q9UMR7 | 12 | 8123617 | 8138607 | 1 | 3929.72 | 1.03 | 0.84 | 1.26 | 0.796 | 0.998 |
| CLEC4C | OID20547 | Inflammat | CLEC4C | Q8WTT0 | 12 | 7729383 | 7751605 | 1 | 11342.25 | 1.00 | 0.90 | 1.12 | 0.949 | 0.998 |
| CLEC4M | OID30405 | Cardiome | CLEC4M | Q9H2X3 | 19 | 7763210 | 7769605 | 1 | 2070.25 | 0.99 | 0.73 | 1.35 | 0.944 | 0.998 |
| CLEC6A | OID21228 | Oncology | CLEC6A | Q6EIG7 | 12 | 8455962 | 8478330 | 1 | 5104.31 | 1.04 | 0.87 | 1.25 | 0.670 | 0.998 |
| CLEC7A | OID20636 | Inflammat | CLEC7A | Q9BXN2 | 12 | 10116777 | 10130258 | 1 | 14436.95 | 0.98 | 0.88 | 1.09 | 0.756 | 0.998 |
| CLMP | OID21419 | Oncology | CLMP | Q9H6B4 | 11 | 123069872 | 123195248 | 1 | 2304.00 | 1.08 | 0.82 | 1.44 | 0.581 | 0.998 |
| CLU | OID30732 | Inflammat | CLU | P10909 | 8 | 27596917 | 27614700 | 1 | 138.06 | 1.12 | 0.41 | 3.04 | 0.831 | 0.998 |
| CLUL1 | OID20198 | Cardiome | CLUL1 | Q15846 | 18 | 596988 | 650334 | 1 | 5890.56 | 0.99 | 0.83 | 1.17 | 0.870 | 0.998 |
| CMC1 | OID30360 | Cardiome | CMC1 | Q7Z7K0 | 3 | 28241584 | 28325142 | 1 | 85.29 | 0.83 | 0.23 | 3.03 | 0.775 | 0.998 |
| CNTN1 | OID20307 | Cardiome | CNTN1 | Q12860 | 12 | 40692439 | 41072415 | 1 | 808.18 | 1.07 | 0.66 | 1.75 | 0.780 | 0.998 |
| CNTN2 | OID21426 | Oncology | CNTN2 | Q02246 | 1 | 205042937 | 205078289 | 1 | 11410.12 | 1.01 | 0.89 | 1.14 | 0.906 | 0.998 |
| CNTNAP2 | OID20556 | Inflammat | CNTNAP2 | Q9UHC6 | 7 | 146116002 | 148420998 | 1 | 4795.56 | 1.02 | 0.85 | 1.23 | 0.800 | 0.998 |
| COL15A1 | OID30396 | Cardiome | COL15A1 | P39059 | 9 | 98943179 | 99070792 | 1 | 558.14 | 1.08 | 0.65 | 1.82 | 0.763 | 0.998 |
| COL18A1 | OID20332 | Cardiome | COL18A1 | P39060 | 21 | 45405165 | 45513720 | 1 | 479.12 | 0.85 | 0.46 | 1.58 | 0.608 | 0.998 |
| COL2A1 | OID30286 | Cardiome | COL2A1 | P02458 | 12 | 47972967 | 48004554 | 1 | 8649.00 | 0.99 | 0.86 | 1.12 | 0.833 | 0.998 |
| COMMD1 | OID30222 | Cardiome | COMMD1 | Q8N668 | 2 | 61888724 | 62147247 | 1 | 503.25 | 1.14 | 0.66 | 1.98 | 0.631 | 0.998 |
| COMP | OID20405 | Cardiome | COMP | P49747 | 19 | 18782773 | 18791305 | 1 | 692.65 | 1.04 | 0.64 | 1.69 | 0.885 | 0.998 |
| COMT | OID20086 | Cardiome | COMT | P21964 | 22 | 19941371 | 19969975 | 1 | 3094.14 | 0.95 | 0.77 | 1.17 | 0.616 | 0.998 |
| CPA1 | OID20280 | Cardiome | CPA1 | P15085 | 7 | 130380339 | 130388114 | 1 | 541.62 | 1.16 | 0.68 | 1.99 | 0.586 | 0.998 |
| CPA4 | OID30606 | Inflammat | CPA4 | Q9UI42 | 7 | 130293134 | 130324180 | 1 | 4472.27 | 0.96 | 0.80 | 1.15 | 0.660 | 0.998 |
| CPPED1 | OID21053 | Neurology | CPPED1 | Q9BRF8 | 16 | 12659799 | 12803887 | 1 | 6855.84 | 1.03 | 0.89 | 1.20 | 0.654 | 0.998 |
| CPVL | OID21435 | Oncology | CPVL | Q9H3G5 | 7 | 28995235 | 29195451 | 1 | 8752.64 | 1.02 | 0.89 | 1.18 | 0.760 | 0.998 |
| CPXM1 | OID21479 | Oncology | CPXM1 | Q96SM3 | 20 | 2794074 | 2800627 | 1 | 1626.78 | 0.93 | 0.69 | 1.26 | 0.645 | 0.998 |
| CR2 | OID20393 | Cardiome | CR2 | P20023 | 1 | 207454230 | 207489895 | 1 | 661.89 | 0.96 | 0.57 | 1.62 | 0.883 | 0.998 |
| CRACR2A | OID21249 | Oncology | CRACR2A | Q9BSW2 | 12 | 3606633 | 3764819 | 1 | 280.56 | 1.22 | 0.58 | 2.56 | 0.607 | 0.998 |
| CRADD | OID20923 | Neurology | CRADD | P78560 | 12 | 93677375 | 93894840 | 1 | 100.00 | 1.18 | 0.32 | 4.37 | 0.808 | 0.998 |
| CREG1 | OID21515 | Oncology | CREG1 | O75629 | 1 | 167529117 | 167553805 | 1 | 645.16 | 1.02 | 0.62 | 1.70 | 0.924 | 0.998 |
| CRELD1 | OID30619 | Inflammat | CRELD1 | Q96HD1 | 3 | 9933793 | 9945413 | 1 | 8545.98 | 1.03 | 0.90 | 1.17 | 0.664 | 0.998 |
| CRELD2 | OID20751 | Inflammat | CRELD2 | Q6UXH1 | 22 | 49918167 | 49927540 | 1 | 1652.42 | 1.02 | 0.74 | 1.42 | 0.897 | 0.998 |
| CRH | OID21257 | Oncology | CRH | P06850 | 8 | 66176376 | 66178464 | 1 | 1051.61 | 0.91 | 0.62 | 1.32 | 0.614 | 0.998 |
| CRHBP | OID20747 | Inflammat | CRHBP | P24387 | 5 | 76953045 | 76981158 | 1 | 5402.25 | 0.99 | 0.84 | 1.17 | 0.886 | 0.998 |
| CRISP3 | OID30715 | Inflammat | CRISP3 | P54108 | 6 | 49727376 | 49744437 | 1 | 6653.90 | 1.01 | 0.86 | 1.19 | 0.875 | 0.998 |
| CRNN | OID21427 | Oncology | CRNN | Q9UBG3 | 1 | 152409243 | 152414263 | 1 | 7188.62 | 1.03 | 0.88 | 1.21 | 0.681 | 0.998 |
| CRTAC1 | OID20279 | Cardiome | CRTAC1 | Q9NQ79 | 10 | 97865000 | 98030828 | 1 | 9457.56 | 0.99 | 0.86 | 1.15 | 0.942 | 0.998 |
| CRTAM | OID20914 | Neurology | CRTAM | O95727 | 11 | 122838500 | 122872643 | 1 | 1530.77 | 1.08 | 0.78 | 1.48 | 0.651 | 0.998 |
| CRYBB1 | OID30319 | Cardiome | CRYBB1 | P53674 | 22 | 26599278 | 26618027 | 1 | 503.04 | 0.83 | 0.45 | 1.52 | 0.549 | 0.998 |
| CRYM | OID30815 | Neurology | CRYM | Q14894 | 16 | 21238874 | 21303083 | 1 | 2126.23 | 1.03 | 0.80 | 1.34 | 0.799 | 0.998 |
| CRYZL1 | OID30272 | Cardiome | CRYZL1 | O95825 | 21 | 33589341 | 33643926 | 1 | 310.43 | 0.81 | 0.41 | 1.61 | 0.548 | 0.998 |
| CSF3R | OID30527 | Inflammat | CSF3R | Q99062 | 1 | 36466043 | 36483278 | 1 | 887.41 | 1.03 | 0.69 | 1.52 | 0.900 | 0.998 |
| CST5 | OID20995 | Neurology | CST5 | P28325 | 20 | 23875934 | 23879748 | 1 | 10905.33 | 0.97 | 0.85 | 1.11 | 0.670 | 0.998 |
| CST6 | OID20293 | Cardiome | CST6 | Q15828 | 11 | 66012008 | 66013505 | 1 | 652.38 | 1.04 | 0.63 | 1.70 | 0.887 | 0.998 |
| CST7 | OID20704 | Inflammat | CST7 | O76096 | 20 | 24949269 | 24959928 | 1 | 8628.35 | 1.02 | 0.89 | 1.17 | 0.776 | 0.998 |
| CSTB | OID20294 | Cardiome | CSTB | P04080 | 21 | 43772511 | 43776330 | 1 | 10302.25 | 0.98 | 0.86 | 1.11 | 0.735 | 0.998 |
| CTF1 | OID20061 | Cardiome | CTF1 | Q16619 | 16 | 30896614 | 30903547 | 1 | 45.56 | 1.67 | 0.27 | 10.24 | 0.580 | 0.998 |
| CTRC | OID20752 | Inflammat | CTRC | Q99895 | 1 | 15438442 | 15449242 | 1 | 2248.34 | 0.99 | 0.75 | 1.32 | 0.960 | 0.998 |
| CTSD | OID20358 | Cardiome | CTSD | P07339 | 11 | 1752752 | 1764573 | 1 | 1785.06 | 1.03 | 0.76 | 1.40 | 0.850 | 0.998 |
| CTSE | OID30628 | Inflammat | CTSE | P14091 | 1 | 206008535 | 206023909 | 1 | 4134.49 | 0.97 | 0.81 | 1.17 | 0.767 | 0.998 |
| CTSF | OID21500 | Oncology | CTSF | Q9UBX1 | 11 | 66563464 | 66568879 | 1 | 735.77 | 0.95 | 0.57 | 1.59 | 0.856 | 0.998 |
| CTSV | OID21405 | Oncology | CTSV | O60911 | 9 | 97029677 | 97156556 | 1 | 815.42 | 1.14 | 0.72 | 1.79 | 0.577 | 0.998 |
| CWC15 | OID31479 | Oncology | CWC15 | Q9P013 | 11 | 94962620 | 94973586 | 1 | 52.56 | 0.77 | 0.13 | 4.48 | 0.774 | 0.998 |
| CXCL1 | OID20762 | Inflammat | CXCL1 | P09341 | 4 | 73869393 | 73871308 | 1 | 5608.35 | 0.96 | 0.82 | 1.14 | 0.659 | 0.998 |
| CXCL11 | OID21042 | Neurology | CXCL11 | O14625 | 4 | 76033682 | 76041415 | 1 | 1024.00 | 1.04 | 0.68 | 1.58 | 0.865 | 0.998 |
| CXCL12 | OID20464 | Inflammat | CXCL12 | P48061 | 10 | 44370165 | 44386493 | 1 | 594.14 | 1.09 | 0.66 | 1.82 | 0.732 | 0.998 |
| CXCL5 | OID20207 | Cardiome | CXCL5 | P42830 | 4 | 73995642 | 73998677 | 1 | 2857.39 | 1.05 | 0.83 | 1.35 | 0.670 | 0.998 |
| CXCL6 | OID20613 | Inflammat | CXCL6 | P80162 | 4 | 73836640 | 73849064 | 1 | 4900.00 | 0.98 | 0.82 | 1.16 | 0.805 | 0.998 |
| CXCL8 | OID20153 | Cardiome | CXCL8 | P10145 | 4 | 73740541 | 73743716 | 1 | 112.11 | 1.13 | 0.36 | 3.59 | 0.836 | 0.998 |
| CYB5R2 | OID31512 | Oncology | CYB5R2 | Q6BCY4 | 11 | 7665100 | 7677222 | 1 | 1029.34 | 1.03 | 0.71 | 1.51 | 0.869 | 0.998 |
| DAG1 | OID20774 | Inflammat | DAG1 | Q14118 | 3 | 49468703 | 49535618 | 1 | 56.25 | 1.65 | 0.30 | 9.12 | 0.564 | 0.998 |
| DAPP1 | OID20524 | Inflammat | DAPP1 | Q9UN19 | 4 | 99816827 | 99870190 | 1 | 156.25 | 0.95 | 0.37 | 2.45 | 0.922 | 0.998 |
| DBNL | OID20681 | Inflammat | DBNL | Q9UJU6 | 7 | 44044640 | 44069456 | 1 | 52.56 | 1.60 | 0.29 | 8.91 | 0.590 | 0.998 |
| DCTPP1 | OID20148 | Cardiome | DCTPP1 | Q9H773 | 16 | 30423615 | 30430030 | 1 | 220.73 | 1.30 | 0.53 | 3.20 | 0.566 | 0.998 |
| DDI2 | OID30602 | Inflammat | DDI2 | Q5TDH0 | 1 | 15617458 | 15669044 | 1 | 56.89 | 1.87 | 0.25 | 13.75 | 0.541 | 0.998 |
| DDR1 | OID21009 | Neurology | DDR1 | Q08345 | 6 | 30876421 | 30900156 | 1 | 2457.33 | 1.06 | 0.80 | 1.39 | 0.698 | 0.998 |
| DGCR6 | OID30859 | Neurology | DGCR6 | Q14129 | 22 | 18906028 | 18914238 | 1 | 268.14 | 0.82 | 0.39 | 1.73 | 0.595 | 0.998 |
| DHRS4L2 | OID31072 | Neurology | DHRS4L2 | Q6PKH6 | 14 | 23969874 | 24006408 | 1 | 1466.44 | 1.10 | 0.78 | 1.55 | 0.595 | 0.998 |
| DKK3 | OID20264 | Cardiome | DKK3 | Q9UBP4 | 11 | 11956207 | 12009769 | 1 | 2162.25 | 1.07 | 0.81 | 1.40 | 0.652 | 0.998 |
| DKKL1 | OID21313 | Oncology | DKKL1 | Q9UK85 | 19 | 49361783 | 49375116 | 1 | 19600.00 | 1.02 | 0.94 | 1.11 | 0.640 | 0.998 |
| DLL1 | OID21525 | Oncology | DLL1 | O00548 | 6 | 170282206 | 170306565 | 1 | 503.04 | 0.95 | 0.52 | 1.75 | 0.867 | 0.998 |
| DNAJA4 | OID31130 | Neurology | DNAJA4 | Q8WW22 | 15 | 78264086 | 78282196 | 1 | 304.31 | 0.82 | 0.37 | 1.83 | 0.636 | 0.998 |
| DNAJB1 | OID21363 | Oncology | DNAJB1 | P25685 | 19 | 14514769 | 14560391 | 1 | 50.57 | 1.21 | 0.21 | 6.95 | 0.831 | 0.998 |
| DNAJB6 | OID30613 | Inflammat | DNAJB6 | O75190 | 7 | 157335381 | 157417439 | 1 | 570.02 | 0.91 | 0.55 | 1.51 | 0.726 | 0.998 |
| DNAJC21 | OID31104 | Neurology | DNAJC21 | Q5F1R6 | 5 | 34929559 | 34958964 | 1 | 178.89 | 0.77 | 0.32 | 1.88 | 0.569 | 0.998 |
| DNAJC6 | OID30247 | Cardiome | DNAJC6 | O75061 | 1 | 65248219 | 65415871 | 1 | 97.52 | 0.72 | 0.21 | 2.39 | 0.586 | 0.998 |
| DNER | OID20712 | Inflammat | DNER | Q8NFT8 | 2 | 229357629 | 229714555 | 1 | 1225.00 | 1.10 | 0.77 | 1.58 | 0.600 | 0.998 |
| DNMBP | OID20956 | Neurology | DNMBP | Q6XZF7 | 10 | 99875577 | 100009947 | 1 | 227.15 | 0.89 | 0.39 | 2.04 | 0.784 | 0.998 |
| DPEP1 | OID20897 | Neurology | DPEP1 | P16444 | 16 | 89613308 | 89638456 | 1 | 9975.02 | 1.00 | 0.89 | 1.13 | 0.938 | 0.998 |
| DPT | OID20278 | Cardiome | DPT | Q07507 | 1 | 168695468 | 168729206 | 1 | 2222.45 | 1.05 | 0.78 | 1.40 | 0.761 | 0.998 |
| DRAXIN | OID20917 | Neurology | DRAXIN | Q8NBI3 | 1 | 11691710 | 11725857 | 1 | 1429.79 | 0.93 | 0.66 | 1.31 | 0.685 | 0.998 |
| DSC2 | OID21079 | Neurology | DSC2 | Q02487 | 18 | 31058840 | 31102522 | 1 | 812.25 | 1.10 | 0.72 | 1.68 | 0.644 | 0.998 |
| DTYMK | OID30308 | Cardiome | DTYMK | P23919 | 2 | 241675747 | 241686944 | 1 | 52.75 | 0.68 | 0.13 | 3.68 | 0.657 | 0.998 |
| DXO | OID31080 | Neurology | DXO | O77932 | 6 | 31969810 | 31972290 | 1 | 2441.26 | 0.95 | 0.73 | 1.23 | 0.689 | 0.998 |
| EBAG9 | OID20918 | Neurology | EBAG9 | O00559 | 8 | 109539711 | 109565996 | 1 | 74.39 | 1.29 | 0.32 | 5.24 | 0.723 | 0.998 |
| ECHS1 | OID30239 | Cardiome | ECHS1 | P30084 | 10 | 133362485 | 133373354 | 1 | 691.38 | 1.06 | 0.68 | 1.67 | 0.791 | 0.998 |
| ECM1 | OID30736 | Inflammat | ECM1 | Q16610 | 1 | 150508062 | 150513789 | 1 | 7161.39 | 0.98 | 0.85 | 1.13 | 0.775 | 0.998 |
| EDIL3 | OID20081 | Cardiome | EDIL3 | O43854 | 5 | 83940554 | 84384880 | 1 | 123.46 | 0.91 | 0.30 | 2.80 | 0.869 | 0.998 |
| EFCAB2 | OID31313 | Oncology | EFCAB2 | Q5VUJ9 | 1 | 244969682 | 245127164 | 1 | 2304.00 | 0.97 | 0.76 | 1.25 | 0.833 | 0.998 |
| EGF | OID20698 | Inflammat | EGF | P01133 | 4 | 109912883 | 110013766 | 1 | 280.56 | 0.88 | 0.43 | 1.80 | 0.733 | 0.998 |
| EGFL7 | OID21252 | Oncology | EGFL7 | Q9UHF1 | 9 | 136658856 | 136672678 | 1 | 414.41 | 1.02 | 0.55 | 1.89 | 0.948 | 0.998 |

| | | | | | | | | | | | | | | |
|---|---|---|---|---|---|---|---|---|---|---|---|---|---|---|
| EGFLAM | OID31496 | Oncology | EGFLAM | Q63HQ2 | 5 | 38258409 | 38465480 | 1 | 4007.44 | 1.06 | 0.87 | 1.29 | 0.576 | 0.998 |
| EGLN1 | OID20572 | Inflammat | EGLN1 | Q9GZT9 | 1 | 231363751 | 231422287 | 1 | 283.92 | 1.10 | 0.52 | 2.32 | 0.796 | 0.998 |
| EIF4G3 | OID31022 | Neurology | EIF4G3 | O43432 | 1 | 20806292 | 21177285 | 1 | 105.80 | 1.48 | 0.40 | 5.57 | 0.559 | 0.998 |
| ENO2 | OID21046 | Neurology | ENO2 | P09104 | 12 | 6913745 | 6923698 | 1 | 328.52 | 1.02 | 0.50 | 2.10 | 0.948 | 0.998 |
| ENO3 | OID30209 | Cardiome | ENO3 | P13929 | 17 | 4948092 | 4957131 | 1 | 126.56 | 1.27 | 0.42 | 3.86 | 0.678 | 0.998 |
| ENPP6 | OID30273 | Cardiome | ENPP6 | Q6UWR7 | 4 | 184088706 | 184221230 | 1 | 1576.75 | 0.99 | 0.72 | 1.37 | 0.963 | 0.998 |
| ENPP7 | OID20689 | Inflammat | ENPP7 | Q6UWV6 | 17 | 79730943 | 79742219 | 1 | 14101.56 | 1.00 | 0.90 | 1.10 | 0.950 | 0.998 |
| ENTPD6 | OID20100 | Cardiome | ENTPD6 | O75354 | 20 | 25195693 | 25228075 | 1 | 7878.68 | 0.98 | 0.85 | 1.13 | 0.769 | 0.998 |
| EPHA2 | OID21342 | Oncology | EPHA2 | P29317 | 1 | 16124337 | 16156069 | 1 | 361.00 | 0.89 | 0.43 | 1.83 | 0.747 | 0.998 |
| EPHA4 | OID30622 | Inflammat | EPHA4 | P54764 | 2 | 221418027 | 221574202 | 1 | 394.31 | 1.20 | 0.61 | 2.38 | 0.595 | 0.998 |
| EPHB6 | OID20982 | Neurology | EPHB6 | O15197 | 7 | 142855061 | 142871094 | 1 | 1248.44 | 0.90 | 0.64 | 1.26 | 0.542 | 0.998 |
| ERBB2 | OID21520 | Oncology | ERBB2 | P04626 | 17 | 39687914 | 39730426 | 1 | 400.00 | 0.89 | 0.46 | 1.75 | 0.738 | 0.998 |
| ERC2 | OID30880 | Neurology | ERC2 | O15083 | 3 | 55508311 | 56469095 | 1 | 46.24 | 1.27 | 0.23 | 7.09 | 0.781 | 0.998 |
| EREG | OID20905 | Neurology | EREG | O14944 | 4 | 74365145 | 74388749 | 1 | 138.06 | 1.04 | 0.36 | 3.04 | 0.943 | 0.998 |
| ERN1 | OID31391 | Oncology | ERN1 | O75460 | 17 | 64039080 | 64130819 | 1 | 571.64 | 0.95 | 0.56 | 1.63 | 0.863 | 0.998 |
| EXTL1 | OID30114 | Cardiome | EXTL1 | Q92935 | 1 | 26019884 | 26036464 | 1 | 2652.25 | 1.03 | 0.81 | 1.33 | 0.790 | 0.998 |
| F11R | OID21151 | Neurology | F11R | Q9Y624 | 1 | 160995211 | 161021343 | 1 | 62.02 | 1.14 | 0.22 | 5.89 | 0.879 | 0.998 |
| F12 | OID30739 | Inflammat | F12 | P00748 | 5 | 177402133 | 177416583 | 1 | 17248.44 | 0.99 | 0.90 | 1.09 | 0.846 | 0.998 |
| F2 | OID30723 | Inflammat | F2 | P00734 | 11 | 46719196 | 46739506 | 1 | 390.33 | 1.18 | 0.66 | 2.12 | 0.578 | 0.998 |
| F7 | OID20372 | Cardiome | F7 | P08709 | 13 | 113105788 | 113120685 | 1 | 4214.17 | 1.03 | 0.85 | 1.24 | 0.773 | 0.998 |
| FABP1 | OID20778 | Inflammat | FABP1 | P07148 | 2 | 88122982 | 88128062 | 1 | 877.64 | 0.88 | 0.57 | 1.35 | 0.554 | 0.998 |
| FABP2 | OID20200 | Cardiome | FABP2 | P12104 | 4 | 119317250 | 119322138 | 1 | 708.89 | 1.01 | 0.63 | 1.62 | 0.966 | 0.998 |
| FABP4 | OID20245 | Cardiome | FABP4 | P15090 | 8 | 81478419 | 81483236 | 1 | 171.90 | 1.17 | 0.39 | 3.48 | 0.780 | 0.998 |
| FABP9 | OID20501 | Inflammat | FABP9 | Q0Z7S8 | 8 | 81458253 | 81461579 | 1 | 138.99 | 1.23 | 0.39 | 3.82 | 0.726 | 0.998 |
| FAM13A | OID31463 | Oncology | FAM13A | O94988 | 4 | 88725955 | 89111398 | 1 | 278.89 | 0.93 | 0.44 | 1.97 | 0.853 | 0.998 |
| FAM20A | OID30348 | Cardiome | FAM20A | Q96MK3 | 17 | 68535113 | 68601367 | 1 | 892.52 | 0.90 | 0.60 | 1.37 | 0.632 | 0.998 |
| FAM3B | OID21372 | Oncology | FAM3B | P58499 | 21 | 41304212 | 41357727 | 1 | 4600.03 | 1.01 | 0.83 | 1.23 | 0.915 | 0.998 |
| FAM3D | OID31514 | Oncology | FAM3D | Q96BQ1 | 3 | 58633946 | 58666834 | 1 | 2000.53 | 1.07 | 0.76 | 1.50 | 0.705 | 0.998 |
| FAP | OID20330 | Cardiome | FAP | Q12884 | 2 | 162170684 | 162245151 | 1 | 844.14 | 1.06 | 0.67 | 1.68 | 0.805 | 0.998 |
| FAS | OID20316 | Cardiome | FAS | P25445 | 10 | 88953813 | 89029605 | 1 | 1347.94 | 1.03 | 0.71 | 1.50 | 0.857 | 0.998 |
| FBN2 | OID30974 | Neurology | FBN2 | P35556 | 5 | 128257909 | 128659185 | 1 | 1515.43 | 1.08 | 0.79 | 1.46 | 0.631 | 0.998 |
| FBP1 | OID20227 | Cardiome | FBP1 | P09467 | 9 | 94603133 | 94640249 | 1 | 1686.86 | 0.93 | 0.69 | 1.27 | 0.655 | 0.998 |
| FCAR | OID20564 | Inflammat | FCAR | P24071 | 19 | 54874235 | 54891420 | 1 | 8794.27 | 1.03 | 0.90 | 1.18 | 0.661 | 0.998 |
| FCN1 | OID30690 | Inflammat | FCN1 | O00602 | 9 | 134903232 | 134917912 | 1 | 3052.56 | 0.98 | 0.77 | 1.26 | 0.901 | 0.998 |
| FCN2 | OID20382 | Cardiome | FCN2 | Q15485 | 9 | 134880810 | 134887523 | 1 | 7844.90 | 1.00 | 0.86 | 1.17 | 0.961 | 0.998 |
| FCRL1 | OID20172 | Cardiome | FCRL1 | Q96LA6 | 1 | 157794403 | 157820120 | 1 | 2615.59 | 0.98 | 0.75 | 1.28 | 0.890 | 0.998 |
| FCRL2 | OID20639 | Inflammat | FCRL2 | Q96LA5 | 1 | 157745733 | 157777132 | 1 | 5882.89 | 0.96 | 0.82 | 1.14 | 0.654 | 0.998 |
| FES | OID21207 | Oncology | FES | P07332 | 15 | 90883695 | 90895776 | 1 | 276.39 | 0.94 | 0.46 | 1.92 | 0.871 | 0.998 |
| FGF21 | OID21402 | Oncology | FGF21 | Q9NSA1 | 19 | 48755524 | 48758333 | 1 | 236.39 | 1.02 | 0.47 | 2.24 | 0.957 | 0.998 |
| FGFBP2 | OID30395 | Cardiome | FGFBP2 | Q9BYJ0 | 4 | 15960245 | 15969309 | 1 | 5169.61 | 0.98 | 0.81 | 1.18 | 0.815 | 0.998 |
| FGFBP3 | OID31108 | Neurology | FGFBP3 | Q8TAT2 | 10 | 91906584 | 91909486 | 1 | 2376.56 | 0.93 | 0.72 | 1.21 | 0.585 | 0.998 |
| FGFR2 | OID21478 | Oncology | FGFR2 | P21802 | 10 | 121478332 | 121598458 | 1 | 260.59 | 0.93 | 0.40 | 2.16 | 0.861 | 0.998 |
| FGFR4 | OID30646 | Inflammat | FGFR4 | P22455 | 5 | 177086905 | 177098144 | 1 | 15907.52 | 1.02 | 0.92 | 1.13 | 0.735 | 0.998 |
| FGL1 | OID30702 | Inflammat | FGL1 | Q08830 | 8 | 17864380 | 17910365 | 1 | 6744.52 | 1.04 | 0.89 | 1.22 | 0.606 | 0.998 |
| FKBP4 | OID20960 | Neurology | FKBP4 | Q02790 | 12 | 2794970 | 2805423 | 1 | 44.89 | 1.42 | 0.22 | 9.14 | 0.709 | 0.998 |
| FLT3 | OID21272 | Oncology | FLT3 | P36888 | 13 | 28003274 | 28100592 | 1 | 110.25 | 0.70 | 0.22 | 2.25 | 0.554 | 0.998 |
| FNDC1 | OID31127 | Neurology | FNDC1 | Q4ZHG4 | 6 | 159169400 | 159272108 | 1 | 2862.25 | 1.04 | 0.81 | 1.34 | 0.750 | 0.998 |
| FOLR1 | OID21529 | Oncology | FOLR1 | P15328 | 11 | 72189558 | 72196323 | 1 | 167.15 | 1.11 | 0.39 | 3.18 | 0.840 | 0.998 |
| FOXJ3 | OID30445 | Inflammat | FOXJ3 | Q9UPW0 | 1 | 42176539 | 42335877 | 1 | 87.68 | 1.38 | 0.36 | 5.27 | 0.633 | 0.998 |
| FSHB | OID30233 | Cardiome | FSHB | P01225 | 11 | 30231014 | 30235261 | 1 | 441.00 | 1.08 | 0.43 | 2.71 | 0.866 | 0.998 |
| FST | OID20770 | Inflammat | FST | P19883 | 5 | 53480626 | 53487134 | 1 | 315.06 | 1.05 | 0.53 | 2.08 | 0.891 | 0.998 |
| FUCA1 | OID20261 | Cardiome | FUCA1 | P04066 | 1 | 23845077 | 23868290 | 1 | 5836.96 | 1.08 | 0.83 | 1.40 | 0.562 | 0.998 |
| FUT3_FUT5 | OID21013 | Neurology | FUT3;FU | P21217;Q | 19;19 | 5842888;586 | 5851471;587 | 1 | 5069.44 | 0.98 | 0.81 | 1.18 | 0.824 | 0.998 |
| FXYD5 | OID20463 | Inflammat | FXYD5 | Q96DB9 | 19 | 35154730 | 35169881 | 1 | 1782.72 | 1.08 | 0.80 | 1.47 | 0.598 | 0.998 |
| GALNT10 | OID21371 | Oncology | GALNT10 | Q86SR1 | 5 | 154190730 | 154420984 | 1 | 2304.00 | 1.04 | 0.81 | 1.34 | 0.761 | 0.998 |
| GALNT2 | OID21423 | Oncology | GALNT2 | Q10471 | 1 | 230057990 | 230282122 | 1 | 1752.02 | 0.90 | 0.65 | 1.25 | 0.536 | 0.998 |
| GALNT3 | OID20471 | Inflammat | GALNT3 | Q14435 | 2 | 165747588 | 165794659 | 1 | 2364.39 | 1.03 | 0.79 | 1.34 | 0.828 | 0.998 |
| GATD3 | OID30253 | Cardiome | GATD3 | P0DPI2 | 21 | 44133610 | 44210114 | 1 | 153.76 | 1.09 | 0.41 | 2.90 | 0.864 | 0.998 |
| GC | OID30780 | Inflammat | GC | P02774 | 4 | 71741696 | 71804041 | 1 | 22575.06 | 0.99 | 0.91 | 1.07 | 0.756 | 0.998 |
| GCLM | OID30059 | Cardiome | GCLM | P48507 | 1 | 93885199 | 93909456 | 1 | 56.25 | 0.80 | 0.17 | 3.85 | 0.782 | 0.998 |
| GCNT1 | OID21379 | Oncology | GCNT1 | Q02742 | 9 | 76419850 | 76651203 | 1 | 3037.23 | 0.99 | 0.79 | 1.24 | 0.904 | 0.998 |
| GFAP | OID21247 | Oncology | GFAP | P14136 | 17 | 44903159 | 44916937 | 1 | 134.17 | 0.82 | 0.25 | 2.66 | 0.739 | 0.998 |
| GFER | OID21259 | Oncology | GFER | P55789 | 16 | 1984193 | 1987749 | 1 | 89.39 | 0.97 | 0.24 | 3.90 | 0.969 | 0.998 |
| GFRA1 | OID21457 | Oncology | GFRA1 | P56159 | 10 | 116056925 | 116276803 | 1 | 918.09 | 0.98 | 0.63 | 1.52 | 0.929 | 0.998 |
| GFRA2 | OID21330 | Oncology | GFRA2 | O00451 | 8 | 21690398 | 21812357 | 1 | 1491.89 | 1.06 | 0.78 | 1.45 | 0.702 | 0.998 |
| GFRA3 | OID20933 | Neurology | GFRA3 | O60609 | 5 | 138252380 | 138274621 | 1 | 132.25 | 1.43 | 0.45 | 4.58 | 0.549 | 0.998 |
| GGACT | OID30290 | Cardiome | GGACT | Q9BVM4 | 13 | 100530164 | 100589528 | 1 | 3094.14 | 1.01 | 0.82 | 1.25 | 0.905 | 0.998 |
| GGT5 | OID20909 | Neurology | GGT5 | P36269 | 22 | 24219654 | 24245108 | 1 | 1336.31 | 0.99 | 0.70 | 1.41 | 0.966 | 0.998 |
| GIMAP7 | OID30639 | Inflammat | GIMAP7 | Q8NHV1 | 7 | 150514872 | 150521073 | 1 | 4548.75 | 0.99 | 0.82 | 1.19 | 0.874 | 0.998 |
| GIMAP8 | OID31194 | Oncology | GIMAP8 | Q8ND71 | 7 | 150450630 | 150479393 | 1 | 735.18 | 0.88 | 0.57 | 1.37 | 0.576 | 0.998 |
| GIPC3 | OID31507 | Oncology | GIPC3 | Q8TF64 | 19 | 3585478 | 3593541 | 1 | 1308.03 | 0.98 | 0.69 | 1.38 | 0.893 | 0.998 |
| GIT1 | OID30533 | Inflammat | GIT1 | Q9Y2X7 | 17 | 29573475 | 29594054 | 1 | 82.34 | 0.85 | 0.22 | 3.34 | 0.815 | 0.998 |
| GLO1 | OID20107 | Cardiome | GLO1 | Q04760 | 6 | 38675925 | 38703145 | 1 | 1711.89 | 0.98 | 0.74 | 1.30 | 0.891 | 0.998 |
| GLRX | OID20195 | Cardiome | GLRX | P35754 | 5 | 95751319 | 95822726 | 1 | 4686.02 | 1.02 | 0.84 | 1.23 | 0.849 | 0.998 |
| GLT8D2 | OID20825 | Neurology | GLT8D2 | Q9H1C3 | 12 | 103988987 | 104064183 | 1 | 506.25 | 1.10 | 0.67 | 1.82 | 0.708 | 0.998 |
| GM2A | OID30413 | Cardiome | GM2A | P17900 | 5 | 151212150 | 151270440 | 1 | 7547.27 | 0.98 | 0.85 | 1.13 | 0.769 | 0.998 |
| GMFG | OID30340 | Cardiome | GMFG | O60234 | 19 | 39328353 | 39342372 | 1 | 75.11 | 1.30 | 0.30 | 5.57 | 0.727 | 0.998 |
| GMPR | OID20641 | Inflammat | GMPR | P36959 | 6 | 16238587 | 16295549 | 1 | 992.25 | 0.90 | 0.61 | 1.32 | 0.585 | 0.998 |
| GORASP2 | OID31438 | Oncology | GORASP | Q9H8Y8 | 2 | 170928464 | 170967130 | 1 | 52.56 | 1.07 | 0.18 | 6.43 | 0.941 | 0.998 |
| GP1BA | OID20381 | Cardiome | GP1BA | P07359 | 17 | 4932277 | 4935023 | 1 | 226.77 | 1.21 | 0.49 | 2.95 | 0.677 | 0.998 |
| GP2 | OID20090 | Cardiome | GP2 | P55259 | 16 | 20309574 | 20327808 | 1 | 1107.94 | 0.94 | 0.62 | 1.41 | 0.761 | 0.998 |
| GPC1 | OID21438 | Oncology | GPC1 | P35052 | 2 | 240435663 | 240468076 | 1 | 2116.00 | 1.09 | 0.81 | 1.45 | 0.577 | 0.998 |
| GPHA2 | OID31437 | Oncology | GPHA2 | Q96T91 | 11 | 64934471 | 64935893 | 1 | 768.52 | 0.87 | 0.55 | 1.39 | 0.571 | 0.998 |
| GPIHBP1 | OID31469 | Oncology | GPIHBP1 | Q8IV16 | 8 | 143213218 | 143217170 | 1 | 1156.00 | 1.03 | 0.72 | 1.47 | 0.874 | 0.998 |
| GPNMB | OID20173 | Cardiome | GPNMB | Q14956 | 7 | 23235967 | 23275108 | 1 | 1849.00 | 0.99 | 0.75 | 1.31 | 0.942 | 0.998 |
| GPR15L | OID31511 | Oncology | C10orf99 | Q6UWK7 | 10 | 84173801 | 84185294 | 1 | 763.14 | 0.92 | 0.59 | 1.46 | 0.733 | 0.998 |
| GPRC5C | OID31340 | Oncology | GPRC5C | Q9NQ84 | 17 | 74424851 | 74451653 | 1 | 231.47 | 0.84 | 0.36 | 1.97 | 0.694 | 0.998 |
| GRP | OID30264 | Cardiome | GRP | P07492 | 18 | 59220158 | 59230774 | 1 | 2134.44 | 0.96 | 0.72 | 1.26 | 0.752 | 0.998 |
| GZMH | OID20096 | Cardiome | GZMH | P20718 | 14 | 24606480 | 24609699 | 1 | 587.73 | 1.20 | 0.65 | 2.20 | 0.566 | 0.998 |
| HAVCR1 | OID21422 | Oncology | HAVCR1 | Q96D42 | 5 | 157029413 | 157059119 | 1 | 4624.00 | 1.03 | 0.85 | 1.26 | 0.738 | 0.998 |
| HBZ | OID30307 | Cardiome | HBZ | P02008 | 16 | 142728 | 154503 | 1 | 16608.77 | 1.00 | 0.91 | 1.10 | 0.969 | 0.998 |
| HCG22 | OID30088 | Cardiome | HCG22 | E2RYF7 | 6 | 31053450 | 31059890 | 1 | 2462.64 | 0.98 | 0.76 | 1.26 | 0.853 | 0.998 |
| HEBP1 | OID20063 | Cardiome | HEBP1 | Q9NRV9 | 12 | 12974870 | 13000265 | 1 | 256.00 | 1.21 | 0.54 | 2.68 | 0.643 | 0.998 |
| HEG1 | OID30579 | Inflammat | HEG1 | Q9ULI3 | 3 | 124965710 | 125055997 | 1 | 242.47 | 0.93 | 0.38 | 2.29 | 0.877 | 0.998 |
| HIP1R | OID31141 | Neurology | HIP1R | O75146 | 12 | 122834453 | 122862961 | 1 | 141.02 | 0.98 | 0.35 | 2.73 | 0.965 | 0.998 |
| HLA-E | OID20532 | Inflammat | HLA-E | P13747 | 6 | 30489509 | 30494194 | 1 | 1610.02 | 0.94 | 0.70 | 1.28 | 0.705 | 0.998 |
| HMOX2 | OID20887 | Neurology | HMOX2 | P30519 | 16 | 4474690 | 4510347 | 1 | 180.75 | 0.89 | 0.36 | 2.20 | 0.805 | 0.998 |
| HPSE | OID30409 | Cardiome | HPSE | Q9Y251 | 4 | 83292461 | 83335153 | 1 | 405.02 | 1.22 | 0.65 | 2.31 | 0.535 | 0.998 |
| HS3ST3B1 | OID21303 | Oncology | HS3ST3B | Q9Y662 | 17 | 14301081 | 14349404 | 1 | 1691.27 | 0.97 | 0.72 | 1.31 | 0.861 | 0.998 |
| HSD17B14 | OID31098 | Neurology | HSD17B1 | Q9BPX1 | 19 | 48813018 | 48836510 | 1 | 7159.04 | 0.96 | 0.83 | 1.11 | 0.600 | 0.998 |
| HSPA2 | OID31424 | Oncology | HSPA2 | P54652 | 14 | 64535905 | 64546173 | 1 | 199.12 | 1.26 | 0.49 | 3.23 | 0.637 | 0.998 |
| HYOU1 | OID20300 | Cardiome | HYOU1 | Q9Y4L1 | 11 | 119044188 | 119057227 | 1 | 523.27 | 0.86 | 0.51 | 1.46 | 0.581 | 0.998 |
| ICAM3 | OID20277 | Cardiome | ICAM3 | P32942 | 19 | 10333776 | 10339661 | 1 | 1580.06 | 0.92 | 0.67 | 1.27 | 0.609 | 0.998 |
| ICAM5 | OID20145 | Cardiome | ICAM5 | Q9UMF0 | 19 | 10289952 | 10296778 | 1 | 8794.27 | 0.97 | 0.85 | 1.11 | 0.705 | 0.998 |
| IDO1 | OID30230 | Cardiome | IDO1 | P14902 | 8 | 39902275 | 39928790 | 1 | 1190.25 | 0.90 | 0.62 | 1.30 | 0.568 | 0.998 |
| IGF1R | OID21274 | Oncology | IGF1R | P08069 | 15 | 98648539 | 98964530 | 1 | 208.64 | 1.05 | 0.43 | 2.58 | 0.909 | 0.998 |
| IGFBP2 | OID20325 | Cardiome | IGFBP2 | P18065 | 2 | 216632828 | 216664436 | 1 | 60.49 | 1.17 | 0.24 | 5.62 | 0.844 | 0.998 |

| | | | | | | | | | | | | | | |
|---|---|---|---|---|---|---|---|---|---|---|---|---|---|---|
| IGFBP6 | OID20413 | Cardiome | IGFBP6 | P24592 | 12 | 53097436 | 53102345 | 1 | 158.76 | 1.43 | 0.44 | 4.69 | 0.551 | 0.998 |
| IGFBP7 | OID20236 | Cardiome | IGFBP7 | Q16270 | 4 | 57030773 | 57110385 | 1 | 2162.25 | 1.06 | 0.79 | 1.41 | 0.699 | 0.998 |
| IGHMBP2 | OID30123 | Cardiome | IGHMBP2 | P38935 | 11 | 68903863 | 68940602 | 1 | 97.52 | 1.13 | 0.34 | 3.79 | 0.847 | 0.998 |
| IGSF3 | OID21312 | Oncology | IGSF3 | O75054 | 1 | 116574399 | 116667755 | 1 | 149.38 | 1.04 | 0.37 | 2.94 | 0.942 | 0.998 |
| IL10 | OID20431 | Inflammat | IL10 | P22301 | 1 | 206767602 | 206774541 | 1 | 164.31 | 1.23 | 0.48 | 3.15 | 0.663 | 0.998 |
| IL12A_IL12B | OID21327 | Oncology | IL12A;IL1: | P29459;P | 3;5 | 159988835;1 | 159996019;1 | 1 | 4607.02 | 1.05 | 0.87 | 1.26 | 0.627 | 0.998 |
| IL12B | OID20666 | Inflammat | IL12B | P29460 | 5 | 159314780 | 159330487 | 1 | 4900.00 | 1.05 | 0.87 | 1.25 | 0.627 | 0.998 |
| IL12RB1 | OID20486 | Inflammat | IL12RB1 | P42701 | 19 | 18058995 | 18098944 | 1 | 2567.11 | 0.98 | 0.76 | 1.27 | 0.896 | 0.998 |
| IL17D | OID20481 | Inflammat | IL17D | Q8TAD2 | 13 | 20702127 | 20723098 | 1 | 2475.06 | 0.98 | 0.75 | 1.29 | 0.910 | 0.998 |
| IL17F | OID20456 | Inflammat | IL17F | Q96PD4 | 6 | 52236681 | 52244500 | 1 | 455.11 | 0.88 | 0.49 | 1.60 | 0.679 | 0.998 |
| IL18 | OID20694 | Inflammat | IL18 | Q14116 | 11 | 112143253 | 112164096 | 1 | 1776.02 | 0.95 | 0.69 | 1.31 | 0.740 | 0.998 |
| IL1R2 | OID20757 | Inflammat | IL1R2 | P27930 | 2 | 101991960 | 102028544 | 1 | 4176.39 | 1.04 | 0.86 | 1.25 | 0.680 | 0.998 |
| IL1RL1 | OID20250 | Cardiome | IL1RL1 | Q01638 | 2 | 102311502 | 102352037 | 1 | 9688.18 | 1.01 | 0.88 | 1.16 | 0.924 | 0.998 |
| IL1RL2 | OID20487 | Inflammat | IL1RL2 | Q9HB29 | 2 | 102187006 | 102240002 | 1 | 3897.33 | 0.96 | 0.77 | 1.19 | 0.712 | 0.998 |
| IL20RB | OID30461 | Inflammat | IL20RB | Q6UXL0 | 3 | 136946230 | 137011085 | 1 | 213.89 | 0.81 | 0.35 | 1.86 | 0.619 | 0.998 |
| IL22 | OID31407 | Oncology | IL22 | Q9GZX6 | 12 | 68248242 | 68253604 | 1 | 175.56 | 0.96 | 0.35 | 2.62 | 0.937 | 0.998 |
| IL22RA1 | OID20449 | Inflammat | IL22RA1 | Q8N6P7 | 1 | 24119771 | 24143140 | 1 | 1011.24 | 1.07 | 0.73 | 1.56 | 0.742 | 0.998 |
| IL31RA | OID30475 | Inflammat | IL31RA | Q8NI17 | 5 | 55851357 | 55922854 | 1 | 6221.27 | 0.99 | 0.84 | 1.17 | 0.943 | 0.998 |
| IL32 | OID20605 | Inflammat | IL32 | P24001 | 16 | 3065297 | 3082192 | 1 | 923.22 | 1.06 | 0.70 | 1.59 | 0.784 | 0.998 |
| IL36A | OID30472 | Inflammat | IL36A | Q9UHA7 | 2 | 113005459 | 113008044 | 1 | 62.02 | 0.70 | 0.13 | 3.67 | 0.674 | 0.998 |
| IL36G | OID30551 | Inflammat | IL36G | Q9NZH8 | 2 | 112973203 | 112985658 | 1 | 114.49 | 1.16 | 0.35 | 3.82 | 0.804 | 0.998 |
| IL4R | OID20576 | Inflammat | IL4R | P24394 | 16 | 27313668 | 27364778 | 1 | 1540.56 | 1.09 | 0.80 | 1.48 | 0.587 | 0.998 |
| IMMT | OID31397 | Oncology | IMMT | Q16891 | 2 | 86143932 | 86195472 | 1 | 274.61 | 1.10 | 0.49 | 2.49 | 0.817 | 0.998 |
| IMPA1 | OID20907 | Neurology | IMPA1 | P29218 | 8 | 81656914 | 81686331 | 1 | 702.25 | 0.88 | 0.54 | 1.43 | 0.596 | 0.998 |
| INHBB | OID30625 | Inflammat | INHBB | P09529 | 2 | 120346136 | 120351803 | 1 | 2718.88 | 0.96 | 0.73 | 1.24 | 0.737 | 0.998 |
| INHBC | OID21093 | Neurology | INHBC | P55103 | 12 | 57434784 | 57452062 | 1 | 5361.49 | 0.96 | 0.81 | 1.13 | 0.620 | 0.998 |
| INPP5D | OID30259 | Cardiome | INPP5D | Q92835 | 2 | 233059967 | 233207903 | 1 | 554.86 | 1.02 | 0.58 | 1.80 | 0.949 | 0.998 |
| INSL5 | OID31448 | Oncology | INSL5 | Q9Y5Q6 | 1 | 66797740 | 66801276 | 1 | 661.22 | 1.18 | 0.69 | 2.02 | 0.538 | 0.998 |
| IPCEF1 | OID20977 | Neurology | IPCEF1 | Q8WWN9 | 6 | 154154496 | 154356803 | 1 | 110.25 | 0.81 | 0.24 | 2.69 | 0.727 | 0.998 |
| IQGAP2 | OID21361 | Oncology | IQGAP2 | Q13576 | 5 | 76403285 | 76708132 | 1 | 328.52 | 1.09 | 0.56 | 2.13 | 0.804 | 0.998 |
| ISM1 | OID20538 | Inflammat | ISM1 | B1AKI9 | 20 | 13221274 | 13300651 | 1 | 546.39 | 0.84 | 0.49 | 1.46 | 0.540 | 0.998 |
| ITGA2 | OID30570 | Inflammat | ITGA2 | P17301 | 5 | 52989340 | 53094779 | 1 | 6006.25 | 1.01 | 0.85 | 1.19 | 0.915 | 0.998 |
| ITGAM | OID21071 | Neurology | ITGAM | P11215 | 16 | 31259967 | 31332892 | 1 | 420.25 | 0.97 | 0.52 | 1.79 | 0.918 | 0.998 |
| ITGAV | OID21416 | Oncology | ITGAV | P06756 | 2 | 186590056 | 186680901 | 1 | 433.40 | 1.02 | 0.52 | 1.98 | 0.962 | 0.998 |
| ITGBL1 | OID30408 | Cardiome | ITGBL1 | O95965 | 13 | 101452593 | 101720856 | 1 | 1626.78 | 1.03 | 0.74 | 1.45 | 0.853 | 0.998 |
| JAM2 | OID21064 | Neurology | JAM2 | P57087 | 21 | 25639258 | 25717562 | 1 | 90.81 | 1.03 | 0.27 | 4.03 | 0.961 | 0.998 |
| KAZALD1 | OID21339 | Oncology | KAZALD1 | Q96I82 | 10 | 101061989 | 101068131 | 1 | 7967.91 | 0.99 | 0.85 | 1.15 | 0.887 | 0.998 |
| KEL | OID20881 | Neurology | KEL | P23276 | 7 | 142941114 | 142962363 | 1 | 1518.56 | 1.12 | 0.76 | 1.67 | 0.565 | 0.998 |
| KHK | OID30241 | Cardiome | KHK | P50053 | 2 | 27086747 | 27100762 | 1 | 2862.25 | 0.96 | 0.76 | 1.21 | 0.733 | 0.998 |
| KIR2DL3 | OID21196 | Oncology | KIR2DL3 | P43628 | 19 | 54738513 | 54753052 | 1 | 2384.69 | 1.01 | 0.76 | 1.34 | 0.927 | 0.998 |
| KIR2DS4 | OID31432 | Oncology | KIR2DS4 | P43632 | 19 | 54832676 | 54848569 | 1 | 21674.38 | 1.02 | 0.94 | 1.11 | 0.651 | 0.998 |
| KIR3DL1 | OID21185 | Oncology | KIR3DL1 | P43629 | 19 | 54816468 | 54830778 | 1 | 13398.06 | 1.01 | 0.91 | 1.12 | 0.907 | 0.998 |
| KIRREL2 | OID20873 | Neurology | KIRREL2 | Q6UWL6 | 19 | 35855861 | 35867136 | 1 | 477.73 | 1.11 | 0.63 | 1.94 | 0.724 | 0.998 |
| KLK12 | OID21201 | Oncology | KLK12 | Q9UKR0 | 19 | 51029092 | 51035230 | 1 | 21279.52 | 0.99 | 0.91 | 1.08 | 0.819 | 0.998 |
| KLK13 | OID21291 | Oncology | KLK13 | Q9UKR3 | 19 | 51055626 | 51065114 | 1 | 3220.56 | 1.06 | 0.85 | 1.32 | 0.598 | 0.998 |
| KLK15 | OID31427 | Oncology | KLK15 | Q9H2R5 | 19 | 50825289 | 50837213 | 1 | 8898.78 | 0.99 | 0.88 | 1.13 | 0.928 | 0.998 |
| KLK8 | OID21517 | Oncology | KLK8 | O60259 | 19 | 50996007 | 51002711 | 1 | 3409.26 | 1.01 | 0.82 | 1.26 | 0.898 | 0.998 |
| KLKB1 | OID30760 | Inflammat | KLKB1 | P03952 | 4 | 186226438 | 186258471 | 1 | 2193.36 | 1.04 | 0.78 | 1.37 | 0.801 | 0.998 |
| KRT19 | OID20596 | Inflammat | KRT19 | P08727 | 17 | 41523617 | 41528308 | 1 | 337.64 | 1.10 | 0.57 | 2.14 | 0.775 | 0.998 |
| KRT5 | OID20860 | Neurology | KRT5 | P13647 | 12 | 52514575 | 52520530 | 1 | 208.18 | 0.83 | 0.32 | 2.17 | 0.707 | 0.998 |
| KYAT1 | OID20232 | Cardiome | KYAT1 | Q16773 | 9 | 128832942 | 128882494 | 1 | 301.89 | 0.97 | 0.45 | 2.07 | 0.935 | 0.998 |
| LAG3 | OID21315 | Oncology | LAG3 | P18627 | 12 | 6772512 | 6778455 | 1 | 478.52 | 1.09 | 0.62 | 1.91 | 0.757 | 0.998 |
| LAIR1 | OID20737 | Inflammat | LAIR1 | Q6GTX8 | 19 | 54351384 | 54370558 | 1 | 5227.29 | 1.04 | 0.88 | 1.23 | 0.647 | 0.998 |
| LAIR2 | OID20965 | Neurology | LAIR2 | Q6ISS4 | 19 | 54497879 | 54510687 | 1 | 7802.78 | 1.01 | 0.88 | 1.16 | 0.889 | 0.998 |
| LAMP1 | OID30416 | Cardiome | LAMP1 | P11279 | 13 | 113297239 | 113323672 | 1 | 55.42 | 1.11 | 0.22 | 5.51 | 0.896 | 0.998 |
| LAMP3 | OID20638 | Inflammat | LAMP3 | Q9UQV4 | 3 | 183122215 | 183163839 | 1 | 848.27 | 1.07 | 0.70 | 1.64 | 0.755 | 0.998 |
| LAYN | OID20970 | Neurology | LAYN | Q6UX15 | 11 | 111540280 | 111561745 | 1 | 3088.18 | 1.05 | 0.82 | 1.35 | 0.676 | 0.998 |
| LBP | OID20355 | Cardiome | LBP | P18428 | 20 | 38346482 | 38377013 | 1 | 7200.73 | 0.98 | 0.85 | 1.13 | 0.798 | 0.998 |
| LCP1 | OID30367 | Cardiome | LCP1 | P13796 | 13 | 46125920 | 46211871 | 1 | 12838.63 | 1.02 | 0.91 | 1.13 | 0.778 | 0.998 |
| LDLRAP1 | OID31068 | Neurology | LDLRAP1 | Q5SW96 | 1 | 25543606 | 25568886 | 1 | 80.10 | 1.45 | 0.36 | 5.83 | 0.597 | 0.998 |
| LECT2 | OID30320 | Cardiome | LECT2 | O14960 | 5 | 135922279 | 135954983 | 1 | 13061.22 | 0.96 | 0.85 | 1.09 | 0.552 | 0.998 |
| LEFTY2 | OID21454 | Oncology | LEFTY2 | O00292 | 1 | 225936598 | 225941383 | 1 | 10374.88 | 1.01 | 0.88 | 1.16 | 0.855 | 0.998 |
| LGALS7_LGA | OID21406 | Oncology | LGALS7B | P47929;P | 19;19 | 38789200;38 | 38791754;38 | 1 | 1033.16 | 0.96 | 0.63 | 1.48 | 0.866 | 0.998 |
| LHB | OID21248 | Oncology | LHB | P01229 | 19 | 49015980 | 49017091 | 1 | 56.25 | 0.75 | 0.14 | 4.04 | 0.740 | 0.998 |
| LHPP | OID20749 | Inflammat | LHPP | Q9H008 | 10 | 124461823 | 124617888 | 1 | 2635.11 | 1.04 | 0.80 | 1.36 | 0.751 | 0.998 |
| LILRA2 | OID21152 | Neurology | LILRA2 | Q8N149 | 19 | 54572920 | 54590287 | 1 | 6350.86 | 0.98 | 0.84 | 1.15 | 0.811 | 0.998 |
| LILRB2 | OID20301 | Cardiome | LILRB2 | Q8N423 | 19 | 54273812 | 54281184 | 1 | 16641.00 | 1.01 | 0.91 | 1.12 | 0.901 | 0.998 |
| LILRB4 | OID20429 | Inflammat | LILRB4 | Q8NHJ6 | 19 | 54643889 | 54670359 | 1 | 1451.61 | 1.07 | 0.74 | 1.57 | 0.713 | 0.998 |
| LILRB5 | OID20324 | Cardiome | LILRB5 | O75023 | 19 | 54249421 | 54257301 | 1 | 21941.02 | 1.01 | 0.93 | 1.10 | 0.801 | 0.998 |
| LMOD1 | OID30212 | Cardiome | LMOD1 | P29536 | 1 | 201896456 | 201946588 | 1 | 816.33 | 1.13 | 0.68 | 1.87 | 0.645 | 0.998 |
| LPL | OID20188 | Cardiome | LPL | P06858 | 8 | 19901717 | 19967259 | 1 | 3499.91 | 0.94 | 0.74 | 1.20 | 0.628 | 0.998 |
| LRIG3 | OID30450 | Inflammat | LRIG3 | Q6UXM1 | 12 | 58872149 | 58920504 | 1 | 100.00 | 0.78 | 0.22 | 2.78 | 0.700 | 0.998 |
| LRRC25 | OID21333 | Oncology | LRRC25 | Q8N386 | 19 | 18391137 | 18397622 | 1 | 2345.33 | 1.08 | 0.81 | 1.43 | 0.616 | 0.998 |
| LRRC38 | OID30924 | Neurology | LRRC38 | Q5VT99 | 1 | 13474973 | 13514003 | 1 | 2013.77 | 0.97 | 0.73 | 1.30 | 0.858 | 0.998 |
| LRRFIP1 | OID31344 | Oncology | LRRFIP1 | Q32MZ4 | 2 | 237627587 | 237813682 | 1 | 62.23 | 0.60 | 0.12 | 3.04 | 0.541 | 0.998 |
| LRTM2 | OID31020 | Neurology | LRTM2 | Q8N967 | 12 | 1820267 | 1836753 | 1 | 379.22 | 0.89 | 0.47 | 1.67 | 0.716 | 0.998 |
| LTA | OID20586 | Inflammat | LTA | P01374 | 6 | 31572054 | 31574324 | 1 | 9906.88 | 0.96 | 0.85 | 1.10 | 0.562 | 0.998 |
| LTA4H | OID21452 | Oncology | LTA4H | P09960 | 12 | 96000753 | 96043520 | 1 | 162.56 | 0.81 | 0.30 | 2.15 | 0.671 | 0.998 |
| LTB | OID31383 | Oncology | LTB | Q06643 | 6 | 31580525 | 31582522 | 1 | 435.77 | 1.09 | 0.60 | 2.00 | 0.772 | 0.998 |
| LTBP2 | OID20254 | Cardiome | LTBP2 | Q14767 | 14 | 74498183 | 74612378 | 1 | 306.25 | 0.84 | 0.33 | 2.10 | 0.702 | 0.998 |
| LUZP2 | OID31402 | Oncology | LUZP2 | Q86TE4 | 11 | 24496970 | 25082638 | 1 | 792.02 | 1.05 | 0.67 | 1.67 | 0.822 | 0.998 |
| LXN | OID21003 | Neurology | LXN | Q9BS40 | 3 | 158645822 | 158672648 | 1 | 435.77 | 1.06 | 0.60 | 1.87 | 0.841 | 0.998 |
| LY6D | OID20708 | Inflammat | LY6D | Q14210 | 8 | 142784882 | 142786539 | 1 | 3066.39 | 0.97 | 0.78 | 1.20 | 0.755 | 0.998 |
| LY9 | OID20670 | Inflammat | LY9 | Q9HBG7 | 1 | 160796074 | 160828261 | 1 | 4444.44 | 0.94 | 0.78 | 1.15 | 0.561 | 0.998 |
| LYVE1 | OID30728 | Inflammat | LYVE1 | Q9Y5Y7 | 11 | 10556966 | 10611689 | 1 | 339.34 | 1.20 | 0.60 | 2.41 | 0.609 | 0.998 |
| M6PR | OID30208 | Cardiome | M6PR | P20645 | 12 | 8940361 | 8949761 | 1 | 463.91 | 1.14 | 0.65 | 1.99 | 0.642 | 0.998 |
| MAMDC2 | OID30309 | Cardiome | MAMDC2 | Q7Z304 | 9 | 70043848 | 70226972 | 1 | 784.00 | 1.12 | 0.69 | 1.80 | 0.646 | 0.998 |
| MAMDC4 | OID31342 | Oncology | MAMDC4 | Q6UXC1 | 9 | 136850943 | 136860799 | 1 | 1031.12 | 1.10 | 0.73 | 1.65 | 0.663 | 0.998 |
| MAN2B2 | OID30306 | Cardiome | MAN2B2 | Q9Y2E5 | 4 | 6575189 | 6623362 | 1 | 13856.65 | 0.99 | 0.88 | 1.11 | 0.890 | 0.998 |
| MANF | OID20707 | Inflammat | MANF | P55145 | 3 | 51385291 | 51389397 | 1 | 79.37 | 0.89 | 0.22 | 3.58 | 0.865 | 0.998 |
| MANSC4 | OID30141 | Cardiome | MANSC4 | A6NHS7 | 12 | 27762427 | 27780236 | 1 | 10862.27 | 0.97 | 0.86 | 1.09 | 0.582 | 0.998 |
| MAP2 | OID31138 | Neurology | MAP2 | P11137 | 2 | 209424047 | 209734147 | 1 | 196.90 | 1.07 | 0.43 | 2.64 | 0.890 | 0.998 |
| MAP4K5 | OID20899 | Neurology | MAP4K5 | Q9Y4K4 | 14 | 50418501 | 50561126 | 1 | 543.47 | 1.12 | 0.64 | 1.96 | 0.702 | 0.998 |
| MAPK9 | OID20557 | Inflammat | MAPK9 | P45984 | 5 | 180233143 | 180292099 | 1 | 252.46 | 1.27 | 0.55 | 2.89 | 0.575 | 0.998 |
| MARCO | OID20210 | Cardiome | MARCO | Q9UEW3 | 2 | 118942194 | 118994660 | 1 | 1156.00 | 1.08 | 0.74 | 1.57 | 0.700 | 0.998 |
| MATN3 | OID20979 | Neurology | MATN3 | O15232 | 2 | 19992052 | 20012668 | 1 | 4970.25 | 0.96 | 0.80 | 1.14 | 0.614 | 0.998 |
| MAVS | OID21204 | Oncology | MAVS | Q7Z434 | 20 | 3846799 | 3876123 | 1 | 150.62 | 0.93 | 0.33 | 2.64 | 0.892 | 0.998 |
| MAX | OID20877 | Neurology | MAX | P61244 | 14 | 65006174 | 65102695 | 1 | 95.06 | 1.39 | 0.41 | 4.74 | 0.599 | 0.998 |
| MBL2 | OID30759 | Inflammat | MBL2 | P11226 | 10 | 52765380 | 52772784 | 1 | 13081.64 | 0.99 | 0.89 | 1.11 | 0.882 | 0.998 |
| MCEE | OID31033 | Neurology | MCEE | Q96PE7 | 2 | 71109684 | 71130239 | 1 | 552.25 | 1.04 | 0.63 | 1.72 | 0.872 | 0.998 |
| MCFD2 | OID20171 | Cardiome | MCFD2 | Q8NI22 | 2 | 46901870 | 46941855 | 1 | 333.06 | 0.83 | 0.41 | 1.70 | 0.618 | 0.998 |
| MDGA1 | OID20951 | Neurology | MDGA1 | Q8NFP4 | 6 | 37630679 | 37699306 | 1 | 23804.08 | 1.00 | 0.92 | 1.10 | 0.928 | 0.998 |
| MDM1 | OID30945 | Neurology | MDM1 | Q8TC05 | 12 | 68272443 | 68332381 | 1 | 637.56 | 1.15 | 0.69 | 1.92 | 0.590 | 0.998 |
| MEGF9 | OID20285 | Cardiome | MEGF9 | Q9H1U4 | 9 | 120600811 | 120714470 | 1 | 1816.89 | 1.06 | 0.78 | 1.44 | 0.708 | 0.998 |
| MELTF | OID30347 | Cardiome | MELTF | P08582 | 3 | 196980590 | 197029835 | 1 | 2601.00 | 1.06 | 0.84 | 1.34 | 0.613 | 0.998 |
| MEPE | OID20753 | Inflammat | MEPE | Q9NQ76 | 4 | 87821398 | 87846814 | 1 | 331.24 | 1.19 | 0.56 | 2.53 | 0.652 | 0.998 |
| MERTK | OID20657 | Inflammat | MERTK | Q12866 | 2 | 111898607 | 112029561 | 1 | 1444.00 | 0.95 | 0.66 | 1.37 | 0.782 | 0.998 |

| | | | | | | | | | | | | | | |
|---|---|---|---|---|---|---|---|---|---|---|---|---|---|---|
| MFGE8 | OID21134 | Neurology | MFGE8 | Q08431 | 15 | 88898683 | 88913381 | 1 | 2173.89 | 1.03 | 0.77 | 1.36 | 0.863 | 0.998 |
| MICB_MICA | OID20593 | Inflammat | MICB;MIC | Q29980;Q | 6;6 | 31494881;31 | 31511124;31 | 1 | 14992.64 | 0.99 | 0.90 | 1.09 | 0.856 | 0.998 |
| MILR1 | OID20482 | Inflammat | MILR1 | Q7Z6M3 | 17 | 64449037 | 64468643 | 1 | 2475.06 | 1.01 | 0.79 | 1.30 | 0.923 | 0.998 |
| MINDY1 | OID31399 | Oncology | MINDY1 | Q8N5J2 | 1 | 150996549 | 151008376 | 1 | 321.95 | 1.16 | 0.61 | 2.19 | 0.656 | 0.998 |
| MITD1 | OID20959 | Neurology | MITD1 | Q8WV92 | 2 | 99161427 | 99181058 | 1 | 87.89 | 0.90 | 0.24 | 3.41 | 0.882 | 0.998 |
| MME | OID21307 | Oncology | MME | P08473 | 3 | 155024124 | 155183704 | 1 | 67.75 | 0.87 | 0.17 | 4.56 | 0.871 | 0.998 |
| MMP8 | OID21019 | Neurology | MMP8 | P22894 | 11 | 102711796 | 102727050 | 1 | 2721.36 | 0.95 | 0.75 | 1.20 | 0.676 | 0.998 |
| MOCS2 | OID30638 | Inflammat | MOCS2 | O96007 | 5 | 53095679 | 53110063 | 1 | 3150.02 | 0.97 | 0.78 | 1.22 | 0.821 | 0.998 |
| MOG | OID21384 | Oncology | MOG | Q16653 | 6 | 29657002 | 29672372 | 1 | 703.81 | 0.98 | 0.60 | 1.59 | 0.924 | 0.998 |
| MPO | OID21100 | Neurology | MPO | P05164 | 17 | 58269855 | 58280935 | 1 | 889.44 | 0.94 | 0.63 | 1.41 | 0.760 | 0.998 |
| MRC1 | OID30694 | Inflammat | MRC1 | P22897 | 10 | 17809348 | 17911164 | 1 | 1269.14 | 0.99 | 0.67 | 1.46 | 0.968 | 0.998 |
| MSLN | OID21380 | Oncology | MSLN | Q13421 | 16 | 760734 | 768865 | 1 | 7018.72 | 1.02 | 0.87 | 1.20 | 0.811 | 0.998 |
| MSMB | OID20275 | Cardiome | MSMB | P08118 | 10 | 46033307 | 46048180 | 1 | 17879.51 | 1.03 | 0.93 | 1.15 | 0.547 | 0.998 |
| MSRA | OID21513 | Oncology | MSRA | Q9UJ68 | 8 | 10054292 | 10428891 | 1 | 50.77 | 0.62 | 0.11 | 3.34 | 0.575 | 0.998 |
| MST1 | OID30713 | Inflammat | MST1 | P26927 | 3 | 49683947 | 49689501 | 1 | 17259.39 | 0.98 | 0.89 | 1.08 | 0.671 | 0.998 |
| MTHFSD | OID31305 | Oncology | MTHFSD | Q2M296 | 16 | 86530178 | 86555235 | 1 | 3364.00 | 1.01 | 0.82 | 1.25 | 0.894 | 0.998 |
| MTSS2 | OID31409 | Oncology | MTSS2 | Q765P7 | 16 | 70661204 | 70686053 | 1 | 93.44 | 0.72 | 0.20 | 2.62 | 0.613 | 0.998 |
| MTUS1 | OID31315 | Oncology | MTUS1 | Q9ULD2 | 8 | 17643795 | 17801094 | 1 | 516.96 | 0.86 | 0.49 | 1.52 | 0.607 | 0.998 |
| MUC16 | OID21378 | Oncology | MUC16 | Q8WXI7 | 19 | 8848844 | 8981342 | 1 | 81.00 | 0.73 | 0.18 | 3.02 | 0.665 | 0.998 |
| MUC2 | OID30377 | Cardiome | MUC2 | Q02817 | 11 | 1074875 | 1110511 | 1 | 1097.27 | 1.01 | 0.65 | 1.57 | 0.960 | 0.998 |
| MYOM3 | OID30633 | Inflammat | MYOM3 | Q5VTT5 | 1 | 24056035 | 24112135 | 1 | 114.49 | 0.94 | 0.27 | 3.30 | 0.929 | 0.998 |
| NAGK | OID30271 | Cardiome | NAGK | Q9UJ70 | 2 | 71064344 | 71079808 | 1 | 777.02 | 0.90 | 0.57 | 1.40 | 0.634 | 0.998 |
| NAGPA | OID30643 | Inflammat | NAGPA | Q9UK23 | 16 | 5024844 | 5034141 | 1 | 1681.00 | 1.02 | 0.75 | 1.38 | 0.899 | 0.998 |
| NAPRT | OID31114 | Neurology | NAPRT | Q6XQN6 | 8 | 143574785 | 143578649 | 1 | 2075.31 | 0.99 | 0.74 | 1.31 | 0.922 | 0.998 |
| NBL1 | OID21486 | Oncology | NBL1 | P41271 | 1 | 19596979 | 19658456 | 1 | 186.78 | 1.28 | 0.51 | 3.21 | 0.598 | 0.998 |
| NCAN | OID21055 | Neurology | NCAN | O14594 | 19 | 19211958 | 19252233 | 1 | 480.01 | 0.97 | 0.52 | 1.82 | 0.919 | 0.998 |
| NCR3LG1 | OID31490 | Oncology | NCR3LG1 | Q68D85 | 11 | 17351800 | 17377341 | 1 | 2659.61 | 0.95 | 0.73 | 1.24 | 0.700 | 0.998 |
| NECTIN2 | OID20222 | Cardiome | NECTIN2 | Q92692 | 19 | 44846175 | 44889223 | 1 | 523.27 | 1.08 | 0.62 | 1.90 | 0.780 | 0.998 |
| NEXN | OID30678 | Inflammat | NEXN | Q0ZGT2 | 1 | 77888513 | 77943895 | 1 | 113.27 | 1.48 | 0.43 | 5.13 | 0.538 | 0.998 |
| NFASC | OID20634 | Inflammat | NFASC | O94856 | 1 | 204828651 | 205022822 | 1 | 2483.36 | 0.97 | 0.74 | 1.26 | 0.798 | 0.998 |
| NFE2 | OID30268 | Cardiome | NFE2 | Q16621 | 12 | 54292111 | 54301015 | 1 | 68.06 | 1.06 | 0.25 | 4.42 | 0.941 | 0.998 |
| NFKB1 | OID30160 | Cardiome | NFKB1 | P19838 | 4 | 102501330 | 102617302 | 1 | 206.64 | 1.26 | 0.54 | 2.91 | 0.590 | 0.998 |
| NID1 | OID20362 | Cardiome | NID1 | P14543 | 1 | 235975830 | 236065109 | 1 | 402.11 | 1.02 | 0.54 | 1.93 | 0.959 | 0.998 |
| NMI | OID31378 | Oncology | NMI | Q13287 | 2 | 151270470 | 151289894 | 1 | 3206.39 | 1.04 | 0.84 | 1.28 | 0.739 | 0.998 |
| NOTCH2 | OID30407 | Cardiome | NOTCH2 | Q04721 | 1 | 119911553 | 120100779 | 1 | 205.44 | 1.06 | 0.44 | 2.55 | 0.895 | 0.998 |
| NPDC1 | OID20205 | Cardiome | NPDC1 | Q9NQX5 | 9 | 137039463 | 137046179 | 1 | 211.37 | 1.27 | 0.54 | 3.00 | 0.590 | 0.998 |
| NPHS1 | OID30540 | Inflammat | NPHS1 | O60500 | 19 | 35825372 | 35869287 | 1 | 2851.56 | 1.01 | 0.80 | 1.26 | 0.964 | 0.998 |
| NPTXR | OID20191 | Cardiome | NPTXR | O95502 | 22 | 38818452 | 38844028 | 1 | 5608.35 | 0.97 | 0.82 | 1.16 | 0.760 | 0.998 |
| NRCAM | OID20229 | Cardiome | NRCAM | Q92823 | 7 | 108147623 | 108456717 | 1 | 1387.56 | 0.90 | 0.64 | 1.28 | 0.573 | 0.998 |
| NT5C | OID30294 | Cardiome | NT5C | Q8TCD5 | 17 | 75130225 | 75131757 | 1 | 5370.80 | 0.99 | 0.83 | 1.19 | 0.945 | 0.998 |
| NT5C3A | OID20553 | Inflammat | NT5C3A | Q9H0P0 | 7 | 33014130 | 33062796 | 1 | 185.64 | 0.89 | 0.36 | 2.23 | 0.805 | 0.998 |
| NTF3 | OID20591 | Inflammat | NTF3 | P20783 | 12 | 5432108 | 5521536 | 1 | 373.78 | 0.86 | 0.46 | 1.62 | 0.638 | 0.998 |
| NTRK3 | OID21057 | Neurology | NTRK3 | Q16288 | 15 | 87859751 | 88256768 | 1 | 615.94 | 1.09 | 0.66 | 1.79 | 0.733 | 0.998 |
| NUDT5 | OID21115 | Neurology | NUDT5 | Q9UKK9 | 10 | 12165330 | 12196144 | 1 | 58.14 | 1.22 | 0.23 | 6.33 | 0.814 | 0.998 |
| OBP2B | OID20980 | Neurology | OBP2B | Q9NPH6 | 9 | 133205277 | 133209250 | 1 | 7744.00 | 1.01 | 0.85 | 1.20 | 0.883 | 0.998 |
| OGA | OID31465 | Oncology | OGA | O60502 | 10 | 101784443 | 101818465 | 1 | 280.56 | 1.14 | 0.53 | 2.46 | 0.739 | 0.998 |
| OMD | OID20725 | Inflammat | OMD | Q99983 | 9 | 92412380 | 92424471 | 1 | 100.00 | 0.72 | 0.19 | 2.70 | 0.626 | 0.998 |
| OPLAH | OID30201 | Cardiome | OPLAH | O14841 | 8 | 144051266 | 144063965 | 1 | 546.39 | 0.86 | 0.51 | 1.43 | 0.556 | 0.998 |
| ORM1 | OID30768 | Inflammat | ORM1 | P02763 | 9 | 114323098 | 114326479 | 1 | 2144.85 | 0.97 | 0.75 | 1.26 | 0.825 | 0.998 |
| OSCAR | OID20776 | Inflammat | OSCAR | Q8IYS5 | 19 | 54094668 | 54102692 | 1 | 4241.27 | 1.02 | 0.85 | 1.22 | 0.835 | 0.998 |
| OSM | OID20574 | Inflammat | OSM | P13725 | 22 | 30262829 | 30266851 | 1 | 56.55 | 0.67 | 0.14 | 3.28 | 0.623 | 0.998 |
| OTOA | OID30825 | Neurology | OTOA | Q7RTW8 | 16 | 21663968 | 21761935 | 1 | 50.77 | 1.55 | 0.28 | 8.46 | 0.614 | 0.998 |
| OXT | OID21047 | Neurology | OXT | P01178 | 20 | 3071620 | 3072517 | 1 | 4853.44 | 1.00 | 0.84 | 1.18 | 0.960 | 0.998 |
| P4HB | OID21509 | Oncology | P4HB | P07237 | 17 | 81843159 | 81860856 | 1 | 75.69 | 1.27 | 0.32 | 5.10 | 0.732 | 0.998 |
| PACS2 | OID30186 | Cardiome | PACS2 | Q86VP3 | 14 | 105300563 | 105398147 | 1 | 495.06 | 1.13 | 0.64 | 2.02 | 0.669 | 0.998 |
| PADI4 | OID20832 | Neurology | PADI4 | Q9UM07 | 1 | 17308195 | 17364004 | 1 | 1032.02 | 1.04 | 0.72 | 1.51 | 0.836 | 0.998 |
| PAEP | OID20925 | Neurology | PAEP | P09466 | 9 | 135561756 | 135566955 | 1 | 9365.94 | 0.98 | 0.85 | 1.12 | 0.744 | 0.998 |
| PALM | OID31515 | Oncology | PALM | O75781 | 19 | 708935 | 748329 | 1 | 397.50 | 1.01 | 0.54 | 1.91 | 0.967 | 0.998 |
| PAM | OID20291 | Cardiome | PAM | P19021 | 5 | 102753981 | 103029730 | 1 | 4179.24 | 1.03 | 0.85 | 1.25 | 0.794 | 0.998 |
| PAMR1 | OID21153 | Neurology | PAMR1 | Q6UXH9 | 11 | 35431823 | 35530300 | 1 | 2265.76 | 0.93 | 0.71 | 1.20 | 0.565 | 0.998 |
| PARK7 | OID21160 | Neurology | PARK7 | Q99497 | 1 | 7954291 | 7985505 | 1 | 436.81 | 1.06 | 0.58 | 1.92 | 0.851 | 0.998 |
| PCBD1 | OID30610 | Inflammat | PCBD1 | P61457 | 10 | 70882280 | 70888565 | 1 | 511.89 | 1.03 | 0.60 | 1.77 | 0.907 | 0.998 |
| PCDH7 | OID31456 | Oncology | PCDH7 | O60245 | 4 | 30720369 | 31146805 | 1 | 210.25 | 1.13 | 0.46 | 2.77 | 0.797 | 0.998 |
| PCDH9 | OID31516 | Oncology | PCDH9 | Q9HC56 | 13 | 66302834 | 67230445 | 1 | 807.51 | 0.98 | 0.63 | 1.52 | 0.919 | 0.998 |
| PCSK9 | OID20235 | Cardiome | PCSK9 | Q8NBP7 | 1 | 55039447 | 55064852 | 1 | 1695.67 | 0.96 | 0.72 | 1.28 | 0.778 | 0.998 |
| PDCD6 | OID20163 | Cardiome | PDCD6 | O75340 | 5 | 271621 | 314974 | 1 | 18520.74 | 0.98 | 0.89 | 1.07 | 0.636 | 0.998 |
| PDGFRB | OID20268 | Cardiome | PDGFRB | P09619 | 5 | 150113839 | 150155872 | 1 | 21719.39 | 1.00 | 0.92 | 1.08 | 0.959 | 0.998 |
| PDIA3 | OID30573 | Inflammat | PDIA3 | P30101 | 15 | 43746394 | 43773279 | 1 | 52.16 | 1.42 | 0.24 | 8.26 | 0.697 | 0.998 |
| PDZD2 | OID30151 | Cardiome | PDZD2 | O15018 | 5 | 31639131 | 32110932 | 1 | 792.02 | 1.14 | 0.72 | 1.79 | 0.575 | 0.998 |
| PEAR1 | OID20257 | Cardiome | PEAR1 | Q5VY43 | 1 | 156893698 | 156916429 | 1 | 708.89 | 1.11 | 0.70 | 1.77 | 0.659 | 0.998 |
| PEBP1 | OID21087 | Neurology | PEBP1 | P30086 | 12 | 118136124 | 118145584 | 1 | 171.82 | 1.14 | 0.42 | 3.14 | 0.796 | 0.998 |
| PECR | OID30228 | Cardiome | PECR | Q9BY49 | 2 | 215996329 | 216082955 | 1 | 1219.62 | 1.08 | 0.77 | 1.53 | 0.643 | 0.998 |
| PEPD | OID30688 | Inflammat | PEPD | P12955 | 19 | 33386950 | 33521823 | 1 | 2430.49 | 0.94 | 0.72 | 1.21 | 0.616 | 0.998 |
| PER3 | OID30511 | Inflammat | PER3 | P56645 | 1 | 7784291 | 7845177 | 1 | 2139.06 | 1.04 | 0.80 | 1.34 | 0.774 | 0.998 |
| PFKFB2 | OID21356 | Oncology | PFKFB2 | O60825 | 1 | 207034366 | 207081024 | 1 | 721.31 | 1.15 | 0.70 | 1.90 | 0.582 | 0.998 |
| PGLYRP2 | OID30742 | Inflammat | PGLYRP2 | Q96PD5 | 19 | 15468645 | 15498956 | 1 | 16157.23 | 1.03 | 0.93 | 1.14 | 0.590 | 0.998 |
| PGLYRP4 | OID30101 | Cardiome | PGLYRP4 | Q96LB8 | 1 | 153330120 | 153348841 | 1 | 2941.47 | 0.96 | 0.77 | 1.20 | 0.734 | 0.998 |
| PIBF1 | OID31061 | Neurology | PIBF1 | Q8WXW3 | 13 | 72782133 | 73016461 | 1 | 60.06 | 1.59 | 0.33 | 7.64 | 0.560 | 0.998 |
| PIK3AP1 | OID20587 | Inflammat | PIK3AP1 | Q6ZUJ8 | 10 | 96593315 | 96720514 | 1 | 653.09 | 1.05 | 0.63 | 1.76 | 0.851 | 0.998 |
| PIK3IP1 | OID21094 | Neurology | PIK3IP1 | Q96FE7 | 22 | 31281594 | 31292534 | 1 | 1670.90 | 0.91 | 0.65 | 1.28 | 0.607 | 0.998 |
| PINLYP | OID30651 | Inflammat | PINLYP | A6NC86 | 19 | 43575801 | 43583964 | 1 | 9192.02 | 1.01 | 0.89 | 1.15 | 0.854 | 0.998 |
| PKD2 | OID30184 | Cardiome | PKD2 | Q13563 | 4 | 88007635 | 88077777 | 1 | 130.34 | 0.96 | 0.32 | 2.84 | 0.939 | 0.998 |
| PLA2G15 | OID21473 | Oncology | PLA2G15 | Q8NCC3 | 16 | 68245304 | 68261058 | 1 | 937.89 | 0.98 | 0.64 | 1.51 | 0.941 | 0.998 |
| PLA2G1B | OID20252 | Cardiome | PLA2G1B | P04054 | 12 | 120322115 | 120327779 | 1 | 57.09 | 0.92 | 0.16 | 5.15 | 0.925 | 0.998 |
| PLAT | OID20247 | Cardiome | PLAT | P00750 | 8 | 42174718 | 42207709 | 1 | 148.20 | 0.81 | 0.29 | 2.29 | 0.692 | 0.998 |
| PLAUR | OID20764 | Inflammat | PLAUR | Q03405 | 19 | 43646095 | 43670547 | 1 | 313.29 | 0.95 | 0.47 | 1.94 | 0.895 | 0.998 |
| PLB1 | OID31110 | Neurology | PLB1 | Q6P1J6 | 2 | 28457145 | 28644142 | 1 | 15221.39 | 1.01 | 0.90 | 1.13 | 0.870 | 0.998 |
| PLCB2 | OID31155 | Neurology | PLCB2 | Q00722 | 15 | 40278176 | 40307935 | 1 | 365.77 | 1.04 | 0.54 | 2.03 | 0.902 | 0.998 |
| PLXDC1 | OID21258 | Oncology | PLXDC1 | Q8IUK5 | 17 | 39063313 | 39154394 | 1 | 883.41 | 1.08 | 0.70 | 1.66 | 0.720 | 0.998 |
| PLXNB2 | OID20317 | Cardiome | PLXNB2 | O15031 | 22 | 50274979 | 50307646 | 1 | 7943.27 | 0.98 | 0.85 | 1.13 | 0.779 | 0.998 |
| PM20D1 | OID20149 | Cardiome | PM20D1 | Q6GTS8 | 1 | 205828025 | 205850132 | 1 | 11025.00 | 1.04 | 0.92 | 1.17 | 0.559 | 0.998 |
| PODXL2 | OID21487 | Oncology | PODXL2 | Q9NZ53 | 3 | 127629185 | 127672802 | 1 | 1203.22 | 0.91 | 0.64 | 1.29 | 0.581 | 0.998 |
| PON2 | OID20088 | Cardiome | PON2 | Q15165 | 7 | 95404862 | 95435329 | 1 | 10066.78 | 1.01 | 0.89 | 1.15 | 0.848 | 0.998 |
| PON3 | OID20777 | Inflammat | PON3 | Q15166 | 7 | 95359872 | 95396375 | 1 | 2088.09 | 1.01 | 0.76 | 1.34 | 0.938 | 0.998 |
| POSTN | OID30410 | Cardiome | POSTN | Q15063 | 13 | 37562583 | 37598844 | 1 | 1024.00 | 0.90 | 0.61 | 1.32 | 0.583 | 0.998 |
| PPCDC | OID21027 | Neurology | PPCDC | Q96CD2 | 15 | 75023586 | 75117462 | 1 | 1995.11 | 1.04 | 0.79 | 1.38 | 0.762 | 0.998 |
| PPM1F | OID30350 | Cardiome | PPM1F | P49593 | 22 | 21919425 | 21952848 | 1 | 49.00 | 0.70 | 0.11 | 4.34 | 0.706 | 0.998 |
| PPP1R14D | OID30997 | Neurology | PPP1R14 | Q9NXH3 | 15 | 40815451 | 40828709 | 1 | 58.14 | 0.84 | 0.16 | 4.33 | 0.837 | 0.998 |
| PPP2R5A | OID31502 | Oncology | PPP2R5A | Q15172 | 1 | 212285410 | 212361853 | 1 | 134.03 | 0.87 | 0.31 | 2.45 | 0.786 | 0.998 |
| PRDX2 | OID30668 | Inflammat | PRDX2 | P32119 | 19 | 12796820 | 12801800 | 1 | 825.26 | 0.92 | 0.59 | 1.43 | 0.718 | 0.998 |
| PRELP | OID20766 | Inflammat | PRELP | P51888 | 1 | 203475806 | 203491352 | 1 | 4210.16 | 1.07 | 0.85 | 1.33 | 0.582 | 0.998 |
| PRG3 | OID30383 | Cardiome | PRG3 | Q9Y2Y8 | 11 | 57376769 | 57381150 | 1 | 260.59 | 0.86 | 0.39 | 1.89 | 0.704 | 0.998 |
| PROK1 | OID20543 | Inflammat | PROK1 | P58294 | 1 | 110451149 | 110457358 | 1 | 3585.02 | 0.95 | 0.72 | 1.24 | 0.696 | 0.998 |
| PRRT3 | OID30276 | Cardiome | PRRT3 | Q5FWE3 | 3 | 9945542 | 9952408 | 1 | 560.11 | 0.90 | 0.54 | 1.51 | 0.699 | 0.998 |
| PRSS2 | OID20364 | Cardiome | PRSS2 | P07478 | 7 | 142760398 | 142774564 | 1 | 301.89 | 0.99 | 0.50 | 1.97 | 0.969 | 0.998 |
| PRSS27 | OID20212 | Cardiome | PRSS27 | Q9BQR3 | 16 | 2712419 | 2720551 | 1 | 886.21 | 1.03 | 0.64 | 1.66 | 0.893 | 0.998 |
| PRSS8 | OID20763 | Inflammat | PRSS8 | Q16651 | 16 | 31131433 | 31135727 | 1 | 206.64 | 1.02 | 0.41 | 2.54 | 0.958 | 0.998 |

| | | | | | | | | | | | | | | |
|---|---|---|---|---|---|---|---|---|---|---|---|---|---|---|
| PRTFDC1 | OID20867 | Neurology | PRTFDC1 | Q9NRG1 | 10 | 24848614 | 24952606 | 1 | 676.00 | 1.07 | 0.66 | 1.73 | 0.792 | 0.998 |
| PRTG | OID21443 | Oncology | PRTG | Q2VWP7 | 15 | 55611544 | 55743152 | 1 | 3150.02 | 0.95 | 0.76 | 1.19 | 0.658 | 0.998 |
| PSCA | OID31493 | Oncology | PSCA | O43653 | 8 | 142670308 | 142682725 | 1 | 21061.27 | 0.99 | 0.91 | 1.08 | 0.865 | 0.998 |
| PSG1 | OID20916 | Neurology | PSG1 | P11464 | 19 | 42866464 | 42879822 | 1 | 11425.23 | 1.02 | 0.90 | 1.15 | 0.752 | 0.998 |
| PSME1 | OID20969 | Neurology | PSME1 | Q06323 | 14 | 24136163 | 24138967 | 1 | 138.06 | 0.85 | 0.27 | 2.69 | 0.783 | 0.998 |
| PSME2 | OID20989 | Neurology | PSME2 | Q9UL46 | 14 | 24143362 | 24147570 | 1 | 169.00 | 0.86 | 0.30 | 2.45 | 0.783 | 0.998 |
| PSMG4 | OID30514 | Inflammat | PSMG4 | Q5JS54 | 6 | 3231403 | 3303373 | 1 | 376.36 | 0.89 | 0.47 | 1.71 | 0.734 | 0.998 |
| PSTPIP2 | OID30596 | Inflammat | PSTPIP2 | Q9H939 | 18 | 45983536 | 46072272 | 1 | 422.12 | 1.11 | 0.60 | 2.07 | 0.734 | 0.998 |
| PTGES2 | OID30561 | Inflammat | PTGES2 | Q9H7Z7 | 9 | 128120693 | 128128462 | 1 | 116.16 | 0.86 | 0.26 | 2.82 | 0.806 | 0.998 |
| PTN | OID20083 | Cardiome | PTN | P21246 | 7 | 137227341 | 137343774 | 1 | 465.33 | 0.91 | 0.48 | 1.73 | 0.784 | 0.998 |
| PTPRC | OID30380 | Cardiome | PTPRC | P08575 | 1 | 198638457 | 198757476 | 1 | 772.46 | 1.16 | 0.71 | 1.89 | 0.548 | 0.998 |
| PTPRF | OID20310 | Cardiome | PTPRF | P10586 | 1 | 43525187 | 43623666 | 1 | 97.79 | 1.37 | 0.38 | 4.87 | 0.630 | 0.998 |
| PTPRH | OID31433 | Oncology | PTPRH | Q9HD43 | 19 | 55181247 | 55209506 | 1 | 6798.75 | 0.97 | 0.83 | 1.13 | 0.663 | 0.998 |
| PTPRK | OID31461 | Oncology | PTPRK | Q15262 | 6 | 127968785 | 128520616 | 1 | 184.96 | 1.21 | 0.49 | 3.01 | 0.676 | 0.998 |
| PTPRM | OID20598 | Inflammat | PTPRM | P28827 | 18 | 7566782 | 8406861 | 1 | 100.00 | 1.36 | 0.33 | 5.57 | 0.672 | 0.998 |
| PTS | OID20847 | Neurology | PTS | Q03393 | 11 | 112226367 | 112269955 | 1 | 687.60 | 0.98 | 0.61 | 1.58 | 0.929 | 0.998 |
| PTX3 | OID20570 | Inflammat | PTX3 | P26022 | 3 | 157436850 | 157443633 | 1 | 62.33 | 0.93 | 0.20 | 4.40 | 0.927 | 0.998 |
| PVALB | OID21420 | Oncology | PVALB | P20472 | 22 | 36800684 | 36819479 | 1 | 10327.64 | 0.96 | 0.85 | 1.09 | 0.580 | 0.998 |
| PXDNL | OID31405 | Oncology | PXDNL | A1KZ92 | 8 | 51319577 | 51809445 | 1 | 826.56 | 1.07 | 0.71 | 1.62 | 0.743 | 0.998 |
| PXN | OID20822 | Neurology | PXN | P49023 | 12 | 120210439 | 120265771 | 1 | 7074.68 | 1.05 | 0.90 | 1.21 | 0.555 | 0.998 |
| PZP | OID30730 | Inflammat | PZP | P20742 | 12 | 9148840 | 9208395 | 1 | 3080.25 | 1.05 | 0.77 | 1.44 | 0.755 | 0.998 |
| QDPR | OID20174 | Cardiome | QDPR | P09417 | 4 | 17460261 | 17512206 | 1 | 2533.44 | 0.94 | 0.74 | 1.19 | 0.598 | 0.998 |
| QPCT | OID20320 | Cardiome | QPCT | Q16769 | 2 | 37342827 | 37373322 | 1 | 1250.33 | 1.09 | 0.76 | 1.56 | 0.655 | 0.998 |
| RAB44 | OID31346 | Oncology | RAB44 | Q7Z6P3 | 6 | 36697826 | 36733184 | 1 | 266.78 | 0.86 | 0.42 | 1.78 | 0.691 | 0.998 |
| RAB6A | OID20608 | Inflammat | RAB6A | P20340 | 11 | 73675638 | 73761137 | 1 | 53.29 | 0.60 | 0.09 | 3.94 | 0.591 | 0.998 |
| RABEP1 | OID30647 | Inflammat | RABEP1 | Q15276 | 17 | 5282265 | 5386340 | 1 | 78.77 | 0.95 | 0.25 | 3.66 | 0.943 | 0.998 |
| RABEPK | OID21163 | Oncology | RABEPK | Q7Z6M1 | 9 | 125200542 | 125234161 | 1 | 354.12 | 1.21 | 0.64 | 2.27 | 0.563 | 0.998 |
| RALY | OID31382 | Oncology | RALY | Q9UKM9 | 20 | 33993646 | 34108308 | 1 | 85.56 | 1.43 | 0.39 | 5.28 | 0.588 | 0.998 |
| RARRES1 | OID21177 | Oncology | RARRES1 | P49788 | 3 | 158696892 | 158732489 | 1 | 3263.27 | 0.99 | 0.80 | 1.22 | 0.918 | 0.998 |
| RARRES2 | OID20373 | Cardiome | RARRES2 | Q99969 | 7 | 150338317 | 150341662 | 1 | 729.00 | 0.89 | 0.54 | 1.47 | 0.641 | 0.998 |
| RBP2 | OID21365 | Oncology | RBP2 | P50120 | 3 | 139452884 | 139480747 | 1 | 72.25 | 1.31 | 0.31 | 5.51 | 0.714 | 0.998 |
| RBP7 | OID31526 | Oncology | RBP7 | Q96R05 | 1 | 9997206 | 10016021 | 1 | 1178.78 | 1.11 | 0.76 | 1.61 | 0.603 | 0.998 |
| RBPMS2 | OID30257 | Cardiome | RBPMS2 | Q6ZRY4 | 15 | 64739891 | 64775589 | 1 | 101.11 | 0.80 | 0.23 | 2.78 | 0.723 | 0.998 |
| REG3G | OID31408 | Oncology | REG3G | Q6UW15 | 2 | 79025686 | 79028505 | 1 | 1016.90 | 0.91 | 0.62 | 1.34 | 0.629 | 0.998 |
| RHOC | OID20950 | Neurology | RHOC | P08134 | 1 | 112701127 | 112707434 | 1 | 110.25 | 0.96 | 0.31 | 2.97 | 0.948 | 0.998 |
| RIDA | OID30635 | Inflammat | RIDA | P52758 | 8 | 98102344 | 98117171 | 1 | 1580.06 | 1.10 | 0.80 | 1.53 | 0.553 | 0.998 |
| RNASE10 | OID30238 | Cardiome | RNASE10 | Q5GAN6 | 14 | 20505537 | 20513884 | 1 | 10075.14 | 0.99 | 0.88 | 1.11 | 0.881 | 0.998 |
| ROR1 | OID20224 | Cardiome | ROR1 | Q01973 | 1 | 63774017 | 64181498 | 1 | 1552.36 | 0.92 | 0.67 | 1.26 | 0.616 | 0.998 |
| RPA2 | OID30453 | Inflammat | RPA2 | P15927 | 1 | 27891524 | 27914746 | 1 | 184.96 | 1.14 | 0.46 | 2.82 | 0.781 | 0.998 |
| RPL14 | OID30284 | Cardiome | RPL14 | P50914 | 3 | 40457292 | 40468587 | 1 | 48.18 | 0.67 | 0.12 | 3.85 | 0.653 | 0.998 |
| RRM2B | OID21174 | Oncology | RRM2B | Q7LG56 | 8 | 102204502 | 102238961 | 1 | 192.02 | 0.90 | 0.36 | 2.24 | 0.822 | 0.998 |
| RTBDN | OID21343 | Oncology | RTBDN | Q9BSG5 | 19 | 12825478 | 12835428 | 1 | 456.89 | 0.95 | 0.50 | 1.77 | 0.861 | 0.998 |
| S100A11 | OID20130 | Cardiome | S100A11 | P31949 | 1 | 152032506 | 152047907 | 1 | 328.01 | 1.03 | 0.48 | 2.18 | 0.946 | 0.998 |
| S100A13 | OID30529 | Inflammat | S100A13 | Q99584 | 1 | 153618787 | 153631360 | 1 | 703.65 | 1.05 | 0.65 | 1.68 | 0.854 | 0.998 |
| S100A4 | OID21519 | Oncology | S100A4 | P26447 | 1 | 153543613 | 153550136 | 1 | 450.94 | 1.08 | 0.60 | 1.96 | 0.796 | 0.998 |
| S100P | OID20103 | Cardiome | S100P | P25815 | 4 | 6693878 | 6697170 | 1 | 1104.58 | 1.05 | 0.71 | 1.54 | 0.819 | 0.998 |
| SAMD9L | OID20536 | Inflammat | SAMD9L | Q8IVG5 | 7 | 93130056 | 93148385 | 1 | 1428.84 | 0.98 | 0.71 | 1.36 | 0.904 | 0.998 |
| SBSN | OID30398 | Cardiome | SBSN | Q6UWP8 | 19 | 35523367 | 35528351 | 1 | 4253.94 | 0.99 | 0.82 | 1.19 | 0.882 | 0.998 |
| SCARF2 | OID21048 | Neurology | SCARF2 | Q96GP6 | 22 | 20424584 | 20437826 | 1 | 2185.56 | 1.08 | 0.79 | 1.49 | 0.618 | 0.998 |
| SCG3 | OID20728 | Inflammat | SCG3 | Q8WXD2 | 15 | 51681492 | 51721026 | 1 | 6188.44 | 1.04 | 0.89 | 1.22 | 0.619 | 0.998 |
| SCGB1A1 | OID20688 | Inflammat | SCGB1A1 | P11684 | 11 | 62405103 | 62423195 | 1 | 1795.64 | 1.03 | 0.76 | 1.38 | 0.866 | 0.998 |
| SCGN | OID20457 | Inflammat | SCGN | O76038 | 6 | 25652201 | 25701783 | 1 | 132.25 | 1.23 | 0.39 | 3.93 | 0.725 | 0.998 |
| SCPEP1 | OID30278 | Cardiome | SCPEP1 | Q9HB40 | 17 | 56978129 | 57006768 | 1 | 1644.75 | 0.99 | 0.74 | 1.33 | 0.964 | 0.998 |
| SCRG1 | OID30333 | Cardiome | SCRG1 | O75711 | 4 | 173384701 | 173406380 | 1 | 289.00 | 1.28 | 0.55 | 2.98 | 0.571 | 0.998 |
| SDK2 | OID30662 | Inflammat | SDK2 | Q58EX2 | 17 | 73334384 | 73644445 | 1 | 3754.35 | 0.96 | 0.79 | 1.17 | 0.701 | 0.998 |
| SEC31A | OID30269 | Cardiome | SEC31A | O94979 | 4 | 82818509 | 82901166 | 1 | 166.84 | 0.91 | 0.35 | 2.39 | 0.848 | 0.998 |
| SELL | OID30752 | Inflammat | SELL | P14151 | 1 | 169690665 | 169711702 | 1 | 2588.27 | 1.06 | 0.83 | 1.37 | 0.629 | 0.998 |
| SELP | OID20379 | Cardiome | SELP | P16109 | 1 | 169588849 | 169630193 | 1 | 1127.84 | 0.98 | 0.68 | 1.41 | 0.905 | 0.998 |
| SELPLG | OID20560 | Inflammat | SELPLG | Q14242 | 12 | 108621895 | 108633894 | 1 | 4329.64 | 0.98 | 0.81 | 1.18 | 0.803 | 0.998 |
| SEMA3F | OID20186 | Cardiome | SEMA3F | Q13275 | 3 | 50155045 | 50189075 | 1 | 309.76 | 0.86 | 0.42 | 1.73 | 0.665 | 0.998 |
| SEMA4D | OID21020 | Neurology | SEMA4D | Q92854 | 9 | 89360787 | 89498130 | 1 | 114.49 | 0.93 | 0.29 | 2.92 | 0.896 | 0.998 |
| SEMA6C | OID30531 | Inflammat | SEMA6C | Q9H3T2 | 1 | 151131685 | 151146631 | 1 | 152.11 | 1.38 | 0.48 | 3.92 | 0.549 | 0.998 |
| SEPTIN9 | OID21279 | Oncology | SEPTIN9 | Q9UHD8 | 17 | 77280569 | 77500596 | 1 | 131.14 | 1.12 | 0.37 | 3.41 | 0.844 | 0.998 |
| SERPINA12 | OID20219 | Cardiome | SERPINA | Q8IW75 | 14 | 94487274 | 94517844 | 1 | 7472.64 | 0.97 | 0.84 | 1.13 | 0.706 | 0.998 |
| SERPINA3 | OID30786 | Inflammat | SERPINA | P01011 | 14 | 94612384 | 94624055 | 1 | 1998.09 | 0.95 | 0.73 | 1.25 | 0.731 | 0.998 |
| SERPINA4 | OID30774 | Inflammat | SERPINA | P29622 | 14 | 94561442 | 94569913 | 1 | 4699.86 | 1.01 | 0.84 | 1.22 | 0.890 | 0.998 |
| SERPINA6 | OID30743 | Inflammat | SERPINA | P08185 | 14 | 94304248 | 94323389 | 1 | 934.61 | 1.03 | 0.66 | 1.59 | 0.911 | 0.998 |
| SERPINA9 | OID21284 | Oncology | SERPINA | Q86WD7 | 14 | 94462717 | 94479689 | 1 | 3875.06 | 0.97 | 0.79 | 1.20 | 0.805 | 0.998 |
| SERPINB1 | OID21088 | Neurology | SERPINB | P30740 | 6 | 2832332 | 2841959 | 1 | 104.04 | 0.77 | 0.23 | 2.63 | 0.675 | 0.998 |
| SERPINB9 | OID20932 | Neurology | SERPINB | P50453 | 6 | 2887270 | 2903309 | 1 | 221.02 | 0.78 | 0.33 | 1.83 | 0.564 | 0.998 |
| SERPINE1 | OID20342 | Cardiome | SERPINE | P05121 | 7 | 101127104 | 101139247 | 1 | 174.83 | 1.09 | 0.40 | 2.99 | 0.865 | 0.998 |
| SERPINF1 | OID30757 | Inflammat | SERPINF | P36955 | 17 | 1762029 | 1777565 | 1 | 1251.39 | 1.08 | 0.76 | 1.54 | 0.654 | 0.998 |
| SERPING1 | OID30777 | Inflammat | SERPING | P05155 | 11 | 57597387 | 57619171 | 1 | 625.00 | 0.97 | 0.59 | 1.58 | 0.901 | 0.998 |
| SERPINH1 | OID31322 | Oncology | SERPINH | P50454 | 11 | 75562056 | 75572783 | 1 | 293.27 | 1.03 | 0.50 | 2.09 | 0.943 | 0.998 |
| SFRP4 | OID30648 | Inflammat | SFRP4 | Q6FHJ7 | 7 | 37905932 | 38025695 | 1 | 724.10 | 1.13 | 0.71 | 1.80 | 0.608 | 0.998 |
| SFTPD | OID20214 | Cardiome | SFTPD | P35247 | 10 | 79937467 | 79982614 | 1 | 11725.80 | 0.98 | 0.87 | 1.12 | 0.788 | 0.998 |
| SGSH | OID30341 | Cardiome | SGSH | P51688 | 17 | 80206716 | 80220923 | 1 | 16020.33 | 0.98 | 0.88 | 1.10 | 0.759 | 0.998 |
| SH2B3 | OID21222 | Oncology | SH2B3 | Q9UQQ2 | 12 | 111405923 | 111451623 | 1 | 70.56 | 1.05 | 0.26 | 4.29 | 0.941 | 0.998 |
| SH3BP1 | OID31439 | Oncology | SH3BP1 | Q9Y3L3 | 22 | 37634654 | 37656117 | 1 | 718.24 | 0.87 | 0.55 | 1.38 | 0.564 | 0.998 |
| SHMT1 | OID20714 | Inflammat | SHMT1 | P34896 | 17 | 18327860 | 18363563 | 1 | 8077.52 | 1.03 | 0.90 | 1.18 | 0.666 | 0.998 |
| SIAE | OID21503 | Oncology | SIAE | Q9HAT2 | 11 | 124633113 | 124695707 | 1 | 770.06 | 0.90 | 0.58 | 1.40 | 0.629 | 0.998 |
| SIGLEC1 | OID20690 | Inflammat | SIGLEC1 | Q9BZZ2 | 20 | 3686970 | 3712600 | 1 | 2769.77 | 0.98 | 0.77 | 1.24 | 0.873 | 0.998 |
| SIGLEC6 | OID21394 | Oncology | SIGLEC6 | O43699 | 19 | 51517819 | 51531856 | 1 | 2673.50 | 0.98 | 0.77 | 1.25 | 0.861 | 0.998 |
| SIGLEC8 | OID30261 | Cardiome | SIGLEC8 | Q9NYZ4 | 19 | 51450847 | 51458456 | 1 | 3866.58 | 1.03 | 0.84 | 1.26 | 0.791 | 0.998 |
| SIGLEC9 | OID21390 | Oncology | SIGLEC9 | Q9Y336 | 19 | 51124906 | 51136651 | 1 | 6700.59 | 1.02 | 0.87 | 1.21 | 0.773 | 0.998 |
| SIL1 | OID30207 | Cardiome | SIL1 | Q9H173 | 5 | 138946724 | 139293557 | 1 | 54.39 | 0.83 | 0.16 | 4.24 | 0.825 | 0.998 |
| SIRPB1 | OID20739 | Inflammat | SIRPB1 | O00241 | 20 | 1561385 | 1620061 | 1 | 7458.68 | 0.96 | 0.83 | 1.10 | 0.539 | 0.998 |
| SKAP1 | OID20906 | Neurology | SKAP1 | Q86WV1 | 17 | 48133442 | 48430275 | 1 | 58.14 | 1.64 | 0.29 | 9.22 | 0.577 | 0.998 |
| SLAMF6 | OID21254 | Oncology | SLAMF6 | Q96DU3 | 1 | 160485030 | 160523262 | 1 | 570.68 | 0.94 | 0.54 | 1.61 | 0.812 | 0.998 |
| SLC27A4 | OID20814 | Neurology | SLC27A4 | Q6P1M0 | 9 | 128340527 | 128361470 | 1 | 45.56 | 0.81 | 0.13 | 4.96 | 0.819 | 0.998 |
| SLC9A3R1 | OID30630 | Inflammat | SLC9A3R | O14745 | 17 | 74748628 | 74769353 | 1 | 132.25 | 1.29 | 0.45 | 3.72 | 0.640 | 0.998 |
| SLC9A3R2 | OID31445 | Oncology | SLC9A3R | Q15599 | 16 | 2025356 | 2039026 | 1 | 216.09 | 1.06 | 0.44 | 2.57 | 0.895 | 0.998 |
| SLITRK1 | OID30510 | Inflammat | SLITRK1 | Q96PX8 | 13 | 83877205 | 83882474 | 1 | 246.49 | 1.03 | 0.46 | 2.32 | 0.940 | 0.998 |
| SLURP1 | OID30404 | Cardiome | SLURP1 | P55000 | 8 | 142740949 | 142742406 | 1 | 3413.90 | 0.95 | 0.76 | 1.20 | 0.684 | 0.998 |
| SMAD3 | OID31418 | Oncology | SMAD3 | P84022 | 15 | 67063763 | 67195173 | 1 | 741.05 | 1.10 | 0.70 | 1.73 | 0.679 | 0.998 |
| SMOC1 | OID21461 | Oncology | SMOC1 | Q9H4F8 | 14 | 69854131 | 70032366 | 1 | 1346.89 | 0.97 | 0.68 | 1.39 | 0.878 | 0.998 |
| SMOC2 | OID20730 | Inflammat | SMOC2 | Q9H3U7 | 6 | 168441151 | 168673445 | 1 | 2437.89 | 0.98 | 0.75 | 1.28 | 0.868 | 0.998 |
| SMPD1 | OID21028 | Neurology | SMPD1 | P17405 | 11 | 6390440 | 6394998 | 1 | 2997.56 | 1.01 | 0.78 | 1.29 | 0.959 | 0.998 |
| SNAP25 | OID30811 | Neurology | SNAP25 | P60880 | 20 | 10172395 | 10308258 | 1 | 165.77 | 1.02 | 0.40 | 2.65 | 0.961 | 0.998 |
| SNCG | OID20971 | Neurology | SNCG | O76070 | 10 | 86958599 | 86963258 | 1 | 13897.79 | 0.98 | 0.88 | 1.08 | 0.681 | 0.998 |
| SNED1 | OID30205 | Cardiome | SNED1 | Q8TER0 | 2 | 240998618 | 241095568 | 1 | 244.14 | 1.07 | 0.50 | 2.30 | 0.858 | 0.998 |
| SNX2 | OID31374 | Oncology | SNX2 | O60749 | 5 | 122775079 | 122834543 | 1 | 144.00 | 0.76 | 0.28 | 2.05 | 0.590 | 0.998 |
| SNX9 | OID20135 | Cardiome | SNX9 | Q9Y5X1 | 6 | 157700387 | 157945077 | 1 | 123.77 | 0.80 | 0.25 | 2.60 | 0.715 | 0.998 |
| SOD3 | OID30683 | Inflammat | SOD3 | P08294 | 4 | 24789912 | 24800842 | 1 | 5170.51 | 0.94 | 0.76 | 1.17 | 0.592 | 0.998 |
| SORD | OID21453 | Oncology | SORD | Q00796 | 15 | 45023147 | 45077185 | 1 | 347.31 | 0.91 | 0.45 | 1.83 | 0.791 | 0.998 |
| SPARCL1 | OID20361 | Cardiome | SPARCL1 | Q14515 | 4 | 87473335 | 87531061 | 1 | 7965.56 | 1.02 | 0.89 | 1.17 | 0.815 | 0.998 |
| SPINK2 | OID30627 | Inflammat | SPINK2 | P20155 | 4 | 56809860 | 56821742 | 1 | 3700.69 | 0.99 | 0.79 | 1.26 | 0.965 | 0.998 |

| | | | | | | | | | | | | | | |
|---|---|---|---|---|---|---|---|---|---|---|---|---|---|---|
| SPINK4 | OID20509 | Inflammat | SPINK4 | O60575 | 9 | 33218365 | 33248567 | 1 | 12409.96 | 1.01 | 0.90 | 1.14 | 0.873 | 0.998 |
| SPINK8 | OID30153 | Cardiome | SPINK8 | P0C7L1 | 3 | 48306842 | 48333661 | 1 | 3320.64 | 0.96 | 0.77 | 1.19 | 0.701 | 0.998 |
| SPINT2 | OID20720 | Inflammat | SPINT2 | O43291 | 19 | 38244035 | 38292615 | 1 | 450.38 | 0.96 | 0.50 | 1.81 | 0.891 | 0.998 |
| SPINT3 | OID31509 | Oncology | SPINT3 | P49223 | 20 | 45512461 | 45515622 | 1 | 331.24 | 1.12 | 0.40 | 3.17 | 0.826 | 0.998 |
| SPON1 | OID20759 | Inflammat | SPON1 | Q9HCB6 | 11 | 13962723 | 14268133 | 1 | 1728.18 | 1.09 | 0.78 | 1.51 | 0.613 | 0.998 |
| SPP1 | OID20322 | Cardiome | SPP1 | P10451 | 4 | 87975667 | 87983532 | 1 | 160.44 | 1.10 | 0.38 | 3.16 | 0.855 | 0.998 |
| SPRED2 | OID30423 | Inflammat | SPRED2 | Q7Z698 | 2 | 65310851 | 65432637 | 1 | 81.00 | 0.83 | 0.22 | 3.16 | 0.785 | 0.998 |
| STX8 | OID20531 | Inflammat | STX8 | Q9UNK0 | 17 | 9250471 | 9576591 | 1 | 128.44 | 0.88 | 0.30 | 2.59 | 0.812 | 0.998 |
| STXBP1 | OID31054 | Neurology | STXBP1 | P61764 | 9 | 127579370 | 127696027 | 1 | 47.27 | 0.65 | 0.11 | 3.78 | 0.635 | 0.998 |
| SUGP1 | OID31524 | Oncology | SUGP1 | Q8IWZ8 | 19 | 19276018 | 19320509 | 1 | 1056.25 | 0.98 | 0.66 | 1.46 | 0.922 | 0.998 |
| SULT2A1 | OID20508 | Inflammat | SULT2A1 | Q06520 | 19 | 47870467 | 47886315 | 1 | 1428.84 | 1.11 | 0.79 | 1.56 | 0.550 | 0.998 |
| SWAP70 | OID31477 | Oncology | SWAP70 | Q9UH65 | 11 | 9664077 | 9752993 | 1 | 328.52 | 1.04 | 0.52 | 2.06 | 0.912 | 0.998 |
| TBC1D17 | OID20844 | Neurology | TBC1D17 | Q9HA65 | 19 | 49877666 | 49888750 | 1 | 3497.88 | 0.99 | 0.79 | 1.25 | 0.935 | 0.998 |
| TBC1D23 | OID21286 | Oncology | TBC1D23 | Q9NUY8 | 3 | 100260992 | 100325251 | 1 | 370.56 | 1.05 | 0.57 | 1.97 | 0.866 | 0.998 |
| TBCA | OID30616 | Inflammat | TBCA | O75347 | 5 | 77691166 | 77868780 | 1 | 306.25 | 0.97 | 0.47 | 2.01 | 0.930 | 0.998 |
| TCL1A | OID20987 | Neurology | TCL1A | P56279 | 14 | 95709947 | 95714196 | 1 | 1573.44 | 0.95 | 0.69 | 1.31 | 0.755 | 0.998 |
| TCN1 | OID30687 | Inflammat | TCN1 | P20061 | 11 | 59852800 | 59866489 | 1 | 3948.03 | 1.05 | 0.85 | 1.28 | 0.671 | 0.998 |
| TCTN3 | OID30326 | Cardiome | TCTN3 | Q6NUS6 | 10 | 95659823 | 95694143 | 1 | 6221.27 | 1.01 | 0.86 | 1.19 | 0.879 | 0.998 |
| TDP1 | OID31403 | Oncology | TDP1 | Q9NUW8 | 14 | 89954939 | 90044764 | 1 | 112.07 | 0.98 | 0.30 | 3.20 | 0.968 | 0.998 |
| TEF | OID30103 | Cardiome | TEF | Q10587 | 22 | 41367333 | 41399326 | 1 | 70.56 | 1.41 | 0.36 | 5.49 | 0.620 | 0.998 |
| TF | OID30783 | Inflammat | TF | P02787 | 3 | 133746040 | 133796641 | 1 | 1691.27 | 0.97 | 0.72 | 1.32 | 0.868 | 0.998 |
| TFF2 | OID20787 | Inflammat | TFF2 | Q03403 | 21 | 42346357 | 42350997 | 1 | 244.14 | 1.20 | 0.51 | 2.81 | 0.673 | 0.998 |
| TGFBR3 | OID20255 | Cardiome | TGFBR3 | Q03167 | 1 | 91680343 | 91906335 | 1 | 272.25 | 0.84 | 0.40 | 1.75 | 0.634 | 0.998 |
| TGOLN2 | OID30691 | Inflammat | TGOLN2 | O43493 | 2 | 85318027 | 85328251 | 1 | 10433.16 | 1.02 | 0.90 | 1.17 | 0.732 | 0.998 |
| THBD | OID20231 | Cardiome | THBD | P07204 | 20 | 23045633 | 23049672 | 1 | 498.78 | 0.83 | 0.46 | 1.50 | 0.538 | 0.998 |
| TIGAR | OID20894 | Neurology | TIGAR | Q9NQ88 | 12 | 4307763 | 4360028 | 1 | 53.78 | 1.12 | 0.21 | 5.93 | 0.897 | 0.998 |
| TIMD4 | OID20375 | Cardiome | TIMD4 | Q96H15 | 5 | 156919292 | 156963226 | 1 | 1098.45 | 0.96 | 0.63 | 1.46 | 0.847 | 0.998 |
| TIMM10 | OID30358 | Cardiome | TIMM10 | P62072 | 11 | 57528464 | 57530803 | 1 | 570.68 | 0.92 | 0.54 | 1.56 | 0.757 | 0.998 |
| TIMP2 | OID30318 | Cardiome | TIMP2 | P16035 | 17 | 78852977 | 78925387 | 1 | 135.14 | 1.22 | 0.44 | 3.36 | 0.706 | 0.998 |
| TINAGL1 | OID20180 | Cardiome | TINAGL1 | Q9GZM7 | 1 | 31576485 | 31587686 | 1 | 112.89 | 1.30 | 0.41 | 4.15 | 0.653 | 0.998 |
| TLR1 | OID30532 | Inflammat | TLR1 | Q15399 | 4 | 38790677 | 38856817 | 1 | 3123.57 | 1.04 | 0.83 | 1.30 | 0.726 | 0.998 |
| TLR3 | OID20612 | Inflammat | TLR3 | O15455 | 4 | 186069155 | 186088073 | 1 | 20700.02 | 0.99 | 0.91 | 1.09 | 0.883 | 0.998 |
| TMEM132A | OID30287 | Cardiome | TMEM132 | Q24JP5 | 11 | 60924460 | 60937159 | 1 | 5871.39 | 0.97 | 0.83 | 1.13 | 0.693 | 0.998 |
| TMPRSS11D | OID31103 | Neurology | TMPRSS1 | O60235 | 4 | 67820876 | 67884002 | 1 | 292.41 | 1.20 | 0.55 | 2.62 | 0.648 | 0.998 |
| TMPRSS15 | OID21364 | Oncology | TMPRSS1 | P98073 | 21 | 18269116 | 18485879 | 1 | 158.76 | 0.94 | 0.35 | 2.53 | 0.898 | 0.998 |
| TNFRSF11A | OID20646 | Inflammat | TNFRSF1 | Q9Y6Q6 | 18 | 62325287 | 62391288 | 1 | 4414.86 | 0.99 | 0.81 | 1.21 | 0.935 | 0.998 |
| TNFRSF11B | OID20735 | Inflammat | TNFRSF1 | O00300 | 8 | 118923557 | 118951885 | 1 | 816.33 | 1.06 | 0.66 | 1.71 | 0.799 | 0.998 |
| TNFRSF13B | OID20702 | Inflammat | TNFRSF1 | O14836 | 17 | 16929816 | 16972118 | 1 | 210.25 | 1.11 | 0.45 | 2.75 | 0.817 | 0.998 |
| TNFRSF14 | OID20783 | Inflammat | TNFRSF1 | Q92956 | 1 | 2555639 | 2565382 | 1 | 269.90 | 1.02 | 0.45 | 2.32 | 0.963 | 0.998 |
| TNFRSF19 | OID21446 | Oncology | TNFRSF1 | Q9NS68 | 13 | 23570370 | 23676104 | 1 | 1048.14 | 0.97 | 0.66 | 1.44 | 0.887 | 0.998 |
| TNFRSF6B | OID20964 | Neurology | TNFRSF6 | O95407 | 20 | 63696652 | 63698684 | 1 | 1491.89 | 0.92 | 0.67 | 1.27 | 0.627 | 0.998 |
| TNFRSF9 | OID20985 | Neurology | TNFRSF9 | Q07011 | 1 | 7915871 | 7943165 | 1 | 464.64 | 0.98 | 0.53 | 1.82 | 0.956 | 0.998 |
| TNFSF10 | OID20611 | Inflammat | TNFSF10 | P50591 | 3 | 172505508 | 172523475 | 1 | 595.80 | 1.09 | 0.64 | 1.84 | 0.760 | 0.998 |
| TNFSF12 | OID20624 | Inflammat | TNFSF12 | O43508 | 17 | 7549058 | 7557890 | 1 | 2162.25 | 0.92 | 0.70 | 1.22 | 0.579 | 0.998 |
| TNFSF13B | OID20272 | Cardiome | TNFSF13 | Q9Y275 | 13 | 108251240 | 108308484 | 1 | 763.40 | 1.13 | 0.72 | 1.79 | 0.589 | 0.998 |
| TNN | OID30412 | Cardiome | TNN | Q9UQP3 | 1 | 175067833 | 175148075 | 1 | 11826.56 | 0.99 | 0.88 | 1.10 | 0.799 | 0.998 |
| TNR | OID20957 | Neurology | TNR | Q92752 | 1 | 175315194 | 175743616 | 1 | 2869.90 | 0.99 | 0.77 | 1.28 | 0.962 | 0.998 |
| TPSAB1 | OID20642 | Inflammat | TPSAB1 | Q15661 | 16 | 1240379 | 1242554 | 1 | 10816.00 | 0.99 | 0.89 | 1.11 | 0.882 | 0.998 |
| TRIM5 | OID20526 | Inflammat | TRIM5 | Q9C035 | 11 | 5663195 | 5938619 | 1 | 1375.74 | 0.91 | 0.66 | 1.26 | 0.572 | 0.998 |
| TSPAN15 | OID30095 | Cardiome | TSPAN15 | O95858 | 10 | 69451465 | 69507666 | 1 | 49.00 | 0.75 | 0.14 | 4.07 | 0.742 | 0.998 |
| TSPAN8 | OID31470 | Oncology | TSPAN8 | P19075 | 12 | 71125085 | 71441898 | 1 | 4624.00 | 0.98 | 0.80 | 1.20 | 0.834 | 0.998 |
| TSPYL1 | OID30512 | Inflammat | TSPYL1 | Q9H0U9 | 6 | 116267760 | 116279930 | 1 | 188.60 | 1.31 | 0.53 | 3.25 | 0.563 | 0.998 |
| TST | OID20868 | Neurology | TST | Q16762 | 22 | 37010859 | 37020183 | 1 | 182.25 | 0.76 | 0.30 | 1.98 | 0.580 | 0.998 |
| TTF2 | OID30942 | Neurology | TTF2 | Q9UNY4 | 1 | 117060326 | 117107453 | 1 | 2126.84 | 0.99 | 0.74 | 1.31 | 0.920 | 0.998 |
| TXNDC5 | OID21012 | Neurology | TXNDC5 | Q8NBS9 | 6 | 7881517 | 7910788 | 1 | 173.36 | 0.75 | 0.29 | 1.91 | 0.543 | 0.998 |
| TXNDC9 | OID31121 | Neurology | TXNDC9 | O14530 | 2 | 99318982 | 99340702 | 1 | 46.02 | 0.77 | 0.13 | 4.76 | 0.780 | 0.998 |
| TYMP | OID20234 | Cardiome | TYMP | P19971 | 22 | 50525752 | 50530032 | 1 | 625.00 | 1.10 | 0.66 | 1.84 | 0.710 | 0.998 |
| TYRP1 | OID30517 | Inflammat | TYRP1 | P17643 | 9 | 12685439 | 12710285 | 1 | 102.52 | 0.87 | 0.26 | 2.95 | 0.826 | 0.998 |
| UBE2L6 | OID30321 | Cardiome | UBE2L6 | O14933 | 11 | 57551656 | 57568284 | 1 | 1008.06 | 1.04 | 0.68 | 1.57 | 0.866 | 0.998 |
| UNC5D | OID30447 | Inflammat | UNC5D | Q6UXZ4 | 8 | 35235475 | 35796550 | 1 | 64.00 | 0.80 | 0.18 | 3.54 | 0.767 | 0.998 |
| UROS | OID30964 | Neurology | UROS | P10746 | 10 | 125784980 | 125823258 | 1 | 235.11 | 0.89 | 0.39 | 2.07 | 0.794 | 0.998 |
| VCAM1 | OID20396 | Cardiome | VCAM1 | P19320 | 1 | 100719742 | 100739045 | 1 | 136.11 | 0.81 | 0.27 | 2.40 | 0.698 | 0.998 |
| VEGFB | OID30554 | Inflammat | VEGFB | P49765 | 11 | 64234584 | 64239264 | 1 | 163.27 | 1.19 | 0.43 | 3.34 | 0.738 | 0.998 |
| VGF | OID31067 | Neurology | VGF | O15240 | 7 | 101162509 | 101165569 | 1 | 69.44 | 1.36 | 0.29 | 6.30 | 0.697 | 0.998 |
| VNN1 | OID30711 | Inflammat | VNN1 | O95497 | 6 | 132680849 | 132714055 | 1 | 12370.38 | 0.97 | 0.87 | 1.08 | 0.612 | 0.998 |
| VSIG10 | OID31400 | Oncology | VSIG10 | Q8N0Z9 | 12 | 118063593 | 118136026 | 1 | 9481.89 | 0.99 | 0.87 | 1.13 | 0.892 | 0.998 |
| VSIR | OID20131 | Cardiome | VSIR | Q9H7M9 | 10 | 71747556 | 71773520 | 1 | 1600.00 | 0.95 | 0.70 | 1.29 | 0.738 | 0.998 |
| VSTM2L | OID20067 | Cardiome | VSTM2L | Q96N03 | 20 | 37903111 | 37945350 | 1 | 213.16 | 1.22 | 0.53 | 2.83 | 0.644 | 0.998 |
| VTCN1 | OID21164 | Oncology | VTCN1 | Q7Z7D3 | 1 | 117143587 | 117210960 | 1 | 96.40 | 0.69 | 0.20 | 2.36 | 0.556 | 0.998 |
| VWA1 | OID21421 | Oncology | VWA1 | Q6PCB0 | 1 | 1434861 | 1442882 | 1 | 2049.62 | 0.98 | 0.75 | 1.28 | 0.874 | 0.998 |
| VWC2 | OID20936 | Neurology | VWC2 | Q2TAL6 | 7 | 49773638 | 49921950 | 1 | 4280.90 | 0.95 | 0.77 | 1.17 | 0.631 | 0.998 |
| WARS | OID21084 | Neurology | WARS1 | P23381 | 14 | 100333790 | 100376805 | 1 | 821.78 | 0.96 | 0.64 | 1.44 | 0.832 | 0.998 |
| WASF1 | OID20091 | Cardiome | WASF1 | Q92558 | 6 | 110099819 | 110180004 | 1 | 1287.02 | 1.07 | 0.76 | 1.49 | 0.710 | 0.998 |
| WASHC3 | OID31123 | Neurology | WASHC3 | Q9Y3C0 | 12 | 102012840 | 102062149 | 1 | 121.00 | 1.18 | 0.37 | 3.74 | 0.781 | 0.998 |
| WFDC12 | OID21462 | Oncology | WFDC12 | Q8WWY7 | 20 | 45123425 | 45124465 | 1 | 3686.22 | 1.01 | 0.80 | 1.26 | 0.964 | 0.998 |
| WFDC2 | OID21505 | Oncology | WFDC2 | Q14508 | 20 | 45469753 | 45481532 | 1 | 114.33 | 0.69 | 0.20 | 2.36 | 0.549 | 0.998 |
| WNT9A | OID20460 | Inflammat | WNT9A | O14904 | 1 | 227918656 | 227947932 | 1 | 339.61 | 0.91 | 0.43 | 1.94 | 0.808 | 0.998 |
| WWP2 | OID20926 | Neurology | WWP2 | O00308 | 16 | 69762328 | 69941741 | 1 | 471.51 | 0.88 | 0.50 | 1.55 | 0.656 | 0.998 |
| ZBP1 | OID30526 | Inflammat | ZBP1 | Q9H171 | 20 | 57603846 | 57620576 | 1 | 1434.52 | 1.06 | 0.76 | 1.48 | 0.730 | 0.998 |
| ZFYVE19 | OID31452 | Oncology | ZFYVE19 | Q96K21 | 15 | 40807086 | 40815084 | 1 | 2185.56 | 1.07 | 0.82 | 1.39 | 0.626 | 0.998 |
| EBI3_IL27 | OID21389 | Oncology | EBI3;IL27 | Q14213;Q | 19;16 | 4229523;284 | 4237528;285 | 2 | 1384.01 | 1.01 | 0.79 | 1.29 | 0.948 | 0.998 |
| LAMA4 | OID20769 | Inflammat | LAMA4 | Q16363 | 6 | 112107931 | 112254939 | 1 | 435.77 | 0.99 | 0.56 | 1.75 | 0.970 | 0.998 |
| BOC | OID20116 | Cardiome | BOC | Q9BWV1 | 3 | 113211003 | 113287459 | 1 | 428.49 | 0.99 | 0.54 | 1.82 | 0.972 | 0.998 |
| CSF1 | OID20719 | Inflammat | CSF1 | P09603 | 1 | 109910242 | 109930992 | 1 | 713.65 | 0.99 | 0.60 | 1.65 | 0.976 | 0.998 |
| CSF2 | OID30072 | Cardiome | CSF2 | P04141 | 5 | 132073789 | 132076170 | 1 | 735.01 | 1.01 | 0.62 | 1.63 | 0.978 | 0.998 |
| FGFBP1 | OID21507 | Oncology | FGFBP1 | Q14512 | 4 | 15935577 | 15938740 | 1 | 180.75 | 0.99 | 0.37 | 2.62 | 0.978 | 0.998 |
| GGH | OID20416 | Cardiome | GGH | Q92820 | 8 | 63014881 | 63039407 | 1 | 2323.24 | 1.00 | 0.77 | 1.30 | 0.978 | 0.998 |
| GPA33 | OID21376 | Oncology | GPA33 | Q99795 | 1 | 167052836 | 167166479 | 1 | 548.21 | 1.01 | 0.57 | 1.79 | 0.975 | 0.998 |
| GSTA1 | OID20166 | Cardiome | GSTA1 | P08263 | 6 | 52791371 | 52803860 | 1 | 2025.00 | 1.00 | 0.74 | 1.36 | 0.976 | 0.998 |
| LYPD3 | OID21530 | Oncology | LYPD3 | O95274 | 19 | 43460787 | 43465608 | 1 | 1892.25 | 1.00 | 0.75 | 1.33 | 0.975 | 0.998 |
| NECTIN4 | OID21490 | Oncology | NECTIN4 | Q96NY8 | 1 | 161070998 | 161089558 | 1 | 1387.56 | 1.01 | 0.72 | 1.40 | 0.973 | 0.998 |
| PDCD5 | OID21021 | Neurology | PDCD5 | O14737 | 19 | 32581190 | 32587453 | 1 | 2304.00 | 1.00 | 0.76 | 1.30 | 0.977 | 0.998 |
| RILP | OID21436 | Oncology | RILP | Q96NA2 | 17 | 1646145 | 1649866 | 1 | 91.61 | 1.02 | 0.26 | 4.07 | 0.978 | 0.998 |
| STAT2 | OID30536 | Inflammat | STAT2 | P52630 | 12 | 56341597 | 56360203 | 1 | 370.23 | 1.01 | 0.53 | 1.93 | 0.976 | 0.998 |
| TAFA5 | OID21321 | Oncology | TAFA5 | Q7Z5A7 | 22 | 48489553 | 48850912 | 1 | 408.94 | 0.99 | 0.54 | 1.81 | 0.978 | 0.998 |
| TIMP4 | OID21147 | Neurology | TIMP4 | Q99727 | 3 | 12153068 | 12158912 | 1 | 1998.09 | 1.00 | 0.76 | 1.31 | 0.978 | 0.998 |
| ANGPTL3 | OID20407 | Cardiome | ANGPTL3 | Q9Y5C1 | 1 | 62597520 | 62606313 | 1 | 1716.33 | 1.00 | 0.71 | 1.41 | 0.981 | 0.999 |
| CD83 | OID20565 | Inflammat | CD83 | Q01151 | 6 | 14117256 | 14136918 | 1 | 493.83 | 0.99 | 0.56 | 1.75 | 0.980 | 0.999 |
| GSTP1 | OID20876 | Neurology | GSTP1 | P09211 | 11 | 67583742 | 67586656 | 1 | 656.64 | 1.01 | 0.63 | 1.60 | 0.981 | 0.999 |
| ST3GAL1 | OID21322 | Oncology | ST3GAL1 | Q11201 | 8 | 133454848 | 133571940 | 1 | 1146.31 | 1.00 | 0.67 | 1.49 | 0.981 | 0.999 |
| ART3 | OID20346 | Cardiome | ART3 | Q13508 | 4 | 76011184 | 76112802 | 1 | 1411.61 | 1.00 | 0.70 | 1.45 | 0.984 | 0.999 |
| FCGR3B | OID20387 | Cardiome | FCGR3B | O75015 | 1 | 161623196 | 161631963 | 1 | 11610.06 | 1.00 | 0.89 | 1.12 | 0.985 | 0.999 |
| PDIA5 | OID31209 | Oncology | PDIA5 | Q14554 | 3 | 123067025 | 123225227 | 1 | 968.77 | 1.00 | 0.68 | 1.47 | 0.985 | 0.999 |
| PRDX3 | OID20483 | Inflammat | PRDX3 | P30048 | 10 | 119167720 | 119178812 | 1 | 66.02 | 0.99 | 0.21 | 4.67 | 0.985 | 0.999 |
| SPTLC1 | OID31086 | Neurology | SPTLC1 | O15269 | 9 | 92000087 | 92115413 | 1 | 484.00 | 0.99 | 0.57 | 1.74 | 0.986 | 0.999 |
| ST13 | OID30624 | Inflammat | ST13 | P50502 | 22 | 40824535 | 40856639 | 1 | 59.51 | 1.02 | 0.18 | 5.82 | 0.985 | 0.999 |
| TNFRSF10B | OID20981 | Neurology | TNFRSF1 | O14763 | 8 | 23020133 | 23069031 | 1 | 2392.10 | 1.00 | 0.75 | 1.34 | 0.985 | 0.999 |

| | | | | | | | | | | | | | | | |
|---|---|---|---|---|---|---|---|---|---|---|---|---|---|---|---|
| CD7 | OID30543 | Inflammat | CD7 | P09564 | | 17 | 82314868 | 82317608 | 1 | 3549.65 | 1.00 | 0.80 | 1.24 | 0.987 | 1.000 |
| CILP | OID30399 | Cardiome | CILP | O75339 | | 15 | 65194760 | 65211473 | 1 | 1332.25 | 1.00 | 0.71 | 1.40 | 0.996 | 1.000 |
| CXADR | OID20561 | Inflammat | CXADR | P78310 | | 21 | 17513043 | 17593579 | 1 | 240.25 | 1.00 | 0.45 | 2.25 | 0.998 | 1.000 |
| DBH | OID30703 | Inflammat | DBH | P09172 | | 9 | 133636363 | 133659329 | 1 | 16900.00 | 1.00 | 0.91 | 1.10 | 0.999 | 1.000 |
| DNPEP | OID31118 | Neurology | DNPEP | Q9ULA0 | | 2 | 219372043 | 219400022 | 1 | 138.90 | 0.99 | 0.33 | 2.97 | 0.988 | 1.000 |
| EIF5 | OID30156 | Cardiome | EIF5 | P55010 | | 14 | 103333544 | 103345025 | 1 | 65.04 | 1.00 | 0.22 | 4.46 | 0.997 | 1.000 |
| ENTPD2 | OID21260 | Oncology | ENTPD2 | Q9Y5L3 | | 9 | 137048107 | 137054061 | 1 | 3057.68 | 1.00 | 0.80 | 1.26 | 0.995 | 1.000 |
| FCGR2A | OID20391 | Cardiome | FCGR2A | P12318 | | 1 | 161505430 | 161524013 | 1 | 15088.03 | 1.00 | 0.90 | 1.11 | 0.994 | 1.000 |
| FGF2 | OID20503 | Inflammat | FGF2 | P09038 | | 4 | 122826708 | 122898236 | 1 | 7898.77 | 1.00 | 0.87 | 1.15 | 0.991 | 1.000 |
| FHIT | OID20872 | Neurology | FHIT | P49789 | | 3 | 59747277 | 61251459 | 1 | 123.77 | 0.99 | 0.33 | 3.02 | 0.992 | 1.000 |
| HGF | OID20656 | Inflammat | HGF | P14210 | | 7 | 81699010 | 81770438 | 1 | 152.11 | 1.00 | 0.35 | 2.84 | 0.996 | 1.000 |
| IGSF9 | OID30330 | Cardiome | IGSF9 | Q9P2J2 | | 1 | 159927039 | 159945613 | 1 | 793.91 | 1.00 | 0.65 | 1.54 | 0.996 | 1.000 |
| MEGF10 | OID20746 | Inflammat | MEGF10 | Q96KG7 | | 5 | 127290796 | 127465737 | 1 | 3232.73 | 1.00 | 0.79 | 1.27 | 0.998 | 1.000 |
| MICALL2 | OID30889 | Neurology | MICALL2 | Q8IY33 | | 7 | 1428465 | 1459470 | 1 | 74.39 | 1.01 | 0.25 | 4.06 | 0.990 | 1.000 |
| NFATC1 | OID20545 | Inflammat | NFATC1 | O95644 | | 18 | 79395856 | 79529325 | 1 | 189.06 | 1.00 | 0.41 | 2.43 | 0.999 | 1.000 |
| NOS2 | OID30087 | Cardiome | NOS2 | P35228 | | 17 | 27756766 | 27800529 | 1 | 82.81 | 1.01 | 0.27 | 3.73 | 0.991 | 1.000 |
| PADI2 | OID20467 | Inflammat | PADI2 | Q9Y2J8 | | 1 | 17066761 | 17119451 | 1 | 232.56 | 1.00 | 0.45 | 2.24 | 1.000 | 1.000 |
| PDIA4 | OID30375 | Cardiome | PDIA4 | P13667 | | 7 | 149003062 | 149028662 | 1 | 136.89 | 0.99 | 0.35 | 2.84 | 0.989 | 1.000 |
| PI16 | OID30676 | Inflammat | PI16 | Q6UXB8 | | 6 | 36948263 | 36964837 | 1 | 1094.08 | 1.00 | 0.68 | 1.46 | 0.999 | 1.000 |
| PTPRB | OID30324 | Cardiome | PTPRB | P23467 | | 12 | 70515870 | 70637440 | 1 | 2104.52 | 1.00 | 0.76 | 1.32 | 0.996 | 1.000 |
| RGMA | OID21065 | Neurology | RGMA | Q96B86 | | 15 | 93035271 | 93089211 | 1 | 1051.61 | 1.00 | 0.66 | 1.52 | 0.999 | 1.000 |
| SPRY2 | OID20475 | Inflammat | SPRY2 | O43597 | | 13 | 80335976 | 80341126 | 1 | 105.06 | 1.00 | 0.31 | 3.31 | 0.994 | 1.000 |
| TACSTD2 | OID21447 | Oncology | TACSTD2 | P09758 | | 1 | 58575433 | 58577252 | 1 | 6798.75 | 1.00 | 0.87 | 1.15 | 0.997 | 1.000 |
| WASF3 | OID20882 | Neurology | WASF3 | Q9UPY6 | | 13 | 26557683 | 26688948 | 1 | 276.39 | 1.00 | 0.49 | 2.03 | 0.994 | 1.000 |
| A1BG | OID30771 | Inflammat | A1BG | P04217 | | 19 | 58345178 | 58353492 | 0 | - | - | - | - | - | - |
| ABL1 | OID21280 | Oncology | ABL1 | P00519 | | 9 | 130713016 | 130887675 | 0 | - | - | - | - | - | - |
| ABRAXAS2 | OID30130 | Cardiome | ABRAXAS | Q15018 | | 10 | 124801819 | 124836667 | 0 | - | - | - | - | - | - |
| ACE2 | OID20105 | Cardiome | ACE2 | Q9BYF1 | X | | 15494566 | 15607236 | 0 | - | - | - | - | - | - |
| ACOT13 | OID31522 | Oncology | ACOT13 | Q9NPJ3 | | 6 | 24667035 | 24705065 | 0 | - | - | - | - | - | - |
| ACSL1 | OID30080 | Cardiome | ACSL1 | P33121 | | 4 | 184755595 | 184826818 | 0 | - | - | - | - | - | - |
| ACTN2 | OID30192 | Cardiome | ACTN2 | P35609 | | 1 | 236664141 | 236764631 | 0 | - | - | - | - | - | - |
| ACTN4 | OID20435 | Inflammat | ACTN4 | O43707 | | 19 | 38647649 | 38731589 | 0 | - | - | - | - | - | - |
| ACY1 | OID20137 | Cardiome | ACY1 | Q03154 | | 3 | 51983340 | 51989197 | 0 | - | - | - | - | - | - |
| ADAM9 | OID31488 | Oncology | ADAM9 | Q13443 | | 8 | 38996754 | 39105445 | 0 | - | - | - | - | - | - |
| ADAMTS1 | OID30528 | Inflammat | ADAMTS1 | Q9UHI8 | | 21 | 26835755 | 26845409 | 0 | - | - | - | - | - | - |
| ADAMTSL4 | OID30275 | Cardiome | ADAMTSL | Q6UY14 | | 1 | 150549369 | 150560937 | 0 | - | - | - | - | - | - |
| ADCYAP1R1 | OID21188 | Oncology | ADCYAP1 | P41586 | | 7 | 31052308 | 31111479 | 0 | - | - | - | - | - | - |
| ADGRF5 | OID30349 | Cardiome | ADGRF5 | Q8IZF2 | | 6 | 46852522 | 46954943 | 0 | - | - | - | - | - | - |
| ADGRG2 | OID20189 | Cardiome | ADGRG2 | Q8IZP9 | X | | 18989307 | 19122637 | 0 | - | - | - | - | - | - |
| ADGRV1 | OID30877 | Neurology | ADGRV1 | Q8WXG9 | | 5 | 90529344 | 91164437 | 0 | - | - | - | - | - | - |
| ADH1B | OID30612 | Inflammat | ADH1B | P00325 | | 4 | 99304971 | 99352760 | 0 | - | - | - | - | - | - |
| ADRA2A | OID30067 | Cardiome | ADRA2A | P08913 | | 10 | 111077029 | 111080907 | 0 | - | - | - | - | - | - |
| AFM | OID30741 | Inflammat | AFM | P43652 | | 4 | 73481745 | 73504001 | 0 | - | - | - | - | - | - |
| AGBL2 | OID30793 | Neurology | AGBL2 | Q5U5Z8 | | 11 | 47659591 | 47715389 | 0 | - | - | - | - | - | - |
| AGR3 | OID21243 | Oncology | AGR3 | Q8TD06 | | 7 | 16859412 | 16881987 | 0 | - | - | - | - | - | - |
| AGRP | OID20658 | Inflammat | AGRP | O00253 | | 16 | 67482571 | 67483547 | 0 | - | - | - | - | - | - |
| AHSA1 | OID31379 | Oncology | AHSA1 | O95433 | | 14 | 77457870 | 77469472 | 0 | - | - | - | - | - | - |
| AHSP | OID21078 | Neurology | AHSP | Q9NZD4 | | 16 | 31527900 | 31528803 | 0 | - | - | - | - | - | - |
| AIDA | OID30956 | Neurology | AIDA | Q96BJ3 | | 1 | 222668013 | 222713210 | 0 | - | - | - | - | - | - |
| AIFM1 | OID21287 | Oncology | AIFM1 | O95831 | X | | 130124666 | 130165879 | 0 | - | - | - | - | - | - |
| AK1 | OID20122 | Cardiome | AK1 | P00568 | | 9 | 127866486 | 127877675 | 0 | - | - | - | - | - | - |
| AK2 | OID31142 | Neurology | AK2 | P54819 | | 1 | 33007986 | 33080996 | 0 | - | - | - | - | - | - |
| AKAP12 | OID30571 | Inflammat | AKAP12 | Q02952 | | 6 | 151239967 | 151358559 | 0 | - | - | - | - | - | - |
| AKR1B10 | OID30955 | Neurology | AKR1B10 | O60218 | | 7 | 134527567 | 134541412 | 0 | - | - | - | - | - | - |
| AKT1S1 | OID20895 | Neurology | AKT1S1 | Q96B36 | | 19 | 49869033 | 49878459 | 0 | - | - | - | - | - | - |
| AKT2 | OID31052 | Neurology | AKT2 | P31751 | | 19 | 40230317 | 40285536 | 0 | - | - | - | - | - | - |
| ALDH2 | OID31380 | Oncology | ALDH2 | P05091 | | 12 | 111766887 | 111817532 | 0 | - | - | - | - | - | - |
| ALMS1 | OID31275 | Oncology | ALMS1 | Q8TCU4 | | 2 | 73385758 | 73625166 | 0 | - | - | - | - | - | - |
| ALPI | OID30497 | Inflammat | ALPI | P09923 | | 2 | 232456125 | 232460753 | 0 | - | - | - | - | - | - |
| AMBP | OID21413 | Oncology | AMBP | P02760 | | 9 | 114060127 | 114078328 | 0 | - | - | - | - | - | - |
| AMDHD2 | OID30366 | Cardiome | AMDHD2 | Q9Y303 | | 16 | 2520357 | 2531422 | 0 | - | - | - | - | - | - |
| AMFR | OID20806 | Neurology | AMFR | Q9UKV5 | | 16 | 56361452 | 56425545 | 0 | - | - | - | - | - | - |
| AMIGO1 | OID30881 | Neurology | AMIGO1 | Q86WK6 | | 1 | 109504178 | 109509738 | 0 | - | - | - | - | - | - |
| AMOT | OID30572 | Inflammat | AMOT | Q4VCS5 | X | | 112774503 | 112840831 | 0 | - | - | - | - | - | - |
| AMPD3 | OID31139 | Neurology | AMPD3 | Q01432 | | 11 | 10308313 | 10507579 | 0 | - | - | - | - | - | - |
| ANGPTL2 | OID20726 | Inflammat | ANGPTL2 | Q9UKU9 | | 9 | 127087348 | 127122635 | 0 | - | - | - | - | - | - |
| ANGPTL7 | OID21412 | Oncology | ANGPTL7 | O43827 | | 1 | 11189355 | 11195981 | 0 | - | - | - | - | - | - |
| ANK2 | OID30161 | Cardiome | ANK2 | Q01484 | | 4 | 112818032 | 113384221 | 0 | - | - | - | - | - | - |
| ANKRA2 | OID31173 | Oncology | ANKRA2 | Q9H9E1 | | 5 | 73552190 | 73565667 | 0 | - | - | - | - | - | - |
| ANP32C | OID30203 | Cardiome | ANP32C | O43423 | | 4 | 164197007 | 16419771 | 0 | - | - | - | - | - | - |
| ANXA1 | OID30617 | Inflammat | ANXA1 | P04083 | | 9 | 73151865 | 73170393 | 0 | - | - | - | - | - | - |
| ANXA10 | OID20886 | Neurology | ANXA10 | Q9UJ72 | | 4 | 168092537 | 168187736 | 0 | - | - | - | - | - | - |
| ANXA3 | OID21036 | Neurology | ANXA3 | P12429 | | 4 | 78551747 | 78610451 | 0 | - | - | - | - | - | - |
| ANXA4 | OID20123 | Cardiome | ANXA4 | P09525 | | 2 | 69644425 | 69827112 | 0 | - | - | - | - | - | - |
| AP2B1 | OID30949 | Neurology | AP2B1 | P63010 | | 17 | 35578046 | 35726413 | 0 | - | - | - | - | - | - |
| AP3B1 | OID31478 | Oncology | AP3B1 | O00203 | | 5 | 78000522 | 78294762 | 0 | - | - | - | - | - | - |
| AP3S2 | OID31117 | Neurology | AP3S2 | P59780 | | 15 | 89830599 | 89894638 | 0 | - | - | - | - | - | - |
| APLP1 | OID20164 | Cardiome | APLP1 | P51693 | | 19 | 35867899 | 35879792 | 0 | - | - | - | - | - | - |
| APOB | OID30673 | Inflammat | APOB | P04114 | | 2 | 21001429 | 21044073 | 0 | - | - | - | - | - | - |
| APRT | OID20927 | Neurology | APRT | P07741 | | 16 | 88809339 | 88811937 | 0 | - | - | - | - | - | - |
| ARAF | OID31260 | Oncology | ARAF | P10398 | X | | 47561205 | 47571908 | 0 | - | - | - | - | - | - |
| ARF6 | OID31523 | Oncology | ARF6 | P62330 | | 14 | 49893079 | 49897054 | 0 | - | - | - | - | - | - |
| ARG2 | OID31339 | Oncology | ARG2 | P78540 | | 14 | 67619920 | 67651708 | 0 | - | - | - | - | - | - |
| ARHGAP1 | OID21202 | Oncology | ARHGAP | Q07960 | | 11 | 46677080 | 46700619 | 0 | - | - | - | - | - | - |
| ARHGAP30 | OID31336 | Oncology | ARHGAP | Q7Z6I6 | | 1 | 161046946 | 161069970 | 0 | - | - | - | - | - | - |
| ARHGAP5 | OID31190 | Oncology | ARHGAP | Q13017 | | 14 | 32076114 | 32159728 | 0 | - | - | - | - | - | - |
| ARHGEF1 | OID31133 | Neurology | ARHGEF | Q92888 | | 19 | 41883173 | 41930150 | 0 | - | - | - | - | - | - |
| ARHGEF12 | OID20554 | Inflammat | ARHGEF | Q9NZN5 | | 11 | 120336413 | 120489937 | 0 | - | - | - | - | - | - |
| ARID3A | OID30861 | Neurology | ARID3A | Q99856 | | 19 | 925781 | 975939 | 0 | - | - | - | - | - | - |
| ARID4B | OID20803 | Neurology | ARID4B | Q4LE39 | | 1 | 235131634 | 235328219 | 0 | - | - | - | - | - | - |
| ARL13B | OID30166 | Cardiome | ARL13B | Q3SXY8 | | 3 | 93980139 | 94055678 | 0 | - | - | - | - | - | - |
| ARMCX2 | OID30792 | Neurology | ARMCX2 | Q7L311 | X | | 101655281 | 101659850 | 0 | - | - | - | - | - | - |
| ARNT | OID20432 | Inflammat | ARNT | P27540 | | 1 | 150809713 | 150876708 | 0 | - | - | - | - | - | - |
| ARNTL | OID30168 | Cardiome | ARNTL | O00327 | | 11 | 13276652 | 13387266 | 0 | - | - | - | - | - | - |
| ARTN | OID20446 | Inflammat | ARTN | Q5T4W7 | | 1 | 43933320 | 43937240 | 0 | - | - | - | - | - | - |
| ASS1 | OID30170 | Cardiome | ASS1 | P00966 | | 9 | 130444961 | 130501274 | 0 | - | - | - | - | - | - |
| ATF2 | OID20798 | Neurology | ATF2 | P15336 | | 2 | 175072250 | 175168382 | 0 | - | - | - | - | - | - |
| ATF4 | OID31162 | Oncology | ATF4 | P18848 | | 22 | 39519695 | 39522683 | 0 | - | - | - | - | - | - |
| ATG16L1 | OID31466 | Oncology | ATG16L1 | Q676U5 | | 2 | 233210051 | 233295674 | 0 | - | - | - | - | - | - |
| ATG4A | OID21246 | Oncology | ATG4A | Q8WYN0 | X | | 108091668 | 108154671 | 0 | - | - | - | - | - | - |
| ATOX1 | OID21476 | Oncology | ATOX1 | O00244 | | 5 | 151742316 | 151772532 | 0 | - | - | - | - | - | - |
| ATP1B1 | OID30124 | Cardiome | ATP1B1 | P05026 | | 1 | 169105697 | 169310992 | 0 | - | - | - | - | - | - |
| ATP1B2 | OID30118 | Cardiome | ATP1B2 | P14415 | | 17 | 7646627 | 7657770 | 0 | - | - | - | - | - | - |
| ATP1B3 | OID30065 | Cardiome | ATP1B3 | P54709 | | 3 | 141876124 | 141926549 | 0 | - | - | - | - | - | - |
| ATP1B4 | OID31230 | Oncology | ATP1B4 | Q9UN42 | X | | 120362085 | 120383249 | 0 | - | - | - | - | - | - |
| ATP2B4 | OID30062 | Cardiome | ATP2B4 | P23634 | | 1 | 203626832 | 203744081 | 0 | - | - | - | - | - | - |
| ATP5F1D | OID30079 | Cardiome | ATP5F1D | P30049 | | 19 | 1241746 | 1244825 | 0 | - | - | - | - | - | - |

| | | | | | | | | | | | | | | | |
|---|---|---|---|---|---|---|---|---|---|---|---|---|---|---|---|
| ATP5PO | OID20811 | Neurology | ATP5PO | P48047 | | 21 | 33903453 | 33915814 | 0 | - | - | - | - | - | - |
| ATP6AP2 | OID21178 | Oncology | ATP6AP2 | O75787 | X | | 40579372 | 40606848 | 0 | - | - | - | - | - | - |
| ATP6V1D | OID21341 | Oncology | ATP6V1D | Q9Y5K8 | | 14 | 67294371 | 67360265 | 0 | - | - | - | - | - | - |
| ATP6V1F | OID20843 | Neurology | ATP6V1F | Q16864 | | 7 | 128862856 | 128865847 | 0 | - | - | - | - | - | - |
| ATP6V1G1 | OID30242 | Cardiome | ATP6V1G | O75348 | | 9 | 114587706 | 114598915 | 0 | - | - | - | - | - | - |
| ATXN2 | OID30850 | Neurology | ATXN2 | Q99700 | | 12 | 111443485 | 111599676 | 0 | - | - | - | - | - | - |
| ATXN2L | OID30980 | Neurology | ATXN2L | Q8WWM | | 16 | 28822999 | 28837237 | 0 | - | - | - | - | - | - |
| AXIN1 | OID20582 | Inflammat | AXIN1 | O15169 | | 16 | 287440 | 352723 | 0 | - | - | - | - | - | - |
| AZI2 | OID31056 | Neurology | AZI2 | Q9H6S1 | | 3 | 28315003 | 28349050 | 0 | - | - | - | - | - | - |
| B2M | OID30725 | Inflammat | B2M | P61769 | | 15 | 44711358 | 44718851 | 0 | - | - | - | - | - | - |
| B3GAT3 | OID30165 | Cardiome | B3GAT3 | O94766 | | 11 | 62615296 | 62622154 | 0 | - | - | - | - | - | - |
| BABAM1 | OID30466 | Inflammat | BABAM1 | Q9NWV8 | | 19 | 17267376 | 17281249 | 0 | - | - | - | - | - | - |
| BACH1 | OID20505 | Inflammat | BACH1 | O14867 | | 21 | 29194071 | 29630751 | 0 | - | - | - | - | - | - |
| BAG4 | OID30474 | Inflammat | BAG4 | O95429 | | 8 | 38176533 | 38213301 | 0 | - | - | - | - | - | - |
| BAG6 | OID20114 | Cardiome | BAG6 | P46379 | | 6 | 31639028 | 31652705 | 0 | - | - | - | - | - | - |
| BAMBI | OID21267 | Oncology | BAMBI | Q13145 | | 10 | 28677510 | 28682932 | 0 | - | - | - | - | - | - |
| BATF | OID30896 | Neurology | BATF | Q16520 | | 14 | 75522455 | 75546993 | 0 | - | - | - | - | - | - |
| BAX | OID20856 | Neurology | BAX | Q07812 | | 19 | 48954815 | 48961798 | 0 | - | - | - | - | - | - |
| BCAT2 | OID30844 | Neurology | BCAT2 | O15382 | | 19 | 48795062 | 48811029 | 0 | - | - | - | - | - | - |
| BCL2 | OID31454 | Oncology | BCL2 | P10415 | | 18 | 63123346 | 63320128 | 0 | - | - | - | - | - | - |
| BCL2L1 | OID31152 | Neurology | BCL2L1 | Q07817 | | 20 | 31664452 | 31723989 | 0 | - | - | - | - | - | - |
| BCL2L11 | OID20513 | Inflammat | BCL2L11 | O43521 | | 2 | 111119378 | 111168445 | 0 | - | - | - | - | - | - |
| BCL7A | OID31218 | Oncology | BCL7A | Q4VC05 | | 12 | 122019422 | 122062044 | 0 | - | - | - | - | - | - |
| BCL7B | OID31421 | Oncology | BCL7B | Q9BQE9 | | 7 | 73536356 | 73557690 | 0 | - | - | - | - | - | - |
| BCR | OID20558 | Inflammat | BCR | P11274 | | 22 | 23179704 | 23318037 | 0 | - | - | - | - | - | - |
| BDNF | OID30373 | Cardiome | BDNF | P23560 | | 11 | 27654893 | 27722058 | 0 | - | - | - | - | - | - |
| BECN1 | OID30220 | Cardiome | BECN1 | Q14457 | | 17 | 42810134 | 42833350 | 0 | - | - | - | - | - | - |
| BEX3 | OID31202 | Oncology | BEX3 | Q00994 | X | | 103376395 | 103378164 | 0 | - | - | - | - | - | - |
| BGN | OID21198 | Oncology | BGN | P21810 | X | | 153494980 | 153509546 | 0 | - | - | - | - | - | - |
| BHLHE40 | OID30873 | Neurology | BHLHE40 | O14503 | | 3 | 4979437 | 4985323 | 0 | - | - | - | - | - | - |
| BHMT2 | OID30051 | Cardiome | BHMT2 | Q9H2M3 | | 5 | 79069767 | 79090069 | 0 | - | - | - | - | - | - |
| BID | OID20517 | Inflammat | BID | P55957 | | 22 | 17734138 | 17774770 | 0 | - | - | - | - | - | - |
| BIRC2 | OID21217 | Oncology | BIRC2 | Q13490 | | 11 | 102347211 | 102378670 | 0 | - | - | - | - | - | - |
| BLOC1S2 | OID30856 | Neurology | BLOC1S2 | Q6QNY1 | | 10 | 100273280 | 100286680 | 0 | - | - | - | - | - | - |
| BLOC1S3 | OID31106 | Neurology | BLOC1S3 | Q6QNY0 | | 19 | 45178784 | 45216933 | 0 | - | - | - | - | - | - |
| BLVRB | OID21141 | Neurology | BLVRB | P30043 | | 19 | 40447765 | 40465764 | 0 | - | - | - | - | - | - |
| BMP4 | OID20994 | Neurology | BMP4 | P12644 | | 14 | 53949736 | 53958761 | 0 | - | - | - | - | - | - |
| BNIP2 | OID30292 | Cardiome | BNIP2 | Q12982 | | 15 | 59659146 | 59689339 | 0 | - | - | - | - | - | - |
| BOLA2_BOLA | OID30267 | Cardiome | BOLA2B;I | Q9H3K6;( | 16;16 | | 30192932;2 | 30194306;2 | 0 | - | - | - | - | - | - |
| BRAP | OID31506 | Oncology | BRAP | Q7Z569 | | 12 | 111642146 | 111685956 | 0 | - | - | - | - | - | - |
| BRD1 | OID31259 | Oncology | BRD1 | O95696 | | 22 | 49773283 | 49827873 | 0 | - | - | - | - | - | - |
| BRD2 | OID30826 | Neurology | BRD2 | P25440 | | 6 | 32968594 | 32981505 | 0 | - | - | - | - | - | - |
| BRD3 | OID30093 | Cardiome | BRD3 | Q15059 | | 9 | 134030305 | 134068535 | 0 | - | - | - | - | - | - |
| BRDT | OID30179 | Cardiome | BRDT | Q58F21 | | 1 | 91949343 | 92014426 | 0 | - | - | - | - | - | - |
| BRK1 | OID20883 | Neurology | BRK1 | Q8WUW1 | | 3 | 10115675 | 10127190 | 0 | - | - | - | - | - | - |
| BRME1 | OID30847 | Neurology | BRME1 | Q0VDD7 | | 19 | 13882348 | 13906452 | 0 | - | - | - | - | - | - |
| BSND | OID31184 | Oncology | BSND | Q8WZ55 | | 1 | 54998933 | 55017172 | 0 | - | - | - | - | - | - |
| BTLA | OID31237 | Oncology | BTLA | Q7Z6A9 | | 3 | 112463966 | 112499472 | 0 | - | - | - | - | - | - |
| BTNL10 | OID31196 | Oncology | BTNL10 | A8MVZ5 | | 1 | 228510425 | 228512305 | 0 | - | - | - | - | - | - |
| C19orf12 | OID20804 | Neurology | C19orf12 | Q9NSK7 | | 19 | 29698937 | 29715789 | 0 | - | - | - | - | - | - |
| C1GALT1C1 | OID30180 | Cardiome | C1GALT1 | Q96EU7 | X | | 120625674 | 120630054 | 0 | - | - | - | - | - | - |
| C1QBP | OID31007 | Neurology | C1QBP | Q07021 | | 17 | 5432777 | 5448830 | 0 | - | - | - | - | - | - |
| C2 | OID20410 | Cardiome | C2 | P06681 | | 6 | 31897785 | 31945672 | 0 | - | - | - | - | - | - |
| C9 | OID30698 | Inflammat | C9 | P02748 | | 5 | 39284140 | 39371324 | 0 | - | - | - | - | - | - |
| C9orf40 | OID31489 | Oncology | C9orf40 | Q8IXQ3 | | 9 | 74946583 | 74952912 | 0 | - | - | - | - | - | - |
| CA7 | OID31265 | Oncology | CA7 | P43166 | | 16 | 66844414 | 66854153 | 0 | - | - | - | - | - | - |
| CA8 | OID30482 | Inflammat | CA8 | P35219 | | 8 | 60185412 | 60281400 | 0 | - | - | - | - | - | - |
| CA9 | OID21417 | Oncology | CA9 | Q16790 | | 9 | 35673928 | 35681159 | 0 | - | - | - | - | - | - |
| CABP2 | OID30916 | Neurology | CABP2 | Q9NPB3 | | 11 | 67518912 | 67524517 | 0 | - | - | - | - | - | - |
| CACNA1C | OID30840 | Neurology | CACNA1( | Q13936 | | 12 | 1970772 | 2697950 | 0 | - | - | - | - | - | - |
| CACNA1H | OID30064 | Cardiome | CACNA1H | O95180 | | 16 | 1153106 | 1221771 | 0 | - | - | - | - | - | - |
| CACNB1 | OID30820 | Neurology | CACNB1 | Q02641 | | 17 | 39173453 | 39197702 | 0 | - | - | - | - | - | - |
| CADPS | OID30915 | Neurology | CADPS | Q9ULU8 | | 3 | 62398346 | 62875416 | 0 | - | - | - | - | - | - |
| CALB1 | OID21306 | Oncology | CALB1 | P05937 | | 8 | 90058608 | 90095475 | 0 | - | - | - | - | - | - |
| CALY | OID30144 | Cardiome | CALY | Q9NYX4 | | 10 | 133324072 | 133336935 | 0 | - | - | - | - | - | - |
| CAMLG | OID30952 | Neurology | CAMLG | P49069 | | 5 | 134738495 | 134752157 | 0 | - | - | - | - | - | - |
| CAMSAP1 | OID31070 | Neurology | CAMSAP1 | Q5T5Y3 | | 9 | 135808487 | 135907546 | 0 | - | - | - | - | - | - |
| CAPN3 | OID31384 | Oncology | CAPN3 | P20807 | | 15 | 42359498 | 42412949 | 0 | - | - | - | - | - | - |
| CAPS | OID31126 | Neurology | CAPS | Q13938 | | 19 | 5912339 | 5916211 | 0 | - | - | - | - | - | - |
| CASC3 | OID30921 | Neurology | CASC3 | O15234 | | 17 | 40140318 | 40172171 | 0 | - | - | - | - | - | - |
| CASP1 | OID20854 | Neurology | CASP1 | P29466 | | 11 | 105025443 | 105035250 | 0 | - | - | - | - | - | - |
| CASP2 | OID20555 | Inflammat | CASP2 | P42575 | | 7 | 143288215 | 143307696 | 0 | - | - | - | - | - | - |
| CASP4 | OID31276 | Oncology | CASP4 | P49662 | | 11 | 104942866 | 104969366 | 0 | - | - | - | - | - | - |
| CASP7 | OID31135 | Neurology | CASP7 | P55210 | | 10 | 113679162 | 113730907 | 0 | - | - | - | - | - | - |
| CASQ2 | OID30063 | Cardiome | CASQ2 | O14958 | | 1 | 115700021 | 115768714 | 0 | - | - | - | - | - | - |
| CBLN1 | OID30851 | Neurology | CBLN1 | P23435 | | 16 | 49277917 | 49281838 | 0 | - | - | - | - | - | - |
| CBX2 | OID30177 | Cardiome | CBX2 | Q14781 | | 17 | 79778148 | 79787983 | 0 | - | - | - | - | - | - |
| CCAR2 | OID31077 | Neurology | CCAR2 | Q8N163 | | 8 | 22604757 | 22620964 | 0 | - | - | - | - | - | - |
| CCDC28A | OID30973 | Neurology | CCDC28A | Q8IWP9 | | 6 | 138773769 | 138793319 | 0 | - | - | - | - | - | - |
| CCER2 | OID30221 | Cardiome | CCER2 | I3L3R5 | | 19 | 38908980 | 38912186 | 0 | - | - | - | - | - | - |
| CCL22 | OID20765 | Inflammat | CCL22 | O00626 | | 16 | 57358783 | 57366189 | 0 | - | - | - | - | - | - |
| CCL26 | OID20546 | Inflammat | CCL26 | Q9Y258 | | 7 | 75769533 | 75789896 | 0 | - | - | - | - | - | - |
| CCND2 | OID31316 | Oncology | CCND2 | P30279 | | 12 | 4269771 | 4305353 | 0 | - | - | - | - | - | - |
| CCNE1 | OID30456 | Inflammat | CCNE1 | P24864 | | 19 | 29811991 | 29824312 | 0 | - | - | - | - | - | - |
| CCT5 | OID21200 | Oncology | CCT5 | P48643 | | 5 | 10249929 | 10266389 | 0 | - | - | - | - | - | - |
| CD1C | OID21483 | Oncology | CD1C | P29017 | | 1 | 158289923 | 158294774 | 0 | - | - | - | - | - | - |
| CD2 | OID30097 | Cardiome | CD2 | P06729 | | 1 | 116754430 | 116769229 | 0 | - | - | - | - | - | - |
| CD244 | OID20628 | Inflammat | CD244 | Q9BZW8 | | 1 | 160830160 | 160862887 | 0 | - | - | - | - | - | - |
| CD248 | OID30394 | Cardiome | CD248 | Q9HCU0 | | 11 | 66314494 | 66317044 | 0 | - | - | - | - | - | - |
| CD3D | OID30827 | Neurology | CD3D | P04234 | | 11 | 118339075 | 118342705 | 0 | - | - | - | - | - | - |
| CD3E | OID31302 | Oncology | CD3E | P07766 | | 11 | 118304730 | 118316175 | 0 | - | - | - | - | - | - |
| CD3G | OID30448 | Inflammat | CD3G | P09693 | | 11 | 118344344 | 118355161 | 0 | - | - | - | - | - | - |
| CD40LG | OID20616 | Inflammat | CD40LG | P29965 | X | | 136648158 | 136660390 | 0 | - | - | - | - | - | - |
| CD63 | OID20988 | Neurology | CD63 | P08962 | | 12 | 55725323 | 55729707 | 0 | - | - | - | - | - | - |
| CD82 | OID31256 | Oncology | CD82 | P27701 | | 11 | 44564427 | 44620358 | 0 | - | - | - | - | - | - |
| CD86 | OID31500 | Oncology | CD86 | P42081 | | 3 | 122055362 | 122121139 | 0 | - | - | - | - | - | - |
| CD93 | OID20397 | Cardiome | CD93 | Q9NPY3 | | 20 | 23079360 | 23086324 | 0 | - | - | - | - | - | - |
| CD99 | OID21110 | Neurology | CD99 | P14209 | X | | 2691187 | 2741309 | 0 | - | - | - | - | - | - |
| CD99L2 | OID21122 | Neurology | CD99L2 | Q8TCZ2 | X | | 150766336 | 150898816 | 0 | - | - | - | - | - | - |
| CDAN1 | OID31076 | Neurology | CDAN1 | Q8IWY9 | | 15 | 42723544 | 42737128 | 0 | - | - | - | - | - | - |
| CDC123 | OID31242 | Oncology | CDC123 | O75794 | | 10 | 12195965 | 12250589 | 0 | - | - | - | - | - | - |
| CDC25A | OID31189 | Oncology | CDC25A | P30304 | | 3 | 48157146 | 48188417 | 0 | - | - | - | - | - | - |
| CDC26 | OID31508 | Oncology | CDC26 | Q8NHZ8 | | 9 | 113255835 | 113275572 | 0 | - | - | - | - | - | - |
| CDC37 | OID21470 | Oncology | CDC37 | Q16543 | | 19 | 10391133 | 10420121 | 0 | - | - | - | - | - | - |
| CDC42BPB | OID31485 | Oncology | CDC42BF | Q9Y5S2 | | 14 | 102932380 | 103057549 | 0 | - | - | - | - | - | - |
| CDH1 | OID20353 | Cardiome | CDH1 | P12830 | | 16 | 68737292 | 68835537 | 0 | - | - | - | - | - | - |
| CDH22 | OID31332 | Oncology | CDH22 | Q9UJ99 | | 20 | 46173739 | 46308498 | 0 | - | - | - | - | - | - |

| | | | | | | | | | | | | | | | |
|---|---|---|---|---|---|---|---|---|---|---|---|---|---|---|---|
| CDH3 | OID21008 | Neurology | CDH3 | P22223 | | 16 | 68636189 | 68727468 | 0 | - | - | - | - | - | - |
| CDH4 | OID30875 | Neurology | CDH4 | P55283 | | 20 | 61252261 | 61940617 | 0 | - | - | - | - | - | - |
| CDK1 | OID31164 | Oncology | CDK1 | P06493 | | 10 | 60778331 | 60794852 | 0 | - | - | - | - | - | - |
| CDK5RAP3 | OID30985 | Neurology | CDK5RAF | Q96JB5 | | 17 | 47967810 | 47981781 | 0 | - | - | - | - | - | - |
| CDKL5 | OID30796 | Neurology | CDKL5 | O76039 | X | | 18425583 | 18653629 | 0 | - | - | - | - | - | - |
| CDKN2D | OID21295 | Oncology | CDKN2D | P55273 | | 19 | 10566460 | 10569059 | 0 | - | - | - | - | - | - |
| CEACAM18 | OID31251 | Oncology | CEACAM | A8MTB9 | | 19 | 51478539 | 51491301 | 0 | - | - | - | - | - | - |
| CEACAM3 | OID21245 | Oncology | CEACAM: | P40198 | | 19 | 41796587 | 41811554 | 0 | - | - | - | - | - | - |
| CEBPA | OID30458 | Inflammat | CEBPA | P49715 | | 19 | 33299934 | 33302534 | 0 | - | - | - | - | - | - |
| CEBPB | OID20052 | Cardiome | CEBPB | P17676 | | 20 | 50190830 | 50192668 | 0 | - | - | - | - | - | - |
| CEND1 | OID30821 | Neurology | CEND1 | Q8N111 | | 11 | 787115 | 790113 | 0 | - | - | - | - | - | - |
| CENPF | OID31329 | Oncology | CENPF | P49454 | | 1 | 214603195 | 214664571 | 0 | - | - | - | - | - | - |
| CENPJ | OID31314 | Oncology | CENPJ | Q9HC77 | | 13 | 24882279 | 24922889 | 0 | - | - | - | - | - | - |
| CEP164 | OID20441 | Inflammat | CEP164 | Q9UPV0 | | 11 | 117314557 | 117413266 | 0 | - | - | - | - | - | - |
| CEP290 | OID31287 | Oncology | CEP290 | O15078 | | 12 | 88049016 | 88142099 | 0 | - | - | - | - | - | - |
| CEP350 | OID31247 | Oncology | CEP350 | Q5VT06 | | 1 | 179954674 | 180114880 | 0 | - | - | - | - | - | - |
| CES2 | OID21366 | Oncology | CES2 | O00748 | | 16 | 66934444 | 66945096 | 0 | - | - | - | - | - | - |
| CETN2 | OID20836 | Neurology | CETN2 | P41208 | X | | 152826994 | 152830757 | 0 | - | - | - | - | - | - |
| CFC1 | OID21261 | Oncology | CFC1 | P0CG37 | | 2 | 130592165 | 130599575 | 0 | - | - | - | - | - | - |
| CFP | OID30784 | Inflammat | CFP | P27918 | X | | 47623172 | 47630305 | 0 | - | - | - | - | - | - |
| CGA | OID21069 | Neurology | CGA | P01215 | | 6 | 87085498 | 87095106 | 0 | - | - | - | - | - | - |
| CGN | OID30894 | Neurology | CGN | Q9P2M7 | | 1 | 151510510 | 151538692 | 0 | - | - | - | - | - | - |
| CHAD | OID30660 | Inflammat | CHAD | O15335 | | 17 | 50464496 | 50468906 | 0 | - | - | - | - | - | - |
| CHEK2 | OID20054 | Cardiome | CHEK2 | O96017 | | 22 | 28687743 | 28742422 | 0 | - | - | - | - | - | - |
| CHM | OID30972 | Neurology | CHM | P24386 | X | | 85861180 | 86047561 | 0 | - | - | - | - | - | - |
| CHP1 | OID30869 | Neurology | CHP1 | Q99653 | | 15 | 41230839 | 41281887 | 0 | - | - | - | - | - | - |
| CHRDL1 | OID20771 | Inflammat | CHRDL1 | Q9BU40 | X | | 110673856 | 110795819 | 0 | - | - | - | - | - | - |
| CHRM1 | OID30930 | Neurology | CHRM1 | P11229 | | 11 | 62908679 | 62921807 | 0 | - | - | - | - | - | - |
| CINP | OID31328 | Oncology | CINP | Q9BW66 | | 14 | 102341102 | 102362916 | 0 | - | - | - | - | - | - |
| CIRBP | OID31503 | Oncology | CIRBP | Q14011 | | 19 | 1259384 | 1274880 | 0 | - | - | - | - | - | - |
| CKB | OID30400 | Cardiome | CKB | P12277 | | 14 | 103519667 | 103522833 | 0 | - | - | - | - | - | - |
| CLASP1 | OID31192 | Oncology | CLASP1 | Q7Z460 | | 2 | 121337776 | 121649476 | 0 | - | - | - | - | - | - |
| CLC | OID20126 | Cardiome | CLC | Q05315 | | 19 | 39731255 | 39738029 | 0 | - | - | - | - | - | - |
| CLEC2L | OID30884 | Neurology | CLEC2L | P0C7M8 | | 7 | 139523685 | 139544985 | 0 | - | - | - | - | - | - |
| CLINT1 | OID31182 | Oncology | CLINT1 | Q14677 | | 5 | 157785743 | 157859145 | 0 | - | - | - | - | - | - |
| CLNS1A | OID30929 | Neurology | CLNS1A | P54105 | | 11 | 77514936 | 77637794 | 0 | - | - | - | - | - | - |
| CLPP | OID20910 | Neurology | CLPP | Q16740 | | 19 | 6361531 | 6370242 | 0 | - | - | - | - | - | - |
| CLSPN | OID20831 | Neurology | CLSPN | Q9HAW4 | | 1 | 35720218 | 35769978 | 0 | - | - | - | - | - | - |
| CLSTN1 | OID20874 | Neurology | CLSTN1 | O94985 | | 1 | 9728926 | 9823984 | 0 | - | - | - | - | - | - |
| CLSTN2 | OID20664 | Inflammat | CLSTN2 | Q9H4D0 | | 3 | 139935185 | 140577397 | 0 | - | - | - | - | - | - |
| CLTA | OID20118 | Cardiome | CLTA | P09496 | | 9 | 36190856 | 36304781 | 0 | - | - | - | - | - | - |
| CMIP | OID31406 | Oncology | CMIP | Q8IY22 | | 16 | 81444808 | 81711762 | 0 | - | - | - | - | - | - |
| CNGB3 | OID30805 | Neurology | CNGB3 | Q9NQW8 | | 8 | 86553977 | 86743675 | 0 | - | - | - | - | - | - |
| CNPY2 | OID20069 | Cardiome | CNPY2 | Q9Y2B0 | | 12 | 56309842 | 56316119 | 0 | - | - | - | - | - | - |
| CNST | OID20127 | Cardiome | CNST | Q6PJW8 | | 1 | 246566444 | 246668595 | 0 | - | - | - | - | - | - |
| CNTF | OID30807 | Neurology | CNTF | P26441 | | 11 | 58622665 | 58625733 | 0 | - | - | - | - | - | - |
| CNTNAP4 | OID30911 | Neurology | CNTNAP4 | Q9C0A0 | | 16 | 76277278 | 76560757 | 0 | - | - | - | - | - | - |
| COL1A1 | OID20335 | Cardiome | COL1A1 | P02452 | | 17 | 50184101 | 50201632 | 0 | - | - | - | - | - | - |
| COL3A1 | OID30379 | Cardiome | COL3A1 | P02461 | | 2 | 188974373 | 189012746 | 0 | - | - | - | - | - | - |
| COL4A4 | OID31333 | Oncology | COL4A4 | P53420 | | 2 | 227002714 | 227164453 | 0 | - | - | - | - | - | - |
| COL9A1 | OID20550 | Inflammat | COL9A1 | P20849 | | 6 | 70216040 | 70303084 | 0 | - | - | - | - | - | - |
| COL9A2 | OID31255 | Oncology | COL9A2 | Q14055 | | 1 | 40300489 | 40317813 | 0 | - | - | - | - | - | - |
| COLEC12 | OID20738 | Inflammat | COLEC12 | Q5KU26 | | 18 | 316737 | 500722 | 0 | - | - | - | - | - | - |
| COMMD9 | OID31390 | Oncology | COMMD9 | Q9P000 | | 11 | 36269284 | 36289449 | 0 | - | - | - | - | - | - |
| COPB2 | OID30173 | Cardiome | COPB2 | P35606 | | 3 | 139353946 | 139389736 | 0 | - | - | - | - | - | - |
| COPE | OID20797 | Neurology | COPE | O14579 | | 19 | 18899514 | 18919387 | 0 | - | - | - | - | - | - |
| CORO1A | OID20286 | Cardiome | CORO1A | P31146 | | 16 | 30182827 | 30189076 | 0 | - | - | - | - | - | - |
| CORO6 | OID30870 | Neurology | CORO6 | Q6QEF8 | | 17 | 29614756 | 29622912 | 0 | - | - | - | - | - | - |
| COX5B | OID21297 | Oncology | COX5B | P10606 | | 2 | 97646062 | 97648383 | 0 | - | - | - | - | - | - |
| COX6B1 | OID30317 | Cardiome | COX6B1 | P14854 | | 19 | 35648323 | 35658782 | 0 | - | - | - | - | - | - |
| CPLX2 | OID31186 | Oncology | CPLX2 | Q6PUV4 | | 5 | 175796310 | 175884021 | 0 | - | - | - | - | - | - |
| CPQ | OID30406 | Cardiome | CPQ | Q9Y646 | | 8 | 96645242 | 97149654 | 0 | - | - | - | - | - | - |
| CPTP | OID30106 | Cardiome | CPTP | Q5TA50 | | 1 | 1324756 | 1328896 | 0 | - | - | - | - | - | - |
| CREB3 | OID31188 | Oncology | CREB3 | O43889 | | 9 | 35732598 | 35736999 | 0 | - | - | - | - | - | - |
| CREBZF | OID31206 | Oncology | CREBZF | Q9NS37 | | 11 | 85657742 | 85682908 | 0 | - | - | - | - | - | - |
| CRHR1 | OID20055 | Cardiome | CRHR1 | P34998 | | 17 | 45784280 | 45835828 | 0 | - | - | - | - | - | - |
| CRKL | OID20742 | Inflammat | CRKL | P46109 | | 22 | 20917407 | 20953747 | 0 | - | - | - | - | - | - |
| CRLF1 | OID20699 | Inflammat | CRLF1 | O75462 | | 19 | 18572220 | 18607741 | 0 | - | - | - | - | - | - |
| CRTAP | OID30902 | Neurology | CRTAP | O75718 | | 3 | 33114014 | 33147773 | 0 | - | - | - | - | - | - |
| CRX | OID20070 | Cardiome | CRX | O43186 | | 19 | 47819779 | 47843330 | 0 | - | - | - | - | - | - |
| CRYBB2 | OID31081 | Neurology | CRYBB2 | P43320 | | 22 | 25212564 | 25231870 | 0 | - | - | - | - | - | - |
| CSDE1 | OID30183 | Cardiome | CSDE1 | O75534 | | 1 | 114716913 | 114758676 | 0 | - | - | - | - | - | - |
| CSF1R | OID30681 | Inflammat | CSF1R | P07333 | | 5 | 150053291 | 150113372 | 0 | - | - | - | - | - | - |
| CSF2RA | OID20915 | Neurology | CSF2RA | P15509 | X | | 1268800 | 1310381 | 0 | - | - | - | - | - | - |
| CSH1 | OID30667 | Inflammat | CSH1 | P0DML2 | | 17 | 63894909 | 63896661 | 0 | - | - | - | - | - | - |
| CSNK1D | OID30492 | Inflammat | CSNK1D | P48730 | | 17 | 82239023 | 82273700 | 0 | - | - | - | - | - | - |
| CSNK2A1 | OID31053 | Neurology | CSNK2A1 | P68400 | | 20 | 472498 | 543835 | 0 | - | - | - | - | - | - |
| CSPG5 | OID30843 | Neurology | CSPG5 | O95196 | | 3 | 47562238 | 47580792 | 0 | - | - | - | - | - | - |
| CSRP3 | OID30176 | Cardiome | CSRP3 | P50461 | | 11 | 19182030 | 19210571 | 0 | - | - | - | - | - | - |
| CTAG1A_CT/ | OID31299 | Oncology | CTAG1A; | P78358;P | X;X | | 154585133;1 | 154586816;1 | 0 | - | - | - | - | - | - |
| CTLA4 | OID30363 | Cardiome | CTLA4 | P16410 | | 2 | 203853888 | 203873965 | 0 | - | - | - | - | - | - |
| CTNNA1 | OID30800 | Neurology | CTNNA1 | P35221 | | 5 | 138610967 | 138935034 | 0 | - | - | - | - | - | - |
| CTSZ | OID20365 | Cardiome | CTSZ | Q9UBR2 | | 20 | 58985686 | 59008238 | 0 | - | - | - | - | - | - |
| CUZD1 | OID30914 | Neurology | CUZD1 | Q86UP6 | | 10 | 122832158 | 122846175 | 0 | - | - | - | - | - | - |
| CXCL10 | OID20697 | Inflammat | CXCL10 | P02778 | | 4 | 76021118 | 76023497 | 0 | - | - | - | - | - | - |
| CXCL3 | OID20788 | Inflammat | CXCL3 | P19876 | | 4 | 74036589 | 74038807 | 0 | - | - | - | - | - | - |
| CYB5A | OID31171 | Oncology | CYB5A | P00167 | | 18 | 74250846 | 74291973 | 0 | - | - | - | - | - | - |
| CYP24A1 | OID30091 | Cardiome | CYP24A1 | Q07973 | | 20 | 54153446 | 54173986 | 0 | - | - | - | - | - | - |
| CYTH3 | OID31208 | Oncology | CYTH3 | O43739 | | 7 | 6161776 | 6272644 | 0 | - | - | - | - | - | - |
| DAAM1 | OID30664 | Inflammat | DAAM1 | Q9Y4D1 | | 14 | 59188646 | 59371405 | 0 | - | - | - | - | - | - |
| DAB2 | OID21428 | Oncology | DAB2 | P98082 | | 5 | 39371675 | 39462300 | 0 | - | - | - | - | - | - |
| DAND5 | OID30420 | Inflammat | DAND5 | Q8N907 | | 19 | 12965159 | 12974760 | 0 | - | - | - | - | - | - |
| DARS1 | OID20903 | Neurology | DARS1 | P14868 | | 2 | 135905881 | 135986100 | 0 | - | - | - | - | - | - |
| DBN1 | OID30549 | Inflammat | DBN1 | Q16643 | | 5 | 177456608 | 177474401 | 0 | - | - | - | - | - | - |
| DCDC2C | OID31235 | Oncology | DCDC2C | A8MYV0 | | 2 | 3703575 | 3848008 | 0 | - | - | - | - | - | - |
| DCLRE1C | OID31240 | Oncology | DCLRE1C | Q96SD1 | | 10 | 14897359 | 14954432 | 0 | - | - | - | - | - | - |
| DCN | OID20106 | Cardiome | DCN | P07585 | | 12 | 91140484 | 91183217 | 0 | - | - | - | - | - | - |
| DCTD | OID30657 | Inflammat | DCTD | P32321 | | 4 | 182890060 | 182917936 | 0 | - | - | - | - | - | - |
| DCTN1 | OID21399 | Oncology | DCTN1 | Q14203 | | 2 | 74361154 | 74392087 | 0 | - | - | - | - | - | - |
| DCTN2 | OID21334 | Oncology | DCTN2 | Q13561 | | 12 | 57529633 | 57547224 | 0 | - | - | - | - | - | - |
| DCTN6 | OID20879 | Neurology | DCTN6 | O00399 | | 8 | 30156319 | 30183639 | 0 | - | - | - | - | - | - |
| DCUN1D1 | OID30830 | Neurology | DCUN1D1 | Q96GG9 | | 3 | 182938074 | 182985953 | 0 | - | - | - | - | - | - |
| DCUN1D2 | OID31231 | Oncology | DCUN1D2 | Q6PH85 | | 13 | 113455819 | 113490951 | 0 | - | - | - | - | - | - |
| DCXR | OID21480 | Oncology | DCXR | Q7Z4W1 | | 17 | 82035136 | 82037709 | 0 | - | - | - | - | - | - |
| DDA1 | OID30126 | Cardiome | DDA1 | Q9BW61 | | 19 | 17309518 | 17323298 | 0 | - | - | - | - | - | - |
| DDX1 | OID31518 | Oncology | DDX1 | Q92499 | | 2 | 15591178 | 15634346 | 0 | - | - | - | - | - | - |
| DDX25 | OID30892 | Neurology | DDX25 | Q9UHL0 | | 11 | 125903348 | 125943702 | 0 | - | - | - | - | - | - |

| | | | | | | | | | | | | | | |
|---|---|---|---|---|---|---|---|---|---|---|---|---|---|---|
| DDX39A | OID30494 | Inflammat | DDX39A | O00148 | 19 | 14408798 | 14419383 | 0 | - | - | - | - | - | - |
| DDX4 | OID30431 | Inflammat | DDX4 | Q9NQI0 | 5 | 55738017 | 55817157 | 0 | - | - | - | - | - | - |
| DDX53 | OID30944 | Neurology | DDX53 | Q86TM3 | X | 22999960 | 23003589 | 0 | - | - | - | - | - | - |
| DECR1 | OID20579 | Inflammat | DECR1 | Q16698 | 8 | 90001405 | 90053633 | 0 | - | - | - | - | - | - |
| DEFB103A_D | OID30534 | Inflammat | DEFB103 | P81534;P | 8;8 | 7428888;78 | 7430348;78 | 0 | - | - | - | - | - | - |
| DEFB116 | OID30823 | Neurology | DEFB116 | Q30KQ4 | 20 | 31303212 | 31308585 | 0 | - | - | - | - | - | - |
| DEFB118 | OID30858 | Neurology | DEFB118 | Q96PH6 | 20 | 31368601 | 31373923 | 0 | - | - | - | - | - | - |
| DENND2B | OID30050 | Cardiome | DENND2B | P78524 | 11 | 8693351 | 8910951 | 0 | - | - | - | - | - | - |
| DENR | OID30493 | Inflammat | DENR | O43583 | 12 | 122752824 | 122771064 | 0 | - | - | - | - | - | - |
| DFFA | OID20620 | Inflammat | DFFA | O00273 | 1 | 10456522 | 10472529 | 0 | - | - | - | - | - | - |
| DGKA | OID30546 | Inflammat | DGKA | P23743 | 12 | 55927319 | 55954027 | 0 | - | - | - | - | - | - |
| DGKZ | OID20466 | Inflammat | DGKZ | Q13574 | 11 | 46332905 | 46380554 | 0 | - | - | - | - | - | - |
| DHODH | OID30137 | Cardiome | DHODH | Q02127 | 16 | 72008588 | 72027664 | 0 | - | - | - | - | - | - |
| DHPS | OID31357 | Oncology | DHPS | P49366 | 19 | 12675717 | 12681880 | 0 | - | - | - | - | - | - |
| DIABLO | OID20184 | Cardiome | DIABLO | Q9NR28 | 12 | 122207663 | 122227456 | 0 | - | - | - | - | - | - |
| DIPK1C | OID30932 | Neurology | DIPK1C | Q0P6D2 | 18 | 74434775 | 74457944 | 0 | - | - | - | - | - | - |
| DIPK2B | OID30582 | Inflammat | DIPK2B | Q9H7Y0 | X | 45148373 | 45200901 | 0 | - | - | - | - | - | - |
| DLG4 | OID31071 | Neurology | DLG4 | P78352 | 17 | 7187187 | 7219841 | 0 | - | - | - | - | - | - |
| DLGAP5 | OID30976 | Neurology | DLGAP5 | Q15398 | 14 | 55148112 | 55191608 | 0 | - | - | - | - | - | - |
| DLL4 | OID30255 | Cardiome | DLL4 | Q9NR61 | 15 | 40929340 | 40939073 | 0 | - | - | - | - | - | - |
| DMD | OID30223 | Cardiome | DMD | P11532 | X | 31097677 | 33339609 | 0 | - | - | - | - | - | - |
| DMP1 | OID30213 | Cardiome | DMP1 | Q13316 | 4 | 87650280 | 87664361 | 0 | - | - | - | - | - | - |
| DNAJA1 | OID31204 | Oncology | DNAJA1 | P31689 | 9 | 33025273 | 33039907 | 0 | - | - | - | - | - | - |
| DNAJA2 | OID20627 | Inflammat | DNAJA2 | O60884 | 16 | 46955362 | 46973674 | 0 | - | - | - | - | - | - |
| DNAJB14 | OID31434 | Oncology | DNAJB14 | Q8TBM8 | 4 | 99896248 | 99946618 | 0 | - | - | - | - | - | - |
| DNAJB2 | OID30661 | Inflammat | DNAJB2 | P25686 | 2 | 219279342 | 219286898 | 0 | - | - | - | - | - | - |
| DNAJB8 | OID20093 | Cardiome | DNAJB8 | Q8NHS0 | 3 | 128462437 | 128466968 | 0 | - | - | - | - | - | - |
| DNAJC9 | OID31472 | Oncology | DNAJC9 | Q8WXX5 | 10 | 73183362 | 73247255 | 0 | - | - | - | - | - | - |
| DNLZ | OID30822 | Neurology | DNLZ | Q5SXM8 | 9 | 136359480 | 136363744 | 0 | - | - | - | - | - | - |
| DNM3 | OID31000 | Neurology | DNM3 | Q9UQ16 | 1 | 171817887 | 172418466 | 0 | - | - | - | - | - | - |
| DOCK9 | OID30836 | Neurology | DOCK9 | Q9BZ29 | 13 | 98793429 | 99086625 | 0 | - | - | - | - | - | - |
| DOK1 | OID31111 | Neurology | DOK1 | Q99704 | 2 | 74549026 | 74557551 | 0 | - | - | - | - | - | - |
| DPEP2 | OID21305 | Oncology | DPEP2 | Q9H4A9 | 16 | 67987394 | 68000586 | 0 | - | - | - | - | - | - |
| DRG2 | OID21335 | Oncology | DRG2 | P55039 | 17 | 18087892 | 18107970 | 0 | - | - | - | - | - | - |
| DSG4 | OID21325 | Oncology | DSG4 | Q86SJ6 | 18 | 31376777 | 31414912 | 0 | - | - | - | - | - | - |
| DTNB | OID31395 | Oncology | DTNB | O60941 | 2 | 25377198 | 25673647 | 0 | - | - | - | - | - | - |
| DTX2 | OID30882 | Neurology | DTX2 | Q86UW9 | 7 | 76461676 | 76505995 | 0 | - | - | - | - | - | - |
| DUOX2 | OID20059 | Cardiome | DUOX2 | Q9NRD8 | 15 | 45092650 | 45114172 | 0 | - | - | - | - | - | - |
| DUSP3 | OID20827 | Neurology | DUSP3 | P51452 | 17 | 43766125 | 43778977 | 0 | - | - | - | - | - | - |
| DUT | OID31457 | Oncology | DUT | P33316 | 15 | 48331011 | 48343373 | 0 | - | - | - | - | - | - |
| DYNC1H1 | OID31341 | Oncology | DYNC1H1 | Q14204 | 14 | 101964573 | 102056443 | 0 | - | - | - | - | - | - |
| DYNLT3 | OID31415 | Oncology | DYNLT3 | P51808 | X | 37836757 | 37847571 | 0 | - | - | - | - | - | - |
| ECE1 | OID20891 | Neurology | ECE1 | P42892 | 1 | 21217247 | 21345572 | 0 | - | - | - | - | - | - |
| ECSCR | OID30831 | Neurology | ECSCR | Q19T08 | 5 | 139448560 | 139462743 | 0 | - | - | - | - | - | - |
| EDA2R | OID21451 | Oncology | EDA2R | Q9HAV5 | X | 66595637 | 66639298 | 0 | - | - | - | - | - | - |
| EDEM2 | OID30136 | Cardiome | EDEM2 | Q9BV94 | 20 | 35115364 | 35147336 | 0 | - | - | - | - | - | - |
| EDF1 | OID31010 | Neurology | EDF1 | O60869 | 9 | 136862119 | 136866308 | 0 | - | - | - | - | - | - |
| EDNRB | OID30421 | Inflammat | EDNRB | P24530 | 13 | 77895481 | 77975529 | 0 | - | - | - | - | - | - |
| EEF1D | OID30335 | Cardiome | EEF1D | P29692 | 8 | 143579697 | 143599541 | 0 | - | - | - | - | - | - |
| EFCAB14 | OID31460 | Oncology | EFCAB14 | O75071 | 1 | 46674659 | 46719146 | 0 | - | - | - | - | - | - |
| EFNA4 | OID20935 | Neurology | EFNA4 | P52798 | 1 | 155063737 | 155069553 | 0 | - | - | - | - | - | - |
| EFNB2 | OID30954 | Neurology | EFNB2 | P52799 | 13 | 106489745 | 106535662 | 0 | - | - | - | - | - | - |
| EIF1AX | OID31125 | Neurology | EIF1AX | P47813 | X | 20124525 | 20141838 | 0 | - | - | - | - | - | - |
| EIF2S2 | OID30270 | Cardiome | EIF2S2 | P20042 | 20 | 34088309 | 34112243 | 0 | - | - | - | - | - | - |
| EIF4B | OID21000 | Neurology | EIF4B | P23588 | 12 | 53006282 | 53042215 | 0 | - | - | - | - | - | - |
| EIF4E | OID30557 | Inflammat | EIF4E | P06730 | 4 | 98879276 | 98929133 | 0 | - | - | - | - | - | - |
| EIF4EBP1 | OID20162 | Cardiome | EIF4EBP1 | Q13541 | 8 | 38030534 | 38060365 | 0 | - | - | - | - | - | - |
| EIF4G1 | OID20625 | Inflammat | EIF4G1 | Q04637 | 3 | 184314495 | 184335358 | 0 | - | - | - | - | - | - |
| EIF5A | OID20423 | Inflammat | EIF5A | P63241 | 17 | 7306999 | 7312463 | 0 | - | - | - | - | - | - |
| ELAC1 | OID31029 | Neurology | ELAC1 | Q9H777 | 18 | 50967991 | 50988121 | 0 | - | - | - | - | - | - |
| ELAVL4 | OID30854 | Neurology | ELAVL4 | P26378 | 1 | 50024029 | 50203772 | 0 | - | - | - | - | - | - |
| ELOB | OID30127 | Cardiome | ELOB | Q15370 | 16 | 2771414 | 2777289 | 0 | - | - | - | - | - | - |
| ENO1 | OID21037 | Neurology | ENO1 | P06733 | 1 | 8861000 | 8879190 | 0 | - | - | - | - | - | - |
| ENOPH1 | OID31349 | Oncology | ENOPH1 | Q9UHY7 | 4 | 82430590 | 82461177 | 0 | - | - | - | - | - | - |
| ENOX2 | OID30152 | Cardiome | ENOX2 | Q16206 | X | 130622325 | 130903234 | 0 | - | - | - | - | - | - |
| EP300 | OID30438 | Inflammat | EP300 | Q09472 | 22 | 41092592 | 41180077 | 0 | - | - | - | - | - | - |
| EPB41L5 | OID30886 | Neurology | EPB41L5 | Q9HCM4 | 2 | 120013077 | 120179119 | 0 | - | - | - | - | - | - |
| EPCAM | OID20743 | Inflammat | EPCAM | P16422 | 2 | 47345158 | 47387601 | 0 | - | - | - | - | - | - |
| EPGN | OID31373 | Oncology | EPGN | Q6UW88 | 4 | 74308470 | 74316789 | 0 | - | - | - | - | - | - |
| EPHA10 | OID20840 | Neurology | EPHA10 | Q5JZY3 | 1 | 37713880 | 37765133 | 0 | - | - | - | - | - | - |
| EPN1 | OID31252 | Oncology | EPN1 | Q9Y6I3 | 19 | 55675226 | 55709858 | 0 | - | - | - | - | - | - |
| EPO | OID20522 | Inflammat | EPO | P01588 | 7 | 100720468 | 100723700 | 0 | - | - | - | - | - | - |
| ERBB3 | OID20705 | Inflammat | ERBB3 | P21860 | 12 | 56076799 | 56103505 | 0 | - | - | - | - | - | - |
| ERBIN | OID21323 | Oncology | ERBIN | Q96RT1 | 5 | 65926556 | 66082546 | 0 | - | - | - | - | - | - |
| ERCC1 | OID31223 | Oncology | ERCC1 | P07992 | 19 | 45407334 | 45478828 | 0 | - | - | - | - | - | - |
| ERP29 | OID30578 | Inflammat | ERP29 | P30040 | 12 | 112013348 | 112023449 | 0 | - | - | - | - | - | - |
| ERP44 | OID21338 | Oncology | ERP44 | Q9BS26 | 9 | 99979185 | 100099052 | 0 | - | - | - | - | - | - |
| ERVV-1 | OID30094 | Cardiome | ERVV-1 | B6SEH8 | 19 | 53013921 | 53016123 | 0 | - | - | - | - | - | - |
| ESAM | OID20270 | Cardiome | ESAM | Q96AP7 | 11 | 124752583 | 124762290 | 0 | - | - | - | - | - | - |
| ESPL1 | OID31176 | Oncology | ESPL1 | Q14674 | 12 | 53268299 | 53293638 | 0 | - | - | - | - | - | - |
| ESR1 | OID30434 | Inflammat | ESR1 | P03372 | 6 | 151656691 | 152129619 | 0 | - | - | - | - | - | - |
| EVI2B | OID31227 | Oncology | EVI2B | P34910 | 17 | 31303770 | 31314105 | 0 | - | - | - | - | - | - |
| EVPL | OID31309 | Oncology | EVPL | Q92817 | 17 | 76004502 | 76027306 | 0 | - | - | - | - | - | - |
| EXOSC10 | OID30083 | Cardiome | EXOSC10 | Q01780 | 1 | 11066618 | 11099869 | 0 | - | - | - | - | - | - |
| F9 | OID20349 | Cardiome | F9 | P00740 | X | 139530739 | 139563459 | 0 | - | - | - | - | - | - |
| FAM171A2 | OID31005 | Neurology | FAM171A | A8MVW0 | 17 | 44353215 | 44363853 | 0 | - | - | - | - | - | - |
| FARSA | OID30887 | Neurology | FARSA | Q9Y285 | 19 | 12922479 | 12934037 | 0 | - | - | - | - | - | - |
| FBLN2 | OID30378 | Cardiome | FBLN2 | P98095 | 3 | 13549125 | 13638422 | 0 | - | - | - | - | - | - |
| FDX1 | OID30078 | Cardiome | FDX1 | P10109 | 11 | 110429948 | 110464884 | 0 | - | - | - | - | - | - |
| FDX2 | OID30147 | Cardiome | FDX2 | Q6P4F2 | 19 | 10310045 | 10316015 | 0 | - | - | - | - | - | - |
| FEN1 | OID21192 | Oncology | FEN1 | P39748 | 11 | 61792911 | 61797238 | 0 | - | - | - | - | - | - |
| FETUB | OID20404 | Cardiome | FETUB | Q9UGM5 | 3 | 186635969 | 186653141 | 0 | - | - | - | - | - | - |
| FGA | OID30788 | Inflammat | FGA | P02671 | 4 | 154583128 | 154590745 | 0 | - | - | - | - | - | - |
| FGD3 | OID31040 | Neurology | FGD3 | Q5JSP0 | 9 | 92947523 | 93036236 | 0 | - | - | - | - | - | - |
| FGF12 | OID30454 | Inflammat | FGF12 | P61328 | 3 | 192139390 | 192767764 | 0 | - | - | - | - | - | - |
| FGF16 | OID30535 | Inflammat | FGF16 | O43320 | X | 77447389 | 77457278 | 0 | - | - | - | - | - | - |
| FGF19 | OID20715 | Inflammat | FGF19 | O95750 | 11 | 69698238 | 69704022 | 0 | - | - | - | - | - | - |
| FGF20 | OID30459 | Inflammat | FGF20 | Q9NP95 | 8 | 16992181 | 17002345 | 0 | - | - | - | - | - | - |
| FGF3 | OID30544 | Inflammat | FGF3 | P11487 | 11 | 69809968 | 69819416 | 0 | - | - | - | - | - | - |
| FGF6 | OID30443 | Inflammat | FGF6 | P10767 | 12 | 4428155 | 4445815 | 0 | - | - | - | - | - | - |
| FGF7 | OID31241 | Oncology | FGF7 | P21781 | 15 | 49423237 | 49488775 | 0 | - | - | - | - | - | - |
| FGF9 | OID31193 | Oncology | FGF9 | P31371 | 13 | 21671073 | 21704498 | 0 | - | - | - | - | - | - |
| FH | OID30839 | Neurology | FH | P07954 | 1 | 241497511 | 241519799 | 0 | - | - | - | - | - | - |
| FHIP2A | OID30187 | Cardiome | FHIP2A | Q5W0V3 | 10 | 114821744 | 114899832 | 0 | - | - | - | - | - | - |
| FKBP14 | OID31065 | Neurology | FKBP14 | Q9NWM8 | 7 | 30010587 | 30026702 | 0 | - | - | - | - | - | - |
| FKBP7 | OID20846 | Neurology | FKBP7 | Q9Y680 | 2 | 178463664 | 178478600 | 0 | - | - | - | - | - | - |
| FKBPL | OID31499 | Oncology | FKBPL | Q9UIM3 | 6 | 32128707 | 32130288 | 0 | - | - | - | - | - | - |

| | | | | | | | | | | | | | | | |
|---|---|---|---|---|---|---|---|---|---|---|---|---|---|---|---|
| FLI1 | OID21239 | Oncology | FLI1 | Q01543 | | 11 | 128686535 | 128813267 | 0 | - | - | - | - | - | - |
| FLT1 | OID21301 | Oncology | FLT1 | P17948 | | 13 | 28300346 | 28495145 | 0 | - | - | - | - | - | - |
| FLT3LG | OID20661 | Inflammat | FLT3LG | P49771 | | 19 | 49474207 | 49486231 | 0 | - | - | - | - | - | - |
| FMNL1 | OID20884 | Neurology | FMNL1 | O95466 | | 17 | 45221444 | 45247319 | 0 | - | - | - | - | - | - |
| FMR1 | OID21233 | Oncology | FMR1 | Q06787 | X | | 147911919 | 147951125 | 0 | - | - | - | - | - | - |
| FNTA | OID31381 | Oncology | FNTA | P49354 | | 8 | 43034194 | 43085788 | 0 | - | - | - | - | - | - |
| FOS | OID31221 | Oncology | FOS | P01100 | | 14 | 75278826 | 75282230 | 0 | - | - | - | - | - | - |
| FOSB | OID20791 | Neurology | FOSB | P53539 | | 19 | 45467995 | 45475179 | 0 | - | - | - | - | - | - |
| FOXO3 | OID21271 | Oncology | FOXO3 | O43524 | | 6 | 108559835 | 108684774 | 0 | - | - | - | - | - | - |
| FRMD4B | OID30155 | Cardiome | FRMD4B | Q9Y2L6 | | 3 | 69168782 | 69542583 | 0 | - | - | - | - | - | - |
| FRMD7 | OID30898 | Neurology | FRMD7 | Q6ZUT3 | X | | 132076990 | 132128020 | 0 | - | - | - | - | - | - |
| FTCD | OID31137 | Neurology | FTCD | O95954 | | 21 | 46136160 | 46155579 | 0 | - | - | - | - | - | - |
| FUS | OID21256 | Oncology | FUS | P35637 | | 16 | 31180138 | 31191605 | 0 | - | - | - | - | - | - |
| FUT1 | OID31295 | Oncology | FUT1 | P19526 | | 19 | 48748011 | 48755390 | 0 | - | - | - | - | - | - |
| FYB1 | OID21140 | Neurology | FYB1 | O15117 | | 5 | 39105252 | 39274528 | 0 | - | - | - | - | - | - |
| FZD10 | OID30931 | Neurology | FZD10 | Q9ULW2 | | 12 | 130162459 | 130165740 | 0 | - | - | - | - | - | - |
| FZD8 | OID30818 | Neurology | FZD8 | Q9H461 | | 10 | 35638247 | 35642296 | 0 | - | - | - | - | - | - |
| GABARAP | OID31414 | Oncology | GABARAI | O95166 | | 17 | 7240008 | 7242449 | 0 | - | - | - | - | - | - |
| GABARAPL1 | OID31253 | Oncology | GABARAI | Q9H0R8 | | 12 | 10212458 | 10223128 | 0 | - | - | - | - | - | - |
| GABRA4 | OID30803 | Neurology | GABRA4 | P48169 | | 4 | 46918900 | 46993581 | 0 | - | - | - | - | - | - |
| GAD1 | OID31166 | Oncology | GAD1 | Q99259 | | 2 | 170813213 | 170861151 | 0 | - | - | - | - | - | - |
| GAD2 | OID30449 | Inflammat | GAD2 | Q05329 | | 10 | 26216665 | 26304558 | 0 | - | - | - | - | - | - |
| GADD45B | OID31261 | Oncology | GADD45E | O75293 | | 19 | 2476122 | 2478259 | 0 | - | - | - | - | - | - |
| GADD45GIP1 | OID30237 | Cardiome | GADD45C | Q8TAE8 | | 19 | 12953119 | 12957223 | 0 | - | - | - | - | - | - |
| GAGE2A | OID31199 | Oncology | GAGE2A | Q6NT46 | X | | 49589496 | 49596824 | 0 | - | - | - | - | - | - |
| GALNT7 | OID21377 | Oncology | GALNT7 | Q86SF2 | | 4 | 173168811 | 173323967 | 0 | - | - | - | - | - | - |
| GAMT | OID30133 | Cardiome | GAMT | Q14353 | | 19 | 1397026 | 1401570 | 0 | - | - | - | - | - | - |
| GAPDH | OID30600 | Inflammat | GAPDH | P04406 | | 12 | 6534512 | 6538374 | 0 | - | - | - | - | - | - |
| GAST | OID31083 | Neurology | GAST | P01350 | | 17 | 41712331 | 41715969 | 0 | - | - | - | - | - | - |
| GATA3 | OID31219 | Oncology | GATA3 | P23771 | | 10 | 8045378 | 8075198 | 0 | - | - | - | - | - | - |
| GBA | OID30966 | Neurology | GBA | P04062 | | 1 | 155234452 | 155244699 | 0 | - | - | - | - | - | - |
| GBP6 | OID30139 | Cardiome | GBP6 | Q6ZN66 | | 1 | 89364059 | 89388160 | 0 | - | - | - | - | - | - |
| GCC1 | OID31147 | Neurology | GCC1 | Q96CN9 | | 7 | 127580628 | 127593611 | 0 | - | - | - | - | - | - |
| GCG | OID21263 | Oncology | GCG | P01275 | | 2 | 162142882 | 162152404 | 0 | - | - | - | - | - | - |
| GDF2 | OID20202 | Cardiome | GDF2 | Q9UK05 | | 10 | 47322454 | 47327588 | 0 | - | - | - | - | - | - |
| GET3 | OID30303 | Cardiome | GET3 | O43681 | | 19 | 12737139 | 12748323 | 0 | - | - | - | - | - | - |
| GFOD2 | OID21216 | Oncology | GFOD2 | Q3B7J2 | | 16 | 67674531 | 67719339 | 0 | - | - | - | - | - | - |
| GGA1 | OID20838 | Neurology | GGA1 | Q9UJY5 | | 22 | 37608725 | 37633564 | 0 | - | - | - | - | - | - |
| GH1 | OID20220 | Cardiome | GH1 | P01241 | | 17 | 63917200 | 63918839 | 0 | - | - | - | - | - | - |
| GH2 | OID21168 | Oncology | GH2 | P01242 | | 17 | 63880215 | 63881944 | 0 | - | - | - | - | - | - |
| GHRHR | OID20841 | Neurology | GHRHR | Q02643 | | 7 | 30938669 | 30993254 | 0 | - | - | - | - | - | - |
| GHRL | OID20175 | Cardiome | GHRL | Q9UBU3 | | 3 | 10285666 | 10292947 | 0 | - | - | - | - | - | - |
| GID8 | OID31041 | Neurology | GID8 | Q9NWU2 | | 20 | 62938147 | 62948475 | 0 | - | - | - | - | - | - |
| GIGYF2 | OID31062 | Neurology | GIGYF2 | Q6Y7W6 | | 2 | 232697299 | 232860605 | 0 | - | - | - | - | - | - |
| GIP | OID30288 | Cardiome | GIP | P09681 | | 17 | 48958554 | 48968596 | 0 | - | - | - | - | - | - |
| GIPR | OID30542 | Inflammat | GIPR | P48546 | | 19 | 45668221 | 45683722 | 0 | - | - | - | - | - | - |
| GJA8 | OID31187 | Oncology | GJA8 | P48165 | | 1 | 147902795 | 147909269 | 0 | - | - | - | - | - | - |
| GKN1 | OID20885 | Neurology | GKN1 | Q9NS71 | | 2 | 68974573 | 68980980 | 0 | - | - | - | - | - | - |
| GLA | OID30594 | Inflammat | GLA | P06280 | X | | 101393273 | 101408012 | 0 | - | - | - | - | - | - |
| GLI2 | OID30429 | Inflammat | GLI2 | P10070 | | 2 | 120735623 | 120992653 | 0 | - | - | - | - | - | - |
| GLOD4 | OID20710 | Inflammat | GLOD4 | Q9HC38 | | 17 | 757097 | 782294 | 0 | - | - | - | - | - | - |
| GLP1R | OID30809 | Neurology | GLP1R | P43220 | | 6 | 39048781 | 39091303 | 0 | - | - | - | - | - | - |
| GLYR1 | OID31364 | Oncology | GLYR1 | Q49A26 | | 16 | 4803203 | 4847288 | 0 | - | - | - | - | - | - |
| GMPR2 | OID30623 | Inflammat | GMPR2 | Q9P2T1 | | 14 | 24232422 | 24239242 | 0 | - | - | - | - | - | - |
| GNE | OID21231 | Oncology | GNE | Q9Y223 | | 9 | 36214441 | 36277042 | 0 | - | - | - | - | - | - |
| GNGT1 | OID30920 | Neurology | GNGT1 | P63211 | | 7 | 93591573 | 93911265 | 0 | - | - | - | - | - | - |
| GOLM2 | OID21044 | Neurology | GOLM2 | Q6P4E1 | | 15 | 44288719 | 44415758 | 0 | - | - | - | - | - | - |
| GOPC | OID20551 | Inflammat | GOPC | Q9HD26 | | 6 | 117560269 | 117602542 | 0 | - | - | - | - | - | - |
| GOT1 | OID30417 | Cardiome | GOT1 | P17174 | | 10 | 99396870 | 99430624 | 0 | - | - | - | - | - | - |
| GP1BB | OID30174 | Cardiome | GP1BB | P13224 | | 22 | 19723539 | 19724771 | 0 | - | - | - | - | - | - |
| GPD1 | OID31358 | Oncology | GPD1 | P21695 | | 12 | 50103982 | 50111313 | 0 | - | - | - | - | - | - |
| GPI | OID30680 | Inflammat | GPI | P06744 | | 19 | 34359480 | 34402413 | 0 | - | - | - | - | - | - |
| GPKOW | OID20890 | Neurology | GPKOW | Q92917 | X | | 49113407 | 49123735 | 0 | - | - | - | - | - | - |
| GPR101 | OID30819 | Neurology | GPR101 | Q96P66 | X | | 137023929 | 137033995 | 0 | - | - | - | - | - | - |
| GRIN2B | OID30802 | Neurology | GRIN2B | Q13224 | | 12 | 13437942 | 13982002 | 0 | - | - | - | - | - | - |
| GRPEL1 | OID21309 | Oncology | GRPEL1 | Q9HAV7 | | 4 | 7058895 | 7068064 | 0 | - | - | - | - | - | - |
| GRSF1 | OID31224 | Oncology | GRSF1 | Q12849 | | 4 | 70815783 | 70839897 | 0 | - | - | - | - | - | - |
| GSAP | OID21173 | Oncology | GSAP | A4D1B5 | | 7 | 77310751 | 77416349 | 0 | - | - | - | - | - | - |
| GSN | OID30714 | Inflammat | GSN | P06396 | | 9 | 121207794 | 121332843 | 0 | - | - | - | - | - | - |
| GTF2IRD1 | OID31264 | Oncology | GTF2IRD | Q9UHL9 | | 7 | 74453790 | 74602605 | 0 | - | - | - | - | - | - |
| GTPBP2 | OID30946 | Neurology | GTPBP2 | Q9BX10 | | 6 | 43605316 | 43629264 | 0 | - | - | - | - | - | - |
| GUCA2A | OID21101 | Neurology | GUCA2A | Q02747 | | 1 | 42162690 | 42164745 | 0 | - | - | - | - | - | - |
| GUCY2C | OID31318 | Oncology | GUCY2C | P25092 | | 12 | 14612632 | 14696599 | 0 | - | - | - | - | - | - |
| GUK1 | OID30148 | Cardiome | GUK1 | Q16774 | | 1 | 228139962 | 228148984 | 0 | - | - | - | - | - | - |
| GUSB | OID20296 | Cardiome | GUSB | P08236 | | 7 | 65960684 | 65982215 | 0 | - | - | - | - | - | - |
| GYS1 | OID20097 | Cardiome | GYS1 | P13807 | | 19 | 48968130 | 48993310 | 0 | - | - | - | - | - | - |
| H2AP | OID31232 | Oncology | H2AP | O75409 | X | | 37990779 | 37991314 | 0 | - | - | - | - | - | - |
| HAGH | OID21459 | Oncology | HAGH | Q16775 | | 16 | 1795620 | 1827157 | 0 | - | - | - | - | - | - |
| HAO1 | OID21395 | Oncology | HAO1 | Q9UJM8 | | 20 | 7882985 | 7940458 | 0 | - | - | - | - | - | - |
| HARS1 | OID21086 | Neurology | HARS1 | P12081 | | 5 | 140673035 | 140691537 | 0 | - | - | - | - | - | - |
| HAVCR2 | OID21137 | Neurology | HAVCR2 | Q8TDQ0 | | 5 | 157085832 | 157142869 | 0 | - | - | - | - | - | - |
| HBEGF | OID21465 | Oncology | HBEGF | Q99075 | | 5 | 140332843 | 140346603 | 0 | - | - | - | - | - | - |
| HDAC8 | OID30436 | Inflammat | HDAC8 | Q9BY41 | X | | 72329516 | 72573101 | 0 | - | - | - | - | - | - |
| HDAC9 | OID30832 | Neurology | HDAC9 | Q9UKV0 | | 7 | 18086949 | 19002416 | 0 | - | - | - | - | - | - |
| HDGFL2 | OID31136 | Neurology | HDGFL2 | Q7Z4V5 | | 19 | 4472297 | 4502208 | 0 | - | - | - | - | - | - |
| HEPACAM2 | OID31151 | Neurology | HEPACAI | A8MVW5 | | 7 | 93188534 | 93226469 | 0 | - | - | - | - | - | - |
| HEPH | OID30337 | Cardiome | HEPH | Q9BQS7 | X | | 66162671 | 66268867 | 0 | - | - | - | - | - | - |
| HGS | OID21175 | Oncology | HGS | O14964 | | 17 | 81683326 | 81703138 | 0 | - | - | - | - | - | - |
| HHEX | OID31107 | Neurology | HHEX | Q03014 | | 10 | 92689955 | 92695647 | 0 | - | - | - | - | - | - |
| HIF1A | OID30425 | Inflammat | HIF1A | Q16665 | | 14 | 61695513 | 61748259 | 0 | - | - | - | - | - | - |
| HIP1 | OID30071 | Cardiome | HIP1 | O00291 | | 7 | 75533298 | 75738962 | 0 | - | - | - | - | - | - |
| HK2 | OID20065 | Cardiome | HK2 | P52789 | | 2 | 74834127 | 74893359 | 0 | - | - | - | - | - | - |
| HMGCL | OID30214 | Cardiome | HMGCL | P35914 | | 1 | 23801885 | 23838620 | 0 | - | - | - | - | - | - |
| HMGCS1 | OID30157 | Cardiome | HMGCS1 | Q01581 | | 5 | 43287470 | 43313512 | 0 | - | - | - | - | - | - |
| HMMR | OID31175 | Oncology | HMMR | O75330 | | 5 | 163460203 | 163491941 | 0 | - | - | - | - | - | - |
| HNF1A | OID30919 | Neurology | HNF1A | P20823 | | 12 | 120978543 | 121002512 | 0 | - | - | - | - | - | - |
| HNRNPK | OID20051 | Cardiome | HNRNPK | P61978 | | 9 | 83968083 | 83980616 | 0 | - | - | - | - | - | - |
| HNRNPUL1 | OID31037 | Neurology | HNRNPUI | Q9BUJ2 | | 19 | 41262496 | 41307787 | 0 | - | - | - | - | - | - |
| HRAS | OID31263 | Oncology | HRAS | P01112 | | 11 | 532242 | 537321 | 0 | - | - | - | - | - | - |
| HRC | OID30211 | Cardiome | HRC | P23327 | | 19 | 49151198 | 49155396 | 0 | - | - | - | - | - | - |
| HS1BP3 | OID30243 | Cardiome | HS1BP3 | Q53T59 | | 2 | 20560448 | 20651130 | 0 | - | - | - | - | - | - |
| HS6ST2 | OID30481 | Inflammat | HS6ST2 | Q96MM7 | X | | 132626015 | 132961395 | 0 | - | - | - | - | - | - |
| HSD11B1 | OID20575 | Inflammat | HSD11B1 | P28845 | | 1 | 209686178 | 209734949 | 0 | - | - | - | - | - | - |
| HSD17B3 | OID30145 | Cardiome | HSD17B3 | P37058 | | 9 | 96235306 | 96302176 | 0 | - | - | - | - | - | - |
| HSP90B1 | OID20829 | Neurology | HSP90B1 | P14625 | | 12 | 103930107 | 103953931 | 0 | - | - | - | - | - | - |
| HSPA1A | OID20718 | Inflammat | HSPA1A | P0DMV8 | | 6 | 31815543 | 31817946 | 0 | - | - | - | - | - | - |
| HSPB6 | OID21408 | Oncology | HSPB6 | O14558 | | 19 | 35754566 | 35758079 | 0 | - | - | - | - | - | - |

| | | | | | | | | | | | | | | | |
|---|---|---|---|---|---|---|---|---|---|---|---|---|---|---|---|
| HTR1A | OID30824 | Neurology | HTR1A | P08908 | | 5 | 63957874 | 63962507 | 0 | - | - | - | - | - | - |
| HTR1B | OID30866 | Neurology | HTR1B | P28222 | | 6 | 77460924 | 77463491 | 0 | - | - | - | - | - | - |
| HTRA2 | OID21471 | Oncology | HTRA2 | O43464 | | 2 | 74529725 | 74533332 | 0 | - | - | - | - | - | - |
| ICA1 | OID20484 | Inflammat | ICA1 | Q05084 | | 7 | 8113184 | 8262687 | 0 | - | - | - | - | - | - |
| ID4 | OID31284 | Oncology | ID4 | P47928 | | 6 | 19837370 | 19842197 | 0 | - | - | - | - | - | - |
| IDS | OID20619 | Inflammat | IDS | P22304 | X | | 149476988 | 149521096 | 0 | - | - | - | - | - | - |
| IFIT1 | OID31310 | Oncology | IFIT1 | P09914 | | 10 | 89392546 | 89406487 | 0 | - | - | - | - | - | - |
| IFNG | OID20495 | Inflammat | IFNG | P01579 | | 12 | 68154768 | 68159740 | 0 | - | - | - | - | - | - |
| IFNL1 | OID20795 | Neurology | IFNL1 | Q8IU54 | | 19 | 39296407 | 39298673 | 0 | - | - | - | - | - | - |
| IFNL2 | OID30791 | Neurology | IFNL2 | Q8IZJ0 | | 19 | 39268396 | 39270188 | 0 | - | - | - | - | - | - |
| IFNW1 | OID30129 | Cardiome | IFNW1 | P05000 | | 9 | 21140632 | 21141832 | 0 | - | - | - | - | - | - |
| IFT20 | OID30962 | Neurology | IFT20 | Q8IY31 | | 17 | 28328325 | 28335489 | 0 | - | - | - | - | - | - |
| IGBP1 | OID31059 | Neurology | IGBP1 | P78318 | X | | 70133447 | 70166324 | 0 | - | - | - | - | - | - |
| IGDCC3 | OID30862 | Neurology | IGDCC3 | Q8IVU1 | | 15 | 65327127 | 65378002 | 0 | - | - | - | - | - | - |
| IGF2BP3 | OID30112 | Cardiome | IGF2BP3 | O00425 | | 7 | 23310209 | 23470491 | 0 | - | - | - | - | - | - |
| IGFBP1 | OID20246 | Cardiome | IGFBP1 | P08833 | | 7 | 45888360 | 45893660 | 0 | - | - | - | - | - | - |
| IGFBP4 | OID21158 | Neurology | IGFBP4 | P22692 | | 17 | 40443450 | 40457725 | 0 | - | - | - | - | - | - |
| IGFL4 | OID30500 | Inflammat | IGFL4 | Q6B9Z1 | | 19 | 46039182 | 46077118 | 0 | - | - | - | - | - | - |
| IGLON5 | OID31169 | Oncology | IGLON5 | A6NGN9 | | 19 | 51311644 | 51330891 | 0 | - | - | - | - | - | - |
| IKBKG | OID20544 | Inflammat | IKBKG | Q9Y6K9 | X | | 154541199 | 154565046 | 0 | - | - | - | - | - | - |
| IKZF2 | OID21213 | Oncology | IKZF2 | Q9UKS7 | | 2 | 212999691 | 213152427 | 0 | - | - | - | - | - | - |
| IL11 | OID20455 | Inflammat | IL11 | P20809 | | 19 | 55364382 | 55370463 | 0 | - | - | - | - | - | - |
| IL12RB2 | OID30422 | Inflammat | IL12RB2 | Q99665 | | 1 | 67307364 | 67398724 | 0 | - | - | - | - | - | - |
| IL13 | OID20430 | Inflammat | IL13 | P35225 | | 5 | 132656263 | 132661110 | 0 | - | - | - | - | - | - |
| IL13RA1 | OID21504 | Oncology | IL13RA1 | P78552 | X | | 118727133 | 118794535 | 0 | - | - | - | - | - | - |
| IL13RA2 | OID30841 | Neurology | IL13RA2 | Q14627 | X | | 115003975 | 115019977 | 0 | - | - | - | - | - | - |
| IL15 | OID20562 | Inflammat | IL15 | P40933 | | 4 | 141636583 | 141733987 | 0 | - | - | - | - | - | - |
| IL17A | OID20469 | Inflammat | IL17A | Q16552 | | 6 | 52186375 | 52190638 | 0 | - | - | - | - | - | - |
| IL18BP | OID20348 | Cardiome | IL18BP | O95998 | | 11 | 71998613 | 72005715 | 0 | - | - | - | - | - | - |
| IL18RAP | OID20900 | Neurology | IL18RAP | O95256 | | 2 | 102418689 | 102452565 | 0 | - | - | - | - | - | - |
| IL1A | OID20488 | Inflammat | IL1A | P01583 | | 2 | 112773925 | 112784493 | 0 | - | - | - | - | - | - |
| IL1B | OID20427 | Inflammat | IL1B | P01584 | | 2 | 112829751 | 112836816 | 0 | - | - | - | - | - | - |
| IL2 | OID20419 | Inflammat | IL2 | P60568 | | 4 | 122451470 | 122456725 | 0 | - | - | - | - | - | - |
| IL20 | OID20453 | Inflammat | IL20 | Q9NYY1 | | 1 | 206865623 | 206869223 | 0 | - | - | - | - | - | - |
| IL20RA | OID20422 | Inflammat | IL20RA | Q9UHF4 | | 6 | 136999971 | 137045180 | 0 | - | - | - | - | - | - |
| IL21R | OID30441 | Inflammat | IL21R | Q9HBE5 | | 16 | 27402174 | 27452042 | 0 | - | - | - | - | - | - |
| IL24 | OID20440 | Inflammat | IL24 | Q13007 | | 1 | 206897443 | 206904139 | 0 | - | - | - | - | - | - |
| IL25 | OID31243 | Oncology | IL25 | Q9H293 | | 14 | 23372809 | 23376403 | 0 | - | - | - | - | - | - |
| IL2RB | OID20450 | Inflammat | IL2RB | P14784 | | 22 | 37125843 | 37175054 | 0 | - | - | - | - | - | - |
| IL2RG | OID31361 | Oncology | IL2RG | P31785 | X | | 71107404 | 71112108 | 0 | - | - | - | - | - | - |
| IL3 | OID31229 | Oncology | IL3 | P08700 | | 5 | 132060655 | 132063204 | 0 | - | - | - | - | - | - |
| IL31 | OID30487 | Inflammat | IL31 | Q6EBC2 | | 12 | 122172029 | 122174221 | 0 | - | - | - | - | - | - |
| IL33 | OID20428 | Inflammat | IL33 | O95760 | | 9 | 6215786 | 6257983 | 0 | - | - | - | - | - | - |
| IL3RA | OID20461 | Inflammat | IL3RA | P26951 | X | | 1336616 | 1382689 | 0 | - | - | - | - | - | - |
| IL4 | OID20426 | Inflammat | IL4 | P05112 | | 5 | 132673986 | 132682678 | 0 | - | - | - | - | - | - |
| IL5 | OID20472 | Inflammat | IL5 | P05113 | | 5 | 132541445 | 132556838 | 0 | - | - | - | - | - | - |
| IL6 | OID20101 | Cardiome | IL6 | P05231 | | 7 | 22725884 | 22732002 | 0 | - | - | - | - | - | - |
| IL9 | OID31298 | Oncology | IL9 | P15248 | | 5 | 135892246 | 135895841 | 0 | - | - | - | - | - | - |
| ILKAP | OID20855 | Neurology | ILKAP | Q9H0C8 | | 2 | 238170402 | 238203708 | 0 | - | - | - | - | - | - |
| IMPACT | OID31112 | Neurology | IMPACT | Q9P2X3 | | 18 | 24426634 | 24453531 | 0 | - | - | - | - | - | - |
| IMPG1 | OID30814 | Neurology | IMPG1 | Q17R60 | | 6 | 75921114 | 76072678 | 0 | - | - | - | - | - | - |
| ING1 | OID20853 | Neurology | ING1 | Q9UK53 | | 13 | 110712736 | 110723339 | 0 | - | - | - | - | - | - |
| INPP1 | OID21345 | Oncology | INPP1 | P49441 | | 2 | 190343570 | 190371665 | 0 | - | - | - | - | - | - |
| INPP5J | OID30975 | Neurology | INPP5J | Q15735 | | 22 | 31122731 | 31134697 | 0 | - | - | - | - | - | - |
| INPPL1 | OID21314 | Oncology | INPPL1 | O15357 | | 11 | 72223701 | 72239147 | 0 | - | - | - | - | - | - |
| INSL3 | OID31129 | Neurology | INSL3 | P51460 | | 19 | 17816512 | 17821574 | 0 | - | - | - | - | - | - |
| INSR | OID30566 | Inflammat | INSR | P06213 | | 19 | 7112255 | 7294414 | 0 | - | - | - | - | - | - |
| IRAG2 | OID20085 | Cardiome | IRAG2 | Q12912 | | 12 | 25004342 | 25108335 | 0 | - | - | - | - | - | - |
| IRAK1 | OID20485 | Inflammat | IRAK1 | P51617 | X | | 154010506 | 154019902 | 0 | - | - | - | - | - | - |
| IRAK4 | OID20577 | Inflammat | IRAK4 | Q9NWZ3 | | 12 | 43758944 | 43789543 | 0 | - | - | - | - | - | - |
| ISM2 | OID30365 | Cardiome | ISM2 | Q6H9L7 | | 14 | 77474394 | 77498816 | 0 | - | - | - | - | - | - |
| ITGA5 | OID21139 | Neurology | ITGA5 | P08648 | | 12 | 54395261 | 54419266 | 0 | - | - | - | - | - | - |
| ITGAX | OID31325 | Oncology | ITGAX | P20702 | | 16 | 31355134 | 31382999 | 0 | - | - | - | - | - | - |
| ITGB1 | OID20376 | Cardiome | ITGB1 | P05556 | | 10 | 32887273 | 33005792 | 0 | - | - | - | - | - | - |
| ITGB1BP1 | OID21268 | Oncology | ITGB1BP: | O14713 | | 2 | 9403475 | 9423528 | 0 | - | - | - | - | - | - |
| ITGB1BP2 | OID20112 | Cardiome | ITGB1BP: | Q9UKP3 | X | | 71301750 | 71305371 | 0 | - | - | - | - | - | - |
| ITM2A | OID20504 | Inflammat | ITM2A | O43736 | X | | 79360384 | 79367667 | 0 | - | - | - | - | - | - |
| ITPRIP | OID30860 | Neurology | ITPRIP | Q8IWB1 | | 10 | 104309698 | 104338465 | 0 | - | - | - | - | - | - |
| IVD | OID20833 | Neurology | IVD | P26440 | | 15 | 40405485 | 40435947 | 0 | - | - | - | - | - | - |
| IZUMO1 | OID31159 | Oncology | IZUMO1 | Q8IYV9 | | 19 | 48740852 | 48746909 | 0 | - | - | - | - | - | - |
| JCHAIN | OID20494 | Inflammat | JCHAIN | P01591 | | 4 | 70655541 | 70681817 | 0 | - | - | - | - | - | - |
| JMJD1C | OID31172 | Oncology | JMJD1C | Q15652 | | 10 | 63167221 | 63521850 | 0 | - | - | - | - | - | - |
| JPT2 | OID31520 | Oncology | JPT2 | Q9H910 | | 16 | 1678256 | 1702280 | 0 | - | - | - | - | - | - |
| JUN | OID20424 | Inflammat | JUN | P05412 | | 1 | 58776845 | 58784048 | 0 | - | - | - | - | - | - |
| KAZN | OID31481 | Oncology | KAZN | Q674X7 | | 1 | 13892792 | 15118043 | 0 | - | - | - | - | - | - |
| KCNC4 | OID30813 | Neurology | KCNC4 | Q03721 | | 1 | 110210314 | 110283100 | 0 | - | - | - | - | - | - |
| KCNH2 | OID30899 | Neurology | KCNH2 | Q12809 | | 7 | 150944961 | 150978321 | 0 | - | - | - | - | - | - |
| KCNIP4 | OID20813 | Neurology | KCNIP4 | Q6PIL6 | | 4 | 20728606 | 21948772 | 0 | - | - | - | - | - | - |
| KCTD5 | OID31148 | Neurology | KCTD5 | Q9NXV2 | | 16 | 2682523 | 2709030 | 0 | - | - | - | - | - | - |
| KDM3A | OID30426 | Inflammat | KDM3A | Q9Y4C1 | | 2 | 86440647 | 86492716 | 0 | - | - | - | - | - | - |
| KHDC3L | OID31245 | Oncology | KHDC3L | Q587J8 | | 6 | 73362658 | 73364171 | 0 | - | - | - | - | - | - |
| KIAA1549 | OID31307 | Oncology | KIAA1549 | Q9HCM3 | | 7 | 138831381 | 138981389 | 0 | - | - | - | - | - | - |
| KIAA1549L | OID31312 | Oncology | KIAA1549 | Q6ZVL6 | | 11 | 33376108 | 33674102 | 0 | - | - | - | - | - | - |
| KIAA2013 | OID31392 | Oncology | KIAA2013 | Q8IYS2 | | 1 | 11919591 | 11926428 | 0 | - | - | - | - | - | - |
| KIF1C | OID30163 | Cardiome | KIF1C | O43896 | | 17 | 4997950 | 5028401 | 0 | - | - | - | - | - | - |
| KIF20B | OID30816 | Neurology | KIF20B | Q96Q89 | | 10 | 89701590 | 89774939 | 0 | - | - | - | - | - | - |
| KIF22 | OID30175 | Cardiome | KIF22 | Q14807 | | 16 | 29790727 | 29805385 | 0 | - | - | - | - | - | - |
| KIFBP | OID21320 | Oncology | KIFBP | Q96EK5 | | 10 | 68988803 | 69043544 | 0 | - | - | - | - | - | - |
| KIRREL1 | OID30806 | Neurology | KIRREL1 | Q96J84 | | 1 | 157993273 | 158100262 | 0 | - | - | - | - | - | - |
| KLF4 | OID30808 | Neurology | KLF4 | O43474 | | 9 | 107484852 | 107490482 | 0 | - | - | - | - | - | - |
| KLHL41 | OID30986 | Neurology | KLHL41 | O60662 | | 2 | 169509702 | 169526258 | 0 | - | - | - | - | - | - |
| KLRB1 | OID20629 | Inflammat | KLRB1 | Q12918 | | 12 | 9594551 | 9607916 | 0 | - | - | - | - | - | - |
| KLRC1 | OID30855 | Neurology | KLRC1 | P26715 | | 12 | 10442264 | 10454685 | 0 | - | - | - | - | - | - |
| KLRF1 | OID30464 | Inflammat | KLRF1 | Q9NZS2 | | 12 | 9827481 | 9845007 | 0 | - | - | - | - | - | - |
| KRT14 | OID20800 | Neurology | KRT14 | P02533 | | 17 | 41582279 | 41586895 | 0 | - | - | - | - | - | - |
| KRT17 | OID30092 | Cardiome | KRT17 | Q04695 | | 17 | 41619442 | 41624842 | 0 | - | - | - | - | - | - |
| KRT18 | OID21296 | Oncology | KRT18 | P05783 | | 12 | 52948871 | 52952906 | 0 | - | - | - | - | - | - |
| KRT6C | OID30169 | Cardiome | KRT6C | P48668 | | 12 | 52468516 | 52473805 | 0 | - | - | - | - | - | - |
| KRT8 | OID30865 | Neurology | KRT8 | P05787 | | 12 | 52897187 | 52949954 | 0 | - | - | - | - | - | - |
| L1CAM | OID21484 | Oncology | L1CAM | P32004 | X | | 153861514 | 153886173 | 0 | - | - | - | - | - | - |
| L3HYPDH | OID30314 | Cardiome | L3HYPDH | Q96EM0 | | 14 | 59460363 | 59484408 | 0 | - | - | - | - | - | - |
| LACTB2 | OID20150 | Cardiome | LACTB2 | Q53H82 | | 8 | 70635318 | 70669185 | 0 | - | - | - | - | - | - |
| LAMA1 | OID30057 | Cardiome | LAMA1 | P25391 | | 18 | 6941742 | 7117797 | 0 | - | - | - | - | - | - |
| LAMP2 | OID21035 | Neurology | LAMP2 | P13473 | X | | 120426148 | 120469365 | 0 | - | - | - | - | - | - |
| LAMTOR5 | OID31396 | Oncology | LAMTOR5 | O43504 | | 1 | 110401249 | 110407942 | 0 | - | - | - | - | - | - |
| LAP3 | OID20436 | Inflammat | LAP3 | P28838 | | 4 | 17577198 | 17607972 | 0 | - | - | - | - | - | - |
| LARP1 | OID31372 | Oncology | LARP1 | Q6PKG0 | | 5 | 154682986 | 154817605 | 0 | - | - | - | - | - | - |

| | | | | | | | | | | | | | | | |
|---|---|---|---|---|---|---|---|---|---|---|---|---|---|---|---|
| LAT | OID20640 | Inflammat | LAT | O43561 | | 16 | 28984826 | 28990784 | 0 | - | - | - | - | - | - |
| LAT2 | OID21292 | Oncology | LAT2 | Q9GZY6 | | 7 | 74199652 | 74229834 | 0 | - | - | - | - | - | - |
| LBR | OID21034 | Neurology | LBR | Q14739 | | 1 | 225401502 | 225428925 | 0 | - | - | - | - | - | - |
| LDLR | OID20240 | Cardiome | LDLR | P01130 | | 19 | 11089462 | 11133820 | 0 | - | - | - | - | - | - |
| LELP1 | OID30845 | Neurology | LELP1 | Q5T871 | | 1 | 153203430 | 153205120 | 0 | - | - | - | - | - | - |
| LEO1 | OID30991 | Neurology | LEO1 | Q8WVC0 | | 15 | 51938025 | 51971778 | 0 | - | - | - | - | - | - |
| LEP | OID20187 | Cardiome | LEP | P41159 | | 7 | 128241278 | 128257629 | 0 | - | - | - | - | - | - |
| LETM1 | OID31018 | Neurology | LETM1 | O95202 | | 4 | 1811479 | 1856156 | 0 | - | - | - | - | - | - |
| LIF | OID20824 | Neurology | LIF | P15018 | | 22 | 30240453 | 30246759 | 0 | - | - | - | - | - | - |
| LILRA4 | OID30162 | Cardiome | LILRA4 | P59901 | | 19 | 54333185 | 54339162 | 0 | - | - | - | - | - | - |
| LMNB1 | OID31501 | Oncology | LMNB1 | P20700 | | 5 | 126776623 | 126837020 | 0 | - | - | - | - | - | - |
| LMNB2 | OID31109 | Neurology | LMNB2 | Q03252 | | 19 | 2427638 | 2456959 | 0 | - | - | - | - | - | - |
| LMOD2 | OID30864 | Neurology | LMOD2 | Q6P5Q4 | | 7 | 123655866 | 123664290 | 0 | - | - | - | - | - | - |
| LPP | OID30190 | Cardiome | LPP | Q93052 | | 3 | 188153284 | 188890671 | 0 | - | - | - | - | - | - |
| LRFN2 | OID30895 | Neurology | LRFN2 | Q9ULH4 | | 6 | 40391591 | 40587364 | 0 | - | - | - | - | - | - |
| LRP1 | OID21293 | Oncology | LRP1 | Q07954 | | 12 | 57128483 | 57213361 | 0 | - | - | - | - | - | - |
| LRP2 | OID30960 | Neurology | LRP2 | P98164 | | 2 | 169127109 | 169362534 | 0 | - | - | - | - | - | - |
| LRP2BP | OID30907 | Neurology | LRP2BP | Q9P2M1 | | 4 | 185363872 | 185395924 | 0 | - | - | - | - | - | - |
| LRRC59 | OID30361 | Cardiome | LRRC59 | Q96AG4 | | 17 | 50375059 | 50397523 | 0 | - | - | - | - | - | - |
| LRTM1 | OID30906 | Neurology | LRTM1 | Q9HBL6 | | 3 | 54918231 | 54967088 | 0 | - | - | - | - | - | - |
| LSM1 | OID21210 | Oncology | LSM1 | O15116 | | 8 | 38163335 | 38176730 | 0 | - | - | - | - | - | - |
| LSM8 | OID31269 | Oncology | LSM8 | O95777 | | 7 | 118184144 | 118204035 | 0 | - | - | - | - | - | - |
| LTO1 | OID20434 | Inflammat | LTO1 | Q8WV07 | | 11 | 69653076 | 69675416 | 0 | - | - | - | - | - | - |
| LYN | OID21350 | Oncology | LYN | P07948 | | 8 | 55879835 | 56014169 | 0 | - | - | - | - | - | - |
| LYPD1 | OID20821 | Neurology | LYPD1 | Q8N2G4 | | 2 | 132643286 | 132671579 | 0 | - | - | - | - | - | - |
| LYPLA2 | OID30817 | Neurology | LYPLA2 | O95372 | | 1 | 23791145 | 23795539 | 0 | - | - | - | - | - | - |
| MAD1L1 | OID20904 | Neurology | MAD1L1 | Q9Y6D9 | | 7 | 1815793 | 2233243 | 0 | - | - | - | - | - | - |
| MAEA | OID21206 | Oncology | MAEA | Q7L5Y9 | | 4 | 1289887 | 1340147 | 0 | - | - | - | - | - | - |
| MAG | OID30967 | Neurology | MAG | P20916 | | 19 | 35292125 | 35313807 | 0 | - | - | - | - | - | - |
| MAGEA3 | OID31331 | Oncology | MAGEA3 | P43357 | X | | 152698767 | 152702347 | 0 | - | - | - | - | - | - |
| MAGED1 | OID21181 | Oncology | MAGED1 | Q9Y5V3 | X | | 51803007 | 51902354 | 0 | - | - | - | - | - | - |
| MANEAL | OID31244 | Oncology | MANEAL | Q5VSG8 | | 1 | 37793847 | 37801137 | 0 | - | - | - | - | - | - |
| MANSC1 | OID21492 | Oncology | MANSC1 | Q9H8J5 | | 12 | 12326056 | 12350242 | 0 | - | - | - | - | - | - |
| MAP1LC3A | OID30868 | Neurology | MAP1LC3 | Q9H492 | | 20 | 34546854 | 34560345 | 0 | - | - | - | - | - | - |
| MAP1LC3B2 | OID30074 | Cardiome | MAP1LC3 | A6NCE7 | | 12 | 116548105 | 116576606 | 0 | - | - | - | - | - | - |
| MAP2K6 | OID20552 | Inflammat | MAP2K6 | P52564 | | 17 | 69414697 | 69553865 | 0 | - | - | - | - | - | - |
| MAP3K5 | OID21253 | Oncology | MAP3K5 | Q99683 | | 6 | 136557046 | 136792477 | 0 | - | - | - | - | - | - |
| MAPRE3 | OID31177 | Oncology | MAPRE3 | Q9UPY8 | | 2 | 26970637 | 27027219 | 0 | - | - | - | - | - | - |
| MAPT | OID20830 | Neurology | MAPT | P10636 | | 17 | 45894527 | 46028334 | 0 | - | - | - | - | - | - |
| MARS1 | OID30658 | Inflammat | MARS1 | P56192 | | 12 | 57475445 | 57517569 | 0 | - | - | - | - | - | - |
| MB | OID20263 | Cardiome | MB | P02144 | | 22 | 35606764 | 35637951 | 0 | - | - | - | - | - | - |
| MCEMP1 | OID30501 | Inflammat | MCEMP1 | Q8IX19 | | 19 | 7677088 | 7679829 | 0 | - | - | - | - | - | - |
| MCTS1 | OID31444 | Oncology | MCTS1 | Q9ULC4 | X | | 120594010 | 120621159 | 0 | - | - | - | - | - | - |
| MDK | OID21489 | Oncology | MDK | P21741 | | 11 | 46380756 | 46383837 | 0 | - | - | - | - | - | - |
| MED18 | OID21236 | Oncology | MED18 | Q9BUE0 | | 1 | 28329002 | 28335965 | 0 | - | - | - | - | - | - |
| MED21 | OID30066 | Cardiome | MED21 | Q13503 | | 12 | 27022546 | 27066343 | 0 | - | - | - | - | - | - |
| MENT | OID30595 | Inflammat | C1orf56 | Q9BUN1 | | 1 | 151047751 | 151051986 | 0 | - | - | - | - | - | - |
| MESD | OID21099 | Neurology | MESD | Q14696 | | 15 | 80946289 | 80989828 | 0 | - | - | - | - | - | - |
| METAP1 | OID20815 | Neurology | METAP1 | P53582 | | 4 | 98995659 | 99062809 | 0 | - | - | - | - | - | - |
| METAP1D | OID20519 | Inflammat | METAP1D | Q6UB28 | | 2 | 171999943 | 172082430 | 0 | - | - | - | - | - | - |
| METAP2 | OID21437 | Oncology | METAP2 | P50579 | | 12 | 95473520 | 95515839 | 0 | - | - | - | - | - | - |
| MFAP3 | OID20057 | Cardiome | MFAP3 | P55082 | | 5 | 154038959 | 154220478 | 0 | - | - | - | - | - | - |
| MFAP3L | OID30933 | Neurology | MFAP3L | O75121 | | 4 | 169986597 | 170033031 | 0 | - | - | - | - | - | - |
| MFAP5 | OID20239 | Cardiome | MFAP5 | Q13361 | | 12 | 8637346 | 8662888 | 0 | - | - | - | - | - | - |
| MINK1 | OID31156 | Neurology | MINK1 | Q8N4C8 | | 17 | 4833340 | 4898061 | 0 | - | - | - | - | - | - |
| MKI67 | OID30455 | Inflammat | MKI67 | P46013 | | 10 | 128096659 | 128126423 | 0 | - | - | - | - | - | - |
| MLLT1 | OID31211 | Oncology | MLLT1 | Q03111 | | 19 | 6210381 | 6279975 | 0 | - | - | - | - | - | - |
| MMP15 | OID30299 | Cardiome | MMP15 | P51511 | | 16 | 58025754 | 58046901 | 0 | - | - | - | - | - | - |
| MN1 | OID30908 | Neurology | MN1 | Q10571 | | 22 | 27748277 | 27801756 | 0 | - | - | - | - | - | - |
| MNAT1 | OID31327 | Oncology | MNAT1 | P51948 | | 14 | 60734742 | 60969965 | 0 | - | - | - | - | - | - |
| MNDA | OID20156 | Cardiome | MNDA | P41218 | | 1 | 158831351 | 158849506 | 0 | - | - | - | - | - | - |
| MORC3 | OID31073 | Neurology | MORC3 | Q14149 | | 21 | 36320189 | 36386148 | 0 | - | - | - | - | - | - |
| MORF4L1 | OID31257 | Oncology | MORF4L1 | Q9UBU8 | | 15 | 78810487 | 78898139 | 0 | - | - | - | - | - | - |
| MORF4L2 | OID31160 | Oncology | MORF4L2 | Q15014 | X | | 103675496 | 103688158 | 0 | - | - | - | - | - | - |
| MORN4 | OID31217 | Oncology | MORN4 | Q8NDC4 | | 10 | 97614553 | 97633500 | 0 | - | - | - | - | - | - |
| MPIG6B | OID20779 | Inflammat | MPIG6B | O95866 | | 6 | 31718594 | 31726714 | 0 | - | - | - | - | - | - |
| MPRIP | OID31420 | Oncology | MPRIP | Q6WCQ1 | | 17 | 17042457 | 17217679 | 0 | - | - | - | - | - | - |
| MRPL24 | OID30178 | Cardiome | MRPL24 | Q96A35 | | 1 | 156737303 | 156741590 | 0 | - | - | - | - | - | - |
| MRPL28 | OID31023 | Neurology | MRPL28 | Q13084 | | 16 | 366969 | 371289 | 0 | - | - | - | - | - | - |
| MRPL46 | OID20858 | Neurology | MRPL46 | Q9H2W6 | | 15 | 88459477 | 88467390 | 0 | - | - | - | - | - | - |
| MRPL52 | OID31239 | Oncology | MRPL52 | Q86TS9 | | 14 | 22829879 | 22835037 | 0 | - | - | - | - | - | - |
| MRPL58 | OID31036 | Neurology | MRPL58 | Q14197 | | 17 | 75012670 | 75021261 | 0 | - | - | - | - | - | - |
| MRPS16 | OID30489 | Inflammat | MRPS16 | Q9Y3D3 | | 10 | 73248843 | 73252693 | 0 | - | - | - | - | - | - |
| MSLNL | OID30939 | Neurology | MSLNL | Q96KJ4 | | 16 | 769428 | 783370 | 0 | - | - | - | - | - | - |
| MSTN | OID20115 | Cardiome | MSTN | O14793 | | 2 | 190055700 | 190062729 | 0 | - | - | - | - | - | - |
| MTDH | OID30538 | Inflammat | MTDH | Q86UE4 | | 8 | 97644184 | 97730260 | 0 | - | - | - | - | - | - |
| MTHFD2 | OID31078 | Neurology | MTHFD2 | P13995 | | 2 | 74186172 | 74217565 | 0 | - | - | - | - | - | - |
| MTPN | OID20080 | Cardiome | MTPN | P58546 | | 7 | 135926760 | 135977359 | 0 | - | - | - | - | - | - |
| MTR | OID30110 | Cardiome | MTR | Q99707 | | 1 | 236795260 | 236921278 | 0 | - | - | - | - | - | - |
| MTSS1 | OID31492 | Oncology | MTSS1 | O43312 | | 8 | 124550784 | 124728473 | 0 | - | - | - | - | - | - |
| MUCL3 | OID31170 | Oncology | MUCL3 | Q3MIW9 | | 6 | 30934523 | 30954221 | 0 | - | - | - | - | - | - |
| MVK | OID20459 | Inflammat | MVK | Q03426 | | 12 | 109573255 | 109598125 | 0 | - | - | - | - | - | - |
| MXRA8 | OID30583 | Inflammat | MXRA8 | Q9BRK3 | | 1 | 1352689 | 1361777 | 0 | - | - | - | - | - | - |
| MYBPC1 | OID30296 | Cardiome | MYBPC1 | Q00872 | | 12 | 101568353 | 101686028 | 0 | - | - | - | - | - | - |
| MYBPC2 | OID30199 | Cardiome | MYBPC2 | Q14324 | | 19 | 50432892 | 50466321 | 0 | - | - | - | - | - | - |
| MYCBP2 | OID30996 | Neurology | MYCBP2 | O75592 | | 13 | 77042474 | 77327094 | 0 | - | - | - | - | - | - |
| MYH4 | OID30061 | Cardiome | MYH4 | Q9Y623 | | 17 | 10443290 | 10469559 | 0 | - | - | - | - | - | - |
| MYH7B | OID31178 | Oncology | MYH7B | A7E2Y1 | | 20 | 34955810 | 35002437 | 0 | - | - | - | - | - | - |
| MYH9 | OID30382 | Cardiome | MYH9 | P35579 | | 22 | 36281280 | 36388010 | 0 | - | - | - | - | - | - |
| MYL1 | OID30120 | Cardiome | MYL1 | P05976 | | 2 | 210290150 | 210315174 | 0 | - | - | - | - | - | - |
| MYL3 | OID30251 | Cardiome | MYL3 | P08590 | | 3 | 46835111 | 46882172 | 0 | - | - | - | - | - | - |
| MYL4 | OID30245 | Cardiome | MYL4 | P12829 | | 17 | 47200446 | 47223679 | 0 | - | - | - | - | - | - |
| MYL6B | OID30936 | Neurology | MYL6B | P14649 | | 12 | 56152256 | 56159647 | 0 | - | - | - | - | - | - |
| MYLPF | OID30917 | Neurology | MYL11 | Q96A32 | | 16 | 30370934 | 30377991 | 0 | - | - | - | - | - | - |
| MYO6 | OID30989 | Neurology | MYO6 | Q9UM54 | | 6 | 75749201 | 75919537 | 0 | - | - | - | - | - | - |
| MYO9B | OID20451 | Inflammat | MYO9B | Q13459 | | 19 | 17075777 | 17214537 | 0 | - | - | - | - | - | - |
| MYOM1 | OID30834 | Neurology | MYOM1 | P52179 | | 18 | 3066807 | 3219968 | 0 | - | - | - | - | - | - |
| MYOM2 | OID30131 | Cardiome | MYOM2 | P54296 | | 8 | 2045046 | 2165552 | 0 | - | - | - | - | - | - |
| MZT1 | OID21186 | Oncology | MZT1 | Q08AG7 | | 13 | 72708367 | 72727629 | 0 | - | - | - | - | - | - |
| NAA10 | OID30188 | Cardiome | NAA10 | P41227 | X | | 153929225 | 153935080 | 0 | - | - | - | - | - | - |
| NAA80 | OID30893 | Neurology | NAA80 | Q93015 | | 3 | 50296402 | 50299416 | 0 | - | - | - | - | - | - |
| NACC1 | OID31393 | Oncology | NACC1 | Q96RE7 | | 19 | 13116862 | 13141147 | 0 | - | - | - | - | - | - |
| NAGA | OID30509 | Inflammat | NAGA | P17050 | | 22 | 42058334 | 42070842 | 0 | - | - | - | - | - | - |
| NAMPT | OID21227 | Oncology | NAMPT | P43490 | | 7 | 106248298 | 106285966 | 0 | - | - | - | - | - | - |
| NARS1 | OID31035 | Neurology | NARS1 | O43776 | | 18 | 57600656 | 57622213 | 0 | - | - | - | - | - | - |
| NBN | OID20442 | Inflammat | NBN | O60934 | | 8 | 89933331 | 90003228 | 0 | - | - | - | - | - | - |
| NCK2 | OID20683 | Inflammat | NCK2 | O43639 | | 2 | 105744912 | 105894274 | 0 | - | - | - | - | - | - |

| | | | | | | | | | | | | | | | |
|---|---|---|---|---|---|---|---|---|---|---|---|---|---|---|---|
| NCLN | OID20448 | Inflammat | NCLN | Q969V3 | | 19 | 3185563 | 3209575 | 0 | - | - | - | - | - | - |
| NDRG1 | OID20792 | Neurology | NDRG1 | Q92597 | | 8 | 133237175 | 133302022 | 0 | - | - | - | - | - | - |
| NDST1 | OID31369 | Oncology | NDST1 | P52848 | | 5 | 150485818 | 150558211 | 0 | - | - | - | - | - | - |
| NDUFA5 | OID30520 | Inflammat | NDUFA5 | Q16718 | | 7 | 123536997 | 123557904 | 0 | - | - | - | - | - | - |
| NDUFB7 | OID31521 | Oncology | NDUFB7 | P17568 | | 19 | 14566078 | 14572066 | 0 | - | - | - | - | - | - |
| NDUFS6 | OID21224 | Oncology | NDUFS6 | O75380 | | 5 | 1801407 | 1816048 | 0 | - | - | - | - | - | - |
| NEB | OID30105 | Cardiome | NEB | P20929 | | 2 | 151485336 | 151734487 | 0 | - | - | - | - | - | - |
| NECAP2 | OID30089 | Cardiome | NECAP2 | Q9NVZ3 | | 1 | 16440721 | 16460078 | 0 | - | - | - | - | - | - |
| NECTIN1 | OID30545 | Inflammat | NECTIN1 | Q15223 | | 11 | 119623408 | 119729200 | 0 | - | - | - | - | - | - |
| NEDD4L | OID30521 | Inflammat | NEDD4L | Q96PU5 | | 18 | 58044226 | 58401540 | 0 | - | - | - | - | - | - |
| NEDD9 | OID30503 | Inflammat | NEDD9 | Q14511 | | 6 | 11183298 | 11382348 | 0 | - | - | - | - | - | - |
| NEFL | OID20871 | Neurology | NEFL | P07196 | | 8 | 24950955 | 24956721 | 0 | - | - | - | - | - | - |
| NEK7 | OID31250 | Oncology | NEK7 | Q8TDX7 | | 1 | 198156994 | 198322420 | 0 | - | - | - | - | - | - |
| NENF | OID31354 | Oncology | NENF | Q9UMX5 | | 1 | 212432920 | 212446379 | 0 | - | - | - | - | - | - |
| NFAT5 | OID30522 | Inflammat | NFAT5 | O94916 | | 16 | 69565094 | 69704666 | 0 | - | - | - | - | - | - |
| NFATC3 | OID20470 | Inflammat | NFATC3 | Q12968 | | 16 | 68084751 | 68229259 | 0 | - | - | - | - | - | - |
| NFIC | OID30988 | Neurology | NFIC | P08651 | | 19 | 3314403 | 3469217 | 0 | - | - | - | - | - | - |
| NFKB2 | OID31258 | Oncology | NFKB2 | Q00653 | | 10 | 102394110 | 102402524 | 0 | - | - | - | - | - | - |
| NFX1 | OID30158 | Cardiome | NFX1 | Q12986 | | 9 | 33290512 | 33371157 | 0 | - | - | - | - | - | - |
| NFYA | OID30134 | Cardiome | NFYA | P23511 | | 6 | 41072974 | 41102403 | 0 | - | - | - | - | - | - |
| NGF | OID20920 | Neurology | NGF | P01138 | | 1 | 115285904 | 115338770 | 0 | - | - | - | - | - | - |
| NGFR | OID30300 | Cardiome | NGFR | P08138 | | 17 | 49495293 | 49515008 | 0 | - | - | - | - | - | - |
| NGRN | OID31115 | Neurology | NGRN | Q9NPE2 | | 15 | 90265659 | 90275778 | 0 | - | - | - | - | - | - |
| NHLRC3 | OID30614 | Inflammat | NHLRC3 | Q5JS37 | | 13 | 39038306 | 39050109 | 0 | - | - | - | - | - | - |
| NIT2 | OID30336 | Cardiome | NIT2 | Q9NQR4 | | 3 | 100334739 | 100361635 | 0 | - | - | - | - | - | - |
| NLGN1 | OID30900 | Neurology | NLGN1 | Q8N2Q7 | | 3 | 173396284 | 174294372 | 0 | - | - | - | - | - | - |
| NLGN2 | OID30878 | Neurology | NLGN2 | Q8NFZ4 | | 17 | 7404874 | 7419860 | 0 | - | - | - | - | - | - |
| NME1 | OID30451 | Inflammat | NME1 | P15531 | | 17 | 51153559 | 51162428 | 0 | - | - | - | - | - | - |
| NME3 | OID20789 | Inflammat | NME3 | Q13232 | | 16 | 1770320 | 1771561 | 0 | - | - | - | - | - | - |
| NMNAT1 | OID20913 | Neurology | NMNAT1 | Q9HAN9 | | 1 | 9943428 | 9985501 | 0 | - | - | - | - | - | - |
| NMRK2 | OID31163 | Oncology | NMRK2 | Q9NPI5 | | 19 | 3933069 | 3942416 | 0 | - | - | - | - | - | - |
| NMT1 | OID31021 | Neurology | NMT1 | P30419 | | 17 | 44957992 | 45109016 | 0 | - | - | - | - | - | - |
| NOP56 | OID30055 | Cardiome | NOP56 | O00567 | | 20 | 2652593 | 2658393 | 0 | - | - | - | - | - | - |
| NOS3 | OID20834 | Neurology | NOS3 | P29474 | | 7 | 150991017 | 151014588 | 0 | - | - | - | - | - | - |
| NOTCH1 | OID20311 | Cardiome | NOTCH1 | P46531 | | 9 | 136494433 | 136546048 | 0 | - | - | - | - | - | - |
| NOTCH3 | OID20253 | Cardiome | NOTCH3 | Q9UM47 | | 19 | 15159038 | 15200995 | 0 | - | - | - | - | - | - |
| NPC2 | OID30418 | Cardiome | NPC2 | P61916 | | 14 | 74476192 | 74494177 | 0 | - | - | - | - | - | - |
| NPHS2 | OID30462 | Inflammat | NPHS2 | Q9NP85 | | 1 | 179550539 | 179575952 | 0 | - | - | - | - | - | - |
| NPL | OID30329 | Cardiome | NPL | Q9BXD5 | | 1 | 182789293 | 182830384 | 0 | - | - | - | - | - | - |
| NPPC | OID20568 | Inflammat | NPPC | P23582 | | 2 | 231921809 | 231926396 | 0 | - | - | - | - | - | - |
| NPR1 | OID30116 | Cardiome | NPR1 | P16066 | | 1 | 153678688 | 153693992 | 0 | - | - | - | - | - | - |
| NPTN | OID21235 | Oncology | NPTN | Q9Y639 | | 15 | 73560014 | 73634134 | 0 | - | - | - | - | - | - |
| NRGN | OID30674 | Inflammat | NRGN | Q92686 | | 11 | 124739942 | 124747210 | 0 | - | - | - | - | - | - |
| NRN1 | OID30983 | Neurology | NRN1 | Q9NPD7 | | 6 | 5997999 | 6007605 | 0 | - | - | - | - | - | - |
| NRXN3 | OID30901 | Neurology | NRXN3 | Q9Y4C0 | | 14 | 78170373 | 79868291 | 0 | - | - | - | - | - | - |
| NT5C1A | OID31102 | Neurology | NT5C1A | Q9BXI3 | | 1 | 39651229 | 39672107 | 0 | - | - | - | - | - | - |
| NTF4 | OID21208 | Oncology | NTF4 | P34130 | | 19 | 49061066 | 49065054 | 0 | - | - | - | - | - | - |
| NTRK2 | OID20136 | Cardiome | NTRK2 | Q16620 | | 9 | 84668375 | 85095751 | 0 | - | - | - | - | - | - |
| NUBP1 | OID31412 | Oncology | NUBP1 | P53384 | | 16 | 10743786 | 10769351 | 0 | - | - | - | - | - | - |
| NUDC | OID20623 | Inflammat | NUDC | Q9Y266 | | 1 | 26900238 | 26946871 | 0 | - | - | - | - | - | - |
| NUDT10 | OID30198 | Cardiome | NUDT10 | Q8NFP7 | X | | 51332231 | 51337525 | 0 | - | - | - | - | - | - |
| NUDT15 | OID30102 | Cardiome | NUDT15 | Q9NV35 | | 13 | 48037726 | 48047221 | 0 | - | - | - | - | - | - |
| NUP50 | OID30100 | Cardiome | NUP50 | Q9UKX7 | | 22 | 45163925 | 45188017 | 0 | - | - | - | - | - | - |
| NXPE4 | OID30076 | Cardiome | NXPE4 | Q6UWF7 | | 11 | 114570591 | 114595786 | 0 | - | - | - | - | - | - |
| NXPH1 | OID20849 | Neurology | NXPH1 | P58417 | | 7 | 8433609 | 8752961 | 0 | - | - | - | - | - | - |
| NXPH3 | OID30565 | Inflammat | NXPH3 | O95157 | | 17 | 49575871 | 49583827 | 0 | - | - | - | - | - | - |
| OFD1 | OID31268 | Oncology | OFD1 | O75665 | X | | 13734743 | 13777955 | 0 | - | - | - | - | - | - |
| OGFR | OID21360 | Oncology | OGFR | Q9NZT2 | | 20 | 62804835 | 62814000 | 0 | - | - | - | - | - | - |
| OGT | OID31300 | Oncology | OGT | O15294 | X | | 71533087 | 71575892 | 0 | - | - | - | - | - | - |
| OPHN1 | OID31087 | Neurology | OPHN1 | O60890 | X | | 67949349 | 68433913 | 0 | - | - | - | - | - | - |
| OPTC | OID21362 | Oncology | OPTC | Q9UBM4 | | 1 | 203494153 | 203508949 | 0 | - | - | - | - | - | - |
| OSBPL2 | OID31085 | Neurology | OSBPL2 | Q9H1P3 | | 20 | 62231922 | 62296213 | 0 | - | - | - | - | - | - |
| OTUD6B | OID31487 | Oncology | OTUD6B | Q8N6M0 | | 8 | 91070196 | 91087095 | 0 | - | - | - | - | - | - |
| OTUD7B | OID31064 | Neurology | OTUD7B | Q6GQQ9 | | 1 | 149937812 | 150010726 | 0 | - | - | - | - | - | - |
| OXCT1 | OID30227 | Cardiome | OXCT1 | P55809 | | 5 | 41730065 | 41870425 | 0 | - | - | - | - | - | - |
| PAFAH1B3 | OID30998 | Neurology | PAFAH1E | Q15102 | | 19 | 42297033 | 42303546 | 0 | - | - | - | - | - | - |
| PAFAH2 | OID31286 | Oncology | PAFAH2 | Q99487 | | 1 | 25959767 | 25998117 | 0 | - | - | - | - | - | - |
| PAGR1 | OID30206 | Cardiome | PAGR1 | Q9BTK6 | | 16 | 29816152 | 29822489 | 0 | - | - | - | - | - | - |
| PAIP2B | OID31228 | Oncology | PAIP2B | Q9ULR5 | | 2 | 71182738 | 71227103 | 0 | - | - | - | - | - | - |
| PAK4 | OID20819 | Neurology | PAK4 | O96013 | | 19 | 39125770 | 39182816 | 0 | - | - | - | - | - | - |
| PALLD | OID30444 | Inflammat | PALLD | Q8WX93 | | 4 | 168497052 | 168928457 | 0 | - | - | - | - | - | - |
| PALM3 | OID30182 | Cardiome | PALM3 | A6NDB9 | | 19 | 14053363 | 14062076 | 0 | - | - | - | - | - | - |
| PBK | OID31158 | Oncology | PBK | Q96KB5 | | 8 | 27809624 | 27838082 | 0 | - | - | - | - | - | - |
| PBXIP1 | OID31435 | Oncology | PBXIP1 | Q96AQ6 | | 1 | 154944076 | 154956123 | 0 | - | - | - | - | - | - |
| PCARE | OID30838 | Neurology | PCARE | A6NGG8 | | 2 | 29060976 | 29074523 | 0 | - | - | - | - | - | - |
| PCBP2 | OID31003 | Neurology | PCBP2 | Q15366 | | 12 | 53452102 | 53481162 | 0 | - | - | - | - | - | - |
| PCNA | OID31161 | Oncology | PCNA | P12004 | | 20 | 5114953 | 5126626 | 0 | - | - | - | - | - | - |
| PCYT2 | OID31519 | Oncology | PCYT2 | Q99447 | | 17 | 81900958 | 81911432 | 0 | - | - | - | - | - | - |
| PDAP1 | OID30323 | Cardiome | PDAP1 | Q13442 | | 7 | 99392048 | 99408597 | 0 | - | - | - | - | - | - |
| PDCL2 | OID31168 | Oncology | PDCL2 | Q8N4E4 | | 4 | 55556519 | 55592245 | 0 | - | - | - | - | - | - |
| PDE1C | OID31201 | Oncology | PDE1C | Q14123 | | 7 | 31751179 | 32428131 | 0 | - | - | - | - | - | - |
| PDE4D | OID30084 | Cardiome | PDE4D | Q08499 | | 5 | 58969038 | 60522120 | 0 | - | - | - | - | - | - |
| PDGFA | OID20313 | Cardiome | PDGFA | P04085 | | 7 | 497258 | 520296 | 0 | - | - | - | - | - | - |
| PDGFB | OID20741 | Inflammat | PDGFB | P01127 | | 22 | 39223359 | 39244982 | 0 | - | - | - | - | - | - |
| PDIA2 | OID31214 | Oncology | PDIA2 | Q13087 | | 16 | 283164 | 287215 | 0 | - | - | - | - | - | - |
| PDLIM5 | OID31063 | Neurology | PDLIM5 | Q96HC4 | | 4 | 94451857 | 94668227 | 0 | - | - | - | - | - | - |
| PDLIM7 | OID20729 | Inflammat | PDLIM7 | Q9NR12 | | 5 | 177483394 | 177497606 | 0 | - | - | - | - | - | - |
| PDP1 | OID21176 | Oncology | PDP1 | Q9P0J1 | | 8 | 93857807 | 93926068 | 0 | - | - | - | - | - | - |
| PDRG1 | OID30912 | Neurology | PDRG1 | Q9NUG6 | | 20 | 31944337 | 31952046 | 0 | - | - | - | - | - | - |
| PDXDC1 | OID31226 | Oncology | PDXDC1 | Q6P996 | | 16 | 14974591 | 15139339 | 0 | - | - | - | - | - | - |
| PECAM1 | OID21131 | Neurology | PECAM1 | P16284 | | 17 | 64319415 | 64413776 | 0 | - | - | - | - | - | - |
| PFDN2 | OID20892 | Neurology | PFDN2 | Q9UHV9 | | 1 | 161100556 | 161118055 | 0 | - | - | - | - | - | - |
| PFDN4 | OID31212 | Oncology | PFDN4 | Q9NQP4 | | 20 | 54208087 | 54228052 | 0 | - | - | - | - | - | - |
| PFDN6 | OID30938 | Neurology | PFDN6 | O15212 | | 6 | 33289302 | 33298401 | 0 | - | - | - | - | - | - |
| PGM2 | OID30904 | Neurology | PGM2 | Q96G03 | | 4 | 37826660 | 37862937 | 0 | - | - | - | - | - | - |
| PGR | OID30460 | Inflammat | PGR | P06401 | | 11 | 101029624 | 101129813 | 0 | - | - | - | - | - | - |
| PHLDB1 | OID31467 | Oncology | PHLDB1 | Q86UU1 | | 11 | 118606440 | 118658031 | 0 | - | - | - | - | - | - |
| PHLDB2 | OID31167 | Oncology | PHLDB2 | Q86SQ0 | | 3 | 111732497 | 111976517 | 0 | - | - | - | - | - | - |
| PHOSPHO1 | OID20922 | Neurology | PHOSPH( | Q8TCT1 | | 17 | 49223366 | 49230787 | 0 | - | - | - | - | - | - |
| PHYKPL | OID30634 | Inflammat | PHYKPL | Q8IUZ5 | | 5 | 178208471 | 178232802 | 0 | - | - | - | - | - | - |
| PIKFYVE | OID30465 | Inflammat | PIKFYVE | Q9Y2I7 | | 2 | 208266255 | 208358746 | 0 | - | - | - | - | - | - |
| PITHD1 | OID31215 | Oncology | PITHD1 | Q9GZP4 | | 1 | 23778418 | 23788232 | 0 | - | - | - | - | - | - |
| PLA2G10 | OID21002 | Neurology | PLA2G10 | O15496 | | 16 | 14672548 | 14694308 | 0 | - | - | - | - | - | - |
| PLA2G7 | OID21105 | Neurology | PLA2G7 | Q13093 | | 6 | 46704201 | 46735693 | 0 | - | - | - | - | - | - |
| PLAU | OID21124 | Neurology | PLAU | P00749 | | 10 | 73909177 | 73917496 | 0 | - | - | - | - | - | - |
| PLCB1 | OID30484 | Inflammat | PLCB1 | Q9NQ66 | | 20 | 8077251 | 8968360 | 0 | - | - | - | - | - | - |
| PLG | OID30785 | Inflammat | PLG | P00747 | | 6 | 160702238 | 160754097 | 0 | - | - | - | - | - | - |

| | | | | | | | | | | | | | | | |
|---|---|---|---|---|---|---|---|---|---|---|---|---|---|---|---|
| PLIN1 | OID20835 | Neurology | PLIN1 | O60240 | | 15 | 89664367 | 89679427 | 0 | - | - | - | - | - | - |
| PLPBP | OID20144 | Cardiome | PLPBP | O94903 | | 8 | 37762595 | 37779768 | 0 | - | - | - | - | - | - |
| PLXNB3 | OID20160 | Cardiome | PLXNB3 | Q9ULL4 | X | | 153764196 | 153779346 | 0 | - | - | - | - | - | - |
| PMCH | OID30172 | Cardiome | PMCH | P20382 | | 12 | 102196459 | 102197833 | 0 | - | - | - | - | - | - |
| PMS1 | OID30943 | Neurology | PMS1 | P54277 | | 2 | 189784085 | 189877629 | 0 | - | - | - | - | - | - |
| PMVK | OID20850 | Neurology | PMVK | Q15126 | | 1 | 154924740 | 154936719 | 0 | - | - | - | - | - | - |
| PNLIP | OID30693 | Inflammat | PNLIP | P16233 | | 10 | 116545931 | 116567855 | 0 | - | - | - | - | - | - |
| PNMA1 | OID30194 | Cardiome | PNMA1 | Q8ND90 | | 14 | 73711783 | 73714384 | 0 | - | - | - | - | - | - |
| PNMA2 | OID30842 | Neurology | PNMA2 | Q9UL42 | | 8 | 26504701 | 26514092 | 0 | - | - | - | - | - | - |
| PNPT1 | OID20445 | Inflammat | PNPT1 | Q8TCS8 | | 2 | 55634061 | 55693863 | 0 | - | - | - | - | - | - |
| POF1B | OID30504 | Inflammat | POF1B | Q8WVV4 | X | | 85277396 | 85379717 | 0 | - | - | - | - | - | - |
| POLR2A | OID30440 | Inflammat | POLR2A | P24928 | | 17 | 7484366 | 7514616 | 0 | - | - | - | - | - | - |
| POLR2F | OID21283 | Oncology | POLR2F | P61218 | | 22 | 37952607 | 38041915 | 0 | - | - | - | - | - | - |
| PPBP | OID30761 | Inflammat | PPBP | P02775 | | 4 | 73986439 | 73988190 | 0 | - | - | - | - | - | - |
| PPIB | OID20331 | Cardiome | PPIB | P23284 | | 15 | 64155740 | 64163134 | 0 | - | - | - | - | - | - |
| PPIE | OID31279 | Oncology | PPIE | Q9UNP9 | | 1 | 39692182 | 39763914 | 0 | - | - | - | - | - | - |
| PPIF | OID30385 | Cardiome | PPIF | P30405 | | 10 | 79347469 | 79355334 | 0 | - | - | - | - | - | - |
| PPM1A | OID21184 | Oncology | PPM1A | P35813 | | 14 | 60245752 | 60299087 | 0 | - | - | - | - | - | - |
| PPM1B | OID30548 | Inflammat | PPM1B | O75688 | | 2 | 44167969 | 44244384 | 0 | - | - | - | - | - | - |
| PPME1 | OID21347 | Oncology | PPME1 | Q9Y570 | | 11 | 74171267 | 74254703 | 0 | - | - | - | - | - | - |
| PPP1CC | OID31360 | Oncology | PPP1CC | P36873 | | 12 | 110719680 | 110742939 | 0 | - | - | - | - | - | - |
| PPP1R12A | OID21475 | Oncology | PPP1R12 | O14974 | | 12 | 79773563 | 79935460 | 0 | - | - | - | - | - | - |
| PPP1R12B | OID31304 | Oncology | PPP1R12 | O60237 | | 1 | 202348699 | 202592706 | 0 | - | - | - | - | - | - |
| PPP1R2 | OID20176 | Cardiome | PPP1R2 | P41236 | | 3 | 195514428 | 195543386 | 0 | - | - | - | - | - | - |
| PPP1R9B | OID20643 | Inflammat | PPP1R9B | Q96SB3 | | 17 | 50133737 | 50150677 | 0 | - | - | - | - | - | - |
| PPP3R1 | OID20902 | Neurology | PPP3R1 | P63098 | | 2 | 68178857 | 68256237 | 0 | - | - | - | - | - | - |
| PPT1 | OID30947 | Neurology | PPT1 | P50897 | | 1 | 40072710 | 40097260 | 0 | - | - | - | - | - | - |
| PPY | OID21516 | Oncology | PPY | P01298 | | 17 | 43940804 | 43942476 | 0 | - | - | - | - | - | - |
| PQBP1 | OID21172 | Oncology | PQBP1 | O60828 | X | | 48890197 | 48903402 | 0 | - | - | - | - | - | - |
| PRAME | OID31165 | Oncology | PRAME | P78395 | | 22 | 22547701 | 22559361 | 0 | - | - | - | - | - | - |
| PRC1 | OID31345 | Oncology | PRC1 | O43663 | | 15 | 90966040 | 90995629 | 0 | - | - | - | - | - | - |
| PRCP | OID20337 | Cardiome | PRCP | P42785 | | 11 | 82822936 | 82970584 | 0 | - | - | - | - | - | - |
| PRDX5 | OID20644 | Inflammat | PRDX5 | P30044 | | 11 | 64318121 | 64321811 | 0 | - | - | - | - | - | - |
| PREB | OID20439 | Inflammat | PREB | Q9HCU5 | | 2 | 27130756 | 27134666 | 0 | - | - | - | - | - | - |
| PRKAG3 | OID30941 | Neurology | PRKAG3 | Q9UGI9 | | 2 | 218822308 | 218832086 | 0 | - | - | - | - | - | - |
| PRKAR1A | OID20146 | Cardiome | PRKAR1A | P10644 | | 17 | 68511780 | 68551319 | 0 | - | - | - | - | - | - |
| PRKAR2A | OID31119 | Neurology | PRKAR2A | P13861 | | 3 | 48744597 | 48847874 | 0 | - | - | - | - | - | - |
| PRKCQ | OID20468 | Inflammat | PRKCQ | Q04759 | | 10 | 6427143 | 6580301 | 0 | - | - | - | - | - | - |
| PRKD2 | OID30140 | Cardiome | PRKD2 | Q9BZL6 | | 19 | 46674275 | 46717127 | 0 | - | - | - | - | - | - |
| PRKRA | OID21225 | Oncology | PRKRA | O75569 | | 2 | 178431292 | 178451512 | 0 | - | - | - | - | - | - |
| PRL | OID21033 | Neurology | PRL | P01236 | | 6 | 22287244 | 22302826 | 0 | - | - | - | - | - | - |
| PROS1 | OID30737 | Inflammat | PROS1 | P07225 | | 3 | 93873051 | 93980003 | 0 | - | - | - | - | - | - |
| PRR5 | OID30457 | Inflammat | PRR5 | P85299 | | 22 | 44668547 | 44737681 | 0 | - | - | - | - | - | - |
| PRUNE2 | OID31270 | Oncology | PRUNE2 | Q8WUY3 | | 9 | 76611376 | 76906114 | 0 | - | - | - | - | - | - |
| PSIP1 | OID20534 | Inflammat | PSIP1 | O75475 | | 9 | 15464066 | 15510995 | 0 | - | - | - | - | - | - |
| PSMA1 | OID21221 | Oncology | PSMA1 | P25786 | | 11 | 14504874 | 14643635 | 0 | - | - | - | - | - | - |
| PSMC3 | OID31200 | Oncology | PSMC3 | P17980 | | 11 | 47418769 | 47426473 | 0 | - | - | - | - | - | - |
| PSMD1 | OID31058 | Neurology | PSMD1 | Q99460 | | 2 | 231056845 | 231173116 | 0 | - | - | - | - | - | - |
| PSMD5 | OID30918 | Neurology | PSMD5 | Q16401 | | 9 | 120815496 | 120842951 | 0 | - | - | - | - | - | - |
| PSMG3 | OID20549 | Inflammat | PSMG3 | Q9BT73 | | 7 | 1567332 | 1571005 | 0 | - | - | - | - | - | - |
| PSPN | OID20511 | Inflammat | PSPN | O60542 | | 19 | 6375148 | 6379058 | 0 | - | - | - | - | - | - |
| PTEN | OID20794 | Neurology | PTEN | P60484 | | 10 | 87862638 | 87971930 | 0 | - | - | - | - | - | - |
| PTH | OID30963 | Neurology | PTH | P01270 | | 11 | 13492054 | 13496181 | 0 | - | - | - | - | - | - |
| PTH1R | OID20571 | Inflammat | PTH1R | Q03431 | | 3 | 46877721 | 46903799 | 0 | - | - | - | - | - | - |
| PTP4A3 | OID30469 | Inflammat | PTP4A3 | O75365 | | 8 | 141391995 | 141432454 | 0 | - | - | - | - | - | - |
| PTPN1 | OID20908 | Neurology | PTPN1 | P18031 | | 20 | 50510321 | 50585241 | 0 | - | - | - | - | - | - |
| PTPN6 | OID20717 | Inflammat | PTPN6 | P29350 | | 12 | 6946468 | 6961316 | 0 | - | - | - | - | - | - |
| PTPN9 | OID30437 | Inflammat | PTPN9 | P43378 | | 15 | 75463251 | 75579315 | 0 | - | - | - | - | - | - |
| PTPRR | OID31099 | Neurology | PTPRR | Q15256 | | 12 | 70638073 | 70920738 | 0 | - | - | - | - | - | - |
| PTRHD1 | OID30659 | Inflammat | PTRHD1 | Q6GMV3 | | 2 | 24789728 | 24793391 | 0 | - | - | - | - | - | - |
| PTTG1 | OID31326 | Oncology | PTTG1 | O95997 | | 5 | 160421855 | 160428739 | 0 | - | - | - | - | - | - |
| RAB10 | OID30357 | Cardiome | RAB10 | P61026 | | 2 | 26034084 | 26137454 | 0 | - | - | - | - | - | - |
| RAB27B | OID30354 | Cardiome | RAB27B | O00194 | | 18 | 54717860 | 54895516 | 0 | - | - | - | - | - | - |
| RAB33A | OID30122 | Cardiome | RAB33A | Q14088 | X | | 130171962 | 130184870 | 0 | - | - | - | - | - | - |
| RAB37 | OID20425 | Inflammat | RAB37 | Q96AX2 | | 17 | 74670578 | 74747335 | 0 | - | - | - | - | - | - |
| RAB39B | OID30069 | Cardiome | RAB39B | Q96DA2 | X | | 155258235 | 155264491 | 0 | - | - | - | - | - | - |
| RAB3GAP1 | OID30829 | Neurology | RAB3GAF | Q15042 | | 2 | 135052289 | 135176667 | 0 | - | - | - | - | - | - |
| RAB6B | OID20817 | Neurology | RAB6B | Q9NRW1 | | 3 | 133824235 | 133895882 | 0 | - | - | - | - | - | - |
| RAC3 | OID30958 | Neurology | RAC3 | P60763 | | 17 | 82031678 | 82034204 | 0 | - | - | - | - | - | - |
| RAD51 | OID31254 | Oncology | RAD51 | Q06609 | | 15 | 40694774 | 40732340 | 0 | - | - | - | - | - | - |
| RANBP2 | OID30812 | Neurology | RANBP2 | P49792 | | 2 | 108719482 | 108785810 | 0 | - | - | - | - | - | - |
| RANGAP1 | OID21203 | Oncology | RANGAP | P46060 | | 22 | 41244779 | 41286187 | 0 | - | - | - | - | - | - |
| RAP1A | OID30452 | Inflammat | RAP1A | P62834 | | 1 | 111542218 | 111716691 | 0 | - | - | - | - | - | - |
| RAPGEF2 | OID31337 | Oncology | RAPGEF | Q9Y4G8 | | 4 | 159103013 | 159360174 | 0 | - | - | - | - | - | - |
| RASA1 | OID20826 | Neurology | RASA1 | P20936 | | 5 | 87267883 | 87391931 | 0 | - | - | - | - | - | - |
| RASGRF1 | OID31213 | Oncology | RASGRF | Q13972 | | 15 | 78959906 | 79090780 | 0 | - | - | - | - | - | - |
| RBFOX3 | OID30984 | Neurology | RBFOX3 | A6NFN3 | | 17 | 79089345 | 79611051 | 0 | - | - | - | - | - | - |
| RBKS | OID20986 | Neurology | RBKS | Q9H477 | | 2 | 27781379 | 27890681 | 0 | - | - | - | - | - | - |
| RBM19 | OID30090 | Cardiome | RBM19 | Q9Y4C8 | | 12 | 113816738 | 113966325 | 0 | - | - | - | - | - | - |
| RBM25 | OID31236 | Oncology | RBM25 | P49756 | | 14 | 73058532 | 73123899 | 0 | - | - | - | - | - | - |
| RBPMS | OID30507 | Inflammat | RBPMS | Q93062 | | 8 | 30384511 | 30572256 | 0 | - | - | - | - | - | - |
| RCC1 | OID31356 | Oncology | RCC1 | P18754 | | 1 | 28505943 | 28539300 | 0 | - | - | - | - | - | - |
| RCOR1 | OID20073 | Cardiome | RCOR1 | Q9UKL0 | | 14 | 102592649 | 102730561 | 0 | - | - | - | - | - | - |
| RELB | OID30463 | Inflammat | RELB | Q01201 | | 19 | 45001461 | 45038192 | 0 | - | - | - | - | - | - |
| REN | OID20201 | Cardiome | REN | P00797 | | 1 | 204154819 | 204190324 | 0 | - | - | - | - | - | - |
| REPS1 | OID30496 | Inflammat | REPS1 | Q96D71 | | 6 | 138903493 | 138988261 | 0 | - | - | - | - | - | - |
| REST | OID31248 | Oncology | REST | Q13127 | | 4 | 56907876 | 56966808 | 0 | - | - | - | - | - | - |
| RETN | OID20327 | Cardiome | RETN | Q9HD89 | | 19 | 7669049 | 7670455 | 0 | - | - | - | - | - | - |
| REXO2 | OID30075 | Cardiome | REXO2 | Q9Y3B8 | | 11 | 114439435 | 114450279 | 0 | - | - | - | - | - | - |
| RFC4 | OID31376 | Oncology | RFC4 | P35249 | | 3 | 186789880 | 186807058 | 0 | - | - | - | - | - | - |
| RGCC | OID31060 | Neurology | RGCC | Q9H4X1 | | 13 | 41457550 | 41470871 | 0 | - | - | - | - | - | - |
| RGL2 | OID31297 | Oncology | RGL2 | O15211 | | 6 | 33291654 | 33298942 | 0 | - | - | - | - | - | - |
| RGS10 | OID30999 | Neurology | RGS10 | O43665 | | 10 | 119499817 | 119542719 | 0 | - | - | - | - | - | - |
| RGS8 | OID20454 | Inflammat | RGS8 | P57771 | | 1 | 182641816 | 182684576 | 0 | - | - | - | - | - | - |
| RICTOR | OID30433 | Inflammat | RICTOR | Q6R327 | | 5 | 38937920 | 39074399 | 0 | - | - | - | - | - | - |
| RIPK4 | OID30138 | Cardiome | RIPK4 | P57078 | | 21 | 41739369 | 41767089 | 0 | - | - | - | - | - | - |
| RLN1 | OID30995 | Neurology | RLN1 | P04808 | | 9 | 5334930 | 5339876 | 0 | - | - | - | - | - | - |
| RNF168 | OID30506 | Inflammat | RNF168 | Q8IYW5 | | 3 | 196468783 | 196503768 | 0 | - | - | - | - | - | - |
| RNF31 | OID30473 | Inflammat | RNF31 | Q96EP0 | | 14 | 24146683 | 24160660 | 0 | - | - | - | - | - | - |
| RNF4 | OID31321 | Oncology | RNF4 | P78317 | | 4 | 2462220 | 2515857 | 0 | - | - | - | - | - | - |
| RNF41 | OID21179 | Oncology | RNF41 | Q9H4P4 | | 12 | 56202179 | 56221933 | 0 | - | - | - | - | - | - |
| RNF5 | OID30218 | Cardiome | RNF5 | Q99942 | | 6 | 32178405 | 32180793 | 0 | - | - | - | - | - | - |
| RP2 | OID21302 | Oncology | RP2 | O75695 | X | | 46837043 | 46882358 | 0 | - | - | - | - | - | - |
| RPGR | OID30959 | Neurology | RPGR | Q92834 | X | | 38269163 | 38327544 | 0 | - | - | - | - | - | - |
| RPS10 | OID30070 | Cardiome | RPS10 | P46783 | | 6 | 34417454 | 34426069 | 0 | - | - | - | - | - | - |
| RRAS | OID30961 | Neurology | RRAS | P10301 | | 19 | 49635292 | 49640143 | 0 | - | - | - | - | - | - |
| RRM2 | OID21187 | Oncology | RRM2 | P31350 | | 2 | 10120698 | 10211725 | 0 | - | - | - | - | - | - |

| | | | | | | | | | | | | | | | |
|---|---|---|---|---|---|---|---|---|---|---|---|---|---|---|---|
| RRP15 | OID30867 | Neurology | RRP15 | Q9Y3B9 | | 1 | 218285293 | 218337983 | 0 | - | - | - | - | - | - |
| RTKN2 | OID30049 | Cardiome | RTKN2 | Q8IZC4 | | 10 | 62183035 | 62268844 | 0 | - | - | - | - | - | - |
| RUVBL1 | OID21211 | Oncology | RUVBL1 | Q9Y265 | | 3 | 128064778 | 128153914 | 0 | - | - | - | - | - | - |
| RYR1 | OID30085 | Cardiome | RYR1 | P21817 | | 19 | 38433691 | 38595273 | 0 | - | - | - | - | - | - |
| S100A3 | OID31352 | Oncology | S100A3 | P33764 | | 1 | 153547329 | 153549258 | 0 | - | - | - | - | - | - |
| S100G | OID31051 | Neurology | S100G | P29377 | X | | 16650158 | 16654670 | 0 | - | - | - | - | - | - |
| SAFB2 | OID31012 | Neurology | SAFB2 | Q14151 | | 19 | 5586999 | 5624046 | 0 | - | - | - | - | - | - |
| SAG | OID30874 | Neurology | SAG | P10523 | | 2 | 233307816 | 233347055 | 0 | - | - | - | - | - | - |
| SAP18 | OID31323 | Oncology | SAP18 | O00422 | | 13 | 21140119 | 21149097 | 0 | - | - | - | - | - | - |
| SART1 | OID30113 | Cardiome | SART1 | O43290 | | 11 | 65961728 | 65980137 | 0 | - | - | - | - | - | - |
| SAT1 | OID30111 | Cardiome | SAT1 | P21673 | X | | 23783173 | 23786210 | 0 | - | - | - | - | - | - |
| SATB1 | OID31038 | Neurology | SATB1 | Q01826 | | 3 | 18345377 | 18445621 | 0 | - | - | - | - | - | - |
| SCARB1 | OID20805 | Neurology | SCARB1 | Q8WTV0 | | 12 | 124776856 | 124882668 | 0 | - | - | - | - | - | - |
| SCG2 | OID21400 | Oncology | SCG2 | P13521 | | 2 | 223596940 | 223602361 | 0 | - | - | - | - | - | - |
| SCGB2A2 | OID30104 | Cardiome | SCGB2A2 | Q13296 | | 11 | 62270158 | 62273160 | 0 | - | - | - | - | - | - |
| SCIN | OID30857 | Neurology | SCIN | Q9Y6U3 | | 7 | 12570577 | 12660182 | 0 | - | - | - | - | - | - |
| SCN2A | OID30833 | Neurology | SCN2A | Q99250 | | 2 | 165194993 | 165392310 | 0 | - | - | - | - | - | - |
| SCN2B | OID30948 | Neurology | SCN2B | O60939 | | 11 | 118162806 | 118176639 | 0 | - | - | - | - | - | - |
| SCN3A | OID30903 | Neurology | SCN3A | Q9NY46 | | 2 | 165087526 | 165204050 | 0 | - | - | - | - | - | - |
| SCN3B | OID30804 | Neurology | SCN3B | Q9NY72 | | 11 | 123629187 | 123655244 | 0 | - | - | - | - | - | - |
| SCP2 | OID21161 | Oncology | SCP2 | P22307 | | 1 | 52927276 | 53051698 | 0 | - | - | - | - | - | - |
| SCRIB | OID30225 | Cardiome | SCRIB | Q14160 | | 8 | 143790920 | 143815773 | 0 | - | - | - | - | - | - |
| SCT | OID30951 | Neurology | SCT | P09683 | | 11 | 626309 | 627181 | 0 | - | - | - | - | - | - |
| SEMA3G | OID30599 | Inflammat | SEMA3G | Q9NS98 | | 3 | 52433035 | 52445103 | 0 | - | - | - | - | - | - |
| SEMA4C | OID21329 | Oncology | SEMA4C | Q9C0C4 | | 2 | 96859718 | 96870757 | 0 | - | - | - | - | - | - |
| SEPTIN3 | OID31025 | Neurology | SEPTIN3 | Q9UH03 | | 22 | 41969443 | 41998221 | 0 | - | - | - | - | - | - |
| SEPTIN7 | OID31288 | Oncology | SEPTIN7 | Q16181 | | 7 | 35800932 | 35907110 | 0 | - | - | - | - | - | - |
| SERPINA7 | OID30724 | Inflammat | SERPINA | P05543 | X | | 106032435 | 106038727 | 0 | - | - | - | - | - | - |
| SERPINC1 | OID30789 | Inflammat | SERPINC | P01008 | | 1 | 173903800 | 173917327 | 0 | - | - | - | - | - | - |
| SERPIND1 | OID30767 | Inflammat | SERPIND | P05546 | | 22 | 20774113 | 20787720 | 0 | - | - | - | - | - | - |
| SH2D1A | OID20514 | Inflammat | SH2D1A | O60880 | X | | 124227868 | 124373197 | 0 | - | - | - | - | - | - |
| SH3BGRL2 | OID30863 | Neurology | SH3BGRL | Q9UJC5 | | 6 | 79631329 | 79703655 | 0 | - | - | - | - | - | - |
| SH3GL3 | OID31334 | Oncology | SH3GL3 | Q99963 | | 15 | 83447228 | 83618743 | 0 | - | - | - | - | - | - |
| SHC1 | OID31222 | Oncology | SHC1 | P29353 | | 1 | 154962298 | 154974395 | 0 | - | - | - | - | - | - |
| SHD | OID30098 | Cardiome | SHD | Q96IW2 | | 19 | 4279266 | 4290722 | 0 | - | - | - | - | - | - |
| SIGLEC15 | OID20812 | Neurology | SIGLEC1! | Q6ZMC9 | | 18 | 45825675 | 45844094 | 0 | - | - | - | - | - | - |
| SIRT1 | OID30439 | Inflammat | SIRT1 | Q96EB6 | | 10 | 67884656 | 67918390 | 0 | - | - | - | - | - | - |
| SIRT2 | OID21375 | Oncology | SIRT2 | Q8IXJ6 | | 19 | 38878555 | 38899862 | 0 | - | - | - | - | - | - |
| SIRT5 | OID20820 | Neurology | SIRT5 | Q9NXA8 | | 6 | 13574227 | 13615158 | 0 | - | - | - | - | - | - |
| SIT1 | OID20462 | Inflammat | SIT1 | Q9Y3P8 | | 9 | 35649295 | 35650931 | 0 | - | - | - | - | - | - |
| SKAP2 | OID20761 | Inflammat | SKAP2 | O75563 | | 7 | 26667068 | 26995239 | 0 | - | - | - | - | - | - |
| SKIV2L | OID30056 | Cardiome | SKIV2L | Q15477 | | 6 | 31959116 | 31969818 | 0 | - | - | - | - | - | - |
| SLA2 | OID31017 | Neurology | SLA2 | Q9H6Q3 | | 20 | 36612318 | 36646196 | 0 | - | - | - | - | - | - |
| SLC12A2 | OID30077 | Cardiome | SLC12A2 | P55011 | | 5 | 128083766 | 128189677 | 0 | - | - | - | - | - | - |
| SLC13A1 | OID31388 | Oncology | SLC13A1 | Q9BZW2 | | 7 | 123113531 | 123199972 | 0 | - | - | - | - | - | - |
| SLC1A4 | OID30926 | Neurology | SLC1A4 | P43007 | | 2 | 64988477 | 65023865 | 0 | - | - | - | - | - | - |
| SLC28A1 | OID31220 | Oncology | SLC28A1 | O00337 | | 15 | 84884654 | 84945798 | 0 | - | - | - | - | - | - |
| SLC34A3 | OID31203 | Oncology | SLC34A3 | Q8N130 | | 9 | 137230757 | 137236555 | 0 | - | - | - | - | - | - |
| SLC39A5 | OID20539 | Inflammat | SLC39A5 | Q6ZMH5 | | 12 | 56230049 | 56237846 | 0 | - | - | - | - | - | - |
| SLC44A4 | OID30897 | Neurology | SLC44A4 | Q53GD3 | | 6 | 31863192 | 31879046 | 0 | - | - | - | - | - | - |
| SLC51B | OID30107 | Cardiome | SLC51B | Q86UW2 | | 15 | 65045387 | 65053397 | 0 | - | - | - | - | - | - |
| SLIRP | OID31174 | Oncology | SLIRP | Q9GZT3 | | 14 | 77708071 | 77761104 | 0 | - | - | - | - | - | - |
| SLITRK2 | OID21409 | Oncology | SLITRK2 | Q9H156 | X | | 145817829 | 145829856 | 0 | - | - | - | - | - | - |
| SLK | OID31282 | Oncology | SLK | Q9H2G2 | | 10 | 103967140 | 104029233 | 0 | - | - | - | - | - | - |
| SMAD1 | OID21180 | Oncology | SMAD1 | Q15797 | | 4 | 145481194 | 145559176 | 0 | - | - | - | - | - | - |
| SMAD2 | OID31353 | Oncology | SMAD2 | Q15796 | | 18 | 47808957 | 47931146 | 0 | - | - | - | - | - | - |
| SMAD5 | OID21353 | Oncology | SMAD5 | Q99717 | | 5 | 136132845 | 136188747 | 0 | - | - | - | - | - | - |
| SMARCA2 | OID20796 | Neurology | SMARCA: | P51531 | | 9 | 1980290 | 2193624 | 0 | - | - | - | - | - | - |
| SMC3 | OID31195 | Oncology | SMC3 | Q9UQE7 | | 10 | 110567684 | 110606048 | 0 | - | - | - | - | - | - |
| SMNDC1 | OID31471 | Oncology | SMNDC1 | O75940 | | 10 | 110290730 | 110304938 | 0 | - | - | - | - | - | - |
| SMPD3 | OID30477 | Inflammat | SMPD3 | Q9NY59 | | 16 | 68358327 | 68448508 | 0 | - | - | - | - | - | - |
| SMPDL3B | OID31387 | Oncology | SMPDL3E | Q92485 | | 1 | 27935000 | 27959157 | 0 | - | - | - | - | - | - |
| SMS | OID30853 | Neurology | SMS | P52788 | X | | 21940709 | 21994837 | 0 | - | - | - | - | - | - |
| SMTN | OID31154 | Neurology | SMTN | P53814 | | 22 | 31064105 | 31104757 | 0 | - | - | - | - | - | - |
| SNAP23 | OID20218 | Cardiome | SNAP23 | O00161 | | 15 | 42491233 | 42545356 | 0 | - | - | - | - | - | - |
| SNAPIN | OID31019 | Neurology | SNAPIN | O95295 | | 1 | 153658703 | 153661852 | 0 | - | - | - | - | - | - |
| SNCA | OID30645 | Inflammat | SNCA | P37840 | | 4 | 89700345 | 89838315 | 0 | - | - | - | - | - | - |
| SNRPB2 | OID31132 | Neurology | SNRPB2 | P08579 | | 20 | 16729961 | 16742564 | 0 | - | - | - | - | - | - |
| SNU13 | OID30052 | Cardiome | SNU13 | P55769 | | 22 | 41673933 | 41690504 | 0 | - | - | - | - | - | - |
| SNX5 | OID30215 | Cardiome | SNX5 | Q9Y5X3 | | 20 | 17941597 | 17968980 | 0 | - | - | - | - | - | - |
| SORBS1 | OID30978 | Neurology | SORBS1 | Q9BX66 | | 10 | 95311771 | 95561414 | 0 | - | - | - | - | - | - |
| SOWAHA | OID30888 | Neurology | SOWAHA | Q2M3V2 | | 5 | 132813302 | 132816786 | 0 | - | - | - | - | - | - |
| SOX2 | OID30981 | Neurology | SOX2 | P48431 | | 3 | 181711925 | 181714436 | 0 | - | - | - | - | - | - |
| SOX9 | OID30905 | Neurology | SOX9 | P48436 | | 17 | 72121020 | 72126416 | 0 | - | - | - | - | - | - |
| SPACA5_SPA | OID30990 | Neurology | SPACA5E | Q96QH8;( | X;X | | 48130626;4{ | 48132620;4{ | 0 | - | - | - | - | - | - |
| SPARC | OID21511 | Oncology | SPARC | P09486 | | 5 | 151661096 | 151686975 | 0 | - | - | - | - | - | - |
| SPART | OID30656 | Inflammat | SPART | Q8N0X7 | | 13 | 36301638 | 36370180 | 0 | - | - | - | - | - | - |
| SPESP1 | OID30298 | Cardiome | SPESP1 | Q6UW49 | | 15 | 68818221 | 68946811 | 0 | - | - | - | - | - | - |
| SPRING1 | OID31197 | Oncology | SPRING1 | Q9H741 | | 12 | 116710171 | 116738070 | 0 | - | - | - | - | - | - |
| SPRR1B | OID31296 | Oncology | SPRR1B | P22528 | | 1 | 153031203 | 153032900 | 0 | - | - | - | - | - | - |
| SPTBN2 | OID30876 | Neurology | SPTBN2 | O15020 | | 11 | 66682497 | 66744670 | 0 | - | - | - | - | - | - |
| SRC | OID21521 | Oncology | SRC | P12931 | | 20 | 37344685 | 37406050 | 0 | - | - | - | - | - | - |
| SRP14 | OID21324 | Oncology | SRP14 | P37108 | | 15 | 40035690 | 40039181 | 0 | - | - | - | - | - | - |
| SRPK2 | OID20502 | Inflammat | SRPK2 | P78362 | | 7 | 105110704 | 105399308 | 0 | - | - | - | - | - | - |
| SRPX | OID30310 | Cardiome | SRPX | P78539 | X | | 38149336 | 38220924 | 0 | - | - | - | - | - | - |
| SSB | OID20818 | Neurology | SSB | P05455 | | 2 | 169791933 | 169812064 | 0 | - | - | - | - | - | - |
| SSBP1 | OID30468 | Inflammat | SSBP1 | Q04837 | | 7 | 141738334 | 141787922 | 0 | - | - | - | - | - | - |
| SSH3 | OID31216 | Oncology | SSH3 | Q8TE77 | | 11 | 67303478 | 67312607 | 0 | - | - | - | - | - | - |
| SSNA1 | OID31453 | Oncology | SSNA1 | O43805 | | 9 | 137188660 | 137190370 | 0 | - | - | - | - | - | - |
| ST8SIA1 | OID30442 | Inflammat | ST8SIA1 | Q92185 | | 12 | 22063773 | 22437041 | 0 | - | - | - | - | - | - |
| STAM | OID31425 | Oncology | STAM | Q92783 | | 10 | 17644151 | 17716824 | 0 | - | - | - | - | - | - |
| STAMBP | OID20955 | Neurology | STAMBP | O95630 | | 2 | 73828916 | 73873659 | 0 | - | - | - | - | - | - |
| STAT5B | OID21277 | Oncology | STAT5B | P51692 | | 17 | 42199177 | 42276707 | 0 | - | - | - | - | - | - |
| STAU1 | OID31210 | Oncology | STAU1 | O95793 | | 20 | 49113339 | 49188367 | 0 | - | - | - | - | - | - |
| STEAP4 | OID30846 | Neurology | STEAP4 | Q687X5 | | 7 | 88270892 | 88306894 | 0 | - | - | - | - | - | - |
| STIP1 | OID21077 | Neurology | STIP1 | P31948 | | 11 | 64185272 | 64204543 | 0 | - | - | - | - | - | - |
| STK11 | OID20077 | Cardiome | STK11 | Q15831 | | 19 | 1177558 | 1228431 | 0 | - | - | - | - | - | - |
| STK24 | OID20837 | Neurology | STK24 | Q9Y6E0 | | 13 | 98445185 | 98577940 | 0 | - | - | - | - | - | - |
| STOML2 | OID31183 | Oncology | STOML2 | Q9UJZ1 | | 9 | 35099776 | 35103195 | 0 | - | - | - | - | - | - |
| STX1B | OID30934 | Neurology | STX1B | P61266 | | 16 | 30989256 | 31010638 | 0 | - | - | - | - | - | - |
| STX3 | OID31090 | Neurology | STX3 | Q13277 | | 11 | 59713456 | 59805882 | 0 | - | - | - | - | - | - |
| STX4 | OID21326 | Oncology | STX4 | Q12846 | | 16 | 31032889 | 31042975 | 0 | - | - | - | - | - | - |
| STX5 | OID30432 | Inflammat | STX5 | Q13190 | | 11 | 62806860 | 62832051 | 0 | - | - | - | - | - | - |
| STX6 | OID21219 | Oncology | STX6 | O43752 | | 1 | 180972712 | 181023121 | 0 | - | - | - | - | - | - |
| STXBP3 | OID21240 | Oncology | STXBP3 | O00186 | | 1 | 108746674 | 108809523 | 0 | - | - | - | - | - | - |
| SUGT1 | OID21229 | Oncology | SUGT1 | Q9Y2Z0 | | 13 | 52652709 | 52700909 | 0 | - | - | - | - | - | - |
| SUOX | OID31031 | Neurology | SUOX | P51687 | | 12 | 55997180 | 56006641 | 0 | - | - | - | - | - | - |

| | | | | | | | | | | | | | | | |
|---|---|---|---|---|---|---|---|---|---|---|---|---|---|---|---|
| SUSD1 | OID20072 | Cardiome | SUSD1 | Q6UWL2 | | 9 | 112040783 | 112175297 | 0 | - | - | - | - | - | - |
| SYAP1 | OID30636 | Inflammat | SYAP1 | Q96A49 | X | | 16719612 | 16765340 | 0 | - | - | - | - | - | - |
| SYNGAP1 | OID30797 | Neurology | SYNGAP | Q96PV0 | | 6 | 33419661 | 33453689 | 0 | - | - | - | - | - | - |
| SYT1 | OID31089 | Neurology | SYT1 | P21579 | | 12 | 78863993 | 79452008 | 0 | - | - | - | - | - | - |
| SYTL4 | OID30339 | Cardiome | SYTL4 | Q96C24 | X | | 100671783 | 100732123 | 0 | - | - | - | - | - | - |
| TAB2 | OID31319 | Oncology | TAB2 | Q9NYJ8 | | 6 | 149218641 | 149411613 | 0 | - | - | - | - | - | - |
| TADA3 | OID31422 | Oncology | TADA3 | O75528 | | 3 | 9779967 | 9793011 | 0 | - | - | - | - | - | - |
| TAGLN3 | OID31225 | Oncology | TAGLN3 | Q9UI15 | | 3 | 111998739 | 112013887 | 0 | - | - | - | - | - | - |
| TALDO1 | OID30414 | Cardiome | TALDO1 | P37837 | | 11 | 747415 | 765012 | 0 | - | - | - | - | - | - |
| TANK | OID20474 | Inflammat | TANK | Q92844 | | 2 | 161136908 | 161236230 | 0 | - | - | - | - | - | - |
| TAP1 | OID31289 | Oncology | TAP1 | Q03518 | | 6 | 32845209 | 32853816 | 0 | - | - | - | - | - | - |
| TARBP2 | OID20870 | Neurology | TARBP2 | Q15633 | | 12 | 53500921 | 53506431 | 0 | - | - | - | - | - | - |
| TARM1 | OID30937 | Neurology | TARM1 | B6A8C7 | | 19 | 54069895 | 54081365 | 0 | - | - | - | - | - | - |
| TARS1 | OID31343 | Oncology | TARS1 | P26639 | | 5 | 33440696 | 33468091 | 0 | - | - | - | - | - | - |
| TAX1BP1 | OID31075 | Neurology | TAX1BP1 | Q86VP1 | | 7 | 27739331 | 27844564 | 0 | - | - | - | - | - | - |
| TBC1D5 | OID20521 | Inflammat | TBC1D5 | Q92609 | | 3 | 17157162 | 18444817 | 0 | - | - | - | - | - | - |
| TBCB | OID20993 | Neurology | TBCB | Q99426 | | 19 | 36114289 | 36125947 | 0 | - | - | - | - | - | - |
| TBL1X | OID21220 | Oncology | TBL1X | O60907 | X | | 9463320 | 9741037 | 0 | - | - | - | - | - | - |
| TBR1 | OID30923 | Neurology | TBR1 | Q16650 | | 2 | 161416297 | 161425870 | 0 | - | - | - | - | - | - |
| TCL1B | OID20058 | Cardiome | TCL1B | O95988 | | 14 | 95686426 | 95692628 | 0 | - | - | - | - | - | - |
| TCOF1 | OID30316 | Cardiome | TCOF1 | Q13428 | | 5 | 150357629 | 150400308 | 0 | - | - | - | - | - | - |
| TCP11 | OID31267 | Oncology | TCP11 | Q8WWU5 | | 6 | 35118071 | 35148610 | 0 | - | - | - | - | - | - |
| TDO2 | OID30910 | Neurology | TDO2 | P48775 | | 4 | 155854738 | 155920406 | 0 | - | - | - | - | - | - |
| TERF1 | OID30480 | Inflammat | TERF1 | P54274 | | 8 | 73008856 | 73048123 | 0 | - | - | - | - | - | - |
| TET2 | OID31266 | Oncology | TET2 | Q6N021 | | 4 | 105145875 | 105279816 | 0 | - | - | - | - | - | - |
| TEX33 | OID31280 | Oncology | TEX33 | O43247 | | 22 | 36991120 | 37007851 | 0 | - | - | - | - | - | - |
| TFAP2A | OID30848 | Neurology | TFAP2A | P05549 | | 6 | 10393186 | 10419659 | 0 | - | - | - | - | - | - |
| TFF3 | OID20371 | Cardiome | TFF3 | Q07654 | | 21 | 42311667 | 42315409 | 0 | - | - | - | - | - | - |
| TGFBR1 | OID30502 | Inflammat | TGFBR1 | P36897 | | 9 | 99104038 | 99154192 | 0 | - | - | - | - | - | - |
| THAP12 | OID31238 | Oncology | THAP12 | O43422 | | 11 | 76349898 | 76381132 | 0 | - | - | - | - | - | - |
| THRAP3 | OID30053 | Cardiome | THRAP3 | Q9Y2W1 | | 1 | 36224432 | 36305357 | 0 | - | - | - | - | - | - |
| THSD1 | OID30476 | Inflammat | THSD1 | Q9NS62 | | 13 | 52377167 | 52416373 | 0 | - | - | - | - | - | - |
| TIA1 | OID20068 | Cardiome | TIA1 | P31483 | | 2 | 70209444 | 70248660 | 0 | - | - | - | - | - | - |
| TIMM8A | OID31143 | Neurology | TIMM8A | O60220 | X | | 101345661 | 101348742 | 0 | - | - | - | - | - | - |
| TIMP1 | OID20418 | Cardiome | TIMP1 | P01033 | X | | 47582408 | 47586789 | 0 | - | - | - | - | - | - |
| TJAP1 | OID21477 | Oncology | TJAP1 | Q5JTD0 | | 6 | 43477523 | 43506556 | 0 | - | - | - | - | - | - |
| TJP3 | OID30798 | Neurology | TJP3 | O95049 | | 19 | 3708362 | 3750813 | 0 | - | - | - | - | - | - |
| TLR2 | OID31290 | Oncology | TLR2 | O60603 | | 4 | 153684070 | 153706260 | 0 | - | - | - | - | - | - |
| TMCO5A | OID30940 | Neurology | TMCO5A | Q8N6Q1 | | 15 | 37921939 | 37967724 | 0 | - | - | - | - | - | - |
| TMED1 | OID31278 | Oncology | TMED1 | Q13445 | | 19 | 10832067 | 10836318 | 0 | - | - | - | - | - | - |
| TMED10 | OID30068 | Cardiome | TMED10 | P49755 | | 14 | 75131469 | 75176612 | 0 | - | - | - | - | - | - |
| TMED4 | OID30154 | Cardiome | TMED4 | Q7Z7H5 | | 7 | 44577894 | 44582287 | 0 | - | - | - | - | - | - |
| TMOD4 | OID31246 | Oncology | TMOD4 | Q9NZQ9 | | 1 | 151169986 | 151176284 | 0 | - | - | - | - | - | - |
| TMPRSS11B | OID30852 | Neurology | TMPRSS | Q86T26 | | 4 | 68226653 | 68245694 | 0 | - | - | - | - | - | - |
| TMSB10 | OID21118 | Neurology | TMSB10 | P63313 | | 2 | 84905656 | 84906671 | 0 | - | - | - | - | - | - |
| TNF | OID20074 | Cardiome | TNF | P01375 | | 6 | 31575565 | 31578336 | 0 | - | - | - | - | - | - |
| TNFAIP2 | OID31446 | Oncology | TNFAIP2 | Q03169 | | 14 | 103121476 | 103137439 | 0 | - | - | - | - | - | - |
| TNFRSF10C | OID20265 | Cardiome | TNFRSF1 | O14798 | | 8 | 23102921 | 23117445 | 0 | - | - | - | - | - | - |
| TNFRSF21 | OID21052 | Neurology | TNFRSF2 | O75509 | | 6 | 47231532 | 47309905 | 0 | - | - | - | - | - | - |
| TNIP1 | OID31120 | Neurology | TNIP1 | Q15025 | | 5 | 151029945 | 151093577 | 0 | - | - | - | - | - | - |
| TNNI3 | OID20050 | Cardiome | TNNI3 | P19429 | | 19 | 55151767 | 55157773 | 0 | - | - | - | - | - | - |
| TNPO1 | OID31330 | Oncology | TNPO1 | Q92973 | | 5 | 72816312 | 72916733 | 0 | - | - | - | - | - | - |
| TOMM20 | OID30353 | Cardiome | TOMM20 | Q15388 | | 1 | 235109341 | 235128837 | 0 | - | - | - | - | - | - |
| TOP1 | OID31449 | Oncology | TOP1 | P11387 | | 20 | 41028822 | 41124487 | 0 | - | - | - | - | - | - |
| TOP1MT | OID31233 | Oncology | TOP1MT | Q969P6 | | 8 | 143304384 | 143359979 | 0 | - | - | - | - | - | - |
| TP53BP1 | OID30470 | Inflammat | TP53BP1 | Q12888 | | 15 | 43403061 | 43510728 | 0 | - | - | - | - | - | - |
| TP53INP1 | OID20120 | Cardiome | TP53INP1 | Q96A56 | | 8 | 94925972 | 94949378 | 0 | - | - | - | - | - | - |
| TP73 | OID30879 | Neurology | TP73 | O15350 | | 1 | 3652516 | 3736201 | 0 | - | - | - | - | - | - |
| TPBGL | OID30928 | Neurology | TPBGL | P0DKB5 | | 11 | 75240774 | 75243704 | 0 | - | - | - | - | - | - |
| TPD52L2 | OID30609 | Inflammat | TPD52L2 | O43399 | | 20 | 63865228 | 63891545 | 0 | - | - | - | - | - | - |
| TPM3 | OID30058 | Cardiome | TPM3 | P06753 | | 1 | 154155308 | 154194648 | 0 | - | - | - | - | - | - |
| TPP1 | OID20750 | Inflammat | TPP1 | O14773 | | 11 | 6612768 | 6619448 | 0 | - | - | - | - | - | - |
| TPPP2 | OID31274 | Oncology | TPPP2 | P59282 | | 14 | 21024109 | 21036276 | 0 | - | - | - | - | - | - |
| TPR | OID30143 | Cardiome | TPR | P12270 | | 1 | 186311652 | 186375693 | 0 | - | - | - | - | - | - |
| TPRKB | OID31234 | Oncology | TPRKB | Q9Y3C4 | | 2 | 73729104 | 73737400 | 0 | - | - | - | - | - | - |
| TPSG1 | OID30424 | Inflammat | TPSG1 | Q9NRR2 | | 16 | 1221651 | 1225268 | 0 | - | - | - | - | - | - |
| TPT1 | OID20476 | Inflammat | TPT1 | P13693 | | 13 | 45333471 | 45341284 | 0 | - | - | - | - | - | - |
| TRAF3 | OID30479 | Inflammat | TRAF3 | Q13114 | | 14 | 102777449 | 102911500 | 0 | - | - | - | - | - | - |
| TRAF3IP2 | OID30054 | Cardiome | TRAF3IP | O43734 | | 6 | 111555381 | 111606906 | 0 | - | - | - | - | - | - |
| TREM2 | OID20731 | Inflammat | TREM2 | Q9NZC2 | | 6 | 41158506 | 41163186 | 0 | - | - | - | - | - | - |
| TREML1 | OID30682 | Inflammat | TREML1 | Q86YW5 | | 6 | 41149337 | 41154347 | 0 | - | - | - | - | - | - |
| TRIAP1 | OID21403 | Oncology | TRIAP1 | O43715 | | 12 | 120443964 | 120446384 | 0 | - | - | - | - | - | - |
| TRIM21 | OID20533 | Inflammat | TRIM21 | P19474 | | 11 | 4384897 | 4393702 | 0 | - | - | - | - | - | - |
| TRIM24 | OID31271 | Oncology | TRIM24 | O15164 | | 7 | 138460259 | 138589996 | 0 | - | - | - | - | - | - |
| TRIM26 | OID31207 | Oncology | TRIM26 | Q12899 | | 6 | 30184455 | 30213427 | 0 | - | - | - | - | - | - |
| TRPV3 | OID30128 | Cardiome | TRPV3 | Q8NET8 | | 17 | 3510502 | 3557812 | 0 | - | - | - | - | - | - |
| TSC1 | OID30478 | Inflammat | TSC1 | Q92574 | | 9 | 132891348 | 132946874 | 0 | - | - | - | - | - | - |
| TSHB | OID20129 | Cardiome | TSHB | P01222 | | 1 | 115029826 | 115034302 | 0 | - | - | - | - | - | - |
| TSLP | OID20056 | Cardiome | TSLP | Q969D9 | | 5 | 111070062 | 111078026 | 0 | - | - | - | - | - | - |
| TSNAX | OID30185 | Cardiome | TSNAX | Q99598 | | 1 | 231528541 | 231566524 | 0 | - | - | - | - | - | - |
| TSPAN1 | OID20078 | Cardiome | TSPAN1 | O60635 | | 1 | 46175073 | 46185962 | 0 | - | - | - | - | - | - |
| TSPAN7 | OID30810 | Neurology | TSPAN7 | P41732 | X | | 38561542 | 38688920 | 0 | - | - | - | - | - | - |
| TTN | OID30108 | Cardiome | TTN | Q8WZ42 | | 2 | 178525989 | 178830802 | 0 | - | - | - | - | - | - |
| TUBB3 | OID30799 | Neurology | TUBB3 | Q13509 | | 16 | 89921392 | 89938761 | 0 | - | - | - | - | - | - |
| TXK | OID31262 | Oncology | TXK | P42681 | | 4 | 48066393 | 48134250 | 0 | - | - | - | - | - | - |
| TXLNA | OID20952 | Neurology | TXLNA | P40222 | | 1 | 32179675 | 32198285 | 0 | - | - | - | - | - | - |
| TXN | OID30670 | Inflammat | TXN | P10599 | | 9 | 110243810 | 110256507 | 0 | - | - | - | - | - | - |
| TXNL1 | OID30872 | Neurology | TXNL1 | O43396 | | 18 | 56597209 | 56651600 | 0 | - | - | - | - | - | - |
| TXNRD1 | OID21135 | Neurology | TXNRD1 | Q16881 | | 12 | 104215779 | 104350307 | 0 | - | - | - | - | - | - |
| UBAC1 | OID21336 | Oncology | UBAC1 | Q9BSL1 | | 9 | 135932969 | 135961373 | 0 | - | - | - | - | - | - |
| UBE2B | OID31368 | Oncology | UBE2B | P63146 | | 5 | 134371184 | 134392108 | 0 | - | - | - | - | - | - |
| UBE2Z | OID30427 | Inflammat | UBE2Z | Q9H832 | | 17 | 48908407 | 48929056 | 0 | - | - | - | - | - | - |
| UBQLN3 | OID30096 | Cardiome | UBQLN3 | Q9H347 | | 11 | 5507300 | 5509958 | 0 | - | - | - | - | - | - |
| UBXN1 | OID30654 | Inflammat | UBXN1 | Q04323 | | 11 | 62676498 | 62679117 | 0 | - | - | - | - | - | - |
| UFD1 | OID31486 | Oncology | UFD1 | Q92890 | | 22 | 19449911 | 19479202 | 0 | - | - | - | - | - | - |
| UGDH | OID30081 | Cardiome | UGDH | O60701 | | 4 | 39498755 | 39528311 | 0 | - | - | - | - | - | - |
| UHRF2 | OID30171 | Cardiome | UHRF2 | Q96PU4 | | 9 | 6413151 | 6507054 | 0 | - | - | - | - | - | - |
| UNC79 | OID31181 | Oncology | UNC79 | Q9P2D8 | | 14 | 93333219 | 93707876 | 0 | - | - | - | - | - | - |
| UNG | OID31419 | Oncology | UNG | P13051 | | 12 | 109097597 | 109126725 | 0 | - | - | - | - | - | - |
| UPB1 | OID30352 | Cardiome | UPB1 | Q9UBR1 | | 22 | 24494107 | 24528390 | 0 | - | - | - | - | - | - |
| UPK3A | OID30794 | Neurology | UPK3A | O75631 | | 22 | 45284949 | 45295874 | 0 | - | - | - | - | - | - |
| UPK3BL1 | OID31281 | Oncology | UPK3BL1 | B0FP48 | | 7 | 102637025 | 102642791 | 0 | - | - | - | - | - | - |
| USO1 | OID21367 | Oncology | USO1 | O60763 | | 4 | 75724577 | 75814286 | 0 | - | - | - | - | - | - |
| USP25 | OID31428 | Oncology | USP25 | Q9UHP3 | | 21 | 15729982 | 15880064 | 0 | - | - | - | - | - | - |
| USP47 | OID30119 | Cardiome | USP47 | Q96K76 | | 11 | 11841423 | 11961887 | 0 | - | - | - | - | - | - |
| VASP | OID30692 | Inflammat | VASP | P50552 | | 19 | 45506579 | 45526989 | 0 | - | - | - | - | - | - |
| VCPKMT | OID31179 | Oncology | VCPKMT | Q9H867 | | 14 | 50108632 | 50116600 | 0 | - | - | - | - | - | - |

| | | | | | | | | | | | | | | | |
|---|---|---|---|---|---|---|---|---|---|---|---|---|---|---|---|
| VEGFC | OID21524 | Oncology | VEGFC | P49767 | | 4 | 176683538 | 176792922 | 0 | - | - | - | - | - | - |
| VEGFD | OID20662 | Inflammat | VEGFD | O43915 | X | | 15345596 | 15384413 | 0 | - | - | - | - | - | - |
| VIM | OID20053 | Cardiome | VIM | P08670 | | 10 | 17228241 | 17237593 | 0 | - | - | - | - | - | - |
| VIPR1 | OID30193 | Cardiome | VIPR1 | P32241 | | 3 | 42489299 | 42537573 | 0 | - | - | - | - | - | - |
| VPS28 | OID31273 | Oncology | VPS28 | Q9UK41 | | 8 | 144423601 | 144428563 | 0 | - | - | - | - | - | - |
| VPS37A | OID21355 | Oncology | VPS37A | Q8NEZ2 | | 8 | 17246931 | 17302427 | 0 | - | - | - | - | - | - |
| VPS4B | OID31386 | Oncology | VPS4B | O75351 | | 18 | 63389190 | 63422483 | 0 | - | - | - | - | - | - |
| VPS53 | OID21281 | Oncology | VPS53 | Q5VIR6 | | 17 | 508503 | 721717 | 0 | - | - | - | - | - | - |
| VSIG4 | OID21144 | Neurology | VSIG4 | Q9Y279 | X | | 66021738 | 66040125 | 0 | - | - | - | - | - | - |
| VSTM2B | OID31272 | Oncology | VSTM2B | A6NLU5 | | 19 | 29525994 | 29564551 | 0 | - | - | - | - | - | - |
| VTA1 | OID21016 | Neurology | VTA1 | Q9NP79 | | 6 | 142147162 | 142224685 | 0 | - | - | - | - | - | - |
| VWA5A | OID31198 | Oncology | VWA5A | O00534 | | 11 | 124115404 | 124147721 | 0 | - | - | - | - | - | - |
| WAS | OID20479 | Inflammat | WAS | P42768 | X | | 48676596 | 48691427 | 0 | - | - | - | - | - | - |
| WASL | OID30467 | Inflammat | WASL | O00401 | | 7 | 123681943 | 123749003 | 0 | - | - | - | - | - | - |
| WDR46 | OID31389 | Oncology | WDR46 | O15213 | | 6 | 33279108 | 33289247 | 0 | - | - | - | - | - | - |
| XG | OID20259 | Cardiome | XG | P55808 | X | | 2752040 | 2816500 | 0 | - | - | - | - | - | - |
| XIAP | OID30490 | Inflammat | XIAP | P98170 | X | | 123859712 | 123913972 | 0 | - | - | - | - | - | - |
| XPNPEP2 | OID21508 | Oncology | XPNPEP2 | O43895 | X | | 129738949 | 129769536 | 0 | - | - | - | - | - | - |
| YARS1 | OID31494 | Oncology | YARS1 | P54577 | | 1 | 32775237 | 32818031 | 0 | - | - | - | - | - | - |
| YJU2 | OID31404 | Oncology | YJU2 | Q9BW85 | | 19 | 4247080 | 4269088 | 0 | - | - | - | - | - | - |
| YTHDF3 | OID20478 | Inflammat | YTHDF3 | Q7Z739 | | 8 | 63168553 | 63212786 | 0 | - | - | - | - | - | - |
| YWHAQ | OID30605 | Inflammat | YWHAQ | P27348 | | 2 | 9583967 | 9630997 | 0 | - | - | - | - | - | - |
| YY1 | OID30446 | Inflammat | YY1 | P25490 | | 14 | 100238298 | 100282788 | 0 | - | - | - | - | - | - |
| ZBTB17 | OID20082 | Cardiome | ZBTB17 | Q13105 | | 1 | 15941869 | 15976132 | 0 | - | - | - | - | - | - |
| ZCCHC8 | OID30117 | Cardiome | ZCCHC8 | Q6NZY4 | | 12 | 122471599 | 122501073 | 0 | - | - | - | - | - | - |
| ZHX2 | OID31140 | Neurology | ZHX2 | Q9Y6X8 | | 8 | 122781655 | 122974510 | 0 | - | - | - | - | - | - |
| ZNF174 | OID30508 | Inflammat | ZNF174 | Q15697 | | 16 | 3401215 | 3409364 | 0 | - | - | - | - | - | - |
| ZNF75D | OID31303 | Oncology | ZNF75D | P51815 | X | | 135248920 | 135344109 | 0 | - | - | - | - | - | - |
| ZNF830 | OID31350 | Oncology | ZNF830 | Q96NB3 | | 17 | 34961540 | 34963777 | 0 | - | - | - | - | - | - |
| ZNRD2 | OID31450 | Oncology | ZNRD2 | O60232 | | 11 | 65570460 | 65573942 | 0 | - | - | - | - | - | - |
| ZNRF4 | OID31347 | Oncology | ZNRF4 | Q8WWF5 | | 19 | 5455417 | 5456856 | 0 | - | - | - | - | - | - |
| ZP4 | OID31185 | Oncology | ZP4 | Q12836 | | 1 | 237877864 | 237890922 | 0 | - | - | - | - | - | - |
| ZPR1 | OID30801 | Neurology | ZPR1 | O75312 | | 11 | 116773799 | 116788039 | 0 | - | - | - | - | - | - |

**Table S8 19 Hazard ratios for incident ADRD associated with the 19 proteins selected by a LASSO Cox regression model**

| | | 95% CI | | |
|---|---|---|---|---|
| Protein | HR | Lower | Upper | P-Value |
| VGF | 0.33 | 0.18 | 0.58 | 1.16E-04 |
| BCAN | 0.94 | 0.57 | 1.54 | 0.793 |
| NPTXR | 0.45 | 0.26 | 0.79 | 0.005 |
| SERPINI1 | 0.43 | 0.25 | 0.75 | 0.003 |
| KIT | 0.59 | 0.39 | 0.90 | 0.013 |
| HPGDS | 0.62 | 0.43 | 0.90 | 0.012 |
| EGFR | 1.02 | 0.43 | 2.44 | 0.962 |
| APOE | 0.54 | 0.40 | 0.74 | 1.06E-04 |
| C1QTNF6 | 0.50 | 0.28 | 0.87 | 0.015 |
| GET3 | 0.62 | 0.45 | 0.86 | 0.004 |
| NPPB | 1.11 | 0.96 | 1.28 | 0.158 |
| NTPROBNP | 0.99 | 0.83 | 1.18 | 0.883 |
| HAVCR1 | 1.27 | 1.02 | 1.57 | 0.030 |
| PSG1 | 1.20 | 1.05 | 1.36 | 0.006 |
| ADGRG1 | 1.06 | 0.92 | 1.23 | 0.422 |
| ELN | 1.78 | 1.24 | 2.54 | 0.002 |
| GDF15 | 1.22 | 0.92 | 1.62 | 0.175 |
| GFAP | 2.19 | 1.77 | 2.72 | 9.28E-13 |
| NEFL | 1.93 | 1.49 | 2.52 | 9.81E-07 |

**Table S9 Parameter estimates associated the 19 proteins selected by a LASSO Cox regression model in a Gomperz regression model**

| | | 95% CI | | |
|---|---|---|---|---|
| | Estimate | Lower | Upeer | SE |
| Shape | 0.12 | 0.08 | 0.16 | 0.02 |
| Rate | 0.00 | 0.00 | 0.00 | 0.00 |
| ADGRG1 | 0.06 | -0.09 | 0.21 | 0.07 |
| APOE | -0.60 | -0.91 | -0.30 | 0.16 |
| BCAN | -0.08 | -0.58 | 0.42 | 0.26 |
| C1QTNF6 | -0.70 | -1.26 | -0.14 | 0.29 |
| EGFR | 0.03 | -0.84 | 0.90 | 0.44 |
| ELN | 0.57 | 0.21 | 0.93 | 0.18 |
| GDF15 | 0.20 | -0.09 | 0.48 | 0.15 |
| GET3 | -0.47 | -0.79 | -0.14 | 0.17 |
| GFAP | 0.79 | 0.57 | 1.00 | 0.11 |
| HAVCR1 | 0.24 | 0.02 | 0.45 | 0.11 |
| HPGDS | -0.47 | -0.85 | -0.10 | 0.19 |
| KIT | -0.54 | -0.96 | -0.12 | 0.21 |
| NEFL | 0.65 | 0.39 | 0.92 | 0.13 |
| NPPB | 0.10 | -0.04 | 0.25 | 0.07 |
| NPTXR | -0.81 | -1.37 | -0.25 | 0.29 |
| NTPROBNP | -0.01 | -0.19 | 0.16 | 0.09 |
| PSG1 | 0.19 | 0.06 | 0.31 | 0.07 |
| SERPINI1 | -0.84 | -1.39 | -0.29 | 0.28 |
| VGF | -1.11 | -1.67 | -0.54 | 0.29 |

**Table S10 Spearman correlations of $PrRS_{MDD-ADRD}$ with T1 structural and T2-weighted brain MRI image-derived phenotypes (IDPs)**

*If two fields from the left and right hemispheres are listed, their average was used to correlate with PrRS.

| IDP Description | Fields* | N | PrRS Correlation | PrRS Correlation Unadjusted P | PrRS Correlation FDR-Adjusted P |
|---|---|---|---|---|---|
| Total volume of peri-ventricular white matter hyperintensities | 24485 | 568 | 0.32 | 4.4E-15 | 3.8E-13 |
| Total volume of white matter hyperintensities (from T1 and T2_FLAIR images) | 25781 | 568 | 0.31 | 4.6E-14 | 2.0E-12 |
| Volume of accumbens (from T1 brain image) | 25023/25024 | 591 | -0.29 | 3.1E-13 | 8.9E-12 |
| Volume of grey matter in Ventral Striatum | 25890/25891 | 591 | -0.28 | 4.5E-12 | 9.6E-11 |
| Volume of grey matter in Amygdala | 25888/25889 | 591 | -0.27 | 1.2E-11 | 2.1E-10 |
| Volume of grey matter in Central Opercular Cortex | 25864/25865 | 591 | -0.26 | 1.1E-10 | 1.5E-09 |
| Volume of grey matter in Heschl's Gyrus (includes H1 and H2) | 25870/25871 | 591 | -0.26 | 2.2E-10 | 2.7E-09 |
| Volume of TotalGray in the whole brain generated by subcortical volumetric segmentation | 26518 | 586 | -0.25 | 4.3E-10 | 4.6E-09 |
| Volume of grey matter in Planum Polare | 25868/25869 | 591 | -0.24 | 3.2E-09 | 3.0E-08 |
| Volume of grey matter in Precentral Gyrus | 25794/25795 | 591 | -0.24 | 3.7E-09 | 3.1E-08 |
| Volume of grey matter in VI Cerebellum | 25897/25899 | 591 | -0.23 | 1.3E-08 | 1.0E-07 |
| Volume of grey matter in Frontal Orbital Cortex | 25846/25847 | 591 | -0.23 | 2.2E-08 | 1.5E-07 |
| Volume of thalamus (from T1 brain image) | 25011/25012 | 591 | -0.22 | 5.4E-08 | 3.5E-07 |
| Volume of putamen (from T1 brain image) | 25015/25016 | 591 | -0.22 | 9.2E-08 | 5.6E-07 |
| Total volume of deep white matter hyperintensities | 24486 | 568 | 0.22 | 2.2E-07 | 1.1E-06 |
| Volume of grey matter in Paracingulate Gyrus | 25836/25837 | 591 | -0.21 | 1.7E-07 | 9.8E-07 |
| Volume of grey matter in VIIIa Cerebellum | 25909/25911 | 591 | -0.21 | 3.4E-07 | 1.7E-06 |
| Volume of grey matter in Frontal Medial Cortex | 25830/25831 | 591 | -0.21 | 4.2E-07 | 2.0E-06 |
| Volume of grey matter in Lingual Gyrus | 25852/25853 | 591 | -0.20 | 5.4E-07 | 2.4E-06 |
| Volume of grey matter in Postcentral Gyrus | 25814/25815 | 591 | -0.20 | 5.7E-07 | 2.4E-06 |
| Volume of grey matter in Superior Temporal Gyrus, posterior division | 25800/25801 | 591 | -0.20 | 6.8E-07 | 2.8E-06 |
| Volume of grey matter in VIIIb Cerebellum | 25912/25914 | 591 | -0.20 | 7.6E-07 | 2.9E-06 |
| Volume of grey matter in Precuneous Cortex | 25842/25843 | 591 | -0.20 | 1.5E-06 | 5.4E-06 |
| Volume of grey matter in Lateral Occipital Cortex, superior division | 25824/25825 | 591 | -0.19 | 4.2E-06 | 1.5E-05 |
| Volume of grey matter in Temporal Pole | 25796/25797 | 591 | -0.19 | 4.3E-06 | 1.5E-05 |
| Volume of grey matter in Hippocampus | 25886/25887 | 591 | -0.19 | 5.2E-06 | 1.7E-05 |
| Volume of grey matter in Temporal Fusiform Cortex, posterior division | 25856/25857 | 591 | -0.19 | 5.3E-06 | 1.7E-05 |
| Volume of grey matter in X Cerebellum | 25918/25920 | 591 | -0.18 | 6.7E-06 | 2.0E-05 |
| Volume of grey matter in Superior Frontal Gyrus | 25786/25787 | 591 | -0.18 | 9.1E-06 | 2.7E-05 |
| Volume of grey matter in Frontal Pole | 25782/25783 | 591 | -0.18 | 1.1E-05 | 3.1E-05 |
| Volume of grey matter in Superior Temporal Gyrus, anterior division | 25798/25799 | 591 | -0.18 | 1.3E-05 | 3.4E-05 |
| Volume of grey matter in Middle Temporal Gyrus, posterior division | 25804/25805 | 591 | -0.18 | 1.3E-05 | 3.4E-05 |
| Volume of grey matter in Lateral Occipital Cortex, inferior division | 25826/25827 | 591 | -0.18 | 1.8E-05 | 4.7E-05 |
| Volume of grey matter in Supramarginal Gyrus, posterior division | 25820/25821 | 591 | -0.17 | 2.2E-05 | 5.5E-05 |
| Volume of grey matter in Crus I Cerebellum | 25900/25902 | 591 | -0.17 | 3.1E-05 | 7.5E-05 |
| Volume of grey matter in Vermis VIIIa Cerebellum | 25910 | 591 | -0.17 | 3.2E-05 | 7.6E-05 |
| Volume of grey matter in Temporal Fusiform Cortex, anterior division | 25854/25855 | 591 | -0.17 | 3.5E-05 | 8.0E-05 |
| Volume of grey matter in VIIb Cerebellum | 25906/25908 | 591 | -0.17 | 4.1E-05 | 9.1E-05 |
| Volume of grey matter in Supracalcarine Cortex | 25874/25875 | 591 | -0.17 | 4.9E-05 | 1.1E-04 |
| Volume of grey matter in V Cerebellum | 25895/25896 | 591 | -0.16 | 1.1E-04 | 2.2E-04 |
| Volume of caudate (from T1 brain image) | 25013/25014 | 591 | -0.16 | 1.3E-04 | 2.7E-04 |
| Volume of grey matter in Inferior Temporal Gyrus, posterior division | 25810/25811 | 591 | -0.15 | 1.7E-04 | 3.5E-04 |
| Volume of grey matter in Occipital Fusiform Gyrus | 25860/25861 | 591 | -0.15 | 2.0E-04 | 3.9E-04 |
| Volume of grey matter in Brain-Stem | 25892 | 591 | -0.15 | 2.7E-04 | 0.001 |
| Volume of grey matter in Parahippocampal Gyrus, anterior division | 25848/25849 | 591 | -0.15 | 2.7E-04 | 0.001 |
| Volume of grey matter in Vermis VI Cerebellum | 25898 | 591 | -0.15 | 2.7E-04 | 0.001 |
| Volume of grey matter in Intracalcarine Cortex | 25828/25829 | 591 | -0.15 | 3.9E-04 | 0.001 |
| Volume of hippocampus (from T1 brain image) | 25019/25020 | 591 | -0.14 | 4.1E-04 | 0.001 |
| Volume of grey matter in Middle Frontal Gyrus | 25788/25789 | 591 | -0.14 | 4.9E-04 | 0.001 |
| Volume of grey matter in Occipital Pole | 25876/25877 | 591 | -0.14 | 0.001 | 0.001 |
| Volume of grey matter in Planum Temporale | 25872/25873 | 591 | -0.14 | 0.001 | 0.001 |
| Volume of grey matter in Insular Cortex | 25784/25785 | 591 | -0.14 | 0.001 | 0.001 |
| Volume of pallidum (from T1 brain image) | 25017/25018 | 591 | -0.14 | 0.001 | 0.001 |
| Volume of grey matter in Vermis VIIb Cerebellum | 25907 | 591 | -0.13 | 0.001 | 0.002 |
| Volume of grey matter in Crus II Cerebellum | 25903/25905 | 591 | -0.13 | 0.001 | 0.002 |
| Volume of grey matter in Frontal Operculum Cortex | 25862/25863 | 591 | -0.13 | 0.001 | 0.002 |
| Volume of grey matter in Putamen | 25882/25883 | 591 | -0.13 | 0.002 | 0.002 |
| Volume of grey matter in Superior Parietal Lobule | 25816/25817 | 591 | -0.13 | 0.002 | 0.002 |
| Volume of grey matter in Vermis VIIIb Cerebellum | 25913 | 591 | -0.13 | 0.002 | 0.003 |
| Volume of grey matter in Inferior Temporal Gyrus, anterior division | 25808/25809 | 591 | -0.12 | 0.002 | 0.003 |
| Volume of grey matter in I-IV Cerebellum | 25893/25894 | 591 | -0.12 | 0.002 | 0.003 |
| Volume of grey matter in Angular Gyrus | 25822/25823 | 591 | -0.12 | 0.004 | 0.005 |
| Volume of grey matter in Inferior Frontal Gyrus, pars opercularis | 25792/25793 | 591 | -0.12 | 0.004 | 0.006 |
| Volume of grey matter in Middle Temporal Gyrus, anterior division | 25802/25803 | 591 | -0.12 | 0.005 | 0.007 |
| Volume of grey matter in Parietal Operculum Cortex | 25866/25867 | 591 | -0.11 | 0.006 | 0.007 |
| Volume of grey matter in Subcallosal Cortex | 25834/25835 | 591 | -0.11 | 0.007 | 0.009 |
| Volume of grey matter in Cuneal Cortex | 25844/25845 | 591 | -0.11 | 0.010 | 0.013 |
| Volume of grey matter in Juxtapositional Lobule Cortex (formerly Supplementary Motor Co | 25832/25833 | 591 | -0.10 | 0.017 | 0.021 |
| Volume of grey matter in Temporal Occipital Fusiform Cortex | 25858/25859 | 591 | -0.09 | 0.024 | 0.029 |
| Volume of grey matter in IX Cerebellum | 25915/25917 | 591 | -0.08 | 0.041 | 0.050 |
| Volume of grey matter in Supramarginal Gyrus, anterior division | 25818/25819 | 591 | -0.08 | 0.043 | 0.052 |
| Volume of grey matter in Vermis X Cerebellum | 25919 | 591 | -0.08 | 0.046 | 0.054 |
| Volume of grey matter in Parahippocampal Gyrus, posterior division | 25850/25851 | 591 | -0.08 | 0.051 | 0.059 |
| Volume of grey matter in Vermis IX Cerebellum | 25916 | 591 | -0.07 | 0.094 | 0.108 |
| Volume of grey matter in Inferior Frontal Gyrus, pars triangularis | 25790/25791 | 591 | -0.07 | 0.103 | 0.117 |
| Volume of grey matter in Pallidum | 25884/25885 | 591 | 0.06 | 0.124 | 0.138 |
| Volume of grey matter in Inferior Temporal Gyrus, temporooccipital part | 25812/25813 | 591 | -0.06 | 0.159 | 0.175 |
| Volume of grey matter in Middle Temporal Gyrus, temporooccipital part | 25806/25807 | 591 | -0.06 | 0.166 | 0.181 |
| Volume of grey matter in Caudate | 25880/25881 | 591 | 0.05 | 0.220 | 0.237 |
| Volume of grey matter in Cingulate Gyrus, anterior division | 25838/25839 | 591 | -0.02 | 0.560 | 0.595 |
| Volume of amygdala (from T1 brain image) | 25021/25022 | 591 | -0.02 | 0.589 | 0.612 |
| Volume of grey matter in Thalamus | 25878/25879 | 591 | -0.02 | 0.590 | 0.612 |
| Volume of grey matter in Vermis Crus II Cerebellum | 25904 | 591 | -0.02 | 0.622 | 0.637 |
| Volume of grey matter in Vermis Crus I Cerebellum | 25901 | 591 | 0.01 | 0.786 | 0.795 |
| Volume of grey matter in Cingulate Gyrus, posterior division | 25840/25841 | 591 | -0.01 | 0.830 | 0.830 |
